# Genomic insights into population stratification, biological adaptation, and archaic introgression at the crossroads of the Himalayas and lowland East Asia

**DOI:** 10.1101/2025.08.25.25334410

**Authors:** Mengge Wang, Shuhan Duan, Qiuxia Sun, Yuntao Sun, Lintao Luo, Yunhui Liu, Renkuan Tang, Libing Yun, Chao Liu, Guanglin He

## Abstract

The Tibetan-Yi Corridor (TYC) has been a crucial agro-pastoral region in the eastern Himalayas, linking the Qinghai-Tibet Plateau with the lowlands of East Asia and facilitating human migration over millennia. Despite its significance, genomic research on TYC populations is limited, restricting understanding of their origins and health. We present genomic data from 1,031 individuals from Austroasiatic and Sino-Tibetan groups, including 147 whole-genome sequences from 13 underrepresented Tibeto-Burman and Austroasiatic communities. Our analysis uncovered approximately 3.3 million new genetic variants and identified four distinct genetic backgrounds among TYC populations. Demographic reconstructions highlight strong genetic connections among Tibeto-Burman groups, Central Plain Sinitic populations, and Yellow River Yangshao farmers, supporting a shared origin for Sino-Tibetan speakers. We observed signatures of high-altitude adaptation common with Tibetans and TYC-specific variants associated with pigmentation and hypoxia response. Differentiation involved mechanisms like HLA-DQB1-rs1049083, related to immune function. Certain rare pathogenic variants, such as CYP21A2-rs6467 and PRX-rs3814290, were especially frequent. Variants influencing warfarin sensitivity showed considerable variation. Archaic human introgression contributed to complexity, affecting cardiovascular and immune-related genes, indicative of adaptation through ancient human interactions. This research advances understanding of human evolution in this region and highlights the importance of broader genetic studies for insights into diversity and tailored medical approaches.

## Introduction

The Tibetan-Yi Corridor (TYC), situated in the eastern Himalayas, links the Qinghai-Tibet Plateau (QTP) with eastern Asia’s lowlands, including most parts of Sichuan, Gansu, Qinghai, and Yunnan provinces. It boasts a rich ecological landscape filled with mountains and rivers. As a crossroads connecting the Hexi Corridor, mainland Southeast Asia (SEA), the QTP, and the Central Plain, this region exhibits significant ethnolinguistic, biological, and genetic diversity. Historically, the linguistic variety in TYC has promoted genetic exchange among different ethnolinguistic groups through population mixing or shared origins of languages such as Sino-Tibetan, Hmong-Mien, Tai-Kadai, and Austroasiatic. The rugged terrain, however, also acts as a barrier, isolating groups like the Amdo Tibetan (ADT), Lahu, Wa, and Blang, who have maintained unique cultural and genetic traits (Bergstrom et al., 2020). Archaeological discoveries at Yuanmou in Yunnan and Piluo in Sichuan reveal Paleolithic human activity within TYC, alongside evidence of extensive Neolithic cultural and demographic diffusion from northern Gansu and Qinghai. Ancient DNA from Yunnan reveals genetic links between early Neolithic populations and older QTP groups, as well as mid-Neolithic communities tied to Austroasiatic origins (Wang et al., 2025d). Additionally, the introduction of agriculture into eastern TYC, evidenced by the Gaoshan and Haimenkou relics of the Baodun culture, occurred around 4,000 years ago (Tao et al., 2023). Interdisciplinary studies have also shed light on the relationships among sublineages of the Sino-Tibetan language family in TYC and surrounding areas. Linguistic research offers competing theories about the origins of Sino-Tibetan, suggesting dates from 4,000 to 6,000 years ago in northern China or around 9,000 years ago in southwest China or northeast India (LaPolla, 2019). Some scholars, like Zhang et al. and Sagart et al., propose that Sino-Tibetan speakers originated in North China and spread through Neolithic millet farmers along the Yellow River (Sagart et al., 2019; Zhang et al., 2019). Archaeological evidence also indicates these millet farmers moved southwestward, interacting with indigenous hunter-gatherers in northwest Sichuan (Liu et al., 2022).

Large-scale genomic resources from Europe (Consortium, 2025), Africa (Choudhury et al., 2020), Oceania (Silcocks et al., 2023), and the Americas (All of Us Research Program Genomics, 2024; Verma et al., 2024) have significantly advanced our understanding of regional evolutionary histories and population health. In contrast, genomic studies of Chinese populations are limited, especially for those from TYC, since most autosomal, Y/MtDNA resources focus on Han Chinese and underrepresent minority ethnic groups (Luo et al., 2025; Wang et al., 2025b; Wang et al., 2025e). Located between the highlands of Tibetans, the lowlands of Han, and surrounding indigenous groups, TYC is predominantly inhabited by Tibeto-Burman speakers but also contains linguistically distinct populations in its eastern and southern parts. The region’ s cultural, ethnic, linguistic, and biological diversity reflects its history as both a Paleolithic migration route and a pathway for the rapid Neolithic expansion of farmers. Although Tibetans on the QTP have been extensively studied (Zhang et al., 2017; Zheng et al., 2023), other Tibeto-Burman-speaking communities within TYC, despite their considerable genetic diversity, have received considerably less attention. Genetic analyses of Tibetan and Deng populations reveal shared ancestry with Han Chinese and close links to ancient northern Chinese millet farmers, with divergence times estimated at 15,000-7,200 years ago (Lu et al., 2016; Ge et al., 2023). Understanding the demographic history of these underrepresented groups is crucial, as migration, gene flow, introgression, and divergence have all influenced their genomes (Kamm et al., 2020). The eastern and southeastern edges of TYC have undergone repeated and complex cultural and genetic interactions. Archaeological and historical records show that ancestors of present-day southern Chinese indigenous peoples include the ancient Baipu and Baiyue tribes, along with the ancestors of Austroasiatic speakers (Quintana-Murci, 2002; Yang et al., 2022b; Wang et al., 2025d). Competing theories suggest the origin of the superlanguage family either in South China or in North India (van Driem, 2001; Sagart, 2021). Evidence linking early rice agriculture to Austroasiatic expansion supports the South China hypothesis (Fuller, 2012). Ancient DNA studies further reveal that Neolithic farmers from Man Bac in mainland Southeast Asia carried ancestry from southern Chinese farmers as well as from deeply divergent indigenous hunter-gatherers (Lipson et al., 2018; McColl et al., 2018). Multiple post-Neolithic migration waves involving Austronesian, Hmong-Mien, and Tai-Kadai speakers from South China later reshaped the genetic landscape of mainland Southeast Asia and eastern TYC (Lipson et al., 2018; McColl et al., 2018; He et al., 2024; Sun et al., 2024a). Y-chromosome phylogeny supports the dispersal of Austroasiatic haplogroup O2a-M95 over the past approximately 5,000 years, likely linked to early rice farmers (Singh et al., 2021). Despite these advances, the detailed genetic structure, demographic history, adaptive traits, and health-related effects of admixture, including archaic introgression from Denisovans and Neanderthals, remain poorly understood for Tibeto-Burman- and Austroasiatic-speaking populations in TYC and their Himalayan neighbors.

Previous genetic studies have examined the population history of the core TYC region. Zhang et al. analyzed low-coverage whole-genome sequencing (WGS) data and identified geographic patterns of genetic affinity among TYC populations (Zhang et al., 2022). Yang et al. showed that population stratification in Yunnan, at the southern end of TYC, was shaped by millet-farming Tibeto-Burman groups and rice-farming populations, including Austroasiatic, Tai-Kadai, Hmong-Mien, and Austronesian speakers (Yang et al., 2022b). Genome-wide SNP analyses also revealed gene flow and biological adaptations that distinguish Yi populations from different geographic regions (Sun et al., 2024b). Ancient DNA has provided additional spatial and temporal insights into population dynamics. Tao et al. reported the first ancient genome-wide data from the eastern margins of TYC, showing that both demic and cultural diffusion from Yangshao/Majiayao-related ancestors contributed to the Neolithic gene pool, supporting a northern China origin (Wang et al., 2021a; Tao et al., 2023). Other studies have examined forensic markers, including STR, SNP, and InDel variation from autosomal and Y-chromosome data, further emphasizing the region’s genetic complexity (Wang et al., 2023; Wang et al., 2025a). Overall, molecular evidence reveals a complex evolutionary history for TYC populations. However, many studies remain limited by small sample sizes, low SNP density, narrow population coverage, or limited geographic scope, hindering a comprehensive understanding of migration, admixture, and health-related patterns. Dense sampling across ethnolinguistically diverse groups is essential to clarify the genetic origins and health implications of TYC populations. Exposed to diverse environments, pathogens, and subsistence strategies, these populations offer a unique opportunity to study how evolutionary pressures have shaped genetic architecture and disease susceptibility. Research on Tibetan, Sherpa, and Deng populations has identified strong signals of high-altitude adaptation, including *EPAS1*, *EGLN1*, *HAL-DQB1*, and *GNPAT* (Simonson et al., 2010; Yi et al., 2010; Zhang et al., 2017; Yang et al., 2022a; Ge et al., 2023; Zheng et al., 2023). Whether these genes play similar roles in other Tibeto-Burman-speaking groups in TYC, and whether additional mechanisms drive rapid adaptation to local environments, remains unresolved. Archaic introgression, most notably EPAS1, has been shown to contribute to high-altitude adaptation in Tibetans from Xizang (Huerta-Sanchez et al., 2014; Zhang et al., 2021b; Zheng et al., 2023). Physical anthropology studies suggest that the Ziyang people exhibit Neanderthal facial traits, implying possible additional gene flow between present-day EAs and archaic humans in TYC (Wu and Yan, 2020). Likewise, the Lahu population carries deeply diverged non-African F lineages, indicating long-term interaction and interbreeding with archaic groups (Bergstrom et al., 2020). Despite these findings, the extent of archaic introgression in TYC populations, the demographic history behind these events, and the biological significance of the introgressed segments remain poorly understood.

Here, we performed whole-genome sequencing of seven Tibeto-Burman-speaking populations from the core TYC regions in Sichuan, including previously unreported Qiang, Baima Tibetan (BMT), and ADT groups, as well as four Tibeto-Burman-speaking populations from Yunnan, Bai, Pumi, Hani, and Lahu. We also sampled Wa and Blang, who speak Austroasiatic languages, from Yunnan (**Fig. 1a,b; Table S1**). In addition, we analyzed genome-wide genotyping data from 884 Sinitic individuals from the Central Plain (CPH), representing a core ancestral region of Sino-Tibetan speakers, together with published data from the 10K Chinese People Genomic Diversity Project (10K_CPGDP) (He et al., 2025) and publicly available WGS resources from the Human Genome Diversity Project (HGDP) (Bergstrom et al., 2020) and the 1000 Genomes Project (1KGP) (Genomes Project et al., 2015) (**Fig. 1a**). This study refines the fine-scale genetic structure of TYC populations, identifies local adaptations, redefines patterns of archaic introgression and their medical relevance, offers insights into population health, and pinpoints archaic-introgressed variants.

**Fig. 1.**
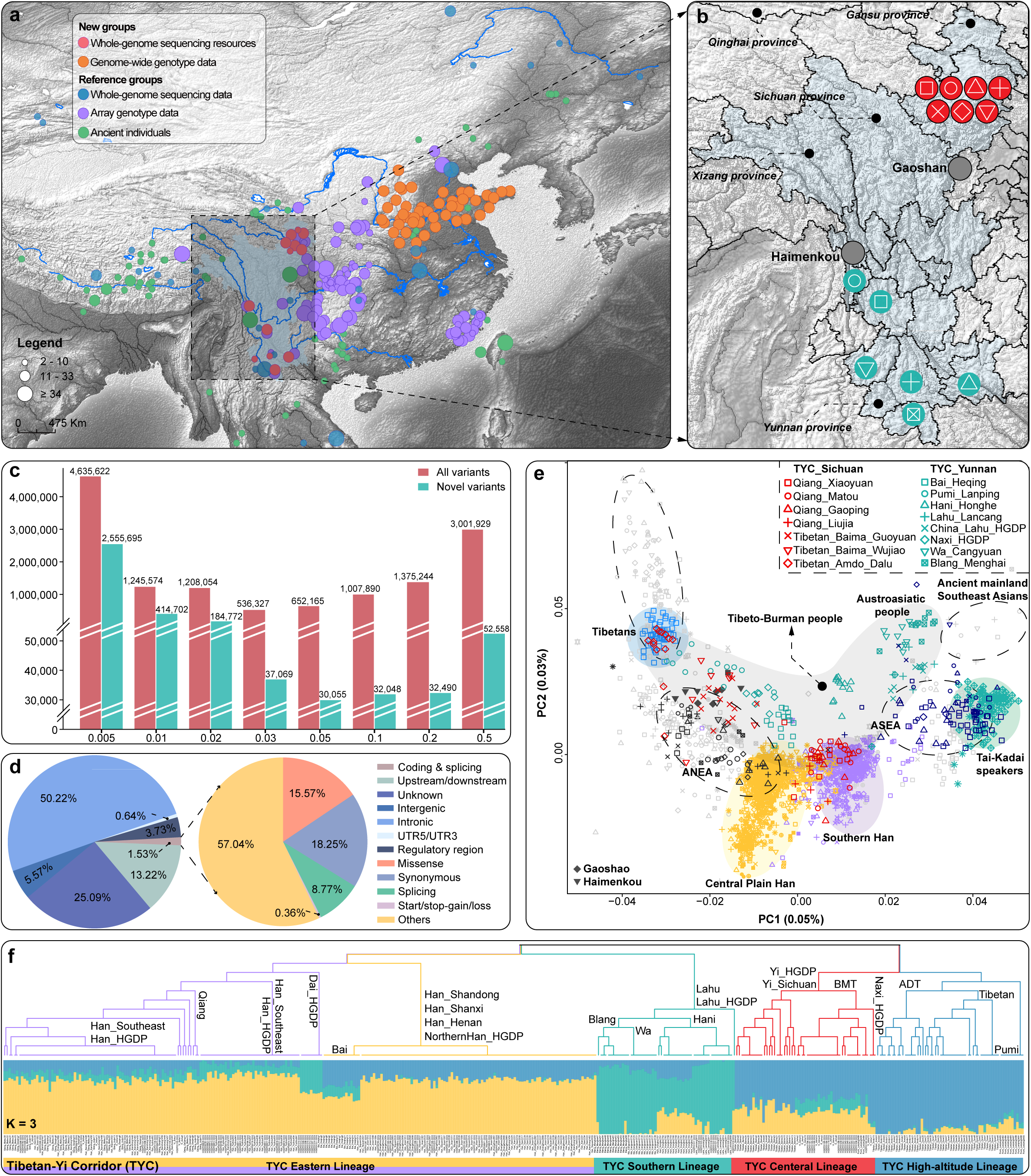
| Sample information, variant discovery, and genetic profile of the TYC population. (**a**) Geographical distribution of newly sampled Tibeto-Burman, Austroasiatic, and Sinitic populations, along with currently available genomic resources in East Asia (EA). (**b**) Geographic location of the Tibetan-Yi Corridor (TYC) populations involved in the whole-genome sequencing study. The symbols representing each population on the map correspond to the legend of the principal component analysis (PCA) shown in **Fig. 1e**, with more detailed individual information provided in **Table S1**. (**c**) The number of known (red) and novel (green) variants detected in the TYC populations, categorized based on dbSNP version 146, with the distribution classified by minor allele frequency. (d) Functional annotation of 13,662,805 biallelic SNPs, including 3.3 million previously unreported SNPs. (**e**) A two-dimensional principal component (PC) plot projects modern and ancient EA populations, as well as Southeast Asians (SEAs), within the EA context, highlighting clustering patterns among geographically and linguistically diverse EA populations. The modern reference EA populations, ancient Qinghai-Tibet Plateau individuals, and ancient mainland SEAs are colored gray. The detailed sample legends are shown in **Fig. S2e**. (**f**) A dendrogram constructed via haplotype-based fineSTRUCTURE and model-based ADMIXTURE results reveals clustering patterns among 23 EA populations on the basis of the Illumina_2240K dataset. The ADMIXTURE results are displayed below, with each column representing an individual. The blue component is maximized in Amdo Tibetan (ADT), the green component is enriched in Lahu, and the yellow component is prominent in Central Plain Han (CPH). ANEA/ASEA, Ancient Northern/Southern East Asians.

## Results

### Whole-genome sequencing, genotyping, and variant discovery

We collected 1,031 blood and saliva samples from indigenous Sinitic, Tibeto-Burman, and Austroasiatic individuals representing diverse ethnic groups from TYC and neighboring regions (**Fig. 1a, b**). Informed consent was obtained from all participants. Whole-genome sequencing was performed on 147 Tibeto-Burman and Austroasiatic individuals from Sichuan and Yunnan Provinces, achieving mean depths of 12.5× and 91.7% coverage, respectively (**Table S1**). After read alignment, variant detection, and strict variant filtering, we identified a set of 13,662,805 biallelic variants with a Ts/Tv ratio of 2.07, along with 1,689,560 small InDels ranging from 1 bp to 50 bp. Of the 13.6 million biallelic SNVs identified, 43% were classified as rare, with a minor allele frequency (MAF) < 0.01. Another 17.5% (2,396,546 SNVs) had an MAF between 0.01 and 0.05. Notably, 24.4% (about 3.3 million) of the SNVs were novel, as they were not listed in dbSNP (Build 146). Most of these novel SNVs were rare, with 89% (2,970,397) having an MAF < 0.01, 7.5% (251,896) between 0.01 and 0.05, and only 3.5% (117,096) being common (**Fig. 1c and Table S2**). To evaluate the biological significance of the novel variants, we annotated them using VEP tools, focusing on medical conditions. Intronic and intergenic regions made up 50.22% and 5.57% of the variants, respectively. Only 1.53% of the variants were located in coding and splicing regions. Within these regions, missense variants accounted for 15.57%, while splicing variants represented 8.77% (**Fig. 1d and Table S3**).

### Insight into the fine-scale genetic profiles of diverse TYC groups

To clarify the genetic relationships between TYC populations and global groups, we analyzed whole-genome data from ethnolinguistically diverse TYC populations, nearby Han Chinese groups, and 185 current worldwide populations. Genetic differences, measured by the fixation index (*F*_ST_), were compared between the TYC populations and reference groups (**Table S4**). The TYC populations, located in southwestern China alongside highland Tibetans and lowland Sinitic speakers, showed the closest genetic affinity to EAs (*F*ST < 0.06), followed by SEAs (F_ST_ < 0.12). Among EA groups (**Fig. S1**), ADT, BMT, Bai, and Pumi had the closest genetic ties to highland Tibetans and Tibeto-Burman populations near the QTP, such as Yi and Naxi (*F*_ST_ < 0.02). In contrast, Qiang groups showed particularly close genetic proximity to southern Han populations (*F*_ST_ = 0.00009). Meanwhile, Lahu, Hani, Wa, and Blang populations demonstrated stronger genetic connections with southern Tai-Kadai groups, like the CDX and Dai in Yunnan (*F*_ST_ < 0.03). These results were supported by principal component analysis (PCA) of the WGS dataset (**Fig. S2a, b**).

To further explore the genetic relationships between the sampled individuals and other Asian populations, we performed additional PCA analysis. Projecting ancient samples onto the first two principal components derived from modern EAs and SEAs showed that PC1 arranged Tibetans, TYC populations, CPH, southern Han Chinese, and Tai-Kadai/Austroasiatic groups along a north-south gradient. PC2 separated the Tibet-Burman and Sinitic groups along the highland-lowland axis, with TYC populations spread across PC1, highlighting their geographic diversity. Notably, these populations appeared structured into four distinct north-south genetic clines among EA groups. The leftmost cline, consisting of highland Tibetan groups, linked QTP populations with ADT and Pumi from TYC (**Fig. S2c**). The middle cline included BMT, Naxi, and ancient individuals from Neolithic Baodun sites, while the lowermost cline comprised Qiang and southern Han groups. The rightmost cline featured Lahu, Hani, Wa, Blang, mainland SEAs, and Ancient Southern East Asians (ASEA) (**Fig. 1e, S2d, e**). The outgroup-*f_3_* statistics further reinforced these genetic patterns within the TYC populations, as well as the haplotype-based IBD analysis (**Fig. S3**). Haplotype-based fineSTRUCTURE analysis, using the Illumina 2240K dataset, identified four distinct genetic clusters within the TYC groups (**Fig. 1f**). The first cluster, the TYC high-altitude lineage (THL, blue), included populations from high-altitude regions, such as the core and peripheral areas of the QTP. The second cluster, the TYC Central Lineage (TCL, red), represented central populations. The third cluster, the TYC Southern Lineage (TSL, green), consisted of Tibeto-Burman speakers like Hani and Lahu, along with Austroasiatic groups from southern TYC. The final cluster, the TYC Eastern Lineage (TEL, indicated by yellow and purple), encompassed populations with close genetic ties to Sinitic groups, including the Qiang and Bai.

### Ancestry makeup and genetic origin of the four ancestrally different TYC lineages

To explore the genetic differences among the four TYC lineages, which are geographically and linguistically close, we performed an unsupervised model-based ADMIXTURE analysis using modern and ancient EA reference populations from the Illumina_2240K dataset, with K values ranging from 2 to 20 (**Fig. 2a**). The model with the lowest cross-validation error, which was considered optimal, identified four ancestral sources (K = 4). The TYC populations mainly shared northern ancestral components, most strongly seen in DevilsCave (green) and Shigatse 2.6K (light blue), along with secondary southern components represented by Yiyang (red). THL populations, including the ADT, Pumi, and Tibetans, showed a strong association with high-altitude ancient QTP populations compared to other TYC groups (Fig. 2b; Wilcoxon, P < 0.05). TCL populations, such as BMT, Yi, and Naxi, displayed relatively high levels of northern EA ancestry, similar to Neolithic Gaoshan individuals. Qiang and Bai, which fall under TEL, shared genetic components with Neolithic Yangshao/Longshan-related groups, especially China_YR_MN and China_YR_LN. In contrast, TSL populations showed a higher proportion of southern modern and ancient ancestry, setting them apart from other TYC groups (**Fig. 2b; Wilcoxon, P < 0.05**). These model-based findings suggest that the genetic structure of TYC populations results from a mixture of high-altitude, northern, and southern ancestral components, with different proportions across the THL, TCL, TSL, and TEL groups (**Fig. 2a, b**). Additionally, the high-resolution WGS dataset uncovered unique demographic histories for the THL and TSL groups, indicated by a distinct ancestral component specific to Wa, Hani, Lahu, ADT, and Pumi (**Fig. S4**).

**Fig. 2.**
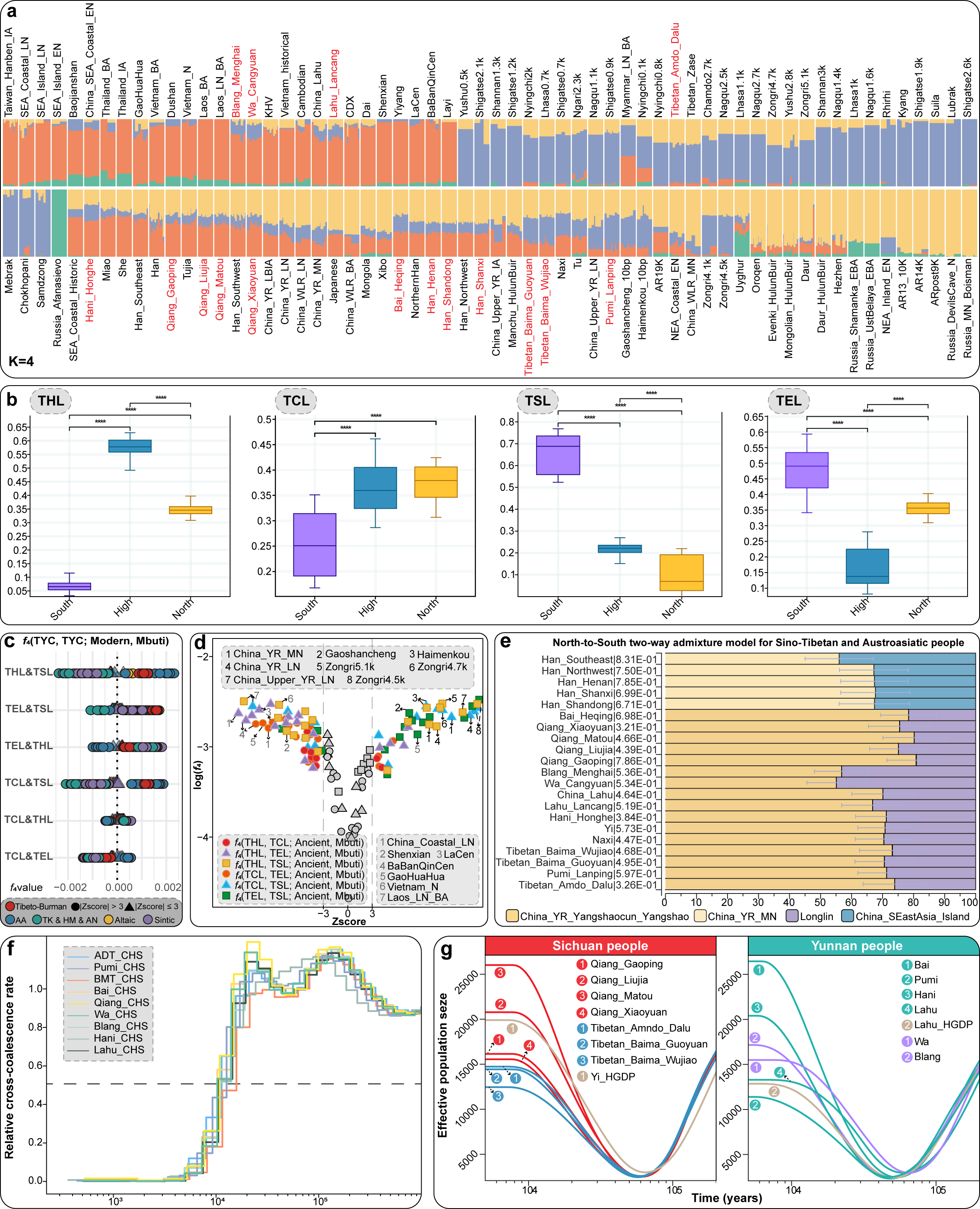
| Genetic admixture and demographic history of TYC populations. (**a**) Individual admixture proportions for TYC populations, Tibetans, present-day EA/SEA populations, ANEA, ASEA, and ancient SEA were estimated via the Illumina_2240K dataset. The minimum CV error for the four ancestral populations was selected, revealing northern ancestral components, including DevilsCave (yellow) and Shigatse 2.6K (blue), as well as southern ancestral components associated with Yiyang (red). (**b**) A Wilcoxon test was performed to assess the contributions of distinct ancestry components. The upper and lower borders represent the 95% confidence intervals. Statistical significance is denoted as follows: * for P ≤ 0.05, ** for P ≤ 0.01, *** for P ≤ 0.001, and **** for P ≤ 0.0001. (**c**) f4-statistics, in the form of f4(TYC1, TYC2; modern reference populations, Mbuti), are presented. Circles represent |Z-values| > 3, whereas triangles signify |Z-values| < 3. The colors correspond to classifications of ethnolinguistically diverse groups. (**d**) f4-statistics for f4(TYC1, TYC2; ancient reference populations, Mbuti) are displayed. The X-axis represents the logarithm of the f4 values, with distinct shapes corresponding to different groups. The colored shapes indicate |Z-values| > 3, whereas the gray shapes denote |Z-values| < 3. (**e**) Two-way admixture models of Tibeto-Burman and Austroasiatic speakers in TYC and Sinitic populations across geographically diverse regions were fitted via qpAdm, with well-fitting models illustrated. (**f**) MSMC2 cross-population results for pairs of TYC populations and Han Chinese (CHS) individuals were generated, assuming 25 years per generation and a mutation rate of 1.25 × 10□□. (**g**) Effective population sizes for TYC populations were inferred via SMC++ and computed via composite likelihoods across all individuals per population. The curves for Qiangs and ADT from Sichuan are shown in red and blue, respectively, whereas Tibeto-Burman and Austroasiatic speakers from Yunnan are represented in green and purple, respectively. The HGDP populations are highlighted with a brown curve. Abbreviations: SEA, Southeast Asia; NEA, Northeast Asia; YR, Yellow River; IA, Iron Age; BA, Bronze Age; N, Neolithic; EN, Early Neolithic; MN, Middle Neolithic; LN, Late Neolithic; LBIA, Late Bronze or Iron Age; RCCR, Relative cross-coalescence rate; TB, Tibeto-Burman; AA, Austroasiatic; TK & HM & AN, Tai-Kadai, Hmong-Mien, and Austronesian.

We further tested potentially differentiated admixtures contributing to the observed genetic differentiation and performed symmetric *f_4_*-statistics in the form of *f_4_*(TYC1, TYC2; modern reference populations, Mbuti) based on the 2240K_HO dataset. Positive Z-scores were observed, indicating that THL and TCL shared more alleles with Tibeto-Burman speakers, such as those previously reported by Ü-Tsang, Kham, and Ando Tibetans, and Ancient Northern East Asians (ANEAs), than did TSL and TEL (**Fig. 2c, d**). Notably, TCL received additional gene flow events from Tai-Kadai/Hmong-Mien/Austronesian-speaking people and the ASEA relative to THL (**Fig. 2c, d and Table S5**). Similarly, compared with the other TYC populations, the TSL population received additional gene flow from modern Austroasiatic speakers and ancient individuals from mainland SEA (**Fig. 2c, d and Table S6**). These results further support the existence of population substructures and the genetic influence of different genetic ancestries among genetically distinct TYC populations.

We then investigated whether the genetically distinct TYC populations originated from a common source. Using the qpWave method, the p_rank0 estimates showed no significant differences, suggesting genetic similarity between Neolithic Gaoshancheng individuals and modern TCL individuals when nearby ancient samples were included (**Fig. S5**). We further examined the genetic uniformity or diversity among these populations, as well as modern Sino-Tibetan groups and ancient QTB populations. qpWave analysis indicated geography-related population substructures. Additional analyses incorporating ancient ancestors, such as Neolithic Yellow River millet farmers, Neolithic rice-farming groups from Fujian and Guangxi, and Paleolithic hunter-gatherers like Zongri5.1k and Hòabìnhian, were performed. These analyses, based on ADMIXTURE results and qpAdm models, helped define the genetic components of current THL, TSL, and TEL populations. Most Tibeto-Burman and Austroasiatic groups mainly inherited ancestry from Middle Neolithic Yangshao-related YRB farmers, with minor contributions from ASEA (**Fig. 2e**). The ancestry and admixture patterns in THL, TCL, and TEL closely matched the model fitted for CPH (China_YR_MN 67%, China_SEastAsia_Island 33%). When two-way models for the post-late Neolithic period were used, the highest Qijia-related (∼96%) ancestry was found in the THL gene pool. Conversely, TSL had less YRB-related ancestry and more from Yangtze rice farmers than other TYC lineages (**Fig. S6a, Table S8**). qpAdm models also clarified the genetic makeup of other Sino-Tibetan and ancient QTB populations, confirming their primary ancestry from millet farmers and revealing diverse admixed gene pools. These findings suggest that TYC Tibeto-Burman-speaking populations share a common origin with ancestors linked to CPH, associated with millet farmers from the YRB in northern China. Furthermore, the four lineages experienced varying levels of genetic input from different millet-farming groups since the late Neolithic. This admixture variability was further supported by qpGraph analysis (Fig. S5b-d) and f4-statistics (Table S7).

### Time depth and demographical mode of TYC population separation

To estimate the timing of genetic divergence between the TYC populations and Sinitic people, we applied MSMC2, which assumes an instantaneous split with no subsequent gene flow between populations. The split time was defined as the point at which the estimated coalescence rate between two populations was half that within a single population. Using a mutation rate of 1.25×10□□ per base pair per generation and a generation time of 29 years, the midpoint estimates indicated a split between Tibeto-Burman and CHS at ∼13-14 thousand years ago (kya), between Austroasiatic and CHS at ∼14 kya, and between Tibeto-Burman and Austroasiatic populations at ∼14-16 kya (**Fig. 2f**). Divergence within the TYC populations revealed the earliest pre-Neolithic split between the TSL and the other TYC groups at approximately 15-21 kya, including splits between the TSL and TCL at ∼21 kya, between the TSL and THL at ∼16 kya, and between the TSL and TEL at ∼15 kya. Pairwise RCCR estimates placed the split between THL and TCL at ∼13 kya, with a later split from TEL at ∼12 kya (**Fig. S7**). These results support a model in which proto-TYC Tibeto-Burman speakers first diverged from proto-Austroasiatic speakers during the Upper Paleolithic, subsequently splitting from the common ancestor of proto-Sino-Tibetan speakers approximately 13-14 kya. This coincides with previous estimates of Tibetan-Han divergence ranging from ∼9 to 15 kya. Additionally, the common ancestor of the Tibeto-Burman groups in TYC further diverged into northern, central, and southern branches, approximately 10-13 kya, reinforcing the idea that TYC was a genetic melting pot.

We also applied nucleotide diversity-based Pi and site frequency spectrum-based SMC++, utilizing data from a larger set of unphased genomes to estimate effective population sizes. Most TEL and TCL populations, including the Qiang, Bai, and Yi populations, which share significant agriculture-related ancestral components, exhibited substantial population growth over the past 11 kya. This trend aligns with a broader pattern of human prehistory, where the effective population size increased as agricultural groups expanded (**Fig. 2g and Fig. S8a**). In contrast, THL and TSL populations, such as ADT, Pumi, Lahu, Wa, and Blang, exhibited modest growth while maintaining high levels of runs of homozygosity (ROH) (Fig. 2g and Fig. S8b) and long segments of identical by descent (IBD) between individuals (**Fig. S3b**). These patterns suggest long-term low but stable effective population sizes, indicating recent population isolation, which contrasts with the expansive growth observed in the TEL and TCL populations.

To further investigate potential historical admixture events in the TYC populations, we applied the haplotype-based method fastGLOBETROTTER, which uses 17 genetically distinct populations as proxies for admixture sources (see Methods). Eight out of the 13 TEL and TSL groups, including Qiang, Bai, Hani, Lahu, Wa, and Blang, could be modeled as mixtures of northern and southern ancestries. Northern ancestry, represented by Han individuals from Shaanxi Province, contributed more than 60% to the Tibeto-Burman-speaking groups, including the Qiang, Bai, and Hani. In contrast, southern ancestry, exemplified by Cambodians, accounted for a significant portion of Austroasiatic groups such as Wa (63%) and Blang (66%). The estimated dates for “one-date” admixture events ranged from 43-48 ± 8-15 generations ago for the TSL groups and approximately 12-21 ± 1-6 generations ago for the Qiang within the TEL groups. Additionally, the ADT, Pumi, and BMT populations could be modeled as admixtures of Tibetan highlanders and lowland Han populations, primarily among the THL and TCL populations. These admixture events were dated to approximately 14-24 ± 1-10 generations ago, which coincided with the Tang Dynasty (**Fig. S9b and Table S9**). We further employed linkage disequilibrium (LD)-based MALDER to explore possible admixture history in Sino-Tibetan populations. The results from the two-way admixture models supported a pattern of admixture between the northern and southern EA source populations in the CPH and TYC groups (**Fig. S9b and Table S9**). In summary, our demographic analysis of genetically diverse TYC populations suggested that prehistorical divergence times, effective population sizes driven by agricultural development, and admixture events during historical periods contributed to shaping genomic diversity.

### Identifying signatures of natural selection

Adaptive selection of genomic loci in response to extreme environments, pathogen exposure, or shifts in dietary habits, such as high altitudes with low oxygen pressure (*EPAS1*, *EGLN1*, and *HLA-DQB1*) and intense ultraviolet radiation (GNPAT), has been well-documented in Tibeto-Burman populations, particularly Tibetans living on the Qinghai-Tibet Plateau (QTP). To investigate potential population-specific adaptations in TYC populations, we conducted a genome-wide scan via population branch statistics (PBS), with Chinese Han individuals from Beijing (CHB) and Europeans from HGDP (EUR) used as ingroup and outgroup reference populations, respectively. SNPs in the top 0.1% of the PBS scores were considered candidates for natural selection, with only protein-coding genes treated as adaptive candidates (**Fig. 3a; Table S10**). In total, 17 genes were identified in the TYC populations, 16 of which were novel to this study. Among these, *OPCML*, previously associated with high-altitude adaptation based on large-scale sequencing of Tibetan genomes, was also detected (**Fig. 3b**). Gene Ontology (GO) analysis revealed that these genes were enriched in pathways related to adaptive physiological traits in TYC individuals, particularly multicellular organismal processes (GO:0032501) and responses to stimuli (GO:0050896) (**Fig. S10a**). Notably, our analysis revealed genetic variants linked to genome-wide association study (GWAS) signals, including blood protein level-related *PKP2*-rs11052294, total cholesterol level-related *AKT1*-rs2494742, and longevity-related *CFAP69*-rs9691522 variants, in at least two or more population lineages (**Fig. 3c; Table S11**). Allele frequency spectrum analysis for these loci revealed higher frequencies of derived alleles in EAs than in other continental populations (**Fig. S10b-d**).

**Fig. 3.**
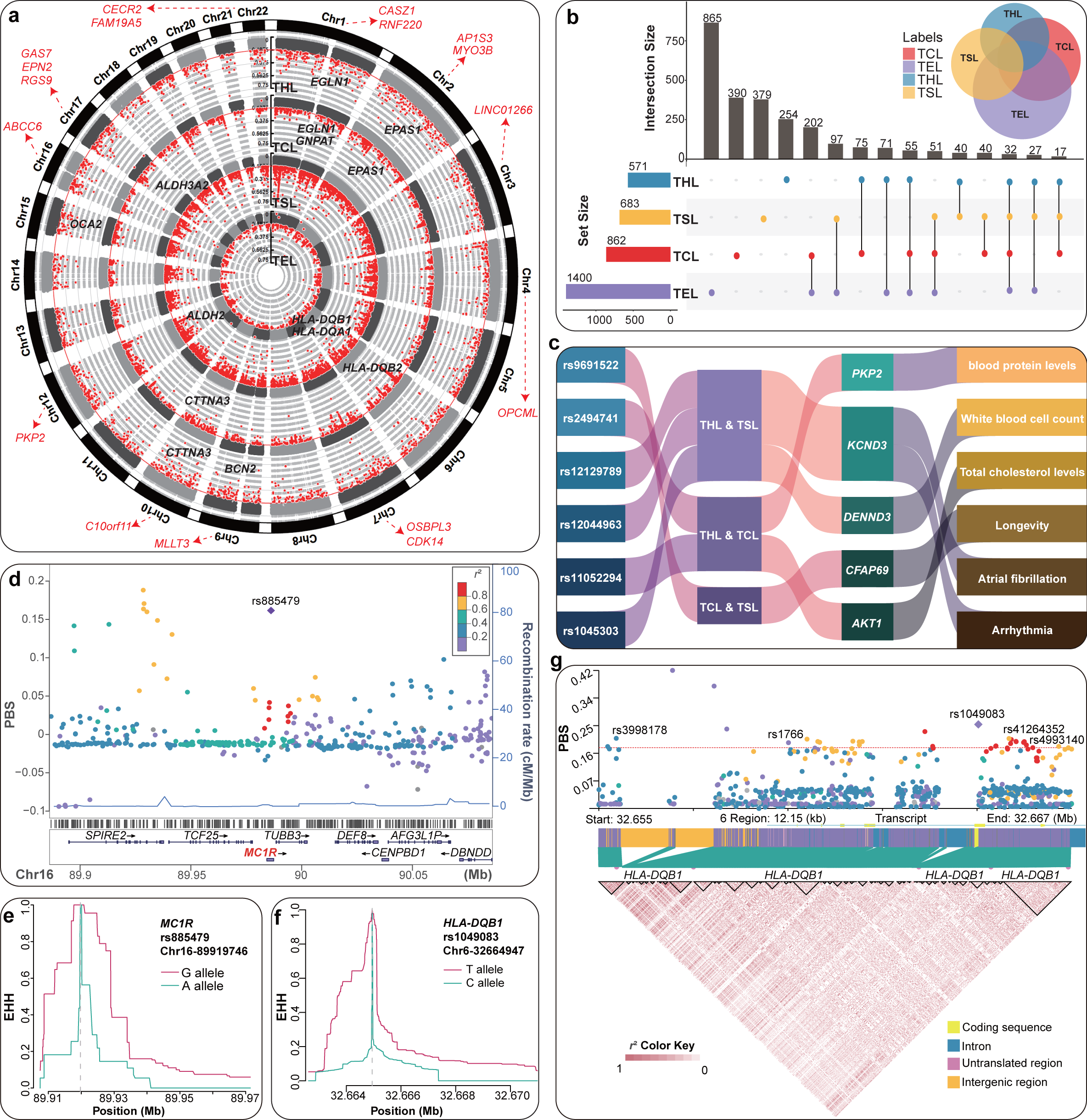
| Selection signals identified in diverse genetic backgrounds of TYC individuals. (**a**) Circular Manhattan plot depicting the population branch statistics (PBS) score distributions across various genetic backgrounds of TYC individuals. Loci within the top 0.1% of the PBS values are highlighted as red dots. Genes marked in red represent shared signatures with outlier scores in THL, TCL, and TSL, whereas those in black indicate population-specific selection signals. (**b**) Upset and Venn diagrams generated from the PBS results, with THL, TCL, TSL, and TEL colored blue, red, yellow, and purple, respectively. (**c**) Sankey diagram illustrating the cluster projection of selective signals from genetically distinct TYC populations correlated with traits annotated via the GWAS Catalog. (**d**) The MC1R genomic region demonstrates strong linkage disequilibrium (LD) between rs885479, which is under positive selection, and adjacent loci. (**e**) Extended haplotype homozygosity (EHH) surrounding MC1R-rs885479 indicates significant positive selection for the G allele. (**f**) Haplotype-based EHH analysis reveals a strong selection sweep acting on the T allele of HLA-DQB1-rs1049083. (**g**) Strong LD is observed between HLA-DQB1-rs1049083 and four previously reported SNPs associated with immunometabolism in TSL patients. Pairwise LD is measured as r² between SNPs, with red squares indicating the strength of the LD (dark red represents the strongest LD, and white represents the weakest).

To identify population-specific genetic signals in THL and Tibetans, two genetically distinct but altitude-similar groups, we conducted a genome-wide scan via the THL-CHB-EUR trio model. This analysis yielded 563 candidate genes, including well-known high-altitude adaptation genes previously identified in Tibetans. Among these variants were the *EPAS1*-rs76347095 (PBS = 0.25) and *EGLN1*-rs58696423 (PBS = 0.25) variants, which have been shown to help maintain lower hemoglobin concentrations, thus preventing polycythemia at high altitudes (Simonson et al., 2010; Yi et al., 2010). Notably, we identified a set of genes showing enriched signals of positive selection related to high-altitude adaptation (**Table S12, 13**). Among them, we discovered a novel missense variant, rs885479, in the *MC1R* gene (**Fig. 3d**). *MC1R*, known for its role in human pigment variation (Jonnalagadda et al., 2022), was functionally predicted based on a SIFT score of < 0.05. Extended haplotype homozygosity (EHH) analysis revealed extended homozygosity for this variant, and its higher frequency in THL populations than in other EA populations suggests that *MC1R*-rs885479-A may have been a target of selection related to skin pigmentation (**Fig. 3e and Fig. S11**).

Different detection methods capture distinct types and timescales of selection. To assess this, we compared candidate signatures exhibiting outlier PBS values with the haplotype-based integrated haplotype homozygosity score (iHS), which identifies more recent sweeps. The genome was divided into 20 kb windows, and windows with |iHS| values greater than 2 were considered candidate SNPs. Regions where more than 20% of the SNPs met these criteria were selected as adaptive regions. A region under positive selection was identified in THL, containing the signal-induced proliferation-associated 1 like 3 (*SIPA1L3*) gene, which encodes a GTPase-activating protein (**Table S13**). Variants within this region, including rs10421944 (iHS = 2.73), rs2384779 (iHS = 2.54), and rs138460641 (iHS = 2.50), presented strong long-range haplotypes indicative of recent positive selection, which was supported by significant iHS values (**Fig. S10f-h and S12**).

To investigate whether geographically diverse TYC populations share similar adaptation mechanisms, we examined similarities and differences in altitude-related selective signals between different TYC groups. These signals, identified in middle-altitude TCL populations, were absent in low-latitude TSL populations (**Table S10**), suggesting that isolated TSLs in forest-grassland ecotones underwent distinct local adaptations. Notably, we detected strong signatures of natural selection in *HLA-DQB1* (PBS = 0.23, iHS = 2.71), HLA-*DQA1* (PBS = 0.50, iHS = 3.09), and *HLA-DRB1* (PBS = 0.21, iHS = 2.44) via the PBS-based TSL-CHB-EUR trio model and haplotype-based iHS approach. These genes, which are part of the major histocompatibility complex class II, have been linked to several autoimmune diseases, including type 1 diabetes, celiac disease, and multiple sclerosis (GO:0002399). We identified *HLA-DQB1*-rs104649083 as a potential target of natural selection, exhibiting a strong EHH signal. Linkage disequilibrium (LD) was observed with upstream SNPs (rs3998178, rs1766) and downstream SNPs (rs41264352, rs4993140), all of which were within the top 0.01% of the PBS values (**Fig. 3f**). These four variants, in LD with HLA-DQB1-rs1049083, have been associated with immunometabolism and the body shape index, as noted in the GWAS catalog (**Fig. 3g and Fig. S10i**). Other immune-related adaptive genes, such as *CD58*, *PELI1*, and *NLRP1*, were also identified. These genes were enriched in pathways related to autoimmune diseases, including interleukin-8 production regulation (GO:0032677) and inflammasome-mediated signaling (GO:0141084). Additionally, metabolic variants under positive selection, such as *KMT2A*-rs514924 and *HLA-DRB5*-rs11967891, were found to be associated with HDL cholesterol levels, which is consistent with GWAS catalog findings.

We further characterized the selection profiles of low-altitude populations, focusing on CPH, Han_Fujian, Han_Shaanxi, and Han_Sichuan, to identify unique selection signals across central, southeastern, northwestern, and southwestern China. Using the PBS approach, we identified 30 shared genes, including *DYM* and *FAM129B*, involved in skeletal system development, and *PSPC1* and *ENOX1*, associated with rhythmic processes. Additionally, 21 genes under natural selection were detected in CPHs from Shaanxi and Han from Sichuan, particularly those related to metabolism (*ABCC11*, *DPYD*, and *ZNF536*), which were absent in southern Han from Fujian (**Fig. S13, Table S14**). These findings suggest that Han Chinese populations from agro-pastoral transitional zones possess distinct adaptive genetic foundations compared with those from southeastern coastal regions of China.

### The context for medically relevant variants

To provide population-specific insights into medically relevant genetic variation in TYC populations, we annotated our dataset via the American College of Medical Genetics and Genomics Secondary Findings gene list (ACMG v3.2) (Miller et al., 2023). Reportable variants were identified in only three individuals, each carrying a singleton variant in *TTR*-rs376790729-C-A (linked to dominantly inherited transthyretin amyloidosis) or the doubleton variant *MUTYH*-rs121918095-G-A (associated with recessively inherited polyposis) (**Fig. S14 and Table S15**). In contrast, 124 individuals (85%) in the TYC variant dataset carried at least one variant classified as “pathogenic” (P, level 5) or “likely pathogenic” (LP, level 4) according to the ClinVar Database (v.202003). Each person carried a median of two alleles (range: 1-5) (**Fig. 4a**). Among the 58 unique variants annotated as P/LP, approximately 69% (40/58) had a derived allele frequency less than 0.05. Among these, 4 variants (6.9%) presented derived allele frequencies above 0.05 across all population groups in the PGG. Han (Gao et al., 2020) (**Fig. 4b**). Notably, we identified *CYP21A2*-rs6467-C-G and *PRX*-rs3814290-C-T, with carrier frequencies exceeding 0.5% for genes associated with classic congenital adrenal hyperplasia due to 21-hydroxylase deficiency and peripheral neuropathy, respectively, in TYC populations (**Fig. 4c and Fig. S15a, b; Table S16**). The site frequency spectra of the predicted damaging variants aligned with expectations of purifying selection while also revealing a significant number of shared and common P or LP variants, underscoring their potential relevance to variant curation efforts.

**Fig. 4.**
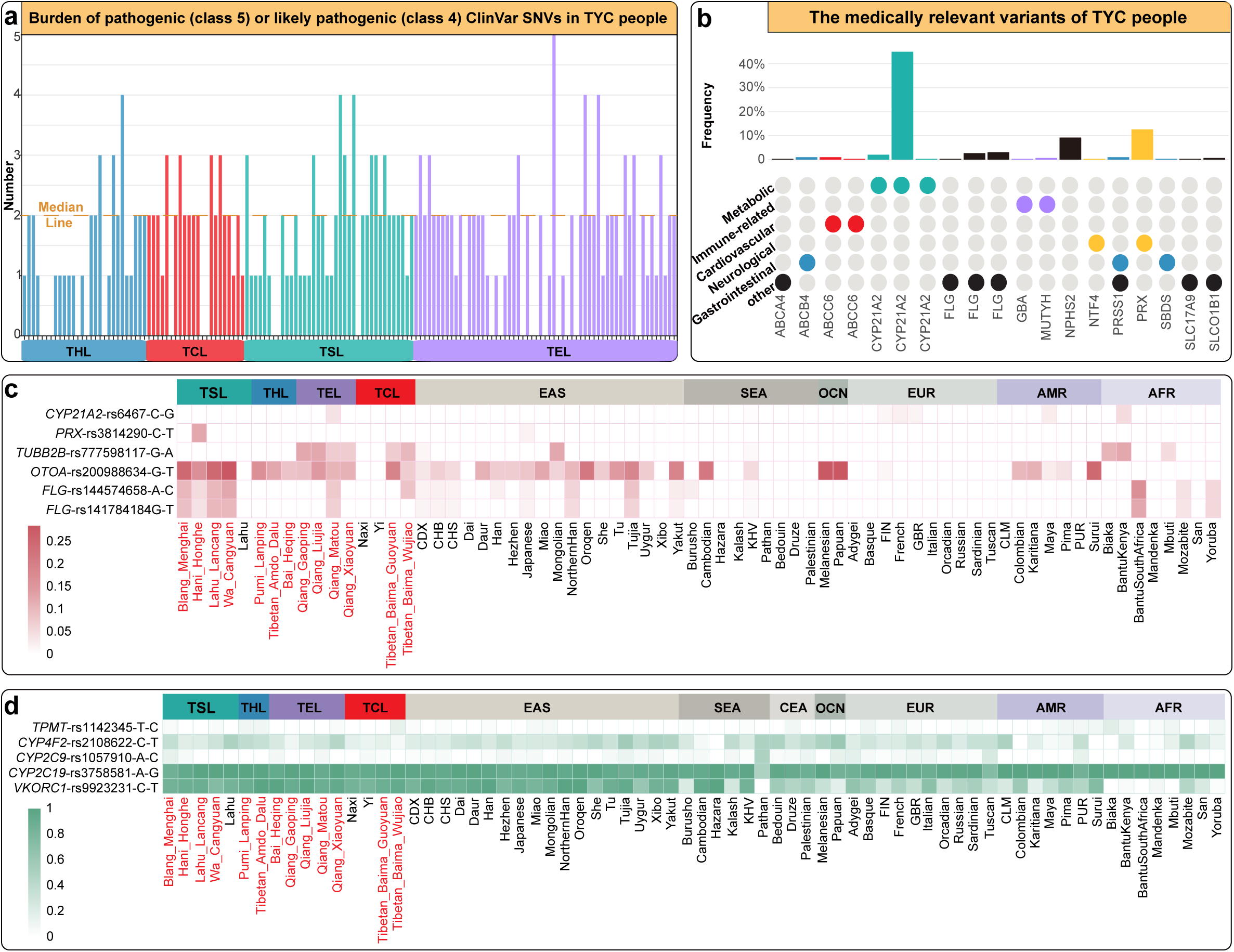
| Carriers of pathogenic/likely pathogenic (P/LP) variants and pharmacogenetic variants in TYC patients. (**a**) The burden of P (class 5) or LP (class 4) ClinVar SNVs per individual in the TYC population. (**b**) Juxtaposition of P/LP variants and the frequency of potential P/LP variants reported in genes from the American College of Medical Genetics and Genomics (ACMG) SF v3.2 list in TYC populations. (**c**) Frequency distribution of variants in genetically distinct TYC populations and worldwide populations from the HGDP and 1KGP resources, which are predicted to be P/LP according to ClinVar. For each population, the cell graphs depict the frequency of disease-associated alleles. (**d**) Population-specific frequencies of adverse drug reactions, predicted from the allele frequency distribution of known variants associated with drug response. EAS, East Asian; SEA, Southeast Asian; CEA, Central Asian; OCN, Oceanian; EUR, European; AMR, American; AFR, African.

Understanding pharmacogenomic diversity, which refers to the variation in allele frequencies that influences individual responses to medication, has significant clinical implications beyond genetic disease risk. To assess this landscape, we identified pharmacogenetic alleles of genes listed in the CPIC drug-gene pair list via PharmGKB level 1 evidence. Every individual in the TYC variant dataset (100%) carried at least one actionable pharmacogenetic variant in 23 genes associated with high-confidence gene-drug interactions, with a median of eight findings per individual (**Table S17**). This high frequency was driven primarily by the *VKORC1*-rs9923231-C-T allele, which is present in 91.8% of individuals, affects sensitivity to the anticoagulant warfarin and is known for its prevalence among Asians, suggesting a reduced dosage (Johnson et al., 2017; GenomeAsia, 2019) (**Fig. 4d**). In genetically distinct TYC populations, however, the allele was observed at relatively low frequencies. Additionally, actionable pharmacophenotypes were identified in *CYP2C19*, a gene crucial for the metabolism of commonly used medications, including the antiplatelet agent clopidogrel, proton pump inhibitors, and selective serotonin reuptake inhibitors. Notably, the frequencies of pharmacogenetic variants varied across different genetic backgrounds, with the TSL population showing the highest prevalence (**Fig. 4d**).

### Archaic introgression and the biological functions of high-confidence introgressed fragments

EAs exhibit introgressed sequences from both Denisovans and Neanderthals (Qin and Stoneking, 2015; Browning et al., 2018), although previous studies have focused largely on Tibetans from the QTP (Huerta-Sanchez et al., 2014; Zhang et al., 2021a). To identify high-confidence sequences likely introgressed from these archaic hominins, we applied IBDmix on an individual basis, intersected the results with introgression segments inferred from SPrime to identify high-confidence Neanderthal introgressed fragments and used SPrime to identify high-confidence Denisovan fragments (see Methods; **Fig. 5a**). In the TYC, after strict quality-control, individuals carried approximately 11.46 Mb of Neanderthal-derived sequences and 4.84 Mb of Denisovan-derived sequences. In total, 413 segments likely introgressed from Neanderthals and 40 from Denisovans were identified, spanning approximately 149.42 Mb and 5.97 Mb of the genome, respectively (**Fig. 5b**). The match rate distribution from SPrime revealed a higher rate of Neanderthal introgression than Denisovan introgression across all the TYC populations. Additionally, two distinct peaks were observed, representing genomic segments with low affinity for Neanderthals and higher affinity for Denisovans (**Fig. 5c**). This bimodal distribution of Denisovan match rates supports the hypothesis of two pulses of admixture with Denisovan-like archaic humans in EA (Browning et al., 2018) (**Fig. S16a**). We also identified the well-characterized Tibetan-*EPAS1* haplotype within the TYC populations. Notably, haplotypes in northern TYC populations were much closer to the Denisovan haplotype than to any modern human haplotype, particularly when considering the five-SNP Tibetan-Denisovan haplotype (**Fig. S16b**), suggesting that TYC individuals shared similar EA-specific Denisovan introgression events, as seen in Tibetans.

**Fig. 5.**
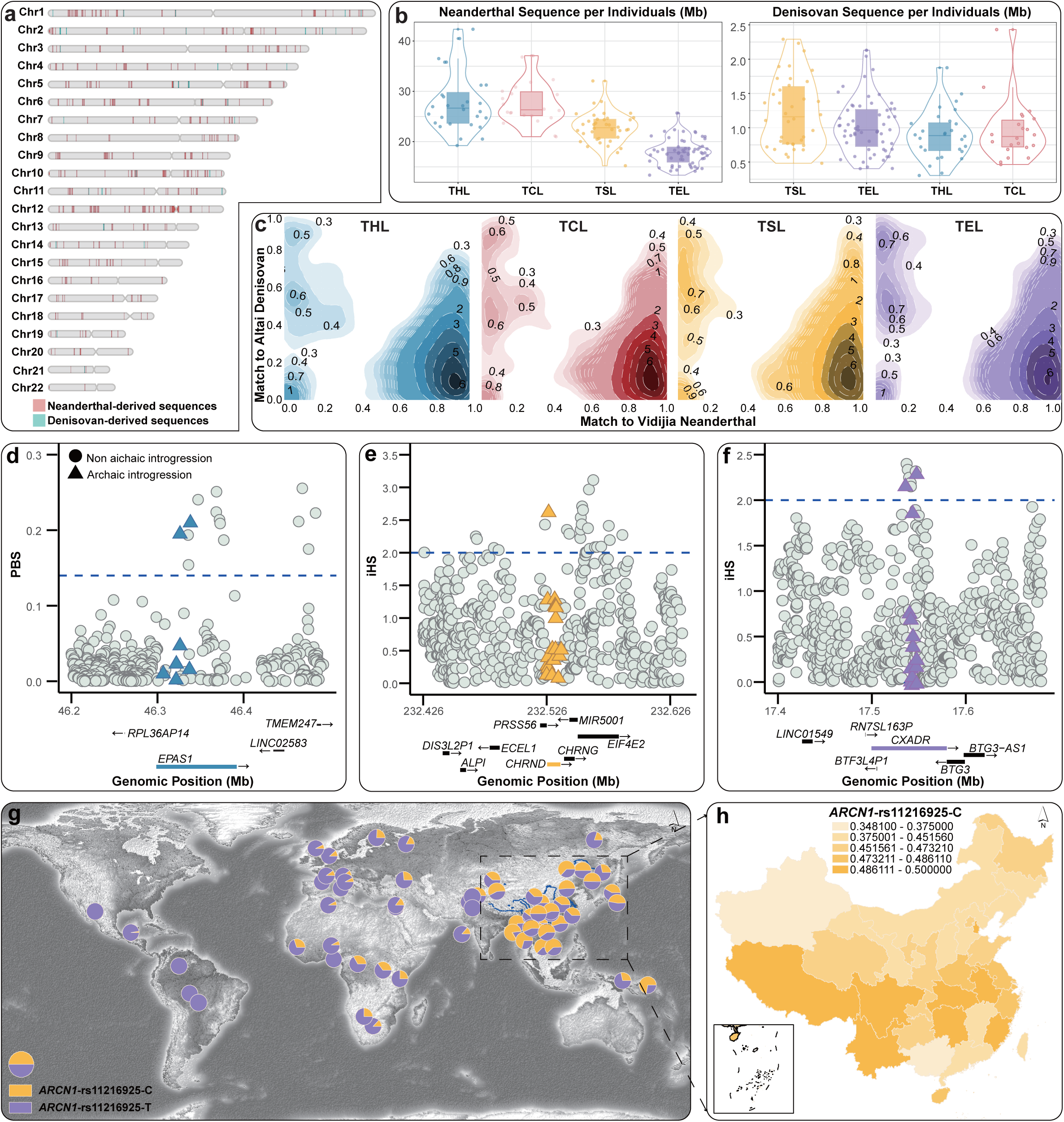
| Genetic profile and phenotypic effects of introgressed sequences from Neanderthal and Denisovan populations. (**a**) Density plot showing the distribution of introgressed sequences across each chromosome. Neanderthal-derived sequences are represented in red, whereas Denisovan-derived sequences are shown in blue. (**b**) Distribution of archaic-introgressed segment lengths on an individual basis. (**c**) Contour plot depicting the genome-wide match rate with archaic individuals in putatively introgressed segments. The x-axis represents the match rate to Neanderthals, and the y-axis represents the match rate to Denisovans. THL, TCL, TSL, and TEL are represented in blue, red, yellow, and purple, respectively. (**d-f**) Variants likely introgressed from Denisovans in the EPAS1, CHRND, and CXADR loci are associated with regulated stress erythropoiesis during hypoxia, the acetylcholine receptor signaling pathway, and cardiac muscle cell adhesion, respectively. The gray circles represent SNPs located within 100 kb upstream and downstream of the target gene, the colored triangles denote Denisovan-derived segments within the target gene, and the blue dashed line highlights regions that underwent positive selection. (**g**) Allele frequency of rs11216925, derived from Neanderthals in ARCN1, across global populations from the 10K_CPGDP, HGDP, and 1KGP genomic sources. (**h**) Frequency of the ancestral allele C, derived from Neanderthals, among Chinese individuals in the Huaxi Biobank.

We analyzed the potential phenotypic effects of the identified introgressed sequences via GWAS summary statistics derived from the integrated IBDmix and SPrime results. A total of 239 archaic segments were associated with various phenotypes, with 215 from Neanderthals and 24 from Denisovans. Among these, 104 Neanderthal-inherited segments and 13 Denisovan-inherited segments were linked to genes associated with body mass index (BMI) and height in TYC individuals (**Fig. S17a and Table S18**). Additionally, a Denisovan-like adaptive haplotype at *DIS3L2*, which is associated with lung function, was identified. Three Neanderthal-like missense variants—*CCDC138*-rs1724184, *CEP83*-rs2271979, and *IARS*-rs471478—were associated with birth weight, pulse pressure, and the COL1A1/OMD protein ratio, respectively, and were highly prevalent in TYC individuals (**Fig. S17b and Table S19**). Distinct adaptive Denisovan-introgressed haplotypes were also identified in differentiated TYC populations. Specifically, THL-specific *EPAS1*-rs7582701 was associated with IgG glycosylation (**Fig. 5d**), which regulates stress erythropoiesis during hypoxia (Wang et al., 2021b). Additionally, the TSL-specific *CHRND*-rs2245601 signal was linked to the acetylcholine receptor signaling pathway (GO:0095500), which modulates metabolic levels (**Fig. 5e and Table S20**). In TEL individuals, the *CXADR*-rs186722413 adaptive introgression was enriched in cardiac muscle cell adhesion (GO:0086042) (**Fig. 5f and Table S20**). Further analysis revealed the Neanderthal-introgressed adaptation profile across genetically distinct TYC populations. Segments at *VPS11*, *EXOC8*, and *USP36* were associated with cardiovascular function in the THL, TCL, and TEL groups, whereas haplotypes at *UBAP2* and *ARCN1* were implicated in metabolic regulation in the TSL group (**Fig. 5g, h, and Table S21**). These findings suggest that archaic introgression contributes to environmental adaptation and that differentiated adaptive introgression has intensified population differentiation within TYC populations.

## Discussion

Although the number of human genomes sequenced in medically motivated genetic studies has rapidly expanded to hundreds of thousands (Howley et al., 2025; Wang et al., 2025c; Yang et al., 2025), anthropologically informed sampling aimed at characterizing human diversity, particularly among ethnolinguistically diverse populations in Southwest China, remains underrepresented (**Fig. 1a**). In this study, 1,031 genomes from the agro-pastoral transitional zone were analyzed, providing a comprehensive genetic profile of these diverse Chinese populations. Four distinct genetic backgrounds of the TYC Tibeto-Burman and Austroasiatic-speaking groups were identified via fineSTRUCTURE analysis (**Fig. 1**). The TYC served as a crucial corridor for interaction between northern and southern ethnic groups in the eastern regions of the QTP, occupying a unique position for population contact. Genetic differentiation along a north-south cline within the TYC was observed in the first principal component, which included Sichuan Tibeto-Burman populations, Yunnan Tibeto-Burman speakers, and Yunnan Austroasiatic speakers (**Fig. 1e**). Genetic affinity and gene flow were further supported by allele-based *f_3_*/*f_4_*statistics and haplotype-based IBD analysis. Additionally, language- and geography-associated genetic substructures were identified among Baima, Ü-Tsang, Amdo, and Kham Tibetans (**Fig. S2c**). The TYC region was found to contain the most genetically diverse populations in China, reflecting its complex biological roots. Notably, the Qiang population sampled in northern Sichuan presented a distinct genetic component compared with previously published data from Qiang individuals in western Sichuan (**Fig. S4b**). These findings underscore the crucial role of anthropological sampling in bridging gaps in our understanding of human genetic diversity.

Compared with the Central Plain, which is recognized as the cradle of the Sino-Tibetan language family and Chinese civilization, archaeological materials from sites unearthed in TYC, such as Gaoshan, Haimenkou, and Jinsha, reveal distinct ethnic characteristics based on craftsmanship. These findings provide compelling evidence that indigenous populations in TYC were distinct from inland populations during the prehistoric period and that ancient DNA has the potential to elucidate their genetic origins and differential evolutionary processes (**Fig. 2**). In this study, well-fitted models using qpAdm demonstrated that both CPH Sinitic populations and genetically distinct Tibeto-Burman speakers inhabiting TYC shared a common origin from the YRB during the Middle Neolithic (**Fig. 2e**), which is consistent with previous evidence from linguistic, archaeological, and ancient DNA studies (Sagart et al., 2019; Zhang et al., 2019; Liu et al., 2022; Tao et al., 2023). Additionally, this work systematically revealed multiple genetic ancestries among genetically differentiated TYC populations since the late Neolithic, with Gaoshan and Haimenkou serving as the best proxy ancestry for the Baodun culture of BMT, Naxi, and Yi, whereas Lajia from the Qijia culture represents the primary proxy ancestry for ADT and Pumi, as supported by *f4*-statistics and qpAdm analyses (**Fig. S6 and Table S8**). These findings align with archaeological evidence indicating two distinct southward migrations of TYC populations (Shi, 2023). Moreover, the divergence times estimated via MSMC2 suggest that the TYC and CHS populations began to diverge at approximately 13-14 kya (**Fig. 2f**), with the earliest separation occurring between Tibeto-Burman and Sinitic speakers, indicating the origin of the proto-TYC populations in the upper YRB, related to millet farmers, which later gave rise to distinct genetic backgrounds in different groups. Additionally, by applying LD decay-based MALDER and haplotype-based fastGLOBETROTTER methodologies (**Table S9**), the north-south migratory movements of proto-TSL/TCL and the eastward migration of highland-related ancestry from the QTP into northern TYC for THL during the historical period were reconstructed. These patterns align with ethnological records (**Fig. S9**). These results fill significant gaps in the genetic understanding of the parallel migration routes of Tibeto-Burman speakers through TYC and the demographic history of populations in this region. These findings underscore the importance of leveraging multi-population WGS resources to disentangle the complex formation patterns of TYC populations.

The underrepresentation of TYC populations in human genetic studies has limited the diversity of individuals in WGS genomic datasets, leading to reduced analysis of genetic variants. In this study, genetic variations were cataloged via dbSNP version 146, revealing approximately 13.6 million biallelic SNPs in the TYC dataset, including 3.3 million novel variants, 89% of which occurred at rare frequencies (**Fig. 1c, d**). Clinically significant genetic variation in TYC populations was also characterized, with risk profiles highlighted for dominant and recessive ACMG reportable variants, as well as P/LP genetic variants annotated in ClinVar (**Table S16**). Previous research has revealed differences in disease burden concentrations between ancestrally diverse populations and those of European descent (Buchanan et al., 2020; Chan et al., 2022). For example, the genetic risk of *CYP21A2*-rs6467-C-G, which is associated with autosomal recessive congenital adrenal 21-hydroxylase deficiency, exceeded 0.5% in TYC individuals (**Fig. 4c**). The site frequency spectra of variants predicted these to be rare in the gnomAD database for EA populations, and nonconformity in these findings is likely due to variant curation efforts. Similarly, *PRX*-rs3814290-C-T, which is rare in EA populations, was frequently carried by TSL individuals (**Fig. 4c**), likely due to private variations within the genetic background. These findings underscore the importance of utilizing WGS resources for underrepresented populations to advance precision medicine in China directly, necessitating more comprehensive, large-scale sampling that considers the fine-scale genetic structure of Chinese populations. Pharmacogenomic analysis further profiled high-confidence gene-drug interactions in TYC patients (**Table S17**). Warfarin, a commonly used anticoagulant, exhibits significant interpatient variability in dosage requirements for effective anticoagulation (Fig. 4d), with lower doses recommended in EA populations to minimize bleeding risk (Johnson et al., 2017). A significantly greater prevalence of *VKORC1*-rs9923231-C-T was identified in eastern China compared to western China, based on large-scale data from the Huaxi biobank (**Fig. S15d**), demonstrating differences in pharmacogenetic variants across populations with distinct genetic backgrounds. This highlights the necessity of conducting pharmacogenomic analyses on underrepresented populations to further advance precision treatment.

The TYC was identified as a geographical transition zone between the high-altitude QTP and the lowland EA, characterized by a topography that slopes from northwest to southeast. This positioning enabled the identification of both shared and population-specific genetic adaptation signatures associated with population differentiation. Shared candidate SNPs in the top 0.1% of scores were mapped to 17 genes (**Fig. 3a**), and GO pathway analysis revealed enrichment in multicellular organismal process (GO:0032501) and response to stimulus (GO:0050896) genes (**Table S12**). Among these genes, the *OPCML* gene, previously associated with high-altitude adaptation, was identified using low-coverage WGS resources (Zheng et al., 2023). These findings suggest that the TYC population exhibited a shared pattern of adaptation to mid-to high-altitude environments. Additionally, different adaptive genes and functional enrichment categories indicated that the unique living environments and habits of the THL and TSL populations likely shaped specific adaptive evolution. A total of 563 candidate high-altitude adaptation genes were identified (**Table S10**), including the well-characterized *EPAS1* and *EGLN1* genes. Allele frequency-based *MC1R* and haplotype-based *SIPA1L3* were detected in THL, with *MC1R* associated with skin pigmentation, suggesting adaptation to high-altitude environments with elevated ultraviolet radiation (**Fig. 3d**). *SIPA1L3*, which encodes a GTPase-activating protein involved in HIF-1 activation, further supports this high-altitude adaptation, as the small GTPase Rac1 is known to be activated under hypoxic conditions, promoting HIF-1 protein expression and transcriptional activity (Hirota and Semenza, 2001). Previous analyses have indicated that the Deng people possess a unique genetic basis for high-altitude adaptation, involving the *HLA-DQB1* gene. This gene was also identified in TSL populations inhabiting the hilly and forested regions of southern TYC. A novel *HLA-DQB1*-rs1049083 signal, which displays strong evidence of positive selection (**Fig. 3f**), was observed, with robust linkage disequilibrium to SNPs with immunometabolism-related functions annotated in the GWAS Catalog (**Fig. 3g**). These findings suggest that the genetic basis of adaptation in TSL reflects a population-specific response to dietary and pathogen pressures rather than convergent adaptation to high-altitude environments. Leveraging large-scale genomic resources from the CPH populations, 30 high-quality genes associated with skeletal system development and rhythmic processes were identified (**Table S14**). These genes were shared with genomes from the northwest, southeast, and southwest Han populations (**Fig. S13**), as documented in the 10K_CPGDP (He et al., 2025). Moreover, distinct adaptive genetic bases associated with metabolic pathways were observed in Sinitic-speaking populations from the agro-pastoral transitional zone of China, including CPH, compared with southeastern coastal Han populations.

This study investigated the genomic and phenotypic impacts of Denisovan and Neanderthal DNA within four distinct genetic backgrounds of TYC populations, each of which has undergone unique environmental adaptations. Using IBDmix and SPrime, 453 introgressed segments were identified, with 413 likely derived from Neanderthals and 40 from Denisovans (**Fig. 5a**). Evidence for two waves of Denisovan admixture and one wave of Neanderthal introgression was observed, which was supported by match rate computations (**Fig. 5c, Fig. S16a**) and aligned with prior findings in EA populations (Browning et al., 2018; Choin et al., 2021). Introgressed Denisovan sequences at the *EPAS1* locus have previously been linked to high-altitude adaptation in Tibetans (Huerta-Sanchez et al., 2014). Similarly, northern TYC populations, including the Qiang, ADT, and BMT, exhibited a closer relationship between the EPAS1 haplotype and the Denisovan haplotype than other modern populations did (**Fig. S16b**). Two SNPs at the *EPAS1* locus (rs10193827 and rs7582701) were identified as contributing to THL and TCL adaptations to hypoxia (**Fig. S17b**). In addition to specific cases such as *EPAS1*, the broader impacts of archaic introgression on human phenotypes remain poorly understood. Adaptive Denisovan-introgressed strains, such as the TSL-specific *CHRND*-rs2245601, associated with metabolism (**Fig. 5e**), and TEL-specific *CXADR*-rs186722413, linked to the cardiovascular system (**Fig. 5f**), have been reported. Additionally, adaptive Neanderthal-derived segments at *VPS11*, *EXOC8*, and *USP36* were found to impact the cardiovascular system in THL, TCL, and TEL. In contrast, segments at *UBAP2* and *ARCN1* influenced the metabolic system in TSL (**Fig. 6g-h, Table S21**). These findings demonstrate that archaic introgression facilitates environmental adaptation in modern humans and that differential adaptive introgression contributes to the genomic diversity of TYC populations. Moreover, the population specificity of significant introgressed variants in EAs was highlighted, suggesting that archaic variant-phenotype associations might be overlooked when relying solely on European data. Further research on genetically diverse EA populations will be necessary to fully understand these associations.

## Conclusion

In summary, this study revealed the genetic characteristics of TYC Indigenous populations, including novel genetic variants, medically relevant variants, and archaic introgressions that were previously undetectable via microarray data. The potential applications of WGS in personalized medicine and clinical settings were emphasized, alongside the importance of extending WGS to diverse populations to better capture genetic characteristics and gain a population-specific understanding of human history. Previous genetic studies, limited by microarray resources, small sample sizes, and restricted geographic regions, have likely missed a substantial portion of population-specific genetic variants, thereby hindering the analysis of fine-scale genetic structure, complex demographic history, local adaptation, medically relevant variants, and archaic introgression. Therefore, large-scale, high-depth WGS projects covering broader sample sizes and geographic regions are essential for elucidating the evolutionary history of the TYC population and other underrepresented ethnolinguistically distinct Chinese groups. Additionally, the adoption of telomere-to-telomere and pangenome reference genomes is recommended for sequence alignment to improve the detection of complex genetic variants.

## Materials and methods

### Population samples and ethical statement

A total of 147 peripheral blood samples were collected from 126 individuals of Tibeto-Burman descent and 22 individuals of Austroasiatic descent. The Tibeto-Burman speakers included 44 Qiang, 16 Amdo Tibetan, and 22 Baima Tibetan individuals from Jiuzhaigou County, Aba Tibetan and Qiang Autonomous Prefecture, Sichuan Province, China, along with 15 Bai, 12 Hani, 5 Lahu, and 12 Pumi individuals from Dali, Honghe, Puer, and Nujiang in Yunnan Province, respectively. The Austroasiatic-speaking individuals consisted of 11 Blang from Xishuangbanna and 10 Wa from Lincang, Yunnan Province, China. Additionally, saliva samples were collected from 884 Sinitic speakers from Shandong, Shanxi, and Henan Provinces through comprehensive geographic sampling. All participants were offspring of nonconsanguineous marriages of indigenous residents within three generations. All procedures adhered to the recommendations of the Declaration of Helsinki, which was revised in 2000 (World Medical Association, 2001). The study protocols were reviewed and approved by the Ministry of Science and Technology of the Human Genetic Resources Administration of China (HGRAC), registration number 2024SQCJ000261, and the Medical Ethics Committee of West China Hospital of Sichuan University, approval number 2024-173.

### Genome sequencing and data processing

Genomic DNA was extracted from the peripheral blood of 147 Tibeto-Burman and Austroasiatic individuals using the QIAamp DNA Mini Kit (QIAGEN, Germany) according to established protocols. WGS was performed on the DNBSEQ-T7 platform (MGI, Shenzhen, China), yielding a mean depth of 12.5× for 150 bp paired-end reads, with standard library preparation. Each TYC sample was processed in a separate lane, generating at least 30 GB of data that passed the filtering process. Quality control ensured that 80% of the bases achieved a base quality score of at least 30. The reads were merged, adaptors were trimmed, and the sequences were aligned to the human reference genome via BWA v.0.7.13 (Li and Durbin, 2010). Variant calling was conducted with the HaplotypeCaller module in the Genome Analysis Toolkit (GATK) (DePristo et al., 2011). For downstream analysis, only biallelic variants were retained, resulting in a final dataset of 26,141,086 single nucleotide polymorphisms (SNPs). Additionally, 884 Han Chinese individuals, reported here for the first time, were genotyped via the Infinium Global Screening Array (Illumina, CA, USA) and aligned to the GRCh37 reference genome. Genetic relatedness between individuals was estimated via PLINK v.1.90 (Chang et al., 2015) and King (Manichaikul et al., 2010). SNPs with a missing call rate greater than 0.05 or those failing Hardy□Weinberg equilibrium testing (--geno 0.05 and --hwe 10-6) were removed, along with individuals with genotyping rates exceeding 0.05 (--mind 0.05).

### Population dataset

i. WGS dataset: Whole-genome sequence data from phase 3 of the 1KGP (Genomes Project et al., 2015) and HGDP (Bergstrom et al., 2020) were used for genotype calling and quality control alongside newly sequenced TYC samples, which were mapped to the human reference genome GRCh38. This resulted in a WGS dataset comprising 964 samples and 26,141,086 variants, which revealed genetic variants, selective signatures, and medically relevant variations.
ii. 2240K dataset: PileupCaller v1.5.2 was used to call 2240K SNVs per sample from BAM files mapped to the human reference genome (GRCh37). To facilitate a comprehensive spatiotemporal analysis of population history, this dataset was merged with the publicly available 1240K dataset from the Allen Ancient DNA Resource (AADR, https://reich.hms.harvard.edu/allen-ancient-dna-resource-aadr-downloadable-genotypes-present-day-and-ancient-dna-data), resulting in the 2240K dataset, which contains 1,135,618 SNPs. Further integration with publicly available HumanOrigin (HO) dataset from the David Reich Laboratory (https://reich.hms.harvard.edu/datasets) has generated the 2240K_HO dataset, which contained 593,050 SNPs.
iii. WGS (GRCh37) dataset: BAM files generated by mapping raw sequencing data to the human reference genome GRCh37 were utilized to simulate demographic history. Additionally, VCF files were used to identify archaic introgression, integrating data from Altai and Vindija Neanderthal (Prufer et al., 2014), as well as Altai Denisovan (Meyer et al., 2012) individuals.
iv. Illumina dataset: Previously reported populations genotyped via the Illumina chip from the 10K_CPGDP dataset were combined with newly studied Han Chinese individuals to form the Illumina dataset, which includes 516,448 SNPs. Further integration with publicly available datasets from the HGDP/Oceania genomic resource, the HO dataset, and the 2240K dataset generated the Illumina_HGDP, Illumina_HO, and Illumina_2240K datasets, which contained 460,678, 55,649, and 146,802 SNPs, respectively.

### Variant quality control and annotations

Following stringent quality control used in 10K_CPGDP (He et al., 2025), variants were categorized into SNPs and InDels via VCFtools (version 0.1.16; http://vcftools.sourceforge.net/) based on the following criteria: (1) sites with a quality score below 30 were excluded, and (2) sites with a P-value less than 1e-10, indicating deviation from HWE, were also removed. After applying these filters, allele counts were calculated, and only biallelic SNPs were retained for further analysis, resulting in 9.8 million identified biallelic SNPs. The clinical significance of the identified SNPs and their GWAS hits were annotated via ClinVar (version 20240419; https://www.ncbi.nlm.nih.gov/clinvar/) and the GWAS catalog (version 2024-04-23; https://www.ebi.ac.uk/gwas/), respectively. The dbSNP dataset (build 156; https://ftp.ncbi.nih.gov/snp/archive/) was consulted to determine whether the identified SNPs in the 147 WGS samples had been previously reported. Variants were classified as known if they matched a dbSNP variant in terms of genomic position, reference allele, and alternative allele; otherwise, they were considered novel.

### Population genetic analysis

#### Principal component analysis

PCA was performed via the smartPCA program from EIGENSOFT (Patterson et al., 2006), with parameters set to numoutlieriter: 0 and lsqproject: YES. To investigate the genetic relationships between TYC individuals and global populations, a worldwide PCA was conducted using a whole-genome sequencing dataset (GRCh38) with biallelic autosomal common variants (MAF > 0.05). Additionally, a series of PCAs were carried out to examine population structure within the context of ancient and modern EA and mainland SEA Tibeto-Burman, Austroasiatic, and Sinitic speakers. In these analyses, the studied populations were iteratively refined by removing “outliers” based on the first two PCs via the Illumina_2240K dataset, which projected selected ancient individuals onto the background of modern populations.

#### Genetic difference analyses

Pairwise fixation indices (*F*_ST_) were calculated via PLINK v.1.90 (Chang et al., 2015) to estimate the *F*_ST_-based genetic distances between the studied TYC/Sinitic individuals and various geographic and linguistic reference populations. These analyses were based on the Illumina_2240K dataset, with all the samples included.

#### Model-based ADMIXTURE analysis

For admixture analysis, quality control was performed on the WGS dataset and the Illumina_2240K dataset using pruned SNPs in strong linkage disequilibrium. The parameters for pruning were set to --indep-pairwise 50 10 0.1, and analyses were conducted in PLINK v.1.90 (Chang et al., 2015). ADMIXTURE v.1.3.0 was run 100 times with a random seed for each value of K from 2 to 20, generating cross-validation error estimates to determine the optimal value of K (Alexander et al., 2009).

#### Testing for the presence of admixture

To investigate and validate genetic drift between the TYC people and reference populations, we calculated outgroup *f3*-statistics via *f3*(Reference, TYC; Mbuti) based on the qp3Pop module in ADMIXTOOLS (Patterson et al., 2012). Admixture *f3*-statistics, in the form of *f3*(Source1, Source2, TYC), were also conducted to explore potential admixture signals in the focal populations, with significantly negative *f_3_* values (Z scores less than −3) indicating evidence of admixture between two predefined ancestry surrogates. Additionally, qpDstat software in ADMIXTOOLS was employed to assess gene flow events via the *f_4_*:YES parameters. Three general models were constructed for analysis. First, the symmetrical *f_4_*(TYC1, TYC2; Reference, Mbuti) was used to examine whether two sublineages of the TYC population formed a clade relative to the reference populations. The asymmetrical *f_4_*(Reference1, TYC; Reference2, Mbuti) and affinity *f_4_*(Reference1, Reference2; Target, Mbuti) methods were applied to test whether the TYC population shares more alleles with Reference1 or Reference2 than other populations do.

#### Runs of homozygosity and nucleotide diversity estimation

To detect potential consanguinity within populations from the Illumina_2240K dataset, we employed PLINK v.1.9 (Chang et al., 2015) to identify ROH. The sliding window size was set to 500 kb, containing a minimum of 50 SNPs, allowing 1 heterozygote and up to 5 missing calls per window. The total ROH length was then calculated to enable a comparative analysis of distribution patterns across populations at the population level. Nucleotide diversity was computed via VCFtools (Danecek et al., 2011), with estimates based on average pairwise differences within each population for every 20 kb window, with no overlap.

#### Demographic history modeling with qpAdm and qpWave

To assess ancestry proportions and admixture patterns among the thirteen populations inhabiting TYC, we conducted qpAdm analyses via ADMIXTOOLS2 (Patterson et al., 2012). These populations were modeled as mixtures of predefined source populations, where the ancestry of target populations was estimated using a set of source populations (left populations) and a set of reference populations (right populations), without requiring an explicit phylogenetic relationship between them. The reference populations, selected for their distinct relationships with the target populations, included Mbuti, Papuan, Russia_Ust_Ishim, Onge, Mixe, Italy_North_Villabruna, Iran_GanjDareh, and Russia_Kostenki14. qpAdm models were accepted when the p-value exceeded 0.05, provided that the admixture proportion and its standard error were positive; models with p-values below 0.01 were rejected. Prior to qpAdm modeling, qpWave analyses were performed to ensure that the reference populations were sufficiently differentiated on the basis of their unique genetic affinities and relationships with the outgroup. The outgroups, as identified in previous studies (Tao et al., 2023), included Mbuti, Russia_Afanasievo, Papuan, Japan_Jomon, China_AR_EN, Longlin, China_NEastAsia_Coastal_EN, China_SEastAsia_Island_EN, and Laos_Hoabinhian.

#### Admixture dating

To provide further insights into the historical context of admixture events, MALDER (v.1.0) (Loh et al., 2013) was utilized to estimate the timing of multiple admixture events within TYC populations. A minimum genetic distance of 0.005 cM was set for curve fitting to account for short-range linkage disequilibrium. Haplotype-sharing patterns were examined via ChromoPainter, which is based on the Illumina_HGDP dataset, to detect potential admixture within TYC populations. Subsequently, fastGLOBETROTTER (Wangkumhang et al., 2022) was applied by modeling the painting profile of a target cluster, as inferred by ChromoPainter, as a mixture of those from a set of reference clusters. The donor populations included Han_Xian, Yao_Gulei, Yao_Wangmo, Li_Ledong, Li_Linshui, Li_Wuzhishan, Tibetan_Zase, Daur, Hezhen, Basque, French, Balochi, Brahui, Pathan, Sindhi, Yakut, and Cambodian. In fastGLOBETROTTER, 100 bootstrap replicates were performed to assess the statistical significance of the admixture event and to establish a 95% confidence interval for dating the admixture.

#### Identification by descent and fine-STRUCTURE analyses

Haplotypes were estimated via SHAPEIT v.5.0, which is based on recommended human genetic maps (Hofmeister et al., 2023). Shared IBD segments within and between individuals or populations were detected via Refine IBD (Browning and Browning, 2013). The coancestry matrix was calculated, and the fine-scale population structure was explored via fineSTRUCTURE v.4.0 with default parameters.

#### Estimation of Ne and divergence time

The historical effective population size (Ne) for each Tibeto-Burman/Austroasiatic group was inferred via the SMC++ program (Terhorst et al., 2017), with analyses performed on all available genomes. MSMC2 was then applied to infer the divergence time of the TYC population (Schiffels and Wang, 2020), with two samples randomly selected from each population. Initially, sample-specific VCF and mask files were generated via bamCaller.py from the MSMC-tools package. Multistep files for two-phased diploid individuals, including target TYC individuals and Han individuals from 1KGP, were then produced, and VCF and mask files were merged via the generate_multihetsep.py script. MSMC2 was subsequently run to estimate coalescence rates, and the combineCrossCoal.py repository was used to create a joint output file. The relative cross-coalescence rate (rCCR) was computed, and the divergence time was estimated when the rCCR reached 0.5. For all the demographic analyses, the results were scaled, assuming a mutation rate of 1.25×10-8 per base per generation and a generation time of 29 years. Both SMC++ and MSMC2 analyses were performed via the WGS (GRCh37) dataset.

### Biological adaptation

#### Identification of positive natural selection

For natural selection analyses, allele frequency-based (PBS) and haplotype-based (iHS) methods were applied via the WGS dataset. The PBS approach was first employed to detect signals of natural selection in the TYC population, using the Chinese Han (CHB) population from Beijing as the ingroup and the WGS data of the French, Basque, Sardinian, Italian, and Tuscan populations from HGDP (EUR) as the outgroup reference. SNPs with extreme branch lengths (top 0.1%) were considered strong candidates for the genetic basis of adaptive evolution. Haplotype-based tests were then performed to confirm these selection signals, with iHS estimated via selscan v.1.2.078 for each lineage of the TYC population. To detect local adaptive signals, we normalized iHS scores across 20 kb windows, excluding windows with fewer than 20 SNVs. Based on the background distribution of normalized scores across all TYC lineages, windows with |iHS| > 2 were selected. Additionally, PBS was conducted on the Sinitic population from the Central Plain to detect selection signals via the Illumina_HGDP dataset, with the Miao_Guizhou and EUR populations serving as ingroup and outgroup references, respectively.

#### Functional annotation of natural selection signatures

Functional enrichment analysis of genes with strongly selective signals was conducted via Metascape (https://metascape.org/gp/index.html#/main/step1), an online tool that integrates widely used functional category ontologies. Previously reported genome-wide associations for genes and variants were identified through the GWAS Catalog database.

#### Medically relevant variants and pharmacogenomic

The WGS dataset was utilized to catalog medically relevant variants. Medically actionable variants were identified by annotating SNVs based on the gene list specified by the ACMG for reporting secondary findings in clinical exome and genome sequencing (Miller et al., 2023). The genetic burden associated with the ACMG gene panel version 3.2 was assessed via the following metrics: total number of observed alleles, median and range of alleles per individual, the proportion (%) of samples carrying at least one allele, and the genes in which variants were detected. Clinical annotation of SNVs was performed via the NCBI ClinVar database (https://www.ncbi.nlm.nih.gov/clinvar/), a publicly accessible resource linking medically relevant variants to phenotypes ranging from 0-Uncertain significance to 5-Pathogenic. The burden of these variants was also calculated, including the total allele count, median (range) number of alleles per sample, and number of samples with at least one allele and genes, which was consistent with the ACMG variant analysis. To explore the pharmacogenomic landscape, 23 CPIC drug□gene pairs were compiled in conjunction with PharmGKB clinical annotation levels 1A/1B, defined as follows: (Level 1A) gene□drug pairs with variant□specific prescribing guidance in clinical guidelines or FDA-approved drug labels, supported by at least one clinical annotation; (Level 1B) gene□drug pairs without variant□specific guidance but backed by high-level evidence from at least two independent studies affirming the association (Whirl-Carrillo et al., 2021).

#### Detection of introgressed sequences and variants

To identify sequences likely introgressed from Neanderthals or Denisovans, Sprime (Browning et al., 2018) and IBDmix (Chen et al., 2020) were applied to the WGS (GRCh37) dataset. Combining multiple methods for detecting archaic introgressed sequences has been shown to improve the detection rate of authentic introgressed segments (Jacobs et al., 2019). To minimize the false positive rate, only shared introgressed segments were retained to establish a high-confidence set of introgressed sequences. SPrime was used without an archaic reference genome to identify introgressed segments. Since SPrime cannot handle missing data, sites with missing genotypes were excluded. A threshold of 140,000 for the SPrime score was set on the basis of score distribution across populations, with only introgressed haplotypes containing more than 30 sites considered, as longer haplotypes provide stronger evidence of archaic origin. Neanderthal-specific segments were defined by a match rate to Neanderthal of 0.6 or higher and a Denisovan match rate below 0.3. Conversely, Denisovan-specific segments were identified with a Denisovan match rate above 0.3 and a Neanderthal match rate below 0.3.

Additionally, IBDMix, a computational approach utilizing an archaic reference genome, was employed to detect introgressed archaic haplotypes within phased genomic data. Phasing was conducted via Beagle 2.0 with default parameters. Prior to the analysis, genomic regions inaccessible in Vindija and Altai Neanderthals, as well as Altai Denisovan, were masked. The following filters were applied: (1) Coverage filter stratified by GC content, (2) minimum coverage of 10, (3) Heng Li’s Mapability 35 (map35_100), (4) mapping quality score threshold of 25, and (5) exclusion of tandem repeats, InDels, and sites located within CpG islands. For the introgressed sequences, a final set of segments was curated using a logarithm of odds threshold greater than 4 and a minimum segment length of 50 kilobases. Pathway analysis was performed to identify biological pathways enriched in genes containing archaic variants using Matascape (https://metascape.org/gp/index.html#/main/step1), and associations with archaic variants were confirmed using GWAS summary statistics (https://www.ebi.ac.uk/gwas/).

## Data Availability

All the data generated or analyzed during this study are included in this published article, its supplementary information files, and publicly available repositories. The genome-wide variation data were obtained from the public dataset of the Allen Ancient DNA Resource (AADR) in the Reich Laboratory (https://dataverse.harvard.edu/dataset.xhtml?persistentId=doi:10.7910/DVN/FFIDCW). The results of the analyses have been submitted to the supplementary materials and deposited into the OMIX database (https://ngdc.cncb.ac.cn/omix/) under accession number OMIX007569. The whole-genome sequencing data from the 147 Tibeto-Burman and Austroasiatic individuals sampled in the Tibetan-Yi Corridor, generated in this study, have been deposited in the Genome Variation Map 92 in the National Genomics Data Center, Beijing Institute of Genomics, Chinese Academy of Sciences and China National Center for Bioinformation 93 under accession number GVM000862 (https://ngdc.cncb.ac.cn/gvm/getProjectDetail?project=GVM000862). The variation data of 884 Sinitic speakers reported in this paper have been deposited in the Genome Variant Map under accession number GVM000886 (https://ngdc.cncb.ac.cn/gvm/getProjectDetail?project=GVM000886). Data access and use are restricted to academic research in population genetics, including research on population origins, ancestry and history. These data are available under restricted access due to the sensitive information they contain, which could compromise the privacy and consent of the research participants. Requests for access to data can be directed to Guanglin He. The access to and use of data must comply with the regulations of the People's Republic of China on the administration of human genetic resources (No. 2025BAT00381 and 2025BAT00386).

https://ngdc.cncb.ac.cn/gvm/getProjectDetail?project=GVM000886

## Declarations

### Ethnic approval and consent to participate

The protocols were reviewed and approved by the Ministry of Science and Technology of the Human Genetic Resources Administration of China (HGRAC; registration number 2024SLCJ000164) and the Medical Ethics Committee of West China Hospital of Sichuan University (approval number 2024-173). The study was conducted in accordance with the principles of the Helsinki Declaration.

### Availability of data and materials

All the data generated or analyzed during this study are included in this published article, its supplementary information files, and publicly available repositories. The genome-wide variation data were obtained from the public dataset of the Allen Ancient DNA Resource (AADR) in the Reich Laboratory (https://dataverse.harvard.edu/dataset.xhtml?persistentId=doi:10.7910/DVN/FFIDCW). The results of the analyses have been submitted to the supplementary materials and deposited into the OMIX database (https://ngdc.cncb.ac.cn/omix/) under accession number OMIX007569. The whole-genome sequencing data from the 147 Tibeto-Burman and Austroasiatic individuals sampled in the Tibetan-Yi Corridor, generated in this study, have been deposited in the Genome Variation Map (Li et al., 2021) in the National Genomics Data Center, Beijing Institute of Genomics, Chinese Academy of Sciences and China National Center for Bioinformation (Members, 2022) under accession number GVM000862 (https://ngdc.cncb.ac.cn/gvm/getProjectDetail?project=GVM000862). The variation data of 884 Sinitic speakers reported in this paper have been deposited in the Genome Variant Map under accession number GVM000886 (https://ngdc.cncb.ac.cn/gvm/getProjectDetail?project=GVM000886). Data access and use is restricted to academic research in population genetics, including research on population origins, ancestry and history. These data are available under restricted access due to the sensitive information they contain, which could potentially compromise the privacy and consent of the research participants and the access to and use of data should comply with the regulations of the People’s Republic of China on the administration of human genetic resources. Requests for access to data can be directed to Guanglin He.

### Credit authorship contribution statement

**Chao Liu, Renkuan Tang, Mengge Wang**, **Libing Yun**, **and Guanglin He**: Conceptualization, project administration, supervision, funding acquisition. **Mengge Wang**, **Guanglin He**: writing - original draft, review & editing Methodology, Data analysis, Investigation, Sampling, Review & Editing. **Qiuxia Sun and Yuntao Sun**: Writing - Review & editing. **Mengge Wang**, **Libing Yun, Qiuxia Sun, and Yuntao Sun:** Collected the samples, genomic DNA extraction and sequencing. **Chao Liu, Renkuan Tang, Mengge Wang**, **Libing Yun**, **and Guanglin He**: Supervision, review and revision of the manuscript.

## Acknowledgments

This study was supported by the National Natural Science Foundation of China (82402203 and 82202078), the Major Project of the National Social Science Foundation of China (23&ZD203), the Open Project of the Key Laboratory of Forensic Genetics of the Ministry of Public Security (2022FGKFKT05), the Center for Archaeological Science of Sichuan University (23SASA01), the 1.3.5 Project for Disciplines of Excellence, West China Hospital, Sichuan University (ZYJC20002), and the Sichuan Science and Technology Program (2024NSFSC1518).

