## Supplementary material for "Genomic insights into population stratification, biological adaptation, and archaic introgression at the crossroads of the Himalayas and lowland East Asia": Figure S

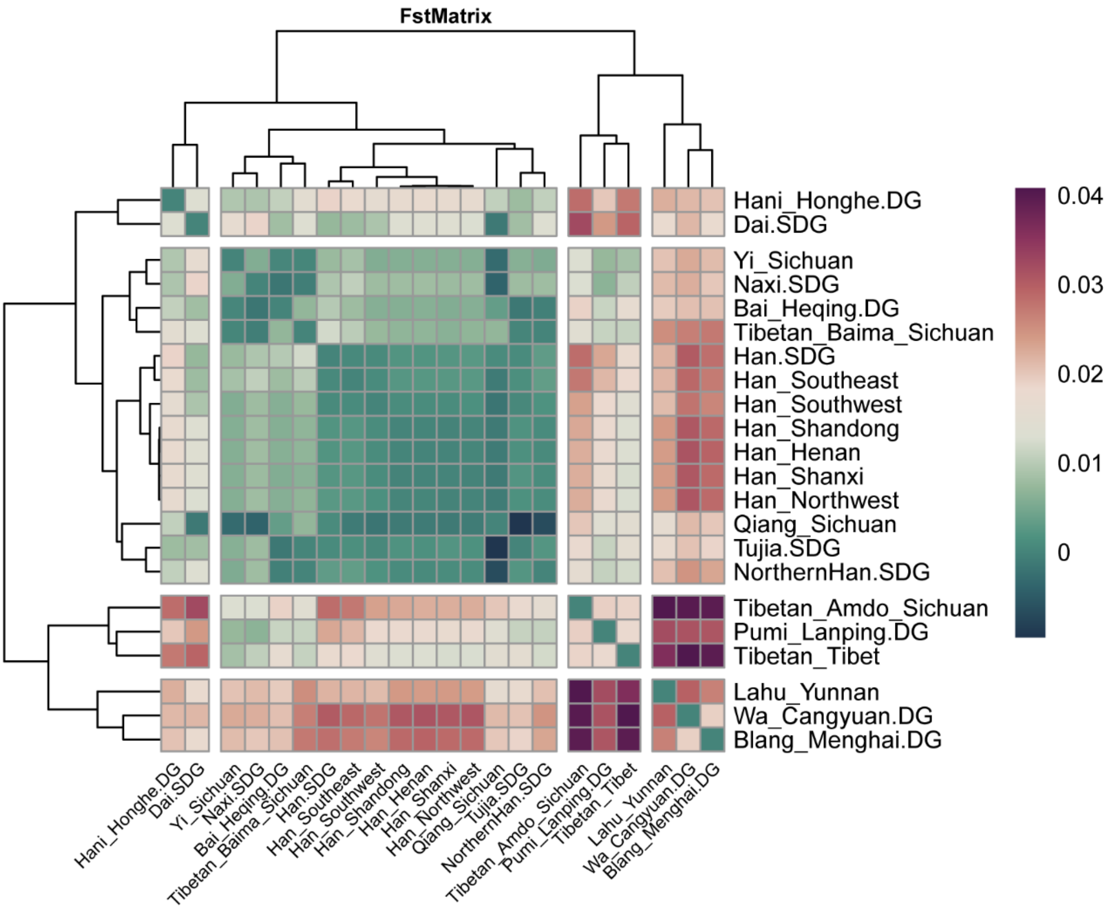


**Fig. S1. The genetic affinity of Sino-Tibetan people based on pairwise fixation index ($F_{ST}$) analysis.** In the context of East Asia, the $F_{ST}$ Matrix revealed that lighter shades of green correspond to lower $F_{ST}$ values, signifying a closer genetic affinity among genetically different TYC populations and Han Chinese from Shandong, Shanxi, and Henan.

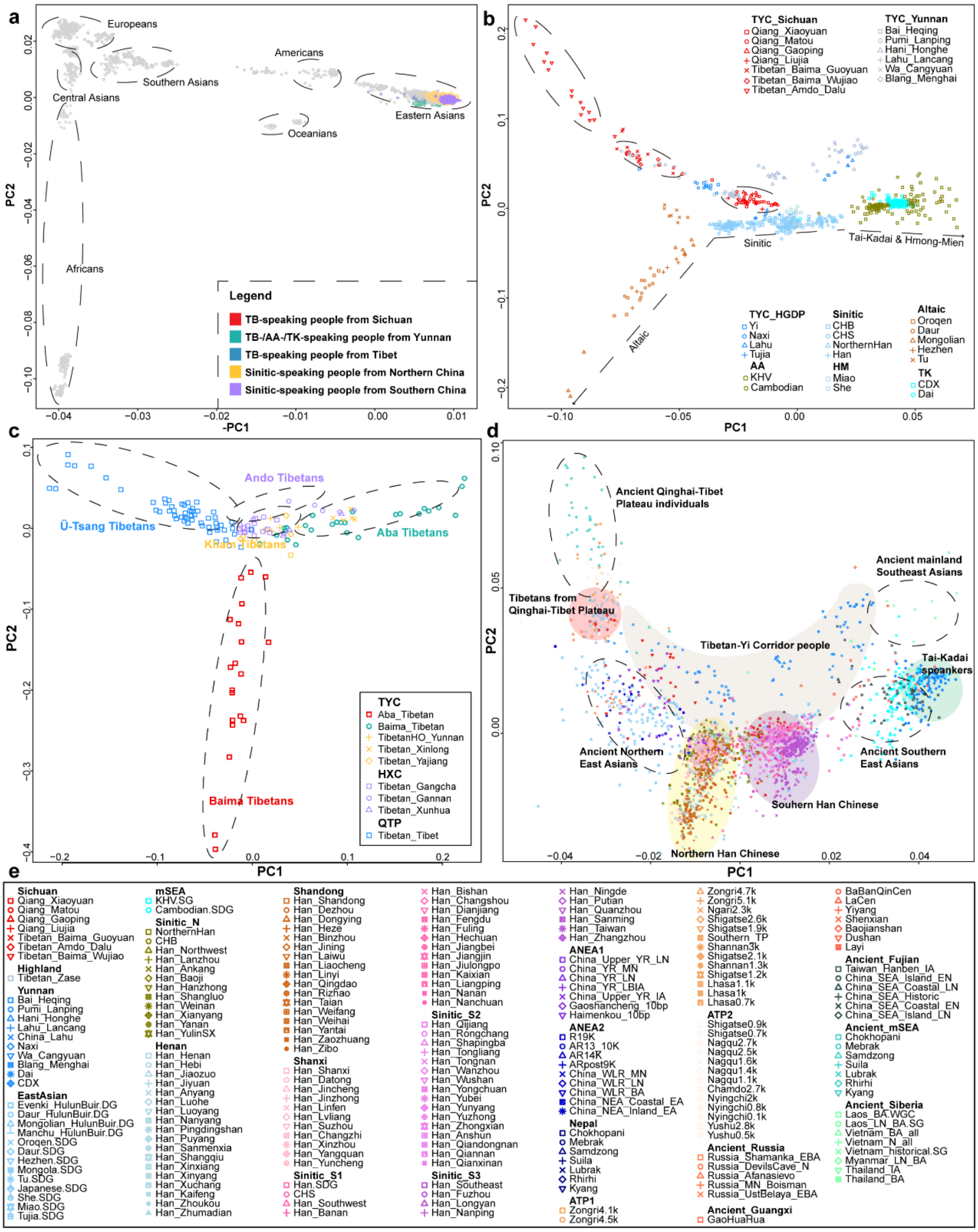


**Fig. S2. The genetic relationship between TYC populations and reference populations summarized by PCA.** (**a**) The PCA plot as defined by worldwide populations based on WGS(GRCh38) dataset, where grey shapes denoted reference populations, and colored shapes represented Tibeto-Burman, Sinitic, and Austroasiatic speakers. (**b**) Plot of the first two principal components in the context of East Asia was constructed utilizing WGS resources from TYC populations, HGDP, and 1KGP. (**c**) The fine-scale genetic profile of geographically/linguistically different Tibetans. (**d-e**) In the two-dimensional PC plot, modern/ancient East Asian populations and Southeast Asians are projected and demonstrate the clustering patterns among geographically/linguistically diverse East Asian populations.

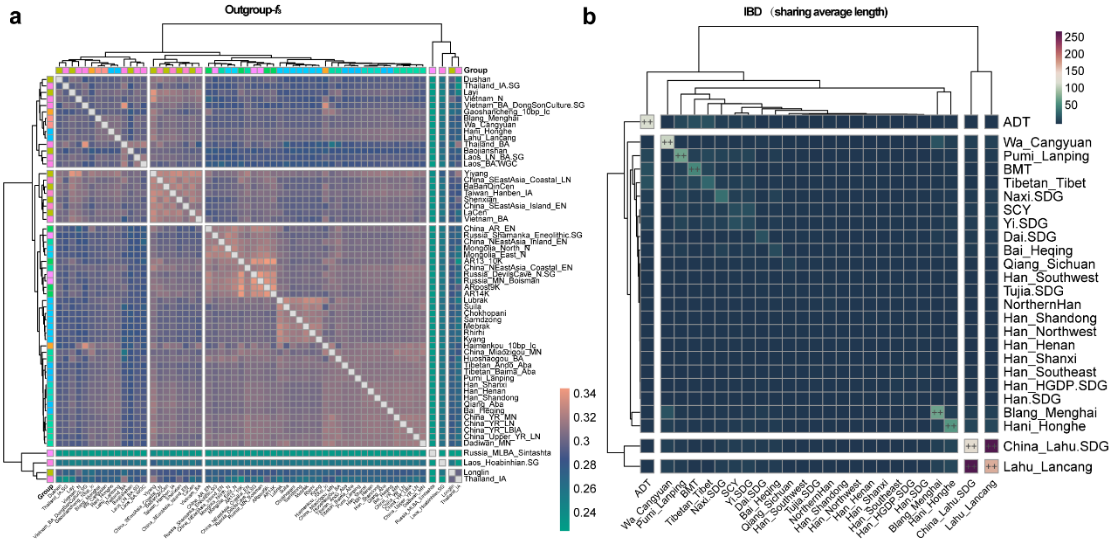


**Fig. S3. Genetic affinity between TYC populations and modern/ancient Chinese populations.** (**a**) Outgroup $f_3$ analysis for $f_3$(X, Y; Mbuti) on the basis of shared genetic drift, where Mbuti is a Central African modern population, and X and Y are TYC subgroups and ancient East Asians. A deeper shade of red in the visualization indicated higher genetic similarity. (**b**) The haplotyped-based IBD analysis revealed longer shared genomic segments between populations, which corresponded to a closer genetic relatedness.

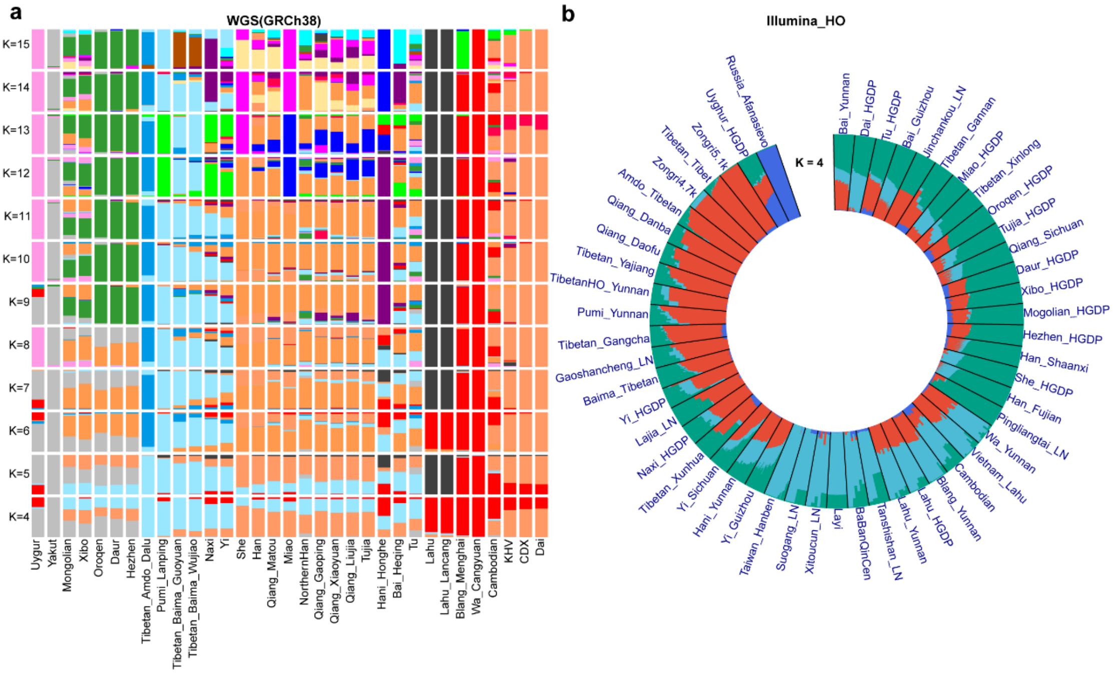


**Fig. S4. The ancestry makeup of TYC populations.** (**a**) ADMIXTURE analysis showed the Wa, Hani, Lahu, ADT, and Pumi-specific ancestral component dominant in other TYC people as ancestral populations (K) increased when utilizing the high-resolution WGS(GRCh38) dataset. (**b**) ADMIXTURE results for K = 4 clusters were shown for 52 ancient and contemporary East Asian populations based on Illumina_HO dataset.

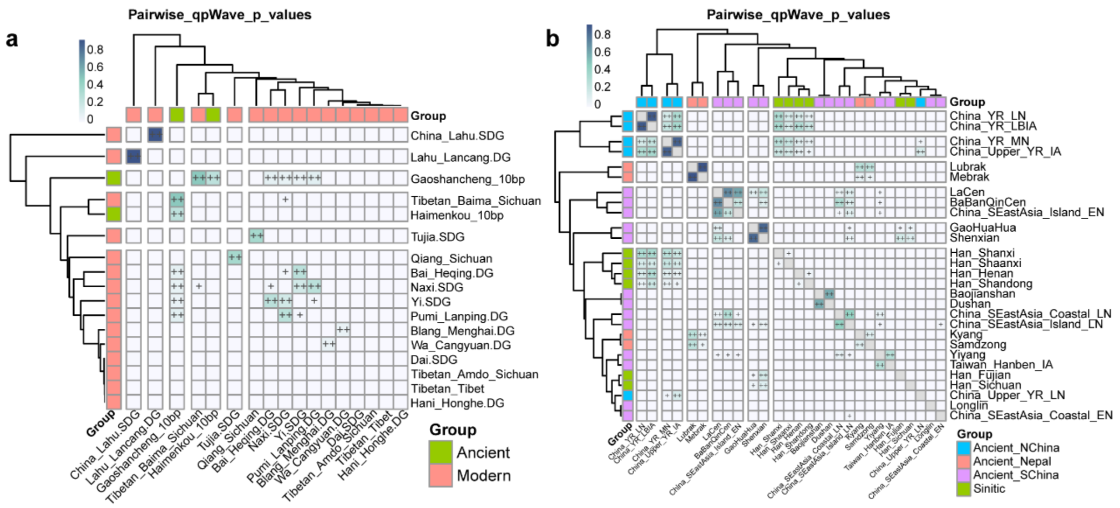

**Fig. S5. Paired qpWave matrix.** (**a**) qpWave modeling to estimate genetically homogeneous between TYC populations and Gaoshan individuals and outgroup included Mbuti, Russia_Afanasievo, Papuan, Japan_Jomon, China_AR_EN, Longlin, China_NEastAsia_Coastal_EN, China_SEastAsia_Island_EN, and Laos_Hoabinhian. (**b**) qpWave analysis was performed to determine the genetic homogeneity among Sinitic people and millet-related farmers from Yellow River Basin, with outgroup including Mbuti, Papuan, Ami, Russia_Ust_Ishim, Onge, Mixe, Ttaly_North_Villabruna, Iran_GanjDareh, and Russia_Kostenki14. ++ indicates p >0.05, suggesting no statistical significance, and the two populations are considered genetically homogeneous. Populations are divided into different groups, represented by distinct colors.

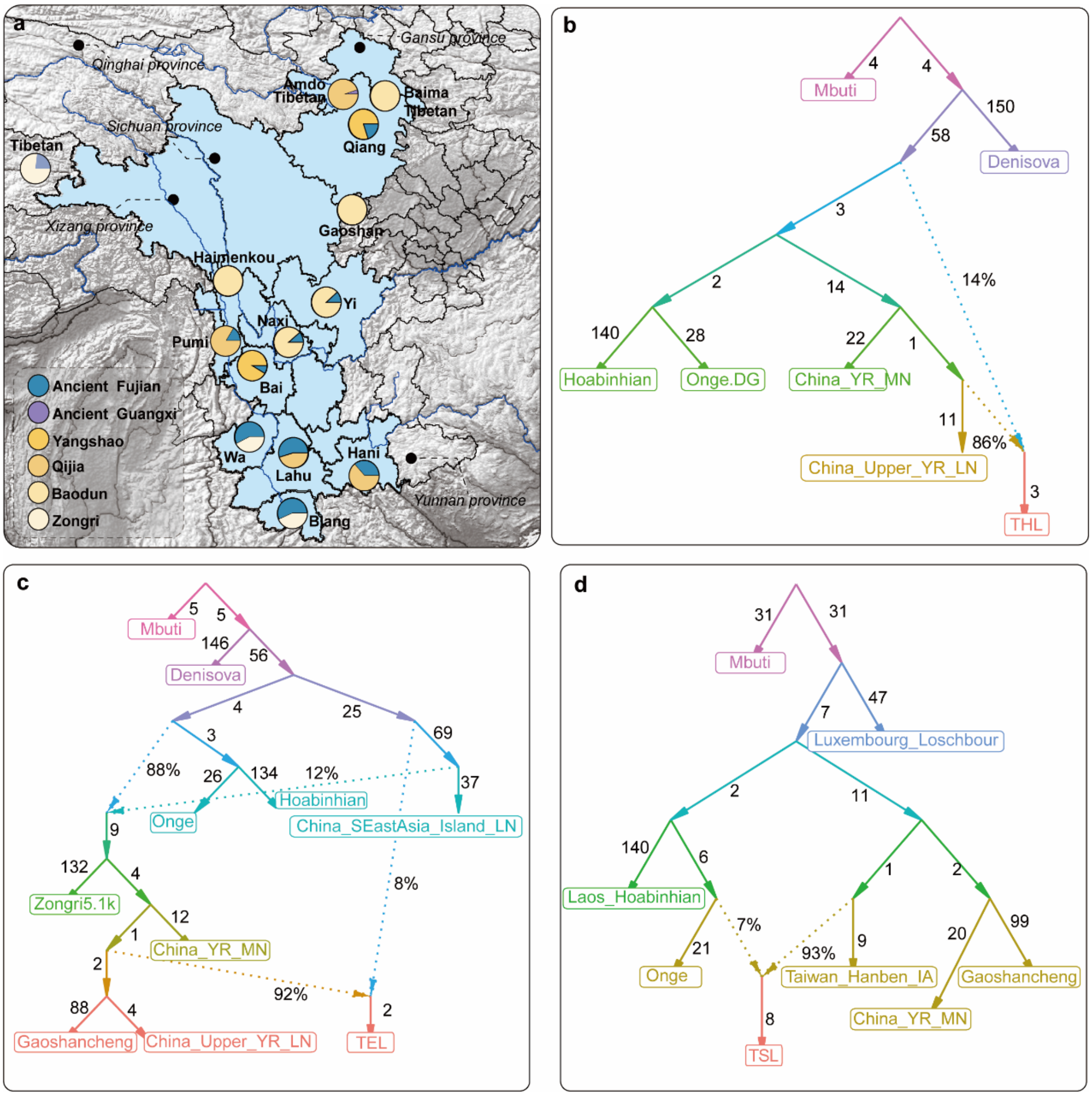


**Fig. S6. Admixture model via qpAdm and qpGraph.** (**a**) Two-way admixture models for genetically different TYC populations, with outgroup including Mbuti, Papuan, Ami, Russia_Ust_Ishim, Onge, Mixe, Ttaly_North_Villabruna, Iran_GanjDareh, and Russia_Kostenki14. Yellow represented millet-related farmers from Yellow River Basin, including individuals affiliated with the Yangshao, Qijia, and Baodun cultures. Bule denoted ancient individuals from Fujian, while purple was used for ancient samples from Guangxi. (**b-d**) qpGraph models of THL, TEL, and TSL. The estimated genetic drift for each branch is provided, and dashed lines indicate the admixture events with the estimated mixture proportions.

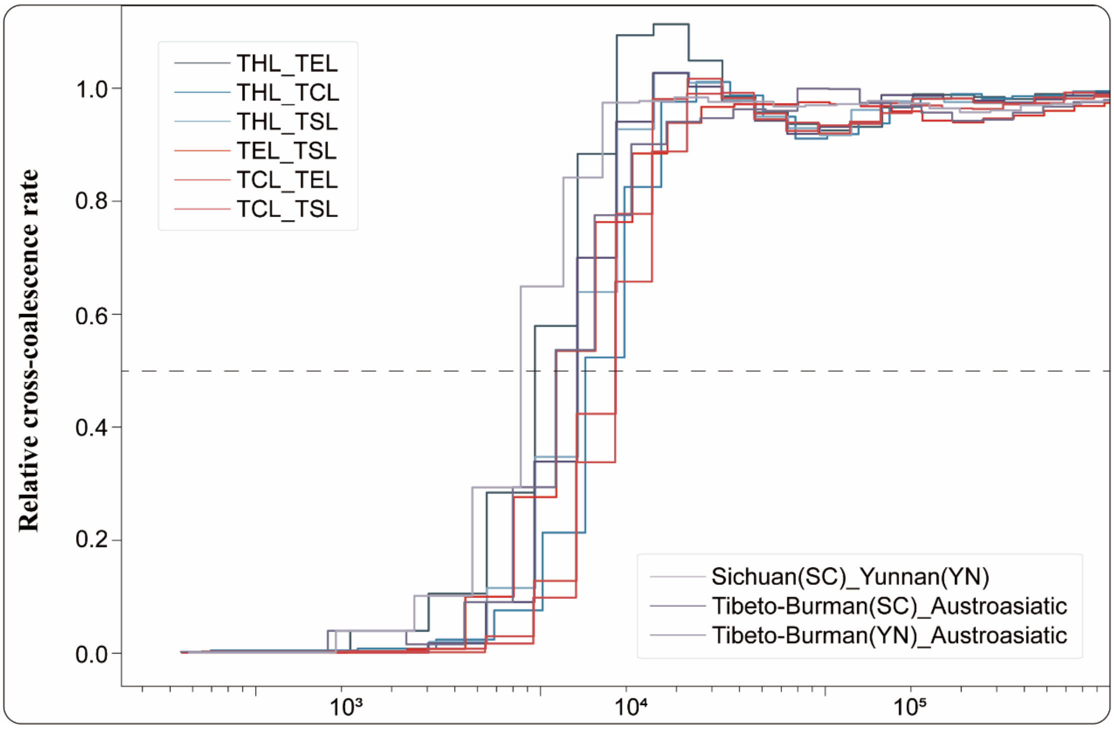


**Fig. S7. Population separations of genetically distinct TYC populations.** MSMC2 cross-population results for pairs among THL, TSL, TCL, and TEL, assuming 25 years per generation and a mutation rate = $1.25\times10^{-8}$.

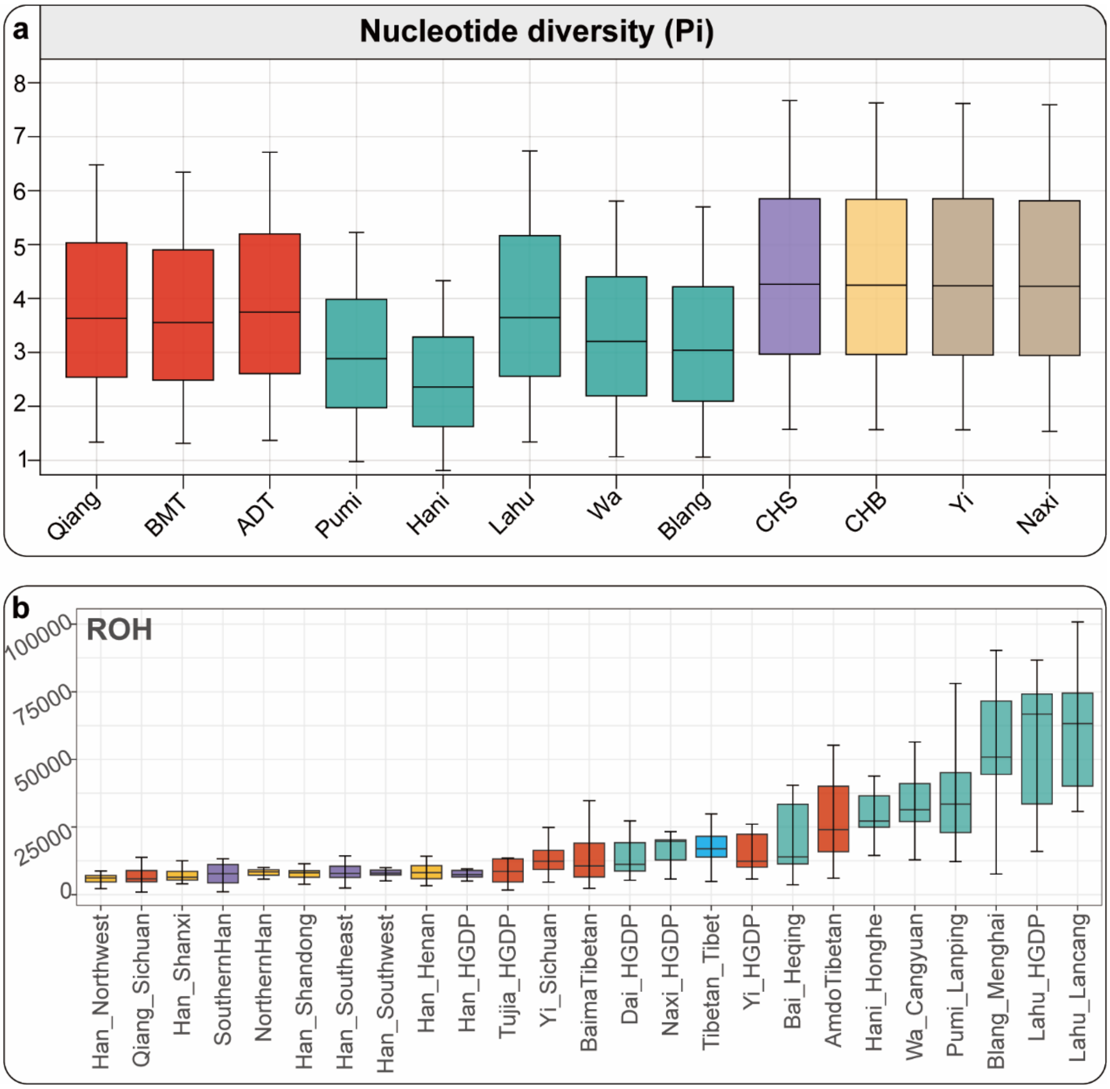


**Fig. S8. Nucleotide diversity and runs of homozygosity.** Isolated populations such as ADT, Pumi, Lahu, Wa, and Blang exhibited a modest growth rate while maintaining relatively high proportions of their genomes in runs of homozygosity.

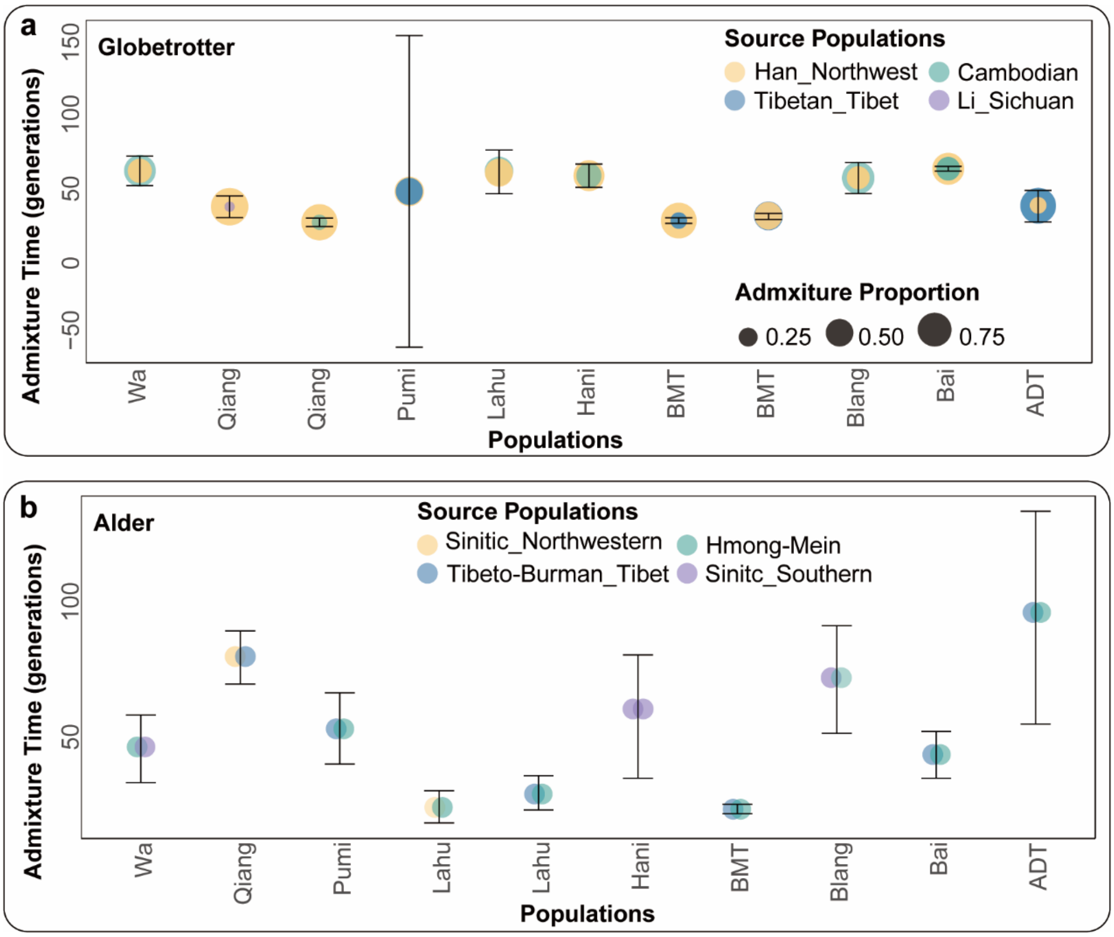


**Fig. S9. Admixture history of TYC populations.** (**a**) GLOBETROTTER analysis results were based on haplotype data that showed admixture events, times, ancestral sources, and proportions for the TYC population. Circle colors denote proxy ancestral sources, while circle sizes reflect admixture proportions. Admixture times are presented as the mean ± standard deviation, derived from 100 iterations of the GLOBETROTTER analysis. (**b**) Admixture events from allele frequency-based MALDER were performed and circle colors denote proxy ancestral sources.

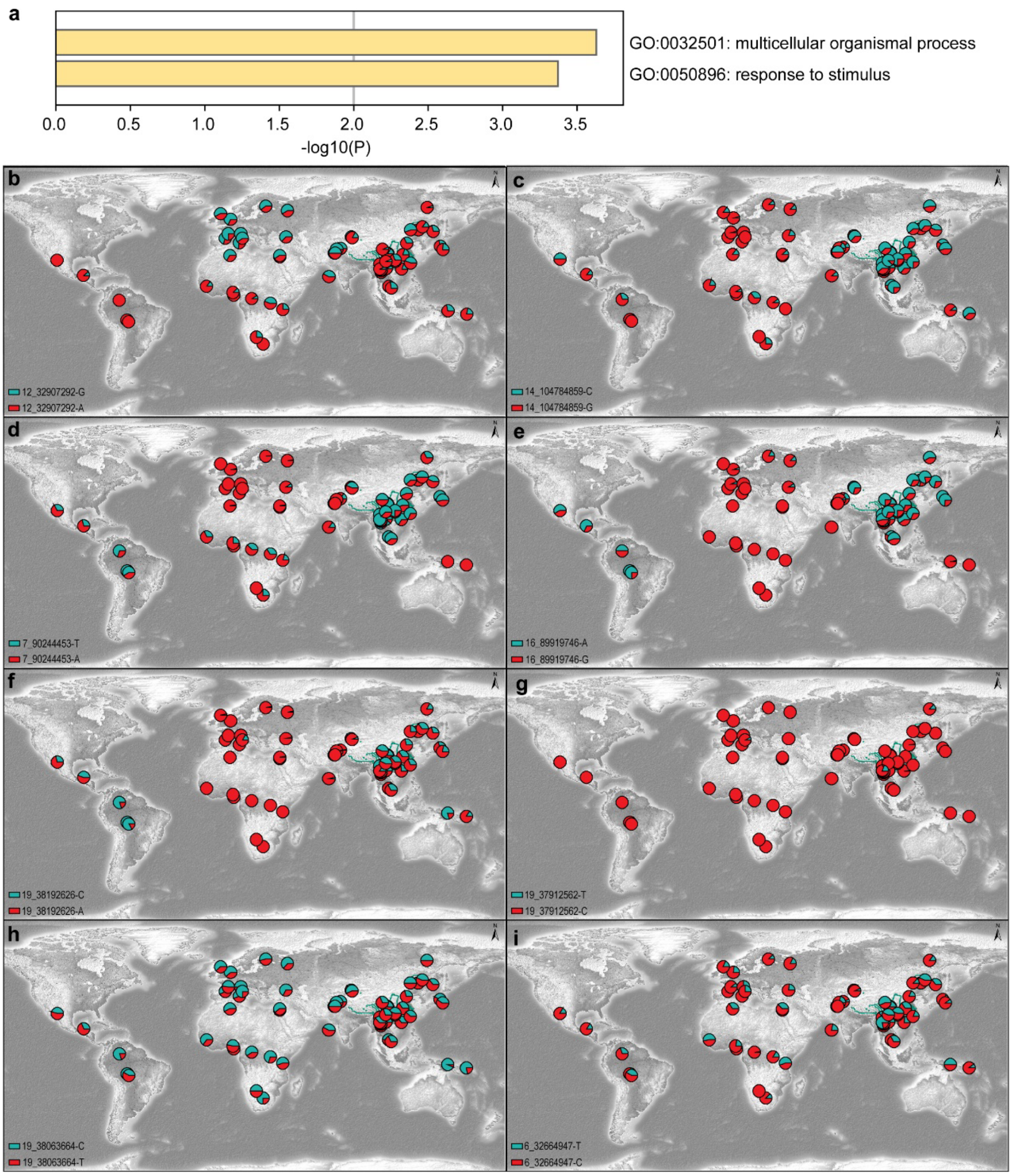


**Fig. S10. The shared and specific selection signatures in TYC populations.** (**a**) Gene Ontology pathway analysis revealed that these putatively population-shared genes were associated with relevant pathways related to adaptive physiological traits in TYC individuals. (**b-i**) Geographic distribution of selective signals in TYC populations based on 10K_CPGDP, HGDP, and 1KGP resources.

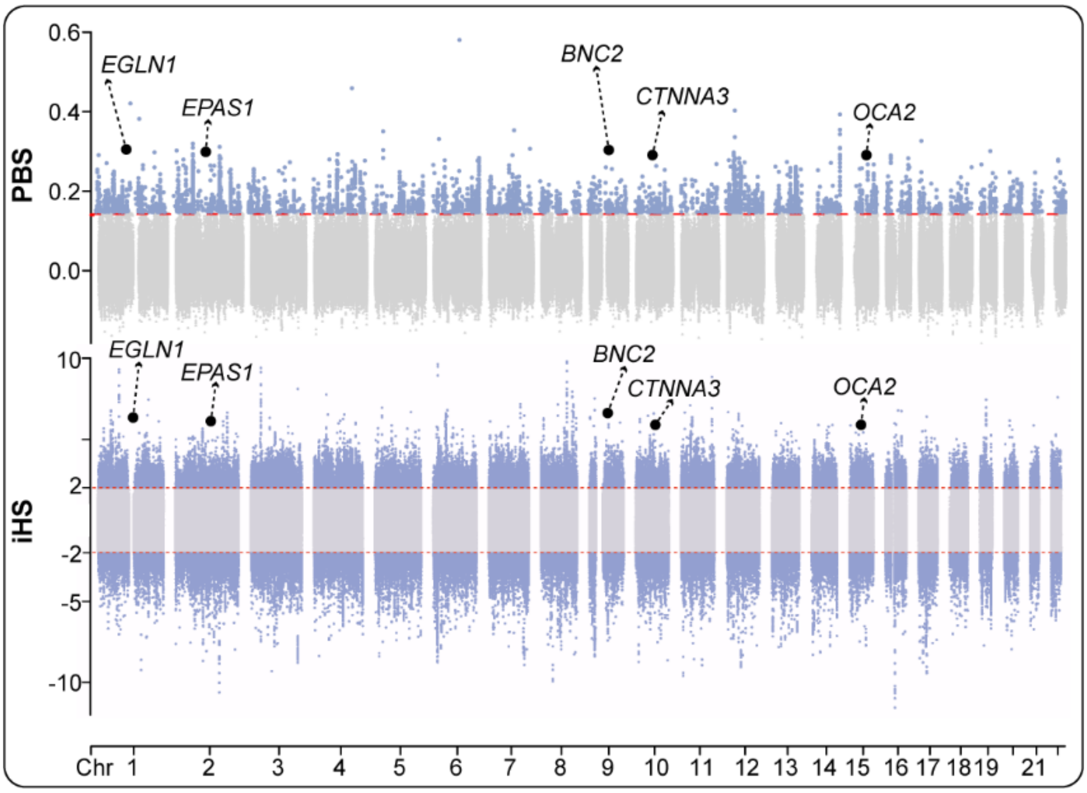


**Fig. S11. Manhattan plots for the Population Branch Statistic (PBS) and integrated haplotype homozygosity score (iHS) in THL.** We divided the whole genome into 20 kb windows, and windows with a |iHS| value > 2 were treated as candidate SNPs and windows with more than 20% candidate SNPs were selected as adaptive regions.

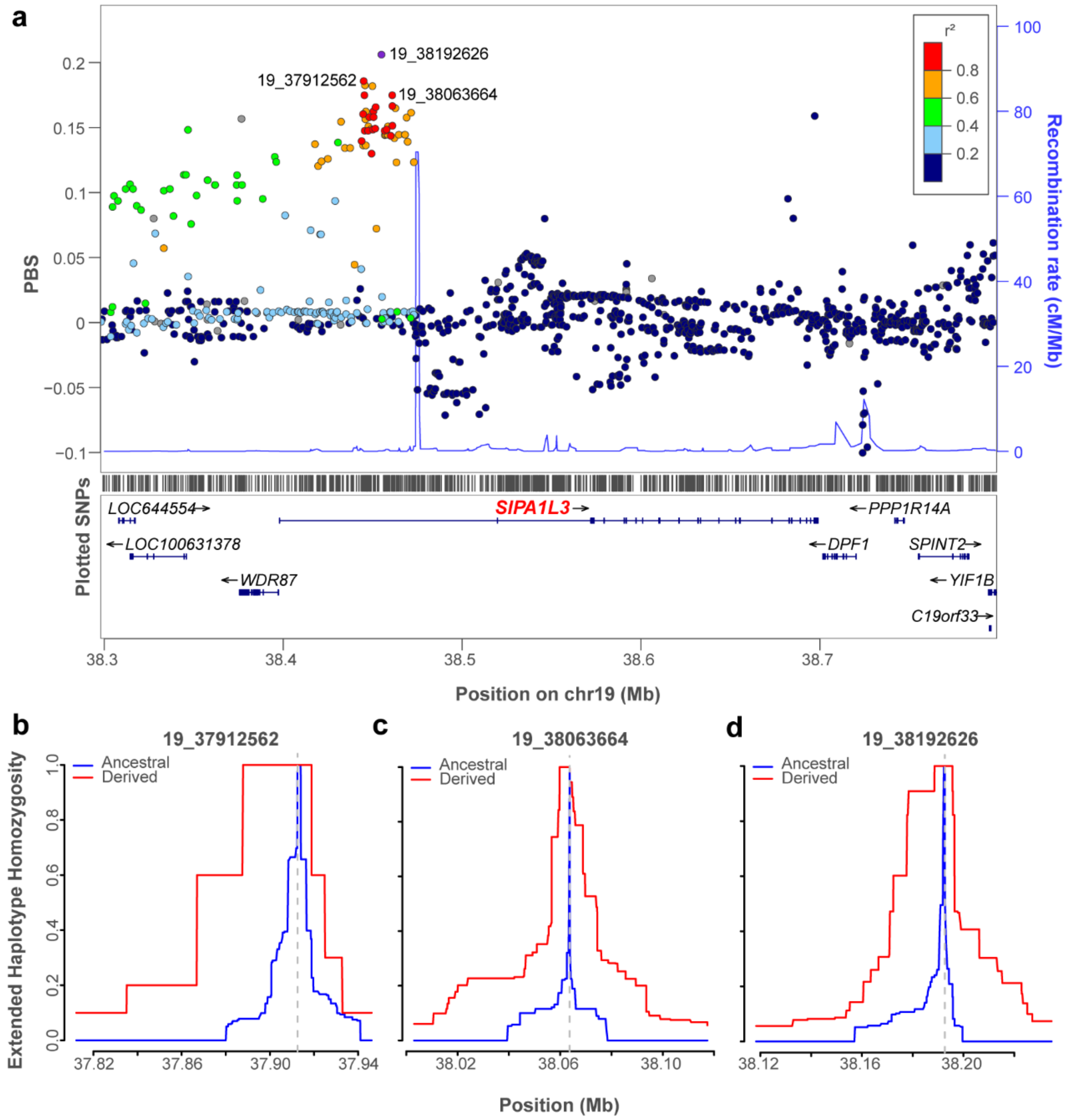


**Fig. S12. Selection signatures at *SIPA1L3* locus.** (**a**) The analysis utilizing LocusZoom revealed significant variants within *SIPA1L3*, specifically at positions 19_37912562, 19_38192626, and 19_38063664. These variants were consistently detected using both PBS and iHS statistical methods. (**b**) Extended haplotype homozygosity (EHH) of the target SNP, with red indicating the derived allele and blue indicating the ancestral allele.

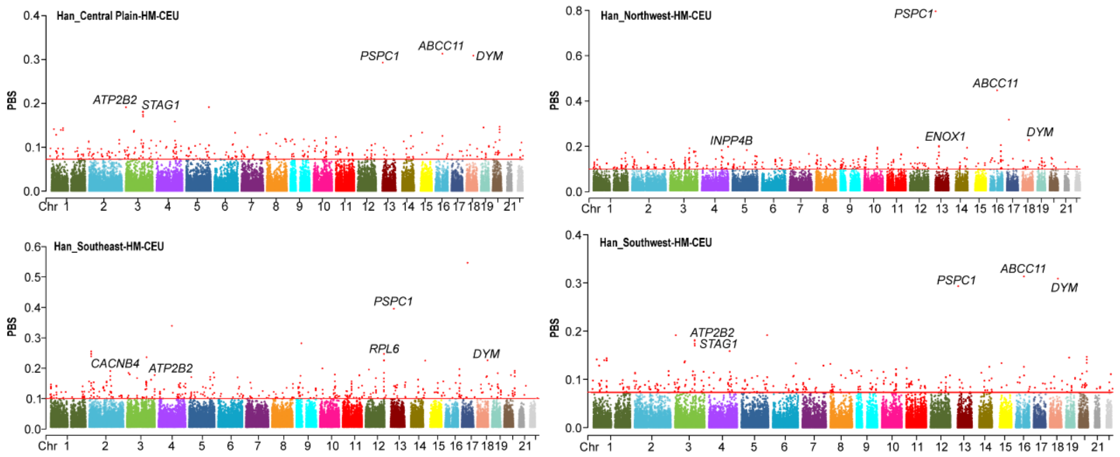


**Fig. S13. Manhattan plots for the Population Branch Statistic (PBS) in geographically different Sinitic people.** PBS was also conducted on Sinitic people to detect selection signals via the Illumina_HGDP dataset, using Miao_Guizhou and EUR serving as ingroup and outgroup, respectively. The SNPs with extreme branch lengths (top 0.1%) were regarded as strong candidates for the genetic basis of adaptive evolution.

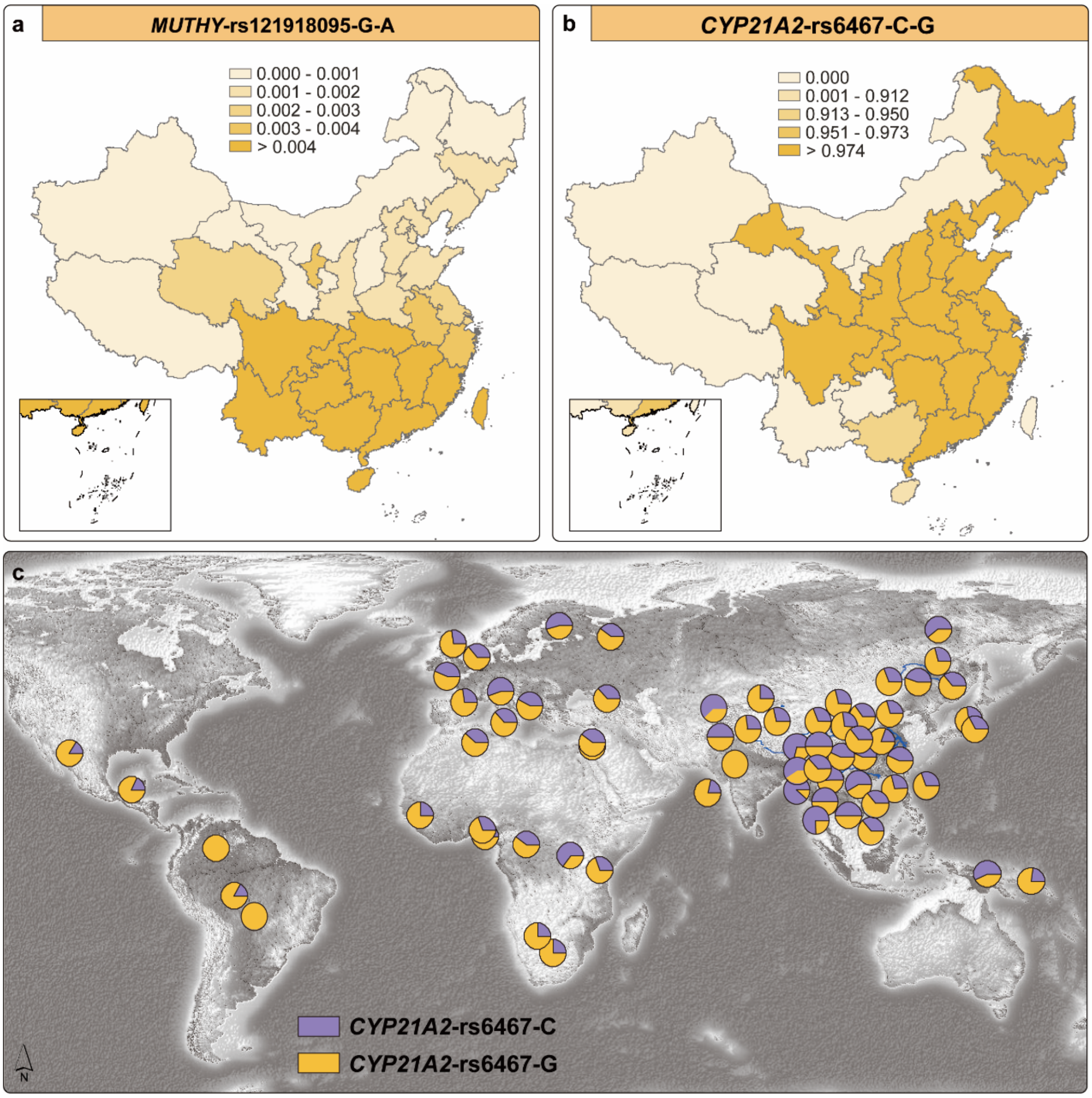


**Fig. S14. Geographic distribution of ACMG reportable and medically relevant variants.** (**a**) The doubleton variant *MUTYH*-rs121918095-G-A, associated with recessively inherited polyposis. Showed high frequency in southern China based on Huaxi Biobank. (**b-c**) Frequency distribution plot of rs6467, which annotated as pathogenic or likely pathogenic in ClinVar Database (v.202003).

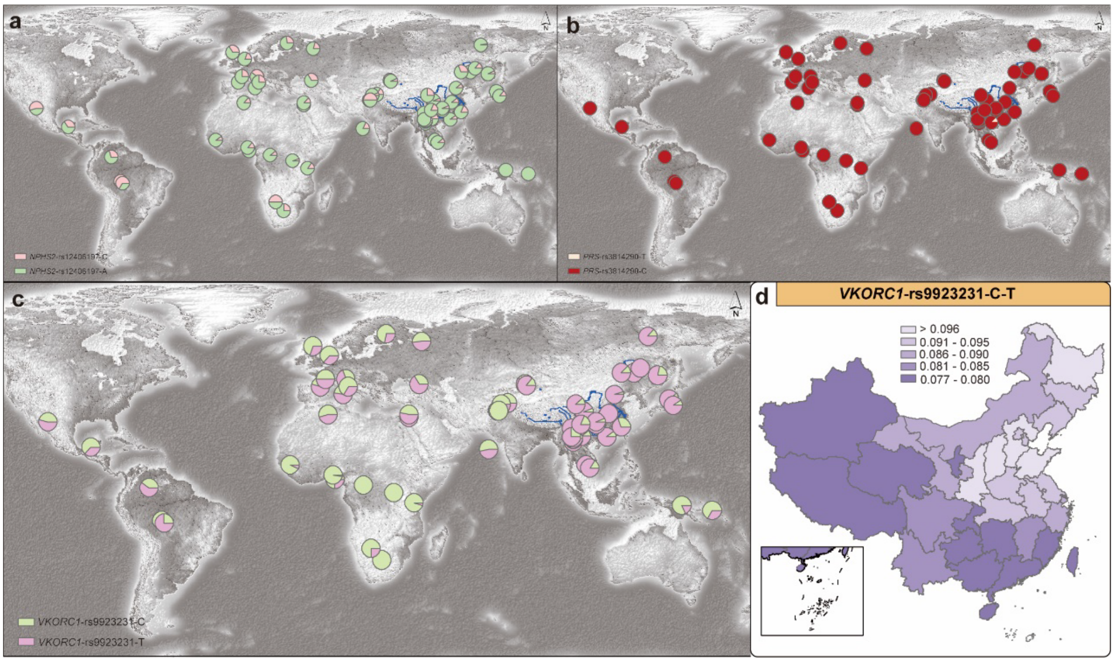


**Fig. S15. Frequency distribution plot of medically relevant variants.** The frequency distribution patterns of *NPHS2*-rs12406197, *PRS*-rs3814290, and *VKORC1*-rs9923231 were plotted, with frequency data obtained from 10K_CPGDP, HGDP, and 1KGP.

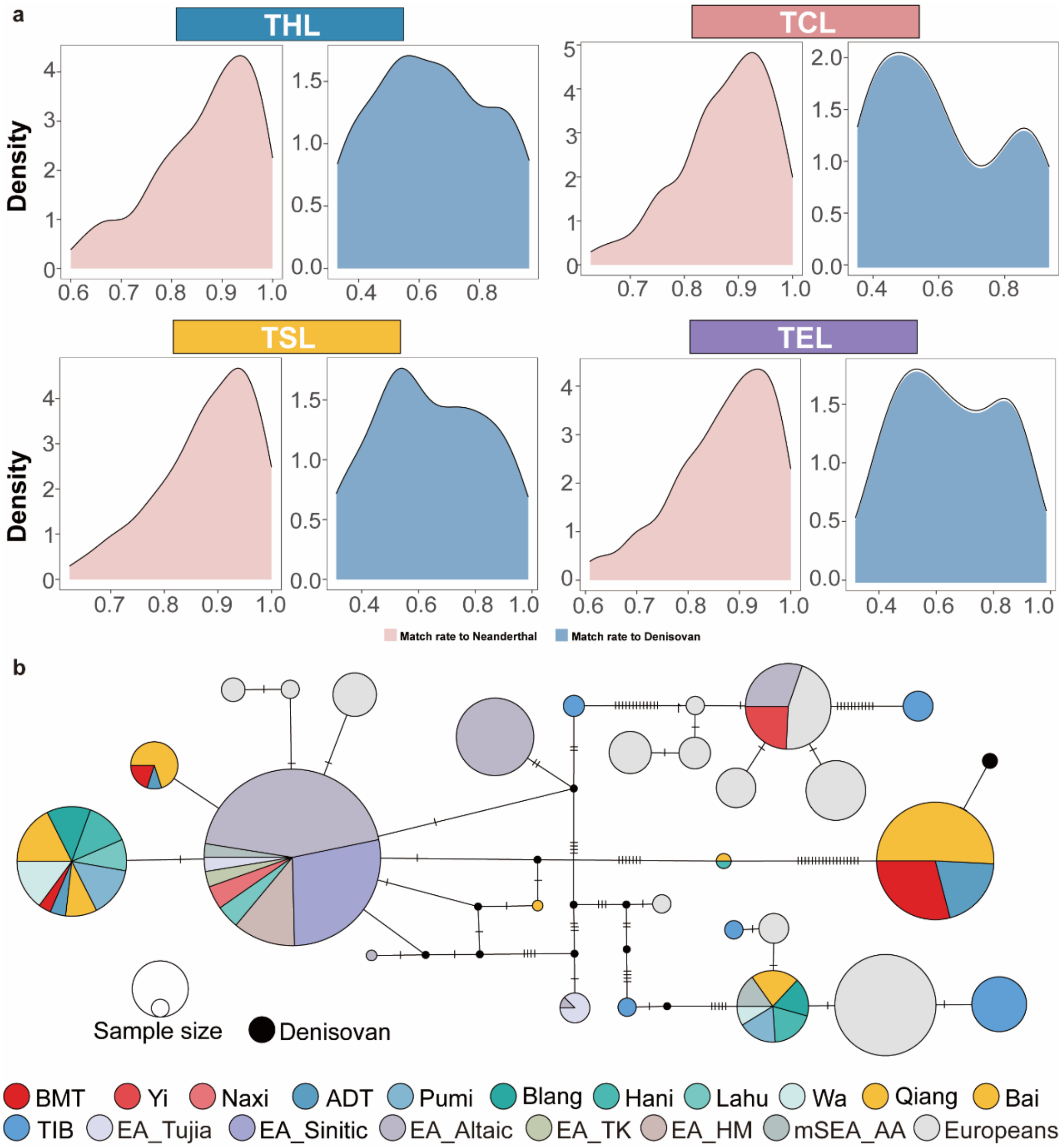


**Fig. S16. Match rates and network of introgressed haplotypes.** (**a**) Fitted density curves for populations with significant unimodal match rate to Neanderthal (red) and bimodal match rate distributions to Denisovan (blue) based on high-confidence archaic haplotypes. (**b**) Median-joining haplotype network constructed within the 32.7 kb *EPAS1* introgressed from Denisovan based on PopArt.

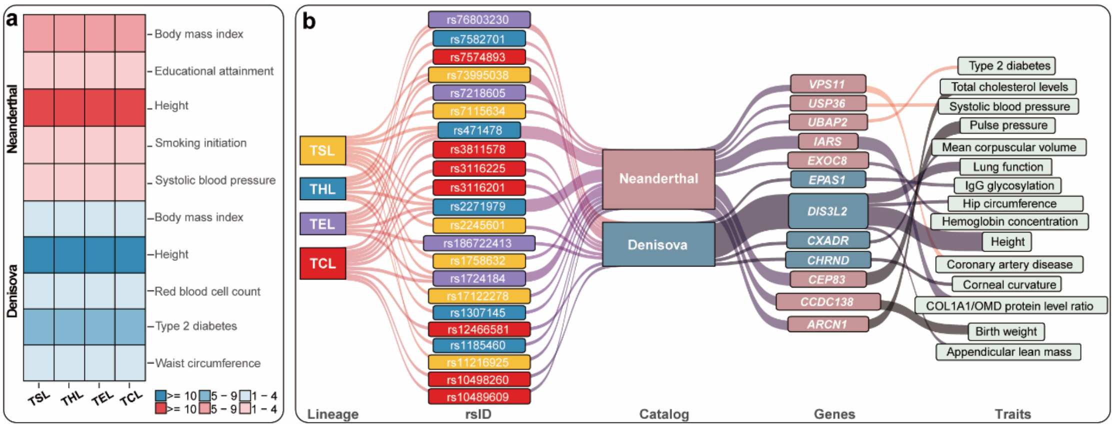


**Fig. S17. The potential phenotypic effects of the identified introgressed sequences from Neanderthal and Denisovans.** (**a**) Heatmap depicting the number of signals derived from high-confidence archaic haplotypes, with annotations informed by the GWAS Catalog. (**b**) Sankey plot of communication from archaic-introgressed signals to traits.
