## Supplementary material for "Genomic insights into population stratification, biological adaptation, and archaic introgression at the crossroads of the Himalayas and lowland East Asia": Table S

**Table S1. The coverage depth of each individual**

| Sample_ID | Depth(x) | Coverage | Group |
|---|---|---|---|
| 10181809195884 | 0.910088 | 8.92263 | Qiang_Xiaoyuan |
| 11641809199794 | 0.914041 | 14.6071 | Tibetan_Baima_Guoyuan |
| 12081809195141 | 0.914303 | 14.2382 | Qiang_Xiaoyuan |
| 13011809195733 | 0.91419 | 19.7621 | Tibetan_Amdo_Dalu |
| 13431809291301 | 0.921036 | 14.5653 | Qiang_Matou |
| 13751811580260 | 0.9142 | 17.9567 | Qiang_Gaoping |
| 14481809290742 | 0.913217 | 10.4175 | Qiang_Gaoping |
| 15781809295696 | NA | NA | Qiang_Gaoping |
| 16421809291286 | 0.919504 | 11.1773 | Qiang_Gaoping |
| 16601809195885 | 0.920039 | 14.2363 | Tibetan_Baima_Wujiao |
| 20161809295225 | 0.920165 | 15.5357 | Qiang_Gaoping |
| 21021809195908 | 0.920681 | 17.7281 | Tibetan_Baima_Wujiao |
| 23721809195902 | 0.913382 | 15.0701 | Tibetan_Baima_Guoyuan |
| 24231809195543 | 0.919449 | 10.9388 | Qiang_Xiaoyuan |
| 25141810065244 | 0.92014 | 15.0233 | Qiang_Liujia |
| 25591809295865 | 0.913459 | 16.0131 | Qiang_Liujia |
| 26581809291226 | 0.914086 | 15.3364 | Tibetan_Baima_Guoyuan |
| 26961809198943 | 0.919903 | 12.7735 | Tibetan_Baima_Wujiao |
| 27621810065314 | 0.919724 | 13.5327 | Qiang_Liujia |
| 28171809195881 | 0.921047 | 16.2647 | Qiang_Matou |
| 28731809183293 | 0.913973 | 14.2538 | Qiang_Matou |
| 28971811580252 | 0.913276 | 13.6993 | Qiang_Gaoping |
| 29711810065856 | 0.920287 | 14.9573 | Tibetan_Amdo_Dalu |
| 32001809291341 | 0.913725 | 14.6386 | Qiang_Gaoping |
| 33141809291307 | 0.9141 | 14.8037 | Qiang_Liujia |
| 36311809291231 | 0.914202 | 15.8612 | Qiang_Matou |
| 36581809291228 | 0.913993 | 14.2624 | Tibetan_Amdo_Dalu |
| 40921809195525 | 0.920076 | 14.2381 | Tibetan_Baima_Guoyuan |
| 41521809183261 | 0.919721 | 14.6137 | Qiang_Gaoping |
| 42211809195255 | 0.9139 | 15.6474 | Tibetan_Baima_Guoyuan |
| 43191809198958 | 0.9133 | 11.4343 | Qiang_Matou |
| 43901809195726 | 0.913574 | 16.2517 | Tibetan_Baima_Guoyuan |
| 44141809290867 | 0.920295 | 15.6674 | Qiang_Liujia |
| 44321809291222 | 0.920017 | 16.3024 | Tibetan_Amdo_Dalu |
| 45561810065173 | 0.919658 | 12.7603 | Qiang_Liujia |
| 45851809199888 | 0.913388 | 15.0854 | Tibetan_Amdo_Dalu |
| 46201809195887 | 0.919739 | 13.4335 | Qiang_Matou |
| 46961809199798 | 0.913675 | 14.4949 | Tibetan_Baima_Wujiao |
| 47341809195883 | 0.913394 | 15.0248 | Qiang_Xiaoyuan |
| 48391809195882 | 0.913832 | 15.4169 | Qiang_Matou |
| 48441809195924 | 0.913328 | 14.9169 | Qiang_Matou |
| 52411809195742 | NA | NA | Qiang_Matou |
| 53331809291198 | NA | NA | Tibetan_Amdo_Dalu |
| 53691809195893 | 0.915619 | 8.62068 | Tibetan_Amdo_Dalu |
| 56651811583022 | NA | NA | Qiang_Gaoping |
| 56871810066170 | NA | NA | Tibetan_Baima_Wujiao |
| 58891809199037 | NA | NA | Tibetan_Amdo_Dalu |
| 59211809198946 | NA | NA | Tibetan_Baima_Guoyuan |
| 60021809195476 | NA | NA | Tibetan_Baima_Guoyuan |
| 60621809195693 | NA | NA | Tibetan_Baima_Guoyuan |
| 61531809199905 | 0.91288 | 11.7487 | Qiang_Matou |
| 62541810066150 | 0.920633 | 17.2014 | Tibetan_Baima_Guoyuan |
| 62661809195909 | 0.920864 | 10.8569 | Tibetan_Baima_Wujiao |
| 62861809195514 | 0.920745 | 11.5396 | Tibetan_Baima_Guoyuan |
| 62861809295598 | 0.921737 | 14.451 | Qiang_Liujia |
| 64651811580012 | 0.914388 | 13.5815 | Qiang_Matou |
| 66681809291160 | 0.920815 | 16.0951 | Qiang_Matou |

| | | | |
|---|---|---|---|
| 67061809290952 | 0.921709 | 15.4484 | Qiang_Xiaoyuan |
| 67941809291241 | 0.921383 | 15.4091 | Qiang_Matou |
| 69861809291332 | 0.91477 | 13.3266 | Qiang_Gaoping |
| 70201809195745 | 0.912682 | 8.68695 | Tibetan_Baima_Wujiao |
| 70311809195932 | 0.921415 | 13.7171 | Tibetan_Amdo_Dalu |
| 72981809291313 | 0.914788 | 13.778 | Qiang_Gaoping |
| 73131809291254 | 0.914309 | 10.7829 | Qiang_Matou |
| 73151809195743 | 0.92054 | 18.0231 | Qiang_Xiaoyuan |
| 73821809195746 | 0.914586 | 12.3782 | Qiang_Xiaoyuan |
| 79291809190102 | 0.921278 | 13.0958 | Tibetan_Baima_Wujiao |
| 80181809190197 | 0.921355 | 14.0149 | Tibetan_Baima_Guoyuan |
| 81351809190272 | 0.914726 | 15.3067 | Tibetan_Amdo_Dalu |
| 81451809190398 | 0.914968 | 14.1251 | Tibetan_Amdo_Dalu |
| 84171809190317 | 0.921186 | 13.3123 | Tibetan_Amdo_Dalu |
| 85961809190477 | 0.921197 | 13.0389 | Tibetan_Amdo_Dalu |
| 86801809190302 | 0.921381 | 14.5014 | Tibetan_Amdo_Dalu |
| 87911809190418 | 0.920799 | 11.4624 | Qiang_Xiaoyuan |
| 88021809190224 | 0.921604 | 15.5468 | Qiang_Matou |
| 88661809190032 | 0.920683 | 12.6237 | Qiang_Xiaoyuan |
| 89031809190043 | NA | NA | Tibetan_Amdo_Dalu |
| 89181809190148 | NA | NA | Qiang_Xiaoyuan |
| 91851809190088 | NA | NA | Qiang_Xiaoyuan |
| 98581809190431 | 0.921021 | 12.0489 | Tibetan_Baima_Guoyuan |
| 98711809190466 | 0.92144 | 14.5821 | Tibetan_Baima_Guoyuan |
| 99761809190256 | 0.914864 | 13.9927 | Tibetan_Amdo_Dalu |
| 2W011709240419 | 0.91657 | 9.82487 | Wa_Cangyuan |
| 2W021709222193 | 0.917853 | 10.5711 | Hani_Honghe |
| 2W031808160321 | 0.917969 | 10.4021 | Pumi_Lanping |
| 2W051709224252 | 0.915807 | 9.25587 | Blang_Menghai |
| 2W051709229263 | 0.918466 | 12.4021 | Blang_Menghai |
| 2W081709222177 | 0.919029 | 13.2982 | Wa_Cangyuan |
| 2W111808160083 | 0.918217 | 11.486 | Bai_Heqing |
| 2W111808212730 | 0.918323 | 10.7603 | Pumi_Lanping |
| 2W121808165387 | 0.919023 | 12.1404 | Bai_Heqing |
| 2W151709221358 | 0.91732 | 9.80761 | Hani_Honghe |
| 2W161709222468 | 0.906188 | 5.75747 | Hani_Honghe |
| 2W161808197987 | 0.919262 | 13.5929 | Bai_Heqing |
| 2W191709240487 | 0.917204 | 9.82858 | Wa_Cangyuan |
| 2W191808160145 | 0.918927 | 14.5118 | Bai_Heqing |
| 2W201709241831 | 0.917739 | 10.2549 | Wa_Cangyuan |
| 2W211709229083 | 0.919358 | 14.0186 | Lahu_Lancang |
| 2W241808160133 | 0.909399 | 6.07159 | Pumi_Lanping |
| 2W261709240049 | 0.915624 | 9.40749 | Wa_Cangyuan |
| 2W261709241932 | 0.918102 | 11.2656 | Wa_Cangyuan |
| 2W281808198052 | 0.917259 | 10.6237 | Bai_Heqing |
| 2W301808190197 | 0.918004 | 11.1991 | Bai_Heqing |
| 2W311808197463 | 0.918268 | 11.6681 | Pumi_Lanping |
| 2W341709229143 | 0.918084 | 11.2481 | Lahu_Lancang |
| 2W351709241811 | 0.915661 | 9.72182 | Blang_Menghai |
| 2W351808160131 | 0.915707 | 9.24019 | Pumi_Lanping |
| 2W391709221278 | 0.917777 | 10.1009 | Hani_Honghe |
| 2W391709241701 | 0.916042 | 9.57231 | Blang_Menghai |
| 2W391808190054 | 0.919004 | 12.6057 | Bai_Heqing |
| 2W401709222976 | 0.919163 | 14.4242 | Blang_Menghai |
| 2W421709240105 | 0.917438 | 10.238 | Blang_Menghai |
| 2W441709229267 | 0.917659 | 11.2006 | Blang_Menghai |
| 2W451808160032 | 0.916312 | 9.10889 | Bai_Heqing |
| 2W501709222338 | 0.913405 | 7.45449 | Hani_Honghe |
| 2W521709229257 | 0.919479 | 14.5697 | Lahu_Lancang |

| | | | |
|---|---|---|---|
| 2W541709222293 | 0.915068 | 8.88591 | Hani_Honghe |
| 2W541808211818 | 0.91646 | 9.12067 | Pumi_Lanping |
| 2W561709221482 | 0.919362 | 13.9373 | Wa_Cangyuan |
| 2W601709240113 | 0.916382 | 9.09759 | Hani_Honghe |
| 2W601808160154 | 0.920015 | 15.7225 | Pumi_Lanping |
| 2W601808195980 | 0.918361 | 12.1147 | Pumi_Lanping |
| 2W621709241844 | 0.917242 | 10.134 | Lahu_Lancang |
| 2W621808190049 | 0.918198 | 11.9267 | Bai_Heqing |
| 2W641709240440 | 0.917987 | 11.9689 | Wa_Cangyuan |
| 2W651709241968 | 0.917659 | 10.9556 | Lahu_Lancang |
| 2W661808190064 | 0.918511 | 11.7591 | Bai_Heqing |
| 2W691709221736 | 0.892983 | 4.46006 | Hani_Honghe |
| 2W691709222283 | 0.919681 | 14.7332 | Hani_Honghe |
| 2W761808198132 | 0.917497 | 10.3561 | Bai_Heqing |
| 2W771709222127 | 0.917502 | 10.3196 | Wa_Cangyuan |
| 2W791709222232 | 0.917341 | 10.0976 | Hani_Honghe |
| 2W791709241491 | 0.919059 | 13.3235 | Blang_Menghai |
| 2W791709241848 | 0.914935 | 8.73722 | Blang_Menghai |
| 2W811709228995 | 0.917358 | 9.50671 | Bai_Heqing |
| 2W811808198136 | 0.91649 | 9.32364 | Pumi_Lanping |
| 2W821808196514 | 0.918289 | 11.1748 | Bai_Heqing |
| 2W821808198142 | 0.918522 | 12.5253 | Pumi_Lanping |
| 2W841709222247 | 0.916299 | 9.54374 | Blang_Menghai |
| 2W871808160336 | 0.918605 | 12.1615 | Bai_Heqing |
| 2W871808198054 | 0.917129 | 9.49132 | Bai_Heqing |
| 2W901709222373 | 0.917034 | 10.0909 | Hani_Honghe |
| 2W911709240519 | 0.917082 | 9.69959 | Hani_Honghe |
| 2W941808211805 | 0.917487 | 10.4048 | Pumi_Lanping |
| 2W951709222764 | 0.918299 | 11.8879 | Wa_Cangyuan |
| 2W951808198103 | 0.918092 | 10.952 | Pumi_Lanping |
| 2W961709240139 | 0.91697 | 9.88615 | Blang_Menghai |

**Table S2. Allele frequency spectrum of all and novel variants**

| MAF | count | type |
|---|---|---|
| 0.005 | 4635622 | all |
| 0.01 | 1245574 | all |
| 0.02 | 1208054 | all |
| 0.03 | 536327 | all |
| 0.05 | 652165 | all |
| 0.1 | 1007890 | all |
| 0.2 | 1375244 | all |
| 0.5 | 3001929 | all |
| 0.005 | 2555695 | Novel |
| 0.01 | 414702 | Novel |
| 0.02 | 184772 | Novel |
| 0.03 | 37069 | Novel |
| 0.05 | 30055 | Novel |
| 0.1 | 32048 | Novel |
| 0.2 | 32490 | Novel |
| 0.5 | 52558 | Novel |

**Table S3. Relationship between bialleic SNPs and the number of variants among diverse functional categories**

| Variant type | Number | Variant |
|---|---|---|
| splice_acceptor_variant | 860 | Novel SNPs |
| splice_donor_variant | 1064 | Novel SNPs |
| stop_gained | 901 | Novel SNPs |
| stop_lost | 90 | Novel SNPs |
| start_lost | 110 | Novel SNPs |
| missense_variant | 48375 | Novel SNPs |
| splice_donor_5th_base_variant | 617 | Novel SNPs |
| splice_region_variant | 10645 | Novel SNPs |
| splice_donor_region_variant | 2232 | Novel SNPs |
| splice_polypyrimidine_tract_variant | 11821 | Novel SNPs |
| incomplete_terminal_codon_variant | 16 | Novel SNPs |
| stop_retained_variant | 30 | Novel SNPs |
| synonymous_variant | 56676 | Novel SNPs |
| coding_sequence_variant | 26 | Novel SNPs |
| mature_miRNA_variant | 41 | Novel SNPs |
| 5_prime_UTR_variant | 23579 | Novel SNPs |
| 3_prime_UTR_variant | 106755 | Novel SNPs |
| non_coding_transcript_exon_variant | 177142 | Novel SNPs |
| intron_variant | 10199169 | Novel SNPs |
| NMD_transcript_variant | 1131317 | Novel SNPs |
| non_coding_transcript_variant | 3965328 | Novel SNPs |
| upstream_gene_variant | 1337549 | Novel SNPs |
| downstream_gene_variant | 1346764 | Novel SNPs |
| TF_binding_site_variant | 136328 | Novel SNPs |
| regulatory_region_variant | 621640 | Novel SNPs |
| intergenic_variant | 1131087 | Novel SNPs |

**Table S4. Pairwise FST results confirmed the genetic relationship between TYC and worldwide populations**

| Population1 | Population2 | Fst |
|---|---|---|
| Blang_Menghai | Han_Yunyang | 0.00003541 |
| Qiang_Matou | Han_Fuling | 0.00008677 |
| Qiang_Matou | Han_Anshun | 0.00015175 |
| Qiang_Liujia | Han_Yubei | 0.00019525 |
| Hani_Honghe | Han_Qijiang | 0.00021173 |
| Bai_Heqing | Yi | 0.00027801 |
| Qiang_Matou | Han_Jiulongpo | 0.00032583 |
| Tibetan_Baima_Wujiao | Han_Dianjiang | 0.00047343 |
| Tibetan_Amdo_Dalu | Han_Yunyang | 0.00048175 |
| Bai_Heqing | Tu | 0.00048232 |
| Qiang_Matou | Han_Hanzhong | 0.00051447 |
| Pumi_Lanping | Han_Liangping | 0.00056367 |
| Qiang_Xiaoyuan | Qiang_Matou | 0.00058445 |
| Qiang_Gaoping | Han_Yanan | 0.00062469 |
| Qiang_Matou | Han_Qiannan | 0.000703 |
| Qiang_Gaoping | Han_Qianxinan | 0.00072512 |
| Qiang_Gaoping | Han_Yuzhong | 0.00082217 |
| Qiang_Gaoping | Han_Banan | 0.00096238 |
| Qiang_Gaoping | Qiang_Liujia | 0.000995 |
| Qiang_Gaoping | Han_Shapingba | 0.00109867 |
| Qiang_Gaoping | Han_Tongliang | 0.00111711 |
| Qiang_Liujia | Han_Bishan | 0.00118608 |
| Lahu_Lancang | Han_Jiangbei | 0.00126133 |
| Qiang_Xiaoyuan | Qiang_Gaoping | 0.00130784 |
| Bai_Heqing | Daur | 0.00133422 |
| Pumi_Lanping | Han_Rongchang | 0.00133506 |
| Qiang_Matou | Han_Xianyang | 0.00137849 |
| Qiang_Liujia | Han_Nanan | 0.00144066 |
| Qiang_Matou | Qiang_Gaoping | 0.00146487 |
| Bai_Heqing | Han_Jiangjin | 0.00151641 |
| Qiang_Liujia | Han_Fengdu | 0.00155251 |
| Qiang_Xiaoyuan | Han_Jiangjin | 0.00158179 |
| Hani_Honghe | Han_Yubei | 0.00159617 |
| Qiang_Gaoping | Han_Shangluo | 0.00177261 |
| Qiang_Liujia | Han_Yongchuan | 0.00180744 |
| Tibetan_Amdo_Dalu | Han_Tongnan | 0.00184755 |
| Qiang_Xiaoyuan | Han_Jiulongpo | 0.00186971 |
| Tibetan_Baima_Guoyuan | Han_Nanan | 0.00189191 |
| Qiang_Liujia | Han_Hechuan | 0.00205111 |
| Qiang_Xiaoyuan | Han_Fuling | 0.00214066 |
| Qiang_Xiaoyuan | Han_Qiannan | 0.0021649 |
| Bai_Heqing | Han_Hanzhong | 0.00221928 |
| Tibetan_Baima_Guoyuan | Han_Hechuan | 0.0022439 |
| Qiang_Matou | Qiang_Liujia | 0.00225217 |
| Tibetan_Baima_Guoyuan | Han_Yanan | 0.00227951 |
| Qiang_Xiaoyuan | Qiang_Liujia | 0.00228834 |
| Qiang_Liujia | Han_Zhongxian | 0.00229019 |
| Bai_Heqing | Han_Xianyang | 0.00230197 |
| Qiang_Gaoping | Han_Ankang | 0.0023258 |
| Tibetan_Baima_Guoyuan | Han_Zhongxian | 0.0023352 |
| Tibetan_Baima_Wujiao | Han_Liangping | 0.00243139 |
| Tibetan_Baima_Guoyuan | Han_Bishan | 0.00252827 |
| Tibetan_Baima_Guoyuan | Han_Fengdu | 0.00254521 |
| Tibetan_Baima_Wujiao | Han_Rongchang | 0.00259708 |
| Qiang_Liujia | Han_Qiandongnan | 0.0026131 |
| Qiang_Xiaoyuan | Han_Hanzhong | 0.00264475 |
| Qiang_Xiaoyuan | Han_Anshun | 0.0026767 |
| Bai_Heqing | Han_Jiulongpo | 0.00272382 |
| Bai_Heqing | Han_Fuling | 0.00275869 |
| Hani_Honghe | Han_Nanan | 0.00286147 |
| Qiang_Xiaoyuan | Han_Xianyang | 0.00297635 |
| Bai_Heqing | Han_Anshun | 0.0029911 |
| Bai_Heqing | Qiang_Xiaoyuan | 0.00305973 |
| Qiang_Matou | Han_Putian | 0.00313091 |

| | | |
|---|---|---|
| Pumi_Lanping | Mongola | 0.00320608 |
| Hani_Honghe | Han_Hechuan | 0.00331769 |
| Qiang_Matou | Han_Kaifeng | 0.00343162 |
| Hani_Honghe | Mongola | 0.00348766 |
| Bai_Heqing | Han_Qiannan | 0.00354544 |
| Hani_Honghe | Han_Bishan | 0.00358127 |
| Pumi_Lanping | Han_Qijiang | 0.00360932 |
| Tibetan_Baima_Guoyuan | Han_Yongchuan | 0.00362353 |
| Bai_Heqing | KHV | 0.0036586 |
| Qiang_Matou | Han_Zhumadian | 0.00369371 |
| Hani_Honghe | CHB | 0.0037769 |
| Qiang_Matou | Han_Zhangzhou | 0.00379241 |
| Bai_Heqing | Miao | 0.0038739 |
| Qiang_Matou | Han_Shangqiu | 0.00389748 |
| Qiang_Matou | Han_Hebi | 0.0039014 |
| Bai_Heqing | Qiang_Matou | 0.00394159 |
| Hani_Honghe | Han_Fengdu | 0.00395096 |
| Hani_Honghe | Han_Qiandongnan | 0.00396119 |
| Qiang_Matou | Han_Datong | 0.00399755 |
| Qiang_Matou | Han_Xinyang | 0.0040051 |
| Qiang_Liujia | Han_Kaixian | 0.00401241 |
| Qiang_Matou | Han_Luohe | 0.00403778 |
| Qiang_Matou | Han_Baoji | 0.00404497 |
| Hani_Honghe | CHS | 0.00405797 |
| Qiang_Matou | Han_Yangquan | 0.00411718 |
| Hani_Honghe | Han_Zhongxian | 0.00412876 |
| Qiang_Matou | Han_Xiamen | 0.00414617 |
| Qiang_Matou | Han_Longyan | 0.00415634 |
| Qiang_Matou | Han_Xuchang | 0.00418962 |
| Qiang_Matou | Han_Luoyang | 0.00419292 |
| Hani_Honghe | Han_Yongchuan | 0.00419468 |
| Wa_Cangyuan | Han_Yunyang | 0.00419627 |
| Hani_Honghe | KHV | 0.00423279 |
| Qiang_Matou | Han_Binzhou | 0.00424866 |
| Qiang_Matou | Han_Jiaozuo | 0.00430974 |
| Qiang_Matou | Han_Xinzhou | 0.0043176 |
| Qiang_Liujia | Han_Wanzhou | 0.00431964 |
| Lahu_Lancang | Han_Nanchuan | 0.00432437 |
| Qiang_Matou | Han_Sanming | 0.00434459 |
| Qiang_Matou | Han_Jining | 0.00434925 |
| Qiang_Matou | Han_Heze | 0.00435612 |
| Tibetan_Baima_Guoyuan | Han_Qiandongnan | 0.00435656 |
| Wa_Cangyuan | Han_Tongnan | 0.00435879 |
| Qiang_Matou | Han_Nanping | 0.0043774 |
| Qiang_Liujia | Han_Changshou | 0.00438321 |
| Qiang_Matou | Han_Taiyuan | 0.00440129 |
| Qiang_Matou | Han_Quanzhou | 0.00440292 |
| Qiang_Matou | Han_Fuzhou | 0.00443583 |
| Qiang_Matou | Han_Jinan | 0.00443869 |
| Qiang_Matou | Han_Linyi | 0.00446115 |
| Qiang_Matou | Han_Weifang | 0.00447549 |
| Qiang_Matou | Han_Dongying | 0.00447855 |
| Qiang_Matou | Han_Puyang | 0.00448158 |
| Qiang_Matou | Han_Jinzhong | 0.0044895 |
| Qiang_Matou | Han_Zibo | 0.00450754 |
| Qiang_Matou | Han_Jincheng | 0.00450897 |
| Qiang_Matou | Han_Ningde | 0.00451731 |
| Qiang_Matou | Han_Yantai | 0.00452897 |
| Qiang_Matou | Han_Linfen | 0.00457755 |
| Tibetan_Baima_Guoyuan | Han_Changshou | 0.0045914 |
| Qiang_Matou | Han_Suzhou | 0.00460317 |
| Qiang_Matou | Han_Pingdingshan | 0.00462963 |
| Bai_Heqing | Qiang_Gaoping | 0.00471305 |
| Lahu_Lancang | Han_Wushan | 0.00471443 |
| Qiang_Matou | Han_Weihai | 0.00472539 |
| Qiang_Matou | Han_Taian | 0.00472841 |

| | | |
|---|---|---|
| Qiang_Matou | Han_Yuncheng | 0.00474521 |
| Qiang_Matou | Han_Sanmenxia | 0.00475339 |
| Tibetan_Baima_Guoyuan | Han_Wanzhou | 0.00475588 |
| Qiang_Matou | Han_Zhengzhou | 0.00481046 |
| Qiang_Matou | Han_Anyang | 0.00483141 |
| Qiang_Matou | Han_Zaozhuang | 0.00488497 |
| Qiang_Matou | Han_Dezhou | 0.00489106 |
| Qiang_Matou | Han_Rizhao | 0.00489188 |
| Bai_Heqing | Qiang_Liujia | 0.00489415 |
| Qiang_Matou | Han_Jiyuan | 0.00496104 |
| Qiang_Matou | Han_Liaocheng | 0.0049615 |
| Tibetan_Baima_Guoyuan | Han_Shangluo | 0.0049864 |
| Bai_Heqing | Han_Kaifeng | 0.00499771 |
| Pumi_Lanping | Han_Yubei | 0.00499953 |
| Qiang_Matou | Han_Nanyang | 0.00501659 |
| Qiang_Gaoping | Han_Jiangjin | 0.00502308 |
| Qiang_Gaoping | Han_Anshun | 0.00504324 |
| Tibetan_Baima_Guoyuan | Han_Kaixian | 0.00512351 |
| Tibetan_Baima_Wujiao | Tibetan_Chamdo | 0.00515892 |
| Qiang_Liujia | Han_Yanan | 0.00516748 |
| Qiang_Matou | Han_Qingdao | 0.0051717 |
| Qiang_Matou | Han_Xinxiang | 0.00517628 |
| Qiang_Matou | Han_Laiwu | 0.00517878 |
| Qiang_Matou | Han_Zhoukou | 0.00518089 |
| Bai_Heqing | Han_Zhumadian | 0.00518351 |
| Bai_Heqing | Han_Baoji | 0.00519455 |
| Qiang_Xiaoyuan | Han_Putian | 0.00525837 |
| Bai_Heqing | Yi_Xichang | 0.00528033 |
| Bai_Heqing | Hezhen | 0.0052904 |
| Bai_Heqing | Han_Luohe | 0.00530687 |
| Qiang_Gaoping | Han_Jiulongpo | 0.00531351 |
| Qiang_Matou | Han_Changzhi | 0.00531846 |
| Bai_Heqing | Han_Datong | 0.00534936 |
| Qiang_Matou | Han_Lvliang | 0.00536154 |
| Qiang_Xiaoyuan | Han_Kaifeng | 0.00536705 |
| Qiang_Matou | Han_YulinSX | 0.00538285 |
| Bai_Heqing | Han_Xinzhou | 0.00540884 |
| Bai_Heqing | Han_Yangquan | 0.00545185 |
| Bai_Heqing | Han_Hebi | 0.00547355 |
| Qiang_Liujia | Han_Yuzhong | 0.00549248 |
| Qiang_Gaoping | Han_Fuling | 0.00559351 |
| Bai_Heqing | Han_Jiaozuo | 0.0056023 |
| Bai_Heqing | Han_Jincheng | 0.00561962 |
| Qiang_Gaoping | Han_Qiannan | 0.00563719 |
| Qiang_Xiaoyuan | Han_Zhumadian | 0.005639 |
| Bai_Heqing | Han_Sanmenxia | 0.00565731 |
| Qiang_Xiaoyuan | Han_Shangqiu | 0.00565995 |
| Bai_Heqing | Han_Taiyuan | 0.00566411 |
| Qiang_Gaoping | Han_Hanzhong | 0.00569643 |
| Bai_Heqing | Han_Binzhou | 0.00569822 |
| Bai_Heqing | Han_Linfen | 0.00570215 |
| Bai_Heqing | Han_Shangqiu | 0.00570419 |
| Bai_Heqing | Han_Jining | 0.00574743 |
| Bai_Heqing | Han_Jinzhong | 0.0058238 |
| Qiang_Xiaoyuan | Han_Xiamen | 0.00582417 |
| Bai_Heqing | Han_Suzhou | 0.00582504 |
| Bai_Heqing | Han_Luoyang | 0.00584478 |
| Bai_Heqing | Han_Heze | 0.00585853 |
| Bai_Heqing | Han_Liaocheng | 0.00587233 |
| Bai_Heqing | Han_Dongying | 0.00588026 |
| Bai_Heqing | Han_Yuncheng | 0.00591777 |
| Hani_Honghe | Han_Kaixian | 0.00592373 |
| Bai_Heqing | Han_Jinan | 0.00593786 |
| Pumi_Lanping | CHB | 0.00597592 |
| Bai_Heqing | CDX | 0.00598187 |
| Qiang_Xiaoyuan | Han_Yangquan | 0.0059952 |

| | | |
|---|---|---|
| Bai_Heqing | Han_Taian | 0.00601563 |
| Bai_Heqing | Han_Weihai | 0.00602083 |
| Qiang_Xiaoyuan | Han_Zhengzhou | 0.00603575 |
| Bai_Heqing | Han_Zhengzhou | 0.00605339 |
| Bai_Heqing | Han_Jiyuan | 0.00606002 |
| Qiang_Xiaoyuan | Han_Baoji | 0.00606567 |
| Qiang_Matou | Han_Weinan | 0.00606942 |
| Qiang_Xiaoyuan | Han_Zhangzhou | 0.00606961 |
| Bai_Heqing | Han_Lvliang | 0.00607605 |
| Qiang_Xiaoyuan | Han_Hebi | 0.00607661 |
| Qiang_Liujia | Han_Shapingba | 0.00608419 |
| Bai_Heqing | Han_Linyi | 0.00609005 |
| Bai_Heqing | Han_YulinSX | 0.00610289 |
| Tibetan_Baima_Guoyuan | Han_Yuzhong | 0.00610411 |
| Bai_Heqing | Han_Weifang | 0.00612501 |
| Qiang_Xiaoyuan | Han_Xuchang | 0.00612876 |
| Bai_Heqing | Han_Yantai | 0.00612904 |
| Qiang_Xiaoyuan | Han_Luohe | 0.00613336 |
| Bai_Heqing | Han_Xuchang | 0.0061439 |
| Qiang_Xiaoyuan | Han_Xinyang | 0.00616463 |
| Bai_Heqing | Han_Zaozhuang | 0.00616677 |
| Bai_Heqing | Han_Puyang | 0.00616911 |
| Qiang_Xiaoyuan | Han_Nanping | 0.00618271 |
| Qiang_Xiaoyuan | Han_Jining | 0.00619528 |
| Qiang_Xiaoyuan | Han_Xinzhou | 0.00620765 |
| Qiang_Xiaoyuan | Han_Weifang | 0.00621365 |
| Bai_Heqing | Han_Anyang | 0.00621677 |
| Blang_Menghai | Han_Dianjiang | 0.00622244 |
| Qiang_Xiaoyuan | Han_Jinzhong | 0.00623659 |
| Bai_Heqing | Mosuo_Yanyuan | 0.00625085 |
| Qiang_Xiaoyuan | Han_Luoyang | 0.00626337 |
| Qiang_Xiaoyuan | Han_Ningde | 0.00626526 |
| Bai_Heqing | Han_Nanyang | 0.00627181 |
| Qiang_Xiaoyuan | Han_Taiyuan | 0.00627431 |
| Qiang_Gaoping | Han_Xianyang | 0.00628335 |
| Bai_Heqing | Han_Pingdingshan | 0.00629504 |
| Bai_Heqing | Han_Laiwu | 0.0063067 |
| Bai_Heqing | Han_Dezhou | 0.00631204 |
| Qiang_Xiaoyuan | Han_Dongying | 0.0063186 |
| Tibetan_Baima_Wujiao | Han_Qijiang | 0.00634087 |
| Hani_Honghe | CDX | 0.00634435 |
| Qiang_Xiaoyuan | Han_Taian | 0.00635057 |
| Qiang_Xiaoyuan | Han_Jiaozuo | 0.00635954 |
| Qiang_Xiaoyuan | Han_Binzhou | 0.0064011 |
| Bai_Heqing | Han_Zibo | 0.00640423 |
| Qiang_Xiaoyuan | Han_Sanming | 0.00641625 |
| Qiang_Xiaoyuan | Han_Jiyuan | 0.00642098 |
| Hani_Honghe | Han_Changshou | 0.00642159 |
| Qiang_Xiaoyuan | Han_Jincheng | 0.00642671 |
| Qiang_Xiaoyuan | Han_Datong | 0.00642709 |
| Qiang_Xiaoyuan | Han_Quanzhou | 0.00642728 |
| Qiang_Xiaoyuan | Han_Linfen | 0.00642993 |
| Bai_Heqing | Han_Changzhi | 0.00644042 |
| Bai_Heqing | Han_Rizhao | 0.0064494 |
| Qiang_Xiaoyuan | Han_Puyang | 0.00644944 |
| Qiang_Liujia | Han_Qianxinan | 0.00645677 |
| Qiang_Liujia | Han_Shangluo | 0.00647534 |
| Qiang_Xiaoyuan | Han_Heze | 0.00647587 |
| Pumi_Lanping | Naxi | 0.00648139 |
| Bai_Heqing | Han_Putian | 0.00649207 |
| Qiang_Liujia | Han_Banan | 0.00649968 |
| Qiang_Xiaoyuan | Han_Fuzhou | 0.00650148 |
| Qiang_Xiaoyuan | Han_Jinan | 0.00652623 |
| Qiang_Xiaoyuan | Han_Yuncheng | 0.00654333 |
| Tibetan_Baima_Wujiao | Tibetan_Nyingchi | 0.00655562 |
| Qiang_Xiaoyuan | Han_Sanmenxia | 0.00656457 |

| | | |
|---|---|---|
| Qiang_Xiaoyuan | Han_Yantai | 0.00658406 |
| Pumi_Lanping | Tibetan_Muli | 0.00660291 |
| Qiang_Liujia | Han_Tongliang | 0.00660307 |
| Qiang_Xiaoyuan | Han_Zibo | 0.00660554 |
| Qiang_Xiaoyuan | Han_Zhoukou | 0.00660842 |
| Hani_Honghe | Han_Wanzhou | 0.00661435 |
| Tibetan_Baima_Guoyuan | Han_Banan | 0.00661765 |
| Bai_Heqing | Han_Qingdao | 0.00662703 |
| Qiang_Xiaoyuan | Han_Longyan | 0.00663326 |
| Qiang_Xiaoyuan | Han_Pingdingshan | 0.00664304 |
| Blang_Menghai | KHV | 0.0066467 |
| Qiang_Xiaoyuan | Han_Nanyang | 0.00666524 |
| Qiang_Matou | Han_Lanzhou | 0.00666822 |
| Qiang_Xiaoyuan | Han_Dezhou | 0.00670268 |
| Qiang_Xiaoyuan | Han_Liaocheng | 0.00670794 |
| Tibetan_Amdo_Dalu | Han_Rongchang | 0.00671401 |
| Bai_Heqing | Han_Zhoukou | 0.00672199 |
| Qiang_Xiaoyuan | Han_Linyi | 0.00672238 |
| Qiang_Xiaoyuan | Han_Suzhou | 0.00675061 |
| Tibetan_Amdo_Dalu | Han_Liangping | 0.00678589 |
| Bai_Heqing | Han_Xinyang | 0.00679006 |
| Qiang_Xiaoyuan | Han_Anyang | 0.00679531 |
| Tibetan_Amdo_Dalu | Han_Dianjiang | 0.0068032 |
| Tibetan_Baima_Guoyuan | Han_Shapingba | 0.00683273 |
| Qiang_Xiaoyuan | Han_Weihai | 0.00683728 |
| Qiang_Xiaoyuan | Han_Zaozhuang | 0.0068491 |
| Bai_Heqing | Han_Xinxiang | 0.00685846 |
| Pumi_Lanping | Han_Nanan | 0.00686175 |
| Qiang_Xiaoyuan | Han_YulinSX | 0.00693595 |
| Qiang_Liujia | Han_Ankang | 0.00693823 |
| Qiang_Xiaoyuan | Han_Lvliang | 0.00698032 |
| Qiang_Xiaoyuan | Han_Qingdao | 0.00702659 |
| Qiang_Xiaoyuan | Han_Changzhi | 0.00703128 |
| Bai_Heqing | Cambodian | 0.00706953 |
| Pumi_Lanping | Han_Bishan | 0.00708947 |
| Qiang_Xiaoyuan | Han_Xinxiang | 0.00709791 |
| Qiang_Xiaoyuan | Han_Rizhao | 0.00709854 |
| Tibetan_Baima_Guoyuan | Han_Ankang | 0.00710912 |
| Hani_Honghe | Han_Yanan | 0.00714994 |
| Blang_Menghai | Han_Rongchang | 0.00715246 |
| Tibetan_Baima_Wujiao | Han_Yubei | 0.0071679 |
| Bai_Heqing | Han_Xiamen | 0.0072326 |
| Tibetan_Baima_Guoyuan | Han_Qianxinan | 0.0072454 |
| Qiang_Xiaoyuan | Han_Laiwu | 0.00725243 |
| Blang_Menghai | Cambodian | 0.00727596 |
| Bai_Heqing | Han_Lanzhou | 0.00730482 |
| Bai_Heqing | Tibetan_Muli | 0.0073052 |
| Tibetan_Baima_Guoyuan | Han_Tongliang | 0.00731101 |
| Pumi_Lanping | Yi | 0.00731246 |
| Hani_Honghe | Han_Banan | 0.00732953 |
| Bai_Heqing | Han_Weinan | 0.00738695 |
| Pumi_Lanping | Han_Yanan | 0.00742622 |
| Bai_Heqing | Han_Sanming | 0.00747216 |
| Bai_Heqing | Han_Ningde | 0.00747766 |
| Bai_Heqing | Han_Zhangzhou | 0.00753126 |
| Hani_Honghe | Han_Shapingba | 0.00753195 |
| Tibetan_Baima_Guoyuan | Tibetan_Chamdo | 0.00762569 |
| Hani_Honghe | Han_Qianxinan | 0.00763997 |
| Bai_Heqing | Han_Longyan | 0.00764508 |
| Pumi_Lanping | Han_Zhongxian | 0.00765352 |
| Tibetan_Baima_Guoyuan | Tibetan_Baima_Wuj | 0.00766596 |
| Hani_Honghe | Han_Tongliang | 0.00766757 |
| Qiang_Matou | Han_Nanchong | 0.00773897 |
| Bai_Heqing | Han_Quanzhou | 0.00774382 |
| Bai_Heqing | Han_Nanping | 0.00775347 |
| Hani_Honghe | Tujia | 0.00782766 |

| | | |
|---|---|---|
| Hani_Honghe | Han_Yuzhong | 0.00787487 |
| Bai_Heqing | Tibetan_Baima_Wuj | 0.00788625 |
| Bai_Heqing | Han_Fuzhou | 0.00790676 |
| Qiang_Xiaoyuan | Tibetan_Baima_Wuj | 0.00795424 |
| Wa_Cangyuan | KHV | 0.00798848 |
| Tibetan_Baima_Wujiao | Tibetan_Nagqu | 0.00801826 |
| Qiang_Xiaoyuan | Han_Weinan | 0.00802378 |
| Bai_Heqing | Tibetan_Chamdo | 0.00802495 |
| Qiang_Xiaoyuan | Han_Lanzhou | 0.00805622 |
| Pumi_Lanping | Han_Hechuan | 0.00807287 |
| Bai_Heqing | Dai | 0.0081199 |
| Bai_Heqing | JPT | 0.00812736 |
| Blang_Menghai | Han_Liangping | 0.00814506 |
| Pumi_Lanping | Xibo | 0.00824819 |
| Qiang_Gaoping | Han_Putian | 0.00828264 |
| Pumi_Lanping | Han_Yongchuan | 0.00835142 |
| Pumi_Lanping | Han_Fengdu | 0.00854299 |
| Qiang_Gaoping | Han_Kaifeng | 0.00865224 |
| Blang_Menghai | CDX | 0.00873569 |
| Qiang_Gaoping | Han_Xiamen | 0.00877211 |
| Qiang_Matou | Han_Xian | 0.00883843 |
| Tibetan_Baima_Guoyuan | Han_Xianyang | 0.00886479 |
| Tibetan_Baima_Guoyuan | Tibetan_Nagqu | 0.0088672 |
| Pumi_Lanping | CHS | 0.00888837 |
| Qiang_Gaoping | Han_Zhumadian | 0.00890455 |
| Pumi_Lanping | Tibetan_Dujiangyan | 0.00894027 |
| Pumi_Lanping | Tu | 0.00895274 |
| Qiang_Gaoping | Han_Shangqiu | 0.00896763 |
| Qiang_Gaoping | Han_Hebi | 0.00898989 |
| Bai_Heqing | She | 0.00902266 |
| Qiang_Xiaoyuan | Tibetan_Baima_Guo | 0.00904805 |
| Hani_Honghe | Naxi | 0.00907476 |
| Qiang_Gaoping | Han_Longyan | 0.00907561 |
| Qiang_Gaoping | Han_Jining | 0.00908215 |
| Wa_Cangyuan | Han_Liangping | 0.00913855 |
| Qiang_Gaoping | Han_Dongying | 0.00915494 |
| Qiang_Gaoping | Han_Yangquan | 0.00916458 |
| Tibetan_Baima_Wujiao | Han_Nanan | 0.00917625 |
| Hani_Honghe | Xibo | 0.00918582 |
| Hani_Honghe | Han_Shangluo | 0.00920669 |
| Qiang_Gaoping | Han_Sanming | 0.00921682 |
| Wa_Cangyuan | Han_Dianjiang | 0.00921897 |
| Qiang_Gaoping | Han_Luohe | 0.00922464 |
| Qiang_Gaoping | Han_Xinyang | 0.00923975 |
| Qiang_Gaoping | Han_Quanzhou | 0.00927347 |
| Qiang_Liujia | Han_Jiangjin | 0.00927679 |
| Qiang_Gaoping | Han_Xuchang | 0.00927983 |
| Qiang_Gaoping | Han_Ningde | 0.00928398 |
| Hani_Honghe | Yi | 0.00931492 |
| Qiang_Gaoping | Han_Jiaozuo | 0.00931612 |
| Qiang_Gaoping | Han_Jinzhong | 0.00931782 |
| Qiang_Gaoping | Han_Jinan | 0.00932014 |
| Tibetan_Baima_Wujiao | Han_Yanan | 0.00932317 |
| Qiang_Gaoping | Han_Baoji | 0.00932793 |
| Qiang_Gaoping | Han_Xinzhou | 0.00932827 |
| Qiang_Gaoping | Han_Datong | 0.00934071 |
| Bai_Heqing | Tibetan_Baima_Guo | 0.00938537 |
| Qiang_Gaoping | Han_Fuzhou | 0.00939347 |
| Tibetan_Baima_Wujiao | Tibetan_Shigatse | 0.00939609 |
| Qiang_Gaoping | Han_Linyi | 0.00940293 |
| Qiang_Gaoping | Han_Taiyuan | 0.00940703 |
| Qiang_Gaoping | Han_Anyang | 0.00941232 |
| Qiang_Gaoping | Han_Zhangzhou | 0.0094131 |
| Tibetan_Baima_Wujiao | Han_Fengdu | 0.00941459 |
| Qiang_Gaoping | Han_Heze | 0.00941751 |
| Pumi_Lanping | Han_Qiandongnan | 0.00942282 |

| | | |
|---|---|---|
| Pumi_Lanping | Tibetan_Chamdo | 0.00943746 |
| Bai_Heqing | Tibetan_Nyingchi | 0.00945238 |
| Bai_Heqing | Tibetan_Dujiangyan | 0.00948323 |
| Qiang_Gaoping | Han_Binzhou | 0.00949492 |
| Qiang_Gaoping | Han_Nanping | 0.00950789 |
| Tibetan_Baima_Wujiao | Tibetan_Shannan | 0.00952827 |
| Tibetan_Baima_Wujiao | Han_Zhongxian | 0.00954362 |
| Qiang_Matou | Tibetan_Baima_Wuj | 0.00954576 |
| Qiang_Gaoping | Han_Weihai | 0.00955802 |
| Qiang_Gaoping | Han_Taian | 0.00956409 |
| Qiang_Gaoping | Han_Linfen | 0.00956605 |
| Qiang_Gaoping | Han_Yantai | 0.00958541 |
| Qiang_Gaoping | Han_Zibo | 0.00958799 |
| Qiang_Gaoping | Han_Puyang | 0.00963546 |
| Qiang_Gaoping | Han_Zhengzhou | 0.00964008 |
| Qiang_Gaoping | Han_Luoyang | 0.00965931 |
| Qiang_Gaoping | Han_Weifang | 0.00966262 |
| Tibetan_Baima_Wujiao | Qiang_Liujia | 0.00966678 |
| Wa_Cangyuan | CDX | 0.0096809 |
| Qiang_Gaoping | Han_Yuncheng | 0.00968929 |
| Qiang_Gaoping | Han_Pingdingshan | 0.00969426 |
| Qiang_Gaoping | Han_YulinSX | 0.00969714 |
| Qiang_Xiaoyuan | Han_Nanchong | 0.0097087 |
| Qiang_Gaoping | Han_Nanyang | 0.00971233 |
| Tibetan_Baima_Guoyuan | Qiang_Liujia | 0.00972998 |
| Pumi_Lanping | Han_Kaixian | 0.00976576 |
| Bai_Heqing | Han | 0.00977021 |
| Qiang_Gaoping | Han_Dezhou | 0.00977442 |
| Hani_Honghe | Cambodian | 0.00977691 |
| Pumi_Lanping | Mosuo_Yanyuan | 0.00977761 |
| Qiang_Gaoping | Han_Jincheng | 0.00978622 |
| Qiang_Gaoping | Han_Liaocheng | 0.00981847 |
| Qiang_Gaoping | Han_Zaozhuang | 0.00982271 |
| Hani_Honghe | Han_Ankang | 0.00982588 |
| Qiang_Gaoping | Han_Laiwu | 0.0098612 |
| Qiang_Gaoping | Han_Sanmenxia | 0.0098625 |
| Qiang_Gaoping | Han_Suzhou | 0.00986943 |
| Pumi_Lanping | Han_Changshou | 0.00989287 |
| Tibetan_Baima_Guoyuan | Qiang_Matou | 0.009893 |
| Qiang_Gaoping | Han_Xinxiang | 0.00993123 |
| Bai_Heqing | Han_Xian | 0.00994115 |
| Tibetan_Baima_Wujiao | Han_Yongchuan | 0.009967 |
| Qiang_Liujia | Han_Anshun | 0.00997317 |
| Qiang_Gaoping | Han_Zhoukou | 0.00998904 |
| Qiang_Gaoping | Han_Rizhao | 0.00999099 |
| Bai_Heqing | Han_Nanchong | 0.00999484 |
| Qiang_Gaoping | Han_Jiyuan | 0.0100175 |
| Tibetan_Baima_Wujiao | Tibetan_Lhasa | 0.0100253 |
| Tibetan_Baima_Wujiao | Han_Hechuan | 0.0100454 |
| Tibetan_Baima_Guoyuan | Han_Hanzhong | 0.01005 |
| Qiang_Gaoping | Han_Qingdao | 0.0101003 |
| Qiang_Gaoping | Tibetan_Baima_Wuj | 0.0101178 |
| Qiang_Xiaoyuan | Han_Taiwan | 0.0102059 |
| Pumi_Lanping | Han_Wanzhou | 0.0102314 |
| Bai_Heqing | Tibetan_Nagqu | 0.0103288 |
| Tibetan_Baima_Wujiao | Han_Bishan | 0.0103339 |
| Qiang_Xiaoyuan | Tibetan_Chamdo | 0.0103481 |
| Qiang_Xiaoyuan | Han_Xian | 0.0103731 |
| Qiang_Gaoping | Han_Lvliang | 0.0103878 |
| Qiang_Liujia | Han_Hanzhong | 0.0104089 |
| Qiang_Gaoping | Han_Changzhi | 0.0104141 |
| Qiang_Liujia | Han_Fuling | 0.0104287 |
| Tibetan_Baima_Guoyuan | Han_Jiangjin | 0.010492 |
| Pumi_Lanping | Han_Shangluo | 0.0105515 |
| Lahu_Lancang | JPT | 0.0105559 |
| Qiang_Gaoping | Han_Weinan | 0.0106365 |

| | | |
|---|---|---|
| Hani_Honghe | NorthernHan | 0.010697 |
| Wa_Cangyuan | Han_Rongchang | 0.0107093 |
| Pumi_Lanping | Yi_Xichang | 0.0107467 |
| Hani_Honghe | Bai_Heqing | 0.010764 |
| Tibetan_Baima_Guoyuan | Tibetan_Nyingchi | 0.0107675 |
| Tibetan_Baima_Guoyuan | Qiang_Gaoping | 0.010804 |
| Pumi_Lanping | Tibetan_Nagqu | 0.0108307 |
| Pumi_Lanping | NorthernHan | 0.0109021 |
| Hani_Honghe | Qiang_Xiaoyuan | 0.0109317 |
| Bai_Heqing | Oroqen | 0.0109929 |
| Qiang_Liujia | Han_Xianyang | 0.0109946 |
| Pumi_Lanping | Tujia | 0.0110828 |
| Qiang_Matou | Tibetan_Chamdo | 0.0111451 |
| Hani_Honghe | Qiang_Matou | 0.0111514 |
| Tibetan_Baima_Guoyuan | Han_Anshun | 0.0111742 |
| Blang_Menghai | Han_Qijiang | 0.011215 |
| Tibetan_Amdo_Dalu | Han_Yubei | 0.0112327 |
| Hani_Honghe | Han_Jiangjin | 0.0112366 |
| Tibetan_Amdo_Dalu | Han_Qijiang | 0.0112471 |
| Pumi_Lanping | Bai_Heqing | 0.0112791 |
| Tibetan_Baima_Guoyuan | Han_Fuling | 0.0112868 |
| Qiang_Liujia | Han_Jiulongpo | 0.0113106 |
| Tibetan_Baima_Wujiao | Han_Qiandongnan | 0.0113335 |
| Pumi_Lanping | Han_Yuzhong | 0.0113419 |
| Qiang_Gaoping | Han_Lanzhou | 0.0113549 |
| Qiang_Liujia | Han_Qiannan | 0.011412 |
| Qiang_Gaoping | Tibetan_Chamdo | 0.0115006 |
| Tibetan_Baima_Guoyuan | Han_Jiulongpo | 0.0115661 |
| Tibetan_Baima_Guoyuan | Han_Hebi | 0.0115727 |
| Hani_Honghe | Han_Jiulongpo | 0.0116397 |
| Hani_Honghe | Tu | 0.0116631 |
| Tibetan_Baima_Guoyuan | Han_Kaifeng | 0.0117244 |
| Pumi_Lanping | Han_Shapingba | 0.0117344 |
| Pumi_Lanping | Han_Banan | 0.0117541 |
| Pumi_Lanping | Han_Ankang | 0.0117626 |
| Qiang_Liujia | Tibetan_Chamdo | 0.0117669 |
| Qiang_Xiaoyuan | Tibetan_Nyingchi | 0.0117681 |
| Tibetan_Baima_Wujiao | Han_Changshou | 0.0118369 |
| Bai_Heqing | Tibetan_Shigatse | 0.0118512 |
| Tibetan_Baima_Guoyuan | Han_Datong | 0.0118529 |
| Bai_Heqing | Han_Taiwan | 0.0119 |
| Pumi_Lanping | Han_Tongliang | 0.011922 |
| Tibetan_Baima_Guoyuan | Han_Qiannan | 0.0119524 |
| Blang_Menghai | Han_Yubei | 0.0119589 |
| Tibetan_Baima_Guoyuan | Han_Yangquan | 0.011973 |
| Blang_Menghai | CHS | 0.0120496 |
| Hani_Honghe | Miao | 0.0120678 |
| Tibetan_Amdo_Dalu | Tibetan_Chamdo | 0.0120803 |
| Tibetan_Baima_Wujiao | Han_Wanzhou | 0.0121074 |
| Pumi_Lanping | Tibetan_Nyingchi | 0.0121117 |
| Pumi_Lanping | Tibetan_Baima_Wuj | 0.0121244 |
| Wa_Cangyuan | CHS | 0.0121247 |
| Hani_Honghe | Han_Hanzhong | 0.0121319 |
| Tibetan_Baima_Guoyuan | Han_Baoji | 0.0121857 |
| Wa_Cangyuan | Cambodian | 0.0121873 |
| Tibetan_Baima_Guoyuan | Han_Taiyuan | 0.0122114 |
| Tibetan_Baima_Guoyuan | Tibetan_Shannan | 0.0122248 |
| Tibetan_Baima_Guoyuan | Han_Zhumadian | 0.0122385 |
| Hani_Honghe | Qiang_Gaoping | 0.0122785 |
| Tibetan_Baima_Guoyuan | Han_Linfen | 0.0122986 |
| Tibetan_Baima_Guoyuan | Han_Jining | 0.0123161 |
| Tibetan_Amdo_Dalu | Han_Yanan | 0.0123499 |
| Tibetan_Baima_Wujiao | Han_Kaixian | 0.0123809 |
| Tibetan_Baima_Wujiao | Han_Shangluo | 0.0123852 |
| Tibetan_Baima_Guoyuan | Tibetan_Shigatse | 0.0123898 |
| Hani_Honghe | Daur | 0.01241 |

| | | |
|---|---|---|
| Tibetan_Baima_Guoyuan | Han_Jinzhong | 0.0124199 |
| Tibetan_Baima_Guoyuan | Han_Suzhou | 0.0124306 |
| Tibetan_Baima_Guoyuan | Han_Jiaozuo | 0.0124316 |
| Hani_Honghe | Han_Anshun | 0.0124388 |
| Qiang_Gaoping | Han_Nanchong | 0.012439 |
| Tibetan_Baima_Guoyuan | Han_Shangqiu | 0.0124565 |
| Pumi_Lanping | Han_Qianxinan | 0.0124862 |
| Tibetan_Baima_Guoyuan | Han_YulinSX | 0.0125154 |
| Tibetan_Baima_Guoyuan | Han_Xinzhou | 0.0125371 |
| Tibetan_Baima_Guoyuan | Han_Luoyang | 0.0125478 |
| Tibetan_Baima_Guoyuan | Han_Zhengzhou | 0.0125607 |
| Tibetan_Baima_Guoyuan | Han_Lvliang | 0.0125999 |
| Tibetan_Baima_Guoyuan | Han_Jincheng | 0.0126081 |
| Tibetan_Baima_Guoyuan | Han_Jinan | 0.0126129 |
| Bai_Heqing | Tibetan_Shannan | 0.0126267 |
| Hani_Honghe | Qiang_Liujia | 0.0126475 |
| Tibetan_Baima_Guoyuan | Han_Luohe | 0.0126493 |
| Tibetan_Baima_Guoyuan | Han_Laiwu | 0.0126719 |
| Tibetan_Baima_Guoyuan | Han_Heze | 0.0126749 |
| Tibetan_Baima_Guoyuan | Han_Binzhou | 0.0126986 |
| Tibetan_Baima_Guoyuan | Han_Sanmenxia | 0.0127008 |
| Tibetan_Baima_Guoyuan | Han_Dongying | 0.0127216 |
| Hani_Honghe | Han_Qiannan | 0.0127269 |
| Tibetan_Baima_Guoyuan | Han_Yuncheng | 0.0127292 |
| Hani_Honghe | Han_Fuling | 0.0127323 |
| Tibetan_Baima_Guoyuan | Han_Puyang | 0.0127355 |
| Tibetan_Baima_Guoyuan | Han_Xuchang | 0.0127513 |
| Qiang_Xiaoyuan | Tibetan_Nagqu | 0.0127573 |
| Tibetan_Baima_Guoyuan | Han_Anyang | 0.0127765 |
| Tibetan_Baima_Guoyuan | Han_Yantai | 0.0128025 |
| Tibetan_Baima_Guoyuan | Han_Linyi | 0.0128035 |
| Tibetan_Baima_Guoyuan | Han_Changzhi | 0.0128252 |
| Qiang_Matou | Tibetan_Nyingchi | 0.0129609 |
| Qiang_Liujia | Han_Zhumadian | 0.0129712 |
| Qiang_Matou | Tibetan_Nagqu | 0.0129712 |
| Tibetan_Baima_Guoyuan | Han_Taian | 0.0129794 |
| Bai_Heqing | Tibetan_Lhasa | 0.0129843 |
| Tibetan_Baima_Guoyuan | Han_Pingdingshan | 0.0130777 |
| Tibetan_Baima_Guoyuan | Han_Xinxiang | 0.0130814 |
| Tibetan_Baima_Guoyuan | Han_Zaozhuang | 0.0130896 |
| Pumi_Lanping | Daur | 0.0130918 |
| Pumi_Lanping | Tibetan_Shigatse | 0.0131385 |
| Tibetan_Baima_Guoyuan | Han_Zibo | 0.0131395 |
| Tibetan_Baima_Guoyuan | Han_Jiyuan | 0.0131623 |
| Hani_Honghe | Dai | 0.0131791 |
| Lahu_Lancang | China_Lahu | 0.0132016 |
| Tibetan_Baima_Guoyuan | Han_Qingdao | 0.0132075 |
| Tibetan_Baima_Guoyuan | Han_Zhoukou | 0.0132435 |
| Qiang_Liujia | Han_Yangquan | 0.0133184 |
| Wa_Cangyuan | CHB | 0.0133333 |
| Tibetan_Baima_Guoyuan | Han_Liaocheng | 0.0133338 |
| Tibetan_Baima_Guoyuan | Han_Dezhou | 0.013335 |
| Tibetan_Baima_Guoyuan | Han_Rizhao | 0.0133553 |
| Qiang_Gaoping | Han_Xian | 0.0133665 |
| Blang_Menghai | Han_Bishan | 0.0134098 |
| Tibetan_Baima_Guoyuan | Han_Weifang | 0.0134199 |
| Tibetan_Baima_Guoyuan | Han_Weihai | 0.01345 |
| Qiang_Liujia | Han_Putian | 0.013469 |
| Tibetan_Baima_Guoyuan | Han_Nanyang | 0.0134938 |
| Qiang_Liujia | Han_Luohe | 0.0135147 |
| Pumi_Lanping | Tibetan_Shannan | 0.0135175 |
| Tibetan_Baima_Wujiao | Han_Banan | 0.0135297 |
| Qiang_Liujia | Han_Hebi | 0.0135321 |
| Tibetan_Baima_Guoyuan | Han_Xinyang | 0.0135339 |
| Tibetan_Baima_Wujiao | Han_Shapingba | 0.0135769 |
| Qiang_Liujia | Han_Kaifeng | 0.0135861 |

| | | |
|---|---|---|
| Hani_Honghe | Han_Xianyang | 0.0135982 |
| Tibetan_Baima_Wujiao | Han_Ankang | 0.0136076 |
| Pumi_Lanping | Han_Xianyang | 0.0136309 |
| Tibetan_Baima_Wujiao | Han_Qianxinan | 0.0137049 |
| Qiang_Liujia | Han_Baoji | 0.0137067 |
| Qiang_Liujia | Han_Jiaozuo | 0.0137709 |
| Blang_Menghai | Han_Qiandongnan | 0.0137856 |
| Qiang_Liujia | Tibetan_Nyingchi | 0.0137958 |
| Qiang_Liujia | Han_Shangqiu | 0.0137969 |
| Qiang_Liujia | Han_Yantai | 0.0138291 |
| Tibetan_Amdo_Dalu | Han_Zhongxian | 0.013836 |
| Qiang_Liujia | Han_Heze | 0.0138397 |
| Pumi_Lanping | Tibetan_Baima_Guo | 0.0138411 |
| Tibetan_Amdo_Dalu | Han_Bishan | 0.0138799 |
| Blang_Menghai | Han_Hechuan | 0.0139226 |
| Blang_Menghai | Han_Yongchuan | 0.0139354 |
| Qiang_Liujia | Han_Rizhao | 0.0139354 |
| Qiang_Liujia | Han_Luoyang | 0.0139367 |
| Tibetan_Amdo_Dalu | Han_Fengdu | 0.0139397 |
| Hani_Honghe | Yi_Xichang | 0.0139511 |
| Qiang_Liujia | Han_Jinzhong | 0.0139586 |
| Tibetan_Baima_Guoyuan | Tibetan_Lhasa | 0.0140103 |
| Qiang_Liujia | Han_Ningde | 0.0140133 |
| Qiang_Liujia | Han_Sanmenxia | 0.0140137 |
| Qiang_Liujia | Han_Jining | 0.0140284 |
| Qiang_Liujia | Han_Pingdingshan | 0.0140334 |
| Qiang_Liujia | Han_Jincheng | 0.0140374 |
| Qiang_Liujia | Han_Jiyuan | 0.0140405 |
| Qiang_Liujia | Han_Zaozhuang | 0.0140406 |
| Blang_Menghai | CHB | 0.0140521 |
| Qiang_Liujia | Han_Datong | 0.0140558 |
| Qiang_Liujia | Han_Nanping | 0.0140632 |
| Pumi_Lanping | Qiang_Xiaoyuan | 0.0140739 |
| Tibetan_Baima_Guoyuan | Han_Weinan | 0.0141055 |
| Qiang_Liujia | Han_Qingdao | 0.0141197 |
| Qiang_Liujia | Han_Xinzhou | 0.0141334 |
| Qiang_Liujia | Han_Linyi | 0.0141359 |
| Qiang_Liujia | Han_Dongying | 0.0141361 |
| Tibetan_Amdo_Dalu | Han_Nanan | 0.0141365 |
| Qiang_Liujia | Han_Taiyuan | 0.0141545 |
| Qiang_Liujia | Han_Jinan | 0.0141623 |
| Qiang_Liujia | Han_Taian | 0.0141763 |
| Tibetan_Baima_Wujiao | Han_Tongliang | 0.0141799 |
| Blang_Menghai | Han_Fengdu | 0.0141899 |
| Qiang_Liujia | Han_Xinyang | 0.0141932 |
| Qiang_Liujia | Han_Liaocheng | 0.0142033 |
| Tibetan_Baima_Guoyuan | Han_Lanzhou | 0.0142084 |
| Qiang_Liujia | Han_Puyang | 0.0142117 |
| Qiang_Liujia | Han_Zibo | 0.0142219 |
| Wa_Cangyuan | Han_Qijiang | 0.0142302 |
| Qiang_Liujia | Han_Zhengzhou | 0.014233 |
| Qiang_Liujia | Han_Dezhou | 0.0142591 |
| Blang_Menghai | Han_Nanan | 0.0142661 |
| Qiang_Liujia | Han_Weifang | 0.0142676 |
| Tibetan_Baima_Wujiao | Han_Yuzhong | 0.0142779 |
| Qiang_Liujia | Han_Weihai | 0.0143 |
| Qiang_Liujia | Han_Longyan | 0.0143001 |
| Qiang_Liujia | Han_Anyang | 0.0143056 |
| Pumi_Lanping | Tibetan_Lhasa | 0.0143064 |
| Qiang_Xiaoyuan | Tibetan_Shigatse | 0.0143558 |
| Qiang_Liujia | Han_Binzhou | 0.0143715 |
| Qiang_Liujia | Han_Nanyang | 0.014384 |
| Qiang_Liujia | Han_Quanzhou | 0.0143985 |
| Qiang_Liujia | Han_Sanming | 0.0144319 |
| Hani_Honghe | JPT | 0.0144375 |
| Qiang_Liujia | Tibetan_Nagqu | 0.0144399 |

| | | |
|---|---|---|
| Tibetan_Amdo_Dalu | Tibetan_Shigatse | 0.0144404 |
| Qiang_Liujia | Han_Suzhou | 0.0144453 |
| Qiang_Liujia | Han_Changzhi | 0.014468 |
| Pumi_Lanping | KHV | 0.0144803 |
| Qiang_Liujia | Han_Linfen | 0.0145068 |
| Qiang_Liujia | Han_Xuchang | 0.0145089 |
| Lahu_Lancang | Han_Yunyang | 0.0145157 |
| Tibetan_Amdo_Dalu | Tibetan_Nagqu | 0.014565 |
| Qiang_Liujia | Han_Laiwu | 0.014572 |
| Qiang_Liujia | Han_Zhoukou | 0.0145961 |
| Qiang_Liujia | Han_Xiamen | 0.0146544 |
| Qiang_Liujia | Han_Fuzhou | 0.0147961 |
| Blang_Menghai | Han_Zhongxian | 0.0148054 |
| Tibetan_Amdo_Dalu | Han_Yongchuan | 0.0148139 |
| Qiang_Xiaoyuan | Tibetan_Shannan | 0.0148178 |
| Pumi_Lanping | Han_Jiangjin | 0.0148374 |
| Qiang_Liujia | Han_Yuncheng | 0.0148912 |
| Tibetan_Amdo_Dalu | Han_Hechuan | 0.0148955 |
| Tibetan_Baima_Guoyuan | Han_Putian | 0.0148993 |
| Qiang_Liujia | Han_Lvliang | 0.0149071 |
| Qiang_Liujia | Han_Xinxiang | 0.0149112 |
| Tibetan_Amdo_Dalu | Han_Shangluo | 0.0149118 |
| Bai_Heqing | Japanese | 0.0149198 |
| Hani_Honghe | Mosuo_Yanyuan | 0.0149276 |
| Qiang_Liujia | Han_Zhangzhou | 0.0149437 |
| Bai_Heqing | China_Lahu | 0.0149887 |
| Pumi_Lanping | Qiang_Matou | 0.0150252 |
| Pumi_Lanping | Han_Hanzhong | 0.0150495 |
| Qiang_Liujia | Han_YulinSX | 0.0150747 |
| Pumi_Lanping | Qiang_Liujia | 0.0151086 |
| Tibetan_Amdo_Dalu | Tibetan_Baima_Wuj | 0.0151382 |
| Wa_Cangyuan | Han_Yubei | 0.0151386 |
| Qiang_Gaoping | Tibetan_Nyingchi | 0.0152442 |
| Qiang_Liujia | Han_Weinan | 0.0152687 |
| Qiang_Liujia | Tibetan_Shigatse | 0.0153546 |
| Tibetan_Amdo_Dalu | Tibetan_Shannan | 0.0153579 |
| Pumi_Lanping | Qiang_Gaoping | 0.0153707 |
| Qiang_Liujia | Han_Lanzhou | 0.015407 |
| Blang_Menghai | Mongola | 0.0154318 |
| Pumi_Lanping | JPT | 0.0154842 |
| Tibetan_Baima_Guoyuan | Han_Zhangzhou | 0.0155869 |
| Qiang_Matou | Tibetan_Shigatse | 0.0155999 |
| Tibetan_Baima_Guoyuan | Han_Sanming | 0.0156997 |
| Qiang_Gaoping | Tibetan_Nagqu | 0.0158137 |
| Qiang_Matou | Tibetan_Shannan | 0.0158152 |
| Hani_Honghe | Han_Putian | 0.0158413 |
| Hani_Honghe | Han_Shangqiu | 0.0158531 |
| Tibetan_Baima_Wujiao | Han_Xianyang | 0.0158551 |
| Hani_Honghe | Han_Baoji | 0.0159033 |
| Hani_Honghe | Han_Kaifeng | 0.015905 |
| Tibetan_Baima_Guoyuan | Han_Xiamen | 0.0159542 |
| Tibetan_Amdo_Dalu | Han_Changshou | 0.0159587 |
| Tibetan_Baima_Guoyuan | Han_Longyan | 0.0159986 |
| Tibetan_Baima_Guoyuan | Han_Nanping | 0.0160324 |
| Tibetan_Baima_Guoyuan | Han_Fuzhou | 0.0160342 |
| Tibetan_Baima_Guoyuan | Han_Ningde | 0.0160498 |
| Pumi_Lanping | Han_Fuling | 0.0160547 |
| Qiang_Gaoping | Tibetan_Shigatse | 0.0160982 |
| Tibetan_Amdo_Dalu | Han_Qiandongnan | 0.0161226 |
| Blang_Menghai | Han_Changshou | 0.0161388 |
| Hani_Honghe | Tibetan_Muli | 0.016199 |
| Wa_Cangyuan | Han_Bishan | 0.0162137 |
| Blang_Menghai | Han_Kaixian | 0.0162511 |
| Wa_Cangyuan | Han_Hechuan | 0.0162646 |
| Tibetan_Baima_Guoyuan | Han_Quanzhou | 0.0162772 |
| Hani_Honghe | Han_Hebi | 0.0162776 |

| | | |
|---|---|---|
| Lahu_Lancang | Han_Tongnan | 0.0162795 |
| Hani_Honghe | Han_Datong | 0.0162961 |
| Pumi_Lanping | Hezhen | 0.0163124 |
| Hani_Honghe | Han_Yangquan | 0.016322 |
| Hani_Honghe | Tibetan_Baima_Wuj | 0.016328 |
| Wa_Cangyuan | Han_Yongchuan | 0.0163553 |
| Hani_Honghe | Han_Luoyang | 0.0163634 |
| Hani_Honghe | Han_Jining | 0.0163717 |
| Hani_Honghe | Han_Zhumadian | 0.0163921 |
| Hani_Honghe | Han_Xinzhou | 0.0164125 |
| Tibetan_Baima_Guoyuan | Han_Xian | 0.0164128 |
| Tibetan_Amdo_Dalu | Tibetan_Nyingchi | 0.0164141 |
| Hani_Honghe | Han_Xinyang | 0.0164281 |
| Qiang_Xiaoyuan | Tibetan_Lhasa | 0.0164386 |
| Hani_Honghe | Han_Pingdingshan | 0.016447 |
| Pumi_Lanping | Han_Kaifeng | 0.0164677 |
| Hani_Honghe | Han_Jinzhong | 0.0164746 |
| Hani_Honghe | Han_Xiamen | 0.016477 |
| Hani_Honghe | Han_Sanming | 0.016483 |
| Pumi_Lanping | Han_Qiannan | 0.0165423 |
| Hani_Honghe | Han_Luohe | 0.016551 |
| Wa_Cangyuan | Mongola | 0.0165552 |
| Tibetan_Amdo_Dalu | Han_Kaixian | 0.0165777 |
| Tibetan_Amdo_Dalu | Han_Wanzhou | 0.0165869 |
| Pumi_Lanping | Han_Jiulongpo | 0.0165905 |
| Blang_Menghai | Han_Wanzhou | 0.0165956 |
| Hani_Honghe | Han_Yuncheng | 0.0166061 |
| Pumi_Lanping | Han_Hebi | 0.0166091 |
| Pumi_Lanping | Han_Yangquan | 0.0166344 |
| Hani_Honghe | Han_Linfen | 0.0166938 |
| Pumi_Lanping | Miao | 0.0166971 |
| Qiang_Liujia | Tibetan_Shannan | 0.0167081 |
| Hani_Honghe | Han_Heze | 0.0167229 |
| Pumi_Lanping | Han_Jincheng | 0.0167321 |
| Pumi_Lanping | Han_Anshun | 0.0167457 |
| Hani_Honghe | Han_Quanzhou | 0.0167519 |
| Hani_Honghe | Han_Taiyuan | 0.0167545 |
| Wa_Cangyuan | Han_Fengdu | 0.016788 |
| Hani_Honghe | Han_Sanmenxia | 0.0168097 |
| Hani_Honghe | Han_Taian | 0.0168249 |
| Pumi_Lanping | Han_Linfen | 0.0168452 |
| Hani_Honghe | Han_Jinan | 0.016874 |
| Hani_Honghe | Han_Jincheng | 0.0169046 |
| Hani_Honghe | Han_Binzhou | 0.0169109 |
| Blang_Menghai | Han_Qianxinan | 0.0169118 |
| Pumi_Lanping | CDX | 0.0169338 |
| Hani_Honghe | Han_Longyan | 0.0169367 |
| Hani_Honghe | Han_Xuchang | 0.0169482 |
| Pumi_Lanping | Han_Xinzhou | 0.0169624 |
| Hani_Honghe | Han_Ningde | 0.0169707 |
| Hani_Honghe | Han_Laiwu | 0.0169751 |
| Pumi_Lanping | Han_Luohe | 0.0169841 |
| Hani_Honghe | Han_Zhangzhou | 0.0169879 |
| Pumi_Lanping | Han_Zhumadian | 0.0170023 |
| Wa_Cangyuan | Han_Nanan | 0.0170093 |
| Hani_Honghe | Han_Nanping | 0.0170102 |
| Hani_Honghe | Han_Jiyuan | 0.0170107 |
| Hani_Honghe | Han_Yantai | 0.0170123 |
| Pumi_Lanping | Han_Jinzhong | 0.0170215 |
| Hani_Honghe | Han_Linyi | 0.0170413 |
| Pumi_Lanping | Han_Sanmenxia | 0.0170496 |
| Hani_Honghe | Han_Jiaozuo | 0.0170519 |
| Hani_Honghe | Han_YulinSX | 0.0170616 |
| Pumi_Lanping | Han_Suzhou | 0.017072 |
| Hani_Honghe | Han_Xinxiang | 0.017086 |
| Hani_Honghe | Han_Anyang | 0.0170902 |

| | | |
|---|---|---|
| Hani_Honghe | Han_Nanyang | 0.0171023 |
| Pumi_Lanping | Han_Taiyuan | 0.0171037 |
| Pumi_Lanping | Han_Yuncheng | 0.0171081 |
| Hani_Honghe | Han_Weihai | 0.0171207 |
| Pumi_Lanping | Han_Datong | 0.0171289 |
| Pumi_Lanping | Han_Baoji | 0.0171415 |
| Hani_Honghe | Han_Dongying | 0.0171425 |
| Tibetan_Baima_Wujiao | Han_Jiangjin | 0.0171583 |
| Hani_Honghe | Han_Dezhou | 0.0171762 |
| Hani_Honghe | Han_Liaocheng | 0.0172411 |
| Pumi_Lanping | Han_Lvliang | 0.0172486 |
| Hani_Honghe | Han_Suzhou | 0.0172808 |
| Hani_Honghe | Han_Lvliang | 0.0172903 |
| Hani_Honghe | Han_Zhengzhou | 0.0173124 |
| Tibetan_Baima_Guoyuan | Han_Nanchong | 0.0173146 |
| Hani_Honghe | Han_Zibo | 0.017315 |
| Tibetan_Baima_Wujiao | Han_Hanzhong | 0.0173154 |
| Qiang_Liujia | Han_Nanchong | 0.0173258 |
| Qiang_Gaoping | Tibetan_Shannan | 0.0173315 |
| Hani_Honghe | Han_Changzhi | 0.017332 |
| Tibetan_Amdo_Dalu | Han_Ankang | 0.0173336 |
| Pumi_Lanping | Han_Binzhou | 0.0173592 |
| Pumi_Lanping | Han_YulinSX | 0.0173618 |
| Blang_Menghai | Dai | 0.0173778 |
| Wa_Cangyuan | Han_Qiandongnan | 0.0173788 |
| Hani_Honghe | Han_Weifang | 0.0173809 |
| Pumi_Lanping | Han_Jining | 0.0173955 |
| Hani_Honghe | Han_Fuzhou | 0.0174063 |
| Pumi_Lanping | Han_Shangqiu | 0.0174078 |
| Pumi_Lanping | Han_Luoyang | 0.0174257 |
| Pumi_Lanping | Han_Anyang | 0.017449 |
| Hani_Honghe | Han_Zhoukou | 0.0174613 |
| Hani_Honghe | Tibetan_Baima_Guo | 0.0174617 |
| Hani_Honghe | Han_Rizhao | 0.0174666 |
| Hani_Honghe | Han_Puyang | 0.0174943 |
| Pumi_Lanping | Han_Jiaozuo | 0.0175193 |
| Hani_Honghe | Han_Zaozhuang | 0.0175246 |
| Pumi_Lanping | Han_Pingdingshan | 0.0175248 |
| Pumi_Lanping | Han_Linyi | 0.0175254 |
| Hani_Honghe | Hezhen | 0.0175306 |
| Qiang_Liujia | Han_Xian | 0.0175659 |
| Wa_Cangyuan | Han_Zhongxian | 0.0175931 |
| Hani_Honghe | Tibetan_Chamdo | 0.0175984 |
| Pumi_Lanping | Han_Zhengzhou | 0.0176615 |
| Tibetan_Baima_Guoyuan | Tibetan_Amdo_Dalu | 0.017683 |
| Pumi_Lanping | Han_Puyang | 0.0176968 |
| Pumi_Lanping | Han_Laiwu | 0.0177202 |
| Pumi_Lanping | Han_Xuchang | 0.0177298 |
| Pumi_Lanping | Han_Weifang | 0.0177389 |
| Hani_Honghe | She | 0.0177553 |
| Tibetan_Amdo_Dalu | Han_Yuzhong | 0.017764 |
| Pumi_Lanping | Han_Taian | 0.0177658 |
| Bai_Heqing | Tibetan_Zase | 0.0177677 |
| Pumi_Lanping | Han_Xinxiang | 0.0177679 |
| Pumi_Lanping | Han_Qingdao | 0.0177754 |
| Pumi_Lanping | Han_Jinan | 0.0177796 |
| Tibetan_Amdo_Dalu | Han_Shapingba | 0.0177952 |
| Pumi_Lanping | Han_Liaocheng | 0.0178097 |
| Pumi_Lanping | Han_Changzhi | 0.0178315 |
| Pumi_Lanping | Han_Dongying | 0.0178484 |
| Pumi_Lanping | Han_Jiyuan | 0.0178707 |
| Pumi_Lanping | Han_Weihai | 0.0178958 |
| Pumi_Lanping | Han_Dezhou | 0.0178992 |
| Blang_Menghai | Han_Banan | 0.0179144 |
| Pumi_Lanping | Han_Zhoukou | 0.0179203 |
| Hani_Honghe | Han_Qingdao | 0.0179403 |

| | | |
|---|---|---|
| Pumi_Lanping | Han_Yantai | 0.0180076 |
| Pumi_Lanping | Han_Nanyang | 0.018012 |
| Hani_Honghe | Han_Weinan | 0.0180441 |
| Qiang_Matou | Tibetan_Lhasa | 0.0180897 |
| Pumi_Lanping | Han_Rizhao | 0.0181077 |
| Tibetan_Amdo_Dalu | Han_Banan | 0.0181223 |
| Hani_Honghe | Han_Lanzhou | 0.0181338 |
| Blang_Menghai | Han_Yuzhong | 0.0181372 |
| Pumi_Lanping | Han_Heze | 0.0181857 |
| Pumi_Lanping | Han_Zaozhuang | 0.0182453 |
| Tibetan_Amdo_Dalu | Han_Tongliang | 0.0182778 |
| Pumi_Lanping | Han_Lanzhou | 0.0183103 |
| Tibetan_Amdo_Dalu | Han_Xianyang | 0.0183326 |
| Tibetan_Amdo_Dalu | Tibetan_Lhasa | 0.0183407 |
| Blang_Menghai | Han_Shapingba | 0.0183792 |
| Hani_Honghe | China_Lahu | 0.0183928 |
| Pumi_Lanping | Han_Zibo | 0.0184269 |
| Qiang_Liujia | Tibetan_Lhasa | 0.0185227 |
| Tibetan_Baima_Wujiao | Han_Qiannan | 0.0185445 |
| Tibetan_Baima_Wujiao | Han_Kaifeng | 0.0185677 |
| Tibetan_Baima_Wujiao | Han_Anshun | 0.0185825 |
| Blang_Menghai | Han_Tongliang | 0.0186006 |
| Tibetan_Baima_Wujiao | Han_Jiulongpo | 0.0186079 |
| Pumi_Lanping | Han_Weinan | 0.0186234 |
| Tibetan_Baima_Wujiao | Han_Yangquan | 0.0186768 |
| Pumi_Lanping | Han_Xinyang | 0.0186773 |
| Qiang_Gaoping | Tibetan_Lhasa | 0.018698 |
| Tibetan_Baima_Wujiao | Han_Fuling | 0.0186993 |
| Tibetan_Baima_Wujiao | Han_Linfen | 0.0187475 |
| Blang_Menghai | Tujia | 0.0188006 |
| Tibetan_Amdo_Dalu | Han_Qianxinan | 0.0188252 |
| Hani_Honghe | Tibetan_Dujiangyan | 0.0188476 |
| Tibetan_Baima_Wujiao | Han_Hebi | 0.0188766 |
| Hani_Honghe | Han | 0.0188781 |
| Tibetan_Baima_Wujiao | Han_Taiyuan | 0.0188886 |
| Tibetan_Baima_Guoyuan | Tibetan_Zase | 0.0188924 |
| Tibetan_Baima_Wujiao | Han_Jinzhong | 0.0189379 |
| Tibetan_Baima_Wujiao | Han_Datong | 0.0189881 |
| Bai_Heqing | Tibetan_Amdo_Dalu | 0.0189888 |
| Wa_Cangyuan | Han_Kaixian | 0.0190367 |
| Tibetan_Baima_Wujiao | Han_Jiaozuo | 0.0190691 |
| Pumi_Lanping | Tibetan_Zase | 0.0190996 |
| Tibetan_Baima_Wujiao | Han_Baoji | 0.0191701 |
| Tibetan_Baima_Wujiao | Han_Suzhou | 0.0192021 |
| Wa_Cangyuan | Blang_Menghai | 0.0192077 |
| Tibetan_Baima_Wujiao | Han_Xinzhou | 0.0192112 |
| Hani_Honghe | Han_Nanchong | 0.0192194 |
| Wa_Cangyuan | Han_Changshou | 0.0192387 |
| Tibetan_Baima_Wujiao | Han_Jincheng | 0.019242 |
| Pumi_Lanping | Tibetan_Amdo_Dalu | 0.0192521 |
| Tibetan_Baima_Wujiao | Han_Zhumadian | 0.0192675 |
| Tibetan_Baima_Wujiao | Han_Xuchang | 0.0193271 |
| Tibetan_Baima_Wujiao | Han_Jining | 0.0193963 |
| Tibetan_Baima_Wujiao | Han_Luoyang | 0.0194039 |
| Tibetan_Baima_Wujiao | Han_Luohe | 0.01943 |
| Tibetan_Baima_Wujiao | Han_Anyang | 0.0194573 |
| Tibetan_Baima_Wujiao | Han_Liaocheng | 0.0194796 |
| Tibetan_Baima_Wujiao | Han_Dongying | 0.0195137 |
| Wa_Cangyuan | Han_Qianxinan | 0.0195532 |
| Tibetan_Baima_Wujiao | Han_Xinxiang | 0.0195594 |
| Tibetan_Baima_Wujiao | Han_Heze | 0.0195626 |
| Wa_Cangyuan | Han_Wanzhou | 0.0195694 |
| Tibetan_Baima_Wujiao | Han_YulinSX | 0.0195724 |
| Tibetan_Baima_Wujiao | Han_Yuncheng | 0.0195775 |
| Tibetan_Baima_Wujiao | Han_Sanmenxia | 0.0195814 |
| Tibetan_Baima_Wujiao | Han_Lvliang | 0.0196588 |

| | | |
|---|---|---|
| Tibetan_Baima_Wujiao | Han_Changzhi | 0.0196819 |
| Tibetan_Baima_Wujiao | Han_Shangqiu | 0.0196921 |
| Tibetan_Baima_Wujiao | Han_Jiyuan | 0.0198297 |
| Tibetan_Baima_Wujiao | Han_Linyi | 0.0198695 |
| Tibetan_Baima_Wujiao | Han_Rizhao | 0.019895 |
| Tibetan_Baima_Wujiao | Han_Binzhou | 0.0199116 |
| Hani_Honghe | Tibetan_Shigatse | 0.0199316 |
| Tibetan_Baima_Wujiao | Han_Zhengzhou | 0.0199347 |
| Tibetan_Baima_Wujiao | Han_Weifang | 0.0199407 |
| Tibetan_Baima_Wujiao | Han_Puyang | 0.0199515 |
| Tibetan_Baima_Wujiao | Han_Jinan | 0.0199597 |
| Pumi_Lanping | Oroqen | 0.0200097 |
| Blang_Menghai | Han_Yanan | 0.020015 |
| Hani_Honghe | Pumi_Lanping | 0.0200168 |
| Hani_Honghe | Tibetan_Nagqu | 0.020021 |
| Blang_Menghai | Naxi | 0.0200259 |
| Tibetan_Baima_Wujiao | Han_Zaozhuang | 0.0200389 |
| Tibetan_Amdo_Dalu | Tibetan_Zase | 0.0200428 |
| Wa_Cangyuan | Han_Banan | 0.0200917 |
| Blang_Menghai | Qiang_Xiaoyuan | 0.0201065 |
| Tibetan_Baima_Wujiao | Han_Yantai | 0.0201189 |
| Tibetan_Baima_Wujiao | Han_Zibo | 0.0201366 |
| Tibetan_Baima_Wujiao | Han_Laiwu | 0.0201445 |
| Tibetan_Baima_Wujiao | Han_Qingdao | 0.0201732 |
| Tibetan_Baima_Wujiao | Han_Taian | 0.0202024 |
| Blang_Menghai | Han_Jiangjin | 0.0202095 |
| Tibetan_Baima_Wujiao | Han_Weinan | 0.0202859 |
| Tibetan_Baima_Wujiao | Han_Zhoukou | 0.020301 |
| Lahu_Lancang | Qiang_Matou | 0.0203028 |
| Pumi_Lanping | Han_Putian | 0.0203224 |
| Tibetan_Baima_Wujiao | Han_Dezhou | 0.020334 |
| Tibetan_Baima_Wujiao | Han_Pingdingshan | 0.0203704 |
| Tibetan_Baima_Wujiao | Han_Nanyang | 0.0203713 |
| Blang_Menghai | Qiang_Matou | 0.0204134 |
| Tibetan_Baima_Wujiao | Han_Lanzhou | 0.0204137 |
| Tibetan_Baima_Wujiao | Han_Weihai | 0.0204266 |
| Hani_Honghe | Blang_Menghai | 0.0204535 |
| Wa_Cangyuan | Han_Yuzhong | 0.0204674 |
| Wa_Cangyuan | Tujia | 0.0204836 |
| Wa_Cangyuan | Han_Tongliang | 0.0205049 |
| Hani_Honghe | Han_Xian | 0.0205363 |
| Tibetan_Amdo_Dalu | Han_Hanzhong | 0.020538 |
| Blang_Menghai | Bai_Heqing | 0.0205651 |
| Wa_Cangyuan | Bai_Heqing | 0.0207109 |
| Qiang_Xiaoyuan | Tibetan_Amdo_Dalu | 0.0207153 |
| Pumi_Lanping | Han_Zhangzhou | 0.0207486 |
| Tibetan_Baima_Wujiao | Han_Xinyang | 0.0208192 |
| Wa_Cangyuan | Han_Shapingba | 0.0208499 |
| Lahu_Lancang | Qiang_Xiaoyuan | 0.0208729 |
| Pumi_Lanping | Han_Xiamen | 0.0209327 |
| Blang_Menghai | Miao | 0.0209663 |
| Pumi_Lanping | Han_Sanming | 0.0209901 |
| Bai_Heqing | Lahu_Lancang | 0.0209979 |
| Pumi_Lanping | Han_Quanzhou | 0.0210152 |
| Blang_Menghai | Yi | 0.0210631 |
| Tibetan_Amdo_Dalu | Han_Jiangjin | 0.021085 |
| Pumi_Lanping | Han_Ningde | 0.021109 |
| Pumi_Lanping | Han_Xian | 0.0211424 |
| Hani_Honghe | Han_Taiwan | 0.0211794 |
| Lahu_Lancang | Qiang_Gaoping | 0.0212155 |
| Tibetan_Amdo_Dalu | Han_Kaifeng | 0.0212276 |
| Lahu_Lancang | Han_Dianjiang | 0.0212324 |
| Blang_Menghai | Han_Ankang | 0.0213655 |
| Tibetan_Amdo_Dalu | Qiang_Matou | 0.0213758 |
| Wa_Cangyuan | Hani_Honghe | 0.0213903 |
| Tibetan_Amdo_Dalu | Han_Hebi | 0.0214149 |

| | | |
|---|---|---|
| Tibetan_Amdo_Dalu | Han_YulinSX | 0.0214166 |
| Pumi_Lanping | Han_Fuzhou | 0.0214488 |
| Tibetan_Amdo_Dalu | Han_Yangquan | 0.0214525 |
| Pumi_Lanping | Cambodian | 0.0214601 |
| Pumi_Lanping | Han_Longyan | 0.0214833 |
| Tibetan_Amdo_Dalu | Han_Jinzhong | 0.0214861 |
| Tibetan_Amdo_Dalu | Han_Baoji | 0.0214992 |
| Wa_Cangyuan | Dai | 0.0215106 |
| Tibetan_Amdo_Dalu | Han_Xinzhou | 0.0215347 |
| Tibetan_Amdo_Dalu | Han_Linfen | 0.0215904 |
| Pumi_Lanping | Han_Nanping | 0.0215915 |
| Tibetan_Amdo_Dalu | Han_Suzhou | 0.0216116 |
| Hani_Honghe | Tibetan_Shannan | 0.0216533 |
| Tibetan_Amdo_Dalu | Han_Datong | 0.02167 |
| Blang_Menghai | Xibo | 0.0217632 |
| Wa_Cangyuan | Qiang_Xiaoyuan | 0.0217981 |
| Tibetan_Amdo_Dalu | Han_Jiaozuo | 0.0218042 |
| Tibetan_Amdo_Dalu | Han_Luoyang | 0.0218649 |
| Blang_Menghai | Qiang_Gaoping | 0.0218702 |
| Wa_Cangyuan | Qiang_Matou | 0.0218898 |
| Tibetan_Amdo_Dalu | Han_Lvliang | 0.021897 |
| Wa_Cangyuan | Han_Yanan | 0.0219018 |
| Lahu_Lancang | Han_Liangping | 0.0219214 |
| Qiang_Matou | Tibetan_Zase | 0.0219257 |
| Blang_Menghai | Han_Shangluo | 0.0219344 |
| Hani_Honghe | Tibetan_Nyingchi | 0.0219841 |
| Tibetan_Amdo_Dalu | Qiang_Liujia | 0.0220221 |
| Tibetan_Amdo_Dalu | Han_Fuling | 0.0220236 |
| Tibetan_Amdo_Dalu | Han_Jincheng | 0.0220241 |
| Tibetan_Amdo_Dalu | Han_Zhumadian | 0.0220811 |
| Tibetan_Amdo_Dalu | Han_Yuncheng | 0.0220826 |
| Tibetan_Amdo_Dalu | Han_Jiyuan | 0.0221589 |
| Tibetan_Amdo_Dalu | Han_Puyang | 0.0221744 |
| Tibetan_Baima_Wujiao | Han_Putian | 0.0221847 |
| Blang_Menghai | Han_Anshun | 0.0221976 |
| Pumi_Lanping | Han_Nanchong | 0.0222174 |
| Wa_Cangyuan | Naxi | 0.022272 |
| Tibetan_Amdo_Dalu | Han_Sanmenxia | 0.0222887 |
| Tibetan_Amdo_Dalu | Han_Jining | 0.0222968 |
| Tibetan_Amdo_Dalu | Han_Zhengzhou | 0.0223091 |
| Tibetan_Amdo_Dalu | Han_Taiyuan | 0.0223093 |
| Tibetan_Amdo_Dalu | Han_Pingdingshan | 0.0223111 |
| Hani_Honghe | Oroqen | 0.0223186 |
| Tibetan_Amdo_Dalu | Han_Dongying | 0.0223444 |
| Tibetan_Amdo_Dalu | Han_Zaozhuang | 0.0223837 |
| Tibetan_Amdo_Dalu | Han_Anyang | 0.0224037 |
| Tibetan_Amdo_Dalu | Han_Zibo | 0.0224113 |
| Tibetan_Amdo_Dalu | Han_Zhoukou | 0.0224117 |
| Tibetan_Amdo_Dalu | Han_Shangqiu | 0.0224285 |
| Tibetan_Amdo_Dalu | Qiang_Gaoping | 0.0224379 |
| Tibetan_Amdo_Dalu | Han_Changzhi | 0.0224471 |
| Tibetan_Amdo_Dalu | Han_Liaocheng | 0.022454 |
| Lahu_Lancang | Qiang_Liujia | 0.0224543 |
| Tibetan_Amdo_Dalu | Han_Xuchang | 0.0224679 |
| Tibetan_Amdo_Dalu | Han_Luohe | 0.0224794 |
| Blang_Menghai | Han_Qiannan | 0.0225107 |
| Tibetan_Amdo_Dalu | Han_Heze | 0.0225367 |
| Blang_Menghai | Han_Jiulongpo | 0.022561 |
| Tibetan_Baima_Wujiao | Han_Zhangzhou | 0.0225637 |
| Tibetan_Amdo_Dalu | Han_Jiulongpo | 0.022594 |
| Tibetan_Amdo_Dalu | Han_Laiwu | 0.0225966 |
| Wa_Cangyuan | Xibo | 0.0226308 |
| Tibetan_Amdo_Dalu | Han_Qingdao | 0.0226639 |
| Tibetan_Amdo_Dalu | Han_Linyi | 0.0226991 |
| Wa_Cangyuan | Yi | 0.0227152 |
| Blang_Menghai | Qiang_Liujia | 0.0227187 |

| | | |
|---|---|---|
| Tibetan_Amdo_Dalu | Han_Lanzhou | 0.0227214 |
| Tibetan_Amdo_Dalu | Han_Rizhao | 0.0227423 |
| Tibetan_Amdo_Dalu | Han_Xinxiang | 0.0227482 |
| Blang_Menghai | Han_Fuling | 0.0227661 |
| Tibetan_Baima_Wujiao | Han_Sanming | 0.0227765 |
| Tibetan_Amdo_Dalu | Han_Weihai | 0.022798 |
| Tibetan_Amdo_Dalu | Han_Anshun | 0.0228024 |
| Tibetan_Amdo_Dalu | Han_Binzhou | 0.022813 |
| Tibetan_Amdo_Dalu | Han_Jinan | 0.0228348 |
| Tibetan_Amdo_Dalu | Han_Weifang | 0.0228553 |
| Tibetan_Amdo_Dalu | Han_Nanyang | 0.0228574 |
| Tibetan_Amdo_Dalu | Han_Taian | 0.0228645 |
| Tibetan_Amdo_Dalu | Han_Qiannan | 0.0228661 |
| Pumi_Lanping | She | 0.022867 |
| Lahu_Lancang | Han_Rongchang | 0.0228913 |
| Tibetan_Baima_Wujiao | Han_Longyan | 0.0229317 |
| Pumi_Lanping | Han | 0.0229466 |
| Tibetan_Baima_Wujiao | Han_Xian | 0.0229776 |
| Tibetan_Amdo_Dalu | Han_Yantai | 0.0230068 |
| Tibetan_Amdo_Dalu | Han_Dezhou | 0.0230115 |
| Tibetan_Baima_Wujiao | Han_Xiamen | 0.0230652 |
| Hani_Honghe | Lahu_Lancang | 0.0230717 |
| Blang_Menghai | NorthernHan | 0.0230766 |
| Qiang_Xiaoyuan | Tibetan_Zase | 0.023181 |
| Wa_Cangyuan | Han_Ankang | 0.0231814 |
| Tibetan_Baima_Wujiao | Han_Nanping | 0.023229 |
| Tibetan_Baima_Wujiao | Han_Ningde | 0.0232505 |
| Tibetan_Amdo_Dalu | Han_Xinyang | 0.0232519 |
| Blang_Menghai | Tu | 0.0233116 |
| Tibetan_Amdo_Dalu | Han_Weinan | 0.0233204 |
| Wa_Cangyuan | Qiang_Gaoping | 0.0233297 |
| Tibetan_Baima_Wujiao | Han_Quanzhou | 0.0233485 |
| Tibetan_Baima_Wujiao | Han_Fuzhou | 0.0234429 |
| Wa_Cangyuan | Han_Jiangjin | 0.0234635 |
| Blang_Menghai | China_Lahu | 0.0235221 |
| Wa_Cangyuan | Miao | 0.0235615 |
| Tibetan_Baima_Wujiao | Tibetan_Zase | 0.0236472 |
| Hani_Honghe | Tibetan_Lhasa | 0.0236573 |
| Blang_Menghai | Yi_Xichang | 0.0237252 |
| Wa_Cangyuan | JPT | 0.0238311 |
| Blang_Menghai | Han_Hanzhong | 0.0240126 |
| Wa_Cangyuan | Qiang_Liujia | 0.0240432 |
| Tibetan_Baima_Wujiao | Han_Nanchong | 0.0241506 |
| Wa_Cangyuan | Han_Shangluo | 0.0241547 |
| Blang_Menghai | JPT | 0.0242359 |
| Pumi_Lanping | Dai | 0.0242819 |
| Wa_Cangyuan | Han_Anshun | 0.024591 |
| Wa_Cangyuan | Tu | 0.0246117 |
| Blang_Menghai | Daur | 0.024641 |
| Wa_Cangyuan | Han_Fuling | 0.0247384 |
| Wa_Cangyuan | NorthernHan | 0.0247395 |
| Wa_Cangyuan | Han_Qiannan | 0.0247787 |
| Pumi_Lanping | Han_Taiwan | 0.024908 |
| Blang_Menghai | Han_Xianyang | 0.0249143 |
| Lahu_Lancang | Han_Qijiang | 0.0250607 |
| Hani_Honghe | Japanese | 0.0253683 |
| Wa_Cangyuan | Han_Jiulongpo | 0.0254146 |
| Tibetan_Amdo_Dalu | Han_Xian | 0.0256024 |
| Wa_Cangyuan | Yi_Xichang | 0.0256245 |
| Lahu_Lancang | Yi_Xichang | 0.025632 |
| Wa_Cangyuan | Daur | 0.0258957 |
| Blang_Menghai | Mosuo_Yanyuan | 0.0260454 |
| Pumi_Lanping | Japanese | 0.0261208 |
| Blang_Menghai | Han_Xiamen | 0.0261856 |
| Blang_Menghai | Han_Putian | 0.0263368 |
| Wa_Cangyuan | Han_Hanzhong | 0.026353 |

| | | |
|---|---|---|
| Blang_Menghai | Lahu_Lancang | 0.0264618 |
| Blang_Menghai | Han_Quanzhou | 0.0266006 |
| Blang_Menghai | Han_Ningde | 0.0266495 |
| Blang_Menghai | Han_Zhangzhou | 0.0266597 |
| Qiang_Gaoping | Tibetan_Zase | 0.0266623 |
| Tibetan_Amdo_Dalu | Han_Putian | 0.0267582 |
| Wa_Cangyuan | Mosuo_Yanyuan | 0.0269841 |
| Blang_Menghai | Han_Longyan | 0.0270191 |
| Tibetan_Amdo_Dalu | Han_Quanzhou | 0.0270496 |
| Blang_Menghai | Han_Fuzhou | 0.0271756 |
| Bai_Heqing | Uyghur | 0.0271791 |
| Wa_Cangyuan | Han_Xianyang | 0.0272047 |
| Blang_Menghai | Han_Sanming | 0.0272241 |
| Wa_Cangyuan | China_Lahu | 0.0272652 |
| Tibetan_Amdo_Dalu | Han_Longyan | 0.027275 |
| Tibetan_Amdo_Dalu | Han_Xiamen | 0.0273109 |
| Tibetan_Amdo_Dalu | Han_Zhangzhou | 0.0273463 |
| Lahu_Lancang | Han_Yubei | 0.0273678 |
| Tibetan_Amdo_Dalu | Han_Nanchong | 0.0273935 |
| Tibetan_Amdo_Dalu | Han_Ningde | 0.0274358 |
| Tibetan_Amdo_Dalu | Han_Fuzhou | 0.0274405 |
| Tibetan_Amdo_Dalu | Han_Nanping | 0.0275006 |
| Tibetan_Amdo_Dalu | Han_Sanming | 0.0275129 |
| Blang_Menghai | Tibetan_Muli | 0.0276054 |
| Blang_Menghai | Han_Nanping | 0.0276324 |
| Lahu_Lancang | Mosuo_Yanyuan | 0.0276389 |
| Blang_Menghai | Han_Xinzhou | 0.0278811 |
| Blang_Menghai | She | 0.0279059 |
| Wa_Cangyuan | Tibetan_Muli | 0.027968 |
| Blang_Menghai | Han_Baoji | 0.0280312 |
| Blang_Menghai | Han_Kaifeng | 0.0280814 |
| Lahu_Lancang | Han_Yongchuan | 0.0280987 |
| Blang_Menghai | Han_Luohe | 0.0281461 |
| Blang_Menghai | Han_Zhumadian | 0.0281877 |
| Lahu_Lancang | Han_Qiandongnan | 0.0282635 |
| Blang_Menghai | Han | 0.0282655 |
| Blang_Menghai | Han_Xinyang | 0.0283125 |
| Blang_Menghai | Han_Yangquan | 0.028349 |
| Blang_Menghai | Han_Luoyang | 0.0283558 |
| Blang_Menghai | Han_Shangqiu | 0.0283851 |
| Wa_Cangyuan | Han_Xiamen | 0.0283856 |
| Lahu_Lancang | Han_Bishan | 0.0284056 |
| Pumi_Lanping | China_Lahu | 0.0284106 |
| Wa_Cangyuan | Han_Putian | 0.0285258 |
| Hani_Honghe | Tibetan_Amdo_Dalu | 0.0285334 |
| Blang_Menghai | Han_Datong | 0.0285969 |
| Blang_Menghai | Tibetan_Baima_Wuj | 0.0286456 |
| Wa_Cangyuan | Han_Quanzhou | 0.0286658 |
| Lahu_Lancang | Han_Hechuan | 0.0286941 |
| Wa_Cangyuan | Han_Zhangzhou | 0.0287228 |
| Blang_Menghai | Han_Hebi | 0.0287318 |
| Wa_Cangyuan | Han_Sanming | 0.0287837 |
| Blang_Menghai | Han_Taiyuan | 0.0287941 |
| Hani_Honghe | Tibetan_Zase | 0.0288065 |
| Blang_Menghai | Hezhen | 0.0288367 |
| Blang_Menghai | Han_Jining | 0.028849 |
| Blang_Menghai | Han_Dongying | 0.0288672 |
| Blang_Menghai | Han_Linfen | 0.02887 |
| Lahu_Lancang | Tibetan_Muli | 0.0288898 |
| Blang_Menghai | Han_Jinan | 0.0288977 |
| Wa_Cangyuan | Tibetan_Baima_Wuj | 0.0289101 |
| Blang_Menghai | Han_Pingdingshan | 0.0289334 |
| Blang_Menghai | Han_Xuchang | 0.02896 |
| Blang_Menghai | Han_Nanyang | 0.0289877 |
| Lahu_Lancang | Han_Nanan | 0.028993 |
| Wa_Cangyuan | Han_Fuzhou | 0.0290159 |

| | | |
|---|---|---|
| Blang_Menghai | Han_Jinzhong | 0.0290188 |
| Blang_Menghai | Han_Jiaozuo | 0.0290555 |
| Wa_Cangyuan | Han_Ningde | 0.0290894 |
| Blang_Menghai | Han_Linyi | 0.0291276 |
| Blang_Menghai | Han_Sanmenxia | 0.0291495 |
| Wa_Cangyuan | Han_Longyan | 0.0291757 |
| Lahu_Lancang | Cambodian | 0.0292025 |
| Wa_Cangyuan | Tibetan_Baima_Guo | 0.0292284 |
| Blang_Menghai | Han_Suzhou | 0.0292521 |
| Blang_Menghai | Han_Zhoukou | 0.0292945 |
| Blang_Menghai | Han_Yuncheng | 0.0293042 |
| Blang_Menghai | Han_YulinSX | 0.0293231 |
| Lahu_Lancang | Han_Zhongxian | 0.0293524 |
| Wa_Cangyuan | Lahu_Lancang | 0.0293553 |
| Blang_Menghai | Han_Weihai | 0.0293834 |
| Blang_Menghai | Han_Lanzhou | 0.0294018 |
| Blang_Menghai | Han_Lvliang | 0.0294291 |
| Blang_Menghai | Han_Taian | 0.0294334 |
| Blang_Menghai | Han_Zhengzhou | 0.0294439 |
| Blang_Menghai | Han_Jincheng | 0.0294635 |
| Lahu_Lancang | Mongola | 0.0294953 |
| Lahu_Lancang | Han_Fengdu | 0.0294985 |
| Blang_Menghai | Han_Jiyuan | 0.0295218 |
| Wa_Cangyuan | Han_Baoji | 0.0295462 |
| Blang_Menghai | Han_Binzhou | 0.0295487 |
| Blang_Menghai | Han_Liaocheng | 0.0296063 |
| Blang_Menghai | Han_Nanchong | 0.0296101 |
| Blang_Menghai | Han_Anyang | 0.0296295 |
| Blang_Menghai | Han_Yantai | 0.0296442 |
| Blang_Menghai | Han_Puyang | 0.0296723 |
| Blang_Menghai | Han_Zibo | 0.0297115 |
| Blang_Menghai | Han_Xinxiang | 0.0297236 |
| Blang_Menghai | Han_Laiwu | 0.0297536 |
| Lahu_Lancang | Tibetan_Baima_Wuj | 0.029762 |
| Blang_Menghai | Tibetan_Chamdo | 0.0298013 |
| Wa_Cangyuan | Han_Nanping | 0.0298482 |
| Blang_Menghai | Han_Heze | 0.0298521 |
| Wa_Cangyuan | Han_Xinzhou | 0.0298887 |
| Blang_Menghai | Han_Changzhi | 0.0298982 |
| Blang_Menghai | Tibetan_Baima_Guo | 0.0299022 |
| Wa_Cangyuan | She | 0.0299566 |
| Blang_Menghai | Han_Dezhou | 0.030006 |
| Blang_Menghai | Han_Weifang | 0.030064 |
| Blang_Menghai | Han_Zaozhuang | 0.030076 |
| Blang_Menghai | Han_Qingdao | 0.0301971 |
| Blang_Menghai | Han_Weinan | 0.0302539 |
| Blang_Menghai | Han_Rizhao | 0.0302746 |
| Wa_Cangyuan | Han_Shangqiu | 0.0302821 |
| Wa_Cangyuan | Han_Jining | 0.0302906 |
| Wa_Cangyuan | Han_Luohe | 0.0303202 |
| Wa_Cangyuan | Han_Xinyang | 0.0303456 |
| Wa_Cangyuan | Han | 0.0304183 |
| Wa_Cangyuan | Han_Hebi | 0.0304508 |
| Wa_Cangyuan | Tibetan_Shigatse | 0.0304571 |
| Lahu_Lancang | Han_Kaixian | 0.030469 |
| Wa_Cangyuan | Tibetan_Chamdo | 0.0305051 |
| Wa_Cangyuan | Han_Zhumadian | 0.0305554 |
| Wa_Cangyuan | Han_Kaifeng | 0.0306085 |
| Blang_Menghai | Tibetan_Shigatse | 0.0306125 |
| Wa_Cangyuan | Han_Luoyang | 0.0306652 |
| Wa_Cangyuan | Han_Linfen | 0.0306783 |
| Blang_Menghai | Tibetan_Nagqu | 0.0307059 |
| Lahu_Lancang | Han_Changshou | 0.0307235 |
| Wa_Cangyuan | Han_Jinzhong | 0.0307525 |
| Wa_Cangyuan | Han_Jinan | 0.0307571 |
| Wa_Cangyuan | Han_Taiyuan | 0.0307683 |

| | | |
|---|---|---|
| Wa_Cangyuan | Han_Datong | 0.0307732 |
| Lahu_Lancang | Tibetan_Baima_Guo | 0.0307733 |
| Wa_Cangyuan | Han_Jiaozuo | 0.0307899 |
| Wa_Cangyuan | Han_Anyang | 0.0307958 |
| Qiang_Liujia | Tibetan_Zase | 0.0307972 |
| Wa_Cangyuan | Han_Heze | 0.0308268 |
| Wa_Cangyuan | Han_Jincheng | 0.0309087 |
| Wa_Cangyuan | Han_Dongying | 0.0309198 |
| Blang_Menghai | Han_Taiwan | 0.0309208 |
| Wa_Cangyuan | Han_Liaocheng | 0.0309258 |
| Wa_Cangyuan | Han_Pingdingshan | 0.0309521 |
| Wa_Cangyuan | Han_Yangquan | 0.030959 |
| Wa_Cangyuan | Han_Nanyang | 0.0309596 |
| Wa_Cangyuan | Han_Taian | 0.0309686 |
| Wa_Cangyuan | Han_Yuncheng | 0.0309772 |
| Wa_Cangyuan | Han_Lanzhou | 0.030985 |
| Wa_Cangyuan | Tibetan_Dujiangyan | 0.0309954 |
| Wa_Cangyuan | Han_Weihai | 0.0310123 |
| Pumi_Lanping | Blang_Menghai | 0.0310282 |
| Wa_Cangyuan | Han_Linyi | 0.0310473 |
| Wa_Cangyuan | Han_Jiyuan | 0.0310642 |
| Wa_Cangyuan | Han_Sanmenxia | 0.031107 |
| Wa_Cangyuan | Han_YulinSX | 0.0311157 |
| Wa_Cangyuan | Han_Suzhou | 0.0311468 |
| Wa_Cangyuan | Han_Xuchang | 0.0311925 |
| Wa_Cangyuan | Han_Puyang | 0.031211 |
| Wa_Cangyuan | Han_Zhengzhou | 0.0312434 |
| Wa_Cangyuan | Han_Zhoukou | 0.0312485 |
| Lahu_Lancang | Tibetan_Chamdo | 0.0312595 |
| Wa_Cangyuan | Hezhen | 0.0312831 |
| Wa_Cangyuan | Pumi_Lanping | 0.0313272 |
| Wa_Cangyuan | Han_Binzhou | 0.0313415 |
| Wa_Cangyuan | Han_Yantai | 0.0314093 |
| Wa_Cangyuan | Han_Xinxiang | 0.0314625 |
| Wa_Cangyuan | Han_Zaozhuang | 0.0314713 |
| Wa_Cangyuan | Han_Laiwu | 0.03148 |
| Wa_Cangyuan | Han_Zibo | 0.0314865 |
| Wa_Cangyuan | Han_Dezhou | 0.0315541 |
| Wa_Cangyuan | Han_Nanchong | 0.0315965 |
| Wa_Cangyuan | Han_Lvliang | 0.0316268 |
| Wa_Cangyuan | Han_Changzhi | 0.0316297 |
| Lahu_Lancang | Han_Wanzhou | 0.0317832 |
| Wa_Cangyuan | Han_Weifang | 0.0318049 |
| Wa_Cangyuan | Han_Rizhao | 0.031832 |
| Blang_Menghai | Tibetan_Dujiangyan | 0.0318748 |
| Wa_Cangyuan | Han_Qingdao | 0.0319253 |
| Wa_Cangyuan | Han_Weinan | 0.0319628 |
| Lahu_Lancang | Tibetan_Shigatse | 0.0320039 |
| Lahu_Lancang | Han_Qianxinan | 0.0320467 |
| Lahu_Lancang | Han_Banan | 0.0320472 |
| Blang_Menghai | Han_Xian | 0.0323833 |
| Lahu_Lancang | Han_Shapingba | 0.0325116 |
| Bai_Heqing | Hazara | 0.0326072 |
| Wa_Cangyuan | Tibetan_Nagqu | 0.0326212 |
| Lahu_Lancang | Tibetan_Dujiangyan | 0.0327348 |
| Lahu_Lancang | Han_Yuzhong | 0.0328691 |
| Blang_Menghai | Tibetan_Shannan | 0.0329757 |
| Lahu_Lancang | Han_Tongliang | 0.033038 |
| Pumi_Lanping | Lahu_Lancang | 0.0331533 |
| Lahu_Lancang | Tujia | 0.0333192 |
| Wa_Cangyuan | Han_Taiwan | 0.0333792 |
| Blang_Menghai | Tibetan_Nyingchi | 0.0334115 |
| Wa_Cangyuan | Tibetan_Shannan | 0.0334395 |
| Lahu_Lancang | Dai | 0.0335787 |
| Lahu_Lancang | Han_Yanan | 0.0337683 |
| Wa_Cangyuan | Han_Xian | 0.0339122 |

| | | |
|---|---|---|
| Lahu_Lancang | Tibetan_Nagqu | 0.0341028 |
| Blang_Menghai | Tibetan_Lhasa | 0.0342448 |
| Blang_Menghai | Oroqen | 0.034367 |
| Lahu_Lancang | Han_Shangluo | 0.0347499 |
| Bai_Heqing | Yakut | 0.0347886 |
| Wa_Cangyuan | Oroqen | 0.0353879 |
| Hani_Honghe | Uyghur | 0.0354004 |
| Pumi_Lanping | Uyghur | 0.0354249 |
| Lahu_Lancang | Han_Ankang | 0.0355078 |
| Lahu_Lancang | Yi | 0.0355213 |
| Lahu_Lancang | Han_Jiangjin | 0.03557 |
| Lahu_Lancang | Tibetan_Shannan | 0.0356177 |
| Wa_Cangyuan | Tibetan_Nyingchi | 0.0356933 |
| Lahu_Lancang | Han_Anshun | 0.0358152 |
| Lahu_Lancang | Naxi | 0.035912 |
| Lahu_Lancang | Han_Jiulongpo | 0.0359927 |
| Lahu_Lancang | Miao | 0.036074 |
| Lahu_Lancang | Han_Fuling | 0.0361555 |
| Lahu_Lancang | Han_Qiannan | 0.0361673 |
| Lahu_Lancang | NorthernHan | 0.0362575 |
| Blang_Menghai | Japanese | 0.0366502 |
| Lahu_Lancang | Xibo | 0.0367587 |
| Lahu_Lancang | Tu | 0.0368703 |
| Lahu_Lancang | Han_Hanzhong | 0.0375445 |
| Lahu_Lancang | Tibetan_Nyingchi | 0.0381321 |
| Wa_Cangyuan | Tibetan_Lhasa | 0.0385664 |
| Wa_Cangyuan | Japanese | 0.0388205 |
| Lahu_Lancang | Han_Xiamen | 0.0391007 |
| Lahu_Lancang | Daur | 0.0391981 |
| Lahu_Lancang | Han_Putian | 0.0394136 |
| Lahu_Lancang | Han_Xianyang | 0.0396019 |
| Lahu_Lancang | Han_Sanming | 0.0397049 |
| Lahu_Lancang | Han_Zhangzhou | 0.0397112 |
| Blang_Menghai | Tibetan_Amdo_Dalu | 0.0397429 |
| Wa_Cangyuan | Tibetan_Amdo_Dalu | 0.0400189 |
| Lahu_Lancang | Han_Quanzhou | 0.0400427 |
| Lahu_Lancang | Han_Ningde | 0.0401481 |
| Lahu_Lancang | Han_Fuzhou | 0.0402565 |
| Lahu_Lancang | Han_Longyan | 0.0402785 |
| Lahu_Lancang | Han_Nanping | 0.0404683 |
| Lahu_Lancang | Han | 0.0406814 |
| Blang_Menghai | Tibetan_Zase | 0.0408573 |
| Lahu_Lancang | Han_Baoji | 0.0409039 |
| Hani_Honghe | Hazara | 0.0409608 |
| Pumi_Lanping | Hazara | 0.0410622 |
| Lahu_Lancang | Han_Zhumadian | 0.0411249 |
| Lahu_Lancang | Han_Pingdingshan | 0.0412368 |
| Lahu_Lancang | Han_Xinzhou | 0.0412662 |
| Lahu_Lancang | Han_Luoyang | 0.0413706 |
| Blang_Menghai | Uyghur | 0.0414111 |
| Lahu_Lancang | Han_Xinyang | 0.0414941 |
| Lahu_Lancang | Han_Hebi | 0.0415199 |
| Lahu_Lancang | Han_Shangqiu | 0.0415924 |
| Lahu_Lancang | Han_Heze | 0.0416166 |
| Lahu_Lancang | Han_Taiyuan | 0.041653 |
| Lahu_Lancang | Han_Linfen | 0.0417056 |
| Lahu_Lancang | Han_Kaifeng | 0.041707 |
| Lahu_Lancang | Han_Luohe | 0.0417158 |
| Lahu_Lancang | Han_Zhoukou | 0.0418176 |
| Lahu_Lancang | Han_Datong | 0.0418204 |
| Lahu_Lancang | Han_Binzhou | 0.0418487 |
| Lahu_Lancang | Han_Jinzhong | 0.0418815 |
| Lahu_Lancang | Han_Nanchong | 0.0418837 |
| Lahu_Lancang | Han_Nanyang | 0.0419231 |
| Lahu_Lancang | Han_Jiyuan | 0.0420246 |
| Lahu_Lancang | Tibetan_Lhasa | 0.0420377 |

| | | |
|---|---|---|
| Lahu_Lancang | Han_Anyang | 0.0420573 |
| Wa_Cangyuan | Tibetan_Zase | 0.0420696 |
| Lahu_Lancang | Han_Yuncheng | 0.0420813 |
| Lahu_Lancang | Han_Taiwan | 0.0420879 |
| Lahu_Lancang | Han_Dongying | 0.0421151 |
| Lahu_Lancang | Han_Xuchang | 0.0421246 |
| Lahu_Lancang | Han_Jinan | 0.0421425 |
| Lahu_Lancang | Han_Jining | 0.0421903 |
| Lahu_Lancang | Han_Weihai | 0.0422006 |
| Lahu_Lancang | Han_Sanmenxia | 0.0422176 |
| Lahu_Lancang | Han_Taian | 0.0422399 |
| Lahu_Lancang | Han_Yangquan | 0.0422931 |
| Lahu_Lancang | Han_Suzhou | 0.0422942 |
| Lahu_Lancang | Han_Liaocheng | 0.0423365 |
| Lahu_Lancang | Han_Zhengzhou | 0.042392 |
| Lahu_Lancang | Han_Laiwu | 0.042425 |
| Lahu_Lancang | Han_Jincheng | 0.0425005 |
| Lahu_Lancang | Han_Lanzhou | 0.0425133 |
| Lahu_Lancang | Han_Linyi | 0.0425293 |
| Lahu_Lancang | Han_Zaozhuang | 0.0425308 |
| Lahu_Lancang | Han_Jiaozuo | 0.042531 |
| Lahu_Lancang | Han_YulinSX | 0.0426002 |
| Lahu_Lancang | Han_Weinan | 0.042639 |
| Lahu_Lancang | Han_Lvliang | 0.0426466 |
| Lahu_Lancang | Han_Weifang | 0.0426485 |
| Lahu_Lancang | Han_Changzhi | 0.0426821 |
| Lahu_Lancang | Han_Xinxiang | 0.0427102 |
| Lahu_Lancang | Han_Yantai | 0.042722 |
| Pumi_Lanping | Yakut | 0.042736 |
| Lahu_Lancang | She | 0.0427472 |
| Lahu_Lancang | Han_Rizhao | 0.0428407 |
| Lahu_Lancang | Han_Puyang | 0.0428534 |
| Lahu_Lancang | Han_Dezhou | 0.0428937 |
| Lahu_Lancang | Tibetan_Amdo_Dalu | 0.0428972 |
| Lahu_Lancang | Han_Zibo | 0.0431744 |
| Lahu_Lancang | Han_Qingdao | 0.0431778 |
| Wa_Cangyuan | Uyghur | 0.0438057 |
| Lahu_Lancang | Hezhen | 0.0438729 |
| Lahu_Lancang | Han_Xian | 0.0443821 |
| Hani_Honghe | Yakut | 0.045557 |
| Blang_Menghai | Hazara | 0.0470283 |
| Wa_Cangyuan | Hazara | 0.048849 |
| Lahu_Lancang | Oroqen | 0.0492583 |
| Lahu_Lancang | Japanese | 0.0502826 |
| Lahu_Lancang | Tibetan_Zase | 0.0549317 |
| Lahu_Lancang | Uyghur | 0.0556563 |
| Blang_Menghai | Yakut | 0.0559858 |
| Wa_Cangyuan | Yakut | 0.0580167 |
| Lahu_Lancang | Hazara | 0.0612248 |
| Bai_Heqing | Bougainville | 0.0657316 |
| Hani_Honghe | Bougainville | 0.0694881 |
| Lahu_Lancang | Yakut | 0.0704064 |
| Bai_Heqing | Burusho | 0.0721597 |
| Blang_Menghai | Bougainville | 0.072282 |
| Wa_Cangyuan | Bougainville | 0.0753495 |
| Pumi_Lanping | Bougainville | 0.075929 |
| Hani_Honghe | Burusho | 0.0778046 |
| Bai_Heqing | Tuscan_2 | 0.0783274 |
| Pumi_Lanping | Burusho | 0.0791041 |
| Blang_Menghai | Burusho | 0.0800904 |
| Bai_Heqing | Sindhi_Pakistan | 0.0806848 |
| Bai_Heqing | Pathan | 0.081716 |
| Wa_Cangyuan | Burusho | 0.0822143 |
| Hani_Honghe | Tuscan_2 | 0.0852372 |
| Hani_Honghe | Sindhi_Pakistan | 0.0855514 |
| Hani_Honghe | Pathan | 0.0864061 |

| | | |
|---|---|---|
| Blang_Menghai | Tuscan_2 | 0.0865363 |
| Blang_Menghai | Sindhi_Pakistan | 0.087044 |
| Pumi_Lanping | Sindhi_Pakistan | 0.0878412 |
| Blang_Menghai | Pathan | 0.0881713 |
| Pumi_Lanping | Tuscan_2 | 0.0885272 |
| Pumi_Lanping | Pathan | 0.0886323 |
| Wa_Cangyuan | Sindhi_Pakistan | 0.088828 |
| Bai_Heqing | Balochi | 0.0900451 |
| Wa_Cangyuan | Pathan | 0.0900771 |
| Wa_Cangyuan | Tuscan_2 | 0.0906824 |
| Bai_Heqing | Brahui | 0.09292 |
| Bai_Heqing | Makrani | 0.0934737 |
| Hani_Honghe | Balochi | 0.09487 |
| Lahu_Lancang | Bougainville | 0.0954988 |
| Lahu_Lancang | Burusho | 0.0955607 |
| Blang_Menghai | Balochi | 0.0961417 |
| Pumi_Lanping | Balochi | 0.097351 |
| Hani_Honghe | Brahui | 0.0977539 |
| Wa_Cangyuan | Balochi | 0.0982027 |
| Hani_Honghe | Makrani | 0.0985837 |
| Lahu_Lancang | CEU | 0.0992272 |
| Blang_Menghai | Brahui | 0.0992784 |
| Bai_Heqing | Adygei | 0.0994496 |
| Blang_Menghai | Makrani | 0.0999312 |
| Bai_Heqing | Russian | 0.0999806 |
| Pumi_Lanping | Brahui | 0.100222 |
| Pumi_Lanping | Makrani | 0.100461 |
| Wa_Cangyuan | Brahui | 0.101247 |
| Wa_Cangyuan | Makrani | 0.101842 |
| Lahu_Lancang | Sindhi_Pakistan | 0.10187 |
| Lahu_Lancang | Pathan | 0.102789 |
| Bai_Heqing | CEU | 0.104403 |
| Bai_Heqing | Mayan | 0.104522 |
| Lahu_Lancang | Tuscan_2 | 0.105134 |
| Hani_Honghe | Russian | 0.106009 |
| Hani_Honghe | Adygei | 0.10606 |
| Bai_Heqing | BedouinA | 0.1062 |
| Wa_Cangyuan | CEU | 0.106276 |
| Hani_Honghe | CEU | 0.106284 |
| Blang_Menghai | CEU | 0.106337 |
| Pumi_Lanping | Russian | 0.107254 |
| Blang_Menghai | Adygei | 0.107582 |
| Pumi_Lanping | CEU | 0.107673 |
| Pumi_Lanping | Adygei | 0.107715 |
| Blang_Menghai | Russian | 0.107757 |
| Bai_Heqing | Palestinian | 0.10818 |
| Wa_Cangyuan | Adygei | 0.109474 |
| Wa_Cangyuan | Russian | 0.109668 |
| Lahu_Lancang | Balochi | 0.110301 |
| Hani_Honghe | BedouinA | 0.111688 |
| Bai_Heqing | Kalash | 0.111944 |
| Bai_Heqing | Italian_North | 0.112155 |
| Pumi_Lanping | Mayan | 0.112758 |
| Bai_Heqing | French | 0.112867 |
| Pumi_Lanping | BedouinA | 0.113011 |
| Blang_Menghai | BedouinA | 0.113176 |
| Hani_Honghe | Palestinian | 0.113288 |
| Bai_Heqing | Orcadian | 0.114041 |
| Bai_Heqing | Druze | 0.114055 |
| Hani_Honghe | Mayan | 0.114194 |
| Lahu_Lancang | Makrani | 0.114296 |
| Blang_Menghai | Palestinian | 0.114488 |
| Wa_Cangyuan | BedouinA | 0.114497 |
| Lahu_Lancang | Brahui | 0.114691 |
| Pumi_Lanping | Palestinian | 0.115133 |
| Bai_Heqing | Mozabite | 0.116292 |

| | | |
|---|---|---|
| Wa_Cangyuan | Palestinian | 0.116294 |
| Hani_Honghe | Kalash | 0.117716 |
| Hani_Honghe | Italian_North | 0.117964 |
| Hani_Honghe | French | 0.118449 |
| Hani_Honghe | Druze | 0.119523 |
| Pumi_Lanping | Kalash | 0.119524 |
| Blang_Menghai | Italian_North | 0.119694 |
| Blang_Menghai | French | 0.119772 |
| Pumi_Lanping | French | 0.119779 |
| Blang_Menghai | Kalash | 0.119851 |
| Pumi_Lanping | Italian_North | 0.11987 |
| Bai_Heqing | Piapoco | 0.120082 |
| Hani_Honghe | Orcadian | 0.120103 |
| Blang_Menghai | Druze | 0.120573 |
| Pumi_Lanping | Druze | 0.121197 |
| Pumi_Lanping | Orcadian | 0.121361 |
| Wa_Cangyuan | Italian_North | 0.121397 |
| Wa_Cangyuan | French | 0.121477 |
| Hani_Honghe | Mozabite | 0.121481 |
| Blang_Menghai | Orcadian | 0.121744 |
| Bai_Heqing | Basque | 0.121992 |
| Wa_Cangyuan | Kalash | 0.122012 |
| Wa_Cangyuan | Druze | 0.122423 |
| Blang_Menghai | Mayan | 0.122477 |
| Lahu_Lancang | Russian | 0.123032 |
| Blang_Menghai | Mozabite | 0.123161 |
| Pumi_Lanping | Mozabite | 0.123281 |
| Wa_Cangyuan | Orcadian | 0.123507 |
| Lahu_Lancang | Adygei | 0.123907 |
| Wa_Cangyuan | Mozabite | 0.124345 |
| Wa_Cangyuan | Mayan | 0.125059 |
| Bai_Heqing | Sardinian | 0.125638 |
| Lahu_Lancang | BedouinA | 0.126324 |
| Hani_Honghe | Basque | 0.128139 |
| Lahu_Lancang | Palestinian | 0.128924 |
| Pumi_Lanping | Basque | 0.129172 |
| Blang_Menghai | Basque | 0.129274 |
| Pumi_Lanping | Piapoco | 0.129277 |
| Hani_Honghe | Piapoco | 0.130417 |
| Wa_Cangyuan | Basque | 0.131419 |
| Hani_Honghe | Sardinian | 0.131584 |
| Blang_Menghai | Sardinian | 0.132711 |
| Bai_Heqing | Nasioi | 0.133003 |
| Pumi_Lanping | Sardinian | 0.133062 |
| Wa_Cangyuan | Sardinian | 0.134737 |
| Lahu_Lancang | French | 0.135125 |
| Lahu_Lancang | Italian_North | 0.135216 |
| Lahu_Lancang | Druze | 0.135465 |
| Bai_Heqing | Pima | 0.13565 |
| Lahu_Lancang | Kalash | 0.136435 |
| Lahu_Lancang | Mozabite | 0.137133 |
| Lahu_Lancang | Orcadian | 0.138102 |
| Hani_Honghe | Nasioi | 0.138314 |
| Blang_Menghai | Piapoco | 0.139269 |
| Blang_Menghai | Nasioi | 0.140893 |
| Lahu_Lancang | Mayan | 0.141219 |
| Wa_Cangyuan | Piapoco | 0.141442 |
| Pumi_Lanping | Nasioi | 0.143486 |
| Wa_Cangyuan | Nasioi | 0.143907 |
| Pumi_Lanping | Pima | 0.144694 |
| Lahu_Lancang | Basque | 0.145623 |
| Hani_Honghe | Pima | 0.146695 |
| Lahu_Lancang | Sardinian | 0.148671 |
| Blang_Menghai | Pima | 0.155882 |
| Wa_Cangyuan | Pima | 0.158038 |
| Bai_Heqing | BantuKenya | 0.158975 |

| | | |
|---|---|---|
| Bai_Heqing | Karitiana | 0.162685 |
| Hani_Honghe | BantuKenya | 0.162982 |
| Bai_Heqing | BantuSA | 0.163893 |
| Lahu_Lancang | Piapoco | 0.16486 |
| Blang_Menghai | BantuKenya | 0.165035 |
| Pumi_Lanping | BantuKenya | 0.166636 |
| Wa_Cangyuan | BantuKenya | 0.167336 |
| Lahu_Lancang | Nasioi | 0.167478 |
| Hani_Honghe | BantuSA | 0.168423 |
| Blang_Menghai | BantuSA | 0.170998 |
| Pumi_Lanping | BantuSA | 0.171973 |
| Bai_Heqing | Papuan | 0.172484 |
| Wa_Cangyuan | BantuSA | 0.172638 |
| Pumi_Lanping | Karitiana | 0.173072 |
| Bai_Heqing | Mandenka | 0.173294 |
| Hani_Honghe | Karitiana | 0.174501 |
| Bai_Heqing | Yoruba | 0.175119 |
| Hani_Honghe | Mandenka | 0.177092 |
| Hani_Honghe | Yoruba | 0.17854 |
| Hani_Honghe | Papuan | 0.178935 |
| Blang_Menghai | Mandenka | 0.179142 |
| Lahu_Lancang | Pima | 0.179913 |
| Pumi_Lanping | Mandenka | 0.179934 |
| Wa_Cangyuan | Mandenka | 0.180741 |
| Blang_Menghai | Yoruba | 0.18114 |
| Blang_Menghai | Papuan | 0.181335 |
| Bai_Heqing | Surui | 0.18155 |
| Pumi_Lanping | Yoruba | 0.181628 |
| Lahu_Lancang | BantuKenya | 0.18206 |
| Wa_Cangyuan | Yoruba | 0.182128 |
| Pumi_Lanping | Papuan | 0.182666 |
| Blang_Menghai | Karitiana | 0.183301 |
| Wa_Cangyuan | Papuan | 0.184942 |
| Wa_Cangyuan | Karitiana | 0.187272 |
| Lahu_Lancang | BantuSA | 0.188583 |
| Pumi_Lanping | Surui | 0.193011 |
| Hani_Honghe | Surui | 0.193461 |
| Lahu_Lancang | Mandenka | 0.194369 |
| Lahu_Lancang | Yoruba | 0.195556 |
| Bai_Heqing | Biaka | 0.201084 |
| Blang_Menghai | Surui | 0.202761 |
| Hani_Honghe | Biaka | 0.205248 |
| Wa_Cangyuan | Surui | 0.206595 |
| Blang_Menghai | Biaka | 0.207562 |
| Pumi_Lanping | Biaka | 0.208326 |
| Wa_Cangyuan | Biaka | 0.209285 |
| Lahu_Lancang | Papuan | 0.210686 |
| Lahu_Lancang | Karitiana | 0.212243 |
| Bai_Heqing | Ju_hoan_North | 0.220054 |
| Bai_Heqing | Mbuti | 0.2204 |
| Lahu_Lancang | Biaka | 0.224433 |
| Hani_Honghe | Mbuti | 0.22565 |
| Hani_Honghe | Ju_hoan_North | 0.226139 |
| Blang_Menghai | Mbuti | 0.228722 |
| Pumi_Lanping | Mbuti | 0.22876 |
| Pumi_Lanping | Ju_hoan_North | 0.228847 |
| Blang_Menghai | Ju_hoan_North | 0.228949 |
| Wa_Cangyuan | Mbuti | 0.23039 |
| Wa_Cangyuan | Ju_hoan_North | 0.231939 |
| Lahu_Lancang | Surui | 0.239434 |
| Lahu_Lancang | Mbuti | 0.250749 |
| Lahu_Lancang | Ju_hoan_North | 0.254551 |

**Table S5. Symmetric *f4*-statistic testing in the form of *f4*(TYC, TYC, Modern, Mbuti)**

| Population Pairs | Reference population | Outgroup | f4 | stderr | Zscore | BABA | ABBA | nsnps |
|---|---|---|---|---|---|---|---|---|
| TCL&TEL | Htin_Mal | Mbuti | -0.001176 | 9.30E-05 | -12.659 | 27121 | 27765 | 547429 |
| TCL&TEL | Mlabri | Mbuti | -0.001006 | 0.00011 | -9.138 | 27100 | 27651 | 547429 |
| TCL&TEL | Mongol_Minorities | Mbuti | -0.000189 | 7.90E-05 | -2.39 | 27754 | 27865 | 585222 |
| TCL&TEL | Mongol_Sainnoyon | Mbuti | -0.000199 | 7.90E-05 | -2.514 | 27721 | 27837 | 585222 |
| TCL&TEL | Mongol_Uuld | Mbuti | -0.000151 | 8.00E-05 | -1.892 | 27712 | 27800 | 585222 |
| TCL&TEL | Mongol_Zasagt | Mbuti | -0.000215 | 8.00E-05 | -2.674 | 27730 | 27856 | 585222 |
| TCL&TEL | Nanai | Mbuti | -0.000239 | 8.50E-05 | -2.795 | 27902 | 28042 | 585222 |
| TCL&TEL | Negidal | Mbuti | -0.000253 | 9.80E-05 | -2.591 | 27872 | 28020 | 585222 |
| TCL&TEL | Ilocano | Mbuti | -0.001386 | 1.00E-04 | -13.842 | 27412 | 28225 | 586876 |
| TCL&TEL | Cong | Mbuti | -0.000816 | 8.40E-05 | -9.687 | 27620 | 28098 | 586882 |
| TCL&TEL | HaNhi | Mbuti | -0.000644 | 8.20E-05 | -7.824 | 27696 | 28074 | 586882 |
| TCL&TEL | LoLo | Mbuti | -0.0012 | 8.70E-05 | -13.851 | 27536 | 28241 | 586882 |
| TCL&TEL | PhuLa | Mbuti | -0.001046 | 8.20E-05 | -12.727 | 27600 | 28214 | 586882 |
| TCL&TEL | Sila | Mbuti | -0.00062 | 8.70E-05 | -7.142 | 27706 | 28070 | 586882 |
| TCL&TEL | Vietnam_Lahu | Mbuti | -0.000832 | 8.60E-05 | -9.705 | 27629 | 28117 | 586882 |
| TCL&TEL | KhoMu | Mbuti | -0.001039 | 8.30E-05 | -12.475 | 27487 | 28097 | 586882 |
| TCL&TEL | Kinh_Vietnam | Mbuti | -0.001227 | 8.40E-05 | -14.685 | 27557 | 28277 | 586882 |
| TCL&TEL | Vietnamese | Mbuti | -0.001168 | 8.50E-05 | -13.733 | 27577 | 28262 | 586882 |
| TCL&TEL | CoLao | Mbuti | -0.001098 | 8.90E-05 | -12.299 | 27674 | 28319 | 586882 |
| TCL&TEL | Dao | Mbuti | -0.001201 | 8.70E-05 | -13.881 | 27610 | 28315 | 586882 |
| TCL&TEL | Dao | Mbuti | -0.001201 | 8.70E-05 | -13.881 | 27610 | 28315 | 586882 |
| TCL&TEL | Dusun | Mbuti | -0.001266 | 8.90E-05 | -14.293 | 27460 | 28203 | 586882 |
| TCL&TEL | Kankanaey | Mbuti | -0.001339 | 9.20E-05 | -14.632 | 27524 | 28310 | 586882 |
| TCL&TEL | Murut | Mbuti | -0.001343 | 9.00E-05 | -14.97 | 27468 | 28256 | 586882 |
| TCL&TEL | PaThen | Mbuti | -0.001029 | 8.80E-05 | -11.68 | 27632 | 28236 | 586882 |
| TCL&TEL | PaThen | Mbuti | -0.001029 | 8.80E-05 | -11.68 | 27632 | 28236 | 586882 |
| TCL&TEL | BoY | Mbuti | -0.001117 | 8.70E-05 | -12.835 | 27607 | 28262 | 586882 |
| TCL&TEL | Cham | Mbuti | -0.001004 | 8.00E-05 | -12.554 | 27385 | 27974 | 586882 |
| TCL&TEL | Ede | Mbuti | -0.0011 | 8.30E-05 | -13.246 | 27268 | 27914 | 586882 |
| TCL&TEL | Giarai | Mbuti | -0.001142 | 8.10E-05 | -14.16 | 27301 | 27972 | 586882 |
| TCL&TEL | LaChi | Mbuti | -0.001297 | 9.10E-05 | -14.301 | 27570 | 28331 | 586882 |
| TCL&TEL | Malay | Mbuti | -0.001071 | 8.00E-05 | -13.347 | 27320 | 27949 | 586882 |
| TCL&TEL | Mang | Mbuti | -0.000917 | 8.60E-05 | -10.667 | 27537 | 28075 | 586882 |
| TCL&TEL | Muong | Mbuti | -0.001255 | 8.50E-05 | -14.712 | 27543 | 28280 | 586882 |
| TCL&TEL | Nung | Mbuti | -0.001282 | 8.60E-05 | -14.879 | 27555 | 28307 | 586882 |
| TCL&TEL | Tagalog | Mbuti | -0.001041 | 8.70E-05 | -12.035 | 27431 | 28042 | 586882 |
| TCL&TEL | Tay | Mbuti | -0.001204 | 8.20E-05 | -14.676 | 27562 | 28269 | 586882 |
| TCL&TEL | Visayan | Mbuti | -0.001176 | 8.90E-05 | -13.167 | 27389 | 28079 | 586882 |
| TCL&TEL | Hmong | Mbuti | -0.001043 | 8.60E-05 | -12.132 | 27771 | 28387 | 590678 |
| TCL&TEL | Hmong | Mbuti | -0.001043 | 8.60E-05 | -12.132 | 27771 | 28387 | 590678 |
| TCL&TEL | Sherpa_Nepal | Mbuti | 0.000344 | 8.40E-05 | 4.096 | 28091 | 27888 | 590899 |
| TCL&TEL | Tibetan_Nagqu | Mbuti | 0.000508 | 8.10E-05 | 6.279 | 28166 | 27866 | 590899 |
| TCL&TEL | Tibetan_Shannan | Mbuti | 0.000466 | 8.10E-05 | 5.787 | 28119 | 27843 | 590899 |
| TCL&TEL | Dong_Hunan | Mbuti | -0.001162 | 8.80E-05 | -13.276 | 27748 | 28435 | 590899 |
| TCL&TEL | Qiang_Danba | Mbuti | 4.00E-04 | 9.10E-05 | 4.421 | 28238 | 28001 | 590900 |
| TCL&TEL | Qiang_Daofu | Mbuti | 0.000407 | 8.30E-05 | 4.884 | 28198 | 27957 | 590900 |
| TCL&TEL | Tamang_Nepal | Mbuti | 6.70E-05 | 8.20E-05 | 0.818 | 27788 | 27748 | 590900 |
| TCL&TEL | Tibetan_Chamdo | Mbuti | 0.00055 | 7.80E-05 | 7.019 | 28193 | 27868 | 590900 |
| TCL&TEL | Tibetan_Gangcha | Mbuti | 0.000315 | 7.70E-05 | 4.105 | 28057 | 27871 | 590900 |
| TCL&TEL | Tibetan_Gannan | Mbuti | -0.000239 | 8.50E-05 | -2.801 | 27906 | 28048 | 590900 |
| TCL&TEL | Tibetan_Lhasa | Mbuti | 0.000453 | 8.00E-05 | 5.633 | 28143 | 27875 | 590900 |
| TCL&TEL | Tibetan_Shigatse | Mbuti | 0.00043 | 8.00E-05 | 5.399 | 28119 | 27865 | 590900 |
| TCL&TEL | Tibetan_Xinlong | Mbuti | -0.000714 | 8.60E-05 | -8.258 | 27890 | 28312 | 590900 |
| TCL&TEL | Tibetan_Xunhua | Mbuti | -1.20E-05 | 9.20E-05 | -0.128 | 28000 | 28007 | 590900 |
| TCL&TEL | Tibetan_Yajiang | Mbuti | 0.000217 | 8.60E-05 | 2.537 | 28128 | 28000 | 590900 |
| TCL&TEL | TibetanHO_Yunnan | Mbuti | 0.000152 | 9.00E-05 | 1.679 | 28116 | 28026 | 590900 |
| TCL&TEL | Gelao_Longlin | Mbuti | -0.001093 | 8.60E-05 | -12.685 | 27762 | 28408 | 590900 |
| TCL&TEL | Maonan_Huanjiang | Mbuti | -0.001313 | 8.30E-05 | -15.858 | 27702 | 28478 | 590900 |
| TCL&TEL | Mulam_Luocheng | Mbuti | -0.001286 | 8.40E-05 | -15.311 | 27701 | 28460 | 590900 |
| TCL&TEL | Han_Hubei | Mbuti | -0.000891 | 8.80E-05 | -10.089 | 27904 | 28431 | 590900 |
| TCL&TEL | Han_Jiangsu | Mbuti | -0.00084 | 8.60E-05 | -9.779 | 27876 | 28373 | 590900 |
| TCL&TEL | Han_Shandong | Mbuti | -0.000661 | 8.30E-05 | -7.993 | 27914 | 28304 | 590900 |
| TCL&TEL | Han_Shanxi | Mbuti | -0.000541 | 8.30E-05 | -6.505 | 27955 | 28275 | 590900 |
| TCL&TEL | Han_SichuanHO | Mbuti | -0.000777 | 8.20E-05 | -9.454 | 27873 | 28332 | 590900 |
| TCL&TEL | Han_Zhejiang | Mbuti | -0.000786 | 8.80E-05 | -8.898 | 27878 | 28342 | 590900 |
| TCL&TEL | Dongxiang_Linxia | Mbuti | -0.000366 | 8.00E-05 | -4.564 | 27788 | 28004 | 590900 |
| TCL&TEL | Han_Taiwan | Mbuti | -0.000923 | 9.30E-05 | -9.931 | 27876 | 28423 | 593034 |
| TCL&TEL | Lue | Mbuti | -0.001251 | 8.60E-05 | -14.541 | 27696 | 28438 | 593044 |
| TCL&TEL | Shan | Mbuti | -0.000903 | 8.40E-05 | -10.73 | 27731 | 28266 | 593045 |
| TCL&TEL | Palaung | Mbuti | -0.000697 | 8.20E-05 | -8.496 | 27782 | 28195 | 593047 |
| TCL&TEL | IuMien | Mbuti | -0.00105 | 8.60E-05 | -12.168 | 27799 | 28421 | 593048 |
| TCL&TEL | IuMien | Mbuti | -0.00105 | 8.60E-05 | -12.168 | 27799 | 28421 | 593048 |
| TCL&TEL | Phuan | Mbuti | -0.001242 | 8.50E-05 | -14.59 | 27665 | 28402 | 593048 |
| TCL&TEL | Yuan | Mbuti | -0.001178 | 8.50E-05 | -13.921 | 27675 | 28374 | 593048 |
| TCL&TEL | Kalueang | Mbuti | -0.001156 | 8.50E-05 | -13.674 | 27582 | 28267 | 593049 |
| TCL&TEL | BlackTai | Mbuti | -0.001233 | 8.80E-05 | -14.013 | 27718 | 28449 | 593049 |
| TCL&TEL | Khmu | Mbuti | -0.001057 | 8.30E-05 | -12.774 | 27628 | 28255 | 593049 |

| | | | | | | | | |
|---|---|---|---|---|---|---|---|---|
| TCL&TEL | Lawa_Eastern | Mbuti | -0.000739 | 8.00E-05 | -9.187 | 27718 | 28156 | 593049 |
| TCL&TEL | Lawa_Western | Mbuti | -0.000717 | 8.20E-05 | -8.734 | 27756 | 28182 | 593049 |
| TCL&TEL | KarenPadaung | Mbuti | -0.000616 | 8.30E-05 | -7.377 | 27789 | 28154 | 593050 |
| TCL&TEL | KarenPwo | Mbuti | -0.000629 | 8.40E-05 | -7.518 | 27785 | 28158 | 593050 |
| TCL&TEL | KarenSkaw | Mbuti | -0.000694 | 8.40E-05 | -8.235 | 27747 | 28158 | 593050 |
| TCL&TEL | Lahu | Mbuti | -0.000875 | 8.40E-05 | -10.456 | 27787 | 28307 | 593050 |
| TCL&TEL | Lahu_HGDP | Mbuti | -0.000895 | 8.70E-05 | -10.294 | 27794 | 28324 | 593050 |
| TCL&TEL | Lisu | Mbuti | -0.000655 | 8.10E-05 | -8.032 | 27852 | 28240 | 593050 |
| TCL&TEL | Naxi_HGDP | Mbuti | -0.000278 | 8.20E-05 | -3.38 | 28006 | 28171 | 593050 |
| TCL&TEL | Tujia_HGDP | Mbuti | -0.000824 | 8.30E-05 | -9.907 | 27909 | 28398 | 593050 |
| TCL&TEL | Cambodian | Mbuti | -0.001005 | 8.10E-05 | -12.483 | 27560 | 28156 | 593050 |
| TCL&TEL | Kinh_HGDP | Mbuti | -0.001148 | 8.40E-05 | -13.588 | 27731 | 28412 | 593050 |
| TCL&TEL | Ami | Mbuti | -0.001342 | 8.80E-05 | -15.162 | 27714 | 28510 | 593050 |
| TCL&TEL | Atayal | Mbuti | -0.001255 | 9.40E-05 | -13.314 | 27726 | 28470 | 593050 |
| TCL&TEL | Dai_HGDP | Mbuti | -0.001316 | 8.60E-05 | -15.299 | 27686 | 28467 | 593050 |
| TCL&TEL | HmongDaw | Mbuti | -0.001064 | 8.90E-05 | -11.913 | 27822 | 28453 | 593050 |
| TCL&TEL | HmongDaw | Mbuti | -0.001064 | 8.90E-05 | -11.913 | 27822 | 28453 | 593050 |
| TCL&TEL | HmongNjua | Mbuti | -0.001046 | 8.80E-05 | -11.921 | 27824 | 28444 | 593050 |
| TCL&TEL | HmongNjua | Mbuti | -0.001046 | 8.80E-05 | -11.921 | 27824 | 28444 | 593050 |
| TCL&TEL | Thai | Mbuti | -0.001144 | 8.00E-05 | -14.236 | 27627 | 28305 | 593050 |
| TCL&TEL | Daur_HGDP | Mbuti | -0.00043 | 8.40E-05 | -5.133 | 27896 | 28151 | 593050 |
| TCL&TEL | Hezhen_HGDP | Mbuti | -0.000454 | 8.60E-05 | -5.308 | 27900 | 28169 | 593050 |
| TCL&TEL | Oroqen_HGDP | Mbuti | -0.000273 | 8.30E-05 | -3.295 | 27962 | 28124 | 593050 |
| TCL&TEL | Tu_HGDP | Mbuti | -0.00027 | 7.80E-05 | -3.448 | 27919 | 28079 | 593050 |
| TCL&TEL | Ulchi | Mbuti | -0.000269 | 8.10E-05 | -3.322 | 27932 | 28091 | 593050 |
| TCL&TEL | Xibo_HGDP | Mbuti | -0.00045 | 8.20E-05 | -5.457 | 27878 | 28145 | 593050 |
| TCL&TEL | Bru | Mbuti | -0.001184 | 8.50E-05 | -14 | 27532 | 28234 | 593050 |
| TCL&TEL | CentralThai | Mbuti | -0.000931 | 7.60E-05 | -12.324 | 27554 | 28107 | 593050 |
| TCL&TEL | HtinPray | Mbuti | -0.001048 | 8.70E-05 | -12.1 | 27582 | 28203 | 593050 |
| TCL&TEL | Khonmueang | Mbuti | -0.001159 | 8.00E-05 | -14.521 | 27680 | 28368 | 593050 |
| TCL&TEL | Khuen | Mbuti | -0.001226 | 8.50E-05 | -14.347 | 27721 | 28448 | 593050 |
| TCL&TEL | LaoIsan | Mbuti | -0.001142 | 8.00E-05 | -14.242 | 27597 | 28274 | 593050 |
| TCL&TEL | Laotian | Mbuti | -0.001194 | 8.00E-05 | -14.898 | 27665 | 28373 | 593050 |
| TCL&TEL | Mon | Mbuti | -0.000754 | 7.20E-05 | -10.438 | 27560 | 28007 | 593050 |
| TCL&TEL | Nyaw | Mbuti | -0.001216 | 8.30E-05 | -14.562 | 27614 | 28335 | 593050 |
| TCL&TEL | Phutai | Mbuti | -0.001141 | 8.70E-05 | -13.133 | 27673 | 28350 | 593050 |
| TCL&TEL | Saek | Mbuti | -0.001204 | 8.30E-05 | -14.591 | 27633 | 28347 | 593050 |
| TCL&TEL | Soa | Mbuti | -0.001169 | 8.10E-05 | -14.364 | 27532 | 28225 | 593050 |
| TCL&TEL | SouthernThai_AN | Mbuti | -0.000969 | 7.60E-05 | -12.816 | 27432 | 28007 | 593050 |
| TCL&TEL | SouthernThai_TK | Mbuti | -0.000876 | 7.60E-05 | -11.484 | 27484 | 28004 | 593050 |
| TCL&THL | Htin_Mal | Mbuti | 0.000235 | 0.000104 | 2.254 | 27394 | 27266 | 547429 |
| TCL&THL | Mlabri | Mbuti | 0.000441 | 0.000117 | 3.755 | 27384 | 27143 | 547429 |
| TCL&THL | Mongol_Minorities | Mbuti | 2.00E-05 | 8.90E-05 | 0.222 | 27749 | 27737 | 585221 |
| TCL&THL | Mongol_Sainnoyon | Mbuti | 2.10E-05 | 9.00E-05 | 0.23 | 27723 | 27711 | 585221 |
| TCL&THL | Mongol_Uuld | Mbuti | 5.20E-05 | 8.60E-05 | 0.601 | 27707 | 27677 | 585221 |
| TCL&THL | Mongol_Zasagt | Mbuti | 3.70E-05 | 8.80E-05 | 0.425 | 27737 | 27715 | 585221 |
| TCL&THL | Nanai | Mbuti | 7.20E-05 | 9.10E-05 | 0.788 | 27925 | 27883 | 585221 |
| TCL&THL | Negidal | Mbuti | 7.50E-05 | 0.000105 | 0.714 | 27903 | 27859 | 585221 |
| TCL&THL | Ilocano | Mbuti | 0.000334 | 0.000111 | 3.005 | 27789 | 27593 | 586875 |
| TCL&THL | Cong | Mbuti | 0.000191 | 9.40E-05 | 2.028 | 27817 | 27705 | 586881 |
| TCL&THL | HaNhi | Mbuti | 5.70E-05 | 9.00E-05 | 0.631 | 27815 | 27781 | 586881 |
| TCL&THL | LoLo | Mbuti | 0.000296 | 9.50E-05 | 3.125 | 27852 | 27678 | 586881 |
| TCL&THL | PhuLa | Mbuti | 0.00024 | 9.20E-05 | 2.603 | 27866 | 27725 | 586881 |
| TCL&THL | Sila | Mbuti | 4.40E-05 | 9.70E-05 | 0.452 | 27813 | 27787 | 586881 |
| TCL&THL | Vietnam_Lahu | Mbuti | 7.00E-05 | 9.30E-05 | 0.749 | 27798 | 27757 | 586881 |
| TCL&THL | KhoMu | Mbuti | 0.000182 | 9.20E-05 | 1.966 | 27736 | 27629 | 586881 |
| TCL&THL | Kinh_Vietnam | Mbuti | 0.00032 | 9.10E-05 | 3.503 | 27889 | 27701 | 586881 |
| TCL&THL | Vietnamese | Mbuti | 0.000396 | 9.30E-05 | 4.254 | 27915 | 27683 | 586881 |
| TCL&THL | CoLao | Mbuti | 0.00032 | 9.90E-05 | 3.235 | 27975 | 27787 | 586881 |
| TCL&THL | Dao | Mbuti | 0.000386 | 9.40E-05 | 4.1 | 27954 | 27727 | 586881 |
| TCL&THL | Dao | Mbuti | 0.000386 | 9.40E-05 | 4.1 | 27954 | 27727 | 586881 |
| TCL&THL | Dusun | Mbuti | 0.000411 | 9.30E-05 | 4.401 | 27825 | 27584 | 586881 |
| TCL&THL | Kankanaey | Mbuti | 0.00044 | 9.80E-05 | 4.485 | 27917 | 27659 | 586881 |
| TCL&THL | Murut | Mbuti | 0.000368 | 9.70E-05 | 3.787 | 27843 | 27627 | 586881 |
| TCL&THL | PaThen | Mbuti | 0.00034 | 9.50E-05 | 3.586 | 27918 | 27718 | 586881 |
| TCL&THL | PaThen | Mbuti | 0.00034 | 9.50E-05 | 3.586 | 27918 | 27718 | 586881 |
| TCL&THL | BoY | Mbuti | 0.000345 | 9.80E-05 | 3.532 | 27922 | 27720 | 586881 |
| TCL&THL | Cham | Mbuti | 0.000304 | 8.60E-05 | 3.545 | 27658 | 27479 | 586881 |
| TCL&THL | Ede | Mbuti | 0.000317 | 8.80E-05 | 3.589 | 27566 | 27380 | 586881 |
| TCL&THL | Giarai | Mbuti | 0.000237 | 9.00E-05 | 2.643 | 27592 | 27453 | 586881 |
| TCL&THL | LaChi | Mbuti | 0.000344 | 9.80E-05 | 3.524 | 27925 | 27723 | 586881 |
| TCL&THL | Malay | Mbuti | 0.000247 | 8.70E-05 | 2.856 | 27596 | 27451 | 586881 |
| TCL&THL | Mang | Mbuti | 0.000232 | 9.40E-05 | 2.476 | 27770 | 27634 | 586881 |
| TCL&THL | Muong | Mbuti | 0.000426 | 9.30E-05 | 4.572 | 27912 | 27661 | 586881 |
| TCL&THL | Nung | Mbuti | 0.000407 | 9.30E-05 | 4.397 | 27922 | 27684 | 586881 |
| TCL&THL | Tagalog | Mbuti | 0.000391 | 9.60E-05 | 4.082 | 27735 | 27506 | 586881 |
| TCL&THL | Tay | Mbuti | 0.000358 | 9.40E-05 | 3.814 | 27899 | 27689 | 586881 |
| TCL&THL | Visayan | Mbuti | 0.000424 | 9.60E-05 | 4.404 | 27735 | 27487 | 586881 |
| TCL&THL | Hmong | Mbuti | 0.000243 | 9.30E-05 | 2.626 | 28038 | 27894 | 590677 |

| | | | | | | | | |
|---|---|---|---|---|---|---|---|---|
| TCL&THL | Hmong | Mbuti | 0.000243 | 9.30E-05 | 2.626 | 28038 | 27894 | 590677 |
| TCL&THL | Sherpa_Nepal | Mbuti | -0.000474 | 9.70E-05 | -4.888 | 27823 | 28103 | 590898 |
| TCL&THL | Tibetan_Nagqu | Mbuti | -0.000377 | 9.00E-05 | -4.175 | 27886 | 28109 | 590898 |
| TCL&THL | Tibetan_Shannan | Mbuti | -0.000383 | 9.00E-05 | -4.261 | 27847 | 28073 | 590898 |
| TCL&THL | Dong_Hunan | Mbuti | 0.00025 | 9.50E-05 | 2.633 | 28047 | 27899 | 590898 |
| TCL&THL | Qiang_Danba | Mbuti | -0.000423 | 1.00E-04 | -4.216 | 27973 | 28222 | 590899 |
| TCL&THL | Qiang_Daofu | Mbuti | -0.000402 | 9.20E-05 | -4.377 | 27934 | 28171 | 590899 |
| TCL&THL | Tamang_Nepal | Mbuti | -0.000272 | 8.80E-05 | -3.096 | 27645 | 27806 | 590899 |
| TCL&THL | Tibetan_Chamdo | Mbuti | -0.00044 | 9.00E-05 | -4.906 | 27883 | 28143 | 590899 |
| TCL&THL | Tibetan_Gangcha | Mbuti | -0.000325 | 8.40E-05 | -3.866 | 27838 | 28030 | 590899 |
| TCL&THL | Tibetan_Gannan | Mbuti | 3.30E-05 | 9.30E-05 | 0.357 | 27915 | 27895 | 590899 |
| TCL&THL | Tibetan_Lhasa | Mbuti | -4.00E-04 | 9.30E-05 | -4.314 | 27870 | 28106 | 590899 |
| TCL&THL | Tibetan_Shigatse | Mbuti | -0.000351 | 9.00E-05 | -3.892 | 27868 | 28075 | 590899 |
| TCL&THL | Tibetan_Xinlong | Mbuti | 0.000108 | 9.50E-05 | 1.128 | 28037 | 27974 | 590899 |
| TCL&THL | Tibetan_Xunhua | Mbuti | -0.00017 | 0.000106 | -1.607 | 27905 | 28005 | 590899 |
| TCL&THL | Tibetan_Yajiang | Mbuti | -0.00044 | 9.30E-05 | -4.739 | 27902 | 28162 | 590899 |
| TCL&THL | TibetanHO_Yunnan | Mbuti | -0.000423 | 1.00E-04 | -4.241 | 27914 | 28164 | 590899 |
| TCL&THL | Gelao_Longlin | Mbuti | 0.000337 | 9.20E-05 | 3.677 | 28064 | 27865 | 590899 |
| TCL&THL | Maonan_Huanjiang | Mbuti | 0.000388 | 9.10E-05 | 4.264 | 28076 | 27847 | 590899 |
| TCL&THL | Mulam_Luocheng | Mbuti | 0.000367 | 9.10E-05 | 4.032 | 28062 | 27845 | 590899 |
| TCL&THL | Han_Hubei | Mbuti | 0.000202 | 9.60E-05 | 2.099 | 28123 | 28004 | 590899 |
| TCL&THL | Han_Jiangsu | Mbuti | 0.00019 | 8.90E-05 | 2.119 | 28084 | 27972 | 590899 |
| TCL&THL | Han_Shandong | Mbuti | 0.000123 | 9.10E-05 | 1.361 | 28055 | 27982 | 590899 |
| TCL&THL | Han_Shanxi | Mbuti | 0.000134 | 9.00E-05 | 1.498 | 28068 | 27989 | 590899 |
| TCL&THL | Han_SichuanHO | Mbuti | 0.000264 | 9.20E-05 | 2.879 | 28078 | 27922 | 590899 |
| TCL&THL | Han_Zhejiang | Mbuti | 0.000316 | 9.70E-05 | 3.268 | 28102 | 27915 | 590899 |
| TCL&THL | Dongxiang_Linxia | Mbuti | 8.80E-05 | 8.90E-05 | 0.991 | 27844 | 27792 | 590899 |
| TCL&THL | Han_Taiwan | Mbuti | 0.000327 | 1.00E-04 | 3.259 | 28139 | 27945 | 593034 |
| TCL&THL | Lue | Mbuti | 0.000319 | 9.20E-05 | 3.449 | 28036 | 27847 | 593043 |
| TCL&THL | Shan | Mbuti | 0.00012 | 8.90E-05 | 1.35 | 27934 | 27863 | 593044 |
| TCL&THL | Palaung | Mbuti | -4.70E-05 | 8.90E-05 | -0.529 | 27887 | 27915 | 593046 |
| TCL&THL | IuMien | Mbuti | 0.000307 | 9.20E-05 | 3.344 | 28087 | 27905 | 593047 |
| TCL&THL | IuMien | Mbuti | 0.000307 | 9.20E-05 | 3.344 | 28087 | 27905 | 593047 |
| TCL&THL | Phuan | Mbuti | 0.000375 | 9.30E-05 | 4.045 | 28017 | 27795 | 593047 |
| TCL&THL | Yuan | Mbuti | 0.000393 | 9.00E-05 | 4.35 | 28017 | 27784 | 593047 |
| TCL&THL | Kalueang | Mbuti | 0.000307 | 9.30E-05 | 3.313 | 27896 | 27714 | 593048 |
| TCL&THL | BlackTai | Mbuti | 0.000368 | 9.60E-05 | 3.845 | 28065 | 27847 | 593048 |
| TCL&THL | Khmu | Mbuti | 0.000212 | 9.40E-05 | 2.269 | 27894 | 27768 | 593048 |
| TCL&THL | Lawa_Eastern | Mbuti | 5.10E-05 | 9.00E-05 | 0.57 | 27860 | 27830 | 593048 |
| TCL&THL | Lawa_Western | Mbuti | 5.80E-05 | 9.30E-05 | 0.627 | 27892 | 27858 | 593048 |
| TCL&THL | KarenPadaung | Mbuti | 6.40E-05 | 9.10E-05 | 0.705 | 27904 | 27866 | 593049 |
| TCL&THL | KarenPwo | Mbuti | 0.000155 | 9.10E-05 | 1.716 | 27926 | 27834 | 593049 |
| TCL&THL | KarenSkaw | Mbuti | 0.000106 | 9.10E-05 | 1.158 | 27894 | 27831 | 593049 |
| TCL&THL | Lahu | Mbuti | 0.000107 | 9.30E-05 | 1.156 | 27975 | 27912 | 593049 |
| TCL&THL | Lahu_HGDP | Mbuti | 6.40E-05 | 9.20E-05 | 0.689 | 27976 | 27938 | 593049 |
| TCL&THL | Lisu | Mbuti | 2.00E-06 | 9.00E-05 | 0.021 | 27960 | 27959 | 593049 |
| TCL&THL | Naxi_HGDP | Mbuti | -0.000204 | 9.00E-05 | -2.251 | 27967 | 28088 | 593049 |
| TCL&THL | Tujia_HGDP | Mbuti | 0.000258 | 9.00E-05 | 2.861 | 28125 | 27972 | 593049 |
| TCL&THL | Cambodian | Mbuti | 0.000261 | 8.80E-05 | 2.985 | 27823 | 27668 | 593049 |
| TCL&THL | Kinh_HGDP | Mbuti | 0.000389 | 9.10E-05 | 4.25 | 28064 | 27833 | 593049 |
| TCL&THL | Ami | Mbuti | 0.000449 | 9.80E-05 | 4.594 | 28114 | 27847 | 593049 |
| TCL&THL | Atayal | Mbuti | 0.000555 | 0.000106 | 5.228 | 28130 | 27801 | 593049 |
| TCL&THL | Dai_HGDP | Mbuti | 0.000316 | 9.20E-05 | 3.427 | 28041 | 27853 | 593049 |
| TCL&THL | HmongDaw | Mbuti | 0.000272 | 9.80E-05 | 2.787 | 28106 | 27945 | 593049 |
| TCL&THL | HmongDaw | Mbuti | 0.000272 | 9.80E-05 | 2.787 | 28106 | 27945 | 593049 |
| TCL&THL | HmongNjua | Mbuti | 0.000263 | 9.70E-05 | 2.707 | 28099 | 27943 | 593049 |
| TCL&THL | HmongNjua | Mbuti | 0.000263 | 9.70E-05 | 2.707 | 28099 | 27943 | 593049 |
| TCL&THL | Thai | Mbuti | 0.000303 | 8.70E-05 | 3.487 | 27936 | 27756 | 593049 |
| TCL&THL | Daur_HGDP | Mbuti | 5.90E-05 | 9.00E-05 | 0.66 | 27965 | 27930 | 593049 |
| TCL&THL | Hezhen_HGDP | Mbuti | 7.40E-05 | 9.10E-05 | 0.816 | 27978 | 27934 | 593049 |
| TCL&THL | Oroqen_HGDP | Mbuti | 8.70E-05 | 9.20E-05 | 0.953 | 27995 | 27943 | 593049 |
| TCL&THL | Tu_HGDP | Mbuti | -9.40E-05 | 8.70E-05 | -1.082 | 27906 | 27962 | 593049 |
| TCL&THL | Ulchi | Mbuti | 3.70E-05 | 8.80E-05 | 0.421 | 27953 | 27931 | 593049 |
| TCL&THL | Xibo_HGDP | Mbuti | 5.80E-05 | 9.10E-05 | 0.641 | 27949 | 27915 | 593049 |
| TCL&THL | Bru | Mbuti | 0.000288 | 9.20E-05 | 3.138 | 27848 | 27677 | 593049 |
| TCL&THL | CentralThai | Mbuti | 0.000243 | 8.10E-05 | 3.016 | 27794 | 27650 | 593049 |
| TCL&THL | HtinPray | Mbuti | 0.000281 | 9.50E-05 | 2.944 | 27866 | 27699 | 593049 |
| TCL&THL | Khonmueang | Mbuti | 0.000296 | 8.80E-05 | 3.35 | 27991 | 27816 | 593049 |
| TCL&THL | Khuen | Mbuti | 0.000362 | 9.00E-05 | 4.01 | 28066 | 27851 | 593049 |
| TCL&THL | LaoIsan | Mbuti | 0.000307 | 8.60E-05 | 3.581 | 27907 | 27725 | 593049 |
| TCL&THL | Laotian | Mbuti | 0.000311 | 8.80E-05 | 3.549 | 27988 | 27804 | 593049 |
| TCL&THL | Mon | Mbuti | 0.000112 | 8.00E-05 | 1.402 | 27721 | 27655 | 593049 |
| TCL&THL | Nyaw | Mbuti | 0.000349 | 9.00E-05 | 3.89 | 27954 | 27747 | 593049 |
| TCL&THL | Phutai | Mbuti | 0.000338 | 9.30E-05 | 3.643 | 27989 | 27788 | 593049 |
| TCL&THL | Saek | Mbuti | 0.000343 | 9.10E-05 | 3.775 | 27968 | 27765 | 593049 |
| TCL&THL | Soa | Mbuti | 0.000277 | 8.90E-05 | 3.109 | 27841 | 27677 | 593049 |
| TCL&THL | SouthernThai_AN | Mbuti | 0.000218 | 8.20E-05 | 2.648 | 27676 | 27547 | 593049 |
| TCL&THL | SouthernThai_TK | Mbuti | 0.000174 | 8.10E-05 | 2.147 | 27695 | 27592 | 593049 |
| TCL&TSL | Htin_Mal | Mbuti | -0.001862 | 0.000112 | -16.563 | 27287 | 28306 | 547419 |

| | | | | | | | | |
|---|---|---|---|---|---|---|---|---|
| TCL&TSL | Mlabri | Mbuti | -0.002085 | 0.000134 | -15.545 | 27174 | 28316 | 547419 |
| TCL&TSL | Mongol_Minorities | Mbuti | 0.001187 | 9.60E-05 | 12.423 | 28435 | 27741 | 585211 |
| TCL&TSL | Mongol_Sainnoyon | Mbuti | 0.001204 | 9.40E-05 | 12.807 | 28410 | 27705 | 585211 |
| TCL&TSL | Mongol_Uuld | Mbuti | 0.00124 | 9.30E-05 | 13.333 | 28394 | 27668 | 585211 |
| TCL&TSL | Mongol_Zasagt | Mbuti | 0.001208 | 9.60E-05 | 12.514 | 28421 | 27714 | 585211 |
| TCL&TSL | Nanai | Mbuti | 0.001379 | 0.000101 | 13.614 | 28647 | 27839 | 585211 |
| TCL&TSL | Negidal | Mbuti | 0.001365 | 0.000112 | 12.218 | 28615 | 27816 | 585211 |
| TCL&TSL | Ilocano | Mbuti | -0.000466 | 0.00012 | -3.883 | 27977 | 28250 | 586865 |
| TCL&TSL | Cong | Mbuti | -0.000614 | 9.90E-05 | -6.222 | 28007 | 28367 | 586871 |
| TCL&TSL | HaNhi | Mbuti | -0.000468 | 9.60E-05 | -4.875 | 28076 | 28351 | 586871 |
| TCL&TSL | LoLo | Mbuti | -0.000829 | 0.000102 | -8.093 | 27963 | 28450 | 586871 |
| TCL&TSL | PhuLa | Mbuti | -0.000666 | 1.00E-04 | -6.631 | 28032 | 28422 | 586871 |
| TCL&TSL | Sila | Mbuti | -0.000369 | 0.000107 | -3.455 | 28108 | 28324 | 586871 |
| TCL&TSL | Vietnam_Lahu | Mbuti | -0.000743 | 0.000101 | -7.325 | 27987 | 28423 | 586871 |
| TCL&TSL | KhoMu | Mbuti | -0.001554 | 9.70E-05 | -15.978 | 27693 | 28606 | 586871 |
| TCL&TSL | Kinh_Vietnam | Mbuti | -0.000522 | 9.70E-05 | -5.368 | 28070 | 28377 | 586871 |
| TCL&TSL | Vietnamese | Mbuti | -0.000492 | 9.80E-05 | -4.997 | 28083 | 28371 | 586871 |
| TCL&TSL | CoLao | Mbuti | -0.000371 | 0.000105 | -3.54 | 28195 | 28413 | 586871 |
| TCL&TSL | Dao | Mbuti | -0.000349 | 0.000103 | -3.4 | 28161 | 28366 | 586871 |
| TCL&TSL | Dao | Mbuti | -0.000349 | 0.000103 | -3.4 | 28161 | 28366 | 586871 |
| TCL&TSL | Dusun | Mbuti | -0.000907 | 0.000101 | -8.946 | 27888 | 28420 | 586871 |
| TCL&TSL | Kankanaey | Mbuti | -0.000297 | 0.000106 | -2.792 | 28124 | 28298 | 586871 |
| TCL&TSL | Murut | Mbuti | -0.000987 | 0.000104 | -9.507 | 27892 | 28471 | 586871 |
| TCL&TSL | PaThen | Mbuti | -1.80E-05 | 0.000105 | -0.168 | 28222 | 28233 | 586871 |
| TCL&TSL | PaThen | Mbuti | -1.80E-05 | 0.000105 | -0.168 | 28222 | 28233 | 586871 |
| TCL&TSL | BoY | Mbuti | -6.50E-05 | 0.000104 | -0.63 | 28208 | 28246 | 586871 |
| TCL&TSL | Cham | Mbuti | -0.001066 | 9.30E-05 | -11.416 | 27704 | 28329 | 586871 |
| TCL&TSL | Ede | Mbuti | -0.001891 | 9.70E-05 | -19.429 | 27399 | 28509 | 586871 |
| TCL&TSL | Giarai | Mbuti | -0.001784 | 9.70E-05 | -18.412 | 27474 | 28521 | 586871 |
| TCL&TSL | LaChi | Mbuti | -0.000693 | 0.000105 | -6.599 | 28059 | 28466 | 586871 |
| TCL&TSL | Malay | Mbuti | -0.001161 | 9.60E-05 | -12.109 | 27630 | 28311 | 586871 |
| TCL&TSL | Mang | Mbuti | -0.001127 | 0.000104 | -10.784 | 27819 | 28481 | 586871 |
| TCL&TSL | Muong | Mbuti | -0.000759 | 1.00E-04 | -7.61 | 28006 | 28451 | 586871 |
| TCL&TSL | Nung | Mbuti | -0.000545 | 0.000101 | -5.409 | 28078 | 28398 | 586871 |
| TCL&TSL | Tagalog | Mbuti | -0.000169 | 9.90E-05 | -1.702 | 27986 | 28085 | 586871 |
| TCL&TSL | Tay | Mbuti | -0.000392 | 1.00E-04 | -3.926 | 28105 | 28335 | 586871 |
| TCL&TSL | Visayan | Mbuti | -0.000369 | 0.000105 | -3.519 | 27927 | 28143 | 586871 |
| TCL&TSL | Hmong | Mbuti | -5.00E-06 | 0.000102 | -0.048 | 28374 | 28377 | 590667 |
| TCL&TSL | Hmong | Mbuti | -5.00E-06 | 0.000102 | -0.048 | 28374 | 28377 | 590667 |
| TCL&TSL | Sherpa_Nepal | Mbuti | 0.001562 | 0.000106 | 14.747 | 28739 | 27816 | 590888 |
| TCL&TSL | Tibetan_Nagqu | Mbuti | 0.001797 | 1.00E-04 | 17.896 | 28833 | 27771 | 590888 |
| TCL&TSL | Tibetan_Shannan | Mbuti | 0.001704 | 9.60E-05 | 17.747 | 28770 | 27763 | 590888 |
| TCL&TSL | Dong_Hunan | Mbuti | -0.000164 | 0.000103 | -1.586 | 28340 | 28437 | 590888 |
| TCL&TSL | Qiang_Danba | Mbuti | 0.00179 | 0.000109 | 16.395 | 28927 | 27870 | 590889 |
| TCL&TSL | Qiang_Daofu | Mbuti | 0.001846 | 1.00E-04 | 18.431 | 28903 | 27812 | 590889 |
| TCL&TSL | Tamang_Nepal | Mbuti | 0.001042 | 9.80E-05 | 10.626 | 28366 | 27751 | 590889 |
| TCL&TSL | Tibetan_Chamdo | Mbuti | 0.001867 | 9.70E-05 | 19.197 | 28866 | 27763 | 590889 |
| TCL&TSL | Tibetan_Gangcha | Mbuti | 0.00163 | 8.90E-05 | 18.215 | 28728 | 27765 | 590889 |
| TCL&TSL | Tibetan_Gannan | Mbuti | 0.001213 | 0.000102 | 11.913 | 28609 | 27893 | 590889 |
| TCL&TSL | Tibetan_Lhasa | Mbuti | 0.001737 | 9.70E-05 | 17.965 | 28807 | 27781 | 590889 |
| TCL&TSL | Tibetan_Shigatse | Mbuti | 0.00168 | 9.40E-05 | 17.813 | 28776 | 27783 | 590889 |
| TCL&TSL | Tibetan_Xinlong | Mbuti | 0.000699 | 0.000102 | 6.826 | 28587 | 28174 | 590889 |
| TCL&TSL | Tibetan_Xunhua | Mbuti | 0.001372 | 0.000106 | 12.896 | 28689 | 27879 | 590889 |
| TCL&TSL | Tibetan_Yajiang | Mbuti | 0.001602 | 0.000103 | 15.621 | 28818 | 27871 | 590889 |
| TCL&TSL | TibetanHO_Yunnan | Mbuti | 0.001311 | 0.00011 | 11.942 | 28748 | 27973 | 590889 |
| TCL&TSL | Gelao_Longlin | Mbuti | -0.000382 | 9.90E-05 | -3.87 | 28279 | 28505 | 590889 |
| TCL&TSL | Maonan_Huanjiang | Mbuti | -0.000513 | 9.80E-05 | -5.226 | 28243 | 28546 | 590889 |
| TCL&TSL | Mulam_Luocheng | Mbuti | -0.000343 | 9.80E-05 | -3.503 | 28277 | 28479 | 590889 |
| TCL&TSL | Han_Hubei | Mbuti | 0.000636 | 0.000104 | 6.106 | 28632 | 28256 | 590889 |
| TCL&TSL | Han_Jiangsu | Mbuti | 0.000834 | 0.000103 | 8.119 | 28643 | 28150 | 590889 |
| TCL&TSL | Han_Shandong | Mbuti | 0.00104 | 1.00E-04 | 10.418 | 28684 | 28069 | 590889 |
| TCL&TSL | Han_Shanxi | Mbuti | 0.001109 | 9.90E-05 | 11.167 | 28713 | 28057 | 590889 |
| TCL&TSL | Han_SichuanHO | Mbuti | 0.000615 | 9.80E-05 | 6.292 | 28565 | 28201 | 590889 |
| TCL&TSL | Han_Zhejiang | Mbuti | 0.000756 | 0.000104 | 7.246 | 28607 | 28160 | 590889 |
| TCL&TSL | Dongxiang_Linxia | Mbuti | 0.000968 | 9.50E-05 | 10.204 | 28461 | 27889 | 590889 |
| TCL&TSL | Han_Taiwan | Mbuti | 0.000578 | 0.000108 | 5.345 | 28595 | 28252 | 593027 |
| TCL&TSL | Lue | Mbuti | -0.000782 | 0.000102 | -7.697 | 28154 | 28618 | 593033 |
| TCL&TSL | Shan | Mbuti | -0.000591 | 0.000102 | -5.809 | 28149 | 28500 | 593034 |
| TCL&TSL | Palaung | Mbuti | -0.000429 | 9.80E-05 | -4.353 | 28186 | 28440 | 593036 |
| TCL&TSL | IuMien | Mbuti | -0.000359 | 0.000101 | -3.568 | 28313 | 28526 | 593037 |
| TCL&TSL | IuMien | Mbuti | -0.000359 | 0.000101 | -3.568 | 28313 | 28526 | 593037 |
| TCL&TSL | Phuan | Mbuti | -0.000821 | 9.80E-05 | -8.333 | 28111 | 28598 | 593037 |
| TCL&TSL | Yuan | Mbuti | -0.00081 | 9.90E-05 | -8.178 | 28108 | 28588 | 593037 |
| TCL&TSL | Kalueang | Mbuti | -0.001364 | 9.80E-05 | -13.858 | 27869 | 28678 | 593038 |
| TCL&TSL | BlackTai | Mbuti | -0.000798 | 0.000101 | -7.891 | 28168 | 28641 | 593038 |
| TCL&TSL | Khmu | Mbuti | -0.001579 | 9.80E-05 | -16.136 | 27832 | 28769 | 593038 |
| TCL&TSL | Lawa_Eastern | Mbuti | -0.000937 | 9.70E-05 | -9.689 | 28004 | 28559 | 593038 |
| TCL&TSL | Lawa_Western | Mbuti | -0.00088 | 9.80E-05 | -8.932 | 28054 | 28576 | 593038 |
| TCL&TSL | KarenPadaung | Mbuti | -0.000676 | 0.000102 | -6.605 | 28114 | 28516 | 593039 |

| | | | | | | | | |
|---|---|---|---|---|---|---|---|---|
| TCL&TSL | KarenPwo | Mbuti | -0.000756 | 9.80E-05 | -7.693 | 28091 | 28539 | 593039 |
| TCL&TSL | KarenSkaw | Mbuti | -0.000734 | 0.000103 | -7.102 | 28076 | 28511 | 593039 |
| TCL&TSL | Lahu | Mbuti | -0.001179 | 0.000103 | -11.454 | 28050 | 28750 | 593039 |
| TCL&TSL | Lahu_HGDP | Mbuti | -0.001022 | 0.000106 | -9.643 | 28101 | 28707 | 593039 |
| TCL&TSL | Lisu | Mbuti | -0.000101 | 1.00E-04 | -1.004 | 28331 | 28391 | 593039 |
| TCL&TSL | Naxi_HGDP | Mbuti | 0.000876 | 9.80E-05 | 8.918 | 28639 | 28120 | 593039 |
| TCL&TSL | Tujia_HGDP | Mbuti | 0.000589 | 9.80E-05 | 5.99 | 28609 | 28260 | 593039 |
| TCL&TSL | Cambodian | Mbuti | -0.001195 | 9.40E-05 | -12.707 | 27848 | 28556 | 593039 |
| TCL&TSL | Kinh_HGDP | Mbuti | -0.000462 | 1.00E-04 | -4.633 | 28243 | 28517 | 593039 |
| TCL&TSL | Ami | Mbuti | -0.000304 | 0.000102 | -2.973 | 28318 | 28498 | 593039 |
| TCL&TSL | Atayal | Mbuti | -0.000239 | 0.000109 | -2.195 | 28325 | 28467 | 593039 |
| TCL&TSL | Dai_HGDP | Mbuti | -0.000679 | 1.00E-04 | -6.783 | 28184 | 28587 | 593039 |
| TCL&TSL | HmongDaw | Mbuti | -5.00E-06 | 0.000105 | -0.043 | 28430 | 28433 | 593039 |
| TCL&TSL | HmongDaw | Mbuti | -5.00E-06 | 0.000105 | -0.043 | 28430 | 28433 | 593039 |
| TCL&TSL | HmongNjua | Mbuti | -2.70E-05 | 0.000106 | -0.254 | 28422 | 28438 | 593039 |
| TCL&TSL | HmongNjua | Mbuti | -2.70E-05 | 0.000106 | -0.254 | 28422 | 28438 | 593039 |
| TCL&TSL | Thai | Mbuti | -0.000779 | 9.30E-05 | -8.409 | 28056 | 28518 | 593039 |
| TCL&TSL | Daur_HGDP | Mbuti | 0.0012 | 9.60E-05 | 12.502 | 28650 | 27938 | 593039 |
| TCL&TSL | Hezhen_HGDP | Mbuti | 0.001097 | 9.80E-05 | 11.138 | 28631 | 27981 | 593039 |
| TCL&TSL | Oroqen_HGDP | Mbuti | 0.001303 | 9.90E-05 | 13.211 | 28703 | 27930 | 593039 |
| TCL&TSL | Tu_HGDP | Mbuti | 0.001162 | 9.40E-05 | 12.42 | 28623 | 27933 | 593039 |
| TCL&TSL | Ulchi | Mbuti | 0.001283 | 9.70E-05 | 13.274 | 28665 | 27904 | 593039 |
| TCL&TSL | Xibo_HGDP | Mbuti | 0.00115 | 9.90E-05 | 11.673 | 28624 | 27942 | 593039 |
| TCL&TSL | Bru | Mbuti | -0.001855 | 0.000101 | -18.394 | 27696 | 28796 | 593039 |
| TCL&TSL | CentralThai | Mbuti | -0.000733 | 8.80E-05 | -8.372 | 27940 | 28374 | 593039 |
| TCL&TSL | HtinPray | Mbuti | -0.001859 | 0.000105 | -17.775 | 27717 | 28819 | 593039 |
| TCL&TSL | Khonmueang | Mbuti | -0.000695 | 9.50E-05 | -7.345 | 28135 | 28547 | 593039 |
| TCL&TSL | Khuen | Mbuti | -0.000611 | 9.90E-05 | -6.189 | 28218 | 28580 | 593039 |
| TCL&TSL | LaoIsan | Mbuti | -0.001215 | 9.30E-05 | -13.029 | 27916 | 28637 | 593039 |
| TCL&TSL | Laotian | Mbuti | -0.001069 | 9.40E-05 | -11.377 | 28034 | 28668 | 593039 |
| TCL&TSL | Mon | Mbuti | -0.000495 | 8.70E-05 | -5.685 | 27960 | 28253 | 593039 |
| TCL&TSL | Nyaw | Mbuti | -0.001267 | 9.80E-05 | -12.946 | 27938 | 28689 | 593039 |
| TCL&TSL | Phutai | Mbuti | -0.001101 | 0.000102 | -10.816 | 28020 | 28673 | 593039 |
| TCL&TSL | Saek | Mbuti | -0.001087 | 9.70E-05 | -11.177 | 28000 | 28644 | 593039 |
| TCL&TSL | Soa | Mbuti | -0.001687 | 9.80E-05 | -17.136 | 27739 | 28739 | 593039 |
| TCL&TSL | SouthernThai_AN | Mbuti | -0.000989 | 8.90E-05 | -11.062 | 27762 | 28348 | 593039 |
| TCL&TSL | SouthernThai_TK | Mbuti | -0.000658 | 8.90E-05 | -7.376 | 27874 | 28264 | 593039 |
| TEL&THL | Htin_Mal | Mbuti | 0.001411 | 9.10E-05 | 15.472 | 27843 | 27071 | 547429 |
| TEL&THL | Mlabri | Mbuti | 0.001447 | 0.000106 | 13.684 | 27784 | 26992 | 547429 |
| TEL&THL | Mongol_Minorities | Mbuti | 0.000209 | 8.20E-05 | 2.56 | 27893 | 27771 | 585221 |
| TEL&THL | Mongol_Sainnoyon | Mbuti | 0.00022 | 7.70E-05 | 2.867 | 27866 | 27738 | 585221 |
| TEL&THL | Mongol_Uuld | Mbuti | 0.000203 | 7.70E-05 | 2.628 | 27839 | 27720 | 585221 |
| TEL&THL | Mongol_Zasagt | Mbuti | 0.000252 | 8.00E-05 | 3.137 | 27887 | 27740 | 585221 |
| TEL&THL | Nanai | Mbuti | 0.000311 | 8.50E-05 | 3.666 | 28085 | 27903 | 585221 |
| TEL&THL | Negidal | Mbuti | 0.000329 | 9.30E-05 | 3.549 | 28074 | 27882 | 585221 |
| TEL&THL | Ilocano | Mbuti | 0.00172 | 0.000101 | 17.101 | 28330 | 27321 | 586875 |
| TEL&THL | Cong | Mbuti | 0.001007 | 8.30E-05 | 12.144 | 28173 | 27582 | 586881 |
| TEL&THL | HaNhi | Mbuti | 0.000701 | 8.10E-05 | 8.702 | 28114 | 27703 | 586881 |
| TEL&THL | LoLo | Mbuti | 0.001497 | 8.50E-05 | 17.693 | 28340 | 27461 | 586881 |
| TEL&THL | PhuLa | Mbuti | 0.001286 | 8.10E-05 | 15.965 | 28297 | 27542 | 586881 |
| TEL&THL | Sila | Mbuti | 0.000664 | 8.70E-05 | 7.633 | 28107 | 27717 | 586881 |
| TEL&THL | Vietnam_Lahu | Mbuti | 0.000902 | 8.40E-05 | 10.684 | 28158 | 27629 | 586881 |
| TEL&THL | KhoMu | Mbuti | 0.001221 | 8.50E-05 | 14.358 | 28163 | 27447 | 586881 |
| TEL&THL | Kinh_Vietnam | Mbuti | 0.001547 | 8.30E-05 | 18.724 | 28384 | 27476 | 586881 |
| TEL&THL | Vietnamese | Mbuti | 0.001563 | 8.20E-05 | 19.001 | 28389 | 27472 | 586881 |
| TEL&THL | CoLao | Mbuti | 0.001418 | 9.00E-05 | 15.693 | 28423 | 27591 | 586881 |
| TEL&THL | Dao | Mbuti | 0.001587 | 8.20E-05 | 19.254 | 28438 | 27507 | 586881 |
| TEL&THL | Dao | Mbuti | 0.001587 | 8.20E-05 | 19.254 | 28438 | 27507 | 586881 |
| TEL&THL | Dusun | Mbuti | 0.001678 | 8.40E-05 | 19.929 | 28332 | 27347 | 586881 |
| TEL&THL | Kankanaey | Mbuti | 0.001779 | 9.00E-05 | 19.876 | 28443 | 27399 | 586881 |
| TEL&THL | Murut | Mbuti | 0.001711 | 8.60E-05 | 19.985 | 28371 | 27367 | 586881 |
| TEL&THL | PaThen | Mbuti | 0.001368 | 8.50E-05 | 16.041 | 28346 | 27543 | 586881 |
| TEL&THL | PaThen | Mbuti | 0.001368 | 8.50E-05 | 16.041 | 28346 | 27543 | 586881 |
| TEL&THL | BoY | Mbuti | 0.001462 | 8.70E-05 | 16.771 | 28377 | 27519 | 586881 |
| TEL&THL | Cham | Mbuti | 0.001308 | 7.70E-05 | 17.015 | 28072 | 27304 | 586881 |
| TEL&THL | Ede | Mbuti | 0.001417 | 7.80E-05 | 18.231 | 28016 | 27184 | 586881 |
| TEL&THL | Giarai | Mbuti | 0.001379 | 7.90E-05 | 17.547 | 28052 | 27243 | 586881 |
| TEL&THL | LaChi | Mbuti | 0.001641 | 8.80E-05 | 18.713 | 28441 | 27478 | 586881 |
| TEL&THL | Malay | Mbuti | 0.001319 | 7.80E-05 | 17.011 | 28034 | 27260 | 586881 |
| TEL&THL | Mang | Mbuti | 0.001149 | 8.40E-05 | 13.753 | 28160 | 27485 | 586881 |
| TEL&THL | Muong | Mbuti | 0.001682 | 8.10E-05 | 20.739 | 28412 | 27425 | 586881 |
| TEL&THL | Nung | Mbuti | 0.001689 | 8.30E-05 | 20.441 | 28434 | 27443 | 586881 |
| TEL&THL | Tagalog | Mbuti | 0.001432 | 8.40E-05 | 17.014 | 28166 | 27326 | 586881 |
| TEL&THL | Tay | Mbuti | 0.001562 | 8.20E-05 | 19.039 | 28383 | 27466 | 586881 |
| TEL&THL | Visayan | Mbuti | 0.0016 | 8.70E-05 | 18.476 | 28210 | 27272 | 586881 |
| TEL&THL | Hmong | Mbuti | 0.001287 | 8.40E-05 | 15.332 | 28473 | 27713 | 590677 |
| TEL&THL | Hmong | Mbuti | 0.001287 | 8.40E-05 | 15.332 | 28473 | 27713 | 590677 |
| TEL&THL | Sherpa_Nepal | Mbuti | -0.000818 | 8.70E-05 | -9.379 | 27788 | 28272 | 590898 |
| TEL&THL | Tibetan_Nagqu | Mbuti | -0.000885 | 8.20E-05 | -10.838 | 27795 | 28318 | 590898 |

| | | | | | | | | |
|---|---|---|---|---|---|---|---|---|
| TEL&THL | Tibetan_Shannan | Mbuti | -0.00085 | 8.10E-05 | -10.463 | 27773 | 28275 | 590898 |
| TEL&THL | Dong_Hunan | Mbuti | 0.001412 | 8.50E-05 | 16.524 | 28523 | 27689 | 590898 |
| TEL&THL | Qiang_Danba | Mbuti | -0.000823 | 9.20E-05 | -8.939 | 27918 | 28405 | 590899 |
| TEL&THL | Qiang_Daofu | Mbuti | -0.000808 | 8.20E-05 | -9.809 | 27879 | 28356 | 590899 |
| TEL&THL | Tamang_Nepal | Mbuti | -0.000339 | 8.00E-05 | -4.235 | 27703 | 27904 | 590899 |
| TEL&THL | Tibetan_Chamdo | Mbuti | -0.000991 | 8.10E-05 | -12.289 | 27782 | 28367 | 590899 |
| TEL&THL | Tibetan_Gangcha | Mbuti | -0.00064 | 7.60E-05 | -8.388 | 27813 | 28191 | 590899 |
| TEL&THL | Tibetan_Gannan | Mbuti | 0.000273 | 8.70E-05 | 3.115 | 28081 | 27920 | 590899 |
| TEL&THL | Tibetan_Lhasa | Mbuti | -0.000853 | 8.40E-05 | -10.107 | 27797 | 28301 | 590899 |
| TEL&THL | Tibetan_Shigatse | Mbuti | -0.000781 | 8.20E-05 | -9.517 | 27802 | 28263 | 590899 |
| TEL&THL | Tibetan_Xinlong | Mbuti | 0.000821 | 8.70E-05 | 9.42 | 28362 | 27876 | 590899 |
| TEL&THL | Tibetan_Xunhua | Mbuti | -0.000158 | 9.20E-05 | -1.716 | 27987 | 28081 | 590899 |
| TEL&THL | Tibetan_Yajiang | Mbuti | -0.000657 | 8.40E-05 | -7.808 | 27911 | 28299 | 590899 |
| TEL&THL | TibetanHO_Yunnan | Mbuti | -0.000574 | 9.30E-05 | -6.177 | 27943 | 28282 | 590899 |
| TEL&THL | Gelao_Longlin | Mbuti | 0.00143 | 8.10E-05 | 17.688 | 28516 | 27671 | 590899 |
| TEL&THL | Maonan_Huanjiang | Mbuti | 0.001701 | 8.20E-05 | 20.794 | 28602 | 27597 | 590899 |
| TEL&THL | Mulam_Luocheng | Mbuti | 0.001652 | 7.90E-05 | 20.789 | 28578 | 27602 | 590899 |
| TEL&THL | Han_Hubei | Mbuti | 0.001093 | 8.40E-05 | 13 | 28504 | 27858 | 590899 |
| TEL&THL | Han_Jiangsu | Mbuti | 0.001029 | 8.20E-05 | 12.489 | 28448 | 27840 | 590899 |
| TEL&THL | Han_Shandong | Mbuti | 0.000784 | 8.20E-05 | 9.607 | 28360 | 27897 | 590899 |
| TEL&THL | Han_Shanxi | Mbuti | 0.000675 | 8.20E-05 | 8.209 | 28335 | 27936 | 590899 |
| TEL&THL | Han_SichuanHO | Mbuti | 0.001042 | 8.20E-05 | 12.664 | 28423 | 27808 | 590899 |
| TEL&THL | Han_Zhejiang | Mbuti | 0.001101 | 8.80E-05 | 12.497 | 28448 | 27798 | 590899 |
| TEL&THL | Dongxiang_Linxia | Mbuti | 0.000454 | 8.00E-05 | 5.67 | 28051 | 27783 | 590899 |
| TEL&THL | Han_Taiwan | Mbuti | 0.00125 | 9.30E-05 | 13.4 | 28534 | 27793 | 593034 |
| TEL&THL | Lue | Mbuti | 0.00157 | 8.00E-05 | 19.536 | 28540 | 27608 | 593043 |
| TEL&THL | Shan | Mbuti | 0.001023 | 7.80E-05 | 13.084 | 28321 | 27714 | 593044 |
| TEL&THL | Palaung | Mbuti | 0.00065 | 8.20E-05 | 7.892 | 28204 | 27819 | 593046 |
| TEL&THL | IuMien | Mbuti | 0.001357 | 8.30E-05 | 16.391 | 28524 | 27719 | 593047 |
| TEL&THL | IuMien | Mbuti | 0.001357 | 8.30E-05 | 16.391 | 28524 | 27719 | 593047 |
| TEL&THL | Phuan | Mbuti | 0.001618 | 8.50E-05 | 19.13 | 28521 | 27562 | 593047 |
| TEL&THL | Yuan | Mbuti | 0.001571 | 8.00E-05 | 19.556 | 28496 | 27564 | 593047 |
| TEL&THL | Kalueang | Mbuti | 0.001464 | 8.30E-05 | 17.596 | 28367 | 27499 | 593048 |
| TEL&THL | BlackTai | Mbuti | 0.001601 | 8.30E-05 | 19.324 | 28568 | 27618 | 593048 |
| TEL&THL | Khmu | Mbuti | 0.001269 | 8.20E-05 | 15.446 | 28332 | 27580 | 593048 |
| TEL&THL | Lawa_Eastern | Mbuti | 0.00079 | 8.10E-05 | 9.715 | 28194 | 27726 | 593048 |
| TEL&THL | Lawa_Western | Mbuti | 0.000775 | 8.00E-05 | 9.634 | 28220 | 27761 | 593048 |
| TEL&THL | KarenPadaung | Mbuti | 0.00068 | 7.90E-05 | 8.613 | 28198 | 27795 | 593049 |
| TEL&THL | KarenPwo | Mbuti | 0.000784 | 8.10E-05 | 9.702 | 28225 | 27760 | 593049 |
| TEL&THL | KarenSkaw | Mbuti | 8.00E-04 | 8.20E-05 | 9.752 | 28211 | 27737 | 593049 |
| TEL&THL | Lahu | Mbuti | 0.000983 | 8.10E-05 | 12.204 | 28358 | 27775 | 593049 |
| TEL&THL | Lahu_HGDP | Mbuti | 0.000959 | 8.20E-05 | 11.689 | 28363 | 27795 | 593049 |
| TEL&THL | Lisu | Mbuti | 0.000656 | 7.90E-05 | 8.299 | 28262 | 27873 | 593049 |
| TEL&THL | Naxi_HGDP | Mbuti | 7.50E-05 | 8.10E-05 | 0.923 | 28147 | 28102 | 593049 |
| TEL&THL | Tujia_HGDP | Mbuti | 0.001082 | 8.20E-05 | 13.198 | 28488 | 27847 | 593049 |
| TEL&THL | Cambodian | Mbuti | 0.001267 | 7.80E-05 | 16.218 | 28243 | 27492 | 593049 |
| TEL&THL | Kinh_HGDP | Mbuti | 0.001537 | 8.30E-05 | 18.584 | 28535 | 27623 | 593049 |
| TEL&THL | Ami | Mbuti | 0.001791 | 8.90E-05 | 20.08 | 28650 | 27588 | 593049 |
| TEL&THL | Atayal | Mbuti | 0.00181 | 9.00E-05 | 20.022 | 28635 | 27562 | 593049 |
| TEL&THL | Dai_HGDP | Mbuti | 0.001632 | 8.30E-05 | 19.76 | 28569 | 27601 | 593049 |
| TEL&THL | HmongDaw | Mbuti | 0.001336 | 8.60E-05 | 15.53 | 28546 | 27753 | 593049 |
| TEL&THL | HmongDaw | Mbuti | 0.001336 | 8.60E-05 | 15.53 | 28546 | 27753 | 593049 |
| TEL&THL | HmongNjua | Mbuti | 0.001308 | 8.90E-05 | 14.724 | 28533 | 27757 | 593049 |
| TEL&THL | HmongNjua | Mbuti | 0.001308 | 8.90E-05 | 14.724 | 28533 | 27757 | 593049 |
| TEL&THL | Thai | Mbuti | 0.001447 | 7.70E-05 | 18.783 | 28406 | 27548 | 593049 |
| TEL&THL | Daur_HGDP | Mbuti | 0.000489 | 7.80E-05 | 6.255 | 28189 | 27899 | 593049 |
| TEL&THL | Hezhen_HGDP | Mbuti | 0.000528 | 8.10E-05 | 6.513 | 28212 | 27899 | 593049 |
| TEL&THL | Oroqen_HGDP | Mbuti | 0.00036 | 7.90E-05 | 4.559 | 28170 | 27957 | 593049 |
| TEL&THL | Tu_HGDP | Mbuti | 0.000176 | 7.70E-05 | 2.283 | 28076 | 27972 | 593049 |
| TEL&THL | Ulchi | Mbuti | 0.000306 | 7.90E-05 | 3.866 | 28125 | 27944 | 593049 |
| TEL&THL | Xibo_HGDP | Mbuti | 0.000508 | 8.10E-05 | 6.299 | 28182 | 27881 | 593049 |
| TEL&THL | Bru | Mbuti | 0.001472 | 8.10E-05 | 18.082 | 28327 | 27454 | 593049 |
| TEL&THL | CentralThai | Mbuti | 0.001174 | 7.30E-05 | 16.065 | 28191 | 27495 | 593049 |
| TEL&THL | HtinPray | Mbuti | 0.001329 | 8.20E-05 | 16.129 | 28300 | 27512 | 593049 |
| TEL&THL | Khonmueang | Mbuti | 0.001454 | 7.70E-05 | 18.78 | 28466 | 27603 | 593049 |
| TEL&THL | Khuen | Mbuti | 0.001589 | 8.30E-05 | 19.024 | 28562 | 27620 | 593049 |
| TEL&THL | LaoIsan | Mbuti | 0.001449 | 7.60E-05 | 19.126 | 28374 | 27514 | 593049 |
| TEL&THL | Laotian | Mbuti | 0.001505 | 7.70E-05 | 19.521 | 28475 | 27583 | 593049 |
| TEL&THL | Mon | Mbuti | 0.000865 | 7.00E-05 | 12.435 | 28058 | 27544 | 593049 |
| TEL&THL | Nyaw | Mbuti | 0.001564 | 8.00E-05 | 19.599 | 28446 | 27518 | 593049 |
| TEL&THL | Phutai | Mbuti | 0.001479 | 8.00E-05 | 18.51 | 28457 | 27580 | 593049 |
| TEL&THL | Saek | Mbuti | 0.001547 | 8.30E-05 | 18.709 | 28460 | 27542 | 593049 |
| TEL&THL | Soa | Mbuti | 0.001446 | 8.10E-05 | 17.869 | 28319 | 27462 | 593049 |
| TEL&THL | SouthernThai_AN | Mbuti | 0.001187 | 7.10E-05 | 16.662 | 28084 | 27380 | 593049 |
| TEL&THL | SouthernThai_TK | Mbuti | 0.001051 | 7.30E-05 | 14.346 | 28072 | 27449 | 593049 |
| TEL&TSL | Htin_Mal | Mbuti | -0.000685 | 8.30E-05 | -8.283 | 27468 | 27843 | 547419 |
| TEL&TSL | Mlabri | Mbuti | -0.001079 | 9.60E-05 | -11.195 | 27305 | 27896 | 547419 |
| TEL&TSL | Mongol_Minorities | Mbuti | 0.001376 | 7.00E-05 | 19.74 | 28312 | 27507 | 585211 |
| TEL&TSL | Mongol_Sainnoyon | Mbuti | 0.001403 | 6.90E-05 | 20.299 | 28287 | 27466 | 585211 |

| | | | | | | | | |
|---|---|---|---|---|---|---|---|---|
| TEL&TSL | Mongol_Uuld | Mbuti | 0.001392 | 6.80E-05 | 20.421 | 28260 | 27445 | 585211 |
| TEL&TSL | Mongol_Zasagt | Mbuti | 0.001422 | 7.10E-05 | 20.17 | 28303 | 27471 | 585211 |
| TEL&TSL | Nanai | Mbuti | 0.001618 | 7.50E-05 | 21.461 | 28538 | 27591 | 585211 |
| TEL&TSL | Negidal | Mbuti | 0.001619 | 8.00E-05 | 20.277 | 28516 | 27568 | 585211 |
| TEL&TSL | Ilocano | Mbuti | 0.000921 | 8.80E-05 | 10.521 | 28251 | 27711 | 586865 |
| TEL&TSL | Cong | Mbuti | 0.000202 | 6.90E-05 | 2.939 | 28092 | 27973 | 586871 |
| TEL&TSL | HaNhi | Mbuti | 0.000176 | 7.10E-05 | 2.497 | 28106 | 28003 | 586871 |
| TEL&TSL | LoLo | Mbuti | 0.000372 | 7.30E-05 | 5.119 | 28178 | 27960 | 586871 |
| TEL&TSL | PhuLa | Mbuti | 0.000381 | 7.20E-05 | 5.255 | 28193 | 27970 | 586871 |
| TEL&TSL | Sila | Mbuti | 0.000251 | 7.70E-05 | 3.239 | 28130 | 27983 | 586871 |
| TEL&TSL | Vietnam_Lahu | Mbuti | 8.90E-05 | 7.20E-05 | 1.236 | 28077 | 28024 | 586871 |
| TEL&TSL | KhoMu | Mbuti | -0.000515 | 7.00E-05 | -7.312 | 27850 | 28152 | 586871 |
| TEL&TSL | Kinh_Vietnam | Mbuti | 0.000705 | 7.10E-05 | 9.993 | 28294 | 27880 | 586871 |
| TEL&TSL | Vietnamese | Mbuti | 0.000676 | 7.10E-05 | 9.587 | 28287 | 27890 | 586871 |
| TEL&TSL | CoLao | Mbuti | 0.000727 | 7.70E-05 | 9.5 | 28374 | 27947 | 586871 |
| TEL&TSL | Dao | Mbuti | 0.000852 | 7.30E-05 | 11.661 | 28375 | 27875 | 586871 |
| TEL&TSL | Dao | Mbuti | 0.000852 | 7.30E-05 | 11.661 | 28375 | 27875 | 586871 |
| TEL&TSL | Dusun | Mbuti | 0.00036 | 7.40E-05 | 4.836 | 28123 | 27912 | 586871 |
| TEL&TSL | Kankanaey | Mbuti | 0.001043 | 7.90E-05 | 13.256 | 28382 | 27770 | 586871 |
| TEL&TSL | Murut | Mbuti | 0.000356 | 7.60E-05 | 4.678 | 28150 | 27941 | 586871 |
| TEL&TSL | PaThen | Mbuti | 0.001011 | 7.60E-05 | 13.331 | 28379 | 27786 | 586871 |
| TEL&TSL | PaThen | Mbuti | 0.001011 | 7.60E-05 | 13.331 | 28379 | 27786 | 586871 |
| TEL&TSL | BoY | Mbuti | 0.001052 | 7.80E-05 | 13.472 | 28394 | 27777 | 586871 |
| TEL&TSL | Cham | Mbuti | -6.20E-05 | 6.80E-05 | -0.919 | 27851 | 27887 | 586871 |
| TEL&TSL | Ede | Mbuti | -0.000791 | 7.10E-05 | -11.127 | 27581 | 28046 | 586871 |
| TEL&TSL | Giarai | Mbuti | -0.000641 | 7.00E-05 | -9.185 | 27666 | 28042 | 586871 |
| TEL&TSL | LaChi | Mbuti | 0.000604 | 7.60E-05 | 7.955 | 28304 | 27950 | 586871 |
| TEL&TSL | Malay | Mbuti | -9.00E-05 | 7.00E-05 | -1.284 | 27799 | 27852 | 586871 |
| TEL&TSL | Mang | Mbuti | -0.00021 | 7.70E-05 | -2.718 | 27939 | 28062 | 586871 |
| TEL&TSL | Muong | Mbuti | 0.000497 | 7.10E-05 | 7.039 | 28237 | 27945 | 586871 |
| TEL&TSL | Nung | Mbuti | 0.000737 | 7.10E-05 | 10.316 | 28319 | 27886 | 586871 |
| TEL&TSL | Tagalog | Mbuti | 0.000872 | 7.40E-05 | 11.778 | 28148 | 27636 | 586871 |
| TEL&TSL | Tay | Mbuti | 0.000812 | 7.20E-05 | 11.226 | 28318 | 27841 | 586871 |
| TEL&TSL | Visayan | Mbuti | 0.000807 | 8.00E-05 | 10.139 | 28135 | 27662 | 586871 |
| TEL&TSL | Hmong | Mbuti | 0.001038 | 7.10E-05 | 14.567 | 28537 | 27924 | 590667 |
| TEL&TSL | Hmong | Mbuti | 0.001038 | 7.10E-05 | 14.567 | 28537 | 27924 | 590667 |
| TEL&TSL | Sherpa_Nepal | Mbuti | 0.001218 | 7.80E-05 | 15.593 | 28431 | 27711 | 590888 |
| TEL&TSL | Tibetan_Nagqu | Mbuti | 0.001289 | 7.20E-05 | 17.869 | 28471 | 27710 | 590888 |
| TEL&TSL | Tibetan_Shannan | Mbuti | 0.001238 | 7.00E-05 | 17.755 | 28424 | 27693 | 590888 |
| TEL&TSL | Dong_Hunan | Mbuti | 0.000998 | 7.30E-05 | 13.621 | 28542 | 27952 | 590888 |
| TEL&TSL | Qiang_Danba | Mbuti | 0.00139 | 7.90E-05 | 17.578 | 28602 | 27781 | 590889 |
| TEL&TSL | Qiang_Daofu | Mbuti | 0.001439 | 7.20E-05 | 20.014 | 28576 | 27726 | 590889 |
| TEL&TSL | Tamang_Nepal | Mbuti | 0.000975 | 6.80E-05 | 14.288 | 28157 | 27581 | 590889 |
| TEL&TSL | Tibetan_Chamdo | Mbuti | 0.001317 | 7.20E-05 | 18.287 | 28492 | 27714 | 590889 |
| TEL&TSL | Tibetan_Gangcha | Mbuti | 0.001315 | 6.50E-05 | 20.083 | 28432 | 27655 | 590889 |
| TEL&TSL | Tibetan_Gannan | Mbuti | 0.001452 | 7.10E-05 | 20.56 | 28506 | 27648 | 590889 |
| TEL&TSL | Tibetan_Lhasa | Mbuti | 0.001284 | 6.90E-05 | 18.608 | 28462 | 27704 | 590889 |
| TEL&TSL | Tibetan_Shigatse | Mbuti | 0.00125 | 6.90E-05 | 18.208 | 28440 | 27701 | 590889 |
| TEL&TSL | Tibetan_Xinlong | Mbuti | 0.001413 | 7.50E-05 | 18.802 | 28639 | 27804 | 590889 |
| TEL&TSL | Tibetan_Xunhua | Mbuti | 0.001384 | 7.80E-05 | 17.66 | 28506 | 27688 | 590889 |
| TEL&TSL | Tibetan_Yajiang | Mbuti | 0.001385 | 7.40E-05 | 18.779 | 28554 | 27736 | 590889 |
| TEL&TSL | TibetanHO_Yunnan | Mbuti | 0.00116 | 7.90E-05 | 14.706 | 28506 | 27821 | 590889 |
| TEL&TSL | Gelao_Longlin | Mbuti | 0.000712 | 7.20E-05 | 9.939 | 28458 | 28038 | 590889 |
| TEL&TSL | Maonan_Huanjiang | Mbuti | 8.00E-04 | 7.00E-05 | 11.417 | 28496 | 28023 | 590889 |
| TEL&TSL | Mulam_Luocheng | Mbuti | 0.000943 | 6.90E-05 | 13.59 | 28522 | 27965 | 590889 |
| TEL&TSL | Han_Hubei | Mbuti | 0.001527 | 7.70E-05 | 19.846 | 28742 | 27839 | 590889 |
| TEL&TSL | Han_Jiangsu | Mbuti | 0.001673 | 7.60E-05 | 21.929 | 28735 | 27746 | 590889 |
| TEL&TSL | Han_Shandong | Mbuti | 0.001701 | 7.20E-05 | 23.78 | 28717 | 27712 | 590889 |
| TEL&TSL | Han_Shanxi | Mbuti | 0.00165 | 7.30E-05 | 22.69 | 28709 | 27734 | 590889 |
| TEL&TSL | Han_SichuanHO | Mbuti | 0.001393 | 7.10E-05 | 19.578 | 28639 | 27816 | 590889 |
| TEL&TSL | Han_Zhejiang | Mbuti | 0.001542 | 7.70E-05 | 20.018 | 28680 | 27770 | 590889 |
| TEL&TSL | Dongxiang_Linxia | Mbuti | 0.001334 | 7.10E-05 | 18.92 | 28397 | 27609 | 590889 |
| TEL&TSL | Han_Taiwan | Mbuti | 0.001502 | 7.90E-05 | 19.087 | 28718 | 27827 | 593027 |
| TEL&TSL | Lue | Mbuti | 0.000469 | 7.30E-05 | 6.401 | 28386 | 28107 | 593033 |
| TEL&TSL | Shan | Mbuti | 0.000312 | 7.10E-05 | 4.418 | 28265 | 28080 | 593034 |
| TEL&TSL | Palaung | Mbuti | 0.000269 | 7.30E-05 | 3.685 | 28233 | 28073 | 593036 |
| TEL&TSL | IuMien | Mbuti | 0.000691 | 7.20E-05 | 9.633 | 28478 | 28068 | 593037 |
| TEL&TSL | IuMien | Mbuti | 0.000691 | 7.20E-05 | 9.633 | 28478 | 28068 | 593037 |
| TEL&TSL | Phuan | Mbuti | 0.000422 | 7.00E-05 | 6.015 | 28344 | 28093 | 593037 |
| TEL&TSL | Yuan | Mbuti | 0.000368 | 7.10E-05 | 5.148 | 28314 | 28096 | 593037 |
| TEL&TSL | Kalueang | Mbuti | -0.000207 | 7.20E-05 | -2.876 | 28069 | 28192 | 593038 |
| TEL&TSL | BlackTai | Mbuti | 0.000435 | 7.30E-05 | 5.974 | 28398 | 28140 | 593038 |
| TEL&TSL | Khmu | Mbuti | -0.000522 | 7.00E-05 | -7.431 | 28000 | 28309 | 593038 |
| TEL&TSL | Lawa_Eastern | Mbuti | -0.000197 | 6.90E-05 | -2.844 | 28066 | 28183 | 593038 |
| TEL&TSL | Lawa_Western | Mbuti | -0.000162 | 7.40E-05 | -2.2 | 28109 | 28205 | 593038 |
| TEL&TSL | KarenPadaung | Mbuti | -6.10E-05 | 7.30E-05 | -0.832 | 28135 | 28171 | 593039 |
| TEL&TSL | KarenPwo | Mbuti | -0.000127 | 7.20E-05 | -1.754 | 28118 | 28193 | 593039 |
| TEL&TSL | KarenSkaw | Mbuti | -4.00E-05 | 7.60E-05 | -0.521 | 28123 | 28146 | 593039 |
| TEL&TSL | Lahu | Mbuti | -0.000304 | 7.40E-05 | -4.077 | 28160 | 28340 | 593039 |

| | | | | | | | | |
|---|---|---|---|---|---|---|---|---|
| TEL&TSL | Lahu_HGDP | Mbuti | -0.000127 | 7.70E-05 | -1.645 | 28217 | 28292 | 593039 |
| TEL&TSL | Lisu | Mbuti | 0.000554 | 7.10E-05 | 7.769 | 28362 | 28033 | 593039 |
| TEL&TSL | Naxi_HGDP | Mbuti | 0.001154 | 7.00E-05 | 16.525 | 28547 | 27862 | 593039 |
| TEL&TSL | Tujia_HGDP | Mbuti | 0.001413 | 7.30E-05 | 19.361 | 28700 | 27862 | 593039 |
| TEL&TSL | Cambodian | Mbuti | -0.00019 | 6.90E-05 | -2.753 | 27998 | 28110 | 593039 |
| TEL&TSL | Kinh_HGDP | Mbuti | 0.000686 | 7.10E-05 | 9.647 | 28443 | 28036 | 593039 |
| TEL&TSL | Ami | Mbuti | 0.001038 | 7.40E-05 | 14.107 | 28582 | 27966 | 593039 |
| TEL&TSL | Atayal | Mbuti | 0.001016 | 8.00E-05 | 12.705 | 28555 | 27952 | 593039 |
| TEL&TSL | Dai_HGDP | Mbuti | 0.000637 | 7.20E-05 | 8.906 | 28439 | 28061 | 593039 |
| TEL&TSL | HmongDaw | Mbuti | 0.00106 | 7.30E-05 | 14.444 | 28598 | 27970 | 593039 |
| TEL&TSL | HmongDaw | Mbuti | 0.00106 | 7.30E-05 | 14.444 | 28598 | 27970 | 593039 |
| TEL&TSL | HmongNjua | Mbuti | 0.001019 | 7.70E-05 | 13.285 | 28584 | 27980 | 593039 |
| TEL&TSL | HmongNjua | Mbuti | 0.001019 | 7.70E-05 | 13.285 | 28584 | 27980 | 593039 |
| TEL&TSL | Thai | Mbuti | 0.000366 | 6.60E-05 | 5.518 | 28255 | 28038 | 593039 |
| TEL&TSL | Daur_HGDP | Mbuti | 0.00163 | 7.00E-05 | 23.25 | 28604 | 27638 | 593039 |
| TEL&TSL | Hezhen_HGDP | Mbuti | 0.001551 | 7.20E-05 | 21.396 | 28596 | 27676 | 593039 |
| TEL&TSL | Oroqen_HGDP | Mbuti | 0.001576 | 7.20E-05 | 21.934 | 28609 | 27675 | 593039 |
| TEL&TSL | Tu_HGDP | Mbuti | 0.001433 | 6.70E-05 | 21.525 | 28522 | 27672 | 593039 |
| TEL&TSL | Ulchi | Mbuti | 0.001552 | 7.20E-05 | 21.559 | 28568 | 27648 | 593039 |
| TEL&TSL | Xibo_HGDP | Mbuti | 0.0016 | 7.20E-05 | 22.36 | 28588 | 27639 | 593039 |
| TEL&TSL | Bru | Mbuti | -0.000671 | 7.50E-05 | -8.995 | 27905 | 28303 | 593039 |
| TEL&TSL | CentralThai | Mbuti | 0.000199 | 6.30E-05 | 3.136 | 28066 | 27949 | 593039 |
| TEL&TSL | HtinPray | Mbuti | -0.000811 | 7.80E-05 | -10.436 | 27881 | 28362 | 593039 |
| TEL&TSL | Khonmueang | Mbuti | 0.000464 | 6.80E-05 | 6.863 | 28337 | 28062 | 593039 |
| TEL&TSL | Khuen | Mbuti | 0.000616 | 7.20E-05 | 8.606 | 28441 | 28076 | 593039 |
| TEL&TSL | LaoIsan | Mbuti | -7.30E-05 | 6.70E-05 | -1.089 | 28113 | 28156 | 593039 |
| TEL&TSL | Laotian | Mbuti | 0.000125 | 6.60E-05 | 1.895 | 28249 | 28174 | 593039 |
| TEL&TSL | Mon | Mbuti | 0.000259 | 6.30E-05 | 4.122 | 28028 | 27874 | 593039 |
| TEL&TSL | Nyaw | Mbuti | -5.10E-05 | 7.10E-05 | -0.72 | 28157 | 28187 | 593039 |
| TEL&TSL | Phutai | Mbuti | 4.00E-05 | 7.30E-05 | 0.556 | 28217 | 28193 | 593039 |
| TEL&TSL | Saek | Mbuti | 0.000117 | 7.20E-05 | 1.626 | 28218 | 28149 | 593039 |
| TEL&TSL | Soa | Mbuti | -0.000518 | 7.10E-05 | -7.287 | 27945 | 28252 | 593039 |
| TEL&TSL | SouthernThai_AN | Mbuti | -2.00E-05 | 6.50E-05 | -0.303 | 27902 | 27914 | 593039 |
| TEL&TSL | SouthernThai_TK | Mbuti | 0.000219 | 6.50E-05 | 3.351 | 27983 | 27853 | 593039 |
| THL&TSL | Htin_Mal | Mbuti | -0.002096 | 0.00011 | -19.065 | 27214 | 28361 | 547419 |
| THL&TSL | Mlabri | Mbuti | -0.002526 | 0.000127 | -19.858 | 27041 | 28424 | 547419 |
| THL&TSL | Mongol_Minorities | Mbuti | 0.001167 | 9.50E-05 | 12.269 | 28427 | 27744 | 585211 |
| THL&TSL | Mongol_Sainnoyon | Mbuti | 0.001183 | 9.20E-05 | 12.924 | 28402 | 27710 | 585211 |
| THL&TSL | Mongol_Uuld | Mbuti | 0.001189 | 9.10E-05 | 13.014 | 28378 | 27682 | 585211 |
| THL&TSL | Mongol_Zasagt | Mbuti | 0.00117 | 9.50E-05 | 12.267 | 28406 | 27721 | 585211 |
| THL&TSL | Nanai | Mbuti | 0.001308 | 9.90E-05 | 13.156 | 28622 | 27857 | 585211 |
| THL&TSL | Negidal | Mbuti | 0.00129 | 0.000107 | 12.059 | 28601 | 27846 | 585211 |
| THL&TSL | Ilocano | Mbuti | -8.00E-04 | 0.000118 | -6.751 | 27862 | 28331 | 586865 |
| THL&TSL | Cong | Mbuti | -0.000804 | 9.50E-05 | -8.436 | 27945 | 28417 | 586871 |
| THL&TSL | HaNhi | Mbuti | -0.000525 | 9.30E-05 | -5.648 | 28058 | 28366 | 586871 |
| THL&TSL | LoLo | Mbuti | -0.001125 | 9.80E-05 | -11.528 | 27863 | 28523 | 586871 |
| THL&TSL | PhuLa | Mbuti | -0.000906 | 9.60E-05 | -9.46 | 27951 | 28483 | 586871 |
| THL&TSL | Sila | Mbuti | -0.000413 | 0.000103 | -4.024 | 28091 | 28333 | 586871 |
| THL&TSL | Vietnam_Lahu | Mbuti | -0.000813 | 9.80E-05 | -8.254 | 27963 | 28440 | 586871 |
| THL&TSL | KhoMu | Mbuti | -0.001736 | 0.000101 | -17.226 | 27630 | 28649 | 586871 |
| THL&TSL | Kinh_Vietnam | Mbuti | -0.000842 | 9.50E-05 | -8.837 | 27964 | 28458 | 586871 |
| THL&TSL | Vietnamese | Mbuti | -0.000887 | 9.30E-05 | -9.497 | 27954 | 28474 | 586871 |
| THL&TSL | CoLao | Mbuti | -0.000691 | 1.00E-04 | -6.871 | 28087 | 28493 | 586871 |
| THL&TSL | Dao | Mbuti | -0.000735 | 9.50E-05 | -7.696 | 28032 | 28463 | 586871 |
| THL&TSL | Dao | Mbuti | -0.000735 | 9.50E-05 | -7.696 | 28032 | 28463 | 586871 |
| THL&TSL | Dusun | Mbuti | -0.001318 | 9.70E-05 | -13.598 | 27751 | 28524 | 586871 |
| THL&TSL | Kankanaey | Mbuti | -0.000737 | 0.000101 | -7.282 | 27975 | 28407 | 586871 |
| THL&TSL | Murut | Mbuti | -0.001355 | 9.90E-05 | -13.644 | 27766 | 28561 | 586871 |
| THL&TSL | PaThen | Mbuti | -0.000357 | 9.90E-05 | -3.6 | 28108 | 28318 | 586871 |
| THL&TSL | PaThen | Mbuti | -0.000357 | 9.90E-05 | -3.6 | 28108 | 28318 | 586871 |
| THL&TSL | BoY | Mbuti | -0.00041 | 0.000104 | -3.959 | 28095 | 28336 | 586871 |
| THL&TSL | Cham | Mbuti | -0.00137 | 9.00E-05 | -15.215 | 27600 | 28404 | 586871 |
| THL&TSL | Ede | Mbuti | -0.002208 | 9.10E-05 | -24.271 | 27293 | 28589 | 586871 |
| THL&TSL | Giarai | Mbuti | -0.002021 | 9.30E-05 | -21.634 | 27392 | 28578 | 586871 |
| THL&TSL | LaChi | Mbuti | -0.001037 | 0.000101 | -10.22 | 27942 | 28550 | 586871 |
| THL&TSL | Malay | Mbuti | -0.001408 | 9.00E-05 | -15.578 | 27546 | 28373 | 586871 |
| THL&TSL | Mang | Mbuti | -0.001359 | 1.00E-04 | -13.565 | 27742 | 28540 | 586871 |
| THL&TSL | Muong | Mbuti | -0.001185 | 9.30E-05 | -12.75 | 27863 | 28559 | 586871 |
| THL&TSL | Nung | Mbuti | -0.000952 | 9.40E-05 | -10.083 | 27942 | 28501 | 586871 |
| THL&TSL | Tagalog | Mbuti | -0.00056 | 9.50E-05 | -5.869 | 27856 | 28184 | 586871 |
| THL&TSL | Tay | Mbuti | -0.00075 | 9.50E-05 | -7.866 | 27984 | 28424 | 586871 |
| THL&TSL | Visayan | Mbuti | -0.000793 | 0.000102 | -7.765 | 27786 | 28252 | 586871 |
| THL&TSL | Hmong | Mbuti | -0.000248 | 9.90E-05 | -2.507 | 28289 | 28436 | 590667 |
| THL&TSL | Hmong | Mbuti | -0.000248 | 9.90E-05 | -2.507 | 28289 | 28436 | 590667 |
| THL&TSL | Sherpa_Nepal | Mbuti | 0.002037 | 0.000108 | 18.867 | 28894 | 27691 | 590888 |
| THL&TSL | Tibetan_Nagqu | Mbuti | 0.002174 | 1.00E-04 | 21.807 | 28960 | 27676 | 590888 |
| THL&TSL | Tibetan_Shannan | Mbuti | 0.002087 | 9.70E-05 | 21.604 | 28900 | 27666 | 590888 |
| THL&TSL | Dong_Hunan | Mbuti | -0.000414 | 9.90E-05 | -4.189 | 28256 | 28501 | 590888 |
| THL&TSL | Qiang_Danba | Mbuti | 0.002213 | 0.000112 | 19.779 | 29071 | 27763 | 590889 |

| | | | | | | | | |
|---|---|---|---|---|---|---|---|---|
| THL&TSL | Qiang_Daofu | Mbuti | 0.002248 | 9.80E-05 | 22.862 | 29037 | 27708 | 590889 |
| THL&TSL | Tamang_Nepal | Mbuti | 0.001314 | 9.50E-05 | 13.884 | 28459 | 27682 | 590889 |
| THL&TSL | Tibetan_Chamdo | Mbuti | 0.002308 | 9.60E-05 | 24.038 | 29015 | 27652 | 590889 |
| THL&TSL | Tibetan_Gangcha | Mbuti | 0.001955 | 8.90E-05 | 22.001 | 28837 | 27682 | 590889 |
| THL&TSL | Tibetan_Gannan | Mbuti | 0.00118 | 1.00E-04 | 11.828 | 28599 | 27902 | 590889 |
| THL&TSL | Tibetan_Lhasa | Mbuti | 0.002136 | 1.00E-04 | 21.357 | 28941 | 27679 | 590889 |
| THL&TSL | Tibetan_Shigatse | Mbuti | 0.00203 | 9.60E-05 | 21.196 | 28895 | 27696 | 590889 |
| THL&TSL | Tibetan_Xinlong | Mbuti | 0.000591 | 0.000102 | 5.772 | 28548 | 28199 | 590889 |
| THL&TSL | Tibetan_Xunhua | Mbuti | 0.001542 | 0.000105 | 14.66 | 28749 | 27838 | 590889 |
| THL&TSL | Tibetan_Yajiang | Mbuti | 0.002042 | 0.000101 | 20.175 | 28964 | 27757 | 590889 |
| THL&TSL | TibetanHO_Yunnan | Mbuti | 0.001734 | 0.000108 | 16.034 | 28890 | 27866 | 590889 |
| THL&TSL | Gelao_Longlin | Mbuti | -0.000718 | 9.30E-05 | -7.714 | 28163 | 28587 | 590889 |
| THL&TSL | Maonan_Huanjiang | Mbuti | -0.000901 | 9.40E-05 | -9.628 | 28113 | 28645 | 590889 |
| THL&TSL | Mulam_Luocheng | Mbuti | -0.00071 | 9.20E-05 | -7.685 | 28153 | 28572 | 590889 |
| THL&TSL | Han_Hubei | Mbuti | 0.000434 | 1.00E-04 | 4.348 | 28560 | 28304 | 590889 |
| THL&TSL | Han_Jiangsu | Mbuti | 0.000644 | 9.70E-05 | 6.625 | 28580 | 28200 | 590889 |
| THL&TSL | Han_Shandong | Mbuti | 0.000917 | 9.50E-05 | 9.632 | 28641 | 28099 | 590889 |
| THL&TSL | Han_Shanxi | Mbuti | 0.000975 | 9.60E-05 | 10.173 | 28668 | 28091 | 590889 |
| THL&TSL | Han_SichuanHO | Mbuti | 0.000351 | 9.50E-05 | 3.71 | 28476 | 28268 | 590889 |
| THL&TSL | Han_Zhejiang | Mbuti | 0.00044 | 0.000103 | 4.263 | 28501 | 28241 | 590889 |
| THL&TSL | Dongxiang_Linxia | Mbuti | 0.00088 | 9.80E-05 | 9.026 | 28432 | 27912 | 590889 |
| THL&TSL | Han_Taiwan | Mbuti | 0.000252 | 0.000106 | 2.379 | 28487 | 28338 | 593027 |
| THL&TSL | Lue | Mbuti | -0.001101 | 9.50E-05 | -11.639 | 28043 | 28696 | 593033 |
| THL&TSL | Shan | Mbuti | -0.000711 | 9.60E-05 | -7.439 | 28108 | 28529 | 593034 |
| THL&TSL | Palaung | Mbuti | -0.000381 | 9.50E-05 | -4.026 | 28200 | 28426 | 593036 |
| THL&TSL | IuMien | Mbuti | -0.000666 | 9.80E-05 | -6.796 | 28210 | 28605 | 593037 |
| THL&TSL | IuMien | Mbuti | -0.000666 | 9.80E-05 | -6.796 | 28210 | 28605 | 593037 |
| THL&TSL | Phuan | Mbuti | -0.001196 | 9.60E-05 | -12.508 | 27983 | 28692 | 593037 |
| THL&TSL | Yuan | Mbuti | -0.001204 | 9.60E-05 | -12.498 | 27971 | 28685 | 593037 |
| THL&TSL | Kalueang | Mbuti | -0.001671 | 9.50E-05 | -17.507 | 27762 | 28753 | 593038 |
| THL&TSL | BlackTai | Mbuti | -0.001165 | 9.50E-05 | -12.209 | 28044 | 28735 | 593038 |
| THL&TSL | Khmu | Mbuti | -0.001791 | 9.70E-05 | -18.536 | 27759 | 28821 | 593038 |
| THL&TSL | Lawa_Eastern | Mbuti | -0.000987 | 9.50E-05 | -10.445 | 27987 | 28573 | 593038 |
| THL&TSL | Lawa_Western | Mbuti | -0.000937 | 9.70E-05 | -9.618 | 28034 | 28590 | 593038 |
| THL&TSL | KarenPadaung | Mbuti | -0.00074 | 9.70E-05 | -7.663 | 28095 | 28534 | 593039 |
| THL&TSL | KarenPwo | Mbuti | -0.000911 | 9.60E-05 | -9.525 | 28042 | 28582 | 593039 |
| THL&TSL | KarenSkaw | Mbuti | -0.000839 | 1.00E-04 | -8.356 | 28041 | 28539 | 593039 |
| THL&TSL | Lahu | Mbuti | -0.001286 | 9.80E-05 | -13.176 | 28014 | 28777 | 593039 |
| THL&TSL | Lahu_HGDP | Mbuti | -0.001086 | 1.00E-04 | -10.836 | 28078 | 28722 | 593039 |
| THL&TSL | Lisu | Mbuti | -0.000103 | 9.50E-05 | -1.077 | 28327 | 28388 | 593039 |
| THL&TSL | Naxi_HGDP | Mbuti | 0.00108 | 9.40E-05 | 11.433 | 28709 | 28069 | 593039 |
| THL&TSL | Tujia_HGDP | Mbuti | 0.000331 | 9.60E-05 | 3.46 | 28521 | 28325 | 593039 |
| THL&TSL | Cambodian | Mbuti | -0.001456 | 9.30E-05 | -15.677 | 27757 | 28620 | 593039 |
| THL&TSL | Kinh_HGDP | Mbuti | -0.000851 | 9.70E-05 | -8.767 | 28111 | 28615 | 593039 |
| THL&TSL | Ami | Mbuti | -0.000752 | 9.70E-05 | -7.727 | 28166 | 28612 | 593039 |
| THL&TSL | Atayal | Mbuti | -0.000793 | 0.000103 | -7.69 | 28136 | 28607 | 593039 |
| THL&TSL | Dai_HGDP | Mbuti | -0.000995 | 9.50E-05 | -10.458 | 28074 | 28664 | 593039 |
| THL&TSL | HmongDaw | Mbuti | -0.000277 | 0.000102 | -2.708 | 28336 | 28500 | 593039 |
| THL&TSL | HmongDaw | Mbuti | -0.000277 | 0.000102 | -2.708 | 28336 | 28500 | 593039 |
| THL&TSL | HmongNjua | Mbuti | -0.000289 | 0.000107 | -2.703 | 28333 | 28504 | 593039 |
| THL&TSL | HmongNjua | Mbuti | -0.000289 | 0.000107 | -2.703 | 28333 | 28504 | 593039 |
| THL&TSL | Thai | Mbuti | -0.001082 | 9.00E-05 | -12.049 | 27952 | 28594 | 593039 |
| THL&TSL | Daur_HGDP | Mbuti | 0.001141 | 9.20E-05 | 12.354 | 28627 | 27950 | 593039 |
| THL&TSL | Hezhen_HGDP | Mbuti | 0.001023 | 9.40E-05 | 10.856 | 28606 | 27999 | 593039 |
| THL&TSL | Oroqen_HGDP | Mbuti | 0.001216 | 9.50E-05 | 12.76 | 28673 | 27952 | 593039 |
| THL&TSL | Tu_HGDP | Mbuti | 0.001257 | 9.20E-05 | 13.639 | 28651 | 27906 | 593039 |
| THL&TSL | Ulchi | Mbuti | 0.001246 | 9.40E-05 | 13.28 | 28652 | 27913 | 593039 |
| THL&TSL | Xibo_HGDP | Mbuti | 0.001092 | 9.50E-05 | 11.458 | 28603 | 27955 | 593039 |
| THL&TSL | Bru | Mbuti | -0.002143 | 9.50E-05 | -22.5 | 27595 | 28866 | 593039 |
| THL&TSL | CentralThai | Mbuti | -0.000975 | 8.30E-05 | -11.81 | 27856 | 28434 | 593039 |
| THL&TSL | HtinPray | Mbuti | -0.00214 | 1.00E-04 | -21.419 | 27623 | 28892 | 593039 |
| THL&TSL | Khonmueang | Mbuti | -0.00099 | 8.90E-05 | -11.159 | 28033 | 28621 | 593039 |
| THL&TSL | Khuen | Mbuti | -0.000973 | 9.50E-05 | -10.242 | 28092 | 28669 | 593039 |
| THL&TSL | LaoIsan | Mbuti | -0.001522 | 8.90E-05 | -17.184 | 27809 | 28712 | 593039 |
| THL&TSL | Laotian | Mbuti | -0.00138 | 8.90E-05 | -15.534 | 27927 | 28745 | 593039 |
| THL&TSL | Mon | Mbuti | -0.000606 | 8.30E-05 | -7.328 | 27921 | 28280 | 593039 |
| THL&TSL | Nyaw | Mbuti | -0.001616 | 9.30E-05 | -17.35 | 27817 | 28775 | 593039 |
| THL&TSL | Phutai | Mbuti | -0.001438 | 9.40E-05 | -15.249 | 27904 | 28757 | 593039 |
| THL&TSL | Saek | Mbuti | -0.00143 | 9.50E-05 | -14.99 | 27885 | 28733 | 593039 |
| THL&TSL | Soa | Mbuti | -0.001964 | 9.50E-05 | -20.72 | 27644 | 28808 | 593039 |
| THL&TSL | SouthernThai_AN | Mbuti | -0.001206 | 8.50E-05 | -14.213 | 27687 | 28402 | 593039 |
| THL&TSL | SouthernThai_TK | Mbuti | -0.000832 | 8.70E-05 | -9.602 | 27815 | 28309 | 593039 |

Table S6. Symmetric *f4*-statistic testing in the form of *f*4(TYC, TYC, Ancient, Mbuti)

| Population1 | Population2 | Reference population | Outgroup | f4 | stderr | Zscore | BABA | ABBA | nsnps |
|---|---|---|---|---|---|---|---|---|---|
| THL | TCL | Laos_LN_BA.SG | Mbuti.SDG | -0.000748 | 0.000195 | -3.843 | 7150 | 7229 | 106250 |
| THL | TCL | Laos_Hoabinhian.SG | Mbuti.SDG | 0.000372 | 0.000232 | 1.605 | 3727 | 3705 | 57253 |
| THL | TCL | Vietnam_N | Mbuti.SDG | -8.00E-04 | 0.00029 | -2.761 | 2251 | 2278 | 33814 |
| THL | TCL | Vietnam_BA | Mbuti.SDG | -0.000478 | 0.000281 | -1.705 | 3031 | 3052 | 44671 |
| THL | TCL | Thailand_BA | Mbuti.SDG | -0.000142 | 6.00E-04 | -0.236 | 497 | 498 | 7465 |
| THL | TCL | Thailand_IA | Mbuti.SDG | -0.002578 | 0.001013 | -2.545 | 172 | 179 | 2632 |
| THL | TCL | China_YR_LN | Mbuti.SDG | -0.000203 | 0.000128 | -1.592 | 10054 | 10084 | 146283 |
| THL | TCL | China_Upper_YR_LN | Mbuti.SDG | 0.000126 | 0.000139 | 0.907 | 10005 | 9987 | 144955 |
| THL | TCL | China_YR_MN | Mbuti.SDG | -0.000115 | 0.000144 | -0.797 | 10021 | 10038 | 145458 |
| THL | TCL | Haimenkou_10bp | Mbuti.SDG | 0.000416 | 0.000236 | 1.768 | 3697 | 3674 | 54038 |
| THL | TCL | Gaoshancheng_10bp | Mbuti.SDG | 2.10E-05 | 0.000211 | 0.097 | 4585 | 4583 | 66804 |
| THL | TCL | China_SEastAsia_Coastal_LN | Mbuti.SDG | -0.000914 | 0.000172 | -5.306 | 5876 | 5955 | 86701 |
| THL | TCL | China_SEastAsia_Island_LN | Mbuti.SDG | -0.000767 | 0.000537 | -1.43 | 644 | 652 | 9685 |
| THL | TCL | Shenxian | Mbuti.SDG | -0.000453 | 0.000217 | -2.088 | 5494 | 5531 | 81077 |
| THL | TCL | China_SEastAsia_Coastal_Historic | Mbuti.SDG | -0.000511 | 0.000293 | -1.745 | 2785 | 2806 | 40518 |
| THL | TCL | Layi | Mbuti.SDG | -0.000854 | 0.000352 | -2.427 | 1702 | 1724 | 25119 |
| THL | TCL | GaoHuaHua | Mbuti.SDG | -0.000614 | 0.000158 | -3.88 | 8595 | 8673 | 125753 |
| THL | TCL | LaCen | Mbuti.SDG | -0.000874 | 0.000194 | -4.501 | 7205 | 7297 | 105702 |
| THL | TCL | Yiyang | Mbuti.SDG | -0.001465 | 0.000296 | -4.944 | 2492 | 2545 | 36553 |
| THL | TCL | Russia_Afanasievo | Mbuti.SDG | 0.000133 | 0.000111 | 1.199 | 9420 | 9401 | 143288 |
| THL | TCL | Zongri5.1k | Mbuti.SDG | 0.000744 | 0.000209 | 3.564 | 7427 | 7346 | 108118 |
| THL | TCL | Zongri4.7k | Mbuti.SDG | 0.000505 | 0.000196 | 2.577 | 7945 | 7887 | 115835 |
| THL | TCL | Zongri4.5k | Mbuti.SDG | 0.000592 | 0.000143 | 4.143 | 9202 | 9123 | 133558 |
| THL | TSL | Laos_LN_BA.SG | Mbuti.SDG | -0.003192 | 0.000252 | -12.691 | 7131 | 7470 | 106250 |
| THL | TSL | Laos_Hoabinhian.SG | Mbuti.SDG | -0.000319 | 0.000284 | -1.124 | 3756 | 3775 | 57253 |
| THL | TSL | Vietnam_N | Mbuti.SDG | -0.002917 | 0.000348 | -8.39 | 2254 | 2353 | 33814 |
| THL | TSL | Vietnam_BA | Mbuti.SDG | -0.001719 | 0.000329 | -5.222 | 3043 | 3120 | 44671 |
| THL | TSL | Thailand_BA | Mbuti.SDG | -0.002593 | 0.000741 | -3.498 | 495 | 514 | 7465 |
| THL | TSL | Thailand_IA | Mbuti.SDG | -0.004361 | 0.001206 | -3.618 | 174 | 186 | 2632 |
| THL | TSL | China_YR_LN | Mbuti.SDG | 0.001683 | 0.000155 | 10.856 | 10303 | 10057 | 146283 |
| THL | TSL | China_Upper_YR_LN | Mbuti.SDG | 0.002416 | 0.000172 | 14.031 | 10278 | 9927 | 144955 |
| THL | TSL | China_YR_MN | Mbuti.SDG | 0.001771 | 0.000178 | 9.941 | 10271 | 10014 | 145458 |
| THL | TSL | Haimenkou_10bp | Mbuti.SDG | 0.00271 | 0.000281 | 9.636 | 3799 | 3653 | 54038 |
| THL | TSL | Gaoshancheng_10bp | Mbuti.SDG | 0.001894 | 0.000257 | 7.356 | 4700 | 4574 | 66804 |
| THL | TSL | China_SEastAsia_Coastal_LN | Mbuti.SDG | -0.001346 | 0.000204 | -6.603 | 5937 | 6053 | 86701 |
| THL | TSL | China_SEastAsia_Island_LN | Mbuti.SDG | -0.001366 | 0.000634 | -2.155 | 649 | 662 | 9685 |
| THL | TSL | Shenxian | Mbuti.SDG | -0.001317 | 0.000265 | -4.965 | 5533 | 5640 | 81077 |
| THL | TSL | China_SEastAsia_Coastal_Historic | Mbuti.SDG | 0.000555 | 0.000338 | 1.644 | 2840 | 2818 | 40518 |
| THL | TSL | Layi | Mbuti.SDG | -0.001853 | 0.000427 | -4.337 | 1714 | 1761 | 25119 |
| THL | TSL | GaoHuaHua | Mbuti.SDG | -0.000745 | 0.000201 | -3.713 | 8700 | 8793 | 125753 |
| THL | TSL | LaCen | Mbuti.SDG | -0.001896 | 0.000233 | -8.136 | 7254 | 7455 | 105702 |
| THL | TSL | Yiyang | Mbuti.SDG | -0.002277 | 0.000375 | -6.073 | 2512 | 2596 | 36553 |
| THL | TSL | Russia_Afanasievo | Mbuti.SDG | 0.000583 | 0.000134 | 4.342 | 9565 | 9481 | 143288 |
| THL | TSL | Zongri5.1k | Mbuti.SDG | 0.003011 | 0.000261 | 11.548 | 7627 | 7301 | 108118 |
| THL | TSL | Zongri4.7k | Mbuti.SDG | 0.002722 | 0.000241 | 11.275 | 8157 | 7842 | 115835 |
| THL | TSL | Zongri4.5k | Mbuti.SDG | 0.002793 | 0.000172 | 16.206 | 9449 | 9076 | 133558 |
| THL | TEL | Laos_LN_BA.SG | Mbuti.SDG | -0.001773 | 0.000205 | -8.663 | 7124 | 7313 | 106250 |
| THL | TEL | Laos_Hoabinhian.SG | Mbuti.SDG | 0.00037 | 0.000253 | 1.463 | 3741 | 3719 | 57253 |
| THL | TEL | Vietnam_N | Mbuti.SDG | -0.001889 | 0.000305 | -6.197 | 2246 | 2310 | 33814 |
| THL | TEL | Vietnam_BA | Mbuti.SDG | -0.002191 | 0.000291 | -7.533 | 3005 | 3103 | 44671 |
| THL | TEL | Thailand_BA | Mbuti.SDG | -0.001404 | 0.000663 | -2.117 | 494 | 505 | 7465 |
| THL | TEL | Thailand_IA | Mbuti.SDG | -0.002073 | 0.001029 | -2.015 | 174 | 179 | 2632 |
| THL | TEL | China_YR_LN | Mbuti.SDG | -0.000702 | 0.000135 | -5.216 | 10053 | 10156 | 146283 |
| THL | TEL | China_Upper_YR_LN | Mbuti.SDG | 0.000189 | 0.000148 | 1.279 | 10043 | 10015 | 144955 |
| THL | TEL | China_YR_MN | Mbuti.SDG | -0.00063 | 0.000153 | -4.116 | 10022 | 10113 | 145458 |
| THL | TEL | Haimenkou_10bp | Mbuti.SDG | 0.000979 | 0.00025 | 3.91 | 3727 | 3674 | 54038 |
| THL | TEL | Gaoshancheng_10bp | Mbuti.SDG | 9.70E-05 | 0.000223 | 0.434 | 4603 | 4597 | 66804 |
| THL | TEL | China_SEastAsia_Coastal_LN | Mbuti.SDG | -0.002363 | 0.000185 | -12.757 | 5842 | 6047 | 86701 |
| THL | TEL | China_SEastAsia_Island_LN | Mbuti.SDG | -0.001887 | 0.000568 | -3.321 | 642 | 661 | 9685 |
| THL | TEL | Shenxian | Mbuti.SDG | -0.001845 | 0.000234 | -7.871 | 5466 | 5615 | 81077 |
| THL | TEL | China_SEastAsia_Coastal_Historic | Mbuti.SDG | -0.001429 | 0.00031 | -4.614 | 2779 | 2836 | 40518 |
| THL | TEL | Layi | Mbuti.SDG | -0.00223 | 0.000371 | -6.013 | 1695 | 1751 | 25119 |
| THL | TEL | GaoHuaHua | Mbuti.SDG | -0.002094 | 0.000171 | -12.263 | 8542 | 8805 | 125753 |
| THL | TEL | LaCen | Mbuti.SDG | -0.002402 | 0.000206 | -11.649 | 7160 | 7414 | 105702 |
| THL | TEL | Yiyang | Mbuti.SDG | -0.002979 | 0.000316 | -9.418 | 2479 | 2588 | 36553 |
| THL | TEL | Russia_Afanasievo | Mbuti.SDG | 0.000196 | 0.000115 | 1.709 | 9454 | 9426 | 143288 |
| THL | TEL | Zongri5.1k | Mbuti.SDG | 0.001655 | 0.000224 | 7.372 | 7490 | 7311 | 108118 |
| THL | TEL | Zongri4.7k | Mbuti.SDG | 0.001093 | 0.00021 | 5.192 | 8001 | 7874 | 115835 |
| THL | TEL | Zongri4.5k | Mbuti.SDG | 0.001032 | 0.00015 | 6.86 | 9258 | 9120 | 133558 |
| TCL | THL | Laos_LN_BA.SG | Mbuti.SDG | 0.000748 | 0.000195 | 3.843 | 7229 | 7150 | 106250 |
| TCL | THL | Laos_Hoabinhian.SG | Mbuti.SDG | -0.000372 | 0.000232 | -1.605 | 3705 | 3727 | 57253 |
| TCL | THL | Vietnam_N | Mbuti.SDG | 8.00E-04 | 0.00029 | 2.761 | 2278 | 2251 | 33814 |
| TCL | THL | Vietnam_BA | Mbuti.SDG | 0.000478 | 0.000281 | 1.705 | 3052 | 3031 | 44671 |
| TCL | THL | Thailand_BA | Mbuti.SDG | 0.000142 | 6.00E-04 | 0.236 | 498 | 497 | 7465 |
| TCL | THL | Thailand_IA | Mbuti.SDG | 0.002578 | 0.001013 | 2.545 | 179 | 172 | 2632 |
| TCL | THL | China_YR_LN | Mbuti.SDG | 0.000203 | 0.000128 | 1.592 | 10084 | 10054 | 146283 |
| TCL | THL | China_Upper_YR_LN | Mbuti.SDG | -0.000126 | 0.000139 | -0.907 | 9987 | 10005 | 144955 |
| TCL | THL | China_YR_MN | Mbuti.SDG | 0.000115 | 0.000144 | 0.797 | 10038 | 10021 | 145458 |
| TCL | THL | Haimenkou_10bp | Mbuti.SDG | -0.000416 | 0.000236 | -1.768 | 3674 | 3697 | 54038 |
| TCL | THL | Gaoshancheng_10bp | Mbuti.SDG | -2.10E-05 | 0.000211 | -0.097 | 4583 | 4585 | 66804 |
| TCL | THL | China_SEastAsia_Coastal_LN | Mbuti.SDG | 0.000914 | 0.000172 | 5.306 | 5955 | 5876 | 86701 |

| | | | | | | | | | |
|---|---|---|---|---|---|---|---|---|---|
| TCL | THL | China_SEastAsia_Island_LN | Mbuti.SDG | 0.000767 | 0.000537 | 1.43 | 652 | 644 | 9685 |
| TCL | THL | Shenxian | Mbuti.SDG | 0.000453 | 0.000217 | 2.088 | 5531 | 5494 | 81077 |
| TCL | THL | China_SEastAsia_Coastal_Historic | Mbuti.SDG | 0.000511 | 0.000293 | 1.745 | 2806 | 2785 | 40518 |
| TCL | THL | Layi | Mbuti.SDG | 0.000854 | 0.000352 | 2.427 | 1724 | 1702 | 25119 |
| TCL | THL | GaoHuaHua | Mbuti.SDG | 0.000614 | 0.000158 | 3.88 | 8673 | 8595 | 125753 |
| TCL | THL | LaCen | Mbuti.SDG | 0.000874 | 0.000194 | 4.501 | 7297 | 7205 | 105702 |
| TCL | THL | Yiyang | Mbuti.SDG | 0.001465 | 0.000296 | 4.944 | 2545 | 2492 | 36553 |
| TCL | THL | Russia_Afanasievo | Mbuti.SDG | -0.000133 | 0.000111 | -1.199 | 9401 | 9420 | 143288 |
| TCL | THL | Zongri5.1k | Mbuti.SDG | -0.000744 | 0.000209 | -3.564 | 7346 | 7427 | 108118 |
| TCL | THL | Zongri4.7k | Mbuti.SDG | -0.000505 | 0.000196 | -2.577 | 7887 | 7945 | 115835 |
| TCL | THL | Zongri4.5k | Mbuti.SDG | -0.000592 | 0.000143 | -4.143 | 9123 | 9202 | 133558 |
| TCL | TSL | Laos_LN_BA.SG | Mbuti.SDG | -0.002445 | 0.000205 | -11.92 | 7165 | 7425 | 106253 |
| TCL | TSL | Laos_Hoabinhian.SG | Mbuti.SDG | -0.000691 | 0.000224 | -3.092 | 3741 | 3780 | 57253 |
| TCL | TSL | Vietnam_N | Mbuti.SDG | -0.002117 | 0.000281 | -7.546 | 2263 | 2334 | 33814 |
| TCL | TSL | Vietnam_BA | Mbuti.SDG | -0.00124 | 0.000272 | -4.556 | 3051 | 3107 | 44671 |
| TCL | TSL | Thailand_BA | Mbuti.SDG | -0.002452 | 0.000565 | -4.342 | 495 | 514 | 7465 |
| TCL | TSL | Thailand_IA | Mbuti.SDG | -0.001784 | 0.001047 | -1.704 | 177 | 182 | 2632 |
| TCL | TSL | China_YR_LN | Mbuti.SDG | 0.001887 | 0.000125 | 15.126 | 10300 | 10024 | 146284 |
| TCL | TSL | China_Upper_YR_LN | Mbuti.SDG | 0.002289 | 0.000139 | 16.458 | 10249 | 9917 | 144956 |
| TCL | TSL | China_YR_MN | Mbuti.SDG | 0.001886 | 0.000142 | 13.328 | 10259 | 9985 | 145459 |
| TCL | TSL | Haimenkou_10bp | Mbuti.SDG | 0.002294 | 0.000242 | 9.497 | 3778 | 3654 | 54038 |
| TCL | TSL | Gaoshancheng_10bp | Mbuti.SDG | 0.001874 | 0.000217 | 8.623 | 4684 | 4559 | 66804 |
| TCL | TSL | China_SEastAsia_Coastal_LN | Mbuti.SDG | -0.000432 | 0.000182 | -2.38 | 5972 | 6009 | 86702 |
| TCL | TSL | China_SEastAsia_Island_LN | Mbuti.SDG | -0.000598 | 0.000513 | -1.168 | 651 | 657 | 9685 |
| TCL | TSL | Shenxian | Mbuti.SDG | -0.000864 | 0.000226 | -3.827 | 5544 | 5614 | 81078 |
| TCL | TSL | China_SEastAsia_Coastal_Historic | Mbuti.SDG | 0.001067 | 0.000283 | 3.765 | 2847 | 2804 | 40519 |
| TCL | TSL | Layi | Mbuti.SDG | -0.000999 | 0.000351 | -2.849 | 1725 | 1750 | 25119 |
| TCL | TSL | GaoHuaHua | Mbuti.SDG | -0.000131 | 0.000163 | -0.801 | 8727 | 8744 | 125754 |
| TCL | TSL | LaCen | Mbuti.SDG | -0.001022 | 0.000199 | -5.141 | 7290 | 7398 | 105703 |
| TCL | TSL | Yiyang | Mbuti.SDG | -0.000812 | 0.000297 | -2.735 | 2539 | 2568 | 36554 |
| TCL | TSL | Russia_Afanasievo | Mbuti.SDG | 0.00045 | 0.00011 | 4.095 | 9538 | 9474 | 143289 |
| TCL | TSL | Zongri5.1k | Mbuti.SDG | 0.002267 | 0.000204 | 11.097 | 7565 | 7320 | 108119 |
| TCL | TSL | Zongri4.7k | Mbuti.SDG | 0.002217 | 0.000195 | 11.384 | 8110 | 7853 | 115836 |
| TCL | TSL | Zongri4.5k | Mbuti.SDG | 0.002202 | 0.000139 | 15.839 | 9387 | 9093 | 133559 |
| TCL | TEL | Laos_LN_BA.SG | Mbuti.SDG | -0.001025 | 0.000157 | -6.524 | 7163 | 7271 | 106251 |
| TCL | TEL | Laos_Hoabinhian.SG | Mbuti.SDG | -2.00E-06 | 0.000184 | -0.012 | 3725 | 3726 | 57253 |
| TCL | TEL | Vietnam_N | Mbuti.SDG | -0.001089 | 0.000231 | -4.711 | 2255 | 2292 | 33814 |
| TCL | TEL | Vietnam_BA | Mbuti.SDG | -0.001712 | 0.000219 | -7.829 | 3014 | 3091 | 44671 |
| TCL | TEL | Thailand_BA | Mbuti.SDG | -0.001262 | 0.00047 | -2.688 | 495 | 505 | 7465 |
| TCL | TEL | Thailand_IA | Mbuti.SDG | 0.000505 | 0.000834 | 0.606 | 177 | 175 | 2632 |
| TCL | TEL | China_YR_LN | Mbuti.SDG | -0.000498 | 9.90E-05 | -5.026 | 10055 | 10128 | 146284 |
| TCL | TEL | China_Upper_YR_LN | Mbuti.SDG | 6.30E-05 | 0.000114 | 0.55 | 10017 | 10008 | 144956 |
| TCL | TEL | China_YR_MN | Mbuti.SDG | -0.000515 | 0.000114 | -4.506 | 10013 | 10088 | 145459 |
| TCL | TEL | Haimenkou_10bp | Mbuti.SDG | 0.000563 | 0.000196 | 2.874 | 3707 | 3677 | 54038 |
| TCL | TEL | Gaoshancheng_10bp | Mbuti.SDG | 7.60E-05 | 0.000172 | 0.444 | 4589 | 4584 | 66804 |
| TCL | TEL | China_SEastAsia_Coastal_LN | Mbuti.SDG | -0.00145 | 0.000144 | -10.086 | 5878 | 6004 | 86702 |
| TCL | TEL | China_SEastAsia_Island_LN | Mbuti.SDG | -0.00112 | 0.00043 | -2.603 | 644 | 655 | 9685 |
| TCL | TEL | Shenxian | Mbuti.SDG | -0.001392 | 0.000187 | -7.436 | 5477 | 5590 | 81078 |
| TCL | TEL | China_SEastAsia_Coastal_Historic | Mbuti.SDG | -0.000918 | 0.000232 | -3.956 | 2786 | 2823 | 40519 |
| TCL | TEL | Layi | Mbuti.SDG | -0.001376 | 0.000289 | -4.755 | 1706 | 1741 | 25119 |
| TCL | TEL | GaoHuaHua | Mbuti.SDG | -0.00148 | 0.000128 | -11.535 | 8572 | 8758 | 125754 |
| TCL | TEL | LaCen | Mbuti.SDG | -0.001528 | 0.000155 | -9.853 | 7200 | 7361 | 105703 |
| TCL | TEL | Yiyang | Mbuti.SDG | -0.001514 | 0.000237 | -6.387 | 2507 | 2562 | 36554 |
| TCL | TEL | Russia_Afanasievo | Mbuti.SDG | 6.30E-05 | 8.80E-05 | 0.719 | 9430 | 9421 | 143289 |
| TCL | TEL | Zongri5.1k | Mbuti.SDG | 0.000911 | 0.000163 | 5.578 | 7431 | 7333 | 108119 |
| TCL | TEL | Zongri4.7k | Mbuti.SDG | 0.000588 | 0.000157 | 3.734 | 7956 | 7888 | 115836 |
| TCL | TEL | Zongri4.5k | Mbuti.SDG | 0.000441 | 0.000115 | 3.824 | 9199 | 9140 | 133559 |
| TSL | THL | Laos_LN_BA.SG | Mbuti.SDG | 0.003192 | 0.000252 | 12.691 | 7470 | 7131 | 106250 |
| TSL | THL | Laos_Hoabinhian.SG | Mbuti.SDG | 0.000319 | 0.000284 | 1.124 | 3775 | 3756 | 57253 |
| TSL | THL | Vietnam_N | Mbuti.SDG | 0.002917 | 0.000348 | 8.39 | 2353 | 2254 | 33814 |
| TSL | THL | Vietnam_BA | Mbuti.SDG | 0.001719 | 0.000329 | 5.222 | 3120 | 3043 | 44671 |
| TSL | THL | Thailand_BA | Mbuti.SDG | 0.002593 | 0.000741 | 3.498 | 514 | 495 | 7465 |
| TSL | THL | Thailand_IA | Mbuti.SDG | 0.004361 | 0.001206 | 3.618 | 186 | 174 | 2632 |
| TSL | THL | China_YR_LN | Mbuti.SDG | -0.001683 | 0.000155 | -10.856 | 10057 | 10303 | 146283 |
| TSL | THL | China_Upper_YR_LN | Mbuti.SDG | -0.002416 | 0.000172 | -14.031 | 9927 | 10278 | 144955 |
| TSL | THL | China_YR_MN | Mbuti.SDG | -0.001771 | 0.000178 | -9.941 | 10014 | 10271 | 145458 |
| TSL | THL | Haimenkou_10bp | Mbuti.SDG | -0.00271 | 0.000281 | -9.636 | 3653 | 3799 | 54038 |
| TSL | THL | Gaoshancheng_10bp | Mbuti.SDG | -0.001894 | 0.000257 | -7.356 | 4574 | 4700 | 66804 |
| TSL | THL | China_SEastAsia_Coastal_LN | Mbuti.SDG | 0.001346 | 0.000204 | 6.603 | 6053 | 5937 | 86701 |
| TSL | THL | China_SEastAsia_Island_LN | Mbuti.SDG | 0.001366 | 0.000634 | 2.155 | 662 | 649 | 9685 |
| TSL | THL | Shenxian | Mbuti.SDG | 0.001317 | 0.000265 | 4.965 | 5640 | 5533 | 81077 |
| TSL | THL | China_SEastAsia_Coastal_Historic | Mbuti.SDG | -0.000555 | 0.000338 | -1.644 | 2818 | 2840 | 40518 |
| TSL | THL | Layi | Mbuti.SDG | 0.001853 | 0.000427 | 4.337 | 1761 | 1714 | 25119 |
| TSL | THL | GaoHuaHua | Mbuti.SDG | 0.000745 | 0.000201 | 3.713 | 8793 | 8700 | 125753 |
| TSL | THL | LaCen | Mbuti.SDG | 0.001896 | 0.000233 | 8.136 | 7455 | 7254 | 105702 |
| TSL | THL | Yiyang | Mbuti.SDG | 0.002277 | 0.000375 | 6.073 | 2596 | 2512 | 36553 |
| TSL | THL | Russia_Afanasievo | Mbuti.SDG | -0.000583 | 0.000134 | -4.342 | 9481 | 9565 | 143288 |
| TSL | THL | Zongri5.1k | Mbuti.SDG | -0.003011 | 0.000261 | -11.548 | 7301 | 7627 | 108118 |
| TSL | THL | Zongri4.7k | Mbuti.SDG | -0.002722 | 0.000241 | -11.275 | 7842 | 8157 | 115835 |
| TSL | THL | Zongri4.5k | Mbuti.SDG | -0.002793 | 0.000172 | -16.206 | 9076 | 9449 | 133558 |
| TSL | TCL | Laos_LN_BA.SG | Mbuti.SDG | 0.002445 | 0.000205 | 11.92 | 7425 | 7165 | 106253 |
| TSL | TCL | Laos_Hoabinhian.SG | Mbuti.SDG | 0.000691 | 0.000224 | 3.092 | 3780 | 3741 | 57253 |
| TSL | TCL | Vietnam_N | Mbuti.SDG | 0.002117 | 0.000281 | 7.546 | 2334 | 2263 | 33814 |

| | | | | | | | | | |
|---|---|---|---|---|---|---|---|---|---|
| TSL | TCL | Vietnam_BA | Mbuti.SDG | 0.00124 | 0.000272 | 4.556 | 3107 | 3051 | 44671 |
| TSL | TCL | Thailand_BA | Mbuti.SDG | 0.002452 | 0.000565 | 4.342 | 514 | 495 | 7465 |
| TSL | TCL | Thailand_IA | Mbuti.SDG | 0.001784 | 0.001047 | 1.704 | 182 | 177 | 2632 |
| TSL | TCL | China_YR_LN | Mbuti.SDG | -0.001887 | 0.000125 | -15.126 | 10024 | 10300 | 146284 |
| TSL | TCL | China_Upper_YR_LN | Mbuti.SDG | -0.002289 | 0.000139 | -16.458 | 9917 | 10249 | 144956 |
| TSL | TCL | China_YR_MN | Mbuti.SDG | -0.001886 | 0.000142 | -13.328 | 9985 | 10259 | 145459 |
| TSL | TCL | Haimenkou_10bp | Mbuti.SDG | -0.002294 | 0.000242 | -9.497 | 3654 | 3778 | 54038 |
| TSL | TCL | Gaoshancheng_10bp | Mbuti.SDG | -0.001874 | 0.000217 | -8.623 | 4559 | 4684 | 66804 |
| TSL | TCL | China_SEastAsia_Coastal_LN | Mbuti.SDG | 0.000432 | 0.000182 | 2.38 | 6009 | 5972 | 86702 |
| TSL | TCL | China_SEastAsia_Island_LN | Mbuti.SDG | 0.000598 | 0.000513 | 1.168 | 657 | 651 | 9685 |
| TSL | TCL | Shenxian | Mbuti.SDG | 0.000864 | 0.000226 | 3.827 | 5614 | 5544 | 81078 |
| TSL | TCL | China_SEastAsia_Coastal_Historic | Mbuti.SDG | -0.001067 | 0.000283 | -3.765 | 2804 | 2847 | 40519 |
| TSL | TCL | Layi | Mbuti.SDG | 0.000999 | 0.000351 | 2.849 | 1750 | 1725 | 25119 |
| TSL | TCL | GaoHuaHua | Mbuti.SDG | 0.000131 | 0.000163 | 0.801 | 8744 | 8727 | 125754 |
| TSL | TCL | LaCen | Mbuti.SDG | 0.001022 | 0.000199 | 5.141 | 7398 | 7290 | 105703 |
| TSL | TCL | Yiyang | Mbuti.SDG | 0.000812 | 0.000297 | 2.735 | 2568 | 2539 | 36554 |
| TSL | TCL | Russia_Afanasievo | Mbuti.SDG | -0.00045 | 0.00011 | -4.095 | 9474 | 9538 | 143289 |
| TSL | TCL | Zongri5.1k | Mbuti.SDG | -0.002267 | 0.000204 | -11.097 | 7320 | 7565 | 108119 |
| TSL | TCL | Zongri4.7k | Mbuti.SDG | -0.002217 | 0.000195 | -11.384 | 7853 | 8110 | 115836 |
| TSL | TCL | Zongri4.5k | Mbuti.SDG | -0.002202 | 0.000139 | -15.839 | 9093 | 9387 | 133559 |
| TSL | TEL | Laos_LN_BA.SG | Mbuti.SDG | 0.001419 | 0.000187 | 7.572 | 7341 | 7190 | 106251 |
| TSL | TEL | Laos_Hoabinhian.SG | Mbuti.SDG | 0.000689 | 0.000208 | 3.305 | 3762 | 3722 | 57253 |
| TSL | TEL | Vietnam_N | Mbuti.SDG | 0.001028 | 0.000247 | 4.164 | 2310 | 2275 | 33814 |
| TSL | TEL | Vietnam_BA | Mbuti.SDG | -0.000472 | 0.000248 | -1.904 | 3058 | 3079 | 44671 |
| TSL | TEL | Thailand_BA | Mbuti.SDG | 0.00119 | 0.000543 | 2.19 | 507 | 498 | 7465 |
| TSL | TEL | Thailand_IA | Mbuti.SDG | 0.002289 | 0.000948 | 2.414 | 182 | 176 | 2632 |
| TSL | TEL | China_YR_LN | Mbuti.SDG | -0.002385 | 0.000112 | -21.311 | 9943 | 10291 | 146284 |
| TSL | TEL | China_Upper_YR_LN | Mbuti.SDG | -0.002227 | 0.000124 | -18.011 | 9873 | 10196 | 144956 |
| TSL | TEL | China_YR_MN | Mbuti.SDG | -0.002401 | 0.000127 | -18.861 | 9904 | 10254 | 145459 |
| TSL | TEL | Haimenkou_10bp | Mbuti.SDG | -0.001731 | 0.000209 | -8.28 | 3653 | 3747 | 54038 |
| TSL | TEL | Gaoshancheng_10bp | Mbuti.SDG | -0.001797 | 0.000186 | -9.673 | 4542 | 4662 | 66804 |
| TSL | TEL | China_SEastAsia_Coastal_LN | Mbuti.SDG | -0.001017 | 0.000159 | -6.392 | 5925 | 6013 | 86702 |
| TSL | TEL | China_SEastAsia_Island_LN | Mbuti.SDG | -0.000521 | 0.000493 | -1.057 | 650 | 655 | 9685 |
| TSL | TEL | Shenxian | Mbuti.SDG | -0.000528 | 0.000205 | -2.574 | 5541 | 5584 | 81078 |
| TSL | TEL | China_SEastAsia_Coastal_Historic | Mbuti.SDG | -0.001984 | 0.000251 | -7.897 | 2775 | 2855 | 40519 |
| TSL | TEL | Layi | Mbuti.SDG | -0.000377 | 0.00031 | -1.215 | 1727 | 1737 | 25119 |
| TSL | TEL | GaoHuaHua | Mbuti.SDG | -0.001349 | 0.000142 | -9.473 | 8619 | 8789 | 125754 |
| TSL | TEL | LaCen | Mbuti.SDG | -0.000506 | 0.000171 | -2.957 | 7294 | 7347 | 105703 |
| TSL | TEL | Yiyang | Mbuti.SDG | -0.000702 | 0.000263 | -2.666 | 2535 | 2560 | 36554 |
| TSL | TEL | Russia_Afanasievo | Mbuti.SDG | -0.000387 | 9.60E-05 | -4.028 | 9431 | 9487 | 143289 |
| TSL | TEL | Zongri5.1k | Mbuti.SDG | -0.001356 | 0.000194 | -7.002 | 7324 | 7470 | 108119 |
| TSL | TEL | Zongri4.7k | Mbuti.SDG | -0.001629 | 0.000173 | -9.398 | 7844 | 8033 | 115836 |
| TSL | TEL | Zongri4.5k | Mbuti.SDG | -0.001761 | 0.000122 | -14.429 | 9074 | 9310 | 133559 |
| TEL | THL | Laos_LN_BA.SG | Mbuti.SDG | 0.001773 | 0.000205 | 8.663 | 7313 | 7124 | 106250 |
| TEL | THL | Laos_Hoabinhian.SG | Mbuti.SDG | -0.00037 | 0.000253 | -1.463 | 3719 | 3741 | 57253 |
| TEL | THL | Vietnam_N | Mbuti.SDG | 0.001889 | 0.000305 | 6.197 | 2310 | 2246 | 33814 |
| TEL | THL | Vietnam_BA | Mbuti.SDG | 0.002191 | 0.000291 | 7.533 | 3103 | 3005 | 44671 |
| TEL | THL | Thailand_BA | Mbuti.SDG | 0.001404 | 0.000663 | 2.117 | 505 | 494 | 7465 |
| TEL | THL | Thailand_IA | Mbuti.SDG | 0.002073 | 0.001029 | 2.015 | 179 | 174 | 2632 |
| TEL | THL | China_YR_LN | Mbuti.SDG | 0.000702 | 0.000135 | 5.216 | 10156 | 10053 | 146283 |
| TEL | THL | China_Upper_YR_LN | Mbuti.SDG | -0.000189 | 0.000148 | -1.279 | 10015 | 10043 | 144955 |
| TEL | THL | China_YR_MN | Mbuti.SDG | 0.00063 | 0.000153 | 4.116 | 10113 | 10022 | 145458 |
| TEL | THL | Haimenkou_10bp | Mbuti.SDG | -0.000979 | 0.00025 | -3.91 | 3674 | 3727 | 54038 |
| TEL | THL | Gaoshancheng_10bp | Mbuti.SDG | -9.70E-05 | 0.000223 | -0.434 | 4597 | 4603 | 66804 |
| TEL | THL | China_SEastAsia_Coastal_LN | Mbuti.SDG | 0.002363 | 0.000185 | 12.757 | 6047 | 5842 | 86701 |
| TEL | THL | China_SEastAsia_Island_LN | Mbuti.SDG | 0.001887 | 0.000568 | 3.321 | 661 | 642 | 9685 |
| TEL | THL | Shenxian | Mbuti.SDG | 0.001845 | 0.000234 | 7.871 | 5615 | 5466 | 81077 |
| TEL | THL | China_SEastAsia_Coastal_Historic | Mbuti.SDG | 0.001429 | 0.00031 | 4.614 | 2836 | 2779 | 40518 |
| TEL | THL | Layi | Mbuti.SDG | 0.00223 | 0.000371 | 6.013 | 1751 | 1695 | 25119 |
| TEL | THL | GaoHuaHua | Mbuti.SDG | 0.002094 | 0.000171 | 12.263 | 8805 | 8542 | 125753 |
| TEL | THL | LaCen | Mbuti.SDG | 0.002402 | 0.000206 | 11.649 | 7414 | 7160 | 105702 |
| TEL | THL | Yiyang | Mbuti.SDG | 0.002979 | 0.000316 | 9.418 | 2588 | 2479 | 36553 |
| TEL | THL | Russia_Afanasievo | Mbuti.SDG | -0.000196 | 0.000115 | -1.709 | 9426 | 9454 | 143288 |
| TEL | THL | Zongri5.1k | Mbuti.SDG | -0.001655 | 0.000224 | -7.372 | 7311 | 7490 | 108118 |
| TEL | THL | Zongri4.7k | Mbuti.SDG | -0.001093 | 0.00021 | -5.192 | 7874 | 8001 | 115835 |
| TEL | THL | Zongri4.5k | Mbuti.SDG | -0.001032 | 0.00015 | -6.86 | 9120 | 9258 | 133558 |
| TEL | TCL | Laos_LN_BA.SG | Mbuti.SDG | 0.001025 | 0.000157 | 6.524 | 7271 | 7163 | 106251 |
| TEL | TCL | Laos_Hoabinhian.SG | Mbuti.SDG | 2.00E-06 | 0.000184 | 0.012 | 3726 | 3725 | 57253 |
| TEL | TCL | Vietnam_N | Mbuti.SDG | 0.001089 | 0.000231 | 4.711 | 2292 | 2255 | 33814 |
| TEL | TCL | Vietnam_BA | Mbuti.SDG | 0.001712 | 0.000219 | 7.829 | 3091 | 3014 | 44671 |
| TEL | TCL | Thailand_BA | Mbuti.SDG | 0.001262 | 0.00047 | 2.688 | 505 | 495 | 7465 |
| TEL | TCL | Thailand_IA | Mbuti.SDG | -0.000505 | 0.000834 | -0.606 | 175 | 177 | 2632 |
| TEL | TCL | China_YR_LN | Mbuti.SDG | 0.000498 | 9.90E-05 | 5.026 | 10128 | 10055 | 146284 |
| TEL | TCL | China_Upper_YR_LN | Mbuti.SDG | -6.30E-05 | 0.000114 | -0.55 | 10008 | 10017 | 144956 |
| TEL | TCL | China_YR_MN | Mbuti.SDG | 0.000515 | 0.000114 | 4.506 | 10088 | 10013 | 145459 |
| TEL | TCL | Haimenkou_10bp | Mbuti.SDG | -0.000563 | 0.000196 | -2.874 | 3677 | 3707 | 54038 |
| TEL | TCL | Gaoshancheng_10bp | Mbuti.SDG | -7.60E-05 | 0.000172 | -0.444 | 4584 | 4589 | 66804 |
| TEL | TCL | China_SEastAsia_Coastal_LN | Mbuti.SDG | 0.00145 | 0.000144 | 10.086 | 6004 | 5878 | 86702 |
| TEL | TCL | China_SEastAsia_Island_LN | Mbuti.SDG | 0.00112 | 0.00043 | 2.603 | 655 | 644 | 9685 |
| TEL | TCL | Shenxian | Mbuti.SDG | 0.001392 | 0.000187 | 7.436 | 5590 | 5477 | 81078 |
| TEL | TCL | China_SEastAsia_Coastal_Historic | Mbuti.SDG | 0.000918 | 0.000232 | 3.956 | 2823 | 2786 | 40519 |
| TEL | TCL | Layi | Mbuti.SDG | 0.001376 | 0.000289 | 4.755 | 1741 | 1706 | 25119 |
| TEL | TCL | GaoHuaHua | Mbuti.SDG | 0.00148 | 0.000128 | 11.535 | 8758 | 8572 | 125754 |

| | | | | | | | | | |
|---|---|---|---|---|---|---|---|---|---|
| TEL | TCL | LaCen | Mbuti.SDG | 0.001528 | 0.000155 | 9.853 | 7361 | 7200 | 105703 |
| TEL | TCL | Yiyang | Mbuti.SDG | 0.001514 | 0.000237 | 6.387 | 2562 | 2507 | 36554 |
| TEL | TCL | Russia_Afanasievo | Mbuti.SDG | -6.30E-05 | 8.80E-05 | -0.719 | 9421 | 9430 | 143289 |
| TEL | TCL | Zongri5.1k | Mbuti.SDG | -0.000911 | 0.000163 | -5.578 | 7333 | 7431 | 108119 |
| TEL | TCL | Zongri4.7k | Mbuti.SDG | -0.000588 | 0.000157 | -3.734 | 7888 | 7956 | 115836 |
| TEL | TCL | Zongri4.5k | Mbuti.SDG | -0.000441 | 0.000115 | -3.824 | 9140 | 9199 | 133559 |
| TEL | TSL | Laos_LN_BA.SG | Mbuti.SDG | -0.001419 | 0.000187 | -7.572 | 7190 | 7341 | 106251 |
| TEL | TSL | Laos_Hoabinhian.SG | Mbuti.SDG | -0.000689 | 0.000208 | -3.305 | 3722 | 3762 | 57253 |
| TEL | TSL | Vietnam_N | Mbuti.SDG | -0.001028 | 0.000247 | -4.164 | 2275 | 2310 | 33814 |
| TEL | TSL | Vietnam_BA | Mbuti.SDG | 0.000472 | 0.000248 | 1.904 | 3079 | 3058 | 44671 |
| TEL | TSL | Thailand_BA | Mbuti.SDG | -0.00119 | 0.000543 | -2.19 | 498 | 507 | 7465 |
| TEL | TSL | Thailand_IA | Mbuti.SDG | -0.002289 | 0.000948 | -2.414 | 176 | 182 | 2632 |
| TEL | TSL | China_YR_LN | Mbuti.SDG | 0.002385 | 0.000112 | 21.311 | 10291 | 9943 | 146284 |
| TEL | TSL | China_Upper_YR_LN | Mbuti.SDG | 0.002227 | 0.000124 | 18.011 | 10196 | 9873 | 144956 |
| TEL | TSL | China_YR_MN | Mbuti.SDG | 0.002401 | 0.000127 | 18.861 | 10254 | 9904 | 145459 |
| TEL | TSL | Haimenkou_10bp | Mbuti.SDG | 0.001731 | 0.000209 | 8.28 | 3747 | 3653 | 54038 |
| TEL | TSL | Gaoshancheng_10bp | Mbuti.SDG | 0.001797 | 0.000186 | 9.673 | 4662 | 4542 | 66804 |
| TEL | TSL | China_SEastAsia_Coastal_LN | Mbuti.SDG | 0.001017 | 0.000159 | 6.392 | 6013 | 5925 | 86702 |
| TEL | TSL | China_SEastAsia_Island_LN | Mbuti.SDG | 0.000521 | 0.000493 | 1.057 | 655 | 650 | 9685 |
| TEL | TSL | Shenxian | Mbuti.SDG | 0.000528 | 0.000205 | 2.574 | 5584 | 5541 | 81078 |
| TEL | TSL | China_SEastAsia_Coastal_Historic | Mbuti.SDG | 0.001984 | 0.000251 | 7.897 | 2855 | 2775 | 40519 |
| TEL | TSL | Layi | Mbuti.SDG | 0.000377 | 0.00031 | 1.215 | 1737 | 1727 | 25119 |
| TEL | TSL | GaoHuaHua | Mbuti.SDG | 0.001349 | 0.000142 | 9.473 | 8789 | 8619 | 125754 |
| TEL | TSL | LaCen | Mbuti.SDG | 0.000506 | 0.000171 | 2.957 | 7347 | 7294 | 105703 |
| TEL | TSL | Yiyang | Mbuti.SDG | 0.000702 | 0.000263 | 2.666 | 2560 | 2535 | 36554 |
| TEL | TSL | Russia_Afanasievo | Mbuti.SDG | 0.000387 | 9.60E-05 | 4.028 | 9487 | 9431 | 143289 |
| TEL | TSL | Zongri5.1k | Mbuti.SDG | 0.001356 | 0.000194 | 7.002 | 7470 | 7324 | 108119 |
| TEL | TSL | Zongri4.7k | Mbuti.SDG | 0.001629 | 0.000173 | 9.398 | 8033 | 7844 | 115836 |
| TEL | TSL | Zongri4.5k | Mbuti.SDG | 0.001761 | 0.000122 | 14.429 | 9310 | 9074 | 133559 |

Table S7. Asymmetry *f4*-statistic testing in the form of *f4*(ADT/BMT, Gaoshancheng/China_Upper_YR_LN, Ancient, Mbuti)

| Population1 | Population2 | Reference population | Outgroup | f4 | stderr | Zscore | BABA | ABBA | nsnps |
|---|---|---|---|---|---|---|---|---|---|
| Tibetan_Amdo_Dalu.SG | China_Upper_YR_LN | China_YR_MN | Mbuti.SDG | -0.001998 | 0.000392 | -5.103 | 9799 | 10087 | 143919 |
| Tibetan_Baima_Guoyuan.SG | China_Upper_YR_LN | Gaoshancheng_10bp | Mbuti.SDG | -0.002941 | 0.000586 | -5.016 | 4457 | 4652 | 66199 |
| Tibetan_Baima_Wujiao.SG | China_Upper_YR_LN | China_YR_MN | Mbuti.SDG | -0.001935 | 0.000401 | -4.828 | 9821 | 10100 | 143919 |
| Tibetan_Baima_Wujiao.SG | China_Upper_YR_LN | Gaoshancheng_10bp | Mbuti.SDG | -0.002878 | 0.000624 | -4.614 | 4451 | 4642 | 66199 |
| Tibetan_Amdo_Dalu.SG | China_Upper_YR_LN | Gaoshancheng_10bp | Mbuti.SDG | -0.002627 | 0.000576 | -4.563 | 4467 | 4641 | 66199 |
| Tibetan_Baima_Guoyuan.SG | China_Upper_YR_LN | China_YR_MN | Mbuti.SDG | -0.001694 | 0.000383 | -4.418 | 9829 | 10073 | 143920 |
| Tibetan_Amdo_Dalu.SG | China_Upper_YR_LN | China_YR_LN | Mbuti.SDG | -0.001483 | 0.000337 | -4.396 | 9865 | 10079 | 144721 |
| Tibetan_Amdo_Dalu.SG | China_Upper_YR_LN | Hezhen.SDG | Mbuti.SDG | -0.001274 | 0.000293 | -4.347 | 9873 | 10058 | 144946 |
| Tibetan_Baima_Guoyuan.SG | Gaoshancheng_10bp | China_Upper_YR_LN | Mbuti.SDG | -0.002881 | 0.000669 | -4.304 | 4461 | 4652 | 66199 |
| Tibetan_Baima_Wujiao.SG | China_Upper_YR_LN | Hezhen.SDG | Mbuti.SDG | -0.001298 | 0.000304 | -4.268 | 9892 | 10080 | 144946 |
| Tibetan_Baima_Guoyuan.SG | China_Upper_YR_LN | Hezhen.SDG | Mbuti.SDG | -0.001181 | 0.000286 | -4.135 | 9886 | 10057 | 144947 |
| Tibetan_Amdo_Dalu.SG | Gaoshancheng_10bp | China_Upper_YR_LN | Mbuti.SDG | -0.002749 | 0.000672 | -4.093 | 4459 | 4641 | 66199 |
| Tibetan_Baima_Wujiao.SG | Gaoshancheng_10bp | China_Upper_YR_LN | Mbuti.SDG | -0.002742 | 0.000679 | -4.037 | 4460 | 4642 | 66199 |
| Tibetan_Baima_Wujiao.SG | China_Upper_YR_LN | Zongri4.5k | Mbuti.SDG | -0.001553 | 0.000389 | -3.997 | 8993 | 9198 | 132308 |
| Tibetan_Baima_Guoyuan.SG | China_Upper_YR_LN | Zongri4.5k | Mbuti.SDG | -0.001392 | 0.000361 | -3.86 | 8998 | 9182 | 132309 |
| Tibetan_Baima_Guoyuan.SG | China_Upper_YR_LN | China_YR_LN | Mbuti.SDG | -0.001214 | 0.000317 | -3.835 | 9893 | 10068 | 144722 |
| Tibetan_Amdo_Dalu.SG | China_Upper_YR_LN | Han_Shandong | Mbuti.SDG | -0.001032 | 0.000278 | -3.708 | 9905 | 10055 | 144955 |
| Tibetan_Amdo_Dalu.SG | China_Upper_YR_LN | Han_Shanxi | Mbuti.SDG | -0.001015 | 0.000277 | -3.665 | 9901 | 10049 | 144955 |
| Tibetan_Amdo_Dalu.SG | China_Upper_YR_LN | Han_Henan | Mbuti.SDG | -0.000988 | 0.000276 | -3.573 | 9905 | 10048 | 144955 |
| Tibetan_Baima_Wujiao.SG | China_Upper_YR_LN | China_YR_LN | Mbuti.SDG | -0.001218 | 0.000344 | -3.543 | 9901 | 10077 | 144721 |
| Tibetan_Amdo_Dalu.SG | China_Upper_YR_LN | Han_Fujian | Mbuti.SDG | -0.000965 | 0.00028 | -3.444 | 9908 | 10048 | 144955 |
| Tibetan_Amdo_Dalu.SG | China_Upper_YR_LN | Oroqen.SDG | Mbuti.SDG | -0.001015 | 0.000297 | -3.418 | 9885 | 10033 | 144948 |
| Tibetan_Amdo_Dalu.SG | China_Upper_YR_LN | Han_Northwest | Mbuti.SDG | -0.00094 | 0.000277 | -3.395 | 9901 | 10037 | 144955 |
| Tibetan_Baima_Guoyuan.SG | Gaoshancheng_10bp | Zongri4.5k | Mbuti.SDG | -0.002289 | 0.000676 | -3.385 | 4169 | 4311 | 61933 |
| Tibetan_Baima_Wujiao.SG | China_Upper_YR_LN | Oroqen.SDG | Mbuti.SDG | -0.001026 | 0.000308 | -3.328 | 9903 | 10052 | 144948 |
| Tibetan_Amdo_Dalu.SG | China_Upper_YR_LN | She_Fujian | Mbuti.SDG | -0.001004 | 0.000305 | -3.289 | 9909 | 10055 | 144955 |
| Tibetan_Baima_Wujiao.SG | Gaoshancheng_10bp | Zongri4.5k | Mbuti.SDG | -0.00227 | 0.000701 | -3.237 | 4165 | 4306 | 61933 |
| Tibetan_Amdo_Dalu.SG | Gaoshancheng_10bp | China_YR_LN | Mbuti.SDG | -0.001851 | 0.000591 | -3.133 | 4508 | 4631 | 66726 |
| Tibetan_Baima_Guoyuan.SG | China_Upper_YR_LN | Oroqen.SDG | Mbuti.SDG | -0.000895 | 0.000287 | -3.121 | 9903 | 10033 | 144949 |
| Tibetan_Amdo_Dalu.SG | Gaoshancheng_10bp | China_YR_MN | Mbuti.SDG | -0.002104 | 0.000679 | -3.1 | 4488 | 4628 | 66507 |
| Tibetan_Amdo_Dalu.SG | China_Upper_YR_LN | Qiang_Gaoping.SG | Mbuti.SDG | -0.000922 | 0.000298 | -3.094 | 9902 | 10035 | 144955 |
| Tibetan_Amdo_Dalu.SG | China_Upper_YR_LN | Daur.SDG | Mbuti.SDG | -0.000904 | 0.000293 | -3.084 | 9890 | 10021 | 144947 |
| Tibetan_Baima_Wujiao.SG | China_Upper_YR_LN | Han_Shanxi | Mbuti.SDG | -0.000852 | 0.000286 | -2.976 | 9934 | 10058 | 144955 |
| Tibetan_Baima_Wujiao.SG | China_Upper_YR_LN | Han_Shandong | Mbuti.SDG | -0.000852 | 0.000288 | -2.96 | 9939 | 10063 | 144955 |
| Tibetan_Baima_Wujiao.SG | China_Upper_YR_LN | Daur.SDG | Mbuti.SDG | -0.000904 | 0.000311 | -2.91 | 9913 | 10044 | 144947 |
| Tibetan_Baima_Wujiao.SG | China_Upper_YR_LN | Han_Henan | Mbuti.SDG | -0.000829 | 0.000286 | -2.898 | 9937 | 10058 | 144955 |
| Tibetan_Amdo_Dalu.SG | China_Upper_YR_LN | Dong_Guizhou | Mbuti.SDG | -0.000827 | 0.000286 | -2.888 | 9909 | 10029 | 144955 |
| Tibetan_Amdo_Dalu.SG | Gaoshancheng_10bp | BaBanQinCen | Mbuti.SDG | -0.002533 | 0.000884 | -2.865 | 3034 | 3149 | 45363 |
| Tibetan_Amdo_Dalu.SG | China_Upper_YR_LN | KHV.SG | Mbuti.SDG | -0.000806 | 0.000284 | -2.842 | 9891 | 10008 | 144955 |
| Tibetan_Amdo_Dalu.SG | China_Upper_YR_LN | BaBanQinCen | Mbuti.SDG | -0.001363 | 0.000491 | -2.777 | 6411 | 6540 | 94245 |
| Tibetan_Amdo_Dalu.SG | China_Upper_YR_LN | Yiyang | Mbuti.SDG | -0.002297 | 0.00083 | -2.767 | 2460 | 2544 | 36252 |
| Tibetan_Baima_Wujiao.SG | China_Upper_YR_LN | Han_Northwest | Mbuti.SDG | -0.000791 | 0.000286 | -2.762 | 9933 | 10048 | 144955 |
| Tibetan_Baima_Guoyuan.SG | Gaoshancheng_10bp | Zongri5.1k | Mbuti.SDG | -0.002665 | 0.000965 | -2.76 | 3413 | 3549 | 50989 |
| Tibetan_Amdo_Dalu.SG | China_Upper_YR_LN | Tujia.SDG | Mbuti.SDG | -0.000786 | 0.000285 | -2.758 | 9913 | 10027 | 144947 |
| Tibetan_Amdo_Dalu.SG | China_Upper_YR_LN | Qiang_Liujia.SG | Mbuti.SDG | -0.000866 | 0.000314 | -2.755 | 9914 | 10040 | 144955 |
| Tibetan_Baima_Wujiao.SG | China_Upper_YR_LN | Tu.SDG | Mbuti.SDG | -0.000807 | 0.000293 | -2.753 | 9902 | 10019 | 144949 |
| Tibetan_Amdo_Dalu.SG | Gaoshancheng_10bp | Cambodian.SDG | Mbuti.SDG | -0.001414 | 0.000527 | -2.684 | 4482 | 4577 | 66803 |
| Tibetan_Amdo_Dalu.SG | Gaoshancheng_10bp | China_SEastAsia_Coastal_LN | Mbuti.SDG | -0.002431 | 0.000916 | -2.655 | 2818 | 2921 | 42135 |
| Tibetan_Amdo_Dalu.SG | China_Upper_YR_LN | Bai_Heqing.DG | Mbuti.SDG | -0.000757 | 0.000289 | -2.621 | 9911 | 10020 | 144915 |
| Tibetan_Baima_Guoyuan.SG | Gaoshancheng_10bp | Laos_Hoabinhian.SG | Mbuti.SDG | -0.003156 | 0.00121 | -2.609 | 1760 | 1849 | 28128 |
| Tibetan_Amdo_Dalu.SG | Gaoshancheng_10bp | Qiang_Liujia.SG | Mbuti.SDG | -0.001406 | 0.000546 | -2.575 | 4535 | 4629 | 66804 |
| Tibetan_Baima_Guoyuan.SG | China_Upper_YR_LN | Han_Shanxi | Mbuti.SDG | -0.000678 | 0.000264 | -2.565 | 9936 | 10034 | 144956 |
| Tibetan_Amdo_Dalu.SG | China_Upper_YR_LN | Qiang_Matou.SG | Mbuti.SDG | -0.000749 | 0.000293 | -2.558 | 9918 | 10026 | 144955 |
| Tibetan_Amdo_Dalu.SG | China_Upper_YR_LN | CDX.SG | Mbuti.SDG | -0.000721 | 0.000284 | -2.54 | 9899 | 10004 | 144955 |
| Tibetan_Amdo_Dalu.SG | China_Upper_YR_LN | Miao_Guizhou | Mbuti.SDG | -0.000722 | 0.000284 | -2.539 | 9910 | 10015 | 144955 |
| Tibetan_Amdo_Dalu.SG | Gaoshancheng_10bp | Zongri4.5k | Mbuti.SDG | -0.001701 | 0.000676 | -2.518 | 4192 | 4297 | 61933 |
| Tibetan_Amdo_Dalu.SG | Gaoshancheng_10bp | China_Lahu.SDG | Mbuti.SDG | -0.001382 | 0.000551 | -2.507 | 4510 | 4602 | 66804 |
| Tibetan_Baima_Guoyuan.SG | Gaoshancheng_10bp | China_YR_MN | Mbuti.SDG | -0.001696 | 0.000677 | -2.505 | 4502 | 4615 | 66507 |
| Tibetan_Amdo_Dalu.SG | China_Upper_YR_LN | Hani_Honghe.DG | Mbuti.SDG | -0.00072 | 0.000292 | -2.472 | 9882 | 9986 | 144908 |
| Tibetan_Baima_Guoyuan.SG | China_Upper_YR_LN | Daur.SDG | Mbuti.SDG | -0.000699 | 0.000283 | -2.471 | 9913 | 10015 | 144948 |
| Tibetan_Baima_Wujiao.SG | Gaoshancheng_10bp | China_YR_MN | Mbuti.SDG | -0.001731 | 0.000707 | -2.45 | 4494 | 4610 | 66507 |
| Tibetan_Baima_Guoyuan.SG | China_Upper_YR_LN | Han_Shandong | Mbuti.SDG | -0.000653 | 0.000267 | -2.443 | 9942 | 10037 | 144956 |
| Tibetan_Baima_Guoyuan.SG | Gaoshancheng_10bp | China_YR_LN | Mbuti.SDG | -0.001414 | 0.00058 | -2.439 | 4529 | 4623 | 66726 |
| Tibetan_Amdo_Dalu.SG | China_Upper_YR_LN | Qiang_Xiaoyuan.SG | Mbuti.SDG | -0.000715 | 0.000293 | -2.439 | 9923 | 10027 | 144955 |
| Tibetan_Baima_Guoyuan.SG | China_Upper_YR_LN | Han_Henan | Mbuti.SDG | -0.000631 | 0.000266 | -2.37 | 9941 | 10032 | 144956 |
| Tibetan_Amdo_Dalu.SG | China_Upper_YR_LN | Lahu_Lancang.DG | Mbuti.SDG | -0.000755 | 0.000321 | -2.352 | 9872 | 9981 | 144900 |
| Tibetan_Amdo_Dalu.SG | Gaoshancheng_10bp | Hani_Honghe.DG | Mbuti.SDG | -0.001247 | 0.000539 | -2.313 | 4515 | 4599 | 66781 |
| Tibetan_Baima_Wujiao.SG | Gaoshancheng_10bp | Zongri4.7k | Mbuti.SDG | -0.002267 | 0.000982 | -2.309 | 3599 | 3722 | 53950 |
| Tibetan_Baima_Guoyuan.SG | China_Upper_YR_LN | Han_Northwest | Mbuti.SDG | -0.000613 | 0.000266 | -2.309 | 9934 | 10023 | 144956 |
| Tibetan_Baima_Guoyuan.SG | China_Upper_YR_LN | Yi.SDG | Mbuti.SDG | -0.000631 | 0.000279 | -2.265 | 9936 | 10027 | 144946 |
| Tibetan_Baima_Guoyuan.SG | China_Upper_YR_LN | Tu.SDG | Mbuti.SDG | -0.000627 | 0.000278 | -2.256 | 9902 | 9993 | 144950 |
| Tibetan_Amdo_Dalu.SG | Gaoshancheng_10bp | Tujia.SDG | Mbuti.SDG | -0.001194 | 0.000535 | -2.23 | 4538 | 4618 | 66801 |
| Tibetan_Amdo_Dalu.SG | Gaoshancheng_10bp | Vietnam_N | Mbuti.SDG | -0.003461 | 0.001565 | -2.211 | 1170 | 1232 | 17832 |
| Tibetan_Baima_Guoyuan.SG | Gaoshancheng_10bp | Zongri4.7k | Mbuti.SDG | -0.002075 | 0.000939 | -2.21 | 3609 | 3721 | 53950 |
| Tibetan_Amdo_Dalu.SG | Gaoshancheng_10bp | CDX.SG | Mbuti.SDG | -0.001133 | 0.000514 | -2.204 | 4528 | 4603 | 66804 |
| Tibetan_Baima_Wujiao.SG | Gaoshancheng_10bp | China_YR_LN | Mbuti.SDG | -0.001333 | 0.000607 | -2.195 | 4530 | 4619 | 66726 |
| Tibetan_Baima_Wujiao.SG | Gaoshancheng_10bp | Laos_Hoabinhian.SG | Mbuti.SDG | -0.002769 | 0.001266 | -2.188 | 1770 | 1848 | 28128 |
| Tibetan_Amdo_Dalu.SG | China_Upper_YR_LN | Dai.SDG | Mbuti.SDG | -0.000652 | 3.00E-04 | -2.175 | 9877 | 9972 | 144944 |
| Tibetan_Amdo_Dalu.SG | Gaoshancheng_10bp | Wa_Cangyuan.DG | Mbuti.SDG | -0.001139 | 0.00054 | -2.11 | 4500 | 4576 | 66784 |
| Tibetan_Amdo_Dalu.SG | Gaoshancheng_10bp | Qiang_Gaoping.SG | Mbuti.SDG | -0.001144 | 0.000556 | -2.057 | 4532 | 4609 | 66804 |
| Tibetan_Amdo_Dalu.SG | China_Upper_YR_LN | Zongri4.5k | Mbuti.SDG | -0.000757 | 0.000368 | -2.057 | 9039 | 9140 | 132308 |
| Tibetan_Baima_Wujiao.SG | China_Upper_YR_LN | Bai_Heqing.DG | Mbuti.SDG | -0.000614 | 3.00E-04 | -2.048 | 9939 | 10028 | 144915 |
| Tibetan_Baima_Wujiao.SG | Gaoshancheng_10bp | Vietnam_N | Mbuti.SDG | -0.003229 | 0.001587 | -2.035 | 1170 | 1227 | 17832 |
| Tibetan_Baima_Wujiao.SG | China_Upper_YR_LN | Haimenkou_10bp | Mbuti.SDG | -0.00145 | 0.000722 | -2.008 | 3629 | 3707 | 53525 |
| Tibetan_Amdo_Dalu.SG | Gaoshancheng_10bp | Dong_Guizhou | Mbuti.SDG | -0.001047 | 0.000522 | -2.008 | 4535 | 4605 | 66804 |
| Tibetan_Amdo_Dalu.SG | Gaoshancheng_10bp | KHV.SG | Mbuti.SDG | -0.001026 | 0.000517 | -1.985 | 4529 | 4597 | 66804 |
| Tibetan_Baima_Guoyuan.SG | China_Upper_YR_LN | Bai_Heqing.DG | Mbuti.SDG | -0.000553 | 0.000279 | -1.983 | 9933 | 10013 | 144915 |
| Tibetan_Baima_Wujiao.SG | China_Upper_YR_LN | Qiang_Liujia.SG | Mbuti.SDG | -0.000622 | 0.000314 | -1.977 | 9956 | 10046 | 144955 |
| Tibetan_Amdo_Dalu.SG | Gaoshancheng_10bp | Han_Fujian | Mbuti.SDG | -0.001002 | 0.000512 | -1.958 | 4540 | 4607 | 66804 |
| Tibetan_Baima_Wujiao.SG | China_Upper_YR_LN | Qiang_Gaoping.SG | Mbuti.SDG | -0.000616 | 0.000315 | -1.955 | 9948 | 10038 | 144955 |
| Tibetan_Amdo_Dalu.SG | Gaoshancheng_10bp | Miao_Guizhou | Mbuti.SDG | -0.000988 | 0.000515 | -1.919 | 4536 | 4602 | 66804 |
| Tibetan_Amdo_Dalu.SG | Gaoshancheng_10bp | Laos_Hoabinhian.SG | Mbuti.SDG | -0.002353 | 0.001243 | -1.893 | 1768 | 1834 | 28128 |
| Tibetan_Amdo_Dalu.SG | China_Upper_YR_LN | China_SEastAsia_Coastal_LN | Mbuti.SDG | -0.000939 | 0.000496 | -1.893 | 5835 | 5916 | 85960 |
| Tibetan_Baima_Wujiao.SG | Gaoshancheng_10bp | China_SEastAsia_Coastal_LN | Mbuti.SDG | -0.001743 | 0.000957 | -1.821 | 2836 | 2909 | 42135 |
| Tibetan_Baima_Wujiao.SG | China_Upper_YR_LN | Han_Fujian | Mbuti.SDG | -0.000524 | 0.000288 | -1.821 | 9962 | 10038 | 144955 |

| | | | | | | | | | |
|---|---|---|---|---|---|---|---|---|---|
| Tibetan_Baima_Guoyuan.SG | Gaoshancheng_10bp | Yi.SDG | Mbuti.SDG | -0.000933 | 0.000514 | -1.817 | 4537 | 4599 | 66802 |
| Tibetan_Amdo_Dalu.SG | Gaoshancheng_10bp | Qiang_Xiaoyuan.SG | Mbuti.SDG | -0.00098 | 0.000546 | -1.795 | 4533 | 4599 | 66804 |
| Tibetan_Baima_Wujiao.SG | China_Upper_YR_LN | Yi_Zhaojue | Mbuti.SDG | -0.000524 | 0.000293 | -1.791 | 9952 | 10028 | 144955 |
| Tibetan_Baima_Guoyuan.SG | Gaoshancheng_10bp | Qiang_Liujia.SG | Mbuti.SDG | -0.00094 | 0.00053 | -1.775 | 4552 | 4615 | 66804 |
| Tibetan_Amdo_Dalu.SG | China_Upper_YR_LN | China_Lahu.SDG | Mbuti.SDG | -0.000534 | 0.000302 | -1.767 | 9887 | 9965 | 144954 |
| Tibetan_Amdo_Dalu.SG | China_Upper_YR_LN | Shenxian | Mbuti.SDG | -0.001002 | 0.000575 | -1.743 | 5436 | 5517 | 80394 |
| Tibetan_Amdo_Dalu.SG | Gaoshancheng_10bp | Han_Shanxi | Mbuti.SDG | -0.000885 | 0.000509 | -1.74 | 4543 | 4603 | 66804 |
| Tibetan_Baima_Wujiao.SG | China_Upper_YR_LN | Yi.SDG | Mbuti.SDG | -0.000529 | 0.000304 | -1.737 | 9961 | 10038 | 144945 |
| Tibetan_Amdo_Dalu.SG | Gaoshancheng_10bp | Bai_Heqing.DG | Mbuti.SDG | -0.000918 | 0.000529 | -1.736 | 4535 | 4596 | 66784 |
| Tibetan_Baima_Wujiao.SG | Gaoshancheng_10bp | Qiang_Liujia.SG | Mbuti.SDG | -0.000976 | 0.000563 | -1.736 | 4552 | 4618 | 66804 |
| Tibetan_Baima_Guoyuan.SG | China_Upper_YR_LN | Zongri5.1k | Mbuti.SDG | -0.000886 | 0.000511 | -1.733 | 7286 | 7381 | 107163 |
| Tibetan_Amdo_Dalu.SG | Gaoshancheng_10bp | Qiang_Matou.SG | Mbuti.SDG | -0.000899 | 0.000527 | -1.708 | 4539 | 4599 | 66804 |
| Tibetan_Baima_Guoyuan.SG | Gaoshancheng_10bp | China_Lahu.SDG | Mbuti.SDG | -0.000917 | 0.000538 | -1.706 | 4528 | 4590 | 66804 |
| Tibetan_Baima_Wujiao.SG | China_Upper_YR_LN | She_Fujian | Mbuti.SDG | -0.000533 | 0.000315 | -1.689 | 9965 | 10042 | 144955 |
| Tibetan_Amdo_Dalu.SG | Gaoshancheng_10bp | Vietnam_BA | Mbuti.SDG | -0.002329 | 0.001387 | -1.679 | 1536 | 1589 | 22955 |
| Tibetan_Amdo_Dalu.SG | Gaoshancheng_10bp | Han_Shandong | Mbuti.SDG | -0.000856 | 0.000514 | -1.667 | 4545 | 4602 | 66804 |
| Tibetan_Baima_Guoyuan.SG | China_Upper_YR_LN | Naxi.SDG | Mbuti.SDG | -0.000463 | 0.000279 | -1.659 | 9950 | 10017 | 144947 |
| Tibetan_Baima_Guoyuan.SG | China_Upper_YR_LN | Yi_Zhaojue | Mbuti.SDG | -0.000446 | 0.000269 | -1.657 | 9943 | 10007 | 144956 |
| Tibetan_Amdo_Dalu.SG | Gaoshancheng_10bp | Han_Northwest | Mbuti.SDG | -0.000842 | 0.00051 | -1.651 | 4540 | 4597 | 66804 |
| Tibetan_Baima_Wujiao.SG | China_Upper_YR_LN | Qiang_Matou.SG | Mbuti.SDG | -0.000497 | 0.000301 | -1.648 | 9955 | 10027 | 144955 |
| Tibetan_Amdo_Dalu.SG | Gaoshancheng_10bp | She_Fujian | Mbuti.SDG | -0.000881 | 0.000537 | -1.64 | 4549 | 4608 | 66804 |
| Tibetan_Baima_Guoyuan.SG | China_Upper_YR_LN | Haimenkou_10bp | Mbuti.SDG | -0.001073 | 0.000661 | -1.623 | 3618 | 3676 | 53525 |
| Tibetan_Amdo_Dalu.SG | Gaoshancheng_10bp | Dai.SDG | Mbuti.SDG | -0.000864 | 0.000533 | -1.622 | 4526 | 4584 | 66801 |
| Tibetan_Amdo_Dalu.SG | Gaoshancheng_10bp | Han_Henan | Mbuti.SDG | -0.000822 | 0.00051 | -1.611 | 4545 | 4600 | 66804 |
| Tibetan_Baima_Guoyuan.SG | Gaoshancheng_10bp | BaBanQinCen | Mbuti.SDG | -0.001434 | 0.000895 | -1.602 | 3059 | 3124 | 45363 |
| Tibetan_Baima_Guoyuan.SG | Gaoshancheng_10bp | Naxi.SDG | Mbuti.SDG | -0.000865 | 0.00054 | -1.602 | 4549 | 4607 | 66802 |
| Tibetan_Baima_Guoyuan.SG | Gaoshancheng_10bp | China_SEastAsia_Coastal_LN | Mbuti.SDG | -0.0015 | 0.000938 | -1.599 | 2843 | 2906 | 42135 |
| Tibetan_Baima_Wujiao.SG | Gaoshancheng_10bp | Hani_Honghe.DG | Mbuti.SDG | -0.000891 | 0.000558 | -1.596 | 4528 | 4588 | 66781 |
| Tibetan_Amdo_Dalu.SG | China_Upper_YR_LN | Tu.SDG | Mbuti.SDG | -0.00045 | 0.000283 | -1.593 | 9909 | 9974 | 144949 |
| Tibetan_Amdo_Dalu.SG | Gaoshancheng_10bp | Hezhen.SDG | Mbuti.SDG | -0.000848 | 0.000533 | -1.591 | 4541 | 4597 | 66802 |
| Tibetan_Amdo_Dalu.SG | Gaoshancheng_10bp | Laos_LN_BA.SG | Mbuti.SDG | -0.001397 | 0.00091 | -1.535 | 3274 | 3343 | 49381 |
| Tibetan_Baima_Guoyuan.SG | Gaoshancheng_10bp | Hani_Honghe.DG | Mbuti.SDG | -0.000812 | 0.00053 | -1.533 | 4532 | 4587 | 66781 |
| Tibetan_Amdo_Dalu.SG | Gaoshancheng_10bp | Oroqen.SDG | Mbuti.SDG | -0.000824 | 0.000539 | -1.529 | 4535 | 4590 | 66802 |
| Tibetan_Baima_Guoyuan.SG | Gaoshancheng_10bp | Russia_Afanasievo | Mbuti.SDG | -0.000838 | 0.00055 | -1.523 | 4285 | 4340 | 65760 |
| Tibetan_Baima_Guoyuan.SG | China_Upper_YR_LN | Qiang_Gaoping.SG | Mbuti.SDG | -0.000436 | 0.000287 | -1.518 | 9946 | 10009 | 144956 |
| Tibetan_Baima_Guoyuan.SG | Gaoshancheng_10bp | Hezhen.SDG | Mbuti.SDG | -0.000795 | 0.000527 | -1.508 | 4543 | 4596 | 66802 |
| Tibetan_Baima_Wujiao.SG | Gaoshancheng_10bp | Zongri5.1k | Mbuti.SDG | -0.001508 | 0.001002 | -1.505 | 3442 | 3519 | 50989 |
| Tibetan_Amdo_Dalu.SG | Gaoshancheng_10bp | Zongri4.7k | Mbuti.SDG | -0.001414 | 0.000944 | -1.498 | 3630 | 3706 | 53950 |
| Tibetan_Baima_Guoyuan.SG | Gaoshancheng_10bp | Oroqen.SDG | Mbuti.SDG | -0.000781 | 0.00053 | -1.472 | 4538 | 4591 | 66802 |
| Tibetan_Baima_Guoyuan.SG | Gaoshancheng_10bp | Wa_Cangyuan.DG | Mbuti.SDG | -0.000769 | 0.000524 | -1.468 | 4516 | 4567 | 66784 |
| Tibetan_Baima_Guoyuan.SG | Gaoshancheng_10bp | Cambodian.SDG | Mbuti.SDG | -0.000747 | 0.000511 | -1.463 | 4506 | 4556 | 66803 |
| Tibetan_Baima_Guoyuan.SG | China_Upper_YR_LN | Laos_Hoabinhian.SG | Mbuti.SDG | -0.000865 | 0.000598 | -1.446 | 3660 | 3709 | 56679 |
| Tibetan_Baima_Wujiao.SG | Gaoshancheng_10bp | Wa_Cangyuan.DG | Mbuti.SDG | -0.000797 | 0.000551 | -1.446 | 4512 | 4566 | 66784 |
| Tibetan_Amdo_Dalu.SG | Gaoshancheng_10bp | Yi.SDG | Mbuti.SDG | -0.000756 | 0.000524 | -1.444 | 4546 | 4597 | 66802 |
| Tibetan_Baima_Guoyuan.SG | Gaoshancheng_10bp | Yi_Zhaojue | Mbuti.SDG | -0.000725 | 0.000504 | -1.439 | 4550 | 4598 | 66804 |
| Tibetan_Amdo_Dalu.SG | Gaoshancheng_10bp | Yiyang | Mbuti.SDG | -0.002199 | 0.001533 | -1.435 | 1209 | 1249 | 17895 |
| Tibetan_Baima_Guoyuan.SG | Gaoshancheng_10bp | Haimenkou_10bp | Mbuti.SDG | -0.001706 | 0.001197 | -1.425 | 1833 | 1879 | 27072 |
| Tibetan_Baima_Wujiao.SG | China_Upper_YR_LN | Naxi.SDG | Mbuti.SDG | -0.00043 | 0.000306 | -1.404 | 9967 | 10030 | 144946 |
| Tibetan_Amdo_Dalu.SG | China_Upper_YR_LN | Haimenkou_10bp | Mbuti.SDG | -0.000945 | 0.000676 | -1.399 | 3629 | 3680 | 53525 |
| Tibetan_Baima_Wujiao.SG | China_Upper_YR_LN | Tujia.SDG | Mbuti.SDG | -0.000412 | 0.000298 | -1.38 | 9962 | 10021 | 144947 |
| Tibetan_Amdo_Dalu.SG | Gaoshancheng_10bp | GaoHuaHua | Mbuti.SDG | -0.001105 | 0.000808 | -1.366 | 3966 | 4030 | 58065 |
| Tibetan_Baima_Wujiao.SG | China_Upper_YR_LN | Dong_Guizhou | Mbuti.SDG | -0.000394 | 0.000289 | -1.365 | 9964 | 10021 | 144955 |
| Tibetan_Baima_Wujiao.SG | Gaoshancheng_10bp | Haimenkou_10bp | Mbuti.SDG | -0.001664 | 0.001224 | -1.36 | 1821 | 1866 | 27072 |
| Tibetan_Amdo_Dalu.SG | Gaoshancheng_10bp | Lahu_Lancang.DG | Mbuti.SDG | -0.000791 | 0.000582 | -1.359 | 4530 | 4583 | 66775 |
| Tibetan_Amdo_Dalu.SG | China_Upper_YR_LN | Cambodian.SDG | Mbuti.SDG | -0.000399 | 0.000293 | -1.359 | 9844 | 9902 | 144946 |
| Tibetan_Baima_Wujiao.SG | Gaoshancheng_10bp | BaBanQinCen | Mbuti.SDG | -0.001235 | 0.000916 | -1.348 | 3069 | 3125 | 45363 |
| Tibetan_Amdo_Dalu.SG | Gaoshancheng_10bp | Haimenkou_10bp | Mbuti.SDG | -0.00157 | 0.001173 | -1.338 | 1833 | 1876 | 27072 |
| Tibetan_Baima_Wujiao.SG | Gaoshancheng_10bp | Hezhen.SDG | Mbuti.SDG | -0.000742 | 0.000555 | -1.337 | 4544 | 4594 | 66802 |
| Tibetan_Baima_Wujiao.SG | Gaoshancheng_10bp | China_Lahu.SDG | Mbuti.SDG | -0.000753 | 0.000567 | -1.33 | 4537 | 4587 | 66804 |
| Tibetan_Baima_Wujiao.SG | Gaoshancheng_10bp | Yi.SDG | Mbuti.SDG | -0.000706 | 0.000533 | -1.325 | 4546 | 4594 | 66802 |
| Tibetan_Baima_Guoyuan.SG | Gaoshancheng_10bp | Bai_Heqing.DG | Mbuti.SDG | -0.000681 | 0.000517 | -1.318 | 4543 | 4589 | 66784 |
| Tibetan_Amdo_Dalu.SG | China_Upper_YR_LN | GaoHuaHua | Mbuti.SDG | -0.000541 | 0.000411 | -1.315 | 8528 | 8595 | 124512 |
| Tibetan_Amdo_Dalu.SG | China_Upper_YR_LN | Yi.SDG | Mbuti.SDG | -0.000378 | 0.000288 | -1.313 | 9952 | 10007 | 144945 |
| Tibetan_Baima_Wujiao.SG | Gaoshancheng_10bp | Vietnam_BA | Mbuti.SDG | -0.001831 | 0.001405 | -1.304 | 1547 | 1589 | 22955 |
| Tibetan_Baima_Wujiao.SG | China_Upper_YR_LN | Laos_Hoabinhian.SG | Mbuti.SDG | -0.000866 | 0.000666 | -1.3 | 3681 | 3730 | 56679 |
| Tibetan_Baima_Guoyuan.SG | Gaoshancheng_10bp | Vietnam_BA | Mbuti.SDG | -0.00176 | 0.001356 | -1.298 | 1544 | 1585 | 22955 |
| Tibetan_Amdo_Dalu.SG | China_Upper_YR_LN | LaCen | Mbuti.SDG | -0.00066 | 0.000518 | -1.274 | 7160 | 7229 | 104781 |
| Tibetan_Baima_Guoyuan.SG | Gaoshancheng_10bp | Vietnam_N | Mbuti.SDG | -0.001973 | 0.001552 | -1.271 | 1174 | 1209 | 17832 |
| Tibetan_Amdo_Dalu.SG | Gaoshancheng_10bp | Russia_Afanasievo | Mbuti.SDG | -0.000701 | 0.000556 | -1.26 | 4286 | 4332 | 65760 |
| Tibetan_Amdo_Dalu.SG | Gaoshancheng_10bp | Yi_Zhaojue | Mbuti.SDG | -0.00065 | 0.000518 | -1.256 | 4552 | 4596 | 66804 |
| Tibetan_Baima_Wujiao.SG | Gaoshancheng_10bp | Naxi.SDG | Mbuti.SDG | -0.000712 | 0.00057 | -1.249 | 4554 | 4602 | 66802 |
| Tibetan_Amdo_Dalu.SG | Gaoshancheng_10bp | Zongri5.1k | Mbuti.SDG | -0.001206 | 0.000969 | -1.245 | 3449 | 3510 | 50989 |
| Tibetan_Baima_Wujiao.SG | Gaoshancheng_10bp | Oroqen.SDG | Mbuti.SDG | -0.000694 | 0.000557 | -1.245 | 4542 | 4589 | 66802 |
| Tibetan_Baima_Wujiao.SG | Gaoshancheng_10bp | Yi_Zhaojue | Mbuti.SDG | -0.000662 | 0.000532 | -1.244 | 4553 | 4597 | 66804 |
| Tibetan_Baima_Wujiao.SG | China_Upper_YR_LN | Hani_Honghe.DG | Mbuti.SDG | -0.000384 | 0.000309 | -1.244 | 9928 | 9984 | 144908 |
| Tibetan_Baima_Guoyuan.SG | Gaoshancheng_10bp | Tu.SDG | Mbuti.SDG | -0.000619 | 0.000503 | -1.231 | 4537 | 4578 | 66802 |
| Tibetan_Baima_Wujiao.SG | Gaoshancheng_10bp | Russia_Afanasievo | Mbuti.SDG | -0.000714 | 0.000587 | -1.217 | 4284 | 4331 | 65760 |
| Tibetan_Baima_Wujiao.SG | Gaoshancheng_10bp | Tu.SDG | Mbuti.SDG | -0.00065 | 0.000535 | -1.215 | 4537 | 4580 | 66802 |
| Tibetan_Baima_Wujiao.SG | Gaoshancheng_10bp | Bai_Heqing.DG | Mbuti.SDG | -0.000662 | 0.000547 | -1.212 | 4546 | 4590 | 66784 |
| Tibetan_Baima_Wujiao.SG | China_Upper_YR_LN | Zongri5.1k | Mbuti.SDG | -0.000648 | 0.000535 | -1.212 | 7322 | 7392 | 107162 |
| Tibetan_Baima_Guoyuan.SG | China_Upper_YR_LN | Pumi_Lanping.DG | Mbuti.SDG | -0.000343 | 0.000285 | -1.202 | 9945 | 9995 | 144913 |
| Tibetan_Baima_Guoyuan.SG | Gaoshancheng_10bp | Pumi_Lanping.DG | Mbuti.SDG | -0.00063 | 0.000534 | -1.181 | 4544 | 4586 | 66783 |
| Tibetan_Baima_Guoyuan.SG | Gaoshancheng_10bp | Tujia.SDG | Mbuti.SDG | -0.000621 | 0.000526 | -1.18 | 4560 | 4601 | 66801 |
| Tibetan_Baima_Wujiao.SG | Gaoshancheng_10bp | Qiang_Gaoping.SG | Mbuti.SDG | -0.00067 | 0.000571 | -1.174 | 4551 | 4595 | 66804 |
| Tibetan_Amdo_Dalu.SG | China_Upper_YR_LN | Yi_Zhaojue | Mbuti.SDG | -0.000334 | 0.000285 | -1.171 | 9948 | 9996 | 144955 |
| Tibetan_Baima_Guoyuan.SG | Gaoshancheng_10bp | Qiang_Gaoping.SG | Mbuti.SDG | -0.000636 | 0.000545 | -1.167 | 4553 | 4595 | 66804 |
| Tibetan_Amdo_Dalu.SG | Gaoshancheng_10bp | Naxi.SDG | Mbuti.SDG | -0.000636 | 0.000548 | -1.161 | 4555 | 4597 | 66802 |
| Tibetan_Amdo_Dalu.SG | China_Upper_YR_LN | Wa_Cangyuan.DG | Mbuti.SDG | -0.000345 | 0.000297 | -1.161 | 9855 | 9905 | 144911 |
| Tibetan_Baima_Wujiao.SG | Gaoshancheng_10bp | Tujia.SDG | Mbuti.SDG | -0.000631 | 0.000553 | -1.139 | 4559 | 4601 | 66801 |
| Tibetan_Baima_Guoyuan.SG | China_Upper_YR_LN | Tibetan_Zase | Mbuti.SDG | -0.000302 | 0.000272 | -1.111 | 9948 | 9992 | 144956 |
| Tibetan_Baima_Guoyuan.SG | China_Upper_YR_LN | Han_Fujian | Mbuti.SDG | -0.000295 | 0.000268 | -1.102 | 9968 | 10011 | 144956 |
| Tibetan_Baima_Wujiao.SG | Gaoshancheng_10bp | Han_Shanxi | Mbuti.SDG | -0.000577 | 0.000527 | -1.094 | 4557 | 4595 | 66804 |
| Tibetan_Baima_Wujiao.SG | China_Upper_YR_LN | Qiang_Xiaoyuan.SG | Mbuti.SDG | -0.00033 | 0.000304 | -1.085 | 9973 | 10021 | 144955 |
| Tibetan_Baima_Guoyuan.SG | Gaoshancheng_10bp | Laos_LN_BA.SG | Mbuti.SDG | -0.000983 | 0.000912 | -1.078 | 3283 | 3332 | 49381 |
| Tibetan_Baima_Guoyuan.SG | China_Upper_YR_LN | Qiang_Liujia.SG | Mbuti.SDG | -0.000322 | 3.00E-04 | -1.075 | 9965 | 10011 | 144955 |
| Tibetan_Baima_Guoyuan.SG | Gaoshancheng_10bp | Tibetan_Zase | Mbuti.SDG | -0.000539 | 0.000503 | -1.071 | 4553 | 4589 | 66804 |
| Tibetan_Amdo_Dalu.SG | China_Upper_YR_LN | Laos_LN_BA.SG | Mbuti.SDG | -0.000553 | 0.000525 | -1.054 | 7092 | 7150 | 105135 |

| | | | | | | | | | |
|---|---|---|---|---|---|---|---|---|---|
| Tibetan_Amdo_Dalu.SG | Gaoshancheng_10bp | Tu.SDG | Mbuti.SDG | -0.000534 | 0.000513 | -1.042 | 4539 | 4575 | 66802 |
| Tibetan_Baima_Wujiao.SG | China_Upper_YR_LN | Pumi_Lanping.DG | Mbuti.SDG | -0.000322 | 0.00031 | -1.041 | 9962 | 10009 | 144913 |
| Tibetan_Baima_Guoyuan.SG | Gaoshancheng_10bp | Uyghur.SDG | Mbuti.SDG | -0.000499 | 0.00048 | -1.04 | 4442 | 4475 | 66804 |
| Tibetan_Baima_Guoyuan.SG | Gaoshancheng_10bp | Han_Shanxi | Mbuti.SDG | -0.000506 | 0.000496 | -1.021 | 4559 | 4593 | 66804 |
| Tibetan_Baima_Wujiao.SG | China_Upper_YR_LN | Yiyang | Mbuti.SDG | -0.000867 | 0.000861 | -1.008 | 2497 | 2528 | 36252 |
| Tibetan_Baima_Wujiao.SG | Gaoshancheng_10bp | Cambodian.SDG | Mbuti.SDG | -0.000541 | 0.000543 | -0.995 | 4514 | 4550 | 66803 |
| Tibetan_Amdo_Dalu.SG | Gaoshancheng_10bp | Daur.SDG | Mbuti.SDG | -0.000527 | 0.000535 | -0.985 | 4545 | 4580 | 66804 |
| Tibetan_Baima_Guoyuan.SG | China_Upper_YR_LN | Hani_Honghe.DG | Mbuti.SDG | -0.000278 | 0.000282 | -0.985 | 9924 | 9964 | 144908 |
| Tibetan_Amdo_Dalu.SG | Gaoshancheng_10bp | Blang_Menghai.DG | Mbuti.SDG | -0.000512 | 0.000528 | -0.971 | 4526 | 4561 | 66782 |
| Tibetan_Baima_Wujiao.SG | Gaoshancheng_10bp | Han_Northwest | Mbuti.SDG | -0.00051 | 0.000529 | -0.966 | 4555 | 4589 | 66804 |
| Tibetan_Baima_Guoyuan.SG | Gaoshancheng_10bp | Han_Northwest | Mbuti.SDG | -0.00048 | 0.000497 | -0.965 | 4555 | 4587 | 66804 |
| Tibetan_Baima_Wujiao.SG | Gaoshancheng_10bp | Han_Shandong | Mbuti.SDG | -0.000508 | 0.000531 | -0.955 | 4560 | 4594 | 66804 |
| Tibetan_Baima_Wujiao.SG | China_Upper_YR_LN | Lahu_Lancang.DG | Mbuti.SDG | -0.000326 | 0.000341 | -0.954 | 9925 | 9972 | 144900 |
| Tibetan_Amdo_Dalu.SG | China_Upper_YR_LN | Naxi.SDG | Mbuti.SDG | -0.000276 | 0.000294 | -0.939 | 9960 | 10000 | 144946 |
| Tibetan_Baima_Wujiao.SG | Gaoshancheng_10bp | Han_Henan | Mbuti.SDG | -0.000489 | 0.000528 | -0.925 | 4559 | 4592 | 66804 |
| Tibetan_Amdo_Dalu.SG | China_Upper_YR_LN | Blang_Menghai.DG | Mbuti.SDG | -0.000265 | 0.000287 | -0.922 | 9881 | 9919 | 144909 |
| Tibetan_Baima_Guoyuan.SG | Gaoshancheng_10bp | Han_Shandong | Mbuti.SDG | -0.000455 | 0.000499 | -0.911 | 4561 | 4592 | 66804 |
| Tibetan_Baima_Guoyuan.SG | Gaoshancheng_10bp | Qiang_Xiaoyuan.SG | Mbuti.SDG | -0.000482 | 0.00053 | -0.91 | 4552 | 4584 | 66804 |
| Tibetan_Baima_Guoyuan.SG | China_Upper_YR_LN | Tujia.SDG | Mbuti.SDG | -0.000253 | 0.000278 | -0.908 | 9963 | 10000 | 144948 |
| Tibetan_Baima_Wujiao.SG | Gaoshancheng_10bp | Qiang_Matou.SG | Mbuti.SDG | -0.000496 | 0.000551 | -0.899 | 4556 | 4589 | 66804 |
| Tibetan_Baima_Wujiao.SG | Gaoshancheng_10bp | Qiang_Xiaoyuan.SG | Mbuti.SDG | -0.000497 | 0.000562 | -0.884 | 4554 | 4587 | 66804 |
| Tibetan_Baima_Guoyuan.SG | China_Upper_YR_LN | Lahu_Lancang.DG | Mbuti.SDG | -0.000281 | 0.00032 | -0.878 | 9920 | 9960 | 144900 |
| Tibetan_Baima_Guoyuan.SG | China_Upper_YR_LN | Qiang_Matou.SG | Mbuti.SDG | -0.000245 | 0.000282 | -0.871 | 9965 | 10001 | 144956 |
| Tibetan_Baima_Guoyuan.SG | Gaoshancheng_10bp | Han_Henan | Mbuti.SDG | -0.00043 | 0.000497 | -0.865 | 4560 | 4589 | 66804 |
| Tibetan_Baima_Wujiao.SG | Gaoshancheng_10bp | GaoHuaHua | Mbuti.SDG | -0.000663 | 0.000831 | -0.798 | 3983 | 4021 | 58065 |
| Tibetan_Baima_Wujiao.SG | Gaoshancheng_10bp | Dong_Guizhou | Mbuti.SDG | -0.000429 | 0.00054 | -0.796 | 4560 | 4588 | 66804 |
| Tibetan_Amdo_Dalu.SG | Gaoshancheng_10bp | Shenxian | Mbuti.SDG | -0.000859 | 0.001096 | -0.784 | 2643 | 2677 | 39069 |
| Tibetan_Baima_Wujiao.SG | China_Upper_YR_LN | Miao_Guizhou | Mbuti.SDG | -0.000227 | 0.000293 | -0.774 | 9969 | 10002 | 144955 |
| Tibetan_Baima_Guoyuan.SG | China_Upper_YR_LN | Qiang_Xiaoyuan.SG | Mbuti.SDG | -0.00021 | 0.000284 | -0.741 | 9971 | 10001 | 144955 |
| Tibetan_Baima_Wujiao.SG | Gaoshancheng_10bp | Uyghur.SDG | Mbuti.SDG | -0.000372 | 0.000512 | -0.726 | 4444 | 4469 | 66804 |
| Tibetan_Amdo_Dalu.SG | Gaoshancheng_10bp | Uyghur.SDG | Mbuti.SDG | -0.000356 | 0.000492 | -0.724 | 4446 | 4470 | 66804 |
| Tibetan_Baima_Wujiao.SG | Gaoshancheng_10bp | Laos_LN_BA.SG | Mbuti.SDG | -0.000671 | 0.000929 | -0.722 | 3294 | 3327 | 49381 |
| Tibetan_Baima_Wujiao.SG | Gaoshancheng_10bp | Han_Fujian | Mbuti.SDG | -0.00038 | 0.000528 | -0.719 | 4566 | 4591 | 66804 |
| Tibetan_Baima_Guoyuan.SG | China_Upper_YR_LN | She_Fujian | Mbuti.SDG | -0.000209 | 0.000293 | -0.712 | 9979 | 10009 | 144956 |
| Tibetan_Baima_Guoyuan.SG | China_Upper_YR_LN | Uyghur.SDG | Mbuti.SDG | -0.000188 | 0.000266 | -0.706 | 9715 | 9743 | 144951 |
| Tibetan_Amdo_Dalu.SG | Gaoshancheng_10bp | China_SEastAsia_Coastal_Historic | Mbuti.SDG | -0.001089 | 0.001546 | -0.704 | 1307 | 1328 | 19186 |
| Tibetan_Baima_Wujiao.SG | China_Upper_YR_LN | Uyghur.SDG | Mbuti.SDG | -0.000197 | 0.000289 | -0.681 | 9730 | 9758 | 144950 |
| Tibetan_Amdo_Dalu.SG | China_Upper_YR_LN | Layi | Mbuti.SDG | -0.000655 | 0.000995 | -0.659 | 1695 | 1711 | 24916 |
| Tibetan_Baima_Guoyuan.SG | Gaoshancheng_10bp | Qiang_Matou.SG | Mbuti.SDG | -0.000339 | 0.000518 | -0.655 | 4562 | 4585 | 66804 |
| Tibetan_Baima_Guoyuan.SG | Gaoshancheng_10bp | Han_Fujian | Mbuti.SDG | -0.000315 | 0.000497 | -0.634 | 4567 | 4588 | 66804 |
| Tibetan_Baima_Wujiao.SG | Gaoshancheng_10bp | Daur.SDG | Mbuti.SDG | -0.000348 | 0.00055 | -0.632 | 4552 | 4575 | 66804 |
| Tibetan_Baima_Wujiao.SG | China_Upper_YR_LN | KHV.SG | Mbuti.SDG | -0.000183 | 0.000291 | -0.629 | 9961 | 9987 | 144955 |
| Tibetan_Baima_Guoyuan.SG | Gaoshancheng_10bp | GaoHuaHua | Mbuti.SDG | -0.000494 | 0.000792 | -0.623 | 3983 | 4011 | 58065 |
| Tibetan_Baima_Wujiao.SG | Gaoshancheng_10bp | Pumi_Lanping.DG | Mbuti.SDG | -0.000349 | 0.000566 | -0.616 | 4555 | 4578 | 66783 |
| Tibetan_Baima_Wujiao.SG | Gaoshancheng_10bp | Miao_Guizhou | Mbuti.SDG | -0.000321 | 0.000532 | -0.603 | 4561 | 4583 | 66804 |
| Tibetan_Baima_Wujiao.SG | China_Upper_YR_LN | Tibetan_Zase | Mbuti.SDG | -0.000175 | 0.000293 | -0.598 | 9973 | 9998 | 144955 |
| Tibetan_Amdo_Dalu.SG | China_Upper_YR_LN | Vietnam_BA | Mbuti.SDG | -0.000436 | 0.00076 | -0.574 | 3013 | 3033 | 44340 |
| Tibetan_Baima_Guoyuan.SG | Gaoshancheng_10bp | Miao_Guizhou | Mbuti.SDG | -0.000283 | 0.000499 | -0.566 | 4563 | 4582 | 66804 |
| Tibetan_Baima_Guoyuan.SG | Gaoshancheng_10bp | Lahu_Lancang.DG | Mbuti.SDG | -0.000317 | 0.000561 | -0.565 | 4544 | 4565 | 66775 |
| Tibetan_Baima_Wujiao.SG | China_Upper_YR_LN | China_SEastAsia_Coastal_LN | Mbuti.SDG | -0.000287 | 0.000509 | -0.564 | 5870 | 5895 | 85960 |
| Tibetan_Baima_Guoyuan.SG | Gaoshancheng_10bp | Dong_Guizhou | Mbuti.SDG | -0.000284 | 0.000507 | -0.561 | 4565 | 4584 | 66804 |
| Tibetan_Baima_Wujiao.SG | Gaoshancheng_10bp | CDX.SG | Mbuti.SDG | -0.000279 | 0.000528 | -0.528 | 4561 | 4579 | 66804 |
| Tibetan_Baima_Wujiao.SG | China_Upper_YR_LN | Vietnam_BA | Mbuti.SDG | -0.000402 | 0.000765 | -0.525 | 3023 | 3041 | 44340 |
| Tibetan_Baima_Wujiao.SG | China_Upper_YR_LN | BaBanQinCen | Mbuti.SDG | -0.000259 | 0.000498 | -0.519 | 6470 | 6494 | 94245 |
| Tibetan_Baima_Wujiao.SG | Gaoshancheng_10bp | She_Fujian | Mbuti.SDG | -0.000274 | 0.000562 | -0.488 | 4575 | 4593 | 66804 |
| Tibetan_Baima_Guoyuan.SG | Gaoshancheng_10bp | Daur.SDG | Mbuti.SDG | -0.000249 | 0.000518 | -0.48 | 4557 | 4573 | 66804 |
| Tibetan_Baima_Wujiao.SG | Gaoshancheng_10bp | Tibetan_Zase | Mbuti.SDG | -0.000256 | 0.000535 | -0.479 | 4565 | 4582 | 66804 |
| Tibetan_Baima_Guoyuan.SG | Gaoshancheng_10bp | Blang_Menghai.DG | Mbuti.SDG | -0.000245 | 0.000514 | -0.476 | 4538 | 4554 | 66782 |
| Tibetan_Baima_Wujiao.SG | Gaoshancheng_10bp | Lahu_Lancang.DG | Mbuti.SDG | -0.00027 | 0.000594 | -0.456 | 4549 | 4567 | 66775 |
| Tibetan_Baima_Guoyuan.SG | China_Upper_YR_LN | Yiyang | Mbuti.SDG | -0.000348 | 0.000799 | -0.435 | 2504 | 2517 | 36253 |
| Tibetan_Baima_Wujiao.SG | China_Upper_YR_LN | Zongri4.7k | Mbuti.SDG | -0.000221 | 0.000513 | -0.43 | 7836 | 7861 | 114753 |
| Tibetan_Baima_Guoyuan.SG | Gaoshancheng_10bp | CDX.SG | Mbuti.SDG | -0.000207 | 0.000497 | -0.416 | 4563 | 4577 | 66804 |
| Tibetan_Baima_Guoyuan.SG | China_Upper_YR_LN | Dong_Guizhou | Mbuti.SDG | -0.000113 | 0.000273 | -0.415 | 9974 | 9990 | 144956 |
| Tibetan_Baima_Wujiao.SG | Gaoshancheng_10bp | KHV.SG | Mbuti.SDG | -0.000218 | 0.00053 | -0.412 | 4559 | 4573 | 66804 |
| Tibetan_Baima_Wujiao.SG | China_Upper_YR_LN | Wa_Cangyuan.DG | Mbuti.SDG | -0.000124 | 0.000304 | -0.408 | 9893 | 9911 | 144911 |
| Tibetan_Amdo_Dalu.SG | China_Upper_YR_LN | Vietnam_N | Mbuti.SDG | -0.000321 | 0.00079 | -0.407 | 2256 | 2266 | 33552 |
| Tibetan_Baima_Guoyuan.SG | Gaoshancheng_10bp | Yiyang | Mbuti.SDG | -0.000575 | 0.001498 | -0.384 | 1225 | 1235 | 17895 |
| Tibetan_Baima_Guoyuan.SG | China_Upper_YR_LN | Laos_LN_BA.SG | Mbuti.SDG | -0.000194 | 0.000515 | -0.376 | 7121 | 7141 | 105136 |
| Tibetan_Baima_Guoyuan.SG | China_Upper_YR_LN | BaBanQinCen | Mbuti.SDG | -0.000171 | 0.000458 | -0.373 | 6471 | 6488 | 94246 |
| Tibetan_Baima_Guoyuan.SG | Gaoshancheng_10bp | KHV.SG | Mbuti.SDG | -0.000175 | 0.000498 | -0.352 | 4561 | 4573 | 66804 |
| Tibetan_Baima_Guoyuan.SG | China_Upper_YR_LN | Shenxian | Mbuti.SDG | -0.00019 | 0.000564 | -0.337 | 5460 | 5475 | 80395 |
| Tibetan_Baima_Wujiao.SG | China_Upper_YR_LN | Laos_LN_BA.SG | Mbuti.SDG | -0.000178 | 0.000541 | -0.329 | 7128 | 7146 | 105135 |
| Tibetan_Baima_Wujiao.SG | Gaoshancheng_10bp | Blang_Menghai.DG | Mbuti.SDG | -0.000171 | 0.000551 | -0.311 | 4537 | 4549 | 66782 |
| Tibetan_Baima_Wujiao.SG | China_Upper_YR_LN | China_Lahu.SDG | Mbuti.SDG | -8.70E-05 | 0.000305 | -0.285 | 9944 | 9957 | 144954 |
| Tibetan_Baima_Wujiao.SG | China_Upper_YR_LN | GaoHuaHua | Mbuti.SDG | -0.000123 | 0.000432 | -0.284 | 8574 | 8590 | 124512 |
| Tibetan_Baima_Guoyuan.SG | China_Upper_YR_LN | China_Lahu.SDG | Mbuti.SDG | -8.20E-05 | 0.000292 | -0.282 | 9932 | 9944 | 144955 |
| Tibetan_Baima_Wujiao.SG | China_Upper_YR_LN | Blang_Menghai.DG | Mbuti.SDG | -8.20E-05 | 0.000303 | -0.272 | 9916 | 9928 | 144909 |
| Tibetan_Amdo_Dalu.SG | China_Upper_YR_LN | Uyghur.SDG | Mbuti.SDG | -7.30E-05 | 0.000272 | -0.268 | 9718 | 9728 | 144950 |
| Tibetan_Amdo_Dalu.SG | China_Upper_YR_LN | China_SEastAsia_Coastal_Historic | Mbuti.SDG | -0.000207 | 0.000817 | -0.253 | 2773 | 2781 | 40198 |
| Tibetan_Baima_Wujiao.SG | China_Upper_YR_LN | CDX.SG | Mbuti.SDG | -6.50E-05 | 0.00029 | -0.224 | 9971 | 9980 | 144955 |
| Tibetan_Baima_Wujiao.SG | China_Upper_YR_LN | LaCen | Mbuti.SDG | -0.000117 | 0.000527 | -0.221 | 7207 | 7219 | 104781 |
| Tibetan_Amdo_Dalu.SG | Gaoshancheng_10bp | LaCen | Mbuti.SDG | -0.000189 | 0.000943 | -0.2 | 3381 | 3390 | 49738 |
| Tibetan_Baima_Guoyuan.SG | Gaoshancheng_10bp | She_Fujian | Mbuti.SDG | -0.000103 | 0.000526 | -0.195 | 4580 | 4587 | 66804 |
| Tibetan_Baima_Wujiao.SG | Gaoshancheng_10bp | Yiyang | Mbuti.SDG | -0.000303 | 0.001558 | -0.194 | 1219 | 1225 | 17895 |
| Tibetan_Baima_Wujiao.SG | Gaoshancheng_10bp | China_SEastAsia_Coastal_Historic | Mbuti.SDG | -0.000276 | 0.001597 | -0.173 | 1313 | 1319 | 19186 |
| Tibetan_Amdo_Dalu.SG | China_Upper_YR_LN | Laos_Hoabinhian.SG | Mbuti.SDG | -9.00E-05 | 0.00063 | -0.143 | 3677 | 3683 | 56679 |
| Tibetan_Amdo_Dalu.SG | Gaoshancheng_10bp | Pumi_Lanping.DG | Mbuti.SDG | -7.70E-05 | 0.000541 | -0.143 | 4564 | 4569 | 66783 |
| Tibetan_Baima_Guoyuan.SG | China_Upper_YR_LN | Zongri4.7k | Mbuti.SDG | -6.60E-05 | 0.000502 | -0.132 | 7829 | 7837 | 114754 |
| Tibetan_Baima_Guoyuan.SG | China_Upper_YR_LN | Miao_Guizhou | Mbuti.SDG | -3.40E-05 | 0.000271 | -0.125 | 9972 | 9977 | 144956 |
| Tibetan_Baima_Guoyuan.SG | China_Upper_YR_LN | Blang_Menghai.DG | Mbuti.SDG | -2.60E-05 | 0.000279 | -0.093 | 9909 | 9913 | 144909 |
| Tibetan_Baima_Wujiao.SG | China_Upper_YR_LN | Shenxian | Mbuti.SDG | -4.50E-05 | 0.000594 | -0.075 | 5492 | 5495 | 80394 |
| Tibetan_Baima_Guoyuan.SG | China_Upper_YR_LN | KHV.SG | Mbuti.SDG | 6.00E-06 | 0.000274 | 0.023 | 9963 | 9962 | 144956 |
| Tibetan_Baima_Wujiao.SG | Gaoshancheng_10bp | Dai.SDG | Mbuti.SDG | 2.00E-05 | 0.000544 | 0.037 | 4560 | 4558 | 66801 |
| Tibetan_Baima_Wujiao.SG | Gaoshancheng_10bp | LaCen | Mbuti.SDG | 0.000102 | 0.000951 | 0.107 | 3393 | 3388 | 49738 |
| Tibetan_Baima_Guoyuan.SG | Gaoshancheng_10bp | Shenxian | Mbuti.SDG | 0.000138 | 0.001092 | 0.127 | 2663 | 2658 | 39069 |
| Tibetan_Baima_Wujiao.SG | China_Upper_YR_LN | Dai.SDG | Mbuti.SDG | 4.00E-05 | 0.000308 | 0.129 | 9954 | 9949 | 144944 |
| Tibetan_Baima_Guoyuan.SG | China_Upper_YR_LN | Wa_Cangyuan.DG | Mbuti.SDG | 3.90E-05 | 0.000287 | 0.135 | 9894 | 9888 | 144911 |

| | | | | | | | | | |
|---|---|---|---|---|---|---|---|---|---|
| Tibetan_Baima_Guoyuan.SG | Gaoshancheng_10bp | Dai.SDG | Mbuti.SDG | 8.80E-05 | 0.000516 | 0.171 | 4563 | 4557 | 66801 |
| Tibetan_Baima_Wujiao.SG | China_Upper_YR_LN | Layi | Mbuti.SDG | 0.000207 | 0.001006 | 0.206 | 1713 | 1708 | 24916 |
| Tibetan_Baima_Guoyuan.SG | China_Upper_YR_LN | GaoHuaHua | Mbuti.SDG | 9.70E-05 | 0.000399 | 0.243 | 8580 | 8568 | 124513 |
| Tibetan_Baima_Wujiao.SG | Gaoshancheng_10bp | Shenxian | Mbuti.SDG | 0.00031 | 0.001107 | 0.28 | 2668 | 2655 | 39069 |
| Tibetan_Amdo_Dalu.SG | China_Upper_YR_LN | Zongri5.1k | Mbuti.SDG | 0.000139 | 0.000493 | 0.282 | 7351 | 7336 | 107162 |
| Tibetan_Baima_Guoyuan.SG | Gaoshancheng_10bp | China_SEastAsia_Coastal_Historic | Mbuti.SDG | 0.000447 | 0.001567 | 0.285 | 1317 | 1308 | 19186 |
| Tibetan_Baima_Wujiao.SG | China_Upper_YR_LN | Russia_Afanasievo | Mbuti.SDG | 0.000128 | 0.000317 | 0.403 | 9358 | 9340 | 141836 |
| Tibetan_Baima_Guoyuan.SG | China_Upper_YR_LN | Vietnam_BA | Mbuti.SDG | 0.000328 | 0.000741 | 0.442 | 3041 | 3027 | 44340 |
| Tibetan_Baima_Wujiao.SG | China_Upper_YR_LN | China_SEastAsia_Coastal_Historic | Mbuti.SDG | 0.000401 | 0.000819 | 0.49 | 2794 | 2778 | 40198 |
| Tibetan_Amdo_Dalu.SG | China_Upper_YR_LN | Pumi_Lanping.DG | Mbuti.SDG | 0.00015 | 0.000296 | 0.507 | 9981 | 9959 | 144913 |
| Tibetan_Baima_Guoyuan.SG | China_Upper_YR_LN | Russia_Afanasievo | Mbuti.SDG | 0.000157 | 0.000302 | 0.521 | 9343 | 9321 | 141837 |
| Tibetan_Amdo_Dalu.SG | China_Upper_YR_LN | Zongri4.7k | Mbuti.SDG | 0.000305 | 0.000521 | 0.586 | 7862 | 7827 | 114753 |
| Tibetan_Baima_Guoyuan.SG | China_Upper_YR_LN | China_SEastAsia_Coastal_LN | Mbuti.SDG | 0.000287 | 0.000478 | 0.602 | 5893 | 5868 | 85961 |
| Tibetan_Baima_Guoyuan.SG | China_Upper_YR_LN | CDX.SG | Mbuti.SDG | 0.000168 | 0.000275 | 0.612 | 9976 | 9952 | 144956 |
| Tibetan_Amdo_Dalu.SG | Gaoshancheng_10bp | Layi | Mbuti.SDG | 0.001355 | 0.001921 | 0.705 | 842 | 826 | 12291 |
| Tibetan_Baima_Guoyuan.SG | Gaoshancheng_10bp | LaCen | Mbuti.SDG | 0.00066 | 0.000934 | 0.707 | 3414 | 3381 | 49738 |
| Tibetan_Baima_Wujiao.SG | China_Upper_YR_LN | Vietnam_N | Mbuti.SDG | 0.000589 | 0.000828 | 0.712 | 2274 | 2254 | 33552 |
| Tibetan_Baima_Guoyuan.SG | China_Upper_YR_LN | LaCen | Mbuti.SDG | 0.000358 | 0.000497 | 0.721 | 7212 | 7174 | 104782 |
| Tibetan_Amdo_Dalu.SG | Gaoshancheng_10bp | Tibetan_Zase | Mbuti.SDG | 0.000375 | 0.000511 | 0.733 | 4588 | 4563 | 66804 |
| Tibetan_Baima_Wujiao.SG | China_Upper_YR_LN | Cambodian.SDG | Mbuti.SDG | 0.000228 | 0.000306 | 0.744 | 9914 | 9881 | 144946 |
| Tibetan_Baima_Guoyuan.SG | China_Upper_YR_LN | Dai.SDG | Mbuti.SDG | 0.000222 | 0.000288 | 0.772 | 9955 | 9923 | 144945 |
| Tibetan_Baima_Guoyuan.SG | China_Upper_YR_LN | Cambodian.SDG | Mbuti.SDG | 0.000234 | 0.000291 | 0.803 | 9901 | 9868 | 144947 |
| Tibetan_Baima_Wujiao.SG | Gaoshancheng_10bp | Layi | Mbuti.SDG | 0.001929 | 0.001863 | 1.036 | 841 | 817 | 12291 |
| Tibetan_Baima_Guoyuan.SG | China_Upper_YR_LN | Layi | Mbuti.SDG | 0.001062 | 0.000984 | 1.079 | 1716 | 1690 | 24916 |
| Tibetan_Baima_Guoyuan.SG | China_Upper_YR_LN | China_SEastAsia_Coastal_Historic | Mbuti.SDG | 0.000921 | 0.000771 | 1.195 | 2789 | 2752 | 40199 |
| Tibetan_Baima_Guoyuan.SG | Gaoshancheng_10bp | Layi | Mbuti.SDG | 0.002406 | 0.001864 | 1.291 | 849 | 820 | 12291 |
| Tibetan_Baima_Guoyuan.SG | China_Upper_YR_LN | Vietnam_N | Mbuti.SDG | 0.001023 | 0.000788 | 1.298 | 2278 | 2243 | 33552 |
| Tibetan_Amdo_Dalu.SG | China_Upper_YR_LN | Russia_Afanasievo | Mbuti.SDG | 0.000451 | 0.000306 | 1.477 | 9359 | 9295 | 141836 |
| Tibetan_Amdo_Dalu.SG | China_Upper_YR_LN | Tibetan_Zase | Mbuti.SDG | 0.000663 | 0.000282 | 2.351 | 10020 | 9924 | 144955 |

**Table S8. The two-way admixture models in the context of the postdate late Neolithic period for TYC populations via qpAdm**

| Target | Source1 | Source2 | prop1 | prop2 | sd | P_value | Group |
|---|---|---|---|---|---|---|---|
| Naxi | Gaoshancheng | Taiwan_Hanben_IA | 0.887 | 0.113 | 0.035 | 0.08362982 | TCL |
| Tibetan_Baima | Gaoshancheng | Taiwan_Hanben_IA | 0.904 | 0.096 | 0.031 | 0.30535313 | TCL |
| Yi_HGDP | Gaoshancheng | Taiwan_Hanben_IA | 0.884 | 0.116 | 0.035 | 0.14138128 | TCL |
| Bai_Heqing | China_YR_MN | Taiwan_Hanben_IA | 0.916 | 0.084 | 0.032 | 0.24340835 | TEL |
| Qiang_Sichuan | China_YR_MN | Taiwan_Hanben_IA | 0.8 | 0.2 | 0.072 | 0.28185965 | TEL |
| Hani_Honghe | China_Upper_YR_LN | Layi | 0.634 | 0.366 | 0.063 | 0.0509541 | TSL |
| Lahu_Lancang | China_Upper_YR_LN | Layi | 0.453 | 0.547 | 0.08 | 0.08818467 | TSL |
| Blang_Menghai | Zongri5.1k | Layi | 0.426 | 0.574 | 0.061 | 0.22599353 | TSL |
| Wa_Cangyuan | Zongri5.1k | Layi | 0.432 | 0.568 | 0.068 | 0.06018349 | TSL |
| Tibetan_Amdo | China_Upper_YR_LN | Laos_Hoabinhian | 0.965 | 0.035 | 0.018 | 0.17419056 | TCL |
| Pumi_Lanping | China_YR_MN | Laos_Hoabinhian | 0.958 | 0.042 | 0.017 | 0.17625557 | TCL |
| Tibetans | Zongri5.1k | Layi | 0.768 | 0.232 | 0.06 | 0.49515409 | TCL |

**Table S9. Simulation of admixture history for TYC populations**

| Methods | test pop | generaion | propotion1 | ref A | propotion2 | ref B | Deviation |
|---|---|---|---|---|---|---|---|
| fastGLOBETROTTER | Bai_Heqing | 58.4709814 | 0.36 | Cambodian | 0.64 | Han_Xian | 1.634413 |
| fastGLOBETROTTER | Blang_Menghai | 78.66015625 | 0.34 | Han_Xian | 0.66 | Cambodian | 10.766994 |
| fastGLOBETROTTER | Wa_Cangyuan | 63.18278664 | 0.37 | Han_Xian | 0.63 | Cambodian | 10.268951 |
| fastGLOBETROTTER | Pumi_Lanping | 7.727227767 | 0.46 | Tibetan_Zase | 0.54 | Han_Xian | 108.168765 |
| fastGLOBETROTTER | Qiang_Gaoping | 27.51156004 | 0.18 | Cambodian | 0.82 | Han_Xian | 2.891062 |
| fastGLOBETROTTER | Qiang_Liujia | 32.42544306 | 0.12 | Li_Linhshui | 0.88 | Han_Xian | 7.544806 |
| fastGLOBETROTTER | Qiang_Matou | 30.20049971 | 0.17 | Cambodian | 0.83 | Han_Xian | 1.021977 |
| fastGLOBETROTTER | Qiang_Xiaoyuan | 24.78077579 | 0.23 | Cambodian | 0.77 | Han_Xian | 6.241314 |
| fastGLOBETROTTER | Tibetan_Amdo_Dalu | 22.63961123 | 0.2 | Han_Xian | 0.8 | Tibetan_Zase | 10.955629 |
| fastGLOBETROTTER | Tibetan_Baima_Guoyuan | 15.27929929 | 0.48 | Han_Xian | 0.52 | Tibetan_Zase | 2.120249 |
| fastGLOBETROTTER | Tibetan_Baima_Wujiao | 13.7537713 | 0.2 | Tibetan_Zase | 0.8 | Han_Xian | 1.94197 |
| fastGLOBETROTTER | Lahu_Lancang | 39.9185588 | 0.49 | Han_Xian | 0.51 | Cambodian | 15.146623 |
| fastGLOBETROTTER | Hani_Honghe | 48.46731913 | 0.4 | Cambodian | 0.6 | Han_Xian | 8.086787 |
| MALDER | Blang_Yunnan | 30.73 | | Han_Gansu | | Han_Sichuan | 6.65 |
| MALDER | Bai_Yunnan | 33.56 | | Tibetan_Tibet | | Miao_Guizhou | 9.38 |
| MALDER | Blang_Yunnan | 63.71 | | Han_Sichuan | | She_Fujian | 21.54 |
| MALDER | Blang_Yunnan | 93.52 | | Han_Fujian | | She_Fujian | 34.49 |
| MALDER | Hani_Yunnan | 48.86 | | Han_Fujian | | Han_Sichuan | 24.7 |
| MALDER | Lahu_Yunnan | 18.37 | | Tibetan_Tibet | | Miao_Sichuan | 6.84 |
| MALDER | Lahu_Yunnan | 12.74 | | Han_Fujian | | Miao_Sichuan | 6.43 |
| MALDER | Lahu_Yunnan | 93.26 | | Han_Shaanxi | | Tujia_Chongqing | 29 |
| MALDER | Pumi_Yunnan | 44.12 | | Tibetan_Tibet | | Miao_Chongqing | 14.23 |
| MALDER | Qiang_Sichuan | 72.51 | | Tibetan_Tibet | | She_Fujian | 10.66 |
| MALDER | Qiang_Sichuan | 94.92 | | Han_Fujian | | Tibetan_Tibet | 13.66 |
| MALDER | Tibetan_Amdo_Sichuan | 88.44 | | Tibetan_Tibet | | She_Fujian | 42.59 |
| MALDER | Tibetan_Baima_Sichuan | 11.87 | | Tibetan_Tibet | | She_Fujian | 1.89 |
| MALDER | Tibetan_Baima_Sichuan | 93.44 | | Han_Shaanxi | | Gaoshan_Fujian | 41.91 |
| MALDER | Wa_Yunnan | 98.4 | | Han_Shaanxi | | Han_Sichuan | 28.4 |
| MALDER | Wa_Yunnan | 35.96 | | Han_Guizhou | | Miao_Chongqing | 13.54 |

**Table S10. The top 0.01% signals selected by PBS (THL/TCL/TSL-CHB-EUR trio)**

| THL | PBS | ANNOT | TCL | PBS | ANNOT | TSL | PBS | ANNOT |
|---|---|---|---|---|---|---|---|---|
| 1 156066766 | 0.377155233 | RAB25 | 2 46625378 | 1.436669371 | CRIPT | 8 72841433 | 0.592443369 | KCNB2 |
| 5 36292004 | 0.345432147 | RANBP3L | 13 112078820 | 1.150006502 | LINC00403 | 6 97469215 | 0.513251702 | LOC101927314 |
| 2 166905163 | 0.306943742 | XIRP2 | 19 42106011 | 1.085679583 | POU2F2 | 6 32637937 | 0.495135289 | HLA-DQA1 |
| 14 104801242 | 0.304641319 | ZBTB42 | 6 133385403 | 1.085580488 | EYA4 | 12_120877827 | 0.491399115 | SPPL3 |
| 14 104792556 | 0.30261411 | AKT1 | 20 32319506 | 1.002236317 | KIF3B | 6 32638351 | 0.485423399 | HLA-DQA1 |
| 5 36297447 | 0.298933057 | RANBP3L | 9 123473990 | 0.881298529 | DENND1A | 6 32643585 | 0.480794928 | HLA-DQA1 |
| 2 12475857 | 0.297780461 | LOC100506457 | 16 57078468 | 0.828587156 | NLRC5 | 20 63420621 | 0.479054125 | KCNQ2 |
| 19 37760016 | 0.296294187 | ZNF573 | 2 32146857 | 0.791344539 | SPAST | 6 32640124 | 0.470134156 | HLA-DQA1 |
| 2 71203963 | 0.288705271 | PAIP2B | 6 137147193 | 0.79075802 | IL22RA2 | 6 32640150 | 0.470134156 | HLA-DQA1 |
| 14 104803442 | 0.287208749 | ZBTB42 | 1 218714548 | 0.737507423 | MIR548F3 | 3 173797696 | 0.462343264 | NLGN1 |
| 1 10746927 | 0.286142104 | CASZ1 | 18 31653439 | 0.732287892 | B4GALT6 | 10 22575929 | 0.454329501 | PIP4K2A |
| 5 36292110 | 0.279041594 | RANBP3L | 2 72565350 | 0.731601346 | EXOC6B | 6 32640129 | 0.447117069 | HLA-DQA1 |
| 12_33436453 | 0.278711904 | SYT10 | 10_12421519 | 0.701950751 | CAMK1D | 6_32638220 | 0.443621007 | HLA-DQA1 |
| 2 170384567 | 0.278576046 | MYO3B | 12 3209966 | 0.701945005 | TSPAN9 | 6 32638225 | 0.443621007 | HLA-DQA1 |
| 16 82887576 | 0.278250745 | CDH13 | 1 41722407 | 0.694925984 | HIVEP3 | 6 32641675 | 0.437866617 | HLA-DQA1 |
| 2_170384519 | 0.2773314 | MYO3B | 13_112078858 | 0.689319022 | LINC00403 | 12_120890553 | 0.437256316 | SPPL3 |
| 2 166912609 | 0.276673948 | XIRP2 | 4 75969032 | 0.678991844 | SDAD1 | 12 120780753 | 0.429168579 | SPPL3 |
| 22 24841137 | 0.27458308 | SGSM1 | 1 36096950 | 0.67708772 | COL8A2 | 2 33158150 | 0.427446746 | LTBP1 |
| 12 32894088 | 0.274235596 | PKP2 | 1 204467820 | 0.667912818 | PIK3C2B | 7 99434063 | 0.426821435 | ATP5J2-PTCD1 |
| 4 89749947 | 0.272173891 | SNCA | 1 36096355 | 0.632543119 | COL8A2 | 7 99434076 | 0.426821435 | ATP5J2-PTCD1 |
| 22 24841141 | 0.271527221 | SGSM1 | 15 27351795 | 0.631719131 | GABRG3 | 6 32638303 | 0.425859878 | HLA-DQA1 |
| 2 12484888 | 0.267433325 | LOC100506457 | 2 24132469 | 0.622224494 | FAM228B | 6 32640276 | 0.425859878 | HLA-DQA1 |
| 2_166908221 | 0.265826887 | XIRP2 | 2_24132471 | 0.622224494 | FAM228B | 7_99434062 | 0.425049826 | ATP5J2-PTCD1 |
| 15 68800700 | 0.262980026 | ANP32A | 2 24132477 | 0.622224494 | FAM228B | 7 99434060 | 0.424441506 | ATP5J2-PTCD1 |
| 15 90046283 | 0.262708231 | ZNF710 | 2 24132468 | 0.622049356 | FAM228B | 6 32638127 | 0.422943217 | HLA-DQA1 |
| 16_82928843 | 0.261646477 | CDH13 | 2_24132467 | 0.621987822 | FAM228B | 6_32639573 | 0.421365222 | HLA-DQA1 |
| 1 169522862 | 0.261022736 | F5 | 14 63688319 | 0.621163411 | SGPP1 | 6 32641669 | 0.416468462 | HLA-DQA1 |
| 7 78031215 | 0.260657795 | MAGI2 | 14 63688320 | 0.621163411 | SGPP1 | 6 32641804 | 0.414816846 | HLA-DQA1 |
| 12_32894274 | 0.259800262 | PKP2 | 1_36096477 | 0.616794013 | COL8A2 | 7_99434035 | 0.414411523 | ATP5J2-PTCD1 |
| 10 76923424 | 0.258783935 | KCNMA1 | 3 181984950 | 0.604029932 | LINC01206 | 4 188094107 | 0.413796137 | TRIML2 |
| 12 54958534 | 0.257994319 | TESPA1 | 1 35495710 | 0.598943621 | KIAA0319L | 6 32640236 | 0.413604821 | HLA-DQA1 |
| 2_166909295 | 0.257335447 | XIRP2 | 1_68118876 | 0.595333427 | GNG12-AS1 | 6_32641021 | 0.413604821 | HLA-DQA1 |
| 2 170384153 | 0.255661463 | MYO3B | 3 181984955 | 0.590187026 | LINC01206 | 6 32637589 | 0.41109281 | HLA-DQA1 |
| 2 170384356 | 0.255661463 | MYO3B | 11 1740307 | 0.589033534 | IFITM10 | 6 32639487 | 0.41109281 | HLA-DQA1 |
| 3 129424674 | 0.255416649 | EFCAB12 | 9 33937601 | 0.587848253 | UBAP2 | 6 32642855 | 0.41109281 | HLA-DQA1 |
| 2 71191767 | 0.255011537 | PAIP2B | 14 92480325 | 0.587757521 | SLC24A4 | 7 99434068 | 0.410190723 | ATP5J2-PTCD1 |
| 6 170254987 | 0.254832698 | LOC154449 | 14 92480318 | 0.586291573 | SLC24A4 | 6 32637673 | 0.408615486 | HLA-DQA1 |
| 13 70135694 | 0.254112429 | ATXN8OS | 17 64596586 | 0.585396679 | SMURF2 | 6 32637681 | 0.408615486 | HLA-DQA1 |
| 15 47318266 | 0.253281854 | SEMA6D | 14 92480317 | 0.581046157 | SLC24A4 | 10 93442041 | 0.40846563 | MYOF |
| 12 54950899 | 0.253249277 | TESPA1 | 4 30887034 | 0.579401677 | PCDH7 | 6 32640035 | 0.406171578 | HLA-DQA1 |
| 15 40300781 | 0.251754938 | PLCB2 | 1 36044395 | 0.563219423 | AGO3 | 12 103644067 | 0.405618453 | STAB2 |
| 1_169512877 | 0.251416241 | F5 | 9_93614991 | 0.561835934 | PHF2 | 6_32640771 | 0.404065614 | HLA-DQA1 |
| 7 78031047 | 0.251228458 | MAGI2 | 14 78702454 | 0.537230679 | NRXN3 | 6 32637827 | 0.403759934 | HLA-DQA1 |
| 16 82863038 | 0.251189623 | CDH13 | 1 35968855 | 0.534876742 | AGO3 | 6 32640376 | 0.402005776 | HLA-DQA1 |
| 2 46362184 | 0.250971068 | EPAS1 | 17 46763843 | 0.531534396 | WNT3 | 2 231313819 | 0.401541008 | ARMC9 |
| 13 107287696 | 0.250274589 | FAM155A | 3 160071920 | 0.531523736 | IL12A-AS1 | 2 231318878 | 0.398118407 | ARMC9 |
| 1 6134520 | 0.249672323 | CHD5 | 6 150903213 | 0.531074899 | MTHFD1L | 11 120136798 | 0.396063359 | TRIM29 |
| 13 70134715 | 0.248639796 | ATXN8OS | 1 36023094 | 0.529216598 | AGO3 | 6 32640339 | 0.395888141 | HLA-DQA1 |
| 13 70136183 | 0.248639796 | ATXN8OS | 3 188288803 | 0.529148575 | LPP | 6 32637813 | 0.389870496 | HLA-DQA1 |
| 12 54950240 | 0.248569835 | TESPA1 | 14 92474866 | 0.528563493 | SLC24A4 | 6 32638168 | 0.389870496 | HLA-DQA1 |
| 12 54954593 | 0.248569835 | TESPA1 | 3 39945492 | 0.527199341 | MYRIP | 6 32638320 | 0.389870496 | HLA-DQA1 |
| 3 18354833 | 0.247756581 | SATB1 | 1 36153474 | 0.519330951 | TRAPPC3 | 6 32640029 | 0.389870496 | HLA-DQA1 |
| 3 108386240 | 0.246475644 | MYH15 | 1 36038107 | 0.514297392 | AGO3 | 7 104420559 | 0.389856571 | LHFPL3 |
| 2 166924958 | 0.246233828 | XIRP2 | 15 48575910 | 0.509296733 | FBN1 | 13 92823978 | 0.386571945 | GPC5 |
| 14 104782985 | 0.245900229 | AKT1 | 3 32121752 | 0.507659166 | GPD1L | 7 99434061 | 0.386019781 | ATP5J2-PTCD1 |
| 6 170260458 | 0.245738857 | LOC154449 | 15 59512472 | 0.503157689 | FAM81A | 20 63431530 | 0.378845552 | KCNQ2 |
| 2 170384201 | 0.245143301 | MYO3B | 13 60557821 | 0.500673515 | TDRD3 | 19 13362078 | 0.376133044 | CACNA1A |
| 1 65397214 | 0.24513609 | DNAJC6 | 12 53530567 | 0.49660151 | ATF7 | 6 32637845 | 0.375586171 | HLA-DQA1 |
| 12 54971436 | 0.245089369 | TESPA1 | 22 46702218 | 0.483345874 | CERK | 15 84604744 | 0.374548569 | ZSCAN2 |
| 13 28300527 | 0.244408682 | FLT1 | 6 100436743 | 0.482215684 | SIM1 | 1 169580391 | 0.372590262 | F5 |
| 13 70107466 | 0.243928008 | ATXN8OS | 1 245574867 | 0.478782473 | KIF26B | 19 5849655 | 0.372089386 | FUT3 |
| 2 201599357 | 0.243836565 | ALS2CR11 | 1 245574870 | 0.478782473 | KIF26B | 6 32638437 | 0.369848816 | HLA-DQA1 |
| 22 32595373 | 0.243791419 | SYN3 | 17 75844191 | 0.477908925 | UNC13D | 5 150785576 | 0.367251083 | SMIM3 |
| 15 68800683 | 0.243667625 | ANP32A | 6 47715998 | 0.475084133 | GPR115 | 6 32638393 | 0.366924461 | HLA-DQA1 |
| 1 6133837 | 0.243251685 | CHD5 | 6 125172311 | 0.472986009 | TPD52L1 | 6 32637775 | 0.366028749 | HLA-DQA1 |
| 20 19215452 | 0.242923931 | SLC24A3 | 1 35505107 | 0.471310051 | KIAA0319L | 6 32640439 | 0.366028749 | HLA-DQA1 |
| 2 166971422 | 0.242657089 | XIRP2 | 12_82821889 | 0.470696487 | TMTC2 | 6_32640440 | 0.366028749 | HLA-DQA1 |
| 7 78027720 | 0.242441359 | MAGI2 | 1 36098196 | 0.47031962 | COL8A2 | 6 30931456 | 0.3654829 | SFTA2 |
| 2 170407669 | 0.24164522 | MYO3B | 9 4802087 | 0.463201148 | RCL1 | 6 32638392 | 0.365083252 | HLA-DQA1 |
| 1_65395191 | 0.241400959 | DNAJC6 | 22_20785068 | 0.458650467 | PI4KA | 4_88069234 | 0.363719667 | PKD2 |
| 12 54975118 | 0.240912627 | TESPA1 | 19 13336598 | 0.456812632 | CACNA1A | 7 71140301 | 0.361985135 | WBSCR17 |
| 6 10412555 | 0.240843919 | TFAP2A | 1 36039292 | 0.452917551 | AGO3 | 18 58872704 | 0.361656969 | ZNF532 |
| 19 45951638 | 0.24083353 | NOVA2 | 6 37377063 | 0.449527204 | RNF8 | 12 40422923 | 0.361074665 | MUC19 |
| 22 32598780 | 0.240538061 | SYN3 | 1 36029761 | 0.44842784 | AGO3 | 5 94988600 | 0.360165976 | MCTP1 |
| 2 46340777 | 0.240407221 | EPAS1 | 2 17640253 | 0.447536489 | VSNL1 | 5 94988602 | 0.360165976 | MCTP1 |
| 3 121580466 | 0.240152248 | ARGFX | 3 50621598 | 0.44707371 | MAPKAPK3 | 7 72316984 | 0.359903731 | CALN1 |
| 15_90053615 | 0.239785475 | ZNF710 | 1_36029458 | 0.446655313 | AGO3 | 17_9598693 | 0.358916162 | WDR16 |
| 12 54950349 | 0.239190762 | TESPA1 | 5 41200897 | 0.446633454 | C6 | 6 29943234 | 0.358440094 | HLA-A |
| 12 54955661 | 0.239190762 | TESPA1 | 3 195965178 | 0.44654698 | SDHAP1 | 1 242267096 | 0.35779796 | PLD5 |
| 12_54957297 | 0.239190762 | TESPA1 | 1_204467828 | 0.446463724 | PIK3C2B | 6_29942500 | 0.352995187 | HLA-A |
| 12 54958821 | 0.239190762 | TESPA1 | 1 35471261 | 0.445216189 | KIAA0319L | 21 32513553 | 0.351853659 | EVA1C |
| 4 89750239 | 0.239007568 | SNCA | 12 109733231 | 0.442936105 | FAM222A | 6 32641627 | 0.350525736 | HLA-DQA1 |
| 2_166918433 | 0.23894248 | XIRP2 | 9_113949392 | 0.44290006 | ZNF618 | 15_30673854 | 0.350204898 | LOC100288637 |
| 2 170384046 | 0.238752508 | MYO3B | 9 113949461 | 0.44290006 | ZNF618 | 1 54727736 | 0.34983171 | MROH7-TTC4 |
| 1 65400984 | 0.238428747 | DNAJC6 | 1 36153223 | 0.441331135 | TRAPPC3 | 6 32640655 | 0.348255445 | HLA-DQA1 |
| 2_170386740 | 0.238357971 | MYO3B | 16_89160390 | 0.440749333 | LINC00304 | 7_71139320 | 0.34613719 | WBSCR17 |
| 1 231421509 | 0.238257392 | EGLN1 | 1 36045899 | 0.439592141 | AGO3 | 6 32638581 | 0.345638573 | HLA-DQA1 |
| 1 62127337 | 0.238220417 | INADL | 1 35188698 | 0.437205668 | SFPQ | 6 30893448 | 0.34444531 | DDR1 |
| 7_90247013 | 0.237887909 | C7orf63 | 8_133070016 | 0.437151919 | SLA | 12_120892982 | 0.343512878 | SPPL3 |
| 15 90047165 | 0.237581523 | ZNF710 | 8 133070018 | 0.43585226 | SLA | 6 30909277 | 0.341552859 | GTF2H4 |
| 12 54958518 | 0.237481959 | TESPA1 | 1 169547030 | 0.434502492 | F5 | 6 32639278 | 0.340657328 | HLA-DQA1 |
| 2 166924342 | 0.235887143 | XIRP2 | 3 195961005 | 0.432710258 | SDHAP1 | 7 104420565 | 0.33927951 | LHFPL3 |
| 1 65398537 | 0.234873839 | DNAJC6 | 2 38492697 | 0.431394771 | LOC101929596 | 16 16204352 | 0.338423535 | ABCC6 |
| 2 167000945 | 0.234777853 | XIRP2 | 1 36053026 | 0.430296874 | AGO3 | 20 63417403 | 0.33790404 | KCNQ2 |
| 5 178463611 | 0.233314395 | COL23A1 | 4 77048967 | 0.429824202 | CCNI | 6 99609905 | 0.337621578 | PRDM13 |

| | | | | | | | | |
|---|---|---|---|---|---|---|---|---|
| 5_178464136 | 0.233314395 | COL23A1 | 14_92480373 | 0.42860473 | SLC24A4 | 7 40846059 | 0.337435048 | SUGCT |
| 15 90048452 | 0.233123755 | ZNF710 | 1 35517621 | 0.427701244 | KIAA0319L | 15 55629395 | 0.337087665 | PRTG |
| 15 90048467 | 0.233123755 | ZNF710 | 9 113945114 | 0.425836587 | ZNF618 | 6 30925350 | 0.336890336 | VARS2 |
| 15_90048239 | 0.233011768 | ZNF710 | 7_74221411 | 0.425441922 | LAT2 | 12_120903692 | 0.335990585 | SPPL3 |
| 2 170402412 | 0.232945352 | MYO3B | 4 89777813 | 0.424953378 | SNCA | 7 104420525 | 0.335586929 | LHFPL3 |
| 16 10794077 | 0.232856482 | TVP23A | 1 35564290 | 0.424360212 | NCDN | 1 54711988 | 0.335333093 | MROH7-TTC4 |
| 14 104786672 | 0.231072946 | AKT1 | 1 35526435 | 0.421676123 | KIAA0319L | 1 35373463 | 0.333556861 | ZMYM4 |
| 9 19740985 | 0.230814835 | SLC24A2 | 1 9939369 | 0.420949086 | LZIC | 6 30921612 | 0.332832206 | VARS2 |
| 7 90258563 | 0.230715446 | C7orf63 | 9 3470503 | 0.420855209 | RFX3 | 6 30931969 | 0.332832206 | SFTA2 |
| 7 104903569 | 0.229937835 | LHFPL3 | 1 35435753 | 0.420654543 | KIAA0319L | 7 40843044 | 0.332779568 | SUGCT |
| 19 56603797 | 0.229903862 | ZNF71 | 1 35466087 | 0.420654543 | KIAA0319L | 6 30899329 | 0.332636508 | DDR1 |
| 2 71225275 | 0.229621596 | PAIP2B | 1 35476333 | 0.420654543 | KIAA0319L | 7 40849441 | 0.331775931 | SUGCT |
| 6 144678939 | 0.229535318 | UTRN | 1 35551875 | 0.420654543 | KIAA0319L | 6_29943088 | 0.331115271 | HLA-A |
| 2 166908328 | 0.228247094 | XIRP2 | 1 35510974 | 0.42045968 | KIAA0319L | 6 99614864 | 0.329841576 | PRDM13 |
| 3 55060445 | 0.228107193 | CACNA2D3 | 12 109725735 | 0.420330759 | FAM222A | 1 1295060 | 0.328602749 | ACAP3 |
| 20 46192922 | 0.225661871 | CDH22 | 1 36168273 | 0.420330759 | MAP7D1 | 12 120781467 | 0.327439407 | SPPL3 |
| 4 6474991 | 0.225438285 | PPP2R2C | 1 35494796 | 0.420040249 | KIAA0319L | 6 29944837 | 0.326119766 | HLA-A |
| 12 54961249 | 0.225224444 | TESPA1 | 12 12205337 | 0.419951588 | LRP6 | 7 40845914 | 0.326038354 | SUGCT |
| 15 68800677 | 0.224997597 | ANP32A | 22 46704869 | 0.41977893 | CERK | 17 5556480 | 0.324366399 | NLRP1 |
| 1 65392468 | 0.224910904 | DNAJC6 | 1 35561470 | 0.419486112 | NCDN | 7 99056948 | 0.324295158 | SMURF1 |
| 1 65397161 | 0.224910904 | DNAJC6 | 2 46340777 | 0.419485349 | EPAS1 | 6 29942547 | 0.322469756 | HLA-A |
| 1 65398515 | 0.224910904 | DNAJC6 | 4 5312437 | 0.417158194 | STK32B | 6 30944637 | 0.322169184 | DPCR1 |
| 4 89732197 | 0.224361253 | SNCA | 14 67296538 | 0.414954412 | MPP5 | 2 231282159 | 0.321509976 | ARMC9 |
| 4 89733003 | 0.224361253 | SNCA | 16 61718649 | 0.414598272 | CDH8 | 2 231295578 | 0.321509976 | ARMC9 |
| 15 90035221 | 0.224313906 | ZNF710 | 16 61726490 | 0.413870456 | CDH8 | 7 28460810 | 0.321181408 | CREB5 |
| 2 46361893 | 0.224039187 | EPAS1 | 2 38492682 | 0.412812998 | LOC101929596 | 17 18759613 | 0.320888415 | FBXW10 |
| 2 166912386 | 0.22402212 | XIRP2 | 1 35446550 | 0.412644975 | KIAA0319L | 7 40843953 | 0.320468987 | SUGCT |
| 2 46625378 | 0.223947397 | CRIPT | 15 68800677 | 0.411300439 | ANP32A | 6 30889295 | 0.320239969 | DDR1 |
| 12 54970710 | 0.223882377 | TESPA1 | 6_148377179 | 0.410792567 | SASH1 | 7_40852085 | 0.320137791 | SUGCT |
| 6 166787354 | 0.22386813 | RPS6KA2 | 6 148377182 | 0.410792567 | SASH1 | 6 30911142 | 0.319528886 | GTF2H4 |
| 2 166998948 | 0.22380627 | XIRP2 | 6 148377177 | 0.410739309 | SASH1 | 7 40854869 | 0.317406918 | SUGCT |
| 15_68800699 | 0.22366854 | ANP32A | 1_36155055 | 0.410655228 | TRAPPC3 | 17_19276667 | 0.316744751 | EPN2 |
| 6 170257033 | 0.222791907 | LOC154449 | 6 148377176 | 0.41062762 | SASH1 | 6 30890211 | 0.316249872 | DDR1 |
| 3 129424716 | 0.222655672 | EFCAB12 | 1 202332917 | 0.410208522 | UBE2T | 6 30890470 | 0.316249872 | DDR1 |
| 16 89160390 | 0.222579912 | LINC00304 | 1 35530769 | 0.408986118 | KIAA0319L | 6 30890477 | 0.316249872 | DDR1 |
| 2 166942654 | 0.221433172 | XIRP2 | 1 35574361 | 0.408986118 | TFAP2E | 6 30909425 | 0.316249872 | GTF2H4 |
| 14 104784859 | 0.220792278 | AKT1 | 15 68800683 | 0.408576269 | ANP32A | 11 118586915 | 0.315757965 | ARCN1 |
| 17 5918271 | 0.220187007 | LOC339166 | 17 531954 | 0.407936331 | VPS53 | 13 92709030 | 0.315446579 | GPC5 |
| 2_71362266 | 0.219941394 | ZNF638 | 12_110133569 | 0.407102367 | IFT81 | 6_30910942 | 0.315427423 | GTF2H4 |
| 1 231387257 | 0.219781536 | EGLN1 | 16 61698128 | 0.404920887 | CDH8 | 1 244578933 | 0.314862168 | C1orf101 |
| 9 114244069 | 0.21964508 | COL27A1 | 7 23002157 | 0.402633866 | FAM126A | 7 40846335 | 0.314327474 | SUGCT |
| 22_32599765 | 0.219539108 | SYN3 | 1_35453860 | 0.402234936 | KIAA0319L | 7_40845629 | 0.314066712 | SUGCT |
| 1 6130155 | 0.219087961 | CHD5 | 11 57603839 | 0.400604353 | SERPING1 | 7 40846977 | 0.314066712 | SUGCT |
| 4 143593396 | 0.218172266 | FREM3 | 12 109723007 | 0.400137295 | FAM222A | 7 40848107 | 0.314066712 | SUGCT |
| 2_71184783 | 0.218161613 | PAIP2B | 1_35459989 | 0.398544403 | KIAA0319L | 12_2066247 | 0.312636916 | CACNA1C |
| 2 71188745 | 0.218161613 | PAIP2B | 1 35545483 | 0.398544403 | KIAA0319L | 6 30909219 | 0.312194472 | GTF2H4 |
| 2 71194076 | 0.218161613 | PAIP2B | 1 35453008 | 0.398340937 | KIAA0319L | 1 246054991 | 0.311632038 | SMYD3 |
| 2_71201818 | 0.218161613 | PAIP2B | 17_75806299 | 0.397827672 | UNK | 3_139116493 | 0.310722605 | BPESC1 |
| 5 178469651 | 0.218112007 | COL23A1 | 16 61731180 | 0.397311255 | CDH8 | 6 29943228 | 0.309624576 | HLA-A |
| 1 153094346 | 0.217856933 | SPRR2E | 12 336284 | 0.396296425 | KDM5A | 6 32640149 | 0.309460066 | HLA-DQA1 |
| 4 175730775 | 0.217766133 | GPM6A | 2 30469577 | 0.394740464 | LCLAT1 | 7 154654457 | 0.308189408 | DPP6 |
| 4 89744105 | 0.217719519 | SNCA | 2 30469578 | 0.394740464 | LCLAT1 | 7 40844643 | 0.307817091 | SUGCT |
| 7 93501505 | 0.217649754 | CALCR | 10 19621375 | 0.393528089 | MALRD1 | 7 40847812 | 0.307817091 | SUGCT |
| 6 89117612 | 0.217409993 | SRSF12 | 2 46361893 | 0.392947355 | EPAS1 | 6 32584222 | 0.307767573 | HLA-DRB1 |
| 12_54961449 | 0.217215899 | TESPA1 | 16_61724999 | 0.390369482 | CDH8 | 9_20375605 | 0.307484923 | MLLT3 |
| 5 106858508 | 0.216945452 | LOC102467213 | 20 35141125 | 0.39036491 | EDEM2 | 20 63420102 | 0.305570283 | KCNQ2 |
| 8 138299395 | 0.216868421 | FAM135B | 14 34714439 | 0.3902132 | CFL2 | 20 63423684 | 0.305570283 | KCNQ2 |
| 7_104888950 | 0.216525015 | LHFPL3 | 5_119142518 | 0.389669912 | DMXL1 | 6_30897948 | 0.304298701 | DDR1 |
| 1 62122048 | 0.216522255 | INADL | 16 61728911 | 0.389570297 | CDH8 | 3 139108375 | 0.304256035 | BPESC1 |
| 19 45951628 | 0.216200722 | NOVA2 | 2 166072839 | 0.389287349 | SCN1A | 7 40846064 | 0.304111836 | SUGCT |
| 1_65397064 | 0.215649702 | DNAJC6 | 2_46366397 | 0.389159812 | EPAS1 | 10_96259933 | 0.30315013 | BLNK |
| 1 65402032 | 0.215649702 | DNAJC6 | 13 33519218 | 0.388081809 | STARD13 | 7 72272060 | 0.302398135 | CALN1 |
| 2 71078682 | 0.215530296 | NAGK | 22 28955053 | 0.387705403 | ZNRF3 | 20 63422401 | 0.301884526 | KCNQ2 |
| 2 167023969 | 0.215265917 | XIRP2 | 6 6189965 | 0.387371751 | F13A1 | 7 40847117 | 0.30138036 | SUGCT |
| 20 46966515 | 0.21494536 | EYA2 | 14 92474795 | 0.386751642 | SLC24A4 | 7 71136633 | 0.301030965 | WBSCR17 |
| 6 170254367 | 0.21494536 | LOC154449 | 19 12051106 | 0.386463341 | ZNF878 | 6 30884970 | 0.30079421 | DDR1 |
| 17 46763843 | 0.214386663 | WNT3 | 19 12051108 | 0.386463341 | ZNF878 | 6 30909983 | 0.30079421 | GTF2H4 |
| 2 46365668 | 0.21379408 | EPAS1 | 19 12051110 | 0.386463341 | ZNF878 | 12 20040535 | 0.300559011 | LOC100506393 |
| 13 70104674 | 0.213734908 | KLHL1 | 19 12051112 | 0.386399908 | ZNF878 | 6 29945609 | 0.299679212 | HLA-A |
| 2 71203588 | 0.213560161 | PAIP2B | 14 47400731 | 0.385725527 | MDGA2 | 6 32642509 | 0.299545795 | HLA-DQA1 |
| 7 104899443 | 0.213445197 | LHFPL3 | 1 36032475 | 0.385507967 | AGO3 | 2 231321254 | 0.299453275 | ARMC9 |
| 18 64369035 | 0.213418452 | LOC284294 | 9 76675834 | 0.385357005 | PRUNE2 | 3 138515725 | 0.29890236 | CEP70 |
| 1 65395657 | 0.213022281 | DNAJC6 | 1 35508447 | 0.384077323 | KIAA0319L | 6 32639082 | 0.298501313 | HLA-DQA1 |
| 7 93483458 | 0.212948944 | CALCR | 12 109790250 | 0.384020748 | TRPV4 | 6 30886313 | 0.298148428 | DDR1 |
| 5 178468382 | 0.212806624 | COL23A1 | 17 73411969 | 0.382666451 | SDK2 | 20 63429411 | 0.297839782 | KCNQ2 |
| 3 314288 | 0.212768479 | CHL1 | 4 188091302 | 0.382437026 | TRIML2 | 2 231320746 | 0.297460312 | ARMC9 |
| 2 166998793 | 0.212537869 | XIRP2 | 4 188091304 | 0.382437026 | TRIML2 | 1 244578791 | 0.297363763 | C1orf101 |
| 11 113333435 | 0.212414911 | TTC12 | 7 74219027 | 0.382056374 | LAT2 | 3 119381995 | 0.297215576 | ARHGAP31 |
| 12 12514615 | 0.212335635 | DUSP16 | 17 75841287 | 0.381744545 | UNC13D | 12 111965983 | 0.295451917 | TMEM116 |
| 12 54953406 | 0.212240712 | TESPA1 | 16 61732469 | 0.380150161 | CDH8 | 6 30899889 | 0.295448127 | DDR1 |
| 1 6132655 | 0.212092279 | CHD5 | 16 61708792 | 0.379371756 | CDH8 | 7 40848999 | 0.295401723 | SUGCT |
| 1 6133177 | 0.212092279 | CHD5 | 16 61721447 | 0.379371756 | CDH8 | 16 16221475 | 0.295127639 | ABCC6 |
| 4 89777813 | 0.211839316 | SNCA | 1 38922419 | 0.3788782 | RHBDL2 | 3 138564310 | 0.295116047 | CEP70 |
| 15 90051033 | 0.211783305 | ZNF710 | 1 35191655 | 0.378238613 | SFPQ | 17 19253591 | 0.295070935 | EPN2 |
| 14 104960032 | 0.211630737 | AHNAK2 | 1 35191684 | 0.378238613 | SFPQ | 7 40843174 | 0.294747365 | SUGCT |
| 11_44831187 | 0.211582694 | TSPAN18 | 10_87905467 | 0.378144152 | PTEN | 6_151406382 | 0.294311121 | RMND1 |
| 13 70097650 | 0.211537515 | KLHL1 | 1 35504840 | 0.377998078 | KIAA0319L | 6_30891743 | 0.294137964 | DDR1 |
| 2 167025497 | 0.211019698 | XIRP2 | 4 188100407 | 0.376419321 | TRIML2 | 2 70068877 | 0.29392979 | PCBP1-AS1 |
| 1 65394101 | 0.210999499 | DNAJC6 | 16 61656199 | 0.376215085 | CDH8 | 20 63418440 | 0.293850546 | KCNQ2 |
| 15 40289342 | 0.210867407 | PLCB2 | 16 61656209 | 0.376215085 | CDH8 | 20 63424386 | 0.293850546 | KCNQ2 |
| 5 109782503 | 0.210505091 | MAN2A1 | 4 108746093 | 0.3752831 | ETNPPL | 6 30918341 | 0.29377692 | VARS2 |
| 9 40998400 | 0.210240742 | LOC100132352 | 4 108748257 | 0.3752831 | ETNPPL | 6 30891080 | 0.292763601 | DDR1 |
| 13 110311329 | 0.210179209 | COL4A2 | 1 27281440 | 0.374804174 | WDTC1 | 6_30910802 | 0.292763601 | GTF2H4 |
| 19 37974514 | 0.210097411 | SIPA1L3 | 16 57078469 | 0.374073066 | NLRC5 | 2 70048354 | 0.292593014 | PCBP1-AS1 |
| 7 78019354 | 0.209793935 | MAGI2 | 14 61576584 | 0.373899748 | FLJ22447 | 5 175960547 | 0.292268336 | THOC3 |
| 14 78913140 | 0.209521054 | NRXN3 | 9_113939095 | 0.373210726 | ZNF618 | 4_56200826 | 0.29210203 | KIAA1211 |
| 13 28217669 | 0.209454407 | PAN3 | 2 241945925 | 0.373127477 | LINC01237 | 2 179474268 | 0.292016714 | ZNF385B |

| | | | | | | | | |
|---|---|---|---|---|---|---|---|---|
| 13_28240518 | 0.209454407 | PAN3 | 12_109720511 | 0.372879414 | FAM222A | 3 181181340 | 0.291007663 | SOX2-OT |
| 7 104893380 | 0.208924266 | LHFPL3 | 12 109721315 | 0.372879414 | FAM222A | 21 14172517 | 0.290684897 | LIPI |
| 2 46366397 | 0.208907194 | EPAS1 | 4 184200580 | 0.372522945 | ENPP6 | 12 111717330 | 0.290492939 | ACAD10 |
| 15_27489030 | 0.208654954 | GABRG3 | 1_20115818 | 0.371751129 | PLA2G2D | 17_19293674 | 0.290237122 | EPN2 |
| 5 13730584 | 0.208516711 | DNAH5 | 1 2233261 | 0.371049804 | SKI | 12 6597046 | 0.290231519 | CHD4 |
| 2 71203196 | 0.20833439 | PAIP2B | 5 119468180 | 0.370951698 | HSD17B4 | 2 62070631 | 0.290224627 | COMMD1 |
| 11 44813214 | 0.208275099 | TSPAN18 | 19 13336602 | 0.370778757 | CACNA1A | 16 16216437 | 0.29021639 | ABCC6 |
| 14 104790244 | 0.207898693 | AKT1 | 16 61715348 | 0.369906197 | CDH8 | 20 46200205 | 0.289902398 | CDH22 |
| 6 144674342 | 0.207624 | UTRN | 16 61718480 | 0.369906197 | CDH8 | 7 154654385 | 0.289268548 | DPP6 |
| 12 53019641 | 0.207282668 | EIF4B | 16 61720415 | 0.369906197 | CDH8 | 7 40834777 | 0.289219381 | SUGCT |
| 19 37965377 | 0.207216861 | SIPA1L3 | 16 61721870 | 0.369906197 | CDH8 | 7 40848146 | 0.289179302 | SUGCT |
| 12 81440808 | 0.207103041 | PPFIA2 | 16 61725975 | 0.369906197 | CDH8 | 7 40851527 | 0.289179302 | SUGCT |
| 9 132335355 | 0.207062126 | SETX | 16 61730429 | 0.369906197 | CDH8 | 12_65771220 | 0.288376038 | RPSAP52 |
| 12 12509518 | 0.20699143 | DUSP16 | 6 129219735 | 0.369276073 | LAMA2 | 5 94988598 | 0.288117835 | MCTP1 |
| 6 170257900 | 0.206937293 | LOC154449 | 1 35471647 | 0.369003756 | KIAA0319L | 2 70027409 | 0.287905873 | PCBP1-AS1 |
| 6 144540445 | 0.206857339 | UTRN | 4 188095924 | 0.368195475 | TRIML2 | 7 120339056 | 0.287670624 | KCND2 |
| 1 62124379 | 0.206795136 | INADL | 13 71841293 | 0.367930971 | DACH1 | 3 133583611 | 0.287274741 | CDV3 |
| 14 32728362 | 0.206747479 | AKAP6 | 9 113961730 | 0.367796213 | ZNF618 | 21 14176693 | 0.286848257 | LIPI |
| 4 89736340 | 0.206404852 | SNCA | 4 92989291 | 0.367607855 | GRID2 | 11 118588987 | 0.286378535 | ARCN1 |
| 1 96371372 | 0.206359527 | LOC101928241 | 17 75825549 | 0.367257558 | UNK | 6 30920195 | 0.286212501 | VARS2 |
| 15 90046425 | 0.206047508 | ZNF710 | 4 188093667 | 0.367078221 | TRIML2 | 1 1307276 | 0.286206586 | ACAP3 |
| 2 170415769 | 0.205861539 | MYO3B | 17 65844870 | 0.366817077 | CEP112 | 14 96075813 | 0.285908687 | C14orf132 |
| 8 142752166 | 0.205811241 | LYPD2 | 16 61721022 | 0.366750299 | CDH8 | 4 56199694 | 0.285825514 | KIAA1211 |
| 12 54981582 | 0.205799593 | TESPA1 | 16 61725848 | 0.366750299 | CDH8 | 7 98626147 | 0.28576025 | NPTX2 |
| 1 65394227 | 0.205781406 | DNAJC6 | 2_46365668 | 0.366688987 | EPAS1 | 3 138553290 | 0.285251476 | CEP70 |
| 1 65399979 | 0.205781406 | DNAJC6 | 17 73412025 | 0.366106627 | SDK2 | 7 40857668 | 0.284674292 | SUGCT |
| 10 52166914 | 0.205768105 | PRKG1 | 4 89732197 | 0.365946542 | SNCA | 12 27260554 | 0.284395631 | STK38L |
| 6 166850641 | 0.205327424 | RPS6KA2 | 3 192342945 | 0.365502734 | FGF12 | 17 65738048 | 0.284346981 | CEP112 |
| 1 169520708 | 0.204940727 | F5 | 4_108756967 | 0.365011167 | ETNPPL | 5_41488479 | 0.284114469 | PLCXD3 |
| 11 132830336 | 0.20491302 | OPCML | 1 35555692 | 0.364610637 | KIAA0319L | 19 56620870 | 0.283040985 | ZNF71 |
| 16 89743812 | 0.204911956 | FANCA | 1 144426562 | 0.363437121 | NBPF15 | 2 231315439 | 0.282975819 | ARMC9 |
| 7_24865254 | 0.2046048 | OSBPL3 | 14_92480355 | 0.363104107 | SLC24A4 | 20_53140534 | 0.282714512 | TSHZ2 |
| 11 130889328 | 0.204597308 | SNX19 | 2 187378134 | 0.362051387 | CALCRL | 2 69995395 | 0.282498483 | PCBP1-AS1 |
| 19 54502908 | 0.204521277 | LAIR2 | 1 35187479 | 0.361751533 | SFPQ | 2 70066363 | 0.282322642 | PCBP1-AS1 |
| 2 71224341 | 0.204161038 | PAIP2B | 1 35510375 | 0.361354369 | KIAA0319L | 2 231320975 | 0.282161395 | ARMC9 |
| 6 166786786 | 0.204065426 | RPS6KA2 | 1 231387257 | 0.361160743 | EGLN1 | 7 40842339 | 0.281900307 | SUGCT |
| 6 47715213 | 0.203887472 | GPR115 | 16 61722961 | 0.361085084 | CDH8 | 10 17881885 | 0.281741668 | MRC1 |
| 1 6134287 | 0.203814434 | CHD5 | 16 61731296 | 0.361085084 | CDH8 | 20 63417594 | 0.281608437 | KCNQ2 |
| 1_96371830 | 0.203666089 | LOC101928241 | 14_61548586 | 0.3608251 | PRKCH | 7_40852213 | 0.281449159 | SUGCT |
| 2 167008385 | 0.203630645 | XIRP2 | 19 2805941 | 0.359447215 | THOP1 | 5 168457149 | 0.281444902 | WWC1 |
| 12 81372709 | 0.203390349 | PPFIA2 | 12 109719428 | 0.358445186 | FAM222A | 6 29944633 | 0.281322433 | HLA-A |
| 1_6132805 | 0.203351645 | CHD5 | 12_109719446 | 0.358445186 | FAM222A | 7_152201625 | 0.28108393 | KMT2C |
| 1 6133244 | 0.203351645 | CHD5 | 12 109719874 | 0.358445186 | FAM222A | 7 152201626 | 0.28108393 | KMT2C |
| 1 6133404 | 0.203351645 | CHD5 | 1 35553423 | 0.357731392 | KIAA0319L | 9 94267817 | 0.281083001 | ZNF169 |
| 1_6133958 | 0.203351645 | CHD5 | 3_55063478 | 0.357425908 | CACNA2D3 | 7_40843835 | 0.280852692 | SUGCT |
| 15 90049165 | 0.2028662 | ZNF710 | 5 1408204 | 0.357190896 | SLC6A3 | 7 40844821 | 0.280852692 | SUGCT |
| 8 10388851 | 0.202812963 | MSRA | 3 54993855 | 0.357093332 | CACNA2D3 | 12 24148710 | 0.280722989 | SOX5 |
| 4_175730693 | 0.20280243 | GPM6A | 3_155610228 | 0.356553052 | PLCH1 | 8_72818977 | 0.280595396 | KCNB2 |
| 7 93497156 | 0.202720121 | CALCR | 15 68800699 | 0.356478396 | ANP32A | 1 244595896 | 0.280511798 | C1orf101 |
| 4 6471273 | 0.202674177 | PPP2R2C | 15 68800700 | 0.356478396 | ANP32A | 1 244595902 | 0.280511798 | C1orf101 |
| 3 11823257 | 0.202310217 | TAMM41 | 3 192342893 | 0.355783231 | FGF12 | 1 244595903 | 0.280511798 | C1orf101 |
| 4 89778628 | 0.202300773 | SNCA | 16 61700102 | 0.35572873 | CDH8 | 7 71135310 | 0.280466715 | WBSCR17 |
| 3 26639475 | 0.202287612 | LRRC3B | 19 42373707 | 0.355103638 | MEGF8 | 6 30139429 | 0.280362215 | TRIM40 |
| 1 62121626 | 0.202264589 | INADL | 1 36037615 | 0.354574048 | AGO3 | 2 70017852 | 0.280258882 | PCBP1-AS1 |
| 4_143691501 | 0.202221023 | FREM3 | 4_8612187 | 0.353933415 | CPZ | 12_24148713 | 0.28010683 | SOX5 |
| 1 65394186 | 0.202152402 | DNAJC6 | 16 25923330 | 0.353271386 | HS3ST4 | 20 63423847 | 0.279896648 | KCNQ2 |
| 1 65394977 | 0.202152402 | DNAJC6 | 4 188100493 | 0.353165237 | TRIML2 | 3 138533326 | 0.279824535 | CEP70 |
| 6_128449614 | 0.202152402 | PTPRK | 13_71838285 | 0.353061746 | DACH1 | 6_32639985 | 0.279193425 | HLA-DQA1 |
| 16 89862104 | 0.202017365 | SPIRE2 | 2 231314071 | 0.353008293 | ARMC9 | 7 152201620 | 0.27907103 | KMT2C |
| 7 94398387 | 0.201946222 | COL1A2 | 9 113937816 | 0.352973559 | ZNF618 | 1 7973926 | 0.278496362 | PARK7 |
| 7_104864247 | 0.201835669 | LHFPL3 | 1_35580543 | 0.352252408 | TFAP2E | 11_118602670 | 0.278209986 | ARCN1 |
| 1 11611017 | 0.201812346 | LOC101929181 | 4 108753304 | 0.351993923 | ETNPPL | 3 138527470 | 0.278205871 | CEP70 |
| 12 113963891 | 0.201809326 | RBM19 | 16 16216437 | 0.351815449 | ABCC6 | 20 63413947 | 0.27778733 | KCNQ2 |
| 2 71405051 | 0.201569641 | ZNF638 | 7 28168382 | 0.350735084 | JAZF1 | 4 56180940 | 0.277711223 | KIAA1211 |
| 6 169664876 | 0.201462623 | WDR27 | 4 188098566 | 0.350054282 | TRIML2 | 15 76276625 | 0.277489942 | ETFA |
| 13 70135712 | 0.201431518 | ATXN8OS | 1 35533818 | 0.350030603 | KIAA0319L | 6 64766567 | 0.277083407 | EYS |
| 13 70120550 | 0.201310921 | ATXN8OS | 4 108744020 | 0.350016043 | ETNPPL | 9 132348370 | 0.277034947 | SETX |
| 2 71208431 | 0.201265596 | PAIP2B | 7 28460810 | 0.349787128 | CREB5 | 9 132348371 | 0.277034947 | SETX |
| 2 71189412 | 0.200959952 | PAIP2B | 16 61678665 | 0.349023914 | CDH8 | 2 70070042 | 0.276730062 | PCBP1-AS1 |
| 2 71190368 | 0.200959952 | PAIP2B | 1 242396674 | 0.348435101 | PLD5 | 11 118574870 | 0.276548239 | ARCN1 |
| 6 166842963 | 0.200837088 | RPS6KA2 | 11 46105260 | 0.348284168 | PHF21A | 9 132348366 | 0.275960926 | SETX |
| 1 111844013 | 0.20073914 | KCND3 | 16 61738817 | 0.348037235 | CDH8 | 9 132348368 | 0.275960926 | SETX |
| 1 111849187 | 0.20073914 | KCND3 | 4 95448230 | 0.346978853 | UNC5C | 1 1298292 | 0.275865652 | ACAP3 |
| 1 111850631 | 0.20073914 | KCND3 | 1 35386402 | 0.346700959 | ZMYM4 | 2 50031071 | 0.275616086 | NRXN1 |
| 4 6471797 | 0.200694722 | PPP2R2C | 17 75846811 | 0.346427427 | WBP2 | 6 155440535 | 0.275365579 | NOX3 |
| 11 132835021 | 0.200485937 | OPCML | 4 146790040 | 0.346175862 | TTC29 | 19 7506429 | 0.274766135 | C19orf45 |
| 6 21802863 | 0.200340116 | CASC15 | 3 100115574 | 0.345449473 | CMSS1 | 18 74446225 | 0.274510591 | FAM69C |
| 2 170380611 | 0.200311212 | MYO3B | 12 109719231 | 0.345168182 | FAM222A | 7 152201619 | 0.274469711 | KMT2C |
| 13 28235676 | 0.200242892 | PAN3 | 12 109728152 | 0.344772471 | FAM222A | 2 70082646 | 0.274261779 | PCBP1-AS1 |
| 1 205714227 | 0.200103626 | NUCKS1 | 4 188091655 | 0.344629206 | TRIML2 | 2 70039041 | 0.274106829 | PCBP1-AS1 |
| 6 144661985 | 0.199873743 | UTRN | 4 188094107 | 0.344601938 | TRIML2 | 1 54727743 | 0.274063459 | MROH7-TTC4 |
| 9 9805496 | 0.199591647 | PTPRD | 1 35543059 | 0.344422805 | KIAA0319L | 6 69055815 | 0.273963465 | BAI3 |
| 1 6138685 | 0.199588741 | CHD5 | 16 61715968 | 0.344013448 | CDH8 | 1 244575290 | 0.273956883 | C1orf101 |
| 19 16114814 | 0.199545066 | RAB8A | 4 108756779 | 0.343998824 | ETNPPL | 12 65771115 | 0.273927345 | RPSAP52 |
| 13 70104246 | 0.199529722 | KLHL1 | 17 75820323 | 0.343879797 | UNK | 6 32638228 | 0.273794648 | HLA-DQA1 |
| 13_70109545 | 0.199529722 | ATXN8OS | 1_201494829 | 0.343542021 | CSRP1 | 3_139109923 | 0.273751862 | BPESC1 |
| 5 168898726 | 0.199433223 | SLIT3 | 7 24841446 | 0.343019236 | OSBPL3 | 3 139111511 | 0.273751862 | BPESC1 |
| 12 49811680 | 0.199265167 | NCKAP5L | 12 109720776 | 0.342606501 | FAM222A | 2 231323081 | 0.273695092 | ARMC9 |
| 7 77347004 | 0.199214677 | GSAP | 1 32214320 | 0.342193388 | DCDC2B | 18 48106758 | 0.273560651 | ZBTB7C |
| 7 78536021 | 0.199107818 | MAGI2 | 2 46370617 | 0.341960052 | EPAS1 | 1 6043858 | 0.272919659 | KCNAB2 |
| 6 21779571 | 0.199079862 | CASC15 | 2 46362184 | 0.341419062 | EPAS1 | 7 143468927 | 0.272860882 | EPHA1-AS1 |
| 1 161038782 | 0.199076248 | TSTD1 | 16 61720703 | 0.341370638 | CDH8 | 17 19276770 | 0.272765241 | EPN2 |
| 17 65164886 | 0.198868927 | RGS9 | 15 27046273 | 0.341303432 | GABRG3 | 7_40840992 | 0.272684409 | SUGCT |
| 22 42511851 | 0.198735132 | RRP7A | 16 83902152 | 0.341299831 | MLYCD | 11 118594580 | 0.27260251 | ARCN1 |
| 7 90252832 | 0.198433472 | C7orf63 | 17 75805856 | 0.341264943 | UNK | 3 181318431 | 0.272572618 | SOX2-OT |
| 9 19740790 | 0.198299982 | SLC24A2 | 4_188101170 | 0.341121364 | TRIML2 | 2_231288494 | 0.272552342 | ARMC9 |
| 1 44549609 | 0.198044883 | RNF220 | 5 73799911 | 0.341076845 | ARHGEF28 | 2 69976112 | 0.272523622 | PCBP1-AS1 |

| | | | | | | | | |
|---|---|---|---|---|---|---|---|---|
| 15_47317874 | 0.197935337 | SEMA6D | 16_61654402 | 0.340951841 | CDH8 | 12_112129490 | 0.272188206 | TRAFD1 |
| 1_162865875 | 0.197856239 | C1orf110 | 4_188097623 | 0.34035859 | TRIML2 | 2_10743995 | 0.272085009 | ATP6V1C2 |
| 13_70104426 | 0.197773406 | KLHL1 | 13_45151912 | 0.340278273 | GTF2F2 | 7_40837187 | 0.27205235 | SUGCT |
| 2_71406429 | 0.19774348 | ZNF638 | 16_61740821 | 0.339582936 | CDH8 | 11_118576441 | 0.271985668 | ARCN1 |
| 2_71366144 | 0.197742998 | ZNF638 | 1_36162458 | 0.339545467 | MAP7D1 | 10_5789120 | 0.271196926 | GDI2 |
| 2_71071187 | 0.197727934 | NAGK | 6_43566664 | 0.339052246 | XPO5 | 8_42284856 | 0.270761798 | IKBKB |
| 10_119305567 | 0.197697693 | GRK5 | 17_75831830 | 0.337852922 | UNC13D | 13_92838587 | 0.27045402 | GPC5 |
| 9_4824375 | 0.197666251 | RCL1 | 17_75832093 | 0.337852922 | UNC13D | 2_157775079 | 0.270219154 | ACVR1 |
| 5_167835704 | 0.197573157 | TENM2 | 17_75834189 | 0.337852922 | UNC13D | 8_72871802 | 0.270160838 | KCNB2 |
| 1_111846685 | 0.197553551 | KCND3 | 4_188103290 | 0.337805443 | TRIML2 | 17_19340113 | 0.270118265 | B9D1 |
| 2_50373395 | 0.197485197 | NRXN1 | 10_25496114 | 0.337768916 | GPR158 | 8_29249432 | 0.270099382 | KIF13B |
| 1_65396396 | 0.197436483 | DNAJC6 | 19_16036749 | 0.337768916 | LINC00905 | 10_96321774 | 0.269929818 | DNTT |
| 1_65396825 | 0.197436483 | DNAJC6 | 1_57155488 | 0.337486908 | DAB1 | 20_63439138 | 0.269891643 | KCNQ2 |
| 12_12502211 | 0.197089841 | DUSP16 | 4_188100765 | 0.337257859 | TRIML2 | 7_40834317 | 0.269889354 | SUGCT |
| 14_104785383 | 0.197077973 | AKT1 | 1_169532462 | 0.337130121 | F5 | 2_70074438 | 0.269863085 | PCBP1-AS1 |
| 13_70133589 | 0.197063258 | ATXN8OS | 14_50189070 | 0.337109543 | SOS2 | 2_55661927 | 0.269730835 | PNPT1 |
| 1_62117855 | 0.197057693 | INADL | 22_46699958 | 0.336996916 | CERK | 4_6355996 | 0.269612263 | PPP2R2C |
| 5_178468752 | 0.196934089 | COL23A1 | 10_54655284 | 0.336594507 | PCDH15 | 11_118534723 | 0.269597274 | TMEM25 |
| 5_178471957 | 0.196934089 | COL23A1 | 17_75843056 | 0.336518312 | UNC13D | 8_96541283 | 0.269368993 | SDC2 |
| 5_178472177 | 0.196934089 | COL23A1 | 16_61705037 | 0.336460438 | CDH8 | 2_231317193 | 0.269158573 | ARMC9 |
| 19_51313635 | 0.196920261 | IGLON5 | 16_61723759 | 0.336460438 | CDH8 | 10_104231630 | 0.268832616 | WDR96 |
| 9_131897343 | 0.196771052 | MED27 | 16_61668662 | 0.336391858 | CDH8 | 10_113096767 | 0.268806169 | TCF7L2 |
| 2_97193330 | 0.196707309 | ANKRD36 | 16_89919746 | 0.336309225 | MC1R | 2_231292630 | 0.26837256 | ARMC9 |
| 12_81365890 | 0.196654195 | PPFIA2 | 4_188092627 | 0.33555436 | TRIML2 | 2_231299776 | 0.26837256 | ARMC9 |
| 12_54950980 | 0.19663255 | TESPA1 | 3_133619123 | 0.335214995 | TOPBP1 | 11_118579031 | 0.268302391 | ARCN1 |
| 6_159228663 | 0.195847244 | FNDC1 | 17_75830530 | 0.334820147 | UNC13D | 20_63428069 | 0.268047918 | KCNQ2 |
| 2_206717111 | 0.195757766 | DYTN | 6_151259472 | 0.334590979 | AKAP12 | 14_61524143 | 0.267984722 | PRKCH |
| 15_90055003 | 0.195615955 | ZNF710 | 4_188096804 | 0.33449681 | TRIML2 | 5_141537492 | 0.267828833 | DIAPH1 |
| 1_65398326 | 0.195613283 | DNAJC6 | 6_165619329 | 0.334219085 | PDE10A | 6_29889244 | 0.267519969 | HLA-H |
| 4_184200580 | 0.1955812 | ENPP6 | 11_46107289 | 0.334182016 | PHF21A | 5_132737310 | 0.267504621 | KIF3A |
| 4_6466258 | 0.195580907 | PPP2R2C | 11_46107989 | 0.334182016 | PHF21A | 6_30887474 | 0.267361366 | DDR1 |
| 1_220749435 | 0.195292939 | 2-Mar | 11_46113256 | 0.334182016 | PHF21A | 7_40857115 | 0.267053645 | SUGCT |
| 16_10788498 | 0.195173428 | TVP23A | 11_46114792 | 0.334182016 | PHF21A | 7_102313218 | 0.266552009 | SH2B2 |
| 12_291902 | 0.195103053 | KDM5A | 5_143026709 | 0.333965512 | ARHGAP26 | 4_54682757 | 0.266001846 | KIT |
| 19_38613810 | 0.195013715 | MAP4K1 | 2_71224341 | 0.33362202 | PAIP2B | 20_63435168 | 0.265701292 | KCNQ2 |
| 2_170427218 | 0.19491682 | MYO3B | 16_61710471 | 0.333584372 | CDH8 | 8_72825893 | 0.265640778 | KCNB2 |
| 8_17723282 | 0.194852982 | MTUS1 | 3_55060445 | 0.333584372 | CACNA2D3 | 7_117319530 | 0.265583218 | WNT2 |
| 7_78029924 | 0.194833749 | MAGI2 | 11_60294913 | 0.333444963 | MS4A4A | 9_99151417 | 0.265522846 | TGFBR1 |
| 15_83546933 | 0.194742689 | SH3GL3 | 16_61667608 | 0.333397105 | CDH8 | 2_69965448 | 0.265424874 | PCBP1-AS1 |
| 4_6469887 | 0.194680845 | PPP2R2C | 1_36090586 | 0.333170364 | ADPRHL2 | 10_52035703 | 0.265162589 | PRKG1 |
| 19_38605337 | 0.194642248 | MAP4K1 | 15_66201776 | 0.332643769 | MEGF11 | 20_63424980 | 0.26458072 | KCNQ2 |
| 5_106867102 | 0.194631533 | LOC102467213 | 2_213128808 | 0.332491484 | IKZF2 | 3_139116442 | 0.264374727 | BPESC1 |
| 22_40491070 | 0.194609398 | MKL1 | 4_95448231 | 0.332255664 | UNC5C | 2_29069702 | 0.264226593 | C2orf71 |
| 20_46970688 | 0.194579547 | EYA2 | 5_73789612 | 0.332196996 | ARHGEF28 | 8_72843434 | 0.264014403 | KCNB2 |
| 21_41451829 | 0.194526693 | MX1 | 16_2260148 | 0.332165534 | RNPS1 | 10_96247101 | 0.263886816 | BLNK |
| 7_78027675 | 0.194460639 | MAGI2 | 13_113033636 | 0.332111192 | MCF2L | 10_124546953 | 0.263630086 | LHPP |
| 3_129411879 | 0.194451085 | EFCAB12 | 10_25457533 | 0.331811504 | GPR158 | 1_52783914 | 0.263422355 | ZYG11B |
| 12_54949635 | 0.194402065 | TESPA1 | 2_231326057 | 0.331435888 | ARMC9 | 19_50694842 | 0.26330034 | SHANK1 |
| 15_90056443 | 0.194142001 | ZNF710 | 12_109714679 | 0.331204907 | FAM222A | 11_118590890 | 0.263054448 | ARCN1 |
| 6_166848939 | 0.194058232 | RPS6KA2 | 11_114088925 | 0.331131941 | ZBTB16 | 19_50694841 | 0.262898874 | SHANK1 |
| 2_170380020 | 0.194055639 | MYO3B | 1_35501313 | 0.330509931 | KIAA0319L | 15_47302625 | 0.262583659 | SEMA6D |
| 1_65395977 | 0.193819051 | DNAJC6 | 1_35524302 | 0.330509931 | KIAA0319L | 14_63374689 | 0.262572473 | PPP2R5E |
| 7_104831880 | 0.193773293 | LHFPL3 | 13_85500052 | 0.330501183 | LINC00351 | 11_118600708 | 0.262339888 | ARCN1 |
| 2_46329206 | 0.193628554 | EPAS1 | 1_154161132 | 0.330278057 | TPM3 | 2_69967694 | 0.262278422 | PCBP1-AS1 |
| 6_166850623 | 0.193628554 | RPS6KA2 | 10_15266867 | 0.330265939 | FAM171A1 | 5_178553100 | 0.262131522 | COL23A1 |
| 8_10424249 | 0.19359146 | MSRA | 2_71190368 | 0.330193929 | PAIP2B | 2_70066243 | 0.262081373 | PCBP1-AS1 |
| 6_159230639 | 0.193578888 | FNDC1 | 4_92989294 | 0.330138637 | GRID2 | 15_47302305 | 0.262070983 | SEMA6D |
| 7_104891759 | 0.193450785 | LHFPL3 | 12_109720816 | 0.330039854 | FAM222A | 2_70172238 | 0.261638295 | C2orf42 |
| 6_144675120 | 0.193388394 | UTRN | 12_109722569 | 0.330039854 | FAM222A | 8_142365637 | 0.261374007 | TSNARE1 |
| 6_21785925 | 0.193388394 | CASC15 | 1_36159501 | 0.330039854 | MAP7D1 | 11_78975342 | 0.261368899 | TENM4 |
| 22_49562790 | 0.193246932 | C22orf34 | 1_36097557 | 0.330010637 | COL8A2 | 3_138517788 | 0.261226421 | CEP70 |
| 16_82912670 | 0.193192449 | CDH13 | 3_192345204 | 0.329979949 | FGF12 | 15_47298549 | 0.261125833 | SEMA6D |
| 19_3697325 | 0.193173971 | PIP5K1C | 17_75800509 | 0.329549487 | UNK | 12_40422942 | 0.260894523 | MUC19 |
| 9_16677805 | 0.193123755 | BNC2 | 17_75842241 | 0.329441833 | UNC13D | 3_133585646 | 0.260894486 | CDV3 |
| 6_88776633 | 0.193062219 | RNGTT | 4_108742215 | 0.329229784 | ETNPPL | 7_40837798 | 0.260804356 | SUGCT |
| 1_243481774 | 0.19303814 | SDCCAG8 | 4_108743739 | 0.329229784 | ETNPPL | 7_40838117 | 0.260455627 | SUGCT |
| 9_40998401 | 0.193030081 | LOC100132352 | 3_155625299 | 0.328959447 | PLCH1 | 2_70081606 | 0.260316065 | PCBP1-AS1 |
| 5_109796760 | 0.192890979 | MAN2A1 | 10_868379 | 0.328901521 | LARP4B | 3_687663 | 0.260104198 | LINC01266 |
| 6_166842353 | 0.192817767 | RPS6KA2 | 15_83052811 | 0.328901521 | BTBD1 | 12_40430732 | 0.26009242 | MUC19 |
| 6_166851534 | 0.19281098 | RPS6KA2 | 4_108747444 | 0.328878739 | ETNPPL | 6_32641509 | 0.259995959 | HLA-DQA1 |
| 17_4210465 | 0.192757046 | ANKFY1 | 2_181977153 | 0.328863038 | PPP1R1C | 3_687664 | 0.259983299 | LINC01266 |
| 1_162863385 | 0.192599383 | C1orf110 | 3_4535427 | 0.328650091 | ITPR1 | 3_138495652 | 0.259958614 | CEP70 |
| 2_71075920 | 0.192597271 | NAGK | 4_188095782 | 0.32845534 | TRIML2 | 2_55593346 | 0.259928825 | SMEK2 |
| 9_19742147 | 0.192431348 | SLC24A2 | 1_93809776 | 0.328429842 | BCAR3 | 6_32442360 | 0.259858278 | HLA-DRA |
| 2_71194492 | 0.192397636 | PAIP2B | 7_149279884 | 0.32841262 | ZNF783 | 13_113033636 | 0.259724098 | MCF2L |
| 1_11611285 | 0.192375976 | LOC101929181 | 3_21583956 | 0.328209592 | ZNF385D | 12_112071071 | 0.259622813 | NAA25 |
| 22_17498704 | 0.192330734 | CECR2 | 10_25458659 | 0.328150765 | GPR158 | 3_33051665 | 0.259529092 | GLB1 |
| 5_109711608 | 0.192328118 | MAN2A1 | 1_201494842 | 0.327973267 | CSRP1 | 2_70086835 | 0.259389541 | PCBP1-AS1 |
| 1_231204650 | 0.192275659 | TRIM67 | 1_201494843 | 0.327973267 | CSRP1 | 17_19289847 | 0.259325131 | EPN2 |
| 1_231206010 | 0.192275659 | TRIM67 | 2_1321878 | 0.326820472 | SNTG2 | 12_3015345 | 0.259251513 | TEAD4 |
| 12_49821241 | 0.192218836 | NCKAP5L | 2_1321883 | 0.326820472 | SNTG2 | 20_46192922 | 0.259201281 | CDH22 |
| 19_2598521 | 0.192208716 | GNG7 | 17_75824854 | 0.326494511 | UNK | 11_118578681 | 0.259104644 | ARCN1 |
| 11_11451898 | 0.192146713 | GALNT18 | 16_61715001 | 0.326317949 | CDH8 | 12_57182709 | 0.258986373 | LRP1 |
| 4_6470377 | 0.192042115 | PPP2R2C | 16_61716694 | 0.326317949 | CDH8 | 2_55681494 | 0.25839439 | PNPT1 |
| 4_7224889 | 0.191902048 | SORCS2 | 1_35848865 | 0.326071148 | AGO4 | 22_26611459 | 0.258326219 | CRYBB1 |
| 2_170430014 | 0.191889319 | MYO3B | 2_158306692 | 0.325897424 | CCDC148 | 11_118549386 | 0.258267286 | IFT46 |
| 16_10794937 | 0.19185688 | TVP23A | 8_71327649 | 0.3253223 | EYA1 | 11_78980648 | 0.25818635 | TENM4 |
| 13_70135663 | 0.191815849 | ATXN8OS | 15_100279147 | 0.324840955 | ADAMTS17 | 1_54711973 | 0.257949703 | MROH7-TTC4 |
| 13_70136101 | 0.191815849 | ATXN8OS | 10_68512722 | 0.32483257 | SLC25A16 | 22_26611460 | 0.257917066 | CRYBB1 |
| 3_129448701 | 0.191812508 | IFT122 | 16_61732075 | 0.324042661 | CDH8 | 4_56189001 | 0.257728552 | KIAA1211 |
| 3_30795044 | 0.191785887 | GADL1 | 7_50783422 | 0.323976174 | GRB10 | 7_40836461 | 0.257481341 | SUGCT |
| 3_30795481 | 0.191785887 | GADL1 | 14_61543519 | 0.323779307 | PRKCH | 7_40836515 | 0.257481341 | SUGCT |
| 1_96371128 | 0.191712349 | LOC101928241 | 1_35908222 | 0.32367961 | AGO1 | 12_111651256 | 0.257280836 | BRAP |
| 4_89765968 | 0.191523178 | SNCA | 20_62178150 | 0.323453537 | SS18L1 | 12_111661715 | 0.257280836 | BRAP |
| 4_89733852 | 0.19143826 | SNCA | 16_61668369 | 0.323353172 | CDH8 | 3_138475135 | 0.257204809 | ESYT3 |
| 5_109724132 | 0.191433539 | MAN2A1 | 14_64014970 | 0.323335262 | SYNE2 | 6_39839319 | 0.256784481 | DAAM2 |
| 2_215100017 | 0.191421007 | ABCA12 | 19_16034843 | 0.323314094 | LINC00905 | 6_30639499 | 0.256757395 | ATAT1 |

| | | | | | | | | |
|---|---|---|---|---|---|---|---|---|
| 2_170415626 | 0.191318757 | MYO3B | 11_46100278 | 0.323213692 | PHF21A | 6_151379906 | 0.256397553 | ZBTB2 |
| 22_32682749 | 0.191292011 | SYN3 | 3_18354833 | 0.322651017 | SATB1 | 16_78405202 | 0.256293019 | WWOX |
| 13_70110779 | 0.191262571 | ATXN8OS | 5_73798785 | 0.322403257 | ARHGEF28 | 8_72838695 | 0.256040229 | KCNB2 |
| 9_133156163 | 0.191236895 | GBGT1 | 12_6806160 | 0.322192905 | CD4 | 8_72843863 | 0.256040229 | KCNB2 |
| 13_110311284 | 0.191212177 | COL4A2 | 17_75778760 | 0.321969879 | H3F3B | 6_151384199 | 0.25601097 | ZBTB2 |
| 2_71214017 | 0.191212177 | PAIP2B | 1_35541010 | 0.321710296 | KIAA0319L | 12_20849727 | 0.255791005 | SLCO1B3 |
| 2_71420333 | 0.191124741 | ZNF638 | 4_188096963 | 0.321310476 | TRIML2 | 2_70047748 | 0.25577896 | PCBP1-AS1 |
| 2_71420334 | 0.191124741 | ZNF638 | 18_23730376 | 0.321289614 | LAMA3 | 12_111855820 | 0.255188826 | MAPKAPK5 |
| 6_159239763 | 0.191112814 | FNDC1 | 2_168171859 | 0.321241872 | STK39 | 7_71133949 | 0.255167488 | WBSCR17 |
| 1_231208537 | 0.191014952 | TRIM67 | 3_124236706 | 0.321107756 | KALRN | 17_10138661 | 0.255167004 | GAS7 |
| 1_96371711 | 0.190958309 | LOC101928241 | 16_61736075 | 0.320697983 | CDH8 | 7_102312998 | 0.255145749 | SH2B2 |
| 17_19240301 | 0.190944418 | EPN2 | 6_150769604 | 0.320622137 | PLEKHG1 | 6_151407337 | 0.255021382 | RMND1 |
| 2_206738750 | 0.190858618 | MDH1B | 9_17202256 | 0.320146003 | CNTLN | 10_96254509 | 0.255019634 | BLNK |
| 2_46322394 | 0.190843701 | EPAS1 | 10_80611196 | 0.320131518 | SH2D4B | 14_66896826 | 0.254833754 | GPHN |
| 7_44695178 | 0.190746486 | OGDH | 8_71329286 | 0.320114795 | EYA1 | 2_70067128 | 0.254735532 | PCBP1-AS1 |
| 5_36292944 | 0.190617273 | RANBP3L | 2_147908051 | 0.320053994 | ACVR2A | 1_244599676 | 0.253789643 | C1orf101 |
| 2_46323630 | 0.190593898 | EPAS1 | 11_46072204 | 0.31949115 | PHF21A | 2_231314675 | 0.253770149 | ARMC9 |
| 2_71361243 | 0.19057312 | ZNF638 | 17_19670564 | 0.318884228 | ALDH3A2 | 15_52241751 | 0.253471886 | MYO5C |
| 7_90261233 | 0.190429963 | C7orf63 | 13_18212633 | 0.318582418 | FAM230C | 6_151406447 | 0.253410834 | RMND1 |
| 7_90261308 | 0.190429963 | C7orf63 | 12_109728420 | 0.318430577 | FAM222A | 3_138447095 | 0.253315817 | ESYT3 |
| 10_66885462 | 0.190306923 | CTNNA3 | 8_48016782 | 0.317873533 | UBE2V2 | 17_19417131 | 0.253196742 | RNF112 |
| 15_90061888 | 0.190300443 | ZNF710 | 1_35508868 | 0.317851103 | KIAA0319L | 6_70297562 | 0.253028831 | COL9A1 |
| 22_32725344 | 0.190258395 | SYN3 | 1_35517288 | 0.317851103 | KIAA0319L | 10_96244941 | 0.252929102 | BLNK |
| 13_28259135 | 0.190066344 | PAN3 | 1_35552461 | 0.317851103 | KIAA0319L | 1_167833748 | 0.252866602 | ADCY10 |
| 7_104898543 | 0.189990869 | LHFPL3 | 2_241945926 | 0.317384624 | LINC01237 | 12_112206642 | 0.252602231 | HECTD4 |
| 7_104898635 | 0.189990869 | LHFPL3 | 16_25917093 | 0.317267853 | HS3ST4 | 4_188094103 | 0.252581217 | TRIML2 |
| 13_70100480 | 0.189921041 | KLHL1 | 1_236020048 | 0.317063715 | NID1 | 6_32444069 | 0.252531942 | HLA-DRA |
| 1_47764474 | 0.189835588 | TRABD2B | 17_75809231 | 0.316880496 | UNK | 2_214143245 | 0.252513113 | SPAG16 |
| 2_71069125 | 0.189704251 | NAGK | 3_42217098 | 0.316708293 | TRAK1 | 9_128172172 | 0.252489741 | CIZ1 |
| 8_10404886 | 0.189693457 | MSRA | 2_71203196 | 0.316647645 | PAIP2B | 3_138590024 | 0.252250108 | CEP70 |
| 12_32895809 | 0.189407906 | PKP2 | 2_71203488 | 0.316647645 | PAIP2B | 7_28173881 | 0.252159573 | JAZF1 |
| 19_37682079 | 0.1893936 | ZNF781 | 2_112000007 | 0.316183565 | MERTK | 1_244560427 | 0.252105879 | C1orf101 |
| 12_95676719 | 0.189348599 | NTN4 | 15_48643656 | 0.316108102 | FBN1 | 11_118561564 | 0.251980735 | IFT46 |
| 15_47315067 | 0.189244626 | SEMA6D | 15_48643661 | 0.316108102 | FBN1 | 1_54733246 | 0.251861825 | MROH7-TTC4 |
| 6_166777097 | 0.189134006 | RPS6KA2 | 19_33108235 | 0.316068341 | GPATCH1 | 11_118599377 | 0.251613471 | ARCN1 |
| 6_166787853 | 0.189134006 | RPS6KA2 | 1_169547269 | 0.316062818 | F5 | 2_55690670 | 0.251402976 | PNPT1 |
| 2_206715426 | 0.189003882 | DYTN | 1_35479285 | 0.316016494 | KIAA0319L | 3_138574449 | 0.251297153 | CEP70 |
| 1_10766562 | 0.188986707 | CASZ1 | 1_35507530 | 0.316016494 | KIAA0319L | 3_138449679 | 0.251290131 | ESYT3 |
| 7_78024793 | 0.188975489 | MAGI2 | 1_35509913 | 0.316016494 | KIAA0319L | 7_40836481 | 0.251233457 | SUGCT |
| 22_42518399 | 0.188933768 | RRP7A | 4_188098634 | 0.31595757 | TRIML2 | 7_40836556 | 0.251233457 | SUGCT |
| 8_72871802 | 0.188845011 | KCNB2 | 9_7122259 | 0.315132572 | KDM4C | 14_67296538 | 0.251206615 | MPP5 |
| 5_31970137 | 0.188762347 | PDZD2 | 1_35523257 | 0.31509228 | KIAA0319L | 11_118581186 | 0.251147011 | ARCN1 |
| 1_231191443 | 0.188700566 | TRIM67 | 16_61734105 | 0.314986759 | CDH8 | 2_70062074 | 0.250953844 | PCBP1-AS1 |
| 1_231201200 | 0.188700566 | TRIM67 | 4_108744043 | 0.314830378 | ETNPPL | 11_118583462 | 0.250897701 | ARCN1 |
| 3_33120406 | 0.188683965 | CRTAP | 4_108749064 | 0.314830378 | ETNPPL | 14_67296416 | 0.25083838 | MPP5 |
| 6_128496437 | 0.188640492 | PTPRK | 4_108751038 | 0.314830378 | ETNPPL | 13_92838340 | 0.250524427 | GPC5 |
| 15_90042627 | 0.188422422 | ZNF710 | 16_61734272 | 0.314367273 | CDH8 | 6_30701985 | 0.250511029 | MDC1 |
| 7_93489203 | 0.188405928 | CALCR | 1_35491196 | 0.314221683 | KIAA0319L | 11_118576865 | 0.25041961 | ARCN1 |
| 7_93516599 | 0.188405928 | CALCR | 4_108743359 | 0.313857776 | ETNPPL | 11_118573229 | 0.250394876 | ARCN1 |
| 13_28289061 | 0.188382884 | PAN3 | 1_35519403 | 0.313825858 | KIAA0319L | 4_54682751 | 0.250350087 | KIT |
| 6_159234370 | 0.188301971 | FNDC1 | 6_166461380 | 0.313556906 | RPS6KA2 | 1_167124776 | 0.250312246 | DUSP27 |
| 15_90055756 | 0.188124732 | ZNF710 | 13_85514170 | 0.313013381 | LINC00351 | 7_40844935 | 0.250226149 | SUGCT |
| 15_47768253 | 0.188106408 | SEMA6D | 14_61544081 | 0.312673297 | PRKCH | 7_40847159 | 0.250226149 | SUGCT |
| 2_71398024 | 0.18805442 | ZNF638 | 5_8370386 | 0.312420273 | LOC729506 | 3_138494450 | 0.250035454 | CEP70 |
| 2_71404910 | 0.18805442 | ZNF638 | 9_3513729 | 0.311923058 | RFX3 | 3_138513563 | 0.250035454 | CEP70 |
| 8_138299003 | 0.187771227 | FAM135B | 1_201494753 | 0.311796946 | CSRP1 | 3_138543972 | 0.250035454 | CEP70 |
| 6_159243290 | 0.18768325 | FNDC1 | 16_61694881 | 0.311482127 | CDH8 | 2_233923743 | 0.249865503 | TRPM8 |
| 1_65395872 | 0.187667743 | DNAJC6 | 15_48642693 | 0.311417666 | FBN1 | 6_32444794 | 0.249844383 | HLA-DRA |
| 12_54950144 | 0.187586774 | TESPA1 | 17_75777230 | 0.310850307 | H3F3B | 19_1766486 | 0.249638052 | ONECUT3 |
| 5_106930109 | 0.187483992 | LOC102467213 | 17_75777231 | 0.310850307 | H3F3B | 8_72818053 | 0.249265387 | KCNB2 |
| 1_44552820 | 0.187405661 | RNF220 | 17_75795734 | 0.310850307 | UNK | 2_231296968 | 0.24921236 | ARMC9 |
| 15_90043119 | 0.187317765 | ZNF710 | 15_48645781 | 0.31051136 | FBN1 | 6_21934712 | 0.249175762 | CASC15 |
| 13_70122385 | 0.187229935 | ATXN8OS | 16_61678849 | 0.310136158 | CDH8 | 3_2525147 | 0.249140783 | CNTN4 |
| 9_133161872 | 0.18704607 | GBGT1 | 16_61714724 | 0.310136158 | CDH8 | 10_96319623 | 0.249025976 | DNTT |
| 6_21789910 | 0.187028236 | CASC15 | 5_73789841 | 0.309886134 | ARHGEF28 | 4_88060685 | 0.248809712 | PKD2 |
| 1_231217716 | 0.18699672 | TRIM67 | 7_28173881 | 0.309640869 | JAZF1 | 11_118447192 | 0.248739611 | KMT2A |
| 1_231191920 | 0.186990822 | TRIM67 | 2_170384356 | 0.309075272 | MYO3B | 6_30908257 | 0.248661424 | GTF2H4 |
| 2_46370617 | 0.186938486 | EPAS1 | 4_188094103 | 0.308689741 | TRIML2 | 11_78975530 | 0.248385324 | TENM4 |
| 19_37955448 | 0.186849325 | SIPA1L3 | 16_7041099 | 0.30847946 | RBFOX1 | 3_32361296 | 0.248113811 | CMTM8 |
| 8_142752157 | 0.186814426 | LYPD2 | 11_113395277 | 0.308139807 | ANKK1 | 1_54733216 | 0.247989317 | MROH7-TTC4 |
| 12_12490415 | 0.186805193 | DUSP16 | 1_244414592 | 0.308139807 | ADSS | 2_69987605 | 0.247921778 | PCBP1-AS1 |
| 6_21762021 | 0.186704713 | CASC15 | 19_13336600 | 0.308066883 | CACNA1A | 3_138567888 | 0.247830242 | CEP70 |
| 11_101031322 | 0.186636255 | PGR | 17_60980533 | 0.307912684 | BCAS3 | 14_63391650 | 0.247476993 | PPP2R5E |
| 7_93502964 | 0.186576227 | CALCR | 7_100604429 | 0.307912684 | PCOLCE | 20_63467130 | 0.247476993 | KCNQ2 |
| 6_89097273 | 0.186525791 | SRSF12 | 12_94365770 | 0.307837291 | CEP83 | 10_96308624 | 0.247443073 | DNTT |
| 6_89097601 | 0.186525791 | SRSF12 | 4_20600414 | 0.307519417 | SLIT2 | 17_19413873 | 0.247144106 | RNF112 |
| 5_109774537 | 0.18645092 | MAN2A1 | 2_6995609 | 0.307480525 | RNF144A | 2_70081520 | 0.247102343 | PCBP1-AS1 |
| 6_47715998 | 0.186287041 | GPR115 | 1_35479841 | 0.307235771 | KIAA0319L | 3_64608807 | 0.24686385 | ADAMTS9 |
| 2_162542357 | 0.186207558 | KCNH7 | 10_20829439 | 0.3071584 | NEBL | 3_64742740 | 0.24674744 | ADAMTS9-AS2 |
| 10_52065427 | 0.186193304 | PRKG1 | 4_188093698 | 0.30703858 | TRIML2 | 10_96317357 | 0.246683992 | DNTT |
| 10_52065428 | 0.186193304 | PRKG1 | 3_124234296 | 0.306617287 | KALRN | 19_56096249 | 0.246438324 | ZNF787 |
| 3_111798201 | 0.186181163 | PHLDB2 | 1_231217716 | 0.306587177 | TRIM67 | 3_138517455 | 0.246217249 | CEP70 |
| 7_93505758 | 0.186132661 | CALCR | 14_61537581 | 0.306425492 | PRKCH | 1_244584540 | 0.246195989 | C1orf101 |
| 4_89750927 | 0.186122069 | SNCA | 7_111111300 | 0.305906131 | IMMP2L | 1_244584555 | 0.246195989 | C1orf101 |
| 3_26639482 | 0.186090714 | LRRC3B | 16_61735892 | 0.30577531 | CDH8 | 1_53439313 | 0.246153404 | SLC25A3P1 |
| 14_104801891 | 0.185951408 | ZBTB42 | 2_71225275 | 0.30545577 | PAIP2B | 3_138547754 | 0.245969261 | CEP70 |
| 2_30469577 | 0.185783202 | LCLAT1 | 6_6192777 | 0.30503385 | F13A1 | 3_138556985 | 0.245889966 | CEP70 |
| 2_30469578 | 0.185783202 | LCLAT1 | 17_65105333 | 0.304845023 | LOC100507002 | 1_1285758 | 0.24574543 | SCNN1D |
| 6_144594199 | 0.185550736 | UTRN | 10_25496259 | 0.30479239 | GPR158 | 10_113108403 | 0.245170784 | TCF7L2 |
| 17_5916877 | 0.185502879 | LOC339166 | 1_35073482 | 0.304344277 | ZMYM1 | 10_20894585 | 0.245060549 | NEBL |
| 4_89766356 | 0.185463171 | SNCA | 1_35518233 | 0.304042497 | KIAA0319L | 6_5654245 | 0.245042165 | FARS2 |
| 5_109707528 | 0.185377523 | MAN2A1 | 1_35542317 | 0.304042497 | KIAA0319L | 17_19266488 | 0.245010375 | EPN2 |
| 5_109730050 | 0.185377523 | MAN2A1 | 4_184193687 | 0.303980073 | ENPP6 | 8_72855743 | 0.244964322 | KCNB2 |
| 5_109730081 | 0.185377523 | MAN2A1 | 17_75804968 | 0.303875097 | UNK | 12_111843698 | 0.244706008 | MAPKAPK5 |
| 15_90050287 | 0.185337505 | ZNF710 | 17_75812699 | 0.303875097 | UNK | 13_22258033 | 0.244663149 | LINC00540 |
| 15_90050691 | 0.185337505 | ZNF710 | 3_155593186 | 0.303875097 | PLCH1 | 2_55590162 | 0.244461496 | SMEK2 |
| 5_106850583 | 0.185193506 | LOC102467213 | 1_35903423 | 0.303807189 | AGO1 | 3_138458092 | 0.244407496 | ESYT3 |

| | | | | | | | | |
|---|---|---|---|---|---|---|---|---|
| 5_13696374 | 0.185017227 | DNAH5 | 6_142210474 | 0.303665198 | VTA1 | 19_56615673 | 0.244357387 | ZNF71 |
| 12_81394672 | 0.184976503 | PPFIA2 | 3_119861520 | 0.303449084 | GSK3B | 10_45487034 | 0.244252309 | 8-Mar |
| 2_206713524 | 0.184958746 | DYTN | 17_80025005 | 0.303419823 | TBC1D16 | 14_96076972 | 0.244082092 | C14orf132 |
| 2_206713533 | 0.184958746 | DYTN | 19_954701 | 0.303414176 | ARID3A | 2_70040129 | 0.243809406 | PCBP1-AS1 |
| 2_206716366 | 0.184958746 | DYTN | 19_16034729 | 0.303132884 | LINC00905 | 1_44597949 | 0.243199203 | RNF220 |
| 20_33761644 | 0.184944475 | ZNF341 | 10_76434345 | 0.302951518 | C10orf11 | 14_67296424 | 0.243013587 | MPP5 |
| 7_88096502 | 0.184943904 | ADAM22 | 1_35488232 | 0.302944599 | KIAA0319L | 10_96261956 | 0.242898998 | BLNK |
| 1_36046566 | 0.184858874 | AGO3 | 4_108585036 | 0.302903139 | RPL34-AS1 | 1_33759319 | 0.242882471 | CSMD2 |
| 7_78027650 | 0.184846849 | MAGI2 | 11_46086807 | 0.302901036 | PHF21A | 8_72817778 | 0.242815838 | KCNB2 |
| 2_167026228 | 0.184639821 | XIRP2 | 11_46107551 | 0.302901036 | PHF21A | 3_64620491 | 0.242801814 | ADAMTS9 |
| 22_32697412 | 0.184550885 | SYN3 | 11_46109587 | 0.302901036 | PHF21A | 2_55604533 | 0.242657915 | SMEK2 |
| 15_83574938 | 0.184424279 | SH3GL3 | 1_155691879 | 0.302760565 | DAP3 | 12_7421023 | 0.242576018 | CD163L1 |
| 8_10411473 | 0.18440913 | MSRA | 2_37123341 | 0.302716387 | EIF2AK2 | 3_64615699 | 0.242554225 | ADAMTS9 |
| 6_159237694 | 0.184361408 | FNDC1 | 9_132348370 | 0.302550784 | SETX | 11_118574759 | 0.242476354 | ARCN1 |
| 6_159241606 | 0.184361408 | FNDC1 | 9_132348371 | 0.302550784 | SETX | 11_118580160 | 0.242476354 | ARCN1 |
| 7_149297150 | 0.184247497 | LOC155060 | 2_71203588 | 0.302539362 | PAIP2B | 11_118576882 | 0.242459037 | ARCN1 |
| 7_103433517 | 0.184201977 | SLC26A5 | 1_61814549 | 0.302391993 | INADL | 18_13533794 | 0.24242983 | LDLRAD4 |
| 5_109750314 | 0.184144984 | MAN2A1 | 22_46692823 | 0.302222976 | CERK | 6_29792759 | 0.242378134 | HCG4 |
| 2_71073080 | 0.184134032 | NAGK | 2_71194076 | 0.302086539 | PAIP2B | 2_70027119 | 0.242131889 | PCBP1-AS1 |
| 1_11610902 | 0.184034386 | LOC101929181 | 2_26459778 | 0.302024759 | OTOF | 11_118534662 | 0.24213058 | TMEM25 |
| 1_11612350 | 0.184034386 | LOC101929181 | 9_113897878 | 0.301932184 | ZNF618 | 13_92777394 | 0.242046968 | GPC5 |
| 2_2229436 | 0.183931385 | MYT1L | 17_75811577 | 0.301853606 | UNK | 4_158948605 | 0.241880083 | C4orf45 |
| 5_106853786 | 0.183834567 | LOC102467213 | 13_35445820 | 0.301764868 | NBEA | 9_129937601 | 0.24186886 | FNBP1 |
| 16_10789856 | 0.183831325 | TVP23A | 9_132348366 | 0.301572115 | SETX | 17_65197676 | 0.241850022 | RGS9 |
| 22_42512283 | 0.183789485 | RRP7A | 9_132348368 | 0.301572115 | SETX | 3_138522007 | 0.241740043 | CEP70 |
| 1_162863439 | 0.183758105 | C1orf110 | 2_30469579 | 0.301517054 | LCLAT1 | 5_37208680 | 0.241528456 | C5orf42 |
| 1_162863810 | 0.183758105 | C1orf110 | 7_149246898 | 0.301453709 | ZNF212 | 16_78360721 | 0.24139131 | WWOX |
| 16_89862446 | 0.183758105 | SPIRE2 | 15_32934599 | 0.301341129 | FMN1 | 8_73320482 | 0.241268122 | RDH10 |
| 22_25762943 | 0.183740608 | MYO18B | 7_39976327 | 0.301157689 | CDK13 | 5_3466359 | 0.241219394 | LINC01019 |
| 6_159231899 | 0.183684788 | FNDC1 | 1_201496929 | 0.300885706 | CSRP1 | 3_33051351 | 0.241207864 | GLB1 |
| 6_6179018 | 0.183637828 | F13A1 | 1_36280931 | 0.300801097 | THRAP3 | 3_138472606 | 0.24120318 | ESYT3 |
| 12_16026371 | 0.183634888 | DERA | 17_75835577 | 0.300718719 | UNC13D | 2_70082845 | 0.24113964 | PCBP1-AS1 |
| 19_37956184 | 0.183620343 | SIPA1L3 | 4_4220394 | 0.300534278 | OTOP1 | 11_118563014 | 0.241092438 | IFT46 |
| 12_12509348 | 0.183570988 | DUSP16 | 15_27046275 | 0.300474518 | GABRG3 | 3_138460943 | 0.240874115 | ESYT3 |
| 7_104899947 | 0.183428855 | LHFPL3 | 1_242396670 | 0.300391618 | PLD5 | 2_55616969 | 0.240828325 | SMEK2 |
| 11_18267994 | 0.183368414 | SAA1 | 13_71840152 | 0.300345953 | DACH1 | 19_49907320 | 0.240231366 | IL4I1 |
| 7_104880408 | 0.183248349 | LHFPL3 | 4_108752684 | 0.300145712 | ETNPPL | 2_69998265 | 0.240166557 | PCBP1-AS1 |
| 15_66153122 | 0.183004651 | MEGF11 | 4_93578593 | 0.299995078 | GRID2 | 6_29830306 | 0.240115983 | HLA-G |
| 1_231192042 | 0.182991577 | TRIM67 | 8_25226259 | 0.29974739 | DOCK5 | 6_29943463 | 0.240087824 | HLA-A |
| 19_37960341 | 0.182988978 | SIPA1L3 | 12_377223 | 0.299326573 | KDM5A | 3_138560708 | 0.240085525 | CEP70 |
| 22_32602033 | 0.182934265 | SYN3 | 3_55873773 | 0.29929353 | ERC2 | 1_35456789 | 0.240001963 | KIAA0319L |
| 16_10786220 | 0.182875037 | TVP23A | 1_155431219 | 0.299282153 | ASH1L | 13_45840194 | 0.239824335 | SIAH3 |
| 7_90251027 | 0.182875037 | C7orf63 | 9_113891446 | 0.299174131 | ZNF618 | 10_96251151 | 0.239742261 | BLNK |
| 7_90256135 | 0.182875037 | C7orf63 | 16_61676684 | 0.29908034 | CDH8 | 6_128238156 | 0.239684222 | PTPRK |
| 8_10407730 | 0.182848576 | MSRA | 7_6126249 | 0.298860446 | USP42 | 8_72829538 | 0.239601106 | KCNB2 |
| 7_103431823 | 0.182826086 | SLC26A5 | 4_89788085 | 0.298857926 | SNCA | 7_95902503 | 0.239545174 | DYNC1I1 |
| 11_113395277 | 0.18265908 | ANKK1 | 10_124833160 | 0.298600102 | FAM175B | 3_64609510 | 0.239472542 | ADAMTS9 |
| 2_71371894 | 0.182609784 | ZNF638 | 11_46051562 | 0.298549929 | PHF21A | 12_120890479 | 0.239329833 | SPPL3 |
| 13_28287004 | 0.182555224 | PAN3 | 1_201498974 | 0.297937367 | CSRP1 | 1_121347210 | 0.23930978 | SRGAP2C |
| 13_28259006 | 0.182484602 | PAN3 | 16_23684974 | 0.297583087 | PLK1 | 10_96311092 | 0.239095602 | DNTT |
| 6_54007004 | 0.182477517 | MLIP-IT1 | 17_75835819 | 0.297551366 | UNC13D | 2_70086436 | 0.239045966 | PCBP1-AS1 |
| 1_96372020 | 0.18247382 | LOC101928241 | 7_139868653 | 0.297505331 | TBXAS1 | 2_69964228 | 0.239014355 | PCBP1-AS1 |
| 13_28203812 | 0.182435991 | PAN3 | 3_1384794 | 0.29736454 | CNTN6 | 3_138541431 | 0.23888105 | CEP70 |
| 6_87801400 | 0.182409941 | LOC101928911 | 16_89160413 | 0.297090586 | LINC00304 | 2_55654681 | 0.238794712 | PNPT1 |
| 7_104854070 | 0.182398647 | LHFPL3 | 2_108752637 | 0.29694898 | RANBP2 | 4_6354044 | 0.238773931 | PPP2R2C |
| 1_10780848 | 0.182303779 | CASZ1 | 4_91297395 | 0.296581833 | CCSER1 | 3_64608927 | 0.238748597 | ADAMTS9 |
| 2_71071407 | 0.182245338 | NAGK | 1_145007803 | 0.296419012 | SRGAP2B | 8_72826305 | 0.238639333 | KCNB2 |
| 19_36753577 | 0.182162274 | ZNF850 | 9_17208123 | 0.296340477 | CNTLN | 19_38335883 | 0.238538781 | CATSPERG |
| 22_32707932 | 0.182151228 | SYN3 | 9_17210475 | 0.296340477 | CNTLN | 17_5554396 | 0.238515148 | NLRP1 |
| 2_170415310 | 0.182123858 | MYO3B | 1_35521225 | 0.296276266 | KIAA0319L | 14_24473099 | 0.238419188 | LOC101927045 |
| 4_143591751 | 0.182114871 | FREM3 | 12_109726650 | 0.296230952 | FAM222A | 1_1302158 | 0.238255292 | ACAP3 |
| 2_30963660 | 0.182093131 | GALNT14 | 12_293599 | 0.29622338 | KDM5A | 12_112085324 | 0.238255292 | NAA25 |
| 8_29344533 | 0.181985489 | DUSP4 | 6_100409689 | 0.296106481 | SIM1 | 2_55666201 | 0.23819245 | PNPT1 |
| 12_12480353 | 0.181982095 | DUSP16 | 3_64800770 | 0.296004665 | ADAMTS9-AS2 | 1_245913376 | 0.238104037 | SMYD3 |
| 1_210139618 | 0.18196176 | SYT14 | 4_6355996 | 0.295900025 | PPP2R2C | 13_94288675 | 0.238054181 | GPC6 |
| 13_70135657 | 0.181949257 | ATXN8OS | 16_61917032 | 0.29567331 | CDH8 | 5_10417463 | 0.238036548 | 6-Mar |
| 5_106874703 | 0.181925159 | LOC102467213 | 4_89778628 | 0.295664391 | SNCA | 12_111978185 | 0.237999121 | TMEM116 |
| 7_78029824 | 0.181924944 | MAGI2 | 15_48642803 | 0.295070935 | FBN1 | 5_178396827 | 0.237965732 | COL23A1 |
| 2_71403699 | 0.181807873 | ZNF638 | 12_109729785 | 0.295031037 | FAM222A | 10_96251962 | 0.237942968 | BLNK |
| 5_109716385 | 0.181732137 | MAN2A1 | 4_108744239 | 0.294494191 | ETNPPL | 10_121598047 | 0.237726592 | FGFR2 |
| 1_162864291 | 0.181525164 | C1orf110 | 14_61575830 | 0.294426756 | FLJ22447 | 13_22261433 | 0.237710948 | LINC00540 |
| 5_106878595 | 0.181477574 | LOC102467213 | 2_201833717 | 0.29436833 | CDK15 | 2_121518290 | 0.237667256 | CLASP1 |
| 8_10409462 | 0.181355675 | MSRA | 17_75786663 | 0.294113677 | UNK | 6_151457049 | 0.237595025 | RMND1 |
| 2_166933887 | 0.181260817 | XIRP2 | 2_170384153 | 0.293554392 | MYO3B | 2_70010340 | 0.237544335 | PCBP1-AS1 |
| 17_65165019 | 0.181220699 | RGS9 | 1_37495707 | 0.293515604 | MEAF6 | 10_96247570 | 0.23738922 | BLNK |
| 2_167001160 | 0.181169771 | XIRP2 | 16_70487291 | 0.293291989 | COG4 | 6_30908375 | 0.237201426 | GTF2H4 |
| 2_71378880 | 0.181120481 | ZNF638 | 10_89757059 | 0.292856615 | KIF20B | 3_138466180 | 0.236973017 | ESYT3 |
| 4_89750336 | 0.181070614 | SNCA | 10_89757061 | 0.292856615 | KIF20B | 3_138523671 | 0.236973017 | CEP70 |
| 1_169525165 | 0.181038137 | F5 | 10_89757062 | 0.292856615 | KIF20B | 3_138574168 | 0.236973017 | CEP70 |
| 21_34518111 | 0.181017417 | RCAN1 | 16_89586896 | 0.292856615 | CPNE7 | 15_28738260 | 0.236884928 | WHAMMP2 |
| 21_34525470 | 0.181017417 | RCAN1 | 17_75803438 | 0.292811594 | UNK | 1_245574867 | 0.236864643 | KIF26B |
| 2_234971228 | 0.181006762 | SH3BP4 | 19_16034723 | 0.29269016 | LINC00905 | 1_245574790 | 0.236864643 | KIF26B |
| 1_36037615 | 0.180973582 | AGO3 | 6_166474260 | 0.292325108 | RPS6KA2 | 8_72846791 | 0.236784928 | KCNB2 |
| 6_166840918 | 0.180896205 | RPS6KA2 | 15_48642431 | 0.292202617 | FBN1 | 12_112140186 | 0.236753082 | TRAFD1 |
| 7_104905852 | 0.180853123 | LHFPL3 | 6_43483455 | 0.292198703 | TJAP1 | 6_99454596 | 0.236721539 | USP45 |
| 8_29350625 | 0.180793207 | DUSP4 | 9_70898763 | 0.292047848 | TRPM3 | 2_231284480 | 0.236568523 | ARMC9 |
| 11_101030247 | 0.180759965 | PGR | 18_63755129 | 0.292032558 | SERPINB7 | 2_69968424 | 0.236530227 | PCBP1-AS1 |
| 1_62156231 | 0.180707148 | INADL | 16_89586409 | 0.291977577 | CPNE7 | 2_69969186 | 0.236530227 | PCBP1-AS1 |
| 4_89756092 | 0.180606452 | SNCA | 7_147236268 | 0.291894332 | CNTNAP2 | 2_231320259 | 0.236472181 | ARMC9 |
| 5_106861209 | 0.180535673 | LOC102467213 | 7_6264515 | 0.291605953 | CYTH3 | 8_72852079 | 0.236328132 | KCNB2 |
| 6_21801373 | 0.180502641 | CASC15 | 9_113910894 | 0.291047522 | ZNF618 | 12_112281206 | 0.236164533 | HECTD4 |
| 1_231208843 | 0.180459201 | TRIM67 | 15_27019873 | 0.290782388 | GABRG3 | 6_32639979 | 0.236161923 | HLA-DQA1 |
| 12_12522994 | 0.18041162 | DUSP16 | 3_48993860 | 0.290667483 | P4HTM | 3_138512421 | 0.236112723 | CEP70 |
| 4_6457888 | 0.180264997 | PPP2R2C | 17_12694882 | 0.290626709 | LOC101928418 | 3_138572727 | 0.236112723 | CEP70 |
| 4_6462547 | 0.180264997 | PPP2R2C | 7_129940895 | 0.290623479 | UBE2H | 3_138579235 | 0.236112723 | CEP70 |
| 4_6462676 | 0.180264997 | PPP2R2C | 12_128117447 | 0.29034128 | LOC100996679 | 2_148747363 | 0.236098134 | EPC2 |
| 12_48347436 | 0.180255743 | ZNF641 | 1_171096951 | 0.290234396 | FMO3 | 10_45574634 | 0.236080367 | 8-Mar |

| | | | | | | | | |
|---|---|---|---|---|---|---|---|---|
| 12_48348706 | 0.180255743 | ZNF641 | 17_76679918 | 0.290195724 | MXRA7 | 6_128486781 | 0.236010873 | PTPRK |
| 4_56243063 | 0.180163642 | KIAA1211 | 17_76702942 | 0.290115249 | MXRA7 | 1_244595310 | 0.235921896 | C1orf101 |
| 4_56243287 | 0.180163642 | KIAA1211 | 1_36046566 | 0.290111125 | AGO3 | 10_96258821 | 0.235883683 | BLNK |
| 1_62123547 | 0.180142485 | INADL | 6_34403930 | 0.289844587 | RPS10-NUDT3 | 10_96261764 | 0.235883683 | BLNK |
| 1_53113730 | 0.180115884 | SLC1A7 | 4_188104147 | 0.289624237 | TRIML2 | 1_1286337 | 0.235808562 | SCNN1D |
| 1_62114235 | 0.180066046 | INADL | 19_45951638 | 0.289597934 | NOVA2 | 12_95720251 | 0.235734592 | NTN4 |
| 1_62121980 | 0.180066046 | INADL | 7_90063136 | 0.289565591 | STEAP2-AS1 | 6_30908461 | 0.235585126 | GTF2H4 |
| 15_40268978 | 0.180026966 | PAK6 | 5_73795324 | 0.288942356 | ARHGEF28 | 2_70020129 | 0.235490877 | PCBP1-AS1 |
| 17_19319911 | 0.180020907 | EPN2 | 4_89769681 | 0.288859754 | SNCA | 2_55604279 | 0.235422463 | SMEK2 |
| 22_40491081 | 0.180005734 | MKL1 | 13_94312794 | 0.288789381 | GPC6 | 2_70073066 | 0.23516785 | PCBP1-AS1 |
| 12_127893379 | 0.179950651 | FLJ37505 | 6_6191288 | 0.288742945 | F13A1 | 2_229142559 | 0.234948029 | PID1 |
| 7_104830765 | 0.179860127 | LHFPL3 | 2_71201818 | 0.288605582 | PAIP2B | 18_13535083 | 0.23481192 | LDLRAD4 |
| 7_104835120 | 0.179860127 | LHFPL3 | 2_71214017 | 0.288605582 | PAIP2B | 14_63389042 | 0.234727872 | PPP2R5E |
| 7_104839685 | 0.179860127 | LHFPL3 | 9_113930403 | 0.288319663 | ZNF618 | 4_39507984 | 0.234718999 | UGDH |
| 8_10397721 | 0.179800398 | MSRA | 17_68105056 | 0.288318664 | LINC00674 | 4_85541089 | 0.234710843 | ARHGAP24 |
| 3_133619123 | 0.179716605 | TOPBP1 | 11_46107488 | 0.288317755 | PHF21A | 10_86507402 | 0.234697688 | WAPAL |
| 3_129433387 | 0.179681591 | MBD4 | 1_156066766 | 0.288094336 | RAB25 | 10_96246589 | 0.234693024 | BLNK |
| 2_71402343 | 0.179624497 | ZNF638 | 16_61664781 | 0.288054044 | CDH8 | 10_96246590 | 0.234693024 | BLNK |
| 4_89733513 | 0.179492129 | SNCA | 5_159077746 | 0.287987885 | EBF1 | 2_231326057 | 0.234368694 | ARMC9 |
| 7_104898289 | 0.179278483 | LHFPL3 | 18_26027701 | 0.287859565 | SS18 | 15_55343361 | 0.234356211 | PIGB |
| 4_172878759 | 0.179244318 | GALNTL6 | 7_6185616 | 0.287813376 | CYTH3 | 8_72846803 | 0.234313364 | KCNB2 |
| 17_65154986 | 0.179239268 | RGS9 | 1_35905818 | 0.287509404 | AGO1 | 11_78978008 | 0.234122754 | TENM4 |
| 18_39399545 | 0.17909579 | LINC00669 | 15_48625851 | 0.287358581 | FBN1 | 2_70041320 | 0.234016854 | PCBP1-AS1 |
| 1_11612444 | 0.178852776 | LOC101929181 | 15_48628146 | 0.287182497 | FBN1 | 2_192135621 | 0.233957595 | TMEFF2 |
| 1_11612552 | 0.178852776 | LOC101929181 | 16_6655428 | 0.287100856 | RBFOX1 | 6_32664947 | 0.233859549 | HLA-DQB1 |
| 1_62202575 | 0.17881096 | L1TD1 | 16_79180537 | 0.287082753 | WWOX | 8_72850662 | 0.233850544 | KCNB2 |
| 10_75854744 | 0.178802563 | C10orf11 | 20_60321544 | 0.28689907 | LOC284757 | 3_138589337 | 0.233724592 | CEP70 |
| 3_33119942 | 0.178782422 | CRTAP | 9_129996068 | 0.28685165 | FNBP1 | 2_55596169 | 0.233685541 | SMEK2 |
| 1_231205823 | 0.178646239 | TRIM67 | 17_75803334 | 0.286679568 | UNK | 2_55681293 | 0.233685541 | PNPT1 |
| 6_159226897 | 0.178576154 | FNDC1 | 11_46038373 | 0.286599288 | PHF21A | 2_69969365 | 0.233674371 | PCBP1-AS1 |
| 2_71148502 | 0.178535246 | MPHOSPH10 | 2_112009145 | 0.286452178 | MERTK | 2_50001450 | 0.233588574 | NRXN1 |
| 2_71148508 | 0.178535246 | MPHOSPH10 | 4_89765968 | 0.28616272 | SNCA | 3_138441644 | 0.233573397 | ESYT3 |
| 5_106828757 | 0.178531848 | LOC102467213 | 20_50875613 | 0.286045267 | BCAS4 | 1_45030996 | 0.233435922 | ZSWIM5 |
| 2_206714302 | 0.178266346 | DYTN | 7_71355470 | 0.286034869 | WBSCR17 | 15_47315026 | 0.233416397 | SEMA6D |
| 6_47711889 | 0.178266346 | GPR115 | 1_35505407 | 0.285970453 | KIAA0319L | 13_22261261 | 0.23323076 | LINC00540 |
| 7_154870989 | 0.178248154 | DPP6 | 1_35542512 | 0.285970453 | KIAA0319L | 3_31924230 | 0.233053295 | OSBPL10 |
| 10_86717093 | 0.178127763 | LDB3 | 6_125172052 | 0.285904446 | TPD52L1 | 20_63436421 | 0.232965834 | KCNQ2 |
| 2_71348283 | 0.178092447 | ZNF638 | 17_75777074 | 0.28585688 | H3F3B | 3_139120800 | 0.232948041 | BPESC1 |
| 5_109701337 | 0.178050558 | MAN2A1 | 19_5869279 | 0.285811768 | FUT5 | 16_78366944 | 0.232927634 | WWOX |
| 5_109701829 | 0.178050558 | MAN2A1 | 3_129424716 | 0.285618667 | EFCAB12 | 12_111878012 | 0.232858384 | MAPKAPK5 |
| 5_109704265 | 0.178050558 | MAN2A1 | 15_52240725 | 0.285419189 | MYO5C | 11_118574560 | 0.232818671 | ARCN1 |
| 5_109707287 | 0.178050558 | MAN2A1 | 10_89757067 | 0.285353472 | KIF20B | 11_118588324 | 0.232818671 | ARCN1 |
| 6_159230271 | 0.17804162 | FNDC1 | 1_35189989 | 0.285343152 | SFPQ | 12_121538271 | 0.232806142 | KDM2B |
| 19_34496637 | 0.177979731 | WTIP | 7_129907996 | 0.285326416 | UBE2H | 19_13409205 | 0.232736484 | CACNA1A |
| 16_89168776 | 0.177960283 | LOC400558 | 8_71323272 | 0.285255526 | EYA1 | 16_78366943 | 0.232584054 | WWOX |
| 17_9215878 | 0.177957333 | NTN1 | 1_35520325 | 0.284877425 | KIAA0319L | 6_32638265 | 0.232495522 | HLA-DQA1 |
| 11_101029985 | 0.177928825 | PGR | 1_169467440 | 0.284630693 | SLC19A2 | 7_98140280 | 0.232469927 | LMTK2 |
| 5_109710509 | 0.177838811 | MAN2A1 | 4_108758282 | 0.28449658 | ETNPPL | 2_55680328 | 0.232342372 | PNPT1 |
| 6_89084175 | 0.177836969 | PNRC1 | 4_108749156 | 0.284343878 | ETNPPL | 3_2556460 | 0.232259328 | CNTN4 |
| 6_89096169 | 0.177836969 | SRSF12 | 7_66696701 | 0.28431789 | RABGEF1 | 10_35259754 | 0.232116644 | CCNY |
| 6_144533593 | 0.177653248 | UTRN | 3_126632660 | 0.284267168 | TXNRD3 | 8_72828241 | 0.232039698 | KCNB2 |
| 1_231564220 | 0.17759148 | TSNAX | 11_46080970 | 0.284231846 | PHF21A | 6_151385435 | 0.232038325 | ZBTB2 |
| 11_44825032 | 0.177445221 | TSPAN18 | 15_52239744 | 0.284072443 | MYO5C | 5_10452556 | 0.232015296 | ROPN1L |
| 9_8879472 | 0.177295584 | PTPRD | 15_52244112 | 0.284072443 | MYO5C | 6_151405165 | 0.231982502 | RMND1 |
| 15_97751769 | 0.177218541 | LINC00923 | 22_40491070 | 0.283938674 | MKL1 | 11_78982776 | 0.23186098 | TENM4 |
| 1_162863595 | 0.177189522 | C1orf110 | 17_75796944 | 0.283729143 | UNK | 1_244572556 | 0.231827621 | C1orf101 |
| 7_93500363 | 0.177159179 | CALCR | 4_188099893 | 0.283454188 | TRIML2 | 12_27333162 | 0.231802158 | ARNTL2 |
| 1_161044859 | 0.177117074 | USF1 | 4_188099947 | 0.283454188 | TRIML2 | 2_64119188 | 0.231710584 | PELI1 |
| 9_8871438 | 0.177092991 | PTPRD | 9_113897863 | 0.283339185 | ZNF618 | 7_30671679 | 0.231563019 | CRHR2 |
| 4_31001014 | 0.177086643 | PCDH7 | 5_73798312 | 0.283070651 | ARHGEF28 | 3_138474981 | 0.231335167 | ESYT3 |
| 10_59703726 | 0.177082253 | SLC16A9 | 19_16034921 | 0.283011872 | LINC00905 | 1_244434624 | 0.231064156 | ADSS |
| 2_71203488 | 0.177070897 | PAIP2B | 10_60514979 | 0.282711406 | ANK3 | 9_20372609 | 0.231054064 | MLLT3 |
| 5_109723507 | 0.177020824 | MAN2A1 | 12_323379 | 0.282500986 | KDM5A | 1_1296817 | 0.23104151 | ACAP3 |
| 1_231203967 | 0.176917653 | TRIM67 | 3_192354989 | 0.282484653 | FGF12 | 2_55655356 | 0.23100207 | PNPT1 |
| 1_231206223 | 0.176917653 | TRIM67 | 2_1429286 | 0.282269303 | TPO | 9_20375578 | 0.230983452 | MLLT3 |
| 12_12520547 | 0.17688581 | DUSP16 | 9_113934438 | 0.282263751 | ZNF618 | 2_50021323 | 0.230905761 | NRXN1 |
| 8_29350328 | 0.17686835 | DUSP4 | 1_2262207 | 0.282024929 | SKI | 6_70279670 | 0.230722905 | COL9A1 |
| 5_106843335 | 0.176828289 | LOC102467213 | 6_161438216 | 0.2818753 | PARK2 | 3_138534773 | 0.230718665 | CEP70 |
| 5_168898316 | 0.17664722 | SLIT3 | 4_26610415 | 0.281828738 | TBC1D19 | 1_244590328 | 0.230673947 | C1orf101 |
| 19_37581427 | 0.176582397 | ZNF540 | 14_61559384 | 0.281689081 | LOC101927780 | 1_244590475 | 0.230673947 | C1orf101 |
| 19_37581429 | 0.176582397 | ZNF540 | 15_48478822 | 0.281526261 | FBN1 | 6_151385753 | 0.230627454 | ZBTB2 |
| 19_54125565 | 0.176496589 | PRPF31 | 13_101797737 | 0.281366918 | FGF14 | 2_50032596 | 0.230620802 | NRXN1 |
| 1_44551761 | 0.176456821 | RNF220 | 7_71318432 | 0.28067699 | WBSCR17 | 8_72834575 | 0.230554791 | KCNB2 |
| 6_169663558 | 0.176451265 | WDR27 | 6_129219744 | 0.280073317 | LAMA2 | 8_72838643 | 0.230554791 | KCNB2 |
| 6_159237427 | 0.176445825 | FNDC1 | 17_75807069 | 0.279549597 | UNK | 8_72846037 | 0.230554791 | KCNB2 |
| 12_33420493 | 0.176424799 | SYT10 | 17_75807277 | 0.279549597 | UNK | 15_83882079 | 0.230543355 | ADAMTSL3 |
| 16_89862193 | 0.176400263 | SPIRE2 | 17_75809532 | 0.279549597 | UNK | 16_29330537 | 0.230425278 | SNX29P2 |
| 8_10389215 | 0.176395794 | MSRA | 11_101032614 | 0.279544644 | PGR | 9_71018350 | 0.230413679 | TRPM3 |
| 15_97742686 | 0.176391418 | LINC00923 | 19_33120653 | 0.279478814 | GPATCH1 | 7_77376796 | 0.230394295 | GSAP |
| 5_106862141 | 0.176391285 | LOC102467213 | 15_48302576 | 0.279446866 | SLC12A1 | 2_55665243 | 0.230386811 | PNPT1 |
| 8_72872701 | 0.176291787 | KCNB2 | 4_89733003 | 0.279199693 | SNCA | 20_46728606 | 0.230147591 | SLC2A10 |
| 7_93500643 | 0.176083404 | CALCR | 17_82267060 | 0.279169719 | CSNK1D | 2_238172075 | 0.230120713 | ILKAP |
| 17_19238269 | 0.176078616 | EPN2 | 5_8370420 | 0.279158675 | LOC729506 | 2_55644150 | 0.229973791 | PNPT1 |
| 17_19239232 | 0.176078616 | EPN2 | 8_43004680 | 0.279156967 | HOOK3 | 11_20609018 | 0.229861221 | SLC6A5 |
| 17_19259739 | 0.176078616 | EPN2 | 19_42091095 | 0.279131922 | POU2F2 | 7_44625083 | 0.229838004 | OGDH |
| 17_19299719 | 0.176078616 | EPN2 | 10_25241413 | 0.279106247 | GPR158 | 6_70285992 | 0.229822522 | COL9A1 |
| 17_19339154 | 0.176078616 | B9D1 | 11_115404845 | 0.279106247 | CADM1 | 20_62744796 | 0.229800419 | NTSR1 |
| 6_159240638 | 0.176070786 | FNDC1 | 1_25886580 | 0.279106247 | STMN1 | 11_118577372 | 0.229735369 | ARCN1 |
| 1_64687657 | 0.17600548 | CACHD1 | 3_192246824 | 0.279079921 | FGF12 | 11_118540998 | 0.229721396 | TMEM25 |
| 19_37955841 | 0.175969684 | SIPA1L3 | 4_108752960 | 0.279062755 | ETNPPL | 17_5558194 | 0.229657464 | NLRP1 |
| 19_37971401 | 0.175969684 | SIPA1L3 | 2_188566796 | 0.279052921 | GULP1 | 21_37895213 | 0.229608782 | KCNJ6 |
| 7_71318432 | 0.175956446 | WBSCR17 | 12_27006141 | 0.279038555 | TM7SF3 | 15_47307854 | 0.229605349 | SEMA6D |
| 4_89746868 | 0.175902953 | SNCA | 3_124152553 | 0.278987006 | KALRN | 17_7487376 | 0.229604298 | POLR2A |
| 1_71895929 | 0.17580139 | NEGR1 | 13_98562918 | 0.278960239 | STK24 | 6_32663523 | 0.229588652 | HLA-DQB1 |
| 1_62118040 | 0.175720509 | INADL | 17_61246472 | 0.278960239 | BCAS3 | 11_12493408 | 0.229551713 | PARVA |
| 6_159234351 | 0.175714946 | FNDC1 | 13_107710811 | 0.27891961 | FAM155A | 17_18776277 | 0.229519107 | FBXW10 |
| 7_103433532 | 0.1756952 | SLC26A5 | 6_98943580 | 0.278904604 | FBXL4 | 1_1287315 | 0.229488101 | SCNN1D |

| | | | | | | | | |
|---|---|---|---|---|---|---|---|---|
| 7_103433546 | 0.1756952 | SLC26A5 | 3_55061691 | 0.278879277 | CACNA2D3 | 3_138541394 | 0.229461735 | CEP70 |
| 5_109737286 | 0.175643008 | MAN2A1 | 16_27248407 | 0.278780407 | NSMCE1 | 18_63718152 | 0.229273377 | SERPINB11 |
| 1_234076989 | 0.175602153 | SLC35F3 | 7_99296149 | 0.278765208 | MYH16 | 11_101917604 | 0.229262116 | KIAA1377 |
| 2_32635960 | 0.175575331 | TTC27 | 17_75798350 | 0.278436623 | UNK | 17_40633824 | 0.229170619 | SMARCE1 |
| 13_70100189 | 0.175525084 | KLHL1 | 1_155795017 | 0.278434339 | GON4L | 2_205583402 | 0.229168429 | PARD3B |
| 12_12537301 | 0.175503259 | DUSP16 | 1_52633845 | 0.278423531 | FAM159A | 5_178579184 | 0.229054682 | COL23A1 |
| 17_19373831 | 0.175496578 | B9D1 | 13_107287696 | 0.278345187 | FAM155A | 2_71373794 | 0.229016375 | ZNF638 |
| 5_109847640 | 0.175484412 | MAN2A1 | 1_231205823 | 0.278272357 | TRIM67 | 2_231143065 | 0.228885844 | PSMD1 |
| 12_81440665 | 0.175414934 | PPFIA2 | 1_116556256 | 0.278058429 | CD58 | 2_55676485 | 0.22885729 | PNPT1 |
| 1_231190090 | 0.175388335 | TRIM67 | 9_17348533 | 0.277981852 | CNTLN | 11_78979435 | 0.22874899 | TENM4 |
| 5_168889745 | 0.1752942 | SLIT3 | 15_100294595 | 0.277810624 | ADAMTS17 | 18_46716090 | 0.228746864 | ST8SIA5 |
| 6_144601329 | 0.175230475 | UTRN | 11_45914484 | 0.277782899 | PEX16 | 8_72820360 | 0.228705281 | KCNB2 |
| 5_106817792 | 0.175203351 | LOC102467213 | 16_7298292 | 0.277651456 | RBFOX1 | 8_72838292 | 0.228578195 | KCNB2 |
| 10_81929369 | 0.175109653 | NRG3 | 2_113925195 | 0.27664789 | ACTR3 | 2_55614885 | 0.22856149 | SMEK2 |
| 16_82811914 | 0.175019675 | CDH13 | 7_66839034 | 0.276609168 | GTF2IRD1P1 | 2_55692356 | 0.22856149 | PNPT1 |
| 12_131211486 | 0.174953657 | LINC01257 | 7_66839026 | 0.2765134 | GTF2IRD1P1 | 1_1285124 | 0.228559144 | SCNN1D |
| 2_170420012 | 0.174885019 | MYO3B | 15_48616960 | 0.276504375 | FBN1 | 6_32442799 | 0.22851231 | HLA-DRA |
| 1_74707573 | 0.174857219 | CRYZ | 15_48621189 | 0.276504375 | FBN1 | 10_96320475 | 0.228453149 | DNTT |
| 12_12536162 | 0.174822027 | DUSP16 | 21_33792857 | 0.276435206 | ITSN1 | 13_71545525 | 0.228172371 | DACH1 |
| 2_166939058 | 0.174789671 | XIRP2 | 14_61573757 | 0.276367715 | FLJ22447 | 7_90247013 | 0.22816322 | C7orf63 |
| 13_28279572 | 0.174689276 | PAN3 | 5_73796000 | 0.276275062 | ARHGEF28 | 11_118601639 | 0.228046139 | ARCN1 |
| 13_28293121 | 0.174689276 | PAN3 | 9_20411653 | 0.276083772 | MLLT3 | 2_55671794 | 0.228041723 | PNPT1 |
| 7_93475043 | 0.174679423 | CALCR | 14_105228992 | 0.275995807 | BRF1 | 3_2550243 | 0.228033616 | CNTN4 |
| 7_93497184 | 0.174679423 | CALCR | 18_52427835 | 0.275660659 | DCC | 7_40089741 | 0.228016522 | CDK13 |
| 7_93500374 | 0.174679423 | CALCR | 15_48478764 | 0.275576488 | FBN1 | 7_90263948 | 0.227953683 | C7orf63 |
| 7_93515271 | 0.174679423 | CALCR | 18_12112112 | 0.275565573 | ANKRD62 | 1_244609827 | 0.227939134 | C1orf101 |
| 15_83574660 | 0.174660477 | SH3GL3 | 1_35025503 | 0.275518429 | ZMYM6 | 1_244570133 | 0.227929631 | C1orf101 |
| 7_104879329 | 0.174641197 | LHFPL3 | 1_10746927 | 0.275483846 | CASZ1 | 6_70296672 | 0.227890716 | COL9A1 |
| 3_129528683 | 0.174637368 | RHO | 8_32723695 | 0.275320134 | NRG1 | 7_99057612 | 0.227880937 | SMURF1 |
| 4_6507511 | 0.174637368 | PPP2R2C | 13_71840493 | 0.275307732 | DACH1 | 2_64115821 | 0.227766729 | PELI1 |
| 16_89919746 | 0.174511414 | MC1R | 6_34382708 | 0.274699915 | NUDT3 | 1_1288005 | 0.227731024 | SCNN1D |
| 1_36044395 | 0.174491611 | AGO3 | 4_89766356 | 0.274662359 | SNCA | 2_64123729 | 0.227686922 | PELI1 |
| 10_81929136 | 0.174358352 | NRG3 | 2_71191767 | 0.274605033 | PAIP2B | 1_45121605 | 0.227449413 | ZSWIM5 |
| 5_106828510 | 0.174354413 | LOC102467213 | 3_125293409 | 0.274534584 | ZNF148 | 1_244578161 | 0.227441756 | C1orf101 |
| 7_93509128 | 0.174288012 | CALCR | 11_113333435 | 0.274243111 | TTC12 | 2_70036388 | 0.227407072 | PCBP1-AS1 |
| 1_44549778 | 0.174284228 | RNF220 | 9_17211721 | 0.274132127 | CNTLN | 3_138533909 | 0.227359481 | CEP70 |
| 1_155687151 | 0.174274898 | YY1AP1 | 3_124236006 | 0.27412283 | KALRN | 10_96259109 | 0.227358031 | BLNK |
| 12_49792286 | 0.174224227 | NCKAP5L | 4_188104320 | 0.273805346 | TRIML2 | 10_96262356 | 0.227358031 | BLNK |
| 12_49805868 | 0.174224227 | NCKAP5L | 12_383119 | 0.273537574 | KDM5A | 7_76418623 | 0.227352983 | ZP3 |
| 3_130461122 | 0.174117609 | COL6A5 | 15_52235996 | 0.2734495 | MYO5C | 7_76418627 | 0.227352983 | ZP3 |
| 1_231187947 | 0.174081152 | TRIM67 | 12_109721681 | 0.273339877 | FAM222A | 12_111676046 | 0.227343541 | BRAP |
| 1_231189883 | 0.174081152 | TRIM67 | 6_138890589 | 0.273198319 | ECT2L | 1_66193972 | 0.227341966 | PDE4B |
| 1_231191282 | 0.174081152 | TRIM67 | 7_105623899 | 0.273006824 | ATXN7L1 | 3_138447990 | 0.227285759 | ESYT3 |
| 1_231192113 | 0.174081152 | TRIM67 | 5_83495995 | 0.273001714 | VCAN | 2_16633741 | 0.22724694 | FAM49A |
| 1_231200663 | 0.174081152 | TRIM67 | 15_100300134 | 0.272983476 | ADAMTS17 | 2_50551086 | 0.227236097 | NRXN1 |
| 1_231200785 | 0.174081152 | TRIM67 | 2_1321874 | 0.272908536 | SNTG2 | 5_19558289 | 0.227139823 | CDH18 |
| 5_140560717 | 0.174006786 | APBB3 | 22_41629984 | 0.272725687 | XRCC6 | 1_45196181 | 0.227125935 | ZSWIM5 |
| 2_44231915 | 0.174001581 | PPM1B | 2_71188745 | 0.272693874 | PAIP2B | 20_20040441 | 0.227030382 | CRNKL1 |
| 6_144641830 | 0.173951712 | UTRN | 1_13590502 | 0.272636113 | PDPN | 3_138557699 | 0.22690315 | CEP70 |
| 10_107133828 | 0.17395165 | SORCS1 | 3_85999270 | 0.272629877 | CADM2 | 6_70295476 | 0.226881941 | COL9A1 |
| 1_85277177 | 0.173937992 | LOC646626 | 22_46692789 | 0.272495958 | CERK | 12_120771499 | 0.226856065 | SPPL3 |
| 5_106840263 | 0.173884053 | LOC102467213 | 11_118118082 | 0.272491637 | TMPRSS4 | 7_76418632 | 0.226724902 | ZP3 |
| 3_56583826 | 0.173812724 | CCDC66 | 3_126401360 | 0.272337438 | CCDC37 | 20_62738100 | 0.22670786 | NTSR1 |
| 12_3015345 | 0.173764614 | TEAD4 | 7_149255221 | 0.272170436 | ZNF212 | 18_74444010 | 0.226632364 | FAM69C |
| 8_10405514 | 0.173759991 | MSRA | 4_108747356 | 0.272114565 | ETNPPL | 21_14172633 | 0.226623582 | LIPI |
| 2_170411526 | 0.173684988 | MYO3B | 5_73796411 | 0.272107359 | ARHGEF28 | 20_63470615 | 0.226617023 | KCNQ2 |
| 7_78029583 | 0.17367925 | MAGI2 | 5_73796444 | 0.272107359 | ARHGEF28 | 12_27338277 | 0.226605353 | ARNTL2 |
| 7_78030350 | 0.17367925 | MAGI2 | 5_73796494 | 0.272107359 | ARHGEF28 | 12_110139664 | 0.22655015 | IFT81 |
| 10_70407131 | 0.17367481 | EIF4EBP2 | 5_73796496 | 0.272107359 | ARHGEF28 | 6_29828592 | 0.226540796 | HLA-G |
| 2_170427397 | 0.173599857 | MYO3B | 17_65105279 | 0.272046384 | LOC100507002 | 2_70034779 | 0.226332207 | PCBP1-AS1 |
| 6_128486909 | 0.173588857 | PTPRK | 14_63957265 | 0.271996243 | SYNE2 | 10_93452882 | 0.226217854 | MYOF |
| 7_7089833 | 0.173475506 | LOC100131257 | 6_34376801 | 0.271712434 | NUDT3 | 9_68409799 | 0.226119886 | PGM5 |
| 5_109704541 | 0.17340686 | MAN2A1 | 7_75926031 | 0.271625972 | POR | 22_18154327 | 0.226058759 | USP18 |
| 5_109707482 | 0.17340686 | MAN2A1 | 19_2598521 | 0.271568616 | GNG7 | 21_42133903 | 0.226021227 | UMODL1 |
| 5_109717432 | 0.17340686 | MAN2A1 | 14_61575581 | 0.271418169 | FLJ22447 | 10_31382016 | 0.226019871 | ZEB1 |
| 17_4359872 | 0.173390058 | UBE2G1 | 5_83496806 | 0.271205456 | VCAN | 11_118492709 | 0.226000695 | KMT2A |
| 15_63557088 | 0.173305727 | USP3 | 11_45922554 | 0.271197871 | GYLTL1B | 14_63381503 | 0.225925158 | PPP2R5E |
| 1_62120426 | 0.173304013 | INADL | 9_113900469 | 0.27116705 | ZNF618 | 1_245575097 | 0.225770257 | KIF26B |
| 13_101111355 | 0.173302361 | NALCN | 18_13315784 | 0.271165905 | LDLRAD4 | 6_29828972 | 0.225760145 | HLA-G |
| 7_93458232 | 0.173278075 | CALCR | 1_35487694 | 0.271160788 | KIAA0319L | 2_172454139 | 0.225714012 | ITGA6 |
| 7_93498428 | 0.173278075 | CALCR | 12_314320 | 0.27106615 | KDM5A | 14_96079682 | 0.2256855 | C14orf132 |
| 7_93500687 | 0.173278075 | CALCR | 2_196912586 | 0.270906796 | PGAP1 | 14_96080493 | 0.2256855 | C14orf132 |
| 7_93504128 | 0.173278075 | CALCR | 12_110285916 | 0.270871898 | ATP2A2 | 15_47317874 | 0.225647067 | SEMA6D |
| 7_93511053 | 0.173278075 | CALCR | 1_98014760 | 0.270669877 | MIR137HG | 6_36508990 | 0.225646254 | STK38 |
| 6_159235181 | 0.173254646 | FNDC1 | 2_196912587 | 0.270469558 | PGAP1 | 8_72821446 | 0.225637344 | KCNB2 |
| 7_104826228 | 0.173136414 | LHFPL3 | 17_1708251 | 0.270267044 | TLCD2 | 2_55533804 | 0.225552617 | CCDC104 |
| 7_90253815 | 0.173127009 | C7orf63 | 12_314793 | 0.270224407 | KDM5A | 6_151386750 | 0.225489091 | ZBTB2 |
| 7_90264874 | 0.173127009 | C7orf63 | 14_61570965 | 0.27020239 | FLJ22447 | 6_151405621 | 0.225489091 | RMND1 |
| 12_81335759 | 0.173083206 | PPFIA2 | 15_66201778 | 0.270196164 | MEGF11 | 19_13391961 | 0.225474659 | CACNA1A |
| 4_80889454 | 0.173001197 | C4orf22 | 3_192341455 | 0.270123058 | FGF12 | 13_102158544 | 0.225466355 | FGF14 |
| 16_82891080 | 0.172999584 | CDH13 | 14_61557049 | 0.27010738 | LOC101927780 | 2_241190416 | 0.225439902 | ANO7 |
| 7_93484812 | 0.172873328 | CALCR | 10_25489393 | 0.270090027 | GPR158 | 12_120890959 | 0.225438285 | SPPL3 |
| 8_29343568 | 0.172789473 | DUSP4 | 11_46038531 | 0.270042558 | PHF21A | 20_63422849 | 0.225408373 | KCNQ2 |
| 7_104840075 | 0.172770807 | LHFPL3 | 13_33163709 | 0.2698821 | STARD13 | 14_100896081 | 0.225379045 | MEG8 |
| 2_71384596 | 0.172622158 | ZNF638 | 5_167649658 | 0.269874369 | TENM2 | 12_112181961 | 0.225363046 | HECTD4 |
| 5_106851351 | 0.172484309 | LOC102467213 | 13_70120550 | 0.269866453 | ATXN8OS | 2_64115043 | 0.225283904 | PELI1 |
| 7_78031062 | 0.172429571 | MAGI2 | 4_176252066 | 0.269843963 | ASB5 | 7_40089729 | 0.22523842 | CDK13 |
| 16_89863721 | 0.172405629 | SPIRE2 | 4_188097459 | 0.269730835 | TRIML2 | 7_40089731 | 0.22523842 | CDK13 |
| 6_166775792 | 0.172400324 | RPS6KA2 | 12_45034202 | 0.269640839 | DBX2 | 7_40089739 | 0.22523842 | CDK13 |
| 6_166777092 | 0.172400324 | RPS6KA2 | 17_75814224 | 0.269624754 | UNK | 11_117651252 | 0.225121369 | DSCAML1 |
| 6_166790842 | 0.172400324 | RPS6KA2 | 8_17657435 | 0.269524019 | MTUS1 | 6_36405898 | 0.225077064 | PXT1 |
| 9_134859278 | 0.172372867 | LOC101448202 | 13_70134715 | 0.269482599 | ATXN8OS | 2_55670430 | 0.22504545 | PNPT1 |
| 1_231186471 | 0.17233752 | LOC149373 | 20_35141127 | 0.269456416 | EDEM2 | 6_151406458 | 0.224947647 | RMND1 |
| 10_86717145 | 0.172227821 | LDB3 | 6_21890444 | 0.269353503 | CASC15 | 1_1292517 | 0.224910904 | ACAP3 |
| 6_128474203 | 0.172210395 | PTPRK | 16_82887576 | 0.269219962 | CDH13 | 8_72837101 | 0.224853883 | KCNB2 |
| 2_71394569 | 0.172204673 | ZNF638 | 7_149275607 | 0.269193239 | ZNF783 | 20_20038252 | 0.224652759 | CRNKL1 |
| 2_71422595 | 0.172204673 | ZNF638 | 9_114244069 | 0.269150905 | COL27A1 | 8_58851189 | 0.22462971 | TOX |

| | | | | | | | | |
|---|---|---|---|---|---|---|---|---|
| 7_104899740 | 0.172127655 | LHFPL3 | 9_3500857 | 0.269097771 | RFX3 | 3_138610733 | 0.224610789 | FAIM |
| 8_10410998 | 0.172119281 | MSRA | 3_129424674 | 0.269032248 | EFCAB12 | 15_33546993 | 0.224521209 | RYR3 |
| 16_82888372 | 0.172068319 | CDH13 | 4_89744105 | 0.268807787 | SNCA | 6_32637480 | 0.22450004 | HLA-DQA1 |
| 7_104905216 | 0.172048967 | LHFPL3 | 4_95448232 | 0.268719237 | UNC5C | 2_70056078 | 0.224490224 | PCBP1-AS1 |
| 8_29341545 | 0.17198648 | DUSP4 | 9_3493586 | 0.268592707 | RFX3 | 2_70062490 | 0.224490224 | PCBP1-AS1 |
| 6_10411162 | 0.171975218 | TFAP2A | 7_147289697 | 0.268586447 | CNTNAP2 | 20_63448386 | 0.224477483 | KCNQ2 |
| 15_97753183 | 0.171938812 | LINC00923 | 10_60421370 | 0.268455892 | ANK3 | 12_12435414 | 0.224356317 | LOH12CR1 |
| 11_123141547 | 0.171803411 | CLMP | 1_35942205 | 0.268437651 | AGO3 | 7_90365267 | 0.224251366 | GTPBP10 |
| 16_89784007 | 0.171802829 | FANCA | 7_54546170 | 0.268297384 | VSTM2A | 10_60321414 | 0.22411173 | ANK3 |
| 1_161043813 | 0.17177257 | USF1 | 16_89862193 | 0.268293412 | SPIRE2 | 6_29669292 | 0.223943627 | MOG |
| 5_109805826 | 0.171770241 | MAN2A1 | 9_135580011 | 0.268246022 | LOC100130954 | 3_133581287 | 0.223899897 | CDV3 |
| 6_159233519 | 0.171711324 | FNDC1 | 17_75804527 | 0.268212382 | UNK | 17_5557205 | 0.223851211 | NLRP1 |
| 6_159234561 | 0.171711324 | FNDC1 | 8_14819664 | 0.268023054 | SGCZ | 7_78027650 | 0.223717019 | MAGI2 |
| 6_159236163 | 0.171711324 | FNDC1 | 16_89846600 | 0.267950968 | SPIRE2 | 20_62817617 | 0.223712946 | COL9A3 |
| 6_159239073 | 0.171711324 | FNDC1 | 17_58730332 | 0.267943572 | RAD51C | 2_54860178 | 0.223624581 | EML6 |
| 1_7443235 | 0.17166488 | CAMTA1 | 2_39102242 | 0.267890684 | SOS1 | 2_64128057 | 0.223454899 | PELI1 |
| 1_231534081 | 0.171625721 | TSNAX | 13_71838807 | 0.267874152 | DACH1 | 6_32642012 | 0.223426232 | HLA-DQA1 |
| 7_104860887 | 0.171483069 | LHFPL3 | 14_49715759 | 0.267831628 | KLHDC1 | 10_14048907 | 0.223424298 | FRMD4A |
| 7_104871129 | 0.171483069 | LHFPL3 | 3_129411879 | 0.267806205 | EFCAB12 | 6_30015524 | 0.223374896 | ZNRD1-AS1 |
| 7_103440341 | 0.171474293 | SLC26A5 | 11_46070020 | 0.267797305 | PHF21A | 6_30889765 | 0.223371544 | DDR1 |
| 9_131883212 | 0.171467779 | MED27 | 11_129963277 | 0.267757896 | PRDM10 | 10_117259615 | 0.223340436 | SLC18A2 |
| 6_159218252 | 0.171448116 | FNDC1 | 4_38485424 | 0.267673861 | LINC01258 | 2_64114589 | 0.223334717 | PELI1 |
| 12_48346797 | 0.171413713 | ZNF641 | 12_44866799 | 0.267571113 | NELL2 | 2_64116098 | 0.223334717 | PELI1 |
| 7_93473582 | 0.171368704 | CALCR | 11_118119288 | 0.267561972 | TMPRSS4 | 8_72838045 | 0.223284856 | KCNB2 |
| 13_107287707 | 0.17136168 | FAM155A | 1_169534412 | 0.267187291 | F5 | 8_72838658 | 0.223284856 | KCNB2 |
| 13_107287708 | 0.17136168 | FAM155A | 16_87215452 | 0.267159386 | LOC101928708 | 6_99432504 | 0.22320129 | USP45 |
| 7_117595164 | 0.171314096 | CFTR | 7_71674350 | 0.266875012 | WBSCR17 | 3_138464118 | 0.223185593 | ESYT3 |
| 15_83575119 | 0.171292802 | SH3GL3 | 7_147291148 | 0.266862281 | CNTNAP2 | 3_138505960 | 0.223185593 | CEP70 |
| 4_89750398 | 0.171211505 | SNCA | 3_177174442 | 0.266798362 | TBL1XR1 | 6_39839312 | 0.223168735 | DAAM2 |
| 2_162559937 | 0.171190343 | KCNH7 | 11_129451171 | 0.266662045 | BARX2 | 2_55639318 | 0.223111841 | PNPT1 |
| 3_129478272 | 0.171125016 | IFT122 | 13_94320258 | 0.266605882 | GPC6 | 8_67136570 | 0.223035535 | CSPP1 |
| 1_111836404 | 0.171064018 | KCND3 | 16_73057548 | 0.266586761 | ZFHX3 | 11_118553293 | 0.222960262 | IFT46 |
| 1_111839043 | 0.171064018 | KCND3 | 9_17405029 | 0.266488109 | CNTLN | 2_69970215 | 0.222885609 | PCBP1-AS1 |
| 12_54980202 | 0.171061562 | TESPA1 | 1_231191282 | 0.266402197 | TRIM67 | 7_90258563 | 0.222880853 | C7orf63 |
| 12_54980210 | 0.171061562 | TESPA1 | 14_92474706 | 0.266259466 | SLC24A4 | 3_64800616 | 0.222790754 | ADAMTS9-AS2 |
| 6_159244660 | 0.170950055 | FNDC1 | 4_188099403 | 0.266202602 | TRIML2 | 2_16632177 | 0.222668049 | FAM49A |
| 2_206745961 | 0.170902017 | MDH1B | 19_949281 | 0.266100286 | ARID3A | 12_76030334 | 0.222667176 | PHLDA1 |
| 21_17599901 | 0.170830707 | BTG3 | 1_74391958 | 0.265861912 | FPGT-TNNI3K | 9_132348312 | 0.222664008 | SETX |
| 5_106848509 | 0.170816678 | LOC102467213 | 18_49392938 | 0.265839509 | DYM | 1_244585928 | 0.222611657 | C1orf101 |
| 1_62122685 | 0.170806475 | INADL | 3_124233756 | 0.265529879 | KALRN | 12_111876531 | 0.222601855 | MAPKAPK5 |
| 1_31037016 | 0.170796331 | PUM1 | 5_73800066 | 0.265390819 | ARHGEF28 | 7_72374423 | 0.222520964 | CALN1 |
| 1_203487911 | 0.170787955 | PRELP | 1_144458954 | 0.265235421 | NBPF15 | 2_70037090 | 0.222490307 | PCBP1-AS1 |
| 9_2142671 | 0.170736397 | SMARCA2 | 9_76675941 | 0.265224656 | PRUNE2 | 2_55661278 | 0.222460657 | PNPT1 |
| 22_49570169 | 0.170717203 | C22orf34 | 13_107757288 | 0.265199069 | FAM155A | 9_77618764 | 0.222410627 | GNA14 |
| 1_162866418 | 0.170707237 | C1orf110 | 3_195119296 | 0.265146993 | XXYLT1 | 3_129544171 | 0.222375146 | H1FOO |
| 9_131885221 | 0.170649685 | MED27 | 3_192342294 | 0.265060526 | FGF12 | 1_247961481 | 0.222370454 | OR2L13 |
| 20_20040441 | 0.170610286 | CRNKL1 | 6_34322705 | 0.264959437 | NUDT3 | 6_110829628 | 0.222339406 | AMD1 |
| 6_16317685 | 0.170564781 | ATXN1 | 4_108752565 | 0.264937005 | ETNPPL | 3_64623523 | 0.222141359 | ADAMTS9 |
| 13_73867599 | 0.17052672 | KLF12 | 17_10137701 | 0.264903532 | GAS7 | 17_5555845 | 0.222135527 | NLRP1 |
| 7_103440678 | 0.170474299 | SLC26A5 | 3_124286079 | 0.264709467 | KALRN | 15_83906823 | 0.22212972 | ADAMTSL3 |
| 7_103441854 | 0.170474299 | SLC26A5 | 12_374956 | 0.264664343 | KDM5A | 2_55616502 | 0.222044451 | SMEK2 |
| 8_10416620 | 0.170407538 | MSRA | 12_378544 | 0.264664343 | KDM5A | 15_33558366 | 0.222032312 | RYR3 |
| 5_126620771 | 0.170397484 | PHAX | 17_81911405 | 0.26465795 | PCYT2 | 3_138544233 | 0.222021842 | CEP70 |
| 5_126620800 | 0.170397484 | PHAX | 1_35909509 | 0.264578657 | AGO1 | 12_112143028 | 0.221820855 | TRAFD1 |
| 7_104831984 | 0.17034372 | LHFPL3 | 7_149278684 | 0.264425873 | ZNF783 | 11_118492154 | 0.221815475 | KMT2A |
| 5_106829597 | 0.170274759 | LOC102467213 | 13_85403073 | 0.264338759 | LINC00351 | 13_71545529 | 0.221591438 | DACH1 |
| 4_89742519 | 0.170264204 | SNCA | 8_116748113 | 0.26403117 | EIF3H | 2_50001228 | 0.221590937 | NRXN1 |
| 4_89753300 | 0.170264204 | SNCA | 8_143003992 | 0.263908121 | LOC100133669 | 10_96267770 | 0.221522821 | BLNK |
| 6_144579773 | 0.170230501 | UTRN | 3_14117633 | 0.263850077 | CHCHD4 | 7_100955297 | 0.221502012 | MUC3A |
| 7_120479798 | 0.170169157 | KCND2 | 3_192352553 | 0.263752712 | FGF12 | 6_155440431 | 0.221485587 | NOX3 |
| 8_10411909 | 0.170146721 | MSRA | 15_100299520 | 0.263735293 | ADAMTS17 | 4_41443205 | 0.221453891 | LIMCH1 |
| 7_94402493 | 0.170141172 | COL1A2 | 9_17352089 | 0.263583544 | CNTLN | 16_56444627 | 0.221384741 | NUDT21 |
| 10_59701772 | 0.170133338 | SLC16A9 | 16_87217466 | 0.263431602 | LOC101928708 | 20_63439051 | 0.221363427 | KCNQ2 |
| 19_37543024 | 0.170098986 | ZNF793 | 13_97300939 | 0.263366966 | MBNL2 | 20_9753347 | 0.221287839 | PAK7 |
| 5_160770779 | 0.170076438 | ATP10B | 15_38542837 | 0.263186784 | RASGRP1 | 8_72839198 | 0.22124458 | KCNB2 |
| 12_12489401 | 0.170058375 | DUSP16 | 7_71673354 | 0.26310714 | WBSCR17 | 6_37638528 | 0.220853121 | MDGA1 |
| 12_12507679 | 0.170058375 | DUSP16 | 12_80471625 | 0.262962268 | PTPRQ | 12_111942777 | 0.220743345 | TMEM116 |
| 12_12507991 | 0.170058375 | DUSP16 | 7_149250607 | 0.262908957 | ZNF212 | 2_71427045 | 0.220594988 | ZNF638 |
| 6_166796735 | 0.170018379 | RPS6KA2 | 3_14127579 | 0.262885457 | TMEM43 | 6_30669686 | 0.220553263 | DHX16 |
| 7_104881030 | 0.169972686 | LHFPL3 | 1_237669336 | 0.262817821 | RYR2 | 21_42133900 | 0.220500742 | UMODL1 |
| 1_161043522 | 0.16993939 | USF1 | 16_89862446 | 0.262716161 | SPIRE2 | 10_17791495 | 0.220215884 | TMEM236 |
| 6_144649071 | 0.169921327 | UTRN | 6_27705964 | 0.262695387 | LINC01012 | 8_100952672 | 0.220128036 | YWHAZ |
| 9_132321956 | 0.169914545 | SETX | 19_4913808 | 0.262626224 | UHRF1 | 12_95704641 | 0.220043925 | NTN4 |
| 22_32667756 | 0.169820465 | SYN3 | 2_231315204 | 0.262621097 | ARMC9 | 3_138547793 | 0.219983295 | CEP70 |
| 19_58232767 | 0.169811445 | ZNF544 | 7_149280117 | 0.26255819 | ZNF783 | 5_151893303 | 0.2199564 | GLRA1 |
| 19_58232770 | 0.169811445 | ZNF544 | 17_75819945 | 0.262253024 | UNK | 4_87976728 | 0.219912865 | SPP1 |
| 7_104890721 | 0.169760114 | LHFPL3 | 14_61557599 | 0.262241987 | LOC101927780 | 3_138553341 | 0.219835495 | CEP70 |
| 5_109702737 | 0.169756538 | MAN2A1 | 8_71318677 | 0.262199941 | EYA1 | 11_78977435 | 0.21980233 | TENM4 |
| 5_109702815 | 0.169756538 | MAN2A1 | 8_71328801 | 0.262199941 | EYA1 | 10_28858500 | 0.219698077 | C10orf126 |
| 6_144593825 | 0.169728614 | UTRN | 12_53587568 | 0.262191316 | ATF7 | 5_10380358 | 0.219690759 | 6-Mar |
| 6_144615807 | 0.169728614 | UTRN | 13_97290972 | 0.262089052 | MBNL2 | 6_29943483 | 0.21964323 | HLA-A |
| 11_101030102 | 0.169657229 | PGR | 17_65105224 | 0.262056058 | LOC100507002 | 9_19014503 | 0.219631727 | FAM154A |
| 3_129469237 | 0.169632544 | IFT122 | 1_13586482 | 0.261976798 | PDPN | 1_54730247 | 0.219617261 | MROH7-TTC4 |
| 2_71352372 | 0.169631987 | ZNF638 | 1_231204650 | 0.261754136 | TRIM67 | 3_138593825 | 0.219484739 | CEP70 |
| 2_71377474 | 0.169631987 | ZNF638 | 11_46062486 | 0.261714093 | PHF21A | 14_63381929 | 0.219408158 | PPP2R5E |
| 2_71427914 | 0.169631987 | ZNF638 | 1_35988415 | 0.261672247 | AGO3 | 2_69976897 | 0.219405866 | PCBP1-AS1 |
| 6_169696893 | 0.169628311 | WDR27 | 19_54584197 | 0.261662661 | LILRA2 | 8_72836632 | 0.219401216 | KCNB2 |
| 3_139116442 | 0.169603206 | BPESC1 | 4_108760111 | 0.261503085 | ETNPPL | 11_78976734 | 0.219370214 | TENM4 |
| 1_53113437 | 0.169544386 | SLC1A7 | 2_77011546 | 0.261308034 | LRRTM4 | 6_39839031 | 0.219074528 | DAAM2 |
| 14_50189070 | 0.169517223 | SOS2 | 19_16021182 | 0.261220946 | LINC00661 | 11_118035071 | 0.21905888 | TMPRSS4-AS1 |
| 16_7298292 | 0.169509191 | RBFOX1 | 11_94614088 | 0.261155564 | PIWIL4 | 1_244603217 | 0.219042773 | C1orf101 |
| 11_121621790 | 0.169471223 | SORL1 | 1_155827874 | 0.260964557 | GON4L | 2_71366405 | 0.21900833 | ZNF638 |
| 7_104898793 | 0.169452264 | LHFPL3 | 1_35983703 | 0.260855709 | AGO3 | 15_52240725 | 0.218935778 | MYO5C |
| 8_72817778 | 0.169425245 | KCNB2 | 22_25773236 | 0.260652378 | MYO18B | 6_151405040 | 0.218829915 | RMND1 |
| 2_30469579 | 0.169368626 | LCLAT1 | 11_101030102 | 0.260573967 | PGR | 3_138496935 | 0.218799143 | CEP70 |
| 5_13717362 | 0.169333213 | DNAH5 | 1_13589065 | 0.260424221 | PDPN | 12_112148558 | 0.218745107 | TRAFD1 |
| 22_42997431 | 0.169208906 | PACSIN2 | 12_315543 | 0.260401724 | KDM5A | 1_77886183 | 0.218717639 | NEXN-AS1 |

| | | | | | | | | |
|---|---|---|---|---|---|---|---|---|
| 1_162866233 | 0.169196113 | C1orf110 | 1_156063053 | 0.260297818 | RAB25 | 2 50013373 | 0.218599915 | NRXN1 |
| 2 71339440 | 0.169170631 | ZNF638 | 5 168889745 | 0.260288494 | SLIT3 | 15 48898363 | 0.218597772 | SHC4 |
| 10 52065816 | 0.169045612 | PRKG1 | 17 75799198 | 0.260195197 | UNK | 15 100323934 | 0.21849735 | ADAMTS17 |
| 2_206738510 | 0.169044813 | MDH1B | 2_71184783 | 0.2600521 | PAIP2B | 12_112267157 | 0.21843714 | HECTD4 |
| 5 106852056 | 0.169003704 | LOC102467213 | 2 71189412 | 0.2600521 | PAIP2B | 6 39840142 | 0.218349488 | DAAM2 |
| 7 104850025 | 0.169000081 | LHFPL3 | 6 34343637 | 0.259872343 | NUDT3 | 7 40089727 | 0.218341884 | CDK13 |
| 7 104853716 | 0.169000081 | LHFPL3 | 19 45951628 | 0.259867369 | NOVA2 | 12 112055305 | 0.218247042 | NAA25 |
| 7 104854113 | 0.169000081 | LHFPL3 | 7 922735 | 0.259856983 | ADAP1 | 10 70886580 | 0.218030402 | PCBD1 |
| 7 104854118 | 0.169000081 | LHFPL3 | 13 71840787 | 0.259856702 | DACH1 | 2 55674317 | 0.217817165 | PNPT1 |
| 2 12494563 | 0.168985751 | LOC100506457 | 10 50315239 | 0.259800784 | SGMS1 | 2 55686835 | 0.217817165 | PNPT1 |
| 9 68281601 | 0.16895362 | CBWD3 | 9 113895784 | 0.259800784 | ZNF618 | 4 188097459 | 0.217817165 | TRIML2 |
| 11 113365142 | 0.168953 | TTC12 | 3 192339529 | 0.259602559 | FGF12 | 2 29069074 | 0.217808968 | C2orf71 |
| 2 166940607 | 0.168909879 | XIRP2 | 19 16034613 | 0.259586452 | LINC00905 | 6_128205884 | 0.217799711 | PTPRK |
| 8 38107872 | 0.168898651 | ASH2L | 19 48676224 | 0.259547148 | SEC1P | 12 111735265 | 0.217777972 | ACAD10 |
| 8 29344706 | 0.168867499 | DUSP4 | 12 32894088 | 0.259499945 | PKP2 | 12 111771193 | 0.217777972 | ALDH2 |
| 8 29350355 | 0.168867499 | DUSP4 | 15 48472745 | 0.259428622 | FBN1 | 12 112294045 | 0.217704416 | HECTD4 |
| 16 82890284 | 0.168779413 | CDH13 | 9 135585096 | 0.259418006 | LOC100130954 | 1 244571310 | 0.21761721 | C1orf101 |
| 13 28199231 | 0.168758316 | PAN3 | 11 94614082 | 0.259415933 | PIWIL4 | 12 111703011 | 0.217612543 | ACAD10 |
| 2 237778130 | 0.168736146 | LRRFIP1 | 2 1072352 | 0.259407342 | SNTG2 | 7 90251027 | 0.21760795 | C7orf63 |
| 2 162513258 | 0.16871953 | KCNH7 | 1 231192042 | 0.259375287 | TRIM67 | 7 90264874 | 0.21760795 | C7orf63 |
| 9 133162643 | 0.168635752 | GBGT1 | 19 16022783 | 0.259373122 | LINC00661 | 11 118595534 | 0.217587684 | ARCN1 |
| 17 19281898 | 0.168628954 | EPN2 | 12 374832 | 0.259299989 | KDM5A | 3 8004008 | 0.217564057 | LOC101927394 |
| 2 221559342 | 0.168606834 | EPHA4 | 1 231215991 | 0.259290243 | TRIM67 | 17 2994391 | 0.217556291 | RAP1GAP2 |
| 3 33119661 | 0.168579163 | CRTAP | 19 12650974 | 0.259133571 | MAN2B1 | 20 63456522 | 0.21745152 | KCNQ2 |
| 3 33119912 | 0.168579163 | CRTAP | 2 37264523 | 0.259067592 | PRKD3 | 13 21008781 | 0.217432688 | LATS2 |
| 1 65370244 | 0.168558836 | DNAJC6 | 4 108565226 | 0.258988171 | RPL34-AS1 | 2 50003778 | 0.217427186 | NRXN1 |
| 5 109714412 | 0.168425227 | MAN2A1 | 1 244424427 | 0.258955267 | ADSS | 18 42599050 | 0.217338135 | LINC00907 |
| 7 104848020 | 0.168418298 | LHFPL3 | 1 244440434 | 0.258955267 | ADSS | 13 28259135 | 0.217292482 | PAN3 |
| 5 167834728 | 0.168415679 | TENM2 | 3_124225570 | 0.258913196 | KALRN | 18_23408179 | 0.21720265 | TMEM241 |
| 1 111938880 | 0.16839042 | KCND3 | 13 107711860 | 0.258858241 | FAM155A | 3 138468250 | 0.217040551 | ESYT3 |
| 5 109732798 | 0.168373829 | MAN2A1 | 8 132939672 | 0.258850548 | TG | 22 30067264 | 0.217027315 | LOC101929664 |
| 5_109776323 | 0.168373829 | MAN2A1 | 3_124224997 | 0.258682414 | KALRN | 12_27265434 | 0.217008687 | STK38L |
| 8 10408112 | 0.168362533 | MSRA | 3 124236125 | 0.258682414 | KALRN | 8 58851187 | 0.217008618 | TOX |
| 8 72883089 | 0.168362151 | KCNB2 | 3 124236683 | 0.258682414 | KALRN | 10 96253745 | 0.216949331 | BLNK |
| 5 11162081 | 0.168300934 | CTNND2 | 8 96521538 | 0.25858972 | SDC2 | 12 27264357 | 0.216945666 | STK38L |
| 13 28261325 | 0.168284954 | PAN3 | 2 108750562 | 0.258382813 | RANBP2 | 2 64129300 | 0.216893484 | PELI1 |
| 5 93830329 | 0.168218171 | FAM172A | 3 192372825 | 0.258129142 | FGF12 | 16 89323224 | 0.216811837 | ANKRD11 |
| 5 109740787 | 0.168212806 | MAN2A1 | 15 48626579 | 0.258088051 | FBN1 | 8 62527887 | 0.216688138 | NKAIN3 |
| 5_109741816 | 0.168212806 | MAN2A1 | 12_53542391 | 0.258018166 | ATF7 | 8_62527897 | 0.216688138 | NKAIN3 |
| 5 109749106 | 0.168212806 | MAN2A1 | 4 88100507 | 0.257997406 | ABCG2 | 21 14201872 | 0.216649197 | LIPI |
| 5 109752655 | 0.168212806 | MAN2A1 | 3 124224893 | 0.257868101 | KALRN | 12 95709926 | 0.216585692 | NTN4 |
| 2_167010237 | 0.168211051 | XIRP2 | 3_124236641 | 0.257868101 | KALRN | 12_112280849 | 0.216309566 | HECTD4 |
| 2 167013107 | 0.168211051 | XIRP2 | 2 113923436 | 0.257777009 | ACTR3 | 1 33759326 | 0.216286749 | CSMD2 |
| 2 167014863 | 0.168211051 | XIRP2 | 8 14819988 | 0.257711142 | SGCZ | 1 33759327 | 0.216286749 | CSMD2 |
| 8_116649873 | 0.168176696 | EIF3H | 4_108754259 | 0.257611585 | ETNPPL | 1_33759329 | 0.216286749 | CSMD2 |
| 17 3924792 | 0.168125631 | ATP2A3 | 4 108756596 | 0.257611585 | ETNPPL | 1 33759330 | 0.216286749 | CSMD2 |
| 14 57621839 | 0.168107255 | SLC35F4 | 7 149280124 | 0.257528544 | ZNF783 | 1 33759331 | 0.216286749 | CSMD2 |
| 12_54974744 | 0.168047833 | TESPA1 | 22_40451844 | 0.257433649 | MKL1 | 10_96317769 | 0.216241633 | DNTT |
| 6 144588611 | 0.168046951 | UTRN | 19 56115355 | 0.257336766 | ZNF787 | 8 40580015 | 0.216205584 | ZMAT4 |
| 12 12473371 | 0.168014055 | DUSP16 | 19 54538543 | 0.257311797 | KIR3DX1 | 3 314288 | 0.216179839 | CHL1 |
| 12 112200156 | 0.167918014 | HECTD4 | 11 46108676 | 0.257246796 | PHF21A | 3 138446698 | 0.216176142 | ESYT3 |
| 8 29341767 | 0.167915739 | DUSP4 | 3 55060528 | 0.257007576 | CACNA2D3 | 18 13536326 | 0.215884566 | LDLRAD4 |
| 5 95858889 | 0.167909645 | C5orf27 | 3 55066891 | 0.257007576 | CACNA2D3 | 1 242271219 | 0.215776098 | PLD5 |
| 3 33120253 | 0.167897582 | CRTAP | 2 170427218 | 0.256974043 | MYO3B | 8 72841608 | 0.215696507 | KCNB2 |
| 10_59703757 | 0.167809521 | SLC16A9 | 5_149287753 | 0.256971628 | AFAP1L1 | 3_138496673 | 0.215692294 | CEP70 |
| 19 15049697 | 0.16779366 | CASP14 | 13 43391303 | 0.256847304 | ENOX1 | 11 2870735 | 0.215664814 | KCNQ1DN |
| 1 161083619 | 0.167773235 | PVRL4 | 1 35517493 | 0.256414605 | KIAA0319L | 19 13410714 | 0.215641408 | CACNA1A |
| 1_15050029 | 0.167766136 | KAZN | 17_76352645 | 0.256088074 | PRPSAP1 | 13_111324135 | 0.215608172 | TEX29 |
| 1 15050034 | 0.167766136 | KAZN | 17 75798132 | 0.256061166 | UNK | 15 90636355 | 0.215501902 | CRTC3 |
| 1 62114529 | 0.167714314 | INADL | 9 76675930 | 0.256017865 | PRUNE2 | 18 42606086 | 0.215386819 | LINC00907 |
| 6_10415074 | 0.16771356 | TFAP2A | 1_147253708 | 0.255974934 | CHD1L | 3_138616757 | 0.215321249 | FAIM |
| 12 12488435 | 0.167707148 | DUSP16 | 19 6254962 | 0.255933261 | MLLT1 | 14 63379170 | 0.215309817 | PPP2R5E |
| 12 12488979 | 0.167707148 | DUSP16 | 10 82062702 | 0.255928045 | NRG3 | 12 7439598 | 0.215276748 | CD163L1 |
| 12 12488985 | 0.167707148 | DUSP16 | 9 135585085 | 0.255922111 | LOC100130954 | 17 19324732 | 0.215197044 | EPN2 |
| 12 12490653 | 0.167707148 | DUSP16 | 9 135585095 | 0.255922111 | LOC100130954 | 17 19344191 | 0.215197044 | B9D1 |
| 19 37971473 | 0.167665041 | SIPA1L3 | 20 59603172 | 0.255807415 | PHACTR3 | 1 244561822 | 0.215182254 | C1orf101 |
| 5 106840470 | 0.167641971 | LOC102467213 | 1 35492253 | 0.255706569 | KIAA0319L | 19 1257538 | 0.215148273 | MIDN |
| 8 72818631 | 0.167521234 | KCNB2 | 15 100271458 | 0.255387015 | ADAMTS17 | 10 96317473 | 0.215146354 | DNTT |
| 1 161042970 | 0.167511733 | USF1 | 8 13181343 | 0.255354874 | DLC1 | 17 78126139 | 0.215138276 | TMC6 |
| 1 111841033 | 0.167453266 | KCND3 | 4 146790189 | 0.255251477 | TTC29 | 4 141068716 | 0.215126072 | RNF150 |
| 1 111841478 | 0.167453266 | KCND3 | 11 118138670 | 0.255247392 | SCN4B | 12 120780588 | 0.215085353 | SPPL3 |
| 4 6476189 | 0.167453266 | PPP2R2C | 15 100297896 | 0.255211852 | ADAMTS17 | 12 120805896 | 0.215084003 | SPPL3 |
| 13 70139745 | 0.167450403 | ATXN8OS | 15 100297898 | 0.255211852 | ADAMTS17 | 12 31601531 | 0.215055728 | DENND5B-AS1 |
| 18 42348490 | 0.167448973 | LINC00907 | 8 40641600 | 0.255206676 | ZMAT4 | 12 121412388 | 0.215032932 | RNF34 |
| 7 104882359 | 0.167448973 | LHFPL3 | 15 25741972 | 0.255128453 | ATP10A | 5 176625941 | 0.215030871 | SNCB |
| 7 104882733 | 0.167448973 | LHFPL3 | 9 113888399 | 0.255123766 | ZNF618 | 4 188103290 | 0.214908365 | TRIML2 |
| 7 104890607 | 0.167448973 | LHFPL3 | 7 5944863 | 0.254949196 | RSPH10B | 10 96259058 | 0.214852145 | BLNK |
| 2 170416232 | 0.167435127 | MYO3B | 7 6158097 | 0.254882405 | USP42 | 12 112083553 | 0.214847296 | NAA25 |
| 2 170416988 | 0.167435127 | MYO3B | 2 112021890 | 0.254805937 | MERTK | 22 23298537 | 0.214818139 | BCR |
| 2 170417196 | 0.167435127 | MYO3B | 15 100272731 | 0.254779462 | ADAMTS17 | 1 244562024 | 0.214755669 | C1orf101 |
| 5 178454762 | 0.167416105 | COL23A1 | 7 53663712 | 0.254594642 | FLJ45974 | 6 30001358 | 0.214737203 | ZNRD1-AS1 |
| 5 178457458 | 0.167416105 | COL23A1 | 7 53663732 | 0.254594642 | FLJ45974 | 12 112294960 | 0.214641063 | HECTD4 |
| 6 21784148 | 0.167416105 | CASC15 | 12 109140231 | 0.254560561 | ACACB | 11 133080661 | 0.214601573 | OPCML |
| 6 21807961 | 0.167294468 | CASC15 | 4 108743664 | 0.254554648 | ETNPPL | 15 30673856 | 0.214527887 | LOC100288637 |
| 9 131909252 | 0.167252086 | MED27 | 12 323477 | 0.254480688 | KDM5A | 2 55599267 | 0.214522399 | SMEK2 |
| 15_90044480 | 0.167250006 | ZNF710 | 12_379335 | 0.254480688 | KDM5A | 2_55668186 | 0.214437918 | PNPT1 |
| 11 123134594 | 0.16719663 | CLMP | 7 5944835 | 0.254440645 | RSPH10B | 6 39825534 | 0.214378047 | DAAM2 |
| 3 123640870 | 0.167000276 | MYLK | 16 89784007 | 0.254431003 | FANCA | 15 49299746 | 0.214319229 | GALK2 |
| 6 128456826 | 0.166976661 | PTPRK | 14 81343308 | 0.254419618 | STON2 | 17 19266411 | 0.21431305 | EPN2 |
| 15 27795075 | 0.166963939 | OCA2 | 6 34367219 | 0.254394482 | NUDT3 | 8 72850825 | 0.214290888 | KCNB2 |
| 4 89773436 | 0.166933749 | SNCA | 15 100299282 | 0.254383563 | ADAMTS17 | 19 6823918 | 0.214245204 | VAV1 |
| 6 54026116 | 0.166933377 | MLIP | 1 169540901 | 0.2543832 | F5 | 12 112173266 | 0.214143051 | HECTD4 |
| 5 109815485 | 0.166930252 | MAN2A1 | 13 71838939 | 0.254311895 | DACH1 | 2_49983459 | 0.21411609 | NRXN1 |
| 5 109815737 | 0.166930252 | MAN2A1 | 17 10136930 | 0.254218069 | GAS7 | 6 151405694 | 0.214080587 | RMND1 |
| 2 170429448 | 0.166882596 | MYO3B | 8 29249432 | 0.254193254 | KIF13B | 10 96264542 | 0.214009377 | BLNK |
| 2 167009344 | 0.166870328 | XIRP2 | 6_34320249 | 0.254123652 | NUDT3 | 19_38557998 | 0.213970264 | RYR1 |
| 5 160770609 | 0.1668206 | ATP10B | 1 147164266 | 0.254111416 | PRKAB2 | 1 244584268 | 0.213710244 | C1orf101 |

| | | | | | | | | |
|---|---|---|---|---|---|---|---|---|
| 19_37962142 | 0.166787482 | SIPA1L3 | 1_35491544 | 0.254079913 | KIAA0319L | 6_70288860 | 0.213658191 | COL9A1 |
| 5_109800324 | 0.166768287 | MAN2A1 | 10_110595915 | 0.253987861 | SMC3 | 5_128970783 | 0.213571343 | SLC27A6 |
| 4_6463074 | 0.166760792 | PPP2R2C | 6_47708186 | 0.253819194 | GPR115 | 16_81359410 | 0.213532263 | GAN |
| 2_179966650 | 0.166759735 | CWC22 | 2_106146870 | 0.253668008 | UXS1 | 3_138556069 | 0.213497955 | CEP70 |
| 5_109833103 | 0.166724049 | MAN2A1 | 12_54420426 | 0.253657368 | LOC102724050 | 22_18127087 | 0.213466201 | TUBA8 |
| 2_71420610 | 0.166625845 | ZNF638 | 4_100494761 | 0.253629713 | EMCN | 17_19350348 | 0.213304428 | B9D1 |
| 5_109815955 | 0.166573322 | MAN2A1 | 11_46091629 | 0.253594993 | PHF21A | 6_29794862 | 0.213294447 | LOC554223 |
| 3_129496202 | 0.166523738 | IFT122 | 6_152306905 | 0.253577588 | SYNE1 | 2_55686762 | 0.213217352 | PNPT1 |
| 7_104847696 | 0.166523738 | LHFPL3 | 15_100294413 | 0.253513171 | ADAMTS17 | 1_244386694 | 0.21318712 | C1orf100 |
| 6_144585697 | 0.166497339 | UTRN | 9_40998400 | 0.253457112 | LOC100132352 | 1_54733760 | 0.213141656 | MROH7-TTC4 |
| 1_162863611 | 0.166454049 | C1orf110 | 13_107768162 | 0.253455041 | FAM155A | 6_29830857 | 0.213124879 | HLA-G |
| 1_162863910 | 0.166454049 | C1orf110 | 14_33340739 | 0.253447684 | NPAS3 | 3_33051426 | 0.21296029 | GLB1 |
| 8_10365920 | 0.16645293 | MSRA | 4_108573927 | 0.253385198 | RPL34-AS1 | 5_37135834 | 0.212895142 | C5orf42 |
| 2_170416627 | 0.166421635 | MYO3B | 9_113906573 | 0.253369142 | ZNF618 | 12_65762357 | 0.212886942 | RPSAP52 |
| 4_6462364 | 0.166419475 | PPP2R2C | 3_55060125 | 0.253337502 | CACNA2D3 | 6_70290507 | 0.212876602 | COL9A1 |
| 2_37264523 | 0.16631576 | PRKD3 | 7_64209344 | 0.253256634 | ZNF735P | 2_69972677 | 0.2128315 | PCBP1-AS1 |
| 1_96373058 | 0.16630404 | LOC101928241 | 8_71328985 | 0.253234902 | EYA1 | 6_99533155 | 0.212733282 | TSTD3 |
| 8_127079368 | 0.166290174 | PCAT2 | 1_13586685 | 0.2530909 | PDPN | 1_247967418 | 0.21257096 | OR2L13 |
| 7_78032816 | 0.166151333 | MAGI2 | 7_24854500 | 0.253079246 | OSBPL3 | 9_110480911 | 0.212565112 | SVEP1 |
| 15_97750038 | 0.16615108 | LINC00923 | 11_7863539 | 0.253038094 | LOC283299 | 5_178572630 | 0.212561998 | COL23A1 |
| 12_32894564 | 0.166131921 | PKP2 | 11_46072994 | 0.252792732 | PHF21A | 3_64613188 | 0.212557564 | ADAMTS9 |
| 8_133569387 | 0.166128981 | ST3GAL1 | 1_2244139 | 0.252791913 | SKI | 12_21039238 | 0.212529526 | SLCO1B7 |
| 14_78901341 | 0.166101354 | NRXN3 | 4_88009736 | 0.252707168 | PKD2 | 6_32641161 | 0.212492596 | HLA-DQA1 |
| 19_37973157 | 0.166008332 | SIPA1L3 | 11_45904894 | 0.252671533 | MAPK8IP1 | 14_63389932 | 0.212457729 | PPP2R5E |
| 2_206745733 | 0.165872155 | MDH1B | 2_178865541 | 0.252517024 | CCDC141 | 11_118559804 | 0.212415911 | IFT46 |
| 7_78031821 | 0.165798222 | MAGI2 | 17_38871150 | 0.252469491 | LASP1 | 11_118579545 | 0.212410195 | ARCN1 |
| 2_166934867 | 0.165795376 | XIRP2 | 12_299534 | 0.25245101 | KDM5A | 2_16638638 | 0.212410174 | FAM49A |
| 4_89754087 | 0.165775452 | SNCA | 2_170384567 | 0.252428049 | MYO3B | 15_49299747 | 0.212317656 | GALK2 |
| 5_109850287 | 0.165771344 | MAN2A1 | 13_70135694 | 0.252382984 | ATXN8OS | 10_96266765 | 0.212297658 | BLNK |
| 11_44821583 | 0.165689303 | TSPAN18 | 4_108745659 | 0.25238002 | ETNPPL | 3_138524388 | 0.212275691 | CEP70 |
| 22_19901968 | 0.165654726 | TXNRD2 | 11_131468729 | 0.252162206 | NTM | 14_63376621 | 0.212265417 | PPP2R5E |
| 13_28234822 | 0.165646189 | PAN3 | 3_195904263 | 0.252104381 | TNK2 | 7_120506163 | 0.212264401 | KCND2 |
| 13_28283073 | 0.165646189 | PAN3 | 7_6140404 | 0.251829321 | USP42 | 21_14191242 | 0.212150105 | LIPI |
| 22_49567866 | 0.16563972 | C22orf34 | 8_42328248 | 0.251817616 | IKBKB | 5_176620760 | 0.212096451 | SNCB |
| 22_49567903 | 0.16563972 | C22orf34 | 22_40489596 | 0.251813524 | MKL1 | 3_139995279 | 0.212079469 | CLSTN2 |
| 22_49568766 | 0.16563972 | C22orf34 | 22_40503950 | 0.251813524 | MKL1 | 14_63388857 | 0.212041073 | PPP2R5E |
| 22_49569242 | 0.16563972 | C22orf34 | 16_89997732 | 0.251751605 | AFG3L1P | 3_55873773 | 0.211923293 | ERC2 |
| 6_159223051 | 0.165636932 | FNDC1 | 11_67460987 | 0.251699981 | CABP4 | 10_60321197 | 0.211839044 | ANK3 |
| 22_49570279 | 0.165623846 | C22orf34 | 9_3495207 | 0.251699593 | RFX3 | 2_69972435 | 0.211718363 | PCBP1-AS1 |
| 1_65374766 | 0.165464177 | DNAJC6 | 1_35190463 | 0.251663797 | SFPQ | 8_72844774 | 0.211676075 | KCNB2 |
| 17_37867926 | 0.165453626 | YWHAEP7 | 1_35191063 | 0.251663797 | SFPQ | 19_38556464 | 0.211660709 | RYR1 |
| 10_17881885 | 0.165421677 | MRC1 | 16_87215357 | 0.251507227 | LOC101928708 | 2_182028788 | 0.211630713 | PPP1R1C |
| 6_159223048 | 0.165378526 | FNDC1 | 4_108758962 | 0.25148353 | ETNPPL | 6_32584308 | 0.21158735 | HLA-DRB1 |
| 2_71418874 | 0.165310358 | ZNF638 | 7_6263794 | 0.251468328 | CYTH3 | 9_9795888 | 0.211580418 | PTPRD |
| 1_65373260 | 0.165306943 | DNAJC6 | 15_25749019 | 0.251416182 | ATP10A | 8_96539617 | 0.211549981 | SDC2 |
| 9_131904942 | 0.165202672 | MED27 | 9_17338752 | 0.251320393 | CNTLN | 17_19416745 | 0.211479036 | RNF112 |
| 6_159219677 | 0.165158013 | FNDC1 | 1_155687151 | 0.251234017 | YY1AP1 | 6_30165050 | 0.211442581 | TRIM15 |
| 1_216995141 | 0.165147378 | ESRRG | 2_113902387 | 0.251108451 | ACTR3 | 15_97751769 | 0.211390167 | LINC00923 |
| 13_98238582 | 0.165134647 | FARP1 | 9_112867474 | 0.251091788 | SNX30 | 10_70095376 | 0.21138593 | H2AFY2 |
| 17_76679918 | 0.165111401 | MXRA7 | 2_232773911 | 0.251083333 | GIGYF2 | 18_13538530 | 0.211322193 | LDLRAD4 |
| 8_3025866 | 0.165073546 | CSMD1 | 22_48687200 | 0.251070949 | FAM19A5 | 2_50001321 | 0.211303014 | NRXN1 |
| 2_231326057 | 0.165021234 | ARMC9 | 14_61571777 | 0.25095362 | FLJ22447 | 1_244560790 | 0.211290313 | C1orf101 |
| 12_48351114 | 0.164995409 | ZNF641 | 6_34382799 | 0.250943587 | NUDT3 | 1_244569034 | 0.211290313 | C1orf101 |
| 12_49796870 | 0.164929219 | NCKAP5L | 15_80444330 | 0.25076486 | ARNT2 | 14_63399207 | 0.211172873 | PPP2R5E |
| 12_49796884 | 0.164929219 | NCKAP5L | 10_19530331 | 0.250735791 | MALRD1 | 4_56257180 | 0.210960232 | KIAA1211 |
| 7_104905016 | 0.164928165 | LHFPL3 | 9_113888684 | 0.250676724 | ZNF618 | 2_46700724 | 0.210952664 | SOCS5 |
| 7_44655631 | 0.164855145 | OGDH | 9_113929653 | 0.250676724 | ZNF618 | 22_48633508 | 0.210901567 | FAM19A5 |
| 15_27783102 | 0.164844242 | OCA2 | 9_113930720 | 0.250676724 | ZNF618 | 12_27336654 | 0.210838279 | ARNTL2 |
| 2_237406908 | 0.164830883 | COL6A3 | 9_113936350 | 0.250676724 | ZNF618 | 11_118573810 | 0.210786897 | ARCN1 |
| 1_162866801 | 0.164795745 | C1orf110 | 10_18463289 | 0.250478008 | CACNB2 | 7_90252832 | 0.210745337 | C7orf63 |
| 1_161044656 | 0.164750692 | USF1 | 1_85258126 | 0.250445596 | C1orf52 | 12_112310802 | 0.210730497 | HECTD4 |
| 4_89750136 | 0.16472264 | SNCA | 7_147256219 | 0.250306027 | CNTNAP2 | 12_112503550 | 0.210730497 | PTPN11 |
| 9_20375605 | 0.164689608 | MLLT3 | 6_34369876 | 0.250266737 | NUDT3 | 19_38557928 | 0.210719242 | RYR1 |
| 3_129544171 | 0.164619067 | H1FOO | 2_113936980 | 0.250240648 | ACTR3 | 8_124504847 | 0.210645349 | TATDN1 |
| 10_82003259 | 0.164548349 | NRG3 | 22_40628850 | 0.250225313 | MKL1 | 2_64120001 | 0.210630819 | PELI1 |
| 6_159228628 | 0.164548349 | FNDC1 | 8_71325586 | 0.25022077 | EYA1 | 3_138550151 | 0.210556513 | CEP70 |
| 6_159230173 | 0.164548349 | FNDC1 | 4_108583580 | 0.250208036 | RPL34-AS1 | 1_244587654 | 0.210527648 | C1orf101 |
| 6_159230215 | 0.164548349 | FNDC1 | 5_170025840 | 0.250144665 | DOCK2 | 1_244591199 | 0.210527648 | C1orf101 |
| 6_159230247 | 0.164548349 | FNDC1 | 1_35899115 | 0.250028266 | AGO1 | 1_244595038 | 0.210527648 | C1orf101 |
| 11_11452915 | 0.164523643 | GALNT18 | 12_6806182 | 0.24988802 | CD4 | 4_56252271 | 0.210511504 | KIAA1211 |
| 6_144593189 | 0.164494953 | UTRN | 2_46551129 | 0.2498193 | RHOQ | 2_55673803 | 0.210498112 | PNPT1 |
| 6_144601640 | 0.164494953 | UTRN | 3_160974797 | 0.249761652 | PPM1L | 2_55390119 | 0.210471243 | CCDC88A |
| 6_144606280 | 0.164494953 | UTRN | 15_48558663 | 0.249599619 | FBN1 | 18_9794980 | 0.210466866 | RAB31 |
| 6_144606880 | 0.164494953 | UTRN | 15_48628860 | 0.249599619 | FBN1 | 10_96320202 | 0.21023367 | DNTT |
| 3_33120237 | 0.164473683 | CRTAP | 15_48487333 | 0.24957847 | FBN1 | 3_24228772 | 0.2102049 | THRB |
| 3_33121040 | 0.164473683 | CRTAP | 2_113933881 | 0.249408138 | ACTR3 | 21_14193717 | 0.210141983 | LIPI |
| 3_33121130 | 0.164473683 | CRTAP | 15_48478956 | 0.249401794 | FBN1 | 5_19553625 | 0.210118615 | CDH18 |
| 8_3027060 | 0.164460785 | CSMD1 | 16_89862104 | 0.249342562 | SPIRE2 | 7_134133934 | 0.210103553 | LRGUK |
| 12_29656339 | 0.16444027 | TMTC1 | 6_107588966 | 0.249304717 | SOBP | 2_10731658 | 0.210085648 | ATP6V1C2 |
| 9_102645346 | 0.164373302 | LINC00587 | 19_46491702 | 0.249290321 | PNMAL2 | 3_10147368 | 0.210047367 | VHL |
| 12_48345980 | 0.16432124 | ZNF641 | 6_43618404 | 0.24926272 | POLH | 11_118538178 | 0.209984529 | TMEM25 |
| 2_170411684 | 0.164275319 | MYO3B | 21_33655552 | 0.249251701 | ITSN1 | 1_244564359 | 0.209952986 | C1orf101 |
| 10_99990754 | 0.164233346 | DNMBP | 1_244410947 | 0.249112572 | ADSS | 1_244583392 | 0.209952986 | C1orf101 |
| 8_120616435 | 0.164228876 | SNTB1 | 9_72933599 | 0.248944916 | ALDH1A1 | 1_244584224 | 0.209952986 | C1orf101 |
| 12_131185491 | 0.164222657 | LINC01257 | 2_113924081 | 0.248878996 | ACTR3 | 1_244585155 | 0.209952986 | C1orf101 |
| 12_131189181 | 0.164222657 | LINC01257 | 1_159075671 | 0.248834358 | AIM2 | 19_15695508 | 0.209939169 | CYP4F12 |
| 6_166835010 | 0.164168307 | RPS6KA2 | 10_25469945 | 0.248796253 | GPR158 | 2_70173364 | 0.209913342 | C2orf42 |
| 1_44548269 | 0.164167818 | RNF220 | 7_6150897 | 0.248723213 | USP42 | 3_138463176 | 0.209910368 | ESYT3 |
| 1_62119982 | 0.164154653 | INADL | 1_204292519 | 0.248653 | PLEKHA6 | 3_138574656 | 0.209910368 | CEP70 |
| 1_62120507 | 0.164154653 | INADL | 4_39506406 | 0.248567363 | UGDH | 6_39838497 | 0.20987955 | DAAM2 |
| 6_159223121 | 0.164123074 | FNDC1 | 9_17161730 | 0.24854583 | CNTLN | 8_18597083 | 0.209834047 | PSD3 |
| 2_179966274 | 0.164112363 | CWC22 | 2_227172535 | 0.248394377 | COL4A3 | 8_72852648 | 0.209807883 | KCNB2 |
| 6_144637674 | 0.164107114 | UTRN | 1_244414450 | 0.248379645 | ADSS | 11_118476148 | 0.20967028 | KMT2A |
| 8_10394205 | 0.164091998 | MSRA | 22_46693377 | 0.248324728 | CERK | 10_96318244 | 0.209612923 | DNTT |
| 5_178499530 | 0.164061614 | COL23A1 | 8_9576472 | 0.248319907 | TNKS | 1_244584037 | 0.209576261 | C1orf101 |
| 5_178501403 | 0.164061614 | COL23A1 | 1_222566646 | 0.248071247 | TAF1A | 2_70002225 | 0.209370694 | PCBP1-AS1 |
| 22_40495044 | 0.164025672 | MKL1 | 4_56257378 | 0.248049747 | KIAA1211 | 15_47302707 | 0.209348972 | SEMA6D |

| | | | | | | | | |
|---|---|---|---|---|---|---|---|---|
| 1_65401836 | 0.163916008 | DNAJC6 | 15_48614260 | 0.248000838 | FBN1 | 2_16638546 | 0.209345041 | FAM49A |
| 6_74240248 | 0.16391348 | LOC101928516 | 2_97175759 | 0.247999175 | ANKRD36 | 2_47069163 | 0.209342382 | TTC7A |
| 13_28235696 | 0.163899583 | PAN3 | 3_55068437 | 0.24786648 | CACNA2D3 | 5_5435668 | 0.2092405 | KIAA0947 |
| 15_66141289 | 0.16388037 | MEGF11 | 12_53508257 | 0.247691275 | ATF7 | 9_77618779 | 0.209231418 | GNA14 |
| 6_166811784 | 0.163848133 | RPS6KA2 | 10_50313152 | 0.247628643 | SGMS1 | 1_244575932 | 0.2091259 | C1orf101 |
| 9_133161541 | 0.163787733 | GBGT1 | 10_50314354 | 0.247628643 | SGMS1 | 21_42133953 | 0.209123837 | UMODL1 |
| 1_169481026 | 0.163783574 | SLC19A2 | 14_88082523 | 0.247524802 | LINC01146 | 6_36595205 | 0.209073026 | SRSF3 |
| 1_231187003 | 0.163779068 | LOC149373 | 22_40523555 | 0.247136224 | LOC101927257 | 8_6611402 | 0.208988938 | MCPH1 |
| 1_231188511 | 0.163779068 | TRIM67 | 20_42208176 | 0.247120547 | PTPRT | 1_244560439 | 0.208981539 | C1orf101 |
| 5_160771419 | 0.163689782 | ATP10B | 9_113912005 | 0.246986888 | ZNF618 | 20_20030359 | 0.208937375 | NAA20 |
| 5_160771421 | 0.163689782 | ATP10B | 15_48527347 | 0.246966704 | FBN1 | 2_64118268 | 0.208861795 | PELI1 |
| 4_6794696 | 0.163659341 | KIAA0232 | 1_35965340 | 0.246923156 | AGO3 | 2_64119530 | 0.208861795 | PELI1 |
| 2_163613534 | 0.163641624 | FIGN | 9_3375692 | 0.246862044 | RFX3 | 2_64123280 | 0.208861795 | PELI1 |
| 2_71434499 | 0.163602841 | ZNF638 | 15_25736579 | 0.246845569 | ATP10A | 19_17424737 | 0.208831987 | MVB12A |
| 4_6459024 | 0.163597738 | PPP2R2C | 7_104905216 | 0.24681495 | LHFPL3 | 8_125122173 | 0.208812064 | NSMCE2 |
| 19_37960872 | 0.163564489 | SIPA1L3 | 12_338207 | 0.246754293 | KDM5A | 7_120350103 | 0.208783902 | KCND2 |
| 7_93504541 | 0.163457181 | CALCR | 10_60421123 | 0.246660467 | ANK3 | 2_54867684 | 0.208760158 | EML6 |
| 7_93504542 | 0.163457181 | CALCR | 13_97297765 | 0.246570631 | MBNL2 | 2_229142424 | 0.208755859 | PID1 |
| 19_37956680 | 0.163454984 | SIPA1L3 | 9_113894922 | 0.246550577 | ZNF618 | 3_54549473 | 0.20871125 | CACNA2D3 |
| 5_109747980 | 0.163438629 | MAN2A1 | 9_113897470 | 0.246550577 | ZNF618 | 12_131906561 | 0.208686007 | ULK1 |
| 6_144555802 | 0.163431771 | UTRN | 6_100407670 | 0.246478865 | SIM1 | 11_118534722 | 0.208672907 | TMEM25 |
| 6_144559412 | 0.163431771 | UTRN | 1_169548147 | 0.246440413 | F5 | 2_70038664 | 0.208672764 | PCBP1-AS1 |
| 7_129155147 | 0.163408501 | TSPAN33 | 10_17881885 | 0.246417251 | MRC1 | 20_63453620 | 0.208648891 | KCNQ2 |
| 14_78906048 | 0.163393946 | NRXN3 | 7_104840075 | 0.246189282 | LHFPL3 | 22_48633512 | 0.208638834 | FAM19A5 |
| 6_87799362 | 0.163358869 | LOC101928911 | 7_24935485 | 0.246159373 | OSBPL3 | 14_63389094 | 0.208609523 | PPP2R5E |
| 1_146811588 | 0.16333711 | HYDIN2 | 5_168907845 | 0.246140026 | SLIT3 | 10_28858549 | 0.208597566 | C10orf126 |
| 6_166837424 | 0.163270041 | RPS6KA2 | 3_71067589 | 0.246128488 | FOXP1 | 2_55553089 | 0.208564637 | SMEK2 |
| 9_14906870 | 0.163254112 | FREM1 | 11_88823142 | 0.246123909 | GRM5 | 4_93270642 | 0.208556664 | GRID2 |
| 6_166822803 | 0.163197604 | RPS6KA2 | 2_30606434 | 0.246084649 | LCLAT1 | 1_234116572 | 0.208546697 | SLC35F3 |
| 21_36260244 | 0.163177538 | DOPEY2 | 1_169529597 | 0.24606873 | F5 | 1_144426562 | 0.208505993 | NBPF15 |
| 13_28270019 | 0.16317493 | PAN3 | 8_96522805 | 0.245892673 | SDC2 | 1_116560027 | 0.208475815 | CD58 |
| 13_28270024 | 0.16317493 | PAN3 | 1_172145324 | 0.245843894 | DNM3 | 1_220036534 | 0.208466635 | EPRS |
| 13_28291260 | 0.16317493 | PAN3 | 13_107724434 | 0.245770853 | FAM155A | 1_161928007 | 0.208444406 | ATF6 |
| 13_99864774 | 0.163126363 | CLYBL | 1_36044355 | 0.245646208 | AGO3 | 9_36984348 | 0.208444247 | PAX5 |
| 6_159236543 | 0.163108499 | FNDC1 | 1_244448260 | 0.24563127 | ADSS | 12_95752338 | 0.2084298 | NTN4 |
| 12_49832220 | 0.163097206 | BCDIN3D-AS1 | 14_33433730 | 0.24563127 | NPAS3 | 7_74908521 | 0.208197549 | PMS2P5 |
| 12_49837539 | 0.163097206 | BCDIN3D | 7_6163740 | 0.24559529 | CYTH3 | 20_63458343 | 0.208190401 | KCNQ2 |
| 12_49838386 | 0.163097206 | BCDIN3D | 22_40499806 | 0.245418239 | MKL1 | 11_78970208 | 0.208175382 | TENM4 |
| 20_19999129 | 0.16309516 | RIN2 | 11_129451131 | 0.245395483 | BARX2 | 18_63712166 | 0.208161147 | SERPINB11 |
| 7_90263948 | 0.163089248 | C7orf63 | 12_304912 | 0.24532927 | KDM5A | 18_63715608 | 0.208161147 | SERPINB11 |
| 7_88065262 | 0.16307715 | ADAM22 | 7_6135445 | 0.245144932 | USP42 | 2_113924081 | 0.208125592 | ACTR3 |
| 10_12582075 | 0.163044225 | CAMK1D | 1_231191920 | 0.245078164 | TRIM67 | 7_149873635 | 0.208117838 | ATP6V0E2 |
| 22_49580040 | 0.163044094 | C22orf34 | 3_155627215 | 0.24505183 | PLCH1 | 6_30342723 | 0.208037998 | TRIM39 |
| 5_109698681 | 0.16302925 | MAN2A1 | 3_155627427 | 0.24505183 | PLCH1 | 7_74221411 | 0.207992504 | LAT2 |
| 5_109702928 | 0.16302925 | MAN2A1 | 3_155630628 | 0.24505183 | PLCH1 | 5_10456581 | 0.207975221 | ROPN1L |
| 4_30999470 | 0.162944403 | PCDH7 | 15_100283402 | 0.244992654 | ADAMTS17 | 10_96321351 | 0.207965104 | DNTT |
| 8_72883955 | 0.162931687 | KCNB2 | 4_186607496 | 0.244929649 | FAT1 | 2_55690967 | 0.207920224 | PNPT1 |
| 15_33851000 | 0.162912051 | RYR3 | 4_108749833 | 0.244673658 | ETNPPL | 18_58872742 | 0.207914964 | ZNF532 |
| 17_79732653 | 0.162892405 | ENPP7 | 15_47749075 | 0.244650932 | SEMA6D | 2_70072594 | 0.207729217 | PCBP1-AS1 |
| 13_28262422 | 0.162882864 | PAN3 | 2_205442291 | 0.244629388 | PARD3B | 9_110439246 | 0.207722748 | SVEP1 |
| 1_162863745 | 0.162761638 | C1orf110 | 16_82928843 | 0.244619556 | CDH13 | 12_27282653 | 0.207717826 | STK38L |
| 1_162865325 | 0.162761638 | C1orf110 | 3_124236611 | 0.244596577 | KALRN | 3_138578642 | 0.207690345 | CEP70 |
| 1_162865417 | 0.162761638 | C1orf110 | 1_244386541 | 0.244511819 | C1orf100 | 8_72848721 | 0.207684999 | KCNB2 |
| 4_108747356 | 0.162723082 | ETNPPL | 7_71672244 | 0.244486383 | WBSCR17 | 8_72852698 | 0.207684999 | KCNB2 |
| 3_61655566 | 0.162664994 | PTPRG | 9_3503793 | 0.244468886 | RFX3 | 6_32665685 | 0.207683039 | HLA-DQB1 |
| 7_143468927 | 0.162659255 | EPHA1-AS1 | 6_34391213 | 0.244428711 | NUDT3 | 11_118446245 | 0.207649149 | KMT2A |
| 20_53949615 | 0.162632104 | BCAS1 | 22_40536037 | 0.244368256 | MKL1 | 2_50043279 | 0.20763366 | NRXN1 |
| 1_9600241 | 0.162627251 | TMEM201 | 22_19205881 | 0.244278433 | CLTCL1 | 20_20021148 | 0.207626673 | NAA20 |
| 10_107116723 | 0.162625236 | SORCS1 | 19_948532 | 0.244204105 | ARID3A | 10_93453921 | 0.207567278 | MYOF |
| 1_231191320 | 0.162600761 | TRIM67 | 3_124237165 | 0.244201791 | KALRN | 6_151357557 | 0.207498373 | ZBTB2 |
| 7_88006908 | 0.162538621 | ADAM22 | 7_149291467 | 0.244108059 | LOC155060 | 2_229142484 | 0.207370625 | PID1 |
| 19_37981953 | 0.162527747 | SIPA1L3 | 7_102446755 | 0.24409196 | ORAI2 | 2_50006214 | 0.207314193 | NRXN1 |
| 7_93513473 | 0.16249809 | CALCR | 19_2596673 | 0.244027498 | GNG7 | 8_72843238 | 0.207271131 | KCNB2 |
| 6_166817763 | 0.16244205 | RPS6KA2 | 8_40644881 | 0.244026235 | ZMAT4 | 2_64127107 | 0.20717762 | PELI1 |
| 3_63416860 | 0.162437917 | SYNPR | 4_108587685 | 0.244010574 | RPL34-AS1 | 21_34527578 | 0.207107684 | RCAN1 |
| 9_130408132 | 0.162412411 | HMCN2 | 1_231203967 | 0.243830449 | TRIM67 | 17_5565330 | 0.207047199 | NLRP1 |
| 2_208145545 | 0.162385316 | CRYGB | 10_50388472 | 0.243729565 | SGMS1 | 7_120492002 | 0.207028292 | KCND2 |
| 5_109853939 | 0.162363188 | MAN2A1 | 15_49171095 | 0.243729565 | GALK2 | 10_82649311 | 0.207017343 | NRG3 |
| 22_32722179 | 0.16232472 | SYN3 | 4_108753764 | 0.243695302 | ETNPPL | 11_118552153 | 0.207016921 | IFT46 |
| 6_159213110 | 0.162280793 | FNDC1 | 22_40491081 | 0.243649107 | MKL1 | 4_88060385 | 0.206995294 | PKD2 |
| 1_65377208 | 0.162279798 | DNAJC6 | 4_38473796 | 0.243452023 | LINC01258 | 7_15495816 | 0.206948594 | AGMO |
| 5_106840295 | 0.162274012 | LOC102467213 | 4_38473802 | 0.243452023 | LINC01258 | 3_64620179 | 0.206895781 | ADAMTS9 |
| 5_106841269 | 0.162274012 | LOC102467213 | 9_98400016 | 0.243312723 | GABBR2 | 19_38549544 | 0.206851902 | RYR1 |
| 2_73291739 | 0.162265536 | EGR4 | 9_17455967 | 0.243223176 | CNTLN | 8_96541728 | 0.206841923 | SDC2 |
| 1_159925320 | 0.162240138 | TAGLN2 | 4_108753768 | 0.243157676 | ETNPPL | 12_7390128 | 0.206777814 | CD163L1 |
| 6_159211218 | 0.162235132 | FNDC1 | 7_71672766 | 0.243120261 | WBSCR17 | 7_72401209 | 0.206777007 | CALN1 |
| 6_170404415 | 0.162204247 | FAM120B | 12_251190 | 0.243117713 | SLC6A13 | 2_75111717 | 0.206756356 | TACR1 |
| 10_59702893 | 0.162200732 | SLC16A9 | 11_101030247 | 0.243067981 | PGR | 2_170465169 | 0.206731031 | MYO3B |
| 7_154871014 | 0.162196534 | DPP6 | 11_101033504 | 0.243067981 | PGR | 7_26846953 | 0.206508578 | SKAP2 |
| 3_129453023 | 0.16218181 | IFT122 | 20_59602560 | 0.242924301 | PHACTR3 | 6_99391503 | 0.206499019 | COQ3 |
| 1_36053026 | 0.162164789 | AGO3 | 1_77475171 | 0.242851563 | AK5 | 1_244386541 | 0.206486345 | C1orf100 |
| 1_155795017 | 0.16213356 | GON4L | 14_61547116 | 0.242686733 | PRKCH | 8_11431917 | 0.206469375 | C8orf12 |
| 9_81607079 | 0.162119146 | TLE1 | 17_10138358 | 0.242612092 | GAS7 | 7_26851208 | 0.206463816 | SKAP2 |
| 2_60387442 | 0.162112529 | MIR4432 | 2_141074589 | 0.242556771 | LRP1B | 3_64634695 | 0.206451105 | ADAMTS9 |
| 1_85276474 | 0.162061322 | BCL10 | 9_17211885 | 0.24246341 | CNTLN | 10_45442799 | 0.206428328 | ALOX5 |
| 1_85277178 | 0.162061322 | LOC646626 | 7_6180012 | 0.242452318 | CYTH3 | 21_13552718 | 0.206388729 | LOC102724188 |
| 15_97749977 | 0.162034113 | LINC00923 | 13_43403847 | 0.242411962 | ENOX1 | 7_91175907 | 0.206378367 | CDK14 |
| 5_106846357 | 0.162024152 | LOC102467213 | 5_3443889 | 0.24233245 | LINC01019 | 1_244577216 | 0.206335092 | C1orf101 |
| 16_89126982 | 0.162004893 | ACSF3 | 12_109767454 | 0.242255161 | FAM222A | 1_244584822 | 0.206335092 | C1orf101 |
| 17_5909114 | 0.161982692 | LOC339166 | 9_3477522 | 0.242153183 | RFX3 | 6_30908661 | 0.206243925 | GTF2H4 |
| 2_206743871 | 0.161978296 | MDH1B | 2_121518290 | 0.242148847 | CLASP1 | 12_112505674 | 0.206226837 | PTPN11 |
| 12_131191630 | 0.161948928 | LINC01257 | 20_61722641 | 0.242137357 | CDH4 | 8_72838511 | 0.206193098 | KCNB2 |
| 6_159238491 | 0.16190953 | FNDC1 | 5_9203802 | 0.242127443 | SEMA5A | 8_20149601 | 0.206157392 | SLC18A1 |
| 2_167009642 | 0.161898208 | XIRP2 | 9_17376115 | 0.242037664 | CNTLN | 12_112356264 | 0.206103984 | HECTD4 |
| 6_21743235 | 0.161888805 | CASC15 | 16_23684971 | 0.24202649 | PLK1 | 7_120496137 | 0.206102986 | KCND2 |
| 6_128486781 | 0.161852413 | PTPRK | 6_34402129 | 0.242025581 | RPS10-NUDT3 | 6_36495850 | 0.206040027 | STK38 |
| 6_144533686 | 0.16178255 | UTRN | 8_17349278 | 0.242012931 | MTMR7 | 12_95706818 | 0.206023813 | NTN4 |

| | | | | | | | | |
|---|---|---|---|---|---|---|---|---|
| 13_28160642 | 0.161778267 | PAN3 | 9_3515185 | 0.241961792 | RFX3 | 2_55655537 | 0.205921555 | PNPT1 |
| 1_83979083 | 0.161769794 | TTLL7 | 7_147251289 | 0.241950061 | CNTNAP2 | 2_55680222 | 0.205892765 | PNPT1 |
| 3_123597574 | 0.161734857 | MYLK-AS1 | 15_48474981 | 0.241752196 | FBN1 | 6_5653545 | 0.205891156 | FARS2 |
| 2_206746079 | 0.161699462 | MDH1B | 5_74732134 | 0.241721386 | GFM2 | 1_204331900 | 0.205756903 | PLEKHA6 |
| 5_109739808 | 0.161694451 | MAN2A1 | 15_48489009 | 0.241684255 | FBN1 | 2_50003021 | 0.205705697 | NRXN1 |
| 6_166820842 | 0.161647841 | RPS6KA2 | 4_89750136 | 0.241664517 | SNCA | 12_27259401 | 0.205683101 | STK38L |
| 6_166823893 | 0.161647841 | RPS6KA2 | 2_132591267 | 0.241654575 | GPR39 | 6_32660495 | 0.205582921 | HLA-DQB1 |
| 7_94417954 | 0.161647841 | COL1A2 | 1_35064311 | 0.241635416 | ZMYM1 | 3_33056183 | 0.205580343 | GLB1 |
| 12_48732127 | 0.161622929 | LINC00935 | 1_48381908 | 0.241579553 | SPATA6 | 17_19241957 | 0.205577414 | EPN2 |
| 2_71135404 | 0.161620917 | MPHOSPH10 | 4_78796454 | 0.24149787 | BMP2K | 2_70080474 | 0.205573939 | PCBP1-AS1 |
| 3_33119863 | 0.161609353 | CRTAP | 15_25736030 | 0.241457847 | ATP10A | 5_151559619 | 0.205559449 | FAT2 |
| 10_16501359 | 0.161608483 | PTER | 12_53580559 | 0.241448876 | ATF7 | 11_118579331 | 0.205443476 | ARCN1 |
| 12_54972163 | 0.161607562 | TESPA1 | 1_231191320 | 0.241421252 | TRIM67 | 1_244588236 | 0.205431195 | C1orf101 |
| 3_33127444 | 0.161576668 | CRTAP | 15_48626081 | 0.241418062 | FBN1 | 5_178390998 | 0.205427278 | COL23A1 |
| 1_244578791 | 0.161542742 | C1orf101 | 6_34300330 | 0.241403163 | NUDT3 | 7_40843277 | 0.205425604 | SUGCT |
| 6_166802946 | 0.161534358 | RPS6KA2 | 20_46262228 | 0.241286767 | CDH22 | 6_5653761 | 0.205371371 | FARS2 |
| 1_62118004 | 0.161524497 | INADL | 1_147155343 | 0.241275354 | PRKAB2 | 2_16638892 | 0.205336272 | FAM49A |
| 4_31002319 | 0.161524108 | PCDH7 | 3_14178523 | 0.241260916 | XPC | 10_54575062 | 0.205317974 | PCDH15 |
| 7_93495438 | 0.161515425 | CALCR | 2_170386740 | 0.241199587 | MYO3B | 12_27344159 | 0.205289434 | ARNTL2 |
| 7_93497033 | 0.161515425 | CALCR | 7_94954863 | 0.24116692 | PPP1R9A | 18_12427698 | 0.205250275 | SLMO1 |
| 7_93513253 | 0.161515425 | CALCR | 5_119521802 | 0.241143116 | HSD17B4 | 1_21892806 | 0.205226928 | HSPG2 |
| 20_46957379 | 0.161487873 | EYA2 | 9_17211720 | 0.241125214 | CNTLN | 13_102162007 | 0.205193206 | FGF14 |
| 20_46957650 | 0.161487873 | EYA2 | 2_170384201 | 0.241035194 | MYO3B | 6_29943579 | 0.205182951 | HLA-A |
| 5_14838800 | 0.161474316 | ANKH | 12_377818 | 0.24096616 | KDM5A | 2_29072523 | 0.205130453 | C2orf71 |
| 19_37955140 | 0.161467615 | SIPA1L3 | 4_108751603 | 0.240877771 | ETNPPL | 3_138621065 | 0.205114859 | FAIM |
| 6_166843892 | 0.161334364 | RPS6KA2 | 7_6151315 | 0.240768191 | USP42 | 8_72845778 | 0.205054542 | KCNB2 |
| 22_32667821 | 0.161299495 | SYN3 | 1_202217148 | 0.240747424 | LGR6 | 8_72850818 | 0.205054542 | KCNB2 |
| 20_52977076 | 0.16124233 | TSHZ2 | 3_33114290 | 0.240711736 | CRTAP | 8_72859286 | 0.205054542 | KCNB2 |
| 1_111855365 | 0.161130247 | KCND3 | 15_80474125 | 0.240694824 | ARNT2 | 6_30052897 | 0.205012724 | ZNRD1-AS1 |
| 8_72818053 | 0.161103637 | KCNB2 | 1_147164191 | 0.240635481 | PRKAB2 | 3_64610544 | 0.205009582 | ADAMTS9 |
| 4_89752690 | 0.161093197 | SNCA | 3_129478272 | 0.24044764 | IFT122 | 4_164055264 | 0.205000497 | 1-Mar |
| 6_155435231 | 0.161055171 | NOX3 | 6_133644638 | 0.240393057 | EYA4-AS1 | 10_19848598 | 0.204964279 | PLXDC2 |
| 7_104875594 | 0.161040014 | LHFPL3 | 1_35192273 | 0.240345616 | SFPQ | 7_120505856 | 0.204879535 | KCND2 |
| 20_46962912 | 0.161033615 | EYA2 | 16_83191538 | 0.240073666 | CDH13 | 13_22262406 | 0.204862531 | LINC00540 |
| 19_13410544 | 0.160985124 | CACNA1A | 4_39520114 | 0.239974432 | UGDH | 20_46197351 | 0.204856671 | CDH22 |
| 19_13410546 | 0.160985124 | CACNA1A | 8_17175229 | 0.239828132 | ZDHHC2 | 11_118565364 | 0.204832913 | IFT46 |
| 2_71391300 | 0.160968305 | ZNF638 | 12_81462288 | 0.23976981 | PPFIA2 | 2_63904496 | 0.204822477 | VPS54 |
| 10_91959510 | 0.160951784 | BTAF1 | 9_3480671 | 0.239769761 | RFX3 | 3_8421876 | 0.20468175 | LMCD1-AS1 |
| 6_144629855 | 0.160895387 | UTRN | 10_79256182 | 0.239742147 | ZMIZ1 | 3_126455995 | 0.204643683 | ZXDC |
| 6_21744747 | 0.160895387 | CASC15 | 6_75768283 | 0.239703232 | MYO6 | 6_16272842 | 0.204579989 | GMPR |
| 7_104830017 | 0.160877911 | LHFPL3 | 3_192365116 | 0.239700441 | FGF12 | 21_21251845 | 0.204523784 | NCAM2 |
| 6_87806124 | 0.160874018 | LOC101928911 | 1_231186471 | 0.239647947 | LOC149373 | 5_5435666 | 0.204444041 | KIAA0947 |
| 5_109755916 | 0.160842696 | MAN2A1 | 3_192362162 | 0.239539823 | FGF12 | 2_16632926 | 0.204443672 | FAM49A |
| 20_53048770 | 0.160758345 | TSHZ2 | 6_34383990 | 0.239423469 | NUDT3 | 6_32665742 | 0.204440578 | HLA-DQB1 |
| 12_12435414 | 0.160754974 | LOH12CR1 | 7_6144205 | 0.239385674 | USP42 | 10_117259666 | 0.204434176 | SLC18A2 |
| 4_6811778 | 0.160744579 | KIAA0232 | 7_6151446 | 0.239385674 | USP42 | 2_231315204 | 0.204403816 | ARMC9 |
| 3_55061845 | 0.160742437 | CACNA2D3 | 18_52611203 | 0.239353442 | DCC | 9_94354060 | 0.204299781 | LOC100132077 |
| 6_159234886 | 0.160724675 | FNDC1 | 15_25733987 | 0.239328509 | ATP10A | 2_70047879 | 0.204288465 | PCBP1-AS1 |
| 6_159235600 | 0.160724675 | FNDC1 | 3_14176372 | 0.239328509 | XPC | 8_98085388 | 0.204234769 | ERICH5 |
| 6_159236996 | 0.160724675 | FNDC1 | 16_87219255 | 0.239281238 | LOC101928708 | 6_151408446 | 0.204138104 | RMND1 |
| 6_159237010 | 0.160724675 | FNDC1 | 16_87219261 | 0.239281238 | LOC101928708 | 14_63379385 | 0.204076746 | PPP2R5E |
| 6_159238449 | 0.160724675 | FNDC1 | 2_231313819 | 0.239166323 | ARMC9 | 1_244589240 | 0.204047295 | C1orf101 |
| 6_159239116 | 0.160724675 | FNDC1 | 1_244436150 | 0.239160109 | ADSS | 1_244591659 | 0.204047295 | C1orf101 |
| 6_159241032 | 0.160724675 | FNDC1 | 13_70097650 | 0.239156306 | KLHL1 | 2_50036167 | 0.204021808 | NRXN1 |
| 6_159241467 | 0.160724675 | FNDC1 | 4_38479802 | 0.239154681 | LINC01258 | 1_244565144 | 0.203962208 | C1orf101 |
| 6_159243484 | 0.160724675 | FNDC1 | 14_61560972 | 0.239112776 | LOC101927780 | 10_96260794 | 0.203883757 | BLNK |
| 18_53277606 | 0.160708197 | DCC | 16_89716148 | 0.239061706 | VPS9D1 | 5_10456224 | 0.203882454 | ROPN1L |
| 16_89868478 | 0.160684797 | SPIRE2 | 1_35989999 | 0.239040963 | AGO3 | 1_22004046 | 0.203861942 | CELA3A |
| 20_46961769 | 0.160677743 | EYA2 | 2_33150633 | 0.239031622 | LTBP1 | 17_75051195 | 0.20380738 | KCTD2 |
| 5_160771099 | 0.160674487 | ATP10B | 10_50448719 | 0.239004043 | SGMS1 | 2_50013117 | 0.20376393 | NRXN1 |
| 5_160771100 | 0.160674487 | ATP10B | 9_113911628 | 0.238907747 | ZNF618 | 8_72843340 | 0.203754769 | KCNB2 |
| 14_78908211 | 0.160645593 | NRXN3 | 16_89868478 | 0.23889614 | SPIRE2 | 11_118465212 | 0.203686375 | KMT2A |
| 17_5911626 | 0.160635857 | LOC339166 | 3_55062901 | 0.238876236 | CACNA2D3 | 2_75122295 | 0.203677604 | TACR1 |
| 6_159221931 | 0.160600747 | FNDC1 | 8_71328913 | 0.23885105 | EYA1 | 6_30013973 | 0.203646511 | ZNRD1-AS1 |
| 3_11823701 | 0.160582856 | TAMM41 | 1_954915 | 0.23882216 | NOC2L | 2_64118511 | 0.203638224 | PELI1 |
| 7_104850790 | 0.160569888 | LHFPL3 | 10_60548761 | 0.238757734 | ANK3 | 2_70032363 | 0.203613354 | PCBP1-AS1 |
| 22_49565404 | 0.160537881 | C22orf34 | 10_50316168 | 0.238528894 | SGMS1 | 2_54865789 | 0.203571377 | EML6 |
| 22_49566581 | 0.160537881 | C22orf34 | 17_65105262 | 0.238525664 | LOC100507002 | 21_13552713 | 0.203539296 | LOC102724188 |
| 9_4739005 | 0.160522153 | AK3 | 17_65105263 | 0.238525664 | LOC100507002 | 6_30259469 | 0.203523707 | HCG17 |
| 12_26825947 | 0.160513678 | ITPR2 | 6_101720722 | 0.238522087 | GRIK2 | 3_41494312 | 0.203478219 | ULK4 |
| 7_93490806 | 0.160512643 | CALCR | 20_58494266 | 0.238380832 | APCDD1L | 3_41494316 | 0.203478219 | ULK4 |
| 7_93493384 | 0.160512643 | CALCR | 20_58494270 | 0.238380832 | APCDD1L | 4_183188686 | 0.203449617 | WWC2 |
| 7_93493649 | 0.160512643 | CALCR | 20_58494274 | 0.238380832 | APCDD1L | 1_245913373 | 0.203448325 | SMYD3 |
| 7_93496643 | 0.160512643 | CALCR | 20_58494275 | 0.238380832 | APCDD1L | 2_70010966 | 0.203438727 | PCBP1-AS1 |
| 7_93498045 | 0.160512643 | CALCR | 17_45797286 | 0.238376219 | CRHR1 | 3_138556728 | 0.203437539 | CEP70 |
| 4_165958980 | 0.160387004 | TLL1 | 14_61556546 | 0.238354518 | LOC101927780 | 4_188091302 | 0.203320632 | TRIML2 |
| 1_38927359 | 0.160362365 | RHBDL2 | 13_107288431 | 0.238333612 | FAM155A | 4_188091304 | 0.203320632 | TRIML2 |
| 2_206746533 | 0.160319312 | MDH1B | 22_40483839 | 0.238253795 | MKL1 | 6_70291111 | 0.203317701 | COL9A1 |
| 9_131873617 | 0.16027335 | MED27 | 1_231208537 | 0.238199684 | TRIM67 | 2_50006468 | 0.20331726 | NRXN1 |
| 8_10413808 | 0.160216914 | MSRA | 1_231187947 | 0.237928951 | TRIM67 | 4_164040095 | 0.203315602 | 1-Mar |
| 5_106854457 | 0.160215687 | LOC102467213 | 1_231190090 | 0.237928951 | TRIM67 | 15_47307958 | 0.203249585 | SEMA6D |
| 22_32603844 | 0.160176475 | SYN3 | 1_231200663 | 0.237928951 | TRIM67 | 10_96312784 | 0.203224523 | DNTT |
| 3_697176 | 0.16016801 | LINC01266 | 1_231200785 | 0.237928951 | TRIM67 | 19_16034613 | 0.203149844 | LINC00905 |
| 5_149598810 | 0.160156494 | ARHGEF37 | 12_10216408 | 0.237875118 | GABARAPL1 | 12_112070638 | 0.2031351 | NAA25 |
| 2_71378103 | 0.160077862 | ZNF638 | 6_100420845 | 0.237789716 | SIM1 | 12_111931943 | 0.203102417 | TMEM116 |
| 6_16314149 | 0.160052392 | ATXN1 | 6_100420903 | 0.237789716 | SIM1 | 12_112006737 | 0.203102417 | TMEM116 |
| 4_175739671 | 0.160051224 | GPM6A | 16_87215517 | 0.237789042 | LOC101928708 | 2_69986381 | 0.203098367 | PCBP1-AS1 |
| 1_205729947 | 0.16004071 | NUCKS1 | 3_125337718 | 0.237773423 | ZNF148 | 10_96255250 | 0.20309091 | BLNK |
| 19_38207620 | 0.160028208 | SIPA1L3 | 11_45922557 | 0.237714916 | GYLTL1B | 20_20049044 | 0.202888366 | CRNKL1 |
| 6_159234641 | 0.160017295 | FNDC1 | 8_14656325 | 0.237706952 | SGCZ | 9_110482151 | 0.202869629 | SVEP1 |
| 20_9423104 | 0.160007131 | PLCB4 | 16_89863721 | 0.237661486 | SPIRE2 | 1_244573850 | 0.202847223 | C1orf101 |
| 3_6668460 | 0.159980564 | LOC101927347 | 1_35908500 | 0.237554534 | AGO1 | 1_244579578 | 0.202847223 | C1orf101 |
| 5_167829895 | 0.159951154 | TENM2 | 16_19666919 | 0.237520367 | C16orf62 | 1_244580655 | 0.202847223 | C1orf101 |
| 5_167835503 | 0.159951154 | TENM2 | 1_35188182 | 0.237434841 | SFPQ | 1_244583603 | 0.202847223 | C1orf101 |
| 3_195961005 | 0.159925547 | SDHAP1 | 1_35189847 | 0.237434841 | SFPQ | 2_55674182 | 0.202781293 | PNPT1 |
| 5_109788177 | 0.159913455 | MAN2A1 | 13_107732964 | 0.237301088 | FAM155A | 2_55684216 | 0.202719705 | PNPT1 |
| 5_106875195 | 0.1598084 | LOC102467213 | 13_107732971 | 0.237301088 | FAM155A | 13_71858839 | 0.202712019 | DACH1 |

| | | | | | | | | |
|---|---|---|---|---|---|---|---|---|
| 9_81607315 | 0.159762292 | TLE1 | 7_104898543 | 0.237255809 | LHFPL3 | 20_63466649 | 0.202677836 | KCNQ2 |
| 17_19374231 | 0.159721297 | B9D1 | 20_20148363 | 0.237211149 | C20orf26 | 10_96260149 | 0.20266828 | BLNK |
| 6_166775581 | 0.159709522 | RPS6KA2 | 1_201506080 | 0.237200624 | CSRP1 | 2_55676837 | 0.202654534 | PNPT1 |
| 20_53996600 | 0.159625806 | BCAS1 | 10_77260658 | 0.237145345 | KCNMA1 | 2_55682856 | 0.202654534 | PNPT1 |
| 5_109759863 | 0.159580792 | MAN2A1 | 22_40488658 | 0.237134171 | MKL1 | 2_100056065 | 0.202624363 | AFF3 |
| 5_109791049 | 0.159580792 | MAN2A1 | 2_37169637 | 0.237081461 | SULT6B1 | 2_49983564 | 0.202580769 | NRXN1 |
| 4_80890133 | 0.159541868 | C4orf22 | 2_37169700 | 0.237081461 | SULT6B1 | 15_83826311 | 0.202573634 | ADAMTSL3 |
| 10_70525095 | 0.159536433 | PALD1 | 20_19910725 | 0.237029826 | RIN2 | 12_112271317 | 0.202564992 | HECTD4 |
| 2_206744919 | 0.159532753 | MDH1B | 9_128441295 | 0.236981419 | MIR1268A | 7_26841233 | 0.202498803 | SKAP2 |
| 3_30786412 | 0.159494217 | GADL1 | 11_44790346 | 0.236980837 | TSPAN18 | 2_70007491 | 0.202434255 | PCBP1-AS1 |
| 7_93493855 | 0.159492794 | CALCR | 2_1292894 | 0.236828401 | SNTG2 | 3_701406 | 0.202389016 | LINC01266 |
| 7_93501685 | 0.159492794 | CALCR | 14_41750345 | 0.236738036 | LRFN5 | 7_90256135 | 0.202342255 | C7orf63 |
| 7_93503679 | 0.159492794 | CALCR | 1_35995080 | 0.236726769 | AGO3 | 6_29945653 | 0.202316771 | HLA-A |
| 7_93503863 | 0.159492794 | CALCR | 7_105628767 | 0.236649812 | ATXN7L1 | 12_111986415 | 0.202303254 | TMEM116 |
| 7_93505683 | 0.159492794 | CALCR | 4_108745595 | 0.236637384 | ETNPPL | 12_111996665 | 0.202303254 | TMEM116 |
| 7_93508770 | 0.159492794 | CALCR | 6_5763769 | 0.236636098 | FARS2 | 12_112033498 | 0.202303254 | NAA25 |
| 7_93509110 | 0.159492794 | CALCR | 10_60538858 | 0.236602466 | ANK3 | 12_112063722 | 0.202303254 | NAA25 |
| 15_48167991 | 0.159485864 | MYEF2 | 11_74355382 | 0.236598478 | PGM2L1 | 8_21752539 | 0.202240442 | GFRA2 |
| 17_19364058 | 0.159449613 | B9D1 | 1_35977613 | 0.236595681 | AGO3 | 13_99800907 | 0.202221845 | CLYBL |
| 6_169665646 | 0.159417944 | WDR27 | 1_244410630 | 0.236479977 | ADSS | 9_110956460 | 0.202195481 | LPAR1 |
| 7_78027296 | 0.159402744 | MAGI2 | 1_244442556 | 0.236479977 | ADSS | 2_64141762 | 0.202186874 | PELI1 |
| 15_97757124 | 0.159381314 | LINC00923 | 11_40789094 | 0.236467019 | LRRC4C | 2_50282493 | 0.202161429 | NRXN1 |
| 2_71333535 | 0.159344843 | ZNF638 | 4_188103756 | 0.236461945 | TRIML2 | 2_55544025 | 0.202001862 | CCDC104 |
| 14_88241383 | 0.159276422 | KCNK10 | 10_21166235 | 0.236422562 | NEBL | 2_50029374 | 0.201886409 | NRXN1 |
| 12_127892081 | 0.159210452 | FLJ37505 | 4_140027130 | 0.236397594 | MAML3 | 11_118581619 | 0.201885342 | ARCN1 |
| 12_12529122 | 0.159194583 | DUSP16 | 14_81518660 | 0.236313572 | SEL1L | 6_39821815 | 0.201884788 | DAAM2 |
| 12_12538794 | 0.159194583 | DUSP16 | 8_97974998 | 0.236284213 | MATN2 | 6_151419832 | 0.20181959 | RMND1 |
| 6_166789532 | 0.159178032 | RPS6KA2 | 13_70100480 | 0.236255821 | KLHL1 | 2_55596347 | 0.201818829 | SMEK2 |
| 6_166795418 | 0.159178032 | RPS6KA2 | 3_55060172 | 0.236255821 | CACNA2D3 | 2_49986540 | 0.201796874 | NRXN1 |
| 6_100649401 | 0.15917756 | ASCC3 | 1_35546895 | 0.23622519 | KIAA0319L | 2_54845796 | 0.201774964 | EML6 |
| 12_54973394 | 0.159169791 | TESPA1 | 22_40581552 | 0.2361507 | MKL1 | 11_118601281 | 0.201755989 | ARCN1 |
| 6_166808688 | 0.159131525 | RPS6KA2 | 6_89978634 | 0.236124054 | BACH2 | 2_229138442 | 0.201674753 | PID1 |
| 19_37958607 | 0.159113895 | SIPA1L3 | 3_14192662 | 0.236122968 | LSM3 | 2_229142020 | 0.201674753 | PID1 |
| 19_37960971 | 0.159113895 | SIPA1L3 | 2_79930249 | 0.236073798 | CTNNA2 | 2_229142430 | 0.201674753 | PID1 |
| 2_166984469 | 0.159012769 | XIRP2 | 3_55073230 | 0.236053033 | CACNA2D3 | 20_63470194 | 0.201663996 | KCNQ2 |
| 1_65373091 | 0.159004952 | DNAJC6 | 22_40489494 | 0.236045605 | MKL1 | 1_244586280 | 0.201628605 | C1orf101 |
| 10_81882278 | 0.158957881 | NRG3 | 22_40490989 | 0.236045605 | MKL1 | 12_112281362 | 0.2016086 | HECTD4 |
| 5_109831140 | 0.158924266 | MAN2A1 | 7_148080605 | 0.236037925 | CNTNAP2 | 12_111804401 | 0.201608252 | ALDH2 |
| 1_162865549 | 0.158912017 | C1orf110 | 22_17472708 | 0.235970034 | CECR2 | 12_7439783 | 0.201578158 | CD163L1 |
| 1_85253927 | 0.158911799 | C1orf52 | 12_27501430 | 0.235935203 | SMCO2 | 12_112427201 | 0.201535563 | PTPN11 |
| 12_127895037 | 0.158909733 | FLJ37505 | 9_3495070 | 0.235916612 | RFX3 | 6_30063679 | 0.201504188 | ZNRD1 |
| 14_77923717 | 0.158907977 | ADCK1 | 9_17243374 | 0.235913286 | CNTLN | 7_27095019 | 0.20144013 | HOXA1 |
| 11_92575367 | 0.158885113 | FAT3 | 4_108758648 | 0.235877969 | ETNPPL | 2_70039930 | 0.20141802 | PCBP1-AS1 |
| 22_49569965 | 0.158823691 | C22orf34 | 13_107271985 | 0.235833196 | FAM155A | 8_72837113 | 0.201398452 | KCNB2 |
| 15_27772290 | 0.15878335 | OCA2 | 11_34326523 | 0.235798675 | ABTB2 | 8_72837677 | 0.201398452 | KCNB2 |
| 19_37979430 | 0.158775579 | SIPA1L3 | 1_35188950 | 0.235736684 | SFPQ | 8_72847165 | 0.201398452 | KCNB2 |
| 2_166925596 | 0.158662291 | XIRP2 | 6_34326851 | 0.235702826 | NUDT3 | 6_99497268 | 0.201394447 | USP45 |
| 1_44551685 | 0.158654635 | RNF220 | 17_17123704 | 0.235625492 | MPRIP | 11_118495241 | 0.201385676 | KMT2A |
| 16_10788775 | 0.158631075 | TVP23A | 1_231184974 | 0.235571426 | LOC149373 | 2_55675661 | 0.201367692 | PNPT1 |
| 4_80889660 | 0.158621322 | C4orf22 | 1_231206223 | 0.235539263 | TRIM67 | 17_19043841 | 0.201321608 | GRAP |
| 6_159229946 | 0.158589297 | FNDC1 | 7_6174123 | 0.235500255 | CYTH3 | 2_70033480 | 0.201298107 | PCBP1-AS1 |
| 7_104893478 | 0.158587277 | LHFPL3 | 1_39639339 | 0.235456903 | HEYL | 2_70083746 | 0.201229951 | PCBP1-AS1 |
| 7_104894219 | 0.158587277 | LHFPL3 | 1_111866275 | 0.235411624 | KCND3 | 17_9488321 | 0.201114291 | STX8 |
| 6_144638559 | 0.158578186 | UTRN | 2_37245384 | 0.235390829 | NDUFAF7 | 2_69995431 | 0.201109506 | PCBP1-AS1 |
| 6_21777100 | 0.158578186 | CASC15 | 1_35502232 | 0.235375646 | KIAA0319L | 10_86508859 | 0.201076619 | WAPAL |
| 6_21777890 | 0.158578186 | CASC15 | 1_35947159 | 0.235355882 | AGO3 | 17_19319567 | 0.201048699 | EPN2 |
| 3_129446514 | 0.15857422 | IFT122 | 22_46699275 | 0.235334954 | CERK | 12_111972531 | 0.201020306 | TMEM116 |
| 3_129481413 | 0.15857422 | IFT122 | 6_108182556 | 0.235318069 | NR2E1 | 12_112067372 | 0.201020306 | NAA25 |
| 7_104861074 | 0.15857422 | LHFPL3 | 13_51647766 | 0.235306553 | WDFY2 | 15_90666212 | 0.201005541 | LOC101926895 |
| 7_104863366 | 0.15857422 | LHFPL3 | 15_100298344 | 0.23521619 | ADAMTS17 | 21_14163568 | 0.200982457 | LIPI |
| 6_21776444 | 0.15855472 | CASC15 | 1_2236091 | 0.235201847 | SKI | 10_45487677 | 0.200982335 | 8-Mar |
| 4_184193687 | 0.158484213 | ENPP6 | 15_100278700 | 0.235201847 | ADAMTS17 | 3_138436012 | 0.200955519 | ESYT3 |
| 3_4494719 | 0.158446253 | ITPR1 | 1_15910126 | 0.235196783 | SPEN | 6_30030199 | 0.200897007 | ZNRD1-AS1 |
| 20_46954769 | 0.158414233 | EYA2 | 9_113906364 | 0.235142134 | ZNF618 | 1_244433176 | 0.200888329 | ADSS |
| 20_46954896 | 0.158414233 | EYA2 | 17_59162727 | 0.235140042 | PRR11 | 11_118486899 | 0.200853789 | KMT2A |
| 20_46957973 | 0.158414233 | EYA2 | 15_100297988 | 0.235119682 | ADAMTS17 | 3_2528799 | 0.200825582 | CNTN4 |
| 3_11813908 | 0.158413088 | TAMM41 | 8_94252982 | 0.235105098 | GEM | 7_78027675 | 0.200807561 | MAGI2 |
| 15_48645781 | 0.15840929 | FBN1 | 7_100593798 | 0.235093033 | FBXO24 | 8_72846882 | 0.200777955 | KCNB2 |
| 14_57621938 | 0.158390589 | SLC35F4 | 10_7842187 | 0.23491564 | TAF3 | 20_63408819 | 0.200719148 | KCNQ2 |
| 4_31087241 | 0.158382416 | PCDH7 | 7_40073623 | 0.234883638 | CDK13 | 6_30163769 | 0.200685168 | TRIM15 |
| 5_109831249 | 0.158331853 | MAN2A1 | 15_100272317 | 0.234802883 | ADAMTS17 | 21_36515879 | 0.200660136 | CLDN14 |
| 5_109841522 | 0.158331853 | MAN2A1 | 9_113921998 | 0.234788249 | ZNF618 | 1_244573194 | 0.200635097 | C1orf101 |
| 22_32681230 | 0.158288516 | SYN3 | 2_231318878 | 0.234751741 | ARMC9 | 1_244573708 | 0.200635097 | C1orf101 |
| 1_205746143 | 0.158288126 | NUCKS1 | 12_342411 | 0.234743397 | KDM5A | 1_10707322 | 0.20055817 | CASZ1 |
| 21_17586862 | 0.158285024 | CXADR | 13_70136183 | 0.234726624 | ATXN8OS | 17_19411963 | 0.200526297 | RNF112 |
| 3_150762029 | 0.158263605 | SIAH2 | 15_48603494 | 0.234726624 | FBN1 | 1_244572955 | 0.200504253 | C1orf101 |
| 13_28274578 | 0.158227357 | PAN3 | 3_14123789 | 0.234726624 | CHCHD4 | 1_244577925 | 0.200504253 | C1orf101 |
| 17_76676748 | 0.158219096 | MXRA7 | 15_100273456 | 0.234712548 | ADAMTS17 | 1_244583749 | 0.200504253 | C1orf101 |
| 5_109700097 | 0.158201163 | MAN2A1 | 4_146821995 | 0.234708802 | TTC29 | 16_16324932 | 0.200396059 | LOC100288162 |
| 17_65166576 | 0.158179834 | RGS9 | 12_44825848 | 0.234632028 | NELL2 | 9_28005434 | 0.200371467 | LINGO2 |
| 17_65166755 | 0.158179834 | RGS9 | 20_42119015 | 0.234582359 | PTPRT | 7_8208375 | 0.200270601 | ICA1 |
| 17_65167053 | 0.158179834 | RGS9 | 9_113910747 | 0.234547166 | ZNF618 | 17_19280001 | 0.200191122 | EPN2 |
| 17_65169391 | 0.158179834 | RGS9 | 5_158969965 | 0.234515549 | EBF1 | 12_112206787 | 0.200180159 | HECTD4 |
| 1_62002731 | 0.158138441 | INADL | 4_78835628 | 0.234434106 | BMP2K | 11_118491699 | 0.200177614 | KMT2A |
| 7_103434808 | 0.158136235 | SLC26A5 | 4_188099479 | 0.234394913 | TRIML2 | 3_64596405 | 0.200171494 | ADAMTS9 |
| 7_103438240 | 0.158136235 | SLC26A5 | 3_192349978 | 0.234259644 | FGF12 | 6_5653478 | 0.20014576 | FARS2 |
| 5_126619314 | 0.158113066 | PHAX | 1_1211863 | 0.234220816 | TNFRSF4 | 3_699540 | 0.200122722 | LINC01266 |
| 6_128474202 | 0.158100808 | PTPRK | 2_71203963 | 0.234220213 | PAIP2B | 6_29614937 | 0.2001167 | GABBR1 |
| 5_109700365 | 0.158077621 | MAN2A1 | 7_149251068 | 0.234168802 | ZNF212 | 6_29794401 | 0.20010865 | LOC554223 |
| 20_46962735 | 0.158062777 | EYA2 | 12_1625980 | 0.234105061 | WNT5B | 22_18154608 | 0.200050996 | USP18 |
| 2_71390743 | 0.158045894 | ZNF638 | 4_88261699 | 0.234104124 | PPM1K | 8_142354839 | 0.200047362 | TSNARE1 |
| 2_167017694 | 0.158026469 | XIRP2 | 10_49688494 | 0.234091593 | C10orf53 | 4_25267340 | 0.20001451 | PI4K2B |
| 2_173274177 | 0.157988103 | MLK7-AS1 | 1_147170781 | 0.234035952 | PRKAB2 | 11_78466278 | 0.199947214 | NARS2 |
| 7_103430598 | 0.157931686 | SLC26A5 | 8_40620677 | 0.233997856 | ZMAT4 | 15_47302042 | 0.199907865 | SEMA6D |
| 7_103431839 | 0.157931686 | SLC26A5 | 3_17532072 | 0.233972337 | TBC1D5 | 17_61896195 | 0.199889087 | INTS2 |
| 7_103431883 | 0.157931686 | SLC26A5 | 16_10786220 | 0.23396229 | TVP23A | 12_95711416 | 0.199872675 | NTN4 |
| 7_103434141 | 0.157931686 | SLC26A5 | 9_84870244 | 0.233947165 | NTRK2 | 3_64609914 | 0.199863528 | ADAMTS9 |

| | | | | | | | | |
|---|---|---|---|---|---|---|---|---|
| 7_103436427 | 0.157931686 | SLC26A5 | 14_94001004 | 0.233934314 | LINC00521 | 12_27341685 | 0.199826903 | ARNTL2 |
| 7_103437295 | 0.157931686 | SLC26A5 | 16_89784625 | 0.233874534 | FANCA | 17_5444256 | 0.199821393 | DHX33 |
| 7_103437855 | 0.157931686 | SLC26A5 | 7_147258596 | 0.233788508 | CNTNAP2 | 2_55656578 | 0.199809216 | PNPT1 |
| 7_103439238 | 0.157931686 | SLC26A5 | 9_92532033 | 0.233571165 | CENPP | 4_122730733 | 0.199743882 | CETN4P |
| 7_103439278 | 0.157931686 | SLC26A5 | 8_40643497 | 0.233511843 | ZMAT4 | 11_118594100 | 0.199731825 | ARCN1 |
| 7_103440168 | 0.157931686 | SLC26A5 | 3_55058520 | 0.233452834 | CACNA2D3 | 11_62590945 | 0.199664057 | TUT1 |
| 7_78020822 | 0.157931686 | MAGI2 | 3_55063072 | 0.233385461 | CACNA2D3 | 10_131141524 | 0.199647619 | TCERG1L |
| 6_159239847 | 0.157919208 | FNDC1 | 3_55063809 | 0.233385461 | CACNA2D3 | 4_127970607 | 0.199599554 | C4orf29 |
| 1_64687258 | 0.157917901 | CACHD1 | 12_379020 | 0.233373084 | KDM5A | 15_47311297 | 0.199592877 | SEMA6D |
| 16_89148345 | 0.157916158 | ACSF3 | 12_333919 | 0.233368544 | KDM5A | 11_118053440 | 0.199453068 | TMPRSS4-AS1 |
| 10_91984716 | 0.157905243 | BTAF1 | 12_365631 | 0.233368544 | KDM5A | 2_70005331 | 0.199432157 | PCBP1-AS1 |
| 17_79737018 | 0.157885441 | ENPP7 | 5_169192576 | 0.233367402 | SLIT3 | 18_42666405 | 0.199427555 | LINC00907 |
| 19_37887133 | 0.157794691 | WDR87 | 15_47768253 | 0.233289649 | SEMA6D | 10_86457521 | 0.199367129 | WAPAL |
| 6_166793409 | 0.15774584 | RPS6KA2 | 7_6148303 | 0.233262731 | USP42 | 7_76413092 | 0.199278021 | ZP3 |
| 1_15050083 | 0.157742219 | KAZN | 7_6250628 | 0.233262731 | CYTH3 | 6_32661673 | 0.199157249 | HLA-DQB1 |
| 6_87820075 | 0.157652232 | LOC101928911 | 19_2824916 | 0.233152034 | ZNF554 | 1_53466256 | 0.199123541 | DMRTB1 |
| 4_31067095 | 0.157650334 | PCDH7 | 20_10267037 | 0.233137424 | SNAP25 | 2_10727899 | 0.199101565 | ATP6V1C2 |
| 7_103440216 | 0.157624608 | SLC26A5 | 18_9356121 | 0.233125307 | TWSG1 | 1_244585675 | 0.199072017 | C1orf101 |
| 5_44375168 | 0.157566775 | FGF10 | 7_6159164 | 0.233125307 | USP42 | 10_93445969 | 0.199029766 | MYOF |
| 7_91176802 | 0.157538127 | CDK14 | 11_101029985 | 0.232868576 | PGR | 1_246619198 | 0.198986025 | CNST |
| 19_37956410 | 0.157506065 | SIPA1L3 | 11_101031322 | 0.232868576 | PGR | 7_100749651 | 0.198948484 | ZAN |
| 22_32599390 | 0.15749701 | SYN3 | 8_14821426 | 0.232848016 | SGCZ | 1_22788753 | 0.198923964 | EPHB2 |
| 1_30948395 | 0.157447179 | PUM1 | 6_100397768 | 0.232847214 | SIM1 | 19_37556173 | 0.19890372 | ZNF540 |
| 14_66837386 | 0.157419088 | GPHN | 4_89750336 | 0.232818805 | SNCA | 10_93463438 | 0.198879627 | MYOF |
| 2_162555020 | 0.157374537 | KCNH7 | 5_173107823 | 0.23281069 | CREBRF | 9_130072364 | 0.198840257 | GPR107 |
| 5_109756113 | 0.157327197 | MAN2A1 | 9_17386384 | 0.232731753 | CNTLN | 8_95546841 | 0.198831394 | C8orf37-AS1 |
| 5_106848113 | 0.157322876 | LOC102467213 | 1_6879614 | 0.232710712 | CAMTA1 | 11_101990738 | 0.198814435 | KIAA1377 |
| 15_90395488 | 0.157285015 | IQGAP1 | 2_73061308 | 0.232710712 | SFXN5 | 15_71762178 | 0.19881294 | THSD4 |
| 6_166779642 | 0.157224301 | RPS6KA2 | 9_40998401 | 0.232672711 | LOC100132352 | 8_72836620 | 0.198763196 | KCNB2 |
| 1_162864490 | 0.157196413 | C1orf110 | 1_169548666 | 0.232649585 | F5 | 8_72837200 | 0.198763196 | KCNB2 |
| 1_162864495 | 0.157196413 | C1orf110 | 1_13584898 | 0.232611929 | PDPN | 8_72844091 | 0.198763196 | KCNB2 |
| 1_111841566 | 0.157165775 | KCND3 | 7_57135633 | 0.232608925 | ZNF479 | 8_72846084 | 0.198763196 | KCNB2 |
| 3_129446680 | 0.157122005 | IFT122 | 13_97294377 | 0.232561409 | MBNL2 | 8_72849970 | 0.198763196 | KCNB2 |
| 3_129454802 | 0.157122005 | IFT122 | 13_107717366 | 0.232550833 | FAM155A | 8_72850051 | 0.198763196 | KCNB2 |
| 3_129474600 | 0.157122005 | IFT122 | 13_107717367 | 0.232550833 | FAM155A | 8_72853278 | 0.198763196 | KCNB2 |
| 3_129485194 | 0.157122005 | IFT122 | 11_18267994 | 0.232549853 | SAA1 | 9_28037763 | 0.198758523 | LINGO2 |
| 3_129487416 | 0.157122005 | IFT122 | 19_58232779 | 0.232515714 | ZNF544 | 2_50034462 | 0.198707654 | NRXN1 |
| 3_129495737 | 0.157122005 | IFT122 | 11_129402101 | 0.232461765 | BARX2 | 3_67529697 | 0.19869658 | SUCLG2 |
| 3_129499054 | 0.157122005 | IFT122 | 5_31935030 | 0.232442662 | PDZD2 | 11_132599295 | 0.198658833 | OPCML |
| 3_129513511 | 0.157122005 | IFT122 | 20_44213153 | 0.232391922 | OSER1-AS1 | 14_105428994 | 0.198606777 | MTA1 |
| 7_104861702 | 0.157122005 | LHFPL3 | 4_100512750 | 0.232256082 | EMCN | 14_105433481 | 0.198606777 | MTA1 |
| 7_104863531 | 0.157122005 | LHFPL3 | 8_14834947 | 0.232230339 | SGCZ | 14_105439736 | 0.198606777 | MTA1 |
| 7_104864834 | 0.157122005 | LHFPL3 | 22_40489495 | 0.232205339 | MKL1 | 14_63385459 | 0.198603441 | PPP2R5E |
| 2_71372023 | 0.157114102 | ZNF638 | 5_173107818 | 0.232128125 | CREBRF | 7_134133973 | 0.198510315 | LRGUK |
| 13_28158164 | 0.157108813 | PAN3 | 4_88010856 | 0.232113896 | PKD2 | 6_70287268 | 0.198493597 | COL9A1 |
| 1_243746100 | 0.157104425 | AKT3 | 9_17379997 | 0.23205774 | CNTLN | 6_32665863 | 0.198486322 | HLA-DQB1 |
| 20_53945555 | 0.157086971 | BCAS1 | 20_19306672 | 0.232055062 | SLC24A3 | 11_118538821 | 0.198461852 | TMEM25 |
| 9_3470503 | 0.157055095 | RFX3 | 12_316577 | 0.231965018 | KDM5A | 11_118538824 | 0.198461852 | TMEM25 |
| 14_96345506 | 0.157023958 | ATG2B | 8_127475946 | 0.231814606 | CASC8 | 6_37666398 | 0.198436517 | MDGA1 |
| 16_10784057 | 0.156992453 | TVP23A | 15_100294903 | 0.231797416 | ADAMTS17 | 17_19240197 | 0.198393453 | EPN2 |
| 4_31120807 | 0.156990818 | PCDH7 | 4_64994097 | 0.231758403 | LOC401134 | 17_19265007 | 0.198393453 | EPN2 |
| 19_16131923 | 0.156954563 | RAB8A | 9_110886534 | 0.231715558 | LPAR1 | 6_45958523 | 0.19836258 | CLIC5 |
| 17_5908558 | 0.156896069 | LOC339166 | 22_40536062 | 0.231668364 | MKL1 | 16_14158989 | 0.198354575 | MKL2 |
| 4_175741597 | 0.156892555 | GPM6A | 8_127476856 | 0.231634431 | CASC8 | 11_100734037 | 0.19833164 | ARHGAP42 |
| 8_142777765 | 0.156875577 | LYNX1 | 13_97296523 | 0.231622882 | MBNL2 | 2_70069149 | 0.198307105 | PCBP1-AS1 |
| 2_172034094 | 0.156783854 | METAP1D | 1_29149716 | 0.231610351 | SRSF4 | 5_10448904 | 0.198253412 | ROPN1L |
| 2_71133774 | 0.156776511 | MPHOSPH10 | 2_227172527 | 0.231607312 | COL4A3 | 12_112309858 | 0.198197731 | HECTD4 |
| 6_144532452 | 0.156725364 | UTRN | 15_83590323 | 0.231591401 | SH3GL3 | 12_112312032 | 0.198197731 | HECTD4 |
| 6_144533571 | 0.156725364 | UTRN | 13_33519222 | 0.231525327 | STARD13 | 1_169566929 | 0.198152852 | F5 |
| 6_144585023 | 0.156725364 | UTRN | 17_65205869 | 0.231499488 | RGS9 | 3_31981279 | 0.19814878 | OSBPL10 |
| 6_144587893 | 0.156725364 | UTRN | 3_14123770 | 0.231428541 | CHCHD4 | 2_47447145 | 0.198148544 | MSH2 |
| 18_42342251 | 0.156666346 | LINC00907 | 17_10140225 | 0.231399082 | GAS7 | 6_32660772 | 0.198119876 | HLA-DQB1 |
| 1_35187479 | 0.156626386 | SFPQ | 17_10140637 | 0.231399082 | GAS7 | 6_32661915 | 0.198119876 | HLA-DQB1 |
| 6_166827565 | 0.156618499 | RPS6KA2 | 17_10141281 | 0.231399082 | GAS7 | 6_70285266 | 0.198118208 | COL9A1 |
| 22_32712072 | 0.156568501 | SYN3 | 20_10296973 | 0.231368421 | SNAP25 | 6_70296395 | 0.198118208 | COL9A1 |
| 14_65441594 | 0.156498727 | FUT8 | 11_134453356 | 0.231285357 | LOC283177 | 4_39504233 | 0.198114155 | UGDH |
| 14_105228992 | 0.156461979 | BRF1 | 11_72999316 | 0.231211195 | FCHSD2 | 12_27372327 | 0.198081886 | ARNTL2 |
| 2_30963821 | 0.156436263 | GALNT14 | 13_85439434 | 0.231187429 | LINC00351 | 2_71418760 | 0.197994381 | ZNF638 |
| 5_126610319 | 0.156420762 | PHAX | 7_91175907 | 0.231068001 | CDK14 | 3_129533950 | 0.19797146 | RHO |
| 19_58232772 | 0.156401385 | ZNF544 | 14_61566413 | 0.230945704 | LOC101927780 | 4_4495567 | 0.197959476 | STX18 |
| 22_42507456 | 0.156400474 | SERHL | 22_46687873 | 0.23085953 | CERK | 4_4495568 | 0.197959476 | STX18 |
| 4_21389987 | 0.156284915 | KCNIP4 | 12_311895 | 0.230802225 | KDM5A | 4_4495572 | 0.197959476 | STX18 |
| 5_106824328 | 0.156281456 | LOC102467213 | 3_124224566 | 0.230794269 | KALRN | 8_72843528 | 0.197943674 | KCNB2 |
| 22_32717389 | 0.156239555 | SYN3 | 7_105626360 | 0.230737926 | ATXN7L1 | 2_16637806 | 0.197886169 | FAM49A |
| 4_89724520 | 0.156219915 | SNCA | 7_6258413 | 0.230737926 | CYTH3 | 9_101482230 | 0.197853876 | TMEM246 |
| 4_89724523 | 0.156219915 | SNCA | 1_244426450 | 0.230716848 | ADSS | 10_35259692 | 0.197836472 | CCNY |
| 4_89743643 | 0.156219915 | SNCA | 1_244442839 | 0.230716848 | ADSS | 2_64112773 | 0.197821674 | PELI1 |
| 11_131475457 | 0.156213422 | NTM | 2_170384046 | 0.230707148 | MYO3B | 6_32666077 | 0.19776883 | HLA-DQB1 |
| 11_126352840 | 0.156165069 | ST3GAL4-AS1 | 22_40627300 | 0.230620277 | MKL1 | 11_118561689 | 0.197757484 | IFT46 |
| 12_12388749 | 0.156153614 | LOH12CR1 | 11_26530586 | 0.230484681 | ANO3 | 1_220035965 | 0.19773827 | EPRS |
| 7_104852141 | 0.156153588 | LHFPL3 | 11_59838849 | 0.230477489 | GIF | 2_46722545 | 0.19773827 | SOCS5 |
| 1_15050283 | 0.156121789 | KAZN | 19_329868 | 0.230477489 | MIER2 | 13_28098318 | 0.197661911 | FLT3 |
| 4_6426569 | 0.156107535 | PPP2R2C | 20_59613299 | 0.230450955 | PHACTR3 | 6_32665869 | 0.197594735 | HLA-DQB1 |
| 6_166836465 | 0.156075967 | RPS6KA2 | 10_25477526 | 0.230374659 | GPR158 | 8_40667846 | 0.197390495 | ZMAT4 |
| 2_240572941 | 0.156065636 | RNPEPL1 | 20_19295817 | 0.230293134 | SLC24A3 | 10_96249576 | 0.197381884 | BLNK |
| 3_131176678 | 0.156059632 | NEK11 | 3_155568047 | 0.23028789 | PLCH1 | 15_52290413 | 0.197356852 | MYO5C |
| 15_68819736 | 0.156054641 | ANP32A | 3_155589178 | 0.23028789 | PLCH1 | 15_88595503 | 0.197286117 | LOC101929743 |
| 9_20375687 | 0.156001285 | MLLT3 | 3_155617000 | 0.23028789 | PLCH1 | 1_244563067 | 0.197245917 | C1orf101 |
| 9_130410013 | 0.155872786 | HMCN2 | 7_6172086 | 0.230194909 | CYTH3 | 13_41218299 | 0.197211416 | LOC101929140 |
| 9_130410021 | 0.155872786 | HMCN2 | 3_192354953 | 0.230151126 | FGF12 | 10_93445470 | 0.197199912 | MYOF |
| 11_78210481 | 0.155838037 | USP35 | 15_25744113 | 0.230144821 | ATP10A | 14_63391090 | 0.1971837 | PPP2R5E |
| 5_106849688 | 0.155833875 | LOC102467213 | 15_48515200 | 0.230144821 | FBN1 | 1_244581420 | 0.197171007 | C1orf101 |
| 13_28156155 | 0.155797896 | PAN3 | 15_48631055 | 0.230144821 | FBN1 | 1_244583848 | 0.197171007 | C1orf101 |
| 5_110712119 | 0.155757642 | TMEM232 | 3_155552012 | 0.230130671 | PLCH1 | 1_244584926 | 0.197171007 | C1orf101 |
| 16_57889755 | 0.155702112 | CNGB1 | 7_6162782 | 0.230015608 | CYTH3 | 1_244584934 | 0.197171007 | C1orf101 |
| 19_37683496 | 0.155642683 | ZNF781 | 17_34021838 | 0.229933761 | ASIC2 | 4_88034451 | 0.197168838 | PKD2 |
| 1_61684869 | 0.155624978 | TM2D1 | 12_32894274 | 0.229853855 | PKP2 | 11_118471070 | 0.197165577 | KMT2A |

| | | | | | | | | |
|---|---|---|---|---|---|---|---|---|
| 19_37942796 | 0.155586524 | SIPA1L3 | 10_50441174 | 0.229829611 | SGMS1 | 2_55688020 | 0.197162119 | PNPT1 |
| 8_48016782 | 0.155582938 | UBE2V2 | 22_46690027 | 0.229692922 | CERK | 2_50037572 | 0.197133185 | NRXN1 |
| 12_1501511 | 0.155561952 | LINC00942 | 11_12783193 | 0.229680757 | TEAD1 | 15_83908577 | 0.197110883 | ADAMTSL3 |
| 6_47708186 | 0.155533533 | GPR115 | 5_21467911 | 0.229644687 | GUSBP1 | 20_46193137 | 0.197097671 | CDH22 |
| 20_46956764 | 0.155443762 | EYA2 | 15_64182054 | 0.22962527 | CSNK1G1 | 2_70076086 | 0.197096738 | PCBP1-AS1 |
| 20_46958080 | 0.155443762 | EYA2 | 12_291045 | 0.229610477 | KDM5A | 15_52239744 | 0.197091076 | MYO5C |
| 15_40268502 | 0.155434579 | PAK6 | 12_314661 | 0.229610477 | KDM5A | 4_147659576 | 0.197076498 | PRMT9 |
| 1_65366331 | 0.155412253 | DNAJC6 | 12_351781 | 0.229610477 | KDM5A | 11_78970010 | 0.197075994 | TENM4 |
| 3_30780755 | 0.155403415 | GADL1 | 12_372784 | 0.229610477 | KDM5A | 20_46195460 | 0.197041635 | CDH22 |
| 17_10140637 | 0.155397189 | GAS7 | 3_55062314 | 0.229595312 | CACNA2D3 | 16_78357633 | 0.197033894 | WWOX |
| 8_10395591 | 0.155385194 | MSRA | 16_89875082 | 0.229588937 | TCF25 | 3_126553409 | 0.197000087 | C3orf22 |
| 18_53259306 | 0.155381769 | DCC | 18_24147084 | 0.229503597 | CABYR | 4_128099286 | 0.196957538 | LARP1B |
| 7_94405377 | 0.15537932 | COL1A2 | 4_108581956 | 0.229441427 | RPL34-AS1 | 2_55684305 | 0.196940099 | PNPT1 |
| 3_150755284 | 0.155376555 | SIAH2 | 4_108584262 | 0.229441427 | RPL34-AS1 | 6_152170896 | 0.196907806 | SYNE1 |
| 5_178455204 | 0.155375031 | COL23A1 | 10_68458528 | 0.229302713 | DNA2 | 1_244586599 | 0.196843556 | C1orf101 |
| 5_31925322 | 0.155375031 | PDZD2 | 6_100397553 | 0.229282827 | SIM1 | 6_32661836 | 0.196838648 | HLA-DQB1 |
| 11_44832840 | 0.155360212 | TSPAN18 | 15_23302982 | 0.22923211 | GOLGA6L22 | 4_93581120 | 0.196799696 | GRID2 |
| 3_65376718 | 0.155353172 | MAGI1 | 11_78268736 | 0.22917329 | GAB2 | 3_138448019 | 0.196791693 | ESYT3 |
| 2_206738166 | 0.155340397 | MDH1B | 19_50522516 | 0.229047645 | LRRC4B | 20_63469275 | 0.196736282 | KCNQ2 |
| 2_206740193 | 0.155340397 | MDH1B | 1_147255322 | 0.229014606 | CHD1L | 20_63469603 | 0.196736282 | KCNQ2 |
| 2_206740886 | 0.155340397 | MDH1B | 2_106194311 | 0.228986073 | UXS1 | 20_63469739 | 0.196736282 | KCNQ2 |
| 2_206741158 | 0.155340397 | MDH1B | 4_89783404 | 0.228964688 | SNCA | 11_118578655 | 0.196733901 | ARCN1 |
| 2_206746185 | 0.155340397 | MDH1B | 20_59646521 | 0.228961273 | PHACTR3 | 15_27461515 | 0.196729304 | GABRG3 |
| 19_35359961 | 0.155334345 | FFAR3 | 16_89804936 | 0.228945055 | FANCA | 7_44629822 | 0.196699234 | OGDH |
| 1_38939437 | 0.15532704 | RHBDL2 | 9_17247673 | 0.228916088 | CNTLN | 1_244585573 | 0.196688534 | C1orf101 |
| 6_166800439 | 0.155283729 | RPS6KA2 | 2_168094841 | 0.228821412 | STK39 | 6_5652545 | 0.196637337 | FARS2 |
| 16_89846600 | 0.155264796 | SPIRE2 | 17_75636571 | 0.228775711 | RECQL5 | 6_5652547 | 0.196637337 | FARS2 |
| 12_127898382 | 0.155238988 | FLJ37505 | 1_13589630 | 0.228737104 | PDPN | 1_52783910 | 0.196532108 | ZYG11B |
| 7_104834016 | 0.155214307 | LHFPL3 | 7_149249630 | 0.228692607 | ZNF212 | 1_54730021 | 0.196521714 | MROH7-TTC4 |
| 1_62119679 | 0.155185781 | INADL | 7_78583906 | 0.228656525 | MAGI2 | 10_19960225 | 0.196491838 | PLXDC2 |
| 1_44536599 | 0.155160283 | RNF220 | 2_240452704 | 0.228608958 | GPC1 | 4_88066763 | 0.196484659 | PKD2 |
| 9_9802798 | 0.155148357 | PTPRD | 8_71320995 | 0.228561694 | EYA1 | 2_238191419 | 0.196439005 | ILKAP |
| 14_63374689 | 0.155133709 | PPP2R5E | 22_40474484 | 0.228539289 | MKL1 | 3_128368448 | 0.196416955 | EEFSEC |
| 2_167012893 | 0.155125697 | XIRP2 | 17_76701304 | 0.228479574 | MXRA7 | 1_52799227 | 0.196390911 | ZYG11B |
| 2_71384001 | 0.155123139 | ZNF638 | 1_147178479 | 0.228442233 | PDIA3P1 | 12_19262835 | 0.196300804 | PLEKHA5 |
| 6_21776316 | 0.155085967 | CASC15 | 1_147224155 | 0.228442233 | FMO5 | 21_40146350 | 0.196297433 | DSCAM |
| 8_16183413 | 0.155080752 | MSR1 | 4_56213401 | 0.22832597 | KIAA1211 | 3_64807995 | 0.196252595 | ADAMTS9-AS2 |
| 3_33120757 | 0.155034946 | CRTAP | 1_244386694 | 0.228305859 | C1orf100 | 5_10434531 | 0.196249348 | 6-Mar |
| 3_33120871 | 0.155034946 | CRTAP | 4_38482627 | 0.228303087 | LINC01258 | 11_34338418 | 0.196226295 | ABTB2 |
| 6_169767522 | 0.155034946 | ERMARD | 10_43448996 | 0.228280506 | ZNF487 | 12_6558398 | 0.196226295 | NOP2 |
| 10_12580681 | 0.15500831 | CAMK1D | 19_426974 | 0.228270881 | SHC2 | 2_10732844 | 0.196220306 | ATP6V1C2 |
| 22_49567700 | 0.154960893 | C22orf34 | 3_155612497 | 0.22823007 | PLCH1 | 17_5450757 | 0.196217164 | DHX33 |
| 22_49568987 | 0.154960893 | C22orf34 | 2_132927405 | 0.228223901 | NCKAP5 | 7_130942189 | 0.196216927 | LINC-PINT |
| 1_201095787 | 0.154928055 | CACNA1S | 8_28947797 | 0.228196423 | HMBOX1 | 15_83823840 | 0.19620483 | ADAMTSL3 |
| 14_57621255 | 0.154892123 | SLC35F4 | 7_149280222 | 0.228106687 | ZNF783 | 17_9503917 | 0.196191456 | STX8 |
| 17_40092101 | 0.154872658 | THRA | 7_6164217 | 0.228011761 | CYTH3 | 3_139121059 | 0.196160957 | BPESC1 |
| 7_103444641 | 0.154840064 | SLC26A5 | 1_155544081 | 0.227985613 | ASH1L | 3_129448701 | 0.196089312 | IFT122 |
| 12_127896972 | 0.154808711 | FLJ37505 | 9_4824375 | 0.227948161 | RCL1 | 19_5828053 | 0.196079887 | NRTN |
| 1_243653334 | 0.154757369 | AKT3 | 22_40628842 | 0.22785722 | MKL1 | 6_90062093 | 0.196041567 | BACH2 |
| 3_129512590 | 0.154693157 | IFT122 | 14_61538930 | 0.227856582 | PRKCH | 15_34795540 | 0.196010129 | ACTC1 |
| 7_104849534 | 0.154693157 | LHFPL3 | 17_16113495 | 0.227852666 | NCOR1 | 10_96325581 | 0.195926858 | DNTT |
| 7_104850553 | 0.154693157 | LHFPL3 | 22_46702823 | 0.22782917 | CERK | 17_10140225 | 0.195902959 | GAS7 |
| 16_89159244 | 0.154684683 | LINC00304 | 13_107773540 | 0.2277548 | FAM155A | 5_176626621 | 0.195830298 | SNCB |
| 13_28216231 | 0.15468403 | PAN3 | 1_185615283 | 0.227677159 | GS1-204I12.1 | 6_128238281 | 0.19577266 | PTPRK |
| 12_21200938 | 0.154672578 | SLCO1B1 | 2_170420012 | 0.227603305 | MYO3B | 2_64129511 | 0.195740657 | PELI1 |
| 6_144604053 | 0.154661489 | UTRN | 2_121732356 | 0.227585955 | NIFK | 16_10794077 | 0.195694507 | TVP23A |
| 18_53305273 | 0.154632953 | DCC | 9_3508284 | 0.227498729 | RFX3 | 18_46716356 | 0.195622512 | ST8SIA5 |
| 7_103442642 | 0.154625672 | SLC26A5 | 2_166905163 | 0.227490407 | XIRP2 | 11_19982259 | 0.195611717 | NAV2 |
| 8_10428300 | 0.154603733 | MSRA | 15_48494997 | 0.22748566 | FBN1 | 6_32660820 | 0.195587676 | HLA-DQB1 |
| 17_79140752 | 0.154586573 | RBFOX3 | 6_34337295 | 0.227479958 | NUDT3 | 8_72836474 | 0.19556609 | KCNB2 |
| 1_231184974 | 0.154570937 | LOC149373 | 8_14819349 | 0.227308468 | SGCZ | 12_112298926 | 0.195561847 | HECTD4 |
| 14_96356488 | 0.154497339 | ATG2B | 20_4185926 | 0.227286842 | SMOX | 14_61488043 | 0.195537373 | PRKCH |
| 6_169582146 | 0.154464223 | WDR27 | 3_698287 | 0.227238283 | LINC01266 | 12_65767809 | 0.195500005 | RPSAP52 |
| 12_81335762 | 0.154395547 | PPFIA2 | 13_59892611 | 0.227180268 | DIAPH3 | 7_78031215 | 0.195488552 | MAGI2 |
| 1_11610672 | 0.154372727 | LOC101929181 | 16_29875864 | 0.227159067 | SEZ6L2 | 2_16636522 | 0.195458823 | FAM49A |
| 11_121585500 | 0.154355564 | SORL1 | 18_69447248 | 0.227110726 | DOK6 | 2_16638691 | 0.195458823 | FAM49A |
| 1_7445781 | 0.15429173 | CAMTA1 | 18_69447254 | 0.227110726 | DOK6 | 3_64629883 | 0.195450741 | ADAMTS9 |
| 1_62002729 | 0.154289988 | INADL | 18_69447260 | 0.227110726 | DOK6 | 6_151304993 | 0.195428773 | AKAP12 |
| 7_78019414 | 0.154288693 | MAGI2 | 11_118118586 | 0.227048532 | TMPRSS4 | 8_72860985 | 0.195405926 | KCNB2 |
| 2_71427854 | 0.154242403 | ZNF638 | 19_948358 | 0.227038602 | ARID3A | 7_140824976 | 0.195401128 | BRAF |
| 15_40370300 | 0.154227758 | DISP2 | 12_342871 | 0.227035913 | KDM5A | 12_111955261 | 0.19539944 | TMEM116 |
| 8_63193976 | 0.15422551 | YTHDF3 | 12_361596 | 0.227035913 | KDM5A | 12_112079028 | 0.19539944 | NAA25 |
| 21_16393298 | 0.154215306 | LINC00478 | 15_100300323 | 0.226978489 | ADAMTS17 | 2_50002671 | 0.195357774 | NRXN1 |
| 12_113923622 | 0.154209799 | RBM19 | 2_113882689 | 0.226947547 | LOC101060091 | 11_21438256 | 0.195353147 | NELL1 |
| 2_162538703 | 0.154177526 | KCNH7 | 13_94891164 | 0.226935615 | LOC101927284 | 3_29393849 | 0.195339746 | RBMS3 |
| 2_170380913 | 0.154175039 | MYO3B | 2_33149221 | 0.226900075 | LTBP1 | 4_90192320 | 0.195326611 | CCSER1 |
| 1_210143812 | 0.154168804 | SYT14 | 2_33149222 | 0.226900075 | LTBP1 | 11_118537318 | 0.19532161 | TMEM25 |
| 1_210143814 | 0.154168804 | SYT14 | 15_80473304 | 0.226886371 | ARNT2 | 11_118559333 | 0.19532161 | IFT46 |
| 15_27781789 | 0.154133181 | OCA2 | 4_39520104 | 0.226883835 | UGDH | 14_96078988 | 0.195300016 | C14orf132 |
| 2_71355125 | 0.154133181 | ZNF638 | 7_71628549 | 0.226849286 | WBSCR17 | 12_112238864 | 0.195226499 | HECTD4 |
| 11_132836451 | 0.154124313 | OPCML | 15_58537252 | 0.226805637 | LIPC | 12_112504549 | 0.195226499 | PTPN11 |
| 2_46329684 | 0.154119326 | EPAS1 | 2_159250092 | 0.226804896 | WDSUB1 | 2_55677391 | 0.195195424 | PNPT1 |
| 3_65425420 | 0.154104252 | MAGI1 | 19_428077 | 0.226717999 | SHC2 | 7_91167295 | 0.195187656 | CDK14 |
| 1_231176453 | 0.154097951 | TRIM67 | 4_108750137 | 0.226696438 | ETNPPL | 6_32666059 | 0.195148574 | HLA-DQB1 |
| 11_113345201 | 0.154083242 | TTC12 | 3_192352798 | 0.226678458 | FGF12 | 17_19319887 | 0.195113163 | EPN2 |
| 1_36045899 | 0.154059779 | AGO3 | 4_146846836 | 0.226631046 | TTC29 | 7_90261308 | 0.195104285 | C7orf63 |
| 1_111840049 | 0.154053007 | KCND3 | 9_17232335 | 0.226587353 | CNTLN | 2_16636067 | 0.195070326 | FAM49A |
| 14_88244219 | 0.153986656 | KCNK10 | 12_296093 | 0.226586357 | KDM5A | 3_64625570 | 0.195047284 | ADAMTS9 |
| 5_109713806 | 0.153956396 | MAN2A1 | 20_20126960 | 0.226585613 | C20orf26 | 20_20038503 | 0.195044277 | CRNKL1 |
| 10_86711454 | 0.153910848 | LDB3 | 8_40623720 | 0.226575141 | ZMAT4 | 15_30766676 | 0.19501883 | LOC100288637 |
| 13_28162307 | 0.153886214 | PAN3 | 2_71194492 | 0.226529074 | PAIP2B | 18_12854430 | 0.194991484 | PTPN2 |
| 13_28163502 | 0.153886214 | PAN3 | 20_10267845 | 0.226492674 | SNAP25 | 19_13409115 | 0.194990419 | CACNA1A |
| 19_2538453 | 0.153882828 | GNG7 | 13_114295800 | 0.226415478 | UPF3A | 2_71345325 | 0.194967217 | ZNF638 |
| 4_6838900 | 0.153818229 | KIAA0232 | 15_48628745 | 0.226412368 | FBN1 | 19_40401699 | 0.194944673 | PRX |
| 13_28255995 | 0.153795513 | PAN3 | 16_27946690 | 0.226410165 | GSG1L | 12_15995871 | 0.194941413 | DERA |
| 2_221557005 | 0.153781592 | EPHA4 | 13_71739657 | 0.226375377 | DACH1 | 11_76966866 | 0.194908543 | ACER3 |
| 2_221557407 | 0.153781592 | EPHA4 | 4_78997652 | 0.226373467 | LINC01088 | 10_45572900 | 0.19489072 | 8-Mar |

| | | | | | | | | |
|---|---|---|---|---|---|---|---|---|
| 17_60966663 | 0.153765616 | BCAS3 | 6_34425871 | 0.226352067 | RPS10 | 15_88595558 | 0.19483716 | LOC101929743 |
| 4_6825433 | 0.153762905 | KIAA0232 | 5_162133256 | 0.226345662 | GABRG2 | 7_43955088 | 0.194797784 | POLR2J4 |
| 6_144557201 | 0.153703425 | UTRN | 4_67537028 | 0.22629841 | CENPC | 4_153356660 | 0.194777473 | MND1 |
| 6_169766234 | 0.15369364 | ERMARD | 6_161380427 | 0.226296475 | PARK2 | 11_118552957 | 0.194775561 | IFT46 |
| 12_49808928 | 0.153691132 | NCKAP5L | 5_119077757 | 0.226261182 | DMXL1 | 8_23163417 | 0.194775561 | TNFRSF10D |
| 7_104881186 | 0.15368663 | LHFPL3 | 1_178349259 | 0.226260913 | RASAL2 | 16_29346679 | 0.194700223 | SNX29P2 |
| 8_16171084 | 0.153633259 | MSR1 | 2_174179221 | 0.226257426 | OLA1 | 4_41443202 | 0.194684378 | LIMCH1 |
| 20_46193137 | 0.153590566 | CDH22 | 1_180817724 | 0.22620161 | XPR1 | 1_1296823 | 0.194675534 | ACAP3 |
| 17_19372886 | 0.15355293 | B9D1 | 9_35105260 | 0.226191685 | FAM214B | 15_83862710 | 0.194672348 | ADAMTSL3 |
| 6_169708971 | 0.153540416 | PHF10 | 12_95206699 | 0.226076202 | FGD6 | 15_52235996 | 0.194653489 | MYO5C |
| 7_104840210 | 0.153540381 | LHFPL3 | 1_202210175 | 0.226019038 | LGR6 | 9_17204516 | 0.194623216 | CNTLN |
| 13_102640719 | 0.153519959 | TPP2 | 11_118116326 | 0.226015958 | TMPRSS4 | 13_28098317 | 0.19461779 | FLT3 |
| 6_87797895 | 0.153504324 | LOC101928911 | 12_310418 | 0.226010251 | KDM5A | 2_223827954 | 0.194610962 | AP1S3 |
| 1_244563237 | 0.153441232 | C1orf101 | 12_316797 | 0.226010251 | KDM5A | 14_63387521 | 0.194596941 | PPP2R5E |
| 6_166805323 | 0.153420407 | RPS6KA2 | 12_321709 | 0.226010251 | KDM5A | 6_38716411 | 0.194575959 | DNAH8 |
| 1_244591199 | 0.153418257 | C1orf101 | 12_362660 | 0.226010251 | KDM5A | 4_56257927 | 0.194575718 | KIAA1211 |
| 2_179991136 | 0.153368431 | CWC22 | 8_70116187 | 0.22600329 | NCOA2 | 6_70289647 | 0.194574337 | COL9A1 |
| 2_163614870 | 0.153364195 | FIGN | 15_68801018 | 0.225991065 | ANP32A | 6_134498914 | 0.194529874 | LINC01010 |
| 16_89830866 | 0.153278348 | SPIRE2 | 11_101917694 | 0.225964091 | KIAA1377 | 21_14166129 | 0.19452891 | LIPI |
| 17_37874488 | 0.153265901 | YWHAEP7 | 2_42553872 | 0.225936717 | MTA3 | 12_111992365 | 0.194498881 | TMEM116 |
| 1_244585928 | 0.153201223 | C1orf101 | 20_59643833 | 0.22591441 | PHACTR3 | 12_112108179 | 0.194498881 | NAA25 |
| 3_131155942 | 0.153190032 | NEK11 | 13_49924989 | 0.225835363 | SPRYD7 | 2_70008474 | 0.194487064 | PCBP1-AS1 |
| 1_96371152 | 0.153182652 | LOC101928241 | 9_20428656 | 0.225755079 | MLLT3 | 2_212003457 | 0.194457387 | ERBB4 |
| 4_6462438 | 0.153166622 | PPP2R2C | 7_6214218 | 0.2257154 | CYTH3 | 20_20049310 | 0.194451492 | CRNKL1 |
| 3_191337828 | 0.153163173 | CCDC50 | 1_180819887 | 0.225693577 | XPR1 | 1_40703448 | 0.194413025 | NFYC |
| 11_44816945 | 0.153125328 | TSPAN18 | 15_48548847 | 0.225584 | FBN1 | 4_88075950 | 0.194407815 | PKD2 |
| 3_720223 | 0.153116116 | LINC01266 | 15_48571077 | 0.225584 | FBN1 | 6_32660018 | 0.194379134 | HLA-DQB1 |
| 22_20785068 | 0.153096415 | PI4KA | 7_6189974 | 0.225556661 | CYTH3 | 2_44352332 | 0.194356929 | PREPL |
| 6_21770553 | 0.15308946 | CASC15 | 10_50317847 | 0.225552914 | SGMS1 | 2_163613534 | 0.194326325 | FIGN |
| 5_106827595 | 0.153075888 | LOC102467213 | 1_35967404 | 0.225528469 | AGO3 | 15_83838005 | 0.194320519 | ADAMTSL3 |
| 2_30962156 | 0.153072167 | GALNT14 | 15_100300285 | 0.225526744 | ADAMTS17 | 2_16639541 | 0.194311025 | FAM49A |
| 11_113331119 | 0.153055734 | TTC12 | 1_244448413 | 0.225269846 | ADSS | 9_10064950 | 0.194297828 | PTPRD |
| 11_113341091 | 0.153055734 | TTC12 | 7_80872924 | 0.225256988 | SEMA3C | 11_118470497 | 0.194290657 | KMT2A |
| 2_106823425 | 0.153038547 | ST6GAL2 | 7_104881186 | 0.225227915 | LHFPL3 | 1_244414592 | 0.194221281 | ADSS |
| 8_3026651 | 0.153010706 | CSMD1 | 5_162127394 | 0.225187675 | GABRG2 | 12_112277230 | 0.194187293 | HECTD4 |
| 9_132324333 | 0.153008218 | SETX | 19_949284 | 0.225101389 | ARID3A | 2_70073359 | 0.19418114 | PCBP1-AS1 |
| 10_118169163 | 0.153002195 | CASC2 | 7_104891759 | 0.225038981 | LHFPL3 | 10_52166914 | 0.194161609 | PRKG1 |
| 15_33854971 | 0.152980916 | RYR3 | 4_145175868 | 0.22498809 | OTUD4 | 20_63444998 | 0.194127127 | KCNQ2 |
| 13_101152930 | 0.152972171 | NALCN | 4_100489233 | 0.224976044 | EMCN | 2_163616060 | 0.19412118 | FIGN |
| 12_131191500 | 0.152948246 | LINC01257 | 4_100491884 | 0.224976044 | EMCN | 2_64129440 | 0.194070642 | PELI1 |
| 12_131191851 | 0.152948246 | LINC01257 | 12_44816876 | 0.224961479 | NELL2 | 6_32642668 | 0.194061496 | HLA-DQA1 |
| 12_29633854 | 0.152945259 | TMTC1 | 7_147245180 | 0.224940859 | CNTNAP2 | 14_67759581 | 0.194025452 | ZFYVE26 |
| 6_169655654 | 0.152942031 | WDR27 | 10_60547720 | 0.224910268 | ANK3 | 1_244561248 | 0.193982888 | C1orf101 |
| 1_231409957 | 0.152920664 | EGLN1 | 1_47311284 | 0.224897201 | STIL | 1_244565177 | 0.193954124 | C1orf101 |
| 12_54974821 | 0.152899914 | TESPA1 | 1_231189883 | 0.224838777 | TRIM67 | 1_244581266 | 0.193954124 | C1orf101 |
| 22_49567530 | 0.152891773 | C22orf34 | 1_231192113 | 0.224838777 | TRIM67 | 1_244583991 | 0.193954124 | C1orf101 |
| 22_49567631 | 0.152891773 | C22orf34 | 15_80477882 | 0.224833235 | ARNT2 | 10_13943777 | 0.193939585 | FRMD4A |
| 22_49568396 | 0.152891773 | C22orf34 | 11_73966846 | 0.224800433 | DNAJB13 | 9_20375687 | 0.193906915 | MLLT3 |
| 22_49568806 | 0.152891773 | C22orf34 | 19_364353 | 0.224742279 | THEG | 12_121412645 | 0.193903315 | RNF34 |
| 22_49569306 | 0.152891773 | C22orf34 | 4_113163782 | 0.224710583 | ANK2 | 1_169593666 | 0.193876701 | SELP |
| 9_132354713 | 0.152878617 | SETX | 4_89746868 | 0.224702611 | SNCA | 6_70289120 | 0.19387662 | COL9A1 |
| 7_66540984 | 0.152855626 | LOC493754 | 16_10784057 | 0.224667409 | TVP23A | 17_37846119 | 0.193868965 | YWHAEP7 |
| 2_71128381 | 0.152819899 | MCEE | 16_81786322 | 0.224647844 | PLCG2 | 3_183300871 | 0.193828637 | MCF2L2 |
| 16_2340431 | 0.152799502 | ABCA3 | 1_35502714 | 0.224602934 | KIAA0319L | 5_5435660 | 0.193821663 | KIAA0947 |
| 5_36255262 | 0.152758458 | RANBP3L | 13_107749320 | 0.224576515 | FAM155A | 2_70045140 | 0.193753055 | PCBP1-AS1 |
| 18_53257495 | 0.152744862 | DCC | 13_97305809 | 0.224559 | MBNL2 | 10_96305051 | 0.193692217 | DNTT |
| 3_129533950 | 0.152724465 | RHO | 17_59868109 | 0.22451786 | TUBD1 | 1_190475849 | 0.193640427 | BRINP3 |
| 6_159234294 | 0.152723069 | FNDC1 | 7_6159441 | 0.224444131 | USP42 | 12_27366199 | 0.193639291 | ARNTL2 |
| 7_117289011 | 0.152720215 | WNT2 | 10_66511482 | 0.22444128 | CTNNA3 | 2_37264523 | 0.193590503 | PRKD3 |
| 2_46169222 | 0.152705122 | PRKCE | 12_48761613 | 0.224400295 | LINC00935 | 1_244576975 | 0.193577847 | C1orf101 |
| 8_72821446 | 0.152675224 | KCNB2 | 3_192341940 | 0.224378644 | FGF12 | 3_688608 | 0.193543919 | LINC01266 |
| 2_167019079 | 0.152641262 | XIRP2 | 11_67423978 | 0.224341718 | CARNS1 | 2_70162337 | 0.19354318 | C2orf42 |
| 7_104840268 | 0.152633641 | LHFPL3 | 9_17347057 | 0.22431046 | CNTLN | 12_12388749 | 0.193516356 | LOH12CR1 |
| 1_243756471 | 0.152618303 | AKT3 | 15_48629852 | 0.224308308 | FBN1 | 2_16636925 | 0.193495733 | FAM49A |
| 8_10421973 | 0.152545709 | MSRA | 4_185172851 | 0.224275636 | KIAA1430 | 7_154692782 | 0.193487288 | DPP6 |
| 8_10421993 | 0.152545709 | MSRA | 8_40640478 | 0.224272293 | ZMAT4 | 12_27258767 | 0.193462223 | STK38L |
| 19_37971517 | 0.152524755 | SIPA1L3 | 5_131819769 | 0.224271397 | LOC728637 | 19_955552 | 0.193454879 | ARID3A |
| 20_5962309 | 0.15250328 | MCM8 | 15_97553346 | 0.224248093 | LOC101927310 | 15_33600318 | 0.193405613 | RYR3 |
| 7_129148389 | 0.152486118 | TSPAN33 | 1_231767264 | 0.224219851 | DISC1 | 15_83831709 | 0.193402318 | ADAMTSL3 |
| 22_32666850 | 0.152427007 | SYN3 | 9_3486257 | 0.224136383 | RFX3 | 9_67197289 | 0.193363726 | CNTNAP3P2 |
| 8_10395426 | 0.152413277 | MSRA | 14_24192500 | 0.224119238 | TM9SF1 | 5_90966675 | 0.193326435 | GPR98 |
| 3_11801979 | 0.152412334 | TAMM41 | 1_202826134 | 0.224074899 | LOC641515 | 6_128486909 | 0.193314381 | PTPRK |
| 1_97180863 | 0.152385096 | DPYD | 20_19273435 | 0.224066652 | LOC100130264 | 9_6883748 | 0.193300731 | KDM4C |
| 1_65375484 | 0.152369038 | DNAJC6 | 4_108572504 | 0.223972156 | RPL34-AS1 | 3_138448124 | 0.193287178 | ESYT3 |
| 13_98210530 | 0.152340152 | FARP1 | 16_7304888 | 0.223688934 | RBFOX1 | 6_148918138 | 0.193245656 | UST |
| 6_159221828 | 0.152337415 | FNDC1 | 18_52598130 | 0.223683926 | DCC | 14_63939968 | 0.193229456 | PPP2R5E |
| 15_43912864 | 0.152265579 | FRMD5 | 15_40268289 | 0.223683566 | PAK6 | 2_49973972 | 0.193222178 | NRXN1 |
| 3_129387825 | 0.152260779 | RPL32P3 | 19_7419955 | 0.223647478 | ARHGEF18 | 2_50012057 | 0.193222178 | NRXN1 |
| 1_231570775 | 0.152260348 | TSNAX-DISC1 | 19_7419956 | 0.223647478 | ARHGEF18 | 15_101485013 | 0.193197164 | PCSK6 |
| 2_221547817 | 0.15224542 | EPHA4 | 16_89783709 | 0.223637407 | FANCA | 8_116876588 | 0.193160731 | RAD21-AS1 |
| 2_221549440 | 0.15224542 | EPHA4 | 7_40025469 | 0.223636083 | CDK13 | 2_70076646 | 0.193151625 | PCBP1-AS1 |
| 2_221556951 | 0.15224542 | EPHA4 | 22_40522708 | 0.223578214 | LOC101927257 | 7_20010381 | 0.193125956 | LOC101927668 |
| 11_26530586 | 0.152241293 | ANO3 | 2_240492703 | 0.223526077 | ANKMY1 | 17_3876759 | 0.193123727 | CAMKK1 |
| 9_3504288 | 0.152237361 | RFX3 | 11_46096409 | 0.223432367 | PHF21A | 17_3876764 | 0.193123727 | CAMKK1 |
| 12_16347514 | 0.15220102 | MGST1 | 12_330464 | 0.223427813 | KDM5A | 17_3876770 | 0.193123727 | CAMKK1 |
| 6_21764778 | 0.152164559 | CASC15 | 12_383245 | 0.223427813 | KDM5A | 4_37842687 | 0.193020756 | PGM2 |
| 2_71354444 | 0.152154602 | ZNF638 | 10_50322076 | 0.223410531 | SGMS1 | 2_172781523 | 0.192989931 | RAPGEF4 |
| 6_21765370 | 0.152146715 | CASC15 | 3_71080728 | 0.223385164 | FOXP1 | 5_10451305 | 0.192945742 | ROPN1L |
| 15_82398639 | 0.152058896 | UBE2Q2P2 | 19_14620471 | 0.223346733 | EMR3 | 2_47073890 | 0.192911101 | TTC7A |
| 4_6797492 | 0.152039302 | KIAA0232 | 11_46037743 | 0.223321341 | PHF21A | 6_32524417 | 0.192910814 | HLA-DRB5 |
| 7_8435517 | 0.152032226 | NXPH1 | 17_75816828 | 0.223264949 | UNK | 13_110543779 | 0.192851541 | RAB20 |
| 5_106873275 | 0.152028762 | LOC102467213 | 9_17377217 | 0.223248092 | CNTLN | 7_120451606 | 0.192849055 | KCND2 |
| 2_173170361 | 0.15200612 | ZAK | 4_108587549 | 0.223234431 | RPL34-AS1 | 6_29945862 | 0.192824381 | HLA-A |
| 19_37958074 | 0.151991791 | SIPA1L3 | 18_24144520 | 0.223173216 | CABYR | 3_138581927 | 0.19279932 | CEP70 |
| 19_37969014 | 0.151991791 | SIPA1L3 | 5_150127140 | 0.223112369 | PDGFRB | 8_72838184 | 0.192774837 | KCNB2 |
| 10_75789413 | 0.151978821 | C10orf11 | 5_60003708 | 0.223061627 | PDE4D | 8_72842295 | 0.192774837 | KCNB2 |
| 16_10788136 | 0.151978611 | TVP23A | 13_97277426 | 0.223043348 | MBNL2 | 8_72848124 | 0.192774837 | KCNB2 |

| | | | | | | | | |
|---|---|---|---|---|---|---|---|---|
| 13_28237742 | 0.151968026 | PAN3 | 6_6202070 | 0.223035522 | F13A1 | 8_72849743 | 0.192774837 | KCNB2 |
| 11_100201623 | 0.151963084 | CNTN5 | 7_6138102 | 0.223001226 | USP42 | 8_72849838 | 0.192774837 | KCNB2 |
| 5_109772237 | 0.151949292 | MAN2A1 | 15_48562066 | 0.222934418 | FBN1 | 8_72857474 | 0.192774837 | KCNB2 |
| 5_109772581 | 0.151949292 | MAN2A1 | 1_244434624 | 0.222902171 | ADSS | 8_72863800 | 0.192774837 | KCNB2 |
| 6_144538372 | 0.151909485 | UTRN | 12_99962811 | 0.222809372 | ANKS1B | 12_32894088 | 0.192768285 | PKP2 |
| 6_144538373 | 0.151909485 | UTRN | 13_85412084 | 0.222796347 | LINC00351 | 19_56032116 | 0.192749342 | NLRP5 |
| 6_144582381 | 0.151909485 | UTRN | 10_12368455 | 0.222764009 | CAMK1D | 12_112372957 | 0.192691559 | HECTD4 |
| 2_231318878 | 0.151905049 | ARMC9 | 10_12368456 | 0.222764009 | CAMK1D | 6_29944733 | 0.192594417 | HLA-A |
| 7_7093524 | 0.151900389 | LOC100131257 | 12_315869 | 0.222742182 | KDM5A | 10_28255857 | 0.192594148 | MPP7 |
| 9_36842687 | 0.151900127 | PAX5 | 12_336571 | 0.222742182 | KDM5A | 15_57479481 | 0.19256505 | CGNL1 |
| 2_37169637 | 0.151886724 | SULT6B1 | 14_61576566 | 0.222707211 | FLJ22447 | 17_19252848 | 0.192494505 | EPN2 |
| 1_47938094 | 0.151861271 | TRABD2B | 2_46383340 | 0.222694711 | EPAS1 | 3_64623558 | 0.192460825 | ADAMTS9 |
| 17_37870305 | 0.151813949 | YWHAEP7 | 14_61542888 | 0.222656185 | PRKCH | 19_15765022 | 0.192455303 | CYP4F24P |
| 17_37870464 | 0.151807726 | YWHAEP7 | 19_14620468 | 0.222647753 | EMR3 | 11_118562137 | 0.192435113 | IFT46 |
| 17_37870365 | 0.151787383 | YWHAEP7 | 15_47706332 | 0.222646158 | SEMA6D | 11_118565041 | 0.192406023 | IFT46 |
| 1_231199611 | 0.151777536 | TRIM67 | 1_35190261 | 0.222584969 | SFPQ | 12_111756923 | 0.192392574 | ACAD10 |
| 2_163616060 | 0.151762262 | FIGN | 15_47726082 | 0.222583515 | SEMA6D | 12_19275075 | 0.192331239 | PLEKHA5 |
| 3_2875771 | 0.151760393 | CNTN4 | 13_43404172 | 0.222569842 | ENOX1 | 3_2473241 | 0.192279061 | CNTN4 |
| 13_28290511 | 0.151736133 | PAN3 | 3_177192284 | 0.222546553 | TBL1XR1 | 6_127307464 | 0.192252674 | ECHDC1 |
| 2_232063138 | 0.151729996 | DIS3L2 | 1_33175005 | 0.222495364 | TRIM62 | 10_73338439 | 0.19223706 | TTC18 |
| 12_32829818 | 0.151711293 | PKP2 | 19_14608629 | 0.222495364 | CLEC17A | 2_50502430 | 0.192234453 | NRXN1 |
| 1_244572955 | 0.151645248 | C1orf101 | 1_160680136 | 0.222461803 | CD48 | 2_217524530 | 0.192234025 | DIRC3 |
| 17_1962297 | 0.151633943 | RTN4RL1 | 19_22601877 | 0.22244686 | GOLGA2P9 | 10_96315737 | 0.192229345 | DNTT |
| 22_46702218 | 0.151578005 | CERK | 9_113909951 | 0.222445275 | ZNF618 | 1_54730235 | 0.192214188 | MROH7-TTC4 |
| 3_2560893 | 0.151573498 | CNTN4 | 13_107710413 | 0.222328686 | FAM155A | 18_43026473 | 0.192124571 | RIT2 |
| 4_108581956 | 0.151511218 | RPL34-AS1 | 13_94913115 | 0.222321661 | LOC101927284 | 12_111718783 | 0.192081722 | ACAD10 |
| 21_16396204 | 0.151506306 | LINC00478 | 20_10286176 | 0.222308603 | SNAP25 | 4_24936732 | 0.192064625 | CCDC149 |
| 6_159224856 | 0.15150224 | FNDC1 | 6_98888247 | 0.222296436 | FBXL4 | 12_48743255 | 0.19205553 | LINC00935 |
| 6_159227026 | 0.15150224 | FNDC1 | 11_118136041 | 0.222211931 | SCN4B | 18_25319761 | 0.192029023 | ZNF521 |
| 6_159227188 | 0.15150224 | FNDC1 | 13_85431896 | 0.2221722 | LINC00351 | 4_38623850 | 0.192017637 | KLF3-AS1 |
| 6_159228713 | 0.15150224 | FNDC1 | 9_17396210 | 0.22215417 | CNTLN | 3_64608450 | 0.19198303 | ADAMTS9 |
| 6_159228715 | 0.15150224 | FNDC1 | 1_231188511 | 0.222057092 | TRIM67 | 3_138516616 | 0.19195323 | CEP70 |
| 1_113414763 | 0.15143894 | MAGI3 | 1_231201200 | 0.22202486 | TRIM67 | 2_113924055 | 0.19194874 | ACTR3 |
| 20_19218974 | 0.151394269 | SLC24A3 | 11_85154137 | 0.221995663 | DLG2 | 8_29249431 | 0.191870387 | KIF13B |
| 12_48340826 | 0.151392014 | ZNF641 | 2_71422595 | 0.221970722 | ZNF638 | 11_118558076 | 0.191864098 | IFT46 |
| 5_109726259 | 0.15138142 | MAN2A1 | 4_161573563 | 0.221957152 | FSTL5 | 19_51716824 | 0.191837632 | HAS1 |
| 5_109742339 | 0.15138142 | MAN2A1 | 7_149267445 | 0.22190337 | ZNF783 | 21_14202076 | 0.19180268 | LIPI |
| 5_109746893 | 0.15138142 | MAN2A1 | 7_151676278 | 0.221859572 | PRKAG2 | 3_55821994 | 0.19180268 | ERC2 |
| 19_16155087 | 0.1513657 | HSH2D | 18_52597876 | 0.22181457 | DCC | 5_178556284 | 0.191785494 | COL23A1 |
| 6_144516982 | 0.151343179 | UTRN | 20_59644709 | 0.221747419 | PHACTR3 | 6_155440112 | 0.191686729 | NOX3 |
| 5_178469679 | 0.151323968 | COL23A1 | 15_42766447 | 0.221701852 | TTBK2 | 18_74443835 | 0.19167916 | FAM69C |
| 9_9786679 | 0.151279534 | PTPRD | 1_220752743 | 0.221668998 | 2-Mar | 16_12347826 | 0.191641771 | SNX29 |
| 4_31085854 | 0.151259487 | PCDH7 | 8_40654417 | 0.221660409 | ZMAT4 | 15_39863065 | 0.19163432 | GPR176 |
| 1_162866771 | 0.151245324 | C1orf110 | 7_2267375 | 0.221625853 | SNX8 | 5_94843257 | 0.191623942 | MCTP1 |
| 3_129490335 | 0.151237039 | IFT122 | 2_26502956 | 0.221601055 | OTOF | 8_47589029 | 0.191621838 | SPIDR |
| 12_95650514 | 0.151190141 | PGAM1P5 | 1_228707128 | 0.221558135 | RHOU | 11_118522681 | 0.19160952 | KMT2A |
| 15_82398637 | 0.151173487 | UBE2Q2P2 | 1_35897874 | 0.221556724 | AGO1 | 6_32665725 | 0.191606529 | HLA-DQB1 |
| 15_82398638 | 0.151173487 | UBE2Q2P2 | 8_17289940 | 0.221546557 | VPS37A | 6_151389942 | 0.191593524 | ZBTB2 |
| 6_159238178 | 0.151134094 | FNDC1 | 1_231187003 | 0.221524503 | LOC149373 | 20_63406396 | 0.191590028 | KCNQ2 |
| 6_159238199 | 0.151134094 | FNDC1 | 6_34322756 | 0.221518712 | NUDT3 | 4_164052146 | 0.191571832 | 1-Mar |
| 6_39810635 | 0.151127999 | DAAM2 | 9_8791718 | 0.221479947 | PTPRD | 7_104834016 | 0.191552959 | LHFPL3 |
| 22_42508933 | 0.151108629 | RRP7A | 7_6173280 | 0.221451648 | CYTH3 | 2_55676612 | 0.191505362 | PNPT1 |
| 7_71874280 | 0.151089268 | CALN1 | 5_16816041 | 0.221414277 | MYO10 | 6_68898015 | 0.191502142 | BAI3 |
| 10_76942095 | 0.1510704 | KCNMA1 | 10_50318996 | 0.221355645 | SGMS1 | 2_50004731 | 0.191477672 | NRXN1 |
| 1_231178113 | 0.151067033 | TRIM67 | 14_50881494 | 0.221345733 | ABHD12B | 1_244586660 | 0.191470227 | C1orf101 |
| 22_49570015 | 0.15101829 | C22orf34 | 9_3487091 | 0.221278449 | RFX3 | 1_244587437 | 0.191470227 | C1orf101 |
| 10_86686130 | 0.151016955 | LDB3 | 1_169525165 | 0.221230523 | F5 | 2_70072022 | 0.191464859 | PCBP1-AS1 |
| 22_42517146 | 0.150967005 | RRP7A | 1_35189164 | 0.221207331 | SFPQ | 6_29943462 | 0.191394833 | HLA-A |
| 17_65154800 | 0.150963996 | RGS9 | 1_35189176 | 0.221207331 | SFPQ | 10_11755187 | 0.191358551 | ECHDC3 |
| 4_6473139 | 0.150962786 | PPP2R2C | 22_40447361 | 0.221166861 | MKL1 | 9_36058329 | 0.191334766 | RECK |
| 6_166814252 | 0.150962786 | RPS6KA2 | 1_9983358 | 0.221160514 | NMNAT1 | 12_27269708 | 0.191308781 | STK38L |
| 4_31099848 | 0.15095909 | PCDH7 | 22_40535999 | 0.221156563 | MKL1 | 3_41493044 | 0.191303408 | ULK4 |
| 3_131055112 | 0.150942564 | NEK11 | 8_27830563 | 0.221082036 | PBK | 2_55688232 | 0.191297638 | PNPT1 |
| 20_58494275 | 0.150932718 | APCDD1L | 15_100299572 | 0.221050265 | ADAMTS17 | 12_112069757 | 0.191209866 | NAA25 |
| 1_111854223 | 0.150916416 | KCND3 | 8_71321208 | 0.22102174 | EYA1 | 2_50011824 | 0.191079569 | NRXN1 |
| 1_111854855 | 0.150916416 | KCND3 | 6_100416242 | 0.220959337 | SIM1 | 2_50012439 | 0.191079569 | NRXN1 |
| 5_109821970 | 0.150892459 | MAN2A1 | 6_100417930 | 0.220959337 | SIM1 | 2_54843371 | 0.190943262 | EML6 |
| 3_55069196 | 0.15083822 | CACNA2D3 | 6_6204108 | 0.220954595 | F13A1 | 10_16955455 | 0.190858095 | CUBN |
| 6_21795214 | 0.150812198 | CASC15 | 14_92487979 | 0.220946459 | SLC24A4 | 10_76407630 | 0.190844288 | C10orf11 |
| 3_55069255 | 0.150771845 | CACNA2D3 | 11_129404139 | 0.22088637 | BARX2 | 1_244561686 | 0.190837024 | C1orf101 |
| 1_155827874 | 0.150771441 | GON4L | 10_50314754 | 0.220883535 | SGMS1 | 4_155366252 | 0.190819163 | MAP9 |
| 1_48579930 | 0.150766351 | AGBL4 | 7_6150149 | 0.220812817 | USP42 | 19_440005 | 0.19080506 | SHC2 |
| 12_127917890 | 0.15075751 | LINC00507 | 7_103744626 | 0.2207395 | RELN | 2_16632494 | 0.190803189 | FAM49A |
| 22_32691268 | 0.150724579 | SYN3 | 4_188101562 | 0.220636143 | TRIML2 | 2_16632496 | 0.190803189 | FAM49A |
| 1_231547622 | 0.150678524 | TSNAX | 15_27497255 | 0.220633169 | GABRG3 | 20_62744775 | 0.190780853 | NTSR1 |
| 6_144569896 | 0.150678524 | UTRN | 13_43400334 | 0.220631368 | ENOX1 | 2_16634576 | 0.190764122 | FAM49A |
| 5_167834377 | 0.150665386 | TENM2 | 10_119135182 | 0.220568965 | FAM45A | 3_56942337 | 0.190741457 | ARHGEF3 |
| 4_6447807 | 0.150644491 | PPP2R2C | 9_135576557 | 0.220559318 | LOC100130954 | 3_138500152 | 0.190732353 | CEP70 |
| 3_55063809 | 0.150636092 | CACNA2D3 | 10_50325003 | 0.220546151 | SGMS1 | 21_14197852 | 0.190718992 | LIPI |
| 8_3027023 | 0.15063515 | CSMD1 | 22_40566804 | 0.220478383 | MKL1 | 12_112333990 | 0.190662654 | HECTD4 |
| 5_109841537 | 0.150624295 | MAN2A1 | 14_67582047 | 0.220472909 | PLEKHH1 | 6_39837446 | 0.190658274 | DAAM2 |
| 20_46200205 | 0.150611733 | CDH22 | 5_119455950 | 0.220373652 | HSD17B4 | 18_9724281 | 0.190630864 | RAB31 |
| 22_32696683 | 0.1505632 | SYN3 | 3_120222474 | 0.220366296 | GPR156 | 18_9724282 | 0.190630864 | RAB31 |
| 10_66876567 | 0.150522599 | CTNNA3 | 2_113870009 | 0.220330218 | LOC101060091 | 4_41443199 | 0.190626723 | LIMCH1 |
| 2_71138675 | 0.150522599 | MPHOSPH10 | 6_101722980 | 0.220329307 | GRIK2 | 18_63714094 | 0.190602469 | SERPINB11 |
| 2_71143095 | 0.150522599 | MPHOSPH10 | 10_123157079 | 0.220285521 | BUB3 | 11_118536355 | 0.190596582 | TMEM25 |
| 6_159214911 | 0.150503282 | FNDC1 | 12_347319 | 0.220285159 | KDM5A | 12_7381963 | 0.190595284 | CD163L1 |
| 17_79140596 | 0.150492007 | RBFOX3 | 7_149274841 | 0.220221261 | ZNF783 | 4_24941546 | 0.190552759 | CCDC149 |
| 15_40362016 | 0.150487038 | DISP2 | 4_182658356 | 0.220181261 | TENM3 | 2_70081023 | 0.190536342 | PCBP1-AS1 |
| 15_40363940 | 0.150487038 | DISP2 | 4_56256823 | 0.220096371 | KIAA1211 | 12_27316340 | 0.190491195 | STK38L |
| 15_40370900 | 0.150487038 | DISP2 | 22_40466970 | 0.220039324 | MKL1 | 10_76418772 | 0.190481998 | C10orf11 |
| 5_109762320 | 0.150477202 | MAN2A1 | 6_101670471 | 0.22002378 | GRIK2 | 1_244576214 | 0.190481998 | C1orf101 |
| 5_109771388 | 0.150477202 | MAN2A1 | 9_121092124 | 0.219962389 | CNTRL | 6_151417816 | 0.190399312 | RMND1 |
| 3_2878039 | 0.150465042 | CNTN4 | 1_116927526 | 0.219916511 | PTGFRN | 17_19246522 | 0.190396479 | EPN2 |
| 13_101155914 | 0.150448973 | NALCN | 1_16457708 | 0.219901073 | NECAP2 | 2_50282833 | 0.190396047 | NRXN1 |
| 5_160770914 | 0.150438003 | ATP10B | 9_3475451 | 0.219899645 | RFX3 | 12_120990800 | 0.190339373 | HNF1A |
| 15_48643656 | 0.150437798 | FBN1 | 6_101711345 | 0.219878048 | GRIK2 | 12_131966910 | 0.190337172 | EP400 |

| | | | | | | | | |
|---|---|---|---|---|---|---|---|---|
| 3_55063478 | 0.150437798 | CACNA2D3 | 6 34345801 | 0.219811763 | NUDT3 | 14 63381557 | 0.190236701 | PPP2R5E |
| 20 46955191 | 0.150422544 | EYA2 | 9 92999747 | 0.219798023 | FGD3 | 11 46872218 | 0.190223594 | LRP4 |
| 5 33570496 | 0.150422544 | ADAMTS12 | 1 202747373 | 0.219790451 | KDM5B | 2 211752506 | 0.190213513 | ERBB4 |
| 1_231215991 | 0.15040974 | TRIM67 | 22_29310262 | 0.219783487 | GAS2L1 | 4_188093698 | 0.190200698 | TRIML2 |
| 17 9215961 | 0.150376699 | NTN1 | 4 146305408 | 0.219702846 | SLC10A7 | 3 64627888 | 0.190153045 | ADAMTS9 |
| 16 84414094 | 0.150370349 | ATP2C2 | 7 6144992 | 0.219695917 | USP42 | 12 120794449 | 0.190145014 | SPPL3 |
| 1 62117689 | 0.150348782 | INADL | 16 87216376 | 0.219694435 | LOC101928708 | 12 27257809 | 0.190143064 | STK38L |
| 3 129503446 | 0.1503434 | IFT122 | 17 16306878 | 0.219662479 | PIGL | 5 19549785 | 0.190138578 | CDH18 |
| 7 143450626 | 0.1503434 | EPHA1-AS1 | 5 170025846 | 0.219659473 | DOCK2 | 15 52244112 | 0.19005942 | MYO5C |
| 8 10396027 | 0.150338159 | MSRA | 4 48852890 | 0.219555531 | OCIAD1 | 12 27257489 | 0.190027515 | STK38L |
| 6 166806047 | 0.150332243 | RPS6KA2 | 9 17210041 | 0.219548301 | CNTLN | 2 50005248 | 0.189949209 | NRXN1 |
| 6 159233455 | 0.150327345 | FNDC1 | 1 52960524 | 0.219521864 | SCP2 | 11 118582966 | 0.189902126 | ARCN1 |
| 19 37961895 | 0.150305329 | SIPA1L3 | 9 17407204 | 0.2194747 | CNTLN | 1_167126644 | 0.189888605 | DUSP27 |
| 20 46961800 | 0.150289524 | EYA2 | 11 45261511 | 0.219468937 | SYT13 | 20 62817705 | 0.189844296 | COL9A3 |
| 7 91156400 | 0.150289524 | CDK14 | 12 53539050 | 0.219450015 | ATF7 | 20 20022103 | 0.189779056 | NAA20 |
| 22 17494000 | 0.150279736 | CECR2 | 5 150111245 | 0.219450015 | CSF1R | 17 10137701 | 0.189759297 | GAS7 |
| 15 27490407 | 0.150262089 | GABRG3 | 3 155547500 | 0.219443943 | PLCH1 | 15 47845511 | 0.189744626 | LOC101928442 |
| 16 89132354 | 0.150246299 | ACSF3 | 2 74292854 | 0.219437672 | SLC4A5 | 3 125128183 | 0.189726852 | SLC12A8 |
| 19 2602241 | 0.150246299 | GNG7 | 9 129996499 | 0.219436154 | FNBP1 | 10 96317405 | 0.189703628 | DNTT |
| 7 103436244 | 0.150240166 | SLC26A5 | 19 16022784 | 0.219424428 | LINC00661 | 2 64121885 | 0.189690167 | PELI1 |
| 7 103437498 | 0.150240166 | SLC26A5 | 19 16022791 | 0.219424428 | LINC00661 | 8 72835274 | 0.189684817 | KCNB2 |
| 7 103440086 | 0.150240166 | SLC26A5 | 2 190314284 | 0.219423944 | HIBCH | 8 72847991 | 0.189684817 | KCNB2 |
| 4 89732553 | 0.150213515 | SNCA | 12 45042651 | 0.219391432 | DBX2 | 8 72849589 | 0.189684817 | KCNB2 |
| 12 131198559 | 0.150191422 | LINC01257 | 3 64800616 | 0.219372959 | ADAMTS9-AS2 | 22 50590618 | 0.189679762 | CHKB-AS1 |
| 14 78898838 | 0.150154256 | NRXN3 | 1 20115969 | 0.219349304 | PLA2G2D | 15 30766675 | 0.189664137 | LOC100288637 |
| 15 47749075 | 0.150154256 | SEMA6D | 5 167357580 | 0.219330198 | TENM2 | 12 19275554 | 0.189642632 | PLEKHA5 |
| 11 113390595 | 0.150147791 | ANKK1 | 12 52938039 | 0.219243328 | KRT8 | 12 19276428 | 0.189642632 | PLEKHA5 |
| 6 170403168 | 0.150142088 | FAM120B | 13 97290056 | 0.219222991 | MBNL2 | 12 27359526 | 0.189624383 | ARNTL2 |
| 6 169653519 | 0.150115379 | WDR27 | 14_61561192 | 0.21909984 | LOC101927780 | 11_118534590 | 0.189589503 | TMEM25 |
| 6 169657764 | 0.150115379 | WDR27 | 17 46763851 | 0.219090299 | WNT3 | 14 63375173 | 0.189585733 | PPP2R5E |
| 2 25449600 | 0.15010259 | DTNB | 18 52607764 | 0.219088377 | DCC | 16 16324951 | 0.189582273 | LOC100288162 |
| 5_109766170 | 0.150088051 | MAN2A1 | 3_55069196 | 0.219084649 | CACNA2D3 | 2_16635589 | 0.189553459 | FAM49A |
| 8 120617932 | 0.150073669 | SNTB1 | 13 34570118 | 0.219022716 | LINC00457 | 8 140398204 | 0.189540075 | TRAPPC9 |
| 2 71204755 | 0.150069377 | PAIP2B | 1 2109014 | 0.219021521 | PRKCZ | 11 118602516 | 0.189481201 | ARCN1 |
| 11 101032614 | 0.150059169 | PGR | 1 12443878 | 0.218992151 | VPS13D | 2 241945925 | 0.189479094 | LINC01237 |
| 10 15363213 | 0.15002031 | FAM171A1 | 10 68424516 | 0.218952573 | DNA2 | 17 40640226 | 0.189425539 | SMARCE1 |
| 9 17358382 | 0.149999735 | CNTLN | 5 150132229 | 0.218879896 | PDGFRB | 9 87528174 | 0.189383149 | DAPK1 |
| 22 49565696 | 0.149979789 | C22orf34 | 22 17998550 | 0.218870433 | MICAL3 | 4 88009736 | 0.189366393 | PKD2 |
| 18_64130436 | 0.149976497 | LINC00305 | 19_16034327 | 0.218836807 | LINC00905 | 2_229141654 | 0.189237558 | PID1 |
| 6 169770465 | 0.14996245 | ERMARD | 3 151874158 | 0.218762824 | AADACL2-AS1 | 20 63468154 | 0.189205533 | KCNQ2 |
| 5 13699019 | 0.149959678 | DNAH5 | 8 132914289 | 0.218762748 | TG | 20 63468155 | 0.189205533 | KCNQ2 |
| 7_2566456 | 0.149940671 | IQCE | 6_75450653 | 0.218678558 | FILIP1 | 12_112223781 | 0.189191986 | HECTD4 |
| 19 37959696 | 0.149923753 | SIPA1L3 | 13 70107466 | 0.218597577 | ATXN8OS | 6 151438057 | 0.189179152 | RMND1 |
| 8 120620923 | 0.149904386 | SNTB1 | 6 98878713 | 0.218468268 | FBXL4 | 18 63708741 | 0.189158629 | SERPINB11 |
| 1_244563357 | 0.149887112 | C1orf101 | 9_113904546 | 0.21836768 | ZNF618 | 18_13535211 | 0.189144176 | LDLRAD4 |
| 1 244579409 | 0.149887112 | C1orf101 | 4 56257406 | 0.218351641 | KIAA1211 | 16 24274197 | 0.189139134 | CACNG3 |
| 1 244584926 | 0.149887112 | C1orf101 | 12 80874992 | 0.218312166 | LIN7A | 2 55675260 | 0.189103969 | PNPT1 |
| 19_38622222 | 0.14988269 | EIF3K | 12_80884133 | 0.218312166 | LIN7A | 3_138615432 | 0.1890632 | FAIM |
| 5 126626106 | 0.149875089 | PHAX | 1 35753408 | 0.218258883 | CLSPN | 14 70940854 | 0.189054702 | PCNX |
| 16 58720984 | 0.149870313 | GOT2 | 4 146790061 | 0.218248705 | TTC29 | 11 118541581 | 0.189036225 | TMEM25 |
| 12 4661582 | 0.149849506 | NDUFA9 | 1 13589965 | 0.218242892 | PDPN | 12 27302561 | 0.189009107 | STK38L |
| 7 24847148 | 0.14983412 | OSBPL3 | 16 10794937 | 0.218204226 | TVP23A | 7 26800297 | 0.188944001 | SKAP2 |
| 2 12485179 | 0.149809497 | LOC100506457 | 7 149280119 | 0.218130205 | ZNF783 | 11 78959946 | 0.188940175 | TENM4 |
| 6 162194303 | 0.149784461 | PARK2 | 3 155671793 | 0.218118439 | PLCH1 | 6 21857795 | 0.188929789 | CASC15 |
| 15_66057426 | 0.149769966 | MEGF11 | 1_53021649 | 0.218113332 | SCP2 | 14_61525094 | 0.188911741 | PRKCH |
| 13 57650986 | 0.149753468 | PCDH17 | 1 44551761 | 0.21810269 | RNF220 | 1 244586792 | 0.18890699 | C1orf101 |
| 13 57672846 | 0.149753468 | PCDH17 | 6 47715213 | 0.218023969 | GPR115 | 1 244587315 | 0.18890699 | C1orf101 |
| 1_44548078 | 0.149725036 | RNF220 | 3_14175789 | 0.217962828 | XPC | 10_45426429 | 0.188802683 | ALOX5 |
| 12 121538271 | 0.149724156 | KDM2B | 12 81440665 | 0.21791323 | PPFIA2 | 7 149866805 | 0.188792434 | ZNF862 |
| 6 144537081 | 0.149721775 | UTRN | 11 129407134 | 0.217901073 | BARX2 | 7 44686436 | 0.188792122 | OGDH |
| 4_65677275 | 0.149672476 | LOC100144602 | 1_244433176 | 0.217884007 | ADSS | 1_121287100 | 0.188791467 | SRGAP2C |
| 8 63191534 | 0.149642749 | YTHDF3 | 16 15997984 | 0.217881194 | ABCC1 | 6 30168200 | 0.188783429 | TRIM15 |
| 6 127307464 | 0.149641177 | ECHDC1 | 10 125879019 | 0.217872627 | DHX32 | 2 70076309 | 0.188768137 | PCBP1-AS1 |
| 6 159227629 | 0.149638546 | FNDC1 | 8 14558611 | 0.217850771 | SGCZ | 3 138612772 | 0.188680345 | FAIM |
| 1 38926997 | 0.149630602 | RHBDL2 | 3 64800754 | 0.217846496 | ADAMTS9-AS2 | 6 31357114 | 0.188636989 | HLA-B |
| 18 14219527 | 0.149596317 | ANKRD20A5P | 5 119501259 | 0.217846496 | HSD17B4 | 3 139995479 | 0.188635885 | CLSTN2 |
| 3 30775012 | 0.149587352 | GADL1 | 4 56211869 | 0.217839662 | KIAA1211 | 2 69974365 | 0.188615431 | PCBP1-AS1 |
| 19 37960652 | 0.149577409 | SIPA1L3 | 8 40625243 | 0.21783371 | ZMAT4 | 3 2528718 | 0.188587561 | CNTN4 |
| 19 37968254 | 0.149577409 | SIPA1L3 | 19 29525361 | 0.217638608 | LOC284395 | 15 83830031 | 0.188566656 | ADAMTSL3 |
| 2 167000718 | 0.149577104 | XIRP2 | 5 150475969 | 0.217625369 | LOC102546298 | 17 19330892 | 0.188560972 | EPN2 |
| 16 10792966 | 0.149567863 | TVP23A | 3 155633658 | 0.217600193 | PLCH1 | 11 118535274 | 0.188560242 | TMEM25 |
| 10 50459489 | 0.1495267 | SGMS1 | 8 76441786 | 0.217576084 | LINC01111 | 21 14163269 | 0.188548752 | LIPI |
| 6 159234052 | 0.14951371 | FNDC1 | 1 197040378 | 0.217526874 | F13B | 6 70291666 | 0.188532452 | COL9A1 |
| 6 159237820 | 0.14951371 | FNDC1 | 7 72374423 | 0.217522856 | CALN1 | 1 234248912 | 0.188488504 | SLC35F3 |
| 6 159245004 | 0.14951371 | FNDC1 | 12 121760130 | 0.217495803 | TMEM120B | 1 30712684 | 0.188456143 | MATN1 |
| 18 53285802 | 0.149463969 | DCC | 2 170384519 | 0.217345132 | MYO3B | 7 26816269 | 0.188442657 | SKAP2 |
| 1 47921093 | 0.149443371 | TRABD2B | 7 147321587 | 0.217261828 | CNTNAP2 | 6 29792252 | 0.188434802 | HCG4 |
| 4 31000529 | 0.149443371 | PCDH7 | 11 118112339 | 0.217212414 | TMPRSS4 | 14 105433330 | 0.188425468 | MTA1 |
| 2 170406200 | 0.149401101 | MYO3B | 22 40435884 | 0.217192202 | MKL1 | 3 32485024 | 0.188403963 | CMTM6 |
| 2 71385716 | 0.149344021 | ZNF638 | 7 6148691 | 0.217165468 | USP42 | 3 38590944 | 0.18840351 | SCN5A |
| 5 109811074 | 0.149325961 | MAN2A1 | 13 97276900 | 0.217131804 | MBNL2 | 3 158584671 | 0.18839023 | MLF1 |
| 4 186403764 | 0.14928399 | F11-AS1 | 1 202789045 | 0.217128196 | KDM5B | 2 238193704 | 0.188368249 | ILKAP |
| 6 162190860 | 0.149278531 | PARK2 | 10 60493544 | 0.217094924 | ANK3 | 1 17206687 | 0.188364182 | PADI1 |
| 7 44609380 | 0.149270267 | OGDH | 10 60535056 | 0.217094924 | ANK3 | 1 17206688 | 0.188364182 | PADI1 |
| 3 121489329 | 0.14920591 | POLQ | 3 55069255 | 0.21708914 | CACNA2D3 | 8 72818631 | 0.188277296 | KCNB2 |
| 5_109751735 | 0.1491845 | MAN2A1 | 4_56199694 | 0.217063786 | KIAA1211 | 12_112363203 | 0.18824299 | HECTD4 |
| 6 166803456 | 0.149176058 | RPS6KA2 | 7 149275715 | 0.217053252 | ZNF783 | 2 16635948 | 0.188231123 | FAM49A |
| 6 166820466 | 0.149176058 | RPS6KA2 | 6 32761307 | 0.217053029 | HLA-DQB2 | 2 16637279 | 0.188231123 | FAM49A |
| 7 94417738 | 0.149176058 | COL1A2 | 4 20906135 | 0.217003695 | KCNIP4 | 15 90541790 | 0.188189533 | CRTC3 |
| 2 206739406 | 0.149090763 | MDH1B | 19 16022773 | 0.217001158 | LINC00661 | 12 111971057 | 0.188082503 | TMEM116 |
| 2 206744815 | 0.149090763 | MDH1B | 7 117292480 | 0.216986174 | WNT2 | 12 112096203 | 0.188082503 | NAA25 |
| 7 93527009 | 0.149082343 | CALCR | 4 165035589 | 0.216872355 | TRIM60 | 5 178386213 | 0.188067713 | COL23A1 |
| 10 133239396 | 0.149050536 | VENTX | 6 34422812 | 0.216850655 | RPS10 | 20_20285289 | 0.188029806 | C20orf26 |
| 2 173078346 | 0.14903978 | ZAK | 15 48603234 | 0.216826527 | FBN1 | 12 19267711 | 0.188029603 | PLEKHA5 |
| 5 106839702 | 0.14903707 | LOC102467213 | 3 14165072 | 0.216826527 | XPC | 12 112293103 | 0.188008083 | HECTD4 |
| 20 20021148 | 0.149035322 | NAA20 | 8_94252477 | 0.216724868 | GEM | 17_9164456 | 0.187997359 | NTN1 |
| 1 65375537 | 0.149030427 | DNAJC6 | 1 53021643 | 0.216698672 | SCP2 | 3 64641641 | 0.187996332 | ADAMTS9 |

| | | | | | | | | |
|---|---|---|---|---|---|---|---|---|
| 2_179580581 | 0.149017074 | ZNF385B | 1_2234084 | 0.21665247 | SKI | 2_213361547 | 0.187989291 | SPAG16 |
| 18_53074551 | 0.149012848 | DCC | 1_2234182 | 0.21665247 | SKI | 16_89160390 | 0.18798675 | LINC00304 |
| 12_12557644 | 0.149012735 | DUSP16 | 12_80878787 | 0.216637326 | LIN7A | 1_17206689 | 0.187963708 | PADI1 |
| 12_12558175 | 0.149012735 | DUSP16 | 12_80878788 | 0.216637326 | LIN7A | 15_83871851 | 0.18794199 | ADAMTSL3 |
| 20_46971613 | 0.148968587 | EYA2 | 16_10794077 | 0.216601148 | TVP23A | 11_132270474 | 0.187861385 | NTM |
| 20_46976341 | 0.148968587 | EYA2 | 2_149341735 | 0.216592936 | LYPD6 | 1_244579409 | 0.18783995 | C1orf101 |
| 13_98214803 | 0.148960748 | FARP1 | 13_70104246 | 0.216552889 | KLHL1 | 6_30898565 | 0.187836994 | DDR1 |
| 7_94415762 | 0.148884219 | COL1A2 | 3_55061845 | 0.216552889 | CACNA2D3 | 15_83826118 | 0.187818557 | ADAMTSL3 |
| 19_10259768 | 0.148843335 | MRPL4 | 9_17346031 | 0.216493007 | CNTLN | 1_244583226 | 0.187800258 | C1orf101 |
| 6_144631798 | 0.148840099 | UTRN | 9_17443820 | 0.216493007 | CNTLN | 18_42908314 | 0.187798866 | RIT2 |
| 7_78585956 | 0.148840099 | MAGI2 | 3_124224658 | 0.216425913 | KALRN | 2_49983270 | 0.187792697 | NRXN1 |
| 6_159239223 | 0.148694099 | FNDC1 | 3_124237230 | 0.216425913 | KALRN | 4_186536368 | 0.187726584 | MTNR1A |
| 6_159244914 | 0.148694099 | FNDC1 | 18_64363516 | 0.216408409 | LOC284294 | 2_70031513 | 0.187719721 | PCBP1-AS1 |
| 15_65977144 | 0.1486856 | MEGF11 | 1_111854223 | 0.216357605 | KCND3 | 20_25215277 | 0.187695566 | ENTPD6 |
| 19_37956035 | 0.148668308 | SIPA1L3 | 1_111854345 | 0.216357605 | KCND3 | 2_50003973 | 0.187688829 | NRXN1 |
| 19_37957834 | 0.148668308 | SIPA1L3 | 9_20428139 | 0.216353978 | MLLT3 | 15_63048506 | 0.187648626 | TPM1 |
| 19_37957953 | 0.148668308 | SIPA1L3 | 11_133463285 | 0.216288899 | OPCML | 16_78360971 | 0.187627166 | WWOX |
| 19_37967169 | 0.148668308 | SIPA1L3 | 2_170402412 | 0.216223688 | MYO3B | 3_33043419 | 0.187607233 | GLB1 |
| 1_35890856 | 0.14863217 | AGO1 | 9_17456201 | 0.216137845 | CNTLN | 2_70007875 | 0.18759713 | PCBP1-AS1 |
| 18_42659978 | 0.148589925 | LINC00907 | 14_61574120 | 0.216125376 | FLJ22447 | 2_16633495 | 0.1875798 | FAM49A |
| 17_65164677 | 0.14856966 | RGS9 | 15_78600993 | 0.216098895 | CHRNA3 | 17_5451208 | 0.187551716 | DHX33 |
| 14_79499432 | 0.148559994 | NRXN3 | 1_147250925 | 0.216052698 | CHD1L | 6_33579048 | 0.18754963 | BAK1 |
| 4_6867708 | 0.148527194 | KIAA0232 | 3_71067661 | 0.216038845 | FOXP1 | 7_120484904 | 0.187529624 | KCND2 |
| 8_6615836 | 0.148490747 | MCPH1 | 14_67330480 | 0.216000572 | MPP5 | 6_106658951 | 0.187526294 | QRSL1 |
| 7_77345231 | 0.14848589 | GSAP | 3_71090648 | 0.215991259 | FOXP1 | 10_50207092 | 0.187520572 | ASAH2 |
| 7_104860718 | 0.148311002 | LHFPL3 | 1_161794462 | 0.215972911 | ATF6 | 13_31252400 | 0.187487054 | B3GALTL |
| 6_39807458 | 0.14830223 | DAAM2 | 15_100277180 | 0.215935778 | ADAMTS17 | 1_54728710 | 0.187445065 | MROH7-TTC4 |
| 3_55060528 | 0.148291957 | CACNA2D3 | 10_25446929 | 0.215899972 | GPR158 | 1_6138685 | 0.187421891 | CHD5 |
| 7_131066563 | 0.148271316 | LINC-PINT | 10_50318967 | 0.215861816 | SGMS1 | 1_54729675 | 0.187387074 | MROH7-TTC4 |
| 5_106823913 | 0.148267926 | LOC102467213 | 3_14128169 | 0.215848994 | TMEM43 | 11_134391018 | 0.187363953 | B3GAT1 |
| 7_93506818 | 0.148258874 | CALCR | 3_89359832 | 0.215814862 | EPHA3 | 12_111746083 | 0.187355413 | ACAD10 |
| 16_10778681 | 0.148235298 | TVP23A | 1_35918614 | 0.215745194 | AGO1 | 12_120169266 | 0.187312868 | GCN1L1 |
| 4_186419433 | 0.148226102 | F11-AS1 | 1_35947728 | 0.215745194 | AGO3 | 3_30668019 | 0.187293874 | TGFBR2 |
| 11_113333759 | 0.148219443 | TTC12 | 10_60540625 | 0.215739973 | ANK3 | 19_37592288 | 0.187287787 | ZNF540 |
| 11_113357068 | 0.148219443 | TTC12 | 9_35107130 | 0.215738134 | FAM214B | 18_10475798 | 0.187283018 | APCDD1 |
| 6_128449016 | 0.148195827 | PTPRK | 1_216886977 | 0.215728341 | ESRRG | 11_118551519 | 0.187252543 | IFT46 |
| 9_98769074 | 0.148193256 | ANKS6 | 3_52078505 | 0.215703432 | POC1A | 3_139043098 | 0.187224743 | PRR23C |
| 4_155365823 | 0.148167549 | MAP9 | 6_100411313 | 0.21561547 | SIM1 | 2_64124274 | 0.187175453 | PELI1 |
| 1_244561822 | 0.148164735 | C1orf101 | 6_34407209 | 0.215493448 | RPS10-NUDT3 | 12_112190136 | 0.187136439 | HECTD4 |
| 1_244562024 | 0.148164735 | C1orf101 | 16_89743812 | 0.215488974 | FANCA | 12_112205808 | 0.187136439 | HECTD4 |
| 1_244583991 | 0.148164735 | C1orf101 | 4_89780995 | 0.215443953 | SNCA | 12_112239475 | 0.187136439 | HECTD4 |
| 1_244584934 | 0.148164735 | C1orf101 | 7_6184221 | 0.215429511 | CYTH3 | 12_112306039 | 0.187136439 | HECTD4 |
| 14_78922708 | 0.148160936 | NRXN3 | 20_45970109 | 0.215425359 | ZNF335 | 8_72839436 | 0.187122655 | KCNB2 |
| 1_36029761 | 0.148150783 | AGO3 | 7_147122264 | 0.215422555 | CNTNAP2 | 8_72843523 | 0.187122655 | KCNB2 |
| 2_71417456 | 0.148127904 | ZNF638 | 9_17442247 | 0.215364447 | CNTLN | 16_89591311 | 0.187115691 | CPNE7 |
| 14_78943162 | 0.148121127 | NRXN3 | 1_35818353 | 0.215349883 | AGO4 | 12_112022343 | 0.187098786 | ERP29 |
| 15_66044803 | 0.14811067 | MEGF11 | 13_71860437 | 0.215337554 | DACH1 | 12_112064295 | 0.187098786 | NAA25 |
| 1_111849738 | 0.148100544 | KCND3 | 7_152070003 | 0.215291836 | GALNT11 | 8_127208721 | 0.187096272 | CCAT1 |
| 1_111853302 | 0.148100544 | KCND3 | 7_104893380 | 0.215271286 | LHFPL3 | 2_50014727 | 0.187092298 | NRXN1 |
| 1_111853353 | 0.148100544 | KCND3 | 7_104898289 | 0.215271286 | LHFPL3 | 3_29392198 | 0.187079509 | RBMS3 |
| 1_111854345 | 0.148100544 | KCND3 | 22_40468844 | 0.215243246 | MKL1 | 1_169580573 | 0.187067904 | F5 |
| 12_15676549 | 0.148095211 | EPS8 | 8_68515401 | 0.215224656 | C8orf34 | 8_72835529 | 0.187065292 | KCNB2 |
| 3_705816 | 0.14808229 | LINC01266 | 3_55062584 | 0.215125358 | CACNA2D3 | 8_72837931 | 0.187065292 | KCNB2 |
| 4_89768920 | 0.148078036 | SNCA | 13_59873683 | 0.215106272 | DIAPH3 | 8_72845598 | 0.187065292 | KCNB2 |
| 4_89769681 | 0.148078036 | SNCA | 1_202776481 | 0.215102527 | KDM5B | 8_72847249 | 0.187065292 | KCNB2 |
| 3_55058520 | 0.148077798 | CACNA2D3 | 4_100414720 | 0.215085643 | EMCN | 8_72852450 | 0.187065292 | KCNB2 |
| 3_55060172 | 0.148077798 | CACNA2D3 | 15_48539667 | 0.215059643 | FBN1 | 8_72863948 | 0.187065292 | KCNB2 |
| 13_73859079 | 0.148049351 | KLF12 | 1_36018347 | 0.215003045 | AGO3 | 6_32661841 | 0.187064911 | HLA-DQB1 |
| 5_109852240 | 0.148015616 | MAN2A1 | 2_71384596 | 0.214997761 | ZNF638 | 6_32666130 | 0.187064911 | HLA-DQB1 |
| 7_7091216 | 0.148005008 | LOC100131257 | 1_57053660 | 0.214862064 | DAB1 | 4_81101722 | 0.187060476 | PRKG2 |
| 12_49795819 | 0.147893566 | NCKAP5L | 3_17592034 | 0.214825139 | TBC1D5 | 16_78370261 | 0.187057198 | WWOX |
| 4_31096906 | 0.147883777 | PCDH7 | 9_35093852 | 0.214776467 | PIGO | 10_13943968 | 0.187006583 | FRMD4A |
| 5_160771819 | 0.147877957 | ATP10B | 22_46698321 | 0.214731567 | CERK | 7_26828899 | 0.186981123 | SKAP2 |
| 5_160771862 | 0.147877957 | ATP10B | 22_40483101 | 0.21472434 | MKL1 | 5_113432271 | 0.186948739 | MCC |
| 5_160771866 | 0.147877957 | ATP10B | 4_20518229 | 0.214704228 | SLIT2 | 12_19273861 | 0.186946463 | PLEKHA5 |
| 5_160771948 | 0.147877957 | ATP10B | 9_17388930 | 0.214691694 | CNTLN | 13_20042728 | 0.186876815 | ZMYM2 |
| 22_42510855 | 0.147874122 | RRP7A | 13_27438492 | 0.214645443 | MTIF3 | 7_44633031 | 0.18687347 | OGDH |
| 11_132837520 | 0.147866277 | OPCML | 10_60562276 | 0.214611855 | ANK3 | 2_64122999 | 0.186837202 | PELI1 |
| 11_132837797 | 0.147866277 | OPCML | 1_203506157 | 0.21459059 | OPTC | 8_109251176 | 0.186788029 | NUDCD1 |
| 2_71114640 | 0.147854178 | MCEE | 16_6256113 | 0.214586806 | RBFOX1 | 11_118456606 | 0.186752111 | KMT2A |
| 2_216276218 | 0.147851923 | 4-Mar | 2_178443793 | 0.214575289 | PRKRA | 14_63381061 | 0.186725106 | PPP2R5E |
| 8_10420738 | 0.147851923 | MSRA | 13_70104426 | 0.214542083 | KLHL1 | 11_118536808 | 0.186722896 | TMEM25 |
| 4_89774357 | 0.147840726 | SNCA | 13_70104674 | 0.214542083 | KLHL1 | 10_14048905 | 0.186650179 | FRMD4A |
| 5_109866331 | 0.147830935 | MAN2A1 | 13_70109545 | 0.214542083 | ATXN8OS | 1_30724198 | 0.186632608 | MATN1-AS1 |
| 20_46963043 | 0.147730377 | EYA2 | 13_70122385 | 0.214542083 | ATXN8OS | 15_97742686 | 0.186583076 | LINC00923 |
| 20_46963160 | 0.147730377 | EYA2 | 9_129937601 | 0.21453497 | FNBP1 | 21_21343287 | 0.186574911 | NCAM2 |
| 14_100732685 | 0.1477229 | DLK1 | 3_84677225 | 0.214507203 | LINC00971 | 12_1819172 | 0.186556739 | CACNA2D4 |
| 10_133239619 | 0.147717792 | VENTX | 13_29254649 | 0.214480962 | MTUS2 | 11_118535615 | 0.186546315 | TMEM25 |
| 17_37866541 | 0.147713342 | YWHAEP7 | 7_98626147 | 0.214454328 | NPTX2 | 8_72854711 | 0.186529792 | KCNB2 |
| 6_144530548 | 0.147685854 | UTRN | 5_173011176 | 0.214390112 | ATP6V0E1 | 12_123642105 | 0.186489045 | GTF2H3 |
| 1_38925252 | 0.147656177 | RHBDL2 | 3_156264644 | 0.214354836 | KCNAB1 | 3_139991674 | 0.18647936 | CLSTN2 |
| 1_38937498 | 0.147656177 | RHBDL2 | 13_85425019 | 0.214350503 | LINC00351 | 12_19263930 | 0.186474793 | PLEKHA5 |
| 14_57619543 | 0.147638461 | SLC35F4 | 4_108744685 | 0.214308715 | ETNPPL | 12_117181604 | 0.18647458 | FBXO21 |
| 6_87811666 | 0.147623091 | LOC101928911 | 18_24148685 | 0.214263437 | CABYR | 6_99433002 | 0.186419402 | USP45 |
| 8_89961436 | 0.147590746 | NBN | 15_43782848 | 0.214223722 | SERF2 | 12_19267674 | 0.186404243 | PLEKHA5 |
| 9_20418325 | 0.147557181 | MLLT3 | 12_53571084 | 0.214162906 | ATF7 | 14_63384641 | 0.186361549 | PPP2R5E |
| 8_141190368 | 0.147554299 | DENND3 | 13_97294966 | 0.214161784 | MBNL2 | 14_63386534 | 0.186361549 | PPP2R5E |
| 11_113346642 | 0.147544193 | TTC12 | 8_18638453 | 0.21416009 | PSD3 | 15_48802066 | 0.18634973 | CEP152 |
| 10_50456663 | 0.14748829 | SGMS1 | 14_35291824 | 0.214101297 | PSMA6 | 12_27485131 | 0.186315016 | SMCO2 |
| 19_37540165 | 0.14739537 | ZNF793 | 1_62150922 | 0.214096106 | INADL | 1_161933480 | 0.186294014 | ATF6 |
| 9_100033055 | 0.147374448 | ERP44 | 1_231252488 | 0.214087503 | GNPAT | 6_37636520 | 0.18625285 | MDGA1 |
| 6_151525686 | 0.147347342 | CCDC170 | 12_80911483 | 0.21407978 | LIN7A | 1_244594120 | 0.186230476 | C1orf101 |
| 7_80750501 | 0.147345484 | SEMA3C | 7_120479798 | 0.214077509 | KCND2 | 11_118585699 | 0.186221862 | ARCN1 |
| 12_12528616 | 0.147328978 | DUSP16 | 16_20951739 | 0.214070898 | DNAH3 | 9_92053134 | 0.186221681 | SPTLC1 |
| 3_65377122 | 0.147307187 | MAGI1 | 13_49704482 | 0.214064326 | KPNA3 | 4_90192517 | 0.186194581 | CCSER1 |
| 19_6314931 | 0.147285905 | ACER1 | 14_91721849 | 0.214039331 | CATSPERB | 18_9724280 | 0.186140915 | RAB31 |
| 1_210133599 | 0.147276146 | SYT14 | 8_75322090 | 0.214030189 | CASC9 | 16_78356712 | 0.186116479 | WWOX |

| | | | | | | | | |
|---|---|---|---|---|---|---|---|---|
| 3_695204 | 0.147225925 | LINC01266 | 7_149280103 | 0.213989616 | ZNF783 | 9_123538757 | 0.186067869 | DENND1A |
| 1_71847870 | 0.147220935 | NEGR1 | 1_231261138 | 0.213961889 | GNPAT | 2_75111254 | 0.186061338 | TACR1 |
| 5_73558326 | 0.147200305 | ANKRA2 | 9_17432095 | 0.213942609 | CNTLN | 2_37038917 | 0.186048769 | HEATR5B |
| 9_131864469 | 0.147186719 | MED27 | 10_50441407 | 0.213879338 | SGMS1 | 12_19270690 | 0.186030468 | PLEKHA5 |
| 9_131893407 | 0.147186719 | MED27 | 13_43407551 | 0.213844568 | ENOX1 | 20_25214382 | 0.186023194 | ENTPD6 |
| 5_109812319 | 0.147186515 | MAN2A1 | 13_43407844 | 0.213844568 | ENOX1 | 2_70008476 | 0.18592763 | PCBP1-AS1 |
| 8_16141977 | 0.147177053 | MSR1 | 1_2138242 | 0.213830221 | PRKCZ | 15_83871977 | 0.185925773 | ADAMTSL3 |
| 1_35188698 | 0.147163833 | SFPQ | 3_17631819 | 0.213810463 | TBC1D5 | 10_60321409 | 0.185924313 | ANK3 |
| 1_35191655 | 0.147163833 | SFPQ | 1_247013736 | 0.213810358 | ZNF670-ZNF695 | 2_75095328 | 0.185896603 | TACR1 |
| 8_29340012 | 0.147124083 | DUSP4 | 4_39508644 | 0.213799134 | UGDH | 1_121370601 | 0.185872761 | SRGAP2-AS1 |
| 17_19826001 | 0.147118617 | ULK2 | 13_97290599 | 0.213789834 | MBNL2 | 2_69999350 | 0.185846383 | PCBP1-AS1 |
| 19_38616601 | 0.147087281 | MAP4K1 | 11_129448080 | 0.213699823 | BARX2 | 8_72843559 | 0.185815403 | KCNB2 |
| 9_135486069 | 0.147087281 | PPP1R26 | 12_112821089 | 0.213685192 | RPH3A | 15_47316671 | 0.185811003 | SEMA6D |
| 8_127076434 | 0.147075398 | PCAT2 | 4_56248322 | 0.213670733 | KIAA1211 | 12_27796857 | 0.185803258 | KLHL42 |
| 6_21807733 | 0.147066128 | CASC15 | 10_25448280 | 0.213601469 | GPR158 | 20_46194171 | 0.18578402 | CDH22 |
| 6_21807973 | 0.147066128 | CASC15 | 9_3477030 | 0.21359452 | RFX3 | 13_92700138 | 0.185729682 | GPC5 |
| 4_31131945 | 0.147042623 | PCDH7 | 6_166469359 | 0.213572289 | RPS6KA2 | 6_7200219 | 0.185663051 | RREB1 |
| 16_89128401 | 0.147041437 | ACSF3 | 22_40441249 | 0.213527001 | MKL1 | 12_112041068 | 0.185661814 | NAA25 |
| 1_15058966 | 0.147041308 | KAZN | 22_40470652 | 0.213527001 | MKL1 | 11_2898062 | 0.185623335 | SLC22A18AS |
| 9_130409662 | 0.147013483 | HMCN2 | 2_37246442 | 0.213472702 | NDUFAF7 | 1_1650452 | 0.185611304 | CDK11B |
| 22_32708529 | 0.146997416 | SYN3 | 17_76279079 | 0.213447004 | QRICH2 | 12_112223183 | 0.185589037 | HECTD4 |
| 18_14222233 | 0.146986799 | ANKRD20A5P | 10_68447561 | 0.213389019 | DNA2 | 12_112302668 | 0.185589037 | HECTD4 |
| 13_94297697 | 0.146966769 | GPC6 | 12_318965 | 0.213364827 | KDM5A | 15_47315067 | 0.185555282 | SEMA6D |
| 7_103441416 | 0.146965746 | SLC26A5 | 12_388792 | 0.213364827 | KDM5A | 1_33759325 | 0.185554967 | CSMD2 |
| 7_103442754 | 0.146965746 | SLC26A5 | 13_107732993 | 0.213364827 | FAM155A | 5_178385610 | 0.185494012 | COL23A1 |
| 7_103443539 | 0.146965746 | SLC26A5 | 3_124236373 | 0.213340358 | KALRN | 14_63380609 | 0.185483448 | PPP2R5E |
| 7_103443553 | 0.146965746 | SLC26A5 | 22_40470759 | 0.213325005 | MKL1 | 7_135483181 | 0.185475791 | CNOT4 |
| 8_70585909 | 0.146921935 | TRAM1 | 2_14309323 | 0.213287624 | LINC00276 | 18_25324152 | 0.185463225 | ZNF521 |
| 3_121454964 | 0.146840175 | POLQ | 2_223756353 | 0.213226165 | AP1S3 | 1_1280815 | 0.185453474 | SCNN1D |
| 5_167833158 | 0.146794908 | TENM2 | 3_125263010 | 0.213207391 | ZNF148 | 12_19232714 | 0.18543324 | PLEKHA5 |
| 21_16381311 | 0.146740167 | LINC00478 | 3_170500150 | 0.213204331 | SLC7A14 | 10_60301289 | 0.185391778 | ANK3 |
| 6_166821798 | 0.146718308 | RPS6KA2 | 14_67289769 | 0.213136051 | MPP5 | 6_12915522 | 0.185383127 | PHACTR1 |
| 6_166822600 | 0.146718308 | RPS6KA2 | 7_6214332 | 0.213125958 | CYTH3 | 6_31696467 | 0.185377585 | ABHD16A |
| 6_144522726 | 0.146683014 | UTRN | 2_178556567 | 0.21309445 | TTN | 11_88334596 | 0.185348032 | CTSC |
| 6_144531639 | 0.146683014 | UTRN | 4_88009624 | 0.213083323 | PKD2 | 2_229619820 | 0.18534488 | DNER |
| 4_72390023 | 0.146681048 | ADAMTS3 | 7_117295509 | 0.213064742 | WNT2 | 18_13534551 | 0.185325456 | LDLRAD4 |
| 11_92577225 | 0.146639043 | FAT3 | 8_96576685 | 0.21306054 | SDC2 | 3_20110692 | 0.185315484 | KAT2B |
| 13_28251618 | 0.146578022 | PAN3 | 6_34318647 | 0.21304629 | NUDT3 | 1_244426450 | 0.185301067 | ADSS |
| 8_3021110 | 0.146559538 | CSMD1 | 8_40643583 | 0.212997772 | ZMAT4 | 20_20040575 | 0.185294663 | CRNKL1 |
| 8_63187935 | 0.146508521 | YTHDF3 | 8_9576476 | 0.212938173 | TNKS | 20_20050098 | 0.185294663 | CRNKL1 |
| 2_206741252 | 0.146496741 | MDH1B | 6_34351615 | 0.212911347 | NUDT3 | 3_128368439 | 0.185265869 | EEFSEC |
| 2_206741955 | 0.146496741 | MDH1B | 9_35071777 | 0.212897385 | VCP | 6_7200220 | 0.185242306 | RREB1 |
| 16_21294347 | 0.146455219 | CRYM | 11_131466180 | 0.212885482 | NTM | 2_141087885 | 0.185235454 | LRP1B |
| 2_170411442 | 0.146451541 | MYO3B | 1_155706701 | 0.212875136 | DAP3 | 6_70289514 | 0.185204087 | COL9A1 |
| 8_72828241 | 0.146443177 | KCNB2 | 9_129051179 | 0.212838473 | FAM73B | 6_70292233 | 0.185204087 | COL9A1 |
| 11_132838852 | 0.146406628 | OPCML | 9_17374956 | 0.212793026 | CNTLN | 6_70293066 | 0.185204087 | COL9A1 |
| 3_65397090 | 0.146393786 | MAGI1 | 1_246054991 | 0.21278495 | SMYD3 | 6_70297015 | 0.185204087 | COL9A1 |
| 20_32575246 | 0.146366731 | NOL4L | 11_118116007 | 0.212776727 | TMPRSS4 | 6_70299607 | 0.185204087 | COL9A1 |
| 5_126608051 | 0.146331562 | PHAX | 10_50316762 | 0.212687754 | SGMS1 | 21_14165406 | 0.185196962 | LIPI |
| 5_126619802 | 0.146331562 | PHAX | 2_26497184 | 0.212675627 | OTOF | 17_65205869 | 0.185111108 | RGS9 |
| 21_41448082 | 0.146287579 | MX1 | 22_40581342 | 0.212670466 | MKL1 | 11_8880356 | 0.185106122 | ST5 |
| 12_127892181 | 0.14626621 | FLJ37505 | 15_47739769 | 0.212653928 | SEMA6D | 12_120662721 | 0.185102545 | CABP1 |
| 13_28301297 | 0.14621948 | FLT1 | 4_39507984 | 0.212634297 | UGDH | 1_1290722 | 0.185069428 | SCNN1D |
| 1_231176121 | 0.146211023 | TRIM67 | 7_104903569 | 0.212633453 | LHFPL3 | 9_9923181 | 0.185067198 | PTPRD |
| 7_148328660 | 0.146197386 | CNTNAP2 | 3_55060080 | 0.212563718 | CACNA2D3 | 1_161947959 | 0.185021768 | ATF6 |
| 10_50464010 | 0.146194983 | SGMS1 | 14_61575644 | 0.212506772 | FLJ22447 | 2_70044551 | 0.185009636 | PCBP1-AS1 |
| 3_129471874 | 0.146194983 | IFT122 | 10_95994153 | 0.212445384 | ENTPD1-AS1 | 2_229127662 | 0.184979888 | PID1 |
| 3_129481929 | 0.146194983 | IFT122 | 17_3783286 | 0.212387908 | ITGAE | 10_121597475 | 0.184972022 | FGFR2 |
| 4_31098944 | 0.146194983 | PCDH7 | 19_54540990 | 0.212386412 | KIR3DX1 | 3_64634197 | 0.184963812 | ADAMTS9 |
| 17_5911991 | 0.146174601 | LOC339166 | 22_40440749 | 0.212348905 | MKL1 | 22_17429168 | 0.184942233 | CECR2 |
| 15_48489009 | 0.146170711 | FBN1 | 22_40446401 | 0.212348905 | MKL1 | 20_63439613 | 0.18494029 | KCNQ2 |
| 8_72853278 | 0.146158537 | KCNB2 | 22_40475461 | 0.212348905 | MKL1 | 1_244560618 | 0.184865325 | C1orf101 |
| 21_16390803 | 0.146111 | LINC00478 | 22_40489706 | 0.212348905 | MKL1 | 1_244561310 | 0.184865325 | C1orf101 |
| 12_48341521 | 0.146103733 | ZNF641 | 3_125128183 | 0.212335793 | SLC12A8 | 1_244564826 | 0.184865325 | C1orf101 |
| 6_6180802 | 0.14607363 | F13A1 | 9_35071776 | 0.21232158 | VCP | 1_244584841 | 0.184865325 | C1orf101 |
| 12_127916203 | 0.146071215 | LINC00507 | 3_71063266 | 0.212305672 | FOXP1 | 7_90261233 | 0.184843085 | C7orf63 |
| 2_15205303 | 0.146060561 | NBAS | 22_40586577 | 0.212283853 | MKL1 | 1_222574833 | 0.184753443 | TAF1A |
| 4_186446675 | 0.146032236 | F11-AS1 | 1_111839043 | 0.212192714 | KCND3 | 3_132694382 | 0.184727799 | NPHP3 |
| 5_109697692 | 0.146028762 | MAN2A1 | 2_65122139 | 0.212178988 | RAB1A | 15_52454751 | 0.184723171 | MYO5A |
| 2_237775694 | 0.146026034 | LRRFIP1 | 9_101111504 | 0.212083108 | LPPR1 | 16_78357983 | 0.184671045 | WWOX |
| 2_237778573 | 0.146006284 | LRRFIP1 | 9_3486704 | 0.212025695 | RFX3 | 17_65728607 | 0.184574514 | CEP112 |
| 2_237779250 | 0.146006284 | LRRFIP1 | 9_3498606 | 0.212025695 | RFX3 | 3_133649014 | 0.184562426 | TOPBP1 |
| 9_131904570 | 0.145935648 | MED27 | 3_181258406 | 0.211974362 | SOX2-OT | 2_64125326 | 0.18455648 | PELI1 |
| 5_14836478 | 0.1459194 | ANKH | 6_138916222 | 0.211889767 | REPS1 | 3_138583694 | 0.184496623 | CEP70 |
| 14_78983868 | 0.145839091 | NRXN3 | 13_97291525 | 0.211771394 | MBNL2 | 12_120871402 | 0.184415952 | SPPL3 |
| 16_89149786 | 0.145828884 | ACSF3 | 6_34407159 | 0.211750966 | RPS10-NUDT3 | 12_27337504 | 0.184405322 | ARNTL2 |
| 16_89151346 | 0.145828884 | ACSF3 | 13_107750338 | 0.211721939 | FAM155A | 3_139996792 | 0.184399244 | CLSTN2 |
| 16_89155387 | 0.145828884 | ACSF3 | 9_3474868 | 0.211719375 | RFX3 | 3_139996961 | 0.184399244 | CLSTN2 |
| 6_146398747 | 0.145828884 | GRM1 | 9_3474872 | 0.211719375 | RFX3 | 3_41591010 | 0.184372222 | ULK4 |
| 6_146408393 | 0.145828884 | GRM1 | 2_168102822 | 0.211718536 | STK39 | 14_34860519 | 0.184312785 | BAZ1A |
| 4_108583580 | 0.14580109 | RPL34-AS1 | 7_6153238 | 0.211656363 | USP42 | 15_90551919 | 0.184309452 | CRTC3 |
| 6_87806056 | 0.145750284 | LOC101928911 | 12_54281329 | 0.211619851 | HNRNPA1 | 2_70047647 | 0.184297452 | PCBP1-AS1 |
| 13_28238417 | 0.145732736 | PAN3 | 15_48528475 | 0.211616259 | FBN1 | 9_20432656 | 0.184246646 | MLLT3 |
| 6_39804677 | 0.145712845 | DAAM2 | 9_17429428 | 0.211616259 | CNTLN | 6_32665349 | 0.184240313 | HLA-DQB1 |
| 13_28291790 | 0.145698705 | PAN3 | 1_4729914 | 0.211592468 | AJAP1 | 10_28881248 | 0.184238646 | C10orf126 |
| 4_100400360 | 0.145696622 | EMCN | 2_15240576 | 0.211583109 | NBAS | 20_63459705 | 0.184237787 | KCNQ2 |
| 6_21746794 | 0.14568112 | CASC15 | 16_68098310 | 0.211545933 | NFATC3 | 7_72323477 | 0.18420457 | CALN1 |
| 6_21771078 | 0.14568112 | CASC15 | 12_4663049 | 0.211510122 | NDUFA9 | 1_234246853 | 0.18419225 | SLC35F3 |
| 19_11553206 | 0.145628265 | ELOF1 | 12_338585 | 0.211384563 | KDM5A | 2_55675281 | 0.184181516 | PNPT1 |
| 22_32699089 | 0.145620805 | SYN3 | 11_45915266 | 0.211368972 | PEX16 | 1_121111967 | 0.184175381 | HIST2H2BA |
| 22_32724671 | 0.145620805 | SYN3 | 2_149338749 | 0.211361385 | LYPD6 | 2_43520725 | 0.184159557 | THADA |
| 18_42665412 | 0.145615928 | LINC00907 | 4_175703321 | 0.211352048 | GPM6A | 18_42606223 | 0.184154142 | LINC00907 |
| 18_42670439 | 0.145615928 | LINC00907 | 1_155711062 | 0.211310003 | DAP3 | 3_64704019 | 0.184138338 | ADAMTS9-AS2 |
| 19_37967759 | 0.145614327 | SIPA1L3 | 10_50459489 | 0.211296945 | SGMS1 | 2_69978350 | 0.184132673 | PCBP1-AS1 |
| 19_37968747 | 0.145614327 | SIPA1L3 | 22_40428005 | 0.211187191 | MKL1 | 1_204843570 | 0.184119526 | NFASC |
| 19_37968851 | 0.145614327 | SIPA1L3 | 22_40428083 | 0.211187191 | MKL1 | 9_132348378 | 0.184064464 | SETX |
| 19_37968877 | 0.145614327 | SIPA1L3 | 22_40431315 | 0.211187191 | MKL1 | 19_37557572 | 0.184047941 | ZNF540 |

| | | | | | | | | |
|---|---|---|---|---|---|---|---|---|
| 19_37975121 | 0.145614327 | SIPA1L3 | 22_40466609 | 0.211187191 | MKL1 | 2_157781963 | 0.183971319 | ACVR1 |
| 19_37978444 | 0.145614327 | SIPA1L3 | 22_40492359 | 0.211187191 | MKL1 | 3_139990466 | 0.18396285 | CLSTN2 |
| 19_37978589 | 0.145614327 | SIPA1L3 | 22_40493904 | 0.211187191 | MKL1 | 3_139990524 | 0.18396285 | CLSTN2 |
| 1_169374141 | 0.145537493 | BLZF1 | 22_40504073 | 0.211187191 | MKL1 | 3_688568 | 0.183960379 | LINC01266 |
| 15_82398640 | 0.145523693 | UBE2Q2P2 | 22_40487595 | 0.211176774 | MKL1 | 8_72835926 | 0.183929134 | KCNB2 |
| 6_68824592 | 0.145503557 | BAI3 | 8_100081910 | 0.211101141 | RGS22 | 8_72844016 | 0.183929134 | KCNB2 |
| 6_146402435 | 0.145475806 | GRM1 | 6_135293407 | 0.211074911 | AHI1 | 8_72844250 | 0.183929134 | KCNB2 |
| 6_146408681 | 0.145475806 | GRM1 | 22_40635023 | 0.211055826 | MKL1 | 19_37575858 | 0.18391315 | ZNF540 |
| 11_92574989 | 0.145457948 | FAT3 | 12_80882377 | 0.210985687 | LIN7A | 19_37575880 | 0.18391315 | ZNF540 |
| 13_24140047 | 0.145399405 | SPATA13 | 1_180699214 | 0.210905607 | XPR1 | 19_37579573 | 0.18391315 | ZNF540 |
| 3_41494312 | 0.145390768 | ULK4 | 1_169482946 | 0.21089087 | SLC19A2 | 19_37697751 | 0.18391315 | ZNF607 |
| 16_15766348 | 0.14536312 | MYH11 | 1_36053125 | 0.210889459 | AGO3 | 10_115244389 | 0.183908926 | ATRNL1 |
| 18_53085739 | 0.145356093 | DCC | 13_97291450 | 0.210881132 | MBNL2 | 3_139990775 | 0.183869895 | CLSTN2 |
| 11_88647956 | 0.145324941 | GRM5 | 7_53679002 | 0.210875942 | FLJ45974 | 19_54125565 | 0.183776635 | PRPF31 |
| 3_33119673 | 0.1453232 | CRTAP | 12_80878760 | 0.210836318 | LIN7A | 1_244567933 | 0.183766574 | C1orf101 |
| 19_37967292 | 0.145315649 | SIPA1L3 | 2_113951899 | 0.210834516 | ACTR3 | 13_98210530 | 0.183762232 | FARP1 |
| 3_131072099 | 0.145315317 | NEK11 | 13_85414806 | 0.210817342 | LINC00351 | 2_213369534 | 0.183721686 | SPAG16 |
| 6_87801418 | 0.145293901 | LOC101928911 | 17_75800685 | 0.210806665 | UNK | 16_89382337 | 0.18369694 | ANKRD11 |
| 3_131120635 | 0.145289165 | NEK11 | 17_73425992 | 0.21078274 | SDK2 | 14_72269583 | 0.183695994 | RGS6 |
| 3_131149738 | 0.145289165 | NEK11 | 8_94252884 | 0.210759179 | GEM | 12_112083440 | 0.183695172 | NAA25 |
| 3_131169747 | 0.145289165 | NEK11 | 8_14820743 | 0.210704934 | SGCZ | 2_70084071 | 0.183659742 | PCBP1-AS1 |
| 3_11826354 | 0.145225682 | TAMM41 | 1_49647855 | 0.210686346 | AGBL4 | 8_95512245 | 0.183627951 | C8orf37-AS1 |
| 1_30962663 | 0.145222046 | PUM1 | 11_16349763 | 0.210569779 | SOX6 | 12_112214719 | 0.183617532 | HECTD4 |
| 11_78241838 | 0.145213579 | GAB2 | 1_147194927 | 0.210556628 | FMO5 | 19_37556065 | 0.183584335 | ZNF540 |
| 1_200067879 | 0.145209265 | NR5A2 | 11_101897326 | 0.210521235 | ANGPTL5 | 12_27257600 | 0.183578268 | STK38L |
| 6_155436694 | 0.145204485 | NOX3 | 15_47742160 | 0.210510603 | SEMA6D | 3_139990964 | 0.183571361 | CLSTN2 |
| 10_76942893 | 0.145172584 | KCNMA1 | 13_43390985 | 0.210493539 | ENOX1 | 7_95897588 | 0.183532018 | DYNC1I1 |
| 10_59684606 | 0.145149348 | SLC16A9 | 12_287743 | 0.210457679 | KDM5A | 12_112180396 | 0.183529108 | HECTD4 |
| 2_173267858 | 0.145147391 | MLK7-AS1 | 13_107725359 | 0.210457679 | FAM155A | 12_112277030 | 0.183529108 | HECTD4 |
| 4_56257180 | 0.145126192 | KIAA1211 | 7_117294146 | 0.210344673 | WNT2 | 12_112291383 | 0.183529108 | HECTD4 |
| 3_168401741 | 0.145121442 | EGFEM1P | 7_147117602 | 0.210333826 | CNTNAP2 | 6_138106727 | 0.183527809 | PERP |
| 3_129446704 | 0.145113757 | IFT122 | 7_90261233 | 0.210325623 | C7orf63 | 3_2832578 | 0.18350911 | CNTN4 |
| 1_30940072 | 0.145112916 | PUM1 | 1_196989168 | 0.210305422 | CFHR5 | 12_111845236 | 0.183508027 | MAPKAPK5 |
| 1_31025772 | 0.145112916 | PUM1 | 13_107296670 | 0.210247853 | FAM155A | 1_65398326 | 0.183508027 | DNAJC6 |
| 2_166995212 | 0.145093921 | XIRP2 | 9_17385296 | 0.210246779 | CNTLN | 7_78536021 | 0.183504385 | MAGI2 |
| 2_14230820 | 0.145066583 | LINC00276 | 17_1178969 | 0.210232541 | ABR | 6_70298297 | 0.183450956 | COL9A1 |
| 1_44555499 | 0.145066158 | RNF220 | 7_147252535 | 0.210230436 | CNTNAP2 | 6_29943451 | 0.183436241 | HLA-A |
| 5_126601994 | 0.144993213 | PHAX | 5_8369363 | 0.2101795 | LOC729506 | 7_824866 | 0.1834087 | SUN1 |
| 12_126991647 | 0.144979068 | LOC101927592 | 5_8372484 | 0.2101795 | LOC729506 | 17_5552441 | 0.183291537 | NLRP1 |
| 5_109824550 | 0.144976608 | MAN2A1 | 3_4835391 | 0.210167789 | ITPR1 | 18_12208891 | 0.183270875 | C18orf61 |
| 13_28261165 | 0.144952585 | PAN3 | 4_89750398 | 0.210167612 | SNCA | 6_151310582 | 0.183263806 | AKAP12 |
| 13_28164963 | 0.144841208 | PAN3 | 15_48570169 | 0.210073649 | FBN1 | 4_41490393 | 0.183257839 | LIMCH1 |
| 9_81613140 | 0.144797667 | TLE1 | 3_64801370 | 0.210031082 | ADAMTS9-AS2 | 15_83859307 | 0.183255504 | ADAMTSL3 |
| 19_37970589 | 0.144766497 | SIPA1L3 | 3_17419372 | 0.209893456 | TBC1D5 | 7_24872454 | 0.183248495 | OSBPL3 |
| 3_131087428 | 0.14475046 | NEK11 | 11_129416017 | 0.209837056 | BARX2 | 1_20918820 | 0.183231186 | EIF4G3 |
| 12_95678275 | 0.14474792 | NTN4 | 3_120229824 | 0.209829851 | GPR156 | 6_32665341 | 0.183230879 | HLA-DQB1 |
| 6_144546780 | 0.144719532 | UTRN | 3_120233326 | 0.209829851 | GPR156 | 13_41571297 | 0.183221206 | VWA8 |
| 1_35192273 | 0.14470641 | SFPQ | 9_8791487 | 0.209810877 | PTPRD | 17_946204 | 0.183194897 | NXN |
| 7_71318465 | 0.144658806 | WBSCR17 | 1_44551685 | 0.209787313 | RNF220 | 10_28063656 | 0.183089404 | MPP7 |
| 8_95520397 | 0.144646357 | C8orf37-AS1 | 14_89406079 | 0.209746988 | FOXN3 | 12_27340862 | 0.1830379 | ARNTL2 |
| 9_131860193 | 0.144642437 | MED27 | 10_50445003 | 0.209655319 | SGMS1 | 12_27343154 | 0.1830379 | ARNTL2 |
| 1_113601782 | 0.14463227 | MAGI3 | 10_60420054 | 0.209602665 | ANK3 | 12_7435031 | 0.183013922 | CD163L1 |
| 8_72825893 | 0.144629507 | KCNB2 | 1_10975424 | 0.209600778 | C1orf127 | 3_64640238 | 0.183006752 | ADAMTS9 |
| 13_94312794 | 0.14461344 | GPC6 | 7_147399162 | 0.209545301 | CNTNAP2 | 1_177938376 | 0.182985934 | SEC16B |
| 15_79233567 | 0.144607953 | LOC729911 | 8_6876857 | 0.209516487 | DEFB1 | 4_121370367 | 0.182983309 | QRFPR |
| 7_103455526 | 0.144599461 | LOC101927870 | 22_40604960 | 0.209500775 | MKL1 | 9_128623856 | 0.182965694 | MIR1268A |
| 19_38616284 | 0.144580418 | MAP4K1 | 22_40511947 | 0.20949879 | MKL1 | 18_46701302 | 0.182946381 | ST8SIA5 |
| 16_21300033 | 0.144564033 | CRYM | 4_161564471 | 0.209470365 | FSTL5 | 6_32663723 | 0.182938523 | HLA-DQB1 |
| 1_243660409 | 0.144556954 | AKT3 | 12_2827464 | 0.209457481 | NRIP2 | 6_29976055 | 0.182914986 | HCG9 |
| 14_94001203 | 0.14455594 | LINC00521 | 4_186605682 | 0.209445474 | FAT1 | 17_19326635 | 0.182883892 | EPN2 |
| 1_62157324 | 0.144551167 | INADL | 20_9311529 | 0.209412874 | PLCB4 | 21_40149904 | 0.182845405 | DSCAM |
| 13_49054723 | 0.144509217 | FNDC3A | 1_74519305 | 0.209407945 | FPGT-TNNI3K | 1_54736400 | 0.18280316 | MROH7-TTC4 |
| 3_160071939 | 0.144491755 | IL12A-AS1 | 14_61570866 | 0.209400851 | FLJ22447 | 14_67289769 | 0.182799713 | MPP5 |
| 14_78901053 | 0.144454127 | NRXN3 | 9_121092151 | 0.209379067 | CNTRL | 20_59756473 | 0.182787063 | PHACTR3 |
| 21_41464802 | 0.144440431 | TMPRSS2 | 15_49150263 | 0.209366916 | COPS2 | 1_17206685 | 0.182767545 | PADI1 |
| 21_41467145 | 0.144440431 | TMPRSS2 | 7_95118819 | 0.209339002 | PPP1R9A | 17_40628675 | 0.182747826 | SMARCE1 |
| 9_14906950 | 0.144436359 | FREM1 | 1_35980236 | 0.209281855 | AGO3 | 20_63470260 | 0.182743528 | KCNQ2 |
| 17_46763851 | 0.144433336 | WNT3 | 7_78031047 | 0.209271668 | MAGI2 | 7_120487860 | 0.182743528 | KCND2 |
| 2_237771415 | 0.144417893 | LRRFIP1 | 17_38876523 | 0.209266517 | LASP1 | 19_45542676 | 0.182734376 | OPA3 |
| 2_71357092 | 0.144408254 | ZNF638 | 1_111872462 | 0.209251831 | KCND3 | 7_143414055 | 0.182641639 | EPHA1-AS1 |
| 17_37868736 | 0.144350421 | YWHAEP7 | 9_3479911 | 0.209248348 | RFX3 | 10_14089629 | 0.182626014 | FRMD4A |
| 11_113329836 | 0.144342071 | TTC12 | 9_92999173 | 0.209234606 | FGD3 | 21_36262805 | 0.182617804 | DOPEY2 |
| 11_113336145 | 0.144342071 | TTC12 | 13_107710517 | 0.209219691 | FAM155A | 17_5520004 | 0.182570471 | NLRP1 |
| 10_50444719 | 0.144315277 | SGMS1 | 8_40639196 | 0.209182526 | ZMAT4 | 6_30260639 | 0.182549482 | HCG17 |
| 17_65156076 | 0.144312788 | RGS9 | 15_48483052 | 0.209050495 | FBN1 | 6_30262145 | 0.182549482 | HCG17 |
| 8_10380482 | 0.144276616 | MSRA | 15_48557575 | 0.209050495 | FBN1 | 17_29584539 | 0.182539764 | GIT1 |
| 4_94987101 | 0.144226775 | BMPR1B | 15_48563922 | 0.209050495 | FBN1 | 2_70075665 | 0.182521481 | PCBP1-AS1 |
| 22_17214059 | 0.144221853 | CECR1 | 9_17400624 | 0.209050495 | CNTLN | 6_99531111 | 0.182509014 | TSTD3 |
| 2_15205317 | 0.144218642 | NBAS | 4_127970595 | 0.209036867 | C4orf29 | 15_88590866 | 0.182496803 | LOC101929743 |
| 6_159227122 | 0.144206479 | FNDC1 | 4_127970607 | 0.20899332 | C4orf29 | 2_69978315 | 0.182454031 | PCBP1-AS1 |
| 17_7775730 | 0.144190462 | DNAH2 | 7_24856694 | 0.208975365 | OSBPL3 | 2_69981322 | 0.182454031 | PCBP1-AS1 |
| 6_159210822 | 0.144177297 | FNDC1 | 8_17259991 | 0.208962427 | VPS37A | 21_14160166 | 0.182353422 | LIPI |
| 2_46169225 | 0.14415633 | PRKCE | 7_157487704 | 0.208931098 | LOC101927914 | 21_14182029 | 0.182353422 | LIPI |
| 2_46169547 | 0.14415633 | PRKCE | 15_53760311 | 0.208925873 | WDR72 | 15_90359228 | 0.182352055 | ZNF774 |
| 6_159209320 | 0.144153455 | FNDC1 | 15_81314908 | 0.208923141 | STARD5 | 15_83827930 | 0.182300485 | ADAMTSL3 |
| 15_40268289 | 0.144066766 | PAK6 | 4_48225702 | 0.208889791 | TEC | 1_225094588 | 0.182281939 | DNAH14 |
| 7_21855133 | 0.14406592 | DNAH11 | 12_112812671 | 0.208881799 | RPH3A | 18_13538970 | 0.182229566 | LDLRAD4 |
| 2_50350869 | 0.144062677 | NRXN1 | 12_112819608 | 0.208881274 | RPH3A | 12_89521109 | 0.182208874 | GALNT4 |
| 1_162866345 | 0.144047073 | C1orf110 | 17_3621561 | 0.208866221 | SHPK | 17_5547922 | 0.18216426 | NLRP1 |
| 9_130404389 | 0.1440408 | HMCN2 | 13_94915527 | 0.208810171 | LOC101927284 | 2_71400409 | 0.182112236 | ZNF638 |
| 18_53304764 | 0.14404073 | DCC | 7_78583438 | 0.208804045 | MAGI2 | 12_111845246 | 0.182099815 | MAPKAPK5 |
| 15_33862188 | 0.144017571 | RYR3 | 4_140026062 | 0.208764492 | MAML3 | 1_244563778 | 0.182025662 | C1orf101 |
| 16_10789147 | 0.144009974 | TVP23A | 6_166462331 | 0.208764492 | RPS6KA2 | 1_244569252 | 0.182025662 | C1orf101 |
| 8_72839198 | 0.14397482 | KCNB2 | 1_111870400 | 0.208760035 | KCND3 | 2_70071264 | 0.182017524 | PCBP1-AS1 |
| 4_7327808 | 0.143944602 | SORCS2 | 11_118174025 | 0.208748832 | SCN2B | 2_70085848 | 0.182017524 | PCBP1-AS1 |
| 4_89769553 | 0.143914063 | SNCA | 12_380228 | 0.208691642 | KDM5A | 18_42606348 | 0.182009501 | LINC00907 |
| 4_89769780 | 0.143914063 | SNCA | 2_113898239 | 0.208691642 | ACTR3 | 15_43379650 | 0.181997506 | TUBGCP4 |

| | | | | | | | | |
|---|---|---|---|---|---|---|---|---|
| 4_89774885 | 0.143914063 | SNCA | 2_113947853 | 0.208691642 | ACTR3 | 11_118592506 | 0.181986822 | ARCN1 |
| 2_167029968 | 0.143867066 | XIRP2 | 2_113948228 | 0.208691642 | ACTR3 | 2_49974409 | 0.181941978 | NRXN1 |
| 11_131472891 | 0.14385596 | NTM | 7_71582513 | 0.208669222 | WBSCR17 | 1_116537544 | 0.181924953 | CD58 |
| 18_53069621 | 0.14385518 | DCC | 2_74294284 | 0.208638921 | SLC4A5 | 11_118534738 | 0.181916445 | TMEM25 |
| 12_131122488 | 0.143825438 | GPR133 | 19_50522546 | 0.208613258 | LRRC4B | 2_64115305 | 0.181902943 | PELI1 |
| 1_201098285 | 0.143809984 | CACNA1S | 12_54256157 | 0.20861263 | CBX5 | 15_83686970 | 0.181897224 | ADAMTSL3 |
| 16_61694881 | 0.14379695 | CDH8 | 1_52636461 | 0.208596788 | FAM159A | 2_25288994 | 0.181873402 | DNMT3A |
| 4_105274384 | 0.143787329 | TET2 | 10_60337026 | 0.208566304 | ANK3 | 1_218801474 | 0.181858785 | MIR548F3 |
| 6_169689104 | 0.143763321 | WDR27 | 8_27802142 | 0.208531834 | ESCO2 | 1_244440434 | 0.181843092 | ADSS |
| 20_15914575 | 0.143757057 | MACROD2 | 15_94051373 | 0.208525571 | LOC101927112 | 21_14160484 | 0.181799885 | LIPI |
| 8_72818977 | 0.143739552 | KCNB2 | 19_2590864 | 0.208493133 | GNG7 | 5_5435662 | 0.181791577 | KIAA0947 |
| 10_49621847 | 0.143725574 | CHAT | 10_50444719 | 0.208483891 | SGMS1 | 17_19280421 | 0.181752459 | EPN2 |
| 5_109743666 | 0.14371538 | MAN2A1 | 6_34402070 | 0.208444402 | RPS10-NUDT3 | 17_19315525 | 0.181752459 | EPN2 |
| 7_154874721 | 0.143703504 | DPP6 | 3_129474600 | 0.208393178 | IFT122 | 17_19325953 | 0.181752459 | EPN2 |
| 7_154874722 | 0.143703504 | DPP6 | 11_101969193 | 0.208326482 | KIAA1377 | 17_19338043 | 0.181752459 | B9D1 |
| 6_151408446 | 0.143677785 | RMND1 | 1_52961061 | 0.208309003 | SCP2 | 12_27357158 | 0.181747538 | ARNTL2 |
| 1_47766994 | 0.143674062 | TRABD2B | 3_123640692 | 0.208307176 | MYLK | 1_54729845 | 0.181746077 | MROH7-TTC4 |
| 12_131182935 | 0.143665309 | LINC01257 | 1_35186362 | 0.20828244 | SFPQ | 13_102158568 | 0.181724161 | FGF14 |
| 5_109781470 | 0.143612671 | MAN2A1 | 3_8449461 | 0.208247436 | LMCD1-AS1 | 14_63387509 | 0.181671279 | PPP2R5E |
| 22_49551087 | 0.143607074 | C22orf34 | 1_222559626 | 0.208209447 | TAF1A | 7_104840210 | 0.181622089 | LHFPL3 |
| 7_77380231 | 0.143607074 | GSAP | 1_222566841 | 0.208209447 | TAF1A | 1_169559342 | 0.181546088 | F5 |
| 3_4620813 | 0.143582498 | ITPR1 | 1_222576806 | 0.208209447 | TAF1A | 1_169564790 | 0.181546088 | F5 |
| 19_38602237 | 0.14356239 | MAP4K1 | 5_11545122 | 0.208199669 | CTNND2 | 4_87978268 | 0.181540747 | SPP1 |
| 10_45572900 | 0.143518675 | 8-Mar | 1_180720050 | 0.208171411 | XPR1 | 4_87978282 | 0.181540747 | SPP1 |
| 12_112505674 | 0.143514921 | PTPN11 | 6_38303872 | 0.208126573 | BTBD9 | 2_127964905 | 0.181509821 | SAP130 |
| 6_21752741 | 0.143499492 | CASC15 | 17_73408090 | 0.208092698 | SDK2 | 21_31524013 | 0.181503793 | TIAM1 |
| 1_231176217 | 0.1434744 | TRIM67 | 6_166460592 | 0.208078946 | RPS6KA2 | 14_63384844 | 0.181478836 | PPP2R5E |
| 11_121603530 | 0.143466086 | SORL1 | 10_50317678 | 0.208076031 | SGMS1 | 12_61845508 | 0.181473889 | FAM19A2 |
| 3_710743 | 0.143466086 | LINC01266 | 17_19276770 | 0.208042661 | EPN2 | 10_50448719 | 0.1814602 | SGMS1 |
| 3_713541 | 0.143466086 | LINC01266 | 10_122020327 | 0.208012384 | TACC2 | 11_88324087 | 0.181435176 | CTSC |
| 3_721988 | 0.143466086 | LINC01266 | 22_40569332 | 0.207997608 | MKL1 | 17_5447451 | 0.181409603 | DHX33 |
| 4_6453822 | 0.14345446 | PPP2R2C | 5_119462462 | 0.207938046 | HSD17B4 | 2_16634738 | 0.181370531 | FAM49A |
| 1_1459658 | 0.143434986 | ATAD3C | 7_53688277 | 0.207927465 | FLJ45974 | 8_72840318 | 0.181310248 | KCNB2 |
| 1_1459662 | 0.143434986 | ATAD3C | 1_35386920 | 0.207919242 | ZMYM4 | 18_7057801 | 0.181298635 | LAMA1 |
| 12_127896345 | 0.143397801 | FLJ37505 | 14_61570579 | 0.207791083 | FLJ22447 | 18_7057802 | 0.181298635 | LAMA1 |
| 7_103453933 | 0.143393227 | LOC101927870 | 14_61571250 | 0.207791083 | FLJ22447 | 17_43373756 | 0.181244342 | LINC00910 |
| 10_25369563 | 0.143369899 | GPR158 | 15_100277409 | 0.207780683 | ADAMTS17 | 17_5554518 | 0.181242893 | NLRP1 |
| 2_137007050 | 0.143369566 | THSD7B | 1_231205765 | 0.207757701 | TRIM67 | 15_33595832 | 0.181191803 | RYR3 |
| 5_14834333 | 0.143369566 | ANKH | 8_18664020 | 0.207756226 | PSD3 | 2_50005128 | 0.181162056 | NRXN1 |
| 5_14840595 | 0.143369566 | ANKH | 1_231191443 | 0.207712482 | TRIM67 | 2_50012953 | 0.181162056 | NRXN1 |
| 13_28153889 | 0.143336241 | PAN3 | 2_87482185 | 0.207710134 | LINC00152 | 22_48687200 | 0.181154658 | FAM19A5 |
| 12_12515516 | 0.143330871 | DUSP16 | 20_10275604 | 0.207689141 | SNAP25 | 7_44615825 | 0.181129024 | OGDH |
| 9_3375692 | 0.143324324 | RFX3 | 7_133432117 | 0.207686052 | EXOC4 | 15_44431604 | 0.181116842 | CTDSPL2 |
| 12_19306657 | 0.143309156 | PLEKHA5 | 16_6874754 | 0.207628298 | RBFOX1 | 8_4421847 | 0.181088004 | CSMD1 |
| 6_169654634 | 0.143261247 | WDR27 | 7_92647412 | 0.207608634 | CDK6 | 3_29389915 | 0.181061275 | RBMS3 |
| 6_169655000 | 0.143261247 | WDR27 | 2_28568537 | 0.207560649 | PLB1 | 10_86507616 | 0.181037694 | WAPAL |
| 6_169655407 | 0.143261247 | WDR27 | 5_119455378 | 0.207555326 | HSD17B4 | 11_88315168 | 0.181037594 | CTSC |
| 6_169655886 | 0.143261247 | WDR27 | 6_158087876 | 0.207548653 | SYNJ2 | 6_32637427 | 0.181027209 | HLA-DQA1 |
| 10_12482773 | 0.143252272 | CAMK1D | 1_169374141 | 0.20753052 | BLZF1 | 1_245568109 | 0.181023606 | KIF26B |
| 20_62178150 | 0.143244965 | SS18L1 | 1_91728105 | 0.207514435 | TGFBR3 | 3_50291012 | 0.181009926 | IFRD2 |
| 3_191331063 | 0.143240556 | CCDC50 | 14_67989636 | 0.20746725 | RAD51B | 15_22797492 | 0.180979643 | NIPA1 |
| 1_244575290 | 0.143236243 | C1orf101 | 18_23455274 | 0.20745758 | RIOK3 | 12_21302151 | 0.180947329 | SLCO1A2 |
| 6_144587941 | 0.143229365 | UTRN | 10_14944674 | 0.207445307 | DCLRE1C | 13_92838480 | 0.180936088 | GPC5 |
| 14_57622381 | 0.143218167 | SLC35F4 | 9_17297199 | 0.207419316 | CNTLN | 2_47066519 | 0.180929023 | TTC7A |
| 11_121602682 | 0.143212269 | SORL1 | 6_77923550 | 0.207401903 | MEI4 | 2_70024721 | 0.180895765 | PCBP1-AS1 |
| 7_134150332 | 0.143196846 | LRGUK | 13_107712256 | 0.207360015 | FAM155A | 15_52552345 | 0.180821282 | ARPP19 |
| 10_112709755 | 0.143188114 | VTI1A | 1_61980478 | 0.207321302 | INADL | 6_30262213 | 0.180772295 | HCG17 |
| 11_113341831 | 0.143125403 | TTC12 | 11_118136215 | 0.207319367 | SCN4B | 11_118539753 | 0.180736251 | TMEM25 |
| 6_159230694 | 0.143117131 | FNDC1 | 1_222574833 | 0.207302423 | TAF1A | 3_68754212 | 0.180704717 | FAM19A4 |
| 10_118195978 | 0.143112338 | CASC2 | 7_40089729 | 0.207282613 | CDK13 | 11_131492753 | 0.180700588 | NTM |
| 17_76676325 | 0.143073419 | MXRA7 | 7_40089731 | 0.207282613 | CDK13 | 16_78366888 | 0.180680216 | WWOX |
| 19_15379674 | 0.143047957 | AKAP8 | 7_40089739 | 0.207282613 | CDK13 | 10_28256797 | 0.180619272 | MPP7 |
| 17_37868823 | 0.143012639 | YWHAEP7 | 10_49686465 | 0.207207612 | C10orf53 | 15_83896354 | 0.180603467 | ADAMTSL3 |
| 19_37972622 | 0.142997442 | SIPA1L3 | 20_3979161 | 0.207170215 | RNF24 | 15_83907224 | 0.180603467 | ADAMTSL3 |
| 6_166834969 | 0.14299164 | RPS6KA2 | 3_59907764 | 0.207117726 | FHIT | 6_30708949 | 0.180584039 | MDC1 |
| 17_19295364 | 0.142923944 | EPN2 | 7_78585956 | 0.207117139 | MAGI2 | 7_130951734 | 0.180555525 | LINC-PINT |
| 20_46953632 | 0.142917512 | EYA2 | 4_56213402 | 0.207111733 | KIAA1211 | 15_83832914 | 0.180543838 | ADAMTSL3 |
| 20_46959708 | 0.142917512 | EYA2 | 14_54706292 | 0.207071693 | SAMD4A | 12_86432506 | 0.180542436 | MGAT4C |
| 20_46959862 | 0.142917512 | EYA2 | 5_146269743 | 0.207063411 | RBM27 | 17_19243817 | 0.180503186 | EPN2 |
| 1_15061701 | 0.142912095 | KAZN | 12_103644101 | 0.207055966 | STAB2 | 17_19296066 | 0.180503186 | EPN2 |
| 22_17497117 | 0.142908084 | CECR2 | 6_101693008 | 0.207055031 | GRIK2 | 12_61854887 | 0.180498536 | FAM19A2 |
| 20_9438369 | 0.142896866 | PLCB4 | 5_167652165 | 0.207025942 | TENM2 | 20_35315551 | 0.180497392 | UQCC1 |
| 4_142467366 | 0.142887088 | INPP4B | 7_146331318 | 0.207011354 | CNTNAP2 | 10_54575067 | 0.180492834 | PCDH15 |
| 1_96372957 | 0.142874995 | LOC101928241 | 2_71390743 | 0.206952918 | ZNF638 | 21_25893395 | 0.180479045 | APP |
| 18_42659973 | 0.142858975 | LINC00907 | 11_20834245 | 0.206952478 | NELL1 | 20_7923190 | 0.180476386 | HAO1 |
| 7_103438042 | 0.142855363 | SLC26A5 | 11_96068703 | 0.206926611 | MAML2 | 1_1516109 | 0.180470328 | ATAD3A |
| 17_4252895 | 0.142847302 | ANKFY1 | 8_14826119 | 0.206908697 | SGCZ | 15_33592558 | 0.180447537 | RYR3 |
| 22_48687200 | 0.142808248 | FAM19A5 | 8_40605637 | 0.206878135 | ZMAT4 | 16_78357481 | 0.180445542 | WWOX |
| 3_131087683 | 0.142799326 | NEK11 | 8_17174890 | 0.206865318 | ZDHHC2 | 13_40651064 | 0.1804259 | FOXO1 |
| 3_131102250 | 0.142799326 | NEK11 | 5_150127273 | 0.206862603 | PDGFRB | 12_112229200 | 0.180425646 | HECTD4 |
| 3_131150110 | 0.142799326 | NEK11 | 17_4182871 | 0.206781643 | ANKFY1 | 12_112236553 | 0.180425646 | HECTD4 |
| 3_131155173 | 0.142799326 | NEK11 | 3_14469253 | 0.206758803 | SLC6A6 | 12_112254871 | 0.180425646 | HECTD4 |
| 6_166800446 | 0.1427888 | RPS6KA2 | 20_42201536 | 0.206734999 | PTPRT | 1_35410785 | 0.180411231 | ZMYM4 |
| 2_37169700 | 0.142759166 | SULT6B1 | 9_17319837 | 0.206726747 | CNTLN | 16_90025521 | 0.180408574 | GAS8 |
| 4_7372395 | 0.142759166 | SORCS2 | 13_97293042 | 0.206719969 | MBNL2 | 3_120762079 | 0.180380506 | GTF2E1 |
| 7_104873177 | 0.142745783 | LHFPL3 | 2_141074587 | 0.2066778 | LRP1B | 1_54730027 | 0.180364453 | MROH7-TTC4 |
| 10_25371015 | 0.142731855 | GPR158 | 4_151222617 | 0.206636652 | SH3D19 | 1_54730155 | 0.180364453 | MROH7-TTC4 |
| 14_88212040 | 0.142727591 | KCNK10 | 1_111841478 | 0.206608322 | KCND3 | 13_102163642 | 0.18035814 | FGF14 |
| 6_169769077 | 0.142726236 | ERMARD | 2_113915194 | 0.206587119 | ACTR3 | 17_28374995 | 0.180356987 | SARM1 |
| 5_167723887 | 0.142720189 | TENM2 | 20_3145432 | 0.20657687 | UBOX5 | 2_69770863 | 0.180351701 | ANXA4 |
| 1_231424474 | 0.142717716 | EGLN1 | 2_170407669 | 0.206511351 | MYO3B | 1_246016381 | 0.180313988 | SMYD3 |
| 19_56608988 | 0.142701225 | ZNF71 | 9_135565363 | 0.206509389 | PAEP | 5_95953930 | 0.180276337 | ELL2 |
| 5_126627064 | 0.142675593 | PHAX | 1_180841891 | 0.206508676 | XPR1 | 20_20285390 | 0.180233785 | C20orf26 |
| 12_12515532 | 0.142669463 | DUSP16 | 7_24862756 | 0.206508676 | OSBPL3 | 8_72839285 | 0.180201063 | KCNB2 |
| 3_190118162 | 0.142656813 | LEPREL1 | 12_92910447 | 0.206491809 | EEA1 | 3_14904450 | 0.180144781 | FGD5 |
| 2_71120761 | 0.142652486 | MCEE | 3_192360974 | 0.206426141 | FGF12 | 20_20022072 | 0.180143621 | NAA20 |
| 7_91153324 | 0.142652486 | CDK14 | 7_91156400 | 0.206378367 | CDK14 | 17_19331162 | 0.180122293 | EPN2 |

| | | |
|---|---|---|
| 1_65379393 | 0.142630075 | DNAJC6 |
| 6 61797284 | 0.142614008 | KHDRBS2 |
| 15 83604620 | 0.142600509 | SH3GL3 |
| 3_11817956 | 0.142564106 | TAMM41 |
| 1 15060947 | 0.142558474 | KAZN |
| 8 63208408 | 0.142529931 | YTHDF3 |
| 2 223773499 | 0.142504511 | AP1S3 |
| 6 21771268 | 0.142496753 | CASC15 |
| 9 20375747 | 0.14248751 | MLLT3 |
| 3 2871180 | 0.142478206 | CNTN4 |
| 15 47315026 | 0.142452773 | SEMA6D |
| 8 63170828 | 0.142444043 | YTHDF3 |
| 17 5841861 | 0.142441626 | LOC339166 |
| 1 169545310 | 0.142435642 | F5 |
| 6 87798296 | 0.142416831 | LOC101928911 |
| 8 139614148 | 0.142389655 | KCNK9 |
| 1 157833770 | 0.142384622 | CD5L |
| 4 175732490 | 0.142367275 | GPM6A |
| 1 66200547 | 0.142356178 | PDE4B |
| 9 131879298 | 0.142351725 | MED27 |
| 7 103436751 | 0.142346059 | SLC26A5 |
| 22 49569465 | 0.142335778 | C22orf34 |
| 9 98626987 | 0.142300029 | GABBR2 |
| 2 232119490 | 0.142240998 | DIS3L2 |
| 1 243831739 | 0.142226397 | AKT3 |
| 2 179968535 | 0.142206486 | CWC22 |
| 2 170426376 | 0.142199977 | MYO3B |
| 1 243652479 | 0.142189283 | AKT3 |
| 1 243781829 | 0.142189283 | AKT3 |
| 1 243783615 | 0.142189283 | AKT3 |
| 22 42511335 | 0.142180177 | RRP7A |
| 13_101152587 | 0.142179661 | NALCN |
| 6 144732283 | 0.142151369 | UTRN |
| 16 10903076 | 0.142150764 | CIITA |
| 12 21207801 | 0.142131384 | SLCO1B1 |
| 5 109818284 | 0.142126177 | MAN2A1 |
| 1 243655768 | 0.142099386 | AKT3 |
| 16 74732450 | 0.142095616 | FA2H |
| 10_50441174 | 0.14209373 | SGMS1 |
| 8 72843523 | 0.142080262 | KCNB2 |
| 13 98224964 | 0.142023752 | FARP1 |
| 4_31096340 | 0.142023752 | PCDH7 |
| 5 38868251 | 0.142023752 | OSMR |
| 10 107134326 | 0.142023708 | SORCS1 |
| 10_12488723 | 0.142023708 | CAMK1D |
| 20 19998735 | 0.141986245 | RIN2 |
| 5 32430006 | 0.141975221 | ZFR |
| 7_94400670 | 0.141969242 | COL1A2 |
| 13 98238060 | 0.141952605 | FARP1 |
| 11 131466180 | 0.141938732 | NTM |
| 16 89137058 | 0.14192265 | ACSF3 |
| 6 159237875 | 0.14191076 | FNDC1 |
| 10 12487038 | 0.141875549 | CAMK1D |
| 4 37971003 | 0.141872744 | TBC1D1 |
| 13_28287929 | 0.141832048 | PAN3 |
| 10 70523507 | 0.141816192 | PALD1 |
| 18 42757836 | 0.141763814 | RIT2 |
| 16_61726490 | 0.141749751 | CDH8 |
| 1 231567179 | 0.141734452 | TSNAX-DISC1 |
| 6 54025055 | 0.141711788 | MLIP |
| 8_14828428 | 0.141698838 | SGCZ |
| 18 53257137 | 0.141697882 | DCC |
| 18 53280100 | 0.141697882 | DCC |
| 3 11793718 | 0.141684804 | TAMM41 |
| 3 11804016 | 0.141684804 | TAMM41 |
| 3 11805244 | 0.141684804 | TAMM41 |
| 12 21202151 | 0.141682335 | SLCO1B1 |
| 8 63188037 | 0.141674012 | YTHDF3 |
| 2 215809249 | 0.141666484 | LINC00607 |
| 22 40430729 | 0.14164709 | MKL1 |
| 14 88236189 | 0.141638482 | KCNK10 |
| 14 88238999 | 0.141638482 | KCNK10 |
| 22 32711970 | 0.141628771 | SYN3 |
| 1 169528688 | 0.141581989 | F5 |
| 4 186446674 | 0.141575437 | F11-AS1 |
| 17 5880219 | 0.141570226 | LOC339166 |
| 10 15366357 | 0.141561797 | FAM171A1 |
| 4 31062840 | 0.14152995 | PCDH7 |
| 16 89875082 | 0.141528688 | TCF25 |
| 10 70524162 | 0.141513463 | PALD1 |
| 5 126608925 | 0.141499369 | PHAX |
| 19 6314911 | 0.141487287 | ACER1 |
| 4 97810301 | 0.141475302 | STPG2 |
| 5 177040008 | 0.141468222 | ZNF346 |
| 2 221561296 | 0.141461947 | EPHA4 |
| 4_30999445 | 0.141447843 | PCDH7 |
| 10 81986738 | 0.14144351 | NRG3 |
| 12 3651089 | 0.14142622 | EFCAB4B |
| 2 162643991 | 0.141406366 | KCNH7 |
| 13 28214609 | 0.14139321 | PAN3 |
| 17 33117073 | 0.14139321 | ASIC2 |
| 1 66187002 | 0.141345201 | PDE4B |
| 1 66187003 | 0.141345201 | PDE4B |
| 13 99863907 | 0.141321513 | CLYBL |
| 16 16216437 | 0.141307093 | ABCC6 |
| 3 129447415 | 0.141307093 | IFT122 |
| 5 36285404 | 0.141307093 | RANBP3L |
| 2_33331447 | 0.206350358 | LTBP1 |
| 20 19306755 | 0.206336055 | SLC24A3 |
| 1 31928217 | 0.206268835 | PTP4A2 |
| 2_174235061 | 0.206259358 | OLA1 |
| 9 135583091 | 0.206255535 | LOC100130954 |
| 1 56696351 | 0.206203793 | PRKAA2 |
| 1 169382754 | 0.206202263 | BLZF1 |
| 10 79176690 | 0.206195209 | ZMIZ1 |
| 17 12592942 | 0.206151318 | LINC00670 |
| 1 231206010 | 0.206130779 | TRIM67 |
| 20 19911297 | 0.206088275 | RIN2 |
| 10 25461450 | 0.206069817 | GPR158 |
| 5 83496530 | 0.206068695 | VCAN |
| 6 34293320 | 0.206015381 | NUDT3 |
| 6 34330210 | 0.206015381 | NUDT3 |
| 20 19930634 | 0.205983188 | RIN2 |
| 7 91153324 | 0.205969702 | CDK14 |
| 7 40089727 | 0.205919218 | CDK13 |
| 11 101034325 | 0.205900056 | PGR |
| 11 132271902 | 0.205893181 | NTM |
| 13 94297697 | 0.205859527 | GPC6 |
| 2 71333535 | 0.205838139 | ZNF638 |
| 7 71618198 | 0.205838139 | WBSCR17 |
| 15 71697824 | 0.205811241 | THSD4 |
| 2 162542357 | 0.205746148 | KCNH7 |
| 2 170415769 | 0.205735298 | MYO3B |
| 10 121597900 | 0.205731538 | FGFR2 |
| 22 25803560 | 0.205718808 | MYO18B |
| 20_59614380 | 0.205704483 | PHACTR3 |
| 7 54542961 | 0.205686932 | VSTM2A |
| 4 89781878 | 0.205571154 | SNCA |
| 15_100300237 | 0.20556149 | ADAMTS17 |
| 4 175701038 | 0.205554199 | GPM6A |
| 15 71695511 | 0.205550541 | THSD4 |
| 1 35937573 | 0.205550456 | AGO3 |
| 11 67424636 | 0.205543272 | CARNS1 |
| 2 30582216 | 0.205539945 | LCLAT1 |
| 1 36023355 | 0.205527702 | AGO3 |
| 7_102452903 | 0.205527653 | ORAI2 |
| 21 40266641 | 0.205526658 | DSCAM |
| 21 40266643 | 0.205526658 | DSCAM |
| 4_146924707 | 0.205494919 | TTC29 |
| 12 80893754 | 0.205423017 | LIN7A |
| 12 80896081 | 0.205423017 | LIN7A |
| 12_80901147 | 0.205423017 | LIN7A |
| 7 6225038 | 0.20539915 | CYTH3 |
| 17 75691702 | 0.205398479 | SAP30BP |
| 12_80873894 | 0.205393395 | LIN7A |
| 1 193047044 | 0.205357116 | UCHL5 |
| 21 40266645 | 0.205325266 | DSCAM |
| 21 40266646 | 0.205325266 | DSCAM |
| 21 40266647 | 0.205325266 | DSCAM |
| 15 60908101 | 0.205322367 | RORA |
| 1 231771101 | 0.205280943 | DISC1 |
| 7_93202107 | 0.205240991 | HEPACAM2 |
| 6 16742370 | 0.205223063 | ATXN1 |
| 7 27745666 | 0.205223063 | TAX1BP1 |
| 19_2394032 | 0.205205494 | TMPRSS9 |
| 1 35897571 | 0.20519167 | AGO1 |
| 7 24865885 | 0.20517086 | OSBPL3 |
| 19_3646644 | 0.205141943 | PIP5K1C |
| 3 124236273 | 0.205137624 | KALRN |
| 7 149296883 | 0.205127071 | LOC155060 |
| 16 80196909 | 0.205029222 | LOC102724084 |
| 13 59854867 | 0.205026126 | DIAPH3 |
| 2 28570304 | 0.205007848 | PLB1 |
| 7 24845450 | 0.204886121 | OSBPL3 |
| 5 162143235 | 0.204829161 | GABRG2 |
| 5 162152766 | 0.204829161 | GABRG2 |
| 3 124236997 | 0.204821866 | KALRN |
| 3 124237551 | 0.204821866 | KALRN |
| 2 6996046 | 0.204796026 | RNF144A |
| 4 39522684 | 0.204775836 | UGDH |
| 4 175739671 | 0.204773547 | GPM6A |
| 16 61705866 | 0.204713642 | CDH8 |
| 15 45064972 | 0.204704572 | SORD |
| 11 66170775 | 0.204643683 | PACS1 |
| 16 78263155 | 0.20460788 | WWOX |
| 7 117595164 | 0.204573773 | CFTR |
| 3 24228772 | 0.204556667 | THRB |
| 9 128172172 | 0.2045468 | CIZ1 |
| 6 149381591 | 0.204530892 | TAB2 |
| 22 23746209 | 0.204471477 | ZNF70 |
| 16 85918030 | 0.204452865 | IRF8 |
| 16 85918041 | 0.204452865 | IRF8 |
| 4_164101513 | 0.204430924 | 1-Mar |
| 20 10272931 | 0.204428243 | SNAP25 |
| 20 10284532 | 0.204428243 | SNAP25 |
| 16 80196919 | 0.204409896 | LOC102724084 |
| 16 68192332 | 0.204365163 | NFATC3 |
| 10 60434391 | 0.204354555 | ANK3 |
| 8 28780167 | 0.204354043 | INTS9 |
| 8 40596932 | 0.204344776 | ZMAT4 |
| 8 40597048 | 0.204344776 | ZMAT4 |
| 7 100811870 | 0.204323467 | EPHB4 |
| 19_15519642 | 0.204309925 | CYP4F22 |
| 3 55067868 | 0.204286522 | CACNA2D3 |
| 1 220003142 | 0.180092367 | EPRS |
| 8 72847777 | 0.180086798 | KCNB2 |
| 12 112226112 | 0.180070714 | HECTD4 |
| 12_111999345 | 0.180068666 | TMEM116 |
| 12 112042516 | 0.180068666 | NAA25 |
| 12 112085014 | 0.180068666 | NAA25 |
| 10 28858901 | 0.180016386 | C10orf126 |
| 12 65775251 | 0.179988228 | RPSAP52 |
| 10 93443947 | 0.179971509 | MYOF |
| 7 120497617 | 0.179965117 | KCND2 |
| 9 132301131 | 0.179960633 | SETX |
| 21 21193091 | 0.17995294 | NCAM2 |
| 3_60326148 | 0.179950651 | FHIT |
| 1 53526568 | 0.179921057 | GLIS1 |
| 19 37738477 | 0.179917989 | ZNF573 |
| 15 52448719 | 0.179914742 | MYO5A |
| 1 41787553 | 0.179861295 | HIVEP3 |
| 19 1390840 | 0.179857381 | NDUFS7 |
| 10 96256750 | 0.179845352 | BLNK |
| 19 56608988 | 0.179842531 | ZNF71 |
| 6 4141980 | 0.179796346 | LOC100507506 |
| 19 37561291 | 0.179771436 | ZNF540 |
| 13 30341793 | 0.179739143 | LINC00426 |
| 6 30044847 | 0.179722012 | ZNRD1-AS1 |
| 12 27261944 | 0.179711816 | STK38L |
| 16 78362328 | 0.179695712 | WWOX |
| 17 10138358 | 0.179680317 | GAS7 |
| 2 70158666 | 0.1796763 | C2orf42 |
| 12_27333696 | 0.179674208 | ARNTL2 |
| 13 22260934 | 0.179655876 | LINC00540 |
| 13 42053554 | 0.179616522 | DGKH |
| 2_70056265 | 0.179579447 | PCBP1-AS1 |
| 2 70059999 | 0.179574942 | PCBP1-AS1 |
| 12 19303994 | 0.179542927 | PLEKHA5 |
| 15 52563724 | 0.179538392 | ARPP19 |
| 7 26822646 | 0.179509938 | SKAP2 |
| 19 45014239 | 0.179505695 | RELB |
| 15 90532282 | 0.179440806 | CRTC3 |
| 7_26535047 | 0.17941815 | KIAA0087 |
| 7 44699381 | 0.179391284 | OGDH |
| 1 32214320 | 0.179380841 | DCDC2B |
| 6_150871519 | 0.179293771 | MTHFD1L |
| 1 66253736 | 0.179278292 | PDE4B |
| 19 49158325 | 0.179254886 | TRPM4 |
| 2_72997596 | 0.179184367 | SFXN5 |
| 3 139997707 | 0.179181503 | CLSTN2 |
| 17 19287352 | 0.179163339 | EPN2 |
| 17_19315119 | 0.179163339 | EPN2 |
| 5 77464255 | 0.179136687 | WDR41 |
| 1 66238490 | 0.179085445 | PDE4B |
| 7 120489605 | 0.17908284 | KCND2 |
| 7 120498782 | 0.17908284 | KCND2 |
| 7 120509095 | 0.17908284 | KCND2 |
| 12 7440280 | 0.179052411 | CD163L1 |
| 3_126504159 | 0.17898869 | UROC1 |
| 10 93478923 | 0.178983958 | MYOF |
| 20 9755040 | 0.178944007 | PAK7 |
| 20_9762854 | 0.178944007 | PAK7 |
| 3 696643 | 0.178870202 | LINC01266 |
| 12 112245711 | 0.178839685 | HECTD4 |
| 12_112277520 | 0.178839685 | HECTD4 |
| 12 112294221 | 0.178839685 | HECTD4 |
| 12 112302614 | 0.178839685 | HECTD4 |
| 12 112310765 | 0.178839685 | HECTD4 |
| 17 19416809 | 0.178817686 | RNF112 |
| 5 36260093 | 0.178803443 | RANBP3L |
| 12 112268198 | 0.17880277 | HECTD4 |
| 11 134453356 | 0.178766846 | LOC283177 |
| 1 234251213 | 0.178728989 | SLC35F3 |
| 16 56374290 | 0.178724589 | AMFR |
| 4 24931413 | 0.17871426 | CCDC149 |
| 6 32666297 | 0.178659159 | HLA-DQB1 |
| 12 111806025 | 0.178647723 | ALDH2 |

| | | |
|---|---|---|
| 15_80115894 | 0.141286297 | ZFAND6 |
| 8_3039002 | 0.141257353 | CSMD1 |
| 15_60908101 | 0.141236976 | RORA |
| 1_111840362 | 0.141230302 | KCND3 |
| 8_127079620 | 0.141200789 | PCAT2 |
| 11_46725066 | 0.141185458 | F2 |
| 2_44181781 | 0.141173451 | PPM1B |
| 20_10153802 | 0.141145866 | SNAP25-AS1 |
| 16_89138474 | 0.141145291 | ACSF3 |
| 5_106840853 | 0.141140496 | LOC102467213 |
| 12_80327981 | 0.141138519 | OTOGL |
| 3_716698 | 0.141124778 | LINC01266 |
| 3_736752 | 0.141124778 | LINC01266 |
| 1_31220005 | 0.141114142 | NKAIN1 |
| 11_121618707 | 0.141102037 | SORL1 |
| 1_231176324 | 0.141101819 | TRIM67 |
| 1_231176343 | 0.141101819 | TRIM67 |
| 17_8012594 | 0.141082478 | GUCY2D |
| 10_106842207 | 0.14108232 | SORCS1 |
| 13_70088194 | 0.141051661 | KLHL1 |
| 3_716605 | 0.141051661 | LINC01266 |
| 8_63188795 | 0.14104682 | YTHDF3 |
| 1_243675328 | 0.141030777 | AKT3 |
| 2_170406416 | 0.141030237 | MYO3B |
| 20_19246994 | 0.14101895 | LOC100130264 |
| 15_83556283 | 0.140995208 | SH3GL3 |
| 1_244587437 | 0.140992993 | C1orf101 |
| 1_243728268 | 0.140977229 | AKT3 |
| 4_56248322 | 0.140960384 | KIAA1211 |
| 2_215825571 | 0.140952032 | LINC00607 |
| 4_6472514 | 0.140947373 | PPP2R2C |
| 3_718559 | 0.140906104 | LINC01266 |
| 11_113363744 | 0.140895819 | TTC12 |
| 11_113363749 | 0.140895819 | TTC12 |
| 5_14848297 | 0.140895156 | ANKH |
| 5_14831317 | 0.14086079 | ANKH |
| 5_14835204 | 0.14086079 | ANKH |
| 5_14846358 | 0.14086079 | ANKH |
| 5_14847519 | 0.14086079 | ANKH |
| 2_167036113 | 0.140860691 | XIRP2 |
| 19_37564710 | 0.140830245 | ZNF540 |
| 10_75888023 | 0.140776774 | C10orf11 |
| 7_94424911 | 0.140775541 | COL1A2 |
| 18_42674908 | 0.140764469 | LINC00907 |
| 11_79231717 | 0.140754774 | TENM4 |
| 8_120606504 | 0.140713031 | SNTB1 |
| 8_120612624 | 0.140713031 | SNTB1 |
| 13_20978396 | 0.140708962 | LATS2 |
| 19_37954304 | 0.140701453 | SIPA1L3 |
| 14_88218732 | 0.140690019 | KCNK10 |
| 7_24865885 | 0.140680826 | OSBPL3 |
| 12_53100396 | 0.140665915 | IGFBP6 |
| 11_63512280 | 0.140661839 | LGALS12 |
| 11_113340607 | 0.140657001 | TTC12 |
| 11_113342347 | 0.140657001 | TTC12 |
| 13_28254975 | 0.140649464 | PAN3 |
| 13_28257935 | 0.140649464 | PAN3 |
| 13_28258850 | 0.140649464 | PAN3 |
| 4_186402577 | 0.140649151 | F11-AS1 |
| 20_50943742 | 0.140648147 | DPM1 |
| 14_57623096 | 0.140647674 | SLC35F4 |
| 14_57624479 | 0.140647674 | SLC35F4 |
| 2_167008911 | 0.140603203 | XIRP2 |
| 18_5967020 | 0.14058167 | L3MBTL4 |
| 21_16395503 | 0.140544909 | LINC00478 |
| 2_237779632 | 0.140529147 | LRRFIP1 |
| 4_6872790 | 0.140524338 | KIAA0232 |
| 3_65370534 | 0.140500009 | MAGI1 |
| 1_152337207 | 0.140499331 | FLG-AS1 |
| 18_53106819 | 0.140493864 | DCC |
| 2_79874195 | 0.140481927 | CTNNA2 |
| 7_90062866 | 0.140469015 | STEAP2-AS1 |
| 6_21730960 | 0.140455044 | CASC15 |
| 6_21730967 | 0.140455044 | CASC15 |
| 9_10413091 | 0.140425802 | PTPRD |
| 17_10141281 | 0.140421942 | GAS7 |
| 1_96372855 | 0.140409723 | LOC101928241 |
| 3_131119623 | 0.140391126 | NEK11 |
| 3_131122257 | 0.140391126 | NEK11 |
| 5_110712124 | 0.140332709 | TMEM232 |
| 2_206741009 | 0.140322035 | MDH1B |
| 18_109093 | 0.14031737 | ROCK1P1 |
| 1_211820412 | 0.140316588 | LPGAT1 |
| 1_211820413 | 0.140316588 | LPGAT1 |
| 5_167746389 | 0.140314786 | TENM2 |

| | | |
|---|---|---|
| 18_52607757 | 0.204251263 | DCC |
| 1_216857363 | 0.204242557 | ESRRG |
| 1_204472310 | 0.204151825 | PIK3C2B |
| 16_80196929 | 0.204126288 | LOC102724084 |
| 1_201503847 | 0.204106208 | CSRP1 |
| 20_5949220 | 0.204058254 | TRMT6 |
| 7_91167295 | 0.20405465 | CDK14 |
| 5_162081497 | 0.204022878 | GABRG2 |
| 7_6166051 | 0.203989172 | CYTH3 |
| 4_39513325 | 0.203976967 | UGDH |
| 20_19309851 | 0.203970952 | SLC24A3 |
| 1_231199611 | 0.203924757 | TRIM67 |
| 1_169522862 | 0.203906415 | F5 |
| 1_147246274 | 0.203874773 | CHD1L |
| 1_147253268 | 0.203874773 | CHD1L |
| 6_155286639 | 0.203858135 | TFB1M |
| 4_2164822 | 0.203857676 | POLN |
| 4_2164823 | 0.203857676 | POLN |
| 9_110918066 | 0.203842017 | LPAR1 |
| 3_8433067 | 0.203747483 | LMCD1-AS1 |
| 2_223772121 | 0.203688536 | AP1S3 |
| 10_95993391 | 0.203654933 | ENTPD1-AS1 |
| 20_19333973 | 0.203651382 | SLC24A3 |
| 3_71190478 | 0.20363818 | FOXP1 |
| 17_1491525 | 0.203590761 | MYO1C |
| 7_39975468 | 0.203585478 | CDK13 |
| 17_4024200 | 0.203574668 | ZZEF1 |
| 1_35898381 | 0.203543758 | AGO1 |
| 8_94253736 | 0.203487916 | GEM |
| 8_27814448 | 0.203482137 | PBK |
| 3_185620660 | 0.203478953 | SENP2 |
| 7_156568004 | 0.203472562 | LINC01006 |
| 18_69460421 | 0.203444444 | DOK6 |
| 6_100427640 | 0.203442666 | SIM1 |
| 7_93546377 | 0.203436198 | CALCR |
| 7_88165967 | 0.203416913 | ADAM22 |
| 9_128215710 | 0.203335531 | DNM1 |
| 13_97333972 | 0.203312473 | MBNL2 |
| 4_89752690 | 0.203254334 | SNCA |
| 4_89743643 | 0.203244448 | SNCA |
| 19_2602437 | 0.203236467 | GNG7 |
| 3_55066661 | 0.203236467 | CACNA2D3 |
| 2_242099015 | 0.203185689 | LOC728323 |
| 4_95448233 | 0.203152449 | UNC5C |
| 11_101922418 | 0.203090241 | KIAA1377 |
| 11_101947742 | 0.203090241 | KIAA1377 |
| 4_89788339 | 0.203090241 | SNCA |
| 4_89788887 | 0.203090241 | SNCA |
| 4_89811652 | 0.203090241 | SNCA |
| 4_89813763 | 0.203090241 | SNCA |
| 21_21131515 | 0.203088055 | NCAM2 |
| 9_33050527 | 0.203068942 | SMU1 |
| 11_88522603 | 0.203051645 | GRM5 |
| 15_81351349 | 0.202973796 | LOC101929655 |
| 15_100299636 | 0.202942866 | ADAMTS17 |
| 2_241871422 | 0.202938781 | RTP5 |
| 8_29173765 | 0.202899242 | KIF13B |
| 20_63913361 | 0.202868097 | DNAJC5 |
| 15_27491961 | 0.202802218 | GABRG3 |
| 6_87009056 | 0.202791491 | HTR1E |
| 19_7419855 | 0.202742415 | ARHGEF18 |
| 12_261267 | 0.202486276 | SLC6A13 |
| 12_304980 | 0.202486276 | KDM5A |
| 9_135587099 | 0.202475913 | LOC100130954 |
| 7_151837660 | 0.202462614 | PRKAG2 |
| 4_100510928 | 0.202449841 | EMCN |
| 8_17218977 | 0.202419831 | ZDHHC2 |
| 22_40560812 | 0.202416078 | MKL1 |
| 6_100407338 | 0.202414967 | SIM1 |
| 6_100411472 | 0.202414967 | SIM1 |
| 11_129401916 | 0.202409456 | BARX2 |
| 9_110701564 | 0.202348755 | MUSK |
| 8_17211775 | 0.202275556 | ZDHHC2 |
| 11_101898281 | 0.20227321 | ANGPTL5 |
| 7_147379391 | 0.202198274 | CNTNAP2 |
| 1_960684 | 0.202179104 | KLHL17 |
| 1_960953 | 0.202179104 | KLHL17 |
| 2_132927740 | 0.202163737 | NCKAP5 |
| 4_108743511 | 0.202148719 | ETNPPL |
| 14_92139411 | 0.202048879 | CPSF2 |
| 9_3500471 | 0.202048879 | RFX3 |
| 12_287072 | 0.202048311 | KDM5A |
| 9_17407002 | 0.201971093 | CNTLN |
| 9_17454401 | 0.201971093 | CNTLN |
| 8_76432981 | 0.201856183 | LINC01111 |
| 21_33446234 | 0.201818551 | TMEM50B |
| 12_291902 | 0.201805263 | KDM5A |
| 13_107711447 | 0.201736226 | FAM155A |
| 1_35890856 | 0.201719921 | AGO1 |
| 2_168087752 | 0.201707565 | STK39 |
| 8_29073862 | 0.201685921 | KIF13B |
| 12_124349494 | 0.201685409 | NCOR2 |
| 15_80865651 | 0.201672963 | CEMIP |
| 8_14819525 | 0.201649947 | SGCZ |
| 11_27672554 | 0.201634849 | BDNF |
| 19_2512878 | 0.201568566 | GNG7 |
| 13_70110779 | 0.201544389 | ATXN8OS |

| | | |
|---|---|---|
| 11_126771636 | 0.201500772 | KIRREL3 |
| 19 7418772 | 0.201490428 | ARHGEF18 |
| 3 124237070 | 0.201453131 | KALRN |
| 3_124237071 | 0.201453131 | KALRN |
| 3 124237495 | 0.201453131 | KALRN |
| 3 124237513 | 0.201453131 | KALRN |
| 10 60394411 | 0.201433531 | ANK3 |
| 17 75551661 | 0.201391446 | LLGL2 |
| 9 17263555 | 0.201356177 | CNTLN |
| 1 111840362 | 0.201340256 | KCND3 |
| 20 10273639 | 0.201278571 | SNAP25 |
| 15 100299771 | 0.201278337 | ADAMTS17 |
| 15 100300443 | 0.201278337 | ADAMTS17 |
| 7 104840019 | 0.201278044 | LHFPL3 |
| 8 17288918 | 0.201260369 | VPS37A |
| 7 103740526 | 0.201225389 | RELN |
| 11 70099952 | 0.201212999 | ANO1 |
| 1 169512877 | 0.201178961 | F5 |
| 17 75554404 | 0.201174736 | LLGL2 |
| 11 101892761 | 0.201151588 | ANGPTL5 |
| 16 15874737 | 0.201131178 | FOPNL |
| 2 168089018 | 0.201111648 | STK39 |
| 3 14146538 | 0.201029915 | XPC |
| 9 17375803 | 0.201029915 | CNTLN |

**Table S11. The annotation by GWAS catalog**

| Group pairs | SNP | Traits | Genes |
|---|---|---|---|
| THL_TCL | rs2494741 | Total cholesterol levels | AKT1 |
| | rs11052294 | blood protein levels | PKP2 |
| THL_TSL | rs1045303 | White blood cell count | DENND3 |
| | rs12129789 | Arrhythmia | KCND3 |
| | rs12044963 | Atrial fibrillation | KCND3 |
| TCL_TSL | rs9691522 | Longevity | CFAP69 |

**Table S12. The enrichment result of metascape**

| GroupID | Category | Term | Description | LogP | Log(q-value) | Genes | Symbols | InTerm_InList |
|---|---|---|---|---|---|---|---|---|
| 1_Summary | GO Biological Processes | GO:0042391 | regulation of membrane potential | -8.557352424 | -4.21351074 | 40,1080,12 | ASIC2,CFT | 36/- |
| 1_Member | GO Biological Processes | GO:0042391 | regulation of membrane potential | -8.557352424 | -4.21351074 | 40,1080,12 | ASIC2,CFT | 22/446 |
| 2_Summary | GO Biological Processes | GO:0098660 | inorganic ion transmembrane transport | -7.146180716 | -3.27946028 | 40,779,108 | ASIC2,CAC | 34/- |
| 2_Member | GO Biological Processes | GO:0098660 | inorganic ion transmembrane transport | -7.146180716 | -3.27946028 | 40,779,108 | ASIC2,CAC | 26/726 |
| 3_Summary | GO Biological Processes | GO:0034330 | cell junction organization | -6.540558261 | -2.79877657 | 40,577,779 | ASIC2,ADC | 25/- |
| 3_Member | GO Biological Processes | GO:0034330 | cell junction organization | -6.540558261 | -2.79877657 | 40,577,779 | ASIC2,ADC | 21/533 |
| 4_Summary | KEGG Pathway | hsa05017 | Spinocerebellar ataxia | -6.426351356 | -2.78147967 | 2911,3709, | GRM1,ITPI | 37/- |
| 4_Member | KEGG Pathway | hsa05017 | Spinocerebellar ataxia | -6.426351356 | -2.78147967 | 2911,3709, | GRM1,ITPI | 11/143 |
| 5_Summary | GO Biological Processes | GO:0010975 | regulation of neuron projection development | -5.644430894 | -2.24164187 | 577,1496,1 | ADGRB3,C | 39/- |
| 5_Member | GO Biological Processes | GO:0010975 | regulation of neuron projection development | -5.644430894 | -2.24164187 | 577,1496,1 | ADGRB3,C | 18/460 |
| 6_Summary | GO Biological Processes | GO:0000902 | cell morphogenesis | -5.400826673 | -2.09837767 | 658,1006,1 | BMPR1B,C | 35/- |
| 6_Member | GO Biological Processes | GO:0000902 | cell morphogenesis | -5.400826673 | -2.09837767 | 658,1006,1 | BMPR1B,C | 22/681 |
| 7_Summary | GO Biological Processes | GO:0007423 | sensory organ development | -4.812372244 | -1.84739634 | 658,779,20 | BMPR1B,C | 19/- |
| 7_Member | GO Biological Processes | GO:0007423 | sensory organ development | -4.812372244 | -1.84739634 | 658,779,20 | BMPR1B,C | 19/581 |
| 8_Summary | GO Biological Processes | GO:0030534 | adult behavior | -4.631463917 | -1.75002023 | 2043,5071, | EPHA4,PR | 26/- |
| 8_Member | GO Biological Processes | GO:0030534 | adult behavior | -4.631463917 | -1.75002023 | 2043,5071, | EPHA4,PR | 9/144 |
| 9_Summary | GO Biological Processes | GO:0086042 | cardiac muscle cell-cardiac muscle cell adhesion | -4.405976278 | -1.61399542 | 1525,5318, | CXADR,PK | 26/- |
| 9_Member | GO Biological Processes | GO:0086042 | cardiac muscle cell-cardiac muscle cell adhesion | -4.405976278 | -1.61399542 | 1525,5318, | CXADR,PK | 3/7 |
| 10_Summary | GO Biological Processes | GO:0098609 | cell-cell adhesion | -4.142457527 | -1.38968045 | 1006,1496, | CDH8,CTN | 17/- |
| 10_Member | GO Biological Processes | GO:0098609 | cell-cell adhesion | -4.142457527 | -1.38968045 | 1006,1496, | CDH8,CTN | 17/542 |
| 11_Summary | WikiPathways | WP5294 | Sildenafil treatment | -4.039480974 | -1.30842314 | 5071,5142, | PRKN,PDE | 4/- |
| 11_Member | WikiPathways | WP5294 | Sildenafil treatment | -4.039480974 | -1.30842314 | 5071,5142, | PRKN,PDE | 4/23 |
| 12_Summary | GO Biological Processes | GO:0006936 | muscle contraction | -3.629973732 | -0.96737328 | 779,3751,3 | CACNA1S, | 11/- |
| 12_Member | GO Biological Processes | GO:0006936 | muscle contraction | -3.629973732 | -0.96737328 | 779,3751,3 | CACNA1S, | 10/238 |
| 13_Summary | WikiPathways | WP4549 | Fragile X syndrome | -3.493785102 | -0.8716634 | 2043,2911, | EPHA4,GR | 17/- |
| 13_Member | WikiPathways | WP4549 | Fragile X syndrome | -3.493785102 | -0.8716634 | 2043,2911, | EPHA4,GR | 7/122 |
| 14_Summary | Canonical Pathways | M275 | PID ALPHA SYNUCLEIN PATHWAY | -3.463264088 | -0.8716634 | 2869,5071, | GRK5,PRK | 17/- |
| 14_Member | Canonical Pathways | M275 | PID ALPHA SYNUCLEIN PATHWAY | -3.463264088 | -0.8716634 | 2869,5071, | GRK5,PRK | 4/32 |
| 15_Summary | GO Biological Processes | GO:0098656 | monoatomic anion transmembrane transport | -3.153524577 | -0.63643008 | 368,1080,2 | ABCC6,CF | 9/- |
| 15_Member | GO Biological Processes | GO:0098656 | monoatomic anion transmembrane transport | -3.153524577 | -0.63643008 | 368,1080,2 | ABCC6,CF | 7/139 |
| 16_Summary | Reactome Gene Sets | R-HSA-1650814 | Collagen biosynthesis and modifying enzymes | -3.147959518 | -0.63643008 | 1284,1293, | COL4A2,C | 7/- |
| 16_Member | Reactome Gene Sets | R-HSA-1650814 | Collagen biosynthesis and modifying enzymes | -3.147959518 | -0.63643008 | 1284,1293, | COL4A2,C | 5/67 |
| 17_Summary | GO Biological Processes | GO:0016310 | phosphorylation | -3.147762857 | -0.63643008 | 658,2043,2 | BMPR1B,E | 24/- |
| 17_Member | GO Biological Processes | GO:0016310 | phosphorylation | -3.147762857 | -0.63643008 | 658,2043,2 | BMPR1B,E | 18/721 |
| 18_Summary | WikiPathways | WP2858 | Ectoderm differentiation | -3.098770435 | -0.61825161 | 1006,1496, | CDH8,CTN | 7/- |
| 18_Member | WikiPathways | WP2858 | Ectoderm differentiation | -3.098770435 | -0.61825161 | 1006,1496, | CDH8,CTN | 7/142 |
| 19_Summary | GO Biological Processes | GO:0043484 | regulation of RNA splicing | -3.076237952 | -0.60529728 | 4858,2306 | NOVA2,SE | 8/- |
| 19_Member | GO Biological Processes | GO:0043484 | regulation of RNA splicing | -3.076237952 | -0.60529728 | 4858,2306 | NOVA2,SE | 8/186 |
| 20_Summary | GO Biological Processes | GO:0009100 | glycoprotein metabolic process | -3.054103974 | -0.60235689 | 658,2530,4 | BMPR1B,F | 13/- |
| 20_Member | GO Biological Processes | GO:0009100 | glycoprotein metabolic process | -3.054103974 | -0.60235689 | 658,2530,4 | BMPR1B,F | 11/333 |

**Table S13. The results of the comparison with previously reported high-altitude genes in THL**

| HAAgene_PBS | THL-PBS | Same | Different | THL-specific | rsID | Gene | coding |
|---|---|---|---|---|---|---|---|
| EPAS1 | RAB25 | EPAS1 | MAN2A1 | ASIC2 | rs17607736 | MYO3B | missing |
| EGLN1 | RANBP3L | EGLN1 | FNDC1 | GRM1 | rs353402 | SIPA1L3 | missing |
| HLA-DQB1 | XIRP2 | KHDRBS2 | LHFPL3 | ADGRB3 | rs353398 | SIPA1L3 | missing |
| TMEM132C | ZBTB42 | NBAS | UTRN | BMPR1B | rs353400 | SIPA1L3 | missing |
| ATP13A3 | AKT1 | PRKCE | RPS6KA2 | EPHA4 | rs1725481 | SIPA1L3 | missing |
| L3MBTL2 | RANBP3L | WBSCR17 | SIPA1L3 | CXADR | rs885479 | MC1R | missing |
| SANBR | LOC100506457 | MKL1 | ZNF638 | CDH8 | rs509522 | SETX | missing |
| SLC52A3 | ZNF573 | FANCA | SNCA | PRKN | rs6914051 | MLIP | missing |
| KHDRBS2 | PAIP2B | LCLAT1 | LOC102467213 | CACNA1S | rs353415 | SIPA1L3 | missing |
| BICDL1 | ZBTB42 | CNTNAP2 | SLC26A5 | GRK5 | rs705500 | SIPA1L3 | missing |
| RP11-525K10.3 | CASZ1 | TLL1 | MYO3B | ABCC6 | rs10145015 | ATG2B | missing |
| DISC1 | RANBP3L | DPP6 | DNAJC6 | COL4A2 | rs10901312 | HMCN2 | missing |
| MUC22 | SYT10 | SLIT3 | TRIM67 | NOVA2 | rs833926 | SIPA1L3 | missing |
| RP11-542G1.1 | MYO3B | XIRP2 | CASC15 | CFTR | rs6575582 | ATG2B | missing |
| RP11-804N13.1 | CDH13 | STPG2 | TESPA1 | CNGB1 | rs3912257 | CWC22 | missing |
| RANBP2 | MYO3B | TENM2 | SYN3 | GABRG3 | rs12053447 | ST6GAL2 | missing |
| ZMYND12 | XIRP2 | MACROD2 | C22orf34 | GRM5 | rs4962177 | SETX | missing |
| PGD | SGSM1 | RCAN1 | IFT122 | KCND2 | rs517076 | SETX | missing |
| SBF2 | PKP2 | FAM171A1 | INADL | KCND3 | rs1643466 | SIPA1L3 | missing |
| IGHG3 | SNCA | CDH13 | EYA2 | KCNMA1 | rs353423 | SIPA1L3 | missing |
| NBAS | SGSM1 | OPCML | ZNF710 | PKP2 | rs2542525 | ZNF638 | missing |
| PRKCE | LOC100506457 | | MAGI2 | SLC1A7 | rs3218651 | POLQ | missing |
| STARD13 | XIRP2 | | PAIP2B | SNCA | rs353427 | SIPA1L3 | missing |
| ABCC4 | ANP32A | | TTC12 | NRXN1 | rs351561 | MSR1 | missing |
| RAB40B | ZNF710 | | KCND3 | AKAP6 | rs10901311 | HMCN2 | missing |
| RP11-834C11.5 | CDH13 | | PCDH7 | CTNNA3 | rs1643468 | SIPA1L3 | missing |
| MMUT | F5 | | KCNB2 | KCNK9 | rs1643467 | SIPA1L3 | missing |
| ZBTB4 | MAGI2 | | C1orf101 | KCNK10 | rs833898 | SIPA1L3 | missing |
| SSH2 | PKP2 | | NEK11 | KCNH7 | rs833897 | SIPA1L3 | missing |
| MOV10L1 | KCNMA1 | | ATXN8OS | NALCN | rs1725482 | SIPA1L3 | missing |
| CXCR5 | TESPA1 | | COL23A1 | SLC26A5 | rs2778916 | GABBR2 | missing |
| NKAIN2 | XIRP2 | | WDR27 | CHAT | rs353407 | SIPA1L3 | missing |
| APBA2 | MYO3B | | DCC | PLCB2 | | | |
| RSU1 | MYO3B | | MED27 | RPS6KA2 | | | |
| JADE2 | EFCAB12 | | ATP10B | SYN3 | | | |
| NFAM1 | PAIP2B | | AKT3 | PPFIA2 | | | |
| UPK2 | LOC154449 | | NRXN3 | KCNB2 | | | |
| KALRN | ATXN8OS | | CACNA2D3 | NRXN3 | | | |
| PKD2 | SEMA6D | | LOC101928241 | GABBR2 | | | |
| RP11-6N13.1 | TESPA1 | | RNF220 | CACNA2D3 | | | |
| DHRS3 | PLCB2 | | ACSF3 | DTNB | | | |
| ITPKB | F5 | | PTPRK | RIT2 | | | |
| JAG2 | MAGI2 | | LINC01257 | UTRN | | | |
| CAMKMT | CDH13 | | KLHL1 | FARP1 | | | |
| PDE11A | EPAS1 | | PPFIA2 | GPM6A | | | |
| WARS | FAM155A | | SPIRE2 | ITPR2 | | | |
| NCMAP | CHD5 | | EPN2 | OCA2 | | | |
| RWDD1 | ATXN8OS | | EPHA4 | RYR3 | | | |
| FSTL5 | ATXN8OS | | SLC35F4 | ATP2C2 | | | |
| WBSCR17 | TESPA1 | | LINC00907 | SLC24A2 | | | |
| ZNF160 | TESPA1 | | YTHDF3 | SLC24A3 | | | |
| PWP1 | SATB1 | | RANBP3L | ANO3 | | | |
| FBN3 | MYH15 | | F5 | KCNIP4 | | | |
| CLCN3 | XIRP2 | | SEMA6D | NALF1 | | | |
| PACRG | AKT1 | | TSPAN18 | MYH11 | | | |
| MKL1 | LOC154449 | | SH3GL3 | MYLK | | | |
| FANCA | MYO3B | | KCNH7 | BCL10 | | | |
| SEPSECS | DNAJC6 | | LINC00923 | POLQ | | | |
| STXBP5-AS1 | TESPA1 | | LRRFIP1 | CLYBL | | | |
| TOB2 | FLT1 | | KCNK10 | DNAH11 | | | |
| RAD51B | ATXN8OS | | PKP2 | FLT1 | | | |
| STXBP5L | ALS2CR11 | | LOC154449 | SAA1 | | | |
| CFHR4 | SYN3 | | GPM6A | PTPRK | | | |
| GLRA1 | ANP32A | | RRP7A | PATJ | | | |
| LCLAT1 | CHD5 | | GADL1 | GPHN | | | |
| PDE7B | SLC24A3 | | MEGF11 | NRG3 | | | |
| ARL15 | XIRP2 | | SLC16A9 | PCDH17 | | | |
| FAM160A1 | MAGI2 | | B9D1 | CNTN5 | | | |
| FOXP1 | MYO3B | | NRG3 | HMCN2 | | | |
| RP11-30J20.1 | DNAJC6 | | PUM1 | CTNNA2 | | | |
| CHIAP1 | TESPA1 | | FARP1 | GPR158 | | | |
| CNTNAP2 | TFAP2A | | SNTB1 | NEGR1 | | | |
| LGR4 | NOVA2 | | HMCN2 | PHLDB2 | | | |
| TGFBR3 | SYN3 | | RHBDL2 | DCC | | | |
| CBX5 | EPAS1 | | MAGI1 | PTPRG | | | |
| VSNL1 | ARGFX | | LINC00478 | ULK2 | | | |
| FGF10 | ZNF710 | | SGMS1 | MAGI2 | | | |
| WBP2NL | TESPA1 | | F11-AS1 | SEMA3C | | | |
| ZC3H12C | TESPA1 | | SRSF12 | SETX | | | |
| DAB1 | TESPA1 | | SETX | SS18L1 | | | |
| PNCK | TESPA1 | | PRKG1 | ULK4 | | | |
| KLF7 | SNCA | | GPR115 | SEMA6D | | | |
| GLYAT | XIRP2 | | GBGT1 | FAT3 | | | |
| NEK7 | MYO3B | | TRABD2B | TRIM67 | | | |
| ADAMTS17 | DNAJC6 | | SYT14 | DAAM2 | | | |
| RP11-370P15.1 | MYO3B | | KIAA1211 | MYO3B | | | |
| GREM2 | EGLN1 | | SORCS1 | NBN | | | |
| FOXJ3 | INADL | | NALCN | SH3GL3 | | | |
| COL4A4 | C7orf63 | | OCA2 | PACSIN2 | | | |

TLL1
DDRGK1
LRCH1
ACSS2
TBX3
GUSBP1
ARAP2
DLGAP2
RP11-982M15.8
RP11-86A5.1
RP4-601P9.2
DEPDC1B
DPP6
KCNAB1
DNAH5
RILPL2
RXFP1
SUPV3L1
CBLL2
SLC4A10
TP53BP1
TSC22D2
KLHL28
NAALADL2
SLIT3
DNAJC10
LOC107986973
PBX3
PRIM2
AC133680.1
OR9Q1
RP11-418J17.1
OR7E62P
DMXL2
C1orf21
ACPL2
FOXN3
LPP
GOPC
COL4A5
FCGR2A
TARSL2
REEP5
hsa-mir-490
RP11-766F14.2
RBFOX1
AK3
PUS7L
ADH7
BAG4
TSPAN9
ACACA
HS3ST5
XIRP2
GALNT9
ENDOU
STXBP3
STPG2
HSD11B1
RP11-466L17.1
LINC00877
PCDH15
TENM2
POC1A
PTPRD
PRDM4
TRABD
NREP
MACROD2
SYMPK
ITCH
ASB10
IDH3G
RCAN1
LRP1B
XDH
HADHA
FAM171A1
UPP2
WDPCP
MYO1E
TMEM182
MALRD1
SETD1B
HAND2-AS1
KIAA1217
PPM1H
MRPL23
LNPEP
ADORA1
RP11-733O18.1

ZNF710
TESPA1
XIRP2
DNAJC6
XIRP2
COL23A1
COL23A1
ZNF710
ZNF710
ZNF710
MYO3B
TVP23A
AKT1
SLC24A2
C7orf63
LHFPL3
ZNF71
PAIP2B
UTRN
XIRP2
CACNA2D3
CDH22
PPP2R2C
TESPA1
ANP32A
DNAJC6
DNAJC6
DNAJC6
SNCA
SNCA
ZNF710
EPAS1
XIRP2
CRIPT
TESPA1
RPS6KA2
XIRP2
ANP32A
LOC154449
EFCAB12
LINC00304
XIRP2
AKT1
LOC339166
ZNF638
EGLN1
COL27A1
SYN3
CHD5
FREM3
PAIP2B
PAIP2B
PAIP2B
PAIP2B
COL23A1
SPRR2E
GPM6A
SNCA
CALCR
SRSF12
TESPA1
LOC102467213
FAM135B
LHFPL3
INADL
NOVA2
DNAJC6
DNAJC6
NAGK
XIRP2
EYA2
LOC154449
WNT3
EPAS1
KLHL1
PAIP2B
LHFPL3
LOC284294
DNAJC6
CALCR
COL23A1
CHL1
XIRP2
TTC12
DUSP16
TESPA1
CHD5
CHD5
SNCA
ZNF710
AHNAK2

CWC22
MLLT3
ERMARD
GRM1
KCNMA1
FAM155A
SLC24A2
FREM3
OSBPL3
NUCKS1
GSAP
CECR2
OGDH
ADAM22
GALNT14
PAK6
C10orf11
ZNF540
C4orf22
ZNF544
MXRA7
FIGN
RYR3
BCAS1
TLE1
PALD1
FAT3
FBN1
CDK14
NTM
MSR1
SLCO1B1
DAAM2
PDE4B
SGSM1
PLCB2
FLT1
SLC24A3
NOVA2
FAM135B
COL4A2
GABRG3
LRRC3B
RBM19
NRXN1
ENPP6
MX1
CTNNA3
NTN4
PDZD2
SLC1A7
NEGR1
PPM1B
CLMP
ATXN1
KLF12
MLIP
ARMC9
TMTC1
CLYBL
EPHA1-AS1
GON4L
TSHZ2
NOX3
BTAF1
ATG2B
TMEM232
CACNA1S
RBFOX3
CCDC50
ZAK
DIS3L2
RPL34-AS1
MAGI3
PARK2
PCDH17
POLQ
VENTX
GPC6
CRYM
LOC101927870
TMPRSS2
CDH8
GPR158
LINC00607
LPGAT1
ZNF573
SATB1
ARGFX
LOC284294
AHNAK2

ADCK1
CRACD
MAN2A1
TENM4
AKT3
PRKG1
RORA
ATXN1
ATXN8OS
PUM1
ATG2B
FBN1
PTPN11
SIPA1L3
B9D1
CECR2
NOX3
BNC2
IFT122
MEGF11
KDM2B
PAX5
KLHL1
PAK6
LATS2
NTN4
SLC19A2
COL6A3
ADAMTS3
P3H2
COL23A1
CDK14
PI4KA
MAP3K20
NEK11
SGMS1
NAA20
TTLL7
AGBL4
METAP1D
GALNTL6
CRYM
MC1R
MSR1
SLCO1B1
OSBPL3
BCAS3
SORCS1
PCDH7
FNDC3A
ZNF638
CWC22
ARB2A
AHNAK2
SRSF12
RBFOX3
FUT8
DPM1
ERP44
GBGT1
GALNT14
ADAMTS12
TMTC1
ST6GAL2
TMPRSS2
SNTB1
PDE4B
MLIP
ADGRF4
ADGRD1

| | | |
|---|---|---|
| ZFHX3 | TSPAN18 | EIF4B |
| KCNS3 | KLHL1 | AKAP6 |
| OAS3 | XIRP2 | GRK5 |
| USP40 | DNAJC6 | ANKRD36 |
| BSND | PLCB2 | 2-Mar |
| CDH13 | MAN2A1 | MTUS1 |
| OPCML | LOC100132352 | BNC2 |
| SP110 | COL4A2 | RNGTT |
| GCH1 | SIPA1L3 | PHLDB2 |
| GFRA4 | MAGI2 | LOC155060 |
| GPR98 | NRXN3 | MYO18B |
| DCDC2C | PAN3 | DERA |
| | PAN3 | SAA1 |
| | LHFPL3 | GALNTL6 |
| | EPAS1 | LINC00669 |
| | GABRG3 | PNRC1 |
| | DNAH5 | SLC35F3 |
| | PAIP2B | TTC27 |
| | TSPAN18 | MC1R |
| | AKT1 | CCDC66 |
| | UTRN | USP3 |
| | EIF4B | CFTR |
| | SIPA1L3 | CRNKL1 |
| | PPFIA2 | KCND2 |
| | SETX | SOS2 |
| | DUSP16 | PACSIN2 |
| | LOC154449 | ASH2L |
| | UTRN | FAM172A |
| | INADL | EIF3H |
| | AKAP6 | HECTD4 |
| | SNCA | MYLK |
| | LOC101928241 | TXNRD2 |
| | ZNF710 | MRC1 |
| | MYO3B | ESRRG |
| | LYPD2 | COL6A3 |
| | TESPA1 | LINC00587 |
| | DNAJC6 | DNMBP |
| | DNAJC6 | LOC101928516 |
| | PRKG1 | SLC19A2 |
| | RPS6KA2 | HYDIN2 |
| | F5 | PTPRG |
| | OPCML | SYNPR |
| | FANCA | BCL10 |
| | OSBPL3 | TTLL7 |
| | SNX19 | MYLK-AS1 |
| | LAIR2 | LINC00935 |
| | PAIP2B | PTER |
| | RPS6KA2 | ITPR2 |
| | GPR115 | ARHGEF37 |
| | CHD5 | SDHAP1 |
| | LOC101928241 | ASCC3 |
| | XIRP2 | C1orf52 |
| | PPFIA2 | ADCK1 |
| | CHD5 | CXADR |
| | CHD5 | MLK7-AS1 |
| | CHD5 | WDR87 |
| | CHD5 | GPHN |
| | ZNF710 | METAP1D |
| | MSRA | FUT8 |
| | GPM6A | BRF1 |
| | CALCR | SERHL |
| | PPP2R2C | KCNIP4 |
| | TAMM41 | CNGB1 |
| | SNCA | TM2D1 |
| | LRRC3B | UBE2V2 |
| | INADL | PPP2R5E |
| | FREM3 | BCAS3 |
| | DNAJC6 | TPP2 |
| | DNAJC6 | PI4KA |
| | PTPRK | ST6GAL2 |
| | SPIRE2 | LOC493754 |
| | COL1A2 | DPYD |
| | LHFPL3 | RPL32P3 |
| | LOC101929181 | ANO3 |
| | RBM19 | CNTN5 |
| | ZNF638 | PAX5 |
| | WDR27 | CALN1 |
| | ATXN8OS | AGBL4 |
| | ATXN8OS | ADAMTS12 |
| | PAIP2B | ATP2C2 |
| | PAIP2B | DTNB |
| | PAIP2B | CNTLN |
| | RPS6KA2 | KDM2B |
| | KCND3 | ECHDC1 |
| | KCND3 | NAA20 |
| | KCND3 | ZNF385B |
| | PPP2R2C | LINC-PINT |
| | OPCML | ANKS6 |
| | CASC15 | 4-Mar |
| | MYO3B | NBN |
| | PAN3 | DENND3 |

| | |
|---|---|
| NUCKS1 | ERP44 |
| UTRN | CCDC170 |
| PTPRD | SEMA3C |
| CHD5 | ANKRA2 |
| RAB8A | ULK2 |
| KLHL1 | ADAMTS3 |
| ATXN8OS | EMCN |
| SLIT3 | BAI3 |
| NCKAP5L | SPATA13 |
| GSAP | ULK4 |
| MAGI2 | MYH11 |
| CASC15 | GRM5 |
| TSTD1 | GAB2 |
| RGS9 | EGFEM1P |
| RRP7A | LOC101927592 |
| C7orf63 | C8orf37-AS1 |
| SLC24A2 | LOC729911 |
| RNF220 | FNDC3A |
| SEMA6D | IL12A-AS1 |
| C1orf110 | BMPR1B |
| KLHL1 | CECR1 |
| ZNF638 | DNAH11 |
| ZNF638 | GPR133 |
| NAGK | CHAT |
| GRK5 | 8-Mar |
| RCL1 | PTPN11 |
| TENM2 | THSD7B |
| KCND3 | PLEKHA5 |
| NRXN1 | SS18L1 |
| DNAJC6 | VTI1A |
| DNAJC6 | INPP4B |
| DUSP16 | FAM19A5 |
| AKT1 | LEPREL1 |
| ATXN8OS | AP1S3 |
| INADL | KCNK9 |
| COL23A1 | GABBR2 |
| COL23A1 | ZFR |
| COL23A1 | TBC1D1 |
| IGLON5 | RIT2 |
| MED27 | TCF25 |
| ANKRD36 | ZNF346 |
| PPFIA2 | ASIC2 |
| TESPA1 | ABCC6 |
| FNDC1 | ZFAND6 |
| DYTN | RORA |
| ZNF710 | OTOGL |
| DNAJC6 | NKAIN1 |
| ENPP6 | TENM4 |
| PPP2R2C | LATS2 |
| 2-Mar | DPM1 |
| TVP23A | CTNNA2 |
| KDM5A | STEAP2-AS1 |
| MAP4K1 | |
| MYO3B | |
| MTUS1 | |
| MAGI2 | |
| SH3GL3 | |
| PPP2R2C | |
| MAP4K1 | |
| LOC102467213 | |
| MKL1 | |
| EYA2 | |
| MX1 | |
| MAGI2 | |
| EFCAB12 | |
| TESPA1 | |
| ZNF710 | |
| RPS6KA2 | |
| MYO3B | |
| DNAJC6 | |
| LHFPL3 | |
| EPAS1 | |
| RPS6KA2 | |
| MSRA | |
| FNDC1 | |
| LHFPL3 | |
| UTRN | |
| CASC15 | |
| C22orf34 | |
| CDH13 | |
| PIP5K1C | |
| BNC2 | |
| RNGTT | |
| SDCCAG8 | |
| LOC100132352 | |
| MAN2A1 | |
| RPS6KA2 | |
| RPS6KA2 | |
| ANKFY1 | |
| C1orf110 | |
| NAGK | |

SLC24A2
PAIP2B
LOC101929181
CECR2
MAN2A1
TRIM67
TRIM67
NCKAP5L
GNG7
GALNT18
PPP2R2C
SORCS2
MYO3B
TVP23A
ATXN8OS
ATXN8OS
IFT122
GADL1
GADL1
LOC101928241
SNCA
SNCA
MAN2A1
ABCA12
MYO3B
SYN3
ATXN8OS
GBGT1
COL4A2
PAIP2B
ZNF638
ZNF638
FNDC1
TRIM67
LOC101928241
EPN2
MDH1B
EPAS1
OGDH
RANBP3L
EPAS1
ZNF638
C7orf63
C7orf63
CTNNA3
ZNF710
SYN3
PAN3
LHFPL3
LHFPL3
KLHL1
TRABD2B
NAGK
MSRA
PKP2
ZNF781
NTN4
SEMA6D
RPS6KA2
RPS6KA2
DYTN
CASZ1
MAGI2
RRP7A
KCNB2
PDZD2
TRIM67
TRIM67
CRTAP
PTPRK
ZNF710
CALCR
CALCR
PAN3
FNDC1
ZNF710
SEMA6D
ZNF638
ZNF638
FAM135B
FNDC1
DNAJC6
TESPA1
LOC102467213
RNF220
ZNF710
ATXN8OS
GBGT1
CASC15
TRIM67
TRIM67

EPAS1
SIPA1L3
LYPD2
DUSP16
CASC15
PGR
CALCR
SRSF12
SRSF12
MAN2A1
GPR115
KCNH7
PRKG1
PRKG1
PHLDB2
CALCR
SNCA
LRRC3B
ZBTB42
LCLAT1
LCLAT1
UTRN
LOC339166
SNCA
MAN2A1
MAN2A1
MAN2A1
ZNF710
ZNF710
LOC102467213
DNAH5
PPFIA2
DYTN
DYTN
DYTN
ZNF341
ADAM22
AGO3
MAGI2
XIRP2
SYN3
SH3GL3
MSRA
FNDC1
FNDC1
LOC155060
SLC26A5
MAN2A1
NAGK
LOC101929181
LOC101929181
MYT1L
LOC102467213
TVP23A
RRP7A
C1orf110
C1orf110
SPIRE2
MYO18B
FNDC1
F13A1
DERA
SIPA1L3
DUSP16
LHFPL3
SAA1
LHFPL3
MEGF11
TRIM67
SIPA1L3
SYN3
TVP23A
C7orf63
C7orf63
MSRA
SLC26A5
ANKK1
ZNF638
PAN3
PAN3
MLIP-IT1
LOC101928241
PAN3
LOC101928911
LHFPL3
CASZ1
NAGK
ZNF850
SYN3
MYO3B
FREM3

GALNT14
DUSP4
DUSP16
SYT14
ATXN8OS
LOC102467213
MAGI2
ZNF638
MAN2A1
C1orf110
LOC102467213
MSRA
XIRP2
RGS9
XIRP2
ZNF638
SNCA
F5
RCAN1
RCAN1
SH3BP4
AGO3
RPS6KA2
LHFPL3
DUSP4
PGR
INADL
SNCA
LOC102467213
CASC15
TRIM67
DUSP16
PPP2R2C
PPP2R2C
PPP2R2C
ZNF641
ZNF641
KIAA1211
KIAA1211
INADL
SLC1A7
INADL
INADL
PAK6
EPN2
MKL1
FLJ37505
LHFPL3
LHFPL3
LHFPL3
MSRA
TOPBP1
MBD4
ZNF638
SNCA
LHFPL3
GALNTL6
RGS9
LINC00669
LOC101929181
LOC101929181
L1TD1
C10orf11
CRTAP
TRIM67
FNDC1
MPHOSPH10
MPHOSPH10
LOC102467213
DYTN
GPR115
DPP6
LDB3
ZNF638
MAN2A1
MAN2A1
MAN2A1
MAN2A1
FNDC1
WTIP
LOC400558
NTN1
PGR
MAN2A1
PNRC1
SRSF12
UTRN
TSNAX
TSPAN18
PTPRD
LINC00923

C1orf110
CALCR
USF1
PTPRD
PCDH7
SLC16A9
PAIP2B
MAN2A1
TRIM67
TRIM67
DUSP16
DUSP4
LOC102467213
SLIT3
ZNF540
ZNF540
PRPF31
RNF220
WDR27
FNDC1
SYT10
SPIRE2
MSRA
LINC00923
LOC102467213
KCNB2
CALCR
EPN2
EPN2
EPN2
EPN2
B9D1
FNDC1
CACHD1
SIPA1L3
SIPA1L3
WBSCR17
SNCA
NEGR1
INADL
FNDC1
SLC26A5
SLC26A5
MAN2A1
SLC35F3
TTC27
KLHL1
DUSP16
B9D1
MAN2A1
PPFIA2
TRIM67
SLIT3
UTRN
LOC102467213
NRG3
CDH13
LINC01257
MYO3B
CRYZ
DUSP16
XIRP2
PAN3
PAN3
CALCR
CALCR
CALCR
CALCR
SH3GL3
LHFPL3
RHO
PPP2R2C
MC1R
AGO3
NRG3
LOC102467213
CALCR
RNF220
YY1AP1
NCKAP5L
NCKAP5L
COL6A5
TRIM67
TRIM67
TRIM67
TRIM67
TRIM67
TRIM67
APBB3
PPM1B
UTRN

SORCS1
LOC646626
LOC102467213
CCDC66
TEAD4
MSRA
MYO3B
MAGI2
MAGI2
EIF4EBP2
MYO3B
PTPRK
LOC100131257
MAN2A1
MAN2A1
MAN2A1
UBE2G1
USP3
INADL
NALCN
CALCR
CALCR
CALCR
CALCR
CALCR
FNDC1
LHFPL3
C7orf63
C7orf63
PPFIA2
C4orf22
CDH13
CALCR
DUSP4
LHFPL3
ZNF638
LOC102467213
MAGI2
SPIRE2
RPS6KA2
RPS6KA2
RPS6KA2
LOC101448202
LOC149373
LDB3
PTPRK
ZNF638
ZNF638
LHFPL3
MSRA
CDH13
LHFPL3
DUSP4
TFAP2A
LINC00923
CLMP
FANCA
USF1
MAN2A1
FNDC1
FNDC1
FNDC1
FNDC1
CAMTA1
TSNAX
LHFPL3
LHFPL3
SLC26A5
MED27
FNDC1
ZNF641
CALCR
FAM155A
FAM155A
CFTR
SH3GL3
SNCA
KCNH7
IFT122
KCND3
KCND3
TESPA1
TESPA1
FNDC1
MDH1B
BTG3
LOC102467213
INADL
PUM1
PRELP
SMARCA2

C22orf34
C1orf110
MED27
CRNKL1
ATXN1
KLF12
SLC26A5
SLC26A5
MSRA
PHAX
PHAX
LHFPL3
LOC102467213
SNCA
SNCA
UTRN
KCND2
MSRA
COL1A2
SLC16A9
ZNF793
ATP10B
DUSP16
DUSP16
DUSP16
RPS6KA2
LHFPL3
USF1
UTRN
SETX
SYN3
ZNF544
ZNF544
LHFPL3
MAN2A1
MAN2A1
UTRN
UTRN
PGR
IFT122
ZNF638
ZNF638
ZNF638
WDR27
BPESC1
SLC1A7
SOS2
RBFOX1
SORL1
LHFPL3
KCNB2
LCLAT1
DNAH5
PACSIN2
C1orf110
ZNF638
PRKG1
MDH1B
LOC102467213
LHFPL3
LHFPL3
LHFPL3
LHFPL3
LOC100506457
CBWD3
TTC12
XIRP2
ASH2L
DUSP4
DUSP4
CDH13
PAN3
LRRFIP1
KCNH7
GBGT1
EPN2
EPHA4
CRTAP
CRTAP
DNAJC6
MAN2A1
LHFPL3
TENM2
KCND3
MAN2A1
MAN2A1
MSRA
KCNB2
CTNND2
PAN3
FAM172A

MAN2A1
MAN2A1
MAN2A1
MAN2A1
XIRP2
XIRP2
XIRP2
EIF3H
ATP2A3
SLC35F4
TESPA1
UTRN
DUSP16
HECTD4
DUSP4
C5orf27
CRTAP
SLC16A9
CASP14
PVRL4
KAZN
KAZN
INADL
TFAP2A
DUSP16
DUSP16
DUSP16
DUSP16
SIPA1L3
LOC102467213
KCNB2
USF1
KCND3
KCND3
PPP2R2C
ATXN8OS
LINC00907
LHFPL3
LHFPL3
LHFPL3
MYO3B
MYO3B
MYO3B
COL23A1
COL23A1
CASC15
CASC15
MED27
ZNF710
CLMP
MYLK
PTPRK
OCA2
SNCA
MLIP
MAN2A1
MAN2A1
MYO3B
XIRP2
ATP10B
SIPA1L3
MAN2A1
PPP2R2C
CWC22
MAN2A1
ZNF638
MAN2A1
IFT122
LHFPL3
UTRN
C1orf110
C1orf110
MSRA
MYO3B
PPP2R2C
PRKD3
LOC101928241
PCAT2
MAGI2
LINC00923
PKP2
ST3GAL1
NRXN3
SIPA1L3
MDH1B
MAGI2
XIRP2
SNCA
MAN2A1
TSPAN18
TXNRD2

PAN3
PAN3
C22orf34
C22orf34
C22orf34
C22orf34
FNDC1
C22orf34
DNAJC6
YWHAEP7
MRC1
FNDC1
ZNF638
DNAJC6
MED27
FNDC1
ESRRG
FARP1
MXRA7
CSMD1
ARMC9
ZNF641
NCKAP5L
NCKAP5L
LHFPL3
OGDH
OCA2
COL6A3
C1orf110
USF1
SNCA
MLLT3
H1FOO
NRG3
FNDC1
FNDC1
FNDC1
FNDC1
GALNT18
UTRN
UTRN
UTRN
UTRN
CRTAP
CRTAP
CRTAP
CSMD1
TMTC1
LINC00587
ZNF641
MYO3B
DNMBP
SNTB1
LINC01257
LINC01257
RPS6KA2
RNF220
INADL
INADL
FNDC1
CWC22
UTRN
MSRA
COL23A1
COL23A1
MKL1
DNAJC6
LOC101928516
PAN3
MEGF11
RPS6KA2
GBGT1
SLC19A2
LOC149373
TRIM67
ATP10B
ATP10B
KIAA0232
FIGN
ZNF638
PPP2R2C
SIPA1L3
CALCR
CALCR
SIPA1L3
MAN2A1
UTRN
UTRN
TSPAN33
NRXN3
LOC101928911

HYDIN2
RPS6KA2
FREM1
RPS6KA2
DOPEY2
PAN3
PAN3
PAN3
CLYBL
FNDC1
BCDIN3D-AS1
BCDIN3D
BCDIN3D
RIN2
C7orf63
ADAM22
CAMK1D
C22orf34
MAN2A1
MAN2A1
PCDH7
KCNB2
RYR3
ENPP7
PAN3
C1orf110
C1orf110
C1orf110
ETNPPL
PTPRG
EPHA1-AS1
BCAS1
TMEM201
SORCS1
TRIM67
ADAM22
SIPA1L3
CALCR
RPS6KA2
SYNPR
HMCN2
CRYGB
MAN2A1
SYN3
FNDC1
DNAJC6
LOC102467213
LOC102467213
EGR4
TAGLN2
FNDC1
FAM120B
SLC16A9
DPP6
IFT122
AGO3
GON4L
TLE1
MIR4432
BCL10
LOC646626
LINC00923
LOC102467213
ACSF3
LOC339166
MDH1B
LINC01257
FNDC1
XIRP2
CASC15
PTPRK
UTRN
PAN3
TTLL7
MYLK-AS1
MDH1B
MAN2A1
RPS6KA2
RPS6KA2
COL1A2
LINC00935
MPHOSPH10
CRTAP
PTER
TESPA1
CRTAP
C1orf101
RPS6KA2
INADL
PCDH7
CALCR

CALCR
CALCR
EYA2
EYA2
ANKH
SIPA1L3
RPS6KA2
SYN3
TSHZ2
KCND3
KCNB2
SNCA
NOX3
LHFPL3
EYA2
CACNA1A
CACNA1A
ZNF638
BTAF1
UTRN
CASC15
LHFPL3
LOC101928911
MAN2A1
TSHZ2
LOH12CR1
KIAA0232
CACNA2D3
FNDC1
FNDC1
FNDC1
FNDC1
FNDC1
FNDC1
FNDC1
FNDC1
FNDC1
DCC
SPIRE2
EYA2
ATP10B
ATP10B
NRXN3
LOC339166
FNDC1
TAMM41
LHFPL3
C22orf34
C22orf34
AK3
ITPR2
CALCR
CALCR
CALCR
CALCR
CALCR
TLL1
RHBDL2
MDH1B
MED27
MSRA
LOC102467213
SYN3
LINC01266
ARHGEF37
ZNF638
ATXN1
GPM6A
NUCKS1
SIPA1L3
FNDC1
PLCB4
LOC101927347
TENM2
TENM2
SDHAP1
MAN2A1
LOC102467213
TLE1
B9D1
RPS6KA2
BCAS1
MAN2A1
MAN2A1
C4orf22
PALD1
MDH1B
GADL1
CALCR
CALCR
CALCR

CALCR
CALCR
CALCR
CALCR
MYEF2
B9D1
WDR27
MAGI2
LINC00923
ZNF638
KCNK10
FLJ37505
DUSP16
DUSP16
RPS6KA2
RPS6KA2
ASCC3
TESPA1
RPS6KA2
SIPA1L3
SIPA1L3
XIRP2
DNAJC6
NRG3
MAN2A1
C1orf110
C1orf52
FLJ37505
ADCK1
FAT3
C22orf34
OCA2
SIPA1L3
XIRP2
RNF220
TVP23A
C4orf22
FNDC1
LHFPL3
LHFPL3
UTRN
CASC15
CASC15
IFT122
IFT122
LHFPL3
LHFPL3
CASC15
ENPP6
ITPR1
EYA2
EYA2
EYA2
TAMM41
FBN1
SLC35F4
PCDH7
MAN2A1
MAN2A1
SYN3
NUCKS1
CXADR
SIAH2
PAN3
MXRA7
MAN2A1
RGS9
RGS9
RGS9
RGS9
INADL
SLC26A5
SLC26A5
PHAX
PTPRK
MAN2A1
EYA2
ZNF638
XIRP2
MLK7-AS1
SLC26A5
SLC26A5
SLC26A5
SLC26A5
SLC26A5
SLC26A5
SLC26A5
SLC26A5
SLC26A5
SLC26A5
MAGI2

FNDC1
CACHD1
ACSF3
BTAF1
ENPP7
WDR87
RPS6KA2
KAZN
LOC101928911
PCDH7
SLC26A5
FGF10
CDK14
SIPA1L3
SYN3
PUM1
GPHN
KCNH7
MAN2A1
LOC102467213
IQGAP1
RPS6KA2
C1orf110
C1orf110
KCND3
IFT122
IFT122
IFT122
IFT122
IFT122
IFT122
IFT122
IFT122
LHFPL3
LHFPL3
LHFPL3
ZNF638
PAN3
AKT3
BCAS1
RFX3
ATG2B
TVP23A
PCDH7
RAB8A
LOC339166
GPM6A
LYNX1
METAP1D
MPHOSPH10
UTRN
UTRN
UTRN
UTRN
LINC00907
SFPQ
RPS6KA2
SYN3
FUT8
BRF1
GALNT14
PHAX
ZNF544
SERHL
KCNIP4
LOC102467213
SYN3
SNCA
SNCA
SNCA
NTM
ST3GAL4-AS1
LOH12CR1
LHFPL3
KAZN
PPP2R2C
RPS6KA2
RNPEPL1
NEK11
ANP32A
MLLT3
HMCN2
HMCN2
USP35
LOC102467213
PAN3
TMEM232
CNGB1
ZNF781
TM2D1
SIPA1L3

UBE2V2
LINC00942
GPR115
EYA2
EYA2
PAK6
DNAJC6
GADL1
GAS7
MSRA
DCC
COL1A2
SIAH2
COL23A1
PDZD2
TSPAN18
MAGI1
MDH1B
MDH1B
MDH1B
MDH1B
MDH1B
FFAR3
RHBDL2
RPS6KA2
SPIRE2
FLJ37505
LHFPL3
INADL
RNF220
PTPRD
PPP2R5E
XIRP2
ZNF638
CASC15
MSR1
CRTAP
CRTAP
ERMARD
CAMK1D
C22orf34
C22orf34
CACNA1S
SLC35F4
THRA
SLC26A5
FLJ37505
AKT3
IFT122
LHFPL3
LHFPL3
LINC00304
PAN3
SLCO1B1
UTRN
DCC
SLC26A5
MSRA
RBFOX3
LOC149373
ATG2B
WDR27
PPFIA2
LOC101929181
SORL1
CAMTA1
INADL
MAGI2
ZNF638
DISP2
YTHDF3
LINC00478
RBM19
KCNH7
MYO3B
SYT14
SYT14
OCA2
ZNF638
OPCML
EPAS1
MAGI1
TRIM67
TTC12
AGO3
KCND3
KCNK10
MAN2A1
LDB3
PAN3
PAN3

GNG7
KIAA0232
PAN3
EPHA4
EPHA4
BCAS3
KIAA0232
UTRN
ERMARD
NCKAP5L
LHFPL3
MSR1
CDH22
B9D1
PHF10
LHFPL3
TPP2
LOC101928911
C1orf101
RPS6KA2
C1orf101
CWC22
FIGN
SPIRE2
YWHAEP7
C1orf101
NEK11
LOC101928241
PPP2R2C
CCDC50
TSPAN18
LINC01266
PI4KA
CASC15
LOC102467213
GALNT14
TTC12
TTC12
ST6GAL2
CSMD1
SETX
CASC2
RYR3
NALCN
LINC01257
LINC01257
TMTC1
WDR27
EGLN1
TESPA1
C22orf34
C22orf34
C22orf34
C22orf34
C22orf34
SETX
LOC493754
MCEE
ABCA3
RANBP3L
DCC
RHO
FNDC1
WNT2
PRKCE
KCNB2
XIRP2
LHFPL3
AKT3
MSRA
MSRA
SIPA1L3
MCM8
TSPAN33
SYN3
MSRA
TAMM41
DPYD
DNAJC6
FARP1
FNDC1
FRMD5
RPL32P3
TSNAX-DISC1
EPHA4
EPHA4
EPHA4
ANO3
RFX3
MGST1
CASC15

ZNF638
CASC15
UBE2Q2P2
KIAA0232
NXPH1
LOC102467213
ZAK
SIPA1L3
SIPA1L3
C10orf11
TVP23A
PAN3
CNTN5
MAN2A1
MAN2A1
UTRN
UTRN
UTRN
ARMC9
LOC100131257
PAX5
SULT6B1
TRABD2B
YWHAEP7
YWHAEP7
YWHAEP7
TRIM67
FIGN
CNTN4
PAN3
DIS3L2
PKP2
C1orf101
RTN4RL1
CERK
CNTN4
RPL34-AS1
LINC00478
FNDC1
FNDC1
FNDC1
FNDC1
FNDC1
MAGI3
SLC24A3
ZNF641
MAN2A1
MAN2A1
MAN2A1
HSH2D
UTRN
COL23A1
PTPRD
PCDH7
C1orf110
IFT122
PGAM1P5
UBE2Q2P2
UBE2Q2P2
FNDC1
FNDC1
DAAM2
RRP7A
CALN1
KCNMA1
TRIM67
C22orf34
LDB3
RRP7A
RGS9
PPP2R2C
RPS6KA2
PCDH7
NEK11
APCDD1L
KCND3
KCND3
MAN2A1
CACNA2D3
CASC15
CACNA2D3
GON4L
AGBL4
LINC00507
SYN3
TSNAX
UTRN
TENM2
PPP2R2C
CACNA2D3
CSMD1

MAN2A1
CDH22
SYN3
CTNNA3
MPHOSPH10
MPHOSPH10
FNDC1
RBFOX3
DISP2
DISP2
DISP2
MAN2A1
MAN2A1
CNTN4
NALCN
ATP10B
FBN1
CACNA2D3
EYA2
ADAMTS12
TRIM67
NTN1
ATP2C2
INADL
IFT122
EPHA1-AS1
MSRA
RPS6KA2
FNDC1
SIPA1L3
EYA2
CDK14
CECR2
GABRG3
ACSF3
GNG7
SLC26A5
SLC26A5
SLC26A5
SNCA
LINC01257
NRXN3
SEMA6D
ANKK1
FAM120B
WDR27
WDR27
DTNB
MAN2A1
SNTB1
PAIP2B
PGR
FAM171A1
CNTLN
C22orf34
LINC00305
ERMARD
DNAH5
IQCE
SIPA1L3
SNTB1
C1orf101
C1orf101
C1orf101
EIF3K
PHAX
GOT2
NDUFA9
OSBPL3
LOC100506457
PARK2
MEGF11
PCDH17
PCDH17
RNF220
KDM2B
UTRN
LOC100144602
YTHDF3
ECHDC1
FNDC1
RHBDL2
ANKRD20A5P
GADL1
SIPA1L3
SIPA1L3
XIRP2
TVP23A
SGMS1
FNDC1
FNDC1

FNDC1
DCC
TRABD2B
PCDH7
MYO3B
ZNF638
MAN2A1
F11-AS1
PARK2
OGDH
POLQ
MAN2A1
RPS6KA2
RPS6KA2
COL1A2
MDH1B
MDH1B
CALCR
VENTX
ZAK
LOC102467213
NAA20
DNAJC6
ZNF385B
DCC
DUSP16
DUSP16
EYA2
EYA2
FARP1
COL1A2
MRPL4
UTRN
MAGI2
FNDC1
FNDC1
MEGF11
SIPA1L3
SIPA1L3
SIPA1L3
SIPA1L3
AGO1
LINC00907
RGS9
NRXN3
KIAA0232
MCPH1
GSAP
LHFPL3
DAAM2
CACNA2D3
LINC-PINT
LOC102467213
CALCR
TVP23A
F11-AS1
TTC12
TTC12
PTPRK
ANKS6
MAP9
C1orf101
C1orf101
C1orf101
C1orf101
NRXN3
AGO3
ZNF638
NRXN3
MEGF11
KCND3
KCND3
KCND3
KCND3
EPS8
LINC01266
SNCA
SNCA
CACNA2D3
CACNA2D3
KLF12
MAN2A1
LOC100131257
NCKAP5L
PCDH7
ATP10B
ATP10B
ATP10B
ATP10B
RRP7A
OPCML

OPCML
MCEE
4-Mar
MSRA
SNCA
MAN2A1
EYA2
EYA2
DLK1
VENTX
YWHAEP7
UTRN
RHBDL2
RHBDL2
SLC35F4
LOC101928911
NBN
MLLT3
DENND3
TTC12
SGMS1
ZNF793
ERP44
CCDC170
SEMA3C
DUSP16
MAGI1
ACER1
SYT14
LINC01266
NEGR1
ANKRA2
MED27
MED27
MAN2A1
MSR1
SFPQ
SFPQ
DUSP4
ULK2
MAP4K1
PPP1R26
PCAT2
CASC15
CASC15
PCDH7
ACSF3
KAZN
HMCN2
SYN3
ANKRD20A5P
GPC6
SLC26A5
SLC26A5
SLC26A5
SLC26A5
TRAM1
POLQ
TENM2
LINC00478
RPS6KA2
RPS6KA2
UTRN
UTRN
ADAMTS3
FAT3
PAN3
CSMD1
YTHDF3
MDH1B
MDH1B
CRYM
MYO3B
KCNB2
OPCML
MAGI1
NOL4L
PHAX
PHAX
MX1
FLJ37505
FLT1
TRIM67
CNTNAP2
SGMS1
IFT122
IFT122
PCDH7
LOC339166
FBN1
KCNB2

LINC00478
ZNF641
F13A1
LINC00507
NBAS
F11-AS1
MAN2A1
LRRFIP1
LRRFIP1
LRRFIP1
MED27
ANKH
NRXN3
ACSF3
ACSF3
ACSF3
GRM1
GRM1
RPL34-AS1
LOC101928911
PAN3
DAAM2
PAN3
EMCN
CASC15
CASC15
ELOF1
SYN3
SYN3
LINC00907
LINC00907
SIPA1L3
SIPA1L3
SIPA1L3
SIPA1L3
SIPA1L3
SIPA1L3
SIPA1L3
BLZF1
UBE2Q2P2
BAI3
GRM1
GRM1
FAT3
SPATA13
ULK4
MYH11
DCC
GRM5
CRTAP
SIPA1L3
NEK11
LOC101928911
NEK11
NEK11
NEK11
TAMM41
PUM1
GAB2
NR5A2
NOX3
KCNMA1
SLC16A9
MLK7-AS1
KIAA1211
EGFEM1P
IFT122
PUM1
PUM1
XIRP2
LINC00276
RNF220
PHAX
LOC101927592
MAN2A1
PAN3
PAN3
TLE1
SIPA1L3
NEK11
NTN4
UTRN
SFPQ
WBSCR17
C8orf37-AS1
MED27
MAGI3
KCNB2
GPC6
LOC729911
LOC101927870

MAP4K1
CRYM
AKT3
LINC00521
INADL
FNDC3A
IL12A-AS1
NRXN3
TMPRSS2
TMPRSS2
FREM1
WNT3
LRRFIP1
ZNF638
YWHAEP7
TTC12
TTC12
SGMS1
RGS9
MSRA
BMPR1B
CECR1
NBAS
FNDC1
DNAH2
FNDC1
PRKCE
PRKCE
FNDC1
PAK6
DNAH11
NRXN1
C1orf110
HMCN2
DCC
RYR3
TVP23A
KCNB2
SORCS2
SNCA
SNCA
SNCA
XIRP2
NTM
DCC
GPR133
CACNA1S
CDH8
TET2
WDR27
MACROD2
KCNB2
CHAT
MAN2A1
DPP6
DPP6
RMND1
TRABD2B
LINC01257
MAN2A1
C22orf34
GSAP
ITPR1
MAP4K1
8-Mar
PTPN11
CASC15
TRIM67
SORL1
LINC01266
LINC01266
LINC01266
PPP2R2C
ATAD3C
ATAD3C
FLJ37505
LOC101927870
GPR158
THSD7B
ANKH
ANKH
PAN3
DUSP16
RFX3
PLEKHA5
WDR27
WDR27
WDR27
WDR27
CAMK1D
SS18L1

CCDC50
C1orf101
UTRN
SLC35F4
SORL1
LRGUK
VTI1A
TTC12
FNDC1
CASC2
MXRA7
AKAP8
YWHAEP7
SIPA1L3
RPS6KA2
EPN2
EYA2
EYA2
EYA2
KAZN
CECR2
PLCB4
INPP4B
LOC101928241
LINC00907
SLC26A5
ANKFY1
FAM19A5
NEK11
NEK11
NEK11
NEK11
RPS6KA2
SULT6B1
SORCS2
LHFPL3
GPR158
KCNK10
ERMARD
TENM2
EGLN1
ZNF71
PHAX
DUSP16
LEPREL1
MCEE
CDK14
DNAJC6
KHDRBS2
SH3GL3
TAMM41
KAZN
YTHDF3
AP1S3
CASC15
MLLT3
CNTN4
SEMA6D
YTHDF3
LOC339166
F5
LOC101928911
KCNK9
CD5L
GPM6A
PDE4B
MED27
SLC26A5
C22orf34
GABBR2
DIS3L2
AKT3
CWC22
MYO3B
AKT3
AKT3
AKT3
RRP7A
NALCN
UTRN
CIITA
SLCO1B1
MAN2A1
AKT3
FA2H
SGMS1
KCNB2
FARP1
PCDH7
OSMR
SORCS1

CAMK1D
RIN2
ZFR
COL1A2
FARP1
NTM
ACSF3
FNDC1
CAMK1D
TBC1D1
PAN3
PALD1
RIT2
CDH8
TSNAX-DISC1
MLIP
SGCZ
DCC
DCC
TAMM41
TAMM41
TAMM41
SLCO1B1
YTHDF3
LINC00607
MKL1
KCNK10
KCNK10
SYN3
F5
F11-AS1
LOC339166
FAM171A1
PCDH7
TCF25
PALD1
PHAX
ACER1
STPG2
ZNF346
EPHA4
PCDH7
NRG3
EFCAB4B
KCNH7
PAN3
ASIC2
PDE4B
PDE4B
CLYBL
ABCC6
IFT122
RANBP3L
ZFAND6
CSMD1
RORA
KCND3
PCAT2
F2
PPM1B
SNAP25-AS1
ACSF3
LOC102467213
OTOGL
LINC01266
LINC01266
NKAIN1
SORL1
TRIM67
TRIM67
GUCY2D
SORCS1
KLHL1
LINC01266
YTHDF3
AKT3
MYO3B
LOC100130264
SH3GL3
C1orf101
AKT3
KIAA1211
LINC00607
PPP2R2C
LINC01266
TTC12
TTC12
ANKH
ANKH
ANKH
ANKH

| |
|---|
| ANKH |
| XIRP2 |
| ZNF540 |
| C10orf11 |
| COL1A2 |
| LINC00907 |
| TENM4 |
| SNTB1 |
| SNTB1 |
| LATS2 |
| SIPA1L3 |
| KCNK10 |
| OSBPL3 |
| IGFBP6 |
| LGALS12 |
| TTC12 |
| TTC12 |
| PAN3 |
| PAN3 |
| PAN3 |
| F11-AS1 |
| DPM1 |
| SLC35F4 |
| SLC35F4 |
| XIRP2 |
| L3MBTL4 |
| LINC00478 |
| LRRFIP1 |
| KIAA0232 |
| MAGI1 |
| FLG-AS1 |
| DCC |
| CTNNA2 |
| STEAP2-AS1 |
| CASC15 |
| CASC15 |
| PTPRD |
| GAS7 |
| LOC101928241 |
| NEK11 |
| NEK11 |
| TMEM232 |
| MDH1B |
| ROCK1P1 |
| LPGAT1 |
| LPGAT1 |
| TENM2 |

**Table S14. The top 0.01% signals of geographically different Sinitic-speaking people**

| CPH | CHR | BP | PBS | Annotation | Han_Northwest | CHR | BP | PBS | Annotation |
|---|---|---|---|---|---|---|---|---|---|
| rs17822931 | 16 | 48258198 | 0.313225758 | ABCC11(0)\|=MISSENSE | rs114727280 | 13 | 20292578 | 0.795810502 | PSPC1(0) |
| rs1943674 | 18 | 46974430 | 0.308793989 | DYM(0) | rs17822931 | 16 | 48258198 | 0.447191063 | ABCC11(0)\|=MISSENSE |
| rs114727280 | 13 | 20292578 | 0.292987419 | PSPC1(0) | rs8082393 | 17 | 14432232 | 0.317491059 | . |
| rs72809096 | 5 | 174468624 | 0.191553293 | . | rs1943674 | 18 | 46974430 | 0.227507003 | DYM(0) |
| rs34907 | 3 | 10452231 | 0.19135373 | ATP2B2(0) | rs4482320 | 16 | 75552991 | 0.204993388 | . |
| rs28889583 | 3 | 136179532 | 0.181379353 | STAG1(0) | rs1329285 | 13 | 44170982 | 0.201539537 | ENOX1(0) |
| rs60614489 | 3 | 135986978 | 0.180002239 | PCCB(0) | rs9594979 | 13 | 44171495 | 0.199441725 | ENOX1(0) |
| rs35375192 | 3 | 136175525 | 0.173951303 | STAG1(0) | rs1410940 | 13 | 44182423 | 0.199424822 | ENOX1(0) |
| rs6439655 | 3 | 136466599 | 0.16995836 | STAG1(0) | rs9562500 | 13 | 44181748 | 0.198693341 | ENOX1(0) |
| rs4484375 | 4 | 143450615 | 0.158503478 | INPP4B(0) | rs12501116 | 4 | 188971489 | 0.198113693 | . |
| rs3092735 | 20 | 45419188 | 0.146854741 | . | rs1927464 | 10 | 94877164 | 0.195107051 | . |
| rs12461519 | 19 | 15523914 | 0.145192592 | AKAP8L(0) | rs1707489 | 12 | 54874839 | 0.194398444 | . |
| rs993568 | 1 | 97571799 | 0.143662559 | DPYD(0)\|DPYD-AS1(0) | rs730570 | 14 | 101142890 | 0.193454811 | . |
| rs6669773 | 1 | 31316495 | 0.141443328 | . | rs1586861 | 3 | 139058496 | 0.191748978 | . |
| rs6018037 | 20 | 45396978 | 0.140814926 | . | rs4918664 | 10 | 94921065 | 0.188631954 | . |
| rs7546194 | 1 | 81870764 | 0.139740017 | . | rs17108282 | 10 | 94971442 | 0.186873132 | . |
| rs291592 | 1 | 97543764 | 0.139090801 | DPYD(0) | rs2545656 | 5 | 103031154 | 0.18355126 | . |
| rs9878602 | 3 | 71535338 | 0.13781224 | FOXP1(0) | rs4484375 | 4 | 143450615 | 0.182939673 | INPP4B(0) |
| rs6771678 | 3 | 68283196 | 0.135097997 | FAM19A1(0) | rs61164110 | 16 | 75545479 | 0.178896085 | . |
| rs3092255 | 20 | 45400247 | 0.134227487 | . | rs6787776 | 3 | 171116588 | 0.178292762 | TNIK(0) |
| rs11639447 | 15 | 29625717 | 0.133549239 | FAM189A1(0) | rs3851741 | 16 | 75558483 | 0.17720488 | #NAME? |
| rs7770330 | 6 | 157279857 | 0.133072051 | ARID1B(0) | rs4859259 | 3 | 182664547 | 0.176815786 | DCUN1D1(0) |
| rs10092924 | 8 | 654796 | 0.131857024 | ERICH1(0) | rs4894599 | 3 | 171119365 | 0.173702618 | TNIK(0) |
| rs111221772 | 7 | 142169980 | 0.128516044 | . | rs2067928 | 2 | 112850581 | 0.173685949 | TMEM87B(0) |
| rs11211480 | 1 | 47693220 | 0.128442839 | TAL1(0) | rs7336364 | 13 | 44166351 | 0.173211633 | ENOX1(0) |
| rs12446160 | 16 | 48358368 | 0.12601016 | LONP2(0)\|MIR548AE2(0) | rs10910527 | 1 | 232599189 | 0.171038021 | SIPA1L2(0) |
| rs2152407 | 14 | 84102999 | 0.125991171 | . | rs6439655 | 3 | 136466599 | 0.170450593 | STAG1(0) |
| rs28377091 | 11 | 1691314 | 0.123646015 | . | rs10509567 | 10 | 90945756 | 0.168214777 | . |
| rs1927464 | 10 | 94877164 | 0.121036194 | . | rs357365 | 7 | 137976868 | 0.166360256 | . |
| rs11775576 | 8 | 137128340 | 0.119056871 | . | rs11224556 | 11 | 100899613 | 0.165444119 | . |
| rs751800 | 9 | 100884564 | 0.118367187 | CORO2A(0) | rs4754718 | 11 | 100893598 | 0.16274447 | . |
| rs10738433 | 9 | 16245170 | 0.117897363 | C9orf92(0) | rs56738524 | 8 | 8420053 | 0.160704013 | . |
| rs10110903 | 8 | 142882029 | 0.117690114 | . | rs2146674 | 13 | 43946257 | 0.159231356 | ENOX1(0) |
| rs6725244 | 2 | 233877065 | 0.116983024 | NGEF(0) | rs2296042 | 13 | 43987329 | 0.157376049 | ENOX1(0) |
| rs9813946 | 3 | 124850245 | 0.11668813 | SLC12A8(0) | rs7641582 | 3 | 182673355 | 0.157366066 | DCUN1D1(0) |
| rs3092206 | 20 | 45424218 | 0.116570237 | . | rs1432655 | 5 | 163028782 | 0.156882199 | . |
| rs7618867 | 3 | 196146192 | 0.115813532 | UBXN7(0) | rs62060554 | 16 | 75521189 | 0.156721765 | CHST6(0) |
| rs72785494 | 10 | 10565646 | 0.115395263 | . | rs72809096 | 5 | 174468624 | 0.155180094 | . |
| rs1132787 | 4 | 145030546 | 0.115260981 | GYPA(0) | rs943099 | 10 | 13727937 | 0.155085966 | FRMD4A(0) |
| rs1412695 | 9 | 3125843 | 0.114258411 | . | rs11100748 | 4 | 143460923 | 0.154702896 | INPP4B(0) |
| rs6432018 | 2 | 9721896 | 0.113798161 | . | rs12471210 | 2 | 3292186 | 0.153210543 | TSSC1(0) |
| rs1776570 | 4 | 37265814 | 0.113708081 | NWD2(0) | rs12638212 | 3 | 108159977 | 0.150044502 | MYH15(0)\|=MISSENSE |
| rs55905780 | 14 | 30579705 | 0.113601141 | . | rs191705 | 20 | 12912855 | 0.149773274 | LOC101929486(0) |
| rs2954668 | 8 | 146034379 | 0.112701265 | ZNF517(0) | rs7663766 | 4 | 153011210 | 0.149707107 | . |
| rs6734469 | 2 | 9735900 | 0.112020944 | YWHAQ(0) | rs12492883 | 3 | 139035356 | 0.14947028 | . |
| rs6074763 | 20 | 14568758 | 0.11190265 | MACROD2(0)\|MACROD2-IT1(0) | rs4998826 | 4 | 151392028 | 0.149194396 | LRBA(0) |
| rs7070947 | 10 | 94967076 | 0.111797942 | . | rs17059735 | 6 | 131190309 | 0.148793196 | EPB41L2(0) |
| rs8139798 | 22 | 37660255 | 0.110865047 | . | rs12435881 | 14 | 84125120 | 0.148513435 | . |
| rs11707053 | 3 | 139035269 | 0.109030886 | . | rs9934011 | 16 | 13915807 | 0.148059703 | . |
| rs10101336 | 8 | 72329553 | 0.108678263 | . | rs10412132 | 19 | 30914411 | 0.147842323 | ZNF536(0) |
| rs11707609 | 3 | 123503424 | 0.108527352 | MYLK(0) | rs7525615 | 1 | 229835413 | 0.146982653 | . |
| rs1579166 | 10 | 95535523 | 0.108334467 | LGI1(0) | rs4269843 | 10 | 90944216 | 0.14696585 | . |
| rs6718480 | 2 | 233879066 | 0.108331487 | . | rs8023044 | 14 | 84118602 | 0.146167917 | . |
| rs12370122 | 12 | 127108055 | 0.108169418 | . | rs60614489 | 3 | 135986978 | 0.146028513 | PCCB(0) |
| rs62032002 | 16 | 49088297 | 0.108103642 | . | rs2285750 | 17 | 42114672 | 0.145613835 | LSM12(0) |
| rs198683 | 6 | 143200547 | 0.107503257 | HIVEP2(0) | rs2687558 | 8 | 34711699 | 0.145492348 | . |
| rs34826583 | 5 | 109220429 | 0.107032141 | LOC100289673(0) | rs2183230 | 1 | 205324725 | 0.145173239 | KLHDC8A(0) |
| rs72708615 | 9 | 32138785 | 0.106745836 | . | rs575377 | 11 | 95942773 | 0.144646682 | MAML2(0) |
| rs4998826 | 4 | 151392028 | 0.106518205 | LRBA(0) | rs6081512 | 20 | 19133128 | 0.144601766 | . |
| rs6508312 | 18 | 21908633 | 0.105865772 | OSBPL1A(0) | rs11945180 | 4 | 101238386 | 0.144130998 | . |
| rs9402997 | 6 | 139058418 | 0.105860388 | LOC100507462(0) | rs1442514 | 9 | 17419319 | 0.144048293 | CNTLN(0) |
| rs7311096 | 12 | 100775956 | 0.105087739 | SLC17A8(0) | rs17579526 | 2 | 171241077 | 0.143972868 | MYO3B(0) |
| rs28669813 | 5 | 174430344 | 0.10497162 | . | rs2247493 | 9 | 130296484 | 0.143866222 | FAM129B(0) |
| rs7199599 | 16 | 20130754 | 0.104312291 | . | rs638920 | 10 | 11227341 | 0.143747236 | CELF2(0) |
| rs7306416 | 12 | 100391088 | 0.104306692 | . | rs644631 | 13 | 32098486 | 0.143515018 | . |
| rs10052184 | 5 | 175298874 | 0.104174085 | CPLX2(0) | rs28889583 | 3 | 136179532 | 0.142905529 | STAG1(0) |
| rs6446540 | 4 | 6759638 | 0.103712188 | . | rs716957 | 14 | 99476306 | 0.142705763 | . |
| rs17197552 | 3 | 135722264 | 0.103361084 | PPP2R3A(0)\|=MISSENSE | rs10072686 | 5 | 10308240 | 0.142378384 | . |
| rs292205 | 5 | 73507467 | 0.10322934 | . | rs4132003 | 6 | 98090084 | 0.141562243 | LOC101927314(0) |
| rs11668189 | 19 | 54754103 | 0.103203841 | . | rs7529591 | 1 | 29755064 | 0.141264656 | . |
| rs1361636 | 10 | 25717911 | 0.103124267 | GPR158(0) | rs4832482 | 2 | 17133401 | 0.141113178 | . |
| rs1979154 | 8 | 102067917 | 0.10266479 | FLJ42969(0) | rs34907 | 3 | 10452231 | 0.141099198 | ATP2B2(0) |
| rs12501116 | 4 | 188971489 | 0.102570569 | . | rs2280331 | 11 | 45923697 | 0.140910395 | MAPK8IP1(0) |
| rs9872542 | 3 | 135720673 | 0.102441924 | PPP2R3A(0)\|=missense | rs201405207 | 16 | 75652021 | 0.140776855 | ADAT1(0) |
| rs11103302 | 9 | 138958552 | 0.102233802 | NACC2(0) | rs4243474 | 6 | 145096340 | 0.14008969 | UTRN(0) |
| rs716957 | 14 | 99476306 | 0.101960598 | . | rs13249144 | 8 | 8394297 | 0.139968689 | . |
| rs34450379 | 19 | 54766055 | 0.10134713 | . | rs17730711 | 16 | 6769752 | 0.139624317 | RBFOX1(0) |
| rs9826146 | 3 | 135983089 | 0.101257979 | PCCB(0) | rs3828376 | 3 | 107900424 | 0.13885502 | IFT57(0) |
| rs1353663 | 4 | 82659400 | 0.101237783 | . | rs2095159 | 10 | 2197103 | 0.138519132 | . |
| rs231358 | 11 | 2702513 | 0.101152724 | KCNQ1(0)\|KCNQ1OT1(0) | rs11142207 | 9 | 90886728 | 0.138514213 | . |
| rs7663766 | 4 | 153011210 | 0.100866707 | . | rs1289764 | 3 | 107936558 | 0.138508134 | IFT57(0) |
| rs7779575 | 7 | 48269820 | 0.100705578 | ABCA13(0)\|=missense | rs12678192 | 8 | 127006346 | 0.138452426 | . |
| rs12629334 | 3 | 26106208 | 0.10001844 | . | rs10794822 | 10 | 2035815 | 0.138381844 | . |
| rs2256229 | 20 | 1517979 | 0.099997628 | SIRPD(0) | rs1061646 | 16 | 89805977 | 0.138072412 | FANCA(0)\|ZNF276(0)\|=missense |
| rs13434196 | 3 | 135643303 | 0.09958809 | . | rs35375192 | 3 | 136175525 | 0.138056002 | STAG1(0) |
| rs9819988 | 3 | 135624570 | 0.099496028 | #NAME? | rs7811918 | 7 | 7132043 | 0.137545223 | LOC100131257(0) |
| rs9579116 | 13 | 28421698 | 0.099164324 | PDX1-AS1(0) | rs4747895 | 10 | 11237509 | 0.136874296 | CELF2(0) |
| rs975136 | 4 | 143450575 | 0.0990002 | INPP4B(0) | rs542701 | 18 | 5982759 | 0.136821431 | L3MBTL4(0) |
| rs2229437 | 11 | 82564294 | 0.098951008 | PRCP(0) | rs1339970 | 6 | 97954709 | 0.136627115 | LOC101927314(0) |
| rs61556700 | 5 | 26647677 | 0.09882654 | . | rs1478892 | 8 | 11448529 | 0.136359093 | . |
| rs34027547 | 3 | 80419795 | 0.098684799 | . | rs4307107 | 5 | 162841338 | 0.136198039 | . |
| rs2183230 | 1 | 205324725 | 0.098558919 | KLHDC8A(0) | rs9320598 | 6 | 98019080 | 0.135899673 | LOC101927314(0) |
| rs344102 | 4 | 77721086 | 0.098281822 | . | rs198683 | 6 | 143200547 | 0.135807611 | HIVEP2(0) |
| rs4506826 | 14 | 84062865 | 0.097893027 | . | rs58273293 | 16 | 75628094 | 0.135626541 | . |
| rs4859259 | 3 | 182664547 | 0.097717549 | DCUN1D1(0) | rs11038706 | 11 | 45917703 | 0.135307771 | MAPK8IP1(0) |
| rs11121360 | 1 | 6220962 | 0.09764161 | CHD5(0) | rs939107 | 11 | 45982351 | 0.13514438 | PHF21A(0) |
| rs6700692 | 1 | 62283504 | 0.097355835 | INADL(0) | rs8094515 | 18 | 57133556 | 0.135031196 | CCBE1(0) |
| rs10135768 | 14 | 51671017 | 0.097270194 | . | rs12620194 | 2 | 17148004 | 0.134902296 | . |
| rs17615605 | 10 | 13723311 | 0.097000471 | FRMD4A(0) | rs10976178 | 9 | 7286273 | 0.134810094 | . |
| rs246491 | 5 | 150708723 | 0.096969269 | SLC36A2(0) | rs9789651 | 2 | 17137284 | 0.134796958 | . |
| rs383915 | 5 | 150695724 | 0.096952059 | SLC36A2(0) | rs7925824 | 11 | 45929961 | 0.134740084 | . |
| rs12471210 | 2 | 3292186 | 0.096848763 | TSSC1(0) | rs3924094 | 6 | 98068593 | 0.134654801 | LOC101927314(0) |
| rs62112421 | 18 | 74387761 | 0.096817791 | . | rs11666325 | 19 | 30907789 | 0.134563674 | ZNF536(0) |
| rs11165761 | 1 | 97488820 | 0.096669509 | . | rs3743599 | 16 | 75646576 | 0.134349419 | ADAT1(0)\|=MISSENSE |
| rs4589755 | 2 | 17191807 | 0.096368334 | . | rs4237080 | 8 | 34780509 | 0.134096981 | . |
| rs6747613 | 2 | 101180138 | 0.09628175 | PDCL3(0) | rs9796793 | 16 | 30495652 | 0.134070499 | ITGAL(0) |
| rs80266172 | 11 | 33718278 | 0.096082239 | . | rs4649389 | 1 | 232725391 | 0.133975435 | . |
| rs10064063 | 5 | 26617201 | 0.096077723 | . | rs8051241 | 16 | 75622993 | 0.133248565 | . |
| rs17730711 | 16 | 6769752 | 0.094094047 | RBFOX1(0) | rs7186971 | 16 | 75655406 | 0.132727273 | ADAT1(0) |
| rs17465171 | 22 | 27899464 | 0.093924628 | . | rs3736508 | 11 | 45975130 | 0.132656271 | PHF21A(0)\|=MISSENSE |
| rs10999372 | 10 | 72283134 | 0.093783949 | PALD1(0) | rs11775576 | 8 | 137128340 | 0.132598767 | . |
| rs9844093 | 3 | 1668078 | 0.093523283 | . | rs9489047 | 6 | 97964889 | 0.132585015 | LOC101927314(0) |
| rs7304531 | 12 | 68521687 | 0.093431438 | . | rs694421 | 19 | 34728620 | 0.132368855 | . |
| rs35739 | 12 | 100948515 | 0.092994411 | NR1H4(0) | rs6814775 | 4 | 143476181 | 0.132026915 | INPP4B(0) |
| rs7203498 | 16 | 20130965 | 0.092976855 | . | rs16854451 | 3 | 108039513 | 0.131712748 | HHLA2(0) |
| rs12458576 | 18 | 375021 | 0.092936246 | COLEC12(0) | rs72610763 | 2 | 17165426 | 0.131694194 | . |
| rs55774585 | 3 | 19422143 | 0.092883732 | KCNH8(0) | rs4718978 | 7 | 70126146 | 0.131193302 | AUTS2(0) |
| rs9525816 | 13 | 44223898 | 0.092825338 | ENOX1(0) | rs3169403 | 16 | 769247 | 0.130968475 | . |
| rs592328 | 11 | 30412174 | 0.092774983 | MPPED2(0) | rs978477 | 9 | 125308131 | 0.130901825 | #NAME? |
| rs10018031 | 4 | 133001527 | 0.092728952 | . | rs12025126 | 1 | 8759554 | 0.130561825 | RERE(0) |
| rs7811918 | 7 | 7132043 | 0.092659321 | LOC100131257(0) | rs229459 | 21 | 22048355 | 0.130038254 | . |
| rs9997579 | 4 | 188990434 | 0.092025914 | . | rs13394 | 19 | 49950298 | 0.129872741 | PIH1D1(0)\|=MISSENSE |
| rs2774245 | 9 | 93679125 | 0.091953464 | . | rs9878775 | 3 | 182659953 | 0.129835347 | . |
| rs221309 | 20 | 35274673 | 0.091823416 | . | rs9817112 | 3 | 27827733 | 0.129678415 | . |
| rs189637716 | 10 | 11916375 | 0.091708968 | PROSER2-AS1(0) | rs1879363 | 11 | 30195891 | 0.129675783 | . |
| rs9893497 | 17 | 56002077 | 0.091536979 | CUEDC1(0) | rs11600112 | 11 | 45931019 | 0.129299828 | . |
| rs2404583 | 1 | 46954554 | 0.091441074 | . | rs68107414 | 6 | 6954054 | 0.129241112 | . |
| rs77391939 | 1 | 202204357 | 0.091424222 | LGR6(0) | rs12590913 | 14 | 50241661 | 0.128796929 | KLHDC2(0) |
| rs10741741 | 11 | 18342081 | 0.091235272 | HPS5(0) | rs6558961 | 8 | 4916964 | 0.128749994 | . |
| rs231351 | 11 | 2711142 | 0.090992658 | KCNQ1(0)\|KCNQ1OT1(0) | rs3892909 | 5 | 175305591 | 0.128536584 | CPLX2(0) |
| rs12951391 | 17 | 69911582 | 0.090940612 | . | rs9882014 | 3 | 139176535 | 0.128348196 | RBP2(0) |
| rs1527452 | 2 | 155392642 | 0.090796831 | . | rs1458 | 19 | 34660146 | 0.12818066 | . |
| rs943099 | 10 | 13727937 | 0.090651618 | FRMD4A(0) | rs1575962 | 9 | 22282452 | 0.127448782 | . |
| rs7646563 | 3 | 136364485 | 0.090618548 | STAG1(0) | rs10056443 | 5 | 88752584 | 0.127251294 | . |
| rs56391641 | 10 | 94960095 | 0.090443526 | . | rs6771609 | 3 | 108056020 | 0.127075196 | HHLA2(0)\|=missense |
| rs17548919 | 4 | 133047434 | 0.090287147 | . | rs55774585 | 3 | 19422143 | 0.126837325 | KCNH8(0) |
| rs6685655 | 1 | 187677162 | 0.090043128 | . | rs7304531 | 12 | 68521687 | 0.126505153 | . |
| rs487336 | 11 | 30402105 | 0.090036605 | . | rs10260787 | 7 | 7129858 | 0.12646359 | LOC100131257(0)\|=missense |
| rs2287889 | 5 | 170178695 | 0.090017817 | . | rs7922269 | 10 | 90981887 | 0.126333377 | LIPA(0) |
| rs10512987 | 3 | 135823077 | 0.089961391 | PPP2R3A(0) | rs8035124 | 15 | 92105708 | 0.126274002 | . |
| rs10945563 | 6 | 159282859 | 0.089961338 | . | rs1981030 | 9 | 94217301 | 0.126151235 | . |
| rs9992794 | 4 | 133082999 | 0.089800813 | . | rs993568 | 1 | 97571799 | 0.125923061 | DPYD(0)\|DPYD-AS1(0) |
| rs10763269 | 10 | 57601709 | 0.089797575 | . | rs11053455 | 12 | 34826346 | 0.125748516 | . |
| rs2014131 | 4 | 155047531 | 0.089718585 | . | rs7196251 | 16 | 58410039 | 0.125679188 | . |
| rs2233381 | 2 | 233897101 | 0.089674229 | #NAME? | rs9997579 | 4 | 188990434 | 0.125209505 | . |
| rs794187 | 3 | 4436172 | 0.089616578 | SUMF1(0) | rs12581323 | 12 | 89848187 | 0.124849278 | POC1B(0) |
| rs10987911 | 9 | 130945298 | 0.089599333 | CIZ1(0) | rs9579116 | 13 | 28421698 | 0.124634544 | PDX1-AS1(0) |
| rs78572338 | 1 | 52851729 | 0.089428529 | ORC1(0) | rs3784576 | 15 | 45421662 | 0.124567298 | DUOXA1(0) |
| rs4551053 | 5 | 157838542 | 0.08940253 | . | rs16932667 | 12 | 5131553 | 0.124001294 | . |
| rs12459960 | 19 | 2510036 | 0.089395187 | . | rs6675857 | 1 | 8652154 | 0.123998943 | RERE(0) |
| rs589757 | 5 | 41572722 | 0.089377651 | . | rs291592 | 1 | 97543764 | 0.12394876 | DPYD(0) |
| rs10741742 | 11 | 18342160 | 0.089342735 | HPS5(0) | rs10092924 | 8 | 654796 | 0.123816611 | ERICH1(0) |
| rs1478892 | 8 | 11448529 | 0.089339784 | . | rs7618867 | 3 | 196146192 | 0.123758869 | UBXN7(0) |
| rs10281574 | 7 | 5483042 | 0.089277968 | . | rs36160296 | 2 | 14311647 | 0.123267645 | . |
| rs292726 | 11 | 37807834 | 0.089227414 | . | rs74550841 | 7 | 13842687 | 0.123266112 | . |
| rs11679606 | 2 | 16795418 | 0.089163622 | FAM49A(0) | rs1610247 | 3 | 104259387 | 0.123014922 | . |
| rs6723770 | 2 | 16795666 | 0.089163622 | FAM49A(0) | rs2913131 | 12 | 106596176 | 0.122987015 | . |
| rs7635628 | 3 | 74783321 | 0.088892266 | . | rs6074763 | 20 | 14568758 | 0.122907782 | MACROD2(0)\|MACROD2-IT1(0) |
| rs16843879 | 3 | 136003194 | 0.088880285 | PCCB(0) | rs4258426 | 12 | 53046630 | 0.122147804 | #NAME? |
| rs2555737 | 6 | 93208564 | 0.08885976 | . | rs1920542 | 3 | 107945810 | 0.121058261 | . |
| rs17468034 | 10 | 94965772 | 0.088842496 | . | rs10839825 | 11 | 7646734 | 0.121052202 | PPFIBP2(0) |
| rs2889678 | 20 | 31189993 | 0.088541356 | LOC149950(0) | rs2287889 | 5 | 170178695 | 0.12094269 | . |
| rs975664 | 2 | 75368797 | 0.088458267 | TACR1(0) | rs2578628 | 5 | 10242162 | 0.120827573 | FAM173B(0) |
| rs12106074 | 20 | 17356566 | 0.088331984 | PCSK2(0) | rs2979245 | 8 | 8939373 | 0.1206541 | . |
| rs757248 | 4 | 7703505 | 0.088236845 | SORCS2(0) | rs3776299 | 5 | 142507651 | 0.120639006 | ARHGAP26(0) |
| rs6073949 | 20 | 44531878 | 0.088112967 | PLTP(0) | rs7073711 | 10 | 10884232 | 0.120608341 | . |
| rs1522699 | 2 | 159764389 | 0.088093783 | . | rs10499012 | 6 | 97767591 | 0.120531811 | LOC101927314(0)\|MIR548H3(0) |
| rs687174 | 5 | 41564562 | 0.087906813 | . | rs9374686 | 6 | 98066596 | 0.120298474 | LOC101927314(0) |
| rs8468 | 15 | 63434110 | 0.087764309 | LACTB(0)\|=missense | rs9313968 | 5 | 162832105 | 0.120256595 | . |
| rs3112931 | 2 | 180427409 | 0.087275977 | ZNF385B(0) | rs2147065 | 9 | 9860780 | 0.120233894 | PTPRD(0) |
| rs7915994 | 10 | 95526168 | 0.087134428 | LGI1(0) | rs879067 | 1 | 244392816 | 0.119836134 | . |
| rs856614 | 1 | 52252006 | 0.086772114 | OSBPL9(0) | rs34510487 | 4 | 189149986 | 0.119793696 | . |
| rs1580678 | 11 | 97009206 | 0.086554779 | . | rs12461519 | 19 | 15523914 | 0.119776914 | AKAP8L(0) |
| rs6851301 | 4 | 140959594 | 0.08647562 | MAML3(0) | rs35325283 | 8 | 92265523 | 0.119721471 | SLC26A7(0) |
| rs4150579 | 11 | 18357180 | 0.086393353 | GTF2H1(0) | rs3846599 | 5 | 10255821 | 0.119386387 | CCT5(0) |
| rs10412132 | 19 | 30914411 | 0.086281907 | ZNF536(0) | rs1522699 | 2 | 159764389 | 0.119127886 | . |
| rs1476331 | 3 | 195959924 | 0.0862772 | SLC51A(0) | rs2964798 | 5 | 10788833 | 0.119023991 | . |
| rs930981 | 11 | 122834265 | 0.086137936 | . | rs592565 | 11 | 119939927 | 0.118853425 | . |
| rs6708276 | 2 | 144442150 | 0.086063532 | ARHGAP15(0) | rs10738433 | 9 | 16245170 | 0.118436948 | C9orf92(0) |
| rs3740711 | 11 | 18379839 | 0.086058659 | GTF2H1(0) | rs4324324 | 2 | 17140637 | 0.118370402 | . |
| rs4150650 | 11 | 18374279 | 0.086058659 | GTF2H1(0) | rs6851797 | 4 | 140942859 | 0.118291041 | MAML3(0) |
| rs4150651 | 11 | 18374425 | 0.086058659 | GTF2H1(0) | rs9426354 | 1 | 29763684 | 0.118246851 | . |
| rs4150673 | 11 | 18383449 | 0.086058659 | GTF2H1(0) | rs2131679 | 1 | 8811768 | 0.118223426 | RERE(0) |
| rs3851518 | 6 | 6336584 | 0.086016572 | . | rs9839541 | 3 | 28650238 | 0.118202253 | LINC00693(0) |
| rs12730665 | 1 | 96683642 | 0.085945583 | . | rs2243955 | 9 | 130303640 | 0.118176549 | FAM129B(0) |
| rs17656368 | 3 | 71474216 | 0.085783979 | FOXP1(0) | rs10758256 | 9 | 3446277 | 0.118026851 | RFX3(0) |
| rs10495563 | 2 | 9662210 | 0.08559147 | ADAM17(0) | rs77739786 | 8 | 34778489 | 0.117981154 | . |
| rs221310 | 20 | 35272778 | 0.08551986 | SLA2(0) | rs9496708 | 6 | 143936264 | 0.11795969 | PHACTR2(0) |
| rs4150641 | 11 | 18370739 | 0.085500981 | GTF2H1(0) | rs6701064 | 1 | 202093735 | 0.117683012 | GPR37L1(0) |
| rs4150615 | 11 | 18365028 | 0.085341186 | GTF2H1(0) | rs6851301 | 4 | 140959594 | 0.117540007 | MAML3(0) |
| rs2116980 | 9 | 23503695 | 0.08533888 | . | rs28693601 | 15 | 92115966 | 0.116681726 | . |
| rs4150612 | 11 | 18364358 | 0.085332474 | GTF2H1(0) | rs16925531 | 11 | 30167146 | 0.116580144 | . |
| rs4150661 | 11 | 18379629 | 0.085332474 | GTF2H1(0) | rs6718480 | 2 | 233879066 | 0.116475772 | . |
| rs17027780 | 4 | 152590337 | 0.085301439 | #NAME? | rs10281574 | 7 | 5483042 | 0.116402214 | . |
| rs1339822 | 10 | 94915615 | 0.085112947 | . | rs9895062 | 17 | 9370623 | 0.116255071 | STX8(0) |
| rs529177 | 2 | 45100208 | 0.085101078 | . | rs2578624 | 5 | 10245114 | 0.116170536 | FAM173B(0) |
| rs74550841 | 7 | 13842687 | 0.084948559 | . | rs2371401 | 2 | 34803638 | 0.115890937 | . |
| rs4150562 | 11 | 18354995 | 0.084936611 | GTF2H1(0) | rs7954506 | 12 | 99175840 | 0.115672211 | ANKS1B(0) |
| rs2979725 | 8 | 25352468 | 0.08490791 | CDCA2(0) | rs1433071 | 19 | 30913098 | 0.115553366 | ZNF536(0) |
| rs7027419 | 9 | 107485577 | 0.084822004 | #NAME? | rs1412695 | 9 | 3125843 | 0.115328781 | . |
| rs7954506 | 12 | 99175840 | 0.084791024 | ANKS1B(0) | rs1759092 | 19 | 34676538 | 0.115103584 | LSM14A(0) |
| rs12541800 | 8 | 11423072 | 0.084610051 | . | rs75702132 | 3 | 108367099 | 0.11509418 | DZIP3(0) |
| rs2585390 | 14 | 49268677 | 0.084595601 | . | rs2890503 | 9 | 3128693 | 0.11505865 | . |
| rs2837957 | 21 | 42491120 | 0.084594652 | . | rs61744949 | 19 | 57967049 | 0.115003992 | VN1R1(0) |
| rs4843708 | 16 | 87857939 | 0.084527887 | . | rs1776570 | 4 | 37265814 | 0.114938718 | NWD2(0) |
| rs6761396 | 2 | 104113458 | 0.084367847 | . | rs1979154 | 8 | 102067917 | 0.114881891 | FLJ42969(0) |
| rs11617361 | 13 | 101758117 | 0.084304353 | NALCN(0) | rs2243191 | 1 | 207015957 | 0.114503281 | IL19(0)\|=MISSENSE |
| rs4150575 | 11 | 18356466 | 0.084208744 | GTF2H1(0) | rs10167958 | 2 | 109921632 | 0.114214778 | SH3RF3(0) |
| rs9833156 | 3 | 34198737 | 0.08420726 | . | rs111221772 | 7 | 142169980 | 0.113775791 | . |
| rs3749427 | 1 | 247150740 | 0.084198385 | ZNF670-ZNF695(0)\|ZNF695(0)\|=missense | rs10135768 | 14 | 51671017 | 0.113677869 | . |
| rs2093830 | 9 | 12756889 | 0.084066027 | . | rs10115104 | 9 | 71540376 | 0.113675442 | PIP5K1B(0) |
| rs191705 | 20 | 12912855 | 0.084058409 | LOC101929486(0) | rs11808688 | 1 | 203689314 | 0.11364944 | ATP2B4(0) |
| rs6673823 | 1 | 234668280 | 0.083996671 | . | rs4855697 | 3 | 108867128 | 0.113644912 | FLJ22763(0) |
| rs12276244 | 11 | 123957157 | 0.083830585 | . | rs28463470 | 1 | 234691034 | 0.113481885 | . |
| rs290861 | 1 | 97542797 | 0.083792818 | . | rs1476331 | 3 | 195959924 | 0.113246568 | SLC51A(0) |
| rs1476873 | 2 | 37949251 | 0.083724498 | . | rs6057602 | 20 | 31181808 | 0.1128905 | LOC149950(0) |
| rs10507397 | 13 | 31715015 | 0.083695759 | HSPH1(0) | rs11626551 | 14 | 90027123 | 0.112764458 | FOXN3(0) |
| rs6540602 | 1 | 210696806 | 0.083673372 | HHAT(0) | rs3803167 | 12 | 111785586 | 0.112379844 | CUX2(0)\|=MISSENSE |
| rs7267720 | 20 | 31166190 | 0.083655499 | NOL4L(0) | rs10814535 | 9 | 37418935 | 0.112359345 | . |
| rs62014517 | 15 | 30233518 | 0.083577643 | . | rs1343367 | 10 | 91700359 | 0.111883146 | LOC101926924(0) |
| rs67211505 | 12 | 125780255 | 0.083571207 | . | rs2293012 | 19 | 49954807 | 0.111643319 | PIH1D1(0) |
| rs9538601 | 13 | 31719128 | 0.083551817 | HSPH1(0) | rs11639447 | 15 | 29625717 | 0.111562391 | FAM189A1(0) |
| rs2106735 | 7 | 93070811 | 0.083148131 | CALCR(0) | rs6432018 | 2 | 9721896 | 0.111433173 | . |
| rs12224084 | 11 | 1639388 | 0.082970303 | . | rs970624 | 3 | 28682296 | 0.111379331 | LINC00693(0) |
| rs28113 | 3 | 10400643 | 0.0829518 | ATP2B2(0) | rs4387021 | 8 | 72726695 | 0.111057936 | . |
| rs876305 | 7 | 24758795 | 0.082888089 | DFNA5(0) | rs911478 | 6 | 109262787 | 0.111042624 | ARMC2(0) |
| rs1432655 | 5 | 163028782 | 0.082887716 | . | rs1325140 | 9 | 12544593 | 0.110990095 | . |
| rs6057599 | 20 | 31168439 | 0.08285499 | NOL4L(0) | rs12513800 | 5 | 53966360 | 0.110914064 | LOC102467080(0) |
| rs231675 | 17 | 3190599 | 0.082816002 | #NAME? | rs6673823 | 1 | 234668280 | 0.110857484 | . |
| rs8103067 | 19 | 32027415 | 0.082609438 | . | rs10492964 | 1 | 8629380 | 0.11066263 | RERE(0) |
| rs17296815 | 12 | 51670202 | 0.082593854 | . | rs73202902 | 3 | 191309675 | 0.110541825 | . |
| rs2499502 | 6 | 88661167 | 0.082588061 | . | rs1355696 | 12 | 17424512 | 0.110311376 | . |
| rs1156992 | 3 | 26028747 | 0.082417277 | . | rs11663375 | 18 | 21058082 | 0.110163259 | RIOK3(0) |

| Han_Southeast | CHR | BP | PBS | Annotation | Han_Southwest | CHR | BP | PBS | Annotation |
|---|---|---|---|---|---|---|---|---|---|
| rs8082393 | 17 | 14432232 | 0.546701189 | . | rs17822931 | 16 | 48258198 | 0.313225758 | ABCC11(0)\|=MISSENSE |
| rs114727280 | 13 | 20292578 | 0.395819272 | PSPC1(0) | rs1943674 | 18 | 46974430 | 0.308793989 | DYM(0) |
| rs10018155 | 4 | 92664886 | 0.33925618 | . | rs114727280 | 13 | 20292578 | 0.292987419 | PSPC1(0) |
| rs1575962 | 9 | 22282452 | 0.281989942 | . | rs72809096 | 5 | 174468624 | 0.191553293 | . |
| rs12620194 | 2 | 17148004 | 0.255757784 | . | rs34907 | 3 | 10452231 | 0.19135373 | ATP2B2(0) |
| rs4832482 | 2 | 17133401 | 0.25518151 | . | rs28889583 | 3 | 136179532 | 0.181379353 | STAG1(0) |
| rs4324324 | 2 | 17140637 | 0.247529681 | . | rs60614489 | 3 | 135986978 | 0.180002239 | PCCB(0) |
| rs2301723 | 12 | 112843363 | 0.247025868 | RPL6(0) | rs35375192 | 3 | 136175525 | 0.173951303 | STAG1(0) |
| rs9789651 | 2 | 17137284 | 0.246721438 | . | rs6439655 | 3 | 136466599 | 0.16995836 | STAG1(0) |
| rs72610763 | 2 | 17165426 | 0.238885257 | . | rs4484375 | 4 | 143450615 | 0.158503478 | INPP4B(0) |
| rs1586861 | 3 | 139058496 | 0.236034743 | . | rs3092735 | 20 | 45419188 | 0.146854741 | . |
| rs1943674 | 18 | 46974430 | 0.226152838 | DYM(0) | rs12461519 | 19 | 15523914 | 0.145192592 | AKAP8L(0) |
| rs11066334 | 12 | 112977311 | 0.22602916 | . | rs993568 | 1 | 97571799 | 0.143662559 | DPYD(0)\|DPYD-AS1(0) |
| rs10850061 | 12 | 113016553 | 0.225160794 | . | rs6669773 | 1 | 31316495 | 0.141443328 | . |
| rs716957 | 14 | 99476306 | 0.225003983 | . | rs6018037 | 20 | 45396978 | 0.140814926 | . |
| rs10794822 | 10 | 2035815 | 0.199222032 | . | rs7546194 | 1 | 81870764 | 0.139740017 | . |
| rs4482320 | 16 | 75552991 | 0.19708932 | . | rs291592 | 1 | 97543764 | 0.139090801 | DPYD(0) |
| rs16932667 | 12 | 5131553 | 0.195580225 | . | rs9878602 | 3 | 71535338 | 0.13781224 | FOXP1(0) |
| rs3092735 | 20 | 45419188 | 0.191325749 | . | rs6771678 | 3 | 68283196 | 0.135097997 | FAM19A1(0) |
| rs4664513 | 2 | 152833825 | 0.190680886 | CACNB4(0) | rs3092255 | 20 | 45400247 | 0.134227487 | . |
| rs2287889 | 5 | 170178695 | 0.185136868 | . | rs11639447 | 15 | 29625717 | 0.133549239 | FAM189A1(0) |
| rs6018037 | 20 | 45396978 | 0.184948026 | . | rs7770330 | 6 | 157279857 | 0.133072051 | ARID1B(0) |
| rs9795633 | 12 | 112976957 | 0.18462883 | . | rs10092924 | 8 | 654796 | 0.131857024 | ERICH1(0) |
| rs34907 | 3 | 10452231 | 0.184296934 | ATP2B2(0) | rs111221772 | 7 | 142169980 | 0.128516044 | . |
| rs7246968 | 19 | 12651854 | 0.18363484 | ZNF564(0) | rs11211480 | 1 | 47693220 | 0.128442839 | TAL1(0) |
| rs55774585 | 3 | 19422143 | 0.179601132 | KCNH8(0) | rs12446160 | 16 | 48358368 | 0.12601016 | LONP2(0)\|MIR548AE2(0) |
| rs10509567 | 10 | 90945756 | 0.179140906 | . | rs2152407 | 14 | 84102999 | 0.125991171 | . |
| rs6081512 | 20 | 19133128 | 0.177392855 | . | rs28377091 | 11 | 1691314 | 0.123646015 | . |
| rs11224556 | 11 | 100899613 | 0.177310185 | . | rs1927464 | 10 | 94877164 | 0.121036194 | . |
| rs7618867 | 3 | 196146192 | 0.17728737 | UBXN7(0) | rs11775576 | 8 | 137128340 | 0.119056871 | . |
| rs3851741 | 16 | 75558483 | 0.177209097 | #NAME? | rs751800 | 9 | 100884564 | 0.118367187 | CORO2A(0) |
| rs4754718 | 11 | 100893598 | 0.176011037 | . | rs10738433 | 9 | 16245170 | 0.117897363 | C9orf92(0) |
| rs61164110 | 16 | 75545479 | 0.175141128 | . | rs10110903 | 8 | 142882029 | 0.117690114 | . |
| rs10738433 | 9 | 16245170 | 0.174978673 | C9orf92(0) | rs6725244 | 2 | 233877065 | 0.116983024 | NGEF(0) |
| rs6748530 | 2 | 152790922 | 0.173994055 | CACNB4(0) | rs9813946 | 3 | 124850245 | 0.11668813 | SLC12A8(0) |
| rs8086803 | 18 | 41839227 | 0.173365273 | . | rs3092206 | 20 | 45424218 | 0.116570237 | . |
| rs3092255 | 20 | 45400247 | 0.173183726 | . | rs7618867 | 3 | 196146192 | 0.115813532 | UBXN7(0) |
| rs10044956 | 5 | 145834529 | 0.171750307 | TCERG1(0)\|=missense | rs72785494 | 10 | 10565646 | 0.115395263 | . |
| rs1370979 | 5 | 10832150 | 0.170680818 | . | rs1132787 | 4 | 145030546 | 0.115260981 | GYPA(0) |
| rs4129493 | 18 | 38139080 | 0.170492206 | . | rs1412695 | 9 | 3125843 | 0.114258411 | . |
| rs17059735 | 6 | 131190309 | 0.169669142 | EPB41L2(0) | rs6432018 | 2 | 9721896 | 0.113798161 | . |
| rs72809096 | 5 | 174468624 | 0.169626865 | . | rs1776570 | 4 | 37265814 | 0.113708081 | NWD2(0) |
| rs7617596 | 3 | 71521494 | 0.167212339 | FOXP1(0) | rs55905780 | 14 | 30579705 | 0.113601141 | . |
| rs1707489 | 12 | 54874839 | 0.16708536 | . | rs2954668 | 8 | 146034379 | 0.112701265 | ZNF517(0) |
| rs3803167 | 12 | 111785586 | 0.166340137 | CUX2(0)\|=MISSENSE | rs6734469 | 2 | 9735900 | 0.112020944 | YWHAQ(0) |
| rs2585390 | 14 | 49268677 | 0.162926721 | . | rs6074763 | 20 | 14568758 | 0.11190265 | MACROD2(0)\|MACROD2-IT1(0) |
| rs768232 | 5 | 145823141 | 0.162653684 | #NAME? | rs7070947 | 10 | 94967076 | 0.111797942 | . |
| rs6787776 | 3 | 171116588 | 0.162273928 | TNIK(0) | rs8139798 | 22 | 37660255 | 0.110865047 | . |
| rs4439672 | 14 | 49288860 | 0.162182517 | . | rs11707053 | 3 | 139035269 | 0.109030886 | . |
| rs7604835 | 2 | 152881908 | 0.161566272 | CACNB4(0) | rs10101336 | 8 | 72329553 | 0.108678263 | . |
| rs4894599 | 3 | 171119365 | 0.161295854 | TNIK(0) | rs11707609 | 3 | 123503424 | 0.108527352 | MYLK(0) |
| rs4269843 | 10 | 90944216 | 0.157288249 | . | rs1579166 | 10 | 95535523 | 0.108334467 | LGI1(0) |
| rs939107 | 11 | 45982351 | 0.157278806 | PHF21A(0) | rs6718480 | 2 | 233879066 | 0.108331487 | . |
| rs8035124 | 15 | 92105708 | 0.156769343 | . | rs12370122 | 12 | 127108055 | 0.108169418 | . |
| rs9917191 | 2 | 1719083 | 0.156359465 | PXDN(0) | rs62032002 | 16 | 49088297 | 0.108103642 | . |
| rs6439655 | 3 | 136466599 | 0.156252604 | STAG1(0) | rs198683 | 6 | 143200547 | 0.107503257 | HIVEP2(0) |
| rs1820556 | 2 | 126290376 | 0.156243941 | . | rs34826583 | 5 | 109220429 | 0.107032141 | LOC100289673(0) |
| rs11066079 | 12 | 112350403 | 0.155721487 | . | rs72708615 | 9 | 32138785 | 0.106745836 | . |
| rs60614489 | 3 | 135986978 | 0.155635857 | PCCB(0) | rs4998826 | 4 | 151392028 | 0.106518205 | LRBA(0) |
| rs2371401 | 2 | 34803638 | 0.153841236 | . | rs6508312 | 18 | 21908633 | 0.105865772 | OSBPL1A(0) |
| rs2607642 | 13 | 102816760 | 0.153697359 | FGF14(0) | rs9402997 | 6 | 139058418 | 0.105860388 | LOC100507462(0) |
| rs77717927 | 2 | 126270880 | 0.15312148 | . | rs7311096 | 12 | 100775956 | 0.105087739 | SLC17A8(0) |
| rs10270171 | 7 | 22813879 | 0.152548773 | . | rs28669813 | 5 | 174430344 | 0.10497162 | . |
| rs28889583 | 3 | 136179532 | 0.152250561 | STAG1(0) | rs7199599 | 16 | 20130754 | 0.104312291 | . |
| rs9882014 | 3 | 139176535 | 0.152185066 | RBP2(0) | rs7306416 | 12 | 100391088 | 0.104306692 | . |
| rs1325140 | 9 | 12544593 | 0.151920253 | . | rs10052184 | 5 | 175298874 | 0.104174085 | CPLX2(0) |
| rs2003155 | 16 | 49690136 | 0.1517589 | ZNF423(0) | rs6446540 | 4 | 6759638 | 0.103712188 | . |
| rs4405491 | 15 | 52515523 | 0.151571606 | MYO5C(0) | rs17197552 | 3 | 135722264 | 0.103361084 | PPP2R3A(0)\|=MISSENSE |
| rs16983067 | 2 | 17182314 | 0.151356837 | . | rs292205 | 5 | 73507467 | 0.10322934 | . |
| rs4143812 | 2 | 17181277 | 0.151293047 | . | rs11668189 | 19 | 54754103 | 0.103203841 | . |
| rs4998826 | 4 | 151392028 | 0.150580089 | LRBA(0) | rs1361636 | 10 | 25717911 | 0.103124267 | GPR158(0) |
| rs11038706 | 11 | 45917703 | 0.15028431 | MAPK8IP1(0) | rs1979154 | 8 | 102067917 | 0.10266479 | FLJ42969(0) |
| rs6564869 | 16 | 81372725 | 0.149681001 | GAN(0) | rs12501116 | 4 | 188971489 | 0.102570569 | . |
| rs12501116 | 4 | 188971489 | 0.149125779 | . | rs9872542 | 3 | 135720673 | 0.102441924 | PPP2R3A(0)\|=missense |
| rs4940842 | 18 | 56802550 | 0.148623763 | . | rs11103302 | 9 | 138958552 | 0.102233802 | NACC2(0) |
| rs11775576 | 8 | 137128340 | 0.148509393 | . | rs716957 | 14 | 99476306 | 0.101960598 | . |
| rs7114162 | 11 | 45916781 | 0.148259062 | MAPK8IP1(0) | rs34450379 | 19 | 54766055 | 0.10134713 | . |
| rs4804203 | 19 | 12723146 | 0.147245702 | ZNF791(0) | rs9826146 | 3 | 135983089 | 0.101257979 | PCCB(0) |
| rs35375192 | 3 | 136175525 | 0.147126603 | STAG1(0) | rs1353663 | 4 | 82659400 | 0.101237783 | . |
| rs6675857 | 1 | 8652154 | 0.146676994 | RERE(0) | rs231358 | 11 | 2702513 | 0.101152724 | KCNQ1(0)\|KCNQ1OT1(0) |
| rs522311 | 18 | 571927 | 0.146343253 | . | rs7663766 | 4 | 153011210 | 0.100866707 | . |
| rs7545384 | 1 | 8580960 | 0.146310655 | RERE(0) | rs7779575 | 7 | 48269820 | 0.100705578 | ABCA13(0)\|=missense |
| rs7247513 | 19 | 12691185 | 0.146156099 | ZNF490(0) | rs12629334 | 3 | 26106208 | 0.10001844 | . |
| rs11930453 | 4 | 141710146 | 0.145804225 | . | rs2256229 | 20 | 1517979 | 0.099997628 | SIRPD(0) |
| rs2146674 | 13 | 43946257 | 0.144865646 | ENOX1(0) | rs13434196 | 3 | 135643303 | 0.09958809 | . |
| rs6814775 | 4 | 143476181 | 0.144799577 | INPP4B(0) | rs9819988 | 3 | 135624570 | 0.099496028 | #NAME? |
| rs4772114 | 13 | 99298233 | 0.144763723 | . | rs9579116 | 13 | 28421698 | 0.099164324 | PDX1-AS1(0) |
| rs1432655 | 5 | 163028782 | 0.14446712 | . | rs975136 | 4 | 143450575 | 0.0990002 | INPP4B(0) |
| rs1836858 | 2 | 126159556 | 0.143021275 | . | rs2229437 | 11 | 82564294 | 0.098951008 | PRCP(0) |
| rs7951628 | 11 | 120161117 | 0.142736006 | POU2F3(0) | rs61556700 | 5 | 26647677 | 0.09882654 | . |
| rs977747 | 1 | 47684677 | 0.142484884 | TAL1(0) | rs34027547 | 3 | 80419795 | 0.098684799 | . |
| rs16983064 | 2 | 17181880 | 0.142456943 | . | rs2183230 | 1 | 205324725 | 0.098558919 | KLHDC8A(0) |
| rs2833653 | 21 | 33471519 | 0.142222194 | . | rs344102 | 4 | 77721086 | 0.098281822 | . |
| rs12606223 | 18 | 77813395 | 0.141626254 | . | rs4506826 | 14 | 84062865 | 0.097893027 | . |
| rs1035055 | 2 | 160057141 | 0.141582788 | TANC1(0) | rs4859259 | 3 | 182664547 | 0.097717549 | DCUN1D1(0) |
| rs9820131 | 3 | 138608009 | 0.141577627 | . | rs11121360 | 1 | 6220962 | 0.09764161 | CHD5(0) |
| rs2274190 | 13 | 31797455 | 0.141266751 | B3GALTL(0) | rs6700692 | 1 | 62283504 | 0.097355835 | INADL(0) |
| rs56156709 | 13 | 21285968 | 0.14120102 | IL17D(0) | rs10135768 | 14 | 51671017 | 0.097270194 | . |
| rs4918664 | 10 | 94921065 | 0.140850815 | . | rs17615605 | 10 | 13723311 | 0.097000471 | FRMD4A(0) |
| rs13117732 | 4 | 177496193 | 0.140710679 | . | rs246491 | 5 | 150708723 | 0.096969269 | SLC36A2(0) |
| rs2991953 | 1 | 234475832 | 0.140572264 | . | rs383915 | 5 | 150695724 | 0.096952059 | SLC36A2(0) |
| rs4484375 | 4 | 143450615 | 0.140440563 | INPP4B(0) | rs12471210 | 2 | 3292186 | 0.096848763 | TSSC1(0) |
| rs8011267 | 14 | 49294826 | 0.139985606 | . | rs62112421 | 18 | 74387761 | 0.096817791 | . |
| rs10138228 | 14 | 49778543 | 0.13992957 | . | rs11165761 | 1 | 97488820 | 0.096669509 | . |
| rs12083555 | 1 | 47703400 | 0.139730588 | . | rs4589755 | 2 | 17191807 | 0.096368334 | . |
| rs4505038 | 11 | 45934697 | 0.139699178 | PEX16(0) | rs6747613 | 2 | 101180138 | 0.09628175 | PDCL3(0) |
| rs8112261 | 19 | 12684559 | 0.139683703 | . | rs80266172 | 11 | 33718278 | 0.096082239 | . |
| rs10492964 | 1 | 8629380 | 0.139520336 | RERE(0) | rs10064063 | 5 | 26617201 | 0.096077723 | . |
| rs4767364 | 12 | 112521448 | 0.139126058 | NAA25(0)\|=missense | rs17730711 | 16 | 6769752 | 0.094094047 | RBFOX1(0) |
| rs7953867 | 12 | 51481215 | 0.139030306 | #NAME? | rs17465171 | 22 | 27899464 | 0.093924628 | . |
| rs2280331 | 11 | 45923697 | 0.13897505 | MAPK8IP1(0) | rs10999372 | 10 | 72283134 | 0.093783949 | PALD1(0) |
| rs7922269 | 10 | 90981887 | 0.138890244 | LIPA(0) | rs9844093 | 3 | 1668078 | 0.093523283 | . |
| rs198683 | 6 | 143200547 | 0.138821232 | HIVEP2(0) | rs7304531 | 12 | 68521687 | 0.093431438 | . |
| rs255332 | 5 | 94150212 | 0.138404015 | MCTP1(0) | rs35739 | 12 | 100948515 | 0.092994411 | NR1H4(0) |
| rs6761396 | 2 | 104113458 | 0.138283479 | . | rs7203498 | 16 | 20130965 | 0.092976855 | . |
| rs2479328 | 9 | 261855 | 0.138134632 | DOCK8(0) | rs12458576 | 18 | 375021 | 0.092936246 | COLEC12(0) |
| rs9556998 | 13 | 99303782 | 0.138108813 | . | rs55774585 | 3 | 19422143 | 0.092883732 | KCNH8(0) |
| rs11100748 | 4 | 143460923 | 0.138021737 | INPP4B(0) | rs9525816 | 13 | 44223898 | 0.092825338 | ENOX1(0) |
| rs10976178 | 9 | 7286273 | 0.137427994 | . | rs592328 | 11 | 30412174 | 0.092774983 | MPPED2(0) |
| rs3087801 | 19 | 12744132 | 0.137413659 | . | rs10018031 | 4 | 133001527 | 0.092728952 | . |
| rs11729469 | 4 | 188959641 | 0.136630673 | . | rs7811918 | 7 | 7132043 | 0.092659321 | LOC100131257(0) |
| rs2131679 | 1 | 8811768 | 0.136475812 | RERE(0) | rs9997579 | 4 | 188990434 | 0.092025914 | . |
| rs7982453 | 13 | 41058246 | 0.136423262 | . | rs2774245 | 9 | 93679125 | 0.091953464 | . |
| rs9572141 | 13 | 31809581 | 0.136351639 | B3GALTL(0) | rs221309 | 20 | 35274673 | 0.091823416 | . |
| rs7663766 | 4 | 153011210 | 0.136047844 | . | rs189637716 | 10 | 11916375 | 0.091708968 | PROSER2-AS1(0) |
| rs58273293 | 16 | 75628094 | 0.135910124 | . | rs9893497 | 17 | 56002077 | 0.091536979 | CUEDC1(0) |
| rs575377 | 11 | 95942773 | 0.1358702 | MAML2(0) | rs2404583 | 1 | 46954554 | 0.091441074 | . |
| rs35035095 | 13 | 39376300 | 0.135551329 | FREM2(0) | rs77391939 | 1 | 202204357 | 0.091424222 | LGR6(0) |
| rs35325283 | 8 | 92265523 | 0.135380532 | SLC26A7(0) | rs10741741 | 11 | 18342081 | 0.091235272 | HPS5(0) |
| rs12654468 | 5 | 113653675 | 0.135099516 | . | rs231351 | 11 | 2711142 | 0.090992658 | KCNQ1(0)\|KCNQ1OT1(0) |
| rs10971220 | 9 | 32928215 | 0.135067459 | . | rs12951391 | 17 | 69911582 | 0.090940612 | . |
| rs6747613 | 2 | 101180138 | 0.135058595 | PDCL3(0) | rs1527452 | 2 | 155392642 | 0.090796831 | . |
| rs6577710 | 8 | 137141233 | 0.134810288 | . | rs943099 | 10 | 13727937 | 0.090651618 | FRMD4A(0) |
| rs245974 | 7 | 29295856 | 0.134671281 | CHN2(0) | rs7646563 | 3 | 136364485 | 0.090618548 | STAG1(0) |
| rs1343367 | 10 | 91700359 | 0.134586529 | LOC101926924(0) | rs56391641 | 10 | 94960095 | 0.090443526 | . |
| rs6745226 | 2 | 26493597 | 0.134477928 | HADHB(0)\|=missense | rs17548919 | 4 | 133047434 | 0.090287147 | . |
| rs6073949 | 20 | 44531878 | 0.134466555 | PLTP(0) | rs6685655 | 1 | 187677162 | 0.090043128 | . |
| rs264963 | 2 | 104254564 | 0.134171806 | . | rs487336 | 11 | 30402105 | 0.090036605 | . |
| rs61731670 | 15 | 52901283 | 0.134150435 | FAM214A(0) | rs2287889 | 5 | 170178695 | 0.090017817 | . |
| rs271828 | 19 | 18342477 | 0.133996309 | PDE4C(0)\|=missense | rs10512987 | 3 | 135823077 | 0.089961391 | PPP2R3A(0) |
| rs10985690 | 9 | 125316850 | 0.133866306 | . | rs10945563 | 6 | 159282859 | 0.089961338 | . |
| rs526509 | 9 | 268740 | 0.133842364 | DOCK8(0) | rs9992794 | 4 | 133082999 | 0.089800813 | . |
| rs3092206 | 20 | 45424218 | 0.133557949 | . | rs10763269 | 10 | 57601709 | 0.089797575 | . |
| rs201405207 | 16 | 75652021 | 0.13340717 | ADAT1(0) | rs2014131 | 4 | 155047531 | 0.089718585 | . |
| rs11743333 | 5 | 145838440 | 0.133328413 | TCERG1(0) | rs2233381 | 2 | 233897101 | 0.089674229 | #NAME? |
| rs12025126 | 1 | 8759554 | 0.133119032 | RERE(0) | rs794187 | 3 | 4436172 | 0.089616578 | SUMF1(0) |
| rs12955326 | 18 | 566905 | 0.13273966 | . | rs10987911 | 9 | 130945298 | 0.089599333 | CIZ1(0) |
| rs7925824 | 11 | 45929961 | 0.132645404 | . | rs78572338 | 1 | 52851729 | 0.089428529 | ORC1(0) |
| rs9402997 | 6 | 139058418 | 0.132539124 | LOC100507462(0) | rs4551053 | 5 | 157838542 | 0.08940253 | . |
| rs12435881 | 14 | 84125120 | 0.1322724 | . | rs12459960 | 19 | 2510036 | 0.089395187 | . |
| rs17730711 | 16 | 6769752 | 0.132017257 | RBFOX1(0) | rs589757 | 5 | 41572722 | 0.089377651 | . |
| rs12492883 | 3 | 139035356 | 0.132002461 | . | rs10741742 | 11 | 18342160 | 0.089342735 | HPS5(0) |
| rs2711390 | 18 | 68046599 | 0.131714436 | . | rs1478892 | 8 | 11448529 | 0.089339784 | . |
| rs978477 | 9 | 125308131 | 0.131398638 | #NAME? | rs10281574 | 7 | 5483042 | 0.089277968 | . |
| rs510178 | 3 | 196704036 | 0.131238025 | . | rs292726 | 11 | 37807834 | 0.089227414 | . |
| rs1026301 | 2 | 35210500 | 0.130794158 | . | rs11679606 | 2 | 16795418 | 0.089163622 | FAM49A(0) |
| rs1776570 | 4 | 37265814 | 0.130537887 | NWD2(0) | rs6723770 | 2 | 16795666 | 0.089163622 | FAM49A(0) |
| rs2465169 | 18 | 68046681 | 0.130345939 | . | rs7635628 | 3 | 74783321 | 0.088892266 | . |
| rs6439233 | 3 | 130135456 | 0.130187093 | COL6A5(0) | rs16843879 | 3 | 136003194 | 0.088880285 | PCCB(0) |
| rs11251927 | 10 | 3556594 | 0.129253099 | . | rs2555737 | 6 | 93208564 | 0.08885976 | . |
| rs8023044 | 14 | 84118602 | 0.129203601 | . | rs17468034 | 10 | 94965772 | 0.088842496 | . |
| rs7895779 | 10 | 82202615 | 0.128743724 | . | rs2889678 | 20 | 31189993 | 0.088541356 | LOC149950(0) |
| rs36160296 | 2 | 14311647 | 0.12841479 | . | rs975664 | 2 | 75368797 | 0.088458267 | TACR1(0) |
| rs886205 | 12 | 112204427 | 0.128207047 | #NAME? | rs12106074 | 20 | 17356566 | 0.088331984 | PCSK2(0) |
| rs7306191 | 12 | 56080164 | 0.127942131 | ITGA7(0) | rs757248 | 4 | 7703505 | 0.088236845 | SORCS2(0) |
| rs59743544 | 9 | 125275414 | 0.12731147 | . | rs6073949 | 20 | 44531878 | 0.088112967 | PLTP(0) |
| rs11600112 | 11 | 45931019 | 0.127201784 | . | rs1522699 | 2 | 159764389 | 0.088093783 | . |
| rs2285750 | 17 | 42114672 | 0.127072025 | LSM12(0) | rs687174 | 5 | 41564562 | 0.087906813 | . |
| rs16960761 | 19 | 34432677 | 0.126938504 | . | rs8468 | 15 | 63434110 | 0.087764309 | LACTB(0)\|=missense |
| rs10167958 | 2 | 109921632 | 0.126874243 | SH3RF3(0) | rs3112931 | 2 | 180427409 | 0.087275977 | ZNF385B(0) |
| rs2067928 | 2 | 112850581 | 0.126773397 | TMEM87B(0) | rs7915994 | 10 | 95526168 | 0.087134428 | LGI1(0) |
| rs7991707 | 13 | 31872181 | 0.126490432 | B3GALTL(0) | rs856614 | 1 | 52252006 | 0.086772114 | OSBPL9(0) |
| rs7186971 | 16 | 75655406 | 0.125470972 | ADAT1(0) | rs1580678 | 11 | 97009206 | 0.086554779 | . |
| rs6141285 | 20 | 30892434 | 0.125442896 | KIF3B(0) | rs6851301 | 4 | 140959594 | 0.08647562 | MAML3(0) |
| rs1404915 | 7 | 110272427 | 0.125012681 | . | rs4150579 | 11 | 18357180 | 0.086393353 | GTF2H1(0) |
| rs10769865 | 11 | 8061100 | 0.124947194 | TUB(0) | rs10412132 | 19 | 30914411 | 0.086281907 | ZNF536(0) |
| rs1800974 | 12 | 56089357 | 0.124736579 | ITGA7(0) | rs1476331 | 3 | 195959924 | 0.0862772 | SLC51A(0) |
| rs17764821 | 16 | 75685746 | 0.124644625 | TERF2IP(0)\|=missense | rs930981 | 11 | 122834265 | 0.086137936 | . |
| rs28773026 | 11 | 13566659 | 0.123827628 | . | rs6708276 | 2 | 144442150 | 0.086063532 | ARHGAP15(0) |
| rs1476331 | 3 | 195959924 | 0.123476854 | SLC51A(0) | rs3740711 | 11 | 18379839 | 0.086058659 | GTF2H1(0) |
| rs6490294 | 12 | 112190438 | 0.123294938 | ACAD10(0)\|=missense | rs4150650 | 11 | 18374279 | 0.086058659 | GTF2H1(0) |
| rs4664089 | 2 | 153045036 | 0.123199205 | . | rs4150651 | 11 | 18374425 | 0.086058659 | GTF2H1(0) |
| rs11126430 | 2 | 74544125 | 0.123122731 | SLC4A5(0)\|=missense | rs4150673 | 11 | 18383449 | 0.086058659 | GTF2H1(0) |
| rs7525615 | 1 | 229835413 | 0.12311893 | . | rs3851518 | 6 | 6336584 | 0.086016572 | . |
| rs11578670 | 1 | 246020370 | 0.123050125 | SMYD3(0) | rs12730665 | 1 | 96683642 | 0.085945583 | . |
| rs757248 | 4 | 7703505 | 0.123000987 | SORCS2(0) | rs17656368 | 3 | 71474216 | 0.085783979 | FOXP1(0) |
| rs1412695 | 9 | 3125843 | 0.122327471 | . | rs10495563 | 2 | 9662210 | 0.08559147 | ADAM17(0) |
| rs10910527 | 1 | 232599189 | 0.122271966 | SIPA1L2(0) | rs221310 | 20 | 35272778 | 0.08551986 | SLA2(0) |
| rs1458 | 19 | 34660146 | 0.122146743 | . | rs4150641 | 11 | 18370739 | 0.085500981 | GTF2H1(0) |
| rs10092924 | 8 | 654796 | 0.12208874 | ERICH1(0) | rs4150615 | 11 | 18365028 | 0.085341186 | GTF2H1(0) |
| rs7590562 | 2 | 1087950 | 0.122015317 | SNTG2(0) | rs2116980 | 9 | 23503695 | 0.08533888 | . |
| rs1807921 | 2 | 159694749 | 0.121813479 | . | rs4150612 | 11 | 18364358 | 0.085332474 | GTF2H1(0) |
| rs4664076 | 2 | 152921491 | 0.121733232 | CACNB4(0) | rs4150661 | 11 | 18379629 | 0.085332474 | GTF2H1(0) |
| rs11102573 | 1 | 113573223 | 0.121698754 | LOC100996251(0) | rs17027780 | 4 | 152590337 | 0.085301439 | #NAME? |
| rs10135768 | 14 | 51671017 | 0.121564412 | . | rs1339822 | 10 | 94915615 | 0.085112947 | . |
| rs12513800 | 5 | 53966360 | 0.121133738 | LOC102467080(0) | rs529177 | 2 | 45100208 | 0.085101078 | . |
| rs1704392 | 15 | 42206976 | 0.120955914 | EHD4(0) | rs74550841 | 7 | 13842687 | 0.084948559 | . |
| rs7546194 | 1 | 81870764 | 0.120928271 | . | rs4150562 | 11 | 18354995 | 0.084936611 | GTF2H1(0) |
| rs1470023 | 5 | 10800150 | 0.120834189 | . | rs2979725 | 8 | 25352468 | 0.08490791 | CDCA2(0) |
| rs10115104 | 9 | 71540376 | 0.120768458 | PIP5K1B(0) | rs7027419 | 9 | 107485577 | 0.084822004 | #NAME? |
| rs1759092 | 19 | 34676538 | 0.12072251 | LSM14A(0) | rs7954506 | 12 | 99175840 | 0.084791024 | ANKS1B(0) |
| rs6417001 | 18 | 14550022 | 0.12064537 | #NAME? | rs12541800 | 8 | 11423072 | 0.084610051 | . |
| rs2545656 | 5 | 103031154 | 0.120610174 | . | rs2585390 | 14 | 49268677 | 0.084595601 | . |
| rs2048219 | 5 | 33382240 | 0.120602295 | . | rs2837957 | 21 | 42491120 | 0.084594652 | . |
| rs2414167 | 15 | 52907428 | 0.120539951 | FAM214A(0)\|=missense | rs4843708 | 16 | 87857939 | 0.084527887 | . |
| rs4810551 | 20 | 45385777 | 0.120424366 | . | rs6761396 | 2 | 104113458 | 0.084367847 | . |
| rs11639447 | 15 | 29625717 | 0.120397795 | FAM189A1(0) | rs11617361 | 13 | 101758117 | 0.084304353 | NALCN(0) |
| rs2147065 | 9 | 9860780 | 0.120371486 | PTPRD(0) | rs4150575 | 11 | 18356466 | 0.084208744 | GTF2H1(0) |
| rs11211480 | 1 | 47693220 | 0.120346733 | TAL1(0) | rs9833156 | 3 | 34198737 | 0.08420726 | . |
| rs12590913 | 14 | 50241661 | 0.12004752 | KLHDC2(0) | rs3749427 | 1 | 247150740 | 0.084198385 | ZNF670-ZNF695(0)\|ZNF695(0)\|=missense |
| rs12604641 | 18 | 5631847 | 0.119566486 | . | rs2093830 | 9 | 12756889 | 0.084066027 | . |
| rs1867343 | 4 | 99719961 | 0.119454259 | . | rs191705 | 20 | 12912855 | 0.084058409 | LOC101929486(0) |
| rs4932554 | 15 | 92097108 | 0.119361073 | . | rs6673823 | 1 | 234668280 | 0.083996671 | . |
| rs1048103 | 12 | 56078189 | 0.119321728 | METTL7B(0) | rs12276244 | 11 | 123957157 | 0.083830585 | . |
| rs8005962 | 14 | 96027153 | 0.119160786 | . | rs290861 | 1 | 97542797 | 0.083792818 | . |
| rs11053455 | 12 | 34826346 | 0.119040556 | . | rs1476873 | 2 | 37949251 | 0.083724498 | . |
| rs3019648 | 11 | 134505872 | 0.118994359 | . | rs10507397 | 13 | 31715015 | 0.083695759 | HSPH1(0) |
| rs12461519 | 19 | 15523914 | 0.118817719 | AKAP8L(0) | rs6540602 | 1 | 210696806 | 0.083673372 | HHAT(0) |
| rs28669813 | 5 | 174430344 | 0.118748052 | . | rs7267720 | 20 | 31166190 | 0.083655499 | NOL4L(0) |
| rs13389867 | 2 | 160042430 | 0.118745351 | TANC1(0) | rs62014517 | 15 | 30233518 | 0.083577643 | . |
| rs11857150 | 15 | 52922216 | 0.118709634 | FAM214A(0)\|=missense | rs67211505 | 12 | 125780255 | 0.083571207 | . |
| rs8007406 | 14 | 96027230 | 0.118644623 | . | rs9538601 | 13 | 31719128 | 0.083551817 | HSPH1(0) |
| rs72790850 | 2 | 34837783 | 0.118630033 | . | rs2106735 | 7 | 93070811 | 0.083148131 | CALCR(0) |
| rs7015856 | 8 | 64247381 | 0.118378466 | . | rs12224084 | 11 | 1639388 | 0.082970303 | . |
| rs2183614 | 14 | 49829709 | 0.118250498 | . | rs28113 | 3 | 10400643 | 0.0829518 | ATP2B2(0) |
| rs3864217 | 4 | 7380740 | 0.118228646 | SORCS2(0) | rs876305 | 7 | 24758795 | 0.082888089 | DFNA5(0) |
| rs4933165 | 10 | 91662902 | 0.11818263 | . | rs1432655 | 5 | 163028782 | 0.082887716 | . |
| rs2578624 | 5 | 10245114 | 0.11762813 | FAM173B(0) | rs6057599 | 20 | 31168439 | 0.08285499 | NOL4L(0) |
| rs2578628 | 5 | 10242162 | 0.11762813 | FAM173B(0) | rs231675 | 17 | 3190599 | 0.082816002 | #NAME? |
| rs11202960 | 10 | 82209807 | 0.117609146 | . | rs8103067 | 19 | 32027415 | 0.082609438 | . |
| rs4911546 | 20 | 30718333 | 0.117320292 | TM9SF4(0) | rs17296815 | 12 | 51670202 | 0.082593854 | . |
| rs7196251 | 16 | 58410039 | 0.117306534 | . | rs2499502 | 6 | 88661167 | 0.082588061 | . |
| rs2890503 | 9 | 3128693 | 0.116875938 | . | rs1156992 | 3 | 26028747 | 0.082417277 | . |

| | | | | |
|---|---|---|---|---|
| rs550915 | 9 | 104191392 | 0.082198027 | ALDOB(0) |
| rs9849293 | 3 | 26085604 | 0.082073186 | . |
| rs7109581 | 11 | 93180531 | 0.08201969 | . |
| rs509362 | 11 | 126036852 | 0.081907223 | . |
| rs2959356 | 18 | 23590203 | 0.081893537 | . |
| rs74918773 | 21 | 41711938 | 0.081863186 | DSCAM(0) |
| rs9596209 | 13 | 50463758 | 0.081723468 | . |
| rs925205 | 5 | 10141127 | 0.08168095 | . |
| rs66495531 | 2 | 206496074 | 0.081674975 | . |
| rs67649296 | 18 | 24171427 | 0.081588335 | KCTD1(0) |
| rs72860574 | 2 | 146078481 | 0.081551799 | . |
| rs12955326 | 18 | 566905 | 0.081537145 | . |
| rs996018 | 3 | 26044946 | 0.081384856 | . |
| rs34089886 | 4 | 37297267 | 0.081313692 | NWD2(0) |
| rs6745500 | 2 | 9640526 | 0.081262383 | ADAM17(0) |
| rs7519350 | 1 | 208712599 | 0.080966816 | . |
| rs13436699 | 5 | 10147758 | 0.08096024 | . |
| rs2521755 | 7 | 24749223 | 0.080740024 | DFNA5(0) |
| rs11142207 | 9 | 90886728 | 0.080733174 | . |
| rs4867365 | 5 | 31678198 | 0.08067672 | . |
| rs4325166 | 1 | 62506473 | 0.080577817 | INADL(0) |
| rs11703784 | 22 | 48483072 | 0.080574665 | . |
| rs8132084 | 21 | 20220881 | 0.080502318 | . |
| rs264963 | 2 | 104254564 | 0.080449015 | . |
| rs72807288 | 10 | 94042448 | 0.080367219 | CPEB3(0) |
| rs11610061 | 12 | 100050148 | 0.080360589 | ANKS1B(0) |
| rs2250606 | 17 | 80662614 | 0.080299561 | . |
| rs2642992 | 1 | 247151557 | 0.080255179 | ZNF670-ZNF695(0)\|ZNF695(0)\|=MISSENSE |
| rs9968731 | 5 | 80498455 | 0.080235512 | RASGRF2(0) |
| rs2551617 | 3 | 120367819 | 0.080232729 | HGD(0) |
| rs926244 | 1 | 3006283 | 0.080128453 | PRDM16(0) |
| rs10267099 | 7 | 87278760 | 0.080096606 | ABCB1(0)\|RUNDC3B(0) |
| rs11752519 | 6 | 159270631 | 0.080022924 | OSTCP1(0) |
| rs6708166 | 2 | 30526780 | 0.080010227 | . |
| rs2247493 | 9 | 130296484 | 0.079962022 | FAM129B(0) |
| rs12444034 | 16 | 83405551 | 0.07994533 | CDH13(0) |
| rs10260787 | 7 | 7129858 | 0.079914626 | LOC100131257(0)\|=missense |
| rs4869095 | 5 | 163063208 | 0.079908136 | . |
| rs911478 | 6 | 109262787 | 0.079874849 | ARMC2(0) |
| rs10470654 | 3 | 26055831 | 0.079710401 | . |
| rs10110604 | 8 | 16503082 | 0.079708984 | . |
| rs3748671 | 1 | 207253031 | 0.079685006 | PFKFB2(0) |
| rs72748753 | 5 | 22803278 | 0.07963518 | CDH12(0) |
| rs13226913 | 7 | 7246846 | 0.079445211 | C1GALT1(0) |
| rs12554403 | 9 | 26489225 | 0.079343024 | . |
| rs2067928 | 2 | 112850581 | 0.079324389 | TMEM87B(0) |
| rs4741226 | 9 | 12502531 | 0.079265891 | . |
| rs3006210 | 6 | 169828915 | 0.079053897 | . |
| rs2521222 | 7 | 24541524 | 0.079052413 | . |
| rs10817739 | 9 | 100308231 | 0.079012723 | TMOD1(0) |
| rs10167958 | 2 | 109921632 | 0.078985889 | SH3RF3(0) |
| rs6583867 | 10 | 94938228 | 0.078972417 | . |
| rs12535 | 2 | 159663616 | 0.078949419 | DAPL1(0) |
| rs11726701 | 4 | 132988240 | 0.07883832 | . |
| rs68055747 | 18 | 75347847 | 0.078821046 | . |
| rs4905772 | 14 | 99372389 | 0.078737463 | . |
| rs10933722 | 3 | 193954976 | 0.078707908 | . |
| rs245974 | 7 | 29295856 | 0.07862484 | CHN2(0) |
| rs17302392 | 8 | 66463308 | 0.078469967 | LINC01299(0) |
| rs2841031 | 10 | 90940003 | 0.078383816 | . |
| rs12692772 | 2 | 166373185 | 0.077852577 | CSRNP3(0) |
| rs2243955 | 9 | 130303640 | 0.077831334 | FAM129B(0) |
| rs727755 | 4 | 158724848 | 0.077668349 | . |
| rs1474249 | 2 | 165419009 | 0.077617732 | GRB14(0) |
| rs10455806 | 6 | 166146245 | 0.077613883 | . |
| rs726972 | 19 | 32034831 | 0.077540119 | . |
| rs10136340 | 14 | 70527933 | 0.077428543 | SLC8A3(0) |
| rs2783197 | 1 | 52436038 | 0.077374361 | RAB3B(0) |
| rs2183625 | 14 | 56417838 | 0.077224508 | . |
| rs17734690 | 8 | 9713196 | 0.077148675 | . |
| rs11666325 | 19 | 30907789 | 0.077101923 | ZNF536(0) |
| rs2069727 | 12 | 68548223 | 0.077099852 | . |
| rs8136304 | 22 | 27917070 | 0.076986824 | . |
| rs7895779 | 10 | 82202615 | 0.076971709 | . |
| rs111862778 | 3 | 135783796 | 0.076782448 | PPP2R3A(0) |
| rs4947040 | 6 | 110258455 | 0.076772503 | . |
| rs3892909 | 5 | 175305591 | 0.076753557 | CPLX2(0) |
| rs10105657 | 8 | 607251 | 0.076633908 | . |
| rs2540499 | 12 | 96399869 | 0.076295006 | LTA4H(0) |
| rs13208135 | 6 | 137681617 | 0.076269394 | . |
| rs7834638 | 8 | 11420295 | 0.075717119 | BLK(0)\|=missense |
| rs6918011 | 6 | 134715654 | 0.075588484 | . |
| rs60326795 | 3 | 107420674 | 0.075126881 | BBX(0) |
| rs17398598 | 1 | 52024908 | 0.075113326 | . |
| rs9878775 | 3 | 182659953 | 0.075104778 | . |
| rs6544598 | 2 | 43031054 | 0.075084947 | . |
| rs2675294 | 3 | 2751092 | 0.075048446 | CNTN4(0) |
| rs11645844 | 16 | 78408972 | 0.075042387 | WWOX(0)\|=missense |
| rs1989830 | 7 | 87205663 | 0.075004687 | ABCB1(0) |
| rs1118972 | 8 | 19086353 | 0.074979109 | LOC100128993(0) |
| rs730570 | 14 | 101142890 | 0.074883337 | . |
| rs35970366 | 7 | 87495087 | 0.074767308 | SLC25A40(0) |
| rs331941 | 4 | 143657023 | 0.074745413 | INPP4B(0) |
| rs7525615 | 1 | 229835413 | 0.074719562 | . |
| rs17536135 | 4 | 20295132 | 0.074710179 | SLIT2(0) |
| rs9856369 | 3 | 25968912 | 0.07470341 | . |
| rs13145422 | 4 | 160185669 | 0.074623886 | . |
| rs56160474 | 1 | 97544258 | 0.074622335 | DPYD(0) |
| rs17194406 | 3 | 2488631 | 0.074499511 | CNTN4(0) |
| rs1820048 | 5 | 107113395 | 0.074478387 | . |
| rs13241157 | 7 | 18860014 | 0.07436295 | HDAC9(0) |
| rs1995296 | 12 | 100082450 | 0.074359259 | ANKS1B(0) |
| rs13389867 | 2 | 160042430 | 0.074345431 | TANC1(0) |
| rs17473802 | 2 | 39752628 | 0.074261429 | LOC728730(0) |
| rs2930895 | 19 | 893080 | 0.074149393 | MED16(0) |
| rs10056443 | 5 | 88752584 | 0.074035881 | . |
| rs1074789 | 9 | 12752340 | 0.073883849 | . |
| rs1888189 | 9 | 10948272 | 0.073778858 | . |
| rs13107700 | 4 | 133111065 | 0.073635885 | . |
| rs644631 | 13 | 32098486 | 0.073598561 | . |
| rs4772114 | 13 | 99298233 | 0.073573011 | . |
| rs28621923 | 8 | 72351333 | 0.073538141 | . |
| rs5015243 | 1 | 217427461 | 0.073500411 | . |
| rs2031340 | 10 | 10586371 | 0.073453965 | . |
| rs13240660 | 7 | 4264920 | 0.073444734 | SDK1(0) |
| rs7306191 | 12 | 56080164 | 0.073428105 | ITGA7(0) |
| rs4855697 | 3 | 108867128 | 0.073379951 | FLJ22763(0) |
| rs791681 | 9 | 12522037 | 0.073375326 | . |

| | | | | |
|---|---|---|---|---|
| rs9405499 | 6 | 1630555 | 0.110123285 | GMDS(0) |
| rs4608676 | 3 | 2912864 | 0.110123012 | CNTN4(0) |
| rs11211480 | 1 | 47693220 | 0.110054136 | TAL1(0) |
| rs5761748 | 22 | 27177355 | 0.109941256 | . |
| rs10432402 | 2 | 5467135 | 0.109934074 | . |
| rs8003022 | 14 | 90038821 | 0.109848639 | FOXN3(0) |
| rs72603568 | 7 | 114299774 | 0.109823888 | FOXP2(0) |
| rs16877147 | 5 | 79030212 | 0.109803546 | CMYA5(0)\|=MISSENSE |
| rs16877150 | 5 | 79030338 | 0.109803546 | CMYA5(0)\|=MISSENSE |
| rs62621915 | 5 | 79027700 | 0.109803546 | CMYA5(0) |
| rs35035095 | 13 | 39376300 | 0.109692361 | FREM2(0) |
| rs7545384 | 1 | 8580960 | 0.109642101 | RERE(0) |
| rs4866602 | 5 | 3689363 | 0.109601215 | . |
| rs751800 | 9 | 100884564 | 0.109583913 | CORO2A(0) |
| rs16877133 | 5 | 79028513 | 0.109497641 | CMYA5(0)\|=MISSENSE |
| rs10209727 | 2 | 121828335 | 0.10880556 | . |
| rs4702678 | 5 | 10222014 | 0.108796314 | . |
| rs11103302 | 9 | 138958552 | 0.108637124 | NACC2(0) |
| rs1035055 | 2 | 160057141 | 0.108581984 | TANC1(0) |
| rs28362542 | 5 | 79032711 | 0.108519484 | CMYA5(0) |
| rs10816216 | 9 | 9861006 | 0.108496397 | PTPRD(0) |
| rs1867343 | 4 | 99719961 | 0.108487157 | . |
| rs4129493 | 18 | 38139080 | 0.10845167 | . |
| rs2148327 | 1 | 232597084 | 0.108319011 | SIPA1L2(0) |
| rs58512728 | 10 | 2212767 | 0.108077974 | . |
| rs4800250 | 18 | 23810767 | 0.108035445 | TAF4B(0) |
| rs4869095 | 5 | 163063208 | 0.107842137 | . |
| rs4845404 | 1 | 155007293 | 0.107800504 | DCST1(0)\|=missense |
| rs11707053 | 3 | 139035269 | 0.107678049 | . |
| rs17579757 | 2 | 171259308 | 0.107651347 | MYO3B(0) |
| rs2107942 | 7 | 70131823 | 0.107587657 | AUTS2(0) |
| rs2052466 | 5 | 167261382 | 0.107503885 | TENM2(0) |
| rs245974 | 7 | 29295856 | 0.107446695 | CHN2(0) |
| rs6655965 | 1 | 203680492 | 0.107404131 | ATP2B4(0) |
| rs1135760 | 7 | 44868447 | 0.107316149 | H2AFV(0) |
| rs11257426 | 10 | 11960602 | 0.10729016 | . |
| rs882383 | 12 | 54601031 | 0.107277671 | . |
| rs11264300 | 1 | 155019710 | 0.107241324 | DCST1(0)\|LOC100505666(0)\|=MISSENSE |
| rs682927 | 9 | 1358244 | 0.107154204 | . |
| rs2183614 | 14 | 49829709 | 0.107139792 | . |
| rs1132787 | 4 | 145030546 | 0.106993999 | GYPA(0) |
| rs6734469 | 2 | 9735900 | 0.106913816 | YWHAQ(0) |
| rs7070947 | 10 | 94967076 | 0.106891502 | . |
| rs11683407 | 2 | 35935526 | 0.106691423 | . |
| rs17029538 | 2 | 65096800 | 0.106549881 | . |
| rs17109031 | 12 | 54598367 | 0.106460249 | . |
| rs250508 | 19 | 8334280 | 0.106283685 | . |
| rs35397 | 5 | 33951116 | 0.106112652 | SLC45A2(0) |
| rs10769865 | 11 | 8061100 | 0.106047338 | TUB(0) |
| rs3092735 | 20 | 45419188 | 0.105936636 | . |
| rs4848908 | 2 | 112723117 | 0.105873653 | MERTK(0)\|=missense |
| rs13389867 | 2 | 160042430 | 0.105849181 | TANC1(0) |
| rs6892937 | 5 | 3684777 | 0.105710999 | . |
| rs9938424 | 16 | 78430875 | 0.105667691 | WWOX(0) |
| rs667631 | 13 | 102823429 | 0.105617027 | FGF14(0) |
| rs1788783 | 18 | 21161134 | 0.105539771 | NPC1(0)\|=missense |
| rs12408012 | 1 | 4379687 | 0.105538448 | . |
| rs6779650 | 3 | 108072704 | 0.105537085 | HHLA2(0)\|=missense |
| rs9525816 | 13 | 44223898 | 0.105291808 | ENOX1(0) |
| rs7861047 | 9 | 124877059 | 0.105247839 | . |
| rs4436441 | 1 | 202087936 | 0.10512356 | . |
| rs789129 | 12 | 28980279 | 0.105060683 | . |
| rs11886265 | 2 | 225191976 | 0.105049537 | . |
| rs16877109 | 5 | 79024779 | 0.105036005 | CMYA5(0)\|=MISSENSE |
| rs2057302 | 1 | 172018734 | 0.104894344 | DNM3(0) |
| rs6725244 | 2 | 233877065 | 0.104783753 | NGEF(0) |
| rs17807624 | 8 | 11463015 | 0.104680317 | . |
| rs2077668 | 10 | 4511571 | 0.104295492 | . |
| rs1028180 | 1 | 169345868 | 0.104217269 | BLZF1(0)\|=MISSENSE |
| rs750467 | 1 | 146589758 | 0.104200659 | #NAME? |
| rs593847 | 18 | 5991270 | 0.104156827 | L3MBTL4(0) |
| rs6448494 | 4 | 11099518 | 0.104070782 | . |
| rs2293119 | 4 | 140941411 | 0.104029588 | MAML3(0) |
| rs10270530 | 7 | 8488691 | 0.103858142 | NXPH1(0) |
| rs7841393 | 8 | 77760070 | 0.103816003 | ZFHX4(0) |
| rs1211610 | 12 | 131607054 | 0.103713907 | GPR133(0) |
| rs2585390 | 14 | 49268677 | 0.103465295 | . |
| rs1370717 | 3 | 32354387 | 0.1034448 | CMTM8(0) |
| rs7787870 | 7 | 115454 | 0.103340722 | . |
| rs57461190 | 1 | 206980130 | 0.103281595 | IL19(0) |
| rs6542487 | 2 | 119648349 | 0.103220793 | . |
| rs1887867 | 9 | 7274696 | 0.103184 | . |
| rs3746236 | 19 | 35298961 | 0.103111753 | . |
| rs1656191 | 4 | 37264580 | 0.103090649 | NWD2(0) |
| rs34974801 | 16 | 75939252 | 0.10306561 | . |
| rs13117732 | 4 | 177496193 | 0.103039633 | . |
| rs12553531 | 9 | 112950754 | 0.103019624 | . |
| rs3824103 | 8 | 27324429 | 0.102815491 | CHRNA2(0) |
| rs11119603 | 1 | 206979430 | 0.102782641 | IL19(0) |
| rs11950944 | 5 | 79422993 | 0.102756124 | SERINC5(0) |
| rs11583200 | 1 | 50559820 | 0.10268587 | ELAVL4(0) |
| rs3852166 | 5 | 10281549 | 0.102591507 | CMBL(0) |
| rs4654839 | 1 | 20467577 | 0.102525154 | PLA2G2F(0) |
| rs10138228 | 14 | 49778543 | 0.102328883 | . |
| rs1564469 | 8 | 136210764 | 0.102218635 | . |
| rs1551491 | 4 | 167727136 | 0.102080348 | SPOCK3(0) |
| rs9402997 | 6 | 139058418 | 0.101979507 | LOC100507462(0) |
| rs11793053 | 9 | 37413413 | 0.101951747 | . |
| rs9826146 | 3 | 135983089 | 0.101910946 | PCCB(0) |
| rs6790456 | 3 | 107739094 | 0.101762167 | . |
| rs2544371 | 2 | 170227070 | 0.101749027 | . |
| rs2778031 | 9 | 90835726 | 0.101742518 | . |
| rs9792575 | 9 | 29371045 | 0.101742397 | . |
| rs4525226 | 11 | 7663887 | 0.101708026 | PPFIBP2(0) |
| rs4895745 | 6 | 125797983 | 0.101638412 | . |
| rs978485 | 3 | 64323801 | 0.101605042 | . |
| rs2209543 | 6 | 9384993 | 0.101579169 | . |
| rs75959802 | 4 | 142041988 | 0.101554848 | RNF150(0) |
| rs9396643 | 6 | 16092993 | 0.101427194 | . |
| rs4897445 | 6 | 130926413 | 0.101392838 | . |
| rs7955104 | 12 | 54888139 | 0.101382087 | . |
| rs12444034 | 16 | 83405551 | 0.101283994 | CDH13(0) |
| rs878394 | 1 | 219073958 | 0.101195666 | . |
| rs6970262 | 7 | 55259763 | 0.101145246 | EGFR(0) |
| rs12470270 | 2 | 65390283 | 0.101091211 | . |
| rs7682543 | 4 | 81154975 | 0.100956959 | . |

| | | | | |
|---|---|---|---|---|
| rs9997579 | 4 | 188990434 | 0.116787342 | . |
| rs3736508 | 11 | 45975130 | 0.116310612 | PHF21A(0)\|=MISSENSE |
| rs2295035 | 20 | 30702144 | 0.116088296 | TM9SF4(0) |
| rs12212533 | 6 | 13258648 | 0.115903248 | PHACTR1(0) |
| rs7099721 | 10 | 5055950 | 0.115874138 | AKR1C2(0) |
| rs4897445 | 6 | 130926413 | 0.115692286 | . |
| rs3864202 | 4 | 7329426 | 0.11547736 | SORCS2(0) |
| rs2497484 | 13 | 21199265 | 0.115373366 | IFT88(0) |
| rs11950944 | 5 | 79422993 | 0.115170657 | SERINC5(0) |
| rs12663976 | 6 | 156730386 | 0.115099191 | . |
| rs1355696 | 12 | 17424512 | 0.115007433 | . |
| rs2762990 | 13 | 21371427 | 0.114866737 | XPO4(0) |
| rs11121181 | 1 | 8477006 | 0.114810554 | RERE(0) |
| rs10110604 | 8 | 16503082 | 0.114669501 | . |
| rs1132787 | 4 | 145030546 | 0.114524282 | GYPA(0) |
| rs3864220 | 4 | 7381431 | 0.114462319 | SORCS2(0) |
| rs10788619 | 10 | 82195355 | 0.114461244 | . |
| rs10802528 | 1 | 236506815 | 0.114388781 | . |
| rs1551491 | 4 | 167727136 | 0.114295015 | SPOCK3(0) |
| rs4869095 | 5 | 163063208 | 0.114074551 | . |
| rs8051241 | 16 | 75622993 | 0.113914321 | . |
| rs153852 | 5 | 94176581 | 0.113509622 | MCTP1(0) |
| rs10909810 | 1 | 3783278 | 0.113444556 | DFFB(0) |
| rs10018031 | 4 | 133001527 | 0.113383203 | . |
| rs3092379 | 20 | 45432808 | 0.113309165 | . |
| rs694421 | 19 | 34728620 | 0.112875513 | . |
| rs3846599 | 5 | 10255821 | 0.112846909 | CCT5(0) |
| rs2964798 | 5 | 10788833 | 0.112810696 | . |
| rs6665323 | 1 | 47953054 | 0.11281059 | . |
| rs4702678 | 5 | 10222014 | 0.112723974 | . |
| rs6708276 | 2 | 144442150 | 0.112631782 | ARHGAP15(0) |
| rs9792575 | 9 | 29371045 | 0.112328368 | . |
| rs229459 | 21 | 22048355 | 0.112200627 | . |
| rs4316481 | 11 | 36712583 | 0.111792805 | #NAME? |
| rs9848247 | 3 | 196078746 | 0.111586249 | . |
| rs2984132 | 10 | 105657892 | 0.111445213 | OBFC1(0) |
| rs6891924 | 5 | 33369693 | 0.111391606 | . |
| rs10169375 | 2 | 230945993 | 0.111357378 | . |
| rs17717812 | 14 | 90017447 | 0.111083176 | FOXN3(0) |
| rs28377091 | 11 | 1691314 | 0.111062028 | . |
| rs6141074 | 20 | 1041340 | 0.110812803 | . |
| rs10961165 | 9 | 13602338 | 0.110798063 | . |
| rs12370122 | 12 | 127108055 | 0.110771134 | . |
| rs7934611 | 11 | 134613171 | 0.110745511 | . |
| rs7529591 | 1 | 29755064 | 0.110693485 | . |
| rs3743599 | 16 | 75646576 | 0.110622886 | ADAT1(0)\|=MISSENSE |
| rs184234 | 15 | 101251719 | 0.110551045 | . |
| rs969348 | 15 | 81847279 | 0.110437463 | . |
| rs10946261 | 6 | 170721995 | 0.11018023 | . |
| rs1920667 | 11 | 15847373 | 0.110144654 | . |
| rs2581851 | 7 | 154844123 | 0.110101201 | . |
| rs28113 | 3 | 10400643 | 0.109777666 | ATP2B2(0) |
| rs1061646 | 16 | 89805977 | 0.109719768 | FANCA(0)\|ZNF276(0)\|=missense |
| rs9878602 | 3 | 71535338 | 0.109711333 | FOXP1(0) |
| rs1588930 | 2 | 17191334 | 0.109695053 | . |
| rs2247493 | 9 | 130296484 | 0.109538276 | FAM129B(0) |
| rs10487868 | 7 | 83477670 | 0.109446853 | . |
| rs9789638 | 2 | 227347232 | 0.109428532 | . |
| rs7070947 | 10 | 94967076 | 0.10931237 | . |
| rs2418589 | 11 | 32279713 | 0.109184336 | . |
| rs6057602 | 20 | 31181808 | 0.109126849 | LOC149950(0) |
| rs2014131 | 4 | 155047531 | 0.109011684 | . |
| rs4348200 | 5 | 107139260 | 0.108820511 | . |
| rs6431311 | 2 | 235808211 | 0.108706671 | . |
| rs7336364 | 13 | 44166351 | 0.108659706 | ENOX1(0) |
| rs9666645 | 11 | 34212883 | 0.10839729 | ABTB2(0) |
| rs900431 | 15 | 81869796 | 0.108370676 | . |
| rs1887867 | 9 | 7274696 | 0.108350628 | . |
| rs10490069 | 2 | 60608117 | 0.108345168 | . |
| rs6585804 | 10 | 123980108 | 0.108253772 | TACC2(0) |
| rs4243474 | 6 | 145096340 | 0.108223428 | UTRN(0) |
| rs6013471 | 20 | 36559239 | 0.108150742 | VSTM2L(0) |
| rs718276 | 15 | 81936035 | 0.108078412 | . |
| rs956706 | 1 | 246097156 | 0.108059635 | SMYD3(0) |
| rs2209543 | 6 | 9384993 | 0.108036165 | . |
| rs616519 | 1 | 10768002 | 0.107960351 | CASZ1(0) |
| rs2302344 | 7 | 20338100 | 0.107803895 | LOC101927769(0) |
| rs10072686 | 5 | 10308240 | 0.10778611 | . |
| rs6671634 | 1 | 234228154 | 0.107573743 | SLC35F3(0) |
| rs7608560 | 2 | 216813103 | 0.107545055 | MREG(0) |
| rs10741741 | 11 | 18342081 | 0.107400466 | HPS5(0) |
| rs1325129 | 9 | 12512973 | 0.107220636 | . |
| rs11042937 | 11 | 10745394 | 0.106924097 | . |
| rs6841127 | 4 | 137937334 | 0.106718738 | . |
| rs7184656 | 16 | 78982440 | 0.106671372 | WWOX(0) |
| rs11196172 | 10 | 114726843 | 0.106442394 | TCF7L2(0) |
| rs1024116 | 18 | 75432386 | 0.106419886 | . |
| rs910257 | 11 | 34201285 | 0.106375443 | ABTB2(0) |
| rs17013593 | 4 | 129593111 | 0.106227776 | . |
| rs12921190 | 16 | 73413743 | 0.106226166 | . |
| rs651202 | 11 | 119881745 | 0.10618328 | . |
| rs6491593 | 13 | 101524160 | 0.106179795 | NALCN-AS1(0) |
| rs1522699 | 2 | 159764389 | 0.106115418 | . |
| rs11707053 | 3 | 139035269 | 0.105953594 | . |
| rs4236315 | 7 | 24540490 | 0.10587386 | . |
| rs2695009 | 12 | 19931411 | 0.105752384 | . |
| rs12215863 | 6 | 167806208 | 0.105732286 | . |
| rs4791028 | 17 | 64828574 | 0.105429175 | . |
| rs12613713 | 2 | 35446100 | 0.105283369 | . |
| rs1367027 | 14 | 37352065 | 0.105187097 | SLC25A21(0) |
| rs6865980 | 5 | 38866372 | 0.105161777 | OSMR(0) |
| rs2010922 | 2 | 171369836 | 0.10511193 | MYO3B(0) |
| rs4594727 | 4 | 7592068 | 0.105100659 | SORCS2(0) |
| rs6851301 | 4 | 140959594 | 0.105082689 | MAML3(0) |
| rs1562384 | 1 | 159562624 | 0.105066819 | . |
| rs2275317 | 1 | 246091465 | 0.105062833 | SMYD3(0) |
| rs12982370 | 19 | 10715154 | 0.105036753 | SLC44A2(0) |
| rs1047911 | 2 | 74699778 | 0.105029798 | MRPL53(0)\|=MISSENSE |
| rs1891617 | 10 | 55561645 | 0.105016703 | . |
| rs6443667 | 3 | 179406022 | 0.105010888 | USP13(0) |
| rs171373 | 1 | 13904497 | 0.104998786 | . |
| rs2041381 | 12 | 4954591 | 0.104946414 | KCNA6(0) |
| rs4649389 | 1 | 232725391 | 0.104893337 | . |
| rs2841031 | 10 | 90940003 | 0.10477967 | . |
| rs4855697 | 3 | 108867128 | 0.104760953 | FLJ22763(0) |

| | | | | |
|---|---|---|---|---|
| rs550915 | 9 | 104191392 | 0.082198027 | ALDOB(0) |
| rs9849293 | 3 | 26085604 | 0.082073186 | . |
| rs7109581 | 11 | 93180531 | 0.08201969 | . |
| rs509362 | 11 | 126036852 | 0.081907223 | . |
| rs2959356 | 18 | 23590203 | 0.081893537 | . |
| rs74918773 | 21 | 41711938 | 0.081863186 | DSCAM(0) |
| rs9596209 | 13 | 50463758 | 0.081723468 | . |
| rs925205 | 5 | 10141127 | 0.08168095 | . |
| rs66495531 | 2 | 206496074 | 0.081674975 | . |
| rs67649296 | 18 | 24171427 | 0.081588335 | KCTD1(0) |
| rs72860574 | 2 | 146078481 | 0.081551799 | . |
| rs12955326 | 18 | 566905 | 0.081537145 | . |
| rs996018 | 3 | 26044946 | 0.081384856 | . |
| rs34089886 | 4 | 37297267 | 0.081313692 | NWD2(0) |
| rs6745500 | 2 | 9640526 | 0.081262383 | ADAM17(0) |
| rs7519350 | 1 | 208712599 | 0.080966816 | . |
| rs13436699 | 5 | 10147758 | 0.08096024 | . |
| rs2521755 | 7 | 24749223 | 0.080740024 | DFNA5(0) |
| rs11142207 | 9 | 90886728 | 0.080733174 | . |
| rs4867365 | 5 | 31678198 | 0.08067672 | . |
| rs4325166 | 1 | 62506473 | 0.080577817 | INADL(0) |
| rs11703784 | 22 | 48483072 | 0.080574665 | . |
| rs8132084 | 21 | 20220881 | 0.080502318 | . |
| rs264963 | 2 | 104254564 | 0.080449015 | . |
| rs72807288 | 10 | 94042448 | 0.080367219 | CPEB3(0) |
| rs11610061 | 12 | 100050148 | 0.080360589 | ANKS1B(0) |
| rs2250606 | 17 | 80662614 | 0.080299561 | . |
| rs2642992 | 1 | 247151557 | 0.080255179 | ZNF670-ZNF695(0)\|ZNF695(0)\|=MISSENSE |
| rs9968731 | 5 | 80498455 | 0.080235512 | RASGRF2(0) |
| rs2551617 | 3 | 120367819 | 0.080232729 | HGD(0) |
| rs926244 | 1 | 3006283 | 0.080128453 | PRDM16(0) |
| rs10267099 | 7 | 87278760 | 0.080096606 | ABCB1(0)\|RUNDC3B(0) |
| rs11752519 | 6 | 159270631 | 0.080022924 | OSTCP1(0) |
| rs6708166 | 2 | 30526780 | 0.080010227 | . |
| rs2247493 | 9 | 130296484 | 0.079962022 | FAM129B(0) |
| rs12444034 | 16 | 83405551 | 0.07994533 | CDH13(0) |
| rs10260787 | 7 | 7129858 | 0.079914626 | LOC100131257(0)\|=missense |
| rs4869095 | 5 | 163063208 | 0.079908136 | . |
| rs911478 | 6 | 109262787 | 0.079874849 | ARMC2(0) |
| rs10470654 | 3 | 26055831 | 0.079710401 | . |
| rs10110604 | 8 | 16503082 | 0.079708984 | . |
| rs3748671 | 1 | 207253031 | 0.079685006 | PFKFB2(0) |
| rs72748753 | 5 | 22803278 | 0.07963518 | CDH12(0) |
| rs13226913 | 7 | 7246846 | 0.079445211 | C1GALT1(0) |
| rs12554403 | 9 | 26489225 | 0.079343024 | . |
| rs2067928 | 2 | 112850581 | 0.079324389 | TMEM87B(0) |
| rs4741226 | 9 | 12502531 | 0.079265891 | . |
| rs3006210 | 6 | 169828915 | 0.079053897 | . |
| rs2521222 | 7 | 24541524 | 0.079052413 | . |
| rs10817739 | 9 | 100308231 | 0.079012723 | TMOD1(0) |
| rs10167958 | 2 | 109921632 | 0.078985889 | SH3RF3(0) |
| rs6583867 | 10 | 94938228 | 0.078972417 | . |
| rs12535 | 2 | 159663616 | 0.078949419 | DAPL1(0) |
| rs11726701 | 4 | 132988240 | 0.07883832 | . |
| rs68055747 | 18 | 75347847 | 0.078821046 | . |
| rs4905772 | 14 | 99372389 | 0.078737463 | . |
| rs10933722 | 3 | 193954976 | 0.078707908 | . |
| rs245974 | 7 | 29295856 | 0.07862484 | CHN2(0) |
| rs17302392 | 8 | 66463308 | 0.078469967 | LINC01299(0) |
| rs2841031 | 10 | 90940003 | 0.078383816 | . |
| rs12692772 | 2 | 166373185 | 0.077852577 | CSRNP3(0) |
| rs2243955 | 9 | 130303640 | 0.077831334 | FAM129B(0) |
| rs727755 | 4 | 158724848 | 0.077668349 | . |
| rs1474249 | 2 | 165419009 | 0.077617732 | GRB14(0) |
| rs10455806 | 6 | 166146245 | 0.077613883 | . |
| rs726972 | 19 | 32034831 | 0.077540119 | . |
| rs10136340 | 14 | 70527933 | 0.077428543 | SLC8A3(0) |
| rs2783197 | 1 | 52436038 | 0.077374361 | RAB3B(0) |
| rs2183625 | 14 | 56417838 | 0.077224508 | . |
| rs17734690 | 8 | 9713196 | 0.077148675 | . |
| rs11666325 | 19 | 30907789 | 0.077101923 | ZNF536(0) |
| rs2069727 | 12 | 68548223 | 0.077099852 | . |
| rs8136304 | 22 | 27917070 | 0.076986824 | . |
| rs7895779 | 10 | 82202615 | 0.076971709 | . |
| rs111862778 | 3 | 135783796 | 0.076782448 | PPP2R3A(0) |
| rs4947040 | 6 | 110258455 | 0.076772503 | . |
| rs3892909 | 5 | 175305591 | 0.076753557 | CPLX2(0) |
| rs10105657 | 8 | 607251 | 0.076633908 | . |
| rs2540499 | 12 | 96399869 | 0.076295006 | LTA4H(0) |
| rs13208135 | 6 | 137681617 | 0.076269394 | . |
| rs7834638 | 8 | 11420295 | 0.075717119 | BLK(0)\|=missense |
| rs6918011 | 6 | 134715654 | 0.075588484 | . |
| rs60326795 | 3 | 107420674 | 0.075126881 | BBX(0) |
| rs17398598 | 1 | 52024908 | 0.075113326 | . |
| rs9878775 | 3 | 182659953 | 0.075104778 | . |
| rs6544598 | 2 | 43031054 | 0.075084947 | . |
| rs2675294 | 3 | 2751092 | 0.075048446 | CNTN4(0) |
| rs11645844 | 16 | 78408972 | 0.075042387 | WWOX(0)\|=missense |
| rs1989830 | 7 | 87205663 | 0.075004687 | ABCB1(0) |
| rs1118972 | 8 | 19086353 | 0.074979109 | LOC100128993(0) |
| rs730570 | 14 | 101142890 | 0.074883337 | . |
| rs35970366 | 7 | 87495087 | 0.074767308 | SLC25A40(0) |
| rs331941 | 4 | 143657023 | 0.074745413 | INPP4B(0) |
| rs7525615 | 1 | 229835413 | 0.074719562 | . |
| rs17536135 | 4 | 20295132 | 0.074710179 | SLIT2(0) |
| rs9856369 | 3 | 25968912 | 0.07470341 | . |
| rs13145422 | 4 | 160185669 | 0.074623886 | . |
| rs56160474 | 1 | 97544258 | 0.074622335 | DPYD(0) |
| rs17194406 | 3 | 2488631 | 0.074499511 | CNTN4(0) |
| rs1820048 | 5 | 107113395 | 0.074478387 | . |
| rs13241157 | 7 | 18860014 | 0.07436295 | HDAC9(0) |
| rs1995296 | 12 | 100082450 | 0.074359259 | ANKS1B(0) |
| rs13389867 | 2 | 160042430 | 0.074345431 | TANC1(0) |
| rs17473802 | 2 | 39752628 | 0.074261429 | LOC728730(0) |
| rs2930895 | 19 | 893080 | 0.074149393 | MED16(0) |
| rs10056443 | 5 | 88752584 | 0.074035881 | . |
| rs1074789 | 9 | 12752340 | 0.073883849 | . |
| rs1888189 | 9 | 10948272 | 0.073778858 | . |
| rs13107700 | 4 | 133111065 | 0.073635885 | . |
| rs644631 | 13 | 32098486 | 0.073598561 | . |
| rs4772114 | 13 | 99298233 | 0.073573011 | . |
| rs28621923 | 8 | 72351333 | 0.073538141 | . |
| rs5015243 | 1 | 217427461 | 0.073500411 | . |
| rs2031340 | 10 | 10586371 | 0.073453965 | . |
| rs13240660 | 7 | 4264920 | 0.073444734 | SDK1(0) |
| rs7306191 | 12 | 56080164 | 0.073428105 | ITGA7(0) |
| rs4855697 | 3 | 108867128 | 0.073379951 | FLJ22763(0) |
| rs791681 | 9 | 12522037 | 0.073375326 | . |

**Table S15. Profile of ACMG reportable variants in the TYC population**

| Ind | *MUTYH* _1_45331223 | *TTR* _18_31598602 |
|---|---|---|
| Qiang_Xiaoyuan | 0/0 | 0/0 |
| Tibetan_Baima_Guoyuan | 0/0 | 0/0 |
| Qiang_Xiaoyuan | 0/0 | 0/0 |
| Tibetan_Amdo_Dalu | 0/0 | 0/0 |
| Qiang_Matou | 0/0 | 0/0 |
| Qiang_Gaoping | 0/0 | 0/0 |
| Qiang_Gaoping | 0/0 | 0/0 |
| Qiang_Gaoping | 0/0 | 0/0 |
| Qiang_Gaoping | 0/0 | 0/0 |
| Tibetan_Baima_Wujiao | 0/0 | 0/0 |
| Qiang_Gaoping | 0/0 | 0/0 |
| Tibetan_Baima_Wujiao | 0/0 | 0/0 |
| Tibetan_Baima_Guoyuan | 0/0 | 0/0 |
| Qiang_Xiaoyuan | 0/0 | 0/0 |
| Qiang_Liujia | 0/0 | 0/0 |
| Qiang_Liujia | 0/0 | 0/0 |
| Tibetan_Baima_Guoyuan | 0/0 | 0/0 |
| Tibetan_Baima_Wujiao | 0/0 | 0/0 |
| Qiang_Liujia | ./. | 0/0 |
| Qiang_Matou | 0/0 | 0/0 |
| Qiang_Matou | 0/0 | 0/0 |
| Qiang_Gaoping | 0/0 | 0/0 |
| Tibetan_Amdo_Dalu | 0/0 | 0/0 |
| Qiang_Gaoping | 0/1 | 0/0 |
| Qiang_Liujia | 0/0 | 0/0 |
| Qiang_Matou | 0/0 | 0/0 |
| Tibetan_Amdo_Dalu | 0/0 | ./. |
| Tibetan_Baima_Guoyuan | 0/0 | 0/0 |
| Qiang_Gaoping | 0/0 | 0/0 |
| Tibetan_Baima_Guoyuan | 0/0 | 0/0 |
| Qiang_Matou | 0/0 | 0/0 |
| Tibetan_Baima_Guoyuan | 0/0 | 0/0 |
| Qiang_Liujia | 0/0 | 0/0 |
| Tibetan_Amdo_Dalu | ./. | 0/0 |
| Qiang_Liujia | 0/0 | 0/0 |
| Tibetan_Amdo_Dalu | 0/0 | 0/0 |
| Qiang_Matou | 0/0 | 0/0 |
| Tibetan_Baima_Wujiao | ./. | 0/0 |
| Qiang_Xiaoyuan | 0/0 | 0/0 |
| Qiang_Matou | 0/0 | 0/0 |
| Qiang_Matou | 0/1 | ./. |
| Qiang_Matou | 0/0 | 0/0 |
| Tibetan_Amdo_Dalu | 0/0 | 0/0 |
| Tibetan_Amdo_Dalu | 0/0 | 0/0 |
| Qiang_Gaoping | 0/0 | 0/0 |
| Tibetan_Baima_Wujiao | 0/0 | 0/0 |
| Tibetan_Amdo_Dalu | 0/0 | 0/0 |
| Tibetan_Baima_Guoyuan | 0/0 | 0/0 |
| Tibetan_Baima_Guoyuan | ./. | 0/0 |
| Tibetan_Baima_Guoyuan | ./. | 0/0 |
| Qiang_Matou | 0/0 | 0/0 |
| Tibetan_Baima_Guoyuan | 0/0 | 0/0 |
| Tibetan_Baima_Wujiao | 0/0 | 0/0 |
| Tibetan_Baima_Guoyuan | 0/0 | ./. |
| Qiang_Liujia | 0/0 | 0/0 |
| Qiang_Matou | ./. | 0/0 |
| Qiang_Matou | 0/0 | 0/0 |

| | | |
|---|---|---|
| Qiang_Xiaoyuan | ./. | 0/0 |
| Qiang_Matou | 0/0 | 0/0 |
| Qiang_Gaoping | 0/0 | 0/0 |
| Tibetan_Baima_Wujiao | 0/0 | 0/0 |
| Tibetan_Amdo_Dalu | 0/0 | 0/0 |
| Qiang_Gaoping | 0/0 | 0/0 |
| Qiang_Matou | 0/0 | 0/0 |
| Qiang_Xiaoyuan | 0/0 | 0/0 |
| Qiang_Xiaoyuan | 0/0 | 0/0 |
| Tibetan_Baima_Wujiao | 0/0 | 0/0 |
| Tibetan_Baima_Guoyuan | 0/0 | ./. |
| Tibetan_Amdo_Dalu | ./. | ./. |
| Tibetan_Amdo_Dalu | 0/0 | 0/0 |
| Tibetan_Amdo_Dalu | 0/0 | 0/0 |
| Tibetan_Amdo_Dalu | 0/0 | 0/0 |
| Tibetan_Amdo_Dalu | 0/0 | 0/0 |
| Qiang_Xiaoyuan | 0/0 | 0/0 |
| Qiang_Matou | ./. | 0/0 |
| Qiang_Xiaoyuan | ./. | ./. |
| Tibetan_Amdo_Dalu | 0/0 | 0/0 |
| Qiang_Xiaoyuan | 0/0 | 0/0 |
| Qiang_Xiaoyuan | 0/0 | 0/0 |
| Tibetan_Baima_Guoyuan | 0/0 | 0/0 |
| Tibetan_Baima_Guoyuan | ./. | 0/0 |
| Tibetan_Amdo_Dalu | 0/0 | 0/0 |
| Wa_Cangyuan | 0/0 | 0/0 |
| Hani_Honghe | 0/0 | 0/0 |
| Pumi_Lanping | 0/0 | 0/0 |
| Blang_Menghai | 0/0 | 0/0 |
| Blang_Menghai | 0/0 | 0/0 |
| Wa_Cangyuan | 0/0 | 0/0 |
| Bai_Heqing | 0/0 | 0/0 |
| Pumi_Lanping | 0/0 | 0/0 |
| Bai_Heqing | 0/0 | 0/0 |
| Hani_Honghe | 0/0 | 0/0 |
| Hani_Honghe | 0/0 | 0/0 |
| Bai_Heqing | 0/0 | 0/0 |
| Wa_Cangyuan | 0/0 | 0/0 |
| Bai_Heqing | 0/0 | 0/0 |
| Wa_Cangyuan | 0/0 | 0/0 |
| Lahu_Lancang | 0/0 | 0/0 |
| Pumi_Lanping | 0/0 | 0/0 |
| Wa_Cangyuan | 0/0 | 0/0 |
| Wa_Cangyuan | 0/0 | 0/0 |
| Bai_Heqing | 0/0 | 0/0 |
| Bai_Heqing | 0/0 | 0/0 |
| Pumi_Lanping | 0/0 | 0/0 |
| Lahu_Lancang | 0/0 | 0/0 |
| Blang_Menghai | 0/0 | 0/0 |
| Pumi_Lanping | 0/0 | 0/0 |
| Hani_Honghe | 0/0 | 0/0 |
| Blang_Menghai | 0/0 | 0/0 |
| Bai_Heqing | 0/0 | 0/0 |
| Blang_Menghai | 0/0 | ./. |
| Blang_Menghai | 0/0 | 0/0 |
| Blang_Menghai | 0/0 | 0/0 |
| Bai_Heqing | 0/0 | 0/0 |
| Hani_Honghe | 0/0 | 0/0 |
| Lahu_Lancang | 0/0 | 0/0 |

| | | |
|---|---|---|
| Hani_Honghe | 0/0 | 0/0 |
| Pumi_Lanping | 0/0 | 0/0 |
| Wa_Cangyuan | 0/0 | 0/0 |
| Hani_Honghe | 0/0 | 0/0 |
| Pumi_Lanping | 0/0 | 0/0 |
| Pumi_Lanping | 0/0 | 0/0 |
| Lahu_Lancang | 0/0 | 0/1 |
| Bai_Heqing | 0/0 | 0/0 |
| Wa_Cangyuan | 0/0 | 0/0 |
| Lahu_Lancang | 0/0 | 0/0 |
| Bai_Heqing | 0/0 | 0/0 |
| Hani_Honghe | 0/0 | 0/0 |
| Hani_Honghe | 0/0 | 0/0 |
| Bai_Heqing | 0/0 | 0/0 |
| Wa_Cangyuan | 0/0 | 0/0 |
| Hani_Honghe | 0/0 | 0/0 |
| Blang_Menghai | 0/0 | 0/0 |
| Blang_Menghai | 0/0 | 0/0 |
| Bai_Heqing | 0/0 | 0/0 |
| Pumi_Lanping | 0/0 | 0/0 |
| Bai_Heqing | 0/0 | 0/0 |
| Pumi_Lanping | 0/0 | 0/0 |
| Blang_Menghai | 0/0 | 0/0 |
| Bai_Heqing | 0/0 | 0/0 |
| Bai_Heqing | 0/0 | 0/0 |
| Hani_Honghe | 0/0 | 0/0 |
| Hani_Honghe | 0/0 | 0/0 |
| Pumi_Lanping | 0/0 | 0/0 |
| Wa_Cangyuan | 0/0 | 0/0 |
| Pumi_Lanping | 0/0 | 0/0 |
| Blang_Menghai | 0/0 | 0/0 |

**Table S16. The catalog of ClinVar-annotated pathogenic or likely pathogenic variants in the TYC population**

| SNP | Gene | Disease | Allele count | Frequency |
|---|---|---|---|---|
| 1_45331223 | MUTYH | Immune-related diseases | 2 | 0.680272109 |
| 1_94005469 | ABCA4 | other | 1 | 0.340136054 |
| 1_152307622 | FLG | other | 1 | 0.340136054 |
| 1_152309169 | FLG | other | 8 | 2.721088435 |
| 1_152309170 | FLG | other | 9 | 3.06122449 |
| 1_155235217 | GBA | Immune-related diseases | 1 | 0.340136054 |
| 1_179575915 | NPHS2 | Urological diseases | 27 | 9.183673469 |
| 6_32039081 | CYP21A2 | Metabolic diseases | 6 | 2.040816327 |
| 6_32039426 | CYP21A2 | Metabolic diseases | 132 | 44.89795918 |
| 6_32040110 | CYP21A2 | Metabolic diseases | 1 | 0.340136054 |
| 7_66994286 | SBDS | Gastrointestinal diseases | 1 | 0.340136054 |
| 7_87452957 | ABCB4 | Gastrointestinal diseases | 3 | 1.020408163 |
| 7_142752899 | PRSS1 | Gastrointestinal diseases | 3 | 1.020408163 |
| 12_21222355 | SLCO1B1 | other | 2 | 0.680272109 |
| 16_16202100 | ABCC6 | Cardiovascular diseases | 3 | 1.020408163 |
| 16_16219555 | ABCC6 | Cardiovascular diseases | 1 | 0.340136054 |
| 19_40396401 | PRX | Neurological diseases | 37 | 12.58503401 |
| 19_49061381 | NTF4 | Neurological diseases | 1 | 0.340136054 |
| 20_62952855 | SLC17A9 | other | 1 | 0.340136054 |

**Table S17. The count of adverse drug reaction-related allele in TYC populations**

| chr | snp | ANNOT | count |
|---|---|---|---|
| 1 | 97234991 | DPYD | 1 |
| 1 | 97305364 | DPYD | 2 |
| 1 | 97305364 | DPYD | 2 |
| 1 | 97450068 | DPYD | 5 |
| 1 | 97515839 | DPYD | 21 |
| 1 | 97883329 | DPYD | 12 |
| 2 | 233759924 | UGT1A3 | 11 |
| 2 | 233760498 | UGT1A1 | 9 |
| 6 | 18130687 | TPMT | 6 |
| 6 | 18130687 | TPMT | 6 |
| 6 | 29945521 | HLA-A | 3 |
| 7 | 99672916 | CYP3A5 | 48 |
| 7 | 117548796 | CFTR | 1 |
| 10 | 94775367 | CYP2C19 | 13 |
| 10 | 94780653 | CYP2C19 | 10 |
| 10 | 94781859 | CYP2C19 | 10 |
| 10 | 94781859 | CYP2C19 | 10 |
| 10 | 94842866 | CYP2C19 | 61 |
| 10 | 94981296 | CYP2C9 | 1 |
| 12 | 21176804 | SLCO1B1 | 45 |
| 12 | 21176868 | SLCO1B1 | 1 |
| 12 | 21178615 | SLCO1B1 | 2 |
| 12 | 21239042 | SLCO1B1 | 1 |
| 16 | 31096368 | VKORC1 | 55 |
| 19 | 15878920 | CYP4F2 | 9 |
| 19 | 15879621 | CYP4F2 | 14 |
| 19 | 15897578 | CYP4F2 | 7 |
| 19 | 38492540 | RYR1 | 1 |
| 19 | 38499696 | RYR1 | 5 |
| 19 | 38499705 | RYR1 | 5 |
| 19 | 38504870 | RYR1 | 21 |
| 19 | 38505933 | RYR1 | 2 |
| 19 | 38534726 | RYR1 | 3 |
| 19 | 39248147 | IFNL4 | 3 |
| 19 | 40991369 | CYP2B6 | 2 |
| 19 | 41006936 | CYP2B6 | 11 |
| 19 | 41009358 | CYP2B6 | 5 |
| 19 | 41016810 | CYP2B6 | 1 |
| 19 | 41016810 | CYP2B6 | 1 |
| 22 | 42127803 | CYP2D6 | 4 |
| 22 | 42127853 | CYP2D6 | 2 |
| 22 | 42127941 | CYP2D6 | 10 |
| 22 | 42128181 | CYP2D6 | 1 |
| 22 | 42128185 | CYP2D6 | 1 |
| 22 | 42129033 | CYP2D6 | 1 |
| 22 | 42129033 | CYP2D6 | 1 |
| 22 | 42129130 | CYP2D6 | 41 |
| 22 | 42129180 | CYP2D6 | 1 |
| 22 | 42130692 | CYP2D6 | 31 |

**Table S18. Potential trait-related loci within archaic-introgressed segments**

| Neanderthals | Pos | Chr | Gene | Traits | Denisovans | Pos | Chr | Gene | Traits |
|---|---|---|---|---|---|---|---|---|---|
| rs2639453 | 11982544 | 1 | KIAA2013 | Serum folate levels | rs11805515 | 10767785 | 1 | CASZ1 | Vaginal microbiome MetaCyc pathway (PWY-7371\|1,4-dihydroxy-6-naphthoate biosynthesis II) |
| rs1208984 | 11995176 | 1 | PLOD1 | Eosinophil counts | rs284247 | 10774856 | 1 | CASZ1 | Uterine fibroids |
| rs2336379 | 12008863 | 1 | PLOD1 | Facial wrinkles (crow's feet) | rs706007 | 10780727 | 1 | CASZ1 | QT interval |
| rs58495963 | 12012980 | 1 | PLOD1 | Serum arsenic levels | rs284254 | 10786314 | 1 | CASZ1 | QT interval |
| rs2273291 | 12025353 | 1 | PLOD1 | Moyamoya disease | rs284278 | 10790536 | 1 | CASZ1 | Current medication use (UKB data field 20003) |
| rs6678553 | 12026090 | 1 | PLOD1 | Platelet count | rs284277 | 10790797 | 1 | CASZ1 | Atrial fibrillation |
| rs3818157 | 12027308 | 1 | PLOD1 | Platelet count | rs17035646 | 10796547 | 1 | CASZ1 | Systolic blood pressure |
| rs2236055 | 12042261 | 1 | MFN2 | Mitochondrial DNA copy number (adjusted) | rs880315 | 10796866 | 1 | CASZ1 | Urinary albumin-to-creatinine ratio |
| rs4846082 | 12042443 | 1 | MFN2 | Plateletcrit | rs682178 | 10798552 | 1 | CASZ1 | Low density lipoprotein cholesterol levels |
| rs4845891 | 12042506 | 1 | MFN2 | BRCA1-A complex subunit Abraxas levels | rs205484 | 10815421 | 1 | CASZ1 | Metabolite levels |
| rs4240897 | 12042755 | 1 | MFN2 | Tuberculosis | rs2073017 | 10822808 | 1 | CASZ1 | Cataracts |
| rs3766744 | 12043717 | 1 | MFN2 | Mitochondrial DNA copy number | rs56218493 | 22902772 | 1 | EPHA8 | Serum alkaline phosphatase levels |
| rs1474868 | 12044164 | 1 | MFN2 | Mitochondrial DNA levels | rs75903500 | 22911195 | 1 | EPHA8 | Serum alkaline phosphatase levels |
| rs2336384 | 12046063 | 1 | MFN2 | Platelet count | rs15940 | 22970642 | 1 | C1QC | Serum levels of protein C1QC |
| rs730123 | 12070292 | 1 | MFN2 | Plasma homocysteine levels | rs12073436 | 22970908 | 1 | C1QC | Complement C1q subcomponent subunit C levels |
| rs61776548 | 12091024 | 1 | MIIP | Atopic dermatitis | rs631090 | 22986403 | 1 | C1QB | Complement C1q subcomponent subunit C levels |
| rs4491070 | 12130051 | 1 | TNFRSF8 | Atopic asthma | rs623607 | 22986716 | 1 | C1QB | Sphingomyelin (d38:2) levels |
| rs35408971 | 19180369 | 1 | TAS1R2 | Healthy aging | rs734445 | 23044828 | 1 | EPHB2 | Severe COVID-19 infection |
| rs28530674 | 19234134 | 1 | IFFO2 | Eosinophilic esophagitis | rs6687487 | 23061551 | 1 | EPHB2 | Blood protein levels |
| rs2024724 | 19237107 | 1 | IFFO2 | Plasma parathyroid hormone levels | rs12026894 | 245431303 | 1 | KIF26B | Insomnia in migraine with aura |
| rs9253 | 37958813 | 1 | MEAF6 | Hemostatic factors and hematological phenotypes | rs6665809 | 245462248 | 1 | KIF26B | Estimated glomerular filtration rate (creatinine) |
| rs4653294 | 37963541 | 1 | MEAF6 | Height | rs11692723 | 33467055 | 2 | LTBP1 | Waist circumference adjusted for body mass index |
| rs72657787 | 38010243 | 1 | SNIP1 | Educational attainment | rs3769528 | 33471192 | 2 | LTBP1 | Height |
| rs11577094 | 38026600 | 1 | DNALI1 | Body mass index | rs817534 | 33473564 | 2 | LTBP1 | Height |
| rs4653301 | 38028374 | 1 | DNALI1 | Intraocular pressure | rs12468099 | 33488419 | 2 | LTBP1 | G_Coprococcus prevalence |
| rs11264074 | 38052061 | 1 | GNL2 | Blood pressure | rs3754829 | 33499107 | 2 | LTBP1 | Waist circumference adjusted for body mass index |
| rs2294875 | 38052445 | 1 | GNL2 | Lung function (FEV1/FVC) | rs6739725 | 33506357 | 2 | LTBP1 | Height |
| rs36043533 | 38079517 | 1 | RSPO1 | R-spondin-1 levels | rs78692521 | 33523943 | 2 | LTBP1 | L1-L4 bone mineral density x serum urate levels interaction |
| rs12039431 | 38082122 | 1 | RSPO1 | Epithelial ovarian cancer | rs3754830 | 33524956 | 2 | LTBP1 | Height |
| rs4074961 | 38092723 | 1 | RSPO1 | Intraocular pressure | rs41464348 | 33527299 | 2 | LTBP1 | Height |
| rs58722170 | 38096421 | 1 | RSPO1 | Epithelial ovarian cancer | rs79518802 | 33531033 | 2 | LTBP1 | Vaginal microbiome MetaCyc pathway (PWY1G-0\|mycothiol biosynthesis) |
| rs10789491 | 47179310 | 1 | EFCAB14 | Response to hepatitis C treatment | rs11903480 | 33550769 | 2 | LTBP1 | Central corneal thickness |
| rs3126085 | 152300817 | 1 | FLG-AS1 | Atopic dermatitis | rs13427209 | 33567416 | 2 | LTBP1 | Height |
| rs1933064 | 152301576 | 1 | FLG-AS1 | Serum 25-Hydroxyvitamin D levels | rs5024713 | 33568137 | 2 | LTBP1 | Height |
| rs3120665 | 152316590 | 1 | FLG-AS1 | Personality dimensions | rs11895736 | 33573090 | 2 | LTBP1 | Overall survival in cancer x bevacizumab treatment interaction |
| rs3120667 | 152318161 | 1 | FLG-AS1 | Eating disorders | rs7557776 | 33577792 | 2 | LTBP1 | Height |
| rs76285340 | 152329529 | 1 | FLG-AS1 | Systemic sclerosis | rs34447926 | 33588869 | 2 | LTBP1 | Intraocular pressure |
| rs1123015 | 162042313 | 1 | NOS1AP | Heel bone mineral density | rs2595205 | 46194216 | 2 | PRKCE | Monocyte count |
| rs1415259 | 162085309 | 1 | NOS1AP | Electrocardiographic traits | rs13396424 | 46205039 | 2 | PRKCE | Metabolite levels (X-11787) |
| rs10494366 | 162085685 | 1 | NOS1AP | Electrocardiographic traits | rs1987070 | 46237446 | 2 | PRKCE | Red blood cell count |
| rs4620512 | 162104303 | 1 | NOS1AP | Mean platelet volume | rs6544870 | 46269969 | 2 | PRKCE | S.enteritidis induced IL-1B level |
| rs4657153 | 162108015 | 1 | NOS1AP | Retinal vascular fractal density | rs78448334 | 46274444 | 2 | PRKCE | Impulse control disorders in Parkinson's disease |
| rs6677140 | 162109096 | 1 | NOS1AP | QRS duration | rs11682804 | 46275648 | 2 | PRKCE | Two-hour glucose in pregnancy (gestational week 30-32) |
| rs16857031 | 162112910 | 1 | NOS1AP | QT interval | rs12712961 | 46278720 | 2 | PRKCE | Corpus callosum posterior volume |
| rs59852339 | 162112966 | 1 | NOS1AP | Electrocardiogram morphology (amplitude at temporal datap | rs34496377 | 46307069 | 2 | PRKCE | Childhood ear infection |
| rs78344479 | 162164883 | 1 | NOS1AP | Mean platelet volume during second trimester of pregnancy | rs1868274 | 46309262 | 2 | PRKCE | Hemoglobin |
| rs12567315 | 162166646 | 1 | NOS1AP | Mean reticulocyte volume | rs3768758 | 46320002 | 2 | PRKCE | Height |
| rs4657172 | 162179632 | 1 | NOS1AP | QT interval | rs3768751 | 46346716 | 2 | PRKCE | Hematological and biochemical traits |
| rs3934467 | 162182677 | 1 | NOS1AP | QT interval | rs12623399 | 46349271 | 2 | PRKCE | Red blood cell count |
| rs73017364 | 162184746 | 1 | NOS1AP | QT interval | rs17799476 | 46352326 | 2 | PRKCE | Hematocrit |
| rs4657175 | 162195738 | 1 | NOS1AP | QT interval | rs10495928 | 46353166 | 2 | PRKCE | Hematocrit |
| rs12737539 | 162198429 | 1 | NOS1AP | QT dynamics during recovery from exercise | rs10168349 | 46360907 | 2 | PRKCE | Red blood cell count |
| rs6658646 | 162212765 | 1 | NOS1AP | Granulocyte percentage of myeloid white cells | rs2053797 | 46370892 | 2 | PRKCE | Red blood cell count |
| rs11584803 | 162213769 | 1 | NOS1AP | Monocyte count | rs17034641 | 46372644 | 2 | PRKCE | Hematocrit |
| rs12742393 | 162224586 | 1 | NOS1AP | DNA methylation variation (age effect) | rs4953320 | 46374529 | 2 | PRKCE | Blood pressure (pleiotropy model 1 DBP adjusted for estimated causal effects x SBP) |
| rs4657180 | 162247738 | 1 | NOS1AP | Alanine aminotransferase levels in non-alcoholic fatty liver ( | rs72808780 | 46409007 | 2 | PRKCE | Red blood cell count |
| rs10800409 | 162260715 | 1 | NOS1AP | Height | rs2594495 | 46409466 | 2 | PRKCE | Carotenoid levels (alpha-carotene) |
| rs57561731 | 162468586 | 1 | UHMK1 | Cervical cancer | rs281508 | 46411207 | 2 | PRKCE | Response to zileuton treatment in asthma (FEV1 change interaction) |
| rs7513662 | 162480145 | 1 | UHMK1 | OTU99_123 (Phascolarctobacterium) prevalence | rs11125068 | 46527816 | 2 | EPAS1 | Kidney cancer |
| rs6702820 | 162603881 | 1 | DDR2 | Height | rs11894252 | 46533376 | 2 | EPAS1 | Renal cell carcinoma |
| rs7512080 | 162609738 | 1 | DDR2 | Choroidal melanoma | rs11684885 | 46533552 | 2 | EPAS1 | QRS duration |
| rs6684509 | 162618152 | 1 | DDR2 | Height | rs4952818 | 46537056 | 2 | EPAS1 | Estimated glomerular filtration rate (creatinine) |
| rs75068199 | 162630046 | 1 | DDR2 | Cervical cancer | rs7579899 | 46537604 | 2 | EPAS1 | Renal cell carcinoma |
| rs10799867 | 162658483 | 1 | DDR2 | Height | rs17034950 | 46538794 | 2 | EPAS1 | JT interval |
| rs6697469 | 162658773 | 1 | DDR2 | Height | rs2034327 | 46549040 | 2 | EPAS1 | Kidney cancer |
| rs4411113 | 162668008 | 1 | DDR2 | Glaucoma (primary open-angle) | rs7582701 | 46550769 | 2 | EPAS1 | IgG glycosylation |
| rs72711557 | 162669776 | 1 | DDR2 | Dupuytren's disease | rs11125071 | 46554533 | 2 | EPAS1 | Red blood cell count |
| rs17486566 | 162670464 | 1 | DDR2 | Primary open angle glaucoma (multi-trait analysis) | rs847151 | 176964904 | 2 | HOXD12 | Height |
| rs17433710 | 162672011 | 1 | DDR2 | Dupuytren's disease | rs863678 | 176974104 | 2 | HOXD11 | Hematocrit |
| rs2806424 | 162691459 | 1 | DDR2 | White blood cell count | rs711830 | 177037311 | 2 | HOXD3 | Low-grade serous and serous borderline ovarian cancer |
| rs12116643 | 196973183 | 1 | CFHR5 | Blood protein levels | rs13028842 | 177042261 | 2 | HOXD-AS1 | Systolic blood pressure |
| rs10922153 | 196978615 | 1 | CFHR5 | Cerebrospinal fluid biomarker levels | rs2072590 | 177042633 | 2 | HOXD-AS1 | Ovarian cancer |
| rs10754210 | 197012111 | 1 | F13B | Childhood body mass index | rs6755777 | 177043226 | 2 | HOXD-AS1 | Epithelial ovarian cancer |
| rs2990510 | 197020658 | 1 | F13B | Erectile dysfunction in type 1 diabetes | rs6710142 | 177054769 | 2 | HOXD1 | Urate levels |
| rs10801586 | 197021189 | 1 | F13B | Complement factor H-related protein 2 levels | rs12466158 | 228373615 | 2 | AGFG1 | Height |
| rs6003 | 197031021 | 1 | F13B | Complement factor H-related protein 5 levels | rs10189795 | 228378861 | 2 | AGFG1 | Cortical surface area |
| rs2298882 | 197034826 | 1 | F13B | Coagulation factor XIII B chain levels | rs6731443 | 228397566 | 2 | AGFG1 | Airway hyperresponsiveness |

| | | | | | | | | | |
|---|---|---|---|---|---|---|---|---|---|
| rs10922162 | 197070776 | 1 | ASPM | End-stage coagulation | rs3116162 | 232860077 | 2 | DIS3L2 | Waist circumference adjusted for BMI in active individuals |
| rs12137359 | 197129981 | 1 | ZBTB41 | Low density lipoprotein cholesterol levels | rs73995038 | 232860323 | 2 | DIS3L2 | Lung function (FVC) and body mass index |
| rs10429910 | 197141253 | 1 | ZBTB41 | Systolic blood pressure | rs3116225 | 232880845 | 2 | DIS3L2 | Height |
| rs10922177 | 197153767 | 1 | ZBTB41 | Complement factor H-related protein 2 levels | rs1453867 | 232917899 | 2 | DIS3L2 | Chronic back pain |
| rs7410943 | 197155021 | 1 | ZBTB41 | End-stage coagulation | rs3116240 | 232920430 | 2 | DIS3L2 | Height |
| rs10922179 | 197159347 | 1 | ZBTB41 | Total cholesterol levels | rs76803230 | 232928700 | 2 | DIS3L2 | Height |
| rs10754218 | 197165014 | 1 | ZBTB41 | Hemopexin levels | rs3100596 | 232978214 | 2 | DIS3L2 | FEV1 |
| rs7516246 | 197165712 | 1 | ZBTB41 | Serum levels of protein F13B | rs13412942 | 232981427 | 2 | DIS3L2 | Height |
| rs10754220 | 197244290 | 1 | CRB1 | Body mass index | rs3103267 | 232988582 | 2 | DIS3L2 | Height |
| rs1009188 | 197257090 | 1 | CRB1 | Body mass index | rs3116168 | 232989831 | 2 | DIS3L2 | Height |
| rs1337005 | 197265641 | 1 | CRB1 | Complement factor H-related protein 2 levels | rs3103261 | 232993574 | 2 | DIS3L2 | Whole body fat free mass (UKB data field 23101) |
| rs1764625 | 197299000 | 1 | CRB1 | Complement factor H-related protein 5 levels | rs10498260 | 233056621 | 2 | DIS3L2 | Height |
| rs2242199 | 203465396 | 1 | OPTC | Ankyrin-2 levels | rs3116201 | 233074205 | 2 | DIS3L2 | Appendicular lean mass |
| rs6661843 | 203625651 | 1 | ATP2B4 | Composite immunoglobulin trait (IgA x IgG/IgM) | rs7574893 | 233090338 | 2 | DIS3L2 | Height |
| rs12030888 | 208410364 | 1 | PLXNA2 | General cognitive ability | rs12466581 | 233093315 | 2 | DIS3L2 | Hip circumference adjusted for BMI |
| rs117462017 | 212587381 | 1 | TMEM206 | Response to risperidone in schizophrenia | rs6437055 | 233094365 | 2 | DIS3L2 | Waist circumference adjusted for body mass index |
| rs1105899 | 212758604 | 1 | ATF3 | TestASV_32 (Ruminococcaceae) prevalence | rs882098 | 233108857 | 2 | DIS3L2 | Height |
| rs12564392 | 212775529 | 1 | ATF3 | Facial morphology (D223) | rs2101435 | 233115158 | 2 | DIS3L2 | Waist circumference adjusted for body mass index |
| rs3123543 | 212790686 | 1 | ATF3 | Blood cell traits (multivariate analysis) | rs2853360 | 233120444 | 2 | DIS3L2 | Multi-trait sex score |
| rs3122713 | 212799870 | 1 | FAM71A | Blood protein levels | rs2633266 | 233120758 | 2 | DIS3L2 | Height |
| rs12026832 | 215214716 | 1 | KCNK2 | Height | rs2853361 | 233120923 | 2 | DIS3L2 | Tibia length to body height ratio |
| rs12088480 | 215246836 | 1 | KCNK2 | Height | rs2853362 | 233122412 | 2 | DIS3L2 | Multi-trait sex score |
| rs10127656 | 215296961 | 1 | KCNK2 | Height | rs2633261 | 233131825 | 2 | DIS3L2 | Whole body fat free mass (UKB data field 23101) |
| rs443071 | 220925241 | 1 | 2-Mar | Facial morphology (D481) | rs13020779 | 233149600 | 2 | DIS3L2 | Multi-trait sex score |
| rs6657944 | 220951135 | 1 | 2-Mar | Total cholesterol levels | rs73092108 | 233153745 | 2 | DIS3L2 | Body surface area |
| rs1389742 | 220967683 | 1 | 1-Mar | LDL (standard GWA) | rs6717918 | 233155110 | 2 | DIS3L2 | Height |
| rs2642438 | 220970028 | 1 | 1-Mar | Total cholesterol levels | rs6718438 | 233155566 | 2 | DIS3L2 | Height |
| rs2642439 | 220970499 | 1 | 1-Mar | 3-Hydroxybutyrate levels | rs4973517 | 233161818 | 2 | DIS3L2 | Hip circumference adjusted for BMI |
| rs2807835 | 220971048 | 1 | 1-Mar | Total bilirubin levels | rs67163108 | 233183967 | 2 | DIS3L2 | Weight |
| rs61830266 | 220973432 | 1 | 1-Mar | High density lipoprotein cholesterol levels | rs3748967 | 233198373 | 2 | DIS3L2 | Height |
| rs2642442 | 220973563 | 1 | 1-Mar | Total cholesterol levels | rs3811578 | 233201328 | 2 | DIS3L2 | Height |
| rs1874124 | 220975337 | 1 | 1-Mar | Total cholesterol levels | rs1550094 | 233385396 | 2 | PRSS56 | Myopia |
| rs6693017 | 220977333 | 1 | 1-Mar | Monocyte early outgrowth colony forming units | rs2853447 | 233389918 | 2 | PRSS56 | Vertical cup-disc ratio |
| rs17596144 | 220978645 | 1 | 1-Mar | Triacylglycerol (54:5) levels | rs2245601 | 233390937 | 2 | CHRND | Corneal curvature |
| rs34621709 | 220980078 | 1 | 1-Mar | Triacylglycerol (53:3) levels | rs1881492 | 233406998 | 2 | CHRNG | Refractive error |
| rs17649913 | 220983402 | 1 | 1-Mar | Waist circumference adjusted for body mass index | rs1190452 | 233417552 | 2 | EIF4E2 | Lymphocyte count |
| rs11118607 | 220984364 | 1 | 1-Mar | LDL cholesterol levels | rs1190449 | 233418920 | 2 | EIF4E2 | Height |
| rs12073837 | 221010205 | 1 | HLX-AS1 | F-cell distribution in sickle cell anaemia | rs1307667 | 233419972 | 2 | EIF4E2 | Height |
| rs17008917 | 221011317 | 1 | HLX-AS1 | Height | rs1656402 | 233426526 | 2 | EIF4E2 | Non-small cell lung cancer (survival) |
| rs17163564 | 223087894 | 1 | DISP1 | Height | rs13393800 | 233442091 | 2 | EIF4E2 | Height |
| rs2609383 | 223116472 | 1 | DISP1 | Height | rs9799721 | 8589436 | 4 | GPR78 | Height |
| rs5744168 | 223285200 | 1 | TLR5 | Monocyte percentage of white cells | rs3775887 | 8591528 | 4 | GPR78 | Lung function (FVC) |
| rs1100886 | 223292632 | 1 | TLR5 | Height | rs4696855 | 8597140 | 4 | CPZ | Height |
| rs4661250 | 223399328 | 1 | SUSD4 | Fasting plasma glucose in prediabetes | rs3756173 | 8598698 | 4 | CPZ | Height |
| rs851140 | 223405374 | 1 | SUSD4 | Height | rs11723641 | 8601298 | 4 | CPZ | Height |
| rs17487717 | 223486421 | 1 | SUSD4 | N6-acetyllysine levels in elite athletes | rs3756171 | 8602225 | 4 | CPZ | Refractive error |
| rs17520554 | 226759861 | 1 | C1orf95 | High-sensitivity cardiac troponin T levels | rs2241069 | 8602798 | 4 | CPZ | Body fat distribution (arm fat ratio) |
| rs41314284 | 226790001 | 1 | C1orf95 | General cognitive ability | rs2302580 | 8608634 | 4 | CPZ | Height |
| rs16846346 | 226838508 | 1 | ITPKB | Alzheimer's disease polygenic risk score (upper quantile vs | rs62288581 | 8609848 | 4 | CPZ | Appendicular lean mass |
| rs16846351 | 226846712 | 1 | ITPKB | Parkinson's disease | rs3796727 | 8613026 | 4 | CPZ | Sporadic neuroblastoma |
| rs10916025 | 226855889 | 1 | ITPKB | Subcutaneous adipose tissue | rs4704171 | 74368254 | 5 | ANKRD31 | Alzheimer's disease |
| rs3820640 | 226868918 | 1 | ITPKB | Number of decayed, missing and filled tooth surfaces or us | rs10038095 | 74637711 | 5 | HMGCR | Low density lipoprotein cholesterol levels |
| rs4653767 | 226916078 | 1 | ITPKB | Parkinson's disease | rs6453131 | 74644706 | 5 | HMGCR | Total lipids in very small VLDL (UKB data field 23517) |
| rs708775 | 226923264 | 1 | ITPKB | Height | rs12654264 | 74648603 | 5 | HMGCR | Cholesterol |
| rs6667260 | 226923938 | 1 | ITPKB | Refractive error | rs3846662 | 74651084 | 5 | HMGCR | Cholesterol, total |
| rs10489609 | 231470650 | 1 | EXOC8 | Hemoglobin concentration | rs3846663 | 74655726 | 5 | HMGCR | LDL cholesterol levels x short total sleep time interaction (2df test) |
| rs2437150 | 231488524 | 1 | SPRTN | Hematocrit | rs12916 | 74656539 | 5 | HMGCR | Total cholesterol levels |
| rs973253 | 231520874 | 1 | EGLN1 | Cutaneous mastocytosis | rs17562686 | 74668589 | 5 | COL4A3BP | Height |
| rs508618 | 231532312 | 1 | EGLN1 | Red blood cell count | rs17671845 | 74701766 | 5 | COL4A3BP | Metabolite levels |
| rs681828 | 233762901 | 1 | KCNK1 | Obsessive-compulsive symptoms | rs10067900 | 74706661 | 5 | COL4A3BP | Low density lipoprotein cholesterol levels |
| rs701230 | 233771275 | 1 | KCNK1 | Thrombin-antithrombin complex levels in ischemic stroke | rs9791146 | 74740950 | 5 | COL4A3BP | Collagen type IV alpha-3-binding protein levels |
| rs701231 | 233771306 | 1 | KCNK1 | Vaginal microbiome MetaCyc pathway (PWY-7376\|cob(II)yr | rs4704219 | 74748505 | 5 | COL4A3BP | Body mass index |
| rs630938 | 233800484 | 1 | KCNK1 | N-acetylcitrulline levels in elite athletes | rs4512110 | 74756221 | 5 | COL4A3BP | Body mass index |
| rs10193252 | 38212781 | 2 | RMDN2 | Metabolite levels | rs4704220 | 74757556 | 5 | COL4A3BP | Cholesteryl esters to total lipids ratio in IDL |
| rs1446296 | 38216416 | 2 | RMDN2 | Verbal aggression in schizophrenia | rs4704221 | 74759183 | 5 | COL4A3BP | Age-related diseases, mortality and associated endophenotypes |
| rs76200754 | 38572041 | 2 | ATL2 | Protein quantitative trait loci (liver) | rs5744552 | 74816944 | 5 | POLK | Total cholines levels |
| rs11687138 | 38573690 | 2 | ATL2 | Height | rs4704228 | 74844362 | 5 | POLK | Free cholesterol to total lipids in small HDL percentage (UKB data field 23647) |
| rs10048745 | 68962137 | 2 | ARHGAP25 | Lumbar spine bone mineral density | rs5744598 | 74846613 | 5 | POLK | Type 2 diabetes |
| rs2280246 | 68979141 | 2 | ARHGAP25 | Liver enzyme levels (alkaline phosphatase) | rs5744680 | 74879890 | 5 | POLK | Age-related diseases, mortality and associated endophenotypes |
| rs2280244 | 68979302 | 2 | ARHGAP25 | Serum alkaline phosphatase levels | rs11748027 | 74909972 | 5 | ANKDD1B | Total cholesterol levels |
| rs13413887 | 68996668 | 2 | ARHGAP25 | Rho GTPase-activating protein 25 levels | rs984976 | 74910870 | 5 | ANKDD1B | Low density lipoprotein cholesterol levels |
| rs4854511 | 68997144 | 2 | ARHGAP25 | Rho GTPase-activating protein 25 levels | rs6861279 | 74919409 | 5 | ANKDD1B | Cholesterol |
| rs10196674 | 68997153 | 2 | ARHGAP25 | Heel bone mineral density | rs16872665 | 74926246 | 5 | ANKDD1B | Metabolite levels |
| rs2311410 | 69008205 | 2 | ARHGAP25 | ARHGAP25/RNF41 protein level ratio | rs34341 | 74934009 | 5 | ANKDD1B | Body fat distribution (arm fat ratio) |
| rs11677065 | 69013007 | 2 | ARHGAP25 | Height | rs34344 | 74935825 | 5 | ANKDD1B | Nonsyndromic orofacial cleft x sex interaction (2df) |
| rs11688027 | 78018164 | 2 | LOC101927967 | Age at first sexual intercourse | rs34350 | 74945108 | 5 | ANKDD1B | Multi-trait sex score |
| rs12621150 | 78028838 | 2 | LOC101927967 | Cannabis dependence symptom count | rs253412 | 74955841 | 5 | ANKDD1B | Type 2 diabetes |
| rs7595103 | 78075244 | 2 | LOC101927967 | Attention deficit hyperactivity disorder and conduct disorder | rs253413 | 74955923 | 5 | ANKDD1B | Medication use (drugs used in diabetes) |
| rs35976024 | 86003971 | 2 | ATOH8 | Blood metabolite levels | rs28745309 | 82418056 | 5 | XRCC4 | Squamous cell lung carcinoma |

| | | | | | | | | | |
|---|---|---|---|---|---|---|---|---|---|
| rs13402783 | 86015121 | 2 | ATOH8 | Metastasis in stage I-III microsatellite instability low/stable c | rs4343818 | 82501422 | 5 | XRCC4 | IDP dMRI TBSS ICVF Fornix cres+Stria terminalis R |
| rs2139199 | 86046509 | 2 | LOC284950 | Red blood cell count | rs78913069 | 82567886 | 5 | XRCC4 | Squamous cell lung carcinoma |
| rs2280314 | 86071864 | 2 | ST3GAL5 | Dihexosylceramide (d18:1/22:0) levels | rs116210670 | 34038758 | 6 | GRM4 | Vaginal microbiome MetaCyc pathway (PWY-5838\|superpathway of menaquinol-8 biosynthesis I) |
| rs2280312 | 86072076 | 2 | ST3GAL5 | Lymphocyte count | rs76210789 | 34042113 | 6 | GRM4 | Vaginal microbiome MetaCyc pathway (PWY-5838\|superpathway of menaquinol-8 biosynthesis I) |
| rs11694145 | 86075582 | 2 | ST3GAL5 | Dihexosylceramide (d18:1/20:0) levels | rs1565355 | 34045973 | 6 | GRM4 | 2-aminooctanoate levels in elite athletes |
| rs17026684 | 86079129 | 2 | ST3GAL5 | Dihexosylceramide (d18:1/20:0) levels | rs75286976 | 34051095 | 6 | GRM4 | Protein quantitative trait loci (liver) |
| rs12478916 | 86086396 | 2 | ST3GAL5 | Mean corpuscular volume | rs77490194 | 34053693 | 6 | GRM4 | Inflammatory skin disease |
| rs3770093 | 86089673 | 2 | ST3GAL5 | Dihexosylceramide (d18:1/24:0) levels | rs3887555 | 34069062 | 6 | GRM4 | Height |
| rs7572209 | 86090072 | 2 | ST3GAL5 | Red cell distribution width | rs2499714 | 34072215 | 6 | GRM4 | Body mass index |
| rs10865479 | 86095432 | 2 | ST3GAL5 | Testosterone levels | rs1109654 | 34074463 | 6 | GRM4 | Height |
| rs6547638 | 86101011 | 2 | ST3GAL5 | Mean spheric corpuscular volume | rs2499724 | 34086671 | 6 | GRM4 | Height |
| rs1878905 | 86111688 | 2 | ST3GAL5 | Mean corpuscular volume | rs11970077 | 34093344 | 6 | GRM4 | Height |
| rs116892869 | 86115842 | 2 | ST3GAL5 | Mean platelet volume during first trimester of pregnancy | rs115675705 | 34094919 | 6 | GRM4 | A body shape index |
| rs2276626 | 86259443 | 2 | POLR1A | Diastolic blood pressure | rs11961715 | 34095696 | 6 | GRM4 | Height |
| rs3770081 | 86280925 | 2 | POLR1A | Facial emotion recognition (sad faces) | rs118012224 | 34205822 | 6 | HMGA1 | Platelet count during first trimester of pregnancy |
| rs10184246 | 86341328 | 2 | PTCD3 | Lung function (FEV1/FVC) | rs114344942 | 34207990 | 6 | HMGA1 | FEV1 |
| rs2241437 | 86360573 | 2 | PTCD3 | Response to methylphenidate treatment in attention-deficit/ | rs73744859 | 34208190 | 6 | HMGA1 | Systolic blood pressure |
| rs3731818 | 86368804 | 2 | PTCD3 | Pulse pressure | rs41269026 | 34208773 | 6 | HMGA1 | Body mass index |
| rs2241432 | 86438146 | 2 | MRPL35 | Systolic blood pressure | rs4713761 | 34209281 | 6 | HMGA1 | Height |
| rs7589790 | 106433909 | 2 | NCK2 | Eosinophil counts | rs41269028 | 34211238 | 6 | HMGA1 | Body mass index |
| rs1724184 | 109408776 | 2 | CCDC138 | Birth weight | rs1150781 | 34214322 | 6 | C6orf1 | Height |
| rs749810 | 109539430 | 2 | EDAR | Facial wrinkles (under eye) | rs4711389 | 34214670 | 6 | C6orf1 | Type 2 diabetes |
| rs3749060 | 109759696 | 2 | SH3RF3 | Height | rs1150779 | 34215919 | 6 | C6orf1 | Spleen volume |
| rs11676676 | 109767173 | 2 | SH3RF3 | Pediatric bone mineral density (spine) | rs1868361 | 17495669 | 7 | LOC102659288 | 5-acetylamino-6-formylamino-3-methyluracil levels |
| rs4676076 | 109812126 | 2 | SH3RF3 | General cognitive ability | rs10225144 | 17502590 | 7 | LOC102659288 | Cerebrospinal P-tau181p levels |
| rs12623119 | 109813592 | 2 | SH3RF3 | Cysteine-glutathione disulfide levels in elite athletes | rs6949509 | 17519261 | 7 | LOC101927630 | Coffee consumption (cups per day) |
| rs11123731 | 109817854 | 2 | SH3RF3 | Height | rs652187 | 17525702 | 7 | LOC101927630 | Response to selenium supplementation (change in plasma selenium concentration) |
| rs6542826 | 110049719 | 2 | SH3RF3 | Neurocognitive impairment in HIV-1 infection (dichotomous) | rs10499504 | 17561583 | 7 | LOC101927630 | Response to antipsychotic treatment |
| rs17285688 | 172239759 | 2 | METTL8 | OTU97_100 (Ruminococcaceae) prevalence | rs11768350 | 17561651 | 7 | LOC101927630 | Tea consumption |
| rs16859373 | 172242130 | 2 | METTL8 | Height | rs1349140 | 17562859 | 7 | LOC101927630 | Coffee consumption |
| rs72880595 | 172253633 | 2 | METTL8 | Hip circumference adjusted for BMI | rs73073176 | 17562952 | 7 | LOC101927630 | Coffee consumption |
| rs10171386 | 172399050 | 2 | CYBRD1 | Hip circumference adjusted for BMI | rs17137805 | 17563650 | 7 | LOC101927630 | Non-albumin protein levels |
| rs2138497 | 236619055 | 2 | AGAP1 | Aspartate aminotransferase levels in non-alcoholic fatty live | rs1031275 | 17566664 | 7 | LOC101927630 | Phospholipids to total lipids ratio in medium LDL |
| rs2696418 | 236636987 | 2 | AGAP1 | Participation in an health questionnaire (not invited vs invite | rs2708240 | 147577537 | 7 | CNTNAP2 | QT interval (drug interaction) |
| rs2696394 | 236669103 | 2 | AGAP1 | Educational attainment | rs2283200 | 2729340 | 11 | KCNQ1 | Height |
| rs72977127 | 236681671 | 2 | AGAP1 | Anterior thalamic nuclei volume | rs77294061 | 2734176 | 11 | KCNQ1 | Oleoyl-arachidonoyl-glycerol (18:1/20:4) [1] levels in elite athletes |
| rs148493191 | 236686646 | 2 | AGAP1 | Vaginal microbiome MetaCyc pathway (RHAMCAT-PWY\|L- | rs2411884 | 2735534 | 11 | KCNQ1 | Blood sugar levels |
| rs12472150 | 236696691 | 2 | AGAP1 | 3-phenylpropionate (hydrocinnamate) levels in elite athletes | rs67886747 | 2740934 | 11 | KCNQ1 | Waist circumference adjusted for body mass index |
| rs72983061 | 236718302 | 2 | AGAP1 | Brain shape (segment 7) | rs2283208 | 2743859 | 11 | KCNQ1 | Obesity without metabolic disease |
| rs72983078 | 236721102 | 2 | AGAP1 | Brain morphology (MOSTest) | rs231907 | 2752130 | 11 | KCNQ1 | Estimated glomerular filtration rate (creatinine) |
| rs908443 | 236741888 | 2 | AGAP1 | Body mass index | rs231906 | 2752609 | 11 | KCNQ1 | QT interval |
| rs4663617 | 236744626 | 2 | AGAP1 | Educational attainment (years of education) | rs2283220 | 2755548 | 11 | KCNQ1 | Type 2 diabetes |
| rs4414632 | 236762815 | 2 | AGAP1 | Body mass index (MTAG) | rs2839990 | 2756001 | 11 | KCNQ1 | Homoarginine levels in elite athletes |
| rs4625852 | 236762851 | 2 | AGAP1 | Body mass index | rs108961 | 2757985 | 11 | KCNQ1 | Plasma amyloid beta peptide concentrations (ABx-40) |
| rs72990123 | 238492155 | 2 | RAB17 | Corneal resistance factor | rs231879 | 2761382 | 11 | KCNQ1 | Height |
| rs3751112 | 238494742 | 2 | RAB17 | Height | rs7114479 | 2763228 | 11 | KCNQ1 | Vertex-wise sulcal depth |
| rs2292873 | 238499444 | 2 | RAB17 | Obesity-related traits | rs231878 | 2763777 | 11 | KCNQ1 | Waist circumference adjusted for body mass index |
| rs2292871 | 238499690 | 2 | RAB17 | Corneal resistance factor (MTAG) | rs2237878 | 2766282 | 11 | KCNQ1 | Protein quantitative trait loci |
| rs6740706 | 238628509 | 2 | LRRFIP1 | Longevity | rs84178 | 2774374 | 11 | KCNQ1 | Glomerular filtration rate (creatinine) |
| rs869434 | 238638531 | 2 | LRRFIP1 | Pulse pressure | rs231889 | 2777192 | 11 | KCNQ1 | Estimated glomerular filtration rate |
| rs12624154 | 238647468 | 2 | LRRFIP1 | Blood pressure (pleiotropy model 2 SBP adjusted for estima | rs6578296 | 2779970 | 11 | KCNQ1 | Height |
| rs2279639 | 238667552 | 2 | LRRFIP1 | Diastolic blood pressure (long-term average) | rs179785 | 2781519 | 11 | KCNQ1 | Gout |
| rs3769053 | 238670792 | 2 | LRRFIP1 | Diastolic blood pressure (baseline) | rs179784 | 2781824 | 11 | KCNQ1 | Kidney cancer |
| rs62194228 | 238674555 | 2 | LRRFIP1 | Self-harm x Caesarean-section interaction | rs231872 | 2787061 | 11 | KCNQ1 | Height |
| rs185944654 | 238674859 | 2 | LRRFIP1 | Vaginal microbiome relative abundance (s_Dialister propior | rs67004488 | 2787804 | 11 | KCNQ1 | Hip circumference adjusted for BMI |
| rs3769050 | 238769680 | 2 | RAMP1 | Adolescent idiopathic scoliosis | rs2237881 | 2788916 | 11 | KCNQ1 | Height |
| rs3769047 | 238769892 | 2 | RAMP1 | Short-chain dicarboxylacylcarnitine levels (additive genetic | rs63934 | 2789062 | 11 | KCNQ1 | Estimated glomerular filtration rate |
| rs895574 | 238888974 | 2 | UBE2F | Azelate (nonanedioate) levels in elite athletes | rs163160 | 2789955 | 11 | KCNQ1 | Glomerular filtration rate in non diabetics (creatinine) |
| rs62194510 | 238895507 | 2 | UBE2F | Educational attainment | rs2075870 | 2790019 | 11 | KCNQ1 | Multi-trait sex score |
| rs13007705 | 239069196 | 2 | FAM132B | Hemoglobin concentration | rs76327973 | 2793413 | 11 | KCNQ1 | Hip circumference adjusted for BMI |
| rs6747952 | 239069926 | 2 | FAM132B | Mean corpuscular hemoglobin concentration | rs233438 | 2794392 | 11 | KCNQ1 | Estimated glomerular filtration rate |
| rs11689193 | 239074665 | 2 | FAM132B | Hemoglobin levels | rs3852527 | 2826603 | 11 | KCNQ1 | Bacterial meningitis |
| rs12472274 | 239095422 | 2 | ILKAP | Phospholipid levels (plasma) | rs11601574 | 2828799 | 11 | KCNQ1 | Plasma cobalt levels |
| rs2304674 | 239181904 | 2 | PER2 | Multiple sclerosis (age of onset) | rs8181588 | 2831541 | 11 | KCNQ1 | Type 2 diabetes |
| rs58574366 | 239194837 | 2 | PER2 | Chronotype | rs163177 | 2838413 | 11 | KCNQ1 | Type 2 diabetes |
| rs7599697 | 239231477 | 2 | TRAF3IP1 | Insomnia | rs2237892 | 2839751 | 11 | KCNQ1 | Type 2 diabetes |
| rs12477345 | 239242967 | 2 | TRAF3IP1 | Morningness | rs11606985 | 2843676 | 11 | KCNQ1 | Glycated hemoglobin levels |
| rs3739070 | 239306268 | 2 | TRAF3IP1 | Iris characteristics | rs233449 | 2843803 | 11 | KCNQ1 | Type 2 diabetes |
| rs77008212 | 239307113 | 2 | TRAF3IP1 | Refractive astigmatism | rs233450 | 2844048 | 11 | KCNQ1 | Type 2 diabetes |
| rs11900963 | 239308049 | 2 | TRAF3IP1 | Morningness | rs163182 | 2844216 | 11 | KCNQ1 | Type 2 diabetes |
| rs3769124 | 239349362 | 2 | ASB1 | Pulmonary function | rs163183 | 2844441 | 11 | KCNQ1 | Height |
| rs11888775 | 239424876 | 2 | LINC01107 | Height | rs163184 | 2847069 | 11 | KCNQ1 | Type 2 diabetes |
| rs58568091 | 239430582 | 2 | LINC01107 | Lung function (FEV1/FVC) | rs2283228 | 2849530 | 11 | KCNQ1 | Type 2 diabetes |
| rs13403657 | 241943450 | 2 | SNED1 | Toxicity response to radiotherapy in prostate cancer (hemat | rs234853 | 2850828 | 11 | KCNQ1 | Type 2 diabetes |
| rs78127965 | 241980093 | 2 | SNED1 | Height | rs234855 | 2851113 | 11 | KCNQ1 | Hemoglobin A1c levels |
| rs7581709 | 242002828 | 2 | SNED1 | Resistance to COVID-19 infection (Exposed negative vs po | rs234857 | 2852529 | 11 | KCNQ1 | Height |
| rs2286321 | 242004866 | 2 | SNED1 | Height | rs2237895 | 2857194 | 11 | KCNQ1 | Type 2 diabetes |
| rs7559967 | 242006736 | 2 | SNED1 | Height | rs60808706 | 2857233 | 11 | KCNQ1 | Body mass index |
| rs6437234 | 242019288 | 2 | SNED1 | Suppressor of cytokine signaling 3 levels (SOCS3.11440.5 | rs234866 | 2857897 | 11 | KCNQ1 | Type 2 diabetes (adjusted for BMI) |

| | | | | |
|---|---|---|---|---|
| rs2302045 | 242021216 | 2 | SNED1 | Pharmacokinetics of antipsychotic drugs in severe mental d |
| rs2108485 | 242021742 | 2 | SNED1 | Suppressor of cytokine signaling 3 level in Chronic kidney c |
| rs16843241 | 242031090 | 2 | MTERFD2 | Height |
| rs2286323 | 242036797 | 2 | MTERFD2 | Carpal tunnel syndrome |
| rs2310876 | 242050740 | 2 | PASK | Hip index |
| rs17382723 | 242053546 | 2 | PASK | Height |
| rs3815305 | 242054403 | 2 | PASK | Hip circumference adjusted for BMI |
| rs2240543 | 242066789 | 2 | PASK | Height |
| rs59345329 | 242083763 | 2 | PASK | Height |
| rs758072 | 242083872 | 2 | PASK | Height |
| rs3771350 | 242110786 | 2 | PPP1R7 | Height |
| rs7565364 | 242114037 | 2 | PPP1R7 | Height |
| rs16843418 | 242118564 | 2 | PPP1R7 | Height |
| rs16843438 | 242137702 | 2 | ANO7 | Prostate cancer |
| rs76832527 | 242157241 | 2 | ANO7 | Prostate cancer |
| rs10933538 | 242162191 | 2 | ANO7 | Whole body fat free mass (UKB data field 23101) |
| rs15129 | 242168976 | 2 | HDLBP | Mean reticulocyte volume |
| rs3755326 | 242173882 | 2 | HDLBP | Height |
| rs75590598 | 242185654 | 2 | HDLBP | Eosinophil counts |
| rs2305075 | 242194583 | 2 | HDLBP | Serum alkaline phosphatase levels |
| rs11675957 | 242208792 | 2 | HDLBP | Height |
| rs10933548 | 242234837 | 2 | HDLBP | Waist circumference adjusted for body mass index |
| rs6743226 | 242236972 | 2 | HDLBP | Waist circumference adjusted for BMI (adjusted for smoking |
| rs59104589 | 242237902 | 2 | HDLBP | Fibrinogen levels |
| rs7599888 | 242248033 | 2 | HDLBP | Lung function (FEV1/FVC) |
| rs1106523 | 242251539 | 2 | HDLBP | Height |
| rs1476698 | 242296449 | 2 | FARP2 | Fibrinogen levels |
| rs60624480 | 242297118 | 2 | FARP2 | Calcium levels |
| rs3771555 | 242322680 | 2 | FARP2 | Fibrinogen levels or factor VII levels or factor XI levels or tis |
| rs2240479 | 242350466 | 2 | FARP2 | Serum alkaline phosphatase levels |
| rs62193221 | 242354632 | 2 | FARP2 | Urate levels |
| rs59916403 | 242370751 | 2 | FARP2 | Low density lipoprotein cholesterol levels |
| rs757978 | 242371101 | 2 | FARP2 | Chronic lymphocytic leukemia |
| rs3771570 | 242382864 | 2 | FARP2 | Prostate cancer |
| rs2055568 | 242393574 | 2 | FARP2 | Height |
| rs2302011 | 242393798 | 2 | FARP2 | Red blood cell count |
| rs764081 | 242394141 | 2 | FARP2 | Disability (impaired activities of daily living) |
| rs2302010 | 242394142 | 2 | FARP2 | Mean corpuscular volume |
| rs13388333 | 242400597 | 2 | FARP2 | Educational attainment (years of education) |
| rs41342147 | 242407588 | 2 | FARP2 | Vitiligo |
| rs3821285 | 242421314 | 2 | FARP2 | Height |
| rs871375 | 242421866 | 2 | FARP2 | White blood cell count |
| rs7596240 | 242444173 | 2 | STK25 | Hypothyroidism |
| rs1995496 | 242445336 | 2 | STK25 | Pulse pressure |
| rs7570235 | 242491353 | 2 | BOK-AS1 | Hip circumference adjusted for BMI |
| rs4675801 | 242493511 | 2 | BOK-AS1 | Weight |
| rs13423154 | 242495346 | 2 | BOK-AS1 | Multi-trait sex score |
| rs6718026 | 242495392 | 2 | BOK-AS1 | Appendicular lean mass |
| rs56211918 | 242497274 | 2 | BOK-AS1 | FEV1 |
| rs76993203 | 242498497 | 2 | BOK | Whole body fat free mass (UKB data field 23101) |
| rs876531 | 242500662 | 2 | BOK | Height |
| rs12479254 | 242502956 | 2 | BOK | Brain structure |
| rs4675917 | 242557785 | 2 | THAP4 | Height |
| rs139302128 | 242594226 | 2 | ATG4B | Carotid intima media thickness |
| rs7601000 | 242610773 | 2 | ATG4B | Body mass index |
| rs7943 | 242613089 | 2 | ATG4B | ITGB1BP2/MITD1 protein level ratio |
| rs7740 | 242613133 | 2 | ATG4B | Platelet count |
| rs7590385 | 242643381 | 2 | ING5 | ITGB1BP2/MED18 protein level ratio |
| rs34119918 | 242665602 | 2 | ING5 | Diverticular disease |
| rs7370843 | 242674104 | 2 | D2HGDH | Atopic asthma |
| rs141343442 | 242690770 | 2 | D2HGDH | Asthma (childhood onset) |
| rs35061832 | 242692490 | 2 | D2HGDH | Allergy |
| rs34290285 | 242698640 | 2 | D2HGDH | Eosinophil percentage of granulocytes |
| rs7569717 | 242701989 | 2 | D2HGDH | Ovarian cancer-related tumor marker CA 125 levels |
| rs4449174 | 242702538 | 2 | D2HGDH | Asthma |
| rs62193063 | 242728098 | 2 | GAL3ST2 | Ovarian cancer-related tumor marker CA 125 levels |
| rs35320439 | 242737341 | 2 | GAL3ST2 | Crohn's disease |
| rs35399295 | 242795942 | 2 | PDCD1 | Atopic dermatitis |
| rs4973668 | 242839363 | 2 | LINC01237 | PDCD1/TNFRSF4 protein level ratio |
| rs78458145 | 242877035 | 2 | LINC01237 | Intracranial aneurysm |
| rs12998513 | 242879499 | 2 | LINC01237 | Myopia (age of diagnosis) |
| rs71421178 | 242889533 | 2 | LINC01237 | Eczema |
| rs715827 | 10311773 | 3 | TATDN2 | ghrelin levels |
| rs164938 | 10315103 | 3 | TATDN2 | Educational attainment (years of education) |
| rs3774203 | 10317489 | 3 | TATDN2 | ghrelin levels |
| rs17032621 | 10325638 | 3 | GHRLOS | ghrelin levels |
| rs171407 | 10326169 | 3 | GHRLOS | ghrelin levels |
| rs35681 | 10329377 | 3 | GHRL | ghrelin levels |
| rs55821288 | 10330822 | 3 | GHRL | ghrelin levels |
| rs76868685 | 2858014 | 11 | KCNQ1 | Type 2 diabetes |
| rs2237896 | 2858440 | 11 | KCNQ1 | Type 2 diabetes |
| rs2237898 | 2863820 | 11 | KCNQ1 | Mosquito bite size |
| rs2237899 | 2866248 | 11 | KCNQ1 | Galactonate levels in elite athletes |
| rs2239897 | 2891965 | 11 | KCNQ1DN | Plateletcrit |
| rs2283231 | 2916715 | 11 | SLC22A18AS | Gout |
| rs10766451 | 2917510 | 11 | SLC22A18AS | Body mass index |
| rs388190 | 2918799 | 11 | SLC22A18AS | Autism spectrum disorders (repetitive sensory-motor behaviour) |
| rs4930027 | 2922493 | 11 | SLC22A18 | Height |
| rs2237902 | 2922705 | 11 | SLC22A18 | Type 2 diabetes (adjusted for BMI) |
| rs11822294 | 2924937 | 11 | SLC22A18 | Serum urate levels |
| rs120738 | 2926094 | 11 | SLC22A18 | Estimated glomerular filtration rate (creatinine) |
| rs450244 | 2940492 | 11 | SLC22A18 | Apolipoprotein A1 levels |
| rs1661052 | 2942593 | 11 | SLC22A18 | Total bilirubin levels |
| rs3758785 | 94132139 | 11 | GPR83 | Response to angiotensin II receptor blocker therapy (opposite direction w/ diuretic therapy) |
| rs7117042 | 94170975 | 11 | MRE11A | Metabolite levels |
| rs2212361 | 94312323 | 11 | PIWIL4 | Attention deficit hyperactivity disorder symptoms (maternal expressed emotions interaction) |
| rs7107978 | 94323965 | 11 | PIWIL4 | Color vision defects (Deutan-Protan) |
| rs591584 | 94325839 | 11 | PIWIL4 | Macrophage Migration Inhibitory Factor levels |
| rs932768 | 72347501 | 13 | DACH1 | Retinal arteriolar tortuosity |
| rs500830 | 72348768 | 13 | DACH1 | Estimated glomerular filtration rate |
| rs2147810 | 72364956 | 13 | DACH1 | Educational attainment |
| rs9592809 | 72365371 | 13 | DACH1 | Height |
| rs9572788 | 72367372 | 13 | DACH1 | Platelet count |
| rs303940 | 72371415 | 13 | DACH1 | Platelet count |
| rs303938 | 72371757 | 13 | DACH1 | Estimated glomerular filtration rate (creatinine) |
| rs303937 | 72372524 | 13 | DACH1 | Estimated glomerular filtration rate |
| rs2038856 | 72430228 | 13 | DACH1 | Vascular endothelial function |
| rs7156735 | 101132229 | 14 | LINC00523 | Protein delta homolog 1 levels |
| rs10413294 | 51993726 | 19 | CEACAM18 | Sialic acid-binding Ig-like lectin 6 levels |
| rs75950899 | 51997187 | 19 | SIGLEC12 | Sialic acid-binding Ig-like lectin 6 levels |
| rs73049701 | 51997320 | 19 | SIGLEC12 | Plasma calcium levels |
| rs12463092 | 51998336 | 19 | SIGLEC12 | Sialic acid-binding Ig-like lectin 6 levels |
| rs2124910 | 52025247 | 19 | SIGLEC6 | Sialic acid-binding Ig-like lectin 6 levels |
| rs10409713 | 52026801 | 19 | SIGLEC6 | Sialic acid-binding Ig-like lectin 6 level in Chronic kidney disease with hypertension and no diabetes (2741_22) |
| rs4146202 | 52032668 | 19 | SIGLEC6 | Sialic acid-binding Ig-like lectin 6 levels |
| rs2005199 | 52033206 | 19 | SIGLEC6 | Carotid intima media thickness |
| rs2305771 | 52033572 | 19 | SIGLEC6 | Blood protein levels |
| rs2824337 | 18905398 | 21 | CXADR | White blood cell count |
| rs186722413 | 18911836 | 21 | CXADR | Mean corpuscular volume |
| rs62239920 | 18944889 | 21 | CXADR | Neutrophil count |
| rs2824372 | 18947037 | 21 | CXADR | Neutrophil count |
| rs62239946 | 18952575 | 21 | CXADR | Ascending aorta distensibility |

| | | | | |
|---|---|---|---|---|
| rs14154 | 10368113 | 3 | ATP2B2 | Educational attainment (years of education) |
| rs26313 | 10376167 | 3 | ATP2B2 | Cysteine-rich with EGF-like domain protein 1 levels |
| rs703912 | 10391426 | 3 | ATP2B2 | Height |
| rs10222649 | 10450220 | 3 | ATP2B2 | Cysteine-rich with EGF-like domain protein 1 levels |
| rs719318 | 10474137 | 3 | ATP2B2 | Cysteine-rich with EGF-like domain protein 1 levels |
| rs17603886 | 10484726 | 3 | ATP2B2 | Amyotrophic lateral sclerosis (sporadic) |
| rs34864 | 10485200 | 3 | ATP2B2 | Cysteine-rich with EGF-like domain protein 1 levels |
| rs56284847 | 10512641 | 3 | ATP2B2 | ghrelin levels |
| rs13067215 | 10514008 | 3 | ATP2B2 | Cysteine-rich with EGF-like domain protein 1 levels |
| rs17200543 | 16973762 | 3 | PLCL2 | Lymphocyte percentage of white cells |
| rs9835432 | 17045728 | 3 | PLCL2 | Neuroticism general factor |
| rs5019428 | 17046866 | 3 | PLCL2 | Rheumatoid arthritis (rheumatoid factor and/or anti-cyclic citrullinated peptide seropositive) |
| rs4452313 | 17047032 | 3 | PLCL2 | Rheumatoid arthritis |
| rs7653834 | 17051328 | 3 | PLCL2 | Adolescent idiopathic scoliosis |
| rs13083189 | 17060203 | 3 | PLCL2 | Serum levels of protein CCL17 |
| rs4618210 | 17124384 | 3 | PLCL2 | Myocardial infarction |
| rs34937985 | 17126246 | 3 | PLCL2 | Schizophrenia |
| rs11926494 | 23258614 | 3 | UBE2E2 | Type 2 diabetes |
| rs73033558 | 23280696 | 3 | UBE2E2 | interferon-related traits |
| rs2359637 | 23280826 | 3 | UBE2E2 | Hematocrit |
| rs9815299 | 23334840 | 3 | UBE2E2 | Height |
| rs7612463 | 23336450 | 3 | UBE2E2 | Type 2 diabetes |
| rs4858496 | 23349496 | 3 | UBE2E2 | Reaction time |
| rs4858505 | 23377366 | 3 | MIR548AC | Drinks per week |
| rs716457 | 23380319 | 3 | MIR548AC | Severe COVID-19 infection |
| rs12633613 | 23390507 | 3 | MIR548AC | Type 2 diabetes |
| rs6795968 | 23417027 | 3 | MIR548AC | Drinks per week |
| rs778486 | 23434534 | 3 | MIR548AC | Acceptance of an invitation to participate in a mental health questionnaire |
| rs1496653 | 23454790 | 3 | MIR548AC | Type 2 diabetes |
| rs35352848 | 23455582 | 3 | MIR548AC | Type 2 diabetes |
| rs13094957 | 23457080 | 3 | MIR548AC | Type 2 diabetes |
| rs13062101 | 23483536 | 3 | MIR548AC | Severe COVID-19 infection |
| rs17013314 | 23510044 | 3 | MIR548AC | Type 2 diabetes |
| rs9830388 | 23559547 | 3 | MIR548AC | RR interval (tricyclic/tetracyclic antidepressant use interaction) |
| rs11916411 | 23567980 | 3 | MIR548AC | Eczema |
| rs11713090 | 23570654 | 3 | MIR548AC | Physiological dysregulation (Mahalanobis distance) |
| rs2036226 | 23597986 | 3 | MIR548AC | Asthma |
| rs55732421 | 23606404 | 3 | MIR548AC | Red blood cell count |
| rs76435632 | 23632174 | 3 | MIR548AC | Type 2 diabetes |
| rs6767406 | 23862463 | 3 | UBE2E1 | Academic attainment (English) |
| rs1473379 | 23873602 | 3 | UBE2E1 | Pyroglutamine levels in elite athletes |
| rs13099267 | 23878817 | 3 | UBE2E1 | Smoking initiation (ever regular vs never regular) (MTAG) |
| rs6550805 | 23892781 | 3 | UBE2E1 | Lysophosphatidylethanolamine (18:1) [sn1] levels |
| rs6779537 | 23901028 | 3 | UBE2E1 | Academic attainment (English) |
| rs7615255 | 23902336 | 3 | UBE2E1 | Smoking initiation |
| rs17787217 | 23907775 | 3 | UBE2E1 | Reaction time |
| rs13085379 | 23911939 | 3 | UBE2E1 | Self-reported math ability |
| rs34476593 | 23943599 | 3 | NKIRAS1 | Intelligence (MTAG) |
| rs9836970 | 23949839 | 3 | NKIRAS1 | General cognitive ability |
| rs2001209 | 23962646 | 3 | RPL15 | Intelligence (MTAG) |
| rs9862416 | 23987842 | 3 | NR1D2 | Educational attainment (MTAG) |
| rs5001573 | 23991717 | 3 | NR1D2 | Highest math class taken (MTAG) |
| rs4561770 | 24143218 | 3 | LINC00691 | Intelligence |
| rs1349265 | 24159387 | 3 | THRB | Intelligence |
| rs826374 | 24163921 | 3 | THRB | Educational attainment |
| rs7652234 | 24190459 | 3 | THRB | Menarche (age at onset) |
| rs4858582 | 24219521 | 3 | THRB | General cognitive ability |
| rs2683541 | 24222782 | 3 | THRB | Height |
| rs1700943 | 24231030 | 3 | THRB | Refractive error |
| rs13096529 | 24232035 | 3 | THRB | Lymphocyte count |
| rs13065854 | 24234232 | 3 | THRB | Lymphocyte count |
| rs2017200 | 24254752 | 3 | THRB | Uterine leiomyomata |
| rs4260345 | 24256698 | 3 | THRB | Spherical equivalent or myopia (age of diagnosis) |
| rs826221 | 24268436 | 3 | THRB | Systolic blood pressure in sickle cell anemia |
| rs826220 | 24268677 | 3 | THRB | Myopia |
| rs826217 | 24269789 | 3 | THRB | Educational attainment |
| rs9829876 | 24277179 | 3 | THRB | Leukotriene B4 levels in elite athletes |
| rs12634582 | 24282432 | 3 | THRB | Asthma |
| rs7633961 | 29891801 | 3 | RBMS3 | Basal ganglia growth |
| rs17553695 | 29932963 | 3 | RBMS3 | Predicted developmental stuttering |
| rs6778759 | 29956943 | 3 | RBMS3 | Atopic dermatitis (moderate to severe) |
| rs6549962 | 30029027 | 3 | RBMS3 | Heel bone mineral density |
| rs9844381 | 32878311 | 3 | TRIM71 | Educational attainment |
| rs34010125 | 32878682 | 3 | TRIM71 | Vertical cup-disc ratio (adjusted for vertical disc diameter) |
| rs9832642 | 32879663 | 3 | TRIM71 | Parental longevity (father's age at death) |
| rs6550168 | 32908632 | 3 | TRIM71 | OTU97_56 (Ruminococcaceae) prevalence |
| rs417387 | 42571546 | 3 | VIPR1 | Cerebrospinal fluid clusterin levels |
| rs55940287 | 42573422 | 3 | VIPR1 | N6-carbamoylthreonyladenosine levels in elite athletes |
| rs342515 | 42574808 | 3 | VIPR1 | CCL17/CXCL3 protein level ratio |

| | | | | |
|---|---|---|---|---|
| rs34801786 | 42630979 | 3 | SEC22C | Cognitive impairment test score |
| rs56978598 | 42769657 | 3 | CCDC13 | Eosinophil counts |
| rs6442057 | 42795050 | 3 | CCDC13 | Glaucoma (primary open-angle) |
| rs3092963 | 46396938 | 3 | CCR2 | CCR2 on plasmacytoid Dendritic Cell |
| rs1800024 | 46412559 | 3 | CCR5 | Granulocyte percentage of myeloid white cells |
| rs11266744 | 46449978 | 3 | CCRL2 | Monocyte percentage of white cells |
| rs7626288 | 46485369 | 3 | LTF | Early-onset schizophrenia |
| rs72622926 | 46485773 | 3 | LTF | Sperm acrosome membrane-associated protein 3 levels (SPACA3.8076.4.3) |
| rs4682816 | 46487775 | 3 | LTF | Lactotransferrin levels (LTF.2780.35.2) |
| rs2239692 | 46488854 | 3 | LTF | Teratocarcinoma-derived growth factor 1 levels |
| rs743659 | 46489015 | 3 | LTF | Serum levels of protein MYCT1 |
| rs34827868 | 46493855 | 3 | LTF | Lactotransferrin levels |
| rs11707471 | 46498865 | 3 | LTF | Blood protein levels |
| rs1126478 | 46501213 | 3 | LTF | Alkaline phosphatase, tissue-nonspecific isozyme levels |
| rs1126477 | 46501268 | 3 | LTF | Blood protein levels |
| rs9862388 | 46504316 | 3 | LTF | PAXIP1-associated protein 1 level in Chronic kidney disease with hypertension and no diabetes (23967_8) |
| rs6441996 | 46505266 | 3 | LTF | Nucleoside diphosphate kinase A levels (NME1.5909.51.3) |
| rs9682509 | 46596264 | 3 | LRRC2 | Teratocarcinoma-derived growth factor 1 levels |
| rs2280410 | 46599937 | 3 | LRRC2 | Metabolite levels |
| rs6783573 | 46600520 | 3 | LRRC2 | Cerebrospinal fluid biomarker levels |
| rs7650998 | 46607255 | 3 | LRRC2 | Height |
| rs3806701 | 46617337 | 3 | TDGF1 | Teratocarcinoma-derived growth factor 1 levels |
| rs3806702 | 46617502 | 3 | TDGF1 | Protein levels in obesity |
| rs2139637 | 46654132 | 3 | LOC100132146 | G_Alloprevotella abundance |
| rs2577814 | 62849213 | 3 | CADPS | Metabolite levels |
| rs6794810 | 63268489 | 3 | SYNPR | General cognitive ability |
| rs17031723 | 63385835 | 3 | SYNPR | Vaginal microbiome MetaCyc pathway (PWY-7664\|oleate biosynthesis IV (anaerobic)) |
| rs16851720 | 141463298 | 3 | RNF7 | Hepatitis C induced liver fibrosis |
| rs11721024 | 141602770 | 3 | ATP1B3 | QT interval |
| rs6785570 | 141603887 | 3 | ATP1B3 | Blood pressure (pleiotropy model 2 SBP adjusted for estimated causal effects x DBP) |
| rs4683645 | 141610807 | 3 | ATP1B3 | Complicated vs uncomplicated appendicitis |
| rs2068229 | 141623304 | 3 | ATP1B3 | Red cell distribution width |
| rs7622417 | 141625999 | 3 | ATP1B3 | Coronary artery disease |
| rs3792411 | 141626123 | 3 | ATP1B3 | Medication use (thyroid preparations) |
| rs6782694 | 141627860 | 3 | ATP1B3 | Pulse pressure |
| rs3804772 | 141634056 | 3 | ATP1B3 | Blond vs. brown/black hair color |
| rs9880808 | 141642781 | 3 | ATP1B3 | Height |
| rs9942 | 141663752 | 3 | TFDP2 | General cognitive ability |
| rs9815587 | 141674548 | 3 | TFDP2 | Creatinine levels |
| rs3792409 | 141682825 | 3 | TFDP2 | Urate levels |
| rs3804771 | 141691407 | 3 | TFDP2 | Body mass index |
| rs2901949 | 141691494 | 3 | TFDP2 | Hippocampal subfield left CA4 volume (head) |
| rs4315664 | 141694097 | 3 | TFDP2 | Dorsomedial frontal area (unadjusted for global measures) |
| rs2271386 | 141712708 | 3 | TFDP2 | Creatinine levels |
| rs3817176 | 141712780 | 3 | TFDP2 | Body mass index |
| rs1511299 | 141716072 | 3 | TFDP2 | Glomerular filtration rate |
| rs2861422 | 141724644 | 3 | TFDP2 | Glomerular filtration rate (creatinine) |
| rs7644383 | 141743951 | 3 | TFDP2 | Mean reticulocyte volume |
| rs1397764 | 141750810 | 3 | TFDP2 | Estimated glomerular filtration rate |
| rs6791514 | 141766305 | 3 | TFDP2 | Estimated glomerular filtration rate |
| rs1355438 | 141767853 | 3 | TFDP2 | Estimated glomerular filtration rate (creatinine) |
| rs77982841 | 141770593 | 3 | TFDP2 | Chemoradiation-induced hearing loss in nasopharyngeal carcinoma |
| rs11705932 | 141818850 | 3 | TFDP2 | Testicular germ cell tumor |
| rs6773343 | 141825598 | 3 | TFDP2 | Serum 25-Hydroxyvitamin D levels |
| rs79513925 | 141853976 | 3 | TFDP2 | Creatinine levels |
| rs6769556 | 141881351 | 3 | GK5 | Vaginal microbiome MetaCyc pathway (PWY-5005\|biotin biosynthesis II) |
| rs62282070 | 141888148 | 3 | GK5 | C-reactive protein levels |
| rs2364921 | 158522463 | 3 | MFSD1 | Insomnia |
| rs16854614 | 169636995 | 3 | SAMD7 | Triglyceride levels |
| rs3755605 | 169756119 | 3 | GPR160 | Testicular germ cell tumor |
| rs6808506 | 169839239 | 3 | PHC3 | Eosinophil counts |
| rs1386332 | 169877827 | 3 | PHC3 | Height |
| rs71277158 | 169999216 | 3 | PRKCI | Prostate cancer |
| rs9682041 | 170091902 | 3 | SKIL | Glomerular filtration rate in non diabetics (creatinine) |
| rs1045210 | 170114078 | 3 | SKIL | Height |
| rs11928222 | 189350265 | 3 | TP63 | Lung cancer in never smokers |
| rs4488809 | 189356261 | 3 | TP63 | Lung cancer |
| rs7636839 | 189356941 | 3 | TP63 | EGFR mutation-positive lung adenocarcinoma |
| rs13080835 | 189357199 | 3 | TP63 | Lung adenocarcinoma |
| rs13314271 | 189357602 | 3 | TP63 | Lung cancer |
| rs10937405 | 189383183 | 3 | TP63 | Lung adenocarcinoma |
| rs17505102 | 189401776 | 3 | TP63 | Acute lymphoblastic leukemia (childhood) |
| rs2378515 | 189410000 | 3 | TP63 | Adipose morphology |
| rs12696596 | 189416987 | 3 | TP63 | Estimated glomerular filtration rate (creatinine) |
| rs10804920 | 189438689 | 3 | TP63 | Age at natural menopause |
| rs11711203 | 189441242 | 3 | TP63 | Age at menopause |
| rs3796159 | 194790434 | 3 | XXYLT1 | Venous thromboembolism or factor VII levels (pleiotropy) |
| rs60902112 | 194800853 | 3 | XXYLT1 | Atrial fibrillation |
| rs73206626 | 194821377 | 3 | XXYLT1 | Height |

| | | | | |
|---|---|---|---|---|
| rs952481 | 194822098 | 3 | XXYLT1 | Height |
| rs903306 | 194833868 | 3 | XXYLT1 | Severe COVID-19 infection |
| rs869519 | 194835189 | 3 | XXYLT1 | Severe COVID-19 infection |
| rs6784890 | 194842145 | 3 | XXYLT1 | Height |
| rs7644208 | 194855954 | 3 | XXYLT1 | 21-hydroxypregnenolone disulfate levels in elite athletes |
| rs1009642 | 194857586 | 3 | XXYLT1 | Body mass index (MTAG) |
| rs2131877 | 194858374 | 3 | XXYLT1 | Non-small cell lung cancer |
| rs4677813 | 194863860 | 3 | XXYLT1 | Adult body size |
| rs7644678 | 194876849 | 3 | XXYLT1 | Body mass index |
| rs7616009 | 194881756 | 3 | XXYLT1 | Body mass index |
| rs77485504 | 194972515 | 3 | XXYLT1 | Vaginal microbiome MetaCyc pathway (PWY-5856\|ubiquinol-9 biosynthesis (prokaryotic)) |
| rs11919965 | 194975071 | 3 | XXYLT1 | Height |
| rs76960093 | 194980147 | 3 | XXYLT1 | Vaginal microbiome MetaCyc pathway (PWY-5863\|superpathway of phylloquinol biosynthesis) |
| rs2410840 | 194983053 | 3 | XXYLT1 | Metabolite levels |
| rs7611238 | 195072918 | 3 | ACAP2 | Body mass index |
| rs2932966 | 23806976 | 4 | PPARGC1A | Smoking initiation |
| rs2970853 | 23823519 | 4 | PPARGC1A | IDP dMRI TBSS ICVF Superior cerebellar peduncle R |
| rs6448226 | 23836653 | 4 | PPARGC1A | Kidney cancer |
| rs10018700 | 23855372 | 4 | PPARGC1A | Severe COVID-19 infection |
| rs7437482 | 23855974 | 4 | PPARGC1A | Diffuse plaques |
| rs73243622 | 23875158 | 4 | PPARGC1A | QRS duration |
| rs10028665 | 23875834 | 4 | PPARGC1A | QT interval |
| rs11734408 | 23882519 | 4 | PPARGC1A | Sex hormone-binding globulin levels |
| rs12500214 | 23885568 | 4 | PPARGC1A | OTU99_166 (Prevotella) prevalence |
| rs2970877 | 23887454 | 4 | PPARGC1A | Sex hormone-binding globulin levels adjusted for BMI |
| rs2970876 | 23888619 | 4 | PPARGC1A | Hematocrit |
| rs2970872 | 23889813 | 4 | PPARGC1A | Height |
| rs2970871 | 23890582 | 4 | PPARGC1A | Sex hormone-binding globulin levels adjusted for BMI |
| rs1006181 | 38004280 | 4 | TBC1D1 | Height |
| rs13132184 | 38013026 | 4 | TBC1D1 | Verbal declarative memory |
| rs13142228 | 38018181 | 4 | TBC1D1 | Height |
| rs7671645 | 38019302 | 4 | TBC1D1 | Lymphocyte-to-monocyte ratio |
| rs6845120 | 38047545 | 4 | TBC1D1 | Blood pressure (pleiotropy model 1 DBP adjusted for estimated causal effects x SBP) |
| rs1863329 | 38060485 | 4 | TBC1D1 | Monocyte percentage of white cells |
| rs1364953 | 38062233 | 4 | TBC1D1 | Metabolite levels |
| rs71606104 | 38100888 | 4 | TBC1D1 | Smoking initiation |
| rs933822 | 38105161 | 4 | TBC1D1 | Neutrophil percentage of white cells |
| rs1344603 | 38109847 | 4 | TBC1D1 | Lymphocyte percentage of white cells |
| rs630428 | 38121363 | 4 | TBC1D1 | Hearing function (2 kHz) |
| rs636487 | 38125811 | 4 | TBC1D1 | Neutrophil percentage of white cells |
| rs633059 | 38132999 | 4 | TBC1D1 | Lymphocyte percentage of white cells |
| rs658349 | 38137353 | 4 | TBC1D1 | Lymphocyte count |
| rs922334 | 38616758 | 4 | KLF3-AS1 | Height |
| rs2126526 | 38618689 | 4 | KLF3-AS1 | Lymphocyte count |
| rs28418927 | 38626130 | 4 | KLF3-AS1 | Red blood cell count |
| rs2381199 | 38636537 | 4 | KLF3-AS1 | Estimated glomerular filtration rate (creatinine) |
| rs337604 | 38648703 | 4 | KLF3-AS1 | Essential tremor |
| rs73134233 | 38652812 | 4 | KLF3-AS1 | Hip circumference adjusted for BMI |
| rs12331049 | 38655772 | 4 | KLF3-AS1 | Mean spheric corpuscular volume |
| rs75504410 | 38657819 | 4 | KLF3-AS1 | Sum eosinophil basophil counts |
| rs337606 | 38659263 | 4 | KLF3-AS1 | Height |
| rs1000571 | 38662895 | 4 | KLF3-AS1 | Eosinophil counts |
| rs12507197 | 38663660 | 4 | KLF3-AS1 | Red blood cell count |
| rs78976775 | 38663828 | 4 | KLF3-AS1 | Eosinophil counts |
| rs112691446 | 38665142 | 4 | KLF3-AS1 | Neutrophil percentage of granulocytes |
| rs337623 | 38673845 | 4 | KLF3 | Monocyte percentage of white cells |
| rs68191221 | 38677062 | 4 | KLF3 | Sum eosinophil basophil counts |
| rs34089598 | 38677227 | 4 | KLF3 | White blood cell count |
| rs9992667 | 38680186 | 4 | KLF3 | Eosinophil percentage of granulocytes |
| rs6531656 | 38684201 | 4 | KLF3 | Lymphocyte count |
| rs59048840 | 38684865 | 4 | KLF3 | Eosinophil counts |
| rs73232890 | 38685041 | 4 | KLF3 | Eosinophil counts |
| rs17428931 | 38689042 | 4 | KLF3 | Neutrophil percentage of granulocytes |
| rs17428945 | 38689745 | 4 | KLF3 | Mean corpuscular hemoglobin |
| rs2242189 | 38691024 | 4 | KLF3 | Central nucleus volume |
| rs1000096 | 38692835 | 4 | KLF3 | Body mass index |
| rs36023504 | 38698924 | 4 | KLF3 | Body mass index |
| rs3209570 | 38699657 | 4 | KLF3 | Body mass index (MTAG) |
| rs11466653 | 38776235 | 4 | TLR10 | Peripheral arterial disease (traffic-related air pollution interaction) |
| rs28393318 | 38784267 | 4 | TLR10 | Asthma or allergic disease (pleiotropy) |
| rs4833095 | 38799710 | 4 | TLR1 | Asthma and hay fever |
| rs5743604 | 38801285 | 4 | TLR1 | Hay fever and/or eczema |
| rs2130296 | 38874802 | 4 | FAM114A1 | Height |
| rs6814341 | 38877682 | 4 | FAM114A1 | Height |
| rs10516875 | 91590266 | 4 | CCSER1 | Age at first sexual intercourse |
| rs10030217 | 96457804 | 4 | UNC5C | Blood protein levels |
| rs10032931 | 96458433 | 4 | UNC5C | Optic nerve measurement (cup-to-disc ratio) |
| rs1880529 | 114379379 | 4 | CAMK2D | White matter integrity (bipolar disorder risk interaction) |
| rs17620390 | 114384328 | 4 | CAMK2D | Heart failure |

| | | | | |
|---|---|---|---|---|
| rs62314976 | 114385676 | 4 | CAMK2D | JT interval |
| rs2285703 | 114388820 | 4 | CAMK2D | P wave duration |
| rs17446418 | 114418006 | 4 | CAMK2D | PR interval |
| rs11098193 | 114429407 | 4 | CAMK2D | ECG cardiac MRI latent space |
| rs3815072 | 114435877 | 4 | CAMK2D | Guanosine levels in elite athletes |
| rs7700110 | 114439894 | 4 | CAMK2D | Chronotype |
| rs35132791 | 114456506 | 4 | CAMK2D | TPE interval (resting) |
| rs10023113 | 114546704 | 4 | CAMK2D | Non-small cell lung cancer (survival) |
| rs2189364 | 114556075 | 4 | CAMK2D | Hip circumference adjusted for BMI |
| rs1859229 | 114661681 | 4 | CAMK2D | Height |
| rs115143758 | 114668476 | 4 | CAMK2D | Calcium/calmodulin-dependent protein kinase type II subunit delta levels |
| rs13110620 | 114822579 | 4 | ARSJ | Height |
| rs17620624 | 114832204 | 4 | ARSJ | COVID-19 (covid vs negative) |
| rs17621017 | 114879781 | 4 | ARSJ | Height |
| rs7677674 | 114889374 | 4 | ARSJ | Height |
| rs11098219 | 114891757 | 4 | ARSJ | Height |
| rs6827199 | 114896904 | 4 | ARSJ | Height |
| rs9286351 | 138441530 | 4 | PCDH18 | Diastolic blood pressure |
| rs7685296 | 153254121 | 4 | FBXW7 | Type 2 diabetes |
| rs2676329 | 153275395 | 4 | FBXW7 | Blood pressure (pleiotropy model 2 SBP adjusted for estimated causal effects x DBP) |
| rs2203644 | 153275701 | 4 | FBXW7 | Eosinophil counts |
| rs11931711 | 153291513 | 4 | FBXW7 | Eosinophil counts |
| rs10021011 | 153410390 | 4 | FBXW7 | Eosinophil counts |
| rs17277029 | 153428649 | 4 | FBXW7 | Height |
| rs6817936 | 167698248 | 4 | SPOCK3 | Height |
| rs13114435 | 183821662 | 4 | DCTD | Obesity-related traits |
| rs13101260 | 183823597 | 4 | DCTD | Retinol (Vitamin A) levels in elite athletes |
| rs10020261 | 184171187 | 4 | WWC2 | Initial alcohol sensitivity |
| rs6854052 | 184187311 | 4 | WWC2 | Lung function (FEV1/FVC) |
| rs6858486 | 184190535 | 4 | WWC2 | Height |
| rs1473070 | 184192037 | 4 | WWC2 | Height |
| rs10015974 | 184196254 | 4 | WWC2 | Height |
| rs12643425 | 184201182 | 4 | WWC2 | COVID-19 (hospitalized vs not hospitalized) |
| rs3814420 | 184204228 | 4 | WWC2 | Aspartate aminotransferase levels |
| rs17291855 | 184206296 | 4 | WWC2 | Height |
| rs11939899 | 184209611 | 4 | WWC2 | Gamma glutamyl transpeptidase |
| rs907366 | 184212220 | 4 | WWC2 | Heel bone mineral density |
| rs955748 | 184215675 | 4 | WWC2 | Height |
| rs72703409 | 184225295 | 4 | WWC2 | Height |
| rs11730648 | 184226187 | 4 | WWC2 | Height |
| rs4862155 | 184236868 | 4 | WWC2 | Height |
| rs6839940 | 184242959 | 4 | CLDN24 | Gamma glutamyl transferase levels |
| rs10866527 | 1891800 | 5 | CTD-2194D22.4 | Prostate cancer |
| rs10866528 | 1891821 | 5 | CTD-2194D22.4 | Prostate cancer |
| rs4975759 | 1892455 | 5 | CTD-2194D22.4 | Prostate cancer |
| rs12653946 | 1895829 | 5 | CTD-2194D22.4 | Prostate cancer |
| rs13177232 | 1896263 | 5 | CTD-2194D22.4 | Prostate cancer |
| rs13177600 | 1897179 | 5 | CTD-2194D22.4 | Prostate cancer |
| rs7709041 | 12702044 | 5 | LINC01194 | Educational attainment |
| rs10085101 | 31820465 | 5 | PDZD2 | Severe COVID-19 infection |
| rs163233 | 34859225 | 5 | TTC23L | Dysphagia |
| rs61287758 | 117734465 | 5 | LOC101927280 | Disease progression in age-related macular degeneration (adjusted for baseline) |
| rs6595129 | 117763622 | 5 | LOC101927280 | Height |
| rs6877715 | 117944805 | 5 | LOC102467225 | Adolescent idiopathic scoliosis |
| rs78814834 | 149191547 | 5 | PPARGC1B | Nontraumatic osteonecrosis of the femoral head |
| rs17110447 | 149192846 | 5 | PPARGC1B | Keratinocyte cancer (MTAG) |
| rs251468 | 149194485 | 5 | PPARGC1B | Freckles |
| rs251466 | 149195603 | 5 | PPARGC1B | Sunburns |
| rs251465 | 149196090 | 5 | PPARGC1B | Squamous cell carcinoma (MTAG) |
| rs251464 | 149196234 | 5 | PPARGC1B | Low tan response |
| rs32579 | 149210848 | 5 | PPARGC1B | Tanning |
| rs32576 | 149213456 | 5 | PPARGC1B | Obesity-related traits |
| rs3797622 | 149827098 | 5 | RPS14 | Educational attainment |
| rs2545341 | 149914401 | 5 | NDST1 | Appendicular lean mass |
| rs7719412 | 150000145 | 5 | SYNPO | Imidazole lactate levels in elite athletes |
| rs72838326 | 167723152 | 5 | WWC1 | Drinks per week |
| rs728668 | 167749878 | 5 | WWC1 | Height |
| rs78954674 | 167792682 | 5 | WWC1 | Type 2 diabetes nephropathy including microalbuminuria |
| rs6555807 | 167826166 | 5 | WWC1 | Nucleus accumbens volume change rate |
| rs9313410 | 167828210 | 5 | WWC1 | interferon-related traits |
| rs3926392 | 167838535 | 5 | WWC1 | Cerebellum cortex volume change rate x age interaction (1df) |
| rs729387 | 167844571 | 5 | WWC1 | Body mass index (MTAG) |
| rs1477304 | 167845338 | 5 | WWC1 | OTU97_74 (Alistipes) abundance |
| rs11134512 | 167847460 | 5 | WWC1 | Body mass index |
| rs11948899 | 167848199 | 5 | WWC1 | Left hippocampal volume |
| rs13172600 | 168333137 | 5 | SLIT3 | Body mass index |
| rs13166368 | 168368994 | 5 | SLIT3 | Obsessive-compulsive symptoms |
| rs12188351 | 168386089 | 5 | SLIT3 | Inflammatory skin disease |
| rs77328981 | 168393227 | 5 | SLIT3 | Atrial fibrillation |

| | | | | |
|---|---|---|---|---|
| rs76647377 | 985196 | 6 | LOC285768 | Buruli ulcer (age at onset) |
| rs845868 | 998813 | 6 | LOC285768 | Colorectal cancer |
| rs845885 | 1019362 | 6 | LOC285768 | Risky substance use |
| rs7772296 | 7114842 | 6 | RREB1 | Height |
| rs1297769 | 7122936 | 6 | RREB1 | Urate levels |
| rs1285886 | 7140831 | 6 | RREB1 | Lymphocyte count |
| rs1285885 | 7142891 | 6 | RREB1 | Monocyte count |
| rs1297770 | 7158225 | 6 | RREB1 | Lymphocyte count |
| rs59431963 | 7189858 | 6 | RREB1 | Cortical thickness (MOSTest) |
| rs2326883 | 7192786 | 6 | RREB1 | Height |
| rs7743323 | 7195684 | 6 | RREB1 | Height |
| rs6936191 | 7201660 | 6 | RREB1 | Neutrophil count |
| rs6921580 | 7203714 | 6 | RREB1 | Estimated glomerular filtration rate |
| rs4960295 | 7205796 | 6 | RREB1 | Vertical cup-disc ratio |
| rs7751325 | 7209132 | 6 | RREB1 | Hip circumference adjusted for BMI |
| rs1334577 | 7211751 | 6 | RREB1 | Neutrophil count |
| rs1334576 | 7211818 | 6 | RREB1 | Chronic obstructive pulmonary disease |
| rs4960297 | 7212084 | 6 | RREB1 | Vertical cup-disc ratio (adjusted for vertical disc diameter) |
| rs17762454 | 7213200 | 6 | RREB1 | Mean corpuscular hemoglobin concentration |
| rs76052955 | 7213907 | 6 | RREB1 | Pulse pressure |
| rs17672692 | 7214499 | 6 | RREB1 | Lung function (FEV1/FVC) |
| rs1413700 | 7216484 | 6 | RREB1 | Multiple system atrophy |
| rs6931262 | 7217517 | 6 | RREB1 | Waist-hip ratio |
| rs9379081 | 7219873 | 6 | RREB1 | Waist-to-hip ratio adjusted for BMI |
| rs57340203 | 7220379 | 6 | RREB1 | Waist circumference adjusted for body mass index |
| rs9379082 | 7220960 | 6 | RREB1 | Waist-hip ratio |
| rs9328401 | 7225674 | 6 | RREB1 | Hip circumference adjusted for BMI |
| rs12192672 | 7229619 | 6 | RREB1 | Hemoglobin |
| rs34561875 | 7231770 | 6 | RREB1 | Waist-to-hip ratio adjusted for BMI |
| rs9379084 | 7231843 | 6 | RREB1 | Breast cancer |
| rs9505086 | 7232186 | 6 | RREB1 | Immature fraction of reticulocytes |
| rs75757892 | 7232389 | 6 | RREB1 | Hematocrit |
| rs2714333 | 7236620 | 6 | RREB1 | Steroid induced osteonecrosis of the femoral head and systemic lupus erythematosus |
| rs2714335 | 7238069 | 6 | RREB1 | Waist-to-hip ratio adjusted for BMI |
| rs77510523 | 7240053 | 6 | RREB1 | Alanine aminotransferase levels |
| rs2714337 | 7240577 | 6 | RREB1 | Fasting blood glucose (BMI interaction) |
| rs35742417 | 7247344 | 6 | RREB1 | Hemoglobin levels |
| rs78588343 | 7249460 | 6 | RREB1 | Triglyceride levels |
| rs3778321 | 7250270 | 6 | RREB1 | Glucose levels |
| rs2714338 | 7250960 | 6 | RREB1 | Balding type 1 |
| rs160631 | 52895230 | 6 | ICK | Smoking initiation (ever regular vs never regular) |
| rs222452 | 52905501 | 6 | ICK | Age of smoking initiation (MTAG) |
| rs2876 | 52963071 | 6 | FBXO9 | Smoking initiation |
| rs209489 | 53177678 | 6 | ELOVL5 | Survival in colorectal cancer (distant metastatic) |
| rs9367523 | 53207450 | 6 | ELOVL5 | Mean platelet volume |
| rs61583240 | 53383788 | 6 | GCLC | Lung adenocarcinoma |
| rs531557 | 53389995 | 6 | GCLC | Lung adenocarcinoma |
| rs534957 | 53406351 | 6 | GCLC | Benign prostatic hyperplasia |
| rs510088 | 53408789 | 6 | GCLC | Height |
| rs1325833 | 53896155 | 6 | MLIP | Height |
| rs4715438 | 53901329 | 6 | MLIP | Weight |
| rs9637973 | 53903152 | 6 | MLIP | Gamma glutamyl transferase levels |
| rs9370258 | 53910988 | 6 | MLIP | Gamma glutamyl transferase levels |
| rs7750294 | 53924170 | 6 | MLIP | LDL levels x SSRI defined daily dose interaction in schizophrenia or bipolar disorder |
| rs9296736 | 53924697 | 6 | MLIP | Liver enzyme levels (gamma-glutamyl transferase) |
| rs625899 | 53957588 | 6 | MLIP | Gamma glutamyl transpeptidase |
| rs499594 | 53975913 | 6 | MLIP | Pulse pressure |
| rs670463 | 53977495 | 6 | MLIP | Systolic blood pressure |
| rs1280946 | 53985339 | 6 | MLIP | Body mass index (MTAG) |
| rs631441 | 53994626 | 6 | MLIP | Pulse pressure |
| rs816366 | 53995504 | 6 | MLIP | Urinary sodium excretion |
| rs816367 | 53995542 | 6 | MLIP | Body mass index |
| rs816372 | 53997693 | 6 | MLIP | Pulse pressure |
| rs142516820 | 54003052 | 6 | MLIP | Creatinine levels |
| rs6915002 | 54028069 | 6 | MLIP | Heart failure (multivariate analysis) |
| rs10948810 | 54176227 | 6 | TINAG | Velopharyngeal dysfunction |
| rs1058768 | 54186147 | 6 | TINAG | Estimated glomerular filtration rate (creatinine) |
| rs9398629 | 121629167 | 6 | TBC1D32 | OTU99_45 (Faecalibacterium) prevalence |
| rs10456939 | 121764006 | 6 | GJA1 | Lung function (FEV1/FVC) |
| rs10456940 | 121764258 | 6 | GJA1 | White blood cell count |
| rs11961755 | 121766286 | 6 | GJA1 | HDL cholesterol levels |
| rs12206156 | 123596860 | 6 | TRDN | Gamma-glutamyltyrosine levels in elite athletes |
| rs12214992 | 123623065 | 6 | TRDN | Metabolite levels |
| rs78279606 | 123665041 | 6 | TRDN | Serum chromium levels |
| rs17737857 | 123711840 | 6 | TRDN | Intraocular pressure |
| rs9388238 | 123731285 | 6 | TRDN | 3-(4-hydroxyphenyl)lactate levels in elite athletes |
| rs12209641 | 123755664 | 6 | TRDN | Electrocardiogram morphology (amplitude at temporal datapoints) |
| rs4613836 | 123788961 | 6 | LOC101927990 | Body mass index |
| rs9320939 | 123818871 | 6 | TRDN | Hypertrophic cardiomyopathy (sarcomere negative) |

| | | | | |
|---|---|---|---|---|
| rs2180341 | 127600630 | 6 | RNF146 | Breast cancer |
| rs3823042 | 127641960 | 6 | ECHDC1 | Diffusing capacity of carbon monoxide |
| rs7775090 | 127785691 | 6 | SOGA3 | Heel bone mineral density |
| rs9372872 | 127808155 | 6 | SOGA3 | Urinary albumin-to-creatinine ratio in non-diabetics |
| rs62425513 | 128077738 | 6 | THEMIS | Uterine fibroids |
| rs12153861 | 128121143 | 6 | THEMIS | Height |
| rs4897233 | 128156888 | 6 | THEMIS | Celiac disease |
| rs9321099 | 128202139 | 6 | THEMIS | Sense of smell |
| rs3933376 | 128292071 | 6 | PTPRK | Allergic disease (asthma, hay fever or eczema) |
| rs55743914 | 128293562 | 6 | PTPRK | Celiac disease |
| rs35469349 | 128294709 | 6 | PTPRK | Allergic disease (asthma, hay fever or eczema) |
| rs9402011 | 128305888 | 6 | PTPRK | Verbal short term memory |
| rs13200150 | 128309768 | 6 | PTPRK | Generalized epilepsy |
| rs12055782 | 128312033 | 6 | PTPRK | Educational attainment |
| rs6936878 | 128312985 | 6 | PTPRK | Verbal short term memory |
| rs3903663 | 128313088 | 6 | PTPRK | Verbal short term memory |
| rs7744108 | 128324030 | 6 | PTPRK | Verbal short term memory |
| rs7742212 | 128328833 | 6 | PTPRK | Schizophrenia |
| rs13197257 | 128333682 | 6 | PTPRK | General cognitive ability |
| rs9321106 | 128355316 | 6 | PTPRK | Menarche (age at onset) |
| rs6938574 | 128390980 | 6 | PTPRK | Menarche (age at onset) |
| rs9484840 | 144382983 | 6 | PLAGL1 | Triacylglycerol (58:8) levels |
| rs7755842 | 144479462 | 6 | STX11 | Pre-treatment viral load in HIV-1 infection |
| rs9390172 | 144903517 | 6 | UTRN | Schizophrenia |
| rs6915747 | 144907979 | 6 | UTRN | Smoking initiation |
| rs9497026 | 144950023 | 6 | UTRN | Refractive error |
| rs4895647 | 144951709 | 6 | UTRN | Smoking initiation |
| rs11155364 | 144952491 | 6 | UTRN | Age when finished full-time education (standard GWA) |
| rs6929638 | 144965158 | 6 | UTRN | Educational attainment |
| rs488133 | 152125444 | 6 | ESR1 | Height |
| rs6914438 | 152129588 | 6 | ESR1 | Weight |
| rs532010 | 152130918 | 6 | ESR1 | Height |
| rs9371557 | 152140209 | 6 | ESR1 | Height |
| rs9479126 | 152146580 | 6 | ESR1 | Height |
| rs11155813 | 152149435 | 6 | ESR1 | Educational attainment (years of education) |
| rs6909023 | 152153697 | 6 | ESR1 | Height |
| rs6902771 | 152157881 | 6 | ESR1 | Waist circumference adjusted for body mass index |
| rs79575945 | 152158847 | 6 | ESR1 | Educational attainment |
| rs4870056 | 152162227 | 6 | ESR1 | Height |
| rs2234693 | 152163335 | 6 | ESR1 | Heel bone mineral density |
| rs9340799 | 152163381 | 6 | ESR1 | Multi-trait sex score |
| rs9322332 | 152166801 | 6 | ESR1 | Fat-free mass |
| rs3853252 | 152170247 | 6 | ESR1 | Hip circumference adjusted for BMI |
| rs827420 | 152177529 | 6 | ESR1 | Multi-trait sex score |
| rs9322334 | 152200041 | 6 | ESR1 | Whole body fat free mass (UKB data field 23101) |
| rs9322335 | 152200129 | 6 | ESR1 | Height |
| rs6557168 | 152201201 | 6 | ESR1 | Anxiety |
| rs2275336 | 154726988 | 6 | CNKSR3 | Parkinson's disease |
| rs11155972 | 154733748 | 6 | CNKSR3 | Height |
| rs9356034 | 162946186 | 6 | PARK2 | Educational attainment |
| rs2803073 | 162962828 | 6 | PARK2 | Peginterferon alfa-2a treatment response in chronic hepatitis B infection |
| rs957906 | 162969924 | 6 | PARK2 | Smoking initiation |
| rs2023016 | 162981266 | 6 | PARK2 | Educational attainment (years of education) |
| rs36007635 | 163009335 | 6 | PARK2 | Body mass index |
| rs9364664 | 163023054 | 6 | PARK2 | Educational attainment (years of education) |
| rs1893119 | 163036511 | 6 | PARK2 | Height |
| rs12190823 | 163092643 | 6 | PARK2 | Depression |
| rs55715230 | 163177570 | 6 | PACRG | Vitamin D insufficiency |
| rs9347687 | 163178326 | 6 | PACRG | Cognitive performance (MTAG) |
| rs1333957 | 163205160 | 6 | PACRG | Post-traumatic stress disorder |
| rs9365499 | 163271833 | 6 | PACRG | Body mass index |
| rs4709655 | 163280204 | 6 | PACRG | Insomnia |
| rs6455854 | 163304290 | 6 | PACRG | Educational attainment |
| rs141682404 | 163457196 | 6 | PACRG | Thromboembolic events in Sickle cell disease |
| rs79057730 | 784943 | 7 | HEATR2 | Initial pursuit acceleration |
| rs17499670 | 38489731 | 7 | AMPH | Height |
| rs3807411 | 38546848 | 7 | AMPH | Oleoyl-oleoyl-glycerol (18:1/18:1) [2] levels in elite athletes |
| rs9648078 | 43320413 | 7 | HECW1 | Obstructive sleep apnea trait (apnea hypopnea index, change over time) |
| rs4509217 | 43361008 | 7 | HECW1 | Obstructive sleep apnea trait (apnea hypopnea index, change over time) |
| rs10245574 | 43584521 | 7 | HECW1 | Caudal anterior-cingulate cortex volume |
| rs12702051 | 43599488 | 7 | HECW1 | Educational attainment |
| rs80271829 | 50443141 | 7 | IKZF1 | Monocyte count |
| rs78699863 | 50457621 | 7 | IKZF1 | Mean spheric corpuscular volume |
| rs7797772 | 50460208 | 7 | IKZF1 | Clostridium difficile infection in multiple myeloma |
| rs62447205 | 50465830 | 7 | IKZF1 | Type 1 diabetes |
| rs11978267 | 50466304 | 7 | IKZF1 | Acute lymphoblastic leukemia (childhood) |
| rs11980379 | 50469981 | 7 | IKZF1 | B cell acute lymphoblastic leukaemia (normal cytogenetics) |
| rs4132601 | 50470604 | 7 | IKZF1 | Acute lymphoblastic leukemia (childhood) |
| rs3807566 | 50564204 | 7 | DDC | Body mass index |

| | | | | |
|---|---|---|---|---|
| rs1037351 | 50565404 | 7 | DDC | Body mass index (MTAG) |
| rs4947581 | 50565967 | 7 | DDC | Body mass index |
| rs1376523 | 50569341 | 7 | DDC | Height |
| rs3823674 | 50571996 | 7 | DDC | Adult body size |
| rs4947584 | 50572709 | 7 | DDC | Body mass index |
| rs6944090 | 50573140 | 7 | DDC | Hemoglobin A1c levels |
| rs7777600 | 50592671 | 7 | DDC | Height |
| rs17133877 | 50608698 | 7 | DDC | Height |
| rs3807553 | 50610613 | 7 | DDC | Clostridium difficile infection in multiple myeloma |
| rs10499694 | 50614173 | 7 | DDC | Body mass index |
| rs11765748 | 50615616 | 7 | DDC | Body mass index |
| rs4947644 | 50618876 | 7 | DDC | Body mass index x sex x age interaction (4df test) |
| rs11555134 | 50659193 | 7 | GRB10 | Height |
| rs4245555 | 50661409 | 7 | GRB10 | Morningness |
| rs9656664 | 50661972 | 7 | GRB10 | Mean corpuscular hemoglobin |
| rs12540874 | 50664922 | 7 | GRB10 | Systemic sclerosis |
| rs2329485 | 50675760 | 7 | GRB10 | 3-methoxytyrosine levels |
| rs11769934 | 50691194 | 7 | GRB10 | Height |
| rs10486757 | 50693858 | 7 | GRB10 | Red blood cell count |
| rs6948959 | 50697051 | 7 | GRB10 | Blood pressure (pleiotropy model 1 DBP adjusted for estimated causal effects x SBP) |
| rs3807546 | 50723256 | 7 | GRB10 | Corpus callosum mid-anterior volume |
| rs2237457 | 50726144 | 7 | GRB10 | Schizophrenia (treatment resistant) |
| rs2237459 | 50729428 | 7 | GRB10 | Severe COVID-19 infection |
| rs2715093 | 50733034 | 7 | GRB10 | Waist circumference adjusted for body mass index |
| rs17449280 | 50734418 | 7 | GRB10 | Eosinophil counts |
| rs2529414 | 50737831 | 7 | GRB10 | Hip circumference adjusted for BMI |
| rs73116822 | 50737852 | 7 | GRB10 | Estimated glomerular filtration rate |
| rs2529411 | 50738482 | 7 | GRB10 | Waist-hip ratio |
| rs17152083 | 50739610 | 7 | GRB10 | Estimated glomerular filtration rate |
| rs73116829 | 50739738 | 7 | GRB10 | Estimated glomerular filtration rate |
| rs886821 | 50743095 | 7 | GRB10 | Waist circumference adjusted for body mass index |
| rs2237474 | 50743240 | 7 | GRB10 | Brain region volumes |
| rs56200772 | 50743947 | 7 | GRB10 | Kidney cancer |
| rs2715133 | 50749425 | 7 | GRB10 | Height |
| rs2715135 | 50750128 | 7 | GRB10 | Waist-hip ratio |
| rs10248619 | 50751090 | 7 | GRB10 | Fasting blood glucose (BMI interaction) |
| rs2282930 | 50754679 | 7 | GRB10 | Femoral neck bone mineral density |
| rs73118816 | 50756760 | 7 | GRB10 | Serum creatinine levels |
| rs933360 | 50758245 | 7 | GRB10 | Insulin levels adjusted for BMI |
| rs2190502 | 50763624 | 7 | GRB10 | Glucose levels |
| rs2299156 | 50764404 | 7 | GRB10 | Educational attainment (years of education) |
| rs73118830 | 50764948 | 7 | GRB10 | Serum uric acid levels |
| rs1978208 | 50785448 | 7 | GRB10 | Alcohol-associated liver disease in non drinkers |
| rs10265504 | 50786050 | 7 | GRB10 | Glucose levels |
| rs2108349 | 50786663 | 7 | GRB10 | Random glucose levels |
| rs6943153 | 50791579 | 7 | GRB10 | Fasting glucose |
| rs6968827 | 50802434 | 7 | GRB10 | Age-related hearing impairment (MTAG) |
| rs1962390 | 50806947 | 7 | GRB10 | Brain region volumes |
| rs12536500 | 50808033 | 7 | GRB10 | Subcortical volume (min-P) |
| rs13236710 | 50809085 | 7 | GRB10 | Height |
| rs17152120 | 50815054 | 7 | GRB10 | Height |
| rs6976572 | 50839798 | 7 | GRB10 | Height |
| rs11238325 | 50853151 | 7 | GRB10 | Hearing problems |
| rs7791286 | 50856792 | 7 | GRB10 | Fasting plasma glucose |
| rs38098 | 78238766 | 7 | MAGI2 | Computer vision syndrome |
| rs73145178 | 78315536 | 7 | MAGI2 | Smoking initiation |
| rs9969351 | 78324243 | 7 | MAGI2 | SSRI response to core syndromal factor measured by HAM-D-21 in major depressive disorder |
| rs1496765 | 78331354 | 7 | MAGI2 | Smoking initiation |
| rs1496766 | 78331465 | 7 | MAGI2 | Acute lymphoblastic leukemia (childhood) |
| rs4727751 | 78336934 | 7 | MAGI2 | Schizophrenia |
| rs7793494 | 78344389 | 7 | MAGI2 | COVID-19 (covid vs negative) |
| rs2237527 | 82012870 | 7 | CACNA2D1 | Severe COVID-19 infection |
| rs17156280 | 82043990 | 7 | CACNA2D1 | Major depressive disorder (stressful life events interaction) |
| rs2299187 | 82047102 | 7 | CACNA2D1 | Survival in head and neck cancer |
| rs6465454 | 94702805 | 7 | PPP1R9A | Height |
| rs9649194 | 94754985 | 7 | PPP1R9A | Self-reported math ability (MTAG) |
| rs10953137 | 94768272 | 7 | PPP1R9A | Self-reported math ability |
| rs854524 | 94898811 | 7 | PPP1R9A | Height |
| rs854534 | 94909495 | 7 | PPP1R9A | Total cholesterol levels in very large HDL |
| rs854555 | 94930391 | 7 | PON1 | Response to TNF antagonist treatment |
| rs2237582 | 94934200 | 7 | PON1 | Diacylglycerol kinase beta levels |
| rs2269829 | 94936129 | 7 | PON1 | Group XIIB secretory phospholipase A2-like protein levels |
| rs662 | 94937446 | 7 | PON1 | Sulfiredoxin-1 levels |
| rs2057681 | 94938257 | 7 | PON1 | Metabolite levels |
| rs3917534 | 94939797 | 7 | PON1 | E3 ubiquitin-protein ligase RNF8 levels |
| rs3917532 | 94940119 | 7 | PON1 | Allergin-1 levels (MILR1.11173.29.3) |
| rs3917529 | 94940235 | 7 | PON1 | Protein S100-P levels |
| rs1157745 | 94941038 | 7 | PON1 | Proteasome subunit beta type-9 levels |
| rs2299257 | 94942765 | 7 | PON1 | Transgelin-2 levels |

| | | | | |
|---|---|---|---|---|
| rs854556 | 94944923 | 7 | PON1 | T-complex protein 11 homolog levels |
| rs854560 | 94946084 | 7 | PON1 | Atopic dermatitis (moderate to severe) |
| rs854562 | 94947969 | 7 | PON1 | Triglycerides in very large HDL |
| rs13226149 | 95025600 | 7 | PON3 | JT interval |
| rs7493 | 95034775 | 7 | PON2 | Yu-Zhi constitution type in type 2 diabetes |
| rs12026 | 95041016 | 7 | PON2 | DNPH1/PSMG3 protein level ratio |
| rs2068604 | 95041542 | 7 | PON2 | QT interval |
| rs1034809 | 95051652 | 7 | PON2 | JT interval |
| rs2158806 | 95051998 | 7 | PON2 | QT interval |
| rs12534274 | 95057510 | 7 | PON2 | Cholesterol to total lipids in very large HDL percentage (UKB data field 23630) |
| rs10270813 | 129256285 | 7 | NRF1 | Alzheimer's disease polygenic risk score (upper quantile vs lower quantile) |
| rs4728175 | 129354911 | 7 | NRF1 | Resistance to COVID-19 infection (Exposed negative vs positive) |
| rs9641855 | 129356206 | 7 | NRF1 | Obesity-related traits |
| rs76585997 | 129361115 | 7 | NRF1 | Insomnia in chronic migraine |
| rs11557288 | 129396174 | 7 | NRF1 | Red cell distribution width |
| rs12668638 | 158871812 | 7 | VIPR2 | CPB1/CTRB1 protein level ratio |
| rs184020833 | 158888728 | 7 | VIPR2 | Ischemic stroke in diabetes mellitus |
| rs7810170 | 158914170 | 7 | VIPR2 | Femur bone mineral density x serum urate levels interaction |
| rs144364340 | 158920618 | 7 | VIPR2 | Spherical equivalent |
| rs435969 | 158928880 | 7 | VIPR2 | 3-hydroxysebacate levels in elite athletes |
| rs11787412 | 3134239 | 8 | CSMD1 | Adolescent idiopathic scoliosis |
| rs9314485 | 3178721 | 8 | CSMD1 | RBC levels of Indoxyl |
| rs7013161 | 3292247 | 8 | CSMD1 | OTU97_8 (Ruminococcaceae) abundance |
| rs17394429 | 3316986 | 8 | CSMD1 | Obesity-related traits |
| rs11777685 | 3344204 | 8 | CSMD1 | COVID-19 (severe respiratory symptoms vs population) |
| rs1530615 | 3348853 | 8 | CSMD1 | Nicotine dependence symptom count |
| rs79021388 | 3363460 | 8 | CSMD1 | Neuroblastoma |
| rs2980438 | 8094825 | 8 | FAM86B3P | Waist-to-hip ratio adjusted for BMI |
| rs12549973 | 8176554 | 8 | SGK223 | Serum phosphate levels |
| rs1548198 | 8178223 | 8 | SGK223 | Body mass index (MTAG) |
| rs2945908 | 8192229 | 8 | SGK223 | Squamous cell lung carcinoma |
| rs11785239 | 8205010 | 8 | SGK223 | Carotid intima media thickness |
| rs4840941 | 8216986 | 8 | SGK223 | Waist circumference adjusted for body mass index |
| rs7005904 | 8218597 | 8 | SGK223 | Appendicular lean mass |
| rs4840338 | 8218967 | 8 | SGK223 | Body mass index |
| rs1914826 | 8221088 | 8 | SGK223 | Tyrosine levels in elite athletes |
| rs78553524 | 15974136 | 8 | MSR1 | Protein quantitative trait loci (liver) |
| rs73665257 | 16049656 | 8 | MSR1 | Galectin-3-binding protein levels |
| rs4921617 | 18782047 | 8 | PSD3 | Neutrophil count |
| rs13279936 | 18800163 | 8 | PSD3 | Severe COVID-19 infection |
| rs1365600 | 18856363 | 8 | PSD3 | Sub-cortical grey matter volume |
| rs7841763 | 123971081 | 8 | ZHX2 | Height |
| rs10108684 | 123973068 | 8 | ZHX2 | Immune response to smallpox vaccine (IL-6) |
| rs11783086 | 123980448 | 8 | ZHX2 | Height |
| rs11779459 | 123980551 | 8 | ZHX2 | Height |
| rs11785093 | 123981872 | 8 | ZHX2 | Mean reticulocyte volume |
| rs3802266 | 123985708 | 8 | ZHX2 | Hemoglobin concentration |
| rs2833 | 123986649 | 8 | ZHX2 | Height |
| rs12542856 | 130773458 | 8 | GSDMC | Hip circumference adjusted for BMI |
| rs12544672 | 130786451 | 8 | GSDMC | Hip circumference adjusted for BMI |
| rs837065 | 130853447 | 8 | FAM49B | Educational attainment (years of education) |
| rs2086641 | 130901909 | 8 | FAM49B | Parkinson's disease or first degree relation to individual with Parkinson's disease |
| rs837080 | 130925116 | 8 | FAM49B | Educational attainment |
| rs401526 | 130929166 | 8 | FAM49B | Educational attainment (years of education) |
| rs298601 | 130932427 | 8 | FAM49B | Educational attainment (years of education) |
| rs16904179 | 130958951 | 8 | FAM49B | Post-traumatic stress disorder (asjusted for relatedness) |
| rs16904181 | 130976074 | 8 | FAM49B | Lymphocyte count |
| rs6470788 | 131005345 | 8 | FAM49B | 3-(4-hydroxyphenyl)lactate levels in elite athletes |
| rs7867822 | 20676454 | 9 | FOCAD | Smoking initiation (ever regular vs never regular) |
| rs7043296 | 20747070 | 9 | FOCAD | Medial orbital frontal cortex volume |
| rs17759355 | 20805089 | 9 | FOCAD | Red blood cell count |
| rs10811408 | 20805270 | 9 | FOCAD | Mean corpuscular volume |
| rs10811412 | 20831382 | 9 | FOCAD | Mean corpuscular hemoglobin |
| rs10811413 | 20831512 | 9 | FOCAD | Mean corpuscular hemoglobin |
| rs76677990 | 20863673 | 9 | FOCAD | Vaginal microbiome MetaCyc pathway (FUC-RHAMCAT-PWY\|superpathway of fucose and rhamnose degradation) |
| rs12379622 | 20890316 | 9 | FOCAD | Red blood cell count |
| rs2383162 | 20908513 | 9 | FOCAD | Cleft lip with or without cleft palate x maternal periconceptional smoking interaction (parent of origin effect) |
| rs10811438 | 20962282 | 9 | FOCAD | Daytime nap |
| rs2275882 | 21008377 | 9 | PTPLAD2 | Erectile dysfunction |
| rs10964794 | 21014003 | 9 | PTPLAD2 | interferon-related traits |
| rs12551341 | 21014628 | 9 | PTPLAD2 | interferon-related traits |
| rs2790732 | 33778629 | 9 | PRSS3 | Physical activity (standard GWA) |
| rs3843935 | 33787871 | 9 | PRSS3 | Triglycerides |
| rs2005617 | 33791164 | 9 | PRSS3 | Trypsin-3 levels |
| rs855719 | 33817617 | 9 | UBE2R2 | Refractive error |
| rs10971712 | 33820938 | 9 | UBE2R2 | Body mass index (MTAG) |
| rs1749268 | 33837330 | 9 | UBE2R2 | Immature fraction of reticulocytes |
| rs307646 | 33932338 | 9 | UBAP2 | Osteoporosis |
| rs307660 | 33942930 | 9 | UBAP2 | Bipolar disorder |

| | | | | |
|---|---|---|---|---|
| rs544169 | 33956791 | 9 | UBAP2 | Estimated glomerular filtration rate |
| rs307680 | 33971546 | 9 | UBAP2 | Mosquito bite size |
| rs307703 | 33985465 | 9 | UBAP2 | Brain morphology (MOSTest) |
| rs12555291 | 34011391 | 9 | UBAP2 | High light scatter reticulocyte count |
| rs1758632 | 34025640 | 9 | UBAP2 | Type 2 diabetes |
| rs11557154 | 34107505 | 9 | DCAF12 | Neutrophil percentage of white cells |
| rs10971938 | 34120720 | 9 | DCAF12 | Height |
| rs2275003 | 34124860 | 9 | DCAF12 | Body mass index |
| rs7020310 | 34583962 | 9 | CNTFR | Free testosterone levels |
| rs6476455 | 34588006 | 9 | CNTFR | Metabolite levels |
| rs13339 | 34610702 | 9 | RPP25L | Vaginal microbiome MetaCyc pathway (TYRFUMCAT-PWY\|L-tyrosine degradation I) |
| rs10972168 | 34619621 | 9 | DCTN3 | Bullous pemphigoid (except dipeptidyl peptidase-4 inhibitor-induced noninflammatory) |
| rs3808869 | 34622389 | 9 | ARID3C | Diastolic blood pressure |
| rs41274865 | 34647802 | 9 | GALT | Serum levels of protein IL11RA |
| rs41274867 | 34648088 | 9 | GALT | Blood protein levels |
| rs2070074 | 34649442 | 9 | GALT | CTACK levels |
| rs11575578 | 34656479 | 9 | IL11RA | Interleukin-11 receptor subunit alpha levels |
| rs11574915 | 34710084 | 9 | CCL21 | C-C motif chemokine 19 levels |
| rs1415221 | 73480718 | 9 | TRPM3 | Electrocardiogram morphology (amplitude at temporal datapoints) |
| rs6560168 | 73482647 | 9 | TRPM3 | PR interval |
| rs17471974 | 73496967 | 9 | TRPM3 | PR interval |
| rs2993001 | 73567782 | 9 | TRPM3 | Diacylglycerol (16:1/18:2 [2], 16:0/18:3 [1]) levels in elite athletes |
| rs1558926 | 73598465 | 9 | TRPM3 | Height |
| rs962001 | 73636237 | 9 | TRPM3 | Metabolite levels |
| rs11526468 | 92003679 | 9 | SEMA4D | High light scatter reticulocyte count |
| rs13287803 | 92006536 | 9 | SEMA4D | Youthful appearance (self-reported) |
| rs6559349 | 92007225 | 9 | SEMA4D | Coronary artery calcification in type 2 diabetes (excluding prevalent cardiovascular disease) |
| rs41287361 | 92017733 | 9 | SEMA4D | Mean spheric corpuscular volume |
| rs56339712 | 92025757 | 9 | SEMA4D | Reticulocyte count |
| rs62547238 | 92027498 | 9 | SEMA4D | Reticulocyte count |
| rs4132699 | 92036427 | 9 | SEMA4D | Nonsyndromic cleft lip with or without cleft palate |
| rs4877094 | 92078518 | 9 | SEMA4D | IgA nephropathy |
| rs1411156 | 92261864 | 9 | UNQ6494 | Height |
| rs11265853 | 92263349 | 9 | UNQ6494 | Hemoglobin concentration |
| rs78343805 | 92264395 | 9 | UNQ6494 | Estimated glomerular filtration rate |
| rs78040893 | 92269038 | 9 | UNQ6494 | Estimated glomerular filtration rate (creatinine) |
| rs7040551 | 92274114 | 9 | UNQ6494 | C-reactive protein levels (MTAG) |
| rs4876993 | 92281403 | 9 | UNQ6494 | Sex hormone-binding globulin levels adjusted for BMI |
| rs61311941 | 92285323 | 9 | UNQ6494 | Cystatin C levels |
| rs883924 | 93181531 | 9 | LOC101927873 | Hepatitis C induced liver fibrosis |
| rs1551751 | 93255874 | 9 | LOC340515 | Asthma exacerbations |
| rs73503347 | 93331998 | 9 | LOC340515 | Cerebellum cortex volume |
| rs553405 | 93384978 | 9 | DIRAS2 | Mortality in COVID-19 |
| rs1319677 | 93565999 | 9 | SYK | Monocyte count |
| rs2780701 | 93567281 | 9 | SYK | Platelet distribution width |
| rs1333634 | 93567503 | 9 | SYK | Platelet count |
| rs1333633 | 93567572 | 9 | SYK | Monocyte count |
| rs919386 | 93567607 | 9 | SYK | Plateletcrit |
| rs1333632 | 93567732 | 9 | SYK | Monocyte count |
| rs571599993 | 93581131 | 9 | SYK | Vaginal microbiome relative abundance (g_Streptococcus) |
| rs2278278 | 93608739 | 9 | SYK | Hippurate levels in elite athletes |
| rs4744016 | 93614898 | 9 | SYK | Tyrosine-protein kinase SYK levels |
| rs56049491 | 93980558 | 9 | AUH | Total bilirubin levels |
| rs6479336 | 94110288 | 9 | AUH | Eosinophil counts |
| rs7020893 | 94121330 | 9 | AUH | Eosinophil counts |
| rs2230578 | 94485928 | 9 | ROR2 | Facial morphology |
| rs10761129 | 94486321 | 9 | ROR2 | Nose size |
| rs10820900 | 94495608 | 9 | ROR2 | Tyrosine-protein kinase transmembrane receptor ROR2 levels |
| rs17587223 | 94600207 | 9 | ROR2 | Height |
| rs140064974 | 94604680 | 9 | ROR2 | Vaginal microbiome MetaCyc pathway (P163-PWY\|L-lysine fermentation to acetate and butanoate) |
| rs115132021 | 94657553 | 9 | ROR2 | wg lh intensity-contrast precuneus |
| rs4275276 | 94662604 | 9 | ROR2 | Sib-shared facial trait 763; Facial segment 38; 3D morphology of the upper vermillion |
| rs76684062 | 94691627 | 9 | ROR2 | Glaucoma |
| rs16908003 | 94691644 | 9 | ROR2 | Glaucoma |
| rs77094301 | 94695329 | 9 | ROR2 | Glaucoma |
| rs10992216 | 94816157 | 9 | SPTLC1 | Glaucoma |
| rs7872515 | 94822540 | 9 | SPTLC1 | Bipolar disorder and schizophrenia |
| rs12235495 | 94829905 | 9 | SPTLC1 | Glaucoma |
| rs79161841 | 94836389 | 9 | SPTLC1 | Glaucoma |
| rs11533009 | 94837832 | 9 | SPTLC1 | Glaucoma |
| rs77450572 | 94839129 | 9 | SPTLC1 | Glaucoma |
| rs59856128 | 94842511 | 9 | SPTLC1 | Glaucoma |
| rs16908135 | 94842715 | 9 | SPTLC1 | Glaucoma |
| rs16908137 | 94844150 | 9 | SPTLC1 | Glaucoma |
| rs59183854 | 94854626 | 9 | SPTLC1 | Glaucoma |
| rs10992230 | 94856517 | 9 | SPTLC1 | Glaucoma |
| rs7862826 | 94856819 | 9 | SPTLC1 | Glaucoma |
| rs10992233 | 94875980 | 9 | SPTLC1 | Glaucoma |
| rs10992252 | 94912524 | 9 | LINC00475 | Glaucoma |

| | | | | |
|---|---|---|---|---|
| rs471478 | 95001109 | 9 | IARS | COL1A1/OMD protein level ratio |
| rs17678735 | 95077827 | 9 | NOL8 | Height |
| rs7042335 | 95122547 | 9 | CENPP | Growth/differentiation factor 2 levels |
| rs10283572 | 95133074 | 9 | CENPP | Lung function (FEV1) |
| rs7045409 | 95201540 | 9 | CENPP | Mimecan levels |
| rs8067 | 95218829 | 9 | ASPN | IGFBP4/OGN protein level ratio |
| rs3174352 | 95219248 | 9 | ASPN | Urate levels |
| rs13301537 | 95229047 | 9 | ASPN | Height |
| rs7869742 | 95256174 | 9 | CENPP | Obstructive sleep apnea |
| rs4743873 | 95270781 | 9 | CENPP | Height |
| rs72754434 | 95283708 | 9 | CENPP | Noncompact myelin-associated protein levels |
| rs2131919 | 95283887 | 9 | CENPP | Triglyceride levels |
| rs10491806 | 95284212 | 9 | CENPP | Noncompact myelin-associated protein levels |
| rs17462606 | 95370664 | 9 | CENPP | Height |
| rs2148537 | 95380726 | 9 | IPPK | Neutrophil count |
| rs12686780 | 95382297 | 9 | IPPK | HDL cholesterol levels |
| rs1323523 | 95393049 | 9 | IPPK | Serum levels of protein NCMAP |
| rs17366134 | 95406157 | 9 | IPPK | Height |
| rs9969804 | 95429120 | 9 | IPPK | Height |
| rs10992433 | 95483503 | 9 | BICD2 | Neutrophil count |
| rs755209 | 95489671 | 9 | BICD2 | Haemorrhoidal disease |
| rs10992437 | 95494447 | 9 | BICD2 | White blood cell count |
| rs10992447 | 95509733 | 9 | BICD2 | Waist-to-hip ratio adjusted for BMI |
| rs12236754 | 95512201 | 9 | BICD2 | Appendicular lean mass |
| rs912262 | 95525557 | 9 | BICD2 | Height |
| rs7033638 | 95883785 | 9 | NINJ1 | Lactate dehydrogenase levels |
| rs3793662 | 95886051 | 9 | NINJ1 | CD25 on B cell |
| rs2275848 | 95887320 | 9 | NINJ1 | Obesity (early onset extreme) |
| rs12235416 | 95888667 | 9 | NINJ1 | Aspartate aminotransferase levels |
| rs60629714 | 95889160 | 9 | NINJ1 | CD40 on CD14+ CD16- monocyte |
| rs7018885 | 95892726 | 9 | NINJ1 | Alanine aminotransferase levels |
| rs12342201 | 95894964 | 9 | NINJ1 | Lactate dehydrogenase levels |
| rs10821083 | 95951477 | 9 | WNK2 | Brain morphology (MOSTest) |
| rs7041800 | 95951769 | 9 | WNK2 | Total cholesterol levels (MTAG) |
| rs10992671 | 95952870 | 9 | WNK2 | Node-level brain connectivity (multivariate analysis) |
| rs12349094 | 95953313 | 9 | WNK2 | Educational attainment |
| rs4743902 | 95965696 | 9 | WNK2 | Smoking initiation |
| rs4617217 | 95988968 | 9 | WNK2 | Coronary artery disease |
| rs13290796 | 95991547 | 9 | WNK2 | Cortical thickness |
| rs10821093 | 95993619 | 9 | WNK2 | Corpus callosum fractional anisotropy (MOSTest) |
| rs12341303 | 96003435 | 9 | WNK2 | Highest math class taken (MTAG) |
| rs10821097 | 96012085 | 9 | WNK2 | Cognitive performance (MTAG) |
| rs10761202 | 96019970 | 9 | WNK2 | Educational attainment (years of education) |
| rs11790388 | 129740283 | 9 | RALGPS1 | Schizophrenia |
| rs2297866 | 129854199 | 9 | ANGPTL2 | Cortical surface area |
| rs13298677 | 129862996 | 9 | ANGPTL2 | Height |
| rs11795066 | 129863168 | 9 | ANGPTL2 | Intraocular pressure |
| rs2787594 | 129863545 | 9 | ANGPTL2 | Hair color |
| rs1974291 | 129874095 | 9 | ANGPTL2 | Primary open angle glaucoma (MTAG) |
| rs1281154 | 129891749 | 9 | RALGPS1 | DNA methylation variation (age effect) |
| rs868852 | 129899510 | 9 | RALGPS1 | Major depressive disorder (broad) |
| rs4130590 | 130107964 | 9 | GARNL3 | Bipolar disorder |
| rs7041940 | 130109746 | 9 | GARNL3 | Bipolar disorder |
| rs7029536 | 130110411 | 9 | GARNL3 | Metabolite levels (HVA) |
| rs10819287 | 130166382 | 9 | SLC2A8 | Rate of cognitive decline in Alzheimer's disease |
| rs13292096 | 130191186 | 9 | ZNF79 | Palmitoyl-oleoyl-glycerol (16:0/18:1) [2] levels in elite athletes |
| rs10987668 | 130251615 | 9 | LRSAM1 | Height |
| rs2244843 | 130271666 | 9 | FAM129B | White blood cell count |
| rs2251551 | 130284652 | 9 | FAM129B | Plateletcrit |
| rs1891732 | 130286428 | 9 | FAM129B | Mean corpuscular hemoglobin |
| rs2247361 | 130297409 | 9 | FAM129B | Alzheimer’s disease polygenic risk score (upper quantile vs lower quantile) |
| rs1891730 | 130309028 | 9 | FAM129B | Systolic blood pressure |
| rs72767980 | 130320557 | 9 | FAM129B | Heel bone mineral density |
| rs2252730 | 130323485 | 9 | FAM129B | Systolic blood pressure |
| rs2249851 | 130324154 | 9 | FAM129B | Asthma in gastrointestinal disease |
| rs2249101 | 130330036 | 9 | FAM129B | Retinal venular tortuosity |
| rs4240418 | 130334178 | 9 | FAM129B | Retinal arteriolar tortuosity |
| rs10760490 | 130335418 | 9 | FAM129B | Motor fluctuations in levodopa treated Parkinson's disease |
| rs6478788 | 130376248 | 9 | STXBP1 | Prostate cancer |
| rs10819303 | 130392954 | 9 | STXBP1 | Educational attainment (MTAG) |
| rs2241167 | 130430116 | 9 | STXBP1 | Prostate cancer |
| rs550045 | 130477160 | 9 | PTRH1 | N-formylmethionine levels |
| rs504434 | 130477768 | 9 | PTRH1 | Serum metabolite levels |
| rs13298768 | 130489743 | 9 | TTC16 | Pulse pressure |
| rs514024 | 130504070 | 9 | SH2D3C | Eating disorders (purging via substances) |
| rs10739694 | 130526605 | 9 | SH2D3C | Granulocyte percentage of myeloid white cells |
| rs10739695 | 130527677 | 9 | SH2D3C | Monocyte percentage of white cells |
| rs4836579 | 130528548 | 9 | SH2D3C | Basophil percentage of white cells |
| rs2417054 | 130532639 | 9 | SH2D3C | Neutrophil percentage of white cells |

| | | | | |
|---|---|---|---|---|
| rs2417055 | 130532772 | 9 | SH2D3C | Monocyte count |
| rs10760500 | 130536717 | 9 | SH2D3C | Height |
| rs10760503 | 130578404 | 9 | ENG | Height |
| rs12001427 | 130585031 | 9 | ENG | Red blood cell count |
| rs11794565 | 130601277 | 9 | ENG | Endoglin levels |
| rs11545664 | 130605385 | 9 | ENG | Hemoglobin levels |
| rs41492548 | 130607359 | 9 | ENG | Monocyte count |
| rs10739700 | 130672221 | 9 | ST6GALNAC4 | Height |
| rs1043060 | 130705120 | 9 | FAM102A | Serum creatinine levels |
| rs62585312 | 130723419 | 9 | FAM102A | Coiled-coil domain-containing protein 80 levels |
| rs7036499 | 130844744 | 9 | SLC25A25 | Height |
| rs6478851 | 131561110 | 9 | TBC1D13 | High density lipoprotein cholesterol levels |
| rs7858732 | 131563540 | 9 | TBC1D13 | Waist-to-hip ratio adjusted for BMI |
| rs3829057 | 131567982 | 9 | TBC1D13 | Plasma 3-indoleglyoxylic acid levels in chronic kidney disease |
| rs45609236 | 131570178 | 9 | TBC1D13 | Phenylpyruvate levels |
| rs15676 | 131572027 | 9 | TBC1D13 | Indolelactate levels |
| rs7029757 | 132566666 | 9 | TOR1B | Alanine aminotransferase levels |
| rs12634 | 132573536 | 9 | TOR1B | Prostate cancer |
| rs1182 | 132576060 | 9 | TOR1A | Prostate cancer |
| rs2296793 | 132585058 | 9 | TOR1A | Bone mineral density (hip) and hip bone size |
| rs73672511 | 132611051 | 9 | USP20 | C-reactive protein levels |
| rs72757229 | 132653055 | 9 | FNBP1 | Physiological dysregulation (Mahalanobis distance) |
| rs2295952 | 132681784 | 9 | FNBP1 | Gut microbiota relative abundance (unassigned genus belonging to family Clostridiales) |
| rs2007126 | 132684007 | 9 | FNBP1 | LDL peak particle diameter (total fat intake interaction) |
| rs139762218 | 132700592 | 9 | FNBP1 | Diastolic blood pressure change trajectory |
| rs12722561 | 6069893 | 10 | IL2RA | Epstein Barr virus nuclear antigen 1 IgG levels or multiple sclerosis |
| rs1323657 | 6072427 | 10 | IL2RA | Progression free survival in serous epithelial ovarian cancer treated with carboplatin and paclitaxel |
| rs11816044 | 6074082 | 10 | IL2RA | Age of onset of adult onset asthma |
| rs7893467 | 6079035 | 10 | IL2RA | Hypothyroidism |
| rs12722515 | 6081230 | 10 | IL2RA | Crohn's disease |
| rs1107345 | 6087295 | 10 | IL2RA | Marginal zone lymphoma or multiple sclerosis |
| rs791590 | 6090322 | 10 | IL2RA | Soluble interleukin-2 receptor subunit alpha |
| rs7090504 | 6091017 | 10 | IL2RA | Lymphocyte count |
| rs10795763 | 6096199 | 10 | IL2RA | Systemic lupus erythematosus |
| rs706779 | 6098824 | 10 | IL2RA | Graves' disease |
| rs706778 | 6098949 | 10 | IL2RA | Rheumatoid arthritis |
| rs2104286 | 6099045 | 10 | IL2RA | Chronic inflammatory diseases (ankylosing spondylitis, Crohn's disease, psoriasis, primary sclerosing cholangitis, ulcerative colitis) (pleiotropy) |
| rs3134883 | 6100725 | 10 | IL2RA | Platelet-to-lymphocyte ratio |
| rs3118469 | 6101129 | 10 | IL2RA | Lymphocyte count |
| rs3118470 | 6101713 | 10 | IL2RA | Alopecia areata |
| rs12722489 | 6102012 | 10 | IL2RA | Blood protein levels |
| rs3118471 | 6102757 | 10 | IL2RA | Chronic inflammatory diseases (ankylosing spondylitis, Crohn's disease, psoriasis, primary sclerosing cholangitis, ulcerative colitis) (pleiotropy) |
| rs17566555 | 12275947 | 10 | CDC123 | Coronary artery disease |
| rs7090277 | 12278021 | 10 | CDC123 | Lung function (FEV1) |
| rs57062879 | 12278525 | 10 | CDC123 | Systolic blood pressure |
| rs3013016 | 12393488 | 10 | CAMK1D | Educational attainment |
| rs10752262 | 12395100 | 10 | CAMK1D | Cognitive ability, years of educational attainment or schizophrenia (pleiotropy) |
| rs80016651 | 12410455 | 10 | CAMK1D | Vaginal microbiome MetaCyc pathway (PWY-5505\|L-glutamate and L-glutamine biosynthesis) |
| rs4451621 | 12431367 | 10 | CAMK1D | Educational attainment |
| rs4750211 | 12435379 | 10 | CAMK1D | Obesity-related traits |
| rs2154248 | 12436894 | 10 | CAMK1D | Gut microbiota (bacterial taxa, hurdle binary method) |
| rs10906142 | 12438783 | 10 | CAMK1D | Obesity-related traits |
| rs10906143 | 12439476 | 10 | CAMK1D | Gut microbiota (bacterial taxa, rank normal transformation method) |
| rs12254030 | 12448088 | 10 | CAMK1D | Height |
| rs72767702 | 12460946 | 10 | CAMK1D | Vaginal microbiome MetaCyc pathway (PWY-6562\|norspermidine biosynthesis) |
| rs6602588 | 12487996 | 10 | CAMK1D | Systemic lupus erythematosus (MTAG) |
| rs1108131 | 12497747 | 10 | CAMK1D | Response to cyclophosphamide in systemic lupus erythematosus with lupus nephritis |
| rs2768367 | 12497770 | 10 | CAMK1D | Cholate levels in elite athletes |
| rs1536531 | 12522144 | 10 | CAMK1D | 3beta-hydroxy-5-cholestenoate levels in elite athletes |
| rs1111370 | 12535377 | 10 | CAMK1D | Pelvic organ prolapse x age interaction |
| rs11257835 | 12544852 | 10 | CAMK1D | Paraxanthine levels in elite athletes |
| rs41355846 | 12547384 | 10 | CAMK1D | Height |
| rs11257872 | 12580664 | 10 | CAMK1D | G_Porphyromonadaceae abundance |
| rs2648708 | 12601439 | 10 | CAMK1D | Calcium levels |
| rs17152263 | 12831794 | 10 | CAMK1D | Color vision defects (Deutan-Protan) |
| rs11258541 | 13749228 | 10 | FRMD4A | Red cell distribution width |
| rs1541010 | 13755544 | 10 | FRMD4A | RR interval (heart rate) |
| rs10906466 | 13790265 | 10 | FRMD4A | Migraine |
| rs4750407 | 13795471 | 10 | FRMD4A | Craniofacial microsomia |
| rs10906496 | 13854618 | 10 | FRMD4A | Resistance to COVID-19 infection (Exposed negative vs positive) |
| rs12263011 | 13865505 | 10 | FRMD4A | Theta electroencephalogram power |
| rs11258652 | 13868855 | 10 | FRMD4A | Daytime nap |
| rs11258654 | 13869173 | 10 | FRMD4A | Resistance to COVID-19 infection (Exposed negative vs positive) |
| rs11258656 | 13869240 | 10 | FRMD4A | Resistance to COVID-19 infection (Exposed negative vs positive) |
| rs11258657 | 13869245 | 10 | FRMD4A | Resistance to COVID-19 infection (Exposed negative vs positive) |
| rs10906522 | 13907236 | 10 | FRMD4A | Breast cancer |
| rs10906545 | 13978235 | 10 | FRMD4A | Chronic obstructive pulmonary disease |
| rs4750440 | 14013954 | 10 | FRMD4A | Adiponectin levels |
| rs2292368 | 14050695 | 10 | FRMD4A | Fibroblast growth factor basic levels |

| | | | | |
|---|---|---|---|---|
| rs2457837 | 14051456 | 10 | FRMD4A | Takayasu arteritis |
| rs55829876 | 14077623 | 10 | FRMD4A | Drinks per week |
| rs1218373 | 14296716 | 10 | FRMD4A | interferon-related traits |
| rs10508477 | 14598740 | 10 | FAM107B | Platelet count |
| rs3740120 | 14602443 | 10 | FAM107B | Inositol 1,4,5-trisphosphate receptor-interacting protein-like 1 levels |
| rs11259186 | 14603429 | 10 | FAM107B | Platelet distribution width |
| rs876654 | 14609487 | 10 | FAM107B | Height |
| rs17758689 | 20216247 | 10 | PLXDC2 | Lung function (FEV1/FVC) |
| rs12776043 | 20221250 | 10 | PLXDC2 | NID1/SORT1 protein level ratio |
| rs7083449 | 20228664 | 10 | PLXDC2 | Cognitive domain scores for executive function |
| rs1575869 | 20239495 | 10 | PLXDC2 | Blood pressure (pleiotropy model 1 DBP adjusted for estimated causal effects x SBP) |
| rs7074936 | 24604087 | 10 | KIAA1217 | Right hippocampal volume |
| rs7087182 | 24616847 | 10 | KIAA1217 | OTU99_24 (Prevotella) prevalence |
| rs7075108 | 24684811 | 10 | KIAA1217 | Central cerebrospinal fluid amyloid beta 42 to 40 ratio (abnormal vs normal) |
| rs3762086 | 24720488 | 10 | KIAA1217 | Diverticular disease |
| rs2274831 | 24721946 | 10 | KIAA1217 | Strabismus |
| rs16924788 | 24752328 | 10 | KIAA1217 | Height |
| rs7913390 | 24906143 | 10 | ARHGAP21 | Waist-hip ratio |
| rs34485271 | 24919321 | 10 | ARHGAP21 | Facial skin hydration |
| rs7909388 | 24957906 | 10 | ARHGAP21 | Height |
| rs6482413 | 24985693 | 10 | ARHGAP21 | Whole body fat free mass (UKB data field 23101) |
| rs12357257 | 24999593 | 10 | ARHGAP21 | Advanced age-related macular degeneration |
| rs7087801 | 25005932 | 10 | ARHGAP21 | Appendicular lean mass |
| rs1334893 | 27693305 | 10 | PTCHD3 | Fasting blood insulin (BMI interaction) |
| rs2484191 | 27694764 | 10 | PTCHD3 | Height |
| rs2815561 | 27978360 | 10 | MKX | Height |
| rs7358151 | 28017572 | 10 | MKX | Diisocyanate-induced asthma |
| rs4749261 | 28113533 | 10 | ARMC4 | Height |
| rs10826354 | 28141445 | 10 | ARMC4 | OTU97_140 (Bacteroides) prevalence |
| rs11006747 | 28150935 | 10 | ARMC4 | Obesity-related traits |
| rs12779675 | 28197526 | 10 | ARMC4 | Lung function (FVC) |
| rs7893462 | 28228865 | 10 | ARMC4 | Diastolic blood pressure |
| rs3802517 | 28233469 | 10 | ARMC4 | Systolic blood pressure |
| rs16928444 | 28277669 | 10 | ARMC4 | Life satisfaction |
| rs12249305 | 28342740 | 10 | MPP7 | Taurochenodeoxycholate levels in elite athletes |
| rs898549 | 44458714 | 10 | LINC00841 | Obesity-related traits |
| rs1670140 | 67708359 | 10 | CTNNA3 | Adolescent idiopathic scoliosis |
| rs1670167 | 67719774 | 10 | CTNNA3 | Educational attainment |
| rs12359372 | 67767951 | 10 | CTNNA3 | Educational attainment (years of education) |
| rs7903919 | 67770946 | 10 | CTNNA3 | Educational attainment |
| rs10509258 | 67783437 | 10 | CTNNA3 | IgA nephropathy |
| rs1993849 | 67796008 | 10 | CTNNA3 | Depression |
| rs6480305 | 69879709 | 10 | MYPN | Psoriasis |
| rs61854624 | 69926319 | 10 | MYPN | Vertical cup-disc ratio |
| rs10998194 | 70217052 | 10 | DNA2 | Weight |
| rs10823229 | 70332580 | 10 | TET1 | Height |
| rs12773594 | 70332672 | 10 | TET1 | Diastolic blood pressure |
| rs12221107 | 70332862 | 10 | TET1 | Waist circumference adjusted for body mass index |
| rs34386000 | 70337915 | 10 | TET1 | Corneal resistance factor (MTAG) |
| rs10998304 | 70342775 | 10 | TET1 | Type 2 diabetes |
| rs2298117 | 70346740 | 10 | TET1 | Chronotype |
| rs2795870 | 70352140 | 10 | TET1 | Height |
| rs12241767 | 70405237 | 10 | TET1 | Height |
| rs10998375 | 70425119 | 10 | TET1 | Height |
| rs2030057 | 70444319 | 10 | TET1 | Hip circumference adjusted for BMI |
| rs10762242 | 70592130 | 10 | STOX1 | Cortical surface area |
| rs7076441 | 70728298 | 10 | DDX21 | Breast dense area |
| rs2255607 | 70748784 | 10 | KIAA1279 | CDKN2D/KIFBP protein level ratio |
| rs2429030 | 70751673 | 10 | KIAA1279 | Schizophrenia |
| rs7078237 | 70774039 | 10 | KIAA1279 | Vertical cup-disc ratio |
| rs444386 | 90528178 | 10 | LIPN | Lobe attachment (rater-scored or self-reported) |
| rs3979139 | 90567045 | 10 | LIPM | Lipase member N levels |
| rs1661281 | 90580330 | 10 | ANKRD22 | Mucinous adenocarcinoma in colorectal cancer |
| rs4934425 | 90609718 | 10 | ANKRD22 | Height |
| rs7912777 | 90658059 | 10 | STAMBPL1 | Brain region volumes |
| rs1926203 | 90727334 | 10 | ACTA2 | Lung cancer |
| rs11202914 | 90730211 | 10 | ACTA2 | Tumor necrosis factor receptor superfamily member 6 levels |
| rs7082720 | 90742049 | 10 | ACTA2 | Arthritis (juvenile idiopathic) |
| rs3758483 | 90748736 | 10 | ACTA2 | OTU97_56 (Ruminococcaceae) abundance |
| rs1800682 | 90749963 | 10 | ACTA2 | Chronic inflammatory diseases (ankylosing spondylitis, Crohn's disease, psoriasis, primary sclerosing cholangitis, ulcerative colitis) (pleiotropy) |
| rs12775501 | 90750909 | 10 | ACTA2 | Lymphocyte percentage of white cells |
| rs6586163 | 90752018 | 10 | FAS | Chronic lymphocytic leukemia |
| rs3824729 | 90752616 | 10 | FAS | Mean corpuscular volume |
| rs4406737 | 90759724 | 10 | FAS | Chronic lymphocytic leukemia |
| rs2147419 | 90759916 | 10 | FAS | Testosterone levels |
| rs7069750 | 90762376 | 10 | FAS | Juvenile idiopathic arthritis (oligoarticular or rheumatoid factor-negative polyarticular) |
| rs7901656 | 90766213 | 10 | FAS | Tumor necrosis factor receptor superfamily member 6 levels |
| rs982764 | 90769998 | 10 | FAS | Blood protein levels |
| rs2234978 | 90771829 | 10 | FAS | Immunoglobulin A |

| | | | | |
|---|---|---|---|---|
| rs11195062 | 112036975 | 10 | MXI1 | Mean corpuscular hemoglobin |
| rs2855469 | 112041579 | 10 | MXI1 | Cardiovascular disease |
| rs1803997 | 112046200 | 10 | MXI1 | Height |
| rs10884945 | 112054225 | 10 | SMNDC1 | Brain morphology (MOSTest) |
| rs17098586 | 121006227 | 10 | GRK5 | Coronary artery aneurysm in Kawasaki disease |
| rs7076992 | 121028387 | 10 | GRK5 | CCL17/CCL22 protein level ratio |
| rs4752269 | 121037952 | 10 | GRK5 | Height |
| rs59348252 | 121053180 | 10 | GRK5 | Neutrophil-to-lymphocyte ratio |
| rs4752321 | 121277598 | 10 | RGS10 | Menstruation quality of life impact (edema) |
| rs12219225 | 121280670 | 10 | RGS10 | Rheumatoid arthritis |
| rs151243716 | 121414663 | 10 | BAG3 | Serum manganese levels |
| rs72840788 | 121415685 | 10 | BAG3 | Left ventricular end-systolic volume |
| rs17099139 | 121419487 | 10 | BAG3 | Hypertrophic cardiomyopathy |
| rs7095308 | 121426312 | 10 | BAG3 | Systolic blood pressure |
| rs17617337 | 121426884 | 10 | BAG3 | Diastolic blood pressure |
| rs196321 | 121427109 | 10 | BAG3 | Heart failure (multivariate analysis) |
| rs2234962 | 121429633 | 10 | BAG3 | Lung function (FEV1/FVC) |
| rs72842207 | 121433675 | 10 | BAG3 | Systolic blood pressure |
| rs7100333 | 123913826 | 10 | TACC2 | Indoleacetylglutamine levels in elite athletes |
| rs7897826 | 123932642 | 10 | TACC2 | Type 2 diabetes (adjusted for BMI) |
| rs11596483 | 124037599 | 10 | BTBD16 | Gut microbiome composition (summer) |
| rs4752678 | 124039657 | 10 | BTBD16 | Heel bone mineral density |
| rs11592545 | 124050575 | 10 | BTBD16 | Vaginal microbiome MetaCyc pathway (PWY-6121\|5-aminoimidazole ribonucleotide biosynthesis I) |
| rs10788275 | 124061501 | 10 | BTBD16 | Height |
| rs61871680 | 124070455 | 10 | BTBD16 | Serine protease HTRA1 levels |
| rs6585812 | 124086757 | 10 | BTBD16 | Superior parietal cortex volume |
| rs71486610 | 124134803 | 10 | PLEKHA1 | Offspring birth weight |
| rs3850765 | 124139910 | 10 | PLEKHA1 | Hypothyroidism |
| rs7088058 | 124149352 | 10 | PLEKHA1 | Autoimmune traits |
| rs2421019 | 124150586 | 10 | PLEKHA1 | Height |
| rs11200607 | 124154626 | 10 | PLEKHA1 | Fat-free mass |
| rs12218910 | 124158082 | 10 | PLEKHA1 | Pleckstrin homology domain-containing family A member 1 levels |
| rs4752693 | 124163611 | 10 | PLEKHA1 | Systolic blood pressure |
| rs10082476 | 124164654 | 10 | PLEKHA1 | Hip circumference adjusted for BMI |
| rs6585827 | 124165615 | 10 | PLEKHA1 | Birth weight |
| rs2421016 | 124167512 | 10 | PLEKHA1 | Birth weight |
| rs7908662 | 124172912 | 10 | PLEKHA1 | Alzheimer's disease |
| rs4311997 | 124179299 | 10 | PLEKHA1 | Central corneal thickness |
| rs2421015 | 124183091 | 10 | PLEKHA1 | Pain (pleiotropy) |
| rs2292626 | 124186714 | 10 | PLEKHA1 | Femoral neck bone mineral density |
| rs1045216 | 124189197 | 10 | PLEKHA1 | Autoimmune thyroid disease |
| rs10490923 | 124214251 | 10 | ARMS2 | Diastolic blood pressure |
| rs2736911 | 124214355 | 10 | ARMS2 | Carpal tunnel syndrome |
| rs10490924 | 124214448 | 10 | ARMS2 | Neovascular age-related macular degeneration |
| rs36212732 | 124215198 | 10 | ARMS2 | Spherical equivalent |
| rs3750848 | 124215315 | 10 | ARMS2 | Age-related macular degeneration (extreme sampling) |
| rs3750846 | 124215565 | 10 | ARMS2 | Advanced age-related macular degeneration |
| rs2736912 | 124215594 | 10 | ARMS2 | Pain (pleiotropy) |
| rs2248799 | 124223944 | 10 | HTRA1 | Age-related macular degeneration (MTAG) |
| rs2672596 | 124226793 | 10 | HTRA1 | Sarcalumenin levels |
| rs60401382 | 124227624 | 10 | HTRA1 | Stroke |
| rs11200643 | 124229203 | 10 | HTRA1 | Height |
| rs12252027 | 124230513 | 10 | HTRA1 | Headache or migraine |
| rs2672592 | 124230750 | 10 | HTRA1 | Migraine |
| rs72834448 | 124232420 | 10 | HTRA1 | Systolic blood pressure |
| rs2142308 | 124234037 | 10 | HTRA1 | Lung function (FEV1/FVC) |
| rs72834453 | 124235226 | 10 | HTRA1 | Systolic blood pressure |
| rs2672587 | 124235355 | 10 | HTRA1 | Height |
| rs28445838 | 125650370 | 10 | CPXM2 | Appendicular lean mass |
| rs7084508 | 133932946 | 10 | JAKMIP3 | Educational attainment (years of education) |
| rs954820 | 133957761 | 10 | JAKMIP3 | Chronic obstructive pulmonary disease-related biomarkers |
| rs10870267 | 133968063 | 10 | JAKMIP3 | Back pain |
| rs2172131 | 133978962 | 10 | JAKMIP3 | Body mass index |
| rs2035806 | 133984916 | 10 | JAKMIP3 | Adult body size |
| rs4242715 | 133986135 | 10 | JAKMIP3 | Drinks per week |
| rs4880341 | 133992689 | 10 | JAKMIP3 | Body mass index |
| rs2814162 | 134001748 | 10 | DPYSL4 | Biological sex |
| rs1873506 | 134002617 | 10 | DPYSL4 | Brain morphology (MOSTest) |
| rs2101613 | 134002767 | 10 | DPYSL4 | Cortical thickness |
| rs902627 | 134018001 | 10 | DPYSL4 | Highest math class taken (MTAG) |
| rs1038804 | 134026447 | 10 | STK32C | General risk tolerance (MTAG) |
| rs870427 | 134039891 | 10 | STK32C | Trauma exposure |
| rs10870273 | 134046136 | 10 | STK32C | Pneumococcal meningitis x streptococcus pneumoniae genotype cluster interaction |
| rs7076997 | 134046708 | 10 | STK32C | Metabolite levels |
| rs1000535 | 134051654 | 10 | STK32C | Spontaneous preterm birth without premature rupture of membranes |
| rs12267905 | 134058824 | 10 | STK32C | Ocular sarcoidosis |
| rs11146283 | 134078458 | 10 | STK32C | Argininate levels in elite athletes |
| rs10870282 | 134079236 | 10 | STK32C | Cerebrospinal fluid sTREM-2 levels |
| rs35033992 | 134095819 | 10 | STK32C | Attention deficit hyperactivity disorder or risk-taking behavior (pleiotropy) |

| | | | | |
|---|---|---|---|---|
| rs10870286 | 134099342 | 10 | STK32C | Smoking initiation |
| rs10870287 | 134105044 | 10 | STK32C | Externalizing behaviour (multivariate analysis) |
| rs10870289 | 134109072 | 10 | STK32C | N-acetylalanine levels in elite athletes |
| rs7099822 | 134109886 | 10 | STK32C | General risk tolerance (MTAG) |
| rs10833529 | 21439476 | 11 | NELL1 | Prudent dietary pattern |
| rs10766821 | 21501695 | 11 | NELL1 | Metabolite levels |
| rs73469144 | 21504900 | 11 | NELL1 | PR interval in Tripanosoma cruzi seropositivity |
| rs7116826 | 21515583 | 11 | NELL1 | Adolescent idiopathic scoliosis |
| rs12279250 | 21565079 | 11 | NELL1 | Thiazide-induced adverse metabolic effects in hypertensive patients |
| rs4922560 | 21588442 | 11 | NELL1 | Creatinine levels in elite athletes |
| rs10833719 | 22226884 | 11 | ANO5 | Myosin-binding protein C, slow-type levels |
| rs4922982 | 22237365 | 11 | ANO5 | High-sensitivity cardiac troponin T levels |
| rs10766929 | 22299077 | 11 | ANO5 | High-sensitivity cardiac troponin I concentration |
| rs1562445 | 22367286 | 11 | SLC17A6 | Systolic blood pressure |
| rs2290046 | 22384166 | 11 | SLC17A6 | Educational attainment |
| rs11027983 | 24555148 | 11 | LUZP2 | Externalizing behaviour (multivariate analysis) |
| rs1509605 | 24577183 | 11 | LUZP2 | Depressive symptoms |
| rs10742044 | 24606921 | 11 | LUZP2 | Smoking initiation |
| rs931964 | 24607322 | 11 | LUZP2 | Smoking initiation |
| rs12361953 | 24611130 | 11 | LUZP2 | Alzheimer's disease (late onset) |
| rs10834400 | 24611220 | 11 | LUZP2 | Smoking initiation |
| rs4561213 | 24678819 | 11 | LUZP2 | Magnesium levels |
| rs7118149 | 24700545 | 11 | LUZP2 | Psychiatric symptomatology and risky substance use |
| rs1174117 | 35686478 | 11 | TRIM44 | Photic sneeze reflex |
| rs7129170 | 35754464 | 11 | TRIM44 | Eicosenoylcarnitine (C20:1) levels in elite athletes |
| rs7947746 | 36033069 | 11 | LDLRAD3 | Height |
| rs4547071 | 36052138 | 11 | LDLRAD3 | Metabolite levels |
| rs78006532 | 36061299 | 11 | LDLRAD3 | Vaginal microbiome MetaCyc pathway (PWY-6612\|superpathway of tetrahydrofolate biosynthesis) |
| rs79874564 | 36075232 | 11 | LDLRAD3 | Alopecia areata |
| rs4756265 | 36078171 | 11 | LDLRAD3 | Lymphocyte count |
| rs10768174 | 36079256 | 11 | LDLRAD3 | Lymphocyte count |
| rs9645614 | 36159947 | 11 | LDLRAD3 | Urine 6-hydroxymelatonin sulfate-to-creatinine ratio |
| rs10768195 | 36169869 | 11 | LDLRAD3 | Height |
| rs12574547 | 36173089 | 11 | LDLRAD3 | Serum levels of protein FJX1 |
| rs11033523 | 36301224 | 11 | COMMD9 | Triglyceride levels |
| rs11820100 | 36307341 | 11 | COMMD9 | High density lipoprotein cholesterol levels |
| rs11033535 | 36309463 | 11 | COMMD9 | High density lipoprotein cholesterol levels |
| rs79627555 | 36322401 | 11 | PRR5L | Triglyceride levels |
| rs61878692 | 36332689 | 11 | PRR5L | Atopic dermatitis |
| rs2218565 | 36336263 | 11 | PRR5L | Inflammatory skin disease |
| rs4290197 | 36339673 | 11 | PRR5L | Severe COVID-19 infection |
| rs7127214 | 36343693 | 11 | PRR5L | Juvenile idiopathic arthritis (oligoarticular or rheumatoid factor-negative polyarticular) |
| rs7130870 | 36344202 | 11 | PRR5L | Asthma |
| rs7108441 | 36351781 | 11 | PRR5L | Asthma |
| rs7114403 | 36361607 | 11 | PRR5L | Waist-to-hip ratio adjusted for BMI |
| rs7128640 | 36361616 | 11 | PRR5L | Waist-to-hip ratio adjusted for BMI |
| rs7101640 | 36361918 | 11 | PRR5L | Waist-to-hip ratio adjusted for BMI |
| rs12364550 | 36362125 | 11 | PRR5L | Waist-hip index |
| rs4755450 | 36363575 | 11 | PRR5L | Multiple sclerosis |
| rs10836538 | 36365253 | 11 | PRR5L | Asthma (childhood onset) |
| rs10501149 | 36376021 | 11 | PRR5L | Hay fever and/or eczema |
| rs11037728 | 43926350 | 11 | ALKBH3 | Hair color |
| rs10768994 | 43936945 | 11 | ALKBH3 | Body mass index (MTAG) |
| rs2074038 | 44087989 | 11 | ACCS | IgA nephropathy |
| rs4551754 | 44171456 | 11 | EXT2 | Male-pattern baldness |
| rs4755234 | 44232435 | 11 | EXT2 | Height |
| rs6485503 | 44233957 | 11 | EXT2 | Height |
| rs12577376 | 44239601 | 11 | EXT2 | Ebbinghaus illusion (underestimation) |
| rs117526668 | 44285431 | 11 | ALX4 | Response to efavirenz-containing treatment in HIV 1 infection (virologic failure) |
| rs10769025 | 44289435 | 11 | ALX4 | Response to antidepressants in depression |
| rs2246920 | 62104937 | 11 | ASRGL1 | Serum metabolite levels |
| rs2513749 | 62105391 | 11 | ASRGL1 | Asparagine levels |
| rs1043789 | 62160729 | 11 | ASRGL1 | Waist-hip index |
| rs3741240 | 62186542 | 11 | SCGB1A1 | Chronic obstructive pulmonary disease-related biomarkers |
| rs4963506 | 62187666 | 11 | SCGB1A1 | Degree of unsaturation (UKB data field 23443) |
| rs11231085 | 62190448 | 11 | SCGB1A1 | Secretoglobin family 3A member 1 levels |
| rs2259606 | 62201131 | 11 | AHNAK | Gamma glutamyl transferase levels |
| rs10897275 | 62203865 | 11 | AHNAK | Hair color |
| rs2509985 | 62216041 | 11 | AHNAK | Waist-to-hip ratio adjusted for BMI |
| rs77684111 | 116620129 | 11 | BUD13 | Free cholesterol to total lipids ratio in small VLDL |
| rs11216129 | 116620256 | 11 | BUD13 | Triacylglycerol(54:4)_[M+NH4]1+ levels |
| rs180327 | 116623659 | 11 | BUD13 | Triglyceride levels (MTAG) |
| rs6589564 | 116624153 | 11 | BUD13 | Bone mineral density (femoral neck) in inflammatory bowel disease |
| rs180326 | 116624703 | 11 | BUD13 | Triglyceride levels |
| rs7930786 | 116624727 | 11 | BUD13 | 1-oleoyl-2-arachidonoyl-GPE (18:1/20:4) levels |
| rs3825041 | 116631707 | 11 | BUD13 | Sphingomyelin(41:1)_[M+H]1+ levels |
| rs11602240 | 116632798 | 11 | BUD13 | Sphingomyelin(38:1)_[M+H]1+ levels |
| rs1263149 | 116639941 | 11 | BUD13 | Sphingomyelin(34:0)_[M+H]1+ levels |
| rs622604 | 116640062 | 11 | BUD13 | Triacylglycerol(54:4)_[M+NH4]1+ levels |

| | | | | |
|---|---|---|---|---|
| rs610675 | 116640094 | 11 | BUD13 | Triacylglycerol(54:4)_[M+NH4]1+ levels |
| rs6589565 | 116640237 | 11 | BUD13 | Cholesteryl esters in medium HDL (UKB data field 23569) |
| rs623908 | 116640368 | 11 | BUD13 | Triacylglycerol(54:4)_[M+NH4]1+ levels |
| rs664059 | 116642137 | 11 | BUD13 | Multi-trait sex score |
| rs75198898 | 116649806 | 11 | ZPR1 | Triglyceride levels |
| rs11604424 | 116651115 | 11 | ZPR1 | Total cholesterol levels |
| rs1942478 | 116651463 | 11 | ZPR1 | Sphingomyelin(38:1)_[M+H]1+ levels |
| rs4417316 | 116652301 | 11 | ZPR1 | Sphingomyelin(38:1)_[M+H]1+ levels |
| rs6589566 | 116652423 | 11 | ZPR1 | NHDL cholesterol to HDL cholesterol ratio |
| rs2075290 | 116653296 | 11 | ZPR1 | Total cholesterol levels |
| rs603446 | 116654435 | 11 | ZPR1 | Triglyceride levels |
| rs10750096 | 116656788 | 11 | ZPR1 | Neurexophilin-2 levels |
| rs3741297 | 116657667 | 11 | ZPR1 | High density lipoprotein cholesterol levels |
| rs33984246 | 116660450 | 11 | APOA5 | Total cholesterol levels |
| rs2266788 | 116660686 | 11 | APOA5 | Total cholesterol levels |
| rs2075291 | 116661392 | 11 | APOA5 | Plasma omega-6 polyunsaturated fatty acid levels (dihomo-gamma-linolenic acid) |
| rs3135507 | 116661488 | 11 | APOA5 | High-density lipoprotein levels (MTAG) |
| rs2072560 | 116661826 | 11 | APOA5 | Cholesteryl ester (22:0) levels |
| rs651821 | 116662579 | 11 | APOA5 | Triglyceride levels |
| rs5093 | 116693354 | 11 | APOA4 | High density lipoprotein cholesterol levels |
| rs2070669 | 116701122 | 11 | APOC3 | Triglyceride levels x long total sleep time interaction (2df test) |
| rs4520 | 116701535 | 11 | APOC3 | Phosphatidylinositol levels |
| rs5130 | 116703146 | 11 | APOC3 | Plateletcrit |
| rs4225 | 116703671 | 11 | APOC3 | Nonalcoholic fatty liver disease (imputed) |
| rs7116797 | 116707338 | 11 | APOA1 | Systolic blood pressure |
| rs12718464 | 116707401 | 11 | APOA1 | BMI (standard GWA) |
| rs5072 | 116707583 | 11 | APOA1 | Beta-endorphin levels |
| rs2070665 | 116707684 | 11 | APOA1 | Serum metabolite levels (CMS) |
| rs12718465 | 116707736 | 11 | APOA1 | High density lipoprotein cholesterol levels |
| rs614944 | 116717663 | 11 | SIK3 | Cholesteryl esters to total lipids ratio in small LDL |
| rs628418 | 116718366 | 11 | SIK3 | Cholesteryl esters to total lipids ratio in large HDL |
| rs61905145 | 116718415 | 11 | SIK3 | Subjective well-being |
| rs10047462 | 116722041 | 11 | SIK3 | Iron status biomarkers |
| rs688456 | 116722551 | 11 | SIK3 | Total cholesterol levels (UKB data field 23400) |
| rs12099358 | 116726048 | 11 | SIK3 | Beta-endorphin levels |
| rs625145 | 116727936 | 11 | SIK3 | Apolipoprotein A1 levels (UKB data field 23440) |
| rs12225230 | 116728630 | 11 | SIK3 | Triglycerides |
| rs541586 | 116729196 | 11 | SIK3 | Total cholesterol levels (MTAG) |
| rs6589574 | 116730638 | 11 | SIK3 | Triacylglycerol (54:3) levels |
| rs2075292 | 116732512 | 11 | SIK3 | Triglycerides |
| rs11216168 | 116741553 | 11 | SIK3 | Subjective well-being |
| rs9919599 | 116745484 | 11 | SIK3 | Serum levels of protein TFRC |
| rs543819 | 116750073 | 11 | SIK3 | Diverticular disease |
| rs482371 | 116752163 | 11 | SIK3 | Multi-trait sum score |
| rs148403721 | 116757765 | 11 | SIK3 | Low-density lipoprotein levels (MTAG) |
| rs17174502 | 116759655 | 11 | SIK3 | Phosphatidylethanolamine (18:1_18:1) levels |
| rs525302 | 116762583 | 11 | SIK3 | Linoleic acid levels |
| rs75158858 | 116762829 | 11 | SIK3 | Low density lipoprotein cholesterol levels |
| rs672058 | 116764021 | 11 | SIK3 | Blood protein levels |
| rs595137 | 116771356 | 11 | SIK3 | Triglyceride levels (MTAG) |
| rs1076485 | 116772441 | 11 | SIK3 | Systolic blood pressure |
| rs141023293 | 116772917 | 11 | SIK3 | Allergic rhinitis |
| rs11216185 | 116782974 | 11 | SIK3 | Infantile hypertrophic pyloric stenosis |
| rs7115583 | 116784376 | 11 | SIK3 | HDL cholesterol |
| rs78692246 | 116791307 | 11 | SIK3 | Eosinophil counts |
| rs645258 | 116801201 | 11 | SIK3 | Triglyceride levels |
| rs510988 | 116813804 | 11 | SIK3 | Phosphatidylethanolamine (16:0_18:2) levels |
| rs4245168 | 116882638 | 11 | SIK3 | Triacylglycerol 56:6 levels |
| rs61905706 | 116883777 | 11 | SIK3 | Triglyceride levels x long total sleep time interaction (2df test) |
| rs7115242 | 116908283 | 11 | SIK3 | Age-related diseases, mortality and associated endophenotypes |
| rs2000614 | 116915742 | 11 | SIK3 | Blood protein levels |
| rs10750100 | 116917244 | 11 | SIK3 | Cholesterol_[M+H-H2O]1+ levels |
| rs73576603 | 116917885 | 11 | SIK3 | Apolipoprotein A1 levels |
| rs4938330 | 116928514 | 11 | SIK3 | Proprotein convertase subtilisin/kexin type 7 levels |
| rs12280210 | 116940449 | 11 | SIK3 | Lobe attachment (rater-scored or self-reported) |
| rs17120280 | 116942132 | 11 | SIK3 | Multi-trait sum score |
| rs59097294 | 116964437 | 11 | SIK3 | Cholesterol levels in medium HDL |
| rs12419272 | 117015516 | 11 | PAFAH1B2 | Medication use (thyroid preparations) |
| rs7940310 | 117024481 | 11 | PAFAH1B2 | Blood protein levels |
| rs1871757 | 117030581 | 11 | PAFAH1B2 | Transgelin levels |
| rs12281009 | 117032959 | 11 | PAFAH1B2 | Body mass index |
| rs7112513 | 117037361 | 11 | PAFAH1B2 | Protein quantitative trait loci |
| rs4938353 | 117046197 | 11 | PAFAH1B2 | Blood protein levels |
| rs4938354 | 117051165 | 11 | SIDT2 | Proprotein convertase subtilisin/kexin type 7 levels |
| rs1242229 | 117062370 | 11 | SIDT2 | High density lipoprotein cholesterol levels |
| rs652455 | 117071449 | 11 | LOC100652768 | Transferrin receptor protein 1 levels |
| rs494356 | 117071970 | 11 | LOC100652768 | Medication use (calcium channel blockers) |
| rs12970 | 117074109 | 11 | TAGLN | Apolipoprotein B levels |
| rs641620 | 117074229 | 11 | TAGLN | Systolic blood pressure |

| | | | | |
|---|---|---|---|---|
| rs11216315 | 117080640 | 11 | PCSK7 | HDL cholesterol |
| rs171052 | 117081763 | 11 | PCSK7 | Proprotein convertase subtilisin/kexin type 7 level in Chronic kidney disease with hypertension and no diabetes (4459_68) |
| rs12418744 | 117084537 | 11 | PCSK7 | Height |
| rs77683187 | 117085210 | 11 | PCSK7 | Breast cancer |
| rs236911 | 117085270 | 11 | PCSK7 | Triglycerides |
| rs236918 | 117091609 | 11 | PCSK7 | Iron status biomarkers |
| rs45462696 | 117092660 | 11 | PCSK7 | Proprotein convertase subtilisin/kexin type 7 levels |
| rs3830008 | 117093764 | 11 | PCSK7 | Serum levels of protein PCSK7 |
| rs59781045 | 117095283 | 11 | PCSK7 | HDL cholesterol levels x short total sleep time interaction (2df test) |
| rs7926958 | 117097222 | 11 | PCSK7 | Height |
| rs2306473 | 117097952 | 11 | PCSK7 | Triglyceride levels (MTAG) |
| rs4938362 | 117100029 | 11 | PCSK7 | Triglyceride levels |
| rs522645 | 117101574 | 11 | PCSK7 | Low-density lipoprotein levels (MTAG) |
| rs2252145 | 117102648 | 11 | PCSK7 | Proprotein convertase subtilisin/kexin type 7 levels |
| rs77535829 | 118311677 | 11 | KMT2A | Gamma glutamyl transpeptidase |
| rs74422681 | 118312870 | 11 | KMT2A | Red blood cell count |
| rs2017122 | 118314849 | 11 | KMT2A | Depression |
| rs7948661 | 118341649 | 11 | KMT2A | Autism spectrum disorder, attention deficit-hyperactivity disorder, bipolar disorder, major depressive disorder, and schizophrenia (combined) |
| rs9332781 | 118346599 | 11 | KMT2A | Neuroticism |
| rs17122278 | 118449370 | 11 | ARCN1 | Total cholesterol levels |
| rs11216909 | 118449746 | 11 | ARCN1 | Non-HDL cholesterol levels |
| rs7115634 | 118466249 | 11 | ARCN1 | Total cholesterol levels |
| rs11216925 | 118471423 | 11 | ARCN1 | Total cholesterol levels |
| rs498872 | 118477367 | 11 | PHLDB1 | Glioma |
| rs7125115 | 118478330 | 11 | PHLDB1 | Adult diffuse glioma (IDH mutation) |
| rs11603023 | 118486067 | 11 | PHLDB1 | Cholesterol, total |
| rs45540840 | 118486110 | 11 | PHLDB1 | Hip minimal joint space width |
| rs11216930 | 118488782 | 11 | PHLDB1 | Serum alkaline phosphatase levels |
| rs10892247 | 118490076 | 11 | PHLDB1 | Blood urea nitrogen levels |
| rs11216934 | 118501459 | 11 | PHLDB1 | Vertex-wise sulcal depth |
| rs483598 | 118509668 | 11 | PHLDB1 | Systemic lupus erythematosus (MTAG) |
| rs613243 | 118514022 | 11 | PHLDB1 | Cortical thickness |
| rs11216938 | 118514625 | 11 | PHLDB1 | Liver enzyme levels (alkaline phosphatase) |
| rs10790256 | 118534082 | 11 | TREH | Serum alkaline phosphatase levels |
| rs12225548 | 118535840 | 11 | TREH | Serum alkaline phosphatase levels |
| rs523604 | 118755738 | 11 | CXCR5 | Multiple sclerosis |
| rs497916 | 118758089 | 11 | CXCR5 | Asthma (childhood onset) |
| rs3922 | 118765600 | 11 | CXCR5 | Height |
| rs75438046 | 118767564 | 11 | BCL9L | Human herpes virus 7 U14 antibody levels |
| rs45510602 | 118917587 | 11 | HYOU1 | CA1/HMBS protein level ratio |
| rs511134 | 118925935 | 11 | HYOU1 | Height |
| rs1185460 | 118943433 | 11 | VPS11 | Coronary artery disease |
| rs3825061 | 118944675 | 11 | VPS11 | Body mass index |
| rs1177562 | 118949331 | 11 | VPS11 | Cholesteryl ester levels in large HDL |
| rs1307145 | 118950217 | 11 | VPS11 | Coronary artery disease |
| rs15818 | 118952173 | 11 | VPS11 | Body mass index |
| rs549893 | 118959732 | 11 | HMBS | HYOU1/THBD protein level ratio |
| rs8551 | 118967524 | 11 | DPAGT1 | Lymphocyte percentage of white cells |
| rs643788 | 118967758 | 11 | DPAGT1 | Mean corpuscular hemoglobin concentration |
| rs519672 | 119984535 | 11 | TRIM29 | Height |
| rs606970 | 119989892 | 11 | TRIM29 | Resistance to COVID-19 infection (Exposed negative vs positive) |
| rs73046547 | 10919313 | 12 | LOC101928162 | Years ovulating |
| rs1913256 | 16343868 | 12 | SLC15A5 | Severe COVID-19 infection |
| rs17401474 | 24138390 | 12 | SOX5 | Ischemic stroke in diabetes mellitus |
| rs1948662 | 24144590 | 12 | SOX5 | Height |
| rs4400891 | 24156664 | 12 | SOX5 | Total testosterone levels |
| rs11047198 | 24156829 | 12 | SOX5 | Hematocrit |
| rs989657 | 24166426 | 12 | SOX5 | Height |
| rs11047209 | 24168711 | 12 | SOX5 | Externalizing behaviour (multivariate analysis) |
| rs16926971 | 24169797 | 12 | SOX5 | Depression |
| rs1353792 | 24180307 | 12 | SOX5 | C-reactive protein levels |
| rs10842262 | 24184544 | 12 | SOX5 | Non-obstructive azoospermia |
| rs16927047 | 24192371 | 12 | SOX5 | Height |
| rs1995181 | 24195048 | 12 | SOX5 | Educational attainment (years of education) |
| rs1391790 | 24203096 | 12 | SOX5 | Sex hormone-binding globulin levels adjusted for BMI |
| rs17485141 | 24213634 | 12 | SOX5 | Suicide attempt |
| rs12322111 | 24217109 | 12 | SOX5 | Brain morphology (MOSTest) |
| rs7296101 | 24234001 | 12 | SOX5 | Vertex-wise sulcal depth |
| rs10842272 | 24235715 | 12 | SOX5 | Cortical surface area |
| rs4963744 | 24240179 | 12 | SOX5 | Smoking initiation |
| rs7979575 | 24278821 | 12 | SOX5 | Response to statin therapy |
| rs1909355 | 24288153 | 12 | SOX5 | Height |
| rs7973572 | 24289366 | 12 | SOX5 | Inflammatory bowel disease |
| rs12367235 | 24290997 | 12 | SOX5 | C-reactive protein levels (MTAG) |
| rs10743498 | 24559370 | 12 | SOX5 | Serum total protein levels |
| rs1396206 | 24576859 | 12 | SOX5 | T wave morphology restitution during recovery from exercise |
| rs4963759 | 24586390 | 12 | SOX5 | ECG latent space |
| rs11047436 | 24587491 | 12 | SOX5 | Insomnia |
| rs11047441 | 24593624 | 12 | SOX5 | Height |

| | | | | |
|---|---|---|---|---|
| rs7307613 | 24595192 | 12 | SOX5 | T wave morphology restitution during exercise (MTAG) |
| rs11047449 | 24601818 | 12 | SOX5 | Spatial QRS-T angle |
| rs3112134 | 24697790 | 12 | SOX5 | Smoking initiation |
| rs6487394 | 24701866 | 12 | SOX5 | Vertex-wise sulcal depth |
| rs10842369 | 24702577 | 12 | SOX5 | Smoking initiation |
| rs11047492 | 24702836 | 12 | SOX5 | Smoking initiation |
| rs16927698 | 24704563 | 12 | SOX5 | Height |
| rs10743502 | 24705338 | 12 | SOX5 | Cortical surface area |
| rs11833062 | 24706338 | 12 | SOX5 | Smoking initiation |
| rs11047498 | 24710424 | 12 | SOX5 | Adventurousness |
| rs4542485 | 40238338 | 12 | SLC2A13 | Alzheimer's disease polygenic risk score (upper quantile vs lower quantile) |
| rs75036080 | 40276303 | 12 | SLC2A13 | Alzheimer's disease in hypertension |
| rs1005956 | 40341085 | 12 | SLC2A13 | Sleep quality |
| rs10877840 | 40352996 | 12 | SLC2A13 | Parkinson's disease |
| rs574918 | 40357647 | 12 | SLC2A13 | Educational attainment |
| rs12810368 | 40367171 | 12 | SLC2A13 | Alzheimer's disease polygenic risk score (upper quantile vs lower quantile) |
| rs28370625 | 40370151 | 12 | SLC2A13 | Beer or cider consumption (glasses per month) (UKB data field 1578, 4420) |
| rs7138679 | 40378647 | 12 | SLC2A13 | Disability (impaired activities of daily living) |
| rs11174608 | 40380583 | 12 | SLC2A13 | Pulse pressure |
| rs28370664 | 40413305 | 12 | SLC2A13 | Parkinson's disease |
| rs2242367 | 40413698 | 12 | SLC2A13 | Survival in progressive supranuclear palsy |
| rs1994090 | 40428561 | 12 | SLC2A13 | Parkinson's disease |
| rs1384236 | 40458384 | 12 | SLC2A13 | Parkinson's disease |
| rs10880557 | 44131697 | 12 | PUS7L | Vaginal microbiome MetaCyc pathway (PRPP-PWY\|superpathway of histidine, purine, and pyrimidine biosynthesis) |
| rs12310585 | 44133972 | 12 | PUS7L | Vaginal microbiome MetaCyc pathway (PRPP-PWY\|superpathway of histidine, purine, and pyrimidine biosynthesis) |
| rs3794262 | 44165430 | 12 | IRAK4 | Vaginal microbiome MetaCyc pathway (PRPP-PWY\|superpathway of histidine, purine, and pyrimidine biosynthesis) |
| rs73087684 | 44416728 | 12 | TMEM117 | X-12013 levels |
| rs4587804 | 47217514 | 12 | SLC38A4 | Ornithine levels |
| rs7967842 | 47218144 | 12 | SLC38A4 | Alanine levels |
| rs832741 | 47589998 | 12 | PCED1B | Height |
| rs12367393 | 47617776 | 12 | PCED1B | Vaginal microbiome MetaCyc pathway (PWY-7187\|pyrimidine deoxyribonucleotides de novo biosynthesis II) |
| rs607860 | 52913029 | 12 | KRT5 | Squamous cell carcinoma (MTAG) |
| rs3803084 | 52942627 | 12 | KRT71 | Fasting insulin levels in prediabetes |
| rs11171681 | 56193233 | 12 | SARNP | Longevity |
| rs7970261 | 94665413 | 12 | PLXNC1 | Dementia with Lewy bodies |
| rs4761470 | 94683204 | 12 | PLXNC1 | Estradiol plasma levels (breast cancer) |
| rs3803070 | 94688688 | 12 | PLXNC1 | Adolescent idiopathic scoliosis |
| rs2271979 | 94772742 | 12 | CEP83 | Pulse pressure |
| rs6538496 | 94775926 | 12 | CEP83 | Height |
| rs10859698 | 94852628 | 12 | CEP83 | Lung function (FVC) |
| rs2596741 | 95903543 | 12 | METAP2 | Height |
| rs3812814 | 95927037 | 12 | USP44 | C-reactive protein levels |
| rs3812813 | 95927762 | 12 | USP44 | Height |
| rs11108094 | 95928113 | 12 | USP44 | Type 2 diabetes |
| rs12579528 | 97859730 | 12 | RMST | Height |
| rs10860215 | 97863578 | 12 | RMST | Educational attainment |
| rs75718479 | 97876906 | 12 | RMST | Prostate cancer |
| rs11109072 | 97901270 | 12 | RMST | Obesity (early onset extreme) |
| rs1420341 | 97913785 | 12 | RMST | Body mass index (MTAG) |
| rs7963257 | 97916472 | 12 | RMST | Kawasaki disease |
| rs7962117 | 97919052 | 12 | RMST | Drinks per week |
| rs17027126 | 97921282 | 12 | RMST | COVID-19 (hospitalized covid vs population) |
| rs61943702 | 97924834 | 12 | RMST | Weight |
| rs59066241 | 97925364 | 12 | RMST | BMI (standard GWA) |
| rs11109986 | 100063310 | 12 | ANKS1B | Height |
| rs10778077 | 101193988 | 12 | ANO4 | Stress sensitivity (neuroticism score x major depressive disorder status interaction) |
| rs4764996 | 101194600 | 12 | ANO4 | Univariate microbial feature (asv: asv013) at the dorsal forearm (dry skin) |
| rs3897727 | 101218303 | 12 | ANO4 | Type 2 diabetes |
| rs12579637 | 101273183 | 12 | ANO4 | Externalizing behaviour (multivariate analysis) |
| rs17030834 | 101375438 | 12 | ANO4 | NK4A144 abundance in stool |
| rs609728 | 101375646 | 12 | ANO4 | Attention deficit hyperactivity disorder (hyperactivity-impulsivity symptoms) |
| rs66861122 | 101383958 | 12 | ANO4 | Response to methotrexate in juvenile idiopathic arthritis |
| rs589202 | 101445260 | 12 | ANO4 | S-adenosylhomocysteine (SAH) levels in elite athletes |
| rs7956620 | 101447641 | 12 | ANO4 | Nausea incidence in varenicline-treated smokers |
| rs729062 | 113687859 | 12 | TPCN1 | Body mass index |
| rs2303621 | 113705529 | 12 | TPCN1 | Height |
| rs12831817 | 113863963 | 12 | SDSL | Serine dehydratase-like levels |
| rs4767120 | 114189961 | 12 | LINC01234 | Self-rated health |
| rs2059598 | 114261237 | 12 | RBM19 | Waist circumference |
| rs879901 | 114284143 | 12 | RBM19 | Externalizing behaviour (multivariate analysis) |
| rs7965253 | 114297642 | 12 | RBM19 | General risk tolerance (MTAG) |
| rs77531286 | 114299548 | 12 | RBM19 | Morningness |
| rs3955311 | 114343818 | 12 | RBM19 | Chronotype |
| rs6489927 | 114349337 | 12 | RBM19 | Height |
| rs3782455 | 114359265 | 12 | RBM19 | Amyotrophic lateral sclerosis (sporadic) |
| rs883079 | 114793240 | 12 | TBX5 | Atrial fibrillation |
| rs3825214 | 114795443 | 12 | TBX5 | QRS duration |
| rs7312625 | 114799974 | 12 | TBX5 | P wave duration |
| rs7135659 | 114801772 | 12 | TBX5 | PR interval |

| | | | | |
|---|---|---|---|---|
| rs1895585 | 114802138 | 12 | TBX5 | PR interval |
| rs3782464 | 114804580 | 12 | TBX5 | 3-hydroxypropylmercapturic acid levels in smokers |
| rs1895582 | 114807035 | 12 | TBX5 | P wave duration |
| rs2236018 | 114832360 | 12 | TBX5 | Receptive language ability |
| rs1061657 | 115108136 | 12 | TBX3 | Invasive breast cancer |
| rs1061651 | 115108361 | 12 | TBX3 | Height |
| rs8853 | 115108907 | 12 | TBX3 | Benign prostatic hyperplasia and lower urinary tract symptoms (prostate cancer excluded) |
| rs59336 | 115116352 | 12 | TBX3 | Colorectal cancer |
| rs2242442 | 115121189 | 12 | TBX3 | Osteolysis in total hip arthroplasty (time to prosthesis revision) |
| rs79133148 | 124723221 | 12 | ZNF664-FAM101A | Hip circumference adjusted for BMI |
| rs79022499 | 124727611 | 12 | ZNF664-FAM101A | COVID-19 |
| rs12372180 | 124746863 | 12 | ZNF664-FAM101A | Waist-to-hip ratio adjusted for BMI |
| rs880333 | 124748860 | 12 | ZNF664-FAM101A | Waist circumference adjusted for body mass index |
| rs10773076 | 124769687 | 12 | ZNF664-FAM101A | Height |
| rs2342468 | 124772515 | 12 | ZNF664-FAM101A | Height |
| rs7306812 | 124776497 | 12 | FAM101A | Waist circumference adjusted for body mass index |
| rs73223550 | 124778323 | 12 | FAM101A | Multi-trait sex score |
| rs114786346 | 124780201 | 12 | FAM101A | Osteoarthrosis (time to event) |
| rs74701928 | 124792225 | 12 | FAM101A | Bone mineral density variability |
| rs34533686 | 124793007 | 12 | FAM101A | Whole body fat free mass (UKB data field 23101) |
| rs117285912 | 124795283 | 12 | FAM101A | Waist-to-hip ratio adjusted for BMI |
| rs7963737 | 124796007 | 12 | FAM101A | Hip circumference adjusted for BMI |
| rs11610586 | 124798413 | 12 | FAM101A | Height |
| rs12318072 | 124798801 | 12 | FAM101A | Hip circumference adjusted for BMI |
| rs4765540 | 124799642 | 12 | FAM101A | Osteoarthritis (hip) |
| rs838886 | 125264287 | 12 | SCARB1 | HDL cholesterol |
| rs3741521 | 125264293 | 12 | SCARB1 | HDL cholesterol levels x short total sleep time interaction (2df test) |
| rs921919 | 125265201 | 12 | SCARB1 | HDL cholesterol levels |
| rs2293440 | 125270773 | 12 | SCARB1 | Metabolite peak levels (QI15953) |
| rs5888 | 125284748 | 12 | SCARB1 | HDL cholesterol levels x long total sleep time interaction (2df test) |
| rs838912 | 125286037 | 12 | SCARB1 | Triglyceride levels in very large HDL |
| rs838909 | 125287648 | 12 | SCARB1 | HDL cholesterol levels x short total sleep time interaction (2df test) |
| rs865716 | 125290856 | 12 | SCARB1 | Linoleic acid levels |
| rs838900 | 125293597 | 12 | SCARB1 | Gut microbiota relative abundance (Ruminococcus belonging to family Lachnospiraceae) |
| rs11057830 | 125307053 | 12 | SCARB1 | Lipoprotein phospholipase A2 activity in cardiovascular disease |
| rs10846744 | 125312425 | 12 | SCARB1 | Mean corpuscular volume |
| rs11057839 | 125316031 | 12 | SCARB1 | Mean corpuscular hemoglobin |
| rs11057840 | 125316055 | 12 | SCARB1 | Coronary artery disease |
| rs11057841 | 125316743 | 12 | SCARB1 | Lipoprotein-associated phospholipase A2 activity and mass |
| rs12580803 | 125317247 | 12 | SCARB1 | High density lipoprotein cholesterol levels |
| rs10846749 | 125318448 | 12 | SCARB1 | Kidney cancer |
| rs4765623 | 125320850 | 12 | SCARB1 | Renal cell carcinoma |
| rs12582221 | 125323240 | 12 | SCARB1 | Hip index |
| rs3924313 | 125328375 | 12 | SCARB1 | Concentration of large HDL particles |
| rs11057853 | 125329313 | 12 | SCARB1 | Hip index |
| rs7488791 | 125334068 | 12 | SCARB1 | C-reactive protein levels |
| rs7954697 | 125335253 | 12 | SCARB1 | Waist-to-hip ratio adjusted for BMI |
| rs10846758 | 125335373 | 12 | SCARB1 | Kidney cancer |
| rs11057864 | 125335950 | 12 | SCARB1 | Response to cyclophosphamide in systemic lupus erythematosus with lupus nephritis |
| rs7296737 | 125336956 | 12 | SCARB1 | Coronary artery disease |
| rs78194510 | 125338336 | 12 | SCARB1 | Alzheimer's disease or HDL levels (pleiotropy) |
| rs10773112 | 125338529 | 12 | SCARB1 | HDL cholesterol levels x long total sleep time interaction (2df test) |
| rs28435470 | 133067473 | 12 | FBRSL1 | Serum 25-Hydroxyvitamin D levels |
| rs7295014 | 133067989 | 12 | FBRSL1 | Prostate cancer |
| rs1882297 | 133069134 | 12 | FBRSL1 | Fasting glucose |
| rs12811407 | 133069698 | 12 | FBRSL1 | Type 2 diabetes |
| rs12423664 | 133069894 | 12 | FBRSL1 | Coronary artery disease and triglyceride levels (multivariate analysis) |
| rs61952071 | 133076439 | 12 | FBRSL1 | Mean corpuscular hemoglobin |
| rs36098511 | 133080449 | 12 | FBRSL1 | Glucose levels |
| rs11610659 | 133081895 | 12 | FBRSL1 | Mean corpuscular volume |
| rs117206641 | 133086888 | 12 | FBRSL1 | Calcific aortic valve stenosis |
| rs74971075 | 133099570 | 12 | FBRSL1 | Systolic blood pressure |
| rs4883568 | 133103263 | 12 | FBRSL1 | Height |
| rs10870464 | 133117489 | 12 | FBRSL1 | Parental longevity (mother's age at death or mother's attained age) |
| rs4883556 | 133121897 | 12 | FBRSL1 | Self-reported math ability |
| rs79009737 | 133124753 | 12 | FBRSL1 | Apolipoprotein A1 levels |
| rs7301141 | 133138503 | 12 | FBRSL1 | Basal cell carcinoma (MTAG) |
| rs12809318 | 133140840 | 12 | FBRSL1 | COVID-19 severity |
| rs12826964 | 133140853 | 12 | FBRSL1 | Low density lipoprotein cholesterol levels |
| rs5023077 | 133141973 | 12 | FBRSL1 | COVID-19 (hospitalized covid vs population) |
| rs6560886 | 133150210 | 12 | FBRSL1 | Atrial fibrillation |
| rs11146953 | 133155907 | 12 | FBRSL1 | Cancer |
| rs3814787 | 24152370 | 13 | TNFRSF19 | Blood protein levels |
| rs7987593 | 24165195 | 13 | TNFRSF19 | Heel bone mineral density |
| rs11616958 | 24173049 | 13 | TNFRSF19 | TGFBR2/TNFRSF19 protein level ratio |
| rs1928117 | 24175452 | 13 | TNFRSF19 | Height |
| rs4411363 | 24191231 | 13 | TNFRSF19 | Overall survival in advanced colorectal cancer |
| rs9510787 | 24205195 | 13 | TNFRSF19 | Nasopharyngeal carcinoma |
| rs9510795 | 24216012 | 13 | TNFRSF19 | Diabetic kidney disease |

| | | | | |
|---|---|---|---|---|
| rs1575667 | 24239825 | 13 | TNFRSF19 | Heel bone mineral density |
| rs7329483 | 24242801 | 13 | TNFRSF19 | Heel bone mineral density |
| rs61948069 | 25255503 | 13 | ATP12A | Estimated glomerular filtration rate (creatinine) |
| rs55641580 | 25257917 | 13 | ATP12A | Systolic blood pressure |
| rs7338804 | 38660394 | 13 | LINC00571 | Educational attainment |
| rs7329174 | 41558110 | 13 | ELF1 | Systemic lupus erythematosus |
| rs73176930 | 41579809 | 13 | ELF1 | Eosinophil percentage of white cells |
| rs7335629 | 41579868 | 13 | ELF1 | Height |
| rs7323613 | 41581666 | 13 | ELF1 | Mean corpuscular hemoglobin |
| rs9566656 | 41581930 | 13 | ELF1 | Mean corpuscular hemoglobin |
| rs9532718 | 41644722 | 13 | WBP4 | Corneal resistance factor (MTAG) |
| rs1411041 | 41724776 | 13 | LOC101929140 | Telomere length |
| rs2151236 | 41769660 | 13 | LOC101929140 | Height |
| rs553817 | 41773356 | 13 | LOC101929140 | Corneal resistance factor (MTAG) |
| rs10467472 | 42033698 | 13 | RGCC | Pre-treatment viral load in HIV-1 infection |
| rs3783196 | 42039288 | 13 | RGCC | Multiple sclerosis |
| rs9555361 | 108087499 | 13 | FAM155A | Vaginal microbiome MetaCyc pathway (PWY-5918\|superpathay of heme biosynthesis from glutamate) |
| rs75244278 | 108123109 | 13 | FAM155A | Central cerebrospinal fluid amyloid beta 42 to 40 ratio |
| rs9520462 | 108134909 | 13 | FAM155A | Reading and spelling |
| rs75943599 | 108145708 | 13 | FAM155A | Vaginal microbiome MetaCyc pathway (GOLPDLCAT-PWY\|superpathway of glycerol degradation to 1,3-propanediol) |
| rs9555371 | 108215404 | 13 | FAM155A | Diverticular disease |
| rs16970678 | 108216200 | 13 | FAM155A | Diverticular disease |
| rs11619828 | 108218865 | 13 | FAM155A | Estimated glomerular filtration rate (creatinine) |
| rs11619840 | 108218958 | 13 | FAM155A | Diverticular disease |
| rs957788 | 108257220 | 13 | FAM155A | Anorexia nervosa |
| rs9520569 | 108402551 | 13 | FAM155A | Borderline personality disorder |
| rs10508196 | 108479966 | 13 | FAM155A | Vitamin D levels |
| rs2036707 | 108484454 | 13 | FAM155A | Obesity-related traits |
| rs1509091 | 108491592 | 13 | FAM155A | Metabolite levels (Pyroglutamine) |
| rs13260 | 110802123 | 13 | COL4A1 | Pulse pressure |
| rs650724 | 110804809 | 13 | COL4A1 | Non-lobar intracerebral hemorrhage (MTAG) |
| rs1192201 | 110806490 | 13 | COL4A1 | Age-related nuclear cataracts |
| rs11617955 | 110818102 | 13 | COL4A1 | Coronary artery disease (myocardial infarction, percutaneous transluminal coronary angioplasty, coronary artery bypass grafting, angina or chromic ischemic heart disease) |
| rs3742207 | 110818598 | 13 | COL4A1 | Arterial stiffness |
| rs58814158 | 110820255 | 13 | COL4A1 | Monocyte count |
| rs75372280 | 110821524 | 13 | COL4A1 | Hip circumference adjusted for BMI |
| rs9588107 | 110823340 | 13 | COL4A1 | Coronary artery calcification |
| rs1975514 | 110828891 | 13 | COL4A1 | Peripheral artery disease |
| rs9521632 | 110832918 | 13 | COL4A1 | Coronary artery disease |
| rs9521634 | 110833899 | 13 | COL4A1 | Stroke |
| rs3783113 | 110834746 | 13 | COL4A1 | Coronary artery disease |
| rs12866570 | 110834966 | 13 | COL4A1 | Coronary artery disease |
| rs7329411 | 110835195 | 13 | COL4A1 | Height |
| rs11622988 | 32897337 | 14 | AKAP6 | Femur bone mineral density x serum urate levels interaction |
| rs10138310 | 32912092 | 14 | AKAP6 | Atrial fibrillation |
| rs34976846 | 32916980 | 14 | AKAP6 | Lung function (FEV1/FVC) |
| rs3784192 | 32923336 | 14 | AKAP6 | PR interval |
| rs3825751 | 32923677 | 14 | AKAP6 | PR interval |
| rs1957021 | 32924505 | 14 | AKAP6 | Atrial fibrillation |
| rs1957032 | 32951556 | 14 | AKAP6 | Colorectal cancer |
| rs2985684 | 50101682 | 14 | DNAAF2 | Carotid intima media thickness |
| rs1980615 | 67958372 | 14 | TMEM229B | Phosphatidylcholine (O-18:2_20:4) levels |
| rs7160601 | 67964098 | 14 | TMEM229B | Alkenylphosphatidylcholine (P-16:0/16:0) levels |
| rs12898179 | 67964171 | 14 | TMEM229B | Glomerular filtration rate in chronic kidney disease |
| rs12897380 | 67964355 | 14 | TMEM229B | Sphingomyelin(44:9)_[M+H]1- levels |
| rs12878807 | 67964688 | 14 | TMEM229B | Basophil percentage of white cells |
| rs10873201 | 67966599 | 14 | TMEM229B | Alkenylphosphatidylcholine (P-17:0/20:4) (b) levels |
| rs10873202 | 67966622 | 14 | TMEM229B | Sphingomyelin(44:9)_[M+H]1- levels |
| rs11622608 | 67969550 | 14 | TMEM229B | Sphingomyelin(44:9)_[M+H]1- levels |
| rs11626910 | 67969761 | 14 | TMEM229B | Sphingomyelin(44:9)_[M+H]1- levels |
| rs3818242 | 67970188 | 14 | TMEM229B | Sphingomyelin(44:9)_[M+H]1- levels |
| rs61988190 | 67971136 | 14 | TMEM229B | Sphingomyelin(44:9)_[M+H]1- levels |
| rs28373080 | 67972763 | 14 | TMEM229B | Sphingomyelin(44:9)_[M+H]1- levels |
| rs17093620 | 67972922 | 14 | TMEM229B | Sphingomyelin(44:9)_[M+H]1- levels |
| rs10873203 | 67974505 | 14 | TMEM229B | Sphingomyelin(44:9)_[M+H]1- levels |
| rs10133655 | 67974928 | 14 | TMEM229B | Sphingomyelin(44:9)_[M+H]1- levels |
| rs2874093 | 67978642 | 14 | TMEM229B | Phosphatidylcholine-O_32:1_[M+H]1+/Phosphatidylcholine-P_32:0_[M+H]1+/Phosphatidylethanolamine-O_35:1_[M+H]1+ levels |
| rs7155830 | 67979557 | 14 | TMEM229B | Sphingomyelin(44:9)_[M+H]1- levels |
| rs7156144 | 67979713 | 14 | TMEM229B | Sphingomyelin(44:9)_[M+H]1- levels |
| rs7155911 | 67979845 | 14 | TMEM229B | Sphingomyelin(44:9)_[M+H]1- levels |
| rs76149837 | 67979974 | 14 | TMEM229B | Vaginal microbiome MetaCyc pathway (PWY-7159\|chlorophyllide a biosynthesis III (aerobic, light independent)) |
| rs12878949 | 67979977 | 14 | TMEM229B | Sphingomyelin(44:9)_[M+H]1- levels |
| rs12878981 | 67980031 | 14 | TMEM229B | Sphingomyelin(44:9)_[M+H]1- levels |
| rs12879556 | 67980074 | 14 | TMEM229B | Sphingomyelin(44:9)_[M+H]1- levels |
| rs10133618 | 67981403 | 14 | TMEM229B | Sphingomyelin(44:9)_[M+H]1- levels |
| rs2273596 | 67981870 | 14 | TMEM229B | Sphingomyelin(44:9)_[M+H]1- levels |
| rs731681 | 68010224 | 14 | PLEKHH1 | Diastolic blood pressure |
| rs12891886 | 68021117 | 14 | PLEKHH1 | Serum uric acid levels |
| rs2236231 | 68039027 | 14 | PLEKHH1 | Plasma kynurenine levels in major depressive disorder |

| | | | | |
|---|---|---|---|---|
| rs10151023 | 68064656 | 14 | PIGH | Adolescent idiopathic scoliosis |
| rs2319844 | 68065034 | 14 | PIGH | Adolescent idiopathic scoliosis |
| rs10131404 | 68065844 | 14 | PIGH | Body mass index |
| rs7149859 | 68066489 | 14 | PIGH | Breast cancer (survival) |
| rs10483801 | 68117006 | 14 | ARG2 | Serum creatinine levels |
| rs7149221 | 68189736 | 14 | RDH12 | Blood protein levels |
| rs12891047 | 68243075 | 14 | ZFYVE26 | Amyotrophic lateral sclerosis (sporadic) |
| rs11160832 | 105482352 | 14 | CDCA4 | Serum levels of protein RNF165 |
| rs7147439 | 105523663 | 14 | GPR132 | Atopic dermatitis |
| rs10149229 | 105609335 | 14 | JAG2 | Brain morphology (MOSTest) |
| rs2272591 | 105611282 | 14 | JAG2 | Brain morphology (MOSTest) |
| rs2242635 | 105614974 | 14 | JAG2 | Cortical surface area |
| rs2242634 | 105615026 | 14 | JAG2 | Cortical thickness |
| rs3784240 | 105623261 | 14 | JAG2 | Fasting plasma glucose |
| rs2072674 | 105623512 | 14 | JAG2 | Vertex-wise sulcal depth |
| rs2239285 | 105630465 | 14 | JAG2 | Vertex-wise cortical thickness |
| rs4983589 | 105633544 | 14 | JAG2 | Cortical surface area (MOSTest) |
| rs138903858 | 105634487 | 14 | JAG2 | Body composition (MOSTest) |
| rs12894531 | 105640076 | 14 | NUDT14 | Immature fraction of reticulocytes |
| rs11625865 | 105644421 | 14 | NUDT14 | High light scatter reticulocyte count |
| rs4983590 | 105646179 | 14 | NUDT14 | White blood cell count |
| rs6576086 | 105873144 | 14 | TEX22 | Exploratory eye movement dysfunction in schizophrenia (responsive search score) |
| rs8021311 | 105906073 | 14 | MTA1 | Bilirubin (Z,Z) levels in elite athletes |
| rs80324969 | 105917816 | 14 | MTA1 | Serum levels of protein ASPRV1 |
| rs4983413 | 105930406 | 14 | MTA1 | Non-albumin protein levels |
| rs28835411 | 52455709 | 15 | GNB5 | COVID-19 |
| rs8038456 | 52467546 | 15 | GNB5 | Uveal melanoma |
| rs3825801 | 52517714 | 15 | MYO5C | Type 2 diabetes |
| rs77102195 | 52860366 | 15 | ARPP19 | Metabolonic lactone sulfate levels |
| rs149663666 | 53062068 | 15 | ONECUT1 | Sex hormone-binding globulin levels adjusted for BMI |
| rs10518694 | 53072673 | 15 | ONECUT1 | Body mass index |
| rs2460 | 53073084 | 15 | ONECUT1 | Leisure sedentary behaviour (television watching) |
| rs2456507 | 53077634 | 15 | ONECUT1 | Body mass index |
| rs62023490 | 53079024 | 15 | ONECUT1 | Low density lipoprotein cholesterol levels |
| rs2075613 | 53081221 | 15 | ONECUT1 | Total cholesterol levels |
| rs3816752 | 56992844 | 15 | ZNF280D | Educational attainment (years of education) |
| rs2278613 | 57179300 | 15 | LOC145783 | Platelet distribution width |
| rs1660958 | 57184307 | 15 | LOC145783 | Platelet distribution width |
| rs1158246 | 57185969 | 15 | LOC145783 | Platelet count |
| rs7175191 | 57186397 | 15 | LOC145783 | Platelet count |
| rs28371400 | 57188761 | 15 | LOC145783 | same-sex sexual behaviour |
| rs1529597 | 57190064 | 15 | LOC145783 | Educational attainment (years of education) |
| rs78557702 | 57592920 | 15 | LINC00926 | Alcohol-associated liver disease in light drinkers |
| rs2243504 | 57597280 | 15 | LINC00926 | Insomnia |
| rs12595188 | 57616306 | 15 | LOC101928611 | OTU97_8 (Ruminococcaceae) prevalence |
| rs7162310 | 63571234 | 15 | APH1B | Myopia |
| rs12898755 | 63574641 | 15 | APH1B | Myopia (age of diagnosis) |
| rs11071738 | 63580155 | 15 | APH1B | Estimated glomerular filtration rate |
| rs745338 | 63598430 | 15 | APH1B | Urea levels |
| rs11634841 | 63641427 | 15 | CA12 | Alzheimer's disease polygenic risk score (upper quantile vs lower quantile) |
| rs139132462 | 63646385 | 15 | CA12 | Vaginal microbiome MetaCyc pathway (ALL-CHORISMATE-PWY\|superpathway of chorismate metabolism) |
| rs16946963 | 63667406 | 15 | CA12 | RS-warfarin levels |
| rs12443113 | 65355468 | 15 | RASL12 | White matter hyperintensity volume |
| rs2585035 | 65496475 | 15 | CILP | ECG latent space |
| rs2585034 | 65496587 | 15 | CILP | ECG cardiac MRI latent space |
| rs2681038 | 65497849 | 15 | CILP | Height |
| rs2019185 | 65500435 | 15 | CILP | Electrocardiogram morphology (amplitude at temporal datapoints) |
| rs11638062 | 87163264 | 15 | AGBL1 | Acute lymphoblastic leukemia |
| rs2469184 | 87191977 | 15 | AGBL1 | Activated partial thromboplastin time |
| rs8027439 | 87252550 | 15 | AGBL1 | RS-warfarin levels |
| rs10520638 | 87253534 | 15 | AGBL1 | Modified Stumvoll Insulin Sensitivity Index (BMI interaction) |
| rs12438353 | 87315080 | 15 | AGBL1 | Ischemic stroke |
| rs11631662 | 87318586 | 15 | AGBL1 | 2,3-dihydroxyisovalerate levels in elite athletes |
| rs16977994 | 87343422 | 15 | AGBL1 | Childhood median systolic blood pressure percentile |
| rs2034633 | 87353775 | 15 | AGBL1 | Childhood median systolic blood pressure percentile |
| rs9944243 | 88514162 | 15 | NTRK3 | Blood protein levels |
| rs28735437 | 88514855 | 15 | NTRK3 | NT-3 growth factor receptor levels |
| rs11073757 | 88523397 | 15 | NTRK3 | Global cognition (Mini Mental State Examination) (longitudinal) |
| rs4887348 | 88571534 | 15 | NTRK3 | NT-3 growth factor receptor levels |
| rs12441487 | 88584310 | 15 | NTRK3 | Height |
| rs2283430 | 89876236 | 15 | POLG | Height |
| rs176647 | 89921969 | 15 | LINC00925 | Insomnia |
| rs208826 | 89926799 | 15 | LINC00925 | Smoking initiation |
| rs150353 | 89928189 | 15 | LINC00925 | Smoking behaviour (cigarette pack-years) |
| rs150294 | 89931148 | 15 | LINC00925 | Smoking status (ever vs never smokers) |
| rs176648 | 89939528 | 15 | LINC00925 | Drusen volume within a central 5 mm circle (model 2) |
| rs2072693 | 90014945 | 15 | RHCG | Number of decayed, missing and filled tooth surfaces or use of dentures |
| rs2289352 | 90022837 | 15 | RHCG | Educational attainment |
| rs17807723 | 90023558 | 15 | RHCG | Triglyceride levels |

| | | | | |
|---|---|---|---|---|
| rs16944405 | 90975517 | 15 | IQGAP1 | Glaucoma (primary open-angle) |
| rs8042861 | 90977333 | 15 | IQGAP1 | Multiple sclerosis |
| rs2074585 | 91009484 | 15 | IQGAP1 | Basophil percentage of white cells |
| rs2238325 | 91011262 | 15 | IQGAP1 | Lymphocyte count |
| rs7173724 | 91036855 | 15 | IQGAP1 | Neutrophil percentage of white cells |
| rs11609 | 91043765 | 15 | IQGAP1 | Height |
| rs3540 | 91045408 | 15 | IQGAP1 | Allergic disease (asthma, hay fever or eczema) |
| rs11637971 | 91079863 | 15 | CRTC3 | Heel bone mineral density |
| rs3862435 | 91082532 | 15 | CRTC3 | Response to exercise (triglyceride level interaction) |
| rs8033595 | 91083353 | 15 | CRTC3 | Height |
| rs12915189 | 91114376 | 15 | CRTC3 | Information processing speed |
| rs16944613 | 91139098 | 15 | CRTC3 | Colorectal cancer |
| rs4932352 | 91145800 | 15 | CRTC3 | Thyroid stimulating hormone levels |
| rs5011652 | 91166608 | 15 | CRTC3 | Eosinophil counts |
| rs7495132 | 91172901 | 15 | CRTC3 | Ulcerative colitis |
| rs7165170 | 91181489 | 15 | CRTC3 | Crohn's disease |
| rs12909526 | 91189024 | 15 | LOC101926895 | Serum total protein levels |
| rs10083558 | 91202504 | 15 | LOC101926895 | Eosinophil counts |
| rs35787687 | 91281211 | 15 | BLM | Daytime sleep phenotypes |
| rs3815003 | 91312823 | 15 | BLM | Delta-5 desaturase activity |
| rs8031341 | 91313224 | 15 | BLM | Delta-5 desaturase activity |
| rs401549 | 91356504 | 15 | BLM | Height |
| rs7174740 | 93531714 | 15 | CHD2 | Body mass index |
| rs7179432 | 93564972 | 15 | CHD2 | IgG glycosylation |
| rs12905032 | 93567527 | 15 | CHD2 | Mean corpuscular hemoglobin |
| rs12906163 | 93567864 | 15 | CHD2 | Mean spheric corpuscular volume |
| rs9672839 | 93568884 | 15 | CHD2 | Sex hormone-binding globulin levels adjusted for BMI |
| rs1044778 | 93570786 | 15 | CHD2 | Mean corpuscular hemoglobin |
| rs3784526 | 101716797 | 15 | CHSY1 | Granulocyte percentage of myeloid white cells |
| rs28364839 | 101718097 | 15 | CHSY1 | Fracture of forearm |
| rs12593811 | 101721892 | 15 | CHSY1 | Corneal astigmatism |
| rs8033531 | 101730181 | 15 | CHSY1 | Height |
| rs67175901 | 101748227 | 15 | CHSY1 | Basophil count |
| rs8043304 | 101751698 | 15 | CHSY1 | Vertical cup-disc ratio |
| rs28623369 | 101753394 | 15 | CHSY1 | Vertical cup-disc ratio |
| rs8042424 | 101762539 | 15 | CHSY1 | Hip circumference adjusted for BMI |
| rs2412005 | 101767591 | 15 | CHSY1 | Prostate cancer |
| rs8036083 | 101767822 | 15 | CHSY1 | Ankylosing spondylitis |
| rs12442366 | 101773563 | 15 | CHSY1 | Dupuytren's disease |
| rs8040150 | 101774366 | 15 | CHSY1 | Height |
| rs72768580 | 101774608 | 15 | CHSY1 | Height |
| rs752092 | 101781934 | 15 | CHSY1 | Central corneal thickness |
| rs28445340 | 101786733 | 15 | CHSY1 | Vertex-wise cortical thickness |
| rs9672951 | 101786747 | 15 | CHSY1 | Cortical thickness |
| rs11636578 | 101787969 | 15 | CHSY1 | Hippocampal subfield left CA1 volume (body) |
| rs116643984 | 101791212 | 15 | CHSY1 | Blood pressure (pleiotropy model 2 SBP adjusted for estimated causal effects x DBP) |
| rs1871975 | 101862970 | 15 | PCSK6 | N-delta-acetylornithine levels in elite athletes |
| rs7182874 | 101865472 | 15 | PCSK6 | Relative hand skill in reading disability |
| rs12900794 | 101874841 | 15 | PCSK6 | Severe COVID-19 infection |
| rs11855415 | 101875123 | 15 | PCSK6 | Handedness in dyslexia |
| rs10152756 | 101890861 | 15 | PCSK6 | Height |
| rs10152471 | 101890913 | 15 | PCSK6 | Triglyceride levels |
| rs7178801 | 101895928 | 15 | PCSK6 | vascular endothelial growth factor D levels |
| rs11247287 | 101905533 | 15 | PCSK6 | Triglycerides |
| rs8029570 | 101905966 | 15 | PCSK6 | Free cholesterol to total lipids ratio in IDL |
| rs3760083 | 3450052 | 16 | ZSCAN32 | OTU97_13 (Subdoligranulum) prevalence |
| rs1878931 | 3450907 | 16 | ZSCAN32 | Body mass index (adult) |
| rs40363 | 3509056 | 16 | NAA60 | Tuberculosis |
| rs757270 | 3534451 | 16 | NAA60 | Systolic blood pressure (standard GWA) |
| rs2240075 | 3535540 | 16 | NAA60 | Diastolic blood pressure |
| rs140345617 | 3576345 | 16 | CLUAP1 | Vaginal microbiome MetaCyc pathway (NAD-BIOSYNTHESIS-II\|NAD salvage pathway II) |
| rs3751836 | 3582969 | 16 | CLUAP1 | Height |
| rs3751837 | 3583173 | 16 | CLUAP1 | Lung function (FVC) |
| rs9790 | 3586230 | 16 | CLUAP1 | Estimated glomerular filtration rate (creatinine) |
| rs12596890 | 3588068 | 16 | CLUAP1 | Educational attainment |
| rs12448257 | 3599655 | 16 | NLRC3 | Body mass index |
| rs12445430 | 3613126 | 16 | NLRC3 | Body mass index |
| rs758747 | 3627358 | 16 | NLRC3 | Body mass index |
| rs12933120 | 3634746 | 16 | SLX4 | Type 2 diabetes |
| rs8061528 | 3656482 | 16 | SLX4 | Type 2 diabetes |
| rs1053874 | 3707747 | 16 | DNASE1 | Body mass index |
| rs11646287 | 3719344 | 16 | TRAP1 | Neutrophil count |
| rs2074800 | 3725682 | 16 | TRAP1 | Non-alcoholic fatty liver disease activity score in non-alcoholic fatty liver disease |
| rs3794702 | 3730613 | 16 | TRAP1 | Smoking initiation |
| rs2158962 | 3738027 | 16 | TRAP1 | Infliximab-induced mucosal healing in Crohn's disease |
| rs2072379 | 3738887 | 16 | TRAP1 | Estimated glomerular filtration rate (creatinine) |
| rs75712687 | 3745273 | 16 | TRAP1 | Sex hormone-binding globulin levels |
| rs1639150 | 3747204 | 16 | TRAP1 | Height |
| rs8048452 | 3748965 | 16 | TRAP1 | Smoking initiation |

| | | | | |
|---|---|---|---|---|
| rs75719766 | 57082024 | 16 | NLRC5 | Low-density lipoprotein levels (MTAG) |
| rs72780017 | 57083353 | 16 | NLRC5 | High density lipoprotein cholesterol levels |
| rs7190199 | 57101340 | 16 | NLRC5 | High density lipoprotein cholesterol levels |
| rs289710 | 57103579 | 16 | NLRC5 | High-density lipoprotein levels (MTAG) |
| rs66807230 | 57112289 | 16 | NLRC5 | High density lipoprotein cholesterol levels |
| rs154045 | 57114924 | 16 | NLRC5 | High-density lipoprotein levels (MTAG) |
| rs8058512 | 57151796 | 16 | CPNE2 | High-density lipoprotein levels (MTAG) |
| rs3829501 | 57155995 | 16 | CPNE2 | Low-density lipoprotein levels (MTAG) |
| rs113552373 | 57180536 | 16 | CPNE2 | Human milk oligosaccharide concentration (lacto-N-fucopentaose III) |
| rs60403102 | 57297312 | 16 | PLLP | Blood urea nitrogen levels |
| rs11640439 | 57303194 | 16 | PLLP | Sleep duration |
| rs117932065 | 57318078 | 16 | PLLP | High density lipoprotein cholesterol levels |
| rs4359426 | 57392733 | 16 | CCL22 | High density lipoprotein cholesterol levels |
| rs223819 | 57394862 | 16 | CCL22 | Asthma |
| rs668100 | 57407646 | 16 | CX3CL1 | Vaginal microbiome MetaCyc pathway (PWY-7328\|superpathway of UDP-glucose-derived O-antigen building blocks biosynthesis) |
| rs170364 | 57409934 | 16 | CX3CL1 | C-C motif chemokine 22 levels |
| rs62037082 | 57410082 | 16 | CX3CL1 | CCL17/CD69 protein level ratio |
| rs671623 | 57412802 | 16 | CX3CL1 | Blood protein levels |
| rs670211 | 57414001 | 16 | CX3CL1 | Blood protein levels in cardiovascular risk |
| rs170361 | 57414623 | 16 | CX3CL1 | CX3CL1/EPHB6 protein level ratio |
| rs8102 | 57418725 | 16 | CX3CL1 | Blood protein levels |
| rs9302690 | 57442968 | 16 | CCL17 | C-C motif chemokine 17 levels |
| rs223896 | 57443146 | 16 | CCL17 | Thymus and reactivation regulated chemokine levels |
| rs16956811 | 57444002 | 16 | CCL17 | Blood protein levels |
| rs223900 | 57445376 | 16 | CCL17 | C-C motif chemokine 17 levels |
| rs623685 | 57515188 | 16 | DOK4 | Idiopathic inflammatory myopathy |
| rs8052123 | 57563671 | 16 | CCDC102A | Cataracts in type 2 diabetes |
| rs4783997 | 57580478 | 16 | GPR114 | Mean corpuscular volume |
| rs1859259 | 57596552 | 16 | GPR114 | Mosaic loss of chromosome Y (Y chromosome dosage) |
| rs187908447 | 57660878 | 16 | GPR56 | Plasma vanadium levels |
| rs2004823 | 57686303 | 16 | GPR56 | Red blood cell count |
| rs1801257 | 57689385 | 16 | GPR56 | Adhesion G-protein coupled receptor G1 levels |
| rs11866053 | 57692456 | 16 | GPR56 | Prostate cancer |
| rs16971384 | 72931085 | 16 | ZFHX3 | LDL cholesterol |
| rs6499595 | 72934443 | 16 | ZFHX3 | Number of sexual partners |
| rs17618144 | 72942424 | 16 | ZFHX3 | Externalizing behaviour (multivariate analysis) |
| rs17680397 | 72944807 | 16 | ZFHX3 | Lifetime smoking |
| rs4788676 | 72950468 | 16 | ZFHX3 | Smoking status |
| rs12930834 | 72951627 | 16 | ZFHX3 | Smoking status (ever vs never smokers) |
| rs4788679 | 72956544 | 16 | ZFHX3 | Height |
| rs12922127 | 72958967 | 16 | ZFHX3 | Lung function (FVC) |
| rs4788488 | 72960283 | 16 | ZFHX3 | Lung function (FEV1/FVC) |
| rs756717 | 72996162 | 16 | ZFHX3 | Body mass index |
| rs7190256 | 72996983 | 16 | ZFHX3 | Age-related diseases and mortality |
| rs12325072 | 73006574 | 16 | ZFHX3 | Heart failure |
| rs7199343 | 73009024 | 16 | ZFHX3 | Kawasaki disease |
| rs17681554 | 73016768 | 16 | ZFHX3 | Urine 6-hydroxymelatonin sulfate-to-creatinine ratio |
| rs16971465 | 73017061 | 16 | ZFHX3 | Schizophrenia |
| rs1858800 | 73024276 | 16 | ZFHX3 | Attention deficit hyperactivity disorder (childhood) |
| rs7193343 | 73029160 | 16 | ZFHX3 | Ischemic stroke (cardioembolic) |
| rs57828240 | 73041940 | 16 | ZFHX3 | Medication use (antithrombotic agents) |
| rs67329386 | 73048367 | 16 | ZFHX3 | Atrial fibrillation |
| rs2106261 | 73051620 | 16 | ZFHX3 | Atrial fibrillation |
| rs2359171 | 73053022 | 16 | ZFHX3 | Atrial fibrillation |
| rs4499262 | 73059159 | 16 | ZFHX3 | Incident atrial fibrillation |
| rs1548373 | 73059861 | 16 | ZFHX3 | Intelligence |
| rs56274700 | 73063054 | 16 | ZFHX3 | Estimated glomerular filtration rate |
| rs876727 | 73067761 | 16 | ZFHX3 | Atrial fibrillation |
| rs879324 | 73068678 | 16 | ZFHX3 | Stroke (ischemic) |
| rs12932445 | 73069888 | 16 | ZFHX3 | Stroke |
| rs4788694 | 73070083 | 16 | ZFHX3 | Body mass index |
| rs34003787 | 73071381 | 16 | ZFHX3 | Intrinsic epigenetic age acceleration |
| rs728646 | 73077574 | 16 | ZFHX3 | Height |
| rs11641701 | 73079212 | 16 | ZFHX3 | Height |
| rs7500280 | 76902847 | 16 | MIR4719 | Seropositivity (peptide co-occurrence module 1: CMV, (others)) |
| rs72477006 | 89950299 | 16 | TCF25 | Red vs. brown/black hair color |
| rs11641790 | 89961957 | 16 | TCF25 | Metabolite levels |
| rs4785736 | 89969593 | 16 | TCF25 | Red vs. brown/black hair color |
| rs11862382 | 89972532 | 16 | TCF25 | Red vs. brown/black hair color |
| rs3212358 | 89984739 | 16 | MC1R | Cys-gly, oxidized levels |
| rs3212371 | 89987201 | 16 | MC1R | Cutaneous malignant melanoma |
| rs2302898 | 89998794 | 16 | TUBB3 | Red vs. brown/black hair color |
| rs7195043 | 90020861 | 16 | DEF8 | Basal cell carcinoma |
| rs8049897 | 90024202 | 16 | DEF8 | Skin reflectance |
| rs74800773 | 90024970 | 16 | DEF8 | Blond vs. brown/black hair color |
| rs8063761 | 90027626 | 16 | DEF8 | Squamous cell carcinoma |
| rs4785751 | 90029417 | 16 | DEF8 | Red vs non-red hair color |
| rs7192165 | 90032048 | 16 | DEF8 | Hemoglobin concentration |
| rs11649211 | 90039450 | 16 | AFG3L1P | Low tan response |

| | | | | |
|---|---|---|---|---|
| rs4408545 | 90044028 | 16 | AFG3L1P | Melanoma |
| rs62054570 | 90063890 | 16 | AFG3L1P | Hip circumference adjusted for BMI |
| rs4785763 | 90066936 | 16 | AFG3L1P | Melanoma |
| rs73283869 | 90067184 | 16 | AFG3L1P | Non-melanoma skin cancer |
| rs66701606 | 90080852 | 16 | DBNDD1 | Waist circumference adjusted for body mass index |
| rs11648785 | 90084561 | 16 | DBNDD1 | Tanning |
| rs7498985 | 90103117 | 16 | GAS8 | Basal cell carcinoma (MTAG) |
| rs3785181 | 90105333 | 16 | GAS8 | Verbal learning |
| rs4785767 | 90106525 | 16 | GAS8 | Cancer (pleiotropy) |
| rs9933627 | 90107353 | 16 | GAS8 | Basal cell carcinoma |
| rs2241033 | 90109372 | 16 | GAS8 | Schizophrenia |
| rs3743825 | 90109711 | 16 | GAS8 | Alzheimer's disease polygenic risk score (upper quantile vs lower quantile) |
| rs4785612 | 90113107 | 16 | URAHP | Self-reported math ability |
| rs72809177 | 12460291 | 17 | LINC00670 | Balding type 1 |
| rs12150051 | 12488114 | 17 | LINC00670 | Survival in coronary artery disease |
| rs9889434 | 12538710 | 17 | LINC00670 | OTU97_74 (Alistipes) abundance |
| rs72811264 | 12576704 | 17 | LOC101928418 | Prostate cancer |
| rs72811270 | 12585459 | 17 | LOC101928418 | Prostate cancer |
| rs8073609 | 12608271 | 17 | LOC101928418 | Blood pressure (pleiotropy model 1 DBP adjusted for estimated causal effects x SBP) |
| rs55941572 | 12612931 | 17 | MYOCD | Atrial fibrillation (MTAG) |
| rs72811294 | 12618680 | 17 | MYOCD | Atrial fibrillation |
| rs12453295 | 12634791 | 17 | MYOCD | Electrocardiographic traits (multivariate) |
| rs35712872 | 12635871 | 17 | MYOCD | PR interval |
| rs7224591 | 12636733 | 17 | MYOCD | Delayed reward discounting |
| rs9905820 | 12637371 | 17 | MYOCD | Testosterone levels |
| rs1984481 | 12637742 | 17 | MYOCD | PR interval |
| rs16946539 | 12638478 | 17 | MYOCD | QRS duration |
| rs9913797 | 12689430 | 17 | LOC100128006 | OTU97_23 (Faecalibacterium) prevalence |
| rs4792287 | 12744186 | 17 | ARHGAP44 | Metabolite levels |
| rs8066891 | 15008430 | 17 | CDRT8 | Carotid plaques in rheumatoid arthritis |
| rs77366811 | 19266620 | 17 | B9D1 | Educational attainment |
| rs2233083 | 19286463 | 17 | MAPK7 | Alzheimer's disease |
| rs157386 | 19699121 | 17 | ULK2 | Color vision defects (Tritan) |
| rs58745116 | 33803592 | 17 | SLFN12L | White blood cell count |
| rs9912354 | 33805730 | 17 | SLFN12L | FSC-A on Natural Killer |
| rs12451746 | 33806863 | 17 | SLFN12L | GZMB/IL15 protein level ratio |
| rs35927274 | 33810883 | 17 | SLFN12L | Ck-beta-8-1 levels |
| rs4796089 | 33814758 | 17 | SLFN12L | Neutrophil count |
| rs8073060 | 33875262 | 17 | SLFN14 | Platelet distribution width |
| rs79007502 | 33880305 | 17 | SLFN14 | LBR/RWDD1 protein level ratio |
| rs116169465 | 33881017 | 17 | SLFN14 | Platelet distribution width |
| rs10512472 | 33884804 | 17 | SLFN14 | Plateletcrit |
| rs11653357 | 33923607 | 17 | AP2B1 | Platelet distribution width |
| rs225295 | 33925755 | 17 | AP2B1 | Reticulocyte fraction of red cells |
| rs225285 | 33931084 | 17 | AP2B1 | Highest math class taken (MTAG) |
| rs225245 | 33946107 | 17 | AP2B1 | High light scatter reticulocyte count |
| rs16971241 | 33968102 | 17 | AP2B1 | Mean platelet volume |
| rs9897552 | 33974481 | 17 | AP2B1 | Platelet-to-lymphocyte ratio |
| rs9915021 | 33976619 | 17 | AP2B1 | High light scatter reticulocyte count |
| rs226090 | 34034059 | 17 | AP2B1 | Morning person |
| rs17638867 | 34052513 | 17 | AP2B1 | Cognitive ability, years of educational attainment or schizophrenia (pleiotropy) |
| rs73990293 | 34065903 | 17 | RASL10B | Clopidogrel active metabolite levels |
| rs12944858 | 34067892 | 17 | RASL10B | C-C motif chemokine 15 levels |
| rs3744374 | 34072555 | 17 | GAS2L2 | Childhood ear infection |
| rs11650416 | 34077428 | 17 | GAS2L2 | C-C motif chemokine 16 levels |
| rs2567524 | 70779765 | 17 | SLC39A11 | Metabolite levels |
| rs4793310 | 70783318 | 17 | SLC39A11 | Metabolite levels |
| rs4796796 | 76614120 | 17 | LOC101928710 | Serum bilirubin levels in metabolic syndrome |
| rs11654527 | 76703516 | 17 | CYTH1 | Gamma glutamyl transpeptidase |
| rs16971669 | 76732338 | 17 | CYTH1 | Red cell distribution width |
| rs17736589 | 76737118 | 17 | CYTH1 | Ulcerative colitis |
| rs62075585 | 76762039 | 17 | CYTH1 | Systolic blood pressure |
| rs7224711 | 76772288 | 17 | CYTH1 | Type 2 diabetes |
| rs112210999 | 76777574 | 17 | CYTH1 | Smoking initiation |
| rs3744803 | 76793533 | 17 | USP36 | Facial phenotypes (left palpebrale fissure height) |
| rs3744802 | 76793600 | 17 | USP36 | Glycated hemoglobin levels |
| rs11077397 | 76796463 | 17 | USP36 | Height |
| rs1057040 | 76799795 | 17 | USP36 | Systolic blood pressure |
| rs11649964 | 76800182 | 17 | USP36 | Whole body fat free mass (UKB data field 23101) |
| rs12942645 | 76804316 | 17 | USP36 | Red cell distribution width |
| rs7218605 | 76811803 | 17 | USP36 | Systolic blood pressure |
| rs900348 | 76833429 | 17 | USP36 | Heel bone mineral density |
| rs9900972 | 76868614 | 17 | TIMP2 | Obesity-related traits |
| rs2009196 | 76870581 | 17 | TIMP2 | Periodontitis (CDC/AAP) |
| rs7342889 | 76874461 | 17 | TIMP2 | Gut microbiota relative abundance (bacterial taxa) |
| rs8066695 | 76913843 | 17 | TIMP2 | Waist-to-hip ratio adjusted for BMI |
| rs3826311 | 76971526 | 17 | LGALS3BP | Serum levels of protein LGALS3BP |
| rs11891 | 76988785 | 17 | CANT1 | Magnesium levels |
| rs329007 | 9522606 | 18 | RALBP1 | LDL cholesterol |

| | | | | |
|---|---|---|---|---|
| rs7240432 | 9590896 | 18 | PPP4R1 | Self-reported math ability (MTAG) |
| rs680478 | 9609957 | 18 | PPP4R1 | High fluorescence immature platelet fraction |
| rs583306 | 9617742 | 18 | LOC101927323 | Mean platelet volume |
| rs667695 | 9618049 | 18 | LOC101927323 | Mean platelet volume during third trimester of pregnancy |
| rs3745060 | 55021725 | 18 | ST8SIA3 | Height |
| rs12606549 | 55022163 | 18 | ST8SIA3 | Ambidextrousness (ambidextrous vs right-handed) |
| rs1153628 | 55026072 | 18 | ST8SIA3 | Bitter alcoholic beverage consumption |
| rs1942964 | 55027212 | 18 | ST8SIA3 | Drinks per week |
| rs79426955 | 72983972 | 18 | TSHZ1 | Time to colon cancer recurrence |
| rs3826609 | 72997677 | 18 | TSHZ1 | Diastolic blood pressure |
| rs55679337 | 72999359 | 18 | TSHZ1 | Diastolic blood pressure |
| rs74540285 | 74972027 | 18 | GALR1 | Height |
| rs77169818 | 74980601 | 18 | GALR1 | Waist-to-hip ratio adjusted for BMI |
| rs79005430 | 29934907 | 19 | LOC284395 | Ferritin levels |
| rs78770294 | 29990066 | 19 | LOC284395 | Protein quantitative trait loci (liver) |
| rs117223925 | 30021446 | 19 | VSTM2B | Crohn's disease |
| rs3786516 | 30100319 | 19 | POP4 | Systolic blood pressure |
| rs7255293 | 30104198 | 19 | POP4 | Heart rate response to exercise |
| rs997669 | 30304483 | 19 | CCNE1 | Cancer (pleiotropy) |
| rs3218036 | 30305684 | 19 | CCNE1 | Height |
| rs7257694 | 30314666 | 19 | CCNE1 | Diastolic blood pressure |
| rs436143 | 34245941 | 19 | CHST8 | 1-palmitoyl-GPC (16:0) levels in elite athletes |
| rs58499745 | 34261220 | 19 | CHST8 | Vaginal microbiome MetaCyc pathway (PWY-7013\|L-1,2-propanediol degradation) |
| rs321946 | 52859828 | 19 | ZNF610 | Dental caries (decayed, missing and filled teeth) |
| rs324126 | 52886137 | 19 | ZNF880 | Colonoscopy-negative controls vs population controls |
| rs16986856 | 56497960 | 19 | NLRP8 | Obesity-related traits |
| rs116895675 | 56546045 | 19 | NLRP5 | Cardio-cerebrovascular disease in dyslipidemia |
| rs16986899 | 56549510 | 19 | NLRP5 | 1-linoleoyl-GPC (18:2) levels in elite athletes |
| rs7255126 | 56551608 | 19 | NLRP5 | Plasma omega-3 polyunsaturated fatty acid levels (alphalinolenic acid) |
| rs79395320 | 56572633 | 19 | NLRP5 | Geographic atrophy within a central 5 mm circle (model 1) |
| rs4077285 | 56599405 | 19 | ZNF787 | Sex hormone-binding globulin levels adjusted for BMI |
| rs73059933 | 56609303 | 19 | ZNF787 | Coronary artery disease |
| rs524829 | 56623139 | 19 | ZNF787 | Mean platelet volume during second trimester of pregnancy |
| rs7255998 | 56657013 | 19 | ZNF444 | Heart rate response to beta blockers (atenolol monotherapy) |
| rs200137244 | 3673296 | 20 | SIGLEC1 | Gamma glutamyl transpeptidase |
| rs3746638 | 3675333 | 20 | SIGLEC1 | Visceral fat |
| rs754526 | 3675899 | 20 | SIGLEC1 | Serum levels of protein COLEC12 |
| rs525339 | 3677014 | 20 | SIGLEC1 | Liver enzyme levels (gamma-glutamyl transferase) |
| rs673114 | 3677736 | 20 | SIGLEC1 | Sialoadhesin levels |
| rs12624922 | 3680280 | 20 | SIGLEC1 | Collectin-12 levels |
| rs1018493 | 3682011 | 20 | SIGLEC1 | Platelet count |
| rs567189 | 3682730 | 20 | SIGLEC1 | Galectin-3-binding protein levels |
| rs540691 | 3683335 | 20 | SIGLEC1 | White blood cell count |
| rs611847 | 3684022 | 20 | SIGLEC1 | Lymphocyte count |
| rs150358287 | 3687141 | 20 | SIGLEC1 | Glutamate receptor ionotropic, delta-2 levels |
| rs910652 | 3727970 | 20 | HSPA12B | Height |
| rs58505239 | 3731139 | 20 | HSPA12B | Spontaneous preterm birth without premature rupture of membranes |
| rs658495 | 3737495 | 20 | C20orf27 | Survival in colon cancer |
| rs6116037 | 3760548 | 20 | SPEF1 | Hippocampal volume |
| rs4815603 | 3772527 | 20 | CDC25B | Height |
| rs3761218 | 3776175 | 20 | CDC25B | Bipolar disorder |
| rs4815605 | 3777946 | 20 | CDC25B | Red blood cell count |
| rs4815606 | 3778042 | 20 | CDC25B | Mean corpuscular volume |
| rs6133067 | 3839754 | 20 | MAVS | OTU97_113 (Bacteroidales) abundance |
| rs80216136 | 40033119 | 20 | CHD6 | Height |
| rs6029662 | 40035965 | 20 | CHD6 | Total cholesterol levels |
| rs7265695 | 40043096 | 20 | CHD6 | Diastolic blood pressure |
| rs4142393 | 40078085 | 20 | CHD6 | Total cholesterol levels |
| rs6029691 | 40094364 | 20 | CHD6 | Refractive astigmatism |
| rs1010305 | 40101541 | 20 | CHD6 | Total cholesterol levels |
| rs1010304 | 40101647 | 20 | CHD6 | Verbal declarative memory |
| rs980984 | 44333052 | 20 | WFDC10B | Metabolic signature |
| rs11697801 | 44429815 | 20 | DNTTIP1 | Ubiquitin-conjugating enzyme E2 C levels |
| rs11537645 | 44441403 | 20 | UBE2C | Height |
| rs4812973 | 44507536 | 20 | ZSWIM3 | 3-hydroxypropylmercapturic acid levels in smokers |
| rs6065904 | 44534651 | 20 | PLTP | Gut microbiota (functional units) |
| rs378114 | 44538427 | 20 | PLTP | Metabolic signature |
| rs435306 | 44538484 | 20 | PLTP | HDL cholesterol |
| rs6065908 | 44569930 | 20 | PCIF1 | Serum metabolite levels |
| rs7679 | 44576502 | 20 | PCIF1 | Gut microbiota (functional units) |
| rs3746503 | 44578961 | 20 | ZNF335 | High density lipoprotein cholesterol levels |
| rs6124760 | 44581453 | 20 | ZNF335 | HDL cholesterol levels |
| rs13040315 | 44585793 | 20 | ZNF335 | Verbal learning (oral presentation) |
| rs3827066 | 44586023 | 20 | ZNF335 | Coronary artery disease |
| rs3746506 | 44586036 | 20 | ZNF335 | Multi-trait sex score |
| rs79645412 | 44588816 | 20 | ZNF335 | High density lipoprotein cholesterol levels |
| rs6073972 | 44590298 | 20 | ZNF335 | Red blood cell count |
| rs3746513 | 44594228 | 20 | ZNF335 | Total cholesterol to total lipids ratio in small LDL |
| rs6032606 | 44596207 | 20 | ZNF335 | Celiac disease |

| | | | | |
|---|---|---|---|---|
| rs3092684 | 44880190 | 20 | CDH22 | Fruit intake (standard GWA) |
| rs2868714 | 44886189 | 20 | CDH22 | Height |
| rs2425817 | 44895286 | 20 | CDH22 | Smoking initiation |
| rs2425818 | 44896562 | 20 | CDH22 | Smoking initiation |
| rs2425819 | 44896846 | 20 | CDH22 | Fruit consumption |
| rs2425836 | 44902046 | 20 | CDH22 | Fresh fruit consumption |
| rs2425840 | 44904838 | 20 | CDH22 | Body mass index |
| rs2425847 | 44907927 | 20 | CDH22 | Self-reported math ability (MTAG) |
| rs2425848 | 44907944 | 20 | CDH22 | Height |
| rs2425856 | 44911954 | 20 | CDH22 | Adult body size |
| rs6065945 | 44921631 | 20 | CDH22 | IgG glycosylation |
| rs6032755 | 44921676 | 20 | CDH22 | Disease progression in age-related macular degeneration |
| rs310614 | 62125233 | 20 | EEF1A2 | Electrocardiogram morphology (amplitude at temporal datapoints) |
| rs310618 | 62127121 | 20 | EEF1A2 | Body mass index (MTAG) |
| rs2297433 | 62293460 | 20 | RTEL1 | Adult diffuse glioma (IDH wildtype) |
| rs6122140 | 62296129 | 20 | RTEL1 | Vaginal microbiome MetaCyc pathway (RHAMCAT-PWY\|L-rhamnose degradation I) |
| rs3848669 | 62300811 | 20 | RTEL1 | Eczema |
| rs79834248 | 62302259 | 20 | RTEL1 | Systemic sclerosis (anti-centromere-positive) |
| rs79981941 | 62307938 | 20 | RTEL1 | Telomere length |
| rs6089956 | 62308364 | 20 | RTEL1 | B-cell lymphoblastic leukemia or inflammatory bowel disease (pleiotropy) |
| rs2738782 | 62308570 | 20 | RTEL1 | Educational attainment |
| rs41309367 | 62309554 | 20 | RTEL1 | Leukocyte telomere length |
| rs6010620 | 62309839 | 20 | RTEL1 | Atopic dermatitis |
| rs2297439 | 62311128 | 20 | RTEL1 | Telomere length |
| rs77552606 | 62311129 | 20 | RTEL1 | Prostate cancer |
| rs2297440 | 62312299 | 20 | RTEL1 | Non-glioblastoma glioma |
| rs7361098 | 62312755 | 20 | RTEL1 | Brain region volumes |
| rs75031349 | 62314054 | 20 | RTEL1 | Lung adenocarcinoma |
| rs3787089 | 62316630 | 20 | RTEL1 | Colorectal cancer |
| rs4809324 | 62318220 | 20 | RTEL1 | Glioma (high-grade) |
| rs6062302 | 62320968 | 20 | RTEL1 | Glioblastoma |
| rs6011033 | 62322699 | 20 | RTEL1 | Allergic disease (asthma, hay fever or eczema) |
| rs2236507 | 62323006 | 20 | RTEL1 | Glioma |
| rs730937 | 50665776 | 22 | TUBGCP6 | RBC levels of [Similar to: gamma-Glutamylcysteine; deltaMass: -146.3080 Da] |
| rs17836662 | 50672154 | 22 | TUBGCP6 | Heel bone mineral density |
| rs7511006 | 50676259 | 22 | TUBGCP6 | Obesity-related traits |
| rs2072875 | 50692028 | 22 | MAPK12 | Mitogen-activated protein kinase 12 levels |
| rs742184 | 50697119 | 22 | MAPK12 | Diverticular disease |
| rs2267986 | 50698731 | 22 | MAPK12 | Prolylglycine levels in elite athletes |
| rs3817818 | 50699331 | 22 | MAPK12 | Mitogen-activated protein kinase 12 levels |
| rs9617128 | 50704267 | 22 | MAPK11 | Mitogen-activated protein kinase 12 levels |
| rs5771118 | 50714289 | 22 | PLXNB2 | Body mass index |
| rs1127000 | 50716167 | 22 | PLXNB2 | Inferior parietal area (unadjusted for global measures) |
| rs28418659 | 50717833 | 22 | PLXNB2 | COVID-19 (severe vs tested, not severe) |
| rs11547731 | 50722134 | 22 | PLXNB2 | High light scatter reticulocyte count |
| rs79966207 | 50722408 | 22 | PLXNB2 | Hair color |
| rs4075185 | 50723501 | 22 | PLXNB2 | Alanine aminotransferase levels |
| rs28573806 | 50727792 | 22 | PLXNB2 | Plexin-B2 levels (PLXNB2.9216.100.3) |
| rs28578714 | 50727921 | 22 | PLXNB2 | Plexin-B2 levels |
| rs28379706 | 50728062 | 22 | PLXNB2 | Plexin-B2 level in Chronic kidney disease with hypertension and no diabetes (10855_55) |
| rs68178377 | 50752150 | 22 | DENND6B | Cognitive ability, years of educational attainment or schizophrenia (pleiotropy) |
| rs28642259 | 50785718 | 22 | PPP6R2 | Height |
| rs12162782 | 50853626 | 22 | PPP6R2 | LDL cholesterol levels |
| rs7410608 | 50858813 | 22 | PPP6R2 | Total cholesterol levels |
| rs12167996 | 50875745 | 22 | PPP6R2 | NFASC/PLXNB2 protein level ratio |
| rs11555194 | 50876662 | 22 | PPP6R2 | Height |
| rs12171249 | 50878927 | 22 | PPP6R2 | LDL cholesterol |
| rs5770980 | 50886220 | 22 | SBF1 | Estimated glomerular filtration rate (creatinine) |
| rs5771001 | 50895020 | 22 | SBF1 | Skin reflectance (Melanin index) |
| rs5771040 | 50902390 | 22 | SBF1 | IgG glycosylation |
| rs762669 | 50943423 | 22 | LMF2 | Height |
| rs128941 | 50944163 | 22 | LMF2 | Mitochondrial DNA heteroplasmy (chrM:567:A:ACCCCCC case-only heteroplasmy) |
| rs3091397 | 50957520 | 22 | NCAPH2 | Uridine levels |
| rs140524 | 50960682 | 22 | NCAPH2 | 5-methyluridine (ribothymidine) levels |
| rs2782 | 50961854 | 22 | NCAPH2 | Mean corpuscular volume |
| rs12148 | 50962208 | 22 | NCAPH2 | Red cell distribution width |
| rs131808 | 50963357 | 22 | SCO2 | Red cell distribution width |
| rs131807 | 50963540 | 22 | SCO2 | 5,6-dihydrothymine levels |
| rs74479613 | 50963905 | 22 | SCO2 | Protein SCO2 homolog, mitochondrial levels |
| rs131806 | 50963965 | 22 | SCO2 | Mean corpuscular volume |
| rs131805 | 50964153 | 22 | SCO2 | Granulocyte percentage of myeloid white cells |
| rs11479 | 50964236 | 22 | SCO2 | Mean corpuscular volume |
| rs131804 | 50964862 | 22 | SCO2 | Mean corpuscular hemoglobin |
| rs470119 | 50966914 | 22 | TYMP | Mean corpuscular volume |
| rs131777 | 50991033 | 22 | SYCE3 | Mean corpuscular hemoglobin |
| rs131761 | 51010788 | 22 | CHKB-CPT1B | 2'-deoxyuridine levels |
| rs131758 | 51017082 | 22 | CHKB-CPT1B | Metabolite levels (5-methyluridine (ribothymidine); ribothymidine) |
| rs5770917 | 51017353 | 22 | CHKB-CPT1B | Narcolepsy |
| rs2269382 | 51018911 | 22 | CHKB | Fibrinogen levels |

| | | | | |
|---|---|---|---|---|
| rs74846249 | 51019142 | 22 | CHKB | Arylsulfatase A levels |
| rs131752 | 51023424 | 22 | CHKB-AS1 | Metabolite levels (2'-deoxyuridine; __deoxyuridine) |
| rs131750 | 51024308 | 22 | CHKB-AS1 | Low myopia |
| rs5770924 | 51024542 | 22 | CHKB-AS1 | Choline/ethanolamine kinase levels |

**Table S19. The distinct adaptive Denisovan-introgressed haplotypes in differentiated TYC individuals**

| THL | Gene | iHS | XPEHH | PBS | Traits | TCL | Gene | iHS | XPEHH | PBS | Traits | TSL | Gene | iHS | XPEHH | PBS | Traits | TEL | Gene | iHS | XPEHH | Traits |
|---|---|---|---|---|---|---|---|---|---|---|---|---|---|---|---|---|---|---|---|---|---|---|
| rs3743319 | APH1B | √ | √ | × | | rs7311847 | TMEM117 | √ | √ | × | | rs11950959 | LOC101927 | √ | √ | × | | rs10880598 | TMEM117 | √ | √ | |
| rs4924986 | B9D1 | √ | × | √ | | rs7165361 | CHSY1 | √ | √ | × | | rs76996117 | UAP1 | √ | √ | × | | rs4768546 | TMEM117 | √ | √ | |
| rs12371795 | SOX5 | √ | √ | × | | rs752092 | CHSY1 | √ | √ | × | Central corr | rs17122269 | ARCN1 | √ | × | √ | | rs10880599 | TMEM117 | √ | √ | |
| rs12366963 | SOX5 | √ | √ | × | | rs7182945 | CHSY1 | √ | √ | × | | rs9735449 | ARCN1 | √ | × | √ | | rs10785482 | TMEM117 | √ | √ | |
| rs1355001 | SOX5 | √ | √ | × | | rs11247277 | CHSY1 | √ | √ | × | | rs3895449 | KCNQ2 | √ | × | √ | | rs10880600 | TMEM117 | √ | √ | |
| rs1396197 | SOX5 | √ | √ | × | | rs8032007 | CHSY1 | √ | √ | × | | rs6090411 | KCNQ2 | √ | × | √ | | rs10880601 | TMEM117 | √ | √ | |
| rs11047450 | SOX5 | √ | √ | × | | rs12621791 | ILKAP | √ | √ | × | | rs3787129 | KCNQ2 | √ | × | √ | | rs6730027 | ANKRD44 | √ | √ | |
| rs11047451 | SOX5 | √ | √ | × | | rs4663861 | ILKAP | √ | √ | × | | rs6766 | RAB17 | √ | √ | × | | rs7584406 | ANKRD44 | √ | √ | |
| rs11047452 | SOX5 | √ | √ | × | | rs7795187 | NRF1 | √ | √ | × | | rs78444319 | TRPM3 | √ | × | √ | | rs7561423 | ANKRD44 | √ | √ | |
| rs34020085 | TBC1D1 | √ | √ | × | | rs11844206 | NUDT14 | √ | √ | × | | rs7165361 | CHSY1 | √ | × | × | | rs4241193 | ANKRD44 | √ | √ | |
| rs17607179 | TBC1D1 | √ | √ | × | | rs11844119 | NUDT14 | √ | √ | × | | rs7795187 | NRF1 | √ | × | × | | rs76290121 | AP2B1 | √ | √ | |
| rs7671645 | TBC1D1 | √ | √ | × | Lymphocyte-to-monocyte r | rs7971487 | TMEM117 | √ | √ | × | | rs1881926 | TMEM117 | √ | × | × | | rs16971207 | AP2B1 | √ | √ | |
| rs34119528 | TBC1D1 | √ | √ | × | | rs11182329 | TMEM117 | √ | √ | × | | rs10785478 | TMEM117 | √ | × | × | | rs4947866 | GRB10 | √ | √ | |
| rs12645411 | TBC1D1 | √ | √ | × | | rs10880591 | TMEM117 | √ | √ | × | | rs10785479 | TMEM117 | √ | × | × | | rs11974751 | GRB10 | √ | √ | |
| rs7311847 | TMEM117 | √ | √ | × | | rs2031056 | TMEM117 | √ | √ | × | | rs9841214 | VIPR1 | √ | × | × | | rs2051896 | GRB10 | √ | √ | |
| rs9930504 | ZFHX3 | √ | √ | × | | rs4499064 | TMEM117 | √ | √ | × | | rs2267537 | VIPR1 | √ | × | × | | rs1014384 | GRB10 | √ | √ | |
| rs75739666 | ACAP2 | √ | × | × | | rs10880592 | TMEM117 | √ | √ | × | | rs2267538 | VIPR1 | √ | × | × | | rs7794164 | GRB10 | √ | √ | |
| rs41266999 | ACAP2 | √ | × | × | | rs10748379 | TMEM117 | √ | √ | × | | rs2239499 | VIPR1 | √ | × | × | | rs4947873 | GRB10 | √ | √ | |
| rs13325966 | ACAP2 | √ | × | × | | rs10748380 | TMEM117 | √ | √ | × | | rs10838199 | ALKBH3 | √ | × | × | | rs6958750 | GRB10 | √ | √ | |
| rs12054139 | ACAP2 | √ | × | × | | rs4238092 | TMEM117 | √ | √ | × | | rs3740990 | ALKBH3 | √ | × | × | | rs7803278 | GRB10 | √ | √ | |
| rs1994880 | ACAP2 | √ | × | × | | rs4238093 | TMEM117 | √ | √ | × | | rs11639408 | CHSY1 | √ | × | × | | rs6593180 | GRB10 | √ | √ | |
| rs4677843 | ACAP2 | √ | × | × | | rs10506232 | TMEM117 | √ | √ | × | | rs8032573 | CHSY1 | √ | × | × | | rs2301925 | GRB10 | √ | √ | |
| rs9817672 | ACAP2 | √ | × | × | | rs4768055 | TMEM117 | √ | √ | × | | rs12369569 | TMEM117 | √ | × | × | | rs1859571 | GRB10 | √ | √ | |
| rs10212206 | ACAP2 | √ | × | × | | rs11182332 | TMEM117 | √ | √ | × | | rs10880589 | TMEM117 | √ | × | × | | rs11238325 | GRB10 | √ | √ | Hearing problems |
| rs77075906 | ACAP2 | √ | × | × | | rs80107150 | TMEM117 | √ | √ | × | | rs3925073 | TMEM117 | √ | × | × | | rs11238326 | GRB10 | √ | √ | |
| rs9821765 | ACAP2 | √ | × | × | | rs10785474 | TMEM117 | √ | √ | × | | rs12371795 | SOX5 | √ | × | × | | rs2051897 | GRB10 | √ | √ | |
| rs6788868 | ACAP2 | √ | × | × | | rs10785475 | TMEM117 | √ | √ | × | | rs1355001 | SOX5 | √ | × | × | | rs4947886 | GRB10 | √ | √ | |
| rs74353694 | ACOT8 | √ | × | × | | rs10785476 | TMEM117 | √ | √ | × | | rs11047450 | SOX5 | √ | × | × | | rs6963498 | GRB10 | √ | √ | |
| rs4934434 | ACTA2 | √ | × | × | | rs10785477 | TMEM117 | √ | √ | × | | rs77075906 | ACAP2 | √ | × | × | | rs4947888 | GRB10 | √ | √ | |
| rs12775501 | ACTA2 | √ | × | × | Lymphocyte percentage of | rs1881927 | TMEM117 | √ | √ | × | | rs11765221 | AP2B1 | √ | × | × | | rs6593182 | GRB10 | √ | √ | |
| rs72762042 | AGBL1 | √ | × | × | | rs1881926 | TMEM117 | √ | √ | × | | rs7677674 | ARSJ | √ | × | × | Height | rs4947890 | GRB10 | √ | √ | |
| rs79428239 | AGBL1 | √ | × | × | | rs7974214 | TMEM117 | √ | √ | × | | rs77526635 | ASRGL1 | √ | × | × | | rs4947891 | GRB10 | √ | √ | |
| rs76262344 | AGBL1 | √ | × | × | | rs10785478 | TMEM117 | √ | √ | × | | rs3741244 | ASRGL1 | √ | × | × | | rs10842259 | SOX5 | √ | √ | |
| rs16977892 | AGBL1 | √ | × | × | | rs10785479 | TMEM117 | √ | √ | × | | rs298722 | AUH | √ | × | × | | rs2418156 | SOX5 | √ | √ | |
| rs16977893 | AGBL1 | √ | × | × | | rs11182337 | TMEM117 | √ | √ | × | | rs77073324 | AUH | √ | × | × | | rs7139365 | SOX5 | √ | √ | |
| rs2447249 | AGBL1 | √ | × | × | | rs12301015 | TMEM117 | √ | √ | × | | rs2072352 | BLM | √ | × | × | | rs11047238 | SOX5 | √ | √ | |
| rs896616 | AGBL1 | √ | × | × | | rs10880598 | TMEM117 | √ | √ | × | | rs2283448 | BLM | √ | × | × | | rs1498882 | SOX5 | √ | √ | |
| rs7172992 | AGBL1 | √ | × | × | | rs75410353 | TMEM117 | √ | √ | × | | rs2283449 | BLM | √ | × | × | | rs12322111 | SOX5 | √ | √ | Brain morphology (MOSTest) |
| rs12595382 | AGBL1 | √ | × | × | | rs4768546 | TMEM117 | √ | √ | × | | rs387833 | BLM | √ | × | × | | rs10785481 | TMEM117 | √ | √ | |
| rs78852371 | AGBL1 | √ | × | × | | rs10880599 | TMEM117 | √ | √ | × | | rs36967092 | BLM | √ | × | × | | rs3925073 | TMEM117 | √ | √ | |
| rs17702874 | AGBL1 | √ | × | × | | rs12303804 | TMEM117 | √ | √ | × | | rs2270132 | BLM | √ | × | × | | rs6582490 | TMEM117 | √ | √ | |
| rs7181054 | AGBL1 | √ | × | × | | rs10785482 | TMEM117 | √ | √ | × | | rs389480 | BLM | √ | × | × | | rs11182339 | TMEM117 | √ | √ | |
| rs2819446 | AHNAK2 | × | × | √ | | rs10880600 | TMEM117 | √ | √ | × | | rs12592875 | BLM | √ | × | × | | rs2040509 | ZFHX3 | √ | √ | |
| rs17171340 | AMPH | √ | × | × | | rs10880601 | TMEM117 | √ | √ | × | | rs401549 | BLM | √ | × | × | Height | rs4924986 | B9D1 | √ | × | |
| rs6967094 | AMPH | √ | × | × | | rs73275939 | TUBB3 | √ | √ | × | | rs2239291 | BLM | √ | × | × | | rs34119528 | TBC1D1 | √ | × | |
| rs2297866 | ANGPTL2 | × | √ | × | Cortical surface area | rs11863403 | TUBB3 | √ | √ | × | | rs6835747 | CAMK2D | √ | × | × | | rs7311847 | TMEM117 | √ | × | |
| rs79196031 | ANGPTL2 | × | √ | × | | rs11486795 | TUBB3 | √ | √ | × | | rs6740879 | CCDC138 | √ | × | × | | rs75739666 | ACAP2 | √ | × | |
| rs2789511 | ANGPTL2 | × | √ | × | | rs2302898 | TUBB3 | √ | √ | × | Red vs. bro | rs1724184 | CCDC138 | √ | × | × | Birth weight | rs13325966 | ACAP2 | √ | × | |
| rs117912038 | ANGPTL2 | × | √ | × | | rs9841214 | VIPR1 | √ | √ | × | | rs1724195 | CCDC138 | √ | × | × | | rs12054139 | ACAP2 | √ | × | |
| rs1281159 | ANGPTL2 | × | √ | × | | rs2267537 | VIPR1 | √ | √ | × | | rs4387742 | CCDC138 | √ | × | × | | rs10212206 | ACAP2 | √ | × | |
| rs141430518 | ANGPTL2 | × | √ | × | | rs407931 | VIPR1 | √ | √ | × | | rs12478500 | CCDC138 | √ | × | × | | rs77075906 | ACAP2 | √ | × | |
| rs184016199 | ANGPTL2 | × | √ | × | | rs2267538 | VIPR1 | √ | √ | × | | rs975597 | CCDC138 | √ | × | × | | rs7172992 | AGBL1 | √ | × | |
| rs370305162 | ANGPTL2 | × | √ | × | | rs2239499 | VIPR1 | √ | √ | × | | rs7564164 | CCDC138 | √ | × | × | | rs77411926 | ANO4 | √ | × | |
| rs11793399 | ANGPTL2 | × | √ | × | | rs2239500 | VIPR1 | √ | √ | × | | rs10865022 | CCDC138 | √ | × | × | | rs2654991 | ANO4 | √ | × | |
| rs60313786 | ANGPTL2 | × | √ | × | | rs406360 | VIPR1 | √ | √ | × | | rs73952526 | CCDC138 | √ | × | × | | rs589657 | ANO4 | √ | × | |
| rs13298677 | ANGPTL2 | × | √ | × | Height | rs2276829 | VIPR1 | √ | √ | × | | rs13009852 | CCDC138 | √ | × | × | | rs649094 | ANO4 | √ | × | |
| rs11795066 | ANGPTL2 | × | √ | × | Intraocular pressure | rs4788697 | ZFHX3 | √ | √ | × | | rs4676209 | CCDC138 | √ | × | × | | rs677871 | ANO4 | √ | × | |
| rs145298783 | ANGPTL2 | × | √ | × | | rs10838199 | ALKBH3 | √ | √ | × | | rs12464296 | CCDC138 | √ | × | × | | rs225299 | AP2B1 | √ | × | |
| rs80116254 | ANGPTL2 | × | √ | × | | rs3740990 | ALKBH3 | √ | √ | × | | rs11681025 | CCDC138 | √ | × | × | | rs225293 | AP2B1 | √ | × | |
| rs79881767 | ANGPTL2 | × | √ | × | | rs787043 | APBB1IP | √ | √ | × | | rs6727368 | CCDC138 | √ | × | × | | rs16971241 | AP2B1 | √ | × | Mean platelet volume |
| rs2787594 | ANGPTL2 | × | √ | × | Hair color | rs4924985 | B9D1 | √ | √ | × | | rs7600513 | CCDC138 | √ | × | × | | rs12220759 | ARHGAP21 | √ | × | |
| rs138219987 | ANGPTL2 | × | √ | × | | rs4924782 | B9D1 | √ | √ | × | | rs2031262 | CENPP | √ | × | × | | rs28602629 | ARSJ | √ | × | |
| rs11792699 | ANGPTL2 | × | √ | × | | rs6587067 | B9D1 | √ | √ | × | | rs10761156 | CENPP | √ | × | × | | rs7677674 | ARSJ | √ | × | Height |
| rs1281157 | ANGPTL2 | × | √ | × | | rs6587069 | B9D1 | √ | √ | × | | rs2025389 | CENPP | √ | × | × | | rs77526635 | ASRGL1 | √ | × | |
| rs372777776 | ANGPTL2 | × | √ | × | | rs8033360 | CHSY1 | √ | √ | × | | rs2271979 | CEP83 | √ | × | × | Pulse press | rs3741244 | ASRGL1 | √ | × | |
| rs75586617 | ANGPTL2 | × | √ | × | | rs11856403 | CHSY1 | √ | √ | × | | rs11107523 | CEP83 | √ | × | × | | rs9553424 | ATP12A | × | √ | |
| rs999092 | ANGPTL2 | × | √ | × | | rs11639408 | CHSY1 | √ | √ | × | | rs11634566 | CHD2 | √ | × | × | | rs689300 | ATPAF1 | √ | × | |
| rs35688381 | ANGPTL2 | × | √ | × | | rs12324414 | CHSY1 | √ | √ | × | | rs3736619 | CHD2 | √ | × | × | | rs582269 | ATPAF1 | √ | × | |
| rs12684750 | ANGPTL2 | × | √ | × | | rs75228678 | CHSY1 | √ | √ | × | | rs3784785 | CHD2 | √ | × | × | | rs298722 | AUH | √ | × | |
| rs77370425 | ANGPTL2 | × | √ | × | | rs12324545 | CHSY1 | √ | √ | × | | rs10520718 | CHD2 | √ | × | × | | rs77073324 | AUH | √ | × | |
| rs117502858 | ANGPTL2 | × | √ | × | | rs8032573 | CHSY1 | √ | √ | × | | rs6497013 | CHD2 | √ | × | × | | rs10991851 | AUH | √ | × | |
| rs1766229 | ANGPTL2 | × | √ | × | | rs28445340 | CHSY1 | √ | √ | × | Vertex-wise | rs1074512 | CHD2 | √ | × | × | | rs11204306 | B9D1 | √ | × | |
| rs58411596 | ANGPTL2 | × | √ | × | | rs9672951 | CHSY1 | √ | √ | × | Cortical thic | rs1577802 | CRB1 | √ | × | × | | rs2072352 | BLM | √ | × | |
| rs1974291 | ANGPTL2 | × | √ | × | Primary open angle glauco | rs9672295 | CHSY1 | √ | √ | × | | rs10922190 | CRB1 | √ | × | × | | rs2283448 | BLM | √ | × | |
| rs565285504 | ANGPTL2 | × | √ | × | | rs11636578 | CHSY1 | √ | √ | × | Hippocampa | rs10494752 | CRB1 | √ | × | × | | rs2283449 | BLM | √ | × | |
| rs62578764 | ANGPTL2 | × | √ | × | | rs11636628 | CHSY1 | √ | √ | × | | rs10801600 | CRB1 | √ | × | × | | rs2380165 | BLM | √ | × | |
| rs374259456 | ANGPTL2 | × | √ | × | | rs3785774 | EPN2 | √ | √ | × | | rs59606758 | CYTH1 | √ | × | × | | rs387833 | BLM | √ | × | |
| rs138269816 | ANGPTL2 | × | √ | × | | rs11621316 | JAG2 | √ | √ | × | | rs11557154 | DCAF12 | √ | × | × | Neutrophil p | rs12592875 | BLM | √ | × | |
| rs116958664 | ANGPTL2 | × | √ | × | | rs10236959 | NRF1 | √ | √ | × | | rs60279027 | DCAF12 | √ | × | × | | rs401549 | BLM | √ | × | Height |
| rs779182915 | ANGPTL2 | × | √ | × | | rs10139450 | NUDT14 | √ | √ | × | | rs10971920 | DCAF12 | √ | × | × | | rs2239291 | BLM | √ | × | |
| rs12686113 | ANGPTL2 | × | √ | × | | rs4422300 | SYNPR | √ | √ | × | | rs9372863 | ECHDC1 | √ | × | × | | rs76444394 | CACNA2D1 | √ | × | |
| rs34521337 | ANGPTL2 | × | √ | × | | rs6806142 | SYNPR | √ | √ | × | | rs761842 | ECHDC1 | √ | × | × | | rs6740879 | CCDC138 | √ | × | |
| rs62578765 | ANGPTL2 | × | √ | × | | rs4768544 | TMEM117 | √ | √ | × | | rs9401948 | ECHDC1 | √ | × | × | | rs1724184 | CCDC138 | √ | × | Birth weight |
| rs193284990 | ANGPTL2 | × | √ | × | | rs11182327 | TMEM117 | √ | √ | × | | rs6936621 | ECHDC1 | √ | × | × | | rs1724195 | CCDC138 | √ | × | |
| rs185467118 | ANGPTL2 | × | √ | × | | rs10880586 | TMEM117 | √ | √ | × | | rs9375500 | ECHDC1 | √ | × | × | | rs4387742 | CCDC138 | √ | × | |
| rs1214562 | ANKRD22 | √ | × | × | | rs10880587 | TMEM117 | √ | √ | × | | rs4141423 | ECHDC1 | √ | × | × | | rs12478500 | CCDC138 | √ | × | |
| rs4934425 | ANKRD22 | √ | × | × | Height | rs10880588 | TMEM117 | √ | √ | × | | rs9372865 | ECHDC1 | √ | × | × | | rs1589911 | CCDC138 | √ | × | |
| rs77411926 | ANO4 | √ | × | × | | rs10785470 | TMEM117 | √ | √ | × | | rs75258130 | ECHDC1 | √ | × | × | | rs975597 | CCDC138 | √ | × | |
| rs34741810 | ANO4 | √ | × | × | | rs7964465 | TMEM117 | √ | √ | × | | rs75973322 | ECHDC1 | √ | × | × | | rs7564164 | CCDC138 | √ | × | |
| rs71464253 | ANO4 | √ | × | × | | rs12369569 | TMEM117 | √ | √ | × | | rs9372866 | ECHDC1 | √ | × | × | | rs73952523 | CCDC138 | √ | × | |
| rs2654991 | ANO4 | √ | × | × | | rs10880589 | TMEM117 | √ | √ | × | | rs9375502 | ECHDC1 | √ | × | × | | rs73952524 | CCDC138 | √ | × | |
| rs589657 | ANO4 | √ | × | × | | rs10785481 | TMEM117 | √ | √ | × | | rs9388562 | ECHDC1 | √ | × | × | | rs10865022 | CCDC138 | √ | × | |
| rs649094 | ANO4 | √ | × | × | | rs3925073 | TMEM117 | √ | √ | × | | rs10865025 | EDAR | √ | × | × | | rs57043792 | CCDC138 | √ | × | |
| rs677871 | ANO4 | √ | × | × | | rs6582490 | TMEM117 | √ | √ | × | | rs209490 | ELOVL5 | √ | × | × | | rs73952526 | CCDC138 | √ | × | |
| rs349661 | ANO4 | √ | × | × | | rs11182339 | TMEM117 | √ | √ | × | | rs11600929 | EXT2 | √ | × | × | | rs13009852 | CCDC138 | √ | × | |
| rs349662 | ANO4 | √ | × | × | | rs4924986 | B9D1 | × | √ | × | | rs11600984 | EXT2 | √ | × | × | | rs4676209 | CCDC138 | √ | × | |
| rs12792259 | ANO5 | √ | × | × | | rs12371795 | SOX5 | √ | × | × | | rs10838240 | EXT2 | √ | × | × | | rs12464296 | CCDC138 | √ | × | |
| rs6483831 | ANO5 | √ | × | × | | rs12366963 | SOX5 | √ | × | × | | rs7118541 | EXT2 | √ | × | × | | rs11681025 | CCDC138 | √ | × | |
| rs4589299 | ANO5 | √ | × | × | | rs1355001 | SOX5 | √ | × | × | | rs2031693 | FAM102A | √ | × | × | | rs6727368 | CCDC138 | √ | × | |
| rs4485117 | ANO5 | √ | × | × | | rs11047450 | SOX5 | √ | × | × | | rs11789803 | FAM102A | √ | × | × | | rs7600513 | CCDC138 | √ | × | |
| rs4627075 | ANO5 | √ | × | × | | rs11047452 | SOX5 | √ | × | × | | rs4837212 | FAM102A | √ | × | × | | rs2031262 | CENPP | √ | × | |
| rs4293118 | ANO5 | √ | × | × | | rs34119528 | TBC1D1 | √ | × | × | | rs7849083 | FAM102A | √ | × | × | | rs10761156 | CENPP | √ | × | |
| rs10766918 | ANO5 | √ | × | × | | rs12645411 | TBC1D1 | √ | × | × | | rs6478804 | FAM102A | √ | × | × | | rs75430031 | CEP83 | √ | × | |
| rs4565894 | ANO5 | √ | × | × | | rs9930504 | ZFHX3 | × | √ | × | | rs7037993 | FAM102A | √ | × | × | | rs79671686 | CEP83 | √ | × | |
| rs6416032 | ANO5 | √ | × | × | | rs75739666 | ACAP2 | √ | × | × | | rs77741711 | FBXW7 | × | √ | × | | rs2271979 | CEP83 | √ | × | Pulse pressure |
| rs7102678 | ANO5 | √ | × | × | | rs41266999 | ACAP2 | √ | × | × | | rs14254953 | FBXW7 | √ | × | × | | rs11107523 | CEP83 | √ | × | |
| rs4620717 | ANO5 | √ | × | × | | rs77075906 | ACAP2 | √ | × | × | | rs1175931 | IARS | √ | × | × | | rs12592212 | CHD2 | √ | × | |
| rs4312063 | ANO5 | √ | × | × | | rs2447249 | AGBL1 | √ | × | × | | rs471478 | IARS | √ | × | × | COL1A1/OI | rs3736619 | CHD2 | √ | × | |
| rs7935686 | ANO5 | √ | × | × | | rs7172992 | AGBL1 | √ | × | × | | rs504708 | IARS | √ | × | × | | rs3784785 | CHD2 | √ | × | |
| rs4509753 | ANO5 | √ | × | × | | rs79196031 | ANGPTL2 | × | √ | × | | rs1152757 | IARS | √ | × | × | | rs10520718 | CHD2 | √ | × | |
| rs4297463 | ANO5 | √ | × | × | | rs2789511 | ANGPTL2 | × | √ | × | | rs3996311 | IARS | √ | × | × | | rs6497013 | CHD2 | √ | × | |
| rs76088483 | ANO5 | √ | × | × | | rs11791203 | ANGPTL2 | × | √ | × | | rs10842374 | LINC00477 | √ | × | × | | rs752092 | CHSY1 | √ | × | Central corneal thickness |
| rs4675823 | ANO7 | √ | × | × | | rs1281159 | ANGPTL2 | × | √ | × | | rs6487400 | LINC00477 | √ | × | × | | rs12026252 | CRB1 | √ | × | |
| rs225299 | AP2B1 | √ | × | × | | rs14143051 | ANGPTL2 | × | √ | × | | rs11241458 | LOC101927 | √ | × | × | | rs1577802 | CRB1 | √ | × | |
| rs225293 | AP2B1 | √ | × | × | | rs18401619 | ANGPTL2 | × | √ | × | | rs7703431 | LOC101927 | √ | × | × | | rs10922190 | CRB1 | √ | × | |
| rs117652210 | AP2B1 | √ | × | × | | rs11793399 | ANGPTL2 | × | √ | × | | rs7708440 | LOC101927 | √ | × | × | | rs10494752 | CRB1 | √ | × | |
| rs16971241 | AP2B1 | √ | × | × | Mean platelet volume | rs60313786 | ANGPTL2 | × | √ | × | | rs6595126 | LOC101927 | × | √ | × | | rs10801600 | CRB1 | √ | × | |
| rs997227519 | AP2B1 | √ | × | × | | rs13298677 | ANGPTL2 | × | √ | × | Height | rs9401672 | LOC101927 | √ | × | × | | rs664767 | CRB1 | √ | × | |
| rs34619890 | AP2B1 | √ | × | × | | rs11795066 | ANGPTL2 | × | √ | × | Intraocular p | rs72613284 | LOC101927 | √ | × | × | | rs1670167 | CTNNA3 | √ | × | Educational attainment |
| rs79231460 | AP2B1 | √ | × | × | | rs14529878 | ANGPTL2 | × | √ | × | | rs4412217 | LOC101927 | √ | × | × | | rs12451377 | CYTH1 | √ | × | |
| rs16971437 | AP2B1 | √ | × | × | | rs80116254 | ANGPTL2 | × | √ | × | | rs56131876 | LRSAM1 | √ | × | × | | rs59606758 | CYTH1 | √ | × | |
| rs140575516 | AP2B1 | √ | × | × | | rs79881767 | ANGPTL2 | × | √ | × | | rs2896706 | LUZP2 | √ | × | × | | rs10758240 | DCAF12 | √ | × | |
| rs226441 | AP2B1 | √ | × | × | | rs2787594 | ANGPTL2 | × | √ | × | Hair color | rs12273206 | LUZP2 | √ | × | × | | rs11557154 | DCAF12 | √ | × | Neutrophil percentage of white cells |
| rs146989027 | AP2B1 | × | √ | × | | rs11792699 | ANGPTL2 | × | √ | × | | rs1509605 | LUZP2 | √ | × | × | Depressive | rs10971920 | DCAF12 | √ | × | |
| rs72829925 | AP2B1 | × | √ | × | | rs1281157 | ANGPTL2 | × | √ | × | | rs12289378 | LUZP2 | √ | × | × | | rs10972169 | DCTN3 | √ | × | |
| rs61515677 | AP2B1 | × | √ | × | | rs37277777 | ANGPTL2 | × | √ | × | | rs11529784 | LUZP2 | √ | × | × | | rs2279522 | DPYSL4 | √ | × | |
| rs226428 | AP2B1 | × | √ | × | | rs75586617 | ANGPTL2 | × | √ | × | | rs9943629 | LUZP2 | √ | × | × | | rs9372863 | ECHDC1 | √ | × | |
| rs226427 | AP2B1 | × | √ | × | | rs999092 | ANGPTL2 | × | √ | × | | rs62256972 | MIR548AC | √ | × | × | | rs761842 | ECHDC1 | √ | × | |
| rs226426 | AP2B1 | × | √ | × | | rs35688381 | ANGPTL2 | × | √ | × | | rs62256975 | MIR548AC | √ | × | × | | rs9401948 | ECHDC1 | √ | × | |
| rs141453915 | AP2B1 | × | √ | × | | rs62578764 | ANGPTL2 | × | √ | × | | rs79562849 | MIR548AC | √ | × | × | | rs6936621 | ECHDC1 | √ | × | |
| rs2240903 | AP2B1 | × | √ | × | | rs37425945 | ANGPTL2 | × | √ | × | | rs62256976 | MIR548AC | √ | × | × | | rs9375500 | ECHDC1 | √ | × | |
| rs77247187 | AP2B1 | × | √ | × | | rs11695866 | ANGPTL2 | × | √ | × | | rs62256977 | MIR548AC | √ | × | × | | rs4141423 | ECHDC1 | √ | × | |
| rs17638867 | AP2B1 | × | √ | × | Cognitive ability, years of e | rs62578765 | ANGPTL2 | × | √ | × | | rs11535912 | MIR548AC | √ | × | × | | rs9372865 | ECHDC1 | √ | × | |
| rs740913 | AP2B1 | × | √ | × | | rs18546711 | ANGPTL2 | × | √ | × | | rs6774454 | MIR548AC | √ | × | × | | rs75258130 | ECHDC1 | √ | × | |
| rs80149721 | APBB1IP | √ | × | × | | rs12792259 | ANO5 | √ | × | × | | rs77832017 | MIR548AC | √ | × | × | | rs75973322 | ECHDC1 | √ | × | |
| rs35408871 | APH1B | √ | × | × | | rs6483831 | ANO5 | √ | × | × | | rs60137984 | MIR548AC | √ | × | × | | rs9372866 | ECHDC1 | √ | × | |
| rs56256221 | ARG2 | √ | × | × | | rs4485117 | ANO5 | √ | × | × | | rs9395899 | MLIP | √ | × | × | | rs9375502 | ECHDC1 | √ | × | |
| rs7152326 | ARG2 | √ | × | × | | rs4627075 | ANO5 | √ | × | × | | rs35796001 | MSR1 | √ | × | × | | rs9388562 | ECHDC1 | √ | × | |
| rs7915768 | ARHGAP21 | √ | × | × | | rs4293118 | ANO5 | √ | × | × | | rs35281051 | MSR1 | √ | × | × | | rs72961085 | ECHDC1 | √ | × | |
| rs2886851 | ARHGAP21 | √ | × | × | | rs10766918 | ANO5 | √ | × | × | | rs918 | MSR1 | √ | × | × | | rs76880977 | ECHDC1 | √ | × | |
| rs7474901 | ARHGAP21 | √ | × | × | | rs4565894 | ANO5 | √ | × | × | | rs13306544 | MSR1 | √ | × | × | | rs3756994 | ECHDC1 | √ | × | |
| rs371812734 | ARHGAP21 | √ | × | × | | rs11765221 | AP2B1 | √ | × | × | | rs12678016 | MSR1 | √ | × | × | | rs10865025 | EDAR | √ | × | |
| rs7090524 | ARHGAP21 | √ | × | × | | rs16971241 | AP2B1 | √ | × | × | Mean platel | rs12679486 | MSR1 | √ | × | × | | rs74763346 | F13B | √ | × | |
| rs7075091 | ARHGAP21 | √ | × | × | | rs35036895 | ARSJ | √ | × | × | | rs34612904 | MSR1 | √ | × | × | | rs59944943 | FAM114A1 | √ | × | |
| rs10828682 | ARHGAP21 | √ | × | × | | rs7677674 | ARSJ | √ | × | × | Height | rs7015682 | MSR1 | √ | × | × | | rs56229790 | FAM114A1 | √ | × | |
| rs12220759 | ARHGAP21 | √ | × | × | | rs77526635 | ASRGL1 | √ | × | × | | rs6995004 | MSR1 | √ | × | × | | rs80227559 | FAM114A1 | √ | × | |
| rs78542723 | ARHGAP21 | √ | × | × | | rs3741244 | ASRGL1 | √ | × | × | | rs6995009 | MSR1 | √ | × | × | | rs11725839 | FAM114A1 | √ | × | |
| rs28602629 | ARSJ | √ | × | × | | rs298722 | AUH | √ | × | × | | rs6982066 | MSR1 | √ | × | × | | rs73236661 | FAM114A1 | √ | × | |
| rs35036895 | ARSJ | √ | × | × | | rs77073324 | AUH | √ | × | × | | rs6982504 | MSR1 | √ | × | × | | rs11096964 | FAM114A1 | √ | × | |
| rs7664471 | ARSJ | √ | × | × | | rs2048230 | B9D1 | × | √ | × | | rs78016136 | MSR1 | √ | × | × | | rs11555334 | FAM114A1 | √ | × | |
| rs9917901 | ARSJ | √ | × | × | | rs60606605 | B9D1 | × | √ | × | | rs76903738 | MSR1 | √ | × | × | | rs11938589 | FAM114A1 | √ | × | |
| rs11098209 | ARSJ | √ | × | × | | rs2072352 | BLM | √ | × | × | | rs76841569 | MSR1 | √ | × | × | | rs11944231 | FAM114A1 | √ | × | |
| rs4631117 | ARSJ | √ | × | × | | rs2283448 | BLM | √ | × | × | | rs12676172 | MSR1 | √ | × | × | | rs12676834 | FAM49B | √ | × | |
| rs4410591 | ARSJ | √ | × | × | | rs2283449 | BLM | √ | × | × | | rs12676591 | MSR1 | √ | × | × | | rs9684643 | FBXW7 | √ | × | |
| rs13111878 | ARSJ | √ | × | × | | rs387833 | BLM | √ | × | × | | rs12678046 | MSR1 | √ | × | × | | rs75026869 | FBXW7 | √ | × | |
| rs10008482 | ARSJ | √ | × | × | | rs36967092 | BLM | √ | × | × | | rs62491230 | MSR1 | √ | × | × | | rs77741711 | FBXW7 | √ | × | |
| rs74975228 | ARSJ | √ | × | × | | rs2270132 | BLM | √ | × | × | | rs753947 | NRF1 | √ | × | × | | rs77481157 | FBXW7 | √ | × | |
| rs7677674 | ARSJ | √ | × | × | Height | rs389480 | BLM | √ | × | × | | rs10245818 | NRF1 | √ | × | × | | rs11742062 | FBXW7 | √ | × | |
| rs11098219 | ARSJ | √ | × | × | Height | rs12592875 | BLM | √ | × | × | | rs28689822 | NRF1 | √ | × | × | | rs8051879 | GPR97 | × | √ | |
| rs12501970 | ARSJ | √ | × | × | | rs401549 | BLM | √ | × | × | Height | rs78086437 | NRF1 | √ | × | × | | rs1175931 | IARS | √ | × | |
| rs6827199 | ARSJ | √ | × | × | Height | rs2239291 | BLM | √ | × | × | | rs12581006 | PCED1B | √ | × | × | | rs471478 | IARS | √ | × | COL1A1/OMD protein level ratio |
| rs12498542 | ARSJ | √ | × | × | | rs1882847 | BRF1 | × | × | √ | | rs10881052 | PCED1B | √ | × | × | | rs504708 | IARS | √ | × | |
| rs76099159 | ASPM | √ | × | × | | rs57581057 | BTBD16 | √ | × | × | | rs1525452 | PCED1B | √ | × | × | | rs1152757 | IARS | √ | × | |
| rs3174352 | ASPN | √ | × | × | Urate levels | rs732074 | C9orf78 | √ | × | × | | rs36124720 | PER2 | √ | × | × | | rs3996311 | IARS | √ | × | |
| rs10437726 | ASRGL1 | √ | × | × | | rs55962905 | C9orf78 | √ | × | × | | rs79644971 | PRSS3 | √ | × | × | | rs10992290 | IARS | √ | × | |
| rs77526635 | ASRGL1 | √ | × | × | | rs7032945 | C9orf78 | √ | × | × | | rs1724203 | RANBP2 | √ | × | × | | rs14570431 | IKZF1 | √ | × | |
| rs3741244 | ASRGL1 | √ | × | × | | rs3818553 | C9orf78 | √ | × | × | | rs3782445 | RBM19 | √ | × | × | | rs10850132 | IQCD | √ | × | |
| rs150476377 | ATG4B | √ | × | × | | rs41278732 | C9orf78 | √ | × | × | | rs9372862 | RNF146 | √ | × | × | | rs4251460 | IRAK4 | × | √ | |
| rs12475263 | ATOH8 | √ | × | × | | rs13290140 | C9orf78 | √ | × | × | | rs9388559 | RNF146 | √ | × | × | | rs4251465 | IRAK4 | × | √ | |
| rs139677844 | ATOH8 | √ | × | × | | rs11631047 | CA12 | × | √ | × | | rs9321072 | RNF146 | √ | × | × | | rs4238087 | IRAK4 | × | √ | |
| rs9553424 | ATP12A | √ | × | × | | rs11807511 | CA12 | × | √ | × | | rs2273013 | SNIP1 | √ | × | × | | rs4251466 | IRAK4 | × | √ | |
| rs146386177 | ATP1B3 | √ | × | × | | rs6835747 | CAMK2D | √ | × | × | | rs7138457 | SOX5 | √ | × | × | | rs1461567 | IRAK4 | × | √ | |
| rs689300 | ATPAF1 | √ | × | × | | rs17593086 | CAMK2D | √ | × | × | | rs13146675 | SPOCK3 | √ | × | × | | rs11809731 | IRAK4 | × | √ | |
| rs582269 | ATPAF1 | √ | × | × | | rs6740879 | CCDC138 | √ | × | × | | rs7691894 | SPOCK3 | √ | × | × | | rs3794262 | IRAK4 | × | √ | Vaginal microbiome MetaCyc pathway (PRPP-PWY\|superpathway of histidine, purine, and pyrimidine biosynthesis) |
| rs2218190 | ATPAF1 | √ | × | × | | rs1724184 | CCDC138 | √ | × | × | Birth weight | rs28469298 | SPOCK3 | √ | × | × | | rs4251473 | IRAK4 | × | √ | |
| rs56049491 | AUH | √ | × | × | Total bilirubin levels | rs1724195 | CCDC138 | √ | × | × | | rs2219234 | SPOCK3 | √ | × | × | | rs18134893 | IRAK4 | × | √ | |
| rs190020242 | AUH | √ | × | × | | rs4387742 | CCDC138 | √ | × | × | | rs3828018 | SPOCK3 | √ | × | × | | rs55973711 | IRAK4 | × | √ | |
| rs773518 | AUH | √ | × | × | | rs12478500 | CCDC138 | √ | × | × | | rs3790358 | SPOCK3 | √ | × | × | | rs4251475 | IRAK4 | × | √ | |
| rs12686257 | AUH | √ | × | × | | rs975597 | CCDC138 | √ | × | × | | rs28699632 | SPOCK3 | √ | × | × | | rs4251481 | IRAK4 | × | √ | |
| rs10991840 | AUH | √ | × | × | | rs56936346 | CCDC138 | √ | × | × | | rs7697656 | SPOCK3 | √ | × | × | | rs76136881 | IRAK4 | × | √ | |
| rs298722 | AUH | √ | × | × | | rs7564164 | CCDC138 | √ | × | × | | rs7693595 | SPOCK3 | √ | × | × | | rs11182259 | IRAK4 | × | √ | |
| rs77073324 | AUH | √ | × | × | | rs10865022 | CCDC138 | √ | × | × | | rs17645162 | SPOCK3 | √ | × | × | | rs11182261 | IRAK4 | × | √ | |
| rs10991851 | AUH | √ | × | × | | rs73952526 | CCDC138 | √ | × | × | | rs10517901 | SPOCK3 | √ | × | × | | rs11699938 | IRAK4 | × | √ | |
| rs1757648 | AUH | √ | × | × | | rs13009852 | CCDC138 | √ | × | × | | rs17645192 | SPOCK3 | √ | × | × | | rs14057814 | IRAK4 | × | √ | |
| rs75120664 | AUH | √ | × | × | | rs4676209 | CCDC138 | √ | × | × | | rs2305593 | SPOCK3 | √ | × | × | | rs79154645 | IRAK4 | × | √ | |
| rs11204306 | B9D1 | × | × | √ | | rs12464296 | CCDC138 | √ | × | × | | rs2389414 | TBC1D32 | √ | × | × | | rs4251513 | IRAK4 | × | √ | |
| rs2048230 | B9D1 | × | × | √ | | rs11681025 | CCDC138 | √ | × | × | | rs9320800 | TBC1D32 | √ | × | × | | rs5020835 | ITPKB | √ | × | |
| rs60606605 | B9D1 | × | × | √ | | rs6727368 | CCDC138 | √ | × | × | | rs7908570 | TET1 | √ | × | × | | rs4784010 | KIFC3 | √ | × | |
| rs60166680 | B9D1 | × | × | √ | | rs7600513 | CCDC138 | √ | × | × | | rs11199028 | TIAL1 | √ | × | × | | rs77414536 | KIFC3 | √ | × | |
| rs965779943 | BCL9L | × | √ | × | | rs1073895 | CCDC138 | √ | × | × | | rs11199030 | TIAL1 | √ | × | × | | rs79949875 | KIFC3 | √ | × | |
| rs976141031 | BCL9L | × | √ | × | | rs17183835 | CCSER1 | √ | × | × | | rs12107025 | TP63 | √ | × | × | | rs337616 | KLF3 | √ | × | |
| rs11217081 | BCL9L | × | √ | × | | rs76176992 | CDCA4 | √ | × | × | | rs61943604 | TPCN1 | √ | × | × | | rs10842374 | LINC00477 | √ | × | |
| rs182557494 | BCL9L | × | √ | × | | rs2031262 | CENPP | √ | × | × | | rs7135901 | TPCN1 | √ | × | × | | rs6487399 | LINC00477 | √ | × | |
| rs11217083 | BCL9L | × | √ | × | | rs10761156 | CENPP | √ | × | × | | rs7300258 | TPCN1 | √ | × | × | | rs6487400 | LINC00477 | √ | × | |
| rs187198056 | BICD2 | √ | × | × | | rs2025389 | CENPP | √ | × | × | | rs7314153 | TPCN1 | √ | × | × | | rs7299236 | LINC00477 | √ | × | |
| rs182153690 | BICD2 | √ | × | × | | rs2271979 | CEP83 | √ | × | × | Pulse press | rs2303618 | TPCN1 | √ | × | × | | rs6421879 | LOC101927 | √ | × | |
| rs117856757 | BICD2 | √ | × | × | | rs11107523 | CEP83 | √ | × | × | | rs2303617 | TPCN1 | √ | × | × | | rs6894176 | LOC101927 | √ | × | |
| rs2072352 | BLM | √ | × | × | | rs12592212 | CHD2 | √ | × | × | | rs7955036 | TPCN1 | √ | × | × | | rs6875260 | LOC101927 | √ | × | |
| rs2283448 | BLM | √ | × | × | | rs11634566 | CHD2 | √ | × | × | | rs58301661 | TRDN | √ | × | × | | rs6894916 | LOC101927 | √ | × | |
| rs2283449 | BLM | √ | × | × | | rs3736619 | CHD2 | √ | × | × | | rs9388248 | TRDN | √ | × | × | | rs7721374 | LOC101927 | √ | × | |
| rs2380165 | BLM | √ | × | × | | rs3784785 | CHD2 | √ | × | × | | rs9385300 | TRDN | √ | × | × | | rs7711344 | LOC101927 | √ | × | |
| rs387833 | BLM | √ | × | × | | rs62023149 | CHD2 | √ | × | × | | rs4460222 | TRDN | √ | × | × | | rs11241458 | LOC101927 | √ | × | |
| rs369670923 | BLM | √ | × | × | | rs10520718 | CHD2 | √ | × | × | | rs1830816 | TRIM44 | √ | × | × | | rs77476803 | LOC101927 | √ | × | |
| rs2270132 | BLM | √ | × | × | | rs61334597 | CHD2 | √ | × | × | | rs1969800 | TRIM44 | √ | × | × | | rs75594839 | LOC101927 | √ | × | |
| rs389480 | BLM | √ | × | × | | rs2053359 | CHD2 | √ | × | × | | rs61881279 | TRIM44 | √ | × | × | | rs77950503 | LOC101927 | √ | × | |
| rs12592875 | BLM | √ | × | × | | rs6497013 | CHD2 | √ | × | × | | rs2970329 | TRIM44 | √ | × | × | | rs7703431 | LOC101927 | √ | × | |
| rs401549 | BLM | √ | × | × | Height | rs12438635 | CHD2 | √ | × | × | | rs68063352 | TRIM44 | √ | × | × | | rs80240958 | LOC101927 | √ | × | |
| rs2239291 | BLM | √ | × | × | | rs1074512 | CHD2 | √ | × | × | | rs62555936 | UBAP2 | √ | × | × | | rs79650909 | LOC101927 | √ | × | |
| rs1882847 | BRF1 | × | × | √ | | rs7095732 | CPXM2 | √ | × | × | | rs16935299 | UBAP2 | √ | × | × | | rs11707061 | LOC101927 | √ | × | |
| rs57581057 | BTBD16 | √ | × | × | | rs12026252 | CRB1 | √ | × | × | | rs3739690 | UBAP2 | √ | × | × | | rs74593842 | LOC101927 | √ | × | |
| rs7072288 | BTBD16 | √ | × | × | | rs1577802 | CRB1 | √ | × | × | | rs62555940 | UBAP2 | √ | × | × | | rs7444011 | LOC101927 | √ | × | |
| rs11596483 | BTBD16 | √ | × | × | Gut microbiome compositio | rs10922190 | CRB1 | √ | × | × | | rs307697 | UBAP2 | √ | × | × | | rs7708440 | LOC101927 | √ | × | |
| rs6585809 | BTBD16 | √ | × | × | | rs10494752 | CRB1 | √ | × | × | | rs12553803 | UBAP2 | √ | × | × | | rs78495315 | LOC101927 | √ | × | |
| rs11592862 | BTBD16 | √ | × | × | | rs10801600 | CRB1 | √ | × | × | | rs10971837 | UBAP2 | √ | × | × | | rs6595126 | LOC101927 | √ | × | |
| rs7907130 | BTBD16 | √ | × | × | | rs1670167 | CTNNA3 | √ | × | × | Educational | rs10758235 | UBAP2 | √ | × | × | | rs11950959 | LOC101927 | √ | × | |
| rs80345398 | BTBD16 | √ | × | × | | rs11657403 | CYTH1 | √ | × | × | | rs56404908 | UBAP2 | √ | × | × | | rs7727130 | LOC101927 | √ | × | |
| rs2849179 | BUD13 | √ | × | × | | rs12451377 | CYTH1 | √ | × | × | | rs7044541 | UBAP2 | √ | × | × | | rs6867291 | LOC101927 | √ | × | |
| rs2282425 | C1orf95 | √ | × | × | | rs11656741 | CYTH1 | √ | × | × | | rs10122409 | UBAP2 | √ | × | × | | rs77788081 | LOC101927 | √ | × | |
| rs1935307 | C1orf95 | √ | × | × | | rs11656742 | CYTH1 | √ | × | × | | rs10814059 | UBAP2 | √ | × | × | | rs77124635 | LOC101927 | √ | × | |
| rs732074 | C9orf78 | √ | × | × | | rs59606758 | CYTH1 | √ | × | × | | rs7029112 | UBAP2 | √ | × | × | | rs78400388 | LOC101927 | √ | × | |
| rs55962905 | C9orf78 | √ | × | × | | rs922838 | CYTH1 | √ | × | × | | rs10758236 | UBAP2 | √ | × | × | | rs74926979 | LOC101927 | √ | × | |
| rs7032945 | C9orf78 | √ | × | × | | rs11557154 | DCAF12 | √ | × | × | Neutrophil p | rs13300356 | UBAP2 | √ | × | × | | rs75622096 | LOC101927 | √ | × | |
| rs7047271 | C9orf78 | √ | × | × | | rs60279027 | DCAF12 | √ | × | × | | rs281354 | ULK2 | √ | × | × | | rs12719243 | LOC101927 | √ | × | |
| rs3818553 | C9orf78 | √ | × | × | | rs10971920 | DCAF12 | √ | × | × | | rs281346 | ULK2 | √ | × | × | | rs6873260 | LOC101927 | √ | × | |
| rs41278732 | C9orf78 | √ | × | × | | rs10972169 | DCTN3 | √ | × | × | | rs11265860 | UNQ6494 | √ | × | × | | rs78195006 | LOC101927 | √ | × | |
| rs35632463 | C9orf78 | √ | × | × | | rs13953395 | DEF8 | × | √ | × | | rs3744793 | USP36 | √ | × | × | | rs7715481 | LOC101927 | √ | × | |
| rs13290140 | C9orf78 | √ | × | × | | rs14419594 | DEF8 | × | √ | × | | rs62290320 | XXYLT1 | √ | × | × | | rs3923756 | LOC101927 | √ | × | |
| rs72748939 | CA12 | × | √ | × | | rs9372863 | ECHDC1 | √ | × | × | | rs62122411 | ZNF787 | √ | × | × | | rs7701418 | LOC101927 | √ | × | |
| rs55656642 | CA12 | × | √ | × | | rs761842 | ECHDC1 | √ | × | × | | rs2437788 | AGBL1 | √ | × | × | | rs10058844 | LOC101927 | √ | × | |
| rs34437213 | CA12 | × | √ | × | | rs9401948 | ECHDC1 | √ | × | × | | rs16977988 | AGBL1 | √ | × | × | | rs6883163 | LOC101927 | √ | × | |
| rs2019377 | CA12 | × | √ | × | | rs6936621 | ECHDC1 | √ | × | × | | rs16977990 | AGBL1 | √ | × | × | | rs4440399 | LOC101927 | √ | × | |
| rs2019976 | CA12 | × | √ | × | | rs9375500 | ECHDC1 | √ | × | × | | rs2881581 | AGBL1 | √ | × | × | | rs9327075 | LOC101927 | √ | × | |
| rs16946922 | CA12 | × | √ | × | | rs4141423 | ECHDC1 | √ | × | × | | rs4887244 | AGBL1 | √ | × | × | | rs6595142 | LOC101927 | √ | × | |
| rs6494402 | CA12 | × | √ | × | | rs9372865 | ECHDC1 | √ | × | × | | rs79545780 | AGBL1 | √ | × | × | | rs6595143 | LOC101927 | √ | × | |
| rs143867050 | CA12 | × | √ | × | | rs75258130 | ECHDC1 | √ | × | × | | rs10768994 | ALKBH3 | √ | × | × | Body mass | rs13360367 | LOC101927 | √ | × | |
| rs11631047 | CA12 | × | √ | × | | rs75973322 | ECHDC1 | √ | × | × | | rs5142 | APOC3 | √ | × | × | | rs7708133 | LOC101927 | √ | × | |
| rs118075114 | CA12 | × | √ | × | | rs9372866 | ECHDC1 | √ | × | × | | rs7908377 | ARHGAP21 | √ | × | × | | rs7708340 | LOC101927 | √ | × | |

| | | | | | |
|---|---|---|---|---|---|
| rs17264844 | CACNA2D1 | √ | × | × | |
| rs2299177 | CACNA2D1 | √ | × | × | |
| rs76065766 | CACNA2D1 | √ | × | × | |
| rs2299178 | CACNA2D1 | √ | × | × | |
| rs12673202 | CACNA2D1 | √ | × | × | |
| rs2299180 | CACNA2D1 | √ | × | × | |
| rs80322429 | CACNA2D1 | √ | × | × | |
| rs76160570 | CACNA2D1 | √ | × | × | |
| rs917188 | CACNA2D1 | √ | × | × | |
| rs76444394 | CACNA2D1 | √ | × | × | |
| rs17151755 | CAMK1D | × | × | √ | |
| rs1525000 | CAMK2D | √ | × | × | |
| rs62314976 | CAMK2D | √ | × | × | JT interval |
| rs13106836 | CAMK2D | √ | × | × | |
| rs12511954 | CAMK2D | √ | × | × | |
| rs6835747 | CAMK2D | √ | × | × | |
| rs6842886 | CAMK2D | √ | × | × | |
| rs12507550 | CAMK2D | √ | × | × | |
| rs12503836 | CAMK2D | √ | × | × | |
| rs11936225 | CAMK2D | √ | × | × | |
| rs17591997 | CAMK2D | √ | × | × | |
| rs72678758 | CAMK2D | √ | × | × | |
| rs72678759 | CAMK2D | √ | × | × | |
| rs10033037 | CAMK2D | √ | × | × | |
| rs3764805 | CAMK2D | √ | × | × | |
| rs72617769 | CAMK2D | √ | × | × | |
| rs77541675 | CAMK2D | √ | × | × | |
| rs72617770 | CAMK2D | √ | × | × | |
| rs72617771 | CAMK2D | √ | × | × | |
| rs35507223 | CAMK2D | √ | × | × | |
| rs72617772 | CAMK2D | √ | × | × | |
| rs2189363 | CAMK2D | √ | × | × | |
| rs17593086 | CAMK2D | √ | × | × | |
| rs117368427 | CAMK2D | √ | × | × | |
| rs146306243 | CCAR1 | √ | × | × | |
| rs117850434 | CCAR1 | √ | × | × | |
| rs12146315 | CCAR1 | √ | × | × | |
| rs3912104 | CCDC13 | √ | × | × | |
| rs11718476 | CCDC13 | √ | × | × | |
| rs339731 | CCDC13 | √ | × | × | |
| rs9830553 | CCDC13 | √ | × | × | |
| rs12633707 | CCDC13 | √ | × | × | |
| rs12491179 | CCDC13 | √ | × | × | |
| rs189907539 | CCDC135 | √ | × | × | |
| rs6740879 | CCDC138 | √ | × | × | |
| rs1724184 | CCDC138 | √ | × | × | Birth weight |
| rs1724195 | CCDC138 | √ | × | × | |
| rs4387742 | CCDC138 | √ | × | × | |
| rs12478500 | CCDC138 | √ | × | × | |
| rs1589911 | CCDC138 | √ | × | × | |
| rs975597 | CCDC138 | √ | × | × | |
| rs56936346 | CCDC138 | √ | × | × | |
| rs7564164 | CCDC138 | √ | × | × | |
| rs73952523 | CCDC138 | √ | × | × | |
| rs73952524 | CCDC138 | √ | × | × | |
| rs10865022 | CCDC138 | √ | × | × | |
| rs57043792 | CCDC138 | √ | × | × | |
| rs73952526 | CCDC138 | √ | × | × | |
| rs75324970 | CCDC138 | √ | × | × | |
| rs13009852 | CCDC138 | √ | × | × | |
| rs4676209 | CCDC138 | √ | × | × | |
| rs12464296 | CCDC138 | √ | × | × | |
| rs11681025 | CCDC138 | √ | × | × | |
| rs6727368 | CCDC138 | √ | × | × | |
| rs7600513 | CCDC138 | √ | × | × | |
| rs1073895 | CCDC138 | √ | × | × | |
| rs17243064 | CCSER1 | √ | × | × | |
| rs17183835 | CCSER1 | √ | × | × | |
| rs12884180 | CDCA4 | √ | × | × | |
| rs76176992 | CDCA4 | √ | × | × | |
| rs2031262 | CENPP | √ | × | × | |
| rs10761156 | CENPP | √ | × | × | |
| rs10125450 | CENPP | √ | × | × | |
| rs10820986 | CENPP | √ | × | × | |
| rs2025389 | CENPP | √ | × | × | |
| rs1973832 | CEP83 | √ | × | × | |
| rs7979054 | CEP83 | √ | × | × | |
| rs138006302 | CEP83 | √ | × | × | |
| rs75430031 | CEP83 | √ | × | × | |
| rs79671686 | CEP83 | √ | × | × | |
| rs2271979 | CEP83 | √ | × | × | Pulse pressure |
| rs11107523 | CEP83 | √ | × | × | |
| rs532965970 | CEP83 | √ | × | × | |
| rs12592212 | CHD2 | √ | × | × | |
| rs78835543 | CHD2 | √ | × | × | |
| rs11634566 | CHD2 | √ | × | × | |
| rs3736619 | CHD2 | √ | × | × | |
| rs28667191 | CHD2 | √ | × | × | |
| rs76621355 | CHD2 | √ | × | × | |
| rs3784785 | CHD2 | √ | × | × | |
| rs62023149 | CHD2 | √ | × | × | |
| rs10520718 | CHD2 | √ | × | × | |
| rs79768810 | CHD2 | √ | × | × | |
| rs61334597 | CHD2 | √ | × | × | |
| rs78964321 | CHD2 | √ | × | × | |
| rs2053359 | CHD2 | √ | × | × | |
| rs7162369 | CHD2 | √ | × | × | |
| rs6497013 | CHD2 | √ | × | × | |
| rs12438635 | CHD2 | √ | × | × | |
| rs1074512 | CHD2 | √ | × | × | |
| rs144257568 | CHD2 | √ | × | × | |
| rs148690433 | CHSY1 | × | √ | × | |
| rs11433 | CHSY1 | × | √ | × | |
| rs3784526 | CHSY1 | × | √ | × | Granulocyte percentage of |
| rs3784523 | CHSY1 | × | √ | × | |
| rs8024370 | CHSY1 | × | √ | × | |
| rs375906150 | CHSY1 | × | √ | × | |
| rs28364839 | CHSY1 | × | √ | × | Fracture of forearm |
| rs3803423 | CHSY1 | × | √ | × | |
| rs3803422 | CHSY1 | × | √ | × | |
| rs76457230 | CHSY1 | × | √ | × | |
| rs2005180 | CHSY1 | × | √ | × | |
| rs3743193 | CHSY1 | × | √ | × | |
| rs74752435 | CHSY1 | × | √ | × | |
| rs7164068 | CHSY1 | × | √ | × | |
| rs373935146 | CHSY1 | × | √ | × | |
| rs60140504 | CHSY1 | × | √ | × | |
| rs142869818 | CHSY1 | × | √ | × | |
| rs57352121 | CHSY1 | × | √ | × | |
| rs75317577 | CHSY1 | × | √ | × | |
| rs188178576 | CHSY1 | × | √ | × | |
| rs148636978 | CHSY1 | × | √ | × | |
| rs116886255 | CHSY1 | × | √ | × | |
| rs117597856 | CHSY1 | × | √ | × | |
| rs72768548 | CHSY1 | × | √ | × | |
| rs79439529 | CHSY1 | × | √ | × | |
| rs77824585 | CHSY1 | × | √ | × | |
| rs12595381 | CHSY1 | × | √ | × | |
| rs117548925 | CHSY1 | × | √ | × | |
| rs12593811 | CHSY1 | × | √ | × | Corneal astigmatism |
| rs12595443 | CHSY1 | × | √ | × | |
| rs4278729 | CHSY1 | × | √ | × | |
| rs8028597 | CHSY1 | × | √ | × | |
| rs61414324 | CHSY1 | × | √ | × | |
| rs56815075 | CHSY1 | × | √ | × | |
| rs76161631 | CHSY1 | × | √ | × | |
| rs11630342 | CHSY1 | × | √ | × | |
| rs150506848 | CHSY1 | × | √ | × | |
| rs80007328 | CHSY1 | × | √ | × | |
| rs569042880 | CHSY1 | × | √ | × | |
| rs79327037 | CHSY1 | × | √ | × | |
| rs140457834 | CHSY1 | × | √ | × | |
| rs79271117 | CHSY1 | × | √ | × | |
| rs7180690 | CHSY1 | × | √ | × | |
| rs76967748 | CHSY1 | × | √ | × | |
| rs76149780 | CHSY1 | × | √ | × | |
| rs117899511 | CHSY1 | × | √ | × | |
| rs7166488 | CHSY1 | × | √ | × | |
| rs111434193 | CHSY1 | × | √ | × | |
| rs74660635 | CHSY1 | × | √ | × | |
| rs8024948 | CHSY1 | × | √ | × | |
| rs56408443 | CHSY1 | × | √ | × | |
| rs79571285 | CHSY1 | × | √ | × | |
| rs35050682 | CHSY1 | × | √ | × | |
| rs8034294 | CHSY1 | × | √ | × | |
| rs79377162 | CHSY1 | × | √ | × | |
| rs571922872 | CHSY1 | × | √ | × | |
| rs150911529 | CHSY1 | × | √ | × | |
| rs149565210 | CHSY1 | × | √ | × | |
| rs2045163 | CHSY1 | × | √ | × | |
| rs140076563 | CHSY1 | × | √ | × | |
| rs35400551 | CHSY1 | × | √ | × | |
| rs79715969 | CHSY1 | × | √ | × | |
| rs151225991 | CHSY1 | × | √ | × | |
| rs140048104 | CHSY1 | × | √ | × | |
| rs149878299 | CHSY1 | × | √ | × | |
| rs147574216 | CHSY1 | × | √ | × | |
| rs75857854 | CHSY1 | × | √ | × | |
| rs148167919 | CHSY1 | × | √ | × | |
| rs10751717 | CHSY1 | × | √ | × | |
| rs6598435 | CHSY1 | × | √ | × | |
| rs12443106 | CHSY1 | × | √ | × | |
| rs7165361 | CHSY1 | √ | × | × | |
| rs752092 | CHSY1 | √ | × | × | Central corneal thickness |
| rs7182945 | CHSY1 | √ | × | × | |
| rs11247277 | CHSY1 | √ | × | × | |
| rs8032007 | CHSY1 | √ | × | × | |
| rs1562174 | COL4A1 | √ | × | × | |
| rs874203 | COL4A1 | √ | × | × | |
| rs874204 | COL4A1 | √ | × | × | |
| rs1891964 | COL4A1 | √ | × | × | |
| rs16975472 | COL4A1 | √ | × | × | |
| rs1975514 | COL4A1 | √ | × | × | Peripheral artery disease |
| rs1562173 | COL4A1 | √ | × | × | |
| rs503053 | COL4A1 | √ | × | × | |
| rs9521632 | COL4A1 | √ | × | × | Coronary artery disease |
| rs9515160 | COL4A1 | √ | × | × | |
| rs9521634 | COL4A1 | √ | × | × | Stroke |
| rs57646953 | CPNE2 | × | √ | × | |
| rs201965476 | CPNE2 | × | √ | × | |
| rs113552373 | CPNE2 | × | √ | × | Human milk oligosaccharid |
| rs11248294 | CPXM2 | √ | × | × | |
| rs10902825 | CPXM2 | √ | × | × | |
| rs2176488 | CPXM2 | √ | × | × | |
| rs17106110 | CPXM2 | √ | × | × | |
| rs17106114 | CPXM2 | √ | × | × | |
| rs7071385 | CPXM2 | √ | × | × | |
| rs7907113 | CPXM2 | √ | × | × | |
| rs7920094 | CPXM2 | √ | × | × | |
| rs6599626 | CPXM2 | √ | × | × | |
| rs6599627 | CPXM2 | √ | × | × | |
| rs28584696 | CPXM2 | √ | × | × | |
| rs28599786 | CPXM2 | √ | × | × | |
| rs28413513 | CPXM2 | √ | × | × | |
| rs28699067 | CPXM2 | √ | × | × | |
| rs7095732 | CPXM2 | √ | × | × | |
| rs12026252 | CRB1 | √ | × | × | |
| rs1577802 | CRB1 | √ | × | × | |
| rs10922190 | CRB1 | √ | × | × | |
| rs10494752 | CRB1 | √ | × | × | |
| rs10801600 | CRB1 | √ | × | × | |
| rs664767 | CRB1 | √ | × | × | |
| rs1415214 | CRB1 | √ | × | × | |
| rs1670167 | CTNNA3 | √ | × | × | Educational attainment |
| rs10822684 | CTNNA3 | √ | × | × | |
| rs2276344 | CXCR5 | × | √ | × | |
| rs10892307 | CXCR5 | × | √ | × | |
| rs3782050 | CXCR5 | × | √ | × | |
| rs3176905 | CXCR5 | × | √ | × | |
| rs523604 | CXCR5 | × | √ | × | Multiple sclerosis |
| rs74918784 | CXCR5 | × | √ | × | |
| rs77329980 | CXCR5 | × | √ | × | |
| rs118182497 | CXCR5 | × | √ | × | |
| rs78031275 | CXCR5 | × | √ | × | |
| rs3922 | CXCR5 | × | √ | × | Height |
| rs11657403 | CYTH1 | √ | × | × | |
| rs12451377 | CYTH1 | √ | × | × | |
| rs11656741 | CYTH1 | √ | × | × | |
| rs11656742 | CYTH1 | √ | × | × | |
| rs59606758 | CYTH1 | √ | × | × | |
| rs922838 | CYTH1 | √ | × | × | |
| rs8076588 | CYTH1 | √ | × | × | |
| rs7221314 | CYTH1 | √ | × | × | |
| rs62075585 | CYTH1 | √ | × | × | Systolic blood pressure |
| rs4675887 | D2HGDH | √ | × | × | |
| rs139238068 | D2HGDH | √ | × | × | |
| rs10758240 | DCAF12 | √ | × | × | |
| rs11557154 | DCAF12 | √ | × | × | Neutrophil percentage of w |
| rs60279027 | DCAF12 | √ | × | × | |
| rs9375502 | ECHDC1 | √ | × | × | |
| rs9388562 | ECHDC1 | √ | × | × | |
| rs76880977 | ECHDC1 | √ | × | × | |
| rs3756994 | ECHDC1 | √ | × | × | |
| rs10865025 | EDAR | √ | × | × | |
| rs12027793 | EGLN1 | √ | × | × | |
| rs209490 | ELOVL5 | √ | × | × | |
| rs7774230 | ESR1 | √ | × | × | |
| rs9322332 | ESR1 | √ | × | × | Fat-free ma |
| rs953 | EXOC8 | × | √ | × | |
| rs2053165 | EXOC8 | × | √ | × | |
| rs2064766 | EXOC8 | × | √ | × | |
| rs10489609 | EXOC8 | × | √ | × | Hemoglobin |
| rs75696360 | EXOC8 | × | √ | × | |
| rs11600929 | EXT2 | √ | × | × | |
| rs11600984 | EXT2 | √ | × | × | |
| rs4379834 | EXT2 | √ | × | × | |
| rs10838240 | EXT2 | √ | × | × | |
| rs7118541 | EXT2 | √ | × | × | |
| rs1495119 | EXT2 | √ | × | × | |
| rs2031693 | FAM102A | √ | × | × | |
| rs11789803 | FAM102A | √ | × | × | |
| rs4837212 | FAM102A | √ | × | × | |
| rs7849083 | FAM102A | √ | × | × | |
| rs6478804 | FAM102A | √ | × | × | |
| rs7037993 | FAM102A | √ | × | × | |
| rs12644477 | FBXW7 | √ | × | × | |
| rs9684643 | FBXW7 | √ | × | × | |
| rs75026869 | FBXW7 | √ | × | × | |
| rs77741711 | FBXW7 | √ | × | × | |
| rs11742062 | FBXW7 | √ | × | × | |
| rs14254953 | FBXW7 | √ | × | × | |
| rs872010 | GAS8 | √ | × | × | |
| rs14351569 | GNPAT | × | √ | × | |
| rs9928982 | GPR97 | √ | × | × | |
| rs1175931 | IARS | √ | × | × | |
| rs471478 | IARS | √ | × | × | COL1A1/Ol |
| rs504708 | IARS | √ | × | × | |
| rs1152757 | IARS | √ | × | × | |
| rs3996311 | IARS | √ | × | × | |
| rs10850132 | IQCD | √ | × | × | |
| rs4251460 | IRAK4 | × | √ | × | |
| rs4251465 | IRAK4 | × | √ | × | |
| rs4238087 | IRAK4 | × | √ | × | |
| rs4251466 | IRAK4 | × | √ | × | |
| rs1461567 | IRAK4 | × | √ | × | |
| rs11809731 | IRAK4 | × | √ | × | |
| rs3794262 | IRAK4 | × | √ | × | Vaginal mic |
| rs4251473 | IRAK4 | × | √ | × | |
| rs18134893 | IRAK4 | × | √ | × | |
| rs55973711 | IRAK4 | × | √ | × | |
| rs4251475 | IRAK4 | × | √ | × | |
| rs4251481 | IRAK4 | × | √ | × | |
| rs76136881 | IRAK4 | × | √ | × | |
| rs11182259 | IRAK4 | × | √ | × | |
| rs11182261 | IRAK4 | × | √ | × | |
| rs11699938 | IRAK4 | × | √ | × | |
| rs14057814 | IRAK4 | × | √ | × | |
| rs79154645 | IRAK4 | × | √ | × | |
| rs4251513 | IRAK4 | × | √ | × | |
| rs1144836 | ITPKB | √ | × | × | |
| rs1144835 | ITPKB | √ | × | × | |
| rs4653762 | ITPKB | √ | × | × | |
| rs2298169 | ITPKB | √ | × | × | |
| rs337616 | KLF3 | √ | × | × | |
| rs6531656 | KLF3 | √ | × | × | Lymphocyte |
| rs168084 | KLF3-AS1 | √ | × | × | |
| rs10842374 | LINC00477 | √ | × | × | |
| rs6487399 | LINC00477 | √ | × | × | |
| rs6487400 | LINC00477 | √ | × | × | |
| rs7299236 | LINC00477 | √ | × | × | |
| rs6421879 | LOC101927 | √ | × | × | |
| rs6894176 | LOC101927 | √ | × | × | |
| rs6875260 | LOC101927 | √ | × | × | |
| rs6894916 | LOC101927 | √ | × | × | |
| rs7721374 | LOC101927 | √ | × | × | |
| rs7711344 | LOC101927 | √ | × | × | |
| rs11241458 | LOC101927 | √ | × | × | |
| rs7703431 | LOC101927 | √ | × | × | |
| rs7708440 | LOC101927 | √ | × | × | |
| rs6595126 | LOC101927 | √ | × | × | |
| rs11950959 | LOC101927 | √ | × | × | |
| rs7727130 | LOC101927 | √ | × | × | |
| rs6867291 | LOC101927 | √ | × | × | |
| rs77788081 | LOC101927 | √ | × | × | |
| rs77124635 | LOC101927 | √ | × | × | |
| rs74926979 | LOC101927 | √ | × | × | |
| rs75622096 | LOC101927 | √ | × | × | |
| rs12719243 | LOC101927 | √ | × | × | |
| rs6873260 | LOC101927 | √ | × | × | |
| rs6883163 | LOC101927 | √ | × | × | |
| rs4440399 | LOC101927 | √ | × | × | |
| rs9327075 | LOC101927 | √ | × | × | |
| rs6595142 | LOC101927 | √ | × | × | |
| rs6595143 | LOC101927 | √ | × | × | |
| rs13360367 | LOC101927 | √ | × | × | |
| rs7708133 | LOC101927 | √ | × | × | |
| rs7708340 | LOC101927 | √ | × | × | |
| rs9490762 | LOC101927 | √ | × | × | |
| rs13205992 | LOC101927 | √ | × | × | |
| rs9401672 | LOC101927 | √ | × | × | |
| rs72613284 | LOC101927 | √ | × | × | |
| rs13192662 | LOC101927 | √ | × | × | |
| rs7744051 | LOC101927 | √ | × | × | |
| rs6911883 | LOC101927 | √ | × | × | |
| rs9490772 | LOC101927 | √ | × | × | |
| rs4406254 | LOC101927 | √ | × | × | |
| rs6909484 | LOC101927 | √ | × | × | |
| rs1343640 | LOC101927 | √ | × | × | |
| rs4428524 | LOC101927 | √ | × | × | |
| rs4412217 | LOC101927 | √ | × | × | |
| rs4613836 | LOC101927 | √ | × | × | Body mass |
| rs9482393 | LOC101927 | √ | × | × | |
| rs17085387 | LOC101927 | √ | × | × | |
| rs12942013 | LOC101928 | √ | × | × | |
| rs66530414 | LOC101928 | √ | × | × | |
| rs2840098 | LOC102723 | × | √ | × | |
| rs7162279 | LOC102723 | × | √ | × | |
| rs2254541 | LRSAM1 | √ | × | × | |
| rs56131876 | LRSAM1 | √ | × | × | |
| rs1539568 | LRSAM1 | √ | × | × | |
| rs2246600 | LRSAM1 | √ | × | × | |
| rs7875862 | LRSAM1 | √ | × | × | |
| rs2246375 | LRSAM1 | √ | × | × | |
| rs2243464 | LRSAM1 | √ | × | × | |
| rs2245677 | LRSAM1 | √ | × | × | |
| rs2202893 | LUZP2 | √ | × | × | |
| rs2136483 | LUZP2 | √ | × | × | |
| rs2896706 | LUZP2 | √ | × | × | |
| rs10834377 | LUZP2 | √ | × | × | |
| rs11028003 | LUZP2 | √ | × | × | |
| rs1912138 | LUZP2 | √ | × | × | |
| rs1912139 | LUZP2 | √ | × | × | |
| rs2403959 | LUZP2 | √ | × | × | |
| rs2036750 | LUZP2 | √ | × | × | |
| rs10834382 | LUZP2 | √ | × | × | |
| rs11028007 | LUZP2 | √ | × | × | |
| rs12273206 | LUZP2 | √ | × | × | |
| rs10767212 | LUZP2 | √ | × | × | |
| rs10834384 | LUZP2 | √ | × | × | |
| rs7107070 | LUZP2 | √ | × | × | |
| rs1509605 | LUZP2 | √ | × | × | Depressive |
| rs10834385 | LUZP2 | √ | × | × | |
| rs12276298 | LUZP2 | √ | × | × | |
| rs12295653 | LUZP2 | √ | × | × | |
| rs1509610 | LUZP2 | √ | × | × | |
| rs1509612 | LUZP2 | √ | × | × | |
| rs1509613 | LUZP2 | √ | × | × | |
| rs11028012 | LUZP2 | √ | × | × | |
| rs10834386 | LUZP2 | √ | × | × | |
| rs7105561 | LUZP2 | √ | × | × | |
| rs7105854 | LUZP2 | √ | × | × | |
| rs7121909 | LUZP2 | √ | × | × | |
| rs6484057 | LUZP2 | √ | × | × | |
| rs1509614 | LUZP2 | √ | × | × | |
| rs1039837 | LUZP2 | √ | × | × | |
| rs11028015 | LUZP2 | √ | × | × | |
| rs6484058 | LUZP2 | √ | × | × | |
| rs7936313 | LUZP2 | √ | × | × | |
| rs1912145 | LUZP2 | √ | × | × | |
| rs11028020 | LUZP2 | √ | × | × | |
| rs11028021 | LUZP2 | √ | × | × | |
| rs34167424 | LUZP2 | √ | × | × | |
| rs4465365 | LUZP2 | √ | × | × | |
| rs12289378 | LUZP2 | √ | × | × | |
| rs7951458 | LUZP2 | √ | × | × | |
| rs11529784 | LUZP2 | √ | × | × | |
| rs9943629 | LUZP2 | √ | × | × | |
| rs60028742 | MIR548AC | √ | × | × | |
| rs62256972 | MIR548AC | √ | × | × | |
| rs62256975 | MIR548AC | √ | × | × | |
| rs79562849 | MIR548AC | √ | × | × | |
| rs62256976 | MIR548AC | √ | × | × | |
| rs62256977 | MIR548AC | √ | × | × | |
| rs11535912 | MIR548AC | √ | × | × | |
| rs6774454 | MIR548AC | √ | × | × | |
| rs77832017 | MIR548AC | √ | × | × | |
| rs60137984 | MIR548AC | √ | × | × | |
| rs9395899 | MLIP | √ | × | × | |
| rs9650391 | MSR1 | √ | × | × | |
| rs35796001 | MSR1 | √ | × | × | |
| rs6986419 | MSR1 | √ | × | × | |
| rs34918250 | MSR1 | √ | × | × | |
| rs35281051 | MSR1 | √ | × | × | |
| rs918 | MSR1 | √ | × | × | |
| rs13306546 | MSR1 | √ | × | × | |
| rs13306544 | MSR1 | √ | × | × | |
| rs12675467 | MSR1 | √ | × | × | |
| rs12678016 | MSR1 | √ | × | × | |
| rs12677608 | MSR1 | √ | × | × | |
| rs12679486 | MSR1 | √ | × | × | |
| rs34612904 | MSR1 | √ | × | × | |
| rs7015682 | MSR1 | √ | × | × | |
| rs6995004 | MSR1 | √ | × | × | |
| rs6995009 | MSR1 | √ | × | × | |
| rs6981908 | MSR1 | √ | × | × | |
| rs6982066 | MSR1 | √ | × | × | |
| rs6982504 | MSR1 | √ | × | × | |
| rs78016136 | MSR1 | √ | × | × | |
| rs76903738 | MSR1 | √ | × | × | |
| rs76841569 | MSR1 | √ | × | × | |
| rs12676117 | MSR1 | √ | × | × | |
| rs12675698 | MSR1 | √ | × | × | |
| rs12676141 | MSR1 | √ | × | × | |
| rs12676172 | MSR1 | √ | × | × | |
| rs12676591 | MSR1 | √ | × | × | |
| rs12678046 | MSR1 | √ | × | × | |
| rs62491230 | MSR1 | √ | × | × | |
| rs41454647 | MSR1 | √ | × | × | |
| rs80002038 | MSR1 | √ | × | × | |
| rs77089638 | MSR1 | √ | × | × | |
| rs75164148 | MXI1 | √ | × | × | |
| rs77120294 | NELL1 | √ | × | × | |
| rs76609379 | NELL1 | √ | × | × | |
| rs79269343 | NELL1 | √ | × | × | |
| rs4923635 | NELL1 | × | √ | × | |
| rs75942970 | NELL1 | × | √ | × | |
| rs56272920 | NELL1 | × | √ | × | |
| rs76882629 | NELL1 | × | √ | × | |
| rs4923645 | NELL1 | × | √ | × | |
| rs80197260 | NELL1 | × | √ | × | |
| rs77386834 | NELL1 | × | √ | × | |
| rs12421933 | NELL1 | × | √ | × | |
| rs12421940 | NELL1 | × | √ | × | |
| rs4441013 | NELL1 | × | √ | × | |
| rs4922558 | NELL1 | × | √ | × | |
| rs60795440 | NELL1 | × | √ | × | |
| rs58530376 | NELL1 | × | √ | × | |
| rs58907183 | NELL1 | × | √ | × | |
| rs7933730 | NELL1 | × | √ | × | |
| rs753947 | NRF1 | × | √ | × | |
| rs10245818 | NRF1 | √ | × | × | |
| rs28689822 | NRF1 | √ | × | × | |
| rs78086437 | NRF1 | √ | × | × | |
| rs4731610 | NRF1 | √ | × | × | |
| rs2402974 | NRF1 | × | √ | × | |
| rs1806626 | NRF1 | × | √ | × | |
| rs61193920 | PACRG | × | √ | × | |
| rs55423697 | PACRG | × | √ | × | |
| rs7082393 | ARHGAP21 | √ | × | × | |
| rs6481497 | ARMC4 | √ | × | × | |
| rs3731847 | ATL2 | √ | × | × | |
| rs7211555 | B9D1 | × | × | √ | |
| rs2072351 | BLM | √ | × | × | |
| rs2181162 | BTBD16 | √ | × | × | |
| rs2181161 | BTBD16 | √ | × | × | |
| rs12500935 | CAMK2D | √ | × | × | |
| rs12570981 | CCAR1 | √ | × | × | |
| rs10202541 | CCDC138 | √ | × | × | |
| rs2272459 | CHD2 | √ | × | × | |
| rs11631553 | CHD2 | √ | × | × | |
| rs3826035 | CHD2 | √ | × | × | |
| rs17610487 | CHD2 | √ | × | × | |
| rs7171515 | CHSY1 | √ | × | × | |
| rs34980490 | COMMD9 | √ | × | × | |
| rs17616836 | CRB1 | √ | × | × | |
| rs4915440 | CRB1 | √ | × | × | |
| rs12022402 | CRB1 | √ | × | × | |
| rs1415343 | CRB1 | √ | × | × | |
| rs7553508 | CRB1 | √ | × | × | |
| rs7530522 | CRB1 | √ | × | × | |
| rs1337005 | CRB1 | √ | × | × | Complemen |
| rs10801601 | CRB1 | √ | × | × | |
| rs4915208 | CRB1 | √ | × | × | |
| rs1337170 | CRB1 | √ | × | × | |
| rs10922197 | CRB1 | √ | × | × | |
| rs2432674 | CYP1B1-AS | √ | × | × | |
| rs400681 | CYP1B1-AS | √ | × | × | |
| rs432252 | CYP1B1-AS | √ | × | × | |
| rs28433797 | DCTD | √ | × | × | |
| rs76052760 | DDR2 | × | √ | × | |
| rs7524537 | DDR2 | × | √ | × | |
| rs14938698 | DDR2 | × | √ | × | |
| rs14490426 | DDR2 | × | √ | × | |
| rs14043807 | DDR2 | × | √ | × | |
| rs6684509 | DDR2 | × | √ | × | Height |
| rs74896112 | DDR2 | × | √ | × | |
| rs74345863 | DDR2 | × | √ | × | |
| rs57245690 | DDR2 | × | √ | × | |
| rs11781009 | DDR2 | × | √ | × | |
| rs79775216 | DDR2 | × | √ | × | |
| rs10799854 | DDR2 | × | √ | × | |
| rs10917578 | DDR2 | × | √ | × | |
| rs6427690 | DDR2 | × | √ | × | |
| rs6427691 | DDR2 | × | √ | × | |
| rs11811703 | DDR2 | × | √ | × | |
| rs7533399 | DDR2 | × | √ | × | |
| rs10799855 | DDR2 | × | √ | × | |
| rs10753582 | DDR2 | × | √ | × | |
| rs7412891 | DDR2 | × | √ | × | |
| rs6667476 | DDR2 | × | √ | × | |
| rs6670722 | DDR2 | × | √ | × | |
| rs6683356 | DDR2 | × | √ | × | |
| rs6694368 | DDR2 | × | √ | × | |
| rs6694717 | DDR2 | × | √ | × | |
| rs4351614 | DDR2 | × | √ | × | |
| rs6930849 | ECHDC1 | √ | × | × | |
| rs12196700 | ELOVL5 | √ | × | × | |
| rs209483 | ELOVL5 | √ | × | × | |
| rs209485 | ELOVL5 | √ | × | × | |
| rs7208857 | EPN2 | × | × | √ | |
| rs9479126 | ESR1 | √ | × | × | Height |
| rs11037881 | EXT2 | √ | × | × | |
| rs7117287 | EXT2 | √ | × | × | |
| rs10819322 | FAM102A | √ | × | × | |
| rs3802355 | FAM102A | √ | × | × | |
| rs2417062 | FAM102A | √ | × | × | |
| rs6415846 | FAM102A | √ | × | × | |
| rs6478807 | FAM102A | √ | × | × | |
| rs2031694 | FAM102A | √ | × | × | |
| rs7026115 | FAM102A | √ | × | × | |
| rs16970167 | FAM155A | √ | × | × | |
| rs1924446 | FAM155A | √ | × | × | |
| rs4652961 | GNL2 | √ | × | × | |
| rs14853943 | GRB10 | √ | × | × | |
| rs11974751 | GRB10 | √ | × | × | |
| rs11792542 | IARS | √ | × | × | |
| rs501814 | IARS | √ | × | × | |
| rs556155 | IARS | √ | × | × | |
| rs4743866 | IARS | √ | × | × | |
| rs4743867 | IARS | √ | × | × | |
| rs11793505 | IARS | √ | × | × | |
| rs1175930 | IARS | √ | × | × | |
| rs54897707 | IARS | √ | × | × | |
| rs472163 | IARS | √ | × | × | |
| rs4743868 | IARS | √ | × | × | |
| rs9409659 | IARS | √ | × | × | |
| rs9409466 | IARS | √ | × | × | |
| rs77321619 | IKZF1 | √ | × | × | |
| rs76579247 | ILKAP | × | × | √ | |
| rs12617118 | ILKAP | × | × | √ | |
| rs37088102 | IRAK4 | √ | × | × | |
| rs11160836 | JAG2 | √ | × | × | |
| rs2239285 | JAG2 | √ | × | × | Vertex-wise |
| rs1882845 | JAG2 | √ | × | × | |
| rs4983589 | JAG2 | √ | × | × | Cortical sur |
| rs56302643 | KIFC3 | √ | × | × | |
| rs58082302 | KIFC3 | √ | × | × | |
| rs9576449 | LINC00571 | √ | × | × | |
| rs8031036 | LINC00928 | √ | × | × | |
| rs4663901 | LINC01107 | √ | × | × | |
| rs11712419 | LOC101927 | × | √ | × | |
| rs7707724 | LOC101927 | × | √ | × | |
| rs7446705 | LOC101927 | × | √ | × | |
| rs75337620 | LOC101927 | × | √ | × | |
| rs78903852 | LOC101927 | × | √ | × | |
| rs56933718 | LOC284395 | √ | × | × | |
| rs4973649 | LOC728323 | √ | × | × | |
| rs12617606 | LOC728323 | √ | × | × | |
| rs10834392 | LUZP2 | √ | × | × | |
| rs11028036 | LUZP2 | √ | × | × | |
| rs12225488 | LUZP2 | √ | × | × | |
| rs6747400 | METTL8 | √ | × | × | |
| rs12616940 | METTL8 | √ | × | × | |
| rs1558963 | METTL8 | √ | × | × | |
| rs62183460 | METTL8 | √ | × | × | |
| rs11810958 | MIR548AC | √ | × | × | |
| rs73045642 | MIR548AC | √ | × | × | |
| rs62256973 | MIR548AC | √ | × | × | |
| rs68093882 | MIR548AC | √ | × | × | |
| rs57859482 | MIR548AC | √ | × | × | |
| rs4513393 | MIR548AC | √ | × | × | |
| rs11745547 | MIR548AC | √ | × | × | |
| rs76435632 | MIR548AC | √ | × | × | Type 2 diab |
| rs991199 | MLIP | √ | × | × | |
| rs80334189 | MLIP | √ | × | × | |
| rs13306538 | MSR1 | √ | × | × | |
| rs13306537 | MSR1 | √ | × | × | |
| rs7005569 | MSR1 | √ | × | × | |
| rs6737057 | MTERFD2 | √ | × | × | |
| rs78807828 | NELL1 | √ | × | × | |
| rs55840704 | NLRP8 | √ | × | × | |
| rs710163 | NOL8 | √ | × | × | |
| rs9409664 | NOL8 | √ | × | × | |
| rs17555614 | NRF1 | √ | × | × | |
| rs80257343 | NRF1 | √ | × | × | |
| rs74734973 | NRF1 | √ | × | × | |
| rs78890318 | NRF1 | √ | × | × | |
| rs2001330 | NRF1 | √ | × | × | |
| rs78841374 | NRF1 | √ | × | × | |
| rs79677010 | NRF1 | √ | × | × | |
| rs79020150 | PARK2 | √ | × | × | |
| rs7566357 | PASK | √ | × | × | |
| rs2408716 | PCED1B | √ | × | × | |
| rs10881048 | PCED1B | √ | × | × | |
| rs78639475 | PER2 | √ | × | × | |
| rs57056606 | PER2 | √ | × | × | |
| rs59737477 | PLCL2 | √ | × | × | |
| rs4684306 | PLCL2 | √ | × | × | |
| rs62246033 | PLCL2 | √ | × | × | |
| rs58750701 | PLCL2 | √ | × | × | |
| rs28717370 | PRELP | √ | × | × | |
| rs191318 | PRR5L | √ | × | × | |
| rs700876 | RANBP2 | √ | × | × | |
| rs1649330 | RANBP2 | √ | × | × | |
| rs1649329 | RANBP2 | √ | × | × | |
| rs1649328 | RANBP2 | √ | × | × | |
| rs56132830 | RANBP2 | √ | × | × | |
| rs1614285 | RANBP2 | √ | × | × | |
| rs2072121 | RNF112 | × | × | √ | |
| rs2072120 | RNF112 | × | × | √ | |
| rs2072118 | RNF112 | × | × | √ | |
| rs9372861 | RNF146 | √ | × | × | |
| rs9401947 | RNF146 | √ | × | × | |
| rs579890 | SIK3 | √ | × | × | |
| rs645258 | SIK3 | √ | × | × | Triglyceride |
| rs2513095 | SIK3 | √ | × | × | |
| rs62293481 | SKIL | √ | × | × | |
| rs11056825 | SLC15A5 | √ | × | × | |
| rs6662367 | SNIP1 | √ | × | × | |
| rs6426023 | SNIP1 | √ | × | × | |
| rs7139365 | SOX5 | √ | × | × | |
| rs11047238 | SOX5 | √ | × | × | |
| rs11047437 | SOX5 | √ | × | × | |
| rs11047459 | SOX5 | √ | × | × | |
| rs7957500 | SOX5 | √ | × | × | |
| rs4963760 | SOX5 | √ | × | × | |
| rs12309160 | SOX5 | √ | × | × | |
| rs9630247 | SOX5 | √ | × | × | |
| rs11929833 | SPOCK3 | √ | × | × | |
| rs28626960 | SPOCK3 | √ | × | × | |
| rs28433769 | SPOCK3 | √ | × | × | |
| rs80297501 | SPTLC1 | √ | × | × | |
| rs68122210 | TET1 | √ | × | × | |
| rs6582495 | TMEM117 | √ | × | × | |
| rs11182382 | TMEM117 | √ | × | × | |
| rs14105304 | TMEM117 | √ | × | × | |
| rs12322164 | TMEM117 | √ | × | × | |
| rs11182384 | TMEM117 | √ | × | × | |
| rs12231403 | TMEM117 | √ | × | × | |
| rs859699 | TMEM117 | √ | × | × | |
| rs842194 | TMEM117 | √ | × | × | |
| rs15037743 | TMEM117 | √ | × | × | |
| rs842193 | TMEM117 | √ | × | × | |
| rs73087684 | TMEM117 | √ | × | × | X-12013 lev |
| rs1701083 | TMEM117 | √ | × | × | |
| rs11182385 | TMEM117 | √ | × | × | |
| rs10880616 | TMEM117 | √ | × | × | |
| rs12296857 | TMEM117 | √ | × | × | |
| rs14036701 | TMEM117 | √ | × | × | |
| rs17094013 | TMEM117 | √ | × | × | |
| rs73087700 | TMEM117 | √ | × | × | |
| rs9919676 | TMEM117 | √ | × | × | |
| rs11182390 | TMEM117 | √ | × | × | |
| rs10880618 | TMEM117 | √ | × | × | |
| rs9919742 | TMEM117 | √ | × | × | |
| rs11182394 | TMEM117 | √ | × | × | |
| rs14567800 | TMEM117 | √ | × | × | |
| rs14837371 | TMEM117 | √ | × | × | |
| rs12313365 | TMEM117 | √ | × | × | |
| rs73089723 | TMEM117 | √ | × | × | |
| rs12099769 | TMEM117 | √ | × | × | |
| rs14470724 | TMEM117 | √ | × | × | |
| rs11182402 | TMEM117 | √ | × | × | |
| rs11182403 | TMEM117 | √ | × | × | |
| rs11182404 | TMEM117 | √ | × | × | |
| rs11182406 | TMEM117 | √ | × | × | |
| rs11182409 | TMEM117 | √ | × | × | |
| rs56198335 | TMEM117 | √ | × | × | |
| rs842199 | TMEM117 | √ | × | × | |
| rs6766775 | TP63 | √ | × | × | |
| rs78394743 | TPCN1 | √ | × | × | |
| rs7133428 | TPCN1 | √ | × | × | |
| rs79778981 | TPCN1 | √ | × | × | |
| rs77303274 | TPCN1 | √ | × | × | |
| rs7296638 | TPCN1 | √ | × | × | |
| rs79423105 | TPCN1 | √ | × | × | |
| rs59205839 | TPCN1 | √ | × | × | |
| rs2873478 | TRDN | √ | × | × | |
| rs4346869 | TRDN | √ | × | × | |
| rs1431290 | TRDN | √ | × | × | |
| rs1346920 | TRDN | √ | × | × | |
| rs1431288 | TRDN | √ | × | × | |
| rs76177651 | TRDN | √ | × | × | |
| rs11033258 | TRIM44 | √ | × | × | |
| rs12222279 | TRIM44 | √ | × | × | |
| rs78647470 | TRIM44 | √ | × | × | |
| rs75667066 | TRIM44 | √ | × | × | |
| rs13205992 | LOC101927 | √ | × | | |
| rs9401672 | LOC101927 | √ | × | | |
| rs72613284 | LOC101927 | √ | × | | |
| rs13192662 | LOC101927 | √ | × | | |
| rs7744051 | LOC101927 | √ | × | | |
| rs6911883 | LOC101927 | √ | × | | |
| rs9490772 | LOC101927 | √ | × | | |
| rs4406254 | LOC101927 | √ | × | | |
| rs6909484 | LOC101927 | √ | × | | |
| rs1343640 | LOC101927 | √ | × | | |
| rs4428524 | LOC101927 | √ | × | | |
| rs4412217 | LOC101927 | √ | × | | |
| rs4613836 | LOC101927 | √ | × | | Body mass index |
| rs9482393 | LOC101927 | √ | × | | |
| rs17085387 | LOC101927 | √ | × | | |
| rs15059623 | LOC101928 | √ | × | | |
| rs2136483 | LUZP2 | √ | × | | |
| rs2896706 | LUZP2 | √ | × | | |
| rs2202894 | LUZP2 | √ | × | | |
| rs11028003 | LUZP2 | √ | × | | |
| rs1912138 | LUZP2 | √ | × | | |
| rs1912139 | LUZP2 | √ | × | | |
| rs2403959 | LUZP2 | √ | × | | |
| rs2036750 | LUZP2 | √ | × | | |
| rs10834382 | LUZP2 | √ | × | | |
| rs11028007 | LUZP2 | √ | × | | |
| rs12273206 | LUZP2 | √ | × | | |
| rs10767212 | LUZP2 | √ | × | | |
| rs10834384 | LUZP2 | √ | × | | |
| rs7107070 | LUZP2 | √ | × | | |
| rs1509605 | LUZP2 | √ | × | | Depressive symptoms |
| rs10834385 | LUZP2 | √ | × | | |
| rs12276298 | LUZP2 | √ | × | | |
| rs12295653 | LUZP2 | √ | × | | |
| rs1509610 | LUZP2 | √ | × | | |
| rs1509612 | LUZP2 | √ | × | | |
| rs1509613 | LUZP2 | √ | × | | |
| rs11028012 | LUZP2 | √ | × | | |
| rs10834386 | LUZP2 | √ | × | | |
| rs7105561 | LUZP2 | √ | × | | |
| rs7105854 | LUZP2 | √ | × | | |
| rs7121909 | LUZP2 | √ | × | | |
| rs6484057 | LUZP2 | √ | × | | |
| rs1509614 | LUZP2 | √ | × | | |
| rs1039837 | LUZP2 | √ | × | | |
| rs11028015 | LUZP2 | √ | × | | |
| rs6484058 | LUZP2 | √ | × | | |
| rs7936313 | LUZP2 | √ | × | | |
| rs1912145 | LUZP2 | √ | × | | |
| rs11028020 | LUZP2 | √ | × | | |
| rs11028021 | LUZP2 | √ | × | | |
| rs34167424 | LUZP2 | √ | × | | |
| rs4465365 | LUZP2 | √ | × | | |
| rs12289378 | LUZP2 | √ | × | | |
| rs7951458 | LUZP2 | √ | × | | |
| rs11529784 | LUZP2 | √ | × | | |
| rs9943629 | LUZP2 | √ | × | | |
| rs7590275 | METTL8 | √ | × | | |
| rs11706875 | MIR548AC | √ | × | | |
| rs62256991 | MIR548AC | √ | × | | |
| rs60028742 | MIR548AC | √ | × | | |
| rs62256972 | MIR548AC | √ | × | | |
| rs62256975 | MIR548AC | √ | × | | |
| rs11237447 | MIR548AC | √ | × | | |
| rs79562849 | MIR548AC | √ | × | | |
| rs62256976 | MIR548AC | √ | × | | |
| rs62256977 | MIR548AC | √ | × | | |
| rs11535912 | MIR548AC | √ | × | | |
| rs6774454 | MIR548AC | √ | × | | |
| rs60137984 | MIR548AC | √ | × | | |
| rs62256982 | MIR548AC | √ | × | | |
| rs28367247 | MIR548AC | √ | × | | |
| rs9395899 | MLIP | √ | × | | |
| rs77708866 | MLIP | √ | × | | |
| rs9650391 | MSR1 | √ | × | | |
| rs35796001 | MSR1 | √ | × | | |
| rs11985261 | MSR1 | √ | × | | |
| rs33936250 | MSR1 | √ | × | | |
| rs6986419 | MSR1 | √ | × | | |
| rs34918250 | MSR1 | √ | × | | |
| rs35281051 | MSR1 | √ | × | | |
| rs918 | MSR1 | √ | × | | |
| rs13306546 | MSR1 | √ | × | | |
| rs13306544 | MSR1 | √ | × | | |
| rs12675467 | MSR1 | √ | × | | |
| rs12678016 | MSR1 | √ | × | | |
| rs12677608 | MSR1 | √ | × | | |
| rs12679486 | MSR1 | √ | × | | |
| rs34612904 | MSR1 | √ | × | | |
| rs7015682 | MSR1 | √ | × | | |
| rs6995004 | MSR1 | √ | × | | |
| rs6995009 | MSR1 | √ | × | | |
| rs6981908 | MSR1 | √ | × | | |
| rs6982066 | MSR1 | √ | × | | |
| rs6982504 | MSR1 | √ | × | | |
| rs78757102 | MSR1 | √ | × | | |
| rs78016136 | MSR1 | √ | × | | |
| rs76903738 | MSR1 | √ | × | | |
| rs76841569 | MSR1 | √ | × | | |
| rs12676117 | MSR1 | √ | × | | |
| rs12675698 | MSR1 | √ | × | | |
| rs12676141 | MSR1 | √ | × | | |
| rs12675748 | MSR1 | √ | × | | |
| rs12676172 | MSR1 | √ | × | | |
| rs12676591 | MSR1 | √ | × | | |
| rs12678046 | MSR1 | √ | × | | |
| rs62491230 | MSR1 | √ | × | | |
| rs62491233 | MSR1 | √ | × | | |
| rs56996957 | MSR1 | √ | × | | |
| rs41454647 | MSR1 | √ | × | | |
| rs41487145 | MSR1 | √ | × | | |
| rs78553524 | MSR1 | √ | × | | Protein quantitative trait loci (liver) |
| rs80002038 | MSR1 | √ | × | | |
| rs77089638 | MSR1 | √ | × | | |
| rs78765574 | MSR1 | √ | × | | |
| rs753947 | NRF1 | √ | × | | |
| rs7795187 | NRF1 | √ | × | | |
| rs10245818 | NRF1 | √ | × | | |
| rs28689822 | NRF1 | √ | × | | |
| rs12666356 | NRF1 | √ | × | | |
| rs11844206 | NUDT14 | √ | × | | |
| rs11844119 | NUDT14 | √ | × | | |
| rs79118964 | PAFAH1B2 | √ | × | | |
| rs10892083 | PAFAH1B2 | √ | × | | |
| rs9365464 | PARK2 | √ | × | | |
| rs10945842 | PARK2 | √ | × | | |
| rs12581006 | PCED1B | √ | × | | |
| rs4768776 | PCED1B | √ | × | | |
| rs10881052 | PCED1B | √ | × | | |
| rs1525452 | PCED1B | √ | × | | |
| rs36124720 | PER2 | √ | × | | |
| rs17103488 | PLEKHA1 | √ | × | | |
| rs4565845 | PLEKHA1 | √ | × | | |
| rs717144 | POLR1A | √ | × | | |
| rs16843395 | PPP1R7 | √ | × | | |
| rs58602684 | PPP1R9A | √ | × | | |
| rs12704790 | PPP1R9A | √ | × | | |
| rs2374983 | PPP1R9A | √ | × | | |
| rs79686315 | PRSS3 | √ | × | | |
| rs79665161 | PRSS3 | √ | × | | |
| rs97445685 | PRSS3 | √ | × | | |
| rs92171535 | PRSS3 | √ | × | | |
| rs87920364 | PRSS3 | √ | × | | |
| rs79644971 | PRSS3 | √ | × | | |
| rs79653051 | PRSS3 | √ | × | | |
| rs37738026 | PRSS3 | √ | × | | |
| rs11774278 | PSD3 | √ | × | | |
| rs56277438 | PSD3 | √ | × | | |
| rs73204534 | PSD3 | √ | × | | |
| rs11783114 | PSD3 | √ | × | | |
| rs826532 | RANBP2 | √ | × | | |
| rs1724203 | RANBP2 | √ | × | | |
| rs9854636 | RBMS3 | √ | × | | |
| rs9837143 | RBMS3 | √ | × | | |
| rs6795528 | RBMS3 | √ | × | | |
| rs2057119 | RDH11 | √ | × | | |
| rs6710923 | RMDN2 | √ | × | | |
| rs12175928 | RNF146 | √ | × | | |
| rs9388558 | RNF146 | √ | × | | |
| rs9372862 | RNF146 | √ | × | | |
| rs9388559 | RNF146 | √ | × | | |
| rs9321072 | RNF146 | √ | × | | |
| rs78238091 | RNF17 | √ | × | | |
| rs79091937 | 45537 | √ | × | | |
| rs11795970 | 45537 | √ | × | | |
| rs10892048 | SIK3 | √ | × | | |
| rs2273013 | SNIP1 | √ | × | | |
| rs12422241 | SOX5 | √ | × | | |
| rs59556093 | SOX5 | √ | × | | |
| rs61517249 | SOX5 | √ | × | | |
| rs10842250 | SOX5 | √ | × | | |
| rs10842252 | SOX5 | √ | × | | |
| rs11047179 | SOX5 | √ | × | | |
| rs11047182 | SOX5 | √ | × | | |
| rs76847583 | SOX5 | √ | × | | |
| rs12229376 | SOX5 | √ | × | | |
| rs11047185 | SOX5 | √ | × | | |
| rs11047191 | SOX5 | √ | × | | |
| rs28740811 | SOX5 | √ | × | | |
| rs7296101 | SOX5 | √ | × | | Vertex-wise sulcal depth |
| rs12231866 | SOX5 | √ | × | | |
| rs12582243 | SOX5 | √ | × | | |
| rs1355004 | SOX5 | √ | × | | |
| rs11047455 | SOX5 | √ | × | | |
| rs9668967 | SOX5 | √ | × | | |
| rs13146675 | SPOCK3 | √ | × | | |
| rs7691894 | SPOCK3 | √ | × | | |
| rs62352806 | SPOCK3 | √ | × | | |
| rs62352808 | SPOCK3 | √ | × | | |
| rs11937180 | SPOCK3 | √ | × | | |
| rs28469298 | SPOCK3 | √ | × | | |
| rs2219234 | SPOCK3 | √ | × | | |
| rs3828018 | SPOCK3 | √ | × | | |
| rs3790358 | SPOCK3 | √ | × | | |
| rs28699632 | SPOCK3 | √ | × | | |
| rs6827226 | SPOCK3 | √ | × | | |
| rs10819410 | SPTAN1 | √ | × | | |
| rs11797217 | SPTLC1 | √ | × | | |
| rs4142550 | STX11 | √ | × | | |
| rs3824523 | STXBP1 | √ | × | | |
| rs11754840 | STXBP1 | √ | × | | |
| rs3856943 | SYNPR | √ | × | | |
| rs1901304 | SYNPR | √ | × | | |
| rs933821 | TBC1D1 | √ | × | | |
| rs35308860 | TBC1D1 | √ | × | | |
| rs67733859 | TBC1D1 | √ | × | | |
| rs59608961 | TBC1D1 | √ | × | | |
| rs35021061 | TBC1D1 | √ | × | | |
| rs12649018 | TBC1D1 | √ | × | | |
| rs1364953 | TBC1D1 | √ | × | | Metabolite levels |
| rs9401367 | TBC1D32 | √ | × | | |
| rs2202065 | TBC1D32 | √ | × | | |
| rs2389414 | TBC1D32 | √ | × | | |
| rs9320799 | TBC1D32 | √ | × | | |
| rs9320800 | TBC1D32 | √ | × | | |
| rs9320801 | TBC1D32 | √ | × | | |
| rs4585597 | TBC1D32 | √ | × | | |
| rs9387942 | TBC1D32 | √ | × | | |
| rs9320803 | TBC1D32 | √ | × | | |
| rs4412244 | TBC1D32 | √ | × | | |
| rs9375022 | TBC1D32 | √ | × | | |
| rs9398629 | TBC1D32 | √ | × | | OTU99_45 (Faecalibacterium) prevalence |
| rs62052185 | TCF25 | √ | × | | |
| rs8044906 | TCF25 | √ | × | | |
| rs2270461 | TCF25 | √ | × | | |
| rs11199028 | TIAL1 | √ | × | | |
| rs11199030 | TIAL1 | √ | × | | |
| rs7971487 | TMEM117 | √ | × | | |
| rs11182329 | TMEM117 | √ | × | | |
| rs10880591 | TMEM117 | √ | × | | |
| rs2031056 | TMEM117 | √ | × | | |
| rs4499064 | TMEM117 | √ | × | | |
| rs10880592 | TMEM117 | √ | × | | |
| rs10748379 | TMEM117 | √ | × | | |
| rs10748380 | TMEM117 | √ | × | | |
| rs4238093 | TMEM117 | √ | × | | |
| rs10506232 | TMEM117 | √ | × | | |
| rs4768055 | TMEM117 | √ | × | | |
| rs10785474 | TMEM117 | √ | × | | |

| | | | | | |
|---|---|---|---|---|---|
| rs10971920 | DCAF12 | √ | × | × | |
| rs4879745 | DCAF12 | √ | × | × | |
| rs10511916 | DCAF12 | √ | × | × | |
| rs2275003 | DCAF12 | √ | × | × | Body mass index |
| rs10972169 | DCTN3 | √ | × | × | |
| rs139533955 | DEF8 | × | √ | × | |
| rs144195940 | DEF8 | × | √ | × | |
| rs62052222 | DEF8 | × | √ | × | |
| rs543250984 | DEF8 | × | √ | × | |
| rs113287072 | DEF8 | × | √ | × | |
| rs376876704 | DEF8 | × | √ | × | |
| rs77522907 | DEF8 | × | √ | × | |
| rs7192275 | DEF8 | × | √ | × | |
| rs7187431 | DEF8 | × | √ | × | |
| rs62052240 | DEF8 | × | √ | × | |
| rs62052241 | DEF8 | × | √ | × | |
| rs8166 | DEF8 | × | √ | × | |
| rs8063761 | DEF8 | × | √ | × | Squamous cell carcinoma |
| rs181992761 | DMXL1 | √ | × | × | |
| rs75889668 | DMXL1 | √ | × | × | |
| rs72784088 | DMXL1 | √ | × | × | |
| rs151123907 | DMXL1 | √ | × | × | |
| rs78391275 | DMXL1 | √ | × | × | |
| rs76083343 | DMXL1 | √ | × | × | |
| rs9327092 | DMXL1 | √ | × | × | |
| rs117049970 | DPM2 | √ | × | × | |
| rs2279522 | DPYSL4 | √ | × | × | |
| rs9372863 | ECHDC1 | √ | × | × | |
| rs117669051 | ECHDC1 | √ | × | × | |
| rs761842 | ECHDC1 | √ | × | × | |
| rs75527034 | ECHDC1 | √ | × | × | |
| rs9401948 | ECHDC1 | √ | × | × | |
| rs6936621 | ECHDC1 | √ | × | × | |
| rs3798856 | ECHDC1 | √ | × | × | |
| rs9375500 | ECHDC1 | √ | × | × | |
| rs4141423 | ECHDC1 | √ | × | × | |
| rs3823043 | ECHDC1 | √ | × | × | |
| rs9372865 | ECHDC1 | √ | × | × | |
| rs75258130 | ECHDC1 | √ | × | × | |
| rs17054536 | ECHDC1 | √ | × | × | |
| rs75973322 | ECHDC1 | √ | × | × | |
| rs9372866 | ECHDC1 | √ | × | × | |
| rs9375502 | ECHDC1 | √ | × | × | |
| rs9388562 | ECHDC1 | √ | × | × | |
| rs72961085 | ECHDC1 | √ | × | × | |
| rs76880977 | ECHDC1 | √ | × | × | |
| rs3756994 | ECHDC1 | √ | × | × | |
| rs10865025 | EDAR | √ | × | × | |
| rs370285919 | EFCAB14 | √ | × | × | |
| rs12029031 | EFCAB14 | √ | × | × | |
| rs10890412 | EFCAB14 | √ | × | × | |
| rs10890413 | EFCAB14 | √ | × | × | |
| rs11211339 | EFCAB14 | √ | × | × | |
| rs2275380 | EFCAB14 | √ | × | × | |
| rs1150064 | EFCAB14 | √ | × | × | |
| rs11211340 | EFCAB14 | √ | × | × | |
| rs12027793 | EGLN1 | √ | × | × | |
| rs12033229 | EGLN1 | × | × | √ | |
| rs144187440 | ELOVL5 | √ | × | × | |
| rs12523973 | ELOVL5 | √ | × | × | |
| rs209490 | ELOVL5 | √ | × | × | |
| rs77578809 | ELOVL5 | √ | × | × | |
| rs7747926 | ELOVL5 | √ | × | × | |
| rs209498 | ELOVL5 | √ | × | × | |
| rs7738788 | ELOVL5 | √ | × | × | |
| rs209501 | ELOVL5 | √ | × | × | |
| rs74804858 | ESR1 | √ | × | × | |
| rs17847065 | ESR1 | √ | × | × | |
| rs61412877 | ESR1 | √ | × | × | |
| rs9340782 | ESR1 | √ | × | × | |
| rs7774230 | ESR1 | √ | × | × | |
| rs9322332 | ESR1 | √ | × | × | Fat-free mass |
| rs953 | EXOC8 | × | √ | × | |
| rs2053165 | EXOC8 | × | √ | × | |
| rs2064766 | EXOC8 | × | √ | × | |
| rs10489609 | EXOC8 | × | √ | × | Hemoglobin concentration |
| rs75696360 | EXOC8 | × | √ | × | |
| rs11600929 | EXT2 | √ | × | × | |
| rs11600984 | EXT2 | √ | × | × | |
| rs4379834 | EXT2 | √ | × | × | |
| rs10838240 | EXT2 | √ | × | × | |
| rs7118541 | EXT2 | √ | × | × | |
| rs117013257 | EXT2 | √ | × | × | |
| rs78181837 | EXT2 | √ | × | × | |
| rs117839205 | EXT2 | √ | × | × | |
| rs1495119 | EXT2 | √ | × | × | |
| rs74763346 | F13B | × | √ | × | |
| rs34630941 | FAM101A | √ | × | × | |
| rs2031693 | FAM102A | √ | × | × | |
| rs11789803 | FAM102A | √ | × | × | |
| rs576261762 | FAM102A | √ | × | × | |
| rs4837212 | FAM102A | √ | × | × | |
| rs7849083 | FAM102A | √ | × | × | |
| rs6478804 | FAM102A | √ | × | × | |
| rs7037993 | FAM102A | √ | × | × | |
| rs7023663 | FAM102A | √ | × | × | |
| rs943392 | FAM102A | √ | × | × | |
| rs3739819 | FAM102A | √ | × | × | |
| rs141545611 | FAM102A | √ | × | × | |
| rs59944943 | FAM114A1 | √ | × | × | |
| rs56229790 | FAM114A1 | √ | × | × | |
| rs80227559 | FAM114A1 | √ | × | × | |
| rs117258396 | FAM114A1 | √ | × | × | |
| rs73236661 | FAM114A1 | √ | × | × | |
| rs11096964 | FAM114A1 | √ | × | × | |
| rs11555334 | FAM114A1 | √ | × | × | |
| rs11938589 | FAM114A1 | √ | × | × | |
| rs11944231 | FAM114A1 | √ | × | × | |
| rs9520451 | FAM155A | √ | × | × | |
| rs9520452 | FAM155A | √ | × | × | |
| rs727042 | FAM155A | √ | × | × | |
| rs78056606 | FAM155A | √ | × | × | |
| rs4611321 | FAM155A | √ | × | × | |
| rs139224429 | FAM155A | √ | × | × | |
| rs16970971 | FAM155A | √ | × | × | |
| rs16970981 | FAM155A | √ | × | × | |
| rs10508186 | FAM155A | √ | × | × | |
| rs16970985 | FAM155A | √ | × | × | |
| rs61966045 | FAM155A | √ | × | × | |
| rs7139609 | FAM155A | √ | × | × | |
| rs933716 | FAM192A | × | √ | × | |
| rs767505 | FAM192A | √ | × | × | |
| rs7204054 | FAM192A | √ | × | × | |
| rs146393411 | FAM49B | √ | × | × | |
| rs12676834 | FAM49B | √ | × | × | |
| rs117912630 | FAM49B | √ | × | × | |
| rs371915574 | FAM49B | √ | × | × | |
| rs144368752 | FAM49B | √ | × | × | |
| rs76939852 | FARP2 | √ | × | × | |
| rs3740286 | FAS | √ | × | × | |
| rs79878664 | FBXW7 | √ | × | × | |
| rs1516822 | FBXW7 | √ | × | × | |
| rs12644477 | FBXW7 | √ | × | × | |
| rs1878342 | FBXW7 | √ | × | × | |
| rs72955060 | FBXW7 | √ | × | × | |
| rs9684643 | FBXW7 | √ | × | × | |
| rs17028897 | FBXW7 | √ | × | × | |
| rs75026869 | FBXW7 | √ | × | × | |
| rs900977 | FBXW7 | √ | × | × | |
| rs139058585 | FBXW7 | √ | × | × | |
| rs77741711 | FBXW7 | √ | × | × | |
| rs548608655 | FBXW7 | √ | × | × | |
| rs77481157 | FBXW7 | √ | × | × | |
| rs117420629 | FBXW7 | √ | × | × | |
| rs72955085 | FBXW7 | √ | × | × | |
| rs12651630 | FBXW7 | √ | × | × | |
| rs142549531 | FBXW7 | √ | × | × | |
| rs142142666 | FBXW7 | √ | × | × | |
| rs3126095 | FLG-AS1 | × | × | √ | |
| rs4837427 | FNBP1 | √ | × | × | |
| rs10448161 | FOCAD | √ | × | × | |
| rs12551126 | FOCAD | √ | × | × | |
| rs4978125 | FOCAD | √ | × | × | |
| rs7029608 | FOCAD | √ | × | × | |
| rs35612917 | FOCAD | √ | × | × | |
| rs13297201 | FOCAD | √ | × | × | |
| rs76677990 | FOCAD | √ | × | × | Vaginal microbiome MetaC |
| rs10987596 | GARNL3 | √ | × | × | |
| rs10987598 | GARNL3 | √ | × | × | |
| rs9650750 | GARNL3 | √ | × | × | |
| rs10760481 | GARNL3 | √ | × | × | |
| rs10987603 | GARNL3 | √ | × | × | |
| rs7037767 | GARNL3 | √ | × | × | |
| rs56128487 | GARNL3 | √ | × | × | |
| rs10819279 | GARNL3 | √ | × | × | |
| rs10819280 | GARNL3 | √ | × | × | |
| rs3802369 | GARNL3 | √ | × | × | |
| rs7871533 | GARNL3 | √ | × | × | |
| rs872010 | GAS8 | √ | × | × | |
| rs2300422 | GCLC | √ | × | × | |
| rs79038744 | GNB5 | √ | × | × | |
| rs12902073 | GNB5 | √ | × | × | |
| rs12901730 | GNB5 | √ | × | × | |
| rs12901891 | GNB5 | √ | × | × | |
| rs12438743 | GNB5 | √ | × | × | |
| rs2076519 | GNL2 | √ | × | × | |
| rs143515658 | GNPAT | × | √ | × | |
| rs11558492 | GNPAT | × | √ | × | |
| rs578945 | GNPAT | × | √ | × | |
| rs116203134 | GNPAT | × | √ | × | |
| rs117777700 | GNPAT | × | √ | × | |
| rs118166232 | GNPAT | × | √ | × | |
| rs1982395 | GPR56 | √ | × | × | |
| rs1868677 | GPR56 | √ | × | × | |
| rs1868678 | GPR56 | √ | × | × | |
| rs9938102 | GPR56 | √ | × | × | |
| rs4471674 | GPR56 | √ | × | × | |
| rs8058865 | GPR56 | √ | × | × | |
| rs935746 | GPR56 | √ | × | × | |
| rs1965226 | GPR56 | √ | × | × | |
| rs4784838 | GPR56 | × | √ | × | |
| rs8051879 | GPR97 | × | √ | × | |
| rs9928982 | GPR97 | √ | × | × | |
| rs72795504 | GPR97 | √ | × | × | |
| rs2006654 | GPR97 | √ | × | × | |
| rs8045126 | GPR97 | √ | × | × | |
| rs1064326 | GPR97 | √ | × | × | |
| rs553659110 | GRB10 | √ | × | × | |
| rs13236710 | GRB10 | √ | × | × | Height |
| rs55842511 | GRB10 | √ | × | × | |
| rs17152120 | GRB10 | √ | × | × | Height |
| rs35574823 | GRB10 | √ | × | × | |
| rs61552283 | GRB10 | √ | × | × | |
| rs740155 | GRB10 | √ | × | × | |
| rs2286153 | GRB10 | √ | × | × | |
| rs2074723 | GRB10 | √ | × | × | |
| rs56852197 | GRB10 | √ | × | × | |
| rs4947861 | GRB10 | √ | × | × | |
| rs7805310 | GRB10 | √ | × | × | |
| rs7790423 | GRB10 | √ | × | × | |
| rs60877714 | GRB10 | √ | × | × | |
| rs10510056 | GRK5 | √ | × | × | |
| rs534603664 | GRK7 | √ | × | × | |
| rs12544672 | GSDMC | √ | × | × | Hip circumference adjuste |
| rs900367 | GSDMC | √ | × | × | |
| rs72718335 | GSDMC | √ | × | × | |
| rs62524986 | GSDMC | √ | × | × | |
| rs12618809 | HDLBP | √ | × | × | |
| rs187669850 | HECW1 | √ | × | × | |
| rs35551485 | HECW1 | √ | × | × | |
| rs1047589 | HES6 | √ | × | × | |
| rs2290973 | HHATL | √ | × | × | |
| rs78256041 | HLX-AS1 | √ | × | × | |
| rs589925 | HMBS | × | √ | × | |
| rs60401382 | HTRA1 | × | √ | × | Stroke |
| rs2672594 | HTRA1 | × | √ | × | |
| rs11200643 | HTRA1 | × | √ | × | Height |
| rs12252027 | HTRA1 | × | √ | × | Headache or migraine |
| rs2672592 | HTRA1 | × | √ | × | Migraine |
| rs116921205 | HTRA1 | × | √ | × | |
| rs11200644 | HTRA1 | × | √ | × | |
| rs75150595 | HTRA1 | × | √ | × | |
| rs932275 | HTRA1 | × | √ | × | |
| rs80143083 | HTRA1 | × | √ | × | |
| rs12259258 | HTRA1 | × | √ | × | |
| rs72834448 | HTRA1 | × | √ | × | Systolic blood pressure |
| rs377694527 | HTRA1 | × | √ | × | |
| rs80232578 | HTRA1 | × | √ | × | |
| rs2736914 | HTRA1 | × | √ | × | |

| | | | | | |
|---|---|---|---|---|---|
| rs57415209 | PACRG | × | √ | × | |
| rs34603926 | PACRG | × | √ | × | |
| rs9347696 | PACRG | × | √ | × | |
| rs36944730 | PACRG | × | √ | × | |
| rs79405081 | PACRG | × | √ | × | |
| rs56250752 | PACRG | × | √ | × | |
| rs9365498 | PACRG | × | √ | × | |
| rs1333961 | PACRG | × | √ | × | |
| rs1333962 | PACRG | × | √ | × | |
| rs12527960 | PACRG | × | √ | × | |
| rs36967989 | PACRG | × | √ | × | |
| rs76658474 | PACRG | × | √ | × | |
| rs13202482 | PACRG | × | √ | × | |
| rs78541421 | PACRG | × | √ | × | |
| rs9365499 | PACRG | × | √ | × | Body mass |
| rs13209887 | PACRG | × | √ | × | |
| rs76742106 | PACRG | × | √ | × | |
| rs74354308 | PACRG | × | √ | × | |
| rs18757616 | PACRG | × | √ | × | |
| rs9356068 | PACRG | × | √ | × | |
| rs9356069 | PACRG | × | √ | × | |
| rs79118964 | PAFAH1B2 | √ | × | × | |
| rs12581006 | PCED1B | √ | × | × | |
| rs4768776 | PCED1B | √ | × | × | |
| rs10881052 | PCED1B | √ | × | × | |
| rs1525452 | PCED1B | √ | × | × | |
| rs36124720 | PER2 | √ | × | × | |
| rs80049518 | PLCL2 | √ | × | × | |
| rs3742868 | PLEKHH1 | √ | × | × | |
| rs7141944 | PLEKHH1 | √ | × | × | |
| rs16843395 | PPP1R7 | √ | × | × | |
| rs80209508 | PPP1R9A | √ | × | × | |
| rs79686315 | PRSS3 | √ | × | × | |
| rs79665161 | PRSS3 | √ | × | × | |
| rs97445685 | PRSS3 | √ | × | × | |
| rs87920364 | PRSS3 | √ | × | × | |
| rs79644971 | PRSS3 | √ | × | × | |
| rs79653051 | PRSS3 | √ | × | × | |
| rs37738026 | PRSS3 | √ | × | × | |
| rs11774278 | PSD3 | √ | × | × | |
| rs11780329 | PSD3 | √ | × | × | |
| rs56277438 | PSD3 | √ | × | × | |
| rs73204530 | PSD3 | √ | × | × | |
| rs1426917 | PSD3 | √ | × | × | |
| rs73204534 | PSD3 | √ | × | × | |
| rs11783114 | PSD3 | √ | × | × | |
| rs62580804 | RALGPS1 | × | √ | × | |
| rs3936030 | RALGPS1 | × | √ | × | |
| rs62580836 | RALGPS1 | × | √ | × | |
| rs17368219 | RALGPS1 | × | √ | × | |
| rs17446684 | RALGPS1 | × | √ | × | |
| rs11720739 | RALGPS1 | × | √ | × | |
| rs13791031 | RALGPS1 | × | √ | × | |
| rs78889029 | RALGPS1 | × | √ | × | |
| rs826532 | RANBP2 | √ | × | × | |
| rs1724203 | RANBP2 | √ | × | × | |
| rs3782445 | RBM19 | √ | × | × | |
| rs9854636 | RBMS3 | √ | × | × | |
| rs9837143 | RBMS3 | √ | × | × | |
| rs9372862 | RNF146 | √ | × | × | |
| rs9388559 | RNF146 | √ | × | × | |
| rs9321072 | RNF146 | √ | × | × | |
| rs2241542 | SEC22C | × | √ | × | |
| rs342535 | SEC22C | × | √ | × | |
| rs77920516 | SIK3 | √ | × | × | |
| rs4652958 | SNIP1 | √ | × | × | |
| rs2273013 | SNIP1 | √ | × | × | |
| rs10842252 | SOX5 | √ | × | × | |
| rs11047179 | SOX5 | √ | × | × | |
| rs76847583 | SOX5 | √ | × | × | |
| rs11047185 | SOX5 | √ | × | × | |
| rs7138457 | SOX5 | √ | × | × | |
| rs12582243 | SOX5 | √ | × | × | |
| rs3803688 | SPIRE2 | × | × | √ | |
| rs13146675 | SPOCK3 | √ | × | × | |
| rs7691894 | SPOCK3 | √ | × | × | |
| rs28469298 | SPOCK3 | √ | × | × | |
| rs2219234 | SPOCK3 | √ | × | × | |
| rs3828018 | SPOCK3 | √ | × | × | |
| rs3790358 | SPOCK3 | √ | × | × | |
| rs28699632 | SPOCK3 | √ | × | × | |
| rs7697656 | SPOCK3 | √ | × | × | |
| rs7693595 | SPOCK3 | √ | × | × | |
| rs17645162 | SPOCK3 | √ | × | × | |
| rs10517901 | SPOCK3 | √ | × | × | |
| rs17645192 | SPOCK3 | √ | × | × | |
| rs2305593 | SPOCK3 | √ | × | × | |
| rs11139828 | SPRTN | × | √ | × | |
| rs78658157 | SPRTN | × | √ | × | |
| rs11719208 | SPRTN | × | √ | × | |
| rs11747319 | SPRTN | × | √ | × | |
| rs11729859 | SPRTN | × | √ | × | |
| rs11122273 | SPRTN | × | √ | × | |
| rs7414807 | SPRTN | × | √ | × | |
| rs2355873 | SPRTN | × | √ | × | |
| rs17312483 | SYNPR | × | √ | × | |
| rs19222998 | TACC2 | × | √ | × | |
| rs14707546 | TACC2 | × | √ | × | |
| rs4752665 | TACC2 | × | √ | × | |
| rs72840870 | TACC2 | × | √ | × | |
| rs7920029 | TACC2 | × | √ | × | |
| rs933821 | TBC1D1 | √ | × | × | |
| rs35308860 | TBC1D1 | √ | × | × | |
| rs67733859 | TBC1D1 | √ | × | × | |
| rs59608961 | TBC1D1 | √ | × | × | |
| rs35021061 | TBC1D1 | √ | × | × | |
| rs12649018 | TBC1D1 | √ | × | × | |
| rs1364953 | TBC1D1 | √ | × | × | Metabolite l |
| rs2389414 | TBC1D32 | √ | × | × | |
| rs9320800 | TBC1D32 | √ | × | × | |
| rs7908570 | TET1 | √ | × | × | |
| rs11733022 | THRB | √ | × | × | |
| rs11199028 | TIAL1 | √ | × | × | |
| rs11199030 | TIAL1 | √ | × | × | |
| rs37517233 | TMEM117 | × | √ | × | |
| rs54328592 | TMEM117 | × | √ | × | |
| rs11182331 | TMEM117 | × | √ | × | |
| rs15052828 | TMEM117 | × | √ | × | |
| rs11182334 | TMEM117 | × | √ | × | |
| rs11742227 | TMEM117 | × | √ | × | |
| rs13792863 | TMEM117 | × | √ | × | |
| rs55672947 | TMEM117 | × | √ | × | |
| rs53984381 | TMEM117 | × | √ | × | |
| rs11182340 | TMEM117 | × | √ | × | |
| rs76302415 | TMEM117 | × | √ | × | |
| rs11688455 | TMEM117 | × | √ | × | |
| rs19994260 | TMEM117 | × | √ | × | |
| rs76096911 | TMEM117 | × | √ | × | |
| rs11829716 | TMEM117 | × | √ | × | |
| rs73085705 | TMEM117 | × | √ | × | |
| rs7953173 | TMEM117 | √ | × | × | |
| rs7967957 | TMEM117 | √ | × | × | |
| rs10506234 | TMEM117 | √ | × | × | |
| rs55782485 | TMEM117 | √ | × | × | |
| rs55977905 | TMEM117 | √ | × | × | |
| rs7968777 | TMEM117 | √ | × | × | |
| rs1168078 | TMEM117 | √ | × | × | |
| rs7980383 | TMEM117 | √ | × | × | |
| rs6582498 | TMEM117 | √ | × | × | |
| rs11613902 | TMEM117 | √ | × | × | |
| rs2638855 | TMEM117 | √ | × | × | |
| rs78459717 | TMEM117 | √ | × | × | |
| rs73288040 | TMEM117 | √ | × | × | |
| rs11837611 | TMEM117 | √ | × | × | |
| rs3780698 | TOR1A | √ | × | × | |
| rs2296793 | TOR1A | √ | × | × | Bone miner |
| rs10988519 | TOR1B | √ | × | × | |
| rs10116248 | TOR1B | √ | × | × | |
| rs2302820 | TOR1B | √ | × | × | |
| rs12107025 | TP63 | √ | × | × | |
| rs9647365 | TP63 | √ | × | × | |
| rs61943604 | TPCN1 | √ | × | × | |
| rs7135901 | TPCN1 | √ | × | × | |
| rs7300258 | TPCN1 | √ | × | × | |
| rs7314153 | TPCN1 | √ | × | × | |
| rs2303618 | TPCN1 | √ | × | × | |
| rs2303617 | TPCN1 | √ | × | × | |
| rs7955036 | TPCN1 | √ | × | × | |
| rs7955400 | TPCN1 | √ | × | × | |
| rs2288365 | TPCN1 | √ | × | × | |
| rs12405 | TPCN1 | √ | × | × | |
| rs6569329 | TRDN | √ | × | × | |
| rs58301661 | TRDN | √ | × | × | |
| rs9388248 | TRDN | √ | × | × | |
| rs9385300 | TRDN | √ | × | × | |
| rs4460222 | TRDN | √ | × | × | |
| rs9375257 | TRDN | √ | × | × | |
| rs9388251 | TRDN | √ | × | × | |
| rs9398732 | TRDN | √ | × | × | |
| rs12528126 | TRDN | √ | × | × | |
| rs9398733 | TRDN | √ | × | × | |
| rs13193333 | TRDN | √ | × | × | |
| rs12192602 | TRDN | √ | × | × | |
| rs9401680 | TRDN | √ | × | × | |
| rs6939909 | TRDN | √ | × | × | |
| rs4996638 | TRDN | √ | × | × | |
| rs963811 | TRIM44 | √ | × | × | |
| rs1830816 | TRIM44 | √ | × | × | |
| rs1007591 | TRIM44 | √ | × | × | |
| rs2985418 | TRIM44 | √ | × | × | |
| rs14735745 | TRIM44 | √ | × | × | |
| rs12226318 | TRIM44 | √ | × | × | |
| rs7129170 | TRIM44 | √ | × | × | Eicosenoylc |
| rs74716568 | TRIM44 | √ | × | × | |
| rs1969800 | TRIM44 | √ | × | × | |
| rs61881279 | TRIM44 | √ | × | × | |
| rs2970329 | TRIM44 | √ | × | × | |
| rs16927901 | TRIM44 | √ | × | × | |
| rs68063352 | TRIM44 | √ | × | × | |
| rs12363437 | TRIM44 | √ | × | × | |
| rs12629685 | TRIM71 | √ | × | × | |
| rs11745660 | TUBB3 | × | √ | × | |
| rs54174424 | TUBB3 | × | √ | × | |
| rs18244275 | TUBB3 | × | √ | × | |
| rs59001570 | TUBB3 | × | √ | × | |
| rs11456676 | TUBB3 | × | √ | × | |
| rs57807714 | TUBB3 | × | √ | × | |
| rs58336812 | TUBB3 | × | √ | × | |
| rs28521461 | TUBB3 | × | √ | × | |
| rs37019719 | TUBB3 | × | √ | × | |
| rs14470718 | TUBB3 | × | √ | × | |
| rs14851307 | TUBB3 | × | √ | × | |
| rs18544785 | TUBB3 | × | √ | × | |
| rs76996117 | UAP1 | √ | × | × | |
| rs62555936 | UBAP2 | √ | × | × | |
| rs16935299 | UBAP2 | √ | × | × | |
| rs3739690 | UBAP2 | √ | × | × | |
| rs62555940 | UBAP2 | √ | × | × | |
| rs307697 | UBAP2 | √ | × | × | |
| rs12553803 | UBAP2 | √ | × | × | |
| rs10971836 | UBAP2 | √ | × | × | |
| rs10971837 | UBAP2 | √ | × | × | |
| rs10758235 | UBAP2 | √ | × | × | |
| rs56404908 | UBAP2 | √ | × | × | |
| rs7044541 | UBAP2 | √ | × | × | |
| rs932969 | UBAP2 | √ | × | × | |
| rs10122409 | UBAP2 | √ | × | × | |
| rs10814059 | UBAP2 | √ | × | × | |
| rs7029112 | UBAP2 | √ | × | × | |
| rs7029122 | UBAP2 | √ | × | × | |
| rs10758236 | UBAP2 | √ | × | × | |
| rs13300356 | UBAP2 | √ | × | × | |
| rs12236865 | UBAP2 | √ | × | × | |
| rs6771166 | UBE2E1 | √ | × | × | |
| rs34919006 | UBE2E2 | √ | × | × | |
| rs9809176 | UBE2E2 | √ | × | × | |
| rs4858484 | UBE2E2 | √ | × | × | |
| rs9841944 | UBE2E2 | √ | × | × | |
| rs6550748 | UBE2E2 | √ | × | × | |
| rs150122 | ULK2 | √ | × | × | |
| rs157408 | ULK2 | √ | × | × | |
| rs157401 | ULK2 | √ | × | × | |
| rs157402 | ULK2 | √ | × | × | |
| rs157403 | ULK2 | √ | × | × | |
| rs169460 | ULK2 | √ | × | × | |
| rs157395 | ULK2 | √ | × | × | |
| rs157396 | ULK2 | √ | × | × | |
| rs205103 | ULK2 | √ | × | × | |
| rs205104 | ULK2 | √ | × | × | |

| | | | | | |
|---|---|---|---|---|---|
| rs11033261 | TRIM44 | √ | × | × | |
| rs11694335 | TRIM44 | √ | × | × | |
| rs3112061 | TRIM44 | √ | × | × | |
| rs9633874 | TRIM44 | √ | × | × | |
| rs77770625 | TRIM44 | √ | × | × | |
| rs11033265 | TRIM44 | √ | × | × | |
| rs307658 | UBAP2 | √ | × | × | |
| rs307678 | UBAP2 | √ | × | × | |
| rs307683 | UBAP2 | √ | × | × | |
| rs4272470 | UBAP2 | √ | × | × | |
| rs834027 | UBAP2 | √ | × | × | |
| rs77191623 | ULK2 | √ | × | × | |
| rs75030060 | UNQ6494 | √ | × | × | |
| rs78401801 | UNQ6494 | √ | × | × | |
| rs78343805 | UNQ6494 | √ | × | × | Estimated g |
| rs7033256 | UNQ6494 | √ | × | × | |
| rs2175476 | WWC2 | √ | × | × | |
| rs7677983 | WWC2 | √ | × | × | |
| rs62122448 | ZNF787 | √ | × | × | |
| rs13325966 | ACAP2 | √ | × | × | |
| rs10212206 | ACAP2 | √ | × | × | |
| rs6788868 | ACAP2 | √ | × | × | |
| rs1214562 | ANKRD22 | √ | × | × | |
| rs4934425 | ANKRD22 | √ | × | × | Height |
| rs2654991 | ANO4 | √ | × | × | |
| rs589657 | ANO4 | √ | × | × | |
| rs225299 | AP2B1 | √ | × | × | |
| rs225293 | AP2B1 | √ | × | × | |
| rs226441 | AP2B1 | √ | × | × | |
| rs226428 | AP2B1 | √ | × | × | |
| rs80149721 | APBB1IP | √ | × | × | |
| rs7915768 | ARHGAP21 | √ | × | × | |
| rs7474901 | ARHGAP21 | √ | × | × | |
| rs12220759 | ARHGAP21 | √ | × | × | |
| rs11098219 | ARSJ | √ | × | × | Height |
| rs6827199 | ARSJ | √ | × | × | Height |
| rs12498542 | ARSJ | √ | × | × | |
| rs12686257 | AUH | √ | × | × | |
| rs10991840 | AUH | √ | × | × | |
| rs1757648 | AUH | √ | × | × | |
| rs2380165 | BLM | √ | × | × | |
| rs12511954 | CAMK2D | √ | × | × | |
| rs6842886 | CAMK2D | √ | × | × | |
| rs12507550 | CAMK2D | √ | × | × | |
| rs1589911 | CCDC138 | √ | × | × | |
| rs73952523 | CCDC138 | √ | × | × | |
| rs57043792 | CCDC138 | √ | × | × | |
| rs75324970 | CCDC138 | √ | × | × | |
| rs2279522 | DPYSL4 | √ | × | × | |
| rs72961085 | ECHDC1 | √ | × | × | |
| rs7747926 | ELOVL5 | √ | × | × | |
| rs209498 | ELOVL5 | √ | × | × | |
| rs7738788 | ELOVL5 | √ | × | × | |
| rs9340782 | ESR1 | √ | × | × | |
| rs7023663 | FAM102A | √ | × | × | |
| rs943392 | FAM102A | √ | × | × | |
| rs727042 | FAM155A | √ | × | × | |
| rs14639341 | FAM49B | √ | × | × | |
| rs14436875 | FAM49B | √ | × | × | |
| rs35612917 | FOCAD | √ | × | × | |
| rs13297201 | FOCAD | √ | × | × | |
| rs78618914 | IARS | √ | × | × | |
| rs77414536 | KIFC3 | √ | × | × | |
| rs79949875 | KIFC3 | √ | × | × | |
| rs75594839 | LOC101927 | × | √ | × | |
| rs77950503 | LOC101927 | × | √ | × | |
| rs78495315 | LOC101927 | × | √ | × | |
| rs7590275 | METTL8 | √ | × | × | |
| rs74600707 | MIR548AC | √ | × | × | |
| rs7640308 | MIR548AC | √ | × | × | |
| rs78757102 | MSR1 | √ | × | × | |
| rs12675748 | MSR1 | √ | × | × | |
| rs62491233 | MSR1 | √ | × | × | |
| rs78765574 | MSR1 | √ | × | × | |
| rs3774401 | NKTR | √ | × | × | |
| rs367471 | NLRP8 | √ | × | × | |
| rs9347662 | PARK2 | √ | × | × | |
| rs9356044 | PARK2 | √ | × | × | |
| rs9365460 | PARK2 | √ | × | × | |
| rs9356045 | PARK2 | √ | × | × | |
| rs9356047 | PARK2 | √ | × | × | |
| rs9356049 | PARK2 | √ | × | × | |
| rs9295192 | PARK2 | √ | × | × | |
| rs9295193 | PARK2 | √ | × | × | |
| rs2023016 | PARK2 | √ | × | × | Educational |
| rs10945841 | PARK2 | √ | × | × | |
| rs9364662 | PARK2 | √ | × | × | |
| rs9346922 | PARK2 | √ | × | × | |
| rs9365464 | PARK2 | √ | × | × | |
| rs9458597 | PARK2 | √ | × | × | |
| rs12194834 | PARK2 | √ | × | × | |
| rs10945842 | PARK2 | √ | × | × | |
| rs4709640 | PARK2 | √ | × | × | |
| rs4709641 | PARK2 | √ | × | × | |
| rs9365467 | PARK2 | √ | × | × | |
| rs9458612 | PARK2 | √ | × | × | |
| rs10945854 | PARK2 | √ | × | × | |
| rs9634259 | PCED1B | √ | × | × | |
| rs11183766 | PCED1B | √ | × | × | |
| rs10151023 | PIGH | √ | × | × | Adolescent |
| rs2319844 | PIGH | √ | × | × | Adolescent |
| rs4565845 | PLEKHA1 | √ | × | × | |
| rs717144 | POLR1A | √ | × | × | |
| rs12534317 | PON3 | √ | × | × | |
| rs2057119 | RDH11 | √ | × | × | |
| rs12175928 | RNF146 | √ | × | × | |
| rs9388558 | RNF146 | √ | × | × | |
| rs12994122 | SNED1 | √ | × | × | |
| rs12422241 | SOX5 | √ | × | × | |
| rs59556093 | SOX5 | √ | × | × | |
| rs61517249 | SOX5 | √ | × | × | |
| rs11047191 | SOX5 | √ | × | × | |
| rs11047192 | SOX5 | √ | × | × | |
| rs28740811 | SOX5 | √ | × | × | |
| rs9668967 | SOX5 | √ | × | × | |
| rs11937180 | SPOCK3 | √ | × | × | |
| rs6827226 | SPOCK3 | √ | × | × | |
| rs10819410 | SPTAN1 | √ | × | × | |
| rs3763599 | SPTAN1 | √ | × | × | |
| rs10988042 | SPTAN1 | √ | × | × | |
| rs17849181 | STXBP1 | √ | × | × | |
| rs11754840 | STXBP1 | √ | × | × | |
| rs36803651 | TBC1D1 | √ | × | × | |
| rs75833459 | TBC1D1 | √ | × | × | |
| rs9401367 | TBC1D32 | √ | × | × | |
| rs2202065 | TBC1D32 | √ | × | × | |
| rs9490138 | TBC1D32 | √ | × | × | |
| rs9482123 | TBC1D32 | √ | × | × | |
| rs9320799 | TBC1D32 | √ | × | × | |
| rs9320801 | TBC1D32 | √ | × | × | |
| rs4585597 | TBC1D32 | √ | × | × | |
| rs3923664 | TBC1D32 | √ | × | × | |
| rs9387942 | TBC1D32 | √ | × | × | |
| rs74775566 | TBC1D32 | √ | × | × | |
| rs78455529 | TBC1D32 | √ | × | × | |
| rs76083652 | TBC1D32 | √ | × | × | |
| rs78187099 | TBC1D32 | √ | × | × | |
| rs4945683 | TBC1D32 | √ | × | × | |
| rs4946560 | TBC1D32 | √ | × | × | |
| rs17083385 | TBC1D32 | √ | × | × | |
| rs79274950 | TBC1D32 | √ | × | × | |
| rs9320803 | TBC1D32 | √ | × | × | |
| rs4412244 | TBC1D32 | √ | × | × | |
| rs9375022 | TBC1D32 | √ | × | × | |
| rs9398629 | TBC1D32 | √ | × | × | OTU99_45 |
| rs12230287 | TMEM117 | √ | × | × | |
| rs9385291 | TRDN | √ | × | × | |
| rs9401665 | TRDN | √ | × | × | |
| rs9401666 | TRDN | √ | × | × | |
| rs6941565 | TRDN | √ | × | × | |
| rs6922718 | TRDN | √ | × | × | |
| rs9375255 | TRDN | √ | × | × | |
| rs9372748 | TRDN | √ | × | × | |
| rs4576276 | TRDN | √ | × | × | |
| rs2185160 | TRDN | √ | × | × | |
| rs9320940 | TRDN | √ | × | × | |
| rs9490789 | TRDN | √ | × | × | |
| rs9490790 | TRDN | √ | × | × | |
| rs9490791 | TRDN | √ | × | × | |
| rs9490792 | TRDN | √ | × | × | |
| rs9398730 | TRDN | √ | × | × | |
| rs9398731 | TRDN | √ | × | × | |
| rs4509143 | TRDN | √ | × | × | |
| rs4509144 | TRDN | √ | × | × | |
| rs9385301 | TRDN | √ | × | × | |
| rs7746262 | TRDN | √ | × | × | |
| rs7746287 | TRDN | √ | × | × | |
| rs9320941 | TRDN | √ | × | × | |
| rs4370389 | TRDN | √ | × | × | |
| rs4523105 | TRDN | √ | × | × | |
| rs9401678 | TRDN | √ | × | × | |
| rs7452290 | TRDN | √ | × | × | |
| rs9401679 | TRDN | √ | × | × | |
| rs9375259 | TRDN | √ | × | × | |
| rs9375260 | TRDN | √ | × | × | |
| rs4897260 | TRDN | √ | × | × | |
| rs4612191 | TRDN | √ | × | × | |
| rs9490794 | TRDN | √ | × | × | |
| rs9375261 | TRDN | √ | × | × | |
| rs12055872 | TRDN | √ | × | × | |
| rs75164591 | TRDN | √ | × | × | |
| rs75083890 | TRDN | √ | × | × | |
| rs4391277 | TRDN | √ | × | × | |
| rs6929960 | TRDN | √ | × | × | |
| rs4998058 | TRDN | √ | × | × | |
| rs11154175 | TRDN | √ | × | × | |
| rs75126813 | TRDN | √ | × | × | |
| rs620602 | TRIM29 | √ | × | × | |
| rs12574811 | TRIM44 | √ | × | × | |
| rs1323856 | TRIM44 | √ | × | × | |
| rs34103243 | TRIM44 | √ | × | × | |
| rs2985152 | TRIM44 | √ | × | × | |
| rs80330422 | TRIM44 | √ | × | × | |
| rs7862440 | TRPM3 | √ | × | × | |
| rs1785506 | UBAP2 | √ | × | × | |
| rs1758632 | UBAP2 | √ | × | × | Type 2 diab |
| rs1785512 | UBAP2 | √ | × | × | |
| rs1785509 | UBAP2 | √ | × | × | |
| rs1760816 | UBAP2 | √ | × | × | |
| rs1629188 | UBAP2 | √ | × | × | |
| rs1785507 | UBAP2 | √ | × | × | |
| rs10814062 | UBAP2 | √ | × | × | |
| rs921095 | UBE2F | √ | × | × | |
| rs62194510 | UBE2F | √ | × | × | Educational |
| rs11849521 | VTI1B | √ | × | × | |
| rs17782603 | VTI1B | √ | × | × | |
| rs4902507 | VTI1B | √ | × | × | |
| rs12859864 | WBP4 | √ | × | × | |
| rs41457144 | WWC2 | √ | × | × | |
| rs73006721 | WWC2 | √ | × | × | |
| rs73006723 | WWC2 | √ | × | × | |
| rs73006725 | WWC2 | √ | × | × | |
| rs73006729 | WWC2 | √ | × | × | |
| rs11932929 | WWC2 | √ | × | × | |
| rs17074589 | WWC2 | √ | × | × | |
| rs17074592 | WWC2 | √ | × | × | |
| rs17074601 | WWC2 | √ | × | × | |
| rs55851513 | ZNF335 | √ | × | × | |
| rs6437373 | ACAP2 | √ | × | × | |
| rs9854941 | ACAP2 | √ | × | × | |
| rs11774778 | ACAP2 | √ | × | × | |
| rs1538768 | ACAP2 | √ | × | × | |
| rs60177727 | ACAP2 | √ | × | × | |
| rs6801315 | ACAP2 | √ | × | × | |
| rs9879305 | ACAP2 | √ | × | × | |
| rs9809730 | ACAP2 | √ | × | × | |
| rs28375276 | ACAP2 | √ | × | × | |
| rs36913970 | ACAP2 | √ | × | × | |
| rs76190566 | AGAP1 | √ | × | × | |
| rs68016591 | AGAP1 | √ | × | × | |
| rs2437807 | AGBL1 | √ | × | × | |
| rs2437802 | AGBL1 | √ | × | × | |
| rs62033661 | AGBL1 | √ | × | × | |
| rs4887494 | AGBL1 | √ | × | × | |
| rs2447254 | AGBL1 | √ | × | × | |
| rs4377131 | AGBL1 | √ | × | × | |
| rs4441241 | AGBL1 | √ | × | × | |
| rs15036713 | AGBL1 | √ | × | × | |
| rs11687458 | AGBL1 | √ | × | × | |

| | | | | |
|---|---|---|---|---|
| rs10785475 | TMEM117 | √ | × | |
| rs10785476 | TMEM117 | √ | × | |
| rs10785477 | TMEM117 | √ | × | |
| rs1881927 | TMEM117 | √ | × | |
| rs1881926 | TMEM117 | √ | × | |
| rs7974214 | TMEM117 | √ | × | |
| rs10785478 | TMEM117 | √ | × | |
| rs10785479 | TMEM117 | √ | × | |
| rs11182337 | TMEM117 | √ | × | |
| rs53984381 | TMEM117 | × | √ | |
| rs11182340 | TMEM117 | × | √ | |
| rs75410353 | TMEM117 | × | √ | |
| rs76302415 | TMEM117 | × | √ | |
| rs12303804 | TMEM117 | × | √ | |
| rs11688455 | TMEM117 | × | √ | |
| rs19994260 | TMEM117 | × | √ | |
| rs76096911 | TMEM117 | × | √ | |
| rs11829716 | TMEM117 | × | √ | |
| rs73085705 | TMEM117 | × | √ | |
| rs12230287 | TMEM117 | × | √ | |
| rs14492975 | TMEM117 | × | √ | |
| rs1182 | TOR1A | √ | × | Prostate cancer |
| rs11787741 | TOR1A | √ | × | |
| rs3780696 | TOR1A | √ | × | |
| rs3780697 | TOR1A | √ | × | |
| rs3780698 | TOR1A | √ | × | |
| rs10988519 | TOR1B | √ | × | |
| rs10116248 | TOR1B | √ | × | |
| rs2302820 | TOR1B | √ | × | |
| rs11789668 | TOR1B | √ | × | |
| rs12634 | TOR1B | √ | × | Prostate cancer |
| rs2303617 | TPCN1 | √ | × | |
| rs361508 | TRDN | √ | × | |
| rs6569329 | TRDN | √ | × | |
| rs6569330 | TRDN | √ | × | |
| rs6920186 | TRDN | √ | × | |
| rs9320922 | TRDN | √ | × | |
| rs9482369 | TRDN | √ | × | |
| rs7754306 | TRDN | √ | × | |
| rs7764953 | TRDN | √ | × | |
| rs9385291 | TRDN | √ | × | |
| rs58301661 | TRDN | √ | × | |
| rs9401658 | TRDN | √ | × | |
| rs1346921 | TRDN | √ | × | |
| rs1808620 | TRDN | √ | × | |
| rs9372743 | TRDN | √ | × | |
| rs9375246 | TRDN | √ | × | |
| rs9385298 | TRDN | √ | × | |
| rs9401659 | TRDN | √ | × | |
| rs9372745 | TRDN | √ | × | |
| rs9388227 | TRDN | √ | × | |
| rs9320928 | TRDN | √ | × | |
| rs9375248 | TRDN | √ | × | |
| rs9401661 | TRDN | √ | × | |
| rs9320929 | TRDN | √ | × | |
| rs2060063 | TRDN | √ | × | |
| rs9401665 | TRDN | √ | × | |
| rs9401666 | TRDN | √ | × | |
| rs9388248 | TRDN | √ | × | |
| rs4460222 | TRDN | √ | × | |
| rs7751903 | TRDN | √ | × | |
| rs6941565 | TRDN | √ | × | |
| rs62420493 | TRDN | √ | × | |
| rs34957028 | TRDN | √ | × | |
| rs9375255 | TRDN | √ | × | |
| rs4576276 | TRDN | √ | × | |
| rs9320940 | TRDN | √ | × | |
| rs9490789 | TRDN | √ | × | |
| rs9490790 | TRDN | √ | × | |
| rs9490791 | TRDN | √ | × | |
| rs9490792 | TRDN | √ | × | |
| rs9398730 | TRDN | √ | × | |
| rs9398731 | TRDN | √ | × | |
| rs4509143 | TRDN | √ | × | |
| rs4509144 | TRDN | √ | × | |
| rs9385301 | TRDN | √ | × | |
| rs7746262 | TRDN | √ | × | |
| rs7746287 | TRDN | √ | × | |
| rs9320941 | TRDN | √ | × | |
| rs4370389 | TRDN | √ | × | |
| rs4523105 | TRDN | √ | × | |
| rs9401678 | TRDN | √ | × | |
| rs4897260 | TRDN | √ | × | |
| rs4612191 | TRDN | √ | × | |
| rs9490794 | TRDN | √ | × | |
| rs620602 | TRIM29 | √ | × | |
| rs963811 | TRIM44 | √ | × | |
| rs1830816 | TRIM44 | √ | × | |
| rs2985418 | TRIM44 | √ | × | |
| rs14735745 | TRIM44 | √ | × | |
| rs12226318 | TRIM44 | √ | × | |
| rs7129170 | TRIM44 | √ | × | Eicosenoylcarnitine (C20:1) levels in elite athletes |
| rs12574811 | TRIM44 | √ | × | |
| rs1323856 | TRIM44 | √ | × | |
| rs1359181 | TRIM44 | √ | × | |
| rs34103243 | TRIM44 | √ | × | |
| rs1969800 | TRIM44 | √ | × | |
| rs61881279 | TRIM44 | √ | × | |
| rs60608746 | TRIM44 | √ | × | |
| rs2970329 | TRIM44 | √ | × | |
| rs7929977 | TRIM44 | √ | × | |
| rs16927901 | TRIM44 | √ | × | |
| rs68063352 | TRIM44 | √ | × | |
| rs80330422 | TRIM44 | √ | × | |
| rs12289152 | TRIM44 | √ | × | |
| rs6560172 | TRPM3 | √ | × | |
| rs72614629 | TRPM3 | √ | × | |
| rs62555936 | UBAP2 | √ | × | |
| rs16935299 | UBAP2 | √ | × | |
| rs12553803 | UBAP2 | √ | × | |
| rs10971837 | UBAP2 | √ | × | |
| rs10758235 | UBAP2 | √ | × | |
| rs7044541 | UBAP2 | √ | × | |
| rs7030063 | UBAP2 | √ | × | |
| rs932969 | UBAP2 | √ | × | |
| rs10122409 | UBAP2 | √ | × | |
| rs10814059 | UBAP2 | √ | × | |
| rs7029112 | UBAP2 | √ | × | |
| rs10758236 | UBAP2 | √ | × | |
| rs13300356 | UBAP2 | √ | × | |
| rs1785506 | UBAP2 | √ | × | |
| rs1785503 | UBAP2 | √ | × | |
| rs1785502 | UBAP2 | √ | × | |
| rs1785500 | UBAP2 | √ | × | |
| rs1758632 | UBAP2 | √ | × | Type 2 diabetes |
| rs1785512 | UBAP2 | √ | × | |
| rs1758633 | UBAP2 | √ | × | |
| rs1785509 | UBAP2 | √ | × | |
| rs1760816 | UBAP2 | √ | × | |
| rs1629188 | UBAP2 | √ | × | |
| rs1785507 | UBAP2 | √ | × | |
| rs10814062 | UBAP2 | √ | × | |
| rs1471061 | UBE2E1 | √ | × | |
| rs7513662 | UHMK1 | √ | × | OTU99_123 (Phascolarctobacterium) prevalence |
| rs157401 | ULK2 | √ | × | |
| rs157402 | ULK2 | √ | × | |
| rs169460 | ULK2 | √ | × | |
| rs157395 | ULK2 | √ | × | |
| rs157396 | ULK2 | √ | × | |
| rs205103 | ULK2 | √ | × | |
| rs205104 | ULK2 | √ | × | |
| rs205105 | ULK2 | √ | × | |
| rs3101815 | ULK2 | √ | × | |
| rs421561 | ULK2 | √ | × | |
| rs281357 | ULK2 | √ | × | |
| rs281354 | ULK2 | √ | × | |
| rs75871414 | ULK2 | √ | × | |
| rs188138 | ULK2 | √ | × | |
| rs281346 | ULK2 | √ | × | |
| rs7176603 | UNC13C | √ | × | |
| rs11265860 | UNQ6494 | √ | × | |
| rs3744793 | USP36 | √ | × | |
| rs9841214 | VIPR1 | √ | × | |
| rs2267538 | VIPR1 | √ | × | |
| rs2239499 | VIPR1 | √ | × | |
| rs2239500 | VIPR1 | √ | × | |
| rs2276829 | VIPR1 | √ | × | |
| rs66942973 | VIPR2 | √ | × | |
| rs11849521 | VTI1B | √ | × | |
| rs17782603 | VTI1B | √ | × | |
| rs4902507 | VTI1B | √ | × | |
| rs12859864 | WBP4 | √ | × | |
| rs41457144 | WWC2 | √ | × | |
| rs73006721 | WWC2 | √ | × | |
| rs73006723 | WWC2 | √ | × | |
| rs73006725 | WWC2 | √ | × | |
| rs73006729 | WWC2 | √ | × | |
| rs11932929 | WWC2 | √ | × | |
| rs17074589 | WWC2 | √ | × | |
| rs17074592 | WWC2 | √ | × | |
| rs17074601 | WWC2 | √ | × | |
| rs13892733 | ZBTB43 | √ | × | |
| rs1008275 | ZFHX3 | × | √ | |
| rs62122411 | ZNF787 | √ | × | |
| rs19112031 | ZSCAN5B | × | √ | |
| rs17753438 | ZSCAN5B | × | √ | |
| rs76381877 | ACAP2 | √ | × | |
| rs74694161 | ACAP2 | √ | × | |
| rs9854941 | ACAP2 | √ | × | |
| rs1538768 | ACAP2 | √ | × | |
| rs60177727 | ACAP2 | √ | × | |
| rs6801315 | ACAP2 | √ | × | |
| rs9879305 | ACAP2 | √ | × | |
| rs9809730 | ACAP2 | √ | × | |
| rs6763335 | ACAP2 | √ | × | |
| rs13317705 | ACAP2 | √ | × | |
| rs62288360 | ACAP2 | √ | × | |
| rs77069909 | AGAP1 | √ | × | |
| rs55650317 | AGBL1 | √ | × | |
| rs62033661 | AGBL1 | √ | × | |
| rs7183919 | AGBL1 | √ | × | |
| rs75476988 | AGBL1 | √ | × | |
| rs58001144 | AGBL1 | √ | × | |
| rs78868610 | AGBL1 | √ | × | |
| rs4887494 | AGBL1 | √ | × | |
| rs74517834 | AGBL1 | √ | × | |
| rs11805173 | AGBL1 | √ | × | |
| rs4887244 | AGBL1 | √ | × | |
| rs13380199 | AGBL1 | √ | × | |
| rs16978012 | AGBL1 | √ | × | |
| rs2347248 | AGBL1 | √ | × | |
| rs2881583 | AGBL1 | √ | × | |
| rs76309253 | AGBL1 | √ | × | |
| rs16978052 | AGBL1 | √ | × | |
| rs16978054 | AGBL1 | √ | × | |
| rs57116355 | AGBL1 | √ | × | |
| rs74027145 | AGBL1 | √ | × | |
| rs76422866 | AGBL1 | √ | × | |
| rs7146912 | AKAP6 | √ | × | |
| rs7147629 | AKAP6 | √ | × | |
| rs6571518 | AKAP6 | √ | × | |
| rs8013463 | AKAP6 | √ | × | |
| rs1951191 | AKAP6 | √ | × | |
| rs3784193 | AKAP6 | √ | × | |
| rs4329831 | AKAP6 | √ | × | |
| rs79687473 | ALDH4A1 | × | √ | |
| rs4912075 | ALDH4A1 | × | √ | |
| rs4911985 | ALDH4A1 | × | √ | |
| rs74999630 | ALDH4A1 | × | √ | |
| rs78551322 | ALDH4A1 | × | √ | |
| rs57372319 | ALDH4A1 | × | √ | |
| rs4592275 | ALDH4A1 | × | √ | |
| rs11781951 | ALDH4A1 | × | √ | |
| rs76932496 | ALDH4A1 | × | √ | |
| rs14856487 | ALDH4A1 | × | √ | |
| rs78346435 | ALDH4A1 | × | √ | |
| rs76465841 | ALDH4A1 | × | √ | |
| rs77532991 | ALDH4A1 | × | √ | |
| rs14140343 | ALDH4A1 | × | √ | |
| rs77065167 | ALDH4A1 | × | √ | |
| rs37248424 | ALDH4A1 | × | √ | |
| rs28666920 | ALDH4A1 | × | √ | |
| rs28543677 | ALDH4A1 | × | √ | |
| rs7534676 | ALDH4A1 | × | √ | |
| rs28709274 | ALDH4A1 | × | √ | |
| rs12478175 | ANKRD44 | √ | × | |
| rs11702754 | ANKRD44 | × | √ | |
| rs11894702 | ANKRD44 | × | √ | |
| rs11694397 | ANKRD44 | × | √ | |

| | | | | | |
|---|---|---|---|---|---|
| rs77802229 | HTRA1 | × | √ | × | |
| rs2142308 | HTRA1 | × | √ | × | Lung function (FEV1/FVC) |
| rs186226638 | HTRA1 | × | √ | × | |
| rs17103569 | HTRA1 | × | √ | × | |
| rs78618914 | IARS | √ | × | × | |
| rs1175931 | IARS | √ | × | × | |
| rs7048808 | IARS | √ | × | × | |
| rs471478 | IARS | √ | × | × | COL1A1/OMD protein leve |
| rs504708 | IARS | √ | × | × | |
| rs1152757 | IARS | √ | × | × | |
| rs3996311 | IARS | √ | × | × | |
| rs10992290 | IARS | √ | × | × | |
| rs145704313 | IKZF1 | √ | × | × | |
| rs555249234 | IKZF1 | √ | × | × | |
| rs4663857 | ILKAP | √ | × | × | |
| rs74653364 | ILKAP | √ | × | × | |
| rs12472274 | ILKAP | √ | × | × | Phospholipid levels (plasm |
| rs12621791 | ILKAP | √ | × | × | |
| rs4663861 | ILKAP | √ | × | × | |
| rs60005892 | IQCD | √ | × | × | |
| rs10850132 | IQCD | √ | × | × | |
| rs4251460 | IRAK4 | × | √ | × | |
| rs4251465 | IRAK4 | × | √ | × | |
| rs4238087 | IRAK4 | × | √ | × | |
| rs4251466 | IRAK4 | × | √ | × | |
| rs1461567 | IRAK4 | × | √ | × | |
| rs118097313 | IRAK4 | × | √ | × | |
| rs3794262 | IRAK4 | × | √ | × | Vaginal microbiome MetaC |
| rs4251473 | IRAK4 | × | √ | × | |
| rs181348939 | IRAK4 | × | √ | × | |
| rs55973711 | IRAK4 | × | √ | × | |
| rs4251475 | IRAK4 | × | √ | × | |
| rs4251481 | IRAK4 | × | √ | × | |
| rs76136881 | IRAK4 | × | √ | × | |
| rs11182259 | IRAK4 | × | √ | × | |
| rs11182261 | IRAK4 | × | √ | × | |
| rs116999385 | IRAK4 | × | √ | × | |
| rs140578147 | IRAK4 | × | √ | × | |
| rs79154645 | IRAK4 | × | √ | × | |
| rs4251513 | IRAK4 | × | √ | × | |
| rs2236604 | ITPKB | × | √ | × | |
| rs77435141 | ITPKB | × | √ | × | |
| rs10495246 | ITPKB | × | √ | × | |
| rs4653458 | ITPKB | × | √ | × | |
| rs1288934 | ITPKB | × | √ | × | |
| rs3729551 | ITPKB | × | √ | × | |
| rs187208271 | ITPKB | × | √ | × | |
| rs17522524 | ITPKB | × | √ | × | |
| rs1151803 | ITPKB | √ | × | × | |
| rs1144836 | ITPKB | √ | × | × | |
| rs1144835 | ITPKB | √ | × | × | |
| rs4653762 | ITPKB | √ | × | × | |
| rs2298169 | ITPKB | √ | × | × | |
| rs5020835 | ITPKB | √ | × | × | |
| rs567047805 | JAKMIP3 | √ | × | × | |
| rs139571269 | KATNB1 | √ | × | × | |
| rs34131744 | KATNB1 | √ | × | × | |
| rs7513709 | KCNK2 | √ | × | × | |
| rs6696333 | KCNK2 | √ | × | × | |
| rs146340216 | KCNK2 | √ | × | × | |
| rs6662353 | KCNK2 | √ | × | × | |
| rs12061916 | KCNK2 | √ | × | × | |
| rs11120496 | KCNK2 | √ | × | × | |
| rs4428856 | KCNK2 | √ | × | × | |
| rs10127656 | KCNK2 | √ | × | × | Height |
| rs10127521 | KCNK2 | √ | × | × | |
| rs10864154 | KCNK2 | √ | × | × | |
| rs10127611 | KCNK2 | √ | × | × | |
| rs6666828 | KCNK2 | √ | × | × | |
| rs4269745 | KCNK2 | √ | × | × | |
| rs4655391 | KCNK2 | √ | × | × | |
| rs72735359 | KCNK2 | √ | × | × | |
| rs61818328 | KCNK2 | √ | × | × | |
| rs6675926 | KCNK2 | √ | × | × | |
| rs12031308 | KCNK2 | √ | × | × | |
| rs4655276 | KCNK2 | √ | × | × | |
| rs2363554 | KCNK2 | √ | × | × | |
| rs4497183 | KCNK2 | √ | × | × | |
| rs2885812 | KCNK2 | √ | × | × | |
| rs2885813 | KCNK2 | √ | × | × | |
| rs11120506 | KCNK2 | √ | × | × | |
| rs11120508 | KCNK2 | √ | × | × | |
| rs10779642 | KCNK2 | √ | × | × | |
| rs4509552 | KCNK2 | √ | × | × | |
| rs4509553 | KCNK2 | √ | × | × | |
| rs4655392 | KCNK2 | √ | × | × | |
| rs10779645 | KCNK2 | √ | × | × | |
| rs4655393 | KCNK2 | √ | × | × | |
| rs11014033 | KIAA1217 | √ | × | × | |
| rs60955659 | KIFC3 | √ | × | × | |
| rs71387192 | KIFC3 | √ | × | × | |
| rs34722062 | KIFC3 | √ | × | × | |
| rs12445947 | KIFC3 | √ | × | × | |
| rs12148935 | KIFC3 | √ | × | × | |
| rs35515996 | KIFC3 | √ | × | × | |
| rs34771108 | KIFC3 | √ | × | × | |
| rs16959373 | KIFC3 | √ | × | × | |
| rs4784010 | KIFC3 | √ | × | × | |
| rs77414536 | KIFC3 | √ | × | × | |
| rs79949875 | KIFC3 | √ | × | × | |
| rs1559405 | KIFC3 | √ | × | × | |
| rs12149880 | KIFC3 | √ | × | × | |
| rs11076204 | KIFC3 | √ | × | × | |
| rs77298026 | KIFC3 | √ | × | × | |
| rs12149857 | KIFC3 | √ | × | × | |
| rs112029634 | KIFC3 | √ | × | × | |
| rs12924788 | KIFC3 | √ | × | × | |
| rs337616 | KLF3 | √ | × | × | |
| rs6531656 | KLF3 | √ | × | × | Lymphocyte count |
| rs4240247 | KLF3 | √ | × | × | |
| rs729063 | KLF3 | √ | × | × | |
| rs168084 | KLF3-AS1 | √ | × | × | |
| rs337629 | KLF3-AS1 | √ | × | × | |
| rs2381199 | KLF3-AS1 | √ | × | × | Estimated glomerular filtrat |
| rs4833077 | KLF3-AS1 | √ | × | × | |
| rs337604 | KLF3-AS1 | √ | × | × | Essential tremor |
| rs59873377 | KLHL30 | √ | × | × | |
| rs74843241 | KLHL40 | × | √ | × | |
| rs140068240 | KLHL40 | × | √ | × | |
| rs149512159 | KLHL40 | × | √ | × | |
| rs138768878 | KLHL40 | × | √ | × | |
| rs376290331 | KLHL40 | × | √ | × | |
| rs533441552 | KLHL40 | × | √ | × | |
| rs72865703 | KLHL40 | × | √ | × | |
| rs339701 | KLHL40 | × | √ | × | |
| rs117955816 | KLHL40 | × | √ | × | |
| rs339700 | KLHL40 | × | √ | × | |
| rs575442079 | KLHL40 | × | √ | × | |
| rs339699 | KLHL40 | × | √ | × | |
| rs558265667 | KLHL40 | × | √ | × | |
| rs17030476 | KLHL40 | × | √ | × | |
| rs339698 | KLHL40 | × | √ | × | |
| rs73085372 | KLHL40 | × | √ | × | |
| rs186947018 | KLHL40 | × | √ | × | |
| rs61163642 | KLHL40 | × | √ | × | |
| rs186485036 | LDLRAD3 | √ | × | × | |
| rs10842374 | LINC00477 | √ | × | × | |
| rs6487399 | LINC00477 | √ | × | × | |
| rs6487400 | LINC00477 | √ | × | × | |
| rs7299236 | LINC00477 | √ | × | × | |
| rs116904393 | LINC01237 | √ | × | × | |
| rs9863495 | LOC100128164 | √ | × | × | |
| rs6421879 | LOC101927280 | √ | × | × | |
| rs6894176 | LOC101927280 | √ | × | × | |
| rs6875260 | LOC101927280 | √ | × | × | |
| rs6894916 | LOC101927280 | √ | × | × | |
| rs7721374 | LOC101927280 | √ | × | × | |
| rs7711344 | LOC101927280 | √ | × | × | |
| rs11241458 | LOC101927280 | √ | × | × | |
| rs77476803 | LOC101927280 | √ | × | × | |
| rs75594839 | LOC101927280 | √ | × | × | |
| rs77950503 | LOC101927280 | √ | × | × | |
| rs7703431 | LOC101927280 | √ | × | × | |
| rs80240958 | LOC101927280 | √ | × | × | |
| rs79650909 | LOC101927280 | √ | × | × | |
| rs117070615 | LOC101927280 | √ | × | × | |
| rs74593842 | LOC101927280 | √ | × | × | |
| rs7444011 | LOC101927280 | √ | × | × | |
| rs7708440 | LOC101927280 | √ | × | × | |
| rs78495315 | LOC101927280 | √ | × | × | |
| rs6595126 | LOC101927280 | √ | × | × | |
| rs11950959 | LOC101927280 | √ | × | × | |
| rs7727130 | LOC101927280 | √ | × | × | |
| rs6867291 | LOC101927280 | √ | × | × | |
| rs77788081 | LOC101927280 | √ | × | × | |
| rs77124635 | LOC101927280 | √ | × | × | |
| rs78400388 | LOC101927280 | √ | × | × | |
| rs74926979 | LOC101927280 | √ | × | × | |
| rs75622096 | LOC101927280 | √ | × | × | |
| rs12719243 | LOC101927280 | √ | × | × | |
| rs6873260 | LOC101927280 | √ | × | × | |
| rs78195006 | LOC101927280 | √ | × | × | |
| rs76327771 | LOC101927280 | √ | × | × | |
| rs75180781 | LOC101927280 | √ | × | × | |
| rs76135883 | LOC101927280 | √ | × | × | |
| rs79638554 | LOC101927280 | √ | × | × | |
| rs7445072 | LOC101927280 | √ | × | × | |
| rs7715481 | LOC101927280 | √ | × | × | |
| rs3923756 | LOC101927280 | √ | × | × | |
| rs78457609 | LOC101927280 | √ | × | × | |
| rs7701418 | LOC101927280 | √ | × | × | |
| rs76667220 | LOC101927280 | √ | × | × | |
| rs10058844 | LOC101927280 | √ | × | × | |
| rs116946524 | LOC101927280 | √ | × | × | |
| rs6883163 | LOC101927280 | √ | × | × | |
| rs4440399 | LOC101927280 | √ | × | × | |
| rs9327075 | LOC101927280 | √ | × | × | |
| rs6595142 | LOC101927280 | √ | × | × | |
| rs6595143 | LOC101927280 | √ | × | × | |
| rs13360367 | LOC101927280 | √ | × | × | |
| rs7708133 | LOC101927280 | √ | × | × | |
| rs7708340 | LOC101927280 | √ | × | × | |
| rs112325484 | LOC101927873 | √ | × | × | |
| rs74964617 | LOC101927873 | √ | × | × | |
| rs10733754 | LOC101927873 | √ | × | × | |
| rs883923 | LOC101927873 | √ | × | × | |
| rs883924 | LOC101927873 | √ | × | × | Hepatitis C induced liver fi |
| rs79374117 | LOC101927990 | √ | × | × | |
| rs9490762 | LOC101927990 | √ | × | × | |
| rs13205992 | LOC101927990 | √ | × | × | |
| rs9401672 | LOC101927990 | √ | × | × | |
| rs72613284 | LOC101927990 | √ | × | × | |
| rs13192662 | LOC101927990 | √ | × | × | |
| rs7744051 | LOC101927990 | √ | × | × | |
| rs6911883 | LOC101927990 | √ | × | × | |
| rs9490772 | LOC101927990 | √ | × | × | |
| rs4406254 | LOC101927990 | √ | × | × | |
| rs6909484 | LOC101927990 | √ | × | × | |
| rs1343640 | LOC101927990 | √ | × | × | |
| rs4428524 | LOC101927990 | √ | × | × | |
| rs4412217 | LOC101927990 | √ | × | × | |
| rs4613836 | LOC101927990 | √ | × | × | Body mass index |
| rs4434492 | LOC101927990 | √ | × | × | |
| rs13192696 | LOC101927990 | √ | × | × | |
| rs9482393 | LOC101927990 | √ | × | × | |
| rs17085387 | LOC101927990 | √ | × | × | |
| rs17739286 | LOC101927990 | √ | × | × | |
| rs150596234 | LOC101928203 | √ | × | × | |
| rs769107 | LOC101928663 | × | √ | × | |
| rs769108 | LOC101928663 | × | √ | × | |
| rs8180584 | LOC101928663 | × | √ | × | |
| rs149659663 | LOC101928663 | × | √ | × | |
| rs563458294 | LOC101928663 | × | √ | × | |
| rs7774664 | LOC101928663 | × | √ | × | |
| rs10080736 | LOC101928663 | × | √ | × | |
| rs12942013 | LOC101928710 | √ | × | × | |
| rs56121534 | LOC101928710 | √ | × | × | |
| rs66530414 | LOC101928710 | √ | × | × | |
| rs2840098 | LOC102723344 | √ | × | × | |
| rs7162279 | LOC102723344 | × | √ | × | |
| rs7025171 | LOC340515 | √ | × | × | |
| rs1930922 | LOC340515 | √ | × | × | |
| rs143438020 | LOC642943 | √ | × | × | |
| rs59473571 | LRRFIP1 | × | × | √ | |
| rs72987103 | LRRFIP1 | × | × | √ | |
| rs72987108 | LRRFIP1 | × | × | √ | |
| rs72987110 | LRRFIP1 | × | × | √ | |
| rs60023357 | LRRFIP1 | × | × | √ | |
| rs2117363 | LRRFIP1 | × | × | √ | |

| | | | | | |
|---|---|---|---|---|---|
| rs205105 | ULK2 | √ | × | × | |
| rs3101815 | ULK2 | √ | × | × | |
| rs421561 | ULK2 | √ | × | × | |
| rs281354 | ULK2 | √ | × | × | |
| rs188138 | ULK2 | √ | × | × | |
| rs281348 | ULK2 | √ | × | × | |
| rs281346 | ULK2 | √ | × | × | |
| rs1126586 | UNQ6494 | √ | × | × | |
| rs3744793 | USP36 | × | √ | × | |
| rs61729065 | USP36 | × | √ | × | |
| rs62075628 | USP36 | × | √ | × | |
| rs301004 | USP44 | √ | × | × | |
| rs18442802 | UTRN | √ | × | × | |
| rs7640893 | VIPR1 | × | √ | × | |
| rs76613688 | VIPR1 | × | √ | × | |
| rs735773 | VIPR1 | × | √ | × | |
| rs3678664 | VIPR1 | × | √ | × | |
| rs2278215 | VIPR1 | × | √ | × | |
| rs2267534 | VIPR1 | × | √ | × | |
| rs97515196 | VIPR1 | × | √ | × | |
| rs6799248 | VIPR1 | × | √ | × | |
| rs2267535 | VIPR1 | × | √ | × | |
| rs77432835 | VIPR1 | × | √ | × | |
| rs5759551 | VIPR1 | × | √ | × | |
| rs14569217 | VIPR1 | × | √ | × | |
| rs450812 | VIPR1 | × | √ | × | |
| rs28684623 | VIPR1 | × | √ | × | |
| rs413042 | VIPR1 | × | √ | × | |
| rs6797853 | VIPR1 | × | √ | × | |
| rs14397794 | VIPR1 | × | √ | × | |
| rs14863796 | VIPR1 | × | √ | × | |
| rs811198 | VIPR1 | × | √ | × | |
| rs2284132 | VIPR1 | × | √ | × | |
| rs55806424 | VIPR1 | × | √ | × | |
| rs14623895 | VIPR1 | × | √ | × | |
| rs37350777 | VIPR1 | × | √ | × | |
| rs78008566 | VIPR1 | × | √ | × | |
| rs18125045 | VIPR1 | × | √ | × | |
| rs18680058 | VIPR1 | × | √ | × | |
| rs75498396 | VIPR1 | × | √ | × | |
| rs580447 | VIPR1 | × | √ | × | |
| rs78829151 | VIPR1 | × | √ | × | |
| rs57015949 | VIPR1 | × | √ | × | |
| rs34664814 | VIPR1 | × | √ | × | |
| rs14467084 | VIPR1 | × | √ | × | |
| rs2239501 | VIPR1 | × | √ | × | |
| rs2229722 | VIPR1 | × | √ | × | |
| rs2040094 | WNK2 | √ | × | × | |
| rs10821084 | WNK2 | √ | × | × | |
| rs6229032 | XXYLT1 | √ | × | × | |
| rs3733058 | ZBTB47 | × | √ | × | |
| rs339721 | ZBTB47 | × | √ | × | |
| rs1008275 | ZFHX3 | × | √ | × | |
| rs719354 | ZFHX3 | √ | × | × | |
| rs62122411 | ZNF787 | √ | × | × | |
| rs19112031 | ZSCAN5B | × | √ | × | |
| rs17753438 | ZSCAN5B | × | √ | × | |
| rs76381877 | ACAP2 | √ | × | × | |
| rs11715578 | ACAP2 | √ | × | × | |
| rs14297917 | ACAP2 | √ | × | × | |
| rs72813468 | AFG3L1P | × | × | √ | |
| rs16977847 | AGBL1 | × | √ | × | |
| rs2437788 | AGBL1 | √ | × | × | |
| rs7168028 | AGBL1 | √ | × | × | |
| rs2469187 | AGBL1 | √ | × | × | |
| rs7176651 | AGBL1 | √ | × | × | |
| rs7180391 | AGBL1 | √ | × | × | |
| rs7183096 | AGBL1 | √ | × | × | |
| rs8025744 | AGBL1 | √ | × | × | |
| rs12595242 | AGBL1 | √ | × | × | |
| rs2123264 | AGBL1 | √ | × | × | |
| rs78119909 | AGBL1 | √ | × | × | |
| rs78919213 | AGBL1 | √ | × | × | |
| rs16977987 | AGBL1 | √ | × | × | |
| rs16977988 | AGBL1 | √ | × | × | |
| rs1697799 | AGBL1 | √ | × | × | |
| rs2881581 | AGBL1 | √ | × | × | |
| rs4887244 | AGBL1 | √ | × | × | |
| rs58008085 | AGBL1 | √ | × | × | |
| rs7954578 | AGBL1 | √ | × | × | |
| rs77842594 | AGBL1 | √ | × | × | |
| rs1236360 | ALKBH3 | × | √ | × | |
| rs2460120 | ALKBH3 | × | √ | × | |
| rs10734519 | ALKBH3 | × | √ | × | |
| rs12790543 | ALKBH3 | × | √ | × | |
| rs12577215 | ALKBH3 | × | √ | × | |
| rs1518813 | ALKBH3 | × | √ | × | |
| rs7128112 | ALKBH3 | × | √ | × | |
| rs11037732 | ALKBH3 | × | √ | × | |
| rs34581514 | ALKBH3 | × | √ | × | |
| rs33913339 | ALKBH3 | × | √ | × | |
| rs2292888 | ALKBH3 | × | √ | × | |
| rs34575084 | ALKBH3 | × | √ | × | |
| rs1179449 | ALKBH3 | × | √ | × | |
| rs3740989 | ALKBH3 | × | √ | × | |
| rs2902378 | ALKBH3 | × | √ | × | |
| rs7130796 | ALKBH3 | × | √ | × | |
| rs1073452 | ALKBH3 | × | √ | × | |
| rs11692795 | ALKBH3 | × | √ | × | |
| rs12421423 | ALKBH3 | × | √ | × | |
| rs61884012 | ALKBH3 | × | √ | × | |
| rs1083820 | ALKBH3 | × | √ | × | |
| rs10768994 | ALKBH3 | × | √ | × | Body mass |
| rs35402462 | ALKBH3 | × | √ | × | |
| rs10838202 | ALKBH3 | × | √ | × | |
| rs77231028 | ANGPTL2 | × | √ | × | |
| rs75846464 | ANKRD44 | × | √ | × | |
| rs2305251 | ANKRD44 | × | √ | × | |
| rs13997152 | ANKRD44 | × | √ | × | |
| rs12478175 | ANKRD44 | × | √ | × | |
| rs6757462 | ANKRD44 | × | √ | × | |
| rs75526987 | ANKRD44 | × | √ | × | |
| rs14517539 | ANKRD44 | × | √ | × | |
| rs7675661 | ANKRD44 | × | √ | × | |
| rs2376211 | ANKRD44 | × | √ | × | |
| rs73066855 | ANKRD44 | × | √ | × | |
| rs2305250 | ANKRD44 | × | √ | × | |
| rs4850745 | ANKRD44 | × | √ | × | |
| rs13027173 | ANKRD44 | × | √ | × | |
| rs4850407 | ANKRD44 | × | √ | × | |
| rs18726401 | ANKRD44 | × | √ | × | |
| rs11110012 | ANKS1B | √ | × | × | |
| rs17030634 | ANO4 | √ | × | × | |
| rs10860644 | ANO4 | √ | × | × | |
| rs11110549 | ANO4 | √ | × | × | |
| rs2138045 | ANO4 | √ | × | × | |
| rs4764767 | ANO4 | √ | × | × | |
| rs4764768 | ANO4 | √ | × | × | |
| rs10507124 | ANO4 | √ | × | × | |
| rs11687854 | ANO4 | √ | × | × | |
| rs78635425 | ANO4 | √ | × | × | |
| rs75671754 | AP2B1 | √ | × | × | |
| rs7828317 | AP2B1 | √ | × | × | |
| rs77578912 | AP2B1 | √ | × | × | |
| rs75993796 | AP2B1 | √ | × | × | |
| rs2301731 | AP2B1 | √ | × | × | |
| rs9303689 | AP2B1 | √ | × | × | |
| rs78135579 | AP2B1 | √ | × | × | |
| rs16971207 | AP2B1 | √ | × | × | |
| rs3785758 | AP2B1 | √ | × | × | |
| rs3785759 | AP2B1 | √ | × | × | |
| rs9899927 | AP2B1 | √ | × | × | |
| rs16971227 | AP2B1 | √ | × | × | |
| rs76564459 | AP2B1 | √ | × | × | |
| rs12604067 | AP2B1 | √ | × | × | |
| rs10491114 | AP2B1 | √ | × | × | |
| rs3785760 | AP2B1 | √ | × | × | |
| rs9890191 | AP2B1 | √ | × | × | |
| rs10491115 | AP2B1 | √ | × | × | |
| rs9915021 | AP2B1 | √ | × | × | High light sc |
| rs3785762 | AP2B1 | √ | × | × | |
| rs77866969 | AP2B1 | √ | × | × | |
| rs9914730 | AP2B1 | √ | × | × | |
| rs9907772 | AP2B1 | √ | × | × | |
| rs3785763 | AP2B1 | √ | × | × | |
| rs9916483 | AP2B1 | √ | × | × | |
| rs2428463 | AP2B1 | √ | × | × | |
| rs2523115 | AP2B1 | √ | × | × | |
| rs3760325 | AP2B1 | √ | × | × | |
| rs226437 | AP2B1 | √ | × | × | |
| rs226436 | AP2B1 | √ | × | × | |
| rs226435 | AP2B1 | √ | × | × | |
| rs226433 | AP2B1 | √ | × | × | |
| rs58771368 | AP2B1 | √ | × | × | |
| rs77049963 | APBB1IP | × | √ | × | |
| rs18176389 | APBB1IP | × | √ | × | |
| rs14311923 | APBB1IP | × | √ | × | |
| rs34358740 | APBB1IP | × | √ | × | |
| rs10741113 | APBB1IP | × | √ | × | |
| rs10764632 | APBB1IP | × | √ | × | |
| rs36019146 | APBB1IP | × | √ | × | |
| rs13817250 | APBB1IP | × | √ | × | |
| rs11015167 | APBB1IP | × | √ | × | |
| rs7091260 | APBB1IP | × | √ | × | |
| rs37034913 | APBB1IP | × | √ | × | |
| rs5142 | APOC3 | √ | × | × | |
| rs7908377 | ARHGAP21 | √ | × | × | |
| rs7082393 | ARHGAP21 | √ | × | × | |
| rs18713394 | ARHGAP21 | √ | × | × | |
| rs12573109 | ARHGAP21 | × | √ | × | |
| rs61855025 | ARHGAP21 | × | √ | × | |
| rs74543706 | ARHGAP21 | × | √ | × | |
| rs11014202 | ARHGAP21 | × | √ | × | |
| rs61032267 | ARHGAP21 | × | √ | × | |
| rs57192491 | ARHGAP21 | × | √ | × | |
| rs14128884 | ARHGAP21 | × | √ | × | |
| rs7071907 | ARHGAP21 | × | √ | × | |
| rs11265797 | ARHGAP21 | × | √ | × | |
| rs18461786 | ARHGAP21 | × | √ | × | |
| rs6482413 | ARHGAP21 | × | √ | × | Whole body |
| rs19272611 | ARHGAP21 | × | √ | × | |
| rs6482414 | ARHGAP21 | × | √ | × | |
| rs6482415 | ARHGAP21 | × | √ | × | |
| rs7897303 | ARHGAP21 | × | √ | × | |
| rs72793521 | ARHGAP21 | × | √ | × | |
| rs10764491 | ARHGAP21 | × | √ | × | |
| rs4748968 | ARHGAP21 | × | √ | × | |
| rs2046532 | ARL2BP | √ | × | × | |
| rs6481497 | ARMC4 | √ | × | × | |
| rs67821209 | ASRGL1 | √ | × | × | |
| rs79486818 | ASRGL1 | √ | × | × | |
| rs76825688 | ASRGL1 | √ | × | × | |
| rs11330431 | ASRGL1 | √ | × | × | |
| rs57066953 | ASRGL1 | √ | × | × | |
| rs11203603 | ASRGL1 | √ | × | × | |
| rs11276447 | ASRGL1 | √ | × | × | |
| rs18586370 | ASRGL1 | √ | × | × | |
| rs4340061 | ASRGL1 | √ | × | × | |
| rs15095231 | ASRGL1 | √ | × | × | |
| rs4502002 | ASRGL1 | √ | × | × | |
| rs11368911 | ASRGL1 | √ | × | × | |
| rs12292642 | ASRGL1 | √ | × | × | |
| rs2302359 | ASRGL1 | √ | × | × | |
| rs4432028 | ASRGL1 | √ | × | × | |
| rs4281490 | ASRGL1 | √ | × | × | |
| rs1043785 | ASRGL1 | √ | × | × | |
| rs3125296 | ATF3 | × | √ | × | |
| rs15120001 | ATG4B | √ | × | × | |
| rs75468430 | ATG4B | √ | × | × | |
| rs72484053 | ATG4B | √ | × | × | |
| rs2305243 | ATL2 | √ | × | × | |
| rs10451646 | ATL2 | √ | × | × | |
| rs6745929 | ATL2 | √ | × | × | |
| rs6744372 | ATL2 | √ | × | × | |
| rs10175905 | ATL2 | √ | × | × | |
| rs12712588 | ATL2 | √ | × | × | |
| rs10177042 | ATL2 | √ | × | × | |
| rs3731847 | ATL2 | √ | × | × | |
| rs4234470 | ATP1B3 | √ | × | × | |
| rs16907293 | AUH | √ | × | × | |
| rs79679424 | AUH | √ | × | × | |
| rs10991902 | AUH | √ | × | × | |
| rs78797942 | B9D1 | × | √ | × | |
| rs72836802 | B9D1 | × | √ | × | |
| rs12603532 | B9D1 | × | √ | × | |
| rs55377079 | B9D1 | × | √ | × | |
| rs7211555 | B9D1 | × | √ | × | |
| rs12937289 | B9D1 | × | √ | × | |
| rs20210929 | B9D1 | × | √ | × | |

| | | | | | |
|---|---|---|---|---|---|
| rs8026630 | AGBL1 | √ | × | × | |
| rs7180902 | AGBL1 | √ | × | × | |
| rs1697805 | AGBL1 | √ | × | × | |
| rs6571518 | AKAP6 | √ | × | × | |
| rs8013463 | AKAP6 | √ | × | × | |
| rs1951191 | AKAP6 | √ | × | × | |
| rs3784191 | AKAP6 | √ | × | × | |
| rs3784193 | AKAP6 | √ | × | × | |
| rs76520308 | AMPH | √ | × | × | |
| rs1214555 | ANKRD22 | √ | × | × | |
| rs1214556 | ANKRD22 | √ | × | × | |
| rs1214557 | ANKRD22 | √ | × | × | |
| rs1214559 | ANKRD22 | √ | × | × | |
| rs1300071 | ANKRD44 | × | √ | × | |
| rs36994602 | ANKRD44 | × | √ | × | |
| rs78588598 | ANKRD44 | × | √ | × | |
| rs14868757 | ANKRD44 | × | √ | × | |
| rs730200 | ANKRD44 | × | √ | × | |
| rs13015419 | ANKRD44 | × | √ | × | |
| rs1042728 | ANKRD44 | × | √ | × | |
| rs10427282 | ANKRD44 | × | √ | × | |
| rs62185248 | ANKRD44 | × | √ | × | |
| rs19193263 | ANKRD44 | × | √ | × | |
| rs2376212 | ANKRD44 | × | √ | × | |
| rs62185249 | ANKRD44 | × | √ | × | |
| rs11712788 | ANKRD44 | × | √ | × | |
| rs75040914 | ANO4 | √ | × | × | |
| rs74907998 | ANO4 | √ | × | × | |
| rs76159236 | ANO4 | √ | × | × | |
| rs78656256 | ANO4 | √ | × | × | |
| rs77919721 | ANO4 | √ | × | × | |
| rs79211681 | ANO4 | √ | × | × | |
| rs34162417 | ANO4 | √ | × | × | |
| rs75790214 | ANO4 | √ | × | × | |
| rs1703091 | ANO4 | √ | × | × | |
| rs583825 | ANO4 | √ | × | × | |
| rs585497 | ANO4 | √ | × | × | |
| rs588567 | ANO4 | √ | × | × | |
| rs620545 | ANO4 | √ | × | × | |
| rs14777041 | ANO4 | √ | × | × | |
| rs74891031 | ANO4 | √ | × | × | |
| rs11818947 | ANO4 | √ | × | × | |
| rs11026459 | ANO5 | √ | × | × | |
| rs1102646 | ANO5 | √ | × | × | |
| rs75178716 | ANO5 | √ | × | × | |
| rs225306 | AP2B1 | √ | × | × | |
| rs225304 | AP2B1 | √ | × | × | |
| rs11653357 | AP2B1 | √ | × | × | Platelet dist |
| rs14875391 | AP2B1 | √ | × | × | |
| rs225297 | AP2B1 | √ | × | × | |
| rs225295 | AP2B1 | √ | × | × | Reticulocyte |
| rs225290 | AP2B1 | √ | × | × | |
| rs225284 | AP2B1 | √ | × | × | |
| rs9913875 | AP2B1 | √ | × | × | |
| rs1671656 | AP2B1 | √ | × | × | |
| rs1634684 | AP2B1 | √ | × | × | |
| rs9899860 | AP2B1 | √ | × | × | |
| rs225245 | AP2B1 | √ | × | × | High light sc |
| rs3890901 | AP2B1 | √ | × | × | |
| rs12949341 | AP2B1 | √ | × | × | |
| rs7406966 | AP2B1 | √ | × | × | |
| rs67296463 | AP2B1 | √ | × | × | |
| rs226088 | AP2B1 | √ | × | × | |
| rs720774 | AP2B1 | √ | × | × | |
| rs2285742 | AP2B1 | √ | × | × | |
| rs226086 | AP2B1 | √ | × | × | |
| rs226085 | AP2B1 | √ | × | × | |
| rs170483 | AP2B1 | √ | × | × | |
| rs12150359 | AP2B1 | √ | × | × | |
| rs4796103 | AP2B1 | √ | × | × | |
| rs6505491 | AP2B1 | √ | × | × | |
| rs6505492 | AP2B1 | √ | × | × | |
| rs8067294 | AP2B1 | √ | × | × | |
| rs9904187 | AP2B1 | √ | × | × | |
| rs226434 | AP2B1 | √ | × | × | |
| rs226432 | AP2B1 | √ | × | × | |
| rs1121690 | ARCN1 | × | × | √ | |
| rs11216907 | ARCN1 | × | × | √ | |
| rs55844025 | ARCN1 | √ | × | × | |
| rs9736327 | ARCN1 | × | × | √ | |
| rs9735421 | ARCN1 | × | × | √ | |
| rs17122278 | ARCN1 | × | × | √ | Total choles |
| rs1121690 | ARCN1 | × | × | √ | Non-HDL c |
| rs1121691 | ARCN1 | × | × | √ | |
| rs11216911 | ARCN1 | × | × | √ | |
| rs11216912 | ARCN1 | × | × | √ | |
| rs17842763 | ARCN1 | × | × | √ | |
| rs7115634 | ARCN1 | × | × | √ | Total choles |
| rs11216925 | ARCN1 | × | × | √ | Total choles |
| rs10892245 | ARCN1 | × | × | √ | |
| rs1047079 | ARCN1 | × | × | √ | |
| rs17742 | ARCN1 | × | × | √ | |
| rs742869 | ARG2 | √ | × | × | |
| rs37314471 | ARHGAP21 | √ | × | × | |
| rs7913390 | ARHGAP21 | √ | × | × | Waist-hip ra |
| rs36813415 | ARHGAP21 | √ | × | × | |
| rs14133543 | ARHGAP21 | √ | × | × | |
| rs14474779 | ARHGAP21 | √ | × | × | |
| rs12571219 | ARHGAP21 | √ | × | × | |
| rs14198959 | ARHGAP21 | √ | × | × | |
| rs13806957 | ARHGAP21 | √ | × | × | |
| rs14729709 | ARHGAP21 | √ | × | × | |
| rs36794523 | ARHGAP21 | √ | × | × | |
| rs77134587 | ARHGAP21 | √ | × | × | |
| rs35814566 | ARHGAP21 | √ | × | × | |
| rs14888444 | ARHGAP21 | √ | × | × | |
| rs1432985 | ARHGAP21 | √ | × | × | |
| rs7923935 | ARHGAP21 | √ | × | × | |
| rs7911476 | ARHGAP21 | √ | × | × | |
| rs12571203 | ARHGAP21 | √ | × | × | |
| rs14149521 | ARHGAP21 | √ | × | × | |
| rs12571877 | ARHGAP21 | √ | × | × | |
| rs12572218 | ARHGAP21 | √ | × | × | |
| rs37566997 | ARHGAP21 | √ | × | × | |
| rs7098723 | ARHGAP21 | √ | × | × | |
| rs76597968 | ARHGAP21 | √ | × | × | |
| rs1513117 | ARHGAP21 | √ | × | × | |
| rs11633239 | ARHGAP21 | √ | × | × | |
| rs1383336 | ARHGAP21 | √ | × | × | |
| rs14693257 | ARID3C | √ | × | × | |
| rs13853758 | ARMC4 | √ | × | × | |
| rs14922643 | ARMC4 | √ | × | × | |
| rs10826379 | ARMC4 | √ | × | × | |
| rs11941355 | ARSJ | √ | × | × | |
| rs7700128 | ARSJ | √ | × | × | |
| rs7700131 | ARSJ | √ | × | × | |
| rs508986 | ASB1 | √ | × | × | |
| rs509903 | ASB1 | √ | × | × | |
| rs514396 | ASB1 | √ | × | × | |
| rs7489760 | ASRGL1 | √ | × | × | |
| rs11231058 | ASRGL1 | √ | × | × | |
| rs1079235 | ASRGL1 | √ | × | × | |
| rs10897266 | ASRGL1 | √ | × | × | |
| rs1271259 | ATL2 | √ | × | × | |
| rs1867766 | ATP12A | √ | × | × | |
| rs2863259 | ATP12A | √ | × | × | |
| rs1867768 | ATP12A | √ | × | × | |
| rs7986361 | ATP12A | √ | × | × | |
| rs7988856 | ATP12A | √ | × | × | |
| rs2070809 | ATP12A | √ | × | × | |
| rs895425 | ATP12A | √ | × | × | |
| rs895424 | ATP12A | √ | × | × | |
| rs735772 | ATP12A | √ | × | × | |
| rs1867769 | ATP12A | √ | × | × | |
| rs735770 | ATP12A | √ | × | × | |
| rs7996324 | ATP12A | √ | × | × | |
| rs7996486 | ATP12A | √ | × | × | |
| rs7997850 | ATP12A | √ | × | × | |
| rs7981616 | ATP12A | √ | × | × | |
| rs2071490 | ATP12A | √ | × | × | High light sc |
| rs2071491 | ATP12A | √ | × | × | |
| rs2071492 | ATP12A | √ | × | × | |
| rs2070811 | ATP12A | √ | × | × | |
| rs2070812 | ATP12A | √ | × | × | |
| rs963985 | ATP12A | √ | × | × | |
| rs963984 | ATP12A | √ | × | × | |
| rs9553420 | ATP12A | √ | × | × | |
| rs9553421 | ATP12A | √ | × | × | |
| rs2289898 | ATP12A | √ | × | × | |
| rs3816332 | ATP12A | √ | × | × | |
| rs78656275 | ATP1B3 | √ | × | × | |
| rs9299391 | AUH | √ | × | × | |
| rs78159864 | AUH | √ | × | × | |
| rs18973973 | AUH | √ | × | × | |
| rs7870604 | AUH | √ | × | × | |
| rs75577693 | AUH | √ | × | × | |
| rs7732481 | AUH | √ | × | × | |
| rs76224506 | AUH | √ | × | × | |
| rs56703453 | AUH | √ | × | × | |
| rs76670503 | AUH | √ | × | × | |
| rs5980017 | AUH | √ | × | × | |
| rs80293005 | AUH | √ | × | × | |
| rs11416582 | AUH | √ | × | × | |
| rs77279577 | AUH | √ | × | × | |
| rs77783396 | AUH | √ | × | × | |
| rs79906249 | AUH | √ | × | × | |
| rs75005175 | AUH | √ | × | × | |
| rs59300141 | AUH | √ | × | × | |
| rs11809449 | AUH | √ | × | × | |
| rs78312679 | AUH | √ | × | × | |
| rs12238691 | BICD2 | √ | × | × | |
| rs7165790 | BLM | √ | × | × | |
| rs7178014 | BLM | √ | × | × | |
| rs7175811 | BLM | √ | × | × | |
| rs79727167 | BLM | √ | × | × | |
| rs8042743 | BLM | √ | × | × | |
| rs8026761 | BLM | √ | × | × | |
| rs72751859 | BLM | √ | × | × | |
| rs28385093 | BLM | √ | × | × | |
| rs28385095 | BLM | √ | × | × | Whole body |
| rs28385096 | BLM | √ | × | × | |
| rs28385099 | BLM | √ | × | × | |
| rs2227933 | BLM | √ | × | × | |
| rs28385106 | BLM | √ | × | × | |
| rs28385111 | BLM | √ | × | × | |
| rs28385112 | BLM | √ | × | × | |
| rs72751868 | BLM | √ | × | × | |
| rs72751869 | BLM | √ | × | × | |
| rs3213195 | BLM | √ | × | × | |
| rs17273842 | BLM | √ | × | × | |
| rs3741301 | BUD13 | √ | × | × | |
| rs3785184 | C16orf3 | √ | × | × | |
| rs1270488 | CACNA2D1 | √ | × | × | |
| rs12670356 | CACNA2D1 | √ | × | × | |
| rs1544461 | CACNA2D1 | √ | × | × | |
| rs1544464 | CACNA2D1 | √ | × | × | |
| rs2237525 | CACNA2D1 | √ | × | × | |
| rs17676882 | CAMK2D | √ | × | × | |
| rs1762039 | CAMK2D | √ | × | × | Heart failure |
| rs79394798 | CAMK2D | √ | × | × | |
| rs80023730 | CAMK2D | √ | × | × | |
| rs77622767 | CAMK2D | √ | × | × | |
| rs73841789 | CAMK2D | √ | × | × | |
| rs11703541 | CAMK2D | √ | × | × | |
| rs7672170 | CAMK2D | √ | × | × | |
| rs77859530 | CAMK2D | √ | × | × | |
| rs11726289 | CAMK2D | √ | × | × | |
| rs11741882 | CAMK2D | √ | × | × | |
| rs6849577 | CAMK2D | √ | × | × | |
| rs78330775 | CAMK2D | √ | × | × | |
| rs11686929 | CAMK2D | √ | × | × | |
| rs11727947 | CAMK2D | √ | × | × | |
| rs78398996 | CAMK2D | √ | × | × | |
| rs11802179 | CAMK2D | √ | × | × | |
| rs41493049 | CAMK2D | √ | × | × | |
| rs73841801 | CAMK2D | √ | × | × | |
| rs10016002 | CAMK2D | √ | × | × | |
| rs916911 | CAMK2D | √ | × | × | |
| rs916912 | CAMK2D | √ | × | × | |
| rs73843441 | CAMK2D | √ | × | × | |
| rs10010256 | CAMK2D | √ | × | × | |
| rs17046295 | CAMK2D | √ | × | × | |
| rs73843443 | CAMK2D | √ | × | × | |
| rs17046302 | CAMK2D | √ | × | × | |
| rs11785415 | CAMK2D | √ | × | × | |
| rs76040080 | CAMK2D | √ | × | × | |
| rs17630730 | CAMK2D | √ | × | × | |
| rs10000007 | CAMK2D | √ | × | × | |
| rs10022752 | CAMK2D | √ | × | × | |

| | | | | |
|---|---|---|---|---|
| rs18726401 | ANKRD44 | × | √ | |
| rs14056725 | ANKRD44 | × | √ | |
| rs1020086 | ANKRD44 | × | √ | |
| rs10497802 | ANKRD44 | × | √ | |
| rs79322940 | ANKRD44 | × | √ | |
| rs37259990 | ANKRD44 | × | √ | |
| rs4850747 | ANKRD44 | × | √ | |
| rs53549976 | ANKRD44 | × | √ | |
| rs12470435 | ANKRD44 | × | √ | |
| rs14553282 | ANKRD44 | × | √ | |
| rs1300071 | ANKRD44 | × | √ | |
| rs36994602 | ANKRD44 | × | √ | |
| rs55565691 | ANKRD44 | √ | × | |
| rs75040914 | ANO4 | √ | × | |
| rs10860644 | ANO4 | √ | × | |
| rs11110549 | ANO4 | √ | × | |
| rs2138045 | ANO4 | √ | × | |
| rs17403347 | ANO4 | √ | × | |
| rs76909971 | ANO4 | √ | × | |
| rs75555839 | ANO4 | √ | × | |
| rs11110553 | ANO4 | √ | × | |
| rs77575623 | ANO4 | √ | × | |
| rs75812299 | ANO4 | √ | × | |
| rs11110555 | ANO4 | √ | × | |
| rs11110556 | ANO4 | √ | × | |
| rs79174636 | ANO4 | √ | × | |
| rs4764762 | ANO4 | √ | × | |
| rs78219128 | ANO4 | √ | × | |
| rs1399446 | ANO4 | √ | × | |
| rs11615271 | ANO4 | √ | × | |
| rs10745899 | ANO4 | √ | × | |
| rs11110559 | ANO4 | √ | × | |
| rs1111056 | ANO4 | √ | × | |
| rs925713 | ANO4 | √ | × | |
| rs1105887 | ANO4 | √ | × | |
| rs10507121 | ANO4 | √ | × | |
| rs78122126 | ANO4 | √ | × | |
| rs17404116 | ANO4 | √ | × | |
| rs74907998 | ANO4 | √ | × | |
| rs1703091 | ANO4 | √ | × | |
| rs78054553 | ANO4 | √ | × | |
| rs78985248 | ANO4 | √ | × | |
| rs2653450 | ANO4 | √ | × | |
| rs694265 | ANO4 | √ | × | |
| rs585497 | ANO4 | √ | × | |
| rs79319837 | ANO5 | √ | × | |
| rs1447436 | ANO5 | √ | × | |
| rs225297 | AP2B1 | √ | × | |
| rs225290 | AP2B1 | √ | × | |
| rs225284 | AP2B1 | √ | × | |
| rs3785758 | AP2B1 | × | √ | |
| rs225249 | AP2B1 | × | √ | |
| rs37001921 | AP2B1 | × | √ | |
| rs3785759 | AP2B1 | √ | × | |
| rs1476553 | AP2B1 | √ | × | |
| rs2301730 | AP2B1 | √ | × | |
| rs3890901 | AP2B1 | √ | × | |
| rs12949341 | AP2B1 | √ | × | |
| rs7406966 | AP2B1 | √ | × | |
| rs67296463 | AP2B1 | √ | × | |
| rs226088 | AP2B1 | √ | × | |
| rs720774 | AP2B1 | √ | × | |
| rs2285742 | AP2B1 | √ | × | |
| rs226086 | AP2B1 | √ | × | |
| rs226085 | AP2B1 | √ | × | |
| rs12150359 | AP2B1 | √ | × | |
| rs4796103 | AP2B1 | √ | × | |
| rs5142 | APOC3 | √ | × | |
| rs2295643 | ARG2 | √ | × | |
| rs7156352 | ARG2 | √ | × | |
| rs12884807 | ARG2 | √ | × | |
| rs34970739 | ARG2 | √ | × | |
| rs10764485 | ARHGAP21 | √ | × | |
| rs514396 | ASB1 | √ | × | |
| rs6700180 | ASPM | √ | × | |
| rs12034362 | ASPM | √ | × | |
| rs12042442 | ASPM | √ | × | |
| rs11608211 | ASRGL1 | √ | × | |
| rs7489760 | ASRGL1 | √ | × | |
| rs7935912 | ASRGL1 | √ | × | |
| rs76328766 | ATP12A | × | √ | |
| rs61948065 | ATP12A | × | √ | |
| rs61948066 | ATP12A | × | √ | |
| rs73158338 | ATP12A | × | √ | |
| rs11300401 | ATP12A | × | √ | |
| rs79981812 | ATP12A | × | √ | |
| rs77775634 | ATP12A | × | √ | |
| rs5564158 | ATP12A | × | √ | Systolic blood pressure |
| rs55840438 | ATP12A | × | √ | |
| rs79516284 | ATP12A | × | √ | |
| rs56083529 | ATP12A | × | √ | |
| rs61948075 | ATP12A | × | √ | |
| rs12867825 | ATP12A | × | √ | |
| rs12868533 | ATP12A | × | √ | |
| rs12860747 | ATP12A | × | √ | |
| rs3783069 | ATP12A | × | √ | |
| rs13378915 | ATP12A | × | √ | |
| rs2277422 | ATP12A | × | √ | |
| rs13378235 | ATP12A | × | √ | |
| rs7995316 | ATP12A | × | √ | |
| rs14609692 | ATP12A | × | √ | |
| rs1867766 | ATP12A | × | √ | |
| rs2863259 | ATP12A | × | √ | |
| rs1867768 | ATP12A | × | √ | |
| rs7986361 | ATP12A | × | √ | |
| rs7988856 | ATP12A | × | √ | |
| rs2070809 | ATP12A | × | √ | |
| rs895425 | ATP12A | × | √ | |
| rs895424 | ATP12A | × | √ | |
| rs14779808 | ATP12A | × | √ | |
| rs3783066 | ATP12A | × | √ | |
| rs735772 | ATP12A | × | √ | |
| rs1867769 | ATP12A | × | √ | |
| rs735770 | ATP12A | × | √ | |
| rs20154472 | ATP12A | × | √ | |
| rs20154986 | ATP12A | × | √ | |
| rs7996324 | ATP12A | × | √ | |
| rs7996486 | ATP12A | × | √ | |
| rs7997850 | ATP12A | × | √ | |
| rs77066059 | ATP12A | × | √ | |
| rs7981616 | ATP12A | × | √ | |
| rs2071490 | ATP12A | × | √ | |
| rs2071491 | ATP12A | × | √ | |
| rs2071492 | ATP12A | × | √ | |
| rs2070811 | ATP12A | × | √ | |
| rs2070812 | ATP12A | × | √ | |
| rs17081048 | ATP12A | × | √ | |
| rs2070814 | ATP12A | × | √ | |
| rs18472691 | ATP12A | × | √ | |
| rs963985 | ATP12A | × | √ | |
| rs963984 | ATP12A | × | √ | |
| rs963983 | ATP12A | × | √ | |
| rs17081055 | ATP12A | × | √ | |
| rs17081058 | ATP12A | × | √ | |
| rs1161673 | ATP12A | × | √ | |
| rs9553420 | ATP12A | × | √ | |
| rs9553421 | ATP12A | × | √ | |
| rs2289898 | ATP12A | × | √ | |
| rs2289899 | ATP12A | × | √ | |
| rs3816332 | ATP12A | × | √ | |
| rs9553423 | ATP12A | × | √ | |
| rs74647871 | ATP12A | × | √ | |
| rs9318723 | ATP12A | × | √ | |
| rs10219963 | ATP12A | × | √ | |
| rs9553425 | ATP12A | × | √ | |
| rs12873366 | ATP12A | × | √ | |
| rs1021997 | ATP12A | × | √ | |
| rs12869327 | ATP12A | × | √ | |
| rs12870584 | ATP12A | × | √ | |
| rs8002339 | ATP12A | × | √ | |
| rs56636137 | ATP12A | × | √ | |
| rs1160226 | ATP12A | × | √ | |
| rs724327 | ATP12A | × | √ | |
| rs9553426 | ATP12A | × | √ | |
| rs1904703 | ATP12A | × | √ | |
| rs631368 | ATPAF1 | √ | × | |
| rs582323 | ATPAF1 | √ | × | |
| rs583649 | ATPAF1 | √ | × | |
| rs604233 | ATPAF1 | √ | × | |
| rs2476157 | ATPAF1 | √ | × | |
| rs2486160 | ATPAF1 | √ | × | |
| rs611919 | ATPAF1 | √ | × | |
| rs655369 | ATPAF1 | √ | × | |
| rs582760 | ATPAF1 | √ | × | |
| rs2218191 | ATPAF1 | √ | × | |
| rs596794 | ATPAF1 | √ | × | |
| rs636082 | ATPAF1 | √ | × | |
| rs636424 | ATPAF1 | √ | × | |
| rs10991891 | AUH | √ | × | |
| rs76284073 | AUH | √ | × | |
| rs10991899 | AUH | √ | × | |
| rs4924985 | B9D1 | √ | × | |
| rs7211555 | B9D1 | √ | × | |
| rs12937289 | B9D1 | √ | × | |
| rs8065263 | B9D1 | √ | × | |
| rs4924782 | B9D1 | √ | × | |
| rs77448885 | BICD2 | √ | × | |
| rs12594305 | BLM | √ | × | |
| rs2072351 | BLM | √ | × | |
| rs28385098 | BLM | √ | × | |
| rs62315055 | CAMK2D | √ | × | |
| rs10488958 | CAMK2D | √ | × | |
| rs12500935 | CAMK2D | √ | × | |
| rs77996534 | CAMK2D | √ | × | |
| rs78986306 | CAMK2D | √ | × | |
| rs57665458 | CAMK2D | √ | × | |
| rs57109326 | CAMK2D | √ | × | |
| rs12510043 | CAMK2D | √ | × | |
| rs17448108 | CAMK2D | √ | × | |
| rs10010383 | CAMK2D | √ | × | |
| rs7437038 | CAMK2D | √ | × | |
| rs12507555 | CAMK2D | √ | × | |
| rs12570981 | CCAR1 | √ | × | |
| rs6743705 | CCDC138 | √ | × | |
| rs10190045 | CCDC138 | √ | × | |
| rs12151574 | CCDC138 | √ | × | |
| rs1997120 | CCDC138 | √ | × | |
| rs35177625 | CCDC138 | √ | × | |
| rs4676208 | CCDC138 | √ | × | |
| rs7594575 | CCDC138 | √ | × | |
| rs10190977 | CCDC138 | √ | × | |
| rs12712023 | CCDC138 | √ | × | |
| rs1117424 | CCDC138 | √ | × | |
| rs10202541 | CCDC138 | √ | × | |
| rs73952532 | CCDC138 | √ | × | |
| rs73952534 | CCDC138 | √ | × | |
| rs73952535 | CCDC138 | √ | × | |
| rs58286020 | CCDC138 | √ | × | |
| rs4584990 | CCDC138 | √ | × | |
| rs4676215 | CCDC138 | √ | × | |
| rs1073893 | CCDC138 | √ | × | |
| rs11410908 | CCSER1 | √ | × | |
| rs13150060 | CCSER1 | √ | × | |
| rs3733456 | CCSER1 | √ | × | |
| rs77173714 | CCSER1 | √ | × | |
| rs6124775 | CDH22 | × | √ | |
| rs13934012 | CDH22 | √ | × | |
| rs12480664 | CDH22 | √ | × | |
| rs2425825 | CDH22 | √ | × | |
| rs3904164 | CDH22 | √ | × | |
| rs10992353 | CENPP | √ | × | |
| rs14814467 | CEP83 | √ | × | |
| rs15033075 | CEP83 | √ | × | |
| rs17610487 | CHD2 | √ | × | |
| rs12440804 | CHSY1 | √ | × | |
| rs11778639 | CHSY1 | √ | × | |
| rs11636578 | CHSY1 | √ | × | Hippocampal subfield left CA1 volume (body) |
| rs60057209 | CLPX | × | √ | |
| rs7175110 | CLPX | × | √ | |
| rs57015448 | CLPX | × | √ | |
| rs11335454 | CLPX | × | √ | |
| rs11071828 | CLPX | × | √ | |
| rs13864244 | CLPX | × | √ | |
| rs14029458 | CLPX | × | √ | |
| rs14905960 | CLPX | × | √ | |
| rs18595433 | CLPX | × | √ | |
| rs52957968 | CLPX | × | √ | |
| rs61878683 | COMMD9 | √ | × | |
| rs7544067 | CRB1 | √ | × | |

| | | | | | |
|---|---|---|---|---|---|
| rs2254541 | LRSAM1 | √ | × | × | |
| rs56131876 | LRSAM1 | √ | × | × | |
| rs1539568 | LRSAM1 | √ | × | × | |
| rs2246600 | LRSAM1 | √ | × | × | |
| rs7875862 | LRSAM1 | √ | × | × | |
| rs2246375 | LRSAM1 | √ | × | × | |
| rs2243464 | LRSAM1 | √ | × | × | |
| rs2245677 | LRSAM1 | √ | × | × | |
| rs1539564 | LRSAM1 | √ | × | × | |
| rs975979 | LUZP2 | √ | × | × | |
| rs10834374 | LUZP2 | √ | × | × | |
| rs4923192 | LUZP2 | √ | × | × | |
| rs10767209 | LUZP2 | √ | × | × | |
| rs2202893 | LUZP2 | √ | × | × | |
| rs2136483 | LUZP2 | √ | × | × | |
| rs4491224 | LUZP2 | √ | × | × | |
| rs2896706 | LUZP2 | √ | × | × | |
| rs10834377 | LUZP2 | √ | × | × | |
| rs10834378 | LUZP2 | √ | × | × | |
| rs1396844 | LUZP2 | √ | × | × | |
| rs11027995 | LUZP2 | √ | × | × | |
| rs2202894 | LUZP2 | √ | × | × | |
| rs11028003 | LUZP2 | √ | × | × | |
| rs1912138 | LUZP2 | √ | × | × | |
| rs1912139 | LUZP2 | √ | × | × | |
| rs2403959 | LUZP2 | √ | × | × | |
| rs2036750 | LUZP2 | √ | × | × | |
| rs10834382 | LUZP2 | √ | × | × | |
| rs11028007 | LUZP2 | √ | × | × | |
| rs12273206 | LUZP2 | √ | × | × | |
| rs10767212 | LUZP2 | √ | × | × | |
| rs10834384 | LUZP2 | √ | × | × | |
| rs7107070 | LUZP2 | √ | × | × | |
| rs1509605 | LUZP2 | √ | × | × | Depressive symptoms |
| rs10834385 | LUZP2 | √ | × | × | |
| rs12276298 | LUZP2 | √ | × | × | |
| rs12295653 | LUZP2 | √ | × | × | |
| rs1509610 | LUZP2 | √ | × | × | |
| rs1509612 | LUZP2 | √ | × | × | |
| rs1509613 | LUZP2 | √ | × | × | |
| rs11028012 | LUZP2 | √ | × | × | |
| rs10834386 | LUZP2 | √ | × | × | |
| rs7105561 | LUZP2 | √ | × | × | |
| rs7105854 | LUZP2 | √ | × | × | |
| rs7121909 | LUZP2 | √ | × | × | |
| rs6484057 | LUZP2 | √ | × | × | |
| rs1509614 | LUZP2 | √ | × | × | |
| rs1039837 | LUZP2 | √ | × | × | |
| rs75579040 | LUZP2 | √ | × | × | |
| rs11028015 | LUZP2 | √ | × | × | |
| rs6484058 | LUZP2 | √ | × | × | |
| rs7936313 | LUZP2 | √ | × | × | |
| rs1912145 | LUZP2 | √ | × | × | |
| rs11028020 | LUZP2 | √ | × | × | |
| rs11028021 | LUZP2 | √ | × | × | |
| rs34167424 | LUZP2 | √ | × | × | |
| rs4465365 | LUZP2 | √ | × | × | |
| rs117328323 | LUZP2 | √ | × | × | |
| rs12289378 | LUZP2 | √ | × | × | |
| rs7951458 | LUZP2 | √ | × | × | |
| rs75817223 | LUZP2 | √ | × | × | |
| rs115213330 | LUZP2 | √ | × | × | |
| rs11529784 | LUZP2 | √ | × | × | |
| rs77957935 | LUZP2 | √ | × | × | |
| rs9943629 | LUZP2 | √ | × | × | |
| rs12143447 | MEAF6 | √ | × | × | |
| rs12125547 | MEAF6 | √ | × | × | |
| rs12126401 | MEAF6 | √ | × | × | |
| rs77089424 | METAP2 | √ | × | × | |
| rs12821148 | METAP2 | × | √ | × | |
| rs73231331 | METAP2 | × | √ | × | |
| rs78076980 | METAP2 | × | √ | × | |
| rs7299439 | METAP2 | × | √ | × | |
| rs2596741 | METAP2 | × | √ | × | Height |
| rs543371347 | METAP2 | × | √ | × | |
| rs71458596 | METAP2 | × | √ | × | |
| rs301026 | METAP2 | × | √ | × | |
| rs7590275 | METTL8 | √ | × | × | |
| rs145809785 | MIR548AC | √ | × | × | |
| rs73049415 | MIR548AC | √ | × | × | |
| rs11717508 | MIR548AC | √ | × | × | |
| rs11706875 | MIR548AC | √ | × | × | |
| rs62256991 | MIR548AC | √ | × | × | |
| rs74709254 | MIR548AC | √ | × | × | |
| rs41400550 | MIR548AC | √ | × | × | |
| rs185779029 | MIR548AC | √ | × | × | |
| rs60028742 | MIR548AC | √ | × | × | |
| rs62256972 | MIR548AC | √ | × | × | |
| rs74600707 | MIR548AC | √ | × | × | |
| rs4858513 | MIR548AC | √ | × | × | |
| rs62256975 | MIR548AC | √ | × | × | |
| rs112374474 | MIR548AC | √ | × | × | |
| rs79562849 | MIR548AC | √ | × | × | |
| rs62256976 | MIR548AC | √ | × | × | |
| rs62256977 | MIR548AC | √ | × | × | |
| rs7640308 | MIR548AC | √ | × | × | |
| rs115359127 | MIR548AC | √ | × | × | |
| rs6774454 | MIR548AC | √ | × | × | |
| rs77832017 | MIR548AC | √ | × | × | |
| rs60137984 | MIR548AC | √ | × | × | |
| rs62256982 | MIR548AC | √ | × | × | |
| rs28367247 | MIR548AC | √ | × | × | |
| rs9395899 | MLIP | √ | × | × | |
| rs6914051 | MLIP | × | × | √ | |
| rs77708866 | MLIP | √ | × | × | |
| rs16884853 | MLIP | √ | × | × | |
| rs9650391 | MSR1 | √ | × | × | |
| rs35796001 | MSR1 | √ | × | × | |
| rs11985261 | MSR1 | √ | × | × | |
| rs33936250 | MSR1 | √ | × | × | |
| rs6986419 | MSR1 | √ | × | × | |
| rs34918250 | MSR1 | √ | × | × | |
| rs35281051 | MSR1 | √ | × | × | |
| rs918 | MSR1 | √ | × | × | |
| rs13306546 | MSR1 | √ | × | × | |
| rs13306544 | MSR1 | √ | × | × | |
| rs12675467 | MSR1 | √ | × | × | |
| rs12678016 | MSR1 | √ | × | × | |
| rs12677608 | MSR1 | √ | × | × | |
| rs12679486 | MSR1 | √ | × | × | |
| rs34612904 | MSR1 | √ | × | × | |
| rs7015682 | MSR1 | √ | × | × | |
| rs6995004 | MSR1 | √ | × | × | |
| rs6995009 | MSR1 | √ | × | × | |
| rs6981908 | MSR1 | √ | × | × | |
| rs6982066 | MSR1 | √ | × | × | |
| rs6982504 | MSR1 | √ | × | × | |
| rs78757102 | MSR1 | √ | × | × | |
| rs78016136 | MSR1 | √ | × | × | |
| rs76903738 | MSR1 | √ | × | × | |
| rs76841569 | MSR1 | √ | × | × | |
| rs12676117 | MSR1 | √ | × | × | |
| rs12675698 | MSR1 | √ | × | × | |
| rs12676141 | MSR1 | √ | × | × | |
| rs12675748 | MSR1 | √ | × | × | |
| rs12676172 | MSR1 | √ | × | × | |
| rs12676591 | MSR1 | √ | × | × | |
| rs12678046 | MSR1 | √ | × | × | |
| rs62491230 | MSR1 | √ | × | × | |
| rs62491233 | MSR1 | √ | × | × | |
| rs56996957 | MSR1 | √ | × | × | |
| rs41454647 | MSR1 | √ | × | × | |
| rs41487145 | MSR1 | √ | × | × | |
| rs78553524 | MSR1 | √ | × | × | Protein quantitative trait loc |
| rs80002038 | MSR1 | √ | × | × | |
| rs77089638 | MSR1 | √ | × | × | |
| rs78765574 | MSR1 | √ | × | × | |
| rs2302047 | MTERFD2 | √ | × | × | |
| rs1062748 | MTERFD2 | √ | × | × | |
| rs61234509 | MTERFD2 | √ | × | × | |
| rs78834190 | MTERFD2 | √ | × | × | |
| rs10884940 | MXI1 | √ | × | × | |
| rs75164148 | MXI1 | √ | × | × | |
| rs7897645 | MXI1 | √ | × | × | |
| rs76158849 | MYPN | √ | × | × | |
| rs77317354 | MYPN | √ | × | × | |
| rs75531686 | MYPN | √ | × | × | |
| rs139351954 | MYPN | √ | × | × | |
| rs10760528 | NAIF1 | × | √ | × | |
| rs4836605 | NAIF1 | × | √ | × | |
| rs7852659 | NAIF1 | × | √ | × | |
| rs1594403 | NCK2 | √ | × | × | |
| rs2377338 | NCK2 | √ | × | × | |
| rs12623430 | NCK2 | √ | × | × | |
| rs3954042 | NCK2 | √ | × | × | |
| rs7589561 | NCK2 | √ | × | × | |
| rs4851877 | NCK2 | √ | × | × | |
| rs2082184 | NCK2 | √ | × | × | |
| rs1945417 | NELL1 | × | √ | × | |
| rs2156806 | NELL1 | × | √ | × | |
| rs4576819 | NELL1 | × | √ | × | |
| rs77120294 | NELL1 | × | √ | × | |
| rs76609379 | NELL1 | × | √ | × | |
| rs10500908 | NELL1 | × | √ | × | |
| rs199926873 | NELL1 | × | √ | × | |
| rs10500909 | NELL1 | × | √ | × | |
| rs7110569 | NELL1 | × | √ | × | |
| rs11026083 | NELL1 | × | √ | × | |
| rs116246800 | NELL1 | × | √ | × | |
| rs1945414 | NELL1 | × | √ | × | |
| rs537945431 | NELL1 | × | √ | × | |
| rs79269343 | NELL1 | × | √ | × | |
| rs61885938 | NELL1 | × | √ | × | |
| rs17305470 | NELL1 | × | √ | × | |
| rs114729427 | NELL1 | × | √ | × | |
| rs146244659 | NELL1 | × | √ | × | |
| rs10833534 | NELL1 | × | √ | × | |
| rs12421932 | NELL1 | √ | × | × | |
| rs78756288 | NELL1 | √ | × | × | |
| rs76305787 | NELL1 | √ | × | × | |
| rs4923635 | NELL1 | √ | × | × | |
| rs75942970 | NELL1 | √ | × | × | |
| rs56272920 | NELL1 | √ | × | × | |
| rs76882629 | NELL1 | √ | × | × | |
| rs4923645 | NELL1 | √ | × | × | |
| rs80197260 | NELL1 | √ | × | × | |
| rs77386834 | NELL1 | √ | × | × | |
| rs12421933 | NELL1 | √ | × | × | |
| rs12421940 | NELL1 | √ | × | × | |
| rs4441013 | NELL1 | √ | × | × | |
| rs4922558 | NELL1 | √ | × | × | |
| rs60795440 | NELL1 | √ | × | × | |
| rs58530376 | NELL1 | √ | × | × | |
| rs58907183 | NELL1 | √ | × | × | |
| rs7933730 | NELL1 | √ | × | × | |
| rs58379880 | NELL1 | √ | × | × | |
| rs61211384 | NELL1 | √ | × | × | |
| rs73471121 | NELL1 | √ | × | × | |
| rs61321874 | NELL1 | √ | × | × | |
| rs7125542 | NELL1 | √ | × | × | |
| rs4343020 | NELL1 | √ | × | × | |
| rs7119525 | NELL1 | √ | × | × | |
| rs68126098 | NELL1 | √ | × | × | |
| rs73014018 | NEU4 | √ | × | × | |
| rs73108258 | NEU4 | √ | × | × | |
| rs2293764 | NEU4 | √ | × | × | |
| rs6793186 | NKTR | √ | × | × | |
| rs6788548 | NKTR | √ | × | × | |
| rs6778982 | NKTR | √ | × | × | |
| rs79712822 | NKTR | √ | × | × | |
| rs77424663 | NKTR | √ | × | × | |
| rs3774401 | NKTR | √ | × | × | |
| rs3774402 | NKTR | √ | × | × | |
| rs74791813 | NKTR | √ | × | × | |
| rs367471 | NLRP8 | √ | × | × | |
| rs4480335 | NOS1AP | √ | × | × | |
| rs12124745 | NOS1AP | √ | × | × | |
| rs7536043 | NOS1AP | √ | × | × | |
| rs4657175 | NOS1AP | √ | × | × | QT interval |
| rs12123267 | NOS1AP | √ | × | × | |
| rs138340875 | NR1D2 | √ | × | × | |
| rs753947 | NRF1 | √ | × | × | |
| rs7795187 | NRF1 | √ | × | × | |
| rs10245818 | NRF1 | √ | × | × | |
| rs75020879 | NRF1 | √ | × | × | |
| rs28689822 | NRF1 | √ | × | × | |
| rs78086437 | NRF1 | √ | × | × | |
| rs4731610 | NRF1 | √ | × | × | |
| rs4731611 | NRF1 | √ | × | × | |

| | | | | | |
|---|---|---|---|---|---|
| rs8065263 | B9D1 | × | √ | × | |
| rs7212549 | B9D1 | × | √ | × | |
| rs1457127 | B9D1 | × | √ | × | |
| rs7221577 | B9D1 | × | √ | × | |
| rs3785773 | B9D1 | × | √ | × | |
| rs5756032 | B9D1 | × | √ | × | |
| rs9519262 | B9D1 | × | √ | × | |
| rs5466806 | B9D1 | × | √ | × | |
| rs7971674 | B9D1 | × | √ | × | |
| rs7839962 | B9D1 | × | √ | × | |
| rs5463577 | B9D1 | × | √ | × | |
| rs7221836 | B9D1 | × | √ | × | |
| rs1378904 | B9D1 | × | √ | × | |
| rs4924988 | B9D1 | √ | × | × | |
| rs1106374 | B9D1 | × | √ | × | |
| rs1168758 | B9D1 | × | √ | × | |
| rs7838681 | B9D1 | × | √ | × | |
| rs7687032 | B9D1 | × | √ | × | |
| rs6587073 | B9D1 | × | √ | × | |
| rs1169139 | B9D1 | × | √ | × | |
| rs7500914 | B9D1 | × | √ | × | |
| rs3886478 | B9D1 | × | √ | × | |
| rs6587075 | B9D1 | × | √ | × | |
| rs5641183 | B9D1 | × | √ | × | |
| rs7283881 | B9D1 | × | √ | × | |
| rs7989626 | B9D1 | × | √ | × | |
| rs6587076 | B9D1 | × | √ | × | |
| rs7283881 | B9D1 | × | √ | × | |
| rs5431455 | B9D1 | × | √ | × | |
| rs7943752 | B9D1 | × | √ | × | |
| rs7502770 | B9D1 | × | √ | × | |
| rs1381973 | B9D1 | × | √ | × | |
| rs5319294 | BICD2 | √ | × | × | |
| rs2518967 | BLM | √ | × | × | |
| rs2072351 | BLM | √ | × | × | |
| rs2181162 | BTBD16 | √ | × | × | |
| rs2181161 | BTBD16 | √ | × | × | |
| rs1159254 | BTBD16 | √ | × | × | Vaginal micr |
| rs1120053 | BTBD16 | √ | × | × | |
| rs1710336 | BTBD16 | √ | × | × | |
| rs7502342 | C1orf115 | √ | × | × | |
| rs1274939 | C1orf95 | √ | × | × | |
| rs4550750 | C9orf78 | √ | × | × | |
| rs6706107 | C9orf78 | √ | × | × | |
| rs3814547 | C9orf78 | √ | × | × | |
| rs1392100 | CA12 | × | √ | × | |
| rs8039774 | CA12 | × | √ | × | |
| rs1694695 | CA12 | × | √ | × | |
| rs1861776 | CA12 | × | √ | × | |
| rs728904 | CA12 | × | √ | × | |
| rs3913344 | CA12 | × | √ | × | |
| rs7731511 | CA12 | × | √ | × | |
| rs7162561 | CA12 | × | √ | × | |
| rs7183733 | CA12 | × | √ | × | |
| rs1694696 | CA12 | × | √ | × | RS-warfarin |
| rs881500 | CA12 | × | √ | × | |
| rs7171715 | CA12 | × | √ | × | |
| rs1386356 | CA12 | × | √ | × | |
| rs1179286 | CA12 | × | √ | × | |
| rs7172837 | CA12 | × | √ | × | |
| rs7173285 | CA12 | × | √ | × | |
| rs7553052 | CA12 | × | √ | × | |
| rs1176378 | CACNA2D1 | √ | × | × | |
| rs1847581 | CACNA2D1 | √ | × | × | |
| rs7439290 | CAMK1D | × | × | √ | |
| rs8001665 | CAMK1D | × | × | √ | Vaginal micr |
| rs1450082 | CAMK2D | √ | × | × | |
| rs6231505 | CAMK2D | √ | × | × | |
| rs1048895 | CAMK2D | √ | × | × | |
| rs1250093 | CAMK2D | √ | × | × | |
| rs5766545 | CAMK2D | √ | × | × | |
| rs5710932 | CAMK2D | √ | × | × | |
| rs1251004 | CAMK2D | √ | × | × | |
| rs1149688 | CCAR1 | √ | × | × | |
| rs1257098 | CCAR1 | √ | × | × | |
| rs3579477 | CCDC138 | √ | × | × | |
| rs1021160 | CCDC138 | √ | × | × | |
| rs7585888 | CCDC138 | √ | × | × | |
| rs1117425 | CCDC138 | √ | × | × | |
| rs1585710 | CCDC138 | √ | × | × | |
| rs2886302 | CCDC138 | √ | × | × | |
| rs1343022 | CCDC138 | √ | × | × | |
| rs1340802 | CCDC138 | √ | × | × | |
| rs7293407 | CCDC138 | √ | × | × | |
| rs1020254 | CCDC138 | √ | × | × | |
| rs1017931 | CCDC138 | √ | × | × | |
| rs7525575 | CCDC138 | √ | × | × | |
| rs1340686 | CCDC138 | √ | × | × | |
| rs7798448 | CCDC138 | √ | × | × | |
| rs5696398 | CCDC138 | √ | × | × | |
| rs1478518 | CCDC138 | √ | × | × | |
| rs4584990 | CCDC138 | √ | × | × | |
| rs4676215 | CCDC138 | √ | × | × | |
| rs1073893 | CCDC138 | √ | × | × | |
| rs3733453 | CCSER1 | √ | × | × | |
| rs3733455 | CCSER1 | √ | × | × | |
| rs1181040 | CDCA4 | √ | × | × | |
| rs1923599 | CENPP | √ | × | × | |
| rs7627552 | CENPP | √ | × | × | |
| rs7275240 | CENPP | √ | × | × | |
| rs7275241 | CENPP | √ | × | × | |
| rs7275241 | CENPP | √ | × | × | |
| rs7275242 | CENPP | √ | × | × | |
| rs7275242 | CENPP | √ | × | × | |
| rs7275242 | CENPP | √ | × | × | |
| rs7674165 | CENPP | √ | × | × | |
| rs7275243 | CENPP | √ | × | × | |
| rs7485149 | CENPP | √ | × | × | |
| rs1895826 | CENPP | √ | × | × | |
| rs1821524 | CENPP | √ | × | × | |
| rs1484582 | CENPP | √ | × | × | |
| rs4130152 | CENPP | √ | × | × | |
| rs4129718 | CENPP | √ | × | × | |
| rs2236529 | CENPP | √ | × | × | |
| rs7952237 | CEP83 | √ | × | × | |
| rs7337031 | CEP83 | √ | × | × | |
| rs7850589 | CEP83 | √ | × | × | |
| rs3952589 | CEP83 | √ | × | × | |
| rs7469433 | CEP83 | √ | × | × | |
| rs7303686 | CEP83 | √ | × | × | |
| rs6538496 | CEP83 | √ | × | × | Height |
| rs1961780 | CEP83 | √ | × | × | |
| rs4761614 | CEP83 | √ | × | × | |
| rs1045073 | CEP83 | × | √ | × | |
| rs1520723 | CEP83 | × | √ | × | |
| rs1843854 | CEP83 | × | √ | × | |
| rs7321827 | CEP83 | × | √ | × | |
| rs1471325 | CEP83 | × | √ | × | |
| rs3700989 | CEP83 | × | √ | × | |
| rs1390722 | CEP83 | × | √ | × | |
| rs2272459 | CHD2 | √ | × | × | |
| rs1163155 | CHD2 | √ | × | × | |
| rs1163925 | CHD2 | √ | × | × | |
| rs7174740 | CHD2 | √ | × | × | Body mass |
| rs3826035 | CHD2 | √ | × | × | |
| rs1761048 | CHD2 | √ | × | × | |
| rs5925311 | CHD2 | √ | × | × | |
| rs2866813 | CHD2 | √ | × | × | |
| rs1290967 | CHD2 | √ | × | × | |
| rs1074513 | CHD2 | √ | × | × | |
| rs1133127 | CHSY1 | × | √ | × | |
| rs7171515 | CHSY1 | × | √ | × | |
| rs1393301 | CHSY1 | × | √ | × | |
| rs1428338 | CHSY1 | × | √ | × | |
| rs1147357 | CHSY1 | × | √ | × | |
| rs7625267 | CHSY1 | × | √ | × | |
| rs1123468 | CHSY1 | × | √ | × | |
| rs1259323 | CHSY1 | × | √ | × | |
| rs1244236 | CHSY1 | × | √ | × | Dupuytren's |
| rs7276857 | CHSY1 | × | √ | × | |
| rs8038694 | CHSY1 | × | √ | × | |
| rs1502431 | CHSY1 | × | √ | × | |
| rs8040150 | CHSY1 | × | √ | × | Height |
| rs8040504 | CHSY1 | × | √ | × | |
| rs7276858 | CHSY1 | × | √ | × | Height |
| rs3743192 | CHSY1 | × | √ | × | |
| rs7165356 | CHSY1 | × | √ | × | |
| rs7507152 | CHSY1 | × | √ | × | |
| rs1132568 | CHSY1 | × | √ | × | |
| rs1163180 | CHSY1 | × | √ | × | |
| rs5667492 | CHSY1 | × | √ | × | |
| rs5800739 | CHSY1 | × | √ | × | |
| rs2857003 | CHSY1 | × | √ | × | |
| rs7868596 | CHSY1 | × | √ | × | |
| rs8043249 | CHSY1 | × | √ | × | |
| rs3777402 | CHSY1 | × | √ | × | |
| rs1244227 | CHSY1 | × | √ | × | |
| rs1404529 | CHSY1 | × | √ | × | |
| rs1185846 | CHSY1 | × | √ | × | |
| rs1163428 | CHSY1 | × | √ | × | |
| rs5813645 | CHSY1 | × | √ | × | |
| rs1179329 | CHSY1 | × | √ | × | |
| rs1885372 | CHSY1 | × | √ | × | |
| rs1176452 | CHSY1 | × | √ | × | |
| rs7734267 | CHSY1 | × | √ | × | |
| rs7512640 | CHSY1 | × | √ | × | |
| rs7775346 | CHSY1 | × | √ | × | |
| rs1175838 | CHSY1 | × | √ | × | |
| rs8038272 | CHSY1 | × | √ | × | |
| rs1503816 | CHSY1 | × | √ | × | |
| rs1408085 | CHSY1 | × | √ | × | |
| rs1163007 | CHSY1 | × | √ | × | |
| rs1163045 | CHSY1 | × | √ | × | |
| rs7276860 | CHSY1 | × | √ | × | |
| rs8023810 | CHSY1 | × | √ | × | |
| rs2849577 | CHSY1 | × | √ | × | |
| rs8031268 | CHSY1 | × | √ | × | |
| rs1488804 | CHSY1 | × | √ | × | |
| rs7794191 | CHSY1 | × | √ | × | |
| rs7404192 | CHSY1 | × | √ | × | |
| rs1166439 | CHSY1 | × | √ | × | Blood press |
| rs7171436 | CHSY1 | × | √ | × | |
| rs5397676 | CHSY1 | × | √ | × | |
| rs1818281 | CHSY1 | × | √ | × | |
| rs7176149 | CHSY1 | × | √ | × | |
| rs7175303 | CHSY1 | × | √ | × | |
| rs1896119 | CHSY1 | × | √ | × | |
| rs3498049 | COMMD9 | √ | × | × | |
| rs1280189 | COMMD9 | √ | × | × | |
| rs3473054 | COMMD9 | √ | × | × | |
| rs3550236 | CPNE2 | √ | × | × | |
| rs7538938 | CPXM2 | √ | × | × | |
| rs1092218 | CRB1 | √ | × | × | |
| rs6722406 | CRB1 | √ | × | × | |
| rs1092218 | CRB1 | √ | × | × | |
| rs3507808 | CRB1 | √ | × | × | |
| rs6673293 | CRB1 | √ | × | × | |
| rs4915406 | CRB1 | √ | × | × | |
| rs7423020 | CRB1 | √ | × | × | |
| rs6740292 | CRB1 | √ | × | × | |
| rs1582118 | CRB1 | √ | × | × | |
| rs1761683 | CRB1 | √ | × | × | |
| rs540544 | CRB1 | √ | × | × | |
| rs1092219 | CRB1 | √ | × | × | |
| rs4915440 | CRB1 | √ | × | × | |
| rs1202240 | CRB1 | √ | × | × | |
| rs1415343 | CRB1 | √ | × | × | |
| rs7553508 | CRB1 | √ | × | × | |
| rs7530522 | CRB1 | √ | × | × | |
| rs1337005 | CRB1 | √ | × | × | Complemen |
| rs1080160 | CRB1 | √ | × | × | |
| rs4915208 | CRB1 | √ | × | × | |
| rs1337170 | CRB1 | √ | × | × | |
| rs1092219 | CRB1 | √ | × | × | |
| rs1204217 | CRB1 | √ | × | × | |
| rs1163832 | CRTC3 | √ | × | × | |
| rs1099678 | CTNNA3 | √ | × | × | |
| rs1099678 | CTNNA3 | √ | × | × | |
| rs1692224 | CTNNA3 | √ | × | × | |
| rs7330604 | CTNNA3 | √ | × | × | |
| rs1099679 | CTNNA3 | √ | × | × | |
| rs7920755 | CTNNA3 | √ | × | × | |
| rs5585025 | CTNNA3 | √ | × | × | |
| rs4291566 | CTNNA3 | √ | × | × | |
| rs1082269 | CTNNA3 | √ | × | × | |
| rs2169664 | CTNNA3 | √ | × | × | |
| rs1903863 | CTNNA3 | √ | × | × | |
| rs5619693 | CTNNA3 | √ | × | × | |
| rs7917997 | CTNNA3 | √ | × | × | |

| | | | | | |
|---|---|---|---|---|---|
| rs2868782 | CAMK2D | √ | × | × | |
| rs7384344 | CAMK2D | √ | × | × | |
| rs1233159 | CAMK2D | √ | × | × | |
| rs5636061 | CAMK2D | √ | × | × | |
| rs5948111 | CAMK2D | √ | × | × | |
| rs2854612 | CAMK2D | √ | × | × | |
| rs2872216 | CAMK2D | √ | × | × | |
| rs2844570 | CAMK2D | √ | × | × | |
| rs1704632 | CAMK2D | √ | × | × | |
| rs7384346 | CAMK2D | √ | × | × | |
| rs5820302 | CAMK2D | √ | × | × | |
| rs7384346 | CAMK2D | √ | × | × | |
| rs7517607 | CAMK2D | √ | × | × | |
| rs1763081 | CAMK2D | √ | × | × | |
| rs1140739 | CAMK2D | √ | × | × | |
| rs7545749 | CAMK2D | √ | × | × | |
| rs1759310 | CAMK2D | √ | × | × | |
| rs7881260 | CAMK2D | √ | × | × | |
| rs5897995 | CAMK2D | √ | × | × | |
| rs5920000 | CAMK2D | √ | × | × | |
| rs5763655 | CAMK2D | √ | × | × | |
| rs1193417 | CAMK2D | √ | × | × | |
| rs1193554 | CAMK2D | √ | × | × | |
| rs1193569 | CAMK2D | √ | × | × | |
| rs7670640 | CAMK2D | √ | × | × | |
| rs1704637 | CAMK2D | √ | × | × | |
| rs1704638 | CAMK2D | √ | × | × | |
| rs7691921 | CAMK2D | √ | × | × | |
| rs1170711 | CAMK2D | √ | × | × | |
| rs6812422 | CAMK2D | √ | × | × | |
| rs7081960 | CCAR1 | √ | × | × | |
| rs3771907 | CCDC13 | √ | × | × | |
| rs5626047 | CCDC13 | √ | × | × | |
| rs6743705 | CCDC138 | √ | × | × | |
| rs1019004 | CCDC138 | √ | × | × | |
| rs1215157 | CCDC138 | √ | × | × | |
| rs1997120 | CCDC138 | √ | × | × | |
| rs3517762 | CCDC138 | √ | × | × | |
| rs4676208 | CCDC138 | √ | × | × | |
| rs7594575 | CCDC138 | √ | × | × | |
| rs1019097 | CCDC138 | √ | × | × | |
| rs1271202 | CCDC138 | √ | × | × | |
| rs1117424 | CCDC138 | √ | × | × | |
| rs7395253 | CCDC138 | √ | × | × | |
| rs7395253 | CCDC138 | √ | × | × | |
| rs223895 | CCL17 | √ | × | × | |
| rs223896 | CCL17 | √ | × | × | Thymus anc |
| rs223897 | CCL17 | √ | × | × | |
| rs223898 | CCL17 | √ | × | × | |
| rs184402 | CCL17 | √ | × | × | |
| rs223900 | CCL17 | √ | × | × | C-C motif cl |
| rs223827 | CCL17 | √ | × | × | |
| rs3218036 | CCNE1 | √ | × | × | Height |
| rs7257694 | CCNE1 | √ | × | × | Diastolic blo |
| rs7558529 | CCSER1 | √ | × | × | |
| rs1718365 | CCSER1 | √ | × | × | |
| rs7670744 | CCSER1 | √ | × | × | |
| rs1718389 | CCSER1 | √ | × | × | |
| rs7831246 | CCSER1 | √ | × | × | |
| rs1701745 | CCSER1 | √ | × | × | |
| rs7478651 | CCSER1 | √ | × | × | |
| rs7661774 | CCSER1 | √ | × | × | |
| rs6231298 | CCSER1 | √ | × | × | |
| rs3774815 | CDH22 | √ | × | × | |
| rs1248066 | CDH22 | √ | × | × | |
| rs1172898 | CDH22 | √ | × | × | |
| rs3904164 | CDH22 | √ | × | × | |
| rs2272457 | CHD2 | √ | × | × | |
| rs1752580 | CHD2 | √ | × | × | |
| rs1052071 | CHD2 | √ | × | × | |
| rs1163058 | CHD2 | √ | × | × | |
| rs140514 | CHKB | √ | × | × | |
| rs131750 | CHKB-AS1 | √ | × | × | Low myopia |
| rs2858794 | CHST15 | √ | × | × | |
| rs6717590 | CHSY1 | √ | × | × | Basophil co |
| rs3608232 | CHSY1 | √ | × | × | |
| rs7817315 | CHSY1 | √ | × | × | |
| rs588430 | CIAPIN1 | √ | × | × | |
| rs7148190 | COMMD9 | √ | × | × | |
| rs1103352 | COMMD9 | √ | × | × | |
| rs1103353 | COMMD9 | √ | × | × | High density |
| rs1906851 | COMMD9 | √ | × | × | |
| rs487372 | COQ9 | √ | × | × | |
| rs686402 | COQ9 | √ | × | × | |
| rs622913 | COQ9 | √ | × | × | |
| rs1075422 | CRB1 | √ | × | × | Body mass |
| rs6496684 | CRTC3 | × | × | √ | |
| rs6656159 | CTNNA3 | √ | × | × | |
| rs1781937 | CTNNA3 | √ | × | × | |
| rs1724121 | CTNNA3 | √ | × | × | |
| rs1786921 | CTNNA3 | √ | × | × | |
| rs1470483 | CTNNA3 | √ | × | × | |
| rs5642143 | CYTH1 | √ | × | × | |
| rs5782340 | CYTH1 | √ | × | × | |
| rs5592573 | CYTH1 | √ | × | × | |
| rs1113567 | CYTH1 | √ | × | × | |
| rs1167756 | DCAF17 | √ | × | × | |
| rs1261286 | DCAF17 | √ | × | × | |
| rs7681966 | DCAF17 | √ | × | × | |
| rs1311659 | DCTD | √ | × | × | |
| rs2709338 | DCTD | √ | × | × | |
| rs6823077 | DCTD | √ | × | × | |
| rs8011305 | DCTN3 | √ | × | × | |
| rs4948205 | DDC | √ | × | × | |
| rs6963996 | DDC | √ | × | × | |
| rs2985689 | DNAAF2 | √ | × | × | |
| rs1226631 | DPYSL4 | √ | × | × | |
| rs3688559 | ECHDC1 | √ | × | × | |
| rs3703284 | ECHDC1 | √ | × | × | |
| rs3823042 | ECHDC1 | √ | × | × | Diffusing ca |
| rs1045749 | ECHDC1 | √ | × | × | |
| rs1045749 | ECHDC1 | √ | × | × | |
| rs7758718 | ECHDC1 | √ | × | × | |
| rs1168983 | EDAR | √ | × | × | |
| rs262086 | EDAR | √ | × | × | |
| rs1112371 | EDAR | √ | × | × | |
| rs260631 | EDAR | √ | × | × | |
| rs260627 | EDAR | √ | × | × | |
| rs5677152 | EFCAB14 | √ | × | × | |
| rs5947999 | EFCAB14 | √ | × | × | |
| rs1320256 | ELOVL5 | √ | × | × | |
| rs9395854 | ELOVL5 | √ | × | × | |
| rs2115564 | ELOVL5 | √ | × | × | |
| rs9370192 | ELOVL5 | √ | × | × | |
| rs9370193 | ELOVL5 | √ | × | × | |
| rs4431421 | ELOVL5 | √ | × | × | |
| rs9395856 | ELOVL5 | √ | × | × | |
| rs1094874 | ELOVL5 | √ | × | × | |
| rs1094874 | ELOVL5 | √ | × | × | |
| rs9395858 | ELOVL5 | √ | × | × | |
| rs9382194 | ELOVL5 | √ | × | × | |
| rs9349662 | ELOVL5 | √ | × | × | |
| rs9474484 | ELOVL5 | √ | × | × | |
| rs9382196 | ELOVL5 | √ | × | × | |
| rs209500 | ELOVL5 | √ | × | × | |
| rs6458918 | ELOVL5 | √ | × | × | |
| rs209507 | ELOVL5 | √ | × | × | |
| rs209512 | ELOVL5 | √ | × | × | |
| rs1856232 | ESR1 | √ | × | × | |
| rs1437821 | ESR1 | √ | × | × | |
| rs9479119 | ESR1 | √ | × | × | |
| rs1336264 | ESR1 | √ | × | × | |
| rs5757823 | ESR1 | √ | × | × | |
| rs1708171 | ESR1 | √ | × | × | |
| rs6930222 | ESR1 | √ | × | × | |
| rs3853248 | ESR1 | √ | × | × | |
| rs3844508 | ESR1 | √ | × | × | |
| rs3844509 | ESR1 | √ | × | × | |
| rs1252377 | ESR1 | √ | × | × | |
| rs1252380 | ESR1 | √ | × | × | |
| rs7962449 | ESR1 | √ | × | × | |
| rs3866461 | ESR1 | √ | × | × | |
| rs7759411 | ESR1 | √ | × | × | |
| rs903511 | EXT2 | √ | × | × | |
| rs1151351 | FAM102A | √ | × | × | |
| rs2296705 | FAM102A | √ | × | × | |
| rs7579186 | FAM102A | √ | × | × | |
| rs7028676 | FAM102A | √ | × | × | |
| rs7913556 | FAM102A | √ | × | × | |
| rs1076051 | FAM102A | √ | × | × | |
| rs1237444 | FAM155A | √ | × | × | |
| rs1925427 | FAM155A | √ | × | × | |
| rs9559086 | FAM155A | √ | × | × | |
| rs6196594 | FAM155A | √ | × | × | |
| rs1697109 | FAM155A | √ | × | × | |
| rs1435128 | FAM155A | √ | × | × | |
| rs1509091 | FAM155A | √ | × | × | Metabolite l |
| rs8032779 | FAM155A | √ | × | × | |
| rs1010341 | FAM49B | √ | × | × | |
| rs1010793 | FAM49B | √ | × | × | |
| rs5391551 | FARP2 | √ | × | × | |
| rs7858847 | FARP2 | √ | × | × | |
| rs1904702 | FBRSL1 | √ | × | × | |
| rs7591386 | FBXW7 | × | √ | × | |
| rs7811214 | FBXW7 | × | √ | × | |
| rs7774109 | FBXW7 | × | √ | × | |
| rs1181876 | FBXW7 | √ | × | × | |
| rs1410017 | FGD3 | √ | × | × | |
| rs7640933 | FGF12 | √ | × | × | |
| rs3735275 | FIGNL1 | √ | × | × | |
| rs3814299 | FLG | √ | × | × | |
| rs1223602 | FOCAD | √ | × | × | |
| rs1328921 | FOCAD | √ | × | × | |
| rs7432091 | FOCAD | √ | × | × | |
| rs2614154 | FRMD4A | × | × | √ | |
| rs2797880 | FRMD4A | × | × | √ | |
| rs3726223 | GMEB2 | √ | × | × | |
| rs4480744 | GNB5 | √ | × | × | |
| rs7166828 | GNB5 | √ | × | × | |
| rs3794546 | GNB5 | √ | × | × | |
| rs2288559 | GNB5 | √ | × | × | |
| rs3753372 | GNL2 | √ | × | × | |
| rs1090835 | GNL2 | √ | × | × | |
| rs6945597 | GRB10 | √ | × | × | |
| rs2876846 | GRB10 | √ | × | × | |
| rs2051896 | GRB10 | √ | × | × | |
| rs1014384 | GRB10 | √ | × | × | |
| rs7794164 | GRB10 | √ | × | × | |
| rs4947873 | GRB10 | √ | × | × | |
| rs6958750 | GRB10 | √ | × | × | |
| rs7803278 | GRB10 | √ | × | × | |
| rs6593180 | GRB10 | √ | × | × | |
| rs2301925 | GRB10 | √ | × | × | |
| rs1859571 | GRB10 | √ | × | × | |
| rs1123832 | GRB10 | √ | × | × | Hearing pro |
| rs1123832 | GRB10 | √ | × | × | |
| rs6963498 | GRB10 | √ | × | × | |
| rs367836 | GSTA4 | √ | × | × | |
| rs673197 | GSTA4 | √ | × | × | |
| rs423475 | GSTA4 | √ | × | × | |
| rs316141 | GSTA4 | √ | × | × | |
| rs1863856 | HDLBP | √ | × | × | |
| rs1901515 | HDLBP | √ | × | × | |
| rs7440065 | IARS | √ | × | × | |
| rs7777123 | IARS | √ | × | × | |
| rs9409661 | IARS | √ | × | × | |
| rs4128021 | IARS | √ | × | × | |
| rs1178957 | IARS | √ | × | × | |
| rs1178769 | IARS | √ | × | × | |
| rs9409465 | IARS | √ | × | × | |
| rs895639 | IARS | √ | × | × | |
| rs9409467 | IARS | √ | × | × | |
| rs1082096 | IARS | √ | × | × | |
| rs1073992 | IARS | √ | × | × | |
| rs7863406 | IARS | √ | × | × | |
| rs6479416 | IARS | √ | × | × | |
| rs2277293 | IFT46 | × | × | √ | |
| rs7929887 | IFT46 | × | × | √ | |
| rs1121689 | IFT46 | × | × | √ | |
| rs1155242 | IFT46 | × | × | √ | |
| rs4326835 | IFT46 | × | × | √ | |
| rs2277291 | IFT46 | × | × | √ | |
| rs1121690 | IFT46 | × | × | √ | |
| rs7595453 | ILKAP | √ | × | × | |
| rs7604350 | ILKAP | √ | × | × | |
| rs6431587 | ILKAP | √ | × | × | |
| rs3437154 | ILKAP | √ | × | × | |
| rs6731358 | IMMT | √ | × | × | |
| rs3821017 | IMMT | √ | × | × | |

| | | | | |
|---|---|---|---|---|
| rs3507808 | CRB1 | √ | × | |
| rs6673293 | CRB1 | √ | × | |
| rs1761683 | CRB1 | √ | × | |
| rs1092219 | CRB1 | √ | × | |
| rs1240832 | CRB1 | √ | × | |
| rs1786918 | CTNNA3 | √ | × | |
| rs1941988 | CTNNA3 | √ | × | |
| rs1786912 | CTNNA3 | √ | × | |
| rs7705058 | CTNNA3 | √ | × | |
| rs5585025 | CTNNA3 | √ | × | |
| rs5614452 | CTNNA3 | √ | × | |
| rs1903865 | CTNNA3 | √ | × | |
| rs1903863 | CTNNA3 | √ | × | |
| rs6480131 | CTNNA3 | √ | × | |
| rs1099685 | CTNNA3 | √ | × | |
| rs1099685 | CTNNA3 | √ | × | |
| rs7081880 | CTNNA3 | √ | × | |
| rs7798799 | CTNNA3 | √ | × | |
| rs1685944 | CYBRD1 | √ | × | |
| rs1247223 | CYBRD1 | √ | × | |
| rs1093047 | CYBRD1 | √ | × | |
| rs7262523 | CYBRD1 | √ | × | |
| rs400681 | CYP1B1-AS | √ | × | |
| rs432252 | CYP1B1-AS | √ | × | |
| rs4879741 | DCAF12 | √ | × | |
| rs1046035 | DCAF17 | √ | × | |
| rs1167756 | DCAF17 | √ | × | |
| rs2842012 | DCAF17 | √ | × | |
| rs1205281 | DCAF17 | √ | × | |
| rs1157546 | DDC | √ | × | |
| rs1267147 | DDC | √ | × | |
| rs1026732 | DDC | √ | × | |
| rs1027134 | DDC | √ | × | |
| rs1027766 | DDC | √ | × | |
| rs7792658 | DDR2 | √ | × | |
| rs7682134 | DDX21 | √ | × | |
| rs5030892 | DDX50 | × | √ | |
| rs1701184 | DISP1 | √ | × | |
| rs1265905 | DMXL1 | × | √ | |
| rs1164591 | DMXL1 | √ | × | |
| rs7613043 | DNALI1 | √ | × | |
| rs1176660 | DPYSL4 | √ | × | |
| rs1399656 | DPYSL4 | √ | × | |
| rs3798850 | ECHDC1 | √ | × | |
| rs7775493 | ECHDC1 | √ | × | |
| rs910720 | ECHDC1 | √ | × | |
| rs6930849 | ECHDC1 | √ | × | |
| rs7453272 | ECHDC1 | √ | × | |
| rs9388561 | ECHDC1 | √ | × | |
| rs9398837 | ECHDC1 | √ | × | |
| rs9398838 | ECHDC1 | √ | × | |
| rs3798852 | ECHDC1 | √ | × | |
| rs3798853 | ECHDC1 | √ | × | |
| rs2057223 | ECHDC1 | √ | × | |
| rs7776136 | ECHDC1 | √ | × | |
| rs6903878 | ECHDC1 | √ | × | |
| rs9285460 | ECHDC1 | √ | × | |
| rs4895822 | ECHDC1 | √ | × | |
| rs6937287 | ECHDC1 | √ | × | |
| rs7756336 | ECHDC1 | √ | × | |
| rs1262395 | EDAR | √ | × | |
| rs1168983 | EDAR | √ | × | |
| rs262086 | EDAR | √ | × | |
| rs7500724 | EDAR | √ | × | |
| rs7564294 | EDAR | √ | × | |
| rs5677152 | EFCAB14 | √ | × | |
| rs7290071 | EFCAB14 | √ | × | |
| rs5947999 | EFCAB14 | √ | × | |
| rs1121134 | EFCAB14 | √ | × | |
| rs7774550 | ELF1 | √ | × | |
| rs209483 | ELOVL5 | √ | × | |
| rs2115564 | ELOVL5 | √ | × | |
| rs209485 | ELOVL5 | √ | × | |
| rs3785774 | EPN2 | √ | × | |
| rs9371227 | ESR1 | √ | × | |
| rs9479126 | ESR1 | √ | × | Height |
| rs7442137 | ESR1 | √ | × | |
| rs1510036 | ESR1 | √ | × | |
| rs1490975 | EXT2 | √ | × | |
| rs1465852 | EXT2 | √ | × | |
| rs1412635 | F13B | √ | × | |
| rs6415846 | FAM102A | √ | × | |
| rs6478807 | FAM102A | √ | × | |
| rs2031694 | FAM102A | √ | × | |
| rs7026115 | FAM102A | √ | × | |
| rs7935609 | FAM102A | √ | × | |
| rs6814341 | FAM114A1 | √ | × | Height |
| rs7323666 | FAM114A1 | √ | × | |
| rs5610645 | FAM114A1 | √ | × | |
| rs5580842 | FAM114A1 | √ | × | |
| rs5606907 | FAM114A1 | √ | × | |
| rs1193967 | FAM114A1 | √ | × | |
| rs1194283 | FAM114A1 | √ | × | |
| rs1194451 | FAM114A1 | √ | × | |
| rs1143816 | FAM155A | √ | × | |
| rs1697016 | FAM155A | √ | × | |
| rs1924446 | FAM155A | √ | × | |
| rs9514694 | FAM155A | √ | × | |
| rs2840289 | FAM155A | √ | × | |
| rs4143295 | FAM155A | √ | × | |
| rs7265724 | FAM155A | √ | × | |
| rs7918871 | FAM155A | √ | × | |
| rs7461284 | FAM155A | √ | × | |
| rs9555418 | FAM155A | √ | × | |
| rs1684359 | FARP2 | √ | × | |
| rs3771549 | FARP2 | √ | × | |
| rs7300214 | FARP2 | √ | × | |
| rs6219320 | FARP2 | √ | × | |
| rs7858847 | FARP2 | √ | × | |
| rs6219320 | FARP2 | √ | × | |
| rs6219320 | FARP2 | √ | × | |
| rs6219321 | FARP2 | √ | × | |
| rs5753002 | FARP2 | √ | × | |
| rs7479590 | FBRSL1 | √ | × | |
| rs1281487 | FBRSL1 | √ | × | |
| rs1178341 | FBRSL1 | √ | × | |
| rs7957803 | FBXW7 | √ | × | |
| rs1023962 | FBXW7 | √ | × | |
| rs1351903 | FBXW7 | √ | × | |
| rs1268460 | FGD3 | √ | × | |
| rs7882839 | FGF12 | × | √ | |
| rs7491040 | FGF12 | × | √ | |
| rs7888928 | FGF12 | × | √ | |
| rs2007126 | FNBP1 | √ | × | LDL peak particle diameter (total fat intake interaction) |
| rs4837432 | FNBP1 | √ | × | |
| rs7039157 | FNBP1 | √ | × | |
| rs1479248 | FOCAD | √ | × | |
| rs1096477 | FOCAD | √ | × | |
| rs1426604 | GALR1 | √ | × | |
| rs7972980 | GALR1 | √ | × | |
| rs6158324 | GCLC | √ | × | Lung adenocarcinoma |
| rs6241686 | GCLC | √ | × | |
| rs3753372 | GNL2 | √ | × | |
| rs1090835 | GNL2 | √ | × | |
| rs522246 | GNPAT | √ | × | |
| rs1298555 | GNPAT | √ | × | |
| rs6244756 | GRB10 | √ | × | |
| rs1485394 | GRB10 | √ | × | |
| rs4947854 | GRB10 | √ | × | |
| rs1123831 | GRB10 | √ | × | |
| rs6593174 | GRB10 | √ | × | |
| rs6966774 | GRB10 | √ | × | |
| rs6945597 | GRB10 | √ | × | |
| rs2876846 | GRB10 | √ | × | |
| rs6161856 | GRB10 | × | √ | |
| rs1173643 | GRB10 | × | √ | |
| rs1879340 | GRB10 | × | √ | |
| rs7451651 | GRB10 | × | √ | |
| rs1891582 | GRB10 | × | √ | |
| rs1825545 | GRB10 | × | √ | |
| rs1011209 | GRB10 | × | √ | |
| rs7474259 | GRB10 | × | √ | |
| rs5742886 | GRB10 | × | √ | |
| rs1441154 | GRB10 | × | √ | |
| rs5598607 | GRB10 | × | √ | |
| rs1205611 | GRB10 | × | √ | |
| rs5329396 | GRB10 | × | √ | |
| rs7791286 | GRB10 | × | √ | Fasting plasma glucose |
| rs7899776 | GRB10 | × | √ | |
| rs1395408 | GRB10 | × | √ | |
| rs6593185 | GRB10 | × | √ | |
| rs5276693 | GRB10 | × | √ | |
| rs7989112 | GRK5 | × | √ | |
| rs7736789 | GRK5 | × | √ | |
| rs1088643 | GRK5 | × | √ | |
| rs3748236 | GRK5 | × | √ | |
| rs7772410 | GRK5 | × | √ | |
| rs672281 | GSTA4 | √ | × | |
| rs1262216 | HDLBP | √ | × | |
| rs6218636 | HDLBP | √ | × | |
| rs6133249 | HDLBP | √ | × | |
| rs8026390 | HDLBP | √ | × | |
| rs7640496 | HDLBP | √ | × | |
| rs1395197 | HDLBP | √ | × | |
| rs6218641 | HDLBP | √ | × | |
| rs5612939 | HDLBP | √ | × | |
| rs6218642 | HDLBP | √ | × | |
| rs1415843 | HLX-AS1 | √ | × | |
| rs1179254 | IARS | √ | × | |
| rs501814 | IARS | √ | × | |
| rs556155 | IARS | √ | × | |
| rs4743866 | IARS | √ | × | |
| rs4743867 | IARS | √ | × | |
| rs1179350 | IARS | √ | × | |
| rs1175930 | IARS | √ | × | |
| rs5489770 | IARS | √ | × | |
| rs472163 | IARS | √ | × | |
| rs4743868 | IARS | √ | × | |
| rs9409659 | IARS | √ | × | |
| rs9409661 | IARS | √ | × | |
| rs4128021 | IARS | √ | × | |
| rs1012547 | IARS | √ | × | |
| rs1178957 | IARS | √ | × | |
| rs1178769 | IARS | √ | × | |
| rs1076115 | IARS | √ | × | |
| rs9409465 | IARS | √ | × | |
| rs895639 | IARS | √ | × | |
| rs9409466 | IARS | √ | × | |
| rs9409467 | IARS | √ | × | |
| rs1466220 | ICK | √ | × | |
| rs1121997 | IFFO2 | × | √ | |
| rs7811050 | IKZF1 | √ | × | |
| rs6731358 | IMMT | √ | × | |
| rs5601077 | ING2 | √ | × | |
| rs7930012 | IPPK | √ | × | |
| rs1407190 | IQCD | √ | × | |
| rs4251569 | IRAK4 | × | √ | |
| rs3708810 | IRAK4 | × | √ | |
| rs1421299 | IRAK4 | × | √ | |
| rs5631211 | IRAK4 | × | √ | |
| rs2072675 | JAG2 | √ | × | |
| rs2072674 | JAG2 | √ | × | Vertex-wise sulcal depth |
| rs1116083 | JAG2 | √ | × | |
| rs2239285 | JAG2 | √ | × | Vertex-wise cortical thickness |
| rs1882845 | JAG2 | √ | × | |
| rs701234 | KCNK1 | √ | × | |
| rs7911355 | KIAA1217 | √ | × | |
| rs8007955 | KIAA1217 | √ | × | |
| rs1388169 | KIAA1217 | × | √ | |
| rs5630264 | KIFC3 | √ | × | |
| rs5808230 | KIFC3 | √ | × | |
| rs7972909 | KRT74 | √ | × | |
| rs262447 | LDLRAD3 | √ | × | |
| rs262445 | LDLRAD3 | √ | × | |
| rs4756265 | LDLRAD3 | √ | × | Lymphocyte count |
| rs1076817 | LDLRAD3 | √ | × | Lymphocyte count |
| rs1084237 | LINC00477 | √ | × | |
| rs4963770 | LINC00477 | √ | × | |
| rs1084237 | LINC00477 | √ | × | |
| rs1170407 | LINC00926 | √ | × | |
| rs8031036 | LINC00928 | √ | × | |
| rs8029479 | LINC00928 | √ | × | |
| rs7721350 | LINC01237 | √ | × | |
| rs7667266 | LINC01237 | √ | × | |
| rs7647863 | LINC01237 | √ | × | |
| rs1339286 | LINC01237 | √ | × | |
| rs7400086 | LINC01237 | √ | × | |
| rs1338991 | LINC01237 | √ | × | |
| rs3934981 | LINC01237 | √ | × | |

| | | | | | | | | | | | | | | | | | | | | | | |
|---|---|---|---|---|---|---|---|---|---|---|---|---|---|---|---|---|---|---|---|---|---|---|
| rs12535925 | NRF1 | √ | × | × | | rs7918256 | CTNNA3 | √ | × | × | | rs18355077 | IPPK | √ | × | × | | rs3934982 | LINC01237 | √ | × | |
| rs12666356 | NRF1 | √ | × | × | | rs4745883 | CTNNA3 | √ | × | × | | rs14071906 | IQCD | √ | × | × | | rs3935095 | LINC01237 | √ | × | |
| rs62489293 | NRF1 | √ | × | × | | rs10996851 | CTNNA3 | √ | × | × | | rs37774373 | IQCD | √ | × | × | | rs12466299 | LINC01237 | √ | × | |
| rs12538553 | NRF1 | √ | × | × | | rs10996852 | CTNNA3 | √ | × | × | | rs56069026 | KCNK2 | √ | × | × | | rs56162244 | LINC01237 | √ | × | |
| rs2402974 | NRF1 | √ | × | × | | rs10996858 | CTNNA3 | √ | × | × | | rs1901622 | KCNK2 | √ | × | × | | rs12517723 | LOC101927 | √ | × | |
| rs1806626 | NRF1 | √ | × | × | | rs10996859 | CTNNA3 | √ | × | × | | rs72762605 | KCNK2 | √ | × | × | | rs74656176 | LOC101927 | √ | × | |
| rs59675752 | NRF1 | √ | × | × | | rs72793421 | CTNNA3 | √ | × | × | | rs11120466 | KCNK2 | √ | × | × | | rs77094617 | LOC101927 | √ | × | |
| rs11844206 | NUDT14 | √ | × | × | | rs10996860 | CTNNA3 | √ | × | × | | rs1452611 | KCNK2 | √ | × | × | | rs76467916 | LOC101927 | √ | × | |
| rs11844119 | NUDT14 | √ | × | × | | rs7893776 | CTNNA3 | √ | × | × | | rs2601621 | KCNK2 | √ | × | × | | rs12055019 | LOC101927 | √ | × | |
| rs143404613 | ONECUT1 | √ | × | × | | rs18709956 | CTNNA3 | √ | × | × | | rs2601622 | KCNK2 | √ | × | × | | rs12518624 | LOC101927 | √ | × | |
| rs117812607 | PACRG | √ | × | × | | rs10996868 | CTNNA3 | √ | × | × | | rs2601623 | KCNK2 | √ | × | × | | rs61287758 | LOC101927 | √ | × | Disease progression in age-related macular degeneration (adjusted for baseline) |
| rs572385015 | PACRG | √ | × | × | | rs10996869 | CTNNA3 | √ | × | × | | rs2802644 | KCNK2 | √ | × | × | | rs12520153 | LOC101927 | √ | × | |
| rs9346934 | PACRG | √ | × | × | | rs7081880 | CTNNA3 | √ | × | × | | rs2841590 | KCNK2 | √ | × | × | | rs75420649 | LOC101927 | √ | × | |
| rs9347692 | PACRG | √ | × | × | | rs7099996 | CTNNA3 | √ | × | × | | rs2841589 | KCNK2 | √ | × | × | | rs12515148 | LOC101927 | √ | × | |
| rs61193920 | PACRG | × | √ | × | | rs59558417 | CTNNA3 | √ | × | × | | rs7519026 | KCNK2 | √ | × | × | | rs77786927 | LOC101927 | √ | × | |
| rs554236978 | PACRG | × | √ | × | | rs12357451 | CTNNA3 | √ | × | × | | rs1156732 | KCNK2 | √ | × | × | | rs12518839 | LOC101927 | √ | × | |
| rs574152099 | PACRG | × | √ | × | | rs10509254 | CTNNA3 | √ | × | × | | rs2841585 | KCNK2 | √ | × | × | | rs12513737 | LOC101927 | √ | × | |
| rs34603926 | PACRG | × | √ | × | | rs10509253 | CTNNA3 | √ | × | × | | rs2841584 | KCNK2 | √ | × | × | | rs34520222 | LOC101927 | √ | × | |
| rs9347696 | PACRG | × | √ | × | | rs10930479 | CYBRD1 | √ | × | × | | rs1452613 | KCNK2 | √ | × | × | | rs77028648 | LOC101927 | √ | × | |
| rs369447301 | PACRG | × | √ | × | | rs72625236 | CYBRD1 | √ | × | × | | rs2841582 | KCNK2 | √ | × | × | | rs79426516 | LOC101927 | √ | × | |
| rs79405081 | PACRG | × | √ | × | | rs2432674 | CYP1B1-AS | √ | × | × | | rs2601626 | KCNK2 | √ | × | × | | rs11712419 | LOC101927 | √ | × | |
| rs562507528 | PACRG | × | √ | × | | rs12105203 | CYP1B1-AS | √ | × | × | | rs2601627 | KCNK2 | √ | × | × | | rs7707724 | LOC101927 | √ | × | |
| rs9365498 | PACRG | × | √ | × | | rs400681 | CYP1B1-AS | √ | × | × | | rs2601628 | KCNK2 | √ | × | × | | rs7446705 | LOC101927 | √ | × | |
| rs1333961 | PACRG | × | √ | × | | rs432252 | CYP1B1-AS | √ | × | × | | rs2167687 | KCNK2 | √ | × | × | | rs75337620 | LOC101927 | √ | × | |
| rs1333962 | PACRG | × | √ | × | | rs13385748 | CYP1B1-AS | √ | × | × | | rs2601629 | KCNK2 | √ | × | × | | rs78903852 | LOC101927 | √ | × | |
| rs12527960 | PACRG | × | √ | × | | rs75556068 | CYP1B1-AS | √ | × | × | | rs1452614 | KCNK2 | √ | × | × | | rs79397614 | LOC101927 | √ | × | |
| rs369679898 | PACRG | × | √ | × | | rs12946167 | CYTH1 | √ | × | × | | rs1452615 | KCNK2 | √ | × | × | | rs78230061 | LOC101927 | √ | × | |
| rs76658474 | PACRG | × | √ | × | | rs55779573 | CYTH1 | √ | × | × | | rs1452616 | KCNK2 | √ | × | × | | rs6887550 | LOC101927 | √ | × | |
| rs13202482 | PACRG | × | √ | × | | rs10512617 | CYTH1 | √ | × | × | | rs2841578 | KCNK2 | √ | × | × | | rs6595129 | LOC101927 | √ | × | Height |
| rs78541421 | PACRG | × | √ | × | | rs4796807 | CYTH1 | √ | × | × | | rs2841576 | KCNK2 | √ | × | × | | rs7448537 | LOC101927 | √ | × | |
| rs182515345 | PACRG | × | √ | × | | rs58567966 | CYTH1 | √ | × | × | | rs1452617 | KCNK2 | √ | × | × | | rs6868999 | LOC101927 | √ | × | |
| rs371071687 | PACRG | × | √ | × | | rs28433797 | DCTD | √ | × | × | | rs1452618 | KCNK2 | √ | × | × | | rs12522494 | LOC101927 | √ | × | |
| rs9365499 | PACRG | × | √ | × | Body mass index | rs13114262 | DCTD | √ | × | × | | rs1452619 | KCNK2 | √ | × | × | | rs4329039 | LOC101927 | √ | × | |
| rs13209887 | PACRG | × | √ | × | | rs17152014 | DDC | √ | × | × | | rs2802655 | KCNK2 | √ | × | × | | rs6859213 | LOC101927 | √ | × | |
| rs76742106 | PACRG | × | √ | × | | rs75495166 | DDC | √ | × | × | | rs2601631 | KCNK2 | √ | × | × | | rs7726074 | LOC101927 | √ | × | |
| rs74354308 | PACRG | × | √ | × | | rs11575453 | DDC | √ | × | × | | rs2802656 | KCNK2 | √ | × | × | | rs35546771 | LOC101927 | √ | × | |
| rs187576164 | PACRG | × | √ | × | | rs11575450 | DDC | √ | × | × | | rs2601632 | KCNK2 | √ | × | × | | rs34273870 | LOC101927 | √ | × | |
| rs9356068 | PACRG | × | √ | × | | rs2329368 | DDC | √ | × | × | | rs1377174 | KCNK2 | √ | × | × | | rs35346599 | LOC101927 | √ | × | |
| rs9356069 | PACRG | × | √ | × | | rs11575441 | DDC | √ | × | × | | rs1562994 | KCNK2 | √ | × | × | | rs6868178 | LOC101927 | √ | × | |
| rs113560866 | PAFAH1B2 | √ | × | × | | rs11575438 | DDC | √ | × | × | | rs1377176 | KCNK2 | √ | × | × | | rs6892913 | LOC101927 | √ | × | |
| rs45625638 | PAFAH1B2 | √ | × | × | | rs59239231 | DDC | √ | × | × | | rs1452621 | KCNK2 | √ | × | × | | rs6892954 | LOC101927 | √ | × | |
| rs4938351 | PAFAH1B2 | √ | × | × | | rs58085392 | DDC | √ | × | × | | rs12097900 | KCNK2 | √ | × | × | | rs4333349 | LOC101927 | √ | × | |
| rs3736120 | PAFAH1B2 | √ | × | × | | rs58024888 | DDC | √ | × | × | | rs1452622 | KCNK2 | √ | × | × | | rs4426942 | LOC101927 | √ | × | |
| rs79118964 | PAFAH1B2 | √ | × | × | | rs11575388 | DDC | √ | × | × | | rs2802652 | KCNK2 | √ | × | × | | rs77500342 | LOC101927 | √ | × | |
| rs6589602 | PAFAH1B2 | √ | × | × | | rs11575387 | DDC | √ | × | × | | rs12064317 | KCNK2 | √ | × | × | | rs78381164 | LOC101927 | √ | × | |
| rs10892083 | PAFAH1B2 | √ | × | × | | rs6951648 | DDC | √ | × | × | | rs12066755 | KCNK2 | √ | × | × | | rs75212422 | LOC101927 | √ | × | |
| rs56371319 | PAFAH1B2 | √ | × | × | | rs11575383 | DDC | √ | × | × | | rs28753797 | KCNK2 | √ | × | × | | rs77130837 | LOC101927 | √ | × | |
| rs558260733 | PAFAH1B2 | √ | × | × | | rs17133853 | DDC | √ | × | × | | rs72735346 | KCNK2 | √ | × | × | | rs61748661 | LOC101927 | √ | × | |
| rs146228126 | PAFAH1B2 | √ | × | × | | rs7791875 | DDC | √ | × | × | | rs72735362 | KCNK2 | √ | × | × | | rs61748662 | LOC101927 | √ | × | |
| rs2846477 | PARK2 | √ | × | × | | rs11575372 | DDC | √ | × | × | | rs72735364 | KCNK2 | √ | × | × | | rs1754076 | LOC101927 | √ | × | |
| rs9347662 | PARK2 | √ | × | × | | rs12671474 | DDC | √ | × | × | | rs72735367 | KCNK2 | √ | × | × | | rs2771102 | LOC101927 | √ | × | |
| rs9356044 | PARK2 | √ | × | × | | rs10249420 | DDC | √ | × | × | | rs73157644 | KCNQ2 | √ | × | × | | rs1329090 | LOC101927 | √ | × | |
| rs9365460 | PARK2 | √ | × | × | | rs4656375 | DDR2 | × | √ | × | | rs6122118 | KCNQ2 | × | × | √ | | rs2798043 | LOC101927 | √ | × | |
| rs9356045 | PARK2 | √ | × | × | | rs6674132 | DDR2 | × | √ | × | | rs12626055 | KCNQ2 | × | × | √ | | rs1648473 | LOC101927 | √ | × | |
| rs9356047 | PARK2 | √ | × | × | | rs76052760 | DDR2 | × | √ | × | | rs3827019 | KCNQ2 | √ | × | × | | rs1777013 | LOC101927 | √ | × | |
| rs9356049 | PARK2 | √ | × | × | | rs7524537 | DDR2 | × | √ | × | | rs3827018 | KCNQ2 | √ | × | × | | rs1343100 | LOC101927 | √ | × | |
| rs9295192 | PARK2 | √ | × | × | | rs14938698 | DDR2 | × | √ | × | | rs3893041 | KCNQ2 | × | × | √ | | rs1754059 | LOC101927 | √ | × | |
| rs9295193 | PARK2 | √ | × | × | | rs14490426 | DDR2 | × | √ | × | | rs4083743 | KCNQ2 | × | × | √ | | rs1343101 | LOC101927 | √ | × | |
| rs2023016 | PARK2 | √ | × | × | Educational attainment (ye | rs14043807 | DDR2 | × | √ | × | | rs3787123 | KCNQ2 | × | × | √ | | rs2798053 | LOC101927 | √ | × | |
| rs10945841 | PARK2 | √ | × | × | | rs6684509 | DDR2 | × | √ | × | Height | rs3787120 | KCNQ2 | × | × | √ | | rs1571452 | LOC101927 | √ | × | |
| rs9364662 | PARK2 | √ | × | × | | rs74896112 | DDR2 | × | √ | × | | rs2302514 | KCNQ2 | × | × | √ | | rs10993252 | LOC101927 | √ | × | |
| rs9346922 | PARK2 | √ | × | × | | rs74345863 | DDR2 | × | √ | × | | rs3787118 | KCNQ2 | × | × | √ | | rs10821371 | LOC101927 | √ | × | |
| rs9365464 | PARK2 | √ | × | × | | rs57245690 | DDR2 | × | √ | × | | rs2270240 | KCNQ2 | × | × | √ | | rs1889205 | LOC101927 | √ | × | |
| rs9458597 | PARK2 | √ | × | × | | rs11781009 | DDR2 | × | √ | × | | rs73146513 | KCNQ2 | × | × | √ | | rs752788 | LOC101927 | √ | × | |
| rs12194834 | PARK2 | √ | × | × | | rs79775216 | DDR2 | × | √ | × | | rs2297384 | KCNQ2 | × | × | √ | | rs2004205 | LOC101927 | √ | × | |
| rs10945842 | PARK2 | √ | × | × | | rs10799854 | DDR2 | × | √ | × | | rs74434764 | KCNQ2 | × | × | √ | | rs10993323 | LOC101927 | √ | × | |
| rs6929604 | PARK2 | √ | × | × | | rs10917578 | DDR2 | × | √ | × | | rs2297385 | KCNQ2 | × | × | √ | | rs883922 | LOC101927 | √ | × | |
| rs6936107 | PARK2 | √ | × | × | | rs6676255 | DDR2 | × | √ | × | | rs77332976 | KCNQ2 | × | × | √ | | rs4744415 | LOC101927 | √ | × | |
| rs6934979 | PARK2 | √ | × | × | | rs6427690 | DDR2 | × | √ | × | | rs2282149 | KCNQ2 | × | × | √ | | rs10821416 | LOC101927 | √ | × | |
| rs4709640 | PARK2 | √ | × | × | | rs6427691 | DDR2 | × | √ | × | | rs61058914 | KCNQ2 | × | × | √ | | rs10821417 | LOC101927 | √ | × | |
| rs4709641 | PARK2 | √ | × | × | | rs11811703 | DDR2 | × | √ | × | | rs78423969 | KCNQ2 | × | × | √ | | rs913619 | LOC101927 | √ | × | |
| rs9365467 | PARK2 | √ | × | × | | rs7533399 | DDR2 | × | √ | × | | rs6122452 | KCNQ2 | × | × | √ | | rs913620 | LOC101927 | √ | × | |
| rs9458612 | PARK2 | √ | × | × | | rs10799855 | DDR2 | × | √ | × | | rs12480417 | KCNQ2 | × | × | √ | | rs913621 | LOC101927 | √ | × | |
| rs141727453 | PARK2 | √ | × | × | | rs10753582 | DDR2 | × | √ | × | | rs73918932 | KCNQ2 | × | × | √ | | rs913622 | LOC101927 | √ | × | |
| rs2846489 | PARK2 | √ | × | × | | rs7412891 | DDR2 | × | √ | × | | rs6122458 | KCNQ2 | × | × | √ | | rs56777946 | LOC101927 | √ | × | |
| rs2849557 | PARK2 | √ | × | × | | rs6667476 | DDR2 | × | √ | × | | rs6122459 | KCNQ2 | × | × | √ | | rs10821428 | LOC101927 | √ | × | |
| rs10945854 | PARK2 | √ | × | × | | rs6670722 | DDR2 | × | √ | × | | rs57946378 | KCNQ2 | × | × | √ | | rs10821429 | LOC101927 | √ | × | |
| rs17382723 | PASK | √ | × | × | Height | rs6683356 | DDR2 | × | √ | × | | rs4809568 | KCNQ2 | × | × | √ | | rs10761365 | LOC101927 | √ | × | |
| rs75369095 | PASK | √ | × | × | | rs6694368 | DDR2 | × | √ | × | | rs4809305 | KCNQ2 | × | × | √ | | rs10761367 | LOC101927 | √ | × | |
| rs76999699 | PASK | √ | × | × | | rs6694717 | DDR2 | × | √ | × | | rs4809569 | KCNQ2 | × | × | √ | | rs10993461 | LOC101927 | √ | × | |
| rs572773616 | PASK | √ | × | × | | rs4351614 | DDR2 | × | √ | × | | rs12481658 | KCNQ2 | × | × | √ | | rs80007285 | LOC101927 | √ | × | |
| rs10219577 | PCED1B | √ | × | × | | rs4567262 | DDR2 | × | √ | × | | rs1932592 | KIAA1217 | √ | × | × | | rs78311679 | LOC101927 | √ | × | |
| rs11183748 | PCED1B | √ | × | × | | rs76229902 | DDR2 | × | √ | × | | rs7077294 | KIAA1217 | √ | × | × | | rs13219865 | LOC101927 | √ | × | |
| rs10219604 | PCED1B | √ | × | × | | rs6689095 | DDR2 | × | √ | × | | rs6482386 | KIAA1217 | √ | × | × | | rs13194849 | LOC101927 | √ | × | |
| rs13377855 | PCED1B | √ | × | × | | rs6697494 | DDR2 | × | √ | × | | rs4418704 | KIAA1217 | √ | × | × | | rs6930651 | LOC101927 | √ | × | |
| rs10506272 | PCED1B | √ | × | × | | rs10047158 | DDR2 | × | √ | × | | rs4330993 | KIAA1217 | √ | × | × | | rs6915179 | LOC101927 | √ | × | |
| rs12581006 | PCED1B | √ | × | × | | rs6687170 | DDR2 | × | √ | × | | rs4434904 | KIAA1217 | √ | × | × | | rs7771575 | LOC101927 | √ | × | |
| rs7484989 | PCED1B | √ | × | × | | rs6698107 | DDR2 | × | √ | × | | rs10828646 | KIAA1217 | √ | × | × | | rs62420461 | LOC101927 | √ | × | |
| rs1917663 | PCED1B | √ | × | × | | rs74876216 | DDR2 | × | √ | × | | rs7906883 | KIAA1217 | √ | × | × | | rs75227599 | LOC101927 | √ | × | |
| rs4768776 | PCED1B | √ | × | × | | rs7553831 | DDR2 | × | √ | × | | rs6482387 | KIAA1217 | √ | × | × | | rs9490771 | LOC101927 | √ | × | |
| rs1404820 | PCED1B | √ | × | × | | rs11775924 | DDR2 | × | √ | × | | rs6482388 | KIAA1217 | √ | × | × | | rs12333042 | LOC101927 | √ | × | |
| rs1404819 | PCED1B | √ | × | × | | rs7519400 | DDR2 | × | √ | × | | rs7091541 | KIAA1217 | √ | × | × | | rs9490776 | LOC101927 | √ | × | |
| rs10881052 | PCED1B | √ | × | × | | rs7554305 | DDR2 | × | √ | × | | rs4456169 | KIAA1217 | √ | × | × | | rs6933435 | LOC101927 | √ | × | |
| rs1525452 | PCED1B | √ | × | × | | rs19140114 | DDR2 | × | √ | × | | rs7475360 | KIAA1217 | √ | × | × | | rs6933479 | LOC101927 | √ | × | |
| rs9634259 | PCED1B | √ | × | × | | rs12563679 | DDR2 | × | √ | × | | rs3858211 | KIAA1217 | √ | × | × | | rs75407261 | LOC101927 | √ | × | |
| rs148060127 | PCED1B | √ | × | × | | rs77072320 | DDR2 | × | √ | × | | rs7896427 | KIAA1217 | √ | × | × | | rs98775316 | LOC101929 | × | √ | |
| rs11183766 | PCED1B | √ | × | × | | rs77926587 | DDR2 | × | √ | × | | rs6482389 | KIAA1217 | √ | × | × | | rs91018295 | LOC101929 | × | √ | |
| rs3784508 | PCSK6 | √ | × | × | | rs35570648 | DDX50 | × | √ | × | | rs7091497 | KIAA1217 | √ | × | × | | rs14185929 | LOC101929 | √ | × | |
| rs80078428 | PCSK6 | √ | × | × | | rs2894071 | DDX50 | × | √ | × | | rs19260681 | KIAA1217 | √ | × | × | | rs18852642 | LOC151174 | × | √ | |
| rs2073590 | PCSK6 | × | √ | × | | rs2394498 | DDX50 | × | √ | × | | rs15069625 | KIFC3 | √ | × | × | | rs73104366 | LOC151174 | × | √ | |
| rs58979969 | PCSK6 | × | √ | × | | rs18081418 | DDX50 | × | √ | × | | rs528056 | KMT2A | × | × | √ | | rs77858523 | LOC151174 | × | √ | |
| rs190284093 | PDCD7 | × | √ | × | | rs5030892 | DDX50 | × | √ | × | | rs561775 | KMT2A | × | × | √ | | rs12151284 | LOC284395 | √ | × | |
| rs36124720 | PER2 | √ | × | × | | rs10762247 | DDX50 | × | √ | × | | rs535327 | KMT2A | × | × | √ | | rs73541109 | LOC284395 | √ | × | |
| rs73088916 | PER2 | √ | × | × | | rs690010 | DIRAS2 | √ | × | × | | rs562780 | KMT2A | × | × | √ | | rs1549508 | LOC340515 | √ | × | |
| rs78832829 | PER2 | √ | × | × | | rs13981470 | DMXL1 | × | √ | × | | rs537975 | KMT2A | × | × | √ | | rs11782726 | LOC642943 | √ | × | |
| rs78445704 | PER2 | √ | × | × | | rs11758451 | DMXL1 | × | √ | × | | rs477106 | KMT2A | × | × | √ | | rs4973649 | LOC728323 | √ | × | |
| rs141670620 | PHLDB1 | √ | × | × | | rs11792931 | DMXL1 | × | √ | × | | rs573971 | KMT2A | × | × | √ | | rs12617606 | LOC728323 | √ | × | |
| rs10151023 | PIGH | √ | × | × | Adolescent idiopathic scoli | rs13878755 | DMXL1 | × | √ | × | | rs1133153 | KRT6A | √ | × | × | | rs57934606 | LRRFIP1 | √ | × | |
| rs28558546 | PIGH | √ | × | × | | rs19217068 | DMXL1 | × | √ | × | | rs28667515 | KRT6A | √ | × | × | | rs11803106 | LRRFIP1 | √ | × | |
| rs2319844 | PIGH | √ | × | × | Adolescent idiopathic scoli | rs56139912 | DMXL1 | × | √ | × | | rs188463 | KRT6A | √ | × | × | | rs2279643 | LRRFIP1 | √ | × | |
| rs79535433 | PLCL2 | √ | × | × | | rs4653301 | DNALI1 | √ | × | × | Intraocular p | rs298117 | KRT6A | √ | × | × | | rs77719131 | LRRFIP1 | √ | × | |
| rs190143802 | PLCL2 | √ | × | × | | rs6781 | DPM2 | √ | × | × | | rs262447 | LDLRAD3 | √ | × | × | | rs12362660 | LUZP2 | √ | × | |
| rs6780191 | PLCL2 | √ | × | × | | rs6930849 | ECHDC1 | √ | × | × | | rs262445 | LDLRAD3 | √ | × | × | | rs12805462 | LUZP2 | √ | × | |
| rs4685409 | PLCL2 | √ | × | × | | rs7453272 | ECHDC1 | √ | × | × | | rs37582137 | LDLRAD3 | √ | × | × | | rs11028026 | LUZP2 | √ | × | |
| rs76903510 | PLCL2 | √ | × | × | | rs9388561 | ECHDC1 | √ | × | × | | rs9566298 | LINC00571 | √ | × | × | | rs1553740 | LUZP2 | √ | × | |
| rs1806556 | PLCL2 | √ | × | × | | rs9398837 | ECHDC1 | √ | × | × | | rs9566305 | LINC00571 | √ | × | × | | rs1553741 | LUZP2 | √ | × | |
| rs7653027 | PLCL2 | √ | × | × | | rs9398838 | ECHDC1 | √ | × | × | | rs11155027 | LINC00698 | √ | × | × | | rs2036752 | LUZP2 | √ | × | |
| rs80049518 | PLCL2 | √ | × | × | | rs2057223 | ECHDC1 | √ | × | × | | rs11800204 | LINC01107 | √ | × | × | | rs1553742 | LUZP2 | √ | × | |
| rs79232960 | PLCL2 | √ | × | × | | rs7776136 | ECHDC1 | √ | × | × | | rs14809418 | LINC01107 | √ | × | × | | rs10834388 | LUZP2 | √ | × | |
| rs6442667 | PLCL2 | √ | × | × | | rs6903878 | ECHDC1 | √ | × | × | | rs67980076 | LINC01107 | √ | × | × | | rs5008808 | LUZP2 | √ | × | |
| rs4563393 | PLCL2 | √ | × | × | | rs9285460 | ECHDC1 | √ | × | × | | rs76532868 | LOC101927 | × | √ | × | | rs1509588 | LUZP2 | √ | × | |
| rs17103488 | PLEKHA1 | √ | × | × | | rs4895822 | ECHDC1 | √ | × | × | | rs77958011 | LOC101927 | × | √ | × | | rs1509589 | LUZP2 | √ | × | |
| rs4565845 | PLEKHA1 | √ | × | × | | rs6937287 | ECHDC1 | √ | × | × | | rs13844868 | LOC101927 | × | √ | × | | rs1509590 | LUZP2 | √ | × | |
| rs3742868 | PLEKHH1 | √ | × | × | | rs7756336 | ECHDC1 | √ | × | × | | rs6595125 | LOC101927 | × | √ | × | | rs981895 | LUZP2 | √ | × | |
| rs7141944 | PLEKHH1 | √ | × | × | | rs13218061 | ECHDC1 | √ | × | × | | rs53508837 | LOC101927 | × | √ | × | | rs10834389 | LUZP2 | √ | × | |
| rs10827903 | PLXDC2 | √ | × | × | | rs9401950 | ECHDC1 | √ | × | × | | rs76139122 | LOC101927 | × | √ | × | | rs11600443 | LUZP2 | √ | × | |
| rs10764179 | PLXDC2 | √ | × | × | | rs9321076 | ECHDC1 | √ | × | × | | rs80154912 | LOC101927 | × | √ | × | | rs10834391 | LUZP2 | √ | × | |
| rs7898627 | PLXDC2 | √ | × | × | | rs12196700 | ELOVL5 | √ | × | × | | rs77786927 | LOC101927 | × | √ | × | | rs11028028 | LUZP2 | √ | × | |
| rs1326233 | PLXDC2 | √ | × | × | | rs209483 | ELOVL5 | √ | × | × | | rs12518839 | LOC101927 | × | √ | × | | rs10767214 | LUZP2 | √ | × | |
| rs10764180 | PLXDC2 | √ | × | × | | rs209485 | ELOVL5 | √ | × | × | | rs12513737 | LOC101927 | × | √ | × | | rs10834392 | LUZP2 | √ | × | |
| rs1575869 | PLXDC2 | √ | × | × | Blood pressure (pleiotropy | rs7208857 | EPN2 | √ | × | × | | rs34520222 | LOC101927 | × | √ | × | | rs11028036 | LUZP2 | √ | × | |
| rs11011697 | PLXDC2 | √ | × | × | | rs9371227 | ESR1 | √ | × | × | | rs77028648 | LOC101927 | × | √ | × | | rs931964 | LUZP2 | √ | × | Smoking initiation |
| rs11011698 | PLXDC2 | √ | × | × | | rs9479126 | ESR1 | √ | × | × | Height | rs79426516 | LOC101927 | × | √ | × | | rs1858156 | LUZP2 | √ | × | |
| rs1926204 | PLXDC2 | √ | × | × | | rs7935138 | EXT2 | √ | × | × | | rs18546190 | LOC101927 | × | √ | × | | rs12225488 | LUZP2 | √ | × | |
| rs7923799 | PLXDC2 | √ | × | × | | rs12801050 | EXT2 | √ | × | × | | rs36874509 | LOC101927 | × | √ | × | | rs20221501 | METAP2 | √ | × | |
| rs10827916 | PLXDC2 | √ | × | × | | rs72901924 | EXT2 | √ | × | × | | rs11951938 | LOC101927 | × | √ | × | | rs6747400 | METTL8 | √ | × | |
| rs717144 | POLR1A | √ | × | × | | rs11037881 | EXT2 | √ | × | × | | rs9327072 | LOC101927 | × | √ | × | | rs12616940 | METTL8 | √ | × | |
| rs79057302 | POLR1A | √ | × | × | | rs11037882 | EXT2 | √ | × | × | | rs11761210 | LOC101927 | × | √ | × | | rs1558963 | METTL8 | √ | × | |
| rs75323994 | POLR1A | √ | × | × | | rs7117287 | EXT2 | √ | × | × | | rs57761190 | LOC101927 | × | √ | × | | rs62183460 | METTL8 | √ | × | |
| rs12534317 | PON3 | √ | × | × | | rs7945703 | EXT2 | √ | × | × | | rs15052376 | LOC101927 | × | √ | × | | rs58618629 | MIR548AC | √ | × | |
| rs891060 | POP4 | √ | × | × | | rs4755782 | EXT2 | √ | × | × | | rs1571452 | LOC101927 | √ | × | × | | rs4858517 | MIR548AC | √ | × | |
| rs2932966 | PPARGC1A | √ | × | × | Smoking initiation | rs4551754 | EXT2 | √ | × | × | Male-pattern | rs10821371 | LOC101927 | √ | × | × | | rs17405137 | MIR548AC | √ | × | |
| rs2970886 | PPARGC1A | √ | × | × | | rs3935072 | EXT2 | √ | × | × | | rs10432691 | LOC101927 | √ | × | × | | rs11493390 | MIR548AC | √ | × | |
| rs2970887 | PPARGC1A | √ | × | × | | rs3935820 | EXT2 | √ | × | × | | rs10207205 | LOC101927 | √ | × | × | | rs73041534 | MIR548AC | √ | × | |
| rs2970845 | PPARGC1A | √ | × | × | | rs7120534 | EXT2 | √ | × | × | | rs10209578 | LOC101927 | √ | × | × | | rs704535 | MIR548AC | √ | × | |
| rs1472095 | PPARGC1A | √ | × | × | | rs4755787 | EXT2 | √ | × | × | | rs4538235 | LOC101927 | √ | × | × | | rs778457 | MIR548AC | √ | × | |
| rs2932970 | PPARGC1A | √ | × | × | | rs12798405 | EXT2 | √ | × | × | | rs4322870 | LOC101927 | √ | × | × | | rs73041557 | MIR548AC | √ | × | |
| rs2932971 | PPARGC1A | √ | × | × | | rs7112070 | EXT2 | √ | × | × | | rs7558985 | LOC101927 | √ | × | × | | rs1695205 | MIR548AC | √ | × | |
| rs2970851 | PPARGC1A | √ | × | × | | rs4755231 | EXT2 | √ | × | × | | rs7588183 | LOC101927 | √ | × | × | | rs14919374 | MIR548AC | √ | × | |
| rs2970852 | PPARGC1A | √ | × | × | | rs903512 | EXT2 | √ | × | × | | rs7575968 | LOC101927 | √ | × | × | | rs778463 | MIR548AC | √ | × | |
| rs2932975 | PPARGC1A | √ | × | × | | rs962848 | EXT2 | √ | × | × | | rs13403346 | LOC101927 | √ | × | × | | rs778469 | MIR548AC | √ | × | |
| rs2932976 | PPARGC1A | √ | × | × | | rs12792891 | EXT2 | √ | × | × | | rs13410776 | LOC101927 | √ | × | × | | rs1695180 | MIR548AC | √ | × | |
| rs57140917 | PPARGC1A | √ | × | × | | rs4755233 | EXT2 | √ | × | × | | rs4268948 | LOC101927 | √ | × | × | | rs1692619 | MIR548AC | √ | × | |
| rs251468 | PPARGC1B | × | √ | × | Freckles | rs12280077 | EXT2 | √ | × | × | | rs6722175 | LOC101927 | √ | × | × | | rs62256973 | MIR548AC | √ | × | |
| rs251466 | PPARGC1B | × | √ | × | Sunburns | rs7120281 | EXT2 | √ | × | × | | rs1979562 | LOC101927 | √ | × | × | | rs68093882 | MIR548AC | √ | × | |
| rs17110463 | PPARGC1B | × | √ | × | | rs11037900 | EXT2 | √ | × | × | | rs12616226 | LOC101927 | √ | × | × | | rs14601739 | MIR548AC | √ | × | |
| rs192884812 | PPARGC1B | × | √ | × | | rs4755234 | EXT2 | √ | × | × | Height | rs12622746 | LOC101927 | √ | × | × | | rs79965216 | MIR548AC | √ | × | |
| rs251465 | PPARGC1B | × | √ | × | Squamous cell carcinoma ( | rs869915 | EXT2 | √ | × | × | | rs875280 | LOC101927 | √ | × | × | | rs14019972 | MIR548AC | √ | × | |
| rs251464 | PPARGC1B | × | √ | × | Low tan response | rs12790182 | EXT2 | √ | × | × | | rs875279 | LOC101927 | √ | × | × | | rs7609845 | MIR548AC | √ | × | |
| rs251460 | PPARGC1B | × | √ | × | | rs1845618 | EXT2 | √ | × | × | | rs1386382 | LOC101927 | √ | × | × | | rs57859482 | MIR548AC | √ | × | |
| rs17110586 | PPARGC1B | × | √ | × | | rs1113132 | EXT2 | √ | × | × | | rs10198550 | LOC101927 | √ | × | × | | rs12486880 | MIR548AC | √ | × | |
| rs32582 | PPARGC1B | × | √ | × | | rs7945827 | EXT2 | √ | × | × | | rs10175035 | LOC101927 | √ | × | × | | rs12676399 | MSR1 | √ | × | |
| rs32581 | PPARGC1B | × | √ | × | | rs13880932 | F13B | √ | × | × | | rs13408424 | LOC101927 | √ | × | × | | rs6981231 | MSR1 | √ | × | |
| rs2003604 | PPARGC1B | × | √ | × | | rs10819322 | FAM102A | √ | × | × | | rs13382961 | LOC101927 | √ | × | × | | rs13306539 | MSR1 | √ | × | |
| rs741582 | PPARGC1B | × | √ | × | | rs3802355 | FAM102A | √ | × | × | | rs17596137 | LOC101927 | √ | × | × | | rs13306538 | MSR1 | √ | × | |
| rs143344511 | PPARGC1B | × | √ | × | | rs2417062 | FAM102A | √ | × | × | | rs7570425 | LOC101927 | √ | × | × | | rs13306537 | MSR1 | √ | × | |
| rs11749124 | PPARGC1B | × | √ | × | | rs6415846 | FAM102A | √ | × | × | | rs7609524 | LOC101927 | √ | × | × | | rs7005569 | MSR1 | √ | × | |
| rs45560442 | PPARGC1B | × | √ | × | | rs6478807 | FAM102A | √ | × | × | | rs1601653 | LOC101927 | × | √ | × | | rs11992232 | MSR1 | √ | × | |
| rs11745113 | PPARGC1B | × | √ | × | | rs2031694 | FAM102A | √ | × | × | | rs7586947 | LOC101927 | × | √ | × | | rs12678689 | MSR1 | √ | × | |
| rs558608743 | PPARGC1B | × | √ | × | | rs7026115 | FAM102A | √ | × | × | | rs10865431 | LOC101927 | × | √ | × | | rs12678705 | MSR1 | √ | × | |
| rs32579 | PPARGC1B | × | √ | × | Tanning | rs79356092 | FAM102A | √ | × | × | | rs78176647 | LOC101927 | × | √ | × | | rs12681374 | MSR1 | √ | × | |
| rs190151814 | PPARGC1B | × | √ | × | | rs12618914 | FAM132B | √ | × | × | | rs4853346 | LOC101927 | × | √ | × | | rs12678776 | MSR1 | √ | × | |
| rs13178617 | PPARGC1B | × | √ | × | | rs34989058 | FAM132B | √ | × | × | | rs18219748 | LOC101927 | × | √ | × | | rs12678780 | MSR1 | √ | × | |
| rs541803390 | PPARGC1B | × | √ | × | | rs16970167 | FAM155A | √ | × | × | | rs10173222 | LOC101927 | × | √ | × | | rs34874653 | MSR1 | √ | × | |
| rs45520937 | PPARGC1B | × | √ | × | | rs1924446 | FAM155A | √ | × | × | | rs10422264 | LOC284395 | √ | × | × | | rs12114258 | MSR1 | √ | × | |
| rs17572019 | PPARGC1B | × | √ | × | | rs14855101 | FAM155A | √ | × | × | | rs4832233 | LOC90784 | √ | × | × | | rs12114597 | MSR1 | √ | × | |
| rs11959820 | PPARGC1B | × | √ | × | | rs79203238 | FAM155A | × | × | √ | | rs2281836 | LRR1 | √ | × | × | | rs12114599 | MSR1 | √ | × | |
| rs12659862 | PPARGC1B | × | √ | × | | rs74112579 | FAM155A | × | × | √ | | rs19262378 | LRR1 | √ | × | × | | rs12114791 | MSR1 | √ | × | |
| rs32576 | PPARGC1B | × | √ | × | Obesity-related traits | rs8000666 | FAM155A | × | × | √ | | rs2354444 | LRR1 | √ | × | × | | rs12114600 | MSR1 | √ | × | |
| rs4549516 | PPARGC1B | × | √ | × | | rs74112582 | FAM155A | × | × | √ | | rs2354445 | LRR1 | √ | × | × | | rs3789014 | MSR1 | √ | × | |
| rs567432040 | PPARGC1B | × | √ | × | | rs74112583 | FAM155A | × | × | √ | | rs4900940 | LRR1 | √ | × | × | | rs9650393 | MSR1 | √ | × | |
| rs12652075 | PPARGC1B | × | √ | × | | rs79164930 | FAM155A | × | × | √ | | rs14912945 | LRRFIP1 | √ | × | × | | rs12114633 | MSR1 | √ | × | |
| rs1024978 | PPARGC1B | × | √ | × | | rs76866938 | FAM155A | × | × | √ | | rs18826223 | LRRFIP1 | √ | × | × | | rs8005552 | MTA1 | × | √ | |
| rs1024979 | PPARGC1B | × | √ | × | | rs77769732 | FAM155A | × | × | √ | | rs19278540 | LRRFIP1 | √ | × | × | | rs18600297 | MTA1 | × | √ | |
| rs32575 | PPARGC1B | × | √ | × | | rs16971011 | FAM155A | × | × | √ | | rs11884774 | LRRFIP1 | √ | × | × | | rs7143366 | MTA1 | × | √ | |
| rs62382346 | PPARGC1B | × | √ | × | | rs77106962 | FAM155A | × | × | √ | | rs79685677 | LRRFIP1 | √ | × | × | | rs74092385 | MTA1 | × | √ | |
| rs962241073 | PPARGC1B | × | √ | × | | rs12583396 | FAM155A | × | × | √ | | rs3769107 | LRRFIP1 | √ | × | × | | rs11357328 | MTA1 | × | √ | |
| rs186462923 | PPARGC1B | × | √ | × | | rs72657240 | FAM155A | × | × | √ | | rs14831285 | LRRFIP1 | √ | × | × | | rs37408746 | MTA1 | × | √ | |
| rs45588534 | PPARGC1B | × | √ | × | | rs16971099 | FAM155A | × | × | √ | | rs14147389 | LRRFIP1 | √ | × | × | | rs37690876 | MTA1 | × | √ | |
| rs45543631 | PPARGC1B | × | √ | × | | rs72657247 | FAM155A | √ | × | × | | rs73096891 | LRRFIP1 | √ | × | × | | rs4983413 | MTA1 | × | √ | Non-albumin protein levels |
| rs10783180 | PPARGC1B | × | √ | × | | rs78070602 | FAM155A | √ | × | × | | rs15013867 | LRRFIP1 | √ | × | × | | rs11845995 | MTA1 | × | √ | |
| rs12659181 | PPARGC1B | × | √ | × | | rs6499867 | FAM192A | √ | × | × | | rs13872385 | LRRFIP1 | √ | × | × | | rs18151939 | MTA1 | × | √ | |
| rs58560614 | PPARGC1B | × | √ | × | | rs72721609 | FBXW7 | √ | × | × | | rs14182928 | LRRFIP1 | √ | × | × | | rs8181908 | MTA1 | × | √ | |
| rs11953848 | PPARGC1B | × | √ | × | | rs79578032 | FBXW7 | √ | × | × | | rs76073889 | LRRFIP1 | √ | × | × | | rs9671696 | MTA1 | × | √ | |
| rs32574 | PPARGC1B | × | √ | × | | rs11756751 | FBXW7 | √ | × | × | | rs77743076 | LRRFIP1 | √ | × | × | | rs58764201 | MTA1 | × | √ | |
| rs188040812 | PPARGC1B | × | √ | × | | rs2714798 | FBXW7 | √ | × | × | | rs10174113 | LRRFIP1 | √ | × | × | | rs19074587 | MTA1 | × | √ | |
| rs77241719 | PPARGC1B | × | √ | × | | rs59584006 | FGF12 | √ | × | × | | rs2249649 | LRSAM1 | √ | × | × | | rs58766661 | MTA1 | × | √ | |
| rs62382349 | PPARGC1B | × | √ | × | | rs11783463 | FIGNL1 | × | √ | × | | rs1539569 | LRSAM1 | √ | × | × | | rs4983414 | MTA1 | × | √ | |
| rs17653642 | PPARGC1B | × | √ | × | | rs1470746 | FIGNL1 | × | √ | × | | rs75339299 | LUZP2 | √ | × | × | | rs6737057 | MTERFD2 | √ | × | |
| rs11953722 | PPARGC1B | × | √ | × | | rs10230307 | FIGNL1 | × | √ | × | | rs13930595 | MAGI2 | √ | × | × | | rs4942030 | MTRF1 | √ | × | |
| rs26127 | PPARGC1B | × | √ | × | | rs10230343 | FIGNL1 | × | √ | × | | rs12407624 | 45353 | √ | × | × | | rs7321509 | NAA16 | √ | × | |
| rs562548277 | PPARGC1B | × | √ | × | | rs10235371 | FIGNL1 | × | √ | × | | rs72657771 | MEAF6 | √ | × | × | | rs9562292 | NAA16 | √ | × | |
| rs144054131 | PPARGC1B | × | √ | × | | rs3735276 | FIGNL1 | × | √ | × | | rs80070498 | METTL8 | √ | × | × | | rs9566753 | NAA16 | √ | × | |
| rs6879670 | PPARGC1B | × | √ | × | | rs10236132 | FIGNL1 | × | √ | × | | rs62181356 | METTL8 | √ | × | × | | rs9566754 | NAA16 | √ | × | |
| rs62383783 | PPARGC1B | × | √ | × | | rs10760644 | FNBP1 | √ | × | × | | rs17405137 | MIR548AC | √ | × | × | | rs3829396 | NAA16 | √ | × | |
| rs58696088 | PPARGC1B | × | √ | × | | rs10760645 | FNBP1 | √ | × | × | | rs77864375 | MIR548AC | √ | × | × | | rs9562293 | NAA16 | √ | × | |
| rs761308897 | PPARGC1B | × | √ | × | | rs10819574 | FNBP1 | √ | × | × | | rs73041534 | MIR548AC | √ | × | × | | rs9562294 | NAA16 | √ | × | |
| rs143268818 | PPARGC1B | × | √ | × | | rs2007126 | FNBP1 | √ | × | × | LDL peak p | rs778485 | MIR548AC | √ | × | × | | rs9566757 | NAA16 | √ | × | |
| rs368936505 | PPARGC1B | × | √ | × | | rs4837430 | FNBP1 | √ | × | × | | rs778486 | MIR548AC | √ | × | × | Acceptance | rs3776839 | NDST1 | √ | × | |
| rs554388706 | PPARGC1B | × | √ | × | | rs41486950 | GARNL3 | × | √ | × | | rs778507 | MIR548AC | √ | × | × | | rs76456883 | NDST1 | √ | × | |
| rs56938831 | PPARGC1B | × | √ | × | | rs37156874 | GARNL3 | × | √ | × | | rs73041557 | MIR548AC | √ | × | × | | rs12657104 | NDST1 | √ | × | |
| rs61408734 | PPARGC1B | × | √ | × | | rs72766253 | GARNL3 | × | √ | × | | rs778459 | MIR548AC | √ | × | × | | rs3822608 | NDST1 | √ | × | |
| rs6579761 | PPARGC1B | × | √ | × | | rs59443037 | GAS8 | √ | × | × | | rs778462 | MIR548AC | √ | × | × | | rs76640203 | NDST1 | √ | × | |
| rs25846 | PPARGC1B | × | √ | × | | rs45476600 | GCM1 | × | √ | × | | rs778471 | MIR548AC | √ | × | × | | rs75541727 | NDST1 | √ | × | |
| rs1549186 | PPARGC1B | × | √ | × | | rs9370179 | GCM1 | × | √ | × | | rs778468 | MIR548AC | √ | × | × | | rs79141975 | NDST1 | √ | × | |
| rs1549188 | PPARGC1B | × | √ | × | | rs7171499 | GNB5 | √ | × | × | | rs14429111 | MIR548AC | √ | × | × | | rs1045538 | NDST1 | √ | × | |
| rs26123 | PPARGC1B | × | √ | × | | rs60705168 | GNB5 | √ | × | × | | rs11688185 | MIR548AC | √ | × | × | | rs1045539 | NDST1 | √ | × | |
| rs26122 | PPARGC1B | × | √ | × | | rs28451227 | GNB5 | √ | × | × | | rs11773415 | MIR548AC | √ | × | × | | rs1045544 | NDST1 | √ | × | |
| rs62383788 | PPARGC1B | × | √ | × | | rs4494485 | GNB5 | √ | × | × | | rs37513901 | MIR548AC | √ | × | × | | rs58575086 | NDST1 | √ | × | |
| rs1107344 | PPARGC1B | × | √ | × | | rs4652961 | GNL2 | √ | × | × | | rs73041601 | MIR548AC | √ | × | × | | rs78527679 | NELL1 | √ | × | |
| rs26121 | PPARGC1B | × | √ | × | | rs75356281 | GNPAT | × | × | √ | | rs7632099 | MIR548AC | √ | × | × | | rs1955066 | NELL1 | √ | × | |
| rs26119 | PPARGC1B | × | √ | × | | rs75761810 | GNPAT | × | × | √ | | rs11775651 | MIR548AC | √ | × | × | | rs79700659 | NELL1 | √ | × | |
| rs2341294 | PPARGC1B | × | √ | × | | rs4947432 | GRB10 | √ | × | × | | rs11688503 | MIR548AC | √ | × | × | | rs78807828 | NELL1 | √ | × | |
| rs888853 | PPARGC1B | × | √ | × | | rs10216232 | GRB10 | √ | × | × | | rs19195706 | MIR548AC | √ | × | × | | rs77024347 | NELL1 | √ | × | |
| rs7712296 | PPARGC1B | × | √ | × | | rs14853943 | GRB10 | √ | × | × | | rs73043677 | MIR548AC | √ | × | × | | rs79127320 | NKTR | √ | × | |
| rs3733665 | PPARGC1B | × | √ | × | | rs11974751 | GRB10 | √ | × | × | | rs4574222 | MIR548AC | √ | × | × | | rs3095258 | NKTR | √ | × | |
| rs536083791 | PPARGC1B | × | √ | × | | rs6593182 | GRB10 | × | √ | × | | rs77689491 | MIR548AC | √ | × | × | | rs339735 | NKTR | √ | × | |
| rs118087243 | PPARGC1B | × | √ | × | | rs4947890 | GRB10 | × | √ | × | | rs17013418 | MIR548AC | √ | × | × | | rs168138 | NKTR | √ | × | |
| rs393499 | PPARGC1B | × | √ | × | | rs7791286 | GRB10 | × | √ | × | Fasting plas | rs73043697 | MIR548AC | √ | × | × | | rs398743 | NKTR | √ | × | |
| rs439598 | PPARGC1B | × | √ | × | | rs80204191 | GRK5 | √ | × | × | | rs79764943 | MIR548AC | √ | × | × | | rs12462690 | NLRP5 | × | √ | |
| rs16843395 | PPP1R7 | √ | × | × | | rs75529647 | GRK5 | √ | × | × | | rs17013422 | MIR548AC | √ | × | × | | rs12462663 | NLRP5 | × | √ | |
| rs10282116 | PPP1R9A | √ | × | × | | rs75839391 | GRK5 | √ | × | × | | rs79937641 | MIR548AC | √ | × | × | | rs513949 | NLRP5 | × | √ | |
| rs35233485 | PPP1R9A | √ | × | × | | rs75094288 | GRK5 | √ | × | × | | rs74431036 | MIR548AC | √ | × | × | | rs74706913 | NLRP5 | × | √ | |
| rs183019886 | PPP1R9A | √ | × | × | | rs79891128 | GRK5 | √ | × | × | | rs17013429 | MIR548AC | √ | × | × | | rs10252513 | NRF1 | √ | × | |
| rs80209508 | PPP1R9A | √ | × | × | | rs77367893 | GRK5 | √ | × | × | | rs11672908 | MIR548AC | √ | × | × | | rs10808240 | NRF1 | √ | × | |
| rs1527722 | PPP1R9A | √ | × | × | | rs77724107 | GRK5 | √ | × | × | | rs75493041 | MIR548AC | √ | × | × | | rs4731608 | NRF1 | √ | × | |
| rs56795270 | PPP1R9A | √ | × | × | | rs79124070 | GRK5 | √ | × | × | | rs11543790 | MIR548AC | √ | × | × | | rs17555614 | NRF1 | √ | × | |
| rs73431071 | PPP1R9A | √ | × | × | | rs76581632 | GRK5 | √ | × | × | | rs15098537 | MIR548AC | √ | × | × | | rs80257343 | NRF1 | √ | × | |
| rs73431078 | PPP1R9A | √ | × | × | | rs76590955 | GRK5 | √ | × | × | | rs78979035 | MIR548AC | √ | × | × | | rs4292630 | NRF1 | √ | × | |

| | | | | | |
|---|---|---|---|---|---|
| rs11981429 | PPP1R9A | √ | × | × | |
| rs58602684 | PPP1R9A | √ | × | × | |
| rs12704790 | PPP1R9A | √ | × | × | |
| rs2374983 | PPP1R9A | √ | × | × | |
| rs1527720 | PPP1R9A | √ | × | × | |
| rs2272364 | PPP1R9A | √ | × | × | |
| rs1342152 | PPP2R5A | √ | × | × | |
| rs12137798 | PPP2R5A | √ | × | × | |
| rs571122744 | PPP6R2 | √ | × | × | |
| rs76542703 | PRELP | × | × | √ | |
| rs516741 | PRKCI | √ | × | × | |
| rs1900337 | PRR5L | √ | × | × | |
| rs4755449 | PRR5L | √ | × | × | |
| rs2467080 | PRR5L | √ | × | × | |
| rs796863158 | PRSS3 | √ | × | × | |
| rs796651612 | PRSS3 | √ | × | × | |
| rs974456855 | PRSS3 | √ | × | × | |
| rs921715354 | PRSS3 | √ | × | × | |
| rs796731252 | PRSS3 | √ | × | × | |
| rs1015380765 | PRSS3 | √ | × | × | |
| rs796106360 | PRSS3 | √ | × | × | |
| rs879203642 | PRSS3 | √ | × | × | |
| rs796449713 | PRSS3 | √ | × | × | |
| rs796530510 | PRSS3 | √ | × | × | |
| rs377380263 | PRSS3 | √ | × | × | |
| rs373120184 | PRSS3 | √ | × | × | |
| rs7001606 | PSD3 | √ | × | × | |
| rs73202193 | PSD3 | √ | × | × | |
| rs11784303 | PSD3 | √ | × | × | |
| rs11774866 | PSD3 | √ | × | × | |
| rs11775869 | PSD3 | √ | × | × | |
| rs59758306 | PSD3 | √ | × | × | |
| rs57334374 | PSD3 | √ | × | × | |
| rs67940952 | PSD3 | √ | × | × | |
| rs11775925 | PSD3 | √ | × | × | |
| rs11776741 | PSD3 | √ | × | × | |
| rs60075374 | PSD3 | √ | × | × | |
| rs56313137 | PSD3 | √ | × | × | |
| rs67830348 | PSD3 | √ | × | × | |
| rs11988566 | PSD3 | √ | × | × | |
| rs67340643 | PSD3 | √ | × | × | |
| rs61363834 | PSD3 | √ | × | × | |
| rs56851678 | PSD3 | √ | × | × | |
| rs35313683 | PSD3 | √ | × | × | |
| rs7012662 | PSD3 | √ | × | × | |
| rs67892585 | PSD3 | √ | × | × | |
| rs67478477 | PSD3 | √ | × | × | |
| rs17384473 | PSD3 | √ | × | × | |
| rs17469017 | PSD3 | √ | × | × | |
| rs17469038 | PSD3 | √ | × | × | |
| rs11774278 | PSD3 | √ | × | × | |
| rs11780329 | PSD3 | √ | × | × | |
| rs2114494 | PSD3 | √ | × | × | |
| rs57280217 | PSD3 | √ | × | × | |
| rs56277438 | PSD3 | √ | × | × | |
| rs73204530 | PSD3 | √ | × | × | |
| rs1426917 | PSD3 | √ | × | × | |
| rs73204534 | PSD3 | √ | × | × | |
| rs11783114 | PSD3 | √ | × | × | |
| rs118097993 | PTCD3 | √ | × | × | |
| rs140197454 | RALGPS1 | √ | × | × | |
| rs60025121 | RALGPS1 | √ | × | × | |
| rs56659927 | RALGPS1 | √ | × | × | |
| rs62580804 | RALGPS1 | × | √ | × | |
| rs3936030 | RALGPS1 | × | √ | × | |
| rs62580836 | RALGPS1 | × | √ | × | |
| rs17368219 | RALGPS1 | × | √ | × | |
| rs17446684 | RALGPS1 | × | √ | × | |
| rs558921798 | RALGPS1 | × | √ | × | |
| rs12554538 | RALGPS1 | × | √ | × | |
| rs4292797 | RALGPS1 | × | √ | × | |
| rs17446804 | RALGPS1 | × | √ | × | |
| rs10819264 | RALGPS1 | × | √ | × | |
| rs10987552 | RALGPS1 | × | √ | × | |
| rs117207399 | RALGPS1 | × | √ | × | |
| rs137910316 | RALGPS1 | × | √ | × | |
| rs78889029 | RALGPS1 | × | √ | × | |
| rs826532 | RANBP2 | √ | × | × | |
| rs1724203 | RANBP2 | √ | × | × | |
| rs7138196 | RBM19 | √ | × | × | |
| rs7138219 | RBM19 | √ | × | × | |
| rs3782445 | RBM19 | √ | × | × | |
| rs4767167 | RBM19 | × | × | √ | |
| rs16943519 | RBM19 | × | × | √ | |
| rs79731175 | RBM44 | √ | × | × | |
| rs78726074 | RBM44 | √ | × | × | |
| rs9854636 | RBMS3 | √ | × | × | |
| rs9837143 | RBMS3 | √ | × | × | |
| rs4680856 | RBMS3 | √ | × | × | |
| rs6795528 | RBMS3 | √ | × | × | |
| rs7647627 | RBMS3 | √ | × | × | |
| rs4902508 | RDH11 | √ | × | × | |
| rs56244142 | RDH11 | √ | × | × | |
| rs2057119 | RDH11 | √ | × | × | |
| rs3808954 | RGS10 | × | √ | × | |
| rs3808955 | RGS10 | × | √ | × | |
| rs10787977 | RGS10 | × | √ | × | |
| rs76021313 | RGS10 | × | √ | × | |
| rs144205189 | RGS10 | × | √ | × | |
| rs79321427 | RMDN2 | √ | × | × | |
| rs6710923 | RMDN2 | √ | × | × | |
| rs1470480 | RMDN2 | √ | × | × | |
| rs7963126 | RMST | √ | × | × | |
| rs199504339 | RNF146 | √ | × | × | |
| rs76406607 | RNF146 | √ | × | × | |
| rs12175928 | RNF146 | √ | × | × | |
| rs9388558 | RNF146 | √ | × | × | |
| rs9401946 | RNF146 | √ | × | × | |
| rs9372862 | RNF146 | √ | × | × | |
| rs9388559 | RNF146 | √ | × | × | |
| rs72961049 | RNF146 | √ | × | × | |
| rs140197363 | RNF146 | √ | × | × | |
| rs79150701 | RNF146 | √ | × | × | |
| rs9321072 | RNF146 | √ | × | × | |
| rs41285264 | RNF146 | √ | × | × | |
| rs78238091 | RNF17 | √ | × | × | |
| rs76239507 | RNF17 | √ | × | × | |
| rs77184361 | RNF17 | √ | × | × | |
| rs79524222 | RNF17 | √ | × | × | |
| rs117676104 | RNF17 | √ | × | × | |
| rs12865323 | RNF17 | √ | × | × | |
| rs77562650 | RNF17 | √ | × | × | |
| rs1596860 | RNF17 | √ | × | × | |
| rs117552532 | RPL15 | √ | × | × | |
| rs2070843 | RPS14 | √ | × | × | |
| rs79096446 | RPS14 | √ | × | × | |
| rs11076181 | RSPRY1 | √ | × | × | |
| rs77353290 | RSPRY1 | √ | × | × | |
| rs7197204 | RSPRY1 | √ | × | × | |
| rs56872751 | RSPRY1 | √ | × | × | |
| rs7190847 | RSPRY1 | √ | × | × | |
| rs74246277 | RSPRY1 | √ | × | × | |
| rs78110752 | RSPRY1 | √ | × | × | |
| rs76203784 | RSPRY1 | √ | × | × | |
| rs75631671 | RSPRY1 | √ | × | × | |
| rs11859493 | RSPRY1 | √ | × | × | |
| rs715040 | RSPRY1 | √ | × | × | |
| rs4784783 | RSPRY1 | √ | × | × | |
| rs35902944 | RTEL1 | × | √ | × | |
| rs34538116 | RTEL1 | × | √ | × | |
| rs2241542 | SEC22C | × | √ | × | |
| rs342535 | SEC22C | × | √ | × | |
| rs188741916 | SEC22C | × | √ | × | |
| rs533938882 | SEC22C | × | √ | × | |
| rs342536 | SEC22C | × | √ | × | |
| rs138539315 | SEC22C | × | √ | × | |
| rs663258 | SEC22C | × | √ | × | |
| rs3774398 | SEC22C | × | √ | × | |
| rs2068278 | SEC22C | × | √ | × | |
| rs192865912 | SEC22C | × | √ | × | |
| rs6441892 | SEC22C | × | √ | × | |
| rs2271181 | SEC22C | × | √ | × | |
| rs2271182 | SEC22C | × | √ | × | |
| rs2271183 | SEC22C | × | √ | × | |
| rs376801834 | SEMA4D | √ | × | × | |
| rs58221985 | SEMA4D | √ | × | × | |
| rs79091937 | 45537 | √ | × | × | |
| rs117959705 | 45537 | √ | × | × | |
| rs6727584 | SH3RF3 | √ | × | × | |
| rs77920516 | SIK3 | √ | × | × | |
| rs10892048 | SIK3 | √ | × | × | |
| rs12574810 | SIK3 | √ | × | × | |
| rs7119185 | SIK3 | √ | × | × | |
| rs549821978 | SIK3 | √ | × | × | |
| rs184357842 | SLC15A5 | √ | × | × | |
| rs2248436 | SLC17A6 | √ | × | × | |
| rs375073736 | SLC25A25 | × | √ | × | |
| rs10739706 | SLC25A25 | × | √ | × | |
| rs138861516 | SLC25A25 | √ | × | × | |
| rs551017139 | SLC39A11 | √ | × | × | |
| rs151245839 | SLC39A11 | √ | × | × | |
| rs12051632 | SLC39A11 | √ | × | × | |
| rs16934253 | SLC8B1 | √ | × | × | |
| rs7953252 | SLC8B1 | √ | × | × | |
| rs7966103 | SLC8B1 | √ | × | × | |
| rs150626285 | SLIT3 | × | √ | × | |
| rs117562612 | SLIT3 | × | √ | × | |
| rs55942286 | SLIT3 | × | √ | × | |
| rs76048769 | SLIT3 | × | √ | × | |
| rs35140226 | SLIT3 | × | √ | × | |
| rs1875972 | SLIT3 | × | √ | × | |
| rs13173661 | SLIT3 | × | √ | × | |
| rs60131218 | SLIT3 | × | √ | × | |
| rs58107910 | SLIT3 | × | √ | × | |
| rs17070562 | SLIT3 | × | √ | × | |
| rs2938769 | SLIT3 | × | √ | × | |
| rs17070563 | SLIT3 | × | √ | × | |
| rs55762992 | SLIT3 | × | √ | × | |
| rs17635676 | SLIT3 | × | √ | × | |
| rs17666252 | SLIT3 | × | √ | × | |
| rs76443289 | SLIT3 | × | √ | × | |
| rs2974437 | SLIT3 | × | √ | × | |
| rs59937048 | SLIT3 | × | √ | × | |
| rs73802404 | SLIT3 | × | √ | × | |
| rs77934415 | SLIT3 | × | √ | × | |
| rs1504955 | SLIT3 | × | √ | × | |
| rs4867642 | SLIT3 | × | √ | × | |
| rs17733776 | SLIT3 | × | √ | × | |
| rs143090984 | SNED1 | √ | × | × | |
| rs74820922 | SNED1 | √ | × | × | |
| rs12994122 | SNED1 | √ | × | × | |
| rs187675909 | SNED1 | √ | × | × | |
| rs4652958 | SNIP1 | √ | × | × | |
| rs2273013 | SNIP1 | √ | × | × | |
| rs10872319 | SOGA3 | √ | × | × | |
| rs12422241 | SOX5 | √ | × | × | |
| rs59556093 | SOX5 | √ | × | × | |
| rs61517249 | SOX5 | √ | × | × | |
| rs10842250 | SOX5 | √ | × | × | |
| rs10842252 | SOX5 | √ | × | × | |
| rs11047179 | SOX5 | √ | × | × | |
| rs11047182 | SOX5 | √ | × | × | |
| rs76847583 | SOX5 | √ | × | × | |
| rs12229376 | SOX5 | √ | × | × | |
| rs11047185 | SOX5 | √ | × | × | |
| rs11047191 | SOX5 | √ | × | × | |
| rs11047192 | SOX5 | √ | × | × | |
| rs28740811 | SOX5 | √ | × | × | |
| rs7296101 | SOX5 | √ | × | × | Vertex-wise sulcal depth |
| rs10842272 | SOX5 | √ | × | × | Cortical surface area |
| rs1021670 | SOX5 | √ | × | × | |
| rs1021669 | SOX5 | √ | × | × | |
| rs10505932 | SOX5 | √ | × | × | |
| rs10505933 | SOX5 | √ | × | × | |
| rs11047251 | SOX5 | √ | × | × | |
| rs11047252 | SOX5 | √ | × | × | |
| rs12231866 | SOX5 | √ | × | × | |
| rs1995183 | SOX5 | √ | × | × | |
| rs7138457 | SOX5 | √ | × | × | |
| rs11047444 | SOX5 | × | √ | × | |
| rs55651930 | SOX5 | × | √ | × | |
| rs79239850 | SOX5 | × | √ | × | |
| rs76832813 | SOX5 | × | √ | × | |
| rs10842350 | SOX5 | × | √ | × | |
| rs78477219 | SOX5 | × | √ | × | |
| rs73285517 | SOX5 | × | √ | × | |
| rs76700244 | SOX5 | × | √ | × | |
| rs12824173 | SOX5 | × | √ | × | |
| rs11047448 | SOX5 | × | √ | × | |

| | | | | | |
|---|---|---|---|---|---|
| rs76104804 | GRK5 | √ | × | × | |
| rs78265082 | GRK5 | √ | × | × | |
| rs76761106 | GRK5 | √ | × | × | |
| rs4752274 | GRK5 | √ | × | × | |
| rs4752275 | GRK5 | √ | × | × | |
| rs12344154 | IARS | √ | × | × | |
| rs72750409 | IARS | √ | × | × | |
| rs11792542 | IARS | √ | × | × | |
| rs501814 | IARS | √ | × | × | |
| rs556155 | IARS | √ | × | × | |
| rs4743866 | IARS | √ | × | × | |
| rs4743867 | IARS | √ | × | × | |
| rs11793505 | IARS | √ | × | × | |
| rs1175930 | IARS | √ | × | × | |
| rs54897707 | IARS | √ | × | × | |
| rs76631712 | IARS | √ | × | × | |
| rs472163 | IARS | √ | × | × | |
| rs4743868 | IARS | √ | × | × | |
| rs9409659 | IARS | √ | × | × | |
| rs9409466 | IARS | √ | × | × | |
| rs3736835 | IARS | √ | × | × | |
| rs14355774 | IARS | √ | × | × | |
| rs15066683 | IARS | √ | × | × | |
| rs79820690 | IFFO2 | √ | × | × | |
| rs77321619 | IKZF1 | √ | × | × | |
| rs12619455 | ILKAP | √ | × | × | |
| rs7568705 | ILKAP | √ | × | × | |
| rs11677736 | ILKAP | √ | × | × | |
| rs35468889 | ILKAP | √ | × | × | |
| rs11688732 | ILKAP | √ | × | × | |
| rs2305171 | ILKAP | √ | × | × | |
| rs10929269 | ILKAP | √ | × | × | |
| rs76579247 | ILKAP | × | √ | × | |
| rs12997158 | ILKAP | × | √ | × | |
| rs74596805 | ILKAP | × | √ | × | |
| rs62194900 | ILKAP | × | √ | × | |
| rs12617118 | ILKAP | × | √ | × | |
| rs62194901 | ILKAP | × | √ | × | |
| rs3795904 | ILKAP | × | √ | × | |
| rs3795903 | ILKAP | × | √ | × | |
| rs15051348 | ILKAP | × | √ | × | |
| rs35393292 | ILKAP | × | √ | × | |
| rs7558210 | ILKAP | × | √ | × | |
| rs7558403 | ILKAP | × | √ | × | |
| rs77873713 | ILKAP | × | √ | × | |
| rs7575100 | ILKAP | × | √ | × | |
| rs72991027 | ILKAP | × | √ | × | |
| rs6707157 | ILKAP | × | √ | × | |
| rs78903918 | ILKAP | × | √ | × | |
| rs80015710 | IMMT | √ | × | × | |
| rs4251569 | IRAK4 | × | √ | × | |
| rs37088102 | IRAK4 | × | √ | × | |
| rs14212993 | IRAK4 | × | √ | × | |
| rs56312115 | IRAK4 | × | √ | × | |
| rs4251522 | IRAK4 | × | √ | × | |
| rs57527157 | IRAK4 | × | √ | × | |
| rs3805198 | IRAK4 | × | √ | × | |
| rs75608281 | IRAK4 | × | √ | × | |
| rs4251527 | IRAK4 | × | √ | × | |
| rs1141168 | IRAK4 | × | √ | × | |
| rs3729552 | ITPKB | √ | × | × | |
| rs1144833 | ITPKB | √ | × | × | |
| rs77297817 | JAG2 | × | √ | × | |
| rs58759755 | JAG2 | × | √ | × | |
| rs78154277 | JAG2 | × | √ | × | |
| rs18533851 | JAG2 | × | √ | × | |
| rs2242635 | JAG2 | × | √ | × | Cortical sur |
| rs2242634 | JAG2 | × | √ | × | Cortical thic |
| rs72707615 | JAG2 | × | √ | × | |
| rs1057744 | JAG2 | × | √ | × | |
| rs2293806 | JAG2 | × | √ | × | |
| rs2238287 | JAG2 | × | √ | × | |
| rs11720302 | JAG2 | × | √ | × | |
| rs78231857 | JAG2 | × | √ | × | |
| rs28774386 | JAG2 | × | √ | × | |
| rs80153838 | JAG2 | × | √ | × | |
| rs57964198 | JAG2 | × | √ | × | |
| rs2072675 | JAG2 | √ | × | × | |
| rs2072674 | JAG2 | √ | × | × | Vertex-wise |
| rs11160836 | JAG2 | √ | × | × | |
| rs2239285 | JAG2 | √ | × | × | Vertex-wise |
| rs1882845 | JAG2 | √ | × | × | |
| rs4983589 | JAG2 | √ | × | × | Cortical sur |
| rs2965797 | KATNB1 | √ | × | × | |
| rs76609534 | KCNK2 | √ | × | × | |
| rs7074936 | KIAA1217 | √ | × | × | Right hippoc |
| rs7911355 | KIAA1217 | √ | × | × | |
| rs56302643 | KIFC3 | √ | × | × | |
| rs58082302 | KIFC3 | √ | × | × | |
| rs1056583 | KLF3-AS1 | √ | × | × | |
| rs7972909 | KRT74 | √ | × | × | |
| rs608690 | KRT74 | √ | × | × | |
| rs1499512 | LDLRAD3 | √ | × | × | |
| rs4756265 | LDLRAD3 | √ | × | × | Lymphocyte |
| rs10768174 | LDLRAD3 | √ | × | × | Lymphocyte |
| rs10842371 | LINC00477 | √ | × | × | |
| rs4963770 | LINC00477 | √ | × | × | |
| rs10842373 | LINC00477 | √ | × | × | |
| rs67462211 | LINC00571 | √ | × | × | |
| rs11840873 | LINC00571 | √ | × | × | |
| rs9576449 | LINC00571 | √ | × | × | |
| rs9576451 | LINC00571 | √ | × | × | |
| rs58037492 | LINC00841 | √ | × | × | |
| rs8031036 | LINC00928 | √ | × | × | |
| rs8029479 | LINC00928 | √ | × | × | |
| rs60084150 | LINC01107 | × | √ | × | |
| rs10188853 | LINC01107 | × | √ | × | |
| rs37058162 | LINC01107 | × | √ | × | |
| rs58568091 | LINC01107 | × | √ | × | Lung functic |
| rs6717371 | LINC01107 | × | √ | × | |
| rs6759921 | LINC01107 | × | √ | × | |
| rs6760257 | LINC01107 | × | √ | × | |
| rs14027906 | LINC01107 | × | √ | × | |
| rs1039887 | LINC01107 | × | √ | × | |
| rs56121087 | LINC01107 | × | √ | × | |
| rs14003182 | LINC01107 | × | √ | × | |
| rs11787803 | LINC01107 | × | √ | × | |
| rs53476129 | LINC01107 | × | √ | × | |
| rs1470592 | LINC01107 | × | √ | × | |
| rs1510513 | LINC01107 | × | √ | × | |
| rs1510514 | LINC01107 | × | √ | × | |
| rs9679172 | LINC01107 | × | √ | × | |
| rs56373856 | LINC01107 | × | √ | × | |
| rs4663901 | LINC01107 | √ | × | × | |
| rs1527550 | LINC01234 | √ | × | × | |
| rs12466299 | LINC01237 | √ | × | × | |
| rs28578916 | LINC01237 | √ | × | × | |
| rs2075547 | LOC10065 | √ | × | × | |
| rs17523113 | LOC10065 | √ | × | × | |
| rs526602 | LOC10065 | √ | × | × | |
| rs11712419 | LOC10192 | √ | × | × | |
| rs7707724 | LOC10192 | √ | × | × | |
| rs7446705 | LOC10192 | √ | × | × | |
| rs75337620 | LOC10192 | √ | × | × | |
| rs78903852 | LOC10192 | √ | × | × | |
| rs79397614 | LOC10192 | √ | × | × | |
| rs78230061 | LOC10192 | √ | × | × | |
| rs6887550 | LOC10192 | √ | × | × | |
| rs6595129 | LOC10192 | √ | × | × | Height |
| rs7448537 | LOC10192 | √ | × | × | |
| rs6868999 | LOC10192 | √ | × | × | |
| rs6595130 | LOC10192 | × | √ | × | |
| rs6595131 | LOC10192 | × | √ | × | |
| rs6595132 | LOC10192 | × | √ | × | |
| rs11737081 | LOC10192 | × | √ | × | |
| rs4329039 | LOC10192 | √ | × | × | |
| rs78110595 | LOC10192 | √ | × | × | |
| rs18476806 | LOC10192 | √ | × | × | |
| rs4440398 | LOC10192 | √ | × | × | |
| rs4544848 | LOC10192 | √ | × | × | |
| rs34273870 | LOC10192 | √ | × | × | |
| rs35346599 | LOC10192 | √ | × | × | |
| rs4333349 | LOC10192 | √ | × | × | |
| rs4426942 | LOC10192 | √ | × | × | |
| rs74823943 | LOC10192 | √ | × | × | |
| rs76914990 | LOC10192 | √ | × | × | |
| rs78381164 | LOC10192 | √ | × | × | |
| rs75212422 | LOC10192 | √ | × | × | |
| rs77130837 | LOC10192 | √ | × | × | |
| rs1754076 | LOC10192 | √ | × | × | |
| rs9482391 | LOC10192 | √ | × | × | |
| rs13219865 | LOC10192 | √ | × | × | |
| rs13220681 | LOC10192 | √ | × | × | |
| rs13194849 | LOC10192 | √ | × | × | |
| rs6930651 | LOC10192 | √ | × | × | |
| rs6915179 | LOC10192 | √ | × | × | |
| rs7771575 | LOC10192 | √ | × | × | |
| rs62420461 | LOC10192 | √ | × | × | |
| rs15129823 | LOC10192 | × | √ | × | |
| rs12333042 | LOC10192 | √ | × | × | |
| rs9490776 | LOC10192 | √ | × | × | |
| rs6933435 | LOC10192 | √ | × | × | |
| rs6933479 | LOC10192 | √ | × | × | |
| rs75407261 | LOC10192 | √ | × | × | |
| rs12714154 | LOC10192 | √ | × | × | |
| rs15117840 | LOC10192 | √ | × | × | |
| rs11729946 | LOC10192 | √ | × | × | |
| rs11782674 | LOC10192 | √ | × | × | |
| rs34680639 | LOC10272 | × | √ | × | |
| rs15040319 | LOC10272 | × | √ | × | |
| rs53570551 | LOC10272 | × | √ | × | |
| rs12908605 | LOC10272 | × | √ | × | |
| rs1002175 | LOC10272 | × | √ | × | |
| rs14580842 | LOC10272 | × | √ | × | |
| rs28449570 | LOC10272 | × | √ | × | |
| rs18107299 | LOC10272 | × | √ | × | |
| rs18555511 | LOC10272 | × | √ | × | |
| rs28678344 | LOC10272 | × | √ | × | |
| rs75991329 | LOC10272 | × | √ | × | |
| rs56855842 | LOC10272 | × | √ | × | |
| rs12885315 | LOC10272 | × | √ | × | |
| rs12885313 | LOC10272 | × | √ | × | |
| rs12886669 | LOC10272 | × | √ | × | |
| rs12886677 | LOC10272 | × | √ | × | |
| rs79319348 | LOC10272 | × | √ | × | |
| rs11160834 | LOC10272 | × | √ | × | |
| rs36171475 | LOC10272 | × | √ | × | |
| rs14340395 | LOC10272 | × | √ | × | |
| rs7155121 | LOC10272 | × | √ | × | |
| rs11850872 | LOC10272 | × | √ | × | |
| rs9743945 | LOC10272 | × | √ | × | |
| rs60849362 | LOC10272 | × | √ | × | |
| rs75206602 | LOC10272 | × | √ | × | |
| rs7160811 | LOC10272 | × | √ | × | |
| rs74896861 | LOC10272 | × | √ | × | |
| rs58770530 | LOC10272 | × | √ | × | |
| rs74001724 | LOC20077 | × | √ | × | |
| rs74001725 | LOC20077 | × | √ | × | |
| rs53995388 | LOC20077 | × | √ | × | |
| rs76679054 | LOC20077 | × | √ | × | |
| rs74001726 | LOC20077 | × | √ | × | |
| rs16962841 | LOC28439 | √ | × | × | |
| rs1529721 | LOC28439 | √ | × | × | |
| rs57249767 | LOC28439 | √ | × | × | |
| rs78635090 | LOC28439 | √ | × | × | |
| rs4805409 | LOC28439 | √ | × | × | |
| rs77713166 | LOC28439 | √ | × | × | |
| rs2866068 | LOC28439 | √ | × | × | |
| rs4805410 | LOC28439 | √ | × | × | |
| rs56933718 | LOC28439 | √ | × | × | |
| rs8103193 | LOC28439 | √ | × | × | |
| rs80121463 | LOC28439 | √ | × | × | |
| rs11505160 | LOC28439 | √ | × | × | |
| rs8101026 | LOC28439 | √ | × | × | |
| rs1865064 | LOC28439 | √ | × | × | |
| rs12975082 | LOC28439 | √ | × | × | |
| rs4973649 | LOC72832 | √ | × | × | |
| rs12617606 | LOC72832 | √ | × | × | |
| rs55814222 | LOC72832 | × | × | √ | |
| rs2491101 | LRSAM1 | √ | × | × | |
| rs2244218 | LRSAM1 | √ | × | × | |
| rs2243509 | LRSAM1 | √ | × | × | |
| rs2253414 | LRSAM1 | √ | × | × | |
| rs2253411 | LRSAM1 | √ | × | × | |
| rs12362660 | LUZP2 | √ | × | × | |
| rs12805462 | LUZP2 | √ | × | × | |
| rs11028026 | LUZP2 | √ | × | × | |
| rs1553740 | LUZP2 | √ | × | × | |
| rs1553741 | LUZP2 | √ | × | × | |

| | | | | | |
|---|---|---|---|---|---|
| rs35982877 | MIR548AC | √ | × | × | |
| rs78111879 | MIR548AC | √ | × | × | |
| rs73045614 | MIR548AC | √ | × | × | |
| rs11015964 | MKX | √ | × | × | |
| rs11015965 | MKX | √ | × | × | |
| rs11015966 | MKX | √ | × | × | |
| rs18162565 | MKX | √ | × | × | |
| rs68142312 | MLPH | × | √ | × | |
| rs73999113 | MLPH | × | √ | × | |
| rs13398593 | MLPH | × | √ | × | |
| rs13395864 | MLPH | × | √ | × | |
| rs13407418 | MLPH | × | √ | × | |
| rs72620833 | MLPH | × | √ | × | |
| rs72620834 | MLPH | × | √ | × | |
| rs3802521 | MPP7 | √ | × | × | |
| rs3802523 | MPP7 | √ | × | × | |
| rs11006821 | MPP7 | × | × | √ | |
| rs11992232 | MSR1 | √ | × | × | |
| rs9650393 | MSR1 | √ | × | × | |
| rs79538977 | MSR1 | √ | × | × | |
| rs75674983 | MSR1 | √ | × | × | |
| rs76917531 | MSR1 | √ | × | × | |
| rs77061383 | MSR1 | √ | × | × | |
| rs80023501 | MSR1 | √ | × | × | |
| rs12114633 | MSR1 | √ | × | × | |
| rs366096 | MSR1 | √ | × | × | |
| rs74303554 | MSR1 | √ | × | × | |
| rs388391 | MSR1 | √ | × | × | |
| rs13306541 | MSR1 | √ | × | × | |
| rs7144389 | MTA1 | × | × | √ | |
| rs8021311 | MTA1 | × | × | √ | Bilirubin (Z,Z |
| rs2240538 | MTERFD2 | √ | × | × | |
| rs12264946 | MXI1 | √ | × | × | |
| rs7072547 | MXI1 | √ | × | × | |
| rs4918487 | MXI1 | √ | × | × | |
| rs35741140 | MXI1 | √ | × | × | |
| rs11812839 | MXI1 | √ | × | × | |
| rs58677130 | MXI1 | √ | × | × | |
| rs3793888 | MXI1 | √ | × | × | |
| rs71479294 | MXI1 | √ | × | × | |
| rs2634705 | MYPN | √ | × | × | |
| rs2673796 | MYPN | √ | × | × | |
| rs2176105 | MYPN | √ | × | × | |
| rs2176104 | MYPN | √ | × | × | |
| rs2673795 | MYPN | √ | × | × | |
| rs2255027 | MYPN | √ | × | × | |
| rs2634709 | MYPN | √ | × | × | |
| rs2673794 | MYPN | √ | × | × | |
| rs2101484 | MYPN | √ | × | × | |
| rs9590589 | NAA16 | √ | × | × | |
| rs75597686 | NAA16 | √ | × | × | |
| rs9590592 | NAA16 | √ | × | × | |
| rs3769499 | NCK2 | √ | × | × | |
| rs77322432 | NDST1 | √ | × | × | |
| rs37254904 | NDST1 | √ | × | × | |
| rs2545342 | NDST1 | √ | × | × | |
| rs1290147 | NDST1 | √ | × | × | |
| rs77013433 | NDST1 | √ | × | × | |
| rs18093771 | NDST1 | √ | × | × | |
| rs7624628 | NKTR | √ | × | × | |
| rs6441928 | NKTR | √ | × | × | |
| rs3846058 | NKTR | √ | × | × | |
| rs3095258 | NKTR | √ | × | × | |
| rs339735 | NKTR | √ | × | × | |
| rs168138 | NKTR | √ | × | × | |
| rs398743 | NKTR | √ | × | × | |
| rs19947600 | NLRC5 | √ | × | × | |
| rs13965881 | NR1D2 | √ | × | × | |
| rs15122031 | NR1D2 | √ | × | × | |
| rs75713041 | NRF1 | √ | × | × | |
| rs75172830 | NRF1 | √ | × | × | |
| rs80256694 | NRF1 | √ | × | × | |
| rs79168780 | NRF1 | √ | × | × | |
| rs78270497 | NRF1 | √ | × | × | |
| rs79847836 | NRF1 | √ | × | × | |
| rs76025913 | NRF1 | √ | × | × | |
| rs6969098 | NRF1 | √ | × | × | |
| rs14467436 | NRF1 | √ | × | × | |
| rs7783235 | NRF1 | √ | × | × | |
| rs7782840 | NRF1 | √ | × | × | |
| rs17160984 | NRF1 | √ | × | × | |
| rs79987037 | NRF1 | √ | × | × | |
| rs3735006 | NRF1 | √ | × | × | |
| rs74598514 | NRF1 | √ | × | × | |
| rs2402964 | NRF1 | √ | × | × | |
| rs10234904 | NRF1 | √ | × | × | |
| rs2456502 | ONECUT1 | √ | × | × | |
| rs4971246 | OPTC | √ | × | × | |
| rs9347686 | PACRG | √ | × | × | |
| rs4709648 | PACRG | √ | × | × | |
| rs9365487 | PACRG | √ | × | × | |
| rs9458643 | PACRG | √ | × | × | |
| rs12215676 | PACRG | √ | × | × | |
| rs10806765 | PACRG | √ | × | × | |
| rs9456811 | PACRG | √ | × | × | |
| rs9365490 | PACRG | √ | × | × | |
| rs6936373 | PACRG | √ | × | × | |
| rs6936932 | PACRG | √ | × | × | |
| rs34936012 | PACRG | √ | × | × | |
| rs1931222 | PACRG | √ | × | × | |
| rs1355929 | PACRG | √ | × | × | |
| rs9365507 | PACRG | √ | × | × | |
| rs2151653 | PACRG | √ | × | × | |
| rs1588787 | PACRG | √ | × | × | |
| rs62427531 | PACRG | √ | × | × | |
| rs6931846 | PACRG | √ | × | × | |
| rs9365514 | PACRG | √ | × | × | |
| rs9356081 | PACRG | √ | × | × | |
| rs7749139 | PACRG | √ | × | × | |
| rs9346941 | PACRG | √ | × | × | |
| rs7769875 | PACRG | √ | × | × | |
| rs73607817 | PACRG | √ | × | × | |
| rs12191527 | PACRG | √ | × | × | |
| rs4296874 | PACRG | √ | × | × | |
| rs10945870 | PACRG | √ | × | × | |
| rs6909464 | PACRG | √ | × | × | |
| rs7940310 | PAFAH1B2 | √ | × | × | Blood protei |
| rs2127905 | PAFAH1B2 | √ | × | × | |
| rs7755248 | PARK2 | √ | × | × | |
| rs9356038 | PARK2 | √ | × | × | |
| rs9347660 | PARK2 | √ | × | × | |
| rs9356039 | PARK2 | √ | × | × | |
| rs9365463 | PARK2 | √ | × | × | |
| rs4636000 | PARK2 | √ | × | × | |
| rs9458589 | PARK2 | √ | × | × | |
| rs9347666 | PARK2 | √ | × | × | |
| rs9295194 | PARK2 | √ | × | × | |
| rs9458595 | PARK2 | √ | × | × | |
| rs9355399 | PARK2 | √ | × | × | |
| rs9347667 | PARK2 | √ | × | × | |
| rs6914377 | PARK2 | √ | × | × | |
| rs10945847 | PARK2 | √ | × | × | |
| rs9458609 | PARK2 | √ | × | × | |
| rs9456797 | PARK2 | √ | × | × | |
| rs9356052 | PARK2 | √ | × | × | |
| rs11810460 | PARK2 | √ | × | × | |
| rs7451729 | PARK2 | √ | × | × | |
| rs1954805 | PARK2 | √ | × | × | |
| rs2023014 | PARK2 | √ | × | × | |
| rs9456802 | PARK2 | √ | × | × | |
| rs9458619 | PARK2 | √ | × | × | |
| rs6941209 | PARK2 | √ | × | × | |
| rs12826679 | PCED1B | √ | × | × | |
| rs10785660 | PCED1B | √ | × | × | |
| rs1880599 | PCED1B | √ | × | × | |
| rs11608610 | PCED1B | √ | × | × | |
| rs12581803 | PCED1B | √ | × | × | |
| rs7960991 | PCED1B | √ | × | × | |
| rs12822740 | PCED1B | √ | × | × | |
| rs11183778 | PCED1B | √ | × | × | |
| rs10881068 | PCED1B | √ | × | × | |
| rs10748466 | PCED1B | √ | × | × | |
| rs12367393 | PCED1B | √ | × | × | Vaginal mic |
| rs11183802 | PCED1B | √ | × | × | |
| rs11744381 | PCSK6 | √ | × | × | |
| rs6875503 | PDZD2 | √ | × | × | |
| rs20010661 | PEX12 | √ | × | × | |
| rs34750359 | PLBD2 | √ | × | × | |
| rs964576 | PLCL2 | √ | × | × | |
| rs34322012 | PLCL2 | √ | × | × | |
| rs1806555 | PLCL2 | √ | × | × | |
| rs4685411 | PLCL2 | √ | × | × | |
| rs17103522 | PLEKHA1 | √ | × | × | |
| rs78753090 | PLEKHH1 | √ | × | × | |
| rs9972155 | PLEKHH1 | √ | × | × | |
| rs10173155 | POLR1A | √ | × | × | |
| rs10208184 | POLR1A | √ | × | × | |
| rs78646493 | POLR1A | √ | × | × | |
| rs311564 | POLR1A | √ | × | × | |
| rs311576 | POLR1A | √ | × | × | |
| rs311578 | POLR1A | √ | × | × | |
| rs62147717 | POLR1A | √ | × | × | |
| rs72847885 | POLR1A | √ | × | × | |
| rs14103104 | PPARGC1A | √ | × | × | |
| rs18516297 | PPARGC1A | √ | × | × | |
| rs2794444 | PRELP | √ | × | × | |
| rs1077291 | PRELP | √ | × | × | |
| rs71277158 | PRKCI | √ | × | × | Prostate ca |
| rs4755450 | PRR5L | √ | × | × | Multiple scle |
| rs10836538 | PRR5L | √ | × | × | Asthma (chi |
| rs7844247 | PSD3 | √ | × | × | |
| rs79479361 | PSD3 | √ | × | × | |
| rs7814302 | PSD3 | √ | × | × | |
| rs77748229 | PSD3 | √ | × | × | |
| rs1025229 | PSD3 | √ | × | × | |
| rs11990985 | PSD3 | √ | × | × | |
| rs73204503 | PSD3 | √ | × | × | |
| rs73204504 | PSD3 | √ | × | × | |
| rs74829276 | PSD3 | √ | × | × | |
| rs76920322 | PSD3 | √ | × | × | |
| rs19106387 | PSD3 | √ | × | × | |
| rs14698696 | PSD3 | √ | × | × | |
| rs37043184 | PSD3 | √ | × | × | |
| rs11127016 | PTCD3 | √ | × | × | |
| rs17618119 | PTCD3 | √ | × | × | |
| rs3764950 | PTCD3 | √ | × | × | |
| rs17584697 | PTCD3 | √ | × | × | |
| rs3731818 | PTCD3 | √ | × | × | Pulse press |
| rs75158612 | RAB17 | × | √ | × | |
| rs52860621 | RAB17 | × | √ | × | |
| rs8184 | RAB17 | × | √ | × | |
| rs2280288 | RAB17 | × | √ | × | |
| rs72990107 | RAB17 | × | √ | × | |
| rs59945834 | RAB17 | × | √ | × | |
| rs2280289 | RAB17 | × | √ | × | |
| rs78478325 | RAB17 | × | √ | × | |
| rs10202062 | RAB17 | × | √ | × | |
| rs60484328 | RAB17 | × | √ | × | |
| rs13414095 | RAB17 | × | √ | × | |
| rs61312223 | RAB17 | × | √ | × | |
| rs59848818 | RAB17 | × | √ | × | |
| rs58094742 | RAB17 | × | √ | × | |
| rs74800521 | RAB17 | × | √ | × | |
| rs72990117 | RAB17 | × | √ | × | |
| rs75983679 | RAB17 | × | √ | × | |
| rs75901095 | RAB17 | × | √ | × | |
| rs78268371 | RAB17 | × | √ | × | |
| rs11759903 | RAB17 | × | √ | × | |
| rs74346400 | RAB17 | × | √ | × | |
| rs72990120 | RAB17 | × | √ | × | |
| rs59044255 | RAB17 | √ | × | × | |
| rs13404216 | RAB17 | √ | × | × | |
| rs72990123 | RAB17 | √ | × | × | Corneal res |
| rs6431547 | RAB17 | √ | × | × | |
| rs7597767 | RAB17 | √ | × | × | |
| rs13390880 | RAB17 | √ | × | × | |
| rs2292875 | RAB17 | √ | × | × | |
| rs57423896 | RAB17 | √ | × | × | |
| rs2292871 | RAB17 | √ | × | × | Corneal res |
| rs3947369 | RALGPS1 | √ | × | × | |
| rs20065820 | RALGPS1 | √ | × | × | |
| rs56013992 | RBM19 | √ | × | × | |
| rs75606772 | RBM19 | √ | × | × | |
| rs16943344 | RBM19 | √ | × | × | |
| rs79149125 | RBM19 | √ | × | × | |
| rs75868956 | RBM19 | √ | × | × | |
| rs10510630 | RBMS3 | √ | × | × | |
| rs2273160 | RDH11 | √ | × | × | |
| rs55790812 | RDH11 | √ | × | × | |

| | | | | |
|---|---|---|---|---|
| rs58003625 | NRF1 | √ | × | |
| rs7778085 | NRF1 | √ | × | |
| rs2402965 | NRF1 | √ | × | |
| rs79497918 | NRF1 | √ | × | |
| rs11773536 | NRF1 | √ | × | |
| rs7802940 | NRF1 | √ | × | |
| rs3800598 | NRF1 | √ | × | |
| rs2402978 | NRF1 | √ | × | |
| rs10258498 | NRF1 | √ | × | |
| rs7796553 | NRF1 | √ | × | |
| rs10224702 | NRF1 | √ | × | |
| rs2402964 | NRF1 | √ | × | |
| rs10234904 | NRF1 | √ | × | |
| rs10139450 | NUDT14 | √ | × | |
| rs75829623 | NUDT14 | × | √ | |
| rs4983590 | NUDT14 | × | √ | White blood cell count |
| rs8017877 | NUDT14 | × | √ | |
| rs28446858 | OPTC | √ | × | |
| rs2306152 | OPTC | √ | × | |
| rs9365507 | PACRG | √ | × | |
| rs7749139 | PACRG | √ | × | |
| rs9346941 | PACRG | √ | × | |
| rs7769875 | PACRG | √ | × | |
| rs9295211 | PACRG | × | √ | |
| rs9347667 | PARK2 | √ | × | |
| rs11807187 | PARK2 | √ | × | |
| rs9355401 | PARK2 | √ | × | |
| rs14569241 | PARK2 | √ | × | |
| rs9347679 | PARK2 | √ | × | |
| rs7566357 | PASK | √ | × | |
| rs2240549 | PASK | √ | × | |
| rs2074831 | PASK | √ | × | |
| rs4675829 | PASK | √ | × | |
| rs12623850 | PASK | √ | × | |
| rs2286326 | PASK | √ | × | |
| rs2286327 | PASK | √ | × | |
| rs2408716 | PCED1B | √ | × | |
| rs10881048 | PCED1B | √ | × | |
| rs56295736 | PCED1B | √ | × | |
| rs12826679 | PCED1B | √ | × | |
| rs11748849 | PCED1B | √ | × | |
| rs80324997 | PCSK6 | √ | × | |
| rs77671416 | PHC3 | √ | × | |
| rs73592669 | PLAGL1 | √ | × | |
| rs71467903 | PLBD2 | √ | × | |
| rs14376111 | PLBD2 | × | √ | |
| rs76543720 | PLBD2 | × | √ | |
| rs11200621 | PLEKHA1 | √ | × | |
| rs10173155 | POLR1A | √ | × | |
| rs10208184 | POLR1A | √ | × | |
| rs14220008 | PPARGC1A | √ | × | |
| rs12053217 | PPP1R7 | √ | × | |
| rs12052806 | PPP1R7 | √ | × | |
| rs2268902 | PPP1R7 | √ | × | |
| rs16843426 | PPP1R7 | √ | × | |
| rs1121819 | PPP1R9A | √ | × | |
| rs2695700 | PPP1R9A | √ | × | |
| rs12704780 | PPP1R9A | √ | × | |
| rs11762113 | PPP1R9A | √ | × | |
| rs12704782 | PPP1R9A | √ | × | |
| rs12704783 | PPP1R9A | √ | × | |
| rs74701306 | PRKCI | √ | × | |
| rs14361553 | PRKCI | √ | × | |
| rs71277158 | PRKCI | √ | × | Prostate cancer |
| rs74892280 | PRSS3 | √ | × | |
| rs93437174 | PRSS3 | √ | × | |
| rs10528394 | PRSS3 | √ | × | |
| rs20200259 | PRSS3 | √ | × | |
| rs1025229 | PSD3 | √ | × | |
| rs12679152 | PSD3 | √ | × | |
| rs11127016 | PTCD3 | √ | × | |
| rs17618119 | PTCD3 | √ | × | |
| rs3764950 | PTCD3 | √ | × | |
| rs17584697 | PTCD3 | √ | × | |
| rs78959772 | PTPRK | × | √ | |
| rs14658063 | PTPRK | × | √ | |
| rs1382449 | PUS7L | × | √ | |
| rs11182246 | PUS7L | × | √ | |
| rs9971895 | PUS7L | × | √ | |
| rs9971859 | PUS7L | × | √ | |
| rs14444038 | PUS7L | × | √ | |
| rs7311465 | PUS7L | × | √ | |
| rs74890729 | PUS7L | × | √ | |
| rs7954870 | PUS7L | × | √ | |
| rs12229685 | PUS7L | × | √ | |
| rs9783418 | PUS7L | × | √ | |
| rs74885123 | PUS7L | √ | × | |
| rs77387647 | PUS7L | √ | × | |
| rs6746529 | RAB17 | √ | × | |
| rs11620947 | RAD51B | √ | × | |
| rs700876 | RANBP2 | √ | × | |
| rs1649330 | RANBP2 | √ | × | |
| rs1649329 | RANBP2 | √ | × | |
| rs1649328 | RANBP2 | √ | × | |
| rs56132830 | RANBP2 | √ | × | |
| rs1614285 | RANBP2 | √ | × | |
| rs4766700 | RBM19 | √ | × | |
| rs2891477 | RBM19 | √ | × | |
| rs79155940 | RBM19 | √ | × | |
| rs77707178 | RBM19 | √ | × | |
| rs75712023 | RBM19 | √ | × | |
| rs75934754 | RBM19 | √ | × | |
| rs74237687 | RBM19 | √ | × | |
| rs75894675 | RBM19 | √ | × | |
| rs3782442 | RBM19 | √ | × | |
| rs3782443 | RBM19 | √ | × | |
| rs59456286 | RBM19 | √ | × | |
| rs61027446 | RBM19 | √ | × | |
| rs4625522 | RBM19 | √ | × | |
| rs75606772 | RBM19 | √ | × | |
| rs4767164 | RBM19 | √ | × | |
| rs16943394 | RBM19 | √ | × | |
| rs11066815 | RBM19 | √ | × | |
| rs10774723 | RBM19 | √ | × | |
| rs10774724 | RBM19 | √ | × | |
| rs7975774 | RBM19 | √ | × | |
| rs2384345 | RBM19 | √ | × | |
| rs10774726 | RBM19 | √ | × | |
| rs6765275 | RBMS3 | √ | × | |
| rs6549962 | RBMS3 | √ | × | Heel bone mineral density |
| rs1868492 | RBMS3 | √ | × | |
| rs3773125 | RBMS3 | √ | × | |
| rs12488708 | RBMS3 | √ | × | |
| rs55790812 | RDH11 | √ | × | |
| rs55711201 | RDH12 | √ | × | |
| rs3783197 | RGCC | √ | × | |
| rs75887931 | RGS10 | × | √ | |
| rs2072693 | RHCG | √ | × | Number of decayed, missing and filled tooth surfaces or use of dentures |
| rs2075833 | RHCG | √ | × | |
| rs1371386 | RHCG | √ | × | |
| rs1043845 | RITA1 | √ | × | |
| rs78924293 | RMDN2 | √ | × | |
| rs13023086 | RMDN2 | √ | × | |
| rs61533771 | RMDN2 | √ | × | |
| rs59527249 | RMDN2 | √ | × | |
| rs58359588 | RMDN2 | √ | × | |
| rs34865785 | RMDN2 | √ | × | |
| rs1446297 | RMDN2 | √ | × | |
| rs35615184 | RMDN2 | √ | × | |
| rs4142168 | RMDN2 | √ | × | |
| rs1596862 | RMDN2 | √ | × | |
| rs1596861 | RMDN2 | √ | × | |
| rs6736956 | RMDN2 | √ | × | |
| rs17021813 | RMDN2 | √ | × | |
| rs7606530 | RMDN2 | √ | × | |
| rs2373327 | RMDN2 | √ | × | |
| rs1992735 | RMDN2 | √ | × | |
| rs6906717 | RNF146 | √ | × | |
| rs9385419 | RNF146 | √ | × | |
| rs2180341 | RNF146 | √ | × | Breast cancer |
| rs9372861 | RNF146 | √ | × | |
| rs9401947 | RNF146 | √ | × | |
| rs4897207 | RNF146 | √ | × | |
| rs2326567 | RNF146 | √ | × | |
| rs6569479 | RNF146 | √ | × | |
| rs9321073 | RNF146 | √ | × | |
| rs9507410 | RNF17 | √ | × | |
| rs76628987 | RNF17 | √ | × | |
| rs1451567 | RNF17 | √ | × | |
| rs2121250 | RNF17 | √ | × | |
| rs9511448 | RNF17 | √ | × | |
| rs9507412 | RNF17 | √ | × | |
| rs9511450 | RNF17 | √ | × | |
| rs9511451 | RNF17 | √ | × | |
| rs7319350 | RNF17 | √ | × | |
| rs6769676 | RNF7 | √ | × | |
| rs18694087 | ROR2 | √ | × | |
| rs13339 | RPP25L | √ | × | Vaginal microbiome MetaCyc pathway (TYRFUMCAT-PWY\|L-tyrosine degradation I) |
| rs6011026 | RTEL1 | × | √ | |
| rs76280092 | RUFY2 | √ | × | |
| rs3812826 | SDS | × | √ | |
| rs10461011 | SEC22C | √ | × | |
| rs2940407 | SEC22C | √ | × | |
| rs637852 | SEC22C | √ | × | |
| rs13064311 | SEC22C | √ | × | |
| rs7856677 | SEMA4D | √ | × | |
| rs12238114 | SEMA4D | √ | × | |
| rs56346836 | SEMA4D | √ | × | |
| rs56339712 | SEMA4D | √ | × | Reticulocyte count |
| rs4877082 | SEMA4D | √ | × | |
| rs4877083 | SEMA4D | √ | × | |
| rs62547238 | SEMA4D | √ | × | Reticulocyte count |
| rs56177051 | SEMA4D | √ | × | |
| rs17529841 | SEMA4D | √ | × | |
| rs3924245 | SEMA4D | √ | × | |
| rs4454378 | SEMA4D | √ | × | |
| rs62547240 | SEMA4D | √ | × | |
| rs4615679 | SEMA4D | √ | × | |
| rs54895503 | SEMA4D | √ | × | |
| rs60209440 | 45537 | √ | × | |
| rs59536819 | 45537 | √ | × | |
| rs41266959 | 45537 | √ | × | |
| rs73002111 | 45537 | √ | × | |
| rs19276680 | SH3RF3 | √ | × | |
| rs645258 | SIK3 | √ | × | Triglyceride levels |
| rs2513094 | SIK3 | √ | × | |
| rs79394589 | SIK3 | √ | × | |
| rs75958227 | SIK3 | √ | × | |
| rs11709172 | SIK3 | √ | × | |
| rs14720322 | SKIL | √ | × | |
| rs11056825 | SLC15A5 | √ | × | |
| rs78968736 | SLC25A25 | √ | × | |
| rs7036499 | SLC25A25 | √ | × | Height |
| rs7036522 | SLC25A25 | √ | × | |
| rs12348948 | SLC25A25 | √ | × | |
| rs12335456 | SLC25A25 | √ | × | |
| rs62586933 | SLC25A25 | √ | × | |
| rs11788116 | SLC25A25 | √ | × | |
| rs7044673 | SLC25A25 | √ | × | |
| rs4837235 | SLC25A25 | √ | × | |
| rs10739707 | SLC25A25 | √ | × | |
| rs10739708 | SLC25A25 | √ | × | |
| rs7854499 | SLC25A25 | √ | × | |
| rs10819348 | SLC25A25 | √ | × | |
| rs10819349 | SLC25A25 | √ | × | |
| rs7967427 | SLC2A13 | √ | × | |
| rs7978427 | SLC2A13 | √ | × | |
| rs1908911 | SLC2A13 | √ | × | |
| rs7969011 | SLC2A13 | √ | × | |
| rs61050676 | SLC2A13 | √ | × | |
| rs57095253 | SLC2A13 | √ | × | |
| rs11772979 | SLC39A11 | √ | × | |
| rs15132243 | SLIT3 | × | √ | |
| rs55620854 | SLIT3 | × | √ | |
| rs1498868 | SOX5 | × | √ | |
| rs11047206 | SOX5 | × | √ | |
| rs77489704 | SOX5 | × | √ | |
| rs10505923 | SOX5 | × | √ | |
| rs10505924 | SOX5 | × | √ | |
| rs7136518 | SOX5 | × | √ | |
| rs41383749 | SOX5 | × | √ | |
| rs12311849 | SOX5 | × | √ | |
| rs11047207 | SOX5 | × | √ | |
| rs77313049 | SOX5 | × | √ | |
| rs18227129 | SOX5 | × | √ | |
| rs12306043 | SOX5 | × | √ | |
| rs2133345 | SOX5 | × | √ | |
| rs28569416 | SOX5 | × | √ | |
| rs16926958 | SOX5 | × | √ | |

| | | | | | |
|---|---|---|---|---|---|
| rs61910419 | SOX5 | × | √ | × | |
| rs138346723 | SOX5 | × | √ | × | |
| rs11047449 | SOX5 | × | √ | × | Spatial QRS-T angle |
| rs12582243 | SOX5 | √ | × | × | |
| rs1355004 | SOX5 | √ | × | × | |
| rs11047455 | SOX5 | √ | × | × | |
| rs9668967 | SOX5 | √ | × | × | |
| rs117617198 | SOX5 | × | √ | × | |
| rs3803688 | SPIRE2 | × | × | √ | |
| rs13146675 | SPOCK3 | √ | × | × | |
| rs7691894 | SPOCK3 | √ | × | × | |
| rs62352806 | SPOCK3 | √ | × | × | |
| rs62352808 | SPOCK3 | √ | × | × | |
| rs79862732 | SPOCK3 | √ | × | × | |
| rs11937180 | SPOCK3 | √ | × | × | |
| rs28469298 | SPOCK3 | √ | × | × | |
| rs2219234 | SPOCK3 | √ | × | × | |
| rs182037746 | SPOCK3 | √ | × | × | |
| rs3828018 | SPOCK3 | √ | × | × | |
| rs3790358 | SPOCK3 | √ | × | × | |
| rs28699632 | SPOCK3 | √ | × | × | |
| rs7697656 | SPOCK3 | √ | × | × | |
| rs7693595 | SPOCK3 | √ | × | × | |
| rs2198734 | SPOCK3 | √ | × | × | |
| rs17645162 | SPOCK3 | √ | × | × | |
| rs10517901 | SPOCK3 | √ | × | × | |
| rs17645192 | SPOCK3 | √ | × | × | |
| rs7658810 | SPOCK3 | √ | × | × | |
| rs2305593 | SPOCK3 | √ | × | × | |
| rs6827226 | SPOCK3 | √ | × | × | |
| rs2318486 | SPOCK3 | √ | × | × | |
| rs111398285 | SPRTN | × | √ | × | |
| rs78658157 | SPRTN | × | √ | × | |
| rs117192083 | SPRTN | × | √ | × | |
| rs117473196 | SPRTN | × | √ | × | |
| rs117298595 | SPRTN | × | √ | × | |
| rs11122273 | SPRTN | × | √ | × | |
| rs7414807 | SPRTN | × | √ | × | |
| rs2355873 | SPRTN | × | √ | × | |
| rs10819410 | SPTAN1 | √ | × | × | |
| rs4395990 | SPTAN1 | √ | × | × | |
| rs10988041 | SPTAN1 | √ | × | × | |
| rs2152833 | SPTAN1 | √ | × | × | |
| rs3763599 | SPTAN1 | √ | × | × | |
| rs10988042 | SPTAN1 | √ | × | × | |
| rs117972175 | SPTLC1 | √ | × | × | |
| rs183426166 | ST3GAL5 | √ | × | × | |
| rs116892869 | ST3GAL5 | √ | × | × | Mean platelet volume durin |
| rs79386899 | STK25 | √ | × | × | |
| rs4472827 | STOX1 | √ | × | × | |
| rs75138397 | STX11 | √ | × | × | |
| rs4142550 | STX11 | √ | × | × | |
| rs148027578 | STXBP1 | × | √ | × | |
| rs191525468 | STXBP1 | × | √ | × | |
| rs3824523 | STXBP1 | × | √ | × | |
| rs7041091 | STXBP1 | × | √ | × | |
| rs17849179 | STXBP1 | × | √ | × | |
| rs13297270 | STXBP1 | × | √ | × | |
| rs368137758 | STXBP1 | × | √ | × | |
| rs17849181 | STXBP1 | × | √ | × | |
| rs2241166 | STXBP1 | × | √ | × | |
| rs2241167 | STXBP1 | × | √ | × | Prostate cancer |
| rs2241168 | STXBP1 | × | √ | × | |
| rs138279872 | STXBP1 | × | √ | × | |
| rs537682074 | STXBP1 | × | √ | × | |
| rs4837174 | STXBP1 | × | √ | × | |
| rs76431389 | STXBP1 | × | √ | × | |
| rs7021441 | STXBP1 | × | √ | × | |
| rs76782343 | STXBP1 | × | √ | × | |
| rs117548400 | STXBP1 | × | √ | × | |
| rs146124614 | STXBP1 | × | √ | × | |
| rs12115218 | STXBP1 | × | √ | × | |
| rs116935017 | STXBP1 | × | √ | × | |
| rs2302662 | STXBP1 | × | √ | × | |
| rs1341291 | SUSD4 | √ | × | × | |
| rs2795063 | SUSD4 | √ | × | × | |
| rs17163773 | SUSD4 | √ | × | × | |
| rs4661253 | SUSD4 | √ | × | × | |
| rs17161996 | SUSD4 | √ | × | × | |
| rs17163791 | SUSD4 | √ | × | × | |
| rs17163794 | SUSD4 | √ | × | × | |
| rs79356211 | SUSD4 | √ | × | × | |
| rs74943508 | SUSD4 | √ | × | × | |
| rs17163798 | SUSD4 | √ | × | × | |
| rs17163802 | SUSD4 | √ | × | × | |
| rs4661255 | SUSD4 | √ | × | × | |
| rs17163812 | SUSD4 | √ | × | × | |
| rs17163814 | SUSD4 | √ | × | × | |
| rs78743027 | SUSD4 | √ | × | × | |
| rs78569743 | SUSD4 | √ | × | × | |
| rs80034327 | SUSD4 | √ | × | × | |
| rs17163819 | SUSD4 | √ | × | × | |
| rs17163821 | SUSD4 | √ | × | × | |
| rs10494038 | SUSD4 | √ | × | × | |
| rs17162000 | SUSD4 | √ | × | × | |
| rs78868457 | SUSD4 | √ | × | × | |
| rs117398609 | SUSD4 | √ | × | × | |
| rs4661256 | SUSD4 | √ | × | × | |
| rs2138971 | SUSD4 | √ | × | × | |
| rs4661257 | SUSD4 | √ | × | × | |
| rs17162003 | SUSD4 | √ | × | × | |
| rs4661258 | SUSD4 | √ | × | × | |
| rs17068265 | SYNPR | √ | × | × | |
| rs17312483 | SYNPR | × | √ | × | |
| rs3856943 | SYNPR | √ | × | × | |
| rs1901304 | SYNPR | √ | × | × | |
| rs11200440 | TACC2 | × | √ | × | |
| rs181038838 | TACC2 | × | √ | × | |
| rs542685914 | TACC2 | × | √ | × | |
| rs12241688 | TACC2 | × | √ | × | |
| rs76748932 | TACC2 | × | √ | × | |
| rs150380034 | TACC2 | × | √ | × | |
| rs183292406 | TACC2 | × | √ | × | |
| rs77167803 | TACC2 | × | √ | × | |
| rs1885516 | TACC2 | × | √ | × | |
| rs117761202 | TACC2 | × | √ | × | |
| rs192229988 | TACC2 | × | √ | × | |
| rs147075462 | TACC2 | × | √ | × | |
| rs78302066 | TACC2 | × | √ | × | |
| rs4752665 | TACC2 | × | √ | × | |
| rs72840870 | TACC2 | × | √ | × | |
| rs7920029 | TACC2 | × | √ | × | |
| rs77132950 | TACC2 | × | √ | × | |
| rs7920046 | TACC2 | × | √ | × | |
| rs59585837 | TACC2 | × | √ | × | |
| rs4752666 | TACC2 | × | √ | × | |
| rs929999944 | TACC2 | × | √ | × | |
| rs72840875 | TACC2 | × | √ | × | |
| rs4751878 | TACC2 | × | √ | × | |
| rs72631110 | TACC2 | × | √ | × | |
| rs1322323 | TACC2 | × | √ | × | |
| rs192753030 | TACC2 | × | √ | × | |
| rs12416127 | TACC2 | × | √ | × | |
| rs4752668 | TACC2 | × | √ | × | |
| rs12411444 | TACC2 | × | √ | × | |
| rs10887101 | TACC2 | × | √ | × | |
| rs548127694 | TACC2 | × | √ | × | |
| rs10732826 | TACC2 | × | √ | × | |
| rs74499951 | TACC2 | × | √ | × | |
| rs911778 | TACC2 | √ | × | × | |
| rs35061398 | TACC2 | √ | × | × | |
| rs963534 | TACC2 | √ | × | × | |
| rs369322006 | TBC1D1 | × | √ | × | |
| rs75935172 | TBC1D1 | × | √ | × | |
| rs79752824 | TBC1D1 | × | √ | × | |
| rs17425501 | TBC1D1 | × | √ | × | |
| rs4832978 | TBC1D1 | × | √ | × | |
| rs75973053 | TBC1D1 | × | √ | × | |
| rs140766190 | TBC1D1 | × | √ | × | |
| rs544895496 | TBC1D1 | × | √ | × | |
| rs6858606 | TBC1D1 | × | √ | × | |
| rs373281161 | TBC1D1 | × | √ | × | |
| rs11931051 | TBC1D1 | × | √ | × | |
| rs7665468 | TBC1D1 | × | √ | × | |
| rs1344602 | TBC1D1 | × | √ | × | |
| rs7670255 | TBC1D1 | × | √ | × | |
| rs78902120 | TBC1D1 | × | √ | × | |
| rs10084926 | TBC1D1 | × | √ | × | |
| rs16995821 | TBC1D1 | × | √ | × | |
| rs538578519 | TBC1D1 | × | √ | × | |
| rs188736882 | TBC1D1 | × | √ | × | |
| rs13137606 | TBC1D1 | × | √ | × | |
| rs80072673 | TBC1D1 | × | √ | × | |
| rs62298551 | TBC1D1 | × | √ | × | |
| rs57524433 | TBC1D1 | × | √ | × | |
| rs59348868 | TBC1D1 | × | √ | × | |
| rs13140144 | TBC1D1 | × | √ | × | |
| rs9999507 | TBC1D1 | × | √ | × | |
| rs13114113 | TBC1D1 | × | √ | × | |
| rs80193966 | TBC1D1 | × | √ | × | |
| rs557902412 | TBC1D1 | × | √ | × | |
| rs1006182 | TBC1D1 | × | √ | × | |
| rs371373344 | TBC1D1 | × | √ | × | |
| rs1006181 | TBC1D1 | × | √ | × | Height |
| rs62298552 | TBC1D1 | × | √ | × | |
| rs2381144 | TBC1D1 | × | √ | × | |
| rs16994139 | TBC1D1 | × | √ | × | |
| rs557756990 | TBC1D1 | × | √ | × | |
| rs4144804 | TBC1D1 | × | √ | × | |
| rs4144803 | TBC1D1 | × | √ | × | |
| rs7672883 | TBC1D1 | × | √ | × | |
| rs4144800 | TBC1D1 | × | √ | × | |
| rs368055006 | TBC1D1 | × | √ | × | |
| rs12643286 | TBC1D1 | × | √ | × | |
| rs4144801 | TBC1D1 | × | √ | × | |
| rs17579337 | TBC1D1 | × | √ | × | |
| rs6531601 | TBC1D1 | × | √ | × | |
| rs78786098 | TBC1D1 | × | √ | × | |
| rs368036510 | TBC1D1 | × | √ | × | |
| rs61731587 | TBC1D1 | × | √ | × | |
| rs10501 | TBC1D1 | × | √ | × | |
| rs16994148 | TBC1D1 | × | √ | × | |
| rs145195654 | TBC1D1 | × | √ | × | |
| rs57940528 | TBC1D1 | × | √ | × | |
| rs13142228 | TBC1D1 | × | √ | × | Height |
| rs112006400 | TBC1D1 | × | √ | × | |
| rs75833459 | TBC1D1 | × | √ | × | |
| rs79254279 | TBC1D1 | × | √ | × | |
| rs35377184 | TBC1D1 | × | √ | × | |
| rs61355869 | TBC1D1 | × | √ | × | |
| rs181267081 | TBC1D1 | × | √ | × | |
| rs933821 | TBC1D1 | √ | × | × | |
| rs35308860 | TBC1D1 | √ | × | × | |
| rs28478131 | TBC1D1 | √ | × | × | |
| rs6827416 | TBC1D1 | × | √ | × | |
| rs67733859 | TBC1D1 | √ | × | × | |
| rs4832981 | TBC1D1 | × | √ | × | |
| rs148145415 | TBC1D1 | × | √ | × | |
| rs12647617 | TBC1D1 | × | √ | × | |
| rs533045117 | TBC1D1 | × | √ | × | |
| rs141092310 | TBC1D1 | × | √ | × | |
| rs6845120 | TBC1D1 | × | √ | × | Blood pressure (pleiotropy |
| rs140454952 | TBC1D1 | × | √ | × | |
| rs58206779 | TBC1D1 | × | √ | × | |
| rs74870874 | TBC1D1 | × | √ | × | |
| rs191313066 | TBC1D1 | × | √ | × | |
| rs7674013 | TBC1D1 | × | √ | × | |
| rs7675606 | TBC1D1 | × | √ | × | |
| rs62299976 | TBC1D1 | × | √ | × | |
| rs7674408 | TBC1D1 | × | √ | × | |
| rs55767827 | TBC1D1 | √ | × | × | |
| rs59608961 | TBC1D1 | √ | × | × | |
| rs35021061 | TBC1D1 | √ | × | × | |
| rs12649018 | TBC1D1 | √ | × | × | |
| rs1364953 | TBC1D1 | √ | × | × | Metabolite levels |
| rs9401367 | TBC1D32 | √ | × | × | |
| rs2202065 | TBC1D32 | √ | × | × | |
| rs9490138 | TBC1D32 | √ | × | × | |
| rs9482123 | TBC1D32 | √ | × | × | |
| rs2389414 | TBC1D32 | √ | × | × | |
| rs9320799 | TBC1D32 | √ | × | × | |
| rs9320800 | TBC1D32 | √ | × | × | |
| rs9320801 | TBC1D32 | √ | × | × | |
| rs9320802 | TBC1D32 | √ | × | × | |
| rs4585597 | TBC1D32 | √ | × | × | |
| rs3923664 | TBC1D32 | √ | × | × | |
| rs76496637 | TBC1D32 | √ | × | × | |
| rs9387942 | TBC1D32 | √ | × | × | |
| rs74775566 | TBC1D32 | √ | × | × | |

| | | | | | |
|---|---|---|---|---|---|
| rs2036752 | LUZP2 | √ | × | × | |
| rs1553742 | LUZP2 | √ | × | × | |
| rs1083438 | LUZP2 | √ | × | × | |
| rs5008808 | LUZP2 | √ | × | × | |
| rs1509588 | LUZP2 | √ | × | × | |
| rs1509589 | LUZP2 | √ | × | × | |
| rs1509590 | LUZP2 | √ | × | × | |
| rs981895 | LUZP2 | √ | × | × | |
| rs1083438 | LUZP2 | √ | × | × | |
| rs1160044 | LUZP2 | √ | × | × | |
| rs1083439 | LUZP2 | √ | × | × | |
| rs1102802 | LUZP2 | √ | × | × | |
| rs1076721 | LUZP2 | √ | × | × | |
| rs1083439 | LUZP2 | √ | × | × | |
| rs1102803 | LUZP2 | √ | × | × | |
| rs1222548 | LUZP2 | √ | × | × | |
| rs7867936 | LUZP2 | √ | × | × | |
| rs4922691 | LUZP2 | √ | × | × | |
| rs451167 | MAGI2 | √ | × | × | |
| rs38118 | MAGI2 | √ | × | × | |
| rs420581 | MAGI2 | √ | × | × | |
| rs413001 | MAGI2 | √ | × | × | |
| rs2022150 | METAP2 | √ | × | × | |
| rs6747400 | METTL8 | √ | × | × | |
| rs1261694 | METTL8 | √ | × | × | |
| rs1558963 | METTL8 | √ | × | × | |
| rs6218346 | METTL8 | √ | × | × | |
| rs1181095 | MIR548AC | √ | × | × | |
| rs7440363 | MIR548AC | √ | × | × | |
| rs778488 | MIR548AC | √ | × | × | |
| rs778469 | MIR548AC | √ | × | × | |
| rs1701342 | MIR548AC | √ | × | × | |
| rs1112912 | MIR548AC | √ | × | × | |
| rs7304564 | MIR548AC | √ | × | × | |
| rs6225697 | MIR548AC | √ | × | × | |
| rs6809388 | MIR548AC | √ | × | × | |
| rs7996521 | MIR548AC | √ | × | × | |
| rs5785948 | MIR548AC | √ | × | × | |
| rs4513393 | MIR548AC | √ | × | × | |
| rs1174554 | MIR548AC | √ | × | × | |
| rs7643563 | MIR548AC | √ | × | × | Type 2 diab |
| rs1325833 | MLIP | √ | × | × | Height |
| rs1688463 | MLIP | √ | × | × | |
| rs991199 | MLIP | √ | × | × | |
| rs8033418 | MLIP | √ | × | × | |
| rs7544699 | MLIP | × | √ | × | |
| rs9370259 | MLIP | √ | × | × | |
| rs9382281 | MLIP | √ | × | × | |
| rs9367551 | MLIP | √ | × | × | |
| rs7296192 | MLIP | √ | × | × | |
| rs1267639 | MSR1 | √ | × | × | |
| rs6981231 | MSR1 | √ | × | × | |
| rs1330653 | MSR1 | √ | × | × | |
| rs1330653 | MSR1 | √ | × | × | |
| rs1330653 | MSR1 | √ | × | × | |
| rs7005569 | MSR1 | √ | × | × | |
| rs351543 | MSR1 | √ | × | × | |
| rs1199049 | MSR1 | √ | × | × | |
| rs6530943 | MSR1 | √ | × | × | |
| rs1268069 | MSR1 | √ | × | × | |
| rs6737057 | MTERFD2 | √ | × | × | |
| rs1257083 | MXI1 | √ | × | × | |
| rs1881392 | MXI1 | √ | × | × | |
| rs2010327 | MXI1 | √ | × | × | |
| rs6188278 | MXI1 | √ | × | × | |
| rs6188278 | MXI1 | √ | × | × | |
| rs6188279 | MXI1 | √ | × | × | |
| rs7321509 | NAA16 | √ | × | × | |
| rs9562292 | NAA16 | √ | × | × | |
| rs9590595 | NAA16 | √ | × | × | |
| rs9566753 | NAA16 | √ | × | × | |
| rs9566754 | NAA16 | √ | × | × | |
| rs3829396 | NAA16 | √ | × | × | |
| rs9562293 | NAA16 | √ | × | × | |
| rs9562294 | NAA16 | √ | × | × | |
| rs9566757 | NAA16 | √ | × | × | |
| rs3776839 | NDST1 | √ | × | × | |
| rs7645688 | NDST1 | √ | × | × | |
| rs1265710 | NDST1 | √ | × | × | |
| rs3822608 | NDST1 | √ | × | × | |
| rs7664020 | NDST1 | √ | × | × | |
| rs7554172 | NDST1 | √ | × | × | |
| rs7914197 | NDST1 | √ | × | × | |
| rs1045538 | NDST1 | √ | × | × | |
| rs1045539 | NDST1 | √ | × | × | |
| rs1045544 | NDST1 | √ | × | × | |
| rs5857508 | NDST1 | √ | × | × | |
| rs5893237 | NELL1 | √ | × | × | |
| rs1955066 | NELL1 | √ | × | × | |
| rs7970065 | NELL1 | √ | × | × | |
| rs1945406 | NELL1 | √ | × | × | |
| rs1690811 | NELL1 | √ | × | × | |
| rs4451725 | NELL1 | √ | × | × | |
| rs7630894 | NELL1 | √ | × | × | |
| rs5788585 | NELL1 | √ | × | × | |
| rs4363588 | NELL1 | √ | × | × | |
| rs7717891 | NELL1 | √ | × | × | |
| rs7880782 | NELL1 | √ | × | × | |
| rs7963031 | NELL1 | √ | × | × | |
| rs7665820 | NELL1 | √ | × | × | |
| rs1257478 | NELL1 | √ | × | × | |
| rs1102612 | NELL1 | × | √ | × | |
| rs7929603 | NELL1 | × | √ | × | |
| rs1171808 | NELL1 | × | √ | × | |
| rs7123743 | NELL1 | × | √ | × | |
| rs7109470 | NELL1 | × | √ | × | |
| rs6483779 | NELL1 | × | √ | × | |
| rs1176538 | NELL1 | × | √ | × | |
| rs6160262 | NELL1 | × | √ | × | |
| rs5337528 | NELL1 | × | √ | × | |
| rs1083355 | NELL1 | × | √ | × | |
| rs5634677 | NELL1 | × | √ | × | |
| rs5593597 | NELL1 | × | √ | × | |
| rs5598386 | NELL1 | × | √ | × | |
| rs5720668 | NELL1 | × | √ | × | |
| rs1490417 | NELL1 | × | √ | × | |
| rs1180430 | NELL1 | × | √ | × | |
| rs7682701 | NELL1 | × | √ | × | |
| rs7450378 | NELL1 | × | √ | × | |
| rs1083356 | NELL1 | × | √ | × | |
| rs1414227 | NELL1 | × | √ | × | |
| rs7114465 | NELL1 | × | √ | × | |
| rs7929246 | NELL1 | × | √ | × | |
| rs7960891 | NELL1 | × | √ | × | |
| rs7927068 | NELL1 | × | √ | × | |
| rs1083356 | NELL1 | × | √ | × | |
| rs1177894 | NELL1 | × | √ | × | |
| rs7609578 | NELL1 | × | √ | × | |
| rs4923643 | NELL1 | × | √ | × | |
| rs4923644 | NELL1 | × | √ | × | |
| rs7638984 | NELL1 | × | √ | × | |
| rs1083356 | NELL1 | × | √ | × | |
| rs1464587 | NELL1 | × | √ | × | |
| rs1102613 | NELL1 | × | √ | × | |
| rs3680225 | NELL1 | × | √ | × | |
| rs7837379 | NELL1 | × | √ | × | |
| rs1181077 | NELL1 | × | √ | × | |
| rs1811426 | NELL1 | × | √ | × | |
| rs7950695 | NELL1 | × | √ | × | |
| rs1227925 | NELL1 | × | √ | × | Thiazide-inc |
| rs7470486 | NELL1 | × | √ | × | |
| rs1083356 | NELL1 | × | √ | × | |
| rs7926887 | NELL1 | × | √ | × | |
| rs7940878 | NELL1 | × | √ | × | |
| rs5707142 | NELL1 | × | √ | × | |
| rs7927517 | NELL1 | × | √ | × | |
| rs7669406 | NELL1 | × | √ | × | |
| rs7931216 | NELL1 | × | √ | × | |
| rs1242193 | NELL1 | × | √ | × | |
| rs4329685 | NELL1 | × | √ | × | |
| rs7551083 | NELL1 | × | √ | × | |
| rs7594394 | NELL1 | × | √ | × | |
| rs7106789 | NELL1 | × | √ | × | |
| rs1455567 | NELL1 | × | √ | × | |
| rs7769071 | NELL1 | × | √ | × | |
| rs1174624 | NELL1 | × | √ | × | |
| rs1918590 | NELL1 | × | √ | × | |
| rs1434396 | NELL1 | × | √ | × | |
| rs7042420 | NINJ1 | √ | × | × | |
| rs7043145 | NINJ1 | √ | × | × | |
| rs1082107 | NINJ1 | √ | × | × | |
| rs7912732 | NKTR | √ | × | × | |
| rs8351 | NKTR | × | √ | × | |
| rs7891376 | NLRC5 | √ | × | × | |
| rs5584070 | NLRP8 | √ | × | × | |
| rs710163 | NOL8 | √ | × | × | |
| rs1174010 | NOL8 | √ | × | × | |
| rs9409664 | NOL8 | √ | × | × | |
| rs7851699 | NOS1AP | √ | × | × | |
| rs1424956 | NOS1AP | √ | × | × | |
| rs7262709 | NR1D2 | √ | × | × | |
| rs6803587 | NR1D2 | √ | × | × | |
| rs6770052 | NR1D2 | √ | × | × | |
| rs6778577 | NR1D2 | √ | × | × | |
| rs1332143 | NR1D2 | √ | × | × | |
| rs1332144 | NR1D2 | √ | × | × | |
| rs1331781 | NR1D2 | √ | × | × | |
| rs3459353 | NR1D2 | √ | × | × | |
| rs7262709 | NR1D2 | √ | × | × | |
| rs9817923 | NR1D2 | √ | × | × | |
| rs7262709 | NR1D2 | √ | × | × | |
| rs3748722 | NR1D2 | √ | × | × | |
| rs7262709 | NR1D2 | √ | × | × | |
| rs7262810 | NR1D2 | √ | × | × | |
| rs7262810 | NR1D2 | √ | × | × | |
| rs7262810 | NR1D2 | √ | × | × | |
| rs1441445 | NRF1 | × | √ | × | |
| rs7861102 | NRF1 | × | √ | × | |
| rs1270689 | NRF1 | × | √ | × | |
| rs1027081 | NRF1 | √ | × | × | Alzheimer's |
| rs1022510 | NRF1 | √ | × | × | |
| rs7787121 | NRF1 | √ | × | × | |
| rs1755561 | NRF1 | √ | × | × | |
| rs8025734 | NRF1 | √ | × | × | |
| rs4292630 | NRF1 | √ | × | × | |
| rs7473497 | NRF1 | √ | × | × | |
| rs7889031 | NRF1 | √ | × | × | |
| rs2001330 | NRF1 | √ | × | × | |
| rs7884137 | NRF1 | √ | × | × | |
| rs1095425 | NRF1 | × | √ | × | |
| rs1173942 | NRF1 | × | √ | × | |
| rs7967701 | NRF1 | × | √ | × | |
| rs1289453 | NUDT14 | × | √ | × | Immature fr |
| rs1830016 | NUDT14 | × | √ | × | |
| rs7444962 | NUDT14 | × | √ | × | |
| rs1022057 | NUDT14 | × | √ | × | |
| rs3809462 | NUDT14 | × | √ | × | |
| rs1428400 | NUDT14 | × | √ | × | |
| rs3803320 | NUDT14 | × | √ | × | |
| rs1014011 | NUDT14 | × | √ | × | |
| rs1162586 | NUDT14 | × | √ | × | High light sc |
| rs6723293 | NUDT14 | × | √ | × | |
| rs3684136 | NUDT14 | × | √ | × | |
| rs6156434 | NUDT14 | × | √ | × | |
| rs7582962 | NUDT14 | × | √ | × | |
| rs4983590 | NUDT14 | × | √ | × | White blood |
| rs8017877 | NUDT14 | × | √ | × | |
| rs2844685 | OPTC | √ | × | × | |
| rs2306152 | OPTC | √ | × | × | |
| rs9456814 | PACRG | × | √ | × | |
| rs6243090 | PARK2 | √ | × | × | |
| rs6242907 | PARK2 | √ | × | × | |
| rs6242907 | PARK2 | √ | × | × | |
| rs1179789 | PARK2 | √ | × | × | |
| rs7574636 | PARK2 | √ | × | × | |
| rs9347679 | PARK2 | √ | × | × | |
| rs7902015 | PARK2 | √ | × | × | |
| rs1453570 | PARK2 | √ | × | × | |
| rs1463433 | PARK2 | √ | × | × | |
| rs7566357 | PASK | √ | × | × | |
| rs7599243 | PASK | × | √ | × | |
| rs7587318 | PASK | × | √ | × | |
| rs740393 | PASK | × | √ | × | |
| rs7508073 | PASK | × | √ | × | |
| rs2240541 | PASK | × | √ | × | |
| rs2240542 | PASK | × | √ | × | |
| rs4675829 | PASK | √ | × | × | |
| rs1262385 | PASK | √ | × | × | |
| rs2286326 | PASK | √ | × | × | |

| | | | | | |
|---|---|---|---|---|---|
| rs1289804 | RDH12 | √ | × | × | |
| rs1214705 | RDH12 | √ | × | × | |
| rs1214761 | RDH12 | √ | × | × | |
| rs1288231 | RDH12 | √ | × | × | |
| rs1289390 | RDH12 | √ | × | × | |
| rs1289390 | RDH12 | √ | × | × | |
| rs3783197 | RGCC | √ | × | × | |
| rs756279 | RGS10 | √ | × | × | |
| rs7919216 | RGS10 | √ | × | × | |
| rs1043845 | RITA1 | √ | × | × | |
| rs6153377 | RMDN2 | √ | × | × | |
| rs1470479 | RMDN2 | √ | × | × | |
| rs5952724 | RMDN2 | √ | × | × | |
| rs5835958 | RMDN2 | √ | × | × | |
| rs3486578 | RMDN2 | √ | × | × | |
| rs1446297 | RMDN2 | √ | × | × | |
| rs4670802 | RMDN2 | √ | × | × | |
| rs3561518 | RMDN2 | √ | × | × | |
| rs6738545 | RMDN2 | √ | × | × | |
| rs4142168 | RMDN2 | √ | × | × | |
| rs1596862 | RMDN2 | √ | × | × | |
| rs1596861 | RMDN2 | √ | × | × | |
| rs6736956 | RMDN2 | √ | × | × | |
| rs1702181 | RMDN2 | √ | × | × | |
| rs7606530 | RMDN2 | √ | × | × | |
| rs6544129 | RMDN2 | √ | × | × | |
| rs8035131 | RMDN2 | √ | × | × | |
| rs2373327 | RMDN2 | √ | × | × | |
| rs2373328 | RMDN2 | √ | × | × | |
| rs1988 | RMDN2 | √ | × | × | |
| rs1562285 | RMDN2 | √ | × | × | |
| rs739829 | RNF112 | × | × | √ | |
| rs7572450 | RNF112 | × | × | √ | |
| rs9507412 | RNF17 | √ | × | × | |
| rs1161634 | RNF17 | √ | × | × | |
| rs1181525 | RNF17 | √ | × | × | |
| rs6131466 | RNF17 | √ | × | × | |
| rs1161792 | RNF17 | √ | × | × | |
| rs5829791 | RNF17 | √ | × | × | |
| rs7512257 | RNF17 | √ | × | × | |
| rs7319350 | RNF17 | √ | × | × | |
| rs1228652 | RNF214 | √ | × | × | |
| rs13339 | RPP25L | √ | × | × | Vaginal micr |
| rs4581611 | RPS29 | √ | × | × | |
| rs1015062 | RPS29 | √ | × | × | |
| rs1335342 | RTP3 | √ | × | × | |
| rs5926041 | SAMD7 | √ | × | × | |
| rs6142079 | SAMD7 | √ | × | × | |
| rs7535320 | SCARB1 | √ | × | × | |
| rs7306507 | SCARB1 | √ | × | × | |
| rs3924313 | SCARB1 | √ | × | × | Concentrati |
| rs1105785 | SCARB1 | √ | × | × | |
| rs1105786 | SCARB1 | √ | × | × | |
| rs1105786 | SCARB1 | √ | × | × | Response t |
| rs1389926 | SEC22C | √ | × | × | |
| rs637852 | SEC22C | √ | × | × | |
| rs1306431 | SEC22C | √ | × | × | |
| rs3111313 | SEC22C | √ | × | × | |
| rs7493585 | SEMA4D | √ | × | × | |
| rs1395445 | SIK3 | √ | × | × | |
| rs1416582 | SIK3 | √ | × | × | |
| rs3517786 | SKIL | √ | × | × | |
| rs6444920 | SKIL | √ | × | × | |
| rs1192447 | SKIL | √ | × | × | |
| rs1093662 | SKIL | √ | × | × | |
| rs1021956 | SLC15A5 | √ | × | × | |
| rs1436122 | SLC15A5 | √ | × | × | |
| rs7036499 | SLC25A25 | √ | × | × | Height |
| rs7036522 | SLC25A25 | √ | × | × | |
| rs1234894 | SLC25A25 | √ | × | × | |
| rs1233545 | SLC25A25 | √ | × | × | |
| rs6258693 | SLC25A25 | √ | × | × | |
| rs1178811 | SLC25A25 | √ | × | × | |
| rs7044673 | SLC25A25 | √ | × | × | |
| rs1073970 | SLC25A25 | √ | × | × | |
| rs1073970 | SLC25A25 | √ | × | × | |
| rs7854499 | SLC25A25 | √ | × | × | |
| rs1081934 | SLC25A25 | √ | × | × | |
| rs5704331 | SLC2A13 | √ | × | × | |
| rs7801694 | SLC2A13 | √ | × | × | |
| rs1117434 | SLC2A13 | √ | × | × | |
| rs1230162 | SLC2A13 | √ | × | × | |
| rs5753523 | SLC2A13 | √ | × | × | |
| rs1117436 | SLC2A13 | √ | × | × | |
| rs953824 | SLC2A13 | √ | × | × | |
| rs1117438 | SLC2A13 | √ | × | × | |
| rs1497049 | SLC2A13 | √ | × | × | |
| rs1051259 | SLC39A11 | √ | × | × | |
| rs5715238 | SLIT3 | × | √ | × | |
| rs7651775 | SLIT3 | × | √ | × | |
| rs1317260 | SLIT3 | × | √ | × | Body mass |
| rs1317688 | SLIT3 | × | √ | × | |
| rs1174872 | SLIT3 | × | √ | × | |
| rs1513224 | SLIT3 | × | √ | × | |
| rs5562085 | SLIT3 | × | √ | × | |
| rs1006649 | SLIT3 | × | √ | × | |
| rs1006650 | SLIT3 | × | √ | × | |
| rs7158984 | SLIT3 | × | √ | × | |
| rs1006663 | SLIT3 | × | √ | × | |
| rs7703544 | SLIT3 | × | √ | × | |
| rs7721574 | SLIT3 | × | √ | × | |
| rs7721721 | SLIT3 | × | √ | × | |
| rs6555838 | SLIT3 | × | √ | × | |
| rs6886126 | SLIT3 | × | √ | × | |
| rs1215339 | SLIT3 | × | √ | × | |
| rs5418804 | SLIT3 | × | √ | × | |
| rs8026207 | SLIT3 | × | √ | × | |
| rs1899595 | SLIT3 | × | √ | × | |
| rs6237722 | SLIT3 | × | √ | × | |
| rs6555841 | SLIT3 | × | √ | × | |
| rs1390360 | SLIT3 | × | √ | × | |
| rs7802595 | SLIT3 | × | √ | × | |
| rs1218835 | SLIT3 | × | √ | × | Inflammator |
| rs1898651 | SLIT3 | × | √ | × | |
| rs7759151 | SLIT3 | × | √ | × | |
| rs6555842 | SLIT3 | × | √ | × | |
| rs1812393 | SLIT3 | × | √ | × | |
| rs1265251 | SLIT3 | × | √ | × | |
| rs7749334 | SLIT3 | × | √ | × | |
| rs7578391 | SLIT3 | × | √ | × | |
| rs1481312 | SLIT3 | × | √ | × | |
| rs1174981 | SLIT3 | × | √ | × | |
| rs3731690 | SOX5 | √ | × | × | |
| rs5332491 | SOX5 | √ | × | × | |
| rs5441035 | SOX5 | √ | × | × | |
| rs1909356 | SOX5 | √ | × | × | |
| rs4262802 | SOX5 | √ | × | × | |
| rs1104743 | SOX5 | √ | × | × | |
| rs1104744 | SOX5 | √ | × | × | Height |
| rs1104745 | SOX5 | √ | × | × | |
| rs7991782 | SPOCK3 | √ | × | × | |
| rs7690787 | SPOCK3 | √ | × | × | |
| rs2168405 | SPOCK3 | √ | × | × | |
| rs2124457 | SPOCK3 | √ | × | × | |
| rs2054146 | SPOCK3 | √ | × | × | |
| rs2054145 | SPOCK3 | √ | × | × | |
| rs2054144 | SPOCK3 | √ | × | × | |
| rs2318488 | SPOCK3 | √ | × | × | |
| rs4602517 | SPOCK3 | √ | × | × | |
| rs1314311 | SPOCK3 | √ | × | × | |
| rs1312715 | SPOCK3 | √ | × | × | |
| rs1250113 | SPOCK3 | √ | × | × | |
| rs1081941 | SPTAN1 | √ | × | × | |
| rs2275258 | SPTAN1 | √ | × | × | |
| rs7046238 | SPTAN1 | √ | × | × | |
| rs1442884 | SPTAN1 | √ | × | × | |
| rs7994082 | SPTAN1 | √ | × | × | |
| rs7357662 | SPTAN1 | √ | × | × | |
| rs7914016 | SPTAN1 | √ | × | × | |
| rs1455664 | SPTAN1 | √ | × | × | |
| rs1170019 | SPTAN1 | √ | × | × | |
| rs3737308 | SPTAN1 | √ | × | × | |
| rs1447718 | SPTLC1 | √ | × | × | |
| rs1174032 | ST3GAL5 | √ | × | × | |
| rs7565401 | ST3GAL5 | √ | × | × | |
| rs3770092 | ST3GAL5 | √ | × | × | |
| rs1169122 | ST3GAL5 | √ | × | × | |
| rs1120287 | STAMBPL1 | √ | × | × | |
| rs7912777 | STAMBPL1 | √ | × | × | Brain regior |
| rs2304803 | STAMBPL1 | √ | × | × | |
| rs1983773 | STX11 | √ | × | × | |
| rs1512297 | SUSD4 | √ | × | × | |
| rs712856 | SUSD4 | √ | × | × | |
| rs1400962 | SUSD4 | √ | × | × | |
| rs1772270 | SUSD4 | √ | × | × | |
| rs1772271 | SUSD4 | √ | × | × | |
| rs3740979 | SYNPO | √ | × | × | |
| rs9387924 | TBC1D32 | √ | × | × | |
| rs9320793 | TBC1D32 | √ | × | × | |
| rs9401380 | TBC1D32 | √ | × | × | |
| rs9387929 | TBC1D32 | √ | × | × | |
| rs4498397 | TBC1D32 | √ | × | × | |
| rs7730932 | TBC1D32 | √ | × | × | |
| rs8000979 | TBC1D32 | √ | × | × | |
| rs7776680 | TBC1D32 | √ | × | × | |
| rs8004117 | TBC1D32 | √ | × | × | |
| rs7500153 | TBC1D32 | √ | × | × | |
| rs7699776 | TBC1D32 | √ | × | × | |
| rs8016466 | TBC1D32 | √ | × | × | |
| rs4245508 | TBC1D32 | √ | × | × | |
| rs1049910 | TBC1D32 | √ | × | × | |
| rs1049910 | TBC1D32 | √ | × | × | |
| rs7965371 | TBC1D32 | √ | × | × | |
| rs7922147 | TBC1D32 | √ | × | × | |
| rs7978880 | TBC1D32 | √ | × | × | |
| rs1708336 | TBC1D32 | √ | × | × | |
| rs7639777 | TBC1D32 | √ | × | × | |
| rs7814615 | TBC1D32 | √ | × | × | |
| rs8006865 | TBC1D32 | √ | × | × | |
| rs7556238 | TBC1D32 | √ | × | × | |
| rs7960459 | TBC1D32 | √ | × | × | |
| rs1708337 | TBC1D32 | √ | × | × | |
| rs7898263 | TBC1D32 | √ | × | × | |
| rs7540935 | TBC1D32 | √ | × | × | |
| rs7696351 | TBC1D32 | √ | × | × | |
| rs7461402 | TBC1D32 | √ | × | × | |
| rs7500456 | TBC1D32 | √ | × | × | |
| rs7877642 | TBC1D32 | √ | × | × | |
| rs1708338 | TBC1D32 | √ | × | × | |
| rs7687361 | TBC1D32 | √ | × | × | |
| rs7590986 | TBC1D32 | √ | × | × | |
| rs4946561 | TBC1D32 | √ | × | × | |
| rs7461365 | TBC1D32 | √ | × | × | |
| rs7904069 | TBC1D32 | √ | × | × | |
| rs7827847 | TBC1D32 | √ | × | × | |
| rs7461890 | TBC1D32 | √ | × | × | |
| rs1177189 | TBC1D32 | √ | × | × | |
| rs7770194 | TBC1D32 | √ | × | × | |
| rs6155091 | TBC1D32 | √ | × | × | |
| rs7605106 | TBC1D32 | √ | × | × | |
| rs7674303 | TBC1D32 | √ | × | × | |
| rs7937144 | TBC1D32 | √ | × | × | |
| rs1708339 | TBC1D32 | √ | × | × | |
| rs7790138 | TBC1D32 | √ | × | × | |
| rs7690330 | TBC1D32 | √ | × | × | |
| rs7962999 | TBC1D32 | √ | × | × | |
| rs4144169 | TBC1D32 | √ | × | × | |
| rs7648458 | TBC1D32 | √ | × | × | |
| rs1049910 | TBC1D32 | √ | × | × | |
| rs1049910 | TBC1D32 | √ | × | × | |
| rs7574735 | TBC1D32 | √ | × | × | |
| rs1833149 | TBC1D32 | √ | × | × | |
| rs7692908 | TBC1D32 | √ | × | × | |
| rs9320808 | TBC1D32 | √ | × | × | |
| rs7069305 | TET1 | √ | × | × | |
| rs2133122 | TET1 | √ | × | × | |
| rs1224335 | TET1 | √ | × | × | |
| rs5604775 | TET1 | √ | × | × | |
| rs7069195 | TET1 | √ | × | × | |
| rs1692556 | TET1 | √ | × | × | |
| rs3565178 | TET1 | √ | × | × | |
| rs1226364 | TET1 | √ | × | × | |
| rs2153813 | TET1 | √ | × | × | |
| rs2185741 | TET1 | √ | × | × | |
| rs3998859 | TET1 | √ | × | × | |
| rs1421624 | TFDP2 | × | √ | × | |
| rs7633951 | TFDP2 | × | √ | × | |
| rs7560098 | TFDP2 | × | √ | × | |
| rs347689 | TFDP2 | × | √ | × | |

| | | | | |
|---|---|---|---|---|
| rs989656 | SOX5 | × | √ | |
| rs989657 | SOX5 | × | √ | Height |
| rs1498869 | SOX5 | × | √ | |
| rs1497715 | SOX5 | × | √ | |
| rs1391786 | SOX5 | × | √ | |
| rs1692696 | SOX5 | × | √ | |
| rs1465986 | SOX5 | × | √ | |
| rs1104720 | SOX5 | × | √ | Externalizing behaviour (multivariate analysis) |
| rs5421584 | SOX5 | × | √ | |
| rs1242458 | SOX5 | × | √ | |
| rs1692697 | SOX5 | × | √ | Depression |
| rs6487369 | SOX5 | × | √ | |
| rs1104721 | SOX5 | × | √ | |
| rs7885627 | SOX5 | × | √ | |
| rs4963735 | SOX5 | × | √ | |
| rs4963737 | SOX5 | × | √ | |
| rs1748118 | SOX5 | × | √ | |
| rs1104721 | SOX5 | × | √ | |
| rs1468386 | SOX5 | × | √ | |
| rs1230755 | SOX5 | × | √ | |
| rs1104721 | SOX5 | × | √ | |
| rs7316782 | SOX5 | × | √ | |
| rs1417273 | SOX5 | × | √ | |
| rs7973213 | SOX5 | × | √ | |
| rs7973352 | SOX5 | × | √ | |
| rs1021671 | SOX5 | × | √ | |
| rs1282333 | SOX5 | × | √ | |
| rs1044441 | SOX5 | × | √ | |
| rs1021672 | SOX5 | × | √ | |
| rs1050592 | SOX5 | × | √ | |
| rs1104721 | SOX5 | × | √ | |
| rs1353792 | SOX5 | × | √ | C-reactive protein levels |
| rs1104721 | SOX5 | × | √ | |
| rs1084226 | SOX5 | × | √ | |
| rs1740368 | SOX5 | × | √ | |
| rs1748214 | SOX5 | × | √ | |
| rs7485933 | SOX5 | × | √ | |
| rs1498870 | SOX5 | × | √ | |
| rs3725161 | SOX5 | × | √ | |
| rs7307316 | SOX5 | × | √ | |
| rs1391788 | SOX5 | × | √ | |
| rs5462017 | SOX5 | × | √ | |
| rs1909351 | SOX5 | × | √ | |
| rs1104721 | SOX5 | × | √ | |
| rs1084226 | SOX5 | × | √ | Non-obstructive azoospermia |
| rs7328383 | SOX5 | × | √ | |
| rs3478991 | SOX5 | × | √ | |
| rs7138690 | SOX5 | × | √ | |
| rs1084226 | SOX5 | × | √ | |
| rs1039229 | SOX5 | × | √ | |
| rs1455373 | SOX5 | × | √ | |
| rs1144278 | SOX5 | × | √ | |
| rs1230064 | SOX5 | × | √ | |
| rs1282392 | SOX5 | × | √ | |
| rs1443212 | SOX5 | × | √ | |
| rs1172195 | SOX5 | × | √ | |
| rs1104722 | SOX5 | × | √ | |
| rs1077105 | SOX5 | × | √ | |
| rs1281338 | SOX5 | × | √ | |
| rs1281291 | SOX5 | × | √ | |
| rs7516792 | SOX5 | × | √ | |
| rs1353793 | SOX5 | × | √ | |
| rs1748310 | SOX5 | × | √ | |
| rs1381660 | SOX5 | × | √ | |
| rs1104722 | SOX5 | × | √ | |
| rs971012 | SOX5 | × | √ | |
| rs3899208 | SOX5 | × | √ | |
| rs7465966 | SOX5 | × | √ | |
| rs1692707 | SOX5 | × | √ | |
| rs1237161 | SOX5 | × | √ | |
| rs1169017 | SOX5 | × | √ | |
| rs7754876 | SOX5 | × | √ | |
| rs7962638 | SOX5 | × | √ | |
| rs7966115 | SOX5 | × | √ | |
| rs1242629 | SOX5 | × | √ | |
| rs7838470 | SOX5 | × | √ | |
| rs3594053 | SOX5 | × | √ | |
| rs7860909 | SOX5 | × | √ | |
| rs997189 | SOX5 | × | √ | |
| rs7634975 | SOX5 | × | √ | |
| rs7570055 | SOX5 | × | √ | |
| rs4963740 | SOX5 | × | √ | |
| rs1391790 | SOX5 | × | √ | Sex hormone-binding globulin levels adjusted for BMI |
| rs3513581 | SOX5 | × | √ | |
| rs3702154 | SOX5 | × | √ | |
| rs7961391 | SOX5 | × | √ | |
| rs4963741 | SOX5 | × | √ | |
| rs2002544 | SOX5 | × | √ | |
| rs3456458 | SOX5 | × | √ | |
| rs7850466 | SOX5 | × | √ | |
| rs7652978 | SOX5 | × | √ | |
| rs1470721 | SOX5 | × | √ | |
| rs1242519 | SOX5 | × | √ | |
| rs1459844 | SOX5 | × | √ | |
| rs1104723 | SOX5 | × | √ | |
| rs1507630 | SOX5 | × | √ | |
| rs7643224 | SOX5 | × | √ | |
| rs7789014 | SOX5 | × | √ | |
| rs1816479 | SOX5 | × | √ | |
| rs1044441 | SOX5 | × | √ | |
| rs3722002 | SOX5 | × | √ | |
| rs1692712 | SOX5 | × | √ | |
| rs1498876 | SOX5 | × | √ | |
| rs7488253 | SOX5 | × | √ | |
| rs1498877 | SOX5 | × | √ | |
| rs1498878 | SOX5 | × | √ | |
| rs1498879 | SOX5 | × | √ | |
| rs1282870 | SOX5 | × | √ | |
| rs1748514 | SOX5 | × | √ | Suicide attempt |
| rs4963565 | SOX5 | × | √ | |
| rs5332491 | SOX5 | × | √ | |
| rs1748523 | SOX5 | × | √ | |
| rs7594883 | SOX5 | × | √ | |
| rs7964057 | SOX5 | × | √ | |
| rs1546227 | SOX5 | × | √ | |
| rs7938453 | SOX5 | × | √ | |
| rs7728233 | SOX5 | × | √ | |
| rs8023507 | SOX5 | × | √ | |
| rs7516531 | SOX5 | × | √ | |
| rs1084226 | SOX5 | × | √ | |
| rs5757906 | SOX5 | × | √ | |
| rs1084226 | SOX5 | × | √ | |
| rs7296480 | SOX5 | × | √ | |
| rs1498883 | SOX5 | × | √ | |
| rs7749761 | SOX5 | × | √ | |
| rs1885250 | SOX5 | × | √ | |
| rs1916954 | SOX5 | × | √ | |
| rs7307217 | SOX5 | × | √ | |
| rs7978583 | SOX5 | √ | × | |
| rs7966757 | SOX5 | √ | × | |
| rs1084227 | SOX5 | √ | × | |
| rs1232212 | SOX5 | √ | × | |
| rs1909356 | SOX5 | √ | × | |
| rs1104745 | SOX5 | √ | × | |
| rs1104745 | SOX5 | √ | × | |
| rs7957500 | SOX5 | √ | × | |
| rs2418176 | SOX5 | √ | × | |
| rs4963760 | SOX5 | √ | × | |
| rs1104746 | SOX5 | √ | × | |
| rs7299191 | SOX5 | √ | × | |
| rs1230916 | SOX5 | √ | × | |
| rs1104746 | SOX5 | √ | × | |
| rs7301819 | SOX5 | √ | × | |
| rs7961598 | SOX5 | √ | × | |
| rs4963765 | SOX5 | √ | × | |
| rs6487389 | SOX5 | √ | × | |
| rs9630247 | SOX5 | √ | × | |
| rs1084236 | SOX5 | √ | × | |
| rs4131686 | SOX5 | √ | × | |
| rs4131685 | SOX5 | √ | × | |
| rs4963766 | SOX5 | √ | × | |
| rs4963767 | SOX5 | √ | × | |
| rs4963768 | SOX5 | √ | × | |
| rs7295669 | SOX5 | √ | × | |
| rs1183306 | SOX5 | √ | × | Smoking initiation |
| rs7974269 | SOX5 | √ | × | |
| rs4963575 | SOX5 | √ | × | |
| rs7132989 | SOX5 | √ | × | |
| rs7693792 | SPOCK3 | √ | × | |
| rs7269559 | SPOCK3 | √ | × | |
| rs7991782 | SPOCK3 | √ | × | |
| rs7269750 | SPOCK3 | √ | × | |
| rs7419786 | SPOCK3 | √ | × | |
| rs2318487 | SPOCK3 | √ | × | |
| rs7576467 | SPTLC1 | √ | × | |
| rs1234932 | SPTLC1 | √ | × | |
| rs1233668 | SPTLC1 | √ | × | |
| rs7351237 | SPTLC1 | √ | × | |
| rs7021408 | SPTLC1 | √ | × | |
| rs1234179 | SPTLC1 | √ | × | |
| rs1233629 | SPTLC1 | √ | × | |
| rs2297568 | SPTLC1 | √ | × | |
| rs1048461 | STX11 | √ | × | |
| rs9311871 | SYNPR | √ | × | |
| rs4499588 | SYNPR | √ | × | |
| rs2887081 | SYNPR | √ | × | |
| rs2367786 | SYNPR | √ | × | |
| rs7645288 | SYNPR | √ | × | |
| rs7648834 | SYNPR | √ | × | |
| rs2367788 | SYNPR | √ | × | |
| rs4377491 | SYNPR | √ | × | |
| rs1192861 | SYNPR | √ | × | |
| rs1554574 | SYNPR | √ | × | |
| rs2056546 | SYNPR | √ | × | |
| rs2056545 | SYNPR | √ | × | |
| rs1113092 | SYNPR | √ | × | |
| rs1105143 | SYNPR | √ | × | |
| rs2367789 | SYNPR | √ | × | |
| rs1916628 | SYNPR | √ | × | |
| rs1113092 | SYNPR | √ | × | |
| rs1523440 | SYNPR | √ | × | |
| rs1523441 | SYNPR | √ | × | |
| rs1308051 | SYNPR | √ | × | |
| rs1308058 | SYNPR | √ | × | |
| rs1874927 | SYNPR | √ | × | |
| rs1874928 | SYNPR | √ | × | |
| rs1309274 | SYNPR | √ | × | |
| rs1608451 | SYNPR | √ | × | |
| rs7626955 | SYNPR | √ | × | |
| rs949610 | SYNPR | √ | × | |
| rs1086606 | SYNPR | √ | × | |
| rs1523437 | SYNPR | √ | × | |
| rs4688392 | SYNPR | √ | × | |
| rs1996917 | SYNPR | √ | × | |
| rs6791221 | SYNPR | √ | × | |
| rs1523436 | SYNPR | √ | × | |
| rs3584372 | SYNPR | √ | × | |
| rs3609311 | SYNPR | √ | × | |
| rs1308584 | SYNPR | √ | × | |
| rs1248937 | SYNPR | √ | × | |
| rs1249655 | SYNPR | √ | × | |
| rs1249652 | SYNPR | √ | × | |
| rs1851666 | SYNPR | √ | × | |
| rs2367791 | SYNPR | √ | × | |
| rs7647850 | SYNPR | √ | × | |
| rs1309680 | SYNPR | √ | × | |
| rs1307658 | SYNPR | √ | × | |
| rs1451312 | SYNPR | √ | × | |
| rs1113092 | SYNPR | √ | × | |
| rs1376482 | SYNPR | √ | × | |
| rs1376481 | SYNPR | √ | × | |
| rs4688166 | SYNPR | √ | × | |
| rs1357522 | SYNPR | √ | × | |
| rs4234675 | SYNPR | √ | × | |
| rs7619968 | SYNPR | × | √ | |
| rs4422300 | SYNPR | √ | × | |
| rs6806142 | SYNPR | √ | × | |
| rs1901302 | SYNPR | √ | × | |
| rs1051089 | SYNPR | √ | × | |
| rs1706839 | SYNPR | √ | × | |
| rs1706843 | SYNPR | √ | × | |
| rs5842436 | SYNPR | √ | × | |
| rs1706844 | SYNPR | √ | × | |
| rs1307500 | SYNPR | √ | × | |
| rs1306681 | SYNPR | √ | × | |
| rs3487044 | SYNPR | √ | × | |
| rs7647290 | SYNPR | √ | × | |
| rs3608239 | SYNPR | √ | × | |
| rs2002943 | SYNPR | √ | × | |

| | | | | | | | | | | | | | | | | | | | | | | |
|---|---|---|---|---|---|---|---|---|---|---|---|---|---|---|---|---|---|---|---|---|---|---|
| rs78455529 | TBC1D32 | √ | × | × | | rs2286327 | PASK | √ | × | × | | rs347690 | TFDP2 | × | √ | × | | rs2005116 | SYNPR | √ | × | |
| rs76083652 | TBC1D32 | √ | × | × | | rs5500542 | PASK | √ | × | × | | rs7730675 | TFDP2 | × | √ | × | | rs9311875 | SYNPR | √ | × | |
| rs78187099 | TBC1D32 | √ | × | × | | rs4768139 | PCED1B | √ | × | × | | rs5404695 | TFDP2 | × | √ | × | | rs8008343 | TACC2 | √ | × | |
| rs4945683 | TBC1D32 | √ | × | × | | rs2408716 | PCED1B | √ | × | × | | rs1920103 | TFDP2 | √ | × | × | | rs6816462 | TBC1D1 | √ | × | |
| rs4946560 | TBC1D32 | √ | × | × | | rs1088104 | PCED1B | √ | × | × | | rs347691 | TFDP2 | × | √ | × | | rs1764432 | TBC1D32 | √ | × | |
| rs17083385 | TBC1D32 | √ | × | × | | rs1078565 | PCED1B | √ | × | × | | rs7323266 | TFDP2 | × | √ | × | | rs9385190 | TBC1D32 | √ | × | |
| rs79274950 | TBC1D32 | √ | × | × | | rs7277073 | PCSK6 | √ | × | × | | rs7323266 | TFDP2 | × | √ | × | | rs1406241 | TBC1D32 | √ | × | |
| rs9320803 | TBC1D32 | √ | × | × | | rs6707850 | PCSK6 | √ | × | × | | rs7521896 | THEMIS | √ | × | × | | rs7730932 | TBC1D32 | √ | × | |
| rs4412244 | TBC1D32 | √ | × | × | | rs5574392 | PCSK6 | √ | × | × | | rs2596626 | THRB | √ | × | × | | rs9387941 | TBC1D32 | √ | × | |
| rs9375022 | TBC1D32 | √ | × | × | | rs7863947 | PER2 | √ | × | × | | rs2683530 | THRB | √ | × | × | | rs9482131 | TBC1D32 | √ | × | |
| rs9398629 | TBC1D32 | √ | × | × | OTU99_45 (Faecalibacter | rs5705660 | PER2 | √ | × | × | | rs1562736 | THRB | √ | × | × | | rs9387943 | TBC1D32 | √ | × | |
| rs73201491 | TBX5 | √ | × | × | | rs2278951 | PEX12 | √ | × | × | | rs2683535 | THRB | √ | × | × | | rs7655047 | TBC1D32 | √ | × | |
| rs16944153 | TBX5 | √ | × | × | | rs1249260 | PHC3 | √ | × | × | | rs1395126 | THRB | √ | × | × | | rs2270460 | TCF25 | √ | × | |
| rs12930606 | TCF25 | × | √ | × | | rs888507 | PHC3 | √ | × | × | | rs4260345 | THRB | √ | × | × | Spherical e | rs1186238 | TCF25 | √ | × | Red vs. brown/black hair color |
| rs62052168 | TCF25 | × | √ | × | | rs879161 | PHC3 | √ | × | × | | rs826251 | THRB | √ | × | × | | rs6205218 | TCF25 | √ | × | |
| rs186838083 | TCF25 | × | √ | × | | rs1685484 | PHC3 | √ | × | × | | rs5887898 | TLR6 | √ | × | × | | rs1415416 | TET1 | √ | × | |
| rs75656929 | TCF25 | × | √ | × | | rs6781454 | PHC3 | √ | × | × | | rs1171940 | TLR6 | √ | × | × | | rs3804771 | TFDP2 | √ | × | Body mass index |
| rs17784386 | TCF25 | × | √ | × | | rs7638400 | PHC3 | √ | × | × | | rs2087465 | TLR6 | √ | × | × | | rs3828397 | TFDP2 | √ | × | |
| rs554000291 | TCF25 | × | √ | × | | rs6808506 | PHC3 | √ | × | × | Eosinophil c | rs7323663 | TLR6 | √ | × | × | | rs6228313 | TFDP2 | √ | × | |
| rs34604714 | TCF25 | × | √ | × | | rs7387917 | PHC3 | √ | × | × | | rs7673124 | TLR6 | √ | × | × | | rs7635444 | TFDP2 | √ | × | |
| rs189400654 | TCF25 | × | √ | × | | rs1080483 | PHC3 | √ | × | × | | rs1179850 | TLR6 | √ | × | × | | rs7323268 | TFDP2 | √ | × | |
| rs536152299 | TCF25 | × | √ | × | | rs6808181 | PHC3 | √ | × | × | | rs7691002 | TLR6 | √ | × | × | | rs347679 | TFDP2 | √ | × | |
| rs72477006 | TCF25 | × | √ | × | Red vs. brown/black hair c | rs7614670 | PHC3 | √ | × | × | | rs1258208 | TMEM117 | √ | × | × | | rs347681 | TFDP2 | √ | × | |
| rs73276534 | TCF25 | × | √ | × | | rs7387917 | PHC3 | √ | × | × | | rs1623829 | TMEM117 | √ | × | × | | rs347682 | TFDP2 | √ | × | |
| rs554983078 | TCF25 | × | √ | × | | rs7387918 | PHC3 | √ | × | × | | rs1731434 | TMEM117 | √ | × | × | | rs347684 | TFDP2 | √ | × | |
| rs62052173 | TCF25 | × | √ | × | | rs7387918 | PHC3 | √ | × | × | | rs1731423 | TMEM117 | √ | × | × | | rs347686 | TFDP2 | √ | × | |
| rs62052174 | TCF25 | × | √ | × | | rs1248898 | PHC3 | √ | × | × | | rs1731433 | TMEM117 | √ | × | × | | rs1170593 | TFDP2 | √ | × | Testicular germ cell tumor |
| rs16966253 | TCF25 | × | √ | × | | rs3772186 | PHC3 | √ | × | × | | rs1731432 | TMEM117 | √ | × | × | | rs6773343 | TFDP2 | √ | × | Serum 25-Hydroxyvitamin D levels |
| rs9806913 | TCF25 | × | √ | × | | rs1723683 | PHC3 | √ | × | × | | rs1731431 | TMEM117 | √ | × | × | | rs2683530 | THRB | √ | × | |
| rs9922515 | TCF25 | × | √ | × | | rs7359266 | PLAGL1 | √ | × | × | | rs2614394 | TMEM117 | √ | × | × | | rs1562736 | THRB | √ | × | |
| rs11642267 | TCF25 | × | √ | × | | rs5608126 | PLAGL1 | √ | × | × | | rs1675764 | TMEM117 | √ | × | × | | rs2683535 | THRB | √ | × | |
| rs4287569 | TCF25 | × | √ | × | | rs5973747 | PLCL2 | √ | × | × | | rs1731459 | TMEM117 | √ | × | × | | rs9395936 | TINAG | √ | × | |
| rs564206127 | TCF25 | × | √ | × | | rs4684306 | PLCL2 | √ | × | × | | rs1731458 | TMEM117 | √ | × | × | | rs1417387 | TINAG | √ | × | |
| rs11645970 | TCF25 | × | √ | × | | rs6224603 | PLCL2 | √ | × | × | | rs1731457 | TMEM117 | √ | × | × | | rs1876606 | TINAG | √ | × | |
| rs7196840 | TCF25 | × | √ | × | | rs5875070 | PLCL2 | √ | × | × | | rs1731456 | TMEM117 | √ | × | × | | rs1221523 | TINAG | √ | × | |
| rs7199685 | TCF25 | × | √ | × | | rs1180205 | PLCL2 | √ | × | × | | rs1358705 | TMEM117 | √ | × | × | | rs9349706 | TINAG | √ | × | |
| rs7186976 | TCF25 | × | √ | × | | rs1498352 | PLCL2 | √ | × | × | | rs1622590 | TMEM117 | √ | × | × | | rs9370291 | TINAG | √ | × | |
| rs7191836 | TCF25 | × | √ | × | | rs6573777 | PLEKHH1 | √ | × | × | | rs1614042 | TMEM117 | √ | × | × | | rs9367570 | TINAG | √ | × | |
| rs7184960 | TCF25 | × | √ | × | | rs941704 | PLEKHH1 | √ | × | × | | rs1675789 | TMEM117 | √ | × | × | | rs6531673 | TLR6 | √ | × | |
| rs141678268 | TCF25 | × | √ | × | | rs7649168 | PLEKHH1 | √ | × | × | | rs1675787 | TMEM117 | √ | × | × | | rs7681628 | TLR6 | √ | × | |
| rs11641790 | TCF25 | × | √ | × | Metabolite levels | rs1014051 | PLEKHH1 | √ | × | × | | rs1675781 | TMEM117 | √ | × | × | | rs7689284 | TLR6 | √ | × | |
| rs2293586 | TCF25 | × | √ | × | | rs3809389 | PLEKHH1 | × | × | √ | | rs1675785 | TMEM117 | √ | × | × | | rs7755629 | TMEM117 | √ | × | |
| rs75110337 | TCF25 | × | √ | × | | rs7675454 | PLXDC2 | √ | × | × | | rs4354753 | TMEM117 | √ | × | × | | rs7907454 | TMEM117 | √ | × | |
| rs77963546 | TCF25 | × | √ | × | | rs1262047 | PLXNA2 | × | √ | × | | rs7955615 | TMEM117 | √ | × | × | | rs7477523 | TMEM117 | √ | × | |
| rs72813432 | TCF25 | × | √ | × | | rs3754979 | PLXNA2 | × | √ | × | | rs1474560 | TMEM117 | √ | × | × | | rs1134512 | TMEM117 | √ | × | |
| rs11648433 | TCF25 | × | √ | × | | rs1861733 | PLXNA2 | × | √ | × | | rs1088057 | TMEM117 | √ | × | × | | rs1118229 | TMEM117 | √ | × | |
| rs190775582 | TCF25 | × | √ | × | | rs1179791 | PLXNA2 | × | √ | × | | rs1088058 | TMEM117 | √ | × | × | | rs1118229 | TMEM117 | √ | × | |
| rs73276564 | TCF25 | × | √ | × | | rs1335045 | PLXNA2 | × | √ | × | | rs4572199 | TMEM117 | √ | × | × | | rs1621279 | TMEM117 | √ | × | |
| rs35542367 | TCF25 | × | √ | × | | rs7759288 | PLXNA2 | × | √ | × | | rs1078546 | TMEM117 | √ | × | × | | rs1118230 | TMEM117 | √ | × | |
| rs10153055 | TCF25 | × | √ | × | | rs5631482 | PLXNA2 | × | √ | × | | rs1078546 | TMEM117 | √ | × | × | | rs1118230 | TMEM117 | √ | × | |
| rs11646448 | TCF25 | × | √ | × | | rs7659340 | PLXNA2 | × | √ | × | | rs1169302 | TMEM117 | √ | × | × | | rs7604888 | TMEM117 | √ | × | |
| rs62052183 | TCF25 | × | √ | × | | rs7533778 | PLXNA2 | × | √ | × | | rs5019975 | TMEM117 | √ | × | × | | rs1118230 | TMEM117 | √ | × | |
| rs62052184 | TCF25 | × | √ | × | | rs7547688 | PLXNA2 | × | √ | × | | rs4537822 | TMEM117 | √ | × | × | | rs1118230 | TMEM117 | √ | × | |
| rs76048225 | TCF25 | × | √ | × | | rs5650615 | PLXNA2 | × | √ | × | | rs4238089 | TMEM117 | √ | × | × | | rs1118230 | TMEM117 | √ | × | |
| rs10153210 | TCF25 | × | √ | × | | rs4844661 | PLXNA2 | × | √ | × | | rs4238090 | TMEM117 | √ | × | × | | rs1118230 | TMEM117 | √ | × | |
| rs144168862 | TCF25 | × | √ | × | | rs4844662 | PLXNA2 | × | √ | × | | rs1827583 | TMEM117 | √ | × | × | | rs3829807 | TMEM117 | √ | × | |
| rs10153196 | TCF25 | × | √ | × | | rs5628417 | PLXNA2 | × | √ | × | | rs7312422 | TMEM117 | √ | × | × | | rs1222949 | TMEM117 | √ | × | |
| rs10153084 | TCF25 | × | √ | × | | rs7915066 | POLR1A | √ | × | × | | rs6582493 | TMEM117 | √ | × | × | | rs1675772 | TMEM117 | √ | × | |
| rs62052185 | TCF25 | × | √ | × | | rs7960397 | POLR1A | √ | × | × | | rs1078549 | TMEM117 | √ | × | × | | rs1057486 | TMEM117 | √ | × | |
| rs75070904 | TCF25 | × | √ | × | | rs5891929 | POLR1A | √ | × | × | | rs1073601 | TMEM117 | √ | × | × | | rs1623829 | TMEM117 | √ | × | |
| rs116083726 | TCF25 | × | √ | × | | rs1173386 | POLR1A | √ | × | × | | rs2407788 | TMEM117 | √ | × | × | | rs1731441 | TMEM117 | √ | × | |
| rs4785736 | TCF25 | × | √ | × | Red vs. brown/black hair c | rs1421536 | PON3 | √ | × | × | | rs7301406 | TMEM117 | √ | × | × | | rs1675775 | TMEM117 | √ | × | |
| rs8044906 | TCF25 | × | √ | × | | rs7301689 | POP4 | √ | × | × | | rs1230150 | TMEM117 | √ | × | × | | rs1731440 | TMEM117 | √ | × | |
| rs2270462 | TCF25 | × | √ | × | | rs1205321 | PPP1R7 | √ | × | × | | rs6582494 | TMEM117 | √ | × | × | | rs1731439 | TMEM117 | √ | × | |
| rs4591146 | TCF25 | × | √ | × | | rs1205322 | PPP1R7 | √ | × | × | | rs1709409 | TMEM117 | √ | × | × | | rs1675774 | TMEM117 | √ | × | |
| rs2270461 | TCF25 | × | √ | × | | rs1205280 | PPP1R7 | √ | × | × | | rs1118242 | TMEM117 | √ | × | × | | rs1402015 | TMEM117 | √ | × | |
| rs72813437 | TCF25 | × | √ | × | | rs7562408 | PPP1R7 | √ | × | × | | rs2062954 | TMEM117 | √ | × | × | | rs1461568 | TMEM117 | √ | × | |
| rs57940434 | TCF25 | × | √ | × | | rs2268902 | PPP1R7 | √ | × | × | | rs1032886 | TMEM117 | √ | × | × | | rs1731434 | TMEM117 | √ | × | |
| rs73265878 | TCF25 | × | √ | × | | rs2268904 | PPP1R7 | √ | × | × | | rs1229793 | TMEM117 | √ | × | × | | rs1675788 | TMEM117 | √ | × | |
| rs11644804 | TCF25 | × | √ | × | | rs1684342 | PPP1R7 | √ | × | × | | rs7977482 | TMEM117 | √ | × | × | | rs1406787 | TMEM117 | √ | × | |
| rs7908570 | TET1 | √ | × | × | | rs8022303 | PPP1R9A | √ | × | × | | rs7969885 | TMEM117 | √ | × | × | | rs1731422 | TMEM117 | √ | × | |
| rs77976327 | TET1 | √ | × | × | | rs7799229 | PPP1R9A | √ | × | × | | rs6697004 | TMEM117 | √ | × | × | | rs1222865 | TMEM117 | √ | × | |
| rs149886530 | TFDP2 | √ | × | × | | rs7752842 | PPP1R9A | √ | × | × | | rs4550550 | TMEM25 | × | × | √ | | rs2699992 | TMEM117 | √ | × | |
| rs13067380 | THRB | √ | × | × | | rs7322005 | PPP1R9A | √ | × | × | | rs4548530 | TMEM25 | × | × | √ | | rs1731423 | TMEM117 | √ | × | |
| rs13096529 | THRB | √ | × | × | Lymphocyte count | rs1527721 | PPP1R9A | √ | × | × | | rs4554823 | TMEM25 | × | × | √ | | rs1731433 | TMEM117 | √ | × | |
| rs13065854 | THRB | √ | × | × | Lymphocyte count | rs7322197 | PPP1R9A | √ | × | × | | rs4552223 | TMEM25 | × | × | √ | | rs1731432 | TMEM117 | √ | × | |
| rs117330227 | THRB | √ | × | × | | rs1253779 | PPP1R9A | √ | × | × | | rs4547490 | TMEM25 | × | × | √ | | rs1731431 | TMEM117 | √ | × | |
| rs851719 | THRB | √ | × | × | | rs7587029 | PPP1R9A | √ | × | × | | rs1170735 | TNFRSF19 | √ | × | × | | rs7515883 | TMEM117 | √ | × | |
| rs10886515 | TIAL1 | √ | × | × | | rs2871737 | PRELP | √ | × | × | | rs1306499 | TP63 | √ | × | × | | rs2614394 | TMEM117 | √ | × | |
| rs11199028 | TIAL1 | √ | × | × | | rs191318 | PRR5L | √ | × | × | | rs1308779 | TP63 | √ | × | × | | rs1675764 | TMEM117 | √ | × | |
| rs11199029 | TIAL1 | √ | × | × | | rs1050114 | PRR5L | √ | × | × | Hay fever a | rs3602150 | TP63 | √ | × | × | | rs1731459 | TMEM117 | √ | × | |
| rs11199030 | TIAL1 | √ | × | × | | rs5856832 | PRR5L | √ | × | × | | rs3583795 | TP63 | √ | × | × | | rs1731458 | TMEM117 | √ | × | |
| rs11816089 | TIAL1 | √ | × | × | | rs1761529 | PRR5L | √ | × | × | | rs3586717 | TP63 | √ | × | × | | rs7309883 | TMEM117 | √ | × | |
| rs3816145 | TIAL1 | √ | × | × | | rs216344 | PRSS3 | √ | × | × | | rs3681349 | TP63 | √ | × | × | | rs1731457 | TMEM117 | √ | × | |
| rs7919868 | TIAL1 | √ | × | × | | rs1177581 | PSD3 | √ | × | × | | rs1210797 | TP63 | √ | × | × | | rs1731456 | TMEM117 | √ | × | |
| rs6833997 | TLR6 | √ | × | × | | rs5978715 | PSD3 | √ | × | × | | rs1210796 | TP63 | √ | × | × | | rs1358705 | TMEM117 | √ | × | |
| rs7971487 | TMEM117 | × | √ | × | | rs5681101 | PSD3 | √ | × | × | | rs1237418 | TP63 | √ | × | × | | rs7547862 | TMEM117 | √ | × | |
| rs11182329 | TMEM117 | × | √ | × | | rs1267915 | PSD3 | √ | × | × | | rs3495182 | TP63 | √ | × | × | | rs1622590 | TMEM117 | √ | × | |
| rs10880591 | TMEM117 | × | √ | × | | rs1267641 | PSD3 | √ | × | × | | rs1309513 | TP63 | √ | × | × | | rs1614042 | TMEM117 | √ | × | |
| rs375172334 | TMEM117 | × | √ | × | | rs7358866 | PSD3 | √ | × | × | | rs7912503 | TP63 | √ | × | × | | rs1675791 | TMEM117 | √ | × | |
| rs2031056 | TMEM117 | × | √ | × | | rs8025114 | PSD3 | √ | × | × | | rs7637305 | TP63 | × | √ | × | | rs1675789 | TMEM117 | √ | × | |
| rs4499064 | TMEM117 | × | √ | × | | rs1712755 | PSD3 | √ | × | × | | rs1171533 | TP63 | × | √ | × | | rs1675787 | TMEM117 | √ | × | |
| rs10880592 | TMEM117 | × | √ | × | | rs7958611 | PSD3 | √ | × | × | | rs7583516 | TP63 | × | √ | × | | rs1675781 | TMEM117 | √ | × | |
| rs10748379 | TMEM117 | × | √ | × | | rs7844906 | PSD3 | √ | × | × | | rs7774458 | TP63 | × | √ | × | | rs1675785 | TMEM117 | √ | × | |
| rs10748380 | TMEM117 | × | √ | × | | rs7844314 | PSD3 | √ | × | × | | rs1093740 | TP63 | × | √ | × | | rs1675786 | TMEM117 | √ | × | |
| rs4238092 | TMEM117 | × | √ | × | | rs7962348 | PSD3 | √ | × | × | | rs1694262 | TPCN1 | √ | × | × | | rs4354753 | TMEM117 | √ | × | |
| rs4238093 | TMEM117 | × | √ | × | | rs7535689 | PTCD3 | √ | × | × | | rs1694263 | TPCN1 | √ | × | × | | rs7955615 | TMEM117 | √ | × | |
| rs10506232 | TMEM117 | × | √ | × | | rs7687057 | PTCD3 | √ | × | × | | rs7138630 | TPCN1 | √ | × | × | | rs7966024 | TMEM117 | √ | × | |
| rs543285921 | TMEM117 | × | √ | × | | rs5905328 | PTCD3 | √ | × | × | | rs1458366 | TPCN1 | √ | × | × | | rs1088057 | TMEM117 | √ | × | |
| rs11182331 | TMEM117 | × | √ | × | | rs6046081 | PTCD3 | √ | × | × | | rs6194358 | TPCN1 | √ | × | × | | rs1088058 | TMEM117 | √ | × | |
| rs4768055 | TMEM117 | × | √ | × | | rs7763085 | PTCD3 | √ | × | × | | rs7962969 | TPCN1 | √ | × | × | | rs4572199 | TMEM117 | √ | × | |
| rs150528285 | TMEM117 | × | √ | × | | rs7651585 | PTCD3 | √ | × | × | | rs7974974 | TPCN1 | √ | × | × | | rs1078546 | TMEM117 | √ | × | |
| rs11182332 | TMEM117 | × | √ | × | | rs7609640 | PTCD3 | √ | × | × | | rs6194359 | TPCN1 | √ | × | × | | rs1078546 | TMEM117 | √ | × | |
| rs80107150 | TMEM117 | × | √ | × | | rs7489072 | PUS7L | × | √ | × | | rs1694266 | TPCN1 | √ | × | × | | rs1169302 | TMEM117 | √ | × | |
| rs10785474 | TMEM117 | × | √ | × | | rs7954870 | PUS7L | × | √ | × | | rs7768853 | TPCN1 | √ | × | × | | rs5019975 | TMEM117 | √ | × | |
| rs11182334 | TMEM117 | × | √ | × | | rs1222968 | PUS7L | × | √ | × | | rs6194359 | TPCN1 | √ | × | × | | rs4537822 | TMEM117 | √ | × | |
| rs10785475 | TMEM117 | × | √ | × | | rs9783418 | PUS7L | × | √ | × | | rs7319282 | TPCN1 | √ | × | × | | rs4238089 | TMEM117 | √ | × | |
| rs10785476 | TMEM117 | × | √ | × | | rs1488256 | RALGPS1 | √ | × | × | | rs1694267 | TPCN1 | √ | × | × | | rs4238090 | TMEM117 | √ | × | |
| rs10785477 | TMEM117 | × | √ | × | | rs1011836 | RALGPS1 | × | √ | × | | rs6194359 | TPCN1 | √ | × | × | | rs1609548 | TMEM117 | √ | × | |
| rs1881927 | TMEM117 | × | √ | × | | rs7767172 | RALGPS1 | × | √ | × | | rs7446105 | TPCN1 | √ | × | × | | rs1609549 | TMEM117 | √ | × | |
| rs117422273 | TMEM117 | × | √ | × | | rs1465910 | RALGPS1 | × | √ | × | | rs7813312 | TPCN1 | √ | × | × | | rs1074837 | TMEM117 | √ | × | |
| rs1881926 | TMEM117 | × | √ | × | | rs7549669 | RALGPS1 | × | √ | × | | rs7505736 | TPCN1 | √ | × | × | | rs1074837 | TMEM117 | √ | × | |
| rs7974214 | TMEM117 | × | √ | × | | rs5388814 | RALGPS1 | × | √ | × | | rs7648897 | TPCN1 | √ | × | × | | rs4768544 | TMEM117 | √ | × | |
| rs137928630 | TMEM117 | × | √ | × | | rs3716176 | RALGPS1 | × | √ | × | | rs2303621 | TPCN1 | √ | × | × | Height | rs1118232 | TMEM117 | √ | × | |
| rs10785478 | TMEM117 | × | √ | × | | rs1477412 | RALGPS1 | × | √ | × | | rs6081316 | TPCN1 | √ | × | × | | rs1088058 | TMEM117 | √ | × | |
| rs10785479 | TMEM117 | × | √ | × | | rs1815801 | RALGPS1 | × | √ | × | | rs5710000 | TPCN1 | √ | × | × | | rs1088058 | TMEM117 | √ | × | |
| rs11182337 | TMEM117 | × | √ | × | | rs1449194 | RALGPS1 | × | √ | × | | rs7930567 | TPCN1 | √ | × | × | | rs1088058 | TMEM117 | √ | × | |
| rs12301015 | TMEM117 | × | √ | × | | rs3522974 | RALGPS1 | × | √ | × | | rs509063 | TRAF3IP1 | √ | × | × | | rs1078547 | TMEM117 | √ | × | |
| rs556729473 | TMEM117 | × | √ | × | | rs1169729 | RALGPS1 | × | √ | × | | rs1431289 | TRDN | √ | × | × | | rs7964465 | TMEM117 | √ | × | |
| rs539843818 | TMEM117 | × | √ | × | | rs1281155 | RALGPS1 | × | √ | × | | rs1367679 | TRDN | √ | × | × | | rs1236956 | TMEM117 | √ | × | |
| rs11182340 | TMEM117 | × | √ | × | | rs1509289 | RALGPS1 | × | √ | × | | rs9385290 | TRDN | √ | × | × | | rs1088058 | TMEM117 | √ | × | |
| rs10880598 | TMEM117 | × | √ | × | | rs1387400 | RALGPS1 | × | √ | × | | rs2217502 | TRDN | √ | × | × | | rs1118233 | TMEM117 | × | √ | |
| rs75410353 | TMEM117 | × | √ | × | | rs1281154 | RALGPS1 | × | √ | × | DNA methyl | rs1217369 | TRDN | √ | × | × | | rs5620961 | TMEM117 | × | √ | |
| rs4768546 | TMEM117 | × | √ | × | | rs1081926 | RALGPS1 | × | √ | × | | rs1367664 | TRDN | √ | × | × | | rs1839660 | TMEM117 | × | √ | |
| rs76302415 | TMEM117 | × | √ | × | | rs1430399 | RALGPS1 | × | √ | × | | rs1367666 | TRDN | √ | × | × | | rs1858127 | TMEM117 | × | √ | |
| rs10880599 | TMEM117 | × | √ | × | | rs1927220 | RALGPS1 | × | √ | × | | rs1708521 | TRDN | √ | × | × | | rs1887153 | TMEM117 | × | √ | |
| rs12303804 | TMEM117 | × | √ | × | | rs700876 | RANBP2 | √ | × | × | | rs9385296 | TRDN | √ | × | × | | rs8025137 | TMEM117 | × | √ | |
| rs116884551 | TMEM117 | × | √ | × | | rs1649330 | RANBP2 | √ | × | × | | rs9385297 | TRDN | √ | × | × | | rs7728020 | TMEM117 | × | √ | |
| rs199942605 | TMEM117 | × | √ | × | | rs1649329 | RANBP2 | √ | × | × | | rs1442170 | TRDN | √ | × | × | | rs1118234 | TMEM117 | × | √ | |
| rs10785482 | TMEM117 | × | √ | × | | rs1649328 | RANBP2 | √ | × | × | | rs9385299 | TRDN | √ | × | × | | rs7622916 | TMEM117 | × | √ | |
| rs10880600 | TMEM117 | × | √ | × | | rs5613283 | RANBP2 | √ | × | × | | rs9401668 | TRDN | √ | × | × | | rs9357253 | TMEM117 | × | √ | |
| rs10880601 | TMEM117 | × | √ | × | | rs1702052 | RANBP2 | √ | × | × | | rs9490744 | TRDN | √ | × | × | | rs1178941 | TMEM117 | × | √ | |
| rs76096911 | TMEM117 | × | √ | × | | rs1614285 | RANBP2 | √ | × | × | | rs9388238 | TRDN | √ | × | × | 3-(4-hydrox | rs7301406 | TMEM117 | × | √ | |
| rs11829716 | TMEM117 | × | √ | × | | rs1442283 | RANBP2 | √ | × | × | | rs9320932 | TRDN | √ | × | × | | rs7852144 | TMEM117 | × | √ | |
| rs73085705 | TMEM117 | × | √ | × | | rs4767135 | RBM19 | √ | × | × | | rs9490746 | TRDN | √ | × | × | | rs7408479 | TMEM117 | × | √ | |
| rs12230287 | TMEM117 | × | √ | × | | rs4766698 | RBM19 | √ | × | × | | rs6937527 | TRDN | √ | × | × | | rs7678594 | TMEM117 | × | √ | |
| rs144929759 | TMEM117 | × | √ | × | | rs7954178 | RBM19 | √ | × | × | | rs4510699 | TRDN | √ | × | × | | rs7136217 | TMEM117 | × | √ | |
| rs17121310 | TMEM117 | × | √ | × | | rs930004 | RBM19 | √ | × | × | | rs1383171 | TRIM29 | √ | × | × | | rs7328625 | TMEM117 | × | √ | |
| rs201310552 | TMEM117 | × | √ | × | | rs1527549 | RBM19 | √ | × | × | | rs7433826 | TRIM44 | √ | × | × | | rs7328625 | TMEM117 | × | √ | |
| rs67713366 | TMEM117 | × | √ | × | | rs1074480 | RBM19 | √ | × | × | | rs1171521 | TRIM44 | √ | × | × | | rs1230150 | TMEM117 | × | √ | |
| rs192326531 | TMEM117 | × | √ | × | | rs4767139 | RBM19 | √ | × | × | | rs2970326 | TRIM44 | √ | × | × | | rs1118236 | TMEM117 | × | √ | |
| rs184453727 | TMEM117 | × | √ | × | | rs964400 | RBM19 | √ | × | × | | rs7585634 | TRPC4 | √ | × | × | | rs1118237 | TMEM117 | × | √ | |
| rs7953173 | TMEM117 | × | √ | × | | rs759905 | RBM19 | √ | × | × | | rs7898354 | TRPM3 | √ | × | × | | rs7483890 | TMEM117 | × | √ | |
| rs141870508 | TMEM117 | × | √ | × | | rs879901 | RBM19 | √ | × | × | Externalizin | rs1888816 | TRPM3 | √ | × | × | | rs7675993 | TMEM117 | × | √ | |
| rs7967957 | TMEM117 | × | √ | × | | rs4767143 | RBM19 | √ | × | × | | rs7716491 | TRPM3 | √ | × | × | | rs6128547 | TMEM117 | × | √ | |
| rs10506234 | TMEM117 | × | √ | × | | rs1169700 | RBM19 | √ | × | × | | rs3568380 | TRPM3 | √ | × | × | | rs8181703 | TMEM117 | × | √ | |
| rs80010022 | TMEM117 | × | √ | × | | rs1569072 | RBM19 | √ | × | × | | rs7273175 | TRPM3 | √ | × | × | | rs2699993 | TMEM117 | × | √ | |
| rs55782485 | TMEM117 | × | √ | × | | rs1074480 | RBM19 | √ | × | × | | rs7705522 | TRPM3 | √ | × | × | | rs7883670 | TMEM117 | × | √ | |
| rs55977905 | TMEM117 | × | √ | × | | rs7965253 | RBM19 | √ | × | × | General risk | rs7499643 | TRPM3 | √ | × | × | | rs1222869 | TMEM117 | × | √ | |
| rs7968777 | TMEM117 | × | √ | × | | rs2049624 | RBM19 | √ | × | × | | rs7800196 | TRPM3 | √ | × | × | | rs5311474 | TMEM117 | × | √ | |
| rs1168078 | TMEM117 | × | √ | × | | rs3782448 | RBM19 | √ | × | × | | rs1171699 | TRPM3 | √ | × | × | | rs5870310 | TMEM117 | × | √ | |
| rs7980383 | TMEM117 | × | √ | × | | rs4625522 | RBM19 | √ | × | × | | rs1168529 | TRPM3 | √ | × | × | | rs1492121 | TMEM117 | × | √ | |
| rs6582498 | TMEM117 | × | √ | × | | rs4767161 | RBM19 | √ | × | × | | rs1179601 | TRPM3 | √ | × | × | | rs6582494 | TMEM117 | × | √ | |
| rs11613902 | TMEM117 | × | √ | × | | rs4767164 | RBM19 | √ | × | × | | rs7949460 | TRPM3 | √ | × | × | | rs6582495 | TMEM117 | × | √ | |
| rs2638855 | TMEM117 | × | √ | × | | rs1458713 | RBM19 | √ | × | × | | rs1705618 | TRPM3 | √ | × | × | | rs7668063 | TMEM117 | × | √ | |
| rs567055666 | TMEM117 | × | √ | × | | rs1473403 | RBM19 | √ | × | × | | rs1393895 | TRPM3 | √ | × | × | | rs1320919 | TMEM117 | × | √ | |
| rs76317381 | TMEM117 | × | √ | × | | rs1478870 | RBM44 | × | √ | × | | rs1170982 | TRPM3 | √ | × | × | | rs3766401 | TMEM117 | × | √ | |
| rs77321351 | TMEM117 | × | √ | × | | rs7308678 | RBM44 | × | √ | × | | rs1200574 | TRPM3 | √ | × | × | | rs3751158 | TMEM117 | × | √ | |
| rs78459717 | TMEM117 | × | √ | × | | rs6219614 | RBM44 | × | √ | × | | rs1255296 | TRPM3 | √ | × | × | | rs3747639 | TMEM117 | × | √ | |
| rs73288040 | TMEM117 | × | √ | × | | rs6219614 | RBM44 | × | √ | × | | rs1173837 | TRPM3 | √ | × | × | | rs2638849 | TMEM117 | × | √ | |
| rs141576644 | TMEM117 | × | √ | × | | rs7511085 | RBM44 | × | √ | × | | rs1177879 | TRPM3 | √ | × | × | | rs1118237 | TMEM117 | × | √ | |
| rs371823542 | TMEM117 | × | √ | × | | rs1190002 | RBM44 | × | √ | × | | rs7942953 | TRPM3 | √ | × | × | | rs1074839 | TMEM117 | × | √ | |
| rs11837611 | TMEM117 | × | √ | × | | rs1198819 | RBM44 | × | √ | × | | rs7892280 | TRPM3 | √ | × | × | | rs1118238 | TMEM117 | × | √ | |
| rs11182419 | TMEM117 | × | √ | × | | rs1866982 | RBM44 | × | √ | × | | rs1411164 | TRPM3 | √ | × | × | | rs1118238 | TMEM117 | × | √ | |
| rs139217151 | TMEM117 | × | √ | × | | rs4533605 | RBMS3 | √ | × | × | | rs1752075 | TRPM3 | √ | × | × | | rs1410530 | TMEM117 | × | √ | |
| rs75048928 | TMEM117 | √ | × | × | | rs9810228 | RBMS3 | √ | × | × | | rs1255192 | TRPM3 | √ | × | × | | rs5554501 | TMEM117 | × | √ | |
| rs145829681 | TMEM117 | √ | × | × | | rs6765275 | RBMS3 | √ | × | × | | rs7848436 | TRPM3 | √ | × | × | | rs7328626 | TMEM117 | × | √ | |
| rs117181959 | TMEM117 | √ | × | × | | rs2072693 | RHCG | √ | × | × | Number of c | rs7532162 | TRPM3 | √ | × | × | | rs1232216 | TMEM117 | × | √ | |
| rs114654422 | TMEM229B | √ | × | × | | rs2075833 | RHCG | √ | × | × | | rs7150591 | TRPM3 | √ | × | × | | rs1118238 | TMEM117 | × | √ | |
| rs544516878 | TNFRSF19 | √ | × | × | | rs1371386 | RHCG | √ | × | × | | rs1705621 | TRPM3 | √ | × | × | | rs1223140 | TMEM117 | × | √ | |
| rs1182 | TOR1A | √ | × | × | Prostate cancer | rs1780785 | RHCG | √ | × | × | | rs1255497 | TRPM3 | √ | × | × | | rs859699 | TMEM117 | × | √ | |
| rs11787741 | TOR1A | √ | × | × | | rs893720 | RHCG | √ | × | × | | rs1255500 | TRPM3 | √ | × | × | | rs3694832 | TMEM117 | × | √ | |
| rs3780696 | TOR1A | √ | × | × | | rs7164075 | RHCG | √ | × | × | | rs3611465 | TRPM3 | √ | × | × | | rs842194 | TMEM117 | × | √ | |
| rs3780697 | TOR1A | √ | × | × | | rs4670805 | RMDN2 | √ | × | × | | rs7458707 | TRPM3 | √ | × | × | | rs1503774 | TMEM117 | × | √ | |
| rs3780698 | TOR1A | √ | × | × | | rs7613838 | RNF112 | × | √ | × | | rs7605983 | TRPM3 | √ | × | × | | rs842193 | TMEM117 | × | √ | |
| rs13297609 | TOR1A | √ | × | × | | rs2072121 | RNF112 | × | √ | × | | rs1255162 | TRPM3 | √ | × | × | | rs7308768 | TMEM117 | × | √ | X-12013 levels |
| rs55946414 | TOR1A | √ | × | × | | rs2072120 | RNF112 | × | √ | × | | rs7903306 | TSG1 | √ | × | × | | rs1701083 | TMEM117 | × | √ | |
| rs2296793 | TOR1A | √ | × | × | Bone mineral density (hip) | rs2072119 | RNF112 | × | √ | × | | rs7411675 | UAP1 | × | √ | × | | rs1118238 | TMEM117 | × | √ | |
| rs10988519 | TOR1B | √ | × | × | | rs2072118 | RNF112 | × | √ | × | | rs1274218 | UAP1 | × | √ | × | | rs1088061 | TMEM117 | × | √ | |
| rs10116248 | TOR1B | √ | × | × | | rs9372861 | RNF146 | √ | × | × | | rs1274235 | UAP1 | × | √ | × | | rs1229685 | TMEM117 | × | √ | |
| rs2302820 | TOR1B | √ | × | × | | rs9401947 | RNF146 | √ | × | × | | rs7632078 | UAP1 | × | √ | × | | rs1403670 | TMEM117 | × | √ | |
| rs11789668 | TOR1B | √ | × | × | | rs7853136 | ROR2 | √ | × | × | | rs7432970 | UAP1 | × | √ | × | | rs1510198 | TMEM117 | × | √ | |
| rs1043186 | TOR1B | √ | × | × | | rs1011879 | ROR2 | √ | × | × | | rs7941313 | UAP1 | × | √ | × | | rs1408729 | TMEM117 | × | √ | |
| rs12634 | TOR1B | √ | × | × | Prostate cancer | rs1011881 | ROR2 | √ | × | × | | rs7543821 | UAP1 | × | √ | × | | rs1709401 | TMEM117 | × | √ | |
| rs12107025 | TP63 | √ | × | × | | rs1881391 | ROR2 | √ | × | × | | rs6695928 | UAP1 | × | √ | × | | rs7308770 | TMEM117 | × | √ | |
| rs114252996 | TP63 | √ | × | × | | rs6479387 | ROR2 | √ | × | × | | rs1181731 | UAP1 | × | √ | × | | rs2007312 | TMEM117 | × | √ | |
| rs9872452 | TP63 | √ | × | × | | rs7863167 | ROR2 | √ | × | × | | rs5959876 | UAP1 | × | √ | × | | rs7742389 | TMEM117 | × | √ | |
| rs34729753 | TP63 | √ | × | × | | rs1333668 | RSPRY1 | √ | × | × | | rs2012062 | UAP1 | × | √ | × | | rs9919676 | TMEM117 | × | √ | |
| rs4362700 | TP63 | √ | × | × | | rs4075286 | RTP5 | × | × | √ | | rs7545344 | UAP1 | × | √ | × | | rs1118239 | TMEM117 | × | √ | |
| rs9835487 | TP63 | √ | × | × | | rs7628009 | RUFY2 | √ | × | × | | rs9919292 | UAP1 | × | √ | × | | rs1118239 | TMEM117 | × | √ | |
| rs12638113 | TP63 | √ | × | × | | rs7071491 | RUFY2 | √ | × | × | | rs1213039 | UAP1 | × | √ | × | | rs2407790 | TMEM117 | × | √ | |
| rs9647365 | TP63 | √ | × | × | | rs6185732 | RUFY2 | √ | × | × | | rs1213117 | UAP1 | × | √ | × | | rs1456780 | TMEM117 | × | √ | |
| rs61943604 | TPCN1 | √ | × | × | | rs1099809 | RUFY2 | √ | × | × | | rs1686350 | UAP1 | × | √ | × | | rs1483737 | TMEM117 | × | √ | |
| rs532003322 | TPCN1 | √ | × | × | | rs7086030 | RUFY2 | √ | × | × | | rs1091756 | UAP1 | × | √ | × | | rs1231336 | TMEM117 | × | √ | |
| rs7135901 | TPCN1 | √ | × | × | | rs5367474 | SEC22C | × | √ | × | | rs7440553 | UAP1 | × | √ | × | | rs1859249 | TMEM117 | × | √ | |
| rs7300258 | TPCN1 | √ | × | × | | rs342534 | SEC22C | × | √ | × | | rs1039869 | UAP1 | × | √ | × | | rs7308972 | TMEM117 | × | √ | |
| rs7314153 | TPCN1 | √ | × | × | | rs7856677 | SEMA4D | × | √ | × | | rs1039868 | UAP1 | × | √ | × | | rs1396658 | TMEM117 | × | √ | |
| rs2303618 | TPCN1 | √ | × | × | | rs9410320 | SEMA4D | × | √ | × | | rs7999458 | UAP1 | × | √ | × | | rs1831716 | TMEM117 | × | √ | |
| rs556892903 | TPCN1 | √ | × | × | | rs7653435 | SEMA4D | × | √ | × | | rs7538393 | UAP1 | × | √ | × | | rs1512540 | TMEM117 | × | √ | |
| rs2303617 | TPCN1 | √ | × | × | | rs2865161 | SEMA4D | × | √ | × | | rs1015923 | UAP1 | × | √ | × | | rs7308973 | TMEM117 | × | √ | |
| rs7955036 | TPCN1 | √ | × | × | | rs1996423 | SIDT2 | √ | × | × | | rs7935330 | UAP1 | × | √ | × | | rs1209976 | TMEM117 | × | √ | |
| rs7955400 | TPCN1 | √ | × | × | | rs1712042 | SIDT2 | √ | × | × | | rs7928402 | UAP1 | × | √ | × | | rs2021761 | TMEM117 | × | √ | |
| rs2232515 | TPCN1 | √ | × | × | | rs579890 | SIK3 | √ | × | × | | rs3767750 | UAP1 | × | √ | × | | rs7408462 | TMEM117 | × | √ | |
| rs2288365 | TPCN1 | √ | × | × | | rs645258 | SIK3 | √ | × | × | Triglyceride | rs7779087 | UAP1 | × | √ | × | | rs1841831 | TMEM117 | × | √ | |
| rs13367 | TPCN1 | √ | × | × | | rs562179 | SIK3 | √ | × | × | | rs1091756 | UAP1 | × | √ | × | | rs1447072 | TMEM117 | × | √ | |
| rs12405 | TPCN1 | √ | × | × | | rs2513095 | SIK3 | √ | × | × | | rs5608646 | UAP1 | × | √ | × | | rs1118240 | TMEM117 | × | √ | |
| rs361508 | TRDN | √ | × | × | | rs7743116 | SIK3 | √ | × | × | | rs1256646 | UAP1 | √ | × | × | | rs1454258 | TMEM117 | × | √ | |
| rs6569329 | TRDN | √ | × | × | | rs6229348 | SKIL | √ | × | × | | rs6672561 | UAP1 | × | √ | × | | rs1118240 | TMEM117 | × | √ | |
| rs6569330 | TRDN | √ | × | × | | rs1105682 | SLC15A5 | √ | × | × | | rs7735296 | UAP1 | × | √ | × | | rs7451163 | TMEM117 | × | √ | |
| rs6920186 | TRDN | √ | × | × | | rs1117386 | SLC2A13 | √ | × | × | | rs1510267 | UAP1 | × | √ | × | | rs1118240 | TMEM117 | × | √ | |
| rs9320922 | TRDN | √ | × | × | | rs4542485 | SLC2A13 | √ | × | × | Alzheimer's | rs5627035 | UAP1 | × | √ | × | | rs866839 | TMEM117 | × | √ | |
| rs9482369 | TRDN | √ | × | × | | rs1117392 | SLC2A13 | √ | × | × | | rs7612015 | UAP1 | × | √ | × | | rs3692920 | TMEM117 | × | √ | |
| rs7754306 | TRDN | √ | × | × | | rs4296099 | SLC2A13 | √ | × | × | | rs7760200 | UAP1 | × | √ | × | | rs1118240 | TMEM117 | × | √ | |
| rs7764953 | TRDN | √ | × | × | | rs4128689 | SLC2A13 | √ | × | × | | rs7842993 | UAP1 | × | √ | × | | rs7838130 | TMEM117 | × | √ | |
| rs9385291 | TRDN | √ | × | × | | rs1077468 | SLC8B1 | √ | × | × | | rs1204066 | UAP1 | × | √ | × | | rs1709409 | TMEM117 | √ | × | |
| rs58301661 | TRDN | √ | × | × | | rs1017172 | SNED1 | × | √ | × | | rs1204073 | UAP1 | × | √ | × | | rs1118242 | TMEM117 | √ | × | |
| rs9401658 | TRDN | √ | × | × | | rs1017193 | SNED1 | × | √ | × | | rs7926288 | UAP1 | × | √ | × | | rs2062954 | TMEM117 | √ | × | |
| rs1346921 | TRDN | √ | × | × | | rs1017468 | SNED1 | × | √ | × | | rs1204075 | UAP1 | × | √ | × | | rs1032886 | TMEM117 | √ | × | |
| rs1808620 | TRDN | √ | × | × | | rs6662367 | SNIP1 | √ | × | × | | rs1204472 | UAP1 | × | √ | × | | rs1229793 | TMEM117 | √ | × | |
| rs9372743 | TRDN | √ | × | × | | rs6426023 | SNIP1 | √ | × | × | | rs1204576 | UAP1 | × | √ | × | | rs7977482 | TMEM117 | √ | × | |
| rs9375246 | TRDN | √ | × | × | | rs7139365 | SOX5 | √ | × | × | | rs7788224 | UAP1 | × | √ | × | | rs1478692 | TMEM117 | √ | × | |
| rs9385298 | TRDN | √ | × | × | | rs1104723 | SOX5 | √ | × | × | | rs1686383 | UAP1 | × | √ | × | | rs7295281 | TMEM117 | √ | × | |
| rs9401659 | TRDN | √ | × | × | | rs1044441 | SOX5 | √ | × | × | | rs1686383 | UAP1 | × | √ | × | | rs1111992 | TMEM206 | √ | × | |
| rs9372745 | TRDN | √ | × | × | | rs1498878 | SOX5 | √ | × | × | | rs1868492 | UAP1 | × | √ | × | | rs7641767 | TMEM229B | √ | × | |
| rs9388227 | TRDN | √ | × | × | | rs1498882 | SOX5 | √ | × | × | | rs1204361 | UAP1 | × | √ | × | | rs1707898 | TNFRSF19 | √ | × | |
| rs9320928 | TRDN | √ | × | × | | rs1232211 | SOX5 | √ | × | × | Brain morph | rs1209099 | UAP1 | × | √ | × | | rs5580361 | TNFRSF19 | √ | × | |
| rs9375248 | TRDN | √ | × | × | | rs1692759 | SOX5 | √ | × | × | | rs307688 | UBAP2 | √ | × | × | | rs5588226 | TNFRSF19 | √ | × | |
| rs9401661 | TRDN | √ | × | × | | rs1546436 | SOX5 | √ | × | × | | rs307648 | UBAP2 | √ | × | × | | rs2149499 | TNFRSF19 | √ | × | |
| rs9320929 | TRDN | √ | × | × | | rs1104743 | SOX5 | √ | × | × | | rs1760820 | UBAP2 | √ | × | × | | rs3794345 | TNFRSF19 | √ | × | |
| rs2060063 | TRDN | √ | × | × | | rs6191041 | SOX5 | √ | × | × | | rs1543605 | UBAP2 | √ | × | × | | rs3751363 | TNFRSF19 | √ | × | |
| rs9401665 | TRDN | √ | × | × | | rs1104745 | SOX5 | √ | × | × | | rs7382517 | UBE2E1 | √ | × | × | | rs913561 | TNFRSF19 | √ | × | |
| rs9401666 | TRDN | √ | × | × | | rs4465447 | SOX5 | √ | × | × | | rs7303355 | UBE2E2 | √ | × | × | interferon-r | rs913562 | TNFRSF19 | √ | × | |
| rs9388248 | TRDN | √ | × | × | | rs4259904 | SOX5 | √ | × | × | | rs1734290 | UBE2E2 | √ | × | × | | rs4396880 | TP63 | √ | × | |
| rs9385300 | TRDN | √ | × | × | | rs7957500 | SOX5 | √ | × | × | | rs1420851 | UBE2E2 | √ | × | × | | rs4381914 | TP63 | √ | × | |
| rs4460222 | TRDN | √ | × | × | | rs7957437 | SOX5 | √ | × | × | | rs7303569 | UBE2E2 | √ | × | × | | rs4396881 | TP63 | √ | × | |
| rs7751903 | TRDN | √ | × | × | | rs2418176 | SOX5 | √ | × | × | | rs7303772 | UBE2E2 | √ | × | × | | rs4327365 | TP63 | √ | × | |
| rs6941565 | TRDN | √ | × | × | | rs1104746 | SOX5 | √ | × | × | | rs3737950 | UBE2E2 | √ | × | × | | rs1332562 | TP63 | √ | × | |
| rs35370772 | TRDN | √ | × | × | | rs4963760 | SOX5 | √ | × | × | | rs1404070 | UBR4 | √ | × | × | | rs1920248 | TP63 | √ | × | |
| rs17739564 | TRDN | √ | × | × | | rs1230916 | SOX5 | √ | × | × | | rs1460421 | UHMK1 | √ | × | × | | rs6229170 | TP63 | √ | × | |

| | | | | |
|---|---|---|---|---|
| rs62420492 | TRDN | √ | × | × |
| rs62420493 | TRDN | √ | × | × |
| rs34957028 | TRDN | √ | × | × |
| rs6922718 | TRDN | √ | × | × |
| rs9375255 | TRDN | √ | × | × |
| rs9372748 | TRDN | √ | × | × |
| rs9375256 | TRDN | √ | × | × |
| rs4576276 | TRDN | √ | × | × |
| rs2185160 | TRDN | √ | × | × |
| rs9375257 | TRDN | √ | × | × |
| rs9320940 | TRDN | √ | × | × |
| rs9490789 | TRDN | √ | × | × |
| rs9490790 | TRDN | √ | × | × |
| rs9490791 | TRDN | √ | × | × |
| rs9490792 | TRDN | √ | × | × |
| rs9398730 | TRDN | √ | × | × |
| rs9398731 | TRDN | √ | × | × |
| rs9388251 | TRDN | √ | × | × |
| rs4509143 | TRDN | √ | × | × |
| rs4509144 | TRDN | √ | × | × |
| rs77566980 | TRDN | √ | × | × |
| rs9385301 | TRDN | √ | × | × |
| rs7746262 | TRDN | √ | × | × |
| rs7746287 | TRDN | √ | × | × |
| rs75910619 | TRDN | √ | × | × |
| rs9320941 | TRDN | √ | × | × |
| rs4370389 | TRDN | √ | × | × |
| rs4523105 | TRDN | √ | × | × |
| rs9401678 | TRDN | √ | × | × |
| rs7452290 | TRDN | √ | × | × |
| rs9398732 | TRDN | √ | × | × |
| rs9401679 | TRDN | √ | × | × |
| rs75584284 | TRDN | √ | × | × |
| rs9375259 | TRDN | √ | × | × |
| rs78585257 | TRDN | √ | × | × |
| rs79085951 | TRDN | √ | × | × |
| rs12528126 | TRDN | √ | × | × |
| rs9375260 | TRDN | √ | × | × |
| rs4897260 | TRDN | √ | × | × |
| rs4612191 | TRDN | √ | × | × |
| rs9490794 | TRDN | √ | × | × |
| rs78336591 | TRDN | √ | × | × |
| rs9375261 | TRDN | √ | × | × |
| rs12055872 | TRDN | √ | × | × |
| rs75164591 | TRDN | √ | × | × |
| rs79490787 | TRDN | √ | × | × |
| rs9398733 | TRDN | √ | × | × |
| rs75083890 | TRDN | √ | × | × |
| rs4391277 | TRDN | √ | × | × |
| rs6929960 | TRDN | √ | × | × |
| rs4998058 | TRDN | √ | × | × |
| rs11154175 | TRDN | √ | × | × |
| rs80019951 | TRDN | √ | × | × |
| rs13193333 | TRDN | √ | × | × |
| rs79789164 | TRDN | √ | × | × |
| rs12192602 | TRDN | √ | × | × |
| rs9401680 | TRDN | √ | × | × |
| rs6939909 | TRDN | √ | × | × |
| rs75126813 | TRDN | √ | × | × |
| rs4996638 | TRDN | √ | × | × |
| rs620602 | TRIM29 | √ | × | × |
| rs963811 | TRIM44 | √ | × | × |
| rs1830816 | TRIM44 | √ | × | × |
| rs2970323 | TRIM44 | √ | × | × |
| rs1007591 | TRIM44 | √ | × | × |
| rs2985418 | TRIM44 | √ | × | × |
| rs146102469 | TRIM44 | √ | × | × |
| rs140099563 | TRIM44 | √ | × | × |
| rs147357459 | TRIM44 | √ | × | × |
| rs12226318 | TRIM44 | √ | × | × |
| rs7129170 | TRIM44 | √ | × | × |
| rs12574811 | TRIM44 | √ | × | × |
| rs117495305 | TRIM44 | √ | × | × |
| rs1323856 | TRIM44 | √ | × | × |
| rs1359181 | TRIM44 | √ | × | × |
| rs74716568 | TRIM44 | √ | × | × |
| rs34103243 | TRIM44 | √ | × | × |
| rs1969800 | TRIM44 | √ | × | × |
| rs61881279 | TRIM44 | √ | × | × |
| rs60608746 | TRIM44 | √ | × | × |
| rs2985152 | TRIM44 | √ | × | × |
| rs2970329 | TRIM44 | √ | × | × |
| rs7929977 | TRIM44 | √ | × | × |
| rs16927901 | TRIM44 | √ | × | × |
| rs2970330 | TRIM44 | √ | × | × |
| rs68063352 | TRIM44 | √ | × | × |
| rs80330422 | TRIM44 | √ | × | × |
| rs11512235 | TRIM44 | √ | × | × |
| rs187351439 | TRIM44 | √ | × | × |
| rs12289152 | TRIM44 | √ | × | × |
| rs12363437 | TRIM44 | √ | × | × |
| rs12629685 | TRIM71 | √ | × | × |
| rs10868904 | TRPM3 | √ | × | × |
| rs1337030 | TRPM3 | √ | × | × |
| rs557767963 | TRPM3 | √ | × | × |
| rs6560160 | TRPM3 | √ | × | × |
| rs6560161 | TRPM3 | √ | × | × |
| rs10124944 | TRPM3 | √ | × | × |
| rs10746856 | TRPM3 | √ | × | × |
| rs1337028 | TRPM3 | √ | × | × |
| rs6560162 | TRPM3 | √ | × | × |
| rs7862440 | TRPM3 | √ | × | × |
| rs10868907 | TRPM3 | √ | × | × |
| rs7847667 | TRPM3 | √ | × | × |
| rs7862799 | TRPM3 | √ | × | × |
| rs7847940 | TRPM3 | √ | × | × |
| rs7863077 | TRPM3 | √ | × | × |
| rs7022926 | TRPM3 | √ | × | × |
| rs10868910 | TRPM3 | √ | × | × |
| rs184249596 | TRPM3 | √ | × | × |
| rs140508855 | TRPM3 | × | √ | × |
| rs112098535 | TRPM3 | × | √ | × |
| rs552181741 | TRPM3 | × | √ | × |
| rs62543183 | TRPM3 | × | √ | × |
| rs79489598 | TRPM3 | × | √ | × |
| rs17056015 | TRPM3 | × | √ | × |
| rs2993024 | TRPM3 | × | √ | × |
| rs1337011 | TRPM3 | × | √ | × |
| rs56226184 | TRPM3 | × | √ | × |
| rs75858509 | TRPM3 | × | √ | × |
| rs1337012 | TRPM3 | × | √ | × |
| rs181639753 | TRPM3 | × | √ | × |
| rs10120894 | TRPM3 | × | √ | × |
| rs11142645 | TRPM3 | × | √ | × |
| rs142637847 | TRPM3 | × | √ | × |
| rs11142648 | TRPM3 | × | √ | × |
| rs13290786 | TRPM3 | × | √ | × |
| rs1337018 | TRPM3 | × | √ | × |
| rs1421155 | TRPM3 | √ | × | × |
| rs6560172 | TRPM3 | √ | × | × |
| rs615507 | TRPM3 | √ | × | × |
| rs72614629 | TRPM3 | √ | × | × |
| rs72731759 | TRPM3 | √ | × | × |
| rs117456609 | TUBB3 | × | √ | × |
| rs541744245 | TUBB3 | × | √ | × |
| rs182442759 | TUBB3 | × | √ | × |
| rs73275939 | TUBB3 | × | √ | × |
| rs11863403 | TUBB3 | × | √ | × |
| rs114867959 | TUBB3 | × | √ | × |
| rs59001570 | TUBB3 | × | √ | × |
| rs114566768 | TUBB3 | × | √ | × |
| rs57807714 | TUBB3 | × | √ | × |
| rs58336812 | TUBB3 | × | √ | × |
| rs28521461 | TUBB3 | × | √ | × |
| rs2302898 | TUBB3 | × | √ | × |
| rs370197198 | TUBB3 | × | √ | × |
| rs144707184 | TUBB3 | × | √ | × |
| rs148513073 | TUBB3 | × | √ | × |
| rs185447857 | TUBB3 | × | √ | × |
| rs75635769 | TUBB3 | × | √ | × |
| rs201856705 | TUBB3 | × | √ | × |
| rs199842533 | TUBB3 | × | √ | × |
| rs371662941 | TUBB3 | × | √ | × |
| rs1135425 | TUBB3 | × | √ | × |
| rs76996117 | UAP1 | √ | × | × |
| rs62555936 | UBAP2 | √ | × | × |
| rs16935299 | UBAP2 | √ | × | × |
| rs3739690 | UBAP2 | √ | × | × |
| rs62555940 | UBAP2 | √ | × | × |
| rs307697 | UBAP2 | √ | × | × |
| rs72727393 | UBAP2 | √ | × | × |
| rs12553803 | UBAP2 | √ | × | × |
| rs10971836 | UBAP2 | √ | × | × |
| rs10971837 | UBAP2 | √ | × | × |
| rs10758235 | UBAP2 | √ | × | × |
| rs56404908 | UBAP2 | √ | × | × |
| rs7044541 | UBAP2 | √ | × | × |
| rs7030063 | UBAP2 | √ | × | × |
| rs932969 | UBAP2 | √ | × | × |
| rs10122409 | UBAP2 | √ | × | × |
| rs10814059 | UBAP2 | √ | × | × |
| rs140606057 | UBAP2 | √ | × | × |
| rs7029112 | UBAP2 | √ | × | × |
| rs7029122 | UBAP2 | √ | × | × |
| rs10758236 | UBAP2 | √ | × | × |
| rs13300356 | UBAP2 | √ | × | × |
| rs1785506 | UBAP2 | √ | × | × |
| rs12236865 | UBAP2 | √ | × | × |
| rs1785503 | UBAP2 | √ | × | × |
| rs1785502 | UBAP2 | √ | × | × |
| rs1785500 | UBAP2 | √ | × | × |
| rs1758632 | UBAP2 | √ | × | × |
| rs1785512 | UBAP2 | √ | × | × |
| rs10448233 | UBAP2 | √ | × | × |
| rs1758633 | UBAP2 | √ | × | × |
| rs1785509 | UBAP2 | √ | × | × |
| rs1758629 | UBAP2 | √ | × | × |
| rs1760816 | UBAP2 | √ | × | × |
| rs1629188 | UBAP2 | √ | × | × |
| rs1785507 | UBAP2 | √ | × | × |
| rs10814062 | UBAP2 | √ | × | × |
| rs1471060 | UBE2E1 | √ | × | × |
| rs11707828 | UBE2E1 | √ | × | × |
| rs6771166 | UBE2E1 | √ | × | × |
| rs6789735 | UBE2E1 | √ | × | × |
| rs6784101 | UBE2E1 | √ | × | × |
| rs1026407 | UBE2E1 | √ | × | × |
| rs1471061 | UBE2E1 | √ | × | × |
| rs34919006 | UBE2E2 | √ | × | × |
| rs9836787 | UBE2E2 | √ | × | × |
| rs17012869 | UBE2E2 | √ | × | × |
| rs12330262 | UBE2E2 | √ | × | × |
| rs12330479 | UBE2E2 | √ | × | × |
| rs9809176 | UBE2E2 | √ | × | × |
| rs4858484 | UBE2E2 | √ | × | × |
| rs9841944 | UBE2E2 | √ | × | × |
| rs12634773 | UBE2E2 | √ | × | × |
| rs6550748 | UBE2E2 | √ | × | × |
| rs1973400 | UBE2E2 | √ | × | × |
| rs11129113 | UBE2E2 | √ | × | × |
| rs9847027 | UBE2E2 | √ | × | × |
| rs6804123 | UBE2E2 | √ | × | × |
| rs4858491 | UBE2E2 | √ | × | × |
| rs9874822 | UBE2E2 | √ | × | × |
| rs9874082 | UBE2E2 | √ | × | × |
| rs80214639 | UBE2E2 | √ | × | × |
| rs4858496 | UBE2E2 | √ | × | × |
| rs80144523 | UBE2E2 | √ | × | × |
| rs9853489 | UBE2E2 | √ | × | × |
| rs4858497 | UBE2E2 | √ | × | × |
| rs11129116 | UBE2E2 | √ | × | × |
| rs142245529 | UBE2E2 | √ | × | × |
| rs9847958 | UBE2E2 | √ | × | × |
| rs1495131 | UBE2E2 | √ | × | × |
| rs921095 | UBE2F | √ | × | × |
| rs62194510 | UBE2F | √ | × | × |
| rs7525201 | UBR4 | √ | × | × |
| rs4657199 | UHMK1 | √ | × | × |
| rs6687743 | UHMK1 | √ | × | × |
| rs10919551 | UHMK1 | √ | × | × |
| rs6676358 | UHMK1 | √ | × | × |
| rs12079305 | UHMK1 | √ | × | × |
| rs7513662 | UHMK1 | √ | × | × |
| rs150122 | ULK2 | √ | × | × |
| rs157408 | ULK2 | √ | × | × |
| rs157401 | ULK2 | √ | × | × |
| rs157402 | ULK2 | √ | × | × |
| rs157403 | ULK2 | √ | × | × |
| rs169460 | ULK2 | √ | × | × |

| | | | | | |
|---|---|---|---|---|---|
| | rs7961598 | SOX5 | √ | × | × |
| | rs4963765 | SOX5 | √ | × | × |
| | rs6487389 | SOX5 | √ | × | × |
| | rs9630247 | SOX5 | √ | × | × |
| | rs1084236 | SOX5 | √ | × | × |
| | rs4131686 | SOX5 | √ | × | × |
| | rs4131685 | SOX5 | √ | × | × |
| | rs4963766 | SOX5 | √ | × | × |
| | rs4963767 | SOX5 | √ | × | × |
| | rs4963768 | SOX5 | √ | × | × |
| | rs3110018 | SOX5 | √ | × | × |
| | rs3112134 | SOX5 | √ | × | × |
| | rs3112135 | SOX5 | √ | × | × |
| | rs3112136 | SOX5 | √ | × | × |
| | rs3112137 | SOX5 | √ | × | × |
| | rs7295669 | SOX5 | √ | × | × |
| | rs10842369 | SOX5 | √ | × | × |
| | rs11047492 | SOX5 | √ | × | × |
| | rs7132960 | SOX5 | √ | × | × |
| | rs16927697 | SOX5 | √ | × | × |
| | rs16927698 | SOX5 | √ | × | × |
| | rs7308618 | SOX5 | √ | × | × |
| | rs11833062 | SOX5 | √ | × | × |
| | rs7974269 | SOX5 | √ | × | × |
| | rs4963575 | SOX5 | √ | × | × |
| | rs7132989 | SOX5 | √ | × | × |
| | rs2242115 | SPOCK3 | √ | × | × |
| | rs17519417 | SPOCK3 | √ | × | × |
| | rs7694002 | SPOCK3 | √ | × | × |
| | rs7678726 | SPOCK3 | √ | × | × |
| | rs11929824 | SPOCK3 | √ | × | × |
| | rs11929833 | SPOCK3 | √ | × | × |
| | rs80336024 | SPOCK3 | √ | × | × |
| | rs28626960 | SPOCK3 | √ | × | × |
| | rs28433769 | SPOCK3 | √ | × | × |
| | rs14631027 | SPRTN | × | √ | × |
| | rs2474632 | SPRTN | × | √ | × |
| | rs2295420 | SPRTN | × | √ | × |
| | rs2250749 | SPRTN | × | √ | × |
| | rs18617547 | SPRTN | × | √ | × |
| | rs18676945 | SPTAN1 | √ | × | × |
| | rs79626263 | SPTAN1 | √ | × | × |
| | rs3750332 | SPTAN1 | × | √ | × |
| | rs10992207 | SPTLC1 | √ | × | × |
| | rs78624938 | SPTLC1 | √ | × | × |
| | rs80297501 | SPTLC1 | √ | × | × |
| | rs75764672 | SPTLC1 | √ | × | × |
| | rs6479405 | SPTLC1 | √ | × | × |
| | rs2297568 | SPTLC1 | √ | × | × |
| | rs6547634 | ST3GAL5 | √ | × | × |
| | rs4553826 | ST3GAL5 | √ | × | × |
| | rs4832015 | ST3GAL5 | √ | × | × |
| | rs6734488 | ST3GAL5 | √ | × | × |
| | rs6721125 | ST3GAL5 | √ | × | × |
| | rs4832213 | ST3GAL5 | √ | × | × |
| | rs2367789 | SYNPR | √ | × | × |
| | rs1916628 | SYNPR | √ | × | × |
| | rs11130921 | SYNPR | √ | × | × |
| | rs1523441 | SYNPR | √ | × | × |
| | rs13080581 | SYNPR | √ | × | × |
| | rs1874927 | SYNPR | √ | × | × |
| | rs949610 | SYNPR | √ | × | × |
| | rs10866065 | SYNPR | √ | × | × |
| | rs1523437 | SYNPR | √ | × | × |
| | rs36093116 | SYNPR | √ | × | × |
| | rs12489378 | SYNPR | √ | × | × |
| | rs13096800 | SYNPR | √ | × | × |
| | rs1376482 | SYNPR | √ | × | × |
| | rs1376481 | SYNPR | √ | × | × |
| | rs4688166 | SYNPR | √ | × | × |
| Eicosenoylcarnitine (C20:1 | rs1357522 | SYNPR | √ | × | × |
| | rs4234675 | SYNPR | √ | × | × |
| | rs2167354 | SYNPR | × | √ | × |
| | rs2367792 | SYNPR | × | √ | × |
| | rs35197431 | SYNPR | × | √ | × |
| | rs14606132 | SYNPR | × | √ | × |
| | rs62251360 | SYNPR | × | √ | × |
| | rs20126728 | SYNPR | × | √ | × |
| | rs15065651 | SYNPR | × | √ | × |
| | rs10510895 | SYNPR | × | √ | × |
| | rs56308794 | SYNPR | × | √ | × |
| | rs55707471 | SYNPR | × | √ | × |
| | rs11720996 | SYNPR | × | √ | × |
| | rs12488438 | SYNPR | × | √ | × |
| | rs62251362 | SYNPR | × | √ | × |
| | rs73108930 | SYNPR | × | √ | × |
| | rs73108933 | SYNPR | × | √ | × |
| | rs11719344 | SYNPR | × | √ | × |
| | rs66667790 | SYNPR | × | √ | × |
| | rs14189939 | SYNPR | × | √ | × |
| | rs1523439 | SYNPR | × | √ | × |
| | rs75351562 | SYNPR | × | √ | × |
| | rs11705961 | SYNPR | × | √ | × |
| | rs56047710 | SYNPR | × | √ | × |
| | rs13060890 | SYNPR | × | √ | × |
| | rs13060806 | SYNPR | × | √ | × |
| | rs1901302 | SYNPR | × | √ | × |
| | rs13061144 | SYNPR | × | √ | × |
| | rs1916627 | SYNPR | × | √ | × |
| | rs1451305 | SYNPR | × | √ | × |
| | rs77909194 | SYNPR | × | √ | × |
| | rs7635844 | SYNPR | × | √ | × |
| | rs35245523 | SYNPR | × | √ | × |
| | rs14533976 | SYNPR | × | √ | × |
| | rs17068288 | SYNPR | × | √ | × |
| | rs13093115 | SYNPR | × | √ | × |
| | rs17068400 | SYNPR | × | √ | × |
| | rs911788 | TACC2 | × | √ | × |
| | rs53667215 | TACC2 | × | √ | × |
| | rs911772 | TACC2 | × | √ | × |
| | rs15061696 | TACC2 | √ | × | × |
| | rs18751511 | TBC1D1 | √ | × | × |
| | rs1344603 | TBC1D1 | × | √ | × |
| | rs17579988 | TBC1D1 | √ | × | × |
| | rs78273921 | TBC1D13 | √ | × | × |
| | rs11794069 | TBC1D13 | √ | × | × |
| | rs11727524 | TBC1D32 | √ | × | × |
| | rs75744292 | TBC1D32 | √ | × | × |
| | rs1273770 | TBC1D32 | √ | × | × |
| | rs73524575 | TBC1D32 | √ | × | × |
| | rs17083281 | TBC1D32 | √ | × | × |
| | rs73524585 | TBC1D32 | √ | × | × |
| | rs9490141 | TBC1D32 | √ | × | × |
| | rs78621169 | TBC1D32 | √ | × | × |
| | rs11379266 | TBC1D32 | √ | × | × |
| | rs73526653 | TBC1D32 | √ | × | × |
| | rs74960643 | TBC1D32 | √ | × | × |
| | rs73526664 | TBC1D32 | √ | × | × |
| | rs57819930 | TBC1D32 | √ | × | × |
| | rs68122210 | TET1 | √ | × | × |
| | rs12204809 | THEMIS | × | √ | × |
| | rs76306959 | THEMIS | × | √ | × |
| | rs68018843 | THEMIS | × | √ | × |
| | rs53331311 | THEMIS | × | √ | × |
| | rs14288600 | THEMIS | × | √ | × |
| | rs18081369 | THEMIS | × | √ | × |
| | rs36754661 | THEMIS | × | √ | × |
| | rs11154427 | THEMIS | × | √ | × |
| | rs18197302 | THEMIS | × | √ | × |
| | rs72618503 | THEMIS | × | √ | × |
| | rs56802662 | THEMIS | × | √ | × |
| | rs62425513 | THEMIS | × | √ | × |
| | rs14581006 | THEMIS | × | √ | × |
| | rs72965789 | THEMIS | × | √ | × |
| Red vs. brown/black hair c | rs14232699 | THEMIS | × | √ | × |
| | rs72965790 | THEMIS | × | √ | × |
| | rs75224818 | THEMIS | × | √ | × |
| | rs37013225 | THEMIS | × | √ | × |
| | rs11752963 | THEMIS | × | √ | × |
| | rs14244433 | THEMIS | × | √ | × |
| | rs9385439 | THEMIS | × | √ | × |
| | rs11961390 | THEMIS | × | √ | × |
| | rs75827423 | THEMIS | × | √ | × |
| | rs14644689 | THEMIS | × | √ | × |
| | rs18933510 | THEMIS | × | √ | × |
| | rs12214409 | THEMIS | × | √ | × |
| | rs12205922 | THEMIS | × | √ | × |
| | rs37457694 | THEMIS | × | √ | × |
| | rs11962700 | THEMIS | × | √ | × |
| | rs54100758 | THEMIS | × | √ | × |
| | rs10872324 | THEMIS | × | √ | × |
| | rs19265201 | THEMIS | × | √ | × |
| | rs74297174 | THEMIS | × | √ | × |
| | rs14607317 | THEMIS | × | √ | × |
| | rs14416013 | THEMIS | × | √ | × |
| | rs77915772 | THRB | √ | × | × |
| | rs75214296 | TMEM117 | × | √ | × |
| | rs7965635 | TMEM117 | × | √ | × |
| | rs18943003 | TMEM117 | × | √ | × |
| | rs77952333 | TMEM117 | × | √ | × |
| | rs80099331 | TMEM117 | × | √ | × |
| | rs77708558 | TMEM117 | × | √ | × |
| | rs74373254 | TMEM117 | × | √ | × |
| | rs11741989 | TMEM117 | × | √ | × |
| | rs79425253 | TMEM117 | × | √ | × |
| | rs73096934 | TMEM117 | × | √ | × |
| | rs80098499 | TMEM117 | × | √ | × |
| | rs77161382 | TMEM117 | × | √ | × |
| | rs11806212 | TMEM117 | × | √ | × |
| | rs15057142 | TMEM117 | × | √ | × |
| | rs78388987 | TMEM117 | × | √ | × |
| Type 2 diabetes | rs76737056 | TMEM117 | × | √ | × |
| | rs1609548 | TMEM117 | √ | × | × |
| | rs1609549 | TMEM117 | √ | × | × |
| | rs4768054 | TMEM117 | × | √ | × |
| | rs7969641 | TMEM117 | × | √ | × |
| | rs11182326 | TMEM117 | × | √ | × |
| | rs75608131 | TMEM117 | × | √ | × |
| | rs11182328 | TMEM117 | × | √ | × |
| | rs14402381 | TMEM117 | × | √ | × |
| | rs11182338 | TMEM117 | × | √ | × |
| | rs56209615 | TMEM117 | × | √ | × |
| | rs18396607 | TMEM117 | × | √ | × |
| | rs18581279 | TMEM117 | × | √ | × |
| | rs18871531 | TMEM117 | × | √ | × |
| | rs80251371 | TMEM117 | × | √ | × |
| | rs77280204 | TMEM117 | × | √ | × |
| | rs11182345 | TMEM117 | × | √ | × |
| | rs6582495 | TMEM117 | × | √ | × |
| | rs76680639 | TMEM117 | × | √ | × |
| | rs1320919 | TMEM117 | × | √ | × |
| | rs37511587 | TMEM117 | × | √ | × |
| | rs37476399 | TMEM117 | × | √ | × |
| | rs2638849 | TMEM117 | × | √ | × |
| | rs11182379 | TMEM117 | × | √ | × |
| | rs10748390 | TMEM117 | × | √ | × |
| | rs11182381 | TMEM117 | × | √ | × |
| | rs11182382 | TMEM117 | × | √ | × |
| | rs14105304 | TMEM117 | × | √ | × |
| | rs55545019 | TMEM117 | × | √ | × |
| | rs73286269 | TMEM117 | × | √ | × |
| | rs12322164 | TMEM117 | × | √ | × |
| | rs11182384 | TMEM117 | × | √ | × |
| | rs12231403 | TMEM117 | × | √ | × |
| | rs859699 | TMEM117 | × | √ | × |
| | rs36948323 | TMEM117 | × | √ | × |
| Reaction time | rs842194 | TMEM117 | × | √ | × |
| | rs15037743 | TMEM117 | × | √ | × |
| | rs842193 | TMEM117 | × | √ | × |
| | rs73087684 | TMEM117 | × | √ | × |
| | rs1701083 | TMEM117 | × | √ | × |
| | rs11182385 | TMEM117 | × | √ | × |
| | rs10880616 | TMEM117 | × | √ | × |
| | rs12296857 | TMEM117 | × | √ | × |
| | rs14036701 | TMEM117 | × | √ | × |
| Educational attainment | rs15101986 | TMEM117 | × | √ | × |
| | rs14087293 | TMEM117 | × | √ | × |
| | rs17094013 | TMEM117 | × | √ | × |
| | rs73087700 | TMEM117 | × | √ | × |
| | rs20073127 | TMEM117 | × | √ | × |
| | rs77423893 | TMEM117 | × | √ | × |
| | rs9919676 | TMEM117 | × | √ | × |
| OTU99_123 (Phascolarcto | rs11182390 | TMEM117 | × | √ | × |
| | rs10880618 | TMEM117 | × | √ | × |
| | rs9919742 | TMEM117 | × | √ | × |
| | rs56227915 | TMEM117 | × | √ | × |
| | rs11182394 | TMEM117 | × | √ | × |
| | rs11182395 | TMEM117 | × | √ | × |
| | rs2407790 | TMEM117 | × | √ | × |

| | | | | | |
|---|---|---|---|---|---|
| | rs16960454 | ULK2 | √ | × | × |
| | rs16960456 | ULK2 | √ | × | × |
| | rs14208044 | UNC5C | √ | × | × |
| | rs12645399 | UNC5C | √ | × | × |
| | rs17386118 | UNC5C | √ | × | × |
| | rs17055104 | UNQ6494 | √ | × | × |
| | rs75147371 | UNQ6494 | √ | × | × |
| | rs57828282 | UNQ6494 | √ | × | × |
| | rs75569459 | UNQ6494 | √ | × | × |
| | rs79212831 | UNQ6494 | √ | × | × |
| | rs4877119 | UNQ6494 | √ | × | × |
| Smoking init | rs11265857 | UNQ6494 | √ | × | × |
| | rs7040277 | UNQ6494 | √ | × | × |
| | rs78227452 | UNQ6494 | √ | × | × |
| | rs58953801 | UNQ6494 | √ | × | × |
| | rs34625605 | UNQ6494 | √ | × | × |
| Smoking init | rs60728828 | UNQ6494 | √ | × | × |
| Smoking init | rs79808925 | UNQ6494 | √ | × | × |
| | rs10908911 | UNQ6494 | √ | × | × |
| | rs1110274 | USP36 | √ | × | × |
| Height | rs7218605 | USP36 | √ | × | × |
| | rs60530618 | UTRN | √ | × | × |
| Smoking init | rs77119056 | UTRN | √ | × | × |
| | rs17793570 | UTRN | √ | × | × |
| | rs917992 | VIPR2 | √ | × | × |
| | rs429893 | VIPR2 | √ | × | × |
| | rs61990129 | VTI1B | √ | × | × |
| | rs9566690 | WBP4 | √ | × | × |
| | rs75784070 | WWC1 | √ | × | × |
| | rs17354970 | WWC2 | √ | × | × |
| | rs72691663 | WWC2 | √ | × | × |
| | rs72691664 | WWC2 | √ | × | × |
| | rs72691674 | WWC2 | √ | × | × |
| | rs72695786 | WWC2 | √ | × | × |
| | rs72695787 | WWC2 | √ | × | × |
| | rs72695790 | WWC2 | √ | × | × |
| | rs17355571 | WWC2 | √ | × | × |
| | rs72695791 | WWC2 | √ | × | × |
| | rs72695792 | WWC2 | √ | × | × |
| | rs72695794 | WWC2 | √ | × | × |
| | rs2309805 | WWC2 | √ | × | × |
| | rs10520547 | WWC2 | √ | × | × |
| | rs74689827 | WWC2 | √ | × | × |
| | rs80172480 | WWC2 | √ | × | × |
| | rs10520546 | WWC2 | √ | × | × |
| | rs72697304 | WWC2 | √ | × | × |
| | rs75036120 | WWC2 | √ | × | × |
| | rs73872929 | WWC2 | √ | × | × |
| | rs73872930 | WWC2 | √ | × | × |
| | rs14521212 | WWC2 | √ | × | × |
| | rs37273188 | WWC2 | √ | × | × |
| | rs10520545 | WWC2 | √ | × | × |
| | rs18768357 | WWC2 | √ | × | × |
| | rs72697306 | WWC2 | √ | × | × |
| | rs72697308 | WWC2 | √ | × | × |
| | rs17090670 | WWC2 | √ | × | × |
| | rs6552641 | WWC2 | √ | × | × |
| | rs75818457 | WWC2 | √ | × | × |
| | rs6552642 | WWC2 | √ | × | × |
| | rs78551452 | WWC2 | √ | × | × |
| | rs12650037 | WWC2 | √ | × | × |
| | rs4579165 | WWC2 | √ | × | × |
| | rs13119486 | WWC2 | √ | × | × |
| | rs6552643 | WWC2 | √ | × | × |
| | rs6552644 | WWC2 | √ | × | × |
| | rs4241761 | WWC2 | √ | × | × |
| | rs4476642 | WWC2 | √ | × | × |
| | rs4582186 | WWC2 | √ | × | × |
| | rs4241762 | WWC2 | √ | × | × |
| | rs6552646 | WWC2 | √ | × | × |
| | rs6552647 | WWC2 | √ | × | × |
| | rs6552648 | WWC2 | √ | × | × |
| | rs4544745 | WWC2 | √ | × | × |
| | rs4260572 | WWC2 | √ | × | × |
| | rs19041614 | WWC2 | √ | × | × |
| | rs18181104 | WWC2 | √ | × | × |
| | rs11654411 | WWC2 | √ | × | × |
| | rs10020186 | WWC2 | √ | × | × |
| | rs11605009 | WWC2 | √ | × | × |
| | rs18798082 | WWC2 | √ | × | × |
| | rs14422589 | WWC2 | √ | × | × |
| | rs4298176 | WWC2 | √ | × | × |
| | rs4298177 | WWC2 | √ | × | × |
| | rs4450976 | WWC2 | √ | × | × |
| | rs6839568 | WWC2 | √ | × | × |
| | rs4485839 | WWC2 | √ | × | × |
| | rs4632701 | WWC2 | √ | × | × |
| | rs11410567 | WWC2 | √ | × | × |
| | rs18377143 | WWC2 | √ | × | × |
| | rs7695850 | WWC2 | √ | × | × |
| | rs4419514 | WWC2 | √ | × | × |
| | rs4490499 | WWC2 | √ | × | × |
| | rs18851868 | WWC2 | √ | × | × |
| | rs4361433 | WWC2 | √ | × | × |
| | rs4479747 | WWC2 | √ | × | × |
| | rs6552651 | WWC2 | √ | × | × |
| | rs11576475 | WWC2 | √ | × | × |
| | rs10008614 | WWC2 | √ | × | × |
| | rs6829968 | WWC2 | √ | × | × |
| | rs18410952 | WWC2 | √ | × | × |
| | rs6856333 | WWC2 | √ | × | × |
| | rs6856341 | WWC2 | √ | × | × |
| | rs37402933 | WWC2 | √ | × | × |
| | rs7664963 | WWC2 | √ | × | × |
| | rs77569159 | WWC2 | √ | × | × |
| | rs77166350 | WWC2 | √ | × | × |
| | rs75921860 | WWC2 | √ | × | × |
| | rs36782137 | WWC2 | √ | × | × |
| | rs11546717 | WWC2 | √ | × | × |
| | rs56625048 | WWC2 | √ | × | × |
| | rs4351063 | WWC2 | √ | × | × |
| | rs4512039 | WWC2 | √ | × | × |
| Lymphocyte | rs4546292 | WWC2 | √ | × | × |
| | rs75380153 | WWC2 | √ | × | × |
| | rs37662338 | WWC2 | √ | × | × |
| | rs76373458 | WWC2 | √ | × | × |
| | rs37274779 | WWC2 | √ | × | × |
| | rs4639121 | WWC2 | √ | × | × |
| | rs4639122 | WWC2 | √ | × | × |
| | rs18780668 | WWC2 | √ | × | × |
| | rs37177947 | WWC2 | √ | × | × |
| | rs74337402 | WWC2 | √ | × | × |
| | rs3924948 | WWC2 | √ | × | × |
| | rs37095005 | WWC2 | √ | × | × |
| | rs37280018 | WWC2 | √ | × | × |
| | rs73870369 | WWC2 | √ | × | × |
| | rs77613838 | WWC2 | √ | × | × |
| | rs6855204 | WWC2 | √ | × | × |
| | rs73870373 | WWC2 | √ | × | × |
| | rs73870374 | WWC2 | √ | × | × |
| | rs73870375 | WWC2 | √ | × | × |
| | rs37073060 | WWC2 | √ | × | × |
| | rs37518160 | WWC2 | √ | × | × |
| | rs37410613 | WWC2 | √ | × | × |
| | rs57306674 | WWC2 | √ | × | × |
| | rs37266331 | WWC2 | √ | × | × |
| | rs79023321 | WWC2 | √ | × | × |
| | rs75377999 | WWC2 | √ | × | × |
| | rs73870380 | WWC2 | √ | × | × |
| | rs1397943 | WWC2 | √ | × | × |
| | rs75558259 | WWC2 | √ | × | × |
| Uterine fibro | rs884387 | WWC2 | √ | × | × |
| | rs73870385 | WWC2 | √ | × | × |
| | rs73870387 | WWC2 | √ | × | × |
| | rs7666191 | WWC2 | √ | × | × |
| | rs12503546 | WWC2 | √ | × | × |
| | rs55631953 | WWC2 | √ | × | × |
| | rs11734558 | WWC2 | √ | × | × |
| | rs62357979 | WWC2 | √ | × | × |
| | rs71620986 | WWC2 | √ | × | × |
| | rs10003798 | WWC2 | √ | × | × |
| | rs6849819 | WWC2 | √ | × | × |
| | rs35306878 | WWC2 | √ | × | × |
| | rs4501261 | WWC2 | √ | × | × |
| | rs35338018 | WWC2 | √ | × | × |
| | rs58876449 | ZFHX3 | √ | × | × |
| | rs14992789 | ZFHX3 | √ | × | × |
| | rs55989215 | ZFHX3 | √ | × | × |
| | rs11860416 | ZFHX3 | √ | × | × |
| | rs74030210 | ZFHX3 | √ | × | × |
| | rs28688024 | ZFHX3 | √ | × | × |
| | rs76438982 | ZHX2 | √ | × | × |
| | rs73622634 | ZNF335 | √ | × | × |
| | rs3746504 | ZNF335 | √ | × | × |
| | rs13040315 | ZNF335 | √ | × | × |
| | rs34293711 | ZNF335 | √ | × | × |
| | rs7873021 | ZNF484 | √ | × | × |
| | rs1323276 | ZNF484 | √ | × | × |
| | rs12338961 | ZNF484 | √ | × | × |
| | rs8039263 | ZNF774 | √ | × | × |
| | rs2601147 | ZNF774 | √ | × | × |
| | rs75898725 | ZNF787 | √ | × | × |
| | rs59924464 | ZNF787 | √ | × | × |
| X-12013 levels | | | | | |

| | | | | | |
|---|---|---|---|---|---|
| | rs10937404 | TP63 | √ | × | |
| | rs10937405 | TP63 | √ | × | Lung adenocarcinoma |
| | rs14428540 | TPCN1 | √ | × | |
| | rs13398327 | TRAF3IP1 | √ | × | |
| | rs9784023 | TRAF3IP1 | √ | × | |
| | rs13391057 | TRAF3IP1 | √ | × | |
| | rs13429066 | TRAF3IP1 | √ | × | |
| | rs11892233 | TRAF3IP1 | √ | × | |
| | rs11893329 | TRAF3IP1 | √ | × | |
| | rs11888517 | TRAF3IP1 | √ | × | |
| | rs7607405 | TRAF3IP1 | √ | × | |
| | rs67789994 | TRAF3IP1 | √ | × | |
| | rs9375239 | TRDN | √ | × | |
| | rs4897172 | TRDN | √ | × | |
| | rs9372741 | TRDN | √ | × | |
| | rs2873478 | TRDN | √ | × | |
| | rs4346869 | TRDN | √ | × | |
| | rs1431289 | TRDN | √ | × | |
| | rs1431290 | TRDN | √ | × | |
| | rs1346920 | TRDN | √ | × | |
| Systolic bloo | rs1431288 | TRDN | √ | × | |
| | rs1367679 | TRDN | √ | × | |
| | rs9385290 | TRDN | √ | × | |
| | rs2217502 | TRDN | √ | × | |
| | rs12173690 | TRDN | √ | × | |
| | rs979849 | TRDN | √ | × | |
| | rs1933899 | TRDN | √ | × | |
| | rs9320931 | TRDN | √ | × | |
| | rs74527900 | TRDN | √ | × | |
| | rs666432 | TRIM29 | √ | × | |
| | rs1174117 | TRIM44 | √ | × | Photic sneeze reflex |
| | rs657292 | TRIM44 | √ | × | |
| | rs14084979 | TRIM44 | √ | × | |
| | rs11033258 | TRIM44 | √ | × | |
| | rs12222279 | TRIM44 | √ | × | |
| | rs78644770 | TRIM44 | √ | × | |
| | rs66522310 | TRIM44 | √ | × | |
| | rs11033260 | TRIM44 | √ | × | |
| | rs16927870 | TRIM44 | √ | × | |
| | rs12576967 | TRIM44 | √ | × | |
| | rs77013261 | TRIM44 | √ | × | |
| | rs75667066 | TRIM44 | √ | × | |
| | rs74338266 | TRIM44 | √ | × | |
| | rs11033261 | TRIM44 | √ | × | |
| | rs7126124 | TRIM44 | √ | × | |
| | rs11694335 | TRIM44 | √ | × | |
| | rs77770625 | TRIM44 | √ | × | |
| | rs11033265 | TRIM44 | √ | × | |
| | rs67667959 | TRIM44 | √ | × | |
| | rs16927891 | TRIM44 | √ | × | |
| | rs10501141 | TRIM44 | √ | × | |
| | rs11033267 | TRIM44 | √ | × | |
| | rs10836443 | TRIM44 | √ | × | |
| | rs11033269 | TRIM44 | √ | × | |
| | rs75871465 | TRIM44 | √ | × | |
| | rs78440146 | TRPC4 | √ | × | |
| | rs3124645 | TRPM3 | √ | × | |
| | rs14044673 | TRPM3 | √ | × | |
| | rs14434533 | TRPM3 | √ | × | |
| | rs12551768 | TRPM3 | √ | × | |
| | rs57244722 | TRPM3 | √ | × | |
| | rs962001 | TRPM3 | √ | × | Metabolite levels |
| | rs35683807 | TRPM3 | √ | × | |
| | rs12552966 | TRPM3 | √ | × | |
| | rs11142687 | TRPM3 | × | √ | |
| | rs9363103 | TSG1 | √ | × | |
| | rs72727346 | UBAP2 | × | √ | |
| | rs12682795 | UBAP2 | × | √ | |
| | rs37284159 | UBAP2 | × | √ | |
| | rs307703 | UBAP2 | √ | × | Brain morphology (MOSTest) |
| | rs834027 | UBAP2 | √ | × | |
| | rs1760820 | UBAP2 | √ | × | |
| | rs2055156 | UBE2E2 | √ | × | |
| | rs2201484 | UBE2E2 | √ | × | |
| | rs17012852 | UBE2E2 | √ | × | |
| | rs75431641 | UBE2E2 | √ | × | |
| | rs2132148 | UBE2E2 | √ | × | |
| | rs11643903 | UBE2E2 | √ | × | |
| | rs76921336 | UBE2E2 | √ | × | |
| | rs17012897 | UBE2E2 | √ | × | |
| | rs13087234 | UBE2E2 | √ | × | |
| | rs77708114 | UBE2E2 | √ | × | |
| | rs17342904 | UBE2E2 | √ | × | |
| | rs34239585 | UBE2E2 | √ | × | |
| | rs11800316 | UBE2E2 | √ | × | |
| | rs34259979 | UBE2E2 | √ | × | |
| | rs17012966 | UBE2E2 | √ | × | |
| | rs14040702 | UBR4 | √ | × | |
| | rs11770659 | ULK2 | √ | × | |
| | rs16960454 | ULK2 | √ | × | |
| | rs77235493 | ULK2 | √ | × | |
| | rs16960456 | ULK2 | √ | × | |
| | rs79534009 | ULK2 | √ | × | |
| | rs77043497 | ULK2 | √ | × | |
| | rs56944681 | UNC13C | √ | × | |
| | rs17386118 | UNC5C | √ | × | |
| | rs14572837 | UNQ6494 | √ | × | |
| | rs10908911 | UNQ6494 | √ | × | |
| | rs1110274 | USP36 | √ | × | |
| | rs7218605 | USP36 | √ | × | Systolic blood pressure |
| | rs1531797 | USP36 | √ | × | |
| | rs917992 | VIPR2 | √ | × | |
| | rs73169238 | VIPR2 | √ | × | |
| | rs73169239 | VIPR2 | √ | × | |
| | rs7797488 | VIPR2 | √ | × | |
| | rs3793227 | VIPR2 | √ | × | |
| | rs73169244 | VIPR2 | √ | × | |
| | rs73169245 | VIPR2 | √ | × | |
| | rs12890397 | VTI1B | √ | × | |
| | rs61990129 | VTI1B | √ | × | |
| | rs11738795 | VTI1B | √ | × | |
| | rs1329520 | WBP4 | √ | × | |
| | rs2766726 | WBP4 | √ | × | |
| | rs9532718 | WBP4 | √ | × | Corneal resistance factor (MTAG) |
| | rs9566690 | WBP4 | √ | × | |
| | rs75450756 | WDR34 | √ | × | |
| | rs16936868 | WDR34 | √ | × | |
| | rs73523650 | WNK2 | √ | × | |
| | rs72645752 | WWC1 | √ | × | |
| | rs78442116 | WWC2 | √ | × | |
| | rs4351062 | WWC2 | × | √ | |
| | rs11930293 | WWC2 | √ | × | |
| | rs80202922 | WWC2 | √ | × | |
| | rs7677983 | WWC2 | √ | × | |
| | rs58714470 | WWC2 | √ | × | |
| | rs3749592 | WWC2 | √ | × | |
| | rs6777904 | XXYLT1 | √ | × | |
| | rs67484513 | XXYLT1 | √ | × | |
| | rs10922177 | ZBTB41 | √ | × | Complement factor H-related protein 2 levels |
| | rs10922179 | ZBTB41 | √ | × | Total cholesterol levels |
| | rs1076158 | ZFHX3 | × | √ | |
| | rs4788481 | ZFHX3 | × | √ | |
| | rs4788482 | ZFHX3 | × | √ | |
| | rs7205502 | ZFHX3 | × | √ | |
| | rs4788485 | ZFHX3 | × | √ | |
| | rs8063923 | ZFHX3 | × | √ | |
| | rs10345662 | ZFHX3 | × | √ | |
| | rs56073718 | ZFHX3 | × | √ | |
| | rs17680205 | ZFHX3 | × | √ | |
| | rs77518531 | ZFHX3 | × | √ | |
| | rs74737973 | ZFHX3 | × | √ | |
| | rs75395117 | ZFHX3 | × | √ | |
| | rs17618144 | ZFHX3 | × | √ | Externalizing behaviour (multivariate analysis) |
| | rs75272187 | ZFHX3 | × | √ | |
| | rs1858803 | ZFHX3 | × | √ | |
| | rs78925069 | ZFHX3 | × | √ | |
| | rs4788672 | ZFHX3 | × | √ | |
| | rs8047422 | ZFHX3 | × | √ | |
| | rs2359128 | ZFHX3 | × | √ | |
| | rs17680397 | ZFHX3 | × | √ | Lifetime smoking |
| | rs9923980 | ZFHX3 | × | √ | |
| | rs78711661 | ZFHX3 | × | √ | |
| | rs79111200 | ZFHX3 | × | √ | |
| | rs11797644 | ZFHX3 | × | √ | |
| | rs75847797 | ZFHX3 | × | √ | |
| | rs77043447 | ZFHX3 | × | √ | |
| | rs60315503 | ZFHX3 | × | √ | |
| | rs14675169 | ZFHX3 | × | √ | |
| | rs721824 | ZFHX3 | × | √ | |
| | rs9923193 | ZFHX3 | × | √ | |
| | rs9922718 | ZFHX3 | × | √ | |
| | rs4788673 | ZFHX3 | × | √ | |
| | rs4788674 | ZFHX3 | × | √ | |
| | rs4788675 | ZFHX3 | × | √ | |
| Verbal learn | rs80060958 | ZFHX3 | × | √ | |
| | rs4788676 | ZFHX3 | × | √ | Smoking status |
| | rs12930834 | ZFHX3 | × | √ | Smoking status (ever vs never smokers) |
| | rs12445751 | ZFHX3 | × | √ | |
| | rs18346254 | ZFHX3 | × | √ | |
| | rs28709625 | ZFHX3 | × | √ | |
| | rs34893494 | ZFHX3 | √ | × | |
| | rs80034230 | ZFHX3 | × | √ | |
| | rs14444228 | ZFHX3 | × | √ | |
| | rs75811565 | ZFHX3 | × | √ | |
| | rs77621608 | ZFHX3 | × | √ | |
| | rs74866145 | ZFHX3 | × | √ | |
| | rs11862948 | ZFHX3 | × | √ | |
| | rs6499599 | ZFHX3 | × | √ | |
| | rs8055870 | ZFHX3 | × | √ | |
| | rs76547147 | ZFHX3 | × | √ | |
| | rs1323276 | ZNF484 | √ | × | |
| | rs75898725 | ZNF787 | √ | × | |
| | rs76717212 | ZNF787 | √ | × | |
| | rs72490744 | ZNF787 | √ | × | |
| | rs59924464 | ZNF787 | √ | × | |
| | rs62122447 | ZNF787 | √ | × | |
| | rs62122448 | ZNF787 | √ | × | |
| | rs62122449 | ZNF787 | √ | × | |
| | rs2279413 | ZNF787 | √ | × | |
| | rs62122452 | ZNF787 | √ | × | |
| | rs58893103 | ZNF787 | √ | × | |

| | | | | | |
|---|---|---|---|---|---|
| rs157395 | ULK2 | √ | × | × | |
| rs157396 | ULK2 | √ | × | × | |
| rs205103 | ULK2 | √ | × | × | |
| rs205104 | ULK2 | √ | × | × | |
| rs205105 | ULK2 | √ | × | × | |
| rs3101815 | ULK2 | √ | × | × | |
| rs421561 | ULK2 | √ | × | × | |
| rs281357 | ULK2 | √ | × | × | |
| rs281354 | ULK2 | √ | × | × | |
| rs75871414 | ULK2 | √ | × | × | |
| rs188138 | ULK2 | √ | × | × | |
| rs281348 | ULK2 | √ | × | × | |
| rs281346 | ULK2 | √ | × | × | |
| rs3101813 | ULK2 | √ | × | × | |
| rs281332 | ULK2 | √ | × | × | |
| rs281327 | ULK2 | √ | × | × | |
| rs10851559 | UNC13C | √ | × | × | |
| rs11634827 | UNC13C | √ | × | × | |
| rs4776215 | UNC13C | √ | × | × | |
| rs719211 | UNC13C | √ | × | × | |
| rs12906904 | UNC13C | √ | × | × | |
| rs8030298 | UNC13C | √ | × | × | |
| rs10518760 | UNC13C | √ | × | × | |
| rs12917522 | UNC13C | √ | × | × | |
| rs8025731 | UNC13C | √ | × | × | |
| rs11856476 | UNC13C | √ | × | × | |
| rs12907862 | UNC13C | √ | × | × | |
| rs7183052 | UNC13C | √ | × | × | |
| rs7162346 | UNC13C | √ | × | × | |
| rs7183952 | UNC13C | √ | × | × | |
| rs2414285 | UNC13C | √ | × | × | |
| rs12901994 | UNC13C | √ | × | × | |
| rs2414288 | UNC13C | √ | × | × | |
| rs7170994 | UNC13C | √ | × | × | |
| rs11071065 | UNC13C | √ | × | × | |
| rs5025252 | UNC13C | √ | × | × | |
| rs12914912 | UNC13C | √ | × | × | |
| rs6493669 | UNC13C | √ | × | × | |
| rs4776222 | UNC13C | √ | × | × | |
| rs10851563 | UNC13C | √ | × | × | |
| rs7169339 | UNC13C | √ | × | × | |
| rs2414292 | UNC13C | √ | × | × | |
| rs7176603 | UNC13C | √ | × | × | |
| rs11265860 | UNQ6494 | √ | × | × | |
| rs3814545 | USP20 | √ | × | × | |
| rs3744793 | USP36 | √ | × | × | |
| rs61729065 | USP36 | × | √ | × | |
| rs62075628 | USP36 | × | √ | × | |
| rs145001834 | USP44 | × | √ | × | |
| rs183347370 | USP44 | × | √ | × | |
| rs188458080 | USP44 | × | √ | × | |
| rs75755929 | USP44 | × | √ | × | |
| rs10859892 | USP44 | × | √ | × | |
| rs370126718 | USP44 | × | √ | × | |
| rs142055127 | USP44 | × | √ | × | |
| rs7974458 | USP44 | × | √ | × | |
| rs7974596 | USP44 | × | √ | × | |
| rs11108086 | USP44 | × | √ | × | |
| rs74762990 | USP44 | × | √ | × | |
| rs12827048 | USP44 | × | √ | × | |
| rs301004 | USP44 | × | √ | × | |
| rs11108087 | USP44 | × | √ | × | |
| rs368548774 | USP44 | × | √ | × | |
| rs301003 | USP44 | × | √ | × | |
| rs188523 | USP44 | × | √ | × | |
| rs301001 | USP44 | × | √ | × | |
| rs301000 | USP44 | × | √ | × | |
| rs11108090 | USP44 | × | √ | × | |
| rs79637347 | USP44 | × | √ | × | |
| rs61939481 | USP44 | × | √ | × | |
| rs539968726 | USP44 | × | √ | × | |
| rs2769470 | USP44 | × | √ | × | |
| rs150739270 | USP44 | × | √ | × | |
| rs184428021 | UTRN | √ | × | × | |
| rs9390172 | UTRN | × | × | √ | Schizophrenia |
| rs191754698 | UTRN | √ | × | × | |
| rs9390178 | UTRN | × | × | √ | |
| rs9373419 | UTRN | × | × | √ | |
| rs2056744 | UTRN | × | × | √ | |
| rs17073995 | UTRN | × | × | √ | |
| rs10499232 | UTRN | × | × | √ | |
| rs12662483 | UTRN | × | × | √ | |
| rs9403572 | UTRN | × | × | √ | |
| rs79847827 | UTRN | × | × | √ | |
| rs7640893 | VIPR1 | × | √ | × | |
| rs76613688 | VIPR1 | × | √ | × | |
| rs735773 | VIPR1 | × | √ | × | |
| rs367866406 | VIPR1 | × | √ | × | |
| rs2278215 | VIPR1 | × | √ | × | |
| rs2267534 | VIPR1 | × | √ | × | |
| rs975151963 | VIPR1 | × | √ | × | |
| rs6799248 | VIPR1 | × | √ | × | |
| rs2267535 | VIPR1 | × | √ | × | |
| rs77432835 | VIPR1 | × | √ | × | |
| rs575955102 | VIPR1 | × | √ | × | |
| rs145692174 | VIPR1 | × | √ | × | |
| rs450812 | VIPR1 | × | √ | × | |
| rs28684623 | VIPR1 | × | √ | × | |
| rs413042 | VIPR1 | × | √ | × | |
| rs6797853 | VIPR1 | × | √ | × | |
| rs143977944 | VIPR1 | × | √ | × | |
| rs148637963 | VIPR1 | × | √ | × | |
| rs811198 | VIPR1 | × | √ | × | |
| rs2284132 | VIPR1 | × | √ | × | |
| rs558064244 | VIPR1 | × | √ | × | |
| rs146238950 | VIPR1 | × | √ | × | |
| rs373507779 | VIPR1 | × | √ | × | |
| rs78008566 | VIPR1 | × | √ | × | |
| rs181250457 | VIPR1 | × | √ | × | |
| rs186800587 | VIPR1 | × | √ | × | |
| rs75498396 | VIPR1 | × | √ | × | |
| rs9841214 | VIPR1 | × | √ | × | |
| rs580447 | VIPR1 | × | √ | × | |
| rs2267537 | VIPR1 | × | √ | × | |
| rs407931 | VIPR1 | × | √ | × | |
| rs78829151 | VIPR1 | × | √ | × | |
| rs2267538 | VIPR1 | × | √ | × | |
| rs2239499 | VIPR1 | × | √ | × | |
| rs570159494 | VIPR1 | × | √ | × | |
| rs34664814 | VIPR1 | × | √ | × | |
| rs144670843 | VIPR1 | × | √ | × | |
| rs2239500 | VIPR1 | × | √ | × | |
| rs406360 | VIPR1 | × | √ | × | |
| rs191842102 | VIPR1 | × | √ | × | |
| rs79133607 | VIPR1 | × | √ | × | |
| rs2239501 | VIPR1 | × | √ | × | |
| rs150035632 | VIPR1 | × | √ | × | |
| rs771678655 | VIPR1 | × | √ | × | |
| rs371882073 | VIPR1 | × | √ | × | |
| rs2276829 | VIPR1 | × | √ | × | |
| rs2229722 | VIPR1 | × | √ | × | |
| rs181293741 | VIPR1 | × | √ | × | |
| rs983400753 | VIPR1 | × | √ | × | |
| rs144611403 | VIPR1 | × | √ | × | |
| rs417387 | VIPR1 | × | √ | × | Cerebrospinal fluid clusteri |
| rs16810 | VIPR1 | × | √ | × | |
| rs9836133 | VIPR1 | × | √ | × | |
| rs14380 | VIPR1 | × | √ | × | |
| rs141898153 | VIPR1 | × | √ | × | |
| rs73169233 | VIPR2 | √ | × | × | |
| rs73169234 | VIPR2 | √ | × | × | |
| rs66942973 | VIPR2 | √ | × | × | |
| rs3793239 | VIPR2 | √ | × | × | |
| rs549833909 | VPS11 | × | √ | × | |
| rs117972538 | VPS11 | × | √ | × | |
| rs1784302 | VPS11 | × | √ | × | |
| rs1784305 | VPS11 | × | √ | × | |
| rs1786690 | VPS11 | × | √ | × | |
| rs148849984 | VPS11 | × | √ | × | |
| rs143519923 | VPS11 | × | √ | × | |
| rs76415576 | VPS11 | × | √ | × | |
| rs782395251 | VPS11 | × | √ | × | |
| rs1614264 | VPS11 | × | √ | × | |
| rs10892328 | VPS11 | × | √ | × | |
| rs1185460 | VPS11 | × | √ | × | Coronary artery disease |
| rs3825061 | VPS11 | × | √ | × | Body mass index |
| rs117412957 | VPS11 | × | √ | × | |
| rs78868054 | VPS11 | × | √ | × | |
| rs114778151 | VPS11 | × | √ | × | |
| rs138206564 | VPS11 | × | √ | × | |
| rs76001104 | VPS11 | × | √ | × | |
| rs1177563 | VPS11 | × | √ | × | |
| rs1177562 | VPS11 | × | √ | × | Cholesteryl ester levels in l |
| rs1168568 | VPS11 | × | √ | × | |
| rs1307145 | VPS11 | × | √ | × | Coronary artery disease |
| rs15818 | VPS11 | × | √ | × | Body mass index |
| rs4614 | VPS11 | × | √ | × | |
| rs11623700 | VTI1B | √ | × | × | |
| rs12050393 | VTI1B | √ | × | × | |
| rs761514 | VTI1B | √ | × | × | |
| rs11849521 | VTI1B | √ | × | × | |
| rs17782603 | VTI1B | √ | × | × | |
| rs4902507 | VTI1B | √ | × | × | |
| rs72723126 | VTI1B | √ | × | × | |
| rs12859864 | WBP4 | √ | × | × | |
| rs200814042 | WBP4 | √ | × | × | |
| rs4524855 | WNK2 | √ | × | × | |
| rs2040094 | WNK2 | √ | × | × | |
| rs10821084 | WNK2 | √ | × | × | |
| rs7856802 | WNK2 | √ | × | × | |
| rs7857039 | WNK2 | √ | × | × | |
| rs10821086 | WNK2 | √ | × | × | |
| rs12349094 | WNK2 | √ | × | × | Educational attainment |
| rs72645751 | WWC1 | √ | × | × | |
| rs149768622 | WWC2 | √ | × | × | |
| rs41457144 | WWC2 | √ | × | × | |
| rs73006721 | WWC2 | √ | × | × | |
| rs73006723 | WWC2 | √ | × | × | |
| rs73006725 | WWC2 | √ | × | × | |
| rs73006729 | WWC2 | √ | × | × | |
| rs11932929 | WWC2 | √ | × | × | |
| rs17074589 | WWC2 | √ | × | × | |
| rs17074592 | WWC2 | √ | × | × | |
| rs17074601 | WWC2 | √ | × | × | |
| rs73206633 | XXYLT1 | √ | × | × | |
| rs62290320 | XXYLT1 | √ | × | × | |
| rs138927338 | ZBTB43 | √ | × | × | |
| rs3733058 | ZBTB47 | × | √ | × | |
| rs339721 | ZBTB47 | × | √ | × | |
| rs146456247 | ZBTB47 | × | √ | × | |
| rs77688187 | ZBTB47 | × | √ | × | |
| rs339720 | ZBTB47 | × | √ | × | |
| rs573588179 | ZBTB47 | × | √ | × | |
| rs150215540 | ZBTB47 | × | √ | × | |
| rs539367074 | ZBTB47 | × | √ | × | |
| rs68190114 | ZBTB47 | × | √ | × | |
| rs11129967 | ZBTB47 | × | √ | × | |
| rs186177120 | ZBTB47 | × | √ | × | |
| rs3821837 | ZBTB47 | × | √ | × | |
| rs3733059 | ZBTB47 | × | √ | × | |
| rs3733060 | ZBTB47 | × | √ | × | |
| rs3733061 | ZBTB47 | × | √ | × | |
| rs2290977 | ZBTB47 | × | √ | × | |
| rs2290976 | ZBTB47 | × | √ | × | |
| rs2290975 | ZBTB47 | × | √ | × | |
| rs1008275 | ZFHX3 | √ | × | × | |
| rs59288483 | ZFHX3 | √ | × | × | |
| rs58084688 | ZFHX3 | √ | × | × | |
| rs57828240 | ZFHX3 | √ | × | × | Medication use (antithromb |
| rs719354 | ZFHX3 | √ | × | × | |
| rs67402452 | ZFHX3 | √ | × | × | |
| rs67329386 | ZFHX3 | √ | × | × | Atrial fibrillation |
| rs2106261 | ZFHX3 | √ | × | × | Atrial fibrillation |
| rs2359171 | ZFHX3 | √ | × | × | Atrial fibrillation |
| rs4499262 | ZFHX3 | √ | × | × | Incident atrial fibrillation |
| rs4788697 | ZFHX3 | × | √ | × | |
| rs34049478 | ZHX2 | √ | × | × | |
| rs12548759 | ZHX2 | × | √ | × | |
| rs3802265 | ZHX2 | × | √ | × | |
| rs28480089 | ZNF280D | √ | × | × | |
| rs55851513 | ZNF335 | √ | × | × | |
| rs77085984 | ZNF484 | √ | × | × | |
| rs75692950 | ZNF484 | √ | × | × | |
| rs10992502 | ZNF484 | √ | × | × | |
| rs11057563 | ZNF664-FAM101A | √ | × | × | |
| rs1979846 | ZNF664-FAM101A | √ | × | × | |
| rs10846646 | ZNF664-FAM101A | √ | × | × | |
| rs2342468 | ZNF664-FAM101A | √ | × | × | Height |
| rs2036306 | ZNF664-FAM101A | √ | × | × | |

| | | | | | |
|---|---|---|---|---|---|
| rs1456780 | TMEM117 | × | √ | × | |
| rs14837371 | TMEM117 | × | √ | × | |
| rs1231336 | TMEM117 | × | √ | × | |
| rs18592491 | TMEM117 | × | √ | × | |
| rs7308972 | TMEM117 | × | √ | × | |
| rs1396658 | TMEM117 | × | √ | × | |
| rs1831716 | TMEM117 | × | √ | × | |
| rs1512540 | TMEM117 | × | √ | × | |
| rs7308973 | TMEM117 | × | √ | × | |
| rs1209976 | TMEM117 | × | √ | × | |
| rs20217617 | TMEM117 | × | √ | × | |
| rs7408462 | TMEM117 | × | √ | × | |
| rs18418312 | TMEM117 | × | √ | × | |
| rs1447072 | TMEM117 | × | √ | × | |
| rs11182402 | TMEM117 | × | √ | × | |
| rs14542582 | TMEM117 | × | √ | × | |
| rs1118240 | TMEM117 | × | √ | × | |
| rs7451163 | TMEM117 | × | √ | × | |
| rs1118240 | TMEM117 | × | √ | × | |
| rs866839 | TMEM117 | × | √ | × | |
| rs36929201 | TMEM117 | × | √ | × | |
| rs1118240 | TMEM117 | × | √ | × | |
| rs7838130 | TMEM117 | × | √ | × | |
| rs1118240 | TMEM117 | × | √ | × | |
| rs5619833 | TMEM117 | × | √ | × | |
| rs842202 | TMEM117 | √ | × | × | |
| rs842199 | TMEM117 | √ | × | × | |
| rs1709406 | TMEM117 | √ | × | × | |
| rs1478692 | TMEM117 | √ | × | × | |
| rs3814790 | TNFRSF19 | √ | × | × | |
| rs1469697 | TNFRSF19 | √ | × | × | |
| rs7333789 | TNFRSF19 | √ | × | × | |
| rs9510796 | TNFRSF19 | √ | × | × | |
| rs6766775 | TP63 | √ | × | × | |
| rs7839474 | TPCN1 | √ | × | × | |
| rs7133428 | TPCN1 | √ | × | × | |
| rs79778981 | TPCN1 | √ | × | × | |
| rs7730327 | TPCN1 | √ | × | × | |
| rs7296638 | TPCN1 | √ | × | × | |
| rs7942310 | TPCN1 | √ | × | × | |
| rs5920583 | TPCN1 | √ | × | × | |
| rs6194363 | TPCN1 | √ | × | × | |
| rs2241544 | TPCN1 | √ | × | × | |
| rs1229865 | TPCN1 | √ | × | × | |
| rs5297443 | TPCN1 | √ | × | × | |
| rs9372738 | TRDN | √ | × | × | |
| rs7835851 | TRDN | √ | × | × | |
| rs6147815 | TRDN | √ | × | × | |
| rs73549391 | TRDN | √ | × | × | |
| rs7354939 | TRDN | √ | × | × | |
| rs1197045 | TRDN | √ | × | × | |
| rs1196666 | TRDN | √ | × | × | |
| rs7801246 | TRDN | √ | × | × | |
| rs76400701 | TRDN | √ | × | × | |
| rs9372741 | TRDN | √ | × | × | |
| rs7771303 | TRDN | √ | × | × | |
| rs7751099 | TRDN | √ | × | × | |
| rs73551207 | TRDN | √ | × | × | |
| rs2873478 | TRDN | √ | × | × | |
| rs4346869 | TRDN | √ | × | × | |
| rs1431290 | TRDN | √ | × | × | |
| rs1346920 | TRDN | √ | × | × | |
| rs1431288 | TRDN | √ | × | × | |
| rs75057871 | TRDN | √ | × | × | |
| rs7355121 | TRDN | √ | × | × | |
| rs14381347 | TRDN | √ | × | × | |
| rs7355121 | TRDN | √ | × | × | |
| rs1196727 | TRDN | √ | × | × | |
| rs1196463 | TRDN | √ | × | × | |
| rs1128315 | TRDN | √ | × | × | |
| rs76177651 | TRDN | √ | × | × | |
| rs12190077 | TRDN | √ | × | × | |
| rs1319670 | TRDN | √ | × | × | |
| rs9320939 | TRDN | √ | × | × | Hypertrophic cardiomyopathy (sarcomere negative) |
| rs3557659 | TRDN | √ | × | × | |
| rs3427939 | TRDN | √ | × | × | |
| rs1219260 | TRDN | √ | × | × | |
| rs1219993 | TRDN | √ | × | × | |
| rs1220624 | TRDN | √ | × | × | |
| rs6939706 | TRDN | √ | × | × | |
| rs6940253 | TRDN | √ | × | × | |
| rs9490795 | TRDN | √ | × | × | |
| rs9490796 | TRDN | √ | × | × | |
| rs9490798 | TRDN | √ | × | × | |
| rs4996639 | TRDN | √ | × | × | |
| rs12196787 | TRDN | √ | × | × | |
| rs1219816 | TRDN | √ | × | × | |
| rs12198167 | TRDN | √ | × | × | |
| rs4897276 | TRDN | √ | × | × | |
| rs7103188 | TRIM44 | √ | × | × | |
| rs16927857 | TRIM44 | √ | × | × | |
| rs7470284 | TRIM44 | √ | × | × | |
| rs1103325 | TRIM44 | √ | × | × | |
| rs1222227 | TRIM44 | √ | × | × | |
| rs7864747 | TRIM44 | √ | × | × | |
| rs6652231 | TRIM44 | √ | × | × | |
| rs1103326 | TRIM44 | √ | × | × | |
| rs1692787 | TRIM44 | √ | × | × | |
| rs12576967 | TRIM44 | √ | × | × | |
| rs77013261 | TRIM44 | √ | × | × | |
| rs7566706 | TRIM44 | √ | × | × | |
| rs2296177 | TRIM44 | √ | × | × | |
| rs11033261 | TRIM44 | √ | × | × | |
| rs1169433 | TRIM44 | √ | × | × | |
| rs3112061 | TRIM44 | √ | × | × | |
| rs9633874 | TRIM44 | √ | × | × | |
| rs7777062 | TRIM44 | √ | × | × | |
| rs1103326 | TRIM44 | √ | × | × | |
| rs6766795 | TRIM44 | √ | × | × | |
| rs16927891 | TRIM44 | √ | × | × | |
| rs10501141 | TRIM44 | √ | × | × | |
| rs11033267 | TRIM44 | √ | × | × | |
| rs1103326 | TRIM44 | √ | × | × | |
| rs7587146 | TRIM44 | √ | × | × | |
| rs4756254 | TRIM44 | √ | × | × | |
| rs1103327 | TRIM44 | √ | × | × | |
| rs7838953 | TRIM44 | √ | × | × | |
| rs4909018 | TRIM71 | √ | × | × | |
| rs7528695 | TRIM71 | √ | × | × | |
| rs79126022 | TRIM71 | √ | × | × | |
| rs7694963 | TRIM71 | √ | × | × | |
| rs1174410 | TRIM71 | √ | × | × | |
| rs1913974 | TRIM71 | × | √ | × | |
| rs7848465 | TRIM71 | × | √ | × | |
| rs75695281 | TRIM71 | × | √ | × | |
| rs6550169 | TRIM71 | × | √ | × | |
| rs4274781 | TRIM71 | × | √ | × | |
| rs7440457 | TRIM71 | × | √ | × | |
| rs7616050 | TRIM71 | × | √ | × | |
| rs1599770 | TRIM71 | × | √ | × | |
| rs1449395 | TRPM3 | √ | × | × | |
| rs1856214 | TRPM3 | √ | × | × | |
| rs2289496 | TSPYL5 | √ | × | × | |
| rs1151201 | TUBB3 | × | √ | × | |
| rs1873536 | TUBB3 | × | √ | × | |
| rs12600051 | TUBB3 | × | √ | × | |
| rs1259614 | TUBB3 | × | √ | × | |
| rs2850807 | TUBB3 | × | √ | × | |
| rs19152112 | TUBB3 | × | √ | × | |
| rs1259850 | TUBB3 | × | √ | × | |
| rs14476497 | TWF1 | × | √ | × | |
| rs1088056 | TWF1 | × | √ | × | |
| rs1992604 | TWF1 | × | √ | × | |
| rs18616067 | TWF1 | × | √ | × | |
| rs7955112 | TWF1 | × | √ | × | |
| rs11726967 | TWF1 | × | √ | × | |
| rs11753042 | TWF1 | × | √ | × | |
| rs307657 | UBAP2 | √ | × | × | |
| rs307658 | UBAP2 | √ | × | × | |
| rs307678 | UBAP2 | √ | × | × | |
| rs307682 | UBAP2 | √ | × | × | |
| rs307683 | UBAP2 | √ | × | × | |
| rs307684 | UBAP2 | √ | × | × | |
| rs4272470 | UBAP2 | √ | × | × | |
| rs2781284 | UBAP2 | √ | × | × | |
| rs307703 | UBAP2 | √ | × | × | Brain morphology (MOSTest) |
| rs834027 | UBAP2 | √ | × | × | |
| rs72627062 | UBE2E1 | √ | × | × | |
| rs4858062 | UBE2E2 | √ | × | × | |
| rs7692133 | UBE2E2 | √ | × | × | |
| rs11691437 | UBE2E2 | √ | × | × | |
| rs4858487 | UBE2E2 | √ | × | × | |
| rs2132146 | UBE2E2 | √ | × | × | |
| rs2101811 | UBE2E2 | √ | × | × | |
| rs9848389 | UBE2E2 | √ | × | × | |
| rs7585649 | UBE2F | √ | × | × | |
| rs9751676 | UBE2F | √ | × | × | |
| rs74398701 | UBE2F | √ | × | × | |
| rs7931787 | UBE2F | √ | × | × | |
| rs2296107 | UBR4 | √ | × | × | |
| rs7538944 | UHMK1 | √ | × | × | |
| rs189820 | UHMK1 | √ | × | × | |
| rs1165670 | ULK2 | √ | × | × | |
| rs1779437 | ULK2 | √ | × | × | |
| rs1179706 | ULK2 | √ | × | × | |
| rs1172391 | ULK2 | √ | × | × | |
| rs1181151 | ULK2 | √ | × | × | |
| rs14597151 | ULK2 | √ | × | × | |
| rs1160661 | ULK2 | √ | × | × | |
| rs75983441 | ULK2 | √ | × | × | |
| rs7719162 | ULK2 | √ | × | × | |
| rs1177065 | ULK2 | √ | × | × | |
| rs1174437 | ULK2 | √ | × | × | |
| rs7723549 | ULK2 | √ | × | × | |
| rs7961161 | ULK2 | √ | × | × | |
| rs7953400 | ULK2 | √ | × | × | |
| rs1173881 | ULK2 | √ | × | × | |
| rs9898591 | ULK2 | √ | × | × | |
| rs7767764 | ULK2 | √ | × | × | |
| rs1499531 | ULK2 | √ | × | × | |
| rs1174116 | ULK2 | √ | × | × | |
| rs11795951 | ULK2 | √ | × | × | |
| rs1181312 | ULK2 | √ | × | × | |
| rs11698947 | ULK2 | √ | × | × | |
| rs1585751 | ULK2 | √ | × | × | |
| rs1178418 | ULK2 | √ | × | × | |
| rs1175324 | ULK2 | √ | × | × | |
| rs7695705 | ULK2 | √ | × | × | |
| rs7174025 | UNC13C | √ | × | × | |
| rs1290219 | UNC13C | √ | × | × | |
| rs7503006 | UNQ6494 | √ | × | × | |
| rs78401801 | UNQ6494 | √ | × | × | |
| rs7834380 | UNQ6494 | √ | × | × | Estimated glomerular filtration rate |
| rs7848364 | UNQ6494 | √ | × | × | |
| rs7349425 | UNQ6494 | √ | × | × | |
| rs7033256 | UNQ6494 | √ | × | × | |
| rs1329471 | USP20 | √ | × | × | |
| rs3758334 | USP20 | √ | × | × | |
| rs3814546 | USP20 | √ | × | × | |
| rs1328475 | USP20 | √ | × | × | |
| rs11793657 | USP20 | √ | × | × | |
| rs1210360 | USP36 | √ | × | × | |
| rs7316923 | VIPR2 | √ | × | × | |
| rs7316923 | VIPR2 | √ | × | × | |
| rs7797488 | VIPR2 | √ | × | × | |
| rs3793228 | VIPR2 | √ | × | × | |
| rs3763427 | VIPR2 | √ | × | × | |
| rs1173879 | VTI1B | √ | × | × | |
| rs7545075 | WDR34 | × | √ | × | |
| rs7366995 | WDR34 | × | √ | × | |
| rs1171011 | WDR34 | × | √ | × | |
| rs73669957 | WDR34 | × | √ | × | |
| rs7571503 | WDR34 | × | √ | × | |
| rs7508262 | WDR34 | × | √ | × | |
| rs1011460 | WDR34 | × | √ | × | |
| rs7366995 | WDR34 | × | √ | × | |
| rs7619404 | WDR34 | × | √ | × | |
| rs945836 | WDR34 | × | √ | × | |
| rs7643929 | WDR34 | × | √ | × | |
| rs2298045 | WDR34 | × | √ | × | |
| rs5783763 | WDR34 | × | √ | × | |
| rs14263971 | WWC2 | × | √ | × | |
| rs18733702 | WWC2 | √ | × | × | |
| rs2175476 | WWC2 | √ | × | × | |
| rs7677983 | WWC2 | √ | × | × | |
| rs5871447 | WWC2 | √ | × | × | |
| rs28756361 | XXYLT1 | √ | × | × | |
| rs12128631 | ZBTB41 | √ | × | × | |
| rs1042991 | ZBTB41 | √ | × | × | Systolic blood pressure |

| | | | | |
|---|---|---|---|---|
| rs62122411 | ZNF787 | √ | × | × |
| rs17815373 | ZNF787 | √ | × | × |
| rs7245587 | ZNF787 | √ | × | × |
| rs67104753 | ZNF787 | √ | × | × |
| rs191120313 | ZSCAN5B | × | √ | × |
| rs17753438 | ZSCAN5B | × | √ | × |

| | | | | | |
|---|---|---|---|---|---|
| rs10922177 | ZBTB41 | √ | × | × | Complement factor H-related protein 2 levels |
| rs10922179 | ZBTB41 | √ | × | × | Total cholesterol levels |
| rs10922181 | ZBTB41 | √ | × | × | |
| rs3846059 | ZBTB47 | × | √ | × | |
| rs82755 | ZBTB47 | × | √ | × | |
| rs14749476 | ZBTB47 | × | √ | × | |
| rs76179470 | ZER1 | √ | × | × | |
| rs4788672 | ZFHX3 | × | √ | × | |
| rs8047422 | ZFHX3 | × | √ | × | |
| rs2359128 | ZFHX3 | × | √ | × | |
| rs17680397 | ZFHX3 | × | √ | × | Lifetime smoking |
| rs9923980 | ZFHX3 | × | √ | × | |
| rs78711661 | ZFHX3 | × | √ | × | |
| rs79111200 | ZFHX3 | × | √ | × | |
| rs2040509 | ZFHX3 | × | √ | × | |
| rs11797644 | ZFHX3 | × | √ | × | |
| rs75847797 | ZFHX3 | × | √ | × | |
| rs77043447 | ZFHX3 | × | √ | × | |
| rs60315503 | ZFHX3 | × | √ | × | |
| rs14675169 | ZFHX3 | × | √ | × | |
| rs721824 | ZFHX3 | × | √ | × | |
| rs9923193 | ZFHX3 | × | √ | × | |
| rs9922718 | ZFHX3 | × | √ | × | |
| rs4788673 | ZFHX3 | × | √ | × | |
| rs4788674 | ZFHX3 | × | √ | × | |
| rs4788675 | ZFHX3 | × | √ | × | |
| rs80060958 | ZFHX3 | × | √ | × | |
| rs4788676 | ZFHX3 | × | √ | × | Smoking status |
| rs12930834 | ZFHX3 | × | √ | × | Smoking status (ever vs never smokers) |
| rs12445751 | ZFHX3 | × | √ | × | |
| rs18346254 | ZFHX3 | × | √ | × | |
| rs28709625 | ZFHX3 | × | √ | × | |
| rs52806655 | ZFHX3 | × | √ | × | |
| rs57644161 | ZNF335 | × | × | √ | |
| rs18319185 | ZNF484 | √ | × | × | |
| rs2589970 | ZNF774 | √ | × | × | |
| rs2589968 | ZNF774 | √ | × | × | |
| rs62122447 | ZNF787 | √ | × | × | |
| rs62122448 | ZNF787 | √ | × | × | |
| rs2279413 | ZNF787 | √ | × | × | |
| rs11673628 | ZNF787 | √ | × | × | |
| rs12380014 | ZNF79 | √ | × | × | |

**Table S20. The potential enrichment pathways of adaptive introgression from Denisovans**

| original_id | TCL | TEL | THL | TSL | Gene ID | Tax ID | Description | Disease & Drugs (ChatGPT) | Biological Process (GO) | TotalEvidenceCount |
|---|---|---|---|---|---|---|---|---|---|---|
| DIS3L2 | 1 | 1 | 1 | 1 | 129563 | H. sapiens | DIS3 like 3'-5' exoribonuclease 2 | Mutations in the DIS3L2 gene have been | GO:1990074 polyuridylation-dependent | 4 |
| AGFG1 | 0 | 1 | 1 | 1 | 3267 | H. sapiens | ArfGAP with FG repeats 1 | There is limited information available on t | GO:0007289 spermatid nucleus differe | 3 |
| CNTNAP2 | 1 | 0 | 1 | 0 | 26047 | H. sapiens | contactin associated protein 2 | CNTNAP2 has been implicated in severa | GO:0071109 superior temporal gyrus d | 2 |
| COL4A3BP | 0 | 1 | 0 | 1 | 10087 | H. sapiens | ceramide transporter 1 | Goodpasture disease is a rare autoimmu | GO:0120012 intermembrane sphingoli | 2 |
| LOC102659288 | 0 | 1 | 0 | 1 | 102659288 | H. sapiens | long intergenic non-protein coding RNA 2888 | | | 2 |
| LTBP1 | 0 | 0 | 1 | 1 | 4052 | H. sapiens | latent transforming growth factor beta binding protein 1 | Mutations in the LTBP1 gene have been a | GO:0035583 sequestering of TGFbeta | 2 |
| C6orf1 | 0 | 1 | 0 | 1 | 221491 | H. sapiens | small integral membrane protein 29 | Unfortunately, there is currently limited information available on the disease impli | | 2 |
| ANKRD31 | 1 | 0 | 0 | 0 | 256006 | H. sapiens | ankyrin repeat domain 31 | There is currently limited information avai | GO:0010780 meiotic DNA double-stran | 1 |
| GPR83 | 1 | 0 | 0 | 0 | 10888 | H. sapiens | G protein-coupled receptor 83 | There is limited information available on t | GO:0007631 feeding behavior;GO:000 | 1 |
| CHRND | 0 | 0 | 0 | 1 | 1144 | H. sapiens | cholinergic receptor nicotinic delta subunit | Defects in the CHRND gene have been a | GO:0048630 skeletal muscle tissue gro | 1 |
| KCNQ1DN | 0 | 0 | 0 | 1 | 55539 | H. sapiens | KCNQ1 downstream neighbor | | GO:0008150 biological_process | 1 |
| EPAS1 | 0 | 0 | 1 | 0 | 2034 | H. sapiens | endothelial PAS domain protein 1 | EPAS1 gene mutations have been linked | GO:0048625 myoblast fate commitmer | 1 |
| HMGA1 | 0 | 0 | 0 | 1 | 3159 | H. sapiens | high mobility group AT-hook 1 | The HMGA1 gene has been implicated in | GO:0090402 oncogene-induced cell se | 1 |
| POLK | 0 | 0 | 0 | 1 | 51426 | H. sapiens | DNA polymerase kappa | There are currently no drugs on the mark | GO:0006297 nucleotide-excision repair | 1 |
| CXADR | 0 | 1 | 0 | 0 | 1525 | H. sapiens | CXADR Ig-like cell adhesion molecule | The CXADR gene has been implicated in | GO:0086072 AV node cell-bundle of Hi | 1 |

**Table S21. The distinct adaptive Neanderthal-introgressed haplotypes in differentiated TYC individuals**

| THL | Gene | iHS | XPEHH | PBS | Traits | TCL | Gene | iHS | XPEHH | PBS | Traits | TSL | Gene | iHS | XPEHH | PBS | Traits | TEL | Gene | iHS | XPEHH | Traits |
|---|---|---|---|---|---|---|---|---|---|---|---|---|---|---|---|---|---|---|---|---|---|---|
| rs7588620 | AGFG1 | √ | √ | × | | rs1903839 | ANKRD31 | √ | √ | × | | rs3455357 | AGFG1 | √ | √ | × | | rs1690030 | XRCC4 | √ | √ | |
| rs10933191 | AGFG1 | √ | √ | × | | rs4360021 | ANKRD31 | √ | √ | × | | rs7523456 | AGFG1 | √ | √ | × | | rs3455357 | AGFG1 | √ | × | |
| rs72980874 | AGFG1 | √ | √ | × | | rs1477934 | ANKRD31 | √ | √ | × | | rs1903839 | ANKRD31 | √ | × | × | | rs7523456 | AGFG1 | × | √ | |
| rs4973181 | AGFG1 | √ | √ | × | | rs7399507 | DIS3L2 | √ | √ | × | | rs4360021 | ANKRD31 | √ | × | × | | rs1903839 | ANKRD31 | √ | × | |
| rs7598597 | AGFG1 | √ | √ | × | | rs4389672 | ANKRD31 | √ | √ | × | | rs1477934 | ANKRD31 | √ | × | × | | rs4360021 | ANKRD31 | √ | × | |
| rs13426457 | AGFG1 | √ | √ | × | | rs1477933 | ANKRD31 | √ | √ | × | | rs1477933 | ANKRD31 | √ | × | × | | rs1477934 | ANKRD31 | √ | × | |
| rs6761651 | AGFG1 | √ | √ | × | | rs1477932 | ANKRD31 | √ | √ | × | | rs1477932 | ANKRD31 | √ | × | × | | rs2247137 | CNTNAP2 | √ | × | |
| rs13382948 | AGFG1 | √ | √ | × | | rs7735937 | ANKRD31 | √ | √ | × | | rs7735937 | ANKRD31 | √ | × | × | | rs2247143 | CNTNAP2 | √ | × | |
| rs4555337 | AGFG1 | √ | √ | × | | rs6888707 | ANKRD31 | √ | √ | × | | rs2247137 | CNTNAP2 | √ | × | × | | rs2247149 | CNTNAP2 | √ | × | |
| rs4312485 | AGFG1 | √ | √ | × | | rs1023337 | CNTNAP2 | √ | × | × | | rs2247143 | CNTNAP2 | √ | × | × | | rs2538984 | CNTNAP2 | √ | × | |
| rs6706723 | AGFG1 | √ | √ | × | | rs2080156 | CNTNAP2 | √ | × | × | | rs2247149 | CNTNAP2 | √ | × | × | | rs2538979 | CNTNAP2 | √ | × | |
| rs10233374 | CNTNAP2 | √ | √ | × | | rs2080154 | CNTNAP2 | √ | × | × | | rs2538984 | CNTNAP2 | √ | × | × | | rs4277518 | AGFG1 | × | √ | |
| rs2080156 | CNTNAP2 | √ | √ | × | | rs2710108 | CNTNAP2 | √ | × | × | | rs2538979 | CNTNAP2 | √ | × | × | | rs4254497 | AGFG1 | × | √ | |
| rs2080154 | CNTNAP2 | √ | √ | × | | rs2710105 | CNTNAP2 | √ | × | × | | rs7594606 | AGFG1 | × | √ | × | | rs4235683 | ANKRD31 | √ | × | |
| rs2710120 | CNTNAP2 | √ | √ | × | | rs2710102 | CNTNAP2 | √ | × | × | | rs4277518 | AGFG1 | × | √ | × | | rs1317704 | ANKRD31 | √ | × | |
| rs2710108 | CNTNAP2 | √ | √ | × | | rs2538994 | CNTNAP2 | √ | × | × | | rs4254497 | AGFG1 | × | √ | × | | rs6885071 | ANKRD31 | √ | × | |
| rs2710105 | CNTNAP2 | √ | √ | × | | rs7806512 | CNTNAP2 | √ | × | × | | rs1046251 | ANKRD31 | √ | × | × | | rs9654429 | ANKRD31 | √ | × | |
| rs2710102 | CNTNAP2 | √ | √ | × | | rs7806519 | CNTNAP2 | √ | × | × | | rs6453112 | ANKRD31 | √ | × | × | | rs7399503 | DIS3L2 | √ | × | |
| rs2538994 | CNTNAP2 | √ | √ | × | | rs2710099 | CNTNAP2 | √ | × | × | | rs4235683 | ANKRD31 | √ | × | × | | rs7399503 | DIS3L2 | √ | × | Lung function (FVC) and body mass index |
| rs7806512 | CNTNAP2 | √ | √ | × | | rs2710098 | CNTNAP2 | √ | × | × | | rs1317704 | ANKRD31 | √ | × | × | | rs2874531 | XRCC4 | √ | × | |
| rs7806519 | CNTNAP2 | √ | √ | × | | rs759179 | CNTNAP2 | √ | × | × | | rs6885071 | ANKRD31 | √ | × | × | | rs2836019 | XRCC4 | √ | × | |
| rs2710099 | CNTNAP2 | √ | √ | × | | rs759178 | CNTNAP2 | √ | × | × | | rs9654429 | ANKRD31 | √ | × | × | | rs1251514 | XRCC4 | √ | × | |
| rs2710098 | CNTNAP2 | √ | √ | × | | rs2708238 | CNTNAP2 | √ | × | × | | rs7399503 | DIS3L2 | √ | × | × | | rs1690026 | XRCC4 | √ | × | |
| rs759179 | CNTNAP2 | √ | √ | × | | rs2708239 | CNTNAP2 | √ | × | × | | rs7399503 | DIS3L2 | √ | × | × | Lung funct | rs1552909 | XRCC4 | √ | × | |
| rs759178 | CNTNAP2 | √ | √ | × | | rs2708240 | CNTNAP2 | √ | × | × | QT interval | rs2874531 | XRCC4 | √ | × | × | | rs1562227 | XRCC4 | √ | × | |
| rs2708238 | CNTNAP2 | √ | √ | × | | rs4140925 | CNTNAP2 | √ | × | × | | rs7979481 | XRCC4 | √ | × | × | | rs3777033 | XRCC4 | √ | × | |
| rs2708239 | CNTNAP2 | √ | √ | × | | rs2538991 | CNTNAP2 | √ | × | × | | rs2836019 | XRCC4 | √ | × | × | | rs1687649 | XRCC4 | √ | × | |
| rs2708240 | CNTNAP2 | √ | √ | × | QT interval | rs2538990 | CNTNAP2 | √ | × | × | | rs1251514 | XRCC4 | √ | × | × | | rs6436698 | AGFG1 | × | √ | |
| rs4140925 | CNTNAP2 | √ | √ | × | | rs2538989 | CNTNAP2 | √ | × | × | | rs1690026 | XRCC4 | √ | × | × | | rs1690016 | XRCC4 | √ | × | |
| rs2538991 | CNTNAP2 | √ | √ | × | | rs2247042 | CNTNAP2 | √ | × | × | | rs1552909 | XRCC4 | √ | × | × | | rs2838313 | XRCC4 | √ | × | |
| rs2538990 | CNTNAP2 | √ | √ | × | | rs2247043 | CNTNAP2 | √ | × | × | | rs1562227 | XRCC4 | √ | × | × | | rs2838313 | XRCC4 | √ | × | |
| rs2538989 | CNTNAP2 | √ | √ | × | | rs2538988 | CNTNAP2 | √ | × | × | | rs3777033 | XRCC4 | √ | × | × | | rs1690016 | XRCC4 | √ | × | |
| rs2247042 | CNTNAP2 | √ | √ | × | | rs2708243 | CNTNAP2 | √ | × | × | | rs1687649 | XRCC4 | √ | × | × | | rs4703944 | XRCC4 | √ | × | |
| rs2247043 | CNTNAP2 | √ | √ | × | | rs2538987 | CNTNAP2 | √ | × | × | | rs1690030 | XRCC4 | √ | × | × | | rs7758428 | XRCC4 | √ | × | |
| rs2538988 | CNTNAP2 | √ | √ | × | | rs2538986 | CNTNAP2 | √ | × | × | | rs6436698 | AGFG1 | × | √ | × | | rs2061783 | XRCC4 | √ | × | |
| rs2708243 | CNTNAP2 | √ | √ | × | | rs2538985 | CNTNAP2 | √ | × | × | | rs7875816 | ANKRD31 | √ | × | × | | rs4458554 | XRCC4 | √ | × | |
| rs2538987 | CNTNAP2 | √ | √ | × | | rs2247137 | CNTNAP2 | √ | × | × | | rs1335914 | ANKRD31 | √ | × | × | | rs925199 | XRCC4 | √ | × | |
| rs2538986 | CNTNAP2 | √ | √ | × | | rs2247143 | CNTNAP2 | √ | × | × | | rs2059159 | ANKRD31 | √ | × | × | | rs2089566 | XRCC4 | √ | × | |
| rs2538985 | CNTNAP2 | √ | √ | × | | rs2247149 | CNTNAP2 | √ | × | × | | rs1003652 | ANKRD31 | √ | × | × | | rs7844564 | XRCC4 | √ | × | |
| rs2247137 | CNTNAP2 | √ | √ | × | | rs2538984 | CNTNAP2 | √ | × | × | | rs6889429 | ANKRD31 | √ | × | × | | rs1251893 | XRCC4 | √ | × | |
| rs2247143 | CNTNAP2 | √ | √ | × | | rs2538983 | CNTNAP2 | √ | × | × | | rs4704188 | ANKRD31 | √ | × | × | | rs4142947 | XRCC4 | √ | × | |
| rs2247149 | CNTNAP2 | √ | √ | × | | rs2538979 | CNTNAP2 | √ | × | × | | rs1005094 | ANKRD31 | √ | × | × | | rs6891117 | XRCC4 | √ | × | |
| rs2538984 | CNTNAP2 | √ | √ | × | | rs569781 | FOLR4 | × | √ | × | | rs1345704 | ANKRD31 | √ | × | × | | rs4547524 | AGFG1 | × | √ | |
| rs2538983 | CNTNAP2 | √ | √ | × | | rs7594606 | AGFG1 | √ | × | × | | rs6722553 | DIS3L2 | √ | × | × | | rs4246652 | AGFG1 | √ | × | |
| rs2538981 | CNTNAP2 | √ | √ | × | | rs3455357 | AGFG1 | √ | × | × | | rs1574528 | DIS3L2 | √ | × | × | | rs4246651 | AGFG1 | √ | × | |
| rs2538979 | CNTNAP2 | √ | √ | × | | rs4277518 | AGFG1 | √ | × | × | | rs209700 | EPHA8 | √ | × | × | | rs4485561 | AGFG1 | √ | × | |
| rs2538976 | CNTNAP2 | √ | √ | × | | rs4254497 | AGFG1 | √ | × | × | | rs7881717 | MRE11A | √ | × | × | | rs1093319 | AGFG1 | √ | × | |
| rs569781 | FOLR4 | √ | √ | × | | rs4324655 | ANKRD31 | √ | × | × | | rs1690016 | XRCC4 | √ | × | × | | rs4246647 | AGFG1 | √ | × | |
| rs4675198 | AGFG1 | √ | × | × | | rs6869345 | ANKRD31 | √ | × | × | | rs2838313 | XRCC4 | √ | × | × | | rs3551122 | AGFG1 | √ | × | |
| rs10194959 | AGFG1 | √ | × | × | | rs1046251 | ANKRD31 | √ | × | × | | rs2838313 | XRCC4 | √ | × | × | | rs1018979 | AGFG1 | √ | × | Cortical surface area |
| rs4675199 | AGFG1 | √ | × | × | | rs6453112 | ANKRD31 | √ | × | × | | rs1690016 | XRCC4 | √ | × | × | | rs7143105 | AGFG1 | √ | × | |
| rs4675200 | AGFG1 | √ | × | × | | rs4235683 | ANKRD31 | √ | × | × | | rs4703944 | XRCC4 | √ | × | × | | rs6737495 | AGFG1 | √ | × | |
| rs78859406 | AGFG1 | √ | × | × | | rs1317704 | ANKRD31 | × | √ | × | | rs7758428 | XRCC4 | √ | × | × | | rs4973499 | AGFG1 | √ | × | |
| rs6715882 | AGFG1 | √ | × | × | | rs6896474 | ANKRD31 | √ | × | × | | rs2061783 | XRCC4 | √ | × | × | | rs1303520 | AGFG1 | √ | × | |
| rs10210812 | AGFG1 | √ | × | × | | rs2837629 | ANKRD31 | √ | × | × | | rs4458554 | XRCC4 | √ | × | × | | rs7578871 | AGFG1 | √ | × | |
| rs7587864 | AGFG1 | √ | × | × | | rs965735 | ANKRD31 | √ | × | × | | rs925199 | XRCC4 | √ | × | × | | rs4349340 | AGFG1 | √ | × | |
| rs7594606 | AGFG1 | √ | × | × | | rs6885071 | ANKRD31 | √ | × | × | | rs2089566 | XRCC4 | √ | × | × | | rs6436700 | AGFG1 | √ | × | |
| rs34553575 | AGFG1 | √ | × | × | | rs7711762 | ANKRD31 | √ | × | × | | rs7844564 | XRCC4 | √ | × | × | | rs7608104 | AGFG1 | √ | × | |
| rs4277518 | AGFG1 | √ | × | × | | rs1363577 | ANKRD31 | √ | × | × | | rs1251893 | XRCC4 | √ | × | × | | rs6707385 | AGFG1 | √ | × | |
| rs4547524 | AGFG1 | √ | × | × | | rs1422696 | ANKRD31 | √ | × | × | | rs4142947 | XRCC4 | √ | × | × | | rs6731443 | AGFG1 | √ | × | Airway hyperresponsiveness |
| rs4254497 | AGFG1 | √ | × | × | | rs9654426 | ANKRD31 | √ | × | × | | rs6891117 | XRCC4 | √ | × | × | | rs3491559 | AGFG1 | √ | × | |
| rs4496327 | AGFG1 | √ | × | × | | rs9654429 | ANKRD31 | √ | × | × | | rs7587864 | AGFG1 | × | √ | × | | rs7633533 | ANKRD31 | √ | × | |
| rs10191597 | AGFG1 | √ | × | × | | rs7669345 | BTG3 | × | √ | × | | rs4547524 | AGFG1 | × | √ | × | | rs9654427 | ANKRD31 | √ | × | |
| rs10182920 | AGFG1 | √ | × | × | | rs5930930 | CASZ1 | × | × | √ | | rs4496327 | AGFG1 | × | √ | × | | rs958535 | XRCC4 | √ | × | |
| rs4246652 | AGFG1 | √ | × | × | | rs1177007 | CNTNAP2 | √ | × | × | | rs1019159 | AGFG1 | × | √ | × | | rs3777021 | XRCC4 | √ | × | |
| rs4246651 | AGFG1 | √ | × | × | | rs1186173 | CNTNAP2 | √ | × | × | | rs1018292 | AGFG1 | × | √ | × | | rs1807027 | AGFG1 | × | √ | |
| rs4075128 | AGFG1 | √ | × | × | | rs1723438 | CNTNAP2 | √ | × | × | | rs4246652 | AGFG1 | × | √ | × | | rs6742120 | AGFG1 | × | √ | |
| rs4075127 | AGFG1 | √ | × | × | | rs6979497 | CNTNAP2 | √ | × | × | | rs4246651 | AGFG1 | × | √ | × | | rs7579575 | AGFG1 | × | √ | |
| rs12466158 | AGFG1 | √ | × | × | Height | rs7399503 | DIS3L2 | × | √ | × | | rs4075127 | AGFG1 | × | √ | × | | rs5847925 | AGFG1 | × | √ | |
| rs4485561 | AGFG1 | √ | × | × | | rs7399503 | DIS3L2 | × | √ | × | Lung funct | rs4485561 | AGFG1 | × | √ | × | | rs1020584 | AGFG1 | × | √ | |
| rs10933190 | AGFG1 | √ | × | × | | rs1262061 | DIS3L2 | × | √ | × | | rs1093319 | AGFG1 | × | √ | × | | rs1477938 | ANKRD31 | √ | × | |
| rs4508591 | AGFG1 | √ | × | × | | rs3100606 | DIS3L2 | × | √ | × | | rs4508591 | AGFG1 | × | √ | × | | rs7894187 | ANKRD31 | √ | × | |
| rs13019694 | AGFG1 | √ | × | × | | rs7680323 | DIS3L2 | √ | × | × | Height | rs1301969 | AGFG1 | × | √ | × | | rs7943520 | ANKRD31 | √ | × | |
| rs7584381 | AGFG1 | √ | × | × | | rs3103286 | DIS3L2 | × | √ | × | | rs7584381 | AGFG1 | × | √ | × | | rs7594163 | ANKRD31 | √ | × | |
| rs4246647 | AGFG1 | √ | × | × | | rs5578485 | FOLR4 | × | √ | × | | rs4246647 | AGFG1 | × | √ | × | | rs1150779 | C6orf1 | √ | × | Spleen volume |
| rs35511227 | AGFG1 | √ | × | × | | rs1880341 | FOLR4 | × | √ | × | | rs3551122 | AGFG1 | × | √ | × | | rs4704220 | COL4A3BI | √ | × | Cholesteryl esters to total lipids ratio in IDL |
| rs10189795 | AGFG1 | √ | × | × | Cortical su | rs2874531 | XRCC4 | √ | × | × | | rs1018979 | AGFG1 | × | √ | × | Cortical su | rs7548350 | DIS3L2 | √ | × | |
| rs10191552 | AGFG1 | √ | × | × | | rs7979481 | XRCC4 | √ | × | × | | rs1019155 | AGFG1 | × | √ | × | | rs2742077 | ECEL1 | × | √ | |
| rs71431054 | AGFG1 | √ | × | × | | rs2836019 | XRCC4 | √ | × | × | | rs7143105 | AGFG1 | × | √ | × | | rs2741282 | ECEL1 | × | √ | |
| rs6737495 | AGFG1 | √ | × | × | | rs1251514 | XRCC4 | √ | × | × | | rs6737495 | AGFG1 | × | √ | × | | rs1190451 | EIF4E2 | √ | × | |
| rs10185822 | AGFG1 | √ | × | × | | rs1690026 | XRCC4 | √ | × | × | | rs1018582 | AGFG1 | × | √ | × | | rs1190450 | EIF4E2 | √ | × | |
| rs4973499 | AGFG1 | √ | × | × | | rs1552909 | XRCC4 | √ | × | × | | rs4973499 | AGFG1 | × | √ | × | | rs1190448 | EIF4E2 | √ | × | |
| rs13035207 | AGFG1 | √ | × | × | | rs1562227 | XRCC4 | √ | × | × | | rs1303520 | AGFG1 | × | √ | × | | rs1201451 | EIF4E2 | √ | × | |
| rs7578871 | AGFG1 | √ | × | × | | rs3777033 | XRCC4 | √ | × | × | | rs7578871 | AGFG1 | × | √ | × | | rs1190447 | EIF4E2 | √ | × | |
| rs4365454 | AGFG1 | √ | × | × | | rs1687649 | XRCC4 | √ | × | × | | rs4365454 | AGFG1 | × | √ | × | | rs1868361 | LOC10265 | √ | × | 5-acetylamino-6-formylamino-3-methyluracil levels |
| rs4349340 | AGFG1 | √ | × | × | | rs1690030 | XRCC4 | √ | × | × | | rs4349340 | AGFG1 | × | √ | × | | rs2029056 | LOC10265 | √ | × | |
| rs6436700 | AGFG1 | √ | × | × | | rs6436698 | AGFG1 | √ | × | × | | rs6436700 | AGFG1 | × | √ | × | | rs1022514 | LOC10265 | √ | × | Cerebrospinal P-tau181p levels |
| rs7608104 | AGFG1 | √ | × | × | | rs1094272 | ANKRD31 | √ | × | × | | rs7608104 | AGFG1 | × | √ | × | | rs7588910 | PRKCE | √ | × | |
| rs7569200 | AGFG1 | √ | × | × | | rs9293642 | ANKRD31 | √ | × | × | | rs6707385 | AGFG1 | × | √ | × | | rs6867727 | TMEM167 | √ | × | |
| rs6707385 | AGFG1 | √ | × | × | | rs9293643 | ANKRD31 | √ | × | × | | rs3404411 | AGFG1 | × | √ | × | | rs1564211 | XRCC4 | √ | × | |
| rs34044110 | AGFG1 | √ | × | × | | rs7720620 | ANKRD31 | √ | × | × | | rs6731443 | AGFG1 | √ | × | × | Airway hyp | rs7703318 | XRCC4 | √ | × | |
| rs6731443 | AGFG1 | √ | × | × | Airway hyp | rs7833467 | ANKRD31 | × | √ | × | | rs3491559 | AGFG1 | √ | × | × | | rs6868817 | XRCC4 | √ | × | |
| rs34915597 | AGFG1 | √ | × | × | | rs4704171 | ANKRD31 | × | √ | × | Alzheimer's | rs1687243 | ANKRD31 | √ | × | × | | rs1690018 | XRCC4 | √ | × | |
| rs61046720 | AGFG1 | × | √ | × | | rs5634213 | ANKRD31 | × | √ | × | | rs7633533 | ANKRD31 | √ | × | × | | rs1051424 | XRCC4 | √ | × | |
| rs13388371 | AGFG1 | × | √ | × | | rs4621540 | ANKRD31 | √ | × | × | | rs9654427 | ANKRD31 | √ | × | × | | rs7399507 | DIS3L2 | √ | × | |
| rs563889066 | AGFG1 | × | √ | × | | rs7753246 | ANKRD31 | × | √ | × | | rs7162708 | ANKRD31 | √ | × | × | | rs4140925 | CNTNAP2 | √ | × | |
| rs142026388 | AGFG1 | × | √ | × | | rs4447963 | ANKRD31 | √ | × | × | | rs1809927 | LTBP1 | × | √ | × | | rs2538991 | CNTNAP2 | √ | × | |
| rs77835902 | AGFG1 | × | √ | × | | rs7717337 | ANKRD31 | √ | × | × | | rs958535 | XRCC4 | √ | × | × | | rs2538990 | CNTNAP2 | √ | × | |
| rs76009459 | AGFG1 | × | √ | × | | rs7735057 | ANKRD31 | √ | × | × | | rs3777021 | XRCC4 | √ | × | × | | rs2538989 | CNTNAP2 | √ | × | |
| rs140403861 | AGFG1 | × | √ | × | | rs4340901 | ANKRD31 | √ | × | × | | rs1017029 | AGFG1 | × | √ | × | | rs2247042 | CNTNAP2 | √ | × | |
| rs149717229 | AGFG1 | × | √ | × | | rs4361493 | ANKRD31 | √ | × | × | | rs7297881 | AGFG1 | × | √ | × | | rs2247043 | CNTNAP2 | √ | × | |
| rs34741105 | AGFG1 | × | √ | × | | rs6862740 | ANKRD31 | √ | × | × | | rs7555737 | AGFG1 | × | √ | × | | rs2538988 | CNTNAP2 | √ | × | |
| rs4635528 | AGFG1 | × | √ | × | | rs7722038 | ANKRD31 | × | √ | × | | rs7143104 | AGFG1 | × | √ | × | | rs2708243 | CNTNAP2 | √ | × | |
| rs572364588 | AGFG1 | × | √ | × | | rs6133429 | ANKRD31 | × | √ | × | | rs3437108 | AGFG1 | × | √ | × | | rs2538987 | CNTNAP2 | √ | × | |
| rs13408988 | AGFG1 | × | √ | × | | rs7875816 | ANKRD31 | × | √ | × | | rs1865016 | AGFG1 | × | √ | × | | rs2538986 | CNTNAP2 | √ | × | |
| rs12866 | AGFG1 | × | √ | × | | rs7486438 | ANKRD31 | × | √ | × | | rs5293149 | AGFG1 | × | √ | × | | rs2538985 | CNTNAP2 | √ | × | |
| rs1359 | AGFG1 | × | √ | × | | rs1336063 | ANKRD31 | √ | × | × | | rs1169312 | AGFG1 | × | √ | × | | rs2538983 | CNTNAP2 | √ | × | |
| rs36138959 | AGFG1 | × | √ | × | | rs1335914 | ANKRD31 | √ | × | × | | rs1834200 | AGFG1 | × | √ | × | | rs6896474 | ANKRD31 | √ | × | |
| rs13404414 | AGFG1 | × | √ | × | | rs5702399 | ANKRD31 | × | √ | × | | rs1807027 | AGFG1 | × | √ | × | | rs2837629 | ANKRD31 | √ | × | |
| rs542415135 | AGFG1 | × | √ | × | | rs9783263 | ANKRD31 | × | √ | × | | rs6742120 | AGFG1 | × | √ | × | | rs965735 | ANKRD31 | √ | × | |
| rs11686517 | AGFG1 | × | √ | × | | rs6893216 | ANKRD31 | × | √ | × | | rs1510499 | AGFG1 | × | √ | × | | rs1363577 | ANKRD31 | √ | × | |
| rs4324655 | ANKRD31 | √ | × | × | | rs1440184 | ANKRD31 | × | √ | × | | rs1162625 | AGFG1 | × | √ | × | | rs1422696 | ANKRD31 | √ | × | |
| rs6869345 | ANKRD31 | √ | × | × | | rs1407660 | ANKRD31 | × | √ | × | | rs7579575 | AGFG1 | × | √ | × | | rs9654426 | ANKRD31 | √ | × | |
| rs12523187 | ANKRD31 | √ | × | × | | rs1422697 | ANKRD31 | √ | × | × | | rs5847925 | AGFG1 | × | √ | × | | rs1177007 | CNTNAP2 | √ | × | |
| rs73127363 | ANKRD31 | √ | × | × | | rs7704104 | ANKRD31 | √ | × | × | | rs1020584 | AGFG1 | × | √ | × | | rs1186173 | CNTNAP2 | √ | × | |
| rs10462512 | ANKRD31 | √ | × | × | | rs2059159 | ANKRD31 | √ | × | × | | rs1484267 | AGFG1 | × | √ | × | | rs1723438 | CNTNAP2 | √ | × | |
| rs6453112 | ANKRD31 | √ | × | × | | rs2335107 | ANKRD31 | √ | × | × | | rs1838109 | AGFG1 | × | √ | × | | rs1262061 | DIS3L2 | √ | × | |
| rs4235683 | ANKRD31 | √ | × | × | | rs1046235 | ANKRD31 | √ | × | × | | rs7826031 | AGFG1 | × | √ | × | | rs7680323 | DIS3L2 | √ | × | Height |
| rs1903839 | ANKRD31 | √ | × | × | | rs1003652 | ANKRD31 | √ | × | × | | rs7460327 | AGFG1 | × | √ | × | | rs7486438 | ANKRD31 | √ | × | |
| rs4360021 | ANKRD31 | √ | × | × | | rs1003905 | ANKRD31 | √ | × | × | | rs7991008 | AGFG1 | × | √ | × | | rs6893216 | ANKRD31 | √ | × | |
| rs1477934 | ANKRD31 | √ | × | × | | rs6889429 | ANKRD31 | √ | × | × | | rs1919819 | AGFG1 | × | √ | × | | rs2643069 | CXADR | √ | × | |
| rs13177044 | ANKRD31 | √ | × | × | | rs4704188 | ANKRD31 | √ | × | × | | rs1019159 | AGFG1 | × | √ | × | | rs2739391 | CXADR | √ | × | |
| rs6896474 | ANKRD31 | √ | × | × | | rs1005094 | ANKRD31 | √ | × | × | | rs5633458 | AGFG1 | × | √ | × | | rs7399504 | DIS3L2 | √ | × | |
| rs28376290 | ANKRD31 | √ | × | × | | rs1345704 | ANKRD31 | √ | × | × | | rs1406636 | AGFG1 | × | √ | × | | rs1247331 | DIS3L2 | √ | × | |
| rs6872586 | ANKRD31 | √ | × | × | | rs9293650 | ANKRD31 | √ | × | × | | rs1340935 | AGFG1 | × | √ | × | | rs209691 | EPHA8 | √ | × | |
| rs965735 | ANKRD31 | √ | × | × | | rs7276872 | BTG3 | × | √ | × | | rs7143105 | AGFG1 | × | √ | × | | rs6884543 | XRCC4 | √ | × | |
| rs6885071 | ANKRD31 | √ | × | × | | rs1776091 | BTG3 | × | √ | × | | rs1018225 | AGFG1 | × | √ | × | | rs2126987 | XRCC4 | √ | × | |
| rs7711762 | ANKRD31 | √ | × | × | | rs2824388 | BTG3 | × | √ | × | | rs7715418 | AGFG1 | × | √ | × | | rs7754748 | XRCC4 | √ | × | |
| rs4703658 | ANKRD31 | √ | × | × | | rs1454680 | BTG3 | × | √ | × | | rs1339482 | AGFG1 | × | √ | × | | rs7588620 | AGFG1 | √ | × | |
| rs79964897 | ANKRD31 | √ | × | × | | rs1380992 | BTG3 | × | √ | × | | rs7958437 | AGFG1 | × | √ | × | | rs1093319 | AGFG1 | √ | × | |
| rs1363577 | ANKRD31 | √ | × | × | | rs2824389 | BTG3 | × | √ | × | | rs1135969 | AGFG1 | × | √ | × | | rs4973181 | AGFG1 | √ | × | |
| rs16872436 | ANKRD31 | √ | × | × | | rs2824391 | BTG3 | × | √ | × | | rs5747835 | AGFG1 | × | √ | × | | rs7598597 | AGFG1 | √ | × | |
| rs16872437 | ANKRD31 | √ | × | × | | rs2824392 | BTG3 | × | √ | × | | rs1823286 | AGFG1 | × | √ | × | | rs1342645 | AGFG1 | √ | × | |
| rs1422696 | ANKRD31 | √ | × | × | | rs2824393 | BTG3 | × | √ | × | | rs7551107 | AGFG1 | × | √ | × | | rs4555337 | AGFG1 | √ | × | |
| rs76335338 | ANKRD31 | √ | × | × | | rs2824394 | BTG3 | × | √ | × | | rs1931677 | AGFG1 | × | √ | × | | rs4312485 | AGFG1 | √ | × | |
| rs9654426 | ANKRD31 | √ | × | × | | rs2824395 | BTG3 | × | √ | × | | rs1120865 | ALPP | × | √ | × | | rs4075128 | AGFG1 | √ | × | |
| rs9654427 | ANKRD31 | √ | × | × | | rs2824396 | BTG3 | × | √ | × | | rs1406884 | ALPP | × | √ | × | | rs1324721 | CNTNAP2 | √ | × | |
| rs9654429 | ANKRD31 | √ | × | × | | rs2824397 | BTG3 | × | √ | × | | rs5709894 | ALPPL2 | × | √ | × | | rs1682857 | DIS3L2 | √ | × | |
| rs577226211 | ANKRD31 | √ | × | × | | rs3792566 | BTG3 | × | √ | × | | rs6728119 | ALPPL2 | × | √ | × | | rs1093338 | DIS3L2 | √ | × | |
| rs10051572 | ANKRD31 | √ | × | × | | rs5330739 | BTG3 | × | √ | × | | rs1439295 | ALPPL2 | × | √ | × | | rs4591730 | XRCC4 | √ | × | |
| rs1422701 | ANKRD31 | √ | × | × | | rs2824398 | BTG3 | × | √ | × | | rs1009934 | ALPPL2 | × | √ | × | | rs1773874 | ANKRD31 | √ | × | |
| rs2161371 | ANKRD31 | √ | × | × | | rs3702057 | BTG3 | × | √ | × | | rs6889839 | ANKRD31 | √ | × | × | | rs1767126 | ANKRD31 | √ | × | |
| rs58067979 | ANKRD31 | √ | × | × | | rs2824399 | BTG3 | × | √ | × | | rs4301212 | ANKRD31 | √ | × | × | | rs7504189 | ANKRD31 | √ | × | |
| rs2112874 | ANKRD31 | √ | × | × | | rs7455112 | BTG3 | × | √ | × | | rs7930753 | ANKRD31 | √ | × | × | | rs1150781 | C6orf1 | √ | × | Height |
| rs57646262 | ANKRD31 | √ | × | × | | rs7845865 | BTG3 | × | √ | × | | rs1477938 | ANKRD31 | √ | × | × | | rs7540696 | CXADR | × | √ | |
| rs71627084 | ANKRD31 | √ | × | × | | rs7597367 | BTG3 | × | √ | × | | rs7312903 | ANKRD31 | √ | × | × | | rs1479360 | CXADR | × | √ | |
| rs76693457 | BTG3 | × | × | √ | | rs1482744 | BTG3 | × | √ | × | | rs7894187 | ANKRD31 | √ | × | × | | rs2824338 | CXADR | × | √ | |
| rs59309307 | CASZ1 | × | × | √ | | rs1783403 | BTG3 | × | √ | × | | rs1687243 | ANKRD31 | √ | × | × | | rs1830982 | CXADR | × | √ | |
| rs3849280 | CASZ1 | × | × | √ | | rs1168648 | BTG3 | × | √ | × | | rs1047443 | ANKRD31 | √ | × | × | | rs1176496 | CXADR | × | √ | |
| rs17035713 | CASZ1 | × | × | √ | | rs1891176 | BTG3 | × | √ | × | | rs7943520 | ANKRD31 | √ | × | × | | rs8126543 | CXADR | × | √ | |
| rs851657 | CNTNAP2 | × | √ | × | | rs851712 | CNTNAP2 | √ | × | × | | rs7745441 | ANKRD31 | √ | × | × | | rs7583394 | CXADR | × | √ | |
| rs2178769 | CNTNAP2 | × | √ | × | | rs851707 | CNTNAP2 | √ | × | × | | rs7779310 | ANKRD31 | √ | × | × | | rs7827214 | CXADR | × | √ | |
| rs2192321 | CNTNAP2 | × | √ | × | | rs1024625 | CNTNAP2 | √ | × | × | | rs7594163 | ANKRD31 | √ | × | × | | rs1867224 | CXADR | × | √ | Mean corpuscular volume |
| rs2110306 | CNTNAP2 | × | √ | × | | rs1323581 | CNTNAP2 | √ | × | × | | rs4711389 | C6orf1 | √ | × | × | Type 2 diat | rs1863468 | CXADR | × | √ | |
| rs76688271 | CNTNAP2 | × | √ | × | | rs2643069 | CXADR | √ | × | × | | rs1150779 | C6orf1 | √ | × | × | Spleen volu | rs2824339 | CXADR | × | √ | |
| rs17170688 | CNTNAP2 | × | √ | × | | rs2739391 | CXADR | √ | × | × | | rs4845952 | CASZ1 | × | × | √ | | rs6517779 | CXADR | × | √ | |
| rs79755166 | CNTNAP2 | × | √ | × | | rs2249449 | CXADR | √ | × | × | | rs2245601 | CHRND | √ | × | × | Corneal cu | rs2824340 | CXADR | × | √ | |
| rs144136510 | CNTNAP2 | × | √ | × | | rs1783728 | DACH1 | × | × | √ | | rs851708 | CNTNAP2 | √ | × | × | | rs764657 | CXADR | √ | × | |
| rs79945735 | CNTNAP2 | × | √ | × | | rs7512804 | DACH1 | × | × | √ | | rs3827608 | COL4A3BI | √ | × | × | | rs211978 | CXADR | √ | × | |
| rs75875112 | CNTNAP2 | × | √ | × | | rs7713319 | DACH1 | × | × | √ | | rs7656771 | COL4A3BI | √ | × | × | | rs7506312 | DIS3L2 | √ | × | |
| rs77627573 | CNTNAP2 | × | √ | × | | rs9564850 | DACH1 | × | × | √ | | rs1205533 | COL4A3BI | √ | × | × | | rs7553245 | DIS3L2 | √ | × | |
| rs117757285 | CNTNAP2 | × | √ | × | | rs9572817 | DACH1 | √ | × | × | | rs6029884 | COL4A3BI | √ | × | × | | rs909431 | ECEL1 | × | √ | |
| rs1357903 | CNTNAP2 | × | √ | × | | rs3100611 | DIS3L2 | × | √ | × | | rs1767184 | COL4A3BI | √ | × | × | Metabolite | rs7834848 | ECEL1 | × | √ | |
| rs13238480 | CNTNAP2 | × | √ | × | | rs6722553 | DIS3L2 | × | √ | × | | rs1051519 | COL4A3BI | √ | × | × | | rs2573246 | ECEL1 | × | √ | |
| rs77658347 | CNTNAP2 | × | √ | × | | rs1982474 | DIS3L2 | × | √ | × | | rs1687255 | COL4A3BI | √ | × | × | | rs7442655 | LOC10265 | √ | × | |
| rs851715 | CNTNAP2 | × | √ | × | | rs3714530 | DIS3L2 | × | √ | × | | rs7659187 | COL4A3BI | √ | × | × | | rs7457038 | LTBP1 | √ | × | |
| rs367930375 | CNTNAP2 | × | √ | × | | rs3762062 | DIS3L2 | × | √ | × | | rs1687255 | COL4A3BI | √ | × | × | | rs6171215 | NPAS3 | √ | × | |
| rs17825525 | CNTNAP2 | × | √ | × | | rs1168934 | DIS3L2 | × | √ | × | | rs5608215 | COL4A3BI | √ | × | × | | rs7592046 | NPAS3 | √ | × | |
| rs851713 | CNTNAP2 | × | √ | × | | rs3100620 | DIS3L2 | × | √ | × | | rs7376309 | COL4A3BI | √ | × | × | | rs2595210 | PRKCE | √ | × | |
| rs17227485 | CNTNAP2 | × | √ | × | | rs3100621 | DIS3L2 | × | √ | × | | rs1454409 | COL4A3BI | √ | × | × | | rs1455423 | PRKCE | √ | × | |
| rs189958191 | CNTNAP2 | × | √ | × | | rs5622583 | DIS3L2 | × | √ | × | | rs1437960 | COL4A3BI | √ | × | × | | rs7349397 | PRKCE | √ | × | |
| rs12532085 | CNTNAP2 | × | √ | × | | rs7298567 | DIS3L2 | × | √ | × | | rs7990177 | COL4A3BI | √ | × | × | | rs7280673 | PRKCE | √ | × | |

| | | | | |
|---|---|---|---|---|
| rs12540078 | CNTNAP2 | × | √ | × |
| rs77525968 | CNTNAP2 | × | √ | × |
| rs116932967 | CNTNAP2 | × | √ | × |
| rs13247212 | CNTNAP2 | × | √ | × |
| rs146416937 | CNTNAP2 | × | √ | × |
| rs1177007 | CNTNAP2 | × | √ | × |
| rs1186173 | CNTNAP2 | × | √ | × |
| rs12532478 | CNTNAP2 | × | √ | × |
| rs762853939 | CNTNAP2 | × | √ | × |
| rs1723438 | CNTNAP2 | × | √ | × |
| rs12154890 | CNTNAP2 | × | √ | × |
| rs12155079 | CNTNAP2 | × | √ | × |
| rs1177009 | CNTNAP2 | × | √ | × |
| rs75298126 | CNTNAP2 | × | √ | × |
| rs62482345 | CNTNAP2 | × | √ | × |
| rs143834093 | CNTNAP2 | × | √ | × |
| rs17170699 | CNTNAP2 | × | √ | × |
| rs116425899 | CNTNAP2 | × | √ | × |
| rs118104701 | CNTNAP2 | × | √ | × |
| rs2110308 | CNTNAP2 | × | √ | × |
| rs2011815 | CNTNAP2 | × | √ | × |
| rs79579313 | CNTNAP2 | × | √ | × |
| rs117348836 | CNTNAP2 | × | √ | × |
| rs34103397 | CNTNAP2 | × | √ | × |
| rs55714492 | CNTNAP2 | × | √ | × |
| rs17825832 | CNTNAP2 | × | √ | × |
| rs182873181 | CNTNAP2 | × | √ | × |
| rs17234187 | CNTNAP2 | × | √ | × |
| rs74798213 | CNTNAP2 | × | √ | × |
| rs75952833 | CNTNAP2 | × | √ | × |
| rs17825924 | CNTNAP2 | × | √ | × |
| rs560863471 | CNTNAP2 | × | √ | × |
| rs55932730 | CNTNAP2 | × | √ | × |
| rs76449654 | CNTNAP2 | × | √ | × |
| rs147617615 | CNTNAP2 | × | √ | × |
| rs12530803 | CNTNAP2 | × | √ | × |
| rs12530825 | CNTNAP2 | × | √ | × |
| rs189316925 | CNTNAP2 | × | √ | × |
| rs79560675 | CNTNAP2 | × | √ | × |
| rs77070965 | CNTNAP2 | × | √ | × |
| rs6464842 | CNTNAP2 | × | √ | × |
| rs7788047 | CNTNAP2 | × | √ | × |
| rs6979528 | CNTNAP2 | × | √ | × |
| rs76096669 | CNTNAP2 | × | √ | × |
| rs6959742 | CNTNAP2 | × | √ | × |
| rs6979497 | CNTNAP2 | × | √ | × |
| rs4305811 | CNTNAP2 | × | √ | × |
| rs62481196 | CNTNAP2 | × | √ | × |
| rs140800453 | CNTNAP2 | × | √ | × |
| rs11765619 | CNTNAP2 | × | √ | × |
| rs10952715 | CNTNAP2 | × | √ | × |
| rs7794345 | CNTNAP2 | × | √ | × |
| rs10252313 | CNTNAP2 | × | √ | × |
| rs142589650 | CNTNAP2 | × | √ | × |
| rs150952558 | CNTNAP2 | × | √ | × |
| rs139830888 | CNTNAP2 | × | √ | × |
| rs10085572 | CNTNAP2 | × | √ | × |
| rs2710139 | CNTNAP2 | × | √ | × |
| rs13234998 | CNTNAP2 | × | √ | × |
| rs56268129 | CNTNAP2 | × | √ | × |
| rs10254338 | CNTNAP2 | × | √ | × |
| rs2710100 | CNTNAP2 | × | √ | × |
| rs75727838 | CNTNAP2 | × | √ | × |
| rs6946545 | CNTNAP2 | × | √ | × |
| rs76298264 | CNTNAP2 | × | √ | × |
| rs79321054 | CNTNAP2 | × | √ | × |
| rs73166262 | CNTNAP2 | × | √ | × |
| rs79962231 | CNTNAP2 | × | √ | × |
| rs10952716 | CNTNAP2 | × | √ | × |
| rs17827310 | CNTNAP2 | × | √ | × |
| rs4397315 | CNTNAP2 | × | √ | × |
| rs116996718 | CNTNAP2 | × | √ | × |
| rs118114254 | CNTNAP2 | × | √ | × |
| rs116916385 | CNTNAP2 | × | √ | × |
| rs2708244 | CNTNAP2 | × | √ | × |
| rs17236239 | CNTNAP2 | × | √ | × |
| rs192854872 | CNTNAP2 | × | √ | × |
| rs11980029 | CNTNAP2 | × | √ | × |
| rs369554431 | CNTNAP2 | × | √ | × |
| rs117894541 | CNTNAP2 | × | √ | × |
| rs10264316 | CNTNAP2 | × | √ | × |
| rs73743371 | CNTNAP2 | × | √ | × |
| rs55717999 | CNTNAP2 | × | √ | × |
| rs10278677 | CNTNAP2 | × | √ | × |
| rs57921419 | CNTNAP2 | × | √ | × |
| rs199863730 | CXADR | √ | × | × |
| rs73995036 | DIS3L2 | √ | × | × |
| rs73995038 | DIS3L2 | √ | × | × |
| rs12620618 | DIS3L2 | √ | × | × |
| rs78316317 | DIS3L2 | √ | × | × |
| rs3100606 | DIS3L2 | √ | × | × |
| rs16828571 | DIS3L2 | √ | × | × |
| rs10933385 | DIS3L2 | √ | × | × |
| rs73995074 | DIS3L2 | × | × | √ |
| rs76803230 | DIS3L2 | √ | × | × |
| rs3100624 | DIS3L2 | √ | × | × |
| rs3100584 | DIS3L2 | √ | × | × |
| rs145298893 | DIS3L2 | √ | × | × |
| rs16828632 | DIS3L2 | × | × | √ |
| rs1869457 | DIS3L2 | √ | × | × |
| rs3103261 | DIS3L2 | √ | × | × |
| rs2880324 | DIS3L2 | √ | × | × |
| rs3116174 | DIS3L2 | √ | × | × |
| rs3103286 | DIS3L2 | √ | × | × |
| rs4973513 | DIS3L2 | √ | × | × |
| rs4973514 | DIS3L2 | √ | × | × |
| rs74269651 | DIS3L2 | √ | × | × |
| rs1190456 | EIF4E2 | √ | × | × |
| rs10193827 | EPAS1 | × | × | √ |
| rs7582701 | EPAS1 | × | × | √ |
| rs55784852 | FOLR4 | × | √ | × |
| rs188034174 | FOLR4 | × | √ | × |
| rs10268888 | LOC101927630 | × | √ | × |
| rs116798463 | LOC101927630 | × | √ | × |
| rs142849695 | LOC101927630 | × | √ | × |
| rs6965300 | LOC101927630 | × | √ | × |
| rs1448653 | LOC101927630 | × | √ | × |
| rs370280053 | LOC101927630 | × | √ | × |
| rs1448654 | LOC101927630 | × | √ | × |
| rs4721627 | LOC101927630 | × | √ | × |
| rs4721628 | LOC101927630 | × | √ | × |
| rs13225976 | LOC102659288 | √ | × | × |
| rs180992764 | LTBP1 | √ | × | × |
| rs76390075 | LTBP1 | √ | × | × |
| rs75486357 | LTBP1 | √ | × | × |
| rs3820912 | LTBP1 | √ | × | × |
| rs3754830 | LTBP1 | √ | × | × |
| rs10150948 | NPAS3 | √ | × | × |
| rs184677330 | NPAS3 | √ | × | × |
| rs75700106 | NPAS3 | √ | × | × |
| rs79540422 | NPAS3 | √ | × | × |
| rs75870771 | PRKCE | √ | × | × |
| rs35332414 | PRKCE | √ | × | × |
| rs13024907 | PRKCE | √ | × | × |
| rs77489679 | PRKCE | × | × | √ |
| rs114263453 | PRKCE | × | × | √ |
| rs733602 | PRSS56 | √ | × | × |
| rs28745310 | XRCC4 | √ | × | × |
| rs4591730 | XRCC4 | √ | × | × |
| rs79794819 | XRCC4 | √ | × | × |
| rs28360199 | XRCC4 | √ | × | × |
| rs12515149 | XRCC4 | √ | × | × |
| rs16900261 | XRCC4 | √ | × | × |
| rs1552909 | XRCC4 | √ | × | × |
| rs1562227 | XRCC4 | √ | × | × |
| rs3777033 | XRCC4 | √ | × | × |
| rs16876496 | XRCC4 | √ | × | × |
| rs16900303 | XRCC4 | √ | × | × |
| rs958535 | XRCC4 | √ | × | × |
| rs3777021 | XRCC4 | √ | × | × |

| | | | | | |
|---|---|---|---|---|---|
| | rs1488589 | DIS3L2 | × | √ | × |
| | rs7787419 | DIS3L2 | × | √ | × |
| | rs5605791 | DIS3L2 | × | √ | × |
| | rs7298567 | DIS3L2 | × | √ | × |
| | rs1338462 | DIS3L2 | × | √ | × |
| | rs3116154 | DIS3L2 | × | √ | × |
| | rs3116160 | DIS3L2 | × | √ | × |
| | rs7298568 | DIS3L2 | × | √ | × |
| | rs1339178 | DIS3L2 | × | √ | × |
| | rs3116162 | DIS3L2 | × | √ | × |
| | rs7298568 | DIS3L2 | × | √ | × |
| | rs1873722 | DIS3L2 | × | √ | × |
| | rs1425487 | DIS3L2 | × | √ | × |
| | rs7298569 | DIS3L2 | × | √ | × |
| | rs7298570 | DIS3L2 | × | √ | × |
| | rs6745958 | DIS3L2 | × | √ | × |
| | rs4973026 | DIS3L2 | × | √ | × |
| | rs1168912 | DIS3L2 | × | √ | × |
| | rs1247043 | DIS3L2 | × | √ | × |
| | rs3116218 | DIS3L2 | × | √ | × |
| | rs3100602 | DIS3L2 | × | √ | × |
| | rs3116222 | DIS3L2 | × | √ | × |
| | rs7399504 | DIS3L2 | × | √ | × |
| | rs1247933 | DIS3L2 | × | √ | × |
| | rs5879601 | DIS3L2 | × | √ | × |
| | rs5568396 | DIS3L2 | × | √ | × |
| | rs5512330 | DIS3L2 | × | √ | × |
| | rs3100603 | DIS3L2 | × | √ | × |
| | rs1169542 | DIS3L2 | × | √ | × |
| | rs1169555 | DIS3L2 | × | √ | × |
| | rs1142893 | DIS3L2 | × | √ | × |
| | rs1188582 | DIS3L2 | × | √ | × |
| | rs3116225 | DIS3L2 | × | √ | × |
| | rs1247331 | DIS3L2 | × | √ | × |
| | rs5967944 | DIS3L2 | × | √ | × |
| | rs3100617 | DIS3L2 | × | √ | × |
| | rs1430681 | DIS3L2 | √ | × | × |
| | rs1501805 | DIS3L2 | × | √ | × |
| | rs1574528 | DIS3L2 | × | √ | × |
| | rs5535621 | DIS3L2 | × | √ | × |
| | rs3744290 | DIS3L2 | × | √ | × |
| | rs3116172 | DIS3L2 | × | √ | × |
| | rs3103258 | DIS3L2 | × | √ | × |
| | rs2880323 | DIS3L2 | × | √ | × |
| | rs1840876 | DIS3L2 | × | √ | × |
| | rs3769872 | DIS3L2 | × | √ | × |
| | rs7300313 | DIS3L2 | × | √ | × |
| | rs3103257 | DIS3L2 | × | √ | × |
| | rs3116173 | DIS3L2 | × | √ | × |
| | rs3116185 | DIS3L2 | × | √ | × |
| | rs1093338 | DIS3L2 | × | √ | × |
| | rs3116188 | DIS3L2 | × | √ | × |
| | rs1341659 | DIS3L2 | × | √ | × |
| | rs7945626 | DIS3L2 | × | √ | × |
| | rs7299231 | DIS3L2 | × | √ | × |
| | rs3116189 | DIS3L2 | × | √ | × |
| | rs3116190 | DIS3L2 | × | √ | × |
| | rs3116191 | DIS3L2 | × | √ | × |
| | rs3547490 | DIS3L2 | × | √ | × |
| | rs1343788 | DIS3L2 | × | √ | × |
| | rs7299233 | DIS3L2 | × | √ | × |
| | rs7299233 | DIS3L2 | × | √ | × |
| | rs7603448 | DIS3L2 | × | √ | × |
| | rs1049826 | DIS3L2 | × | √ | × |
| | rs7299235 | DIS3L2 | × | √ | × |
| | rs7299235 | DIS3L2 | × | √ | × |
| | rs1682866 | DIS3L2 | × | √ | × |
| | rs3103290 | DIS3L2 | × | √ | × |
| | rs3116193 | DIS3L2 | × | √ | × |
| | rs3116194 | DIS3L2 | × | √ | × |
| | rs3103289 | DIS3L2 | × | √ | × |
| | rs3116196 | DIS3L2 | × | √ | × |
| | rs1156539 | DIS3L2 | × | √ | × |
| | rs1817873 | DIS3L2 | × | √ | × |
| | rs1887985 | DIS3L2 | × | √ | × |
| | rs6742242 | DIS3L2 | × | √ | × |
| | rs4973508 | DIS3L2 | × | √ | × |
| | rs7686173 | DIS3L2 | × | √ | × |
| | rs1340326 | DIS3L2 | × | √ | × |
| | rs1167545 | DIS3L2 | × | √ | × |
| | rs998985 | DIS3L2 | × | √ | × |
| | rs3116201 | DIS3L2 | × | √ | × |
| | rs5875230 | DIS3L2 | × | √ | × |
| | rs5612841 | DIS3L2 | × | √ | × |
| | rs7299237 | DIS3L2 | × | √ | × |
| | rs7299237 | DIS3L2 | × | √ | × |
| | rs1153974 | DIS3L2 | × | √ | × |
| Lung funct | rs1246604 | DIS3L2 | × | √ | × |
| | rs2129777 | DIS3L2 | × | √ | × |
| | rs5662161 | DIS3L2 | × | √ | × |
| | rs7602768 | DIS3L2 | × | √ | × |
| | rs1169191 | DIS3L2 | × | √ | × |
| | rs1190197 | DIS3L2 | × | √ | × |
| | rs1817903 | DIS3L2 | × | √ | × |
| Height | rs923333 | DIS3L2 | × | √ | × |
| | rs1431162 | DIS3L2 | × | √ | × |
| | rs1168986 | DIS3L2 | × | √ | × |
| | rs5598984 | DIS3L2 | × | √ | × |
| | rs7299425 | DIS3L2 | × | √ | × |
| | rs1432852 | DIS3L2 | × | √ | × |
| Whole bod | rs3725662 | DIS3L2 | × | √ | × |
| | rs7299425 | DIS3L2 | × | √ | × |
| | rs7299425 | DIS3L2 | × | √ | × |
| | rs7299426 | DIS3L2 | × | √ | × |
| | rs6125034 | DIS3L2 | × | √ | × |
| | rs7574893 | DIS3L2 | × | √ | × |
| | rs1916720 | DIS3L2 | × | √ | × |
| | rs1156412 | DIS3L2 | × | √ | × |
| | rs1839123 | DIS3L2 | × | √ | × |
| IgG glycosy | rs5613160 | DIS3L2 | × | √ | × |
| | rs7688371 | DIS3L2 | × | √ | × |
| | rs7299427 | DIS3L2 | × | √ | × |
| | rs7579970 | DIS3L2 | × | √ | × |
| | rs1246658 | DIS3L2 | × | √ | × |
| | rs6437055 | DIS3L2 | × | √ | × |
| | rs1397754 | DIS3L2 | × | √ | × |
| | rs7299428 | DIS3L2 | × | √ | × |
| | rs7299428 | DIS3L2 | × | √ | × |
| | rs3755099 | DIS3L2 | × | √ | × |
| | rs5394771 | DIS3L2 | × | √ | × |
| | rs1414331 | DIS3L2 | × | √ | × |
| | rs1190438 | DIS3L2 | × | √ | × |
| | rs1411283 | DIS3L2 | × | √ | × |
| | rs7299429 | DIS3L2 | × | √ | × |
| | rs7299429 | DIS3L2 | × | √ | × |
| | rs2087229 | DIS3L2 | × | √ | × |
| Height | rs2853358 | DIS3L2 | × | √ | × |
| | rs7664883 | DIS3L2 | × | √ | × |
| | rs1865912 | DIS3L2 | × | √ | × |
| | rs1818392 | DIS3L2 | × | √ | × |
| | rs3484539 | DIS3L2 | × | √ | × |
| | rs1169596 | DIS3L2 | × | √ | × |
| | rs1403667 | DIS3L2 | × | √ | × |
| | rs2101435 | DIS3L2 | × | √ | × |
| | rs2853361 | DIS3L2 | × | √ | × |
| | rs1168614 | DIS3L2 | × | √ | × |
| | rs7893383 | DIS3L2 | × | √ | × |
| | rs2853362 | DIS3L2 | × | √ | × |
| | rs2633265 | DIS3L2 | × | √ | × |
| | rs2633264 | DIS3L2 | × | √ | × |
| | rs4973038 | DIS3L2 | × | √ | × |
| | rs1262234 | DIS3L2 | × | √ | × |
| | rs1838174 | DIS3L2 | × | √ | × |
| | rs1190307 | DIS3L2 | × | √ | × |
| | rs1247110 | DIS3L2 | × | √ | × |
| | rs2633263 | DIS3L2 | × | √ | × |
| | rs1682873 | DIS3L2 | × | √ | × |
| | rs1484740 | DIS3L2 | × | √ | × |
| | rs7299811 | DIS3L2 | × | √ | × |
| | rs1300474 | DIS3L2 | × | √ | × |
| | rs1491478 | DIS3L2 | × | √ | × |
| | rs2633262 | DIS3L2 | × | √ | × |
| | rs2633261 | DIS3L2 | × | √ | × |
| | rs7299812 | DIS3L2 | × | √ | × |
| | rs1682873 | DIS3L2 | × | √ | × |
| | rs2633260 | DIS3L2 | × | √ | × |
| | rs1390173 | DIS3L2 | × | √ | × |
| | rs1443778 | DIS3L2 | × | √ | × |
| | rs7299813 | DIS3L2 | × | √ | × |
| | rs7598284 | DIS3L2 | × | √ | × |
| | rs1146931 | DIS3L2 | × | √ | × |
| | rs1416033 | DIS3L2 | × | √ | × |
| | rs2633254 | DIS3L2 | × | √ | × |
| | rs2837182 | DIS3L2 | × | √ | × |
| | rs7599108 | DIS3L2 | × | √ | × |
| | rs1832288 | DIS3L2 | × | √ | × |
| | rs1474407 | DIS3L2 | × | √ | × |
| | rs1499438 | DIS3L2 | × | √ | × |
| | rs1169364 | DIS3L2 | × | √ | × |
| | rs7573322 | DIS3L2 | × | √ | × |
| | rs1247617 | DIS3L2 | × | √ | × |
| | rs3116224 | DIS3L2 | × | √ | × |
| | rs4973039 | DIS3L2 | × | √ | × |
| | rs7963152 | DIS3L2 | × | √ | × |

| | | | | | | |
|---|---|---|---|---|---|---|
| | rs7376309 | COL4A3BP | √ | × | × | |
| | rs7376310 | COL4A3BP | √ | × | × | |
| | rs1464270 | COL4A3BP | √ | × | × | |
| | rs1441958 | COL4A3BP | √ | × | × | |
| | rs1389727 | COL4A3BP | √ | × | × | |
| | rs7952143 | COL4A3BP | √ | × | × | |
| | rs7376310 | COL4A3BP | √ | × | × | |
| | rs7466238 | COL4A3BP | √ | × | × | |
| | rs1171476 | COL4A3BP | √ | × | × | |
| Waist circu | rs5636126 | COL4A3BP | √ | × | × | |
| | rs1382197 | COL4A3BP | √ | × | × | |
| | rs9791146 | COL4A3BP | √ | × | × | |
| | rs7376490 | COL4A3BP | √ | × | × | |
| | rs6108064 | COL4A3BP | √ | × | × | |
| | rs4704219 | COL4A3BP | √ | × | × | |
| | rs7376490 | COL4A3BP | √ | × | × | |
| | rs7376490 | COL4A3BP | √ | × | × | |
| | rs4704220 | COL4A3BP | √ | × | × | |
| | rs4704221 | COL4A3BP | √ | × | × | |
| | rs7376491 | COL4A3BP | √ | × | × | |
| | rs7376491 | COL4A3BP | √ | × | × | |
| | rs6100156 | COL4A3BP | √ | × | × | |
| | rs7526951 | COL4A3BP | √ | × | × | |
| | rs7376492 | COL4A3BP | √ | × | × | |
| | rs7376492 | COL4A3BP | √ | × | × | |
| | rs5936713 | COL4A3BP | √ | × | × | |
| | rs7376492 | COL4A3BP | √ | × | × | |
| | rs7926790 | COL4A3BP | √ | × | × | |
| | rs7930299 | DACH1 | × | × | √ | |
| | rs9542760 | DACH1 | √ | × | × | |
| | rs7878430 | DIS3L2 | √ | × | × | |
| | rs1188666 | DIS3L2 | √ | × | × | |
| Height | rs1682856 | DIS3L2 | √ | × | × | |
| | rs6042176 | DIS3L2 | √ | × | × | |
| | rs1247532 | DIS3L2 | √ | × | × | |
| | rs3611114 | DIS3L2 | √ | × | × | |
| | rs7808399 | DIS3L2 | √ | × | × | |
| | rs7450041 | DIS3L2 | √ | × | × | |
| | rs1247537 | DIS3L2 | √ | × | × | |
| | rs7548350 | DIS3L2 | √ | × | × | |
| | rs8005439 | DIS3L2 | √ | × | × | |
| | rs1246611 | DIS3L2 | √ | × | × | |
| | rs7308783 | DIS3L2 | √ | × | × | |
| | rs7308783 | DIS3L2 | √ | × | × | |
| | rs7903650 | DIS3L2 | √ | × | × | |
| | rs7878999 | DIS3L2 | √ | × | × | |
| | rs1188718 | DIS3L2 | √ | × | × | |
| | rs3116157 | DIS3L2 | √ | × | × | |
| | rs1247854 | DIS3L2 | √ | × | × | |
| | rs7575236 | DIS3L2 | √ | × | × | |
| | rs5883206 | DIS3L2 | √ | × | × | |
| | rs1246341 | DIS3L2 | √ | × | × | |
| | rs1727205 | DIS3L2 | √ | × | × | |
| | rs2742077 | ECEL1 | √ | × | × | |
| | rs2741282 | ECEL1 | √ | × | × | |
| | rs1190451 | EIF4E2 | √ | × | × | |
| | rs1190450 | EIF4E2 | √ | × | × | |
| | rs1190448 | EIF4E2 | √ | × | × | |
| | rs1201451 | EIF4E2 | √ | × | × | |
| | rs1190447 | EIF4E2 | √ | × | × | |
| | rs1190440 | EIF4E2 | √ | × | × | |
| | rs1190439 | EIF4E2 | √ | × | × | |
| | rs3108614 | EIF4E2 | √ | × | × | |
| Height | rs3791715 | EIF4E2 | √ | × | × | |
| | rs7527700 | EIF4E2 | √ | × | × | |
| | rs7585327 | EIF4E2 | √ | × | × | |
| | rs7930959 | EIF4E2 | √ | × | × | |
| | rs1180122 | HMGA1 | √ | × | × | |
| | rs4713761 | HMGA1 | √ | × | × | |
| | rs5665795 | HMGA1 | √ | × | × | |
| | rs2239897 | KCNQ1DN | × | × | √ | |
| | rs8032385 | KIF26B | √ | × | × | |
| | rs1202655 | KIF26B | √ | × | × | |
| | rs1203360 | KIF26B | √ | × | × | |
| | rs1203459 | KIF26B | √ | × | × | |
| | rs1275342 | KIF26B | √ | × | × | |
| | rs1092415 | KIF26B | √ | × | × | |
| | rs1273642 | KIF26B | √ | × | × | |
| | rs1202823 | KIF26B | √ | × | × | |
| | rs3606090 | KIF26B | √ | × | × | |
| | rs1171871 | KIF26B | √ | × | × | |
| Appendicul | rs1181168 | KIF26B | √ | × | × | |
| | rs1155845 | LOC10265 | √ | × | × | |
| | rs1868361 | LOC10265 | √ | × | × | |
| | rs2029056 | LOC10265 | √ | × | × | |
| | rs1687157 | LOC10265 | √ | × | × | |
| | rs1022514 | LOC10265 | √ | × | × | |
| | rs3769528 | LTBP1 | × | √ | × | |
| | rs711234 | LTBP1 | × | √ | × | |
| | rs7817756 | LTBP1 | × | √ | × | |
| | rs1262029 | LTBP1 | × | √ | × | |
| | rs7963590 | LTBP1 | × | √ | × | |
| | rs817534 | LTBP1 | × | √ | × | |
| | rs921705 | LTBP1 | × | √ | × | |
| | rs3769530 | LTBP1 | × | √ | × | |
| | rs7827058 | LTBP1 | × | √ | × | |
| | rs1412759 | LTBP1 | × | √ | × | |
| | rs7467205 | LTBP1 | × | √ | × | |
| | rs8031094 | LTBP1 | × | √ | × | |
| | rs3731575 | LTBP1 | × | √ | × | |
| | rs7795218 | LTBP1 | × | √ | × | |
| | rs7813934 | LTBP1 | × | √ | × | |
| | rs1169188 | LTBP1 | × | √ | × | |
| | rs1051477 | LTBP1 | × | √ | × | |
| | rs5650899 | LTBP1 | × | √ | × | |
| Height | rs1168696 | LTBP1 | × | √ | × | |
| | rs7993377 | LTBP1 | × | √ | × | |
| | rs7431971 | LTBP1 | × | √ | × | |
| | rs7569139 | LTBP1 | × | √ | × | |
| | rs6175424 | LTBP1 | × | √ | × | |
| | rs1811668 | LTBP1 | × | √ | × | |
| | rs1167570 | LTBP1 | × | √ | × | |
| | rs3769532 | LTBP1 | × | √ | × | |
| Hip circum | rs817532 | LTBP1 | × | √ | × | |
| Waist circu | rs3769534 | LTBP1 | × | √ | × | |
| | rs3769535 | LTBP1 | × | √ | × | |
| | rs1829120 | LTBP1 | × | √ | × | |
| | rs1168957 | LTBP1 | × | √ | × | |
| | rs3769536 | LTBP1 | × | √ | × | |
| | rs1161932 | LTBP1 | × | √ | × | |
| | rs1871224 | LTBP1 | × | √ | × | |
| | rs1510576 | LTBP1 | × | √ | × | |
| | rs1467575 | LTBP1 | × | √ | × | |
| | rs1246809 | LTBP1 | × | √ | × | G_Coprococcus prevalence |
| | rs7805592 | LTBP1 | × | √ | × | |
| | rs971793 | LTBP1 | × | √ | × | |
| | rs1141736 | LTBP1 | × | √ | × | |
| | rs5585236 | LTBP1 | × | √ | × | |
| | rs1453845 | LTBP1 | × | √ | × | |
| | rs1758193 | LTBP1 | × | √ | × | |
| | rs8009097 | LTBP1 | × | √ | × | |
| | rs817529 | LTBP1 | × | √ | × | |
| | rs817531 | LTBP1 | × | √ | × | |
| Waist circu | rs3754829 | LTBP1 | × | √ | × | Waist circumference adjusted for body mass index |
| Tibia length | rs6214888 | LTBP1 | × | √ | × | |
| | rs2305503 | LTBP1 | × | √ | × | |
| | rs3816479 | LTBP1 | × | √ | × | |
| Multi-trait s | rs1811109 | LTBP1 | × | √ | × | |
| | rs554715 | MRE11A | √ | × | × | |
| | rs604845 | MRE11A | √ | × | × | |
| | rs4246978 | NPAS3 | √ | × | × | |
| | rs7614043 | NPAS3 | √ | × | × | |
| | rs1829011 | NPAS3 | √ | × | × | |
| | rs4549504 | POLK | √ | × | × | |
| | rs1859143 | POLK | √ | × | × | |
| | rs5744552 | POLK | √ | × | × | Total cholines levels |
| | rs5744557 | POLK | √ | × | × | |
| | rs7376492 | POLK | √ | × | × | |
| | rs5744566 | POLK | √ | × | × | |
| | rs5744567 | POLK | √ | × | × | |
| | rs1687258 | POLK | √ | × | × | |
| | rs6085410 | POLK | √ | × | × | |
| Whole bod | rs5744578 | POLK | √ | × | × | |
| | rs5744591 | POLK | √ | × | × | |
| | rs5744612 | POLK | √ | × | × | |
| | rs7376492 | POLK | √ | × | × | |
| | rs1988728 | POLK | √ | × | × | |
| | rs5744661 | POLK | √ | × | × | |
| | rs1018119 | POLK | √ | × | × | |
| | rs767677 | POLK | √ | × | × | |
| | rs3685300 | PRKCE | √ | × | × | |
| | rs3701627 | PRKCE | √ | × | × | |
| | rs1389649 | PRKCE | √ | × | × | |
| | rs3738686 | PRKCE | √ | × | × | |
| | rs3708369 | PRKCE | √ | × | × | |
| | rs3750847 | PRKCE | √ | × | × | |
| | rs7588910 | PRKCE | √ | × | × | |
| | rs3749964 | PRKCE | √ | × | × | |
| | rs3757085 | PRKCE | √ | × | × | |
| | rs3767231 | PRKCE | √ | × | × | |
| | rs3476897 | PRKCE | √ | × | × | |
| | rs3693855 | PRKCE | √ | × | × | |
| | rs3738590 | PRKCE | √ | × | × | |
| | rs1470598 | PRKCE | √ | × | × | |

| | | | | |
|---|---|---|---|---|
| | rs3754569 | PRKCE | √ | × |
| | rs3001978 | RPTN | × | √ |
| | rs7549782 | RPTN | × | √ |
| | rs2902665 | SIGLEC6 | √ | × |
| | rs2168643 | SIGLEC6 | √ | × |
| | rs4801874 | SIGLEC6 | √ | × |
| | rs6896548 | XRCC4 | √ | × |
| | rs6896953 | XRCC4 | √ | × |
| | rs1011979 | XRCC4 | √ | × |
| | rs1728421 | XRCC4 | √ | × |
| | rs1382374 | XRCC4 | √ | × |
| Collagen ty | rs1007161 | XRCC4 | √ | × |
| | rs1382373 | XRCC4 | √ | × |
| | rs1007605 | XRCC4 | √ | × |
| Body mass | rs1195125 | XRCC4 | √ | × |
| | rs767771 | XRCC4 | √ | × |
| | rs1478482 | XRCC4 | √ | × |
| Cholestery | rs1542539 | XRCC4 | √ | × |
| Age-relatec | rs1543847 | XRCC4 | √ | × |
| | rs6897075 | XRCC4 | √ | × |
| | rs1006597 | XRCC4 | √ | × |
| | rs1215344 | XRCC4 | √ | × |
| | rs1215289 | XRCC4 | √ | × |
| | rs6452507 | XRCC4 | √ | × |
| | rs6452508 | XRCC4 | √ | × |
| | rs7717964 | XRCC4 | √ | × |
| | rs1006940 | XRCC4 | √ | × |
| | rs7710630 | XRCC4 | √ | × |
| | rs6237437 | XRCC4 | √ | × |
| | rs7721416 | XRCC4 | √ | × |
| | rs6887846 | XRCC4 | √ | × |
| | rs6452526 | XRCC4 | √ | × |
| | rs6452527 | XRCC4 | √ | × |
| | rs1005584 | XRCC4 | √ | × |
| | rs1007786 | XRCC4 | √ | × |
| | rs9293332 | XRCC4 | √ | × |
| | rs1004201 | XRCC4 | √ | × |
| | rs1003641 | XRCC4 | √ | × |
| | rs1004285 | XRCC4 | √ | × |
| | rs1003787 | XRCC4 | √ | × |
| | rs6237437 | XRCC4 | √ | × |
| | rs1382372 | XRCC4 | √ | × |
| | rs1382371 | XRCC4 | √ | × |
| | rs1382370 | XRCC4 | √ | × |
| | rs1382369 | XRCC4 | √ | × |
| | rs1728428 | XRCC4 | √ | × |
| | rs9986131 | XRCC4 | √ | × |
| | rs1174047 | XRCC4 | √ | × |
| | rs1031902 | XRCC4 | √ | × |
| | rs1047409 | XRCC4 | √ | × |
| | rs1382368 | XRCC4 | √ | × |
| | rs1382365 | XRCC4 | √ | × |
| | rs1382364 | XRCC4 | √ | × |
| | rs1382363 | XRCC4 | √ | × |
| | rs9293334 | XRCC4 | √ | × |
| | rs1174189 | XRCC4 | √ | × |
| | rs1335836 | XRCC4 | √ | × |
| | rs1007600 | XRCC4 | √ | × |
| | rs6452528 | XRCC4 | √ | × |
| | rs7726564 | XRCC4 | √ | × |
| | rs6452531 | XRCC4 | √ | × |
| | rs6890735 | XRCC4 | √ | × |
| | rs1174142 | XRCC4 | √ | × |
| | rs4401555 | XRCC4 | √ | × |
| | rs4616871 | XRCC4 | √ | × |
| | rs6237440 | XRCC4 | √ | × |
| | rs1004036 | XRCC4 | √ | × |
| Platelet cou | rs1005154 | XRCC4 | √ | × |
| Height | rs1005155 | XRCC4 | √ | × |
| | rs1194947 | XRCC4 | √ | × |
| Plateletcrit | rs4541638 | XRCC4 | √ | × |
| | rs2836010 | XRCC4 | √ | × |
| | rs4290995 | XRCC4 | √ | × |
| | rs2974443 | XRCC4 | √ | × |
| | rs2974444 | XRCC4 | √ | × |
| | rs2974445 | XRCC4 | √ | × |
| | rs965674 | XRCC4 | √ | × |
| | rs2940543 | XRCC4 | √ | × |
| | rs2974450 | XRCC4 | √ | × |
| | rs2974449 | XRCC4 | √ | × |
| | rs2662241 | XRCC4 | √ | × |
| | rs7710486 | XRCC4 | √ | × |
| | rs2836028 | XRCC4 | × | √ |
| 5-acetylam | rs7655017 | XRCC4 | × | √ |
| | rs7686221 | XRCC4 | × | √ |
| | rs449837 | XRCC4 | × | √ |
| Cerebrospi | rs7276934 | XRCC4 | × | √ |
| Height | rs1389826 | XRCC4 | × | √ |
| | rs2062227 | XRCC4 | × | √ |
| | rs307316 | XRCC4 | × | √ |
| | rs8031570 | XRCC4 | × | √ |
| | rs307317 | XRCC4 | × | √ |
| Height | rs6707879 | XRCC4 | × | √ |
| | rs7726864 | XRCC4 | × | √ |
| | rs2874647 | XRCC4 | × | √ |
| | rs2874647 | XRCC4 | × | √ |
| | rs2836028 | XRCC4 | × | √ |
| | rs301278 | XRCC4 | × | √ |
| | rs2836028 | XRCC4 | × | √ |
| | rs301279 | XRCC4 | × | √ |
| | rs7776504 | XRCC4 | × | √ |

| | | | | | | | | | | |
|---|---|---|---|---|---|---|---|---|---|---|
| rs1395420 | DIS3L2 | × | √ | × | | rs1485714 | PRKCE | √ | × | × |
| rs1868618 | DIS3L2 | × | √ | × | | rs3731040 | PRKCE | √ | × | × |
| rs1431394 | DIS3L2 | × | √ | × | | rs3727319 | PRKCE | √ | × | × |
| rs1247323 | DIS3L2 | × | √ | × | | rs1154948 | PRKCE | √ | × | × |
| rs5337704 | DIS3L2 | × | √ | × | | rs1169347 | PRKCE | √ | × | × |
| rs1395041 | DIS3L2 | × | √ | × | | rs4953321 | PRKCE | √ | × | × |
| rs6713534 | DIS3L2 | × | √ | × | | rs1020502 | PRKCE | √ | × | × |
| rs3731785 | DIS3L2 | × | √ | × | | rs2278775 | PRKCE | √ | × | × |
| rs1442972 | DIS3L2 | × | √ | × | | rs870139 | PRKCE | √ | × | × |
| rs3811578 | DIS3L2 | × | √ | × | Height | rs870140 | PRKCE | √ | × | × |
| rs209741 | EPHA8 | √ | × | × | | rs2345960 | PRKCE | √ | × | × |
| rs209691 | EPHA8 | √ | × | × | | rs2853446 | PRSS56 | √ | × | × |
| rs209700 | EPHA8 | √ | × | × | | rs1083287 | SLC22A18 | × | × | √ |
| rs1097 | GPR83 | × | √ | × | | rs1871203 | TMEM167 | √ | × | × |
| rs1241781 | GPR83 | × | √ | × | | rs7722924 | TMEM167 | √ | × | × |
| rs7933335 | GPR83 | × | √ | × | | rs6867727 | TMEM167 | √ | × | × |
| rs7924609 | GPR83 | × | √ | × | | rs2306336 | TMEM167 | √ | × | × |
| rs7905892 | GPR83 | × | √ | × | | rs3899556 | TMEM167 | √ | × | × |
| rs1692023 | GPR83 | × | √ | × | | rs6452502 | TMEM167 | √ | × | × |
| rs3740864 | GPR83 | × | √ | × | | rs6452503 | TMEM167 | √ | × | × |
| rs3740865 | GPR83 | × | √ | × | | rs1993948 | TMEM167 | √ | × | × |
| rs3740866 | GPR83 | × | √ | × | | rs2075685 | TMEM167 | √ | × | × |
| rs3740867 | GPR83 | × | √ | × | | rs1478485 | XRCC4 | √ | × | × |
| rs3740868 | GPR83 | × | √ | × | | rs7794966 | XRCC4 | √ | × | × |
| rs1456287 | GPR83 | × | √ | × | | rs1813190 | XRCC4 | √ | × | × |
| rs5556410 | GPR83 | × | √ | × | | rs1564211 | XRCC4 | √ | × | × |
| rs2096762 | GPR83 | × | √ | × | | rs7703318 | XRCC4 | √ | × | × |
| rs2096763 | GPR83 | × | √ | × | | rs2874531 | XRCC4 | √ | × | × |
| rs7555612 | GPR83 | × | √ | × | | rs2617853 | XRCC4 | √ | × | × |
| rs4753604 | GPR83 | × | √ | × | | rs6868817 | XRCC4 | √ | × | × |
| rs1147215 | GPR83 | × | √ | × | | rs1690018 | XRCC4 | √ | × | × |
| rs1172447 | GPR83 | × | √ | × | | rs1051424 | XRCC4 | √ | × | × |
| rs7480817 | GPR83 | × | √ | × | | rs2731866 | XRCC4 | √ | × | × |
| rs8034922 | GPR83 | × | √ | × | | rs301276 | XRCC4 | √ | × | × |
| rs7558247 | GPR83 | × | √ | × | | | | | | |
| rs7126380 | GPR83 | × | √ | × | | | | | | |
| rs1222496 | GPR83 | × | √ | × | | | | | | |
| rs5967891 | GPR83 | × | √ | × | | | | | | |
| rs1692027 | GPR83 | × | √ | × | | | | | | |
| rs6483341 | GPR83 | × | √ | × | | | | | | |
| rs5610880 | GPR83 | × | √ | × | | | | | | |
| rs1083122 | GPR83 | × | √ | × | | | | | | |
| rs7951268 | GPR83 | × | √ | × | | | | | | |
| rs3758785 | GPR83 | × | √ | × | Response to angiotensin II receptor blocker therapy (opposite direction w/ diuretic therapy) | | | | | |
| rs3758786 | GPR83 | × | √ | × | | | | | | |
| rs675478 | GPR83 | × | √ | × | | | | | | |
| rs3758789 | GPR83 | × | √ | × | | | | | | |
| rs7675194 | KIF26B | √ | × | × | | | | | | |
| rs6461317 | LOC10265 | √ | × | × | | | | | | |
| rs1407705 | LOC10265 | √ | × | × | | | | | | |
| rs1177159 | LOC10265 | √ | × | × | | | | | | |
| rs563315 | LOC10265 | √ | × | × | | | | | | |
| rs1267010 | LOC10265 | √ | × | × | | | | | | |
| rs510903 | LOC10265 | √ | × | × | | | | | | |
| rs2155209 | MRE11A | × | √ | × | | | | | | |
| rs622961 | MRE11A | × | √ | × | | | | | | |
| rs1344772 | MRE11A | × | √ | × | | | | | | |
| rs1344772 | MRE11A | × | √ | × | | | | | | |
| rs7881717 | MRE11A | × | √ | × | | | | | | |
| rs584707 | MRE11A | × | √ | × | | | | | | |
| rs598599 | MRE11A | × | √ | × | | | | | | |
| rs518276 | MRE11A | × | √ | × | | | | | | |
| rs1121112 | MRE11A | × | √ | × | | | | | | |
| rs601341 | MRE11A | × | √ | × | | | | | | |
| rs601391 | MRE11A | × | √ | × | | | | | | |
| rs1344771 | MRE11A | × | √ | × | | | | | | |
| rs530569 | MRE11A | × | √ | × | | | | | | |
| rs1411045 | MRE11A | √ | × | × | | | | | | |
| rs4616202 | NPAS3 | × | √ | × | | | | | | |
| rs1878970 | NPAS3 | × | √ | × | | | | | | |
| rs2383519 | NPAS3 | × | √ | × | | | | | | |
| rs3749396 | NPAS3 | × | √ | × | | | | | | |
| rs8008036 | NPAS3 | × | √ | × | | | | | | |
| rs7909425 | NPAS3 | × | √ | × | | | | | | |
| rs1180274 | NPAS3 | × | √ | × | | | | | | |
| rs7772554 | NPAS3 | × | √ | × | | | | | | |
| rs7149836 | NPAS3 | × | √ | × | | | | | | |
| rs7473299 | NPAS3 | × | √ | × | | | | | | |
| rs1447242 | NPAS3 | × | √ | × | | | | | | |
| rs5517710 | NPAS3 | × | √ | × | | | | | | |
| rs2383520 | NPAS3 | × | √ | × | | | | | | |
| rs7140900 | NPAS3 | × | √ | × | | | | | | |
| rs7881961 | NPAS3 | × | √ | × | | | | | | |
| rs1228050 | PIWIL4 | × | × | √ | | | | | | |
| rs6143361 | PIWIL4 | × | × | √ | | | | | | |
| rs2008727 | PRKCE | √ | × | × | | | | | | |
| rs1017104 | TM4SF20 | × | √ | × | | | | | | |
| rs9353296 | TM4SF20 | × | √ | × | | | | | | |
| rs1019483 | TM4SF20 | × | √ | × | | | | | | |
| rs6112916 | TM4SF20 | × | √ | × | | | | | | |
| rs1690016 | XRCC4 | √ | × | × | | | | | | |
| rs2838313 | XRCC4 | √ | × | × | | | | | | |
| rs2838313 | XRCC4 | √ | × | × | | | | | | |
| rs1690016 | XRCC4 | √ | × | × | | | | | | |
| rs4703944 | XRCC4 | √ | × | × | | | | | | |
| rs6884543 | XRCC4 | √ | × | × | | | | | | |
| rs7758428 | XRCC4 | √ | × | × | | | | | | |
| rs2126987 | XRCC4 | √ | × | × | | | | | | |
| rs5766634 | XRCC4 | √ | × | × | | | | | | |
| rs2061783 | XRCC4 | √ | × | × | | | | | | |
| rs7929105 | XRCC4 | √ | × | × | | | | | | |
| rs7754748 | XRCC4 | √ | × | × | | | | | | |
| rs4458554 | XRCC4 | √ | × | × | | | | | | |
| rs925199 | XRCC4 | √ | × | × | | | | | | |
| rs2089566 | XRCC4 | √ | × | × | | | | | | |
| rs7844564 | XRCC4 | √ | × | × | | | | | | |
| rs1251893 | XRCC4 | √ | × | × | | | | | | |
| rs4142947 | XRCC4 | √ | × | × | | | | | | |
| rs6891117 | XRCC4 | √ | × | × | | | | | | |
